# Identification of 16 novel Alzheimer’s disease susceptibility loci using multi-ancestry meta-analyses of clinical Alzheimer’s disease and AD-by-proxy cases from four whole genome sequencing datasets

**DOI:** 10.1101/2024.09.11.24313439

**Authors:** Julian Daniel Sunday Willett, Mohammad Waqas, Younjung Choi, Tiffany Ngai, Kristina Mullin, Rudolph E. Tanzi, Dmitry Prokopenko

## Abstract

Alzheimer’s disease (AD) is the most prevalent form of dementia. While many AD-associated genetic determinants have been previously identified, few studies have analyzed individuals of non-European ancestry. Here, we describe a multi-ancestry genome-wide association study of clinically-diagnosed AD and AD-by-proxy using whole genome sequencing data from NIAGADS, NIMH, UKB, and All of Us (AoU) consisting of 49,149 cases (12,074 clinically-diagnosed and 37,075 AD-by-proxy) and 383,225 controls. Nearly half of NIAGADS and AoU participants are of non-European ancestry. For clinically-diagnosed AD, we identified 14 new loci - five common (*FBN2,/SCL27A6, AC090115.1, DYM, KCNG1/AL121785.1, TIAM1*) and nine rare (*VWA5B1, RNU6-755P/LMX1A, MOB1A, MORC1-AS1, LINC00989, PDE4D, RNU2-49P/CDO1, NEO1,* and *SLC35G3/AC022916.1)*. Meta-analysis of UKB and AoU AD-by-proxy cases yielded two new rare loci (*RPL23/LASP1* and *CEBPA*/AC008738.6) which were also nominally significant in NIAGADS. In summary, we provide evidence for 16 novel AD loci and advocate for more studies using WGS-based GWAS of diverse cohorts.

## Introduction

Alzheimer’s disease (AD) affects 315 million individuals globally - 22% of individuals over age 50^1^, with prevalence dramatically increasing over the past three decades^2^. AD is highly heritable, estimated to be 70% based on twin studies. Genome-wide association studies (GWAS) of clinically diagnosed AD (clinical AD) and AD-by-proxy have identified over 70 genomic loci in predominantly European ancestry individuals^3,4^.

Genetic cohorts of clinical AD used for most GWAS, e.g. the Alzheimer’s Disease Sequencing Project (ADSP) from The National Institute on Aging Genetics of Alzheimer’s Disease Data Storage Site (NIAGADS)^5^ and the family-based AD dataset from the National Institute of Mental Health (NIMH)^6^, have limited sample size. While larger datasets from biobanks, such as UK Biobank contain a limited number of confirmed AD cases, recent studies have increased sample size by using an AD-by-proxy phenotype. AD-by-proxy is based on family history, such as those with a first-degree affected family member, or in some cases, even affected grandparents with AD, as cases^4,7^. AD-by-proxy has been reported to correlate with AD case status in populations of European genetic ancestry^4,8,9^.

A previous GWAS investigating the genetic determinants of clinical AD and AD-by-proxy in more diverse cohorts identified two novel AD loci on chromosome 3^10^. However, in most studies, individuals of non-European ancestry have remained underrepresented^11^. AD has been observed to be more prevalent among individuals who identify as non-Hispanic African Americans versus non-Hispanic Whites^12^. Yet, results obtained in these populations are limited.

Here, we report a comprehensive multi-ancestry WGS single-variant meta-analysis of clinical AD and AD-by-proxy using four cohorts: NIAGADS, NIMH^6^, UK Biobank, and All of Us (AoU) (**Figure 1**). The AoU cohort was initiated in 2018 by the National Institutes of Health to study biomedical and genetic determinants in underrepresented individuals, with presently 315,000 participants, 78% from groups historically underrecruited in biomedical research and roughly half self-reporting to be non-White^13^. Intake collects comprehensive personal, medical, and family history, alongside short-read whole-genome sequencing (WGS) data^14^.

**Figure 1.**
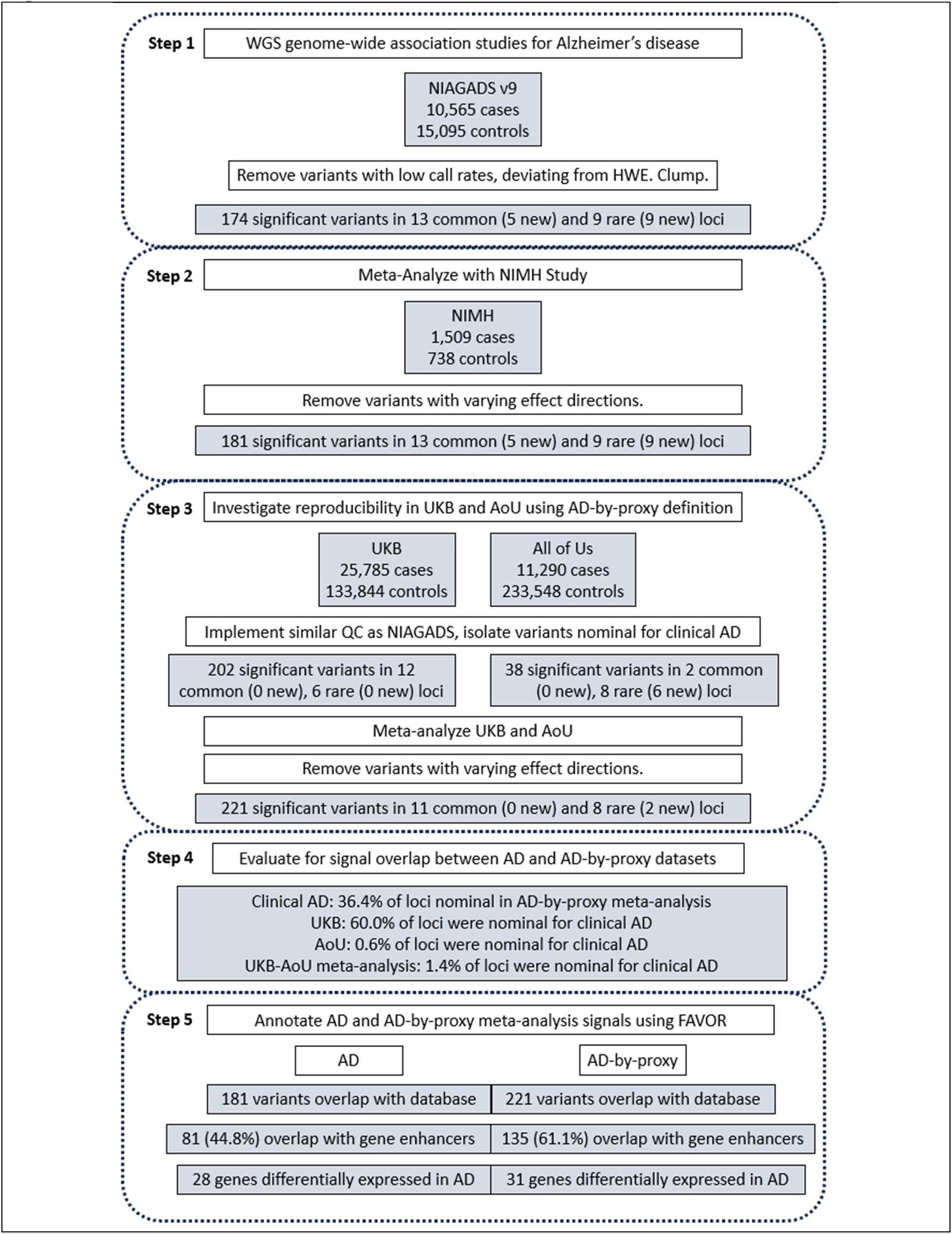
Study design.

Using the most recent release (v9) of NIAGADS we carried out a WGS-based GWAS meta-analysis and identified 5 new common and 9 new rare loci for clinical AD. We also performed a WGS-based GWAS meta-analysis of AD-by-proxy cases in UKB and AoU. We limited the results to genome-wide significant variants that were also nominally significant in NIAGADS. This yielded two new rare loci. Interestingly, there was little overlap in loci between the clinically diagnosed AD and AD-by-proxy meta-analyses. While these results suggest limited generalizability of AD-by-proxy results from diverse cohorts, versus predominantly European ancestry cohorts with clinical AD^4^, they provide complementary evidence for 16 novel AD-associated loci and advocate for using WGS-based GWAS of diverse cohorts to discover novel AD loci.

## Results

### Sample and variant-level quality control

For NIAGADS, we started with 336,500,060 split variants in 34,438 samples. Following quality control, 61,864,192 variants and 25,660 samples remained, including 10,565 AD cases. For the family-based cohorts (referred hereafter as NIMH), we combined two WGS familial cohorts with 1393 (NIMH; AD: n = 966) and 854 (NIAGADS families; AD: n = 543) individuals, as described previously^6^. These datasets included 15,905,393 variants observed across 2,247 samples, with 1,509 AD cases. For UKB, we started with 88,331,742 multiallelic variants, 584,065,627 biallelic variants, and 200,004 samples. Following QC and splitting multi-allelics, 262,394,351 variants and 159,629 subjects remained, with 25,785 AD-by-proxy cases, defined by having AD or an affected parent. For AoU, we started with 1,346,414,851 split variants and 245,394 samples. Following QC, 109,317,793 variants and 244,838 samples remained with 11,290 AD-by-proxy cases, defined by having AD or an affected first-degree relative or grandparent (**Supplemental Table 1**).

Referring to genetic ancestry, NIAGADS v9 was similarly diverse to AoU, with a similar proportion of non-European ancestry individuals. UKB predominantly included individuals of European ancestry. While cases in all cohorts were predominantly European ancestry, a meaningful proportion of cases with diverse genetic ancestry were included in NIAGADS and AoU (**Supplemental Table 1**). The age distributions of cases and controls were similar across UKB and AoU, with the ages of NIAGADS participants being generally greater than UKB and AoU (**Figure 2A**). The broader age distribution in AoU was anticipated given its focus on recruiting underrepresented individuals, that also included younger participants. The lower case:control ratio in AoU, compared to UKB, could be due to the inclusion of a greater number of younger participants (with younger relatives) and AD being under-detected and under-reported in non-whites^15^, with almost half of AoU participants not being European ancestry (**Supplemental Table 1**). While NIAGADS was also relatively diverse, it focused on recruiting individuals with clinical AD. AoU cases included more females than did the controls; this ratio was similar in the other cohorts (**Figure 2B**). The results for all variants that were genome-wide significant across these cohorts are available in the Supplementary Materials (**Supplementary Table 2**).

**Figure 2.**
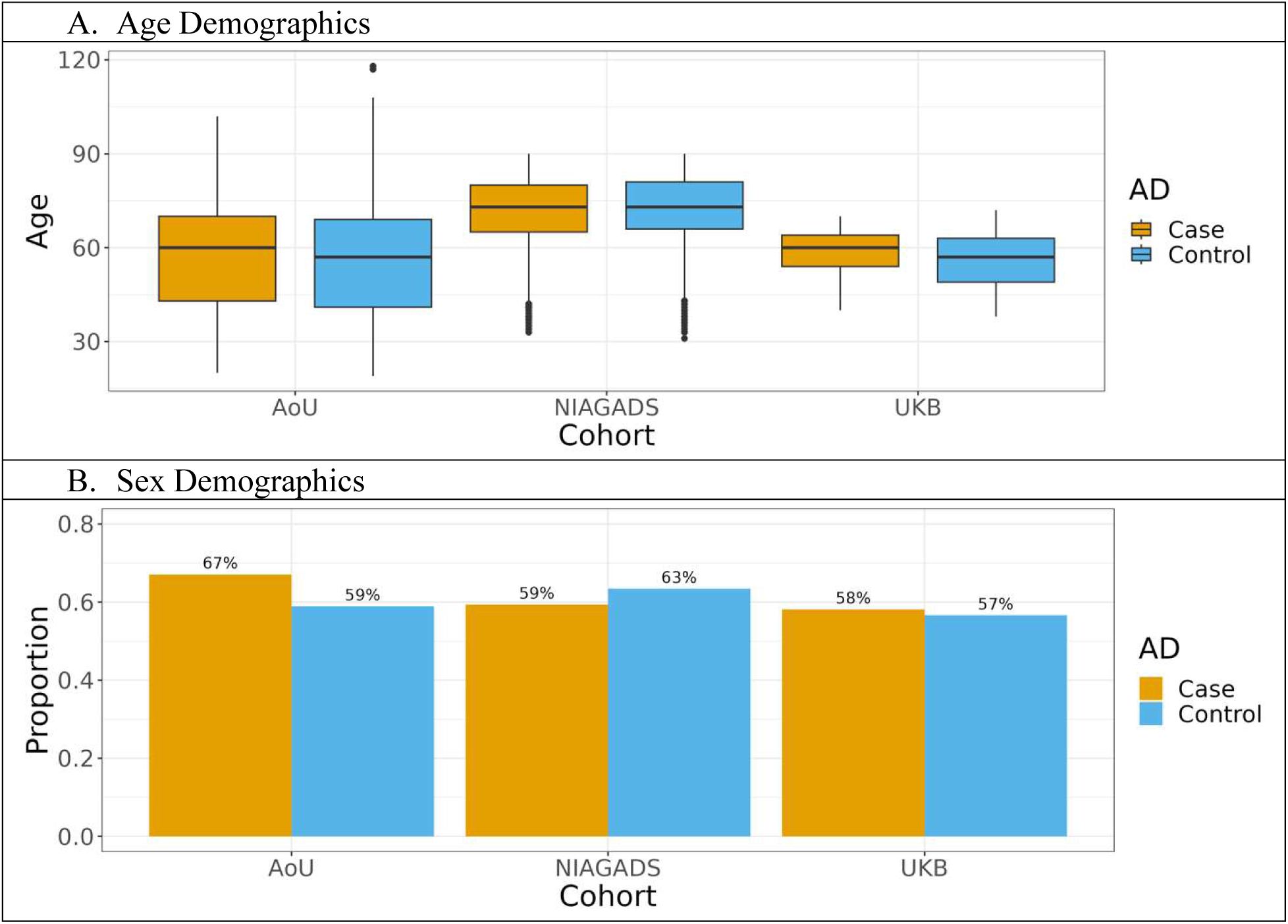
Age (A) and sex (B) demographics of individuals representing AD cases compared to controls in each cohort. NIAGADS used clinical AD as a case definition, with UKB and AoU using AD-by-proxy as case definitions.

### Clinical AD GWAS and meta-analysis

We initially conducted a WGS-based GWAS on clinical AD in NIAGADS v9 using Plink2^16^. We identified 22 genome-wide significant loci (p ≤ 5 ∗ 10^-8^), of which, 14 were novel (5 common and 9 rare) (**Figure 3**, **Table 1**). Following meta-analysis of NIAGADS with NIMH, we observed minor changes in p-values and identified 7 new genome-wide significant variants in known GWAS AD loci. (**Table 1**, **Figure 1**). The genomic inflation factors of NIAGADS and the meta-analysis were 1.00 and 1.01, with λ_1000_ 1.00 and 1.00 (**Supplementary Figure 1**). Data for all variants in these loci is available in the Supplementary Materials (**Supplementary Table 3**).

**Figure 3.**
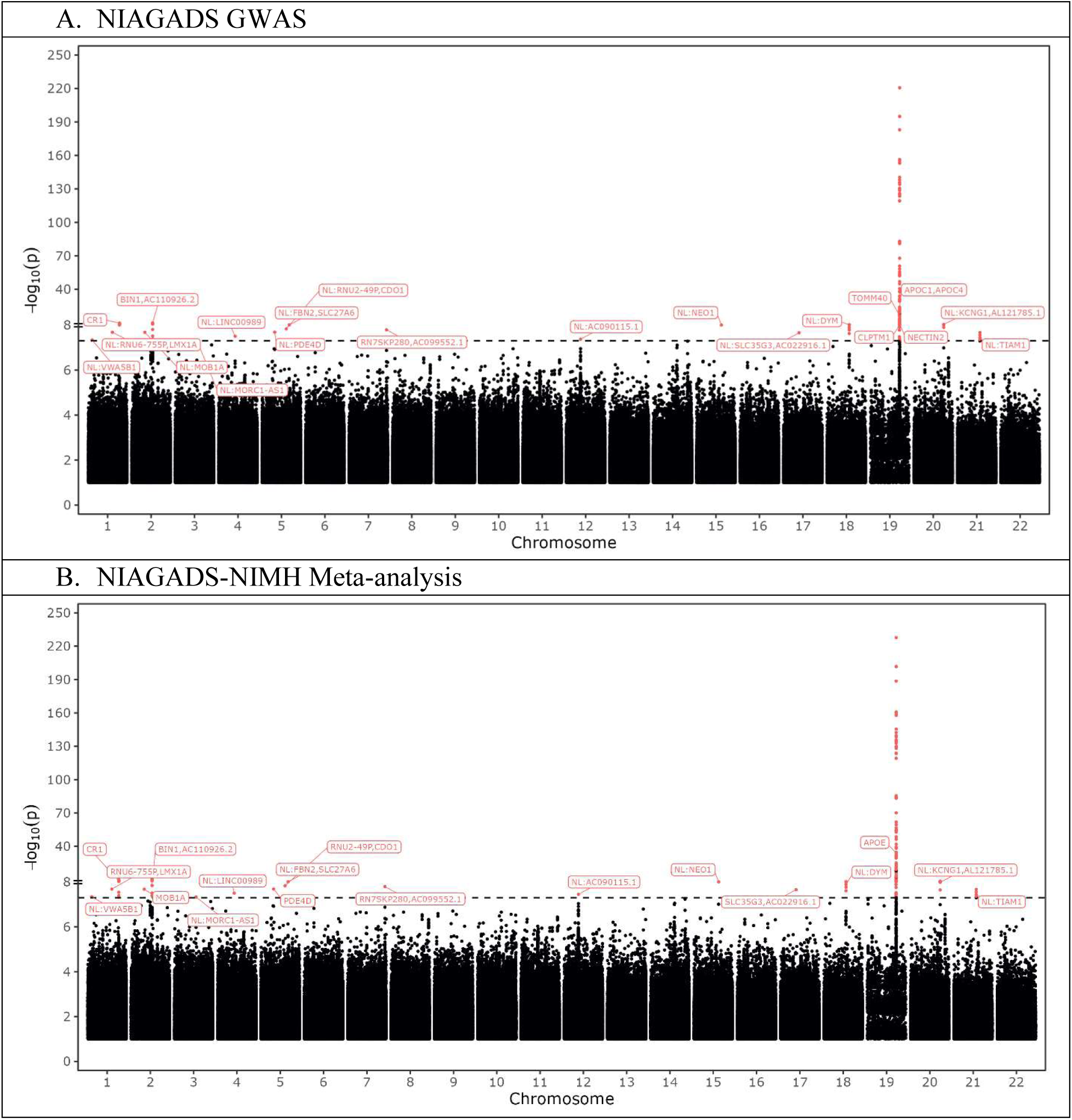
Manhattan plots of AD from NIAGADS alone and after meta-analyzing with NIMH. Red genome-wide significant variants represent variants that passed all quality-control testing. NL signifies a new locus.

**Table 1.** Lead variants for AD in NIAGADS and the NIAGADS-NIMH meta-analysis. Each character in the direction column corresponds to the effect direction in NIAGADS, NIMH, UKB, and AoU, respectively. Known (old) locus refers to a variant proximal to a locus previously linked to AD. For variants proximal to multiple genes, the two closest genes are shown. ND stands for not detected. Variants are ordered by chromosome and position. We focused on variants with an MAF ≥ 0.1% to emphasize higher-confidence results.

| CHR | Rsid | Gene | MAF | Effect allele | Direction | Cohorts with clinically diagnosed AD |  |  | Meta analysis for AD-by-proxy cohorts |  | New/Old |
| --- | --- | --- | --- | --- | --- | --- | --- | --- | --- | --- | --- |
|  |  |  |  |  |  | NIA P | NIMH P | Meta analysis P | AD-by-proxy Z | AD-by-proxy P |  |
| 1 | rs141744862 | VWA5B1 | 0.0031 | T | +??- | 4.6E-08 | ND | 4.6E-08 | -1.347 | 0.18 | New |
| 1 | rs573181360 | RNU6-755P, LMX1A | 0.0005 | G | +??- | 2.09E-08 | ND | 2.09E-08 | -0.018 | 0.99 | New |
| 1 | rs6661489 | CR1 | 0.1425 | T | +?++ | 1.47E-10 | ND | 1.47E-10 | 2.269 | 0.023 | Old |
| 2 | rs150214656 | MOB1A | 0.0003 | G | +?+- | 2.1E-08 | ND | 2.1E-08 | 0.54 | 0.59 | New |
| 2 | rs6733839 | BIN1, AC110926.2 | 0.4168 | T | ++++ | 1.19E-10 | 0.039 | 4.67E-11 | 6.841 | 6.14E-12 | Old |
| 3 | rs557495347 | MORC1-AS1 | 0.0008 | T | +?+- | 4.71E-08 | ND | 4.71E-08 | 0.059 | 0.95 | New |
| 4 | rs117010230 | LINC00989 | 0.0009 | A | +?+- | 3.14E-08 | ND | 3.14E-08 | -0.476 | 0.63 | New |
| 5 | rs182525847 | PDE4D | 0.0005 | G | +?++ | 2.07E-08 | ND | 2.07E-08 | 1.779 | 0.075 | New |
| 5 | rs56918975 | RNU2-49P, CDO1 | 0.0005 | C | +?++ | 1.48E-08 | ND | 1.48E-08 | 1.107 | 0.27 | New |
| 5 | rs147450666 | FBN2, SLC27A6 | 0.0104 | A | ---+ | 7.67E-09 | 0.53 | 7.17E-09 | -0.154 | 0.88 | New |
| 7 | rs186724723 | RN7SKP280, AC099552.1 | 0.0158 | C | -?-- | 1.62E-08 | ND | 1.62E-08 | -0.483 | 0.63 | Old |
| 12 | rs56098445 | AC090115.1 | 0.094 | A | +++- | 4.25E-08 | 0.45 | 3.57E-08 | -0.407 | 0.69 | New |
| 15 | rs541189631 | NEO1 | 0.0008 | T | +?+- | 8.84E-09 | ND | 8.84E-09 | 0.431 | 0.67 | New |
| 17 | rs553129131 | SLC35G3, AC022916.1 | 0.0006 | G | +?++ | 2.21E-08 | ND | 2.21E-08 | 1.125 | 0.26 | New |
| 18 | rs200388554 | DYM | 0.0258 | G | -?++ | 5.99E-09 | ND | 5.99E-09 | 1.93 | 0.054 | New |
| 19 | rs8105340 | NECTIN2 | 0.1069 | C | ++++ | 7.74E-10 | 0.66 | 7.02E-10 | -1.75 | 0.079 | Old |
| 19 | rs157588 | TOMM40 | 0.4373 | C | ++++ | 3.14E-17 | 9.29E-3 | 6.81E-18 | -7.37 | 1.5E-13 | Old |
| 19 | rs429358 | APOE | 0.2104 | C | ++++ | 1.9E-310 | 2.22E-16 | 7.2E-305 | 41.238 | 3.3E-305 | Old |
| 19 | rs141622900 | APOC1, APOC4 | 0.0429 | A | ---- | 1.72E-24 | 2.76E-4 | 5.36E-25 | -9.634 | 4.79E-22 | Old |
| 19 | rs116949436 | CLPTM1 | 0.0115 | A | ++++ | 6.78E-09 | 0.015 | 4.63E-09 | 4.38 | 1.18E-05 | Old |
| 20 | rs4809823 | KCNG1, AL121785.1 | 0.2908 | G | --?- | 2.75E-09 | 0.46 | 2.23E-09 | -0.746 | 0.45 | New |
| 21 | rs77589046 | TIAM1 | 0.0326 | C | -??+ | 2.16E-08 | ND | 2.16E-08 | 1.082 | 0.28 | New |

We replicated association of AD with several genes in the *APOE* locus including *NECTIN2*, *TOMM40*, *APOE*, and *APOC1* (**Table 1**). The strongest genome-wide significant variants defining the five new common AD loci were driven by NIAGADS and all were protective against AD, except for rs56098445 on chromosome 12 proximal to lncRNA *AC090115.1* and *ZNF641* (**Table 1**). rs147450666 on chromosome 5 is proximal to *FBN2* and *SLC27A6*^17^. rs200388554 on chromosome 18 is proximal to *DYM*^17^. rs4809823 on chromosome 20 is proximal to *KCNG1* and the lncRNA *AL121785.1*^17^. rs77589046 on chromosome 21 is proximal to *TIAM1*^17^. We also identified nine new rare variant loci showing genome-wide significant association with clinical AD within or proximal to the following genes: *VWA5B1, RNU6-755P/LMX1A, MOB1A, MORC1-AS1, LINC00989, PDE4D, RNU2-49P/CDO1, NEO1,* and *SLC35G3/AC022916.1* (**Table 1**).

### AD-by-proxy GWAS and meta-analysis

We next performed a WGS-based GWAS on AD-by-proxy in UKB and AoU using Regenie^18^. First, we removed variants deviating from Hardy-Weinberg equilibrium in single genetic ancestry populations given that the statistic is sensitive to population stratification, we meta-analyzed the results with inverse standard error weighting per fixed effects using METAL^19^, including a total of 37,075 AD-by-proxy cases and 367,392 controls. The λ_GC_ and λ_GC,1000_ values were 1.25 and 1.01 for UKB, 1.12 and 1.01 for AoU, and 1.22 and 1.00 for the UKB-AoU meta-analysis, respectively (**Supplementary Figure 1**). We were able to replicate six lead variants (p ≤ 0.05; same direction) of 22 loci identified in our clinical AD meta-analysis, in either our AD-by-proxy meta-analysis or one of the AD-by-proxy datasets (**Table 1, Supplemental Table 2**). Variants that were genome-wide significant in our AD-by-proxy studies and not nominally significant in our clinical-AD analyses are available in the Supplementary materials (**Supplementary Table 4**).

We identified 30 genome-wide significant loci - 21 common and nine rare – in the UKB dataset (**Supplementary Table 5**). We identified 1,558 genome-wide significant loci - 112 common and 1,446 rare - in the AoU dataset (**Supplementary Table 6**). When we removed indels and multiallelic sites from these AoU signals, we were left with 640 genome-wide significant loci – 51 common and 589 rare. Of these, 6 common and 109 rare loci contained more than one genome-significant variant. 18 genome-wide significant loci - 12 common and 6 rare – in UKB, and 10 genome-wide significant loci - two common and eight rare – in AoU showed nominally significant replication (same direction, p ≤ 0.05) in clinical AD in NIAGADS. None of the 18 genome-wide significant loci in UKB were novel, with 15/18 located in the vicinity of ApoE (**Supplementary Figure 2**). Of the AoU genome-wide significant loci with an effect direction matching the clinical AD study, 2/10 were common (all previously known), with 2/8 of the rare loci being novel (**Supplementary Figure 2**). While there were in total 5 common and 27 rare independent loci in the AD-by-proxy AoU data that were also nominally significant in the clinical AD meta-analysis, many of these signals had an opposite direction of effect than clinical AD. Several of these loci were nominally significant or genome-wide significant in individuals of Admixed American, African, or European genetic ancestry (**Supplemental Figure 3**).

8/22 (36.4%) of genome-wide significant loci in the clinical AD meta-analysis had at least one variant that was nominally significant in the AD-by-proxy meta-analysis. However, only 10/1558 (0.6%) of the genome-wide significant loci in the similarly diverse AoU cohort had at least one variant that was nominally significant in the clinical AD meta-analysis. This number was 9/640 when we filtered for indels and multiallelic variants. In UKB, 18/30 of the genome-wide significant loci had at least one variant that was nominally significant in the clinical AD meta-analysis. The greater degree of overlap between the clinical AD meta-analysis and UKB (60.0%) versus 0.6% in AoU was most likely due to UKB being predominantly composed of individuals of European ancestry (**Figure 1**).

We replicated several previously known common loci in our AD-by-proxy meta-analysis, including genes in the *APOE* AD locus (**Table 2**). We also identified 19 genome-wide significant loci (within or near 18 genes), in which the variants were also replicated with nominal significance in clinical AD in NIAGADS (**Table 2**). Some loci represented the same nearest genes due to multiple rare and common genome-wide independent signals within those genes. Of 19 loci, three were only nominally significant or detected in AoU and not UKB. There were 11/19 loci that were nominally significant in both datasets, but none of these were novel AD loci. Thus, out of 19 loci in our AD-by-proxy meta-analysis that were also nominally significant in the clinical AD meta-analysis, we identified 2 novel genome-wide significant AD loci – both involving rare variants (**Figure 4B**, **Table 2**). These loci were proximal to or within *RPL23/LASP1*, *AC008738.6* (**Table 2**).

**Figure 4.**
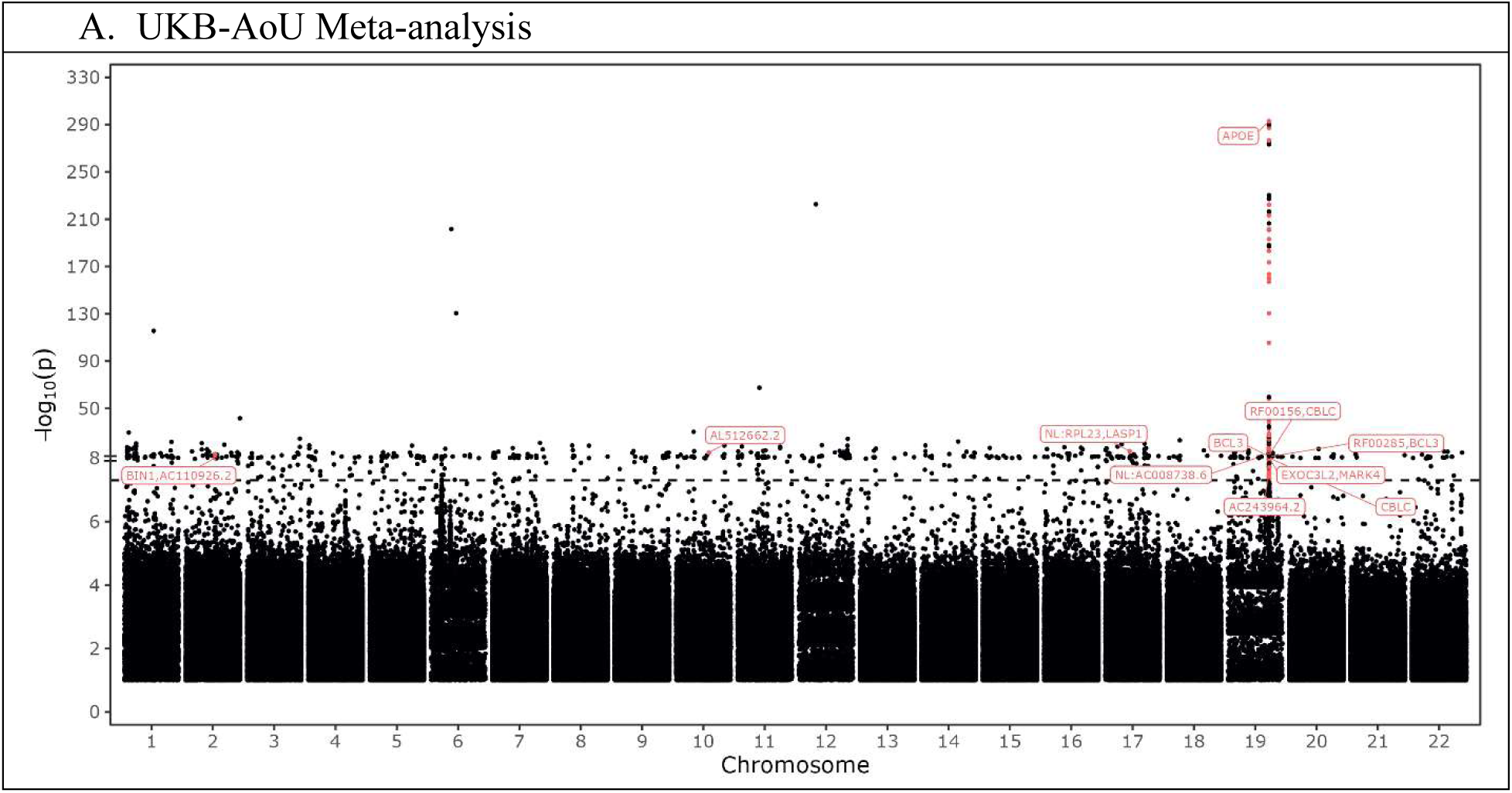
Manhattan plot of AD-by-proxy meta-analysis, highlighting variants that were nominally significant (p ≤ 0.05) in the NIAGADS-NIMH meta-analysis. Given the larger number of independent loci proximal to ApoE, only one locus in this region was shown. NL signifies a new locus.

**Table 2.**
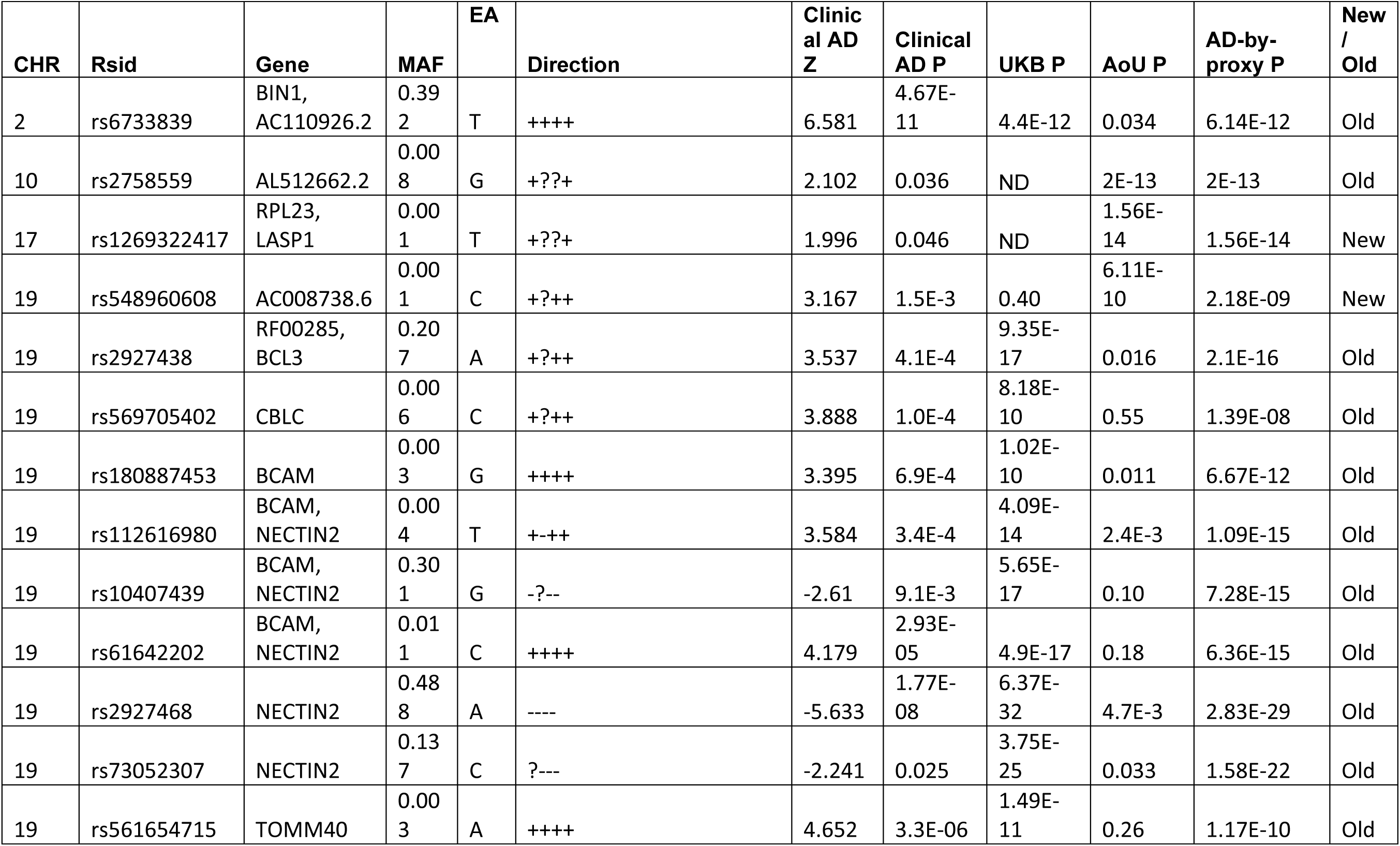
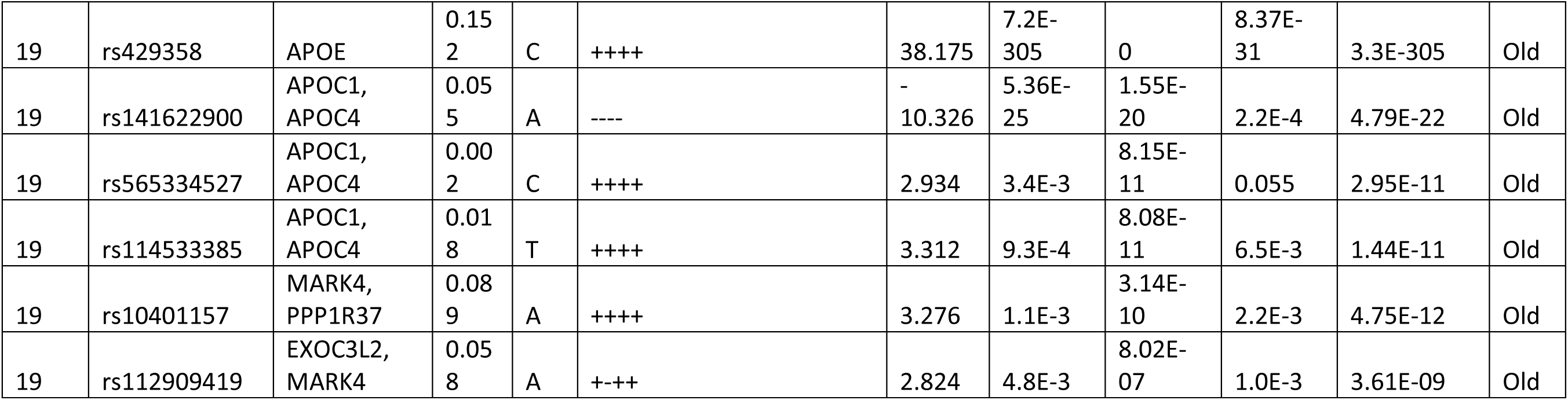
Lead genome-wide significant variants for AD-by-proxy in the meta-analysis between UKB and AoU, highlighting variants that were nominally (p ≤ 0.05) significant in our clinically AD meta-analysis and with an allele frequency of at least 0.1%. Known locus refers to a variant proximal to a locus previously linked to AD. ND stands for not detected. Each character in the direction column corresponds to the effect direction in NIAGADS, NIMH, UKB, and AoU, respectively.

### Replication of previously reported genomic associations and influence of HWE and population diversity

Next, we set out to test for replication of known AD loci from a recent large meta-analysis reported in Bellenguez et al.^4^, which focused on European genetic ancestry participants, in the AoU dataset. Of the 83 AD-associated loci listed in Bellenguez et al.’s Supplementary Table 5^4^, 65 of the variants were detected in UKB and all 83 in AoU before filtering for deviations from HWE (**Supplementary Table 7**). Our AoU study was determined to have sufficient power (Power ≥ 0.8) to detect 7 of the 83 lead variants from Bellenguez et al. Supplementary Table 5^4^, with variants of less power being rare or having a smaller effect size (**Supplemental Table 5**). We did not compute the power of our clinical AD study or the UKB-AoU meta-analysis because the subject data overlapped with the Bellenguez et al. study^4^.

To control for population stratification or genotyping errors^20^, traditional GWAS uniformly removes variants that deviate from HWE^21,22^ (p ≤ 1e-15 in a prior AoU study^23^. Population stratification, inherent in mixed ancestry populations, yields more significant HWE p-values that can lead to the loss of many variants. With a uniform HWE filter only 21/83 in UKB and 22/83 in AoU are retained as passing. When we used single-ancestry HWE p-values versus those derived from the multi-ancestry population, 59/83 and 73/83 of the Bellenguez et al. variants were retained in the UKB and AoU datasets, respectively. Since the Bellenguez et al. dataset was predominantly of European genetic ancestry^4^, we next applied European ancestry HWE statistics from AoU to filter results, in which case, 64/83 and 82/83 variants were retained in the UKB and AoU datasets, respectively.

We were able to replicate 9/83 signals in AoU (p ≤ 0.05, matching direction of effect), with calculated power to detect seven (**Supplementary Table 7**). This number increased to 40/83 when applied to the AD-by-proxy meta-analysis, however UKB was one of the datasets used in Bellenguez et al^4^. Here, we have not reported results from the clinical AD analysis because the NIAGADS European participants and UKB were previously evaluated in Bellenguez et al^4^. Our results corroborate the findings of Bellenguez et al., which is unsurprising given the majority of AD-by-proxy cases in AoU are of European ancestry; but they also demonstrate the difference in results when studying more diverse populations and evaluating different phenotype definitions^4^.

### Functional annotation of identified loci and differential expression of variants’ proximal genes by AD outcomes

We next used a functional annotation of variants based on the online resource (FAVOR). FAVOR provides annotations for specific SNPs and indels across the human genome for whole genome sequencing data^17^. To determine whether the novel AD-associated genomic variants identified here represent biologically plausible candidates, we used FAVOR to identify the genes proximal to the genetic variants, along with their enhancer-relevant annotations, using the Genehancer and SuperEnhancer databases. These databases predict whether variants overlap or are predicted to influence gene expression through enhancers. We also assessed differential expression of the cognate genes associated with these variants using single-cell differential gene expression statistics for individuals with and without cognitive impairment and pathological evidence of AD, available from Mathys et al. 2023^24^.

For the clinical AD meta-analysis, 181/181 genome-wide significant signals overlapped with the FAVOR database and 81 had enhancer-relevant annotations and were proximal to 28 genes differentially expressed with AD phenotypes (FDR ≤ 0.01). While none of those 81 enhancer-linked variants were in novel loci, gene expression and function can be modulated by elements beyond enhancers. Differentially expressed genes proximal to our genome-wide significant AD-associated variants included most genes in the novel loci implicated. *FBN2* was upregulated in inhibitory neurons in individuals with cognitive impairment and pathological evidence of AD. *SLC27A6* was upregulated in excitatory and inhibitory neurons. *AC090115.1* was downregulated in excitatory and inhibitory neurons. *DYM* was upregulated in excitatory and inhibitory neurons. *KCNG1* was downregulated in excitatory neurons and upregulated in astrocytes. Finally, *TIAM1* was upregulated in excitatory neurons and downregulated in inhibitory neurons and oligodendrocyte precursor cells (**Table 3**)^24^.

**Table 3.**
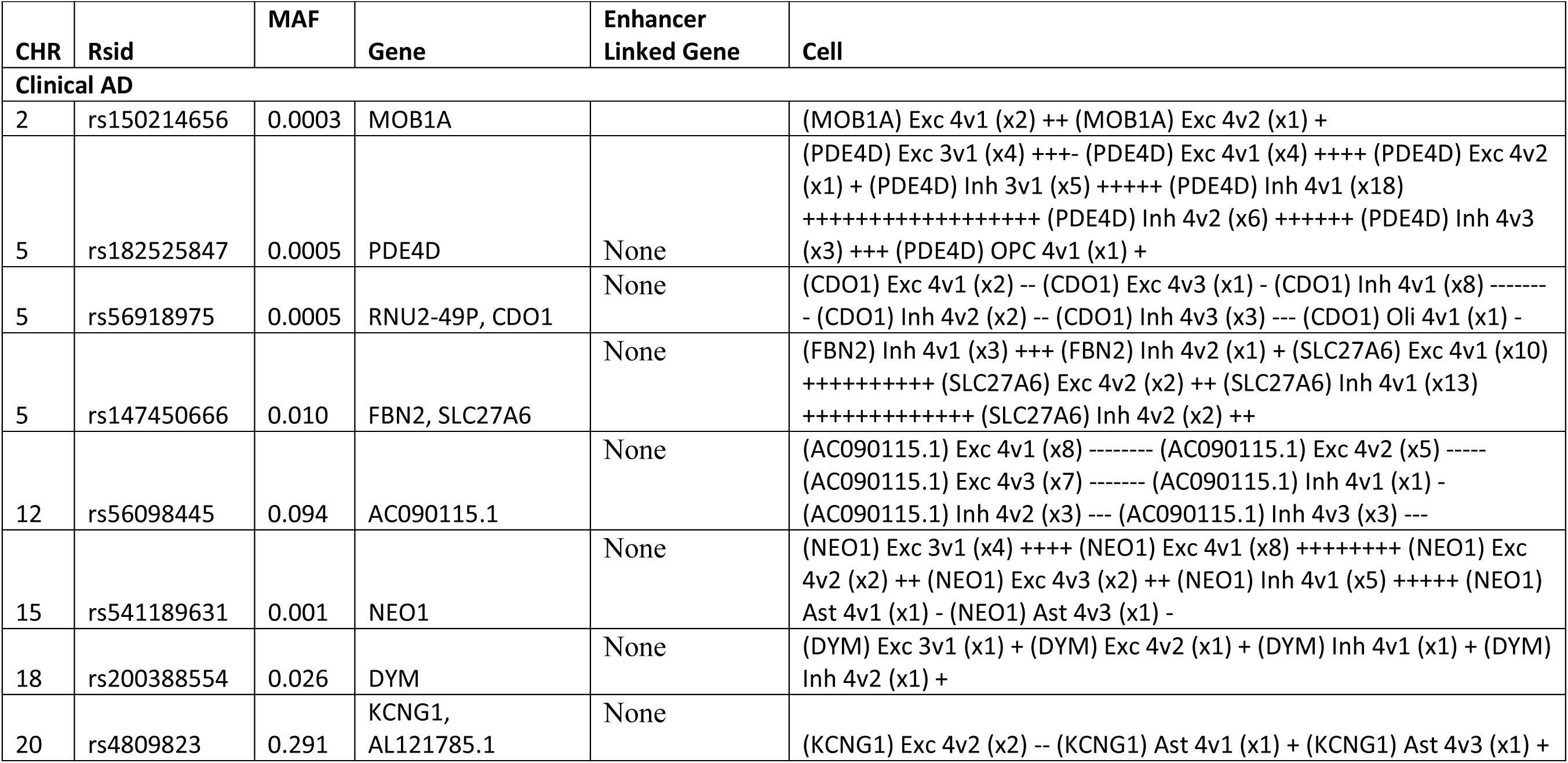
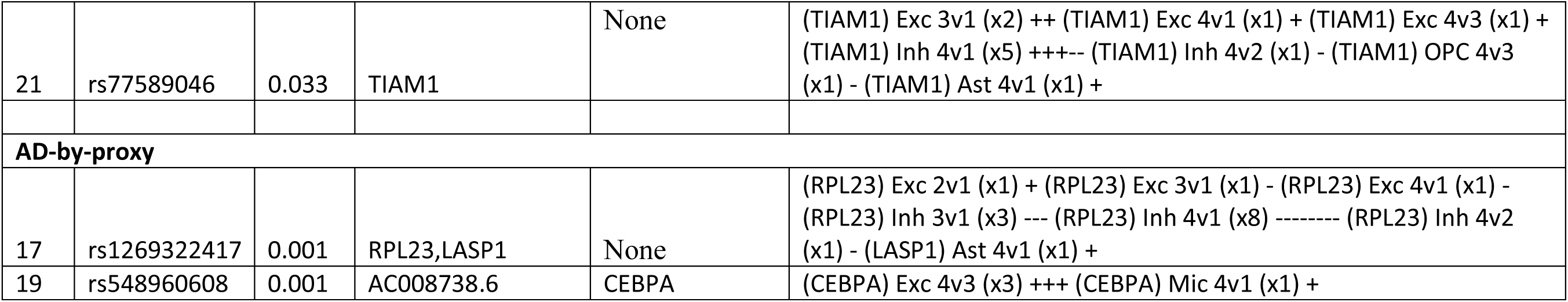
Variants in AD and AD-by-proxy novel loci proximal to or overlapping enhancers of genes whose expression significantly (FDR ≤ 0.01) changed in brain cells, comparing groups separated by pathological evidence of or symptoms of AD. When multiple variants in a locus were observed, the variant with the most significant p-value was highlighted. Enhancer annotations (Enhancer Linked Gene column) were obtained from SuperEnhancer or GeneHancer databases, logged in FAVOR. For cells, Exc represents excitatory neurons, Inh inhibitory neurons, Oli oligodendrocytes, Ast astrocytes. Group comparisons (n v m) include group 1 (no AD pathological evidence or cognitive impairment), group 2 (no pathological evidence with cognitive impairment), group 3 (pathological evidence without cognitive impairment), group 4 (pathological evidence and cognitive impairment). Scores (xN) represent the number of unique cell subpopulations enriched for the cognate gene. Plus and minus signs correspond to whether the log-fold change was positive or negative for the comparison.

For the AD-by-proxy meta-analysis, 221 of 221 genome-wide significant signals overlapped with FAVOR and 135 had enhancer-relevant annotations and were proximal to 31 genes differentially expressed with AD phenotypes. The 221 variants included 2 at novel rare AD loci where the effect direction matched AD-by-proxy, including *RPL23/LASP1* (rs1269322417 on chromosome 17) with the genes being differentially expressed in excitatory and inhibitory neurons (**Table 3**). rs548960608 on chromosome 19 is linked by enhancer to *CEBPA* that is upregulated in excitatory neurons (**Table 3**).

We next asked whether any of these genes are differentially expressed with varying severity of cognitive impairment in the setting of AD pathological features, or in individuals considered resilient to AD versus those with mild or severe AD. Mathys et al. 2023 also studied the role of global AD pathology, amyloid, neurofibrillary tangles (NFTs), and neuritic plaques quantitative burden on cognitive impairment severity^24^. By pathology, genes in the novel loci were only differentially expressed when considering burden of neurofibrillary tangles and neuritic plaques (**Table 4**). Regarding clinical AD, we observed significantly increased (FDR < 1%) expression of *FBN2* and *SCL27A6* in previously resilient subjects that later suffered with mild cognitive impairment in the presence of NFTs^24^. Expression of these genes was further increased when comparing individuals with severe versus mild cognitive impairment in the presence of neuritic plaques (**Table 4**). Other genes proximal to or within our novel loci demonstrated differential expression with AD phenotypes (**Table 4**). Our results support the association of AD with these genes, and their expression profiles.

**Table 4.**
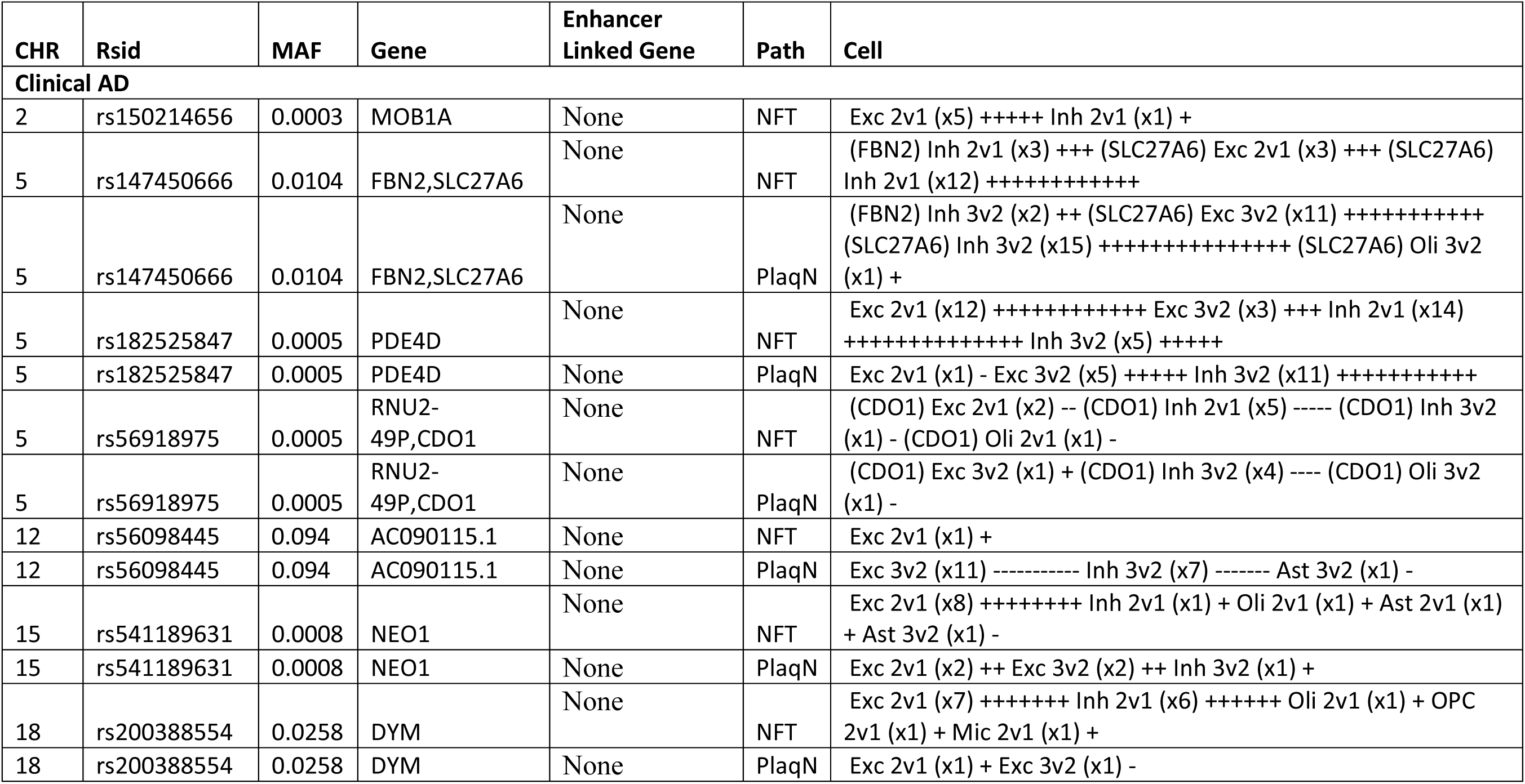
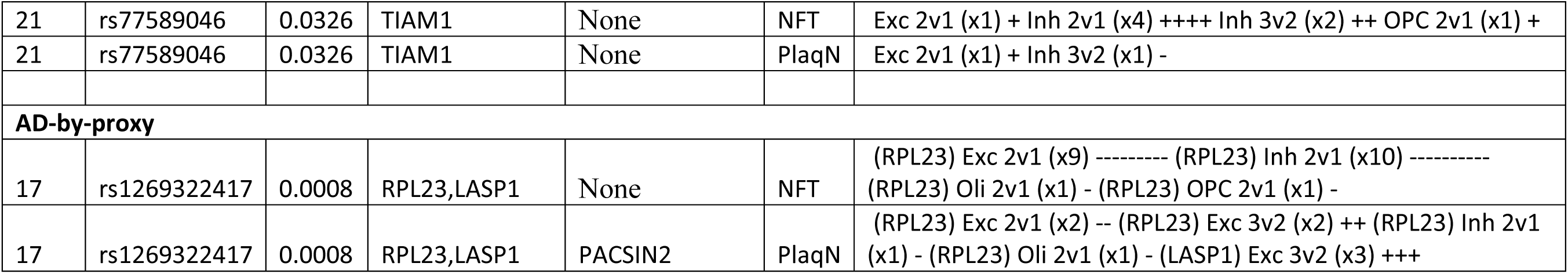
Variants in AD-by-proxy novel loci proximal to or overlapping enhancers of genes with expression that significantly (FDR ≤ 0.01) changed in brain cells for with cognitive impairment, in the setting of pathology (Path). Enhancer annotations (in Enhancer Linked Gene column) were obtained from SuperEnhancer or GeneHancer databases, logged in FAVOR. For progression-relevant outcomes, NFT represents neurofibrillary tangle burden, PlaqD diffuse plaque burden, PlaqN neuritic plaque burden, tangles tangle density. For cells, Exc represents excitatory neurons, Inh inhibitory neurons, Oli oligodendrocytes, OPC oligodendrocyte precursor cell, 2v1 represents comparison of gene expression between individuals with mild cognitive impairment to those with no cognitive impairment, 3v2 comparing individuals with AD dementia and mild cognitive impairment. Scores represent the number of unique cell subpopulations enriched for the cognate gene.

## Discussion

Here, we report 16 novel loci associated with clinical AD or AD-by-proxy emanating from a GWAS of more than 430,000 WGS samples from four cohorts including subjects with diverse genetic ancestry. While we observed that genetic associations using clinical AD datasets were reasonably reproduced in AD-by-proxy datasets, few genetic associations derived from AD-by-proxy studies were reproduced in clinical AD datasets consisting of more diverse cohorts with multi-ancestral genetics. Among AD-by-proxy signals those that were nominally significant in the clinical AD study, particularly in the diverse AoU dataset, several had effect directions that were the opposite as in clinical AD. Many of the novel AD-associated genomic variants were within or proximal to genes that differentially expressed in single-cell brain populations, particularly excitatory and inhibitory neurons, in individuals with cognitive impairment and pathological evidence of disease. Overall, these findings underscore the importance of implementing WGS samples in GWAS to determine the genetic underpinnings of disease, in diverse cohorts. This is valuable for not only validating previous results, but also for yielding new genetic findings that could benefit diverse populations.

We identified five novel common loci in our clinical AD analysis, with plausible roles in mediating disease (**Table 1**) based on differential expression in subjects with and without cognitive impairment and AD neuropathological hallmarks (**Table 3**). Four of the five new common loci contained variants that were protective against AD, with the exception being rs56098445 on chromosome 12 proximal to lncRNA *AC090115.1* and *ZNF641*. rs147450666 is proximal to *FBN2* and *SLC27A6*^17^. *FBN2* is expressed in the choroid plexus with roles in connective tissue structure^25^ with a different gene variant linked by GWAS to vascular dementia^26^. *SCL27A6* is not documented in the Human Protein Atlas and protein dysfunction has been linked to neurodegeneration in C. elegans^27^. rs200388554 is proximal to *DYM*^17^, which is enriched in oligodendrocytes and excitatory and inhibitory neurons, believed to play a role in early brain development and protein secretory pathways^25^; DYM protein levels were previously observed to significantly vary in AD patient plasma versus matched controls^28^. rs4809823 is proximal to *KCNG1* and the lncRNA *AL121785.1*^17^. *KCNG1* contributes to neural synaptic function through voltage-gated potassium channels^25^, and is predicted to be a hub gene for AD-relevant immune pathways^29^. rs77589046 is proximal to *TIAM1*^17^, which plays roles in DNA binding in brain tissues with enriched expression in the cerebellum and inhibitory neurons, oligodendrocytes, and oligodendrocyte precursor cells, with additional protein function as a guanyl nucleotide exchange factor^25^. In addition, we identified nine new rare variant loci showing genome-wide significant association to clinical AD, including *VWA5B1, RNU6-755P/LMX1A, MOB1A, MORC1-AS1, LINC00989, PDE4D, RNU2-49P/CDO1, NEO1,* and *SLC35G3/AC022916.1*.

Based on AD-by-proxy analyses of UKB and AoU datasets, we identified 2 novel rare genome-wide significant AD loci that were nominally significant in our clinical AD and AD-by-proxy analyses (**Figure 4**, **Table 2**). These loci are proximal to or linked by enhancers to genes differentially expressed in AD, specifically *RPL23/LASP1* and AC008738.6/*CEBPA* (**Table 3**). *RPL23* encodes ribosomal protein L23, which is enriched among genes expressed in the brain linked with ribosome function with Shigemizu et al. reported that blood RNA levels of *RPL23* were significantly decreased in AD patients^25,30,31^. *LASP1* encodes LIM And SH3 Protein 1, expression, of which is enriched among genes expressed in the brain in sub-cortical regions to mediate actin-based cytoskeletal activities. LIM And SH3 Protein 1, is a component of synapses and dendritic spines in the CNS and a polymorphism in *LASP1* has been reported to affect cognitive function in schizophrenia^32^. *CEBPA* encodes CCAAT enhancer binding protein alpha, which is enriched among genes expressed in the brain involved with macrophage and microglial immune response. This gene regulates proliferation arrest and the differentiation of myeloid progenitors as well as other cell types, functioning as a DNA-binding activator protein^25^. Interestingly, enrichment of AD heritability at variants within CEBPA has been previously reported^33^.

We also assessed the novel AD-associated genomic variants identified here for proximity to or overlapping enhancers of genes, for which expression is significantly changed in brain cells, when comparing groups separated by pathological evidence of or symptoms of AD, or cognitive impairment, in the setting of pathology (**Tables 3-4**). Differentially expressed genes proximal to genome-wide significant variants associated with clinical AD include *FBN2* (inhibitory neurons), *SLC27A6*, *AC090115.1, DYM* (excitatory and inhibitory neurons), and *KCNG1* (excitatory neurons and astrocytes), and *TIAM1* (excitatory neurons, inhibitory neurons and oligodendrocyte precursor cells). (**Table 3**). These data further support the role of these gene in AD pathogenesis and could represent new pharmacological and biological targets for treatment and prevention.

Our study has limitations based on using AD-by-proxy phenotype for WGS-based GWAS in the AoU and UKB datasets. As AD is highly heritable, we relied on family history of AD in direct relatives (relatedness to first degree relatives is identical to a parent, or 50%) and grandparents to call AD-by-proxy cases. While relatedness with a grandparent is expected to be at minimum 25%, we chose a less conservative definition to align with prior studies^8,9^ and to increase sample size. AD-by-proxy has been previously shown to strongly correlate with definitive AD status. Despite concerns of introducing biases, AD-by-proxy offers the advantage of increasing statistical power in population-based biobanks^34^. Our results suggest that the correlation of AD-by-proxy with definitive AD is limited to comparisons among more homogeneous biobanks, such as UKB, and perhaps to individuals of European genetic ancestry (**Figure 1**). Our findings also underscore the advantages of incorporating diverse admixed cohorts, such as AoU, in genetic studies, as well as the importance of well-defined phenotyping in such cohorts. We observed many variants showing genome-wide significant association with AD in the AoU dataset versus UKB dataset and other clinical AD datasets. Some of those variants could be potential false-positives due to sequencing artifacts or unobserved confounding. An unusually large amount (59%) of genome-wide significant variants in AoU were indels and multiallelic variants. Filtering for indels, multiallelic variants and using more conservative allele frequency filters removes many of those signals. Although we only emphasized significant AD-by-proxy findings which showed a nominal replication in a clinical AD meta-analysis (**Figure 4**), this underscores the importance of validating results in independent datasets with a clinically-defined phenotype.

We were able to replicate roughly 11% of known AD GWAS loci in AoU. (**Supplementary Table 7**)^4^. This number increases to roughly 20% on stratifying AoU results by population. Thus, this discrepancy is perhaps due to population differences between the previous large AD GWAS and the more diverse analysis carried out here. The NIAGADS and AoU cohorts are made of up diverse individuals (nearly half non-European ancestry) that are traditionally underrepresented in AD genetics studies (**Supplemental Table 1**)^11^. In contrast, Bellenguez et al. analyzed a cohort predominantly made up of individuals with European ancestry from the European Alzheimer and Dementia Biobank and UKB^4^. Thus, while our AD-by-proxy AoU results reproduced some of the most robust AD-associated GWAS loci reported in Bellenguez et al, the missing variants could be due to lower power and more diverse cohorts investigated here.

In conclusion, we carried out WGS-based GWAS of both clinical AD and AD-by-proxy cohorts with more diverse genetic ancestry than prior studies, which have most commonly employed GWAS with common variant arrays focusing on populations with largely European ancestry. As a result, we identified 16 novel loci exhibiting genome-wide significance with AD in at least one dataset and nominal significance for association (in the same direction) in an independent cohort. Our results emphasize:

1. WGS-based GWAS can capture disease-associated loci, which could be missed by GWAS using only common variant arrays.
2. While AD-by-proxy case definitions of disease can be limiting, they can also enable plausible discoveries of novel disease loci especially when used alongside datasets with clinical diagnoses.

## Methods

### Cohorts

The NIAGADS dataset includes sequencing data and harmonized phenotypes from cohorts sequenced by the Alzheimer’s Disease Sequencing Project and other AD and Related Dementia’s studies. Full details can be found on NIAGADS at https://dss.niagads.org/datasets/ng00067/, and elsewhere^35^.

We used the NG00067.v9 release for this paper. The UK Biobank, a long-term study based in the United Kingdom, has gathered an extensive array of healthcare data from 502,357 individuals. This includes short-read whole-genome sequencing data from 200,004 of these participants, which were employed in this study. The All of Us cohort is a study including individuals traditionally underrepresented in biomedical research from the United States of America, providing short-read whole-genome sequencing calls for a total of 245,388 individuals in the current release 7^11,13^.

### Outcomes

In NIAGADS, AD cases were defined based on NINCDS-ADRDA criteria for possible, probable, or definite AD, had documented age at onset or age at death (for pathologically verified cases), and APOE genotyping. Controls were free of dementia and had an age over 60. In UKB and AoU, we cases were defined by either having an ICD-9 or ICD-10 code of Alzheimer’s disease (AD) or representing an AD-by-proxy case^4^, defined by an individual having a family history of AD in a first-degree relative or grandparent. While others have defined AD-by-proxy solely by having an affected parent^4^, with quantitative definitions suggesting this approach could limit bias, the genetic similarity among any first-degree relative is comparable. Affected grandparents are at minimum 25% genetically identical to participants, with precedence for their inclusion in the literature^8,9^. We could not use a quantitative definition in AoU as parental age and age of parental death were unavailable, compared to UKB. Although the quantitative definition for UKB was available, we used the binary AD-by-proxy phenotype as in AoU.

### Genome sequencing data processing

WGS variant calls for biallelic variants in vcf format were downloaded for the NIAGADS dataset from NG00067.v9 release. The dataset contained 34,438 subjects, however not all had an AD phenotype available. We excluded subjects which were technical replicates (the one which had less missing variant calls), those with a missing AD diagnosis, outliers based on HET/HOM ratio (6 standard deviations from the mean), subjects who had a high missingness rate (>5%), subjects from pairs which were second degree relatives or closer, based on running KING^36^ on the full dataset). The final NIAGADS dataset contained 25,660 subjects.

For the UKB data, WGS files were initially converted from their original VCF format to biallelic PGEN format using PLINK2 (with original multiallelic variants being split using bcftools) to make them compatible with subsequent Regenie analysis. The files were filtered for variants with more than 10% missingness, samples with more than 10% missingness, Hardy-Weinberg p-values less than 1 x 10^-15^, variants occurring in less than 3 individuals, and spanning deletions^4,18^.

For AoU data, Hail Variant Dataset (VDS) files were processed similarly to Wang et al. 2023, which studied cardiometabolic traits in an earlier AoU release, with analyses performed by other groups on UKB^18,23^. First, 550 samples flagged by AoU were removed from the data. Second, the VDS file was converted to a dense matrix format, removing variants flagged by AoU through internal quality-control and allele-specific variant quality score recalibration and monomorphic variants, including 33,526,160 variants without high-quality genotyping, 3,064,830 that were low quality, and 659,051 with excess heterozygosity. This filtered dataset was output to a Plink2 bgen file, leaving 1,085,790,733 variants and 244,845 samples.

### Genome-wide association (GWAS) analysis

For NIAGADS, we performed a logistic regression (with option “firth-fallback”) for case/control status as implemented in Plink2^16^. We included sex, sequencing center, sample set and 5 Jaccard principal components with standardized variance as covariates^37^.

Regenie v3.4.1 was used to conduct GWAS with settings recommended for UK Biobank analysis (https://rgcgithub.github.io/regenie/recommendations/)^18^. Covariates for the UKB analysis included age at enrollment into UKB, sex, the first twenty PCs, and sequencing center^38^. Covariates for the AoU analysis included age of enrollment, sex, and the first twenty principal components (PCs)^23^. Sequencing center information for each AoU sample was unavailable and was accomplished at three sites using identical reagents, instruments, and sample processing^11^. Outcomes were those defined above. Step 1 was accomplished using array data, similarly processed to Wang et al. 2023, filtered for variants with a MAF ≥ 1%, minimum allele count of 100, variant missingness ≤ 10%, unflagged samples, and samples shared with the Hail VDS dataset of samples sequenced by WGS, pruned for independent variants using 100 kb windows with a step-size of 1 and r^2^ threshold of 0.1, with sex chromosomes removed^23^. Step 2 was accomplished using Plink2 files filtered for variants with variant-level missingness ≤ 10% using array step 1 predictions. We used Firth penalized regression to variants with a p-value less than 0.01 and a minimum minor allele count of 20 for AoU. Reported AoU GWAS data was filtered for any variants where the MAF x N for the minor allele was less than 20, per AoU’s privacy policy. We did not filter by sample missingness in AoU given the internal sample and variant flagging. We ran GWAS in AoU for all individuals and for individuals of AFR, AMR, and EUR genetic ancestry, unable to do GWAS for EAS, MID, or SAS due to convergence issues from small numbers of cases in the cohorts (**Supplemental Table 1**). Step 2 for UKB employed a Firth penalized regression to variants with a P-value less than 0.05. Summary statistics for every genome-significant variant detected across our studies in the other studies is available (**Supplementary Table 3**).

### Principal component calculation

PCs for UKB were obtained from pre-calculated metrics. For AoU, array data that had been processed using the above quality control was input into Plink2 to yield the first twenty PCs^16^. PCs in NIAGADS were calculated based on a LD-pruned set of rare variants using the Jaccard index^37^.

### Meta analysis and quality control

Meta-analyzed results were processed using METAL^19^ (the most recent version, released on 25 March 2011) using default settings, aside from using inverse standard error values as weights for the AD-by-proxy analysis and outputting the average allele frequency across studies. The results were filtered for any genetic variants with a frequency amplitude less than 0.4 (or difference between the maximum and minimum allele frequency between studies^4^), variants that were genome-wide significant and had a HWE p-value larger than 1 ∗ 10^-15^ in all AoU ancestry-stratified populations (EUR, AFR, AMR, EAS, SAS, or MID) (**Supplementary Table 4**). Following this, we used Plink2 to clump every significant variant by chromosome in our study using AoU genetic data, using a threshold R^2^ of 1%^16^. We removed any meta-analysis signals that had varying directions of effect across studies where the variant was at minimum nominally significant. While the case proportion was greater in UKB compared to AoU, Firth correction can address imbalances^18^.

### Defining genomic loci and new versus old loci

To identify novel loci, we defined new loci as those whose variants were at least 500 kb from the transcriptional start site (TSS) of genes previously linked to AD^6,39^ or genome-significant variants from summary statistics available from Bellenguez et al. 2022 (GRCh38) and Wightman et al. 2021 (GRCh37, which we stepped-up using UCSC LiftOver)^4,40,41^. We obtained positional information of TSS from Gencode v44^42^.

### Evaluating our analyses’ power to detect associations from other studies

To evaluate our study’s power to detect associations observed by Bellenguez et al., we calculated power using formulas with code obtained from the University of Helsinki (https://www.mv.helsinki.fi/home/mjxpirin/GWAS_course/material/GWAS3.html)^4,43^. For calculations, we used Bellenguez et al.’s effect size and allele frequency^4^. Power was calculated for each variant individually.

### Genome-wide significant variant annotation

Variants that were genome-wide significant were evaluated for annotations in FAVOR, which provides distance from proximal genes and annotations on relationships to enhancer sequences, provided by GeneHancer and SuperEnhancer documentation^17^. A more detailed description of each score is available in its documentation, available at: favor.genohub.org.

### Cell-Based Gene Enrichment Studies

To evaluate the functional role of genes proximal to our study signals or linked by enhancer sequences using FAVOR GeneHancer or SuperEnhancer annotations in combination with the dbSuper database^17,44^, we evaluated brain single-cell data from Mathys et al. 2023 that studied differential gene expression by cognitive impairment and AD pathologic evidence status and resilience to cognitive impairment, given AD pathologic features^24^. To limit false positives, we used an FDR cutoff of 1%.

### Data availability

NIAGADS data access is available through the DSS NIAGADS under accession number: NG00067. The NIAGADS dataset contains data in part obtained from the Alzheimer’s Disease Neuroimaging Initiative (ADNI) database (adni.loni.usc.edu). As such, the investigators within the ADNI contributed to the design and implementation of ADNI and/or provided data but did not participate in analysis or writing of this report. UKB data access is available through application at https://ukbiobank.dnanexus.com/landing. Access to individual-level data from the All of Us research program was obtained through an MGB-signed data use agreement with All of Us (https://www.researchallofus.org/register/). This research was conducted using the UKB resource (application number 81874). Full GWAS data for this manuscript, edited to comply with privacy requirements for AoU, is available through Zenodo (10.5281/zenodo.13743529).

### Code availability

Code is publicly available on Github: https://github.com/juliandwillett/NIA_UKB_AoU_AlzheimersGWAS.

### Disclosures

None to disclose.

## Data Availability

All data produced in the present study are available upon reasonable request to the authors, with statistics for variants with a p-value less than 1e-5 available on Zenodo.

doi:10.5281/zenodo.13743529

## Acknowledgements

This work was supported by Cure Alzheimer’s Fund; NIGMS T32GM007748 (JW). We value the participants who volunteered for both the UKB and AoU. We appreciate the assistance of Cynthia Morton and James Gusella towards providing feedback for this study. The computations in this paper were run in part on the FASRC Cannon cluster supported by the FAS Division of Science Research Computing Group at Harvard University. The funding body has no role in the design of the study and collection, analysis, and interpretation of data and in writing the manuscript. Please refer to the Supplementary Note for full acknowledgements.

## Author Contributions (CRediT Author Statement)

Julian Willett: conceptualization, methodology, software, validation, formal analysis, investigation, data curation, writing – original draft, writing – review and editing, visualization, supervision. Mohammad Waqas: software, formal analysis, investigation, writing – review and editing. Younjung Choi: formal analysis and data curation. Tiffany Ngai: writing – review and editing. Kristina Mullin: project administration, and data curation, Rudolph Tanzi: conceptualization, validation, resources, writing – review and editing, supervision, funding acquisition, Dmitry Prokopenko: conceptualization, methodology, validation, formal analysis, resources, data curation, writing – review and editing, supervision, project administration, funding acquisition.

