## Supplementary Table 1 for "Identification of 16 novel Alzheimer’s disease susceptibility loci using multi-ancestry meta-analyses of clinical Alzheimer’s disease and AD-by-proxy cases from four whole genome sequencing datasets"

**Supplementary table 1**: Demographic information for subjects included in our study. NA is used for genetic ancestries that were not available in select cohorts. MAF - minor allele frequency; SD - standard deviation

| Cohort | NIAGADS v9 | UK Biobank | All of Us |
|---|---|---|---|
| AD Cases | 10,565 | 924 | 151 |
| AD-By-Proxy Cases | 0 | 25,785 | 11,290 |
| Controls | 15,095 | 133,844 | 233,548 |
| Total Sample Size | 25,660 | 159,629 | 244,838 |
| Case Percentage | 41.10% | 16.20% | 4.60% |
| African Ancestry | 6,261 (24.4%), 2,323 Cases | 10 (0.1%), 0 Cases | 56,702 (23.2%), 999 Cases |
| Admixed-American Ancestry | 1,413 (5.5%), 277 Cases | 622 (0.4%), 86 Cases | 44,886 (18.3%), 1,107 Cases |
| European Ancestry | 13,270 (51.7%), 7,395 Cases | 148,860 (93.3%), 24,542 Cases | 133,469 (54.5%), 8,878 Cases |
| Central/East/South Asian Ancestry | 4,686 (18.3%), 567 Cases | 2,476 (1.6%), 217 Cases | 8,852 (2.3%), 285 Cases |
| Middle-Eastern Ancestry | NA | 511 (0.3%), 51 Cases | 929 (0.4%), 21 Cases |
| Oceanic Ancestry | NA | 221 (0.1%), 28 Cases | NA |
| Age: Mean (SD) | 73.0 (10.0) | 56.3 (8.0) | 55.5 (17.0) |
| Female Sex Percentage | 61.90% | 56.90% | 59.30% |
| rs429358 MAF (ApoE4) | 21.00% | 15.30% | 14.90% |
| rs7412 MAF (ApoE2) | 5.20% | 7.90% | 7.80% |
