## Supplementary Table 2 for "Identification of 16 novel Alzheimer’s disease susceptibility loci using multi-ancestry meta-analyses of clinical Alzheimer’s disease and AD-by-proxy cases from four whole genome sequencing datasets"

**Supplementary table 2**: Summary statistics for every genome-significant variant from each study across studies.
MAF - minor allele frequency; P - p-value; HWE_PASS - variant passes for Hardy-Weinberg equilibrium filter in combined AoU analysis (p<=1e-15); AFR - African; AMR - admixed American; EUR - European; DIR - direction

| | | Cohorts with clinically diagnosed AD | | | | | | Cohorts with AD-by-proxy | | | | | | | | | | | | |
|---|---|---|---|---|---|---|---|---|---|---|---|---|---|---|---|---|---|---|---|---|
| | | NIAGADS | | NIMH | | Meta analysis | | UK Biobank | | All of US | | | Meta analysis | | Stratified by population analysis in All of US | | | | | |
| Variant ID | Effect allele | MAF | P | MAF | P | P | DIRECTION (NIA, NIMH) | MAF | P | MAF | P | HWE_PASS | P | DIRECTION (UKB, AoU) | AFR_P | AFR_DIR | AMR_P | AMR_DIR | EUR_P | EUR_DIR |
| 1-20328237-TG-T | T | 0.003059 | 4.6E-08 | | | 4.604E-08 | +? | | | 0.00966 | 0.177948 | TRUE | 0.1779 | ?- | 0.570254 | 1 | 0.731466 | -1 | 0.309025 | -1 |
| 1-165164548-A-G | G | 0.000487 | 2.09E-08 | | | 2.093E-08 | -? | | | 0.000137 | 0.985705 | TRUE | 0.9857 | ?+ | | | 0.845853 | -1 | | |
| 1-185620013-AATGTG | A | 0.002365 | 2.57E-08 | | | 2.57E-08 | +? | | | | | FALSE | | | | | | | | |
| 1-207510847-G-T | T | 0.127097 | 1.6E-09 | 0.195 | 0.059625 | 8.17E-10 | ++ | 0.825692 | 0.024721 | 0.868626 | 0.069156 | TRUE | 0.00398 | ++ | 0.260104 | 1 | 0.022021 | -1 | 0.106244 | -1 |
| 1-207512441-C-T | T | 0.145073 | 3E-10 | 0.209 | 0.163644 | 1.819E-10 | ++ | 0.814206 | 0.081946 | 0.845134 | 0.135176 | TRUE | 0.02247 | ++ | 0.1818 | 1 | 0.071771 | -1 | 0.154206 | -1 |
| 1-207524699-C-T | T | 0.142515 | 1.47E-10 | | | 1.468E-10 | +? | 0.816057 | 0.104165 | 0.846456 | 0.098274 | TRUE | 0.02257 | ++ | 0.186238 | 1 | 0.127518 | -1 | 0.075362 | -1 |
| 1-207564732-C-T | T | 0.139055 | 9.15E-10 | 0.201 | 0.017061 | 3.684E-10 | ++ | | | 0.849659 | 0.055024 | FALSE | 0.05502 | ?+ | 0.311722 | 1 | 0.06953 | -1 | 0.054832 | -1 |
| 1-207573951-G-A | A | 0.134092 | 3.63E-10 | 0.197 | 0.046681 | 1.726E-10 | ++ | 0.822916 | 0.083761 | 0.854964 | 0.041653 | TRUE | 0.009715 | ++ | 0.215188 | 1 | 0.042934 | -1 | 0.047587 | -1 |
| 1-207577223-C-T | T | 0.124561 | 2.23E-09 | 0.195 | 0.036457 | 1.052E-09 | ++ | | | 0.86938 | 0.031414 | TRUE | 0.03141 | ?+ | 0.413322 | 1 | 0.036254 | -1 | 0.046555 | -1 |
| 1-207611623-G-A | A | 0.222528 | 4.72E-08 | 0.241 | 0.139348 | 2.885E-08 | ++ | 0.816038 | 0.090461 | 0.760843 | 0.034112 | TRUE | 0.008675 | ++ | 0.709622 | -1 | 0.091626 | -1 | 0.071575 | -1 |
| 1-207613197-G-A | A | 0.163295 | 7.68E-09 | 0.215 | 0.07148 | 4.063E-09 | ++ | 0.820232 | 0.090651 | 0.821317 | 0.064459 | TRUE | 0.01409 | ++ | 0.667372 | 1 | 0.123263 | -1 | 0.05709 | -1 |
| 1-207621975-G-A | A | 0.164679 | 2.19E-09 | 0.215 | 0.081119 | 1.164E-09 | ++ | 0.820721 | 0.112267 | 0.819612 | 0.034585 | TRUE | 0.01154 | ++ | 0.858164 | -1 | 0.088832 | -1 | 0.047231 | -1 |
| 1-207623552-T-A | A | 0.13246 | 3.2E-10 | 0.198 | 0.081085 | 1.683E-10 | ++ | 0.823236 | 0.099784 | 0.857079 | 0.076805 | TRUE | 0.01818 | ++ | 0.442355 | 1 | 0.098717 | -1 | 0.065917 | -1 |
| 1-207625349-T-C | C | 0.164724 | 2.63E-09 | | | 2.627E-09 | -? | 0.82063 | 0.119668 | 0.819486 | 0.029305 | TRUE | 0.01112 | -- | 0.719056 | -1 | 0.079057 | -1 | 0.044034 | -1 |
| 1-207626529-C-A | A | 0.164607 | 2.34E-09 | 0.216 | 0.084246 | 1.253E-09 | ++ | 0.820689 | 0.107895 | 0.819542 | 0.027028 | TRUE | 0.009343 | ++ | 0.687779 | -1 | 0.088892 | -1 | 0.043427 | -1 |
| 1-207627210-C-T | T | 0.142267 | 2.42E-09 | 0.206 | 0.072722 | 1.268E-09 | ++ | 0.822393 | 0.093444 | 0.844761 | 0.094667 | TRUE | 0.01954 | ++ | 0.081446 | 1 | 0.093597 | -1 | 0.052879 | -1 |
| 1-207629207-C-A | A | 0.123275 | 3.62E-09 | 0.195 | 0.066785 | 1.907E-09 | ++ | 0.828495 | 0.052599 | 0.870525 | 0.029028 | TRUE | 0.004591 | ++ | 0.615408 | 1 | 0.044784 | -1 | 0.037963 | -1 |
| 1-207630796-C-A | A | 0.141925 | 2.71E-09 | 0.206 | 0.079471 | 1.447E-09 | ++ | 0.822153 | 0.101841 | 0.84493 | 0.080048 | TRUE | 0.01894 | ++ | 0.085375 | 1 | 0.094936 | -1 | 0.047587 | -1 |
| 1-207633385-A-G | G | 0.141602 | 2.86E-09 | 0.206 | 0.079471 | 1.528E-09 | -- | 0.8224 | 0.101234 | 0.845188 | 0.075036 | TRUE | 0.01796 | -- | 0.096848 | 1 | 0.091239 | -1 | 0.045782 | -1 |
| 1-207612944-G-A | A | 0.222586 | 7.86E-08 | 0.241 | 0.12557 | 4.733E-08 | ++ | 0.815997 | 0.099564 | 0.760782 | 0.0358 | TRUE | 0.009994 | ++ | 0.717579 | -1 | 0.092698 | -1 | 0.07392 | -1 |
| 1-207613483-G-A | A | 0.222642 | 6.61E-08 | 0.241 | 0.139348 | 4.052E-08 | ++ | 0.816035 | 0.099344 | 0.760843 | 0.038228 | TRUE | 0.01044 | ++ | 0.783975 | -1 | 0.092493 | -1 | 0.072713 | -1 |
| 1-8599804-C-CTGAGC | CTGAGCCTTTTTA | | | | | | | 3.45E-05 | 4.56E-08 | | | FALSE | 4.56E-08 | +? | | | | | | |
| 1-8599806-T-TCA | TCA | | | | | | | 3.45E-05 | 4.56E-08 | | | FALSE | 4.56E-08 | -? | | | | | | |
| 1-908962-G-GGTCGG | GGTCGGGGTC | | | | | | | | | 0.104825 | 8.69E-16 | FALSE | 8.69E-16 | ?+ | 0.909036 | -1 | 0.020259 | -1 | 9.24E-14 | -1 |
| 1-1277184-A-ATG | ATG | | | | | | | | | 0.000567 | 3.11E-08 | TRUE | 3.11E-08 | ?- | 0.964781 | 1 | 0.018289 | 1 | 3.59E-08 | 1 |
| 1-1280044-T-C | C | 0.494464 | 0.169393 | 0.299 | 0.762001 | 0.762 | ?+ | | | 0.470416 | 1.71E-10 | FALSE | 1.71E-10 | ?- | 0.365628 | 1 | 0.062507 | -1 | 5.71E-07 | 1 |
| 1-1370032-C-CAG | CAG | | | | | | | | | 0.000824 | 3.22E-10 | TRUE | 3.22E-10 | ?+ | 1.49E-06 | 1 | 8.61E-05 | 1 | 0.000496 | 1 |
| 1-1500804-AAAAT-A | A | 0.00141 | 0.111768 | | | 0.1118 | -? | 0.001867 | 0.87353 | 0.003974 | 1.75E-15 | TRUE | 1.39E-10 | ++ | 0.010021 | 1 | 0.002039 | 1 | 6.24E-12 | 1 |
| 1-1718301-C-T | T | | | | | | | | | 0.772999 | 2.29E-17 | FALSE | 2.29E-17 | ?- | 0.309078 | -1 | 0.494763 | -1 | 9.49E-21 | -1 |
| 1-1718311-A-T | T | | | | | | | | | 0.77949 | 8.13E-21 | FALSE | 8.13E-21 | ?+ | 0.172632 | -1 | 0.415792 | -1 | 8.18E-25 | -1 |
| 1-1718313-T-G | G | | | | | | | | | 0.779091 | 2.66E-14 | FALSE | 2.66E-14 | ?+ | 0.754689 | -1 | 0.590245 | -1 | 1.02E-18 | -1 |
| 1-3270621-T-G | G | | | | | | | | | 0.420679 | 1.07E-09 | FALSE | 1.07E-09 | ?- | 0.077591 | 1 | 0.703861 | -1 | 0.00018 | 1 |
| 1-3351529-T-TCCC | TCCC | | | | | | | | | 0.231635 | 1.87E-10 | FALSE | 1.87E-10 | ?+ | 0.001142 | -1 | 0.195221 | -1 | 8.25E-09 | -1 |
| 1-3351529-TGTC-T | T | | | | | | | | | 0.660588 | 1.26E-09 | FALSE | 1.26E-09 | ?+ | 0.009543 | 1 | 0.140674 | 1 | 1.65E-08 | 1 |
| 1-3798817-C-CAAA | CAAA | | | | | | | | | 0.000822 | 3.43E-15 | TRUE | 3.43E-15 | ?+ | 0.001452 | 1 | 0.001481 | 1 | 8.82E-10 | 1 |
| 1-3929441-C-CA | CA | | | | | | | | | 0.093927 | 6.41E-09 | FALSE | 6.41E-09 | ?- | 0.350768 | -1 | 0.064722 | -1 | 3.65E-07 | -1 |
| 1-5674860-GAAT-G | G | 0.000117 | 0.547418 | | | 0.5474 | -? | | | 0.00256 | 2.83E-08 | TRUE | 2.83E-08 | ?+ | 0.189001 | 1 | 0.500396 | -1 | 4.22E-10 | 1 |
| 1-5829844-A-G | G | | | | | | | | | 0.215173 | 2.06E-08 | FALSE | 2.06E-08 | ?+ | 0.247932 | -1 | 0.09994 | -1 | 1.13E-07 | -1 |
| 1-5884481-AGC-A | A | | | | | | | | | 0.002532 | 1.36E-08 | TRUE | 1.36E-08 | ?+ | 0.054161 | 1 | 0.369574 | -1 | 7.71E-09 | 1 |
| 1-7197400-A-G | G | | | | | | | 0.004764 | 0.552694 | 0.002554 | 2.24E-18 | TRUE | 8.1E-08 | -- | 0.000607 | 1 | 0.00953 | 1 | 1.15E-15 | 1 |
| 1-7542553-AGCCTCCT | A | | | | | | | 0.358142 | 0.617994 | 0.048984 | 2.94E-16 | FALSE | 0.000556 | ++ | 0.385159 | 1 | 0.208748 | 1 | 1.48E-17 | 1 |
| 1-8774498-CAT-C | C | | | | | | | | | 0.001151 | 4.74E-08 | TRUE | 4.74E-08 | ?- | 0.0063 | 1 | 0.408039 | -1 | 4.31E-07 | 1 |
| 1-8774520-A-AT | AT | | | | | | | 0.000407 | 0.027594 | 0.000518 | 2.49E-12 | TRUE | 2.96E-11 | -- | 0.022477 | 1 | 0.412396 | 1 | 3.55E-13 | 1 |
| 1-9158215-A-G | G | 0.000176 | 0.931184 | | | 0.9312 | +? | 9.71E-05 | 0.299098 | 0.000918 | 4.41E-10 | TRUE | 4.43E-10 | -- | 0.00015 | 1 | 0.230258 | 1 | 8.04E-06 | 1 |
| 1-9158256-AG-A | A | | | | | | | | | 0.000492 | 2.92E-09 | TRUE | 2.92E-09 | ?+ | 0.018125 | 1 | 0.001403 | 1 | 1.39E-06 | 1 |
| 1-9623342-C-CCT | CCT | | | | | | | | | 0.002957 | 1.73E-25 | TRUE | 1.73E-25 | ?+ | 2.39E-07 | 1 | 0.055264 | 1 | 4.5E-18 | 1 |
| 1-9623353-G-A | A | | | | | | | | | 0.000499 | 4.55E-08 | TRUE | 4.55E-08 | ?+ | 0.009319 | 1 | 0.003315 | 1 | 5.89E-06 | 1 |
| 1-9702141-A-T | T | | | | | | | | | 0.001653 | 6.5E-18 | TRUE | 6.5E-18 | ?- | 0.004281 | 1 | 0.000209 | 1 | 1.43E-13 | 1 |
| 1-9702143-A-T | T | | | | | | | | | 0.001718 | 3.16E-15 | TRUE | 3.15E-15 | ?- | 0.009023 | 1 | 0.011535 | 1 | 1.67E-13 | 1 |

| | | | | | | | | | | | | | | | | | | | | |
|---|---|---|---|---|---|---|---|---|---|---|---|---|---|---|---|---|---|---|---|---|
| 1-9830445-T-C | C | 0.311916 | 0.979624 | 0.196 | 0.32962 | 0.9323 | ++ | | | 0.317745 | 2.58E-13 | TRUE | 2.58E-13 | ?- | 0.172053 | 1 | 0.815398 | -1 | 0.000217 | 1 |
| 1-10152033-C-T | T | | | | | | | 4.39E-05 | 0.421838 | 0.001069 | 1.6E-09 | TRUE | 9.64E-09 | -+ | 0.033329 | 1 | 0.177081 | -1 | 1.16E-10 | 1 |
| 1-10152040-C-CCT | CCT | | | | | | | | | 0.003317 | 7.74E-24 | TRUE | 7.74E-24 | ?+ | 0.133177 | 1 | 0.231019 | 1 | 9.56E-24 | 1 |
| 1-10152050-A-AATTT | AATTT | | | | | | | | | 0.001179 | 3.35E-10 | TRUE | 3.35E-10 | ?- | 0.818478 | 1 | 0.03743 | 1 | 1.22E-09 | 1 |
| 1-10237828-G-A | A | | | | | | | 0.000132 | 0.550187 | 0.000802 | 2E-10 | TRUE | 1.05E-09 | ++ | 0.658855 | 1 | 0.000277 | 1 | 1.48E-08 | 1 |
| 1-10508359-A-G | G | | | | | | | | | 0.000234 | 1.19E-10 | TRUE | 1.19E-10 | ?- | 0.017875 | 1 | | | 2.09E-09 | 1 |
| 1-10508363-T-C | C | | | | | | | | | 0.000207 | 1.9E-10 | TRUE | 1.9E-10 | ?- | 0.011153 | 1 | | | 1.05E-07 | 1 |
| 1-11709990-G-T | T | 7.8E-05 | 0.805963 | | | 0.806 | +? | 0.000113 | 0.464033 | 0.001305 | 3.38E-10 | TRUE | 5.12E-10 | ++ | 0.020388 | 1 | 0.050116 | 1 | 3.29E-09 | 1 |
| 1-13530355-G-A | A | | | | | | | 0.000138 | 0.96944 | 0.0011 | 6.63E-10 | TRUE | 6.94E-09 | -+ | 0.598718 | -1 | 4.29E-05 | 1 | 1.68E-07 | 1 |
| 1-15860102-G-A | A | 0.000117 | 0.130664 | | | 0.1307 | +? | 0.001425 | 0.992715 | 0.001759 | 2.2E-11 | TRUE | 2.69E-06 | -+ | 0.235722 | 1 | 0.000846 | 1 | 1.27E-08 | 1 |
| 1-15860133-G-T | T | 0.000216 | 0.11596 | | | 0.116 | +? | 0.002888 | 0.865415 | 0.006362 | 2.18E-45 | TRUE | 2.4E-30 | ++ | 1.06E-07 | 1 | 1.5E-09 | 1 | 1.1E-30 | 1 |
| 1-15860146-C-T | T | | | | | | | 0.002644 | 0.901398 | 0.003768 | 1.87E-26 | TRUE | 2.2E-15 | ++ | 2.59E-05 | 1 | 2E-06 | 1 | 2.99E-17 | 1 |
| 1-15869308-GC-G | G | | | | | | | | | 0.000545 | 1.84E-08 | TRUE | 1.84E-08 | ?+ | 0.296672 | 1 | 0.051654 | 1 | 5.51E-07 | 1 |
| 1-15977137-T-A | A | | | | | | | 7.52E-05 | 0.322022 | 0.000567 | 7.93E-09 | TRUE | 3.52E-07 | -+ | 0.022332 | 1 | 0.591736 | 1 | 1.44E-08 | 1 |
| 1-15977177-CAA-C | C | | | | | | | | | 0.001313 | 5.38E-10 | TRUE | 5.38E-10 | ?- | 0.0196 | 1 | 0.022136 | 1 | 8.74E-09 | 1 |
| 1-16382341-C-CCA | CCA | | | | | | | | | 0.000537 | 7.08E-11 | TRUE | 7.08E-11 | ?+ | 3.5E-07 | 1 | 0.000194 | 1 | 1.92E-05 | 1 |
| 1-16429296-C-CCCGG | CCCGG | 0.000117 | 0.415779 | | | 0.4158 | +? | | | 0.00137 | 1.31E-10 | TRUE | 1.31E-10 | ?+ | 0.262951 | 1 | 0.002716 | 1 | 6.88E-09 | 1 |
| 1-16516729-GC-G | G | 0.012558 | 0.819185 | | | 0.8192 | +? | 0.00311 | 0.114071 | 0.004008 | 7.42E-10 | TRUE | 0.07768 | +- | 0.079678 | -1 | 0.410479 | -1 | 6.13E-09 | -1 |
| 1-16732342-G-A | A | 0.000942 | 0.669616 | | | 0.6696 | -? | | | 0.006786 | 4.05E-09 | TRUE | 4.05E-09 | ?+ | 0.332965 | 1 | 0.126088 | 1 | 4.4E-08 | 1 |
| 1-17006395-CATGCAC | C | | | | | | | | | 0.002502 | 4.9E-25 | TRUE | 4.9E-25 | ?- | 1.18E-07 | 1 | 0.16377 | 1 | 2.12E-20 | 1 |
| 1-17054484-CT-C | C | 0.001427 | 0.520824 | | | 0.5208 | +? | | | 0.00229 | 1.66E-08 | TRUE | 1.66E-08 | ?- | 0.047014 | 1 | 0.629902 | 1 | 4.92E-08 | 1 |
| 1-17543895-G-GCCCA | GCCCA | | | | | | | | | 0.001037 | 2.25E-08 | TRUE | 2.25E-08 | ?- | 0.60562 | -1 | 0.131607 | 1 | 3.75E-08 | 1 |
| 1-17544491-C-T | T | | | | | | | 0.000993 | 0.112974 | 0.001771 | 5.15E-13 | TRUE | 3.83E-11 | ++ | 0.015906 | 1 | 0.007713 | 1 | 3.68E-10 | 1 |
| 1-17544499-G-T | T | 0.073602 | 0.74189 | | | 0.7419 | +? | 0.080753 | 1.52E-05 | 0.089157 | 2.37E-16 | TRUE | 7.19E-17 | ++ | 0.014437 | 1 | 0.000134 | 1 | 3.93E-12 | 1 |
| 1-17544542-A-ACCTG | ACCTG | | | | | | | | | 0.000607 | 1.47E-10 | TRUE | 1.47E-10 | ?- | 0.007355 | 1 | 0.001595 | 1 | 2.52E-06 | 1 |
| 1-20463538-A-ACC | ACC | | | | | | | | | 0.011826 | 5.43E-15 | TRUE | 5.42E-15 | ?- | 0.145378 | 1 | 0.073589 | 1 | 9.32E-15 | 1 |
| 1-20647193-AC-A | A | | | | | | | | | 0.000308 | 1.37E-08 | TRUE | 1.37E-08 | ?+ | 0.47175 | 1 | 9.18E-05 | 1 | 7.32E-06 | 1 |
| 1-20796590-C-CCT | CCT | | | | | | | | | 0.006205 | 1.85E-12 | TRUE | 1.85E-12 | ?+ | 0.2303 | 1 | 0.031523 | 1 | 7.87E-11 | 1 |
| 1-22058633-C-CCACC/ | CCACCA | | | | | | | | | 0.001004 | 1.07E-10 | TRUE | 1.08E-10 | ?+ | 0.301661 | 1 | 0.036594 | 1 | 1.01E-08 | 1 |
| 1-22429164-C-CCT | CCT | | | | | | | | | 0.000515 | 3.24E-08 | TRUE | 3.24E-08 | ?+ | 0.402425 | -1 | 0.964018 | 1 | 1.89E-09 | 1 |
| 1-23898335-C-CCAA | CCAA | | | | | | | | | 0.080728 | 7.44E-36 | FALSE | 7.44E-36 | ?+ | 0.003711 | 1 | 0.003131 | 1 | 2.57E-29 | 1 |
| 1-23898335-CTTTTTT- | C | | | | | | | | | 0.788816 | 8.13E-42 | FALSE | 8.13E-42 | ?+ | 0.028409 | -1 | 0.042259 | -1 | 4.8E-40 | -1 |
| 1-24812865-CAT-C | C | | | | | | | | | 0.001779 | 1.69E-16 | TRUE | 1.7E-16 | ?- | 7.36E-05 | 1 | 0.026432 | 1 | 1.88E-13 | 1 |
| 1-24812868-G-GCA | GCA | | | | | | | | | 0.001782 | 6.34E-14 | TRUE | 6.34E-14 | ?- | 0.00557 | 1 | 0.058634 | 1 | 1.64E-12 | 1 |
| 1-25925787-AGTGC-A | A | | | | | | | | | 0.000544 | 3.09E-10 | TRUE | 3.09E-10 | ?+ | 0.060533 | 1 | 0.53606 | 1 | 8.55E-10 | 1 |
| 1-26108552-TG-T | T | 0.001884 | 0.147344 | | | 0.1473 | -? | 0.000179 | 0.447481 | 0.001783 | 1.91E-10 | TRUE | 3.98E-10 | ++ | 1.26E-05 | 1 | 0.175742 | 1 | 8.6E-08 | 1 |
| 1-26334705-A-ACC | ACC | | | | | | | | | 0.001288 | 4.5E-11 | TRUE | 4.5E-11 | ?- | 0.00135 | 1 | 0.387991 | 1 | 6.83E-10 | 1 |
| 1-26334710-GTCAA-G | G | | | | | | | | | 0.000543 | 9.13E-10 | TRUE | 9.13E-10 | ?+ | 0.005178 | 1 | 0.829592 | 1 | 6.02E-09 | 1 |
| 1-26648341-CCT-C | C | 0.000234 | 0.168332 | | | 0.1683 | +? | | | 0.00098 | 1.09E-08 | TRUE | 1.09E-08 | ?- | 0.344697 | 1 | 0.822032 | 1 | 1.18E-09 | 1 |
| 1-26651469-AGC-A | A | | | | | | | | | 0.000799 | 4.22E-10 | TRUE | 4.22E-10 | ?+ | 0.790529 | 1 | 0.018964 | 1 | 7.86E-10 | 1 |
| 1-26651472-C-CTT | CTT | | | | | | | | | 0.000624 | 1.55E-08 | TRUE | 1.55E-08 | ?+ | 0.509302 | 1 | 0.041007 | 1 | 6.91E-08 | 1 |
| 1-27232615-C-CA | CA | | | | | | | | | 0.002862 | 9.46E-09 | TRUE | 9.46E-09 | ?+ | 0.105123 | 1 | 0.41594 | -1 | 9.81E-10 | 1 |
| 1-27865762-ATT-A | A | 0.000685 | 0.459284 | | | 0.4593 | +? | 0.0012 | 0.184637 | 0.00154 | 3.18E-08 | TRUE | 0.003287 | -+ | 0.050376 | 1 | 0.001607 | 1 | 3.22E-05 | 1 |
| 1-28350749-GTA-G | G | | | | | | | | | 0.000669 | 4.1E-10 | TRUE | 4.1E-10 | ?+ | 0.000295 | 1 | 0.776998 | -1 | 1.74E-09 | 1 |
| 1-28391247-AAAT-A | A | 0.000235 | 0.072814 | | | 0.07281 | +? | | | 0.005998 | 3.3E-09 | TRUE | 3.3E-09 | ?+ | 0.013611 | 1 | 0.06203 | 1 | 7.78E-08 | 1 |
| 1-28649458-C-CAG | CAG | | | | | | | | | 0.003734 | 3.4E-15 | TRUE | 3.4E-15 | ?+ | 0.098805 | 1 | 0.0002 | 1 | 1.46E-11 | 1 |
| 1-28649461-GAT-G | G | 0.000352 | 0.59874 | | | 0.5987 | -? | | | 0.003142 | 3.36E-14 | TRUE | 3.36E-14 | ?+ | 0.023501 | 1 | 7.28E-05 | 1 | 1.14E-10 | 1 |
| 1-28649645-GGCA-G | G | | | | | | | | | 0.000838 | 1.64E-14 | TRUE | 1.64E-14 | ?+ | 5.83E-06 | 1 | 0.213783 | 1 | 3.33E-11 | 1 |
| 1-28715380-C-CCT | CCT | | | | | | | | | 0.000843 | 3.47E-08 | TRUE | 3.47E-08 | ?+ | 0.60847 | 1 | 0.525888 | -1 | 7.22E-09 | 1 |
| 1-28733121-A-AT | AT | | | | | | | | | 0.000358 | 3.39E-12 | TRUE | 3.39E-12 | ?- | 1.69E-05 | 1 | 0.096013 | 1 | 3.5E-09 | 1 |
| 1-28994406-C-CAA | CAA | | | | | | | | | 0.001177 | 1.23E-08 | TRUE | 1.23E-08 | ?+ | 0.330207 | 1 | 0.5876 | 1 | 6.58E-08 | 1 |
| 1-29175504-G-C | C | 0.000137 | 0.11169 | | | 0.1117 | -? | 0.003978 | 0.682523 | 0.001859 | 5.22E-14 | TRUE | 0.000228 | -+ | 0.00118 | 1 | 0.002311 | 1 | 5.15E-11 | 1 |
| 1-29175505-G-C | C | | | | | | | 0.003978 | 0.682523 | 0.001622 | 5.25E-12 | TRUE | 0.001454 | -+ | 0.006342 | 1 | 0.013848 | 1 | 1.6E-10 | 1 |
| 1-31578988-T-C | C | | | | | | | | | 0.99201 | 1.58E-16 | TRUE | 1.58E-16 | ?+ | 0.012156 | -1 | 0.597919 | -1 | 9.6E-17 | -1 |
| 1-31579040-G-A | A | | | | | | | | | 0.985892 | 2.62E-27 | FALSE | 2.62E-27 | ?- | 0.008458 | -1 | 0.007642 | -1 | 9.07E-22 | -1 |
| 1-32738644-G-C | C | | | | | | | | | 0.419017 | 2.27E-09 | FALSE | 2.27E-09 | ?- | 0.427298 | -1 | 0.044574 | -1 | 2.26E-08 | -1 |
| 1-35117555-CAG-C | C | | | | | | | | | 0.000953 | 3.86E-11 | TRUE | 3.86E-11 | ?- | 0.000354 | 1 | 0.489856 | -1 | 1.97E-09 | 1 |
| 1-35579988-CCT-C | C | 7.8E-05 | 0.845527 | | | 0.8455 | +? | | | 0.000777 | 4.69E-09 | TRUE | 4.69E-09 | ?- | 0.001937 | 1 | 0.219915 | -1 | 2.29E-09 | 1 |
| 1-36102439-C-A | A | | | | | | | 0.001049 | 0.750459 | 0.001962 | 1.18E-14 | TRUE | 4.66E-10 | ++ | 0.079737 | 1 | 0.007705 | 1 | 4.92E-12 | 1 |

| | | | | | | | | | | | | | | | | | | | | |
|---|---|---|---|---|---|---|---|---|---|---|---|---|---|---|---|---|---|---|---|---|
| 1-36102460-CTGGCTA | C | | | | | | | 0.000777 | 0.252577 | 0.000958 | 5.5E-14 | TRUE | 1.04E-06 | +- | 0.104175 | 1 | 0.002221 | 1 | 3.41E-11 | 1 |
| 1-36571154-CT-C | C | | | | | | | 2.51E-05 | 0.486388 | 0.001305 | 2.47E-09 | TRUE | 1.99E-09 | -- | 0.316476 | 1 | 0.571858 | 1 | 1.56E-09 | 1 |
| 1-37884763-A-AAT | AAT | 0.000332 | 0.243233 | | | 0.2432 | +? | | | 0.000791 | 4.6E-09 | TRUE | 4.6E-09 | ?- | 0.93299 | -1 | 0.00274 | 1 | 1.01E-06 | 1 |
| 1-37884766-ATG-A | A | | | | | | | | | 0.00066 | 4.22E-09 | TRUE | 4.22E-09 | ?+ | 0.304242 | -1 | 0.001036 | 1 | 9.06E-07 | 1 |
| 1-37935848-C-CCA | CCA | | | | | | | 0.000648 | 0.518856 | 0.006101 | 1.27E-27 | TRUE | 1.63E-23 | -+ | 0.000249 | 1 | 0.012818 | 1 | 2.23E-21 | 1 |
| 1-37941708-G-GT | GT | 0.000626 | 0.76761 | | | 0.7676 | +? | 0.001262 | 0.767317 | 0.00281 | 1.18E-08 | TRUE | 3.08E-06 | -- | 0.009176 | 1 | 0.009481 | 1 | 7.22E-07 | 1 |
| 1-38869984-T-A | A | | | | | | | 0.000194 | 0.745338 | 0.000963 | 6.08E-15 | TRUE | 2.11E-12 | -+ | 0.010064 | 1 | 0.19057 | 1 | 2.19E-14 | 1 |
| 1-39067815-T-G | G | | | | | | | 1.57E-05 | 0.844572 | 0.000101 | 3.03E-09 | TRUE | 1.05E-08 | -- | | | | | 2.11E-07 | 1 |
| 1-39179844-G-A | A | | | | | | | 0.000692 | 0.82374 | 0.002105 | 5.74E-23 | TRUE | 7.82E-18 | ++ | 0.044106 | 1 | 0.013968 | 1 | 1.73E-20 | 1 |
| 1-39504208-GAT-G | G | 0.00117 | 0.169388 | | | 0.1694 | -? | 0.002907 | 0.781916 | 0.003843 | 1.42E-19 | TRUE | 3.52E-11 | ++ | 0.002526 | 1 | 0.003018 | 1 | 1.07E-15 | 1 |
| 1-39533298-T-C | C | | | | | | | 0.002182 | 0.943951 | 0.001981 | 1.28E-10 | TRUE | 3.49E-05 | -- | 0.103701 | 1 | 0.006985 | 1 | 1.16E-07 | 1 |
| 1-39533299-C-A | A | | | | | | | 0.002188 | 0.884992 | 0.001911 | 7.91E-10 | TRUE | 5.98E-05 | ++ | 0.010247 | 1 | 0.127023 | 1 | 7.49E-08 | 1 |
| 1-39579706-CGTG-C | C | | | | | | | | | 0.000649 | 7.97E-09 | TRUE | 7.97E-09 | ?- | 0.049081 | 1 | 0.418685 | 1 | 7.4E-07 | 1 |
| 1-39606520-C-T | T | 0.000944 | 0.417655 | | | 0.4177 | -? | 0.000119 | 0.775827 | 0.00118 | 1.16E-08 | TRUE | 1.05E-07 | -+ | 0.352765 | 1 | 0.188803 | 1 | 3.87E-08 | 1 |
| 1-39606527-A-C | C | 0.000706 | 0.067856 | | | 0.06786 | +? | 0.000135 | 0.929636 | 0.001565 | 1.85E-11 | TRUE | 1.65E-10 | +- | 0.005186 | 1 | 0.521646 | 1 | 5.58E-10 | 1 |
| 1-39606529-C-T | T | 0.000628 | 0.019746 | | | 0.01975 | -? | | | 0.000874 | 1.9E-11 | TRUE | 1.9E-11 | ?+ | 0.02238 | 1 | 0.318583 | 1 | 1.77E-10 | 1 |
| 1-39993465-TTC-T | T | | | | | | | | | 0.00106 | 3.34E-09 | TRUE | 3.34E-09 | ?+ | 0.711236 | 1 | 0.005696 | 1 | 8.74E-08 | 1 |
| 1-40590743-C-CAA | CAA | | | | | | | | | 0.000539 | 8.47E-09 | TRUE | 8.47E-09 | ?+ | 0.007708 | 1 | 0.002225 | 1 | 1.85E-06 | 1 |
| 1-40590746-TATCG-T | T | | | | | | | | | 0.000314 | 3.29E-08 | TRUE | 3.29E-08 | ?+ | 0.006932 | 1 | 0.000917 | 1 | 1.26E-05 | 1 |
| 1-42614311-ACCT-A | A | | | | | | | | | 0.002108 | 1.45E-14 | TRUE | 1.45E-14 | ?+ | 0.017618 | 1 | 0.256096 | 1 | 1.36E-12 | 1 |
| 1-43248985-C-A | A | | | | | | | 0.000708 | 0.789124 | 0.002292 | 2.94E-15 | TRUE | 9.17E-12 | ++ | 3.34E-05 | 1 | 0.012843 | 1 | 2.33E-10 | 1 |
| 1-44148272-C-CTACT | CTACT | | | | | | | | | 0.000714 | 4.14E-08 | TRUE | 4.15E-08 | ?+ | 0.002367 | 1 | 0.204726 | 1 | 5.15E-07 | 1 |
| 1-44952753-T-C | C | 9.76E-05 | 0.430127 | | | 0.4301 | -? | 0.004222 | 0.125232 | 0.003845 | 3.51E-26 | TRUE | 2.9E-16 | -- | 0.020138 | 1 | 0.002719 | 1 | 1.04E-20 | 1 |
| 1-44952782-T-G | G | | | | | | | 0.005945 | 0.974855 | 0.006977 | 2.69E-35 | TRUE | 5.09E-18 | -- | 0.027994 | 1 | 0.000163 | 1 | 1.8E-30 | 1 |
| 1-46311457-GC-G | G | | | | | | | | | 0.003276 | 1.41E-10 | TRUE | 1.41E-10 | ?+ | 0.075994 | 1 | 0.00615 | 1 | 1.85E-07 | 1 |
| 1-46323003-A-ACC | ACC | | | | | | | | | 0.001229 | 3.65E-10 | TRUE | 3.65E-10 | ?- | 0.390474 | 1 | 0.444828 | -1 | 1.09E-11 | 1 |
| 1-46337062-C-A | A | | | | | | | 0.001632 | 0.861843 | 0.002961 | 8.51E-20 | TRUE | 6.43E-13 | ++ | 0.005538 | 1 | 0.444994 | 1 | 2.03E-18 | 1 |
| 1-46337077-G-A | A | | | | | | | 0.001873 | 0.650541 | 0.003641 | 8.98E-31 | TRUE | 3.97E-21 | ++ | 0.019163 | 1 | 0.09866 | 1 | 2.58E-30 | 1 |
| 1-46337088-G-A | A | | | | | | | 0.001958 | 0.984205 | 0.004053 | 7.11E-29 | TRUE | 5.95E-19 | -+ | 0.004346 | 1 | 0.032549 | 1 | 6.88E-26 | 1 |
| 1-49135071-T-G | G | | | | | | | 0.003452 | 0.588821 | 0.001832 | 6.87E-16 | TRUE | 6.02E-07 | -- | 0.009661 | 1 | 0.00176 | 1 | 1.65E-12 | 1 |
| 1-50369313-A-ATGAT | ATGATGATGGTAGTAATGACGGTGATGATGATGATGGTGATGATGATGGTGGTGATGATGGTAATGGTGATCAT | | | | | | | | | 0.567276 | 8.54E-12 | FALSE | 8.55E-12 | ?- | 0.000204 | 1 | 0.020455 | 1 | 3.35E-07 | 1 |
| 1-52623572-C-T | T | | | | | | | | | 0.000114 | 1.16E-08 | TRUE | 1.17E-08 | ?+ | | | | | 8E-09 | 1 |
| 1-52910506-T-C | C | | | | | | | | | 0.000481 | 1.31E-09 | TRUE | 1.31E-09 | ?- | 0.006525 | 1 | 0.016103 | 1 | 1.04E-06 | 1 |
| 1-52910512-GCAC-G | G | | | | | | | | | 0.000605 | 3.37E-09 | TRUE | 3.37E-09 | ?+ | 0.10069 | 1 | 0.066301 | 1 | 4.98E-08 | 1 |
| 1-53283749-C-CCA | CCA | | | | | | | | | 0.213206 | 1.28E-09 | FALSE | 1.28E-09 | ?+ | 0.021794 | 1 | 0.265686 | 1 | 1.99E-08 | 1 |
| 1-54466273-A-ACC | ACC | | | | | | | | | 0.000311 | 4.74E-10 | TRUE | 4.74E-10 | ?- | 0.000122 | 1 | 0.601611 | 1 | 1.47E-07 | 1 |
| 1-54466274-TTG-T | T | | | | | | | | | 0.000453 | 3.69E-14 | TRUE | 3.69E-14 | ?+ | 6.5E-05 | 1 | 0.183269 | 1 | 5.23E-11 | 1 |
| 1-54504924-CA-C | C | | | | | | | | | 0.000822 | 1.51E-08 | TRUE | 1.51E-08 | ?- | 0.039113 | 1 | 0.010631 | 1 | 3.09E-07 | 1 |
| 1-54505400-C-CA | CA | 0.000198 | 0.378909 | | | 0.3789 | -? | 0.001554 | 0.11527 | 0.001664 | 2.2E-08 | TRUE | 8.53E-07 | ++ | 0.016633 | 1 | 0.205367 | 1 | 1.32E-06 | 1 |
| 1-61155454-A-ACC | ACC | 0.000801 | 0.307135 | | | 0.3071 | +? | | | 0.003414 | 4.31E-12 | TRUE | 4.31E-12 | ?- | 0.00073 | 1 | 0.047935 | 1 | 7.84E-09 | 1 |
| 1-63237607-C-CA | CA | | | | | | | | | 0.000739 | 1.78E-09 | TRUE | 1.78E-09 | ?+ | 0.003455 | 1 | 0.01646 | 1 | 1.87E-06 | 1 |
| 1-77000060-C-T | T | | | | | | | | | 0.52769 | 3.32E-08 | FALSE | 3.32E-08 | ?- | 0.03236 | 1 | 0.173068 | -1 | 1.35E-09 | -1 |
| 1-85367115-G-A | A | | | | | | | | | 0.000251 | 4.81E-09 | TRUE | 4.81E-09 | ?+ | 1.64E-05 | 1 | 0.085953 | 1 | 3.14E-06 | 1 |
| 1-86848155-A-T | T | | | | | | | | | 0.000533 | 9.51E-10 | TRUE | 9.51E-10 | ?- | 0.00844 | 1 | 0.00631 | 1 | 2E-06 | 1 |
| 1-88806458-C-CCG | CCG | | | | | | | | | 0.000965 | 1.52E-08 | TRUE | 1.52E-08 | ?+ | 0.00557 | 1 | 0.016357 | 1 | 2.46E-06 | 1 |
| 1-90050171-A-AGG | AGG | | | | | | | | | 0.000173 | 6.99E-10 | TRUE | 6.99E-10 | ?- | | | | | 1.24E-07 | 1 |
| 1-100385985-G-A | A | | | | | | | 0.001171 | 0.210958 | 0.001182 | 3.53E-11 | TRUE | 0.000263 | -+ | 0.014538 | 1 | 0.016614 | 1 | 1.16E-09 | 1 |
| 1-101553030-T-A | A | | | | | | | | | 0.777359 | 1.72E-13 | FALSE | 1.72E-13 | ?+ | 0.578709 | 1 | 0.215764 | 1 | 4.67E-13 | 1 |
| 1-101553030-T-TATA | TATACATGTATATTATAGATATATACATGTATATACATGTATATATTATAGAATATACATGTATATACATGTATATAT | | | | | | | | | 0.062422 | 1.43E-12 | FALSE | 1.43E-12 | ?+ | 0.194081 | -1 | 0.941497 | 1 | 1.39E-12 | -1 |
| 1-112574467-G-A | A | | | 0.001 | 0.317311 | 0.3173 | ?+ | 0.001068 | 0.436548 | 0.001749 | 4.55E-09 | TRUE | 6.48E-05 | -+ | 0.000819 | 1 | 0.305026 | 1 | 1.24E-07 | 1 |
| 1-112574476-T-G | G | | | 0.001 | 1 | 1 | ?0 | 0.001172 | 0.021861 | 0.002804 | 1.85E-10 | TRUE | 1.81E-10 | -- | 0.002086 | 1 | 0.19185 | 1 | 5.39E-08 | 1 |
| 1-121400518-GAA-G | G | | | | | | | | | 0.00281 | 1.18E-08 | TRUE | 1.19E-08 | ?+ | 0.014146 | 1 | 0.001061 | 1 | 0.000265 | 1 |
| 1-125068456-T-G | G | | | | | | | | | 0.000508 | 1.18E-09 | TRUE | 1.18E-09 | ?- | 0.0019 | 1 | 0.002293 | 1 | 8.06E-06 | 1 |
| 1-125176581-G-T | T | | | | | | | | | 0.000737 | 1.79E-08 | TRUE | 1.79E-08 | ?+ | 0.025947 | 1 | 0.01524 | 1 | 3.95E-06 | 1 |
| 1-125176624-A-G | G | | | | | | | | | 0.802676 | 1.46E-17 | FALSE | 1.46E-17 | ?+ | 4.41E-11 | -1 | 0.049348 | -1 | 1.21E-10 | -1 |
| 1-143195687-T-TGAA | TGAATGGAATCATCATC | | | | | | | | | 0.001191 | 1.41E-11 | TRUE | 1.41E-11 | ?- | 0.632804 | -1 | 0.29426 | 1 | 3.51E-12 | 1 |
| 1-143202364-G-A | A | | | | | | | | | 0.993996 | 6.47E-12 | TRUE | 6.47E-12 | ?- | 0.7989 | 1 | 0.158141 | -1 | 2.94E-13 | -1 |
| 1-143210077-A-T | T | | | | | | | 0.999859 | 0.728836 | 0.987881 | 9.7E-118 | TRUE | 2.5E-116 | -+ | 1.52E-23 | -1 | 2.49E-05 | -1 | 2.39E-90 | -1 |
| 1-143210080-A-T | T | | | | | | | 0.999518 | 0.353217 | 0.981214 | 1.8E-152 | FALSE | 6.3E-151 | ++ | 4.6E-22 | -1 | 1.39E-09 | -1 | 5.7E-124 | -1 |

| | | | | | | | | | | | | | | | | | | | | |
|---|---|---|---|---|---|---|---|---|---|---|---|---|---|---|---|---|---|---|---|---|
| 1-143241011-AT-A | A | | | | | | | | | 0.968028 | 3.83E-11 | FALSE | 3.83E-11 | ?- | 0.004878 | -1 | 0.002055 | -1 | 6.34E-07 | -1 |
| 1-143242279-ATCGAA | A | | | | | | | | | 0.001308 | 5.39E-14 | FALSE | 5.39E-14 | ?+ | 0.019614 | 1 | 0.000836 | 1 | 2.89E-08 | 1 |
| 1-143253866-C-T | T | | | | | | | | | 0.003083 | 2.19E-11 | TRUE | 2.19E-11 | ?+ | 0.02231 | 1 | 0.615739 | 1 | 3.67E-08 | 1 |
| 1-143254157-T-A | A | | | | | | | | | 0.81265 | 7.98E-09 | FALSE | 7.98E-09 | ?+ | 0.010915 | 1 | 0.151104 | 1 | 1.66E-07 | 1 |
| 1-143272094-A-G | G | | | | | | | 4.09E-05 | 0.149567 | 0.000792 | 1.14E-11 | TRUE | 1.79E-10 | +- | 0.587126 | 1 | 0.001434 | 1 | 1.7E-09 | 1 |
| 1-143272167-CG-C | C | | | | | | | | | 0.000825 | 3.94E-08 | TRUE | 3.94E-08 | ?- | 0.164962 | 1 | 0.694287 | -1 | 4.01E-08 | 1 |
| 1-143273397-A-G | G | | | | | | | 0.001197 | 0.997336 | 0.000556 | 4.13E-08 | TRUE | 0.002372 | +- | 2.84E-05 | 1 | 0.173011 | 1 | 0.000233 | 1 |
| 1-144565462-T-C | C | 0.000156 | 0.315235 | | | 0.3152 | +? | | | 0.001018 | 1.88E-11 | TRUE | 1.88E-11 | ?- | 0.107943 | 1 | 0.000835 | 1 | 2.72E-09 | 1 |
| 1-144636407-C-CA | CA | | | | | | | | | 0.010089 | 8.17E-12 | TRUE | 8.17E-12 | ?+ | 0.995938 | 1 | 0.005894 | 1 | 1.88E-09 | 1 |
| 1-144636407-C-CTTC | CTTCA | | | | | | | | | 0.987006 | 1.34E-10 | TRUE | 1.34E-10 | ?- | 0.831926 | -1 | 0.003952 | -1 | 2.1E-08 | -1 |
| 1-146944372-CCA-C | C | | | | | | | | | 0.048897 | 3.93E-08 | FALSE | 3.93E-08 | ?- | 0.151763 | 1 | 0.495758 | -1 | 1.17E-09 | 1 |
| 1-147519765-G-GA | GA | | | | | | | | | 0.956188 | 7.89E-10 | FALSE | 7.89E-10 | ?+ | 0.751102 | -1 | 0.175771 | -1 | 1.53E-10 | -1 |
| 1-147519767-A-AGT | AGT | | | | | | | | | 0.017402 | 3.7E-08 | FALSE | 3.7E-08 | ?- | 0.48247 | -1 | 0.039626 | 1 | 8.05E-08 | 1 |
| 1-147519767-A-T | T | | | | | | | | | 0.97914 | 4.61E-08 | FALSE | 4.61E-08 | ?+ | 0.583285 | 1 | 0.024067 | -1 | 1.15E-07 | -1 |
| 1-148017581-AC-A | A | | | | | | | | | 0.000967 | 4.53E-09 | TRUE | 4.53E-09 | ?+ | 0.088701 | 1 | 0.192865 | 1 | 1.74E-08 | 1 |
| 1-148017587-AAT-A | A | | | | | | | | | 0.00089 | 3.67E-08 | TRUE | 3.67E-08 | ?+ | 0.643951 | -1 | 0.365961 | 1 | 7.57E-09 | 1 |
| 1-148066853-G-GCAT | GCAT | | | | | | | | | 0.000441 | 4.13E-10 | TRUE | 4.13E-10 | ?- | 0.007427 | 1 | 0.303552 | 1 | 2.33E-08 | 1 |
| 1-148066859-G-GCA | GCA | | | | | | | | | 0.000315 | 1.35E-08 | TRUE | 1.35E-08 | ?- | 0.000407 | 1 | 0.567598 | 1 | 8.93E-06 | 1 |
| 1-149920006-GCTGG | G | | | | | | | | | 0.006186 | 7.48E-28 | TRUE | 7.48E-28 | ?+ | 0.025911 | 1 | 8.68E-05 | 1 | 4.89E-23 | 1 |
| 1-149945859-C-G | G | | | | | | | 5.32E-05 | 0.626237 | 0.000292 | 3.27E-08 | TRUE | 7.79E-07 | +- | 0.386463 | 1 | 0.622356 | 1 | 1.83E-07 | 1 |
| 1-150304119-TG-T | T | | | | | | | 3.45E-05 | 0.497618 | 0.000907 | 4.53E-10 | TRUE | 2.23E-09 | -+ | 0.21742 | 1 | 0.042446 | 1 | 1.95E-08 | 1 |
| 1-150331239-C-A | A | | | | | | | 0.000482 | 0.037236 | 0.001247 | 3.23E-13 | TRUE | 1.29E-07 | -+ | 0.485092 | 1 | 0.07022 | 1 | 3.07E-12 | 1 |
| 1-150526800-ATT-A | A | | | | | | | | | 0.000764 | 3.68E-12 | TRUE | 3.68E-12 | ?+ | 0.028619 | 1 | 0.060781 | 1 | 1.17E-09 | 1 |
| 1-150541207-A-AAG | AAG | | | | | | | | | 0.00325 | 1.58E-09 | FALSE | 1.58E-09 | ?- | 0.041965 | 1 | 0.002935 | 1 | 1.88E-07 | 1 |
| 1-150671030-AG-A | A | | | | | | | | | 0.002072 | 1.31E-14 | TRUE | 1.31E-14 | ?+ | 0.892725 | -1 | 0.061529 | 1 | 8.05E-16 | 1 |
| 1-150676899-T-C | C | | | | | | | 0.004395 | 0.303001 | 0.002178 | 9.78E-18 | TRUE | 3.71E-08 | -- | 0.000439 | 1 | 0.694796 | 1 | 5.55E-15 | 1 |
| 1-150960054-CA-C | C | | | | | | | | | 0.004188 | 1.28E-16 | TRUE | 1.28E-16 | ?- | 0.07039 | 1 | 0.113542 | 1 | 1.05E-14 | 1 |
| 1-151083907-A-ATTT | ATTT | | | | | | | | | 0.001984 | 6.95E-09 | TRUE | 6.95E-09 | ?- | 0.01327 | 1 | 0.176855 | 1 | 2.66E-08 | 1 |
| 1-152109643-G-GT | GT | | | | | | | | | 0.001553 | 8.29E-09 | TRUE | 8.29E-09 | ?- | 0.013272 | 1 | 0.721217 | 1 | 2.63E-08 | 1 |
| 1-152109644-C-CTT | CTT | | | | | | | | | 0.001947 | 1.88E-10 | TRUE | 1.88E-10 | ?+ | 0.012136 | 1 | 0.649802 | 1 | 5.47E-10 | 1 |
| 1-152218911-A-G | G | 0.009097 | 0.044873 | | | 0.04487 | +? | 0.001046 | 0.386072 | 0.01166 | 4.93E-09 | TRUE | 8.8E-07 | +- | 0.234908 | 1 | 0.50896 | 1 | 3.98E-14 | 1 |
| 1-152911204-A-G | G | 0.28606 | 0.628795 | | | 0.6288 | +? | | | 0.286404 | 1.54E-10 | FALSE | 1.54E-10 | ?- | 0.107914 | -1 | 0.008298 | -1 | 0.000205 | 1 |
| 1-153704580-A-G | G | 0.103964 | 0.117132 | 0.088 | 0.434633 | 0.1264 | -+ | 0.061495 | 0.213027 | 0.02799 | 5.74E-09 | FALSE | 0.1315 | +- | 0.02356 | 1 | 0.791558 | -1 | 5.46E-10 | 1 |
| 1-155274178-ATTAG- | A | | | | | | | | | 0.001409 | 1.81E-13 | TRUE | 1.81E-13 | ?+ | 0.028333 | 1 | 0.043806 | 1 | 1.82E-12 | 1 |
| 1-155298329-T-C | C | | | 0.308 | 0.833165 | 0.8332 | ?+ | | | 0.428239 | 3.44E-12 | FALSE | 3.44E-12 | ?- | 0.050579 | 1 | 0.514978 | 1 | 2.89E-05 | 1 |
| 1-155819544-C-CTA | CTA | | | | | | | | | 0.000932 | 1.25E-10 | TRUE | 1.25E-10 | ?+ | 0.000984 | 1 | 0.121201 | 1 | 1.48E-08 | 1 |
| 1-155819546-AAG-A | A | | | | | | | | | 0.000909 | 1.59E-10 | TRUE | 1.59E-10 | ?+ | 0.000137 | 1 | 0.093291 | 1 | 1.64E-07 | 1 |
| 1-155825397-AG-A | A | 0.001063 | 0.725814 | | | 0.7258 | -? | 0.000698 | 0.990467 | 0.001687 | 1.71E-10 | TRUE | 1.32E-07 | ++ | 0.412568 | 1 | 0.039903 | 1 | 1.02E-09 | 1 |
| 1-155877614-C-CT | CT | | | | | | | 0.000789 | 0.342595 | 0.001705 | 4.13E-19 | TRUE | 3.48E-15 | ++ | 0.207838 | 1 | 0.010407 | 1 | 2.53E-17 | 1 |
| 1-155877639-C-T | T | | | | | | | 4.39E-05 | 0.105511 | 0.000611 | 2.44E-09 | TRUE | 8.87E-08 | -+ | 0.000143 | 1 | 0.179843 | 1 | 1.66E-07 | 1 |
| 1-155878634-GAC-G | G | | | | | | | | | 0.000829 | 5.36E-10 | TRUE | 5.36E-10 | ?+ | 0.004955 | 1 | 0.00049 | 1 | 2.49E-06 | 1 |
| 1-155948467-T-TA | TA | | | | | | | | | 0.001303 | 4.41E-10 | TRUE | 4.41E-10 | ?- | 0.013942 | 1 | 0.02229 | 1 | 1.16E-07 | 1 |
| 1-155955307-T-TAG | TAG | | | | | | | | | 0.000681 | 3.18E-08 | TRUE | 3.18E-08 | ?- | 0.295658 | -1 | 0.834742 | 1 | 2.78E-08 | 1 |
| 1-155963917-GGT-G | G | | | | | | | | | 0.001357 | 1.09E-09 | TRUE | 1.09E-09 | ?+ | 0.368589 | 1 | 0.051359 | 1 | 6.11E-09 | 1 |
| 1-155963921-C-CAG | CAG | | | | | | | | | 0.001265 | 1.78E-09 | TRUE | 1.78E-09 | ?+ | 0.209543 | 1 | 0.020177 | 1 | 5.26E-08 | 1 |
| 1-161348819-CAG-C | C | | | | | | | | | 0.000746 | 9.94E-14 | TRUE | 9.94E-14 | ?- | 0.019572 | 1 | 0.001418 | 1 | 3.85E-11 | 1 |
| 1-161372350-ATC-A | A | | | | | | | | | 0.0015 | 1.53E-14 | TRUE | 1.53E-14 | ?+ | 0.003486 | 1 | 0.914603 | 1 | 1.62E-12 | 1 |
| 1-161415716-A-C | C | | | 0.356 | 0.942066 | 0.9421 | ?- | 0.324741 | 0.882243 | 0.400509 | 9.77E-10 | FALSE | 0.000303 | -- | 0.295075 | 1 | 0.006826 | 1 | 8.52E-07 | 1 |
| 1-168208100-A-G | G | | | | | | | | | 0.498357 | 1E-10 | FALSE | 1E-10 | ?- | 0.038001 | 1 | 0.088363 | 1 | 0.000208 | 1 |
| 1-171621577-G-T | T | | | | | | | 4.39E-05 | 0.790158 | 0.001147 | 1.85E-13 | TRUE | 8.54E-13 | -+ | 0.016216 | 1 | 0.109818 | 1 | 6.27E-10 | 1 |
| 1-171621582-A-G | G | | | | | | | | | 0.000985 | 7.35E-13 | TRUE | 7.35E-13 | ?- | 0.008009 | 1 | 0.003448 | 1 | 6.57E-10 | 1 |
| 1-173835689-CCT-C | C | | | | | | | | | 0.00056 | 3.1E-08 | TRUE | 3.1E-08 | ?- | 0.226244 | 1 | 0.944931 | -1 | 5.01E-07 | 1 |
| 1-175240593-C-CAT | CAT | | | | | | | | | 0.206601 | 1.44E-11 | FALSE | 1.44E-11 | ?+ | 0.383705 | 1 | 0.015137 | 1 | 2.84E-11 | 1 |
| 1-176942866-A-T | T | | | | | | | | | 0.5681 | 4.58E-08 | FALSE | 4.58E-08 | ?- | 0.699958 | 1 | 0.100956 | 1 | 8.53E-08 | 1 |
| 1-178078516-G-T | T | | | | | | | 0.001096 | 0.809812 | 0.000745 | 3.01E-13 | TRUE | 1.62E-06 | ++ | 0.000166 | 1 | 0.086585 | 1 | 1.18E-08 | 1 |
| 1-179070132-G-A | A | 0.000452 | 0.985259 | | | 0.9853 | -? | 0.000219 | 0.688914 | 0.001445 | 3.31E-11 | TRUE | 3.65E-10 | ++ | 0.030771 | 1 | 0.000371 | 1 | 7.23E-09 | 1 |
| 1-179070191-A-ACC | ACC | | | | | | | | | 0.000983 | 2.49E-08 | TRUE | 2.49E-08 | ?- | 0.024865 | 1 | 0.912378 | 1 | 1.75E-07 | 1 |
| 1-179174275-A-AG | AG | | | | | | | | | 0.000726 | 3.76E-09 | TRUE | 3.76E-09 | ?- | 0.00014 | 1 | 0.003006 | 1 | 9.08E-06 | 1 |
| 1-190628841-ATTCTG | A | 0.011906 | 0.068205 | | | 0.06821 | -? | | | 0.044956 | 5.67E-11 | FALSE | 5.67E-11 | ?+ | 0.667366 | 1 | 0.666377 | -1 | 4.45E-12 | 1 |

| | | | | | | | | | | | | | | | | | | | | |
|---|---|---|---|---|---|---|---|---|---|---|---|---|---|---|---|---|---|---|---|---|
| 1-192857217-GGGGT( | G | | | | | | | | | 0.089762 | 1.05E-08 | FALSE | 1.05E-08 | ?- | 0.676993 | 1 | 0.519505 | -1 | 2.66E-09 | -1 |
| 1-192857225-C-CG | CG | | | | | | | | | 0.089461 | 9.74E-09 | FALSE | 9.74E-09 | ?- | 0.411676 | 1 | 0.564431 | -1 | 1.44E-09 | -1 |
| 1-192857228-C-T | T | | | | | | | | | 0.102882 | 1.25E-11 | FALSE | 1.26E-11 | ?- | 0.517714 | 1 | 0.422708 | -1 | 9.72E-13 | -1 |
| 1-197576732-T-C | C | | | | | | | | | 0.000664 | 9.26E-11 | TRUE | 9.26E-11 | ?- | 0.020549 | 1 | 0.00462 | 1 | 5.6E-07 | 1 |
| 1-199555777-TA-T | T | | | | | | | | | 0.008948 | 5.41E-10 | FALSE | 5.41E-10 | ?+ | 0.002571 | 1 | 0.885089 | 1 | 0.000214 | 1 |
| 1-200258342-T-A | A | | | | | | | | | 0.000592 | 3.07E-08 | TRUE | 3.07E-08 | ?+ | 0.381251 | 1 | 0.464923 | 1 | 1.01E-08 | 1 |
| 1-200284721-CTG-C | C | | | | | | | | | 0.000933 | 1.45E-08 | TRUE | 1.45E-08 | ?- | 0.019657 | 1 | 0.007434 | 1 | 1.83E-06 | 1 |
| 1-200345290-C-CGG | CGG | | | | | | | | | 0.000904 | 2.28E-11 | TRUE | 2.28E-11 | ?+ | 0.009228 | 1 | 0.004554 | 1 | 9.59E-10 | 1 |
| 1-200345291-ACC-A | A | | | | | | | | | 0.000839 | 1.51E-10 | TRUE | 1.51E-10 | ?+ | 0.006073 | 1 | 0.007284 | 1 | 5.69E-09 | 1 |
| 1-200386443-C-T | T | | | | | | | 0.001685 | 0.343766 | 0.000664 | 4.77E-08 | TRUE | 0.05078 | -+ | 0.087813 | 1 | 0.011451 | 1 | 7.94E-07 | 1 |
| 1-202740096-GCA-G | G | | | | | | | | | 0.00368 | 1.51E-09 | TRUE | 1.51E-09 | ?+ | 0.907409 | 1 | 0.025272 | 1 | 2.23E-09 | 1 |
| 1-203503088-A-T | T | | | | | | | | | 0.000696 | 1.11E-09 | TRUE | 1.11E-09 | ?- | 0.015946 | 1 | 0.204397 | 1 | 3.4E-08 | 1 |
| 1-203921617-A-AGG | AGG | | | | | | | | | 0.000258 | 1.36E-08 | TRUE | 1.36E-08 | ?- | 0.07336 | 1 | 0.586592 | 1 | 2.56E-08 | 1 |
| 1-210976986-C-G | G | 0.001826 | 0.581026 | | | 0.581 | +? | | | 0.000619 | 4.53E-08 | TRUE | 4.53E-08 | ?- | 0.348308 | 1 | 0.040642 | 1 | 4.7E-08 | 1 |
| 1-214897967-G-A | A | | | | | | | | | 0.001107 | 3.77E-15 | TRUE | 3.77E-15 | ?+ | 0.006309 | 1 | 0.9573 | -1 | 4.28E-14 | 1 |
| 1-218455409-G-C | C | 9.76E-05 | 0.981578 | | | 0.9816 | -? | 0.000179 | 0.209681 | 0.000744 | 2.23E-09 | TRUE | 1.69E-06 | -+ | 0.241874 | 1 | 0.000453 | 1 | 3.51E-06 | 1 |
| 1-218455927-G-A | A | 0.001616 | 0.860515 | | | 0.8605 | +? | 0.000119 | 0.163756 | 0.002061 | 2.23E-10 | TRUE | 8.97E-11 | ++ | 0.005918 | 1 | 1.88E-05 | 1 | 8.35E-05 | 1 |
| 1-218561848-G-C | C | 0.00098 | 0.676435 | | | 0.6764 | +? | 0.001322 | 0.372117 | 0.004746 | 2.65E-32 | TRUE | 2.01E-22 | -+ | 0.002124 | 1 | 0.000643 | 1 | 1.14E-27 | 1 |
| 1-220048846-C-CAA | CAA | | | | | | | | | 0.000639 | 1.5E-10 | TRUE | 1.5E-10 | ?+ | 0.246191 | 1 | 0.005118 | 1 | 1.74E-08 | 1 |
| 1-220048851-T-A | A | | | | | | | | | 0.00036 | 7.16E-09 | TRUE | 7.16E-09 | ?+ | 0.080595 | 1 | 0.410477 | 1 | 9.47E-08 | 1 |
| 1-220048860-C-T | T | | | | | | | 5.01E-05 | 0.6469 | 0.000446 | 6.14E-09 | TRUE | 1.39E-08 | ++ | 0.253285 | 1 | 0.326591 | 1 | 4.34E-09 | 1 |
| 1-220140384-TGA-T | T | 0.003535 | 0.161887 | | | 0.1619 | -? | | | 0.00349 | 2.69E-08 | TRUE | 2.69E-08 | ?+ | 0.035321 | 1 | 0.159103 | 1 | 3.38E-07 | 1 |
| 1-223118344-G-T | T | | | | | | | 0.000345 | 0.372023 | 0.000969 | 9.8E-11 | TRUE | 3.99E-07 | -+ | 0.014776 | 1 | 0.000582 | 1 | 4.61E-07 | 1 |
| 1-223118351-G-A | A | 9.76E-05 | 0.242815 | | | 0.2428 | +? | 0.00037 | 0.256513 | 0.000628 | 1.03E-08 | TRUE | 0.000114 | -+ | 0.027389 | 1 | 0.001729 | 1 | 1.64E-05 | 1 |
| 1-223119009-C-A | A | 0.000176 | 0.636964 | | | 0.637 | +? | 0.001128 | 0.970535 | 0.001771 | 2.89E-14 | TRUE | 7.22E-09 | ++ | 0.000295 | 1 | 0.692083 | -1 | 2.62E-12 | 1 |
| 1-224206818-C-CCT | CCT | | | | | | | | | 0.000727 | 1.35E-08 | TRUE | 1.35E-08 | ?+ | 0.507671 | 1 | 0.207057 | 1 | 1.49E-08 | 1 |
| 1-226142102-C-CCT | CCT | | | | | | | | | 0.001268 | 4.08E-16 | TRUE | 4.08E-16 | ?+ | 0.400953 | 1 | 0.001728 | 1 | 6.96E-14 | 1 |
| 1-226330686-C-A | A | 0.000156 | 0.633551 | | | 0.6336 | -? | 0.000495 | 0.019132 | 0.000907 | 3.52E-17 | TRUE | 2.02E-09 | -+ | 0.001629 | 1 | 0.003583 | 1 | 2.09E-13 | 1 |
| 1-228892872-T-C | C | | | | | | | | | 0.001086 | 3.29E-08 | TRUE | 3.29E-08 | ?- | 0.244649 | 1 | 0.455286 | 1 | 6.06E-08 | 1 |
| 1-239732335-GT-G | G | | | | | | | | | 0.882318 | 9.43E-11 | FALSE | 9.43E-11 | ?- | 0.648493 | -1 | 0.647642 | -1 | 7.71E-14 | -1 |
| 1-241063951-AGGTG- | A | | | | | | | | | 0.000547 | 1.18E-11 | TRUE | 1.18E-11 | ?+ | 0.042065 | 1 | 0.292487 | 1 | 5.79E-10 | 1 |
| 1-246376445-G-T | T | | | | | | | | | 0.000629 | 5.79E-09 | TRUE | 5.79E-09 | ?+ | 0.89202 | -1 | 0.003771 | 1 | 2.19E-07 | 1 |
| 1-246387888-CTCCTT( | C | | | | | | | | | 0.117022 | 1.22E-08 | FALSE | 1.22E-08 | ?- | 0.120287 | 1 | 0.564996 | 1 | 2.54E-08 | 1 |
| 1-246940554-GCA-G | G | 0.021943 | 0.485563 | | | 0.4856 | -? | | | 0.030764 | 1.76E-11 | FALSE | 1.76E-11 | ?+ | 0.640605 | 1 | 0.181909 | 1 | 3.75E-20 | 1 |
| 1-246940557-T-TGG | TGG | | | | | | | | | 0.031822 | 3.31E-14 | FALSE | 3.31E-14 | ?- | 0.323439 | 1 | 0.245708 | 1 | 1.56E-20 | 1 |
| 1-78398388-C-T | T | | | | | | | 1.88E-05 | 0.374818 | 0.000112 | 0.978477 | FALSE | 2.8E-305 | +- | 0.833051 | 1 | | | | |
| 1-85498204-G-A | A | | | | | | | 9.4E-06 | 0.767059 | | | FALSE | 1E-305 | -? | | | | | | |
| 1-59405581-G-A | A | | | | | | | 9.4E-06 | 0.391446 | | | FALSE | 3.9E-305 | -? | | | | | | |
| 2-74159003-A-G | G | 0.000292 | 2.1E-08 | | | 2.102E-08 | -? | 9.71E-05 | 0.065013 | 0.000167 | 0.272479 | TRUE | 0.5875 | -+ | | | | | | |
| 2-102178470-C-G | G | 0.065613 | 2.98E-08 | 0.008 | 0.396276 | 2.777E-08 | ++ | 0.000379 | 0.461009 | 0.039561 | 0.914537 | FALSE | 0.9625 | +- | 0.733703 | 1 | 0.117875 | -1 | 0.589693 | 1 |
| 2-127090354-G-A | A | 0.244777 | 1.47E-09 | 0.361 | 0.251863 | 9.432E-10 | ++ | 0.346829 | 4.01E-05 | 0.253745 | 0.520575 | TRUE | 0.000167 | ++ | 0.31289 | 1 | 0.969025 | -1 | 0.478762 | 1 |
| 2-127124439-T-A | A | 0.117146 | 1.57E-08 | 0.175 | 0.687451 | 1.444E-08 | ++ | 0.171975 | 0.026462 | 0.123486 | 0.039566 | TRUE | 0.002833 | ++ | 0.794535 | 1 | 0.448554 | 1 | 0.059507 | 1 |
| 2-127130409-C-A | A | 0.120922 | 4.21E-08 | 0.169 | 0.811491 | 4.111E-08 | ++ | 0.157823 | 0.002916 | 0.128036 | 0.035713 | TRUE | 0.000271 | ++ | 0.20156 | 1 | 0.337946 | 1 | 0.128956 | 1 |
| 2-127131181-A-C | C | 0.266082 | 4.34E-10 | 0.369 | 0.2664 | 2.797E-10 | -- | 0.333538 | 8.96E-07 | 0.280489 | 0.453545 | TRUE | 7.87E-06 | -- | 0.212911 | 1 | 0.758028 | 1 | 0.619751 | 1 |
| 2-127131542-C-T | T | 0.106747 | 3.07E-08 | 0.164 | 0.897678 | 3.124E-08 | ++ | | | 0.11192 | 0.04924 | TRUE | 0.04924 | ?+ | 0.20512 | 1 | 0.34204 | 1 | 0.128901 | 1 |
| 2-127133851-A-C | C | 0.478981 | 1.77E-10 | 0.468 | 0.077272 | 8.114E-11 | -- | 0.420562 | 2.51E-10 | 0.459002 | 0.01738 | TRUE | 6.35E-11 | -- | 0.205212 | 1 | 0.442321 | 1 | 0.086425 | 1 |
| 2-127135234-C-T | T | 0.416654 | 1.19E-10 | 0.434 | 0.039481 | 4.673E-11 | ++ | 0.390226 | 4.4E-12 | 0.39655 | 0.033562 | TRUE | 6.14E-12 | ++ | 0.323925 | 1 | 0.356582 | 1 | 0.176635 | 1 |
| 2-127128582-C-T | T | 0.299957 | 1.94E-05 | 0.311 | 0.112468 | 0.00001185 | ++ | 0.299872 | 4.4E-08 | 0.28398 | 0.024032 | TRUE | 7.46E-09 | ++ | 0.409844 | -1 | 0.089187 | 1 | 0.080175 | 1 |
| 2-127128657-C-T | T | 0.299673 | 2.04E-05 | 0.311 | 0.100686 | 0.00001229 | ++ | 0.299709 | 4.17E-08 | 0.283029 | 0.019492 | TRUE | 5.42E-09 | ++ | 0.387981 | -1 | 0.084132 | 1 | 0.081231 | 1 |
| 2-1113154-C-CTTTTGT | CTTTTGTACTAA | | | | | | | | | 0.720336 | 5.77E-52 | FALSE | 5.77E-52 | ?- | 1.93E-05 | -1 | 0.001372 | -1 | 1.08E-44 | -1 |
| 2-1113158-G-GGCTTA | GGCTTAACCCTTATAGTCCTTTGAGGAGAATCATGT | | | | | | | | | 0.733192 | 4.95E-56 | FALSE | 4.95E-56 | ?+ | 2.37E-06 | -1 | 0.00685 | -1 | 2.55E-49 | -1 |
| 2-1113158-G-T | T | | | | | | | | | 0.243449 | 3.73E-50 | FALSE | 3.74E-50 | ?+ | 2.18E-06 | 1 | 0.012827 | 1 | 4.27E-44 | 1 |
| 2-1597315-C-CCA | CCA | | | | | | | | | 0.691428 | 2.1E-08 | FALSE | 2.1E-08 | ?+ | 0.331529 | 1 | 0.220908 | 1 | 1.18E-05 | 1 |
| 2-3140897-C-G | G | | | | | | | | | 0.902822 | 2.14E-10 | FALSE | 2.14E-10 | ?+ | 0.527858 | -1 | 0.377937 | -1 | 2.83E-12 | -1 |
| 2-3181093-C-A | A | | | | | | | | | 0.135575 | 8.95E-26 | FALSE | 8.95E-26 | ?+ | 0.225367 | 1 | 0.037961 | 1 | 1.87E-25 | 1 |
| 2-3181093-C-T | T | | | | | | | | | 0.690049 | 1.77E-24 | FALSE | 1.77E-24 | ?- | 0.154331 | -1 | 0.140549 | -1 | 2.82E-23 | -1 |
| 2-3218610-A-G | G | | | | | | | | | 0.947881 | 1.02E-15 | FALSE | 1.02E-15 | ?+ | 0.31648 | -1 | 0.748673 | -1 | 2.86E-19 | -1 |
| 2-3220957-T-C | C | | | | | | | | | 0.873669 | 2.08E-09 | FALSE | 2.08E-09 | ?- | 0.076047 | 1 | 0.004749 | 1 | 9.06E-07 | 1 |
| 2-3241125-G-C | C | | | | | | | | | 0.863277 | 2.52E-22 | FALSE | 2.52E-22 | ?- | 0.018128 | -1 | 0.031846 | -1 | 2.75E-19 | -1 |

| | | | | | | | | | | | | | | | | | | | | |
|---|---|---|---|---|---|---|---|---|---|---|---|---|---|---|---|---|---|---|---|---|
| 2-3241125-G-GC | GC | | | | | | | | | 0.058534 | 1.4E-36 | FALSE | 1.4E-36 | ?- | 0.001654 | 1 | 6.11E-05 | 1 | 1.05E-27 | 1 |
| 2-6420447-CATGAGG | C | | | | | | | | | 0.005244 | 1.36E-08 | TRUE | 1.36E-08 | ?- | 0.742263 | 1 | 0.512618 | 1 | 2.68E-09 | 1 |
| 2-9503812-TG-T | T | | | | | | | | | 0.000432 | 4.69E-10 | TRUE | 4.69E-10 | ?+ | 0.003852 | 1 | 0.014466 | 1 | 3.26E-08 | 1 |
| 2-11052875-T-TCG | TCG | 9.83E-05 | 0.989994 | | | 0.99 | +? | | | 0.000935 | 1.68E-08 | TRUE | 1.68E-08 | ?- | 0.077007 | 1 | 0.623422 | 1 | 7.28E-08 | 1 |
| 2-11052877-GCA-G | G | | | | | | | | | 0.000812 | 2E-08 | TRUE | 2E-08 | ?+ | 0.170114 | 1 | 0.967565 | 1 | 2.86E-08 | 1 |
| 2-16395352-G-A | A | | | | | | | | | 0.543511 | 1.82E-31 | FALSE | 1.82E-31 | ?+ | 0.735457 | 1 | 0.322751 | 1 | 4.62E-34 | 1 |
| 2-16395352-G-GCACA | GCACACACCATAGATCCTCACACACCATAGAGACACACACACA | | | | | | | | | 0.284789 | 2.06E-53 | FALSE | 2.06E-53 | ?+ | 0.345668 | -1 | 0.098261 | -1 | 5.44E-54 | -1 |
| 2-24152050-G-T | T | | | | | | | | | 0.000133 | 4.32E-08 | TRUE | 4.32E-08 | ?+ | | | | | 5.88E-06 | 1 |
| 2-25534570-CT-C | C | | | | | | | | | 0.003511 | 7.17E-11 | TRUE | 7.17E-11 | ?- | 0.061306 | 1 | 0.006346 | 1 | 9.18E-08 | 1 |
| 2-26010180-T-A | A | | | | | | | 9.71E-05 | 0.752837 | 0.000757 | 4.56E-08 | TRUE | 5.09E-07 | -+ | 0.01689 | 1 | 0.133654 | 1 | 2.67E-06 | 1 |
| 2-27126918-A-G | G | 0.016075 | 0.772173 | | | 0.7722 | +? | 0.993162 | 0.251618 | 0.977023 | 4.73E-19 | TRUE | 2.23E-14 | ++ | 0.147322 | -1 | 0.015244 | -1 | 1.88E-21 | -1 |
| 2-27141043-T-A | A | | | | | | | | | 0.000273 | 2.48E-08 | TRUE | 2.48E-08 | ?+ | 0.100182 | 1 | | | 1.85E-07 | 1 |
| 2-27273508-GAT-G | G | 0.011638 | 0.036199 | | | 0.0362 | -? | | | 0.005787 | 2.09E-11 | TRUE | 2.09E-11 | ?+ | 0.297986 | 1 | 0.418802 | -1 | 5.68E-13 | 1 |
| 2-27511478-CCT-C | C | | | | | | | | | 0.000994 | 1.47E-10 | TRUE | 1.47E-10 | ?- | 0.006363 | 1 | 0.030811 | 1 | 3.62E-09 | 1 |
| 2-33306351-CTT-C | C | 0.000117 | 0.706064 | | | 0.7061 | -? | | | 0.000927 | 3.71E-09 | TRUE | 3.71E-09 | ?- | 0.150489 | 1 | 0.158728 | 1 | 1.64E-08 | 1 |
| 2-34321033-CA-C | C | 0.000721 | 0.708755 | | | 0.7088 | -? | | | 0.003578 | 1.08E-10 | TRUE | 1.08E-10 | ?- | 0.976544 | 1 | 0.0089 | 1 | 5.23E-09 | 1 |
| 2-38696978-G-T | T | | | | | | | 3.76E-05 | 0.867679 | 0.000289 | 2.54E-09 | TRUE | 1.06E-08 | ++ | 0.002163 | 1 | 0.061245 | 1 | 5.73E-08 | 1 |
| 2-39089972-CAAAAAA | C | | | | | | | | | 0.000669 | 3.23E-11 | TRUE | 3.23E-11 | ?- | 0.012234 | 1 | 0.001085 | 1 | 1.02E-07 | 1 |
| 2-39126426-AT-A | A | | | | | | | | | 0.001659 | 7.9E-11 | TRUE | 7.91E-11 | ?+ | 0.208599 | 1 | 0.040927 | 1 | 9.29E-09 | 1 |
| 2-42606050-C-CGGCT | CGGCTGGCCGGGTGGGGGGCTGACCCCCCCTCTCCCTCCCGGACGGGGT | | | | | | | | | 0.468659 | 9.69E-20 | FALSE | 9.69E-20 | ?+ | 0.002297 | 1 | 0.680834 | 1 | 2.4E-18 | 1 |
| 2-42606050-C-T | T | | | | | | | | | 0.264857 | 1.21E-13 | FALSE | 1.21E-13 | ?- | 0.008113 | -1 | 0.593863 | -1 | 9.44E-13 | -1 |
| 2-46273971-C-CAAAG | CAAAGAAAGA | | | | | | | | | 0.00072 | 1.98E-09 | TRUE | 1.98E-09 | ?+ | 0.959086 | -1 | 0.009398 | 1 | 1.05E-08 | 1 |
| 2-47353179-AT-A | A | | | | | | | | | 0.001044 | 1.06E-10 | TRUE | 1.06E-10 | ?+ | 0.018627 | 1 | 0.0077 | 1 | 8.3E-08 | 1 |
| 2-47768888-T-TGG | TGG | | | | | | | | | 0.001377 | 2.16E-09 | TRUE | 2.16E-09 | ?- | 0.002516 | 1 | 0.010475 | 1 | 2.98E-06 | 1 |
| 2-52905737-G-A | A | 0.000137 | 0.932279 | | | 0.9323 | -? | 0.000486 | 0.742768 | 0.000856 | 2.9E-11 | TRUE | 3.81E-07 | -+ | 0.066572 | 1 | 0.905337 | 1 | 4.76E-11 | 1 |
| 2-53970819-CGAA-C | C | | | | | | | | | 0.00645 | 2.31E-12 | TRUE | 2.31E-12 | ?- | 0.037608 | 1 | 0.089576 | 1 | 4.46E-11 | 1 |
| 2-53978264-A-ACTT | ACTT | | | | | | | | | 0.000554 | 1.16E-08 | TRUE | 1.16E-08 | ?- | 0.000543 | 1 | 0.027075 | 1 | 0.000332 | 1 |
| 2-54004201-G-T | T | | | | | | | | | 0.000344 | 4.07E-08 | TRUE | 4.07E-08 | ?+ | 0.391092 | 1 | 0.001612 | 1 | 2.81E-05 | 1 |
| 2-54095998-C-CG | CG | | | | | | | 0.99694 | 0.681293 | 0.913185 | 6.22E-15 | FALSE | 2.08E-13 | -+ | 0.778773 | 1 | 0.589308 | -1 | 3.13E-17 | 1 |
| 2-54486169-G-A | A | | | | | | | 0.496939 | 0.256764 | 0.62948 | 2.92E-12 | FALSE | 0.008587 | -+ | 0.03573 | 1 | 0.324938 | 1 | 6E-09 | 1 |
| 2-61148337-C-CAA | CAA | | | | | | | | | 0.000328 | 2.49E-08 | TRUE | 2.49E-08 | ?+ | 0.016087 | 1 | 0.019982 | 1 | 7.12E-06 | 1 |
| 2-61153373-T-C | C | 0.385048 | 0.041525 | 0.407 | 0.062059 | 0.02948 | -- | 0.401502 | 0.13846 | 0.440206 | 2.95E-10 | FALSE | 0.01944 | +- | 0.061775 | 1 | 0.377993 | 1 | 1.24E-09 | 1 |
| 2-62167572-ATT-A | A | 0.000234 | 0.175657 | | | 0.1757 | +? | | | 0.001794 | 8.19E-12 | TRUE | 8.19E-12 | ?+ | 0.284851 | 1 | 0.184109 | 1 | 1.04E-11 | 1 |
| 2-65157396-T-TA | TA | 0.000409 | 0.299023 | | | 0.299 | -? | | | 0.000815 | 6.16E-10 | TRUE | 6.16E-10 | ?- | 0.02463 | 1 | 0.300435 | 1 | 4.78E-09 | 1 |
| 2-65157939-A-T | T | 0.000295 | 0.922667 | | | 0.9227 | +? | 0.005243 | 0.268906 | 0.003151 | 1.97E-13 | TRUE | 0.001583 | +- | 0.306167 | 1 | 0.870525 | 1 | 5.62E-15 | 1 |
| 2-65168546-C-CCT | CCT | 0.004132 | 0.074886 | | | 0.07489 | -? | | | 0.004198 | 4.06E-10 | TRUE | 4.06E-10 | ?+ | 0.003347 | 1 | 0.051612 | 1 | 4.07E-07 | 1 |
| 2-65280182-A-ACC | ACC | | | | | | | | | 0.001411 | 9E-10 | TRUE | 9E-10 | ?- | 0.017296 | 1 | 0.02041 | 1 | 7.93E-08 | 1 |
| 2-65446107-C-A | A | | | | | | | | | 0.001441 | 2.54E-21 | TRUE | 2.54E-21 | ?+ | 0.005444 | 1 | 0.014023 | 1 | 2.72E-18 | 1 |
| 2-65446110-C-G | G | | | | | | | | | 0.001215 | 5.69E-16 | TRUE | 5.69E-16 | ?- | 0.037379 | 1 | 0.049452 | 1 | 7.49E-14 | 1 |
| 2-70188373-T-A | A | 0.000474 | 0.936254 | | | 0.9363 | +? | 0.000633 | 0.846904 | 0.001597 | 1.11E-14 | TRUE | 9.12E-11 | ++ | 0.001374 | 1 | 0.862554 | 1 | 7.8E-14 | 1 |
| 2-79011165-CT-C | C | | | | | | | | | 0.001459 | 2.39E-12 | TRUE | 2.39E-12 | ?- | 0.000319 | 1 | 0.01918 | 1 | 5.05E-09 | 1 |
| 2-79818457-C-CAT | CAT | | | | | | | | | 0.000842 | 1.88E-10 | TRUE | 1.88E-10 | ?+ | 0.000214 | 1 | 0.698038 | -1 | 4.3E-08 | 1 |
| 2-85532753-C-CACCG | CACCG | | | | | | | | | 0.001925 | 4.04E-13 | TRUE | 4.04E-13 | ?+ | 0.001521 | 1 | 0.018337 | 1 | 9.62E-10 | 1 |
| 2-86191352-A-ATT | ATT | | | | | | | | | 0.000922 | 1.12E-09 | TRUE | 1.12E-09 | ?- | 0.426917 | 1 | 0.086475 | 1 | 3.56E-09 | 1 |
| 2-86191352-A-ATTAAT | ATTAAT | | | | | | | | | 0.00063 | 8.72E-09 | TRUE | 8.73E-09 | ?- | 0.078211 | 1 | 0.004381 | 1 | 1.16E-06 | 1 |
| 2-87407432-C-CG | CG | | | | | | | | | 0.990219 | 2.53E-10 | FALSE | 2.53E-10 | ?- | 0.772236 | -1 | 0.995996 | 1 | 4E-11 | -1 |
| 2-87743577-CAAAA-C | C | | | | | | | | | 0.3185 | 1.66E-08 | FALSE | 1.66E-08 | ?- | 0.010215 | 1 | 0.335003 | -1 | 8.45E-07 | 1 |
| 2-87798773-G-C | C | | | | | | | 0.00331 | 0.165882 | 0.001108 | 1.23E-09 | TRUE | 0.08589 | -+ | 0.76516 | -1 | 0.114249 | 1 | 6.26E-10 | 1 |
| 2-87957236-T-TGG | TGG | | | | | | | | | 0.731438 | 1.27E-09 | FALSE | 1.27E-09 | ?+ | 0.656414 | 1 | 0.108824 | -1 | 1.07E-09 | -1 |
| 2-88005024-A-G | G | 0.000196 | 0.539168 | | | 0.5392 | +? | 3.45E-05 | 0.714893 | 0.001002 | 2.18E-12 | TRUE | 3.08E-12 | -- | 0.064472 | 1 | 1.22E-05 | 1 | 4.31E-09 | 1 |
| 2-88005034-A-T | T | | | | | | | | | 0.000932 | 3.7E-12 | TRUE | 3.7E-12 | ?- | 0.011703 | 1 | 0.012215 | 1 | 3E-09 | 1 |
| 2-88005040-C-T | T | 0.000157 | 0.396865 | | | 0.3969 | +? | | | 0.000706 | 3.1E-16 | TRUE | 3.1E-16 | ?+ | 0.001273 | 1 | 0.00271 | 1 | 1.22E-12 | 1 |
| 2-88005607-CA-C | C | 7.8E-05 | 0.404625 | | | 0.4046 | +? | | | 0.000475 | 7.6E-09 | TRUE | 7.6E-09 | ?- | 0.939459 | 1 | 0.167304 | 1 | 1.5E-09 | 1 |
| 2-89636593-G-GA | GA | | | | | | | | | 0.453434 | 3.98E-09 | TRUE | 3.98E-09 | ?+ | 0.135602 | -1 | 0.835748 | 1 | 4.93E-10 | -1 |
| 2-89636593-GAAAA-G | G | | | | | | | | | 0.331274 | 2.66E-27 | TRUE | 2.66E-27 | ?+ | 0.002724 | 1 | 0.121074 | 1 | 3.8E-24 | 1 |
| 2-90331433-C-G | G | | | | | | | | | 0.002022 | 2.82E-09 | TRUE | 2.82E-09 | ?- | 0.006935 | 1 | 0.470412 | -1 | 3.37E-09 | 1 |
| 2-90362218-ATTTTTTT | A | | | | | | | | | 0.066933 | 2.77E-26 | FALSE | 2.77E-26 | ?- | 0.032001 | -1 | 0.039424 | -1 | 1.5E-23 | -1 |
| 2-90385048-G-A | A | | | | | | | | | 0.734418 | 5.67E-33 | FALSE | 5.68E-33 | ?- | 0.166063 | -1 | 0.031562 | -1 | 1.9E-32 | -1 |
| 2-90385048-G-T | T | | | | | | | | | 0.129467 | 3.01E-47 | FALSE | 3.02E-47 | ?+ | 0.000158 | 1 | 8.42E-06 | 1 | 6.73E-38 | 1 |

| | | | | | | | | | | | | | | | | | | | | |
|---|---|---|---|---|---|---|---|---|---|---|---|---|---|---|---|---|---|---|---|---|
| 2-90392259-G-C | C | | | | | | | | | 0.956186 | 4.35E-29 | FALSE | 4.35E-29 | ?- | 0.000797 | -1 | 0.001721 | -1 | 2.2E-23 | -1 |
| 2-90392259-G-T | T | | | | | | | | | 0.043075 | 1.88E-28 | FALSE | 1.88E-28 | ?+ | 0.001984 | 1 | 0.002066 | 1 | 3.32E-23 | 1 |
| 2-90396303-T-C | C | | | | | | | | | 0.99456 | 4.07E-11 | TRUE | 4.07E-11 | ?- | 0.000124 | 1 | 0.02313 | 1 | 5.13E-07 | 1 |
| 2-94286195-G-A | A | | | | | | | | | 0.788496 | 1.99E-12 | FALSE | 1.99E-12 | ?- | 0.711961 | -1 | 0.234304 | -1 | 8.58E-12 | -1 |
| 2-94499647-C-A | A | 0.000157 | 0.425499 | | | 0.4255 | +? | 0.00041 | 0.40263 | 0.000815 | 1.67E-09 | TRUE | 7.75E-08 | ++ | 0.253142 | 1 | 0.000225 | 1 | 1.85E-07 | 1 |
| 2-97063436-A-G | G | | | | | | | | | 0.521171 | 1.05E-14 | FALSE | 1.05E-14 | ?- | 0.893215 | -1 | 0.001354 | 1 | 8.86E-15 | 1 |
| 2-97068767-A-AT | AT | | | | | | | 0.727709 | 0.534541 | 0.536404 | 1.64E-11 | FALSE | 0.000498 | -+ | 0.550202 | -1 | 0.228231 | 1 | 3.19E-13 | -1 |
| 2-99650376-T-G | G | | | | | | | 4.7E-05 | 0.59274 | 0.000703 | 1.9E-14 | TRUE | 4.03E-14 | -- | 0.004709 | 1 | 0.012854 | 1 | 4.56E-11 | 1 |
| 2-101207215-ATT-A | A | | | | | | | | | 0.001142 | 6.71E-12 | TRUE | 6.71E-12 | ?+ | 0.001696 | 1 | 0.003624 | 1 | 1.99E-08 | 1 |
| 2-101578677-A-T | T | | | | | | | | | 0.001113 | 3.71E-08 | TRUE | 3.71E-08 | ?- | 0.006986 | 1 | 0.016705 | 1 | 2.6E-06 | 1 |
| 2-106888365-A-T | T | | | | | | | 0.00052 | 0.910539 | 0.000914 | 7.3E-11 | TRUE | 2.15E-07 | -- | 0.000992 | 1 | 0.057942 | 1 | 3.22E-08 | 1 |
| 2-106888375-T-G | G | | | | | | | | | 0.000486 | 2.52E-08 | TRUE | 2.52E-08 | ?- | 0.000265 | 1 | 0.010739 | 1 | 5.52E-05 | 1 |
| 2-109658261-G-C | C | | | | | | | 0.002387 | 0.330235 | 0.00155 | 1.84E-12 | TRUE | 6.43E-07 | ++ | 0.001524 | 1 | 0.004188 | 1 | 1.25E-08 | 1 |
| 2-111295877-ATTTTTT | A | | | | | | | | | 0.0791 | 1.17E-12 | TRUE | 1.17E-12 | ?- | 0.333747 | -1 | 0.782229 | -1 | 1.15E-14 | -1 |
| 2-111880255-C-T | T | | | | | | | 0.000113 | 0.006346 | 0.000959 | 3.03E-09 | TRUE | 6.68E-11 | ++ | 9.58E-05 | 1 | 0.41306 | 1 | 1.35E-07 | 1 |
| 2-113864927-TA-T | T | | | | | | | | | 0.002304 | 4.84E-13 | TRUE | 4.84E-13 | ?+ | 0.102912 | 1 | 0.008987 | 1 | 5.64E-11 | 1 |
| 2-117077346-AT-A | A | | | | | | | | | 0.037344 | 5.94E-10 | FALSE | 5.94E-10 | ?+ | 0.000283 | 1 | 0.855917 | 1 | 2.25E-07 | 1 |
| 2-117077348-C-CCTAT | CCTATT | | | | | | | | | 0.301913 | 4.84E-08 | FALSE | 4.84E-08 | ?- | 0.096906 | -1 | 0.749647 | 1 | 3.62E-08 | -1 |
| 2-117077349-CTAT-C | C | | | | | | | | | 0.034476 | 1.72E-09 | FALSE | 1.72E-09 | ?- | 0.001081 | 1 | 0.941136 | 1 | 1.48E-07 | 1 |
| 2-117077919-C-CCA | CCA | | | | | | | | | 0.066458 | 1.44E-12 | FALSE | 1.44E-12 | ?- | 0.013301 | -1 | 0.690092 | -1 | 5.34E-12 | -1 |
| 2-118093497-AGATGA | A | | | | | | | | | 0.328279 | 2.98E-14 | TRUE | 2.98E-14 | ?- | 0.321686 | -1 | 0.106298 | -1 | 2.85E-13 | -1 |
| 2-120197492-A-ACT | ACT | 0.000331 | 0.151033 | | | 0.151 | +? | | | 0.003486 | 1.02E-18 | TRUE | 1.02E-18 | ?- | 0.000375 | 1 | 0.084326 | 1 | 2.11E-13 | 1 |
| 2-123234486-GTATA-( | G | | | | | | | | | 0.106162 | 1.44E-09 | FALSE | 1.44E-09 | ?+ | 0.046671 | 1 | 0.486254 | 1 | 1.48E-08 | 1 |
| 2-126871297-A-G | G | | | | | | | 0.000244 | 0.539144 | 0.001066 | 2.48E-15 | TRUE | 5.94E-12 | +- | 1.01E-05 | 1 | 0.394764 | 1 | 1.01E-11 | 1 |
| 2-126871303-G-T | T | | | | | | | 0.000247 | 0.770985 | 0.001068 | 9.19E-10 | TRUE | 9.84E-08 | -+ | 0.005649 | 1 | 0.1307 | 1 | 3.46E-08 | 1 |
| 2-127858638-C-CTG | CTG | 0.000878 | 0.250372 | | | 0.2504 | +? | | | 0.002637 | 2.36E-15 | TRUE | 2.36E-15 | ?+ | 0.396822 | 1 | 0.088418 | 1 | 7.8E-14 | 1 |
| 2-127858655-G-C | C | 0.000375 | 0.105478 | | | 0.1055 | +? | | | 0.000892 | 8.83E-11 | TRUE | 8.83E-11 | ?+ | 0.16044 | 1 | 0.010647 | 1 | 2.4E-08 | 1 |
| 2-128059121-C-CTT | CTT | | | | | | | | | 0.001829 | 1.65E-12 | TRUE | 1.65E-12 | ?+ | 0.045828 | 1 | 0.07 | 1 | 3.13E-11 | 1 |
| 2-128082350-GCACA- | G | | | | | | | | | 0.002087 | 7.5E-09 | TRUE | 7.5E-09 | ?+ | 0.343488 | 1 | 0.137874 | 1 | 9.95E-09 | 1 |
| 2-130469139-C-CA | CA | | | | | | | | | 0.030043 | 2.28E-11 | TRUE | 2.28E-11 | ?+ | 0.012161 | 1 | 0.010096 | 1 | 1.9E-07 | 1 |
| 2-130863966-C-CTA | CTA | | | | | | | | | 0.002147 | 7.07E-17 | TRUE | 7.08E-17 | ?+ | 0.56583 | 1 | 0.366536 | 1 | 3.06E-18 | 1 |
| 2-131020549-T-C | C | | | | | | | | | 0.804462 | 2.05E-18 | FALSE | 2.05E-18 | ?- | 0.072909 | 1 | 0.700284 | 1 | 6.8E-21 | 1 |
| 2-131634219-C-G | G | 0.000117 | 0.516526 | | | 0.5165 | -? | 0.005315 | 0.690714 | 0.001496 | 5.03E-09 | TRUE | 0.03352 | +- | 0.043253 | 1 | 0.01305 | 1 | 3.38E-06 | 1 |
| 2-131634249-T-C | C | | | | | | | 0.003749 | 0.446437 | 0.000662 | 2.73E-08 | TRUE | 0.006057 | -- | 0.013221 | 1 | 0.899302 | -1 | 5.08E-08 | 1 |
| 2-131634722-C-G | G | | | | | | | 0.000188 | 0.167768 | 0.001088 | 1.5E-10 | TRUE | 1.6E-10 | -- | 0.010237 | 1 | 0.007431 | 1 | 2.62E-07 | 1 |
| 2-131636079-C-A | A | | | | | | | 0.00021 | 0.431759 | 0.001247 | 4.03E-08 | TRUE | 1.11E-07 | ++ | 0.013317 | 1 | 0.3187 | 1 | 8.42E-08 | 1 |
| 2-131756355-A-G | G | | | | | | | | | 0.004142 | 9.49E-09 | TRUE | 9.49E-09 | ?- | 0.965042 | 1 | 0.003744 | 1 | 1.39E-08 | 1 |
| 2-132075422-A-T | T | | | | | | | 0.000652 | 0.477468 | 0.000671 | 3.15E-11 | TRUE | 1.69E-07 | -- | 0.020136 | 1 | 0.020286 | 1 | 9.95E-10 | 1 |
| 2-133308914-G-T | T | | | | | | | 0.000539 | 0.527445 | 0.001462 | 6.25E-12 | TRUE | 1.42E-09 | ++ | 0.002465 | 1 | 0.835901 | -1 | 3.86E-10 | 1 |
| 2-133308921-A-G | G | | | | | | | 0.000529 | 0.609481 | 0.00115 | 1.04E-11 | TRUE | 8.12E-09 | -- | 0.014988 | 1 | 0.529367 | -1 | 6.69E-11 | 1 |
| 2-133308945-C-T | T | | | | | | | 0.001099 | 0.960418 | 0.001179 | 5.64E-15 | TRUE | 4.21E-08 | -+ | 0.002481 | 1 | 0.010182 | 1 | 2.7E-10 | 1 |
| 2-133308946-A-G | G | | | | | | | 0.001143 | 0.931604 | 0.00162 | 3.95E-20 | TRUE | 8.11E-12 | +- | 0.000171 | 1 | 0.259856 | 1 | 8.65E-16 | 1 |
| 2-133308953-A-G | G | | | | | | | 0.001103 | 0.946634 | 0.001406 | 1.55E-15 | TRUE | 8.23E-09 | +- | 0.004579 | 1 | 0.141102 | 1 | 5.2E-12 | 1 |
| 2-134152483-C-G | G | | | | | | | 0.000147 | 0.563896 | 0.000734 | 4.07E-08 | TRUE | 2.51E-06 | +- | 0.011735 | 1 | 0.328397 | -1 | 1.22E-07 | 1 |
| 2-134710815-T-C | C | 0.306557 | 0.513896 | 0.234 | 0.209716 | 0.4644 | ++ | | | 0.322076 | 4.4E-08 | TRUE | 4.4E-08 | ?- | 0.351965 | 1 | 0.298182 | 1 | 2.46E-05 | 1 |
| 2-135470542-CT-C | C | | | | | | | | | 0.001057 | 6.69E-14 | TRUE | 6.69E-14 | ?- | 0.016428 | 1 | 0.14835 | 1 | 6.77E-12 | 1 |
| 2-137484865-GTTGT-( | G | | | | | | | 0.79205 | 0.866816 | 0.652445 | 1.03E-11 | FALSE | 8.45E-05 | -+ | 0.033105 | 1 | 0.198373 | 1 | 2.04E-10 | 1 |
| 2-141583822-T-C | C | 0.473006 | 0.089478 | 0.415 | 0.674296 | 0.0844 | ++ | 0.392278 | 0.086385 | 0.493753 | 1.99E-11 | FALSE | 3.08E-07 | -- | 0.097555 | 1 | 0.337643 | 1 | 1.64E-09 | 1 |
| 2-151415128-A-ACC | ACC | 0.000136 | 0.199483 | | | 0.1995 | +? | | | 0.002331 | 8.08E-14 | TRUE | 8.08E-14 | ?- | 0.01675 | 1 | 0.001396 | 1 | 7.15E-11 | 1 |
| 2-156856973-T-A | A | | | | | | | 0.000132 | 0.448627 | 0.000295 | 3.41E-10 | TRUE | 4.81E-07 | -+ | 0.001393 | 1 | 0.021363 | 1 | 2.16E-08 | 1 |
| 2-157549149-T-A | A | | | | | | | | | 0.000194 | 3.5E-09 | TRUE | 3.5E-09 | ?+ | | | 0.004845 | 1 | 2.58E-06 | 1 |
| 2-157549175-GA-G | G | | | | | | | | | 0.000103 | 3.02E-09 | TRUE | 3.02E-09 | ?+ | | | | | 1.45E-07 | 1 |
| 2-167563224-A-G | G | | | | | | | | | 0.567631 | 1.5E-09 | FALSE | 1.5E-09 | ?- | 0.511558 | 1 | 0.277575 | 1 | 2.2E-05 | 1 |
| 2-169608315-G-C | C | | | | | | | 0.000276 | 0.32132 | 0.003184 | 1.41E-20 | TRUE | 5.1E-20 | ++ | 0.000518 | 1 | 0.079713 | 1 | 4.34E-18 | 1 |
| 2-174296329-T-TA | TA | | | | | | | 0.001585 | 0.506122 | 0.002503 | 6.78E-23 | TRUE | 1.77E-15 | -- | 0.000805 | 1 | 0.034565 | 1 | 2.9E-20 | 1 |
| 2-179792562-A-G | G | | | | | | | | | 0.00059 | 3.76E-08 | TRUE | 3.77E-08 | ?- | 0.007998 | 1 | 0.9451 | 1 | 1.31E-06 | 1 |
| 2-179792566-A-G | G | | | | | | | | | 0.000642 | 4.46E-10 | TRUE | 4.46E-10 | ?- | 0.010241 | 1 | 0.993611 | -1 | 1E-09 | 1 |
| 2-183479004-CTG-C | C | | | | | | | | | 0.001858 | 5.77E-11 | TRUE | 5.77E-11 | ?- | 0.454436 | 1 | 0.001787 | 1 | 3.68E-08 | 1 |
| 2-190430864-C-T | T | | | | | | | | | 0.060785 | 3.33E-08 | FALSE | 3.33E-08 | ?+ | 0.789774 | 1 | 0.333677 | 1 | 1.09E-08 | 1 |

| | | | | | | | | | | | | | | | | | | | | |
|---|---|---|---|---|---|---|---|---|---|---|---|---|---|---|---|---|---|---|---|---|
| 2-198215952-C-CCT | CCT | | | | | | | | | 0.002003 | 1.39E-11 | TRUE | 1.39E-11 | ?+ | 0.074564 | 1 | 0.029453 | 1 | 2.37E-08 | 1 |
| 2-202003854-CCT-C | C | | | | | | | | | 0.000346 | 3.38E-09 | TRUE | 3.38E-09 | ?- | 0.681605 | 1 | 0.005336 | 1 | 1.56E-07 | 1 |
| 2-202294096-CCT-C | C | | | | | | | | | 0.001152 | 2.61E-09 | TRUE | 2.61E-09 | ?- | 0.024924 | 1 | 0.131645 | 1 | 3.99E-08 | 1 |
| 2-202567965-C-CGT | CGT | | | | | | | | | 0.000453 | 2.45E-08 | TRUE | 2.45E-08 | ?+ | 0.011584 | 1 | 0.013753 | 1 | 3.79E-06 | 1 |
| 2-202813151-T-C | C | | | | | | | | | 0.000444 | 2.83E-13 | TRUE | 2.83E-13 | ?- | 0.006314 | 1 | 0.167235 | 1 | 2.42E-11 | 1 |
| 2-202861728-A-AAT | AAT | | | | | | | | | 0.000389 | 2.83E-08 | TRUE | 2.83E-08 | ?- | 0.145093 | 1 | 0.011832 | 1 | 5.9E-07 | 1 |
| 2-205992418-A-ATGG | ATGG | | | | | | | | | 0.000395 | 5.67E-10 | TRUE | 5.67E-10 | ?- | 0.024365 | 1 | 0.195635 | 1 | 1.17E-07 | 1 |
| 2-205992420-AGGG-A | A | | | | | | | | | 0.000176 | 4.47E-09 | TRUE | 4.47E-09 | ?+ | | | | | 3.45E-07 | 1 |
| 2-206050883-G-C | C | | | | | | | 0.000172 | 0.802356 | 0.000794 | 4.28E-13 | TRUE | 1.18E-10 | -+ | 4.7E-05 | 1 | 5.46E-05 | 1 | 1.09E-07 | 1 |
| 2-213584240-T-TA | TA | | | | | | | 0.000122 | 0.7376 | 0.00253 | 5.92E-20 | TRUE | 9.46E-19 | +- | 0.004144 | 1 | 0.041975 | 1 | 1.57E-17 | 1 |
| 2-215134553-T-TATA | TATAC | | | | | | | 0.004333 | 0.427791 | 0.055274 | 4.85E-08 | FALSE | 1.59E-06 | +- | 0.469609 | 1 | 0.966882 | -1 | 6.26E-08 | 1 |
| 2-219315661-G-A | A | | | | | | | 0.009382 | 0.545234 | 0.001565 | 3.58E-10 | TRUE | 0.006004 | ++ | 0.048251 | 1 | 0.768058 | 1 | 7.91E-09 | 1 |
| 2-219315696-T-A | A | | | | | | | 0.00193 | 0.69334 | 0.001399 | 8.09E-11 | TRUE | 1.43E-05 | ++ | 0.864932 | -1 | 0.0184 | 1 | 9.58E-11 | 1 |
| 2-219581029-AGGTG- | A | | | | | | | | | 0.000585 | 1.22E-08 | TRUE | 1.22E-08 | ?+ | 8.98E-06 | 1 | 0.974472 | 1 | 1.03E-05 | 1 |
| 2-219581037-A-AGGT | AGGTG | | | | | | | | | 0.000666 | 1.91E-12 | TRUE | 1.91E-12 | ?- | 1.82E-05 | 1 | 0.024319 | 1 | 6.51E-09 | 1 |
| 2-221892456-A-AG | AG | | | | | | | 0.999749 | 0.050605 | 0.991685 | 4.63E-09 | TRUE | 1.27E-07 | -+ | 0.000619 | -1 | 0.042477 | -1 | 2.84E-06 | -1 |
| 2-223825837-G-T | T | | | | | | | 0.0014 | 0.238963 | 0.001246 | 3.5E-19 | TRUE | 1.16E-07 | -+ | 0.001014 | 1 | 0.06051 | 1 | 2.3E-17 | 1 |
| 2-224529423-C-A | A | | | | | | | | | 0.000857 | 4.38E-10 | TRUE | 4.38E-10 | ?+ | 0.98017 | -1 | 0.280243 | 1 | 8.7E-10 | 1 |
| 2-224755811-C-CCT | CCT | | | | | | | | | 0.00262 | 1.43E-08 | TRUE | 1.43E-08 | ?+ | 0.065116 | 1 | 0.514445 | 1 | 2.17E-07 | 1 |
| 2-231440470-A-ATG | ATG | 0.000371 | 0.53422 | | | 0.5342 | +? | | | 0.001465 | 1.26E-16 | TRUE | 1.26E-16 | ?- | 0.002564 | 1 | 0.021079 | 1 | 8.43E-13 | 1 |
| 2-231440472-ACG-A | A | 0.000296 | 0.451381 | | | 0.4514 | -? | | | 0.001044 | 1.31E-09 | TRUE | 1.31E-09 | ?+ | 0.004821 | 1 | 0.009273 | 1 | 8.23E-07 | 1 |
| 2-231501097-G-C | C | | | | | | | | | 0.000398 | 1.32E-09 | TRUE | 1.32E-09 | ?+ | 0.000133 | 1 | 0.497542 | -1 | 7.25E-08 | 1 |
| 2-231565270-G-A | A | 0.000117 | 0.824962 | | | 0.825 | -? | | | 0.000565 | 3.38E-09 | TRUE | 3.38E-09 | ?+ | 0.011723 | 1 | 0.194853 | 1 | 4.98E-07 | 1 |
| 2-231565277-G-A | A | 0.000137 | 0.598121 | | | 0.5981 | -? | 0.00031 | 0.618733 | 0.000771 | 9.46E-11 | TRUE | 1.24E-08 | ++ | 0.000551 | 1 | 0.255456 | 1 | 6.58E-09 | 1 |
| 2-231565279-G-C | C | 7.83E-05 | 0.410995 | | | 0.411 | -? | 0.000273 | 0.508329 | 0.001006 | 7.88E-16 | TRUE | 9.5E-14 | ++ | 0.001067 | 1 | 0.061525 | 1 | 7.03E-13 | 1 |
| 2-234316318-G-A | A | 0.001326 | 0.16144 | | | 0.1614 | +? | 0.000269 | 0.197389 | 0.001023 | 1.09E-11 | TRUE | 1.47E-08 | -+ | 0.013013 | 1 | 0.000182 | 1 | 1.6E-07 | 1 |
| 2-236272485-G-A | A | | | | | | | 0.000733 | 0.894413 | 0.001419 | 3.55E-08 | TRUE | 1.06E-05 | ++ | 0.01473 | 1 | 0.514083 | 1 | 9.07E-07 | 1 |
| 2-238119146-GAA-G | G | | | | | | | | | 0.985719 | 1.6E-09 | TRUE | 1.6E-09 | ?- | 0.124588 | -1 | 0.327588 | -1 | 2.9E-08 | -1 |
| 2-238794552-C-T | T | | | | | | | | | 0.000857 | 1.09E-11 | TRUE | 1.09E-11 | ?+ | 0.002255 | 1 | 0.008809 | 1 | 1.62E-08 | 1 |
| 2-238902978-T-TG | TG | | | | | | | | | 0.688204 | 1.61E-12 | FALSE | 1.61E-12 | ?- | 0.013891 | 1 | 0.984142 | -1 | 1.58E-12 | 1 |
| 2-238902978-T-TGTG | TGTGTGTGATGTG | | | | | | | | | 0.268022 | 2.28E-20 | FALSE | 2.28E-20 | ?+ | 0.010892 | -1 | 0.478033 | -1 | 1.14E-19 | -1 |
| 2-239719907-A-G | G | | | | | | | | | 0.054377 | 1.4E-08 | FALSE | 1.4E-08 | ?- | 0.285093 | 1 | 0.436976 | 1 | 4.25E-07 | 1 |
| 2-239815908-G-GA | GA | | | | | | | | | 0.006904 | 1.22E-10 | TRUE | 1.22E-10 | ?- | 0.102867 | 1 | 0.466246 | 1 | 3.23E-09 | 1 |
| 2-240961860-ACACTC | A | | | | | | | | | 0.062818 | 5.9E-16 | FALSE | 5.9E-16 | ?+ | 0.177156 | 1 | 0.24754 | 1 | 2.44E-16 | 1 |
| 2-241488609-C-G | G | | | | | | | 0.0002 | 0.717588 | 0.003266 | 1.67E-44 | TRUE | 2.18E-42 | -- | 2.21E-06 | 1 | 0.000288 | 1 | 1.5E-37 | 1 |
| 2-242066139-GTGA-G | G | | | | | | | | | 0.197841 | 7.52E-09 | FALSE | 7.52E-09 | ?+ | 0.082423 | 1 | 0.019147 | -1 | 9.67E-14 | 1 |
| 2-193847374-G-T | T | | | | | | | 0.000749 | 0.137945 | 0.008827 | 0.070637 | FALSE | Inf | +- | 0.294149 | -1 | 0.088396 | -1 | 0.516364 | -1 |
| 2-186273999-G-A | A | | | | | | | 4.39E-05 | 0.709561 | | | FALSE | 1.88E-24 | +? | | | | | | |
| 2-44498127-T-A | A | | | | | | | 9.4E-06 | 0.487109 | | | FALSE | 2.42E-22 | -? | | | | | | |
| 2-217648932-T-A | A | | | | | | | 6.27E-05 | 0.002357 | | | FALSE | 5.89E-12 | -? | | | | | | |
| 2-20112214-AGTTTTG | AGTTTT | | | | | | | 4.07E-05 | 0.237363 | | | FALSE | Inf | -? | | | | | | |
| 2-127128840-C-A | A | | | 0.311 | 0.105174 | 0.1052 | ?- | 0.299819 | 5.39E-08 | 0.283309 | 0.023627 | TRUE | 8.69E-09 | -- | 0.424213 | -1 | 0.092485 | 1 | 0.081349 | 1 |
| 2-201749788-T-A | A | | | | | | | 9.4E-06 | 0.394871 | | | FALSE | 1.8E-23 | +? | | | | | | |
| 3-109106242-C-T | T | 0.000779 | 4.71E-08 | | | 4.707E-08 | +? | 0.000191 | 0.569856 | 0.000102 | 0.284813 | TRUE | 0.9534 | +- | | | | | | |
| 3-3430731-C-T | T | | | | | | | 0.000564 | 0.484819 | 0.000316 | 4.32E-08 | TRUE | 5.51E-05 | ++ | | | 0.174979 | 1 | 4.42E-08 | 1 |
| 3-3431293-G-T | T | 7.8E-05 | 0.971288 | | | 0.9713 | -? | 0.001613 | 0.122209 | 0.00176 | 4.34E-21 | TRUE | 1.75E-08 | -+ | 0.000196 | 1 | 0.347116 | 1 | 1.87E-17 | 1 |
| 3-3431321-T-C | C | | | | | | | 1.25E-05 | 0.386242 | 0.000223 | 1.14E-08 | TRUE | 5.79E-08 | +- | 0.323172 | 1 | | | 8.72E-07 | 1 |
| 3-9803104-T-C | C | 0.000235 | 0.782904 | | | 0.7829 | +? | 0.000319 | 0.900714 | 0.00113 | 3.77E-09 | TRUE | 5.22E-07 | +- | 0.162544 | 1 | 0.957328 | -1 | 2.68E-09 | 1 |
| 3-9803125-C-T | T | 0.00041 | 0.245625 | | | 0.2456 | -? | 5.32E-05 | 0.708871 | 0.001008 | 1.9E-09 | TRUE | 3.02E-09 | ++ | 0.000884 | 1 | 0.018516 | 1 | 1.02E-06 | 1 |
| 3-10030681-CCT-C | C | | | | | | | | | 0.003487 | 2.14E-27 | TRUE | 2.14E-27 | ?- | 0.00011 | 1 | 4.01E-05 | 1 | 2.68E-21 | 1 |
| 3-10102824-C-CA | CA | | | | | | | | | 0.00055 | 7.12E-11 | TRUE | 7.12E-11 | ?+ | 0.001815 | 1 | 0.017388 | 1 | 5.04E-07 | 1 |
| 3-10239992-C-CAA | CAA | | | | | | | | | 0.005683 | 1.07E-17 | TRUE | 1.07E-17 | ?+ | 5.06E-06 | 1 | 0.033628 | 1 | 1.12E-11 | 1 |
| 3-11166550-C-CG | CG | | | | | | | 0.000398 | 0.677911 | 0.001116 | 3.01E-08 | TRUE | 1.26E-05 | -+ | 0.820532 | -1 | 0.747128 | 1 | 1.87E-08 | 1 |
| 3-11823413-G-A | A | 0.000256 | 0.042238 | | | 0.04224 | -? | 0.007236 | 0.285145 | 0.00167 | 2.94E-14 | TRUE | 4.89E-05 | ++ | 0.007602 | 1 | 0.002413 | 1 | 7.68E-11 | 1 |
| 3-12502323-ATC-A | A | 0.000156 | 0.914638 | | | 0.9146 | -? | | | 0.001639 | 1.15E-11 | TRUE | 1.15E-11 | ?+ | 0.417849 | 1 | 0.474374 | 1 | 4.97E-11 | 1 |
| 3-15374074-C-CTA | CTA | | | | | | | | | 0.813203 | 4.59E-14 | FALSE | 4.59E-14 | ?- | 0.101428 | -1 | 0.014881 | -1 | 1.77E-11 | -1 |
| 3-15374076-AAG-A | A | | | | | | | | | 0.829085 | 1.68E-13 | FALSE | 1.68E-13 | ?- | 0.305668 | -1 | 0.019417 | -1 | 5.13E-12 | -1 |
| 3-15491870-C-T | T | 7.8E-05 | 0.482274 | | | 0.4823 | -? | | | 0.000745 | 6.47E-09 | TRUE | 6.47E-09 | ?+ | 0.09238 | 1 | 0.013924 | 1 | 1.28E-07 | 1 |
| 3-15491876-G-A | A | | | | | | | 6.26E-05 | 0.979003 | 0.000872 | 1.35E-13 | TRUE | 7.72E-13 | ++ | 0.00899 | 1 | 0.002304 | 1 | 6.47E-11 | 1 |

| | | | | | | | | | | | | | | | | | | |
|---|---|---|---|---|---|---|---|---|---|---|---|---|---|---|---|---|---|---|
| 3-15569473-T-C | C | | | | | | | 0.000959 | 8.02E-10 | TRUE | 8.02E-10 | ?- | 0.710559 | 1 | 0.000326 | 1 | 1.07E-07 | 1 |
| 3-15569482-G-A | A | | | | | 9.4E-06 | 0.593871 | 0.000627 | 9.45E-09 | TRUE | 1.47E-08 | -+ | 0.961701 | 1 | 0.06542 | 1 | 7.3E-08 | 1 |
| 3-15569486-T-C | C | | | | | 9.4E-06 | 0.408286 | 0.000489 | 5.66E-09 | TRUE | 1.55E-08 | +- | 0.178538 | 1 | 0.139751 | 1 | 5.27E-08 | 1 |
| 3-15592997-CATGTGT | C | | | | | | | 0.009524 | 1.52E-08 | TRUE | 1.52E-08 | ?- | 0.643778 | 1 | 0.103301 | 1 | 2.93E-08 | 1 |
| 3-15593380-AT-A | A | | | | | | | 0.001262 | 4.92E-14 | TRUE | 4.93E-14 | ?+ | 0.138226 | 1 | 0.006116 | 1 | 4.8E-12 | 1 |
| 3-15593389-ATTTT-A | A | | | | | | | 0.001204 | 9.19E-12 | TRUE | 9.19E-12 | ?+ | 0.434215 | 1 | 0.00077 | 1 | 9.54E-09 | 1 |
| 3-16422426-G-C | C | 9.77E-05 | 0.142088 | 0.1421 | +? | 0.000357 | 0.248755 | 0.001222 | 2.28E-10 | TRUE | 1.25E-06 | -+ | 0.001458 | 1 | 0.633254 | 1 | 5.61E-09 | 1 |
| 3-21988145-A-G | G | | | | | | | 0.026028 | 3.37E-10 | FALSE | 3.37E-10 | ?- | 0.009206 | 1 | 0.126984 | 1 | 1.37E-07 | 1 |
| 3-29143871-C-A | A | | | | | 4.39E-05 | 0.893975 | 0.000354 | 3.12E-08 | TRUE | 1.79E-07 | -+ | 0.039768 | 1 | 0.154342 | 1 | 3.07E-06 | 1 |
| 3-36963928-C-T | T | 0.001217 | 0.450887 | 0.4509 | -? | | | 0.001413 | 5.88E-09 | TRUE | 5.88E-09 | ?+ | 0.01855 | 1 | 0.321854 | 1 | 9.08E-08 | 1 |
| 3-36963941-G-T | T | | | | | 4.7E-05 | 0.528123 | 0.000705 | 3.73E-08 | TRUE | 4.13E-08 | ++ | 0.618852 | 1 | 0.072089 | 1 | 2.01E-08 | 1 |
| 3-40344008-A-AGATG | AGATG | | | | | | | 0.000511 | 5.17E-12 | TRUE | 5.17E-12 | ?- | 0.851659 | 1 | 0.004671 | 1 | 1.31E-11 | 1 |
| 3-41061991-A-AT | AT | 0.000156 | 0.222265 | 0.2223 | -? | | | 0.001072 | 2.6E-10 | TRUE | 2.6E-10 | ?- | 0.179704 | 1 | 0.013194 | 1 | 2.06E-09 | 1 |
| 3-41061993-AG-A | A | | | | | | | 0.001347 | 1.06E-10 | TRUE | 1.06E-10 | ?+ | 0.072477 | 1 | 0.02591 | 1 | 1.28E-09 | 1 |
| 3-41971464-CA-C | C | 0.000585 | 0.823928 | 0.8239 | +? | | | 0.001884 | 6.17E-11 | TRUE | 6.17E-11 | ?- | 0.003429 | 1 | 0.16101 | 1 | 2.56E-09 | 1 |
| 3-47755019-GAT-G | G | 0.000917 | 0.858589 | 0.8586 | +? | | | 0.001969 | 6.48E-11 | TRUE | 6.48E-11 | ?+ | 0.004365 | 1 | 0.752425 | -1 | 9.64E-10 | 1 |
| 3-47755022-G-GCA | GCA | | | | | | | 0.001906 | 1.48E-10 | TRUE | 1.48E-10 | ?- | 0.01789 | 1 | 0.626579 | -1 | 2.03E-10 | 1 |
| 3-47789462-C-G | G | | | | | 0.000526 | 0.897148 | 0.000237 | 3.8E-08 | TRUE | 0.000934 | -- | 0.411261 | 1 | | | 2.58E-06 | 1 |
| 3-47789474-G-T | T | | | | | | | 8.79E-05 | 3.78E-08 | TRUE | 3.78E-08 | ?+ | | | | | 1.98E-06 | 1 |
| 3-48201876-A-G | G | 0.000531 | 0.799837 | 0.7998 | -? | 0.006496 | 0.983892 | 0.00184 | 7.41E-13 | TRUE | 0.001856 | +- | 0.113718 | 1 | 0.073144 | 1 | 5.72E-13 | 1 |
| 3-48201926-C-G | G | 0.000235 | 0.261266 | 0.2613 | -? | 0.002885 | 0.009898 | 0.001361 | 1.3E-08 | TRUE | 3.53E-07 | -- | 0.012511 | 1 | 0.540823 | 1 | 2.56E-08 | 1 |
| 3-48876877-CCT-C | C | | | | | | | 0.000693 | 1.65E-11 | TRUE | 1.65E-11 | ?- | 0.510591 | 1 | 0.769017 | -1 | 1.39E-12 | 1 |
| 3-48925935-C-G | G | | | | | | | 0.000846 | 1.54E-09 | TRUE | 1.54E-09 | ?- | 0.151959 | 1 | 0.799577 | -1 | 8.67E-09 | 1 |
| 3-48939528-CGGGTG( | C | | | | | | | 0.002864 | 7.08E-09 | TRUE | 7.08E-09 | ?- | 0.354303 | 1 | 0.015823 | 1 | 9.21E-08 | 1 |
| 3-49210704-T-TCTAA | TCTAA | | | | | | | 0.00133 | 7.16E-11 | TRUE | 7.16E-11 | ?- | 0.002945 | 1 | 0.009845 | 1 | 7.71E-08 | 1 |
| 3-49399235-C-T | T | | | | | 0.000269 | 0.318581 | 0.000538 | 6.73E-09 | TRUE | 3.98E-05 | -+ | 0.004345 | 1 | 0.008332 | 1 | 2.38E-05 | 1 |
| 3-49947243-C-CATGA( | CATGAGAA | | | | | | | 0.00191 | 2.7E-16 | TRUE | 2.7E-16 | ?+ | 0.004926 | 1 | 0.097111 | 1 | 2.03E-15 | 1 |
| 3-50307554-G-A | A | 0.087834 | 0.162401 | 0.1624 | -? | | | 0.075205 | 2.98E-08 | FALSE | 2.98E-08 | ?+ | 0.87054 | -1 | 0.226249 | 1 | 0.050426 | 1 |
| 3-51353784-CCTCT-C | C | 9.75E-05 | 0.922664 | 0.9227 | +? | 2.19E-05 | 0.18184 | 0.002089 | 8.99E-18 | TRUE | 5.87E-17 | +- | 0.113409 | 1 | 0.00926 | 1 | 4.85E-16 | 1 |
| 3-61672000-A-G | G | | | | | | | 0.074192 | 1.15E-10 | FALSE | 1.16E-10 | ?- | 0.913855 | 1 | 0.890912 | 1 | 1.54E-11 | 1 |
| 3-71313726-C-T | T | 0.000237 | 0.874471 | 0.8745 | -? | 0.002875 | 0.937193 | 0.001882 | 7.92E-10 | TRUE | 0.000451 | -+ | 0.699178 | 1 | 0.386596 | 1 | 2.71E-10 | 1 |
| 3-73151147-G-A | A | 0.000215 | 0.46787 | 0.4679 | -? | 1.57E-05 | 0.46582 | 0.000141 | 1.56E-09 | TRUE | 1.43E-08 | -+ | 2.49E-06 | 1 | | | | |
| 3-75612176-C-CG | CG | | | | | | | 0.108114 | 1.23E-10 | TRUE | 1.23E-10 | ?+ | 0.633207 | 1 | 0.976334 | 1 | 1.98E-12 | 1 |
| 3-75616418-A-ATG | ATG | | | | | 0.000658 | 0.413409 | 0.001656 | 1.33E-20 | TRUE | 8.46E-17 | -- | 0.010401 | 1 | 0.018298 | 1 | 6.09E-19 | 1 |
| 3-75674095-TCAAATA | T | | | | | | | 0.003354 | 4.02E-13 | TRUE | 4.02E-13 | ?+ | 0.004004 | 1 | 0.014769 | 1 | 2.12E-10 | 1 |
| 3-75734134-T-A | A | | | | | | | 0.000118 | 1.47E-08 | TRUE | 1.47E-08 | ?+ | | | | | 9.16E-08 | 1 |
| 3-77292683-GGGTAA( | G | | | | | | | 0.261225 | 1.24E-10 | TRUE | 1.24E-10 | ?- | 0.631302 | -1 | 0.939375 | -1 | 2.93E-10 | -1 |
| 3-77294296-G-GTAAA | GTAAAATTGACGGTTAAACGGGTAAGCTGAGGCAGATCACCAAAGACATAAAA | | | | | | | 0.300102 | 1.15E-11 | FALSE | 1.15E-11 | ?+ | 0.579059 | -1 | 0.596997 | -1 | 1.34E-10 | -1 |
| 3-80420455-G-T | T | | | | | 4.07E-05 | 0.34103 | 0.000629 | 8.47E-09 | TRUE | 1.06E-07 | -+ | 0.0138 | 1 | 0.657531 | 1 | 4.07E-07 | 1 |
| 3-90283097-C-CT | CT | | | | | | | 0.001818 | 6.09E-09 | TRUE | 6.09E-09 | ?+ | 0.010815 | 1 | 0.481179 | 1 | 6.73E-08 | 1 |
| 3-90291826-T-C | C | | | | | 0.001218 | 0.116525 | 0.000577 | 1.24E-08 | TRUE | 0.0554 | +- | 0.012992 | 1 | 0.190317 | 1 | 4.26E-08 | 1 |
| 3-90498243-C-G | G | | | | | | | 0.000962 | 2.01E-08 | TRUE | 2.01E-08 | ?- | 0.010938 | 1 | 0.001929 | 1 | 1.72E-05 | 1 |
| 3-90498313-G-T | T | | | | | 5.95E-05 | 0.385347 | 0.000807 | 1.62E-09 | TRUE | 1.52E-09 | ++ | 7.92E-05 | 1 | 0.016943 | 1 | 1.77E-05 | 1 |
| 3-103419699-GT-G | G | | | | | 0.004269 | 0.094139 | 0.004396 | 1.66E-31 | TRUE | 7.12E-20 | ++ | 0.005162 | 1 | 2.56E-07 | 1 | 1.28E-23 | 1 |
| 3-109285911-C-CGT | CGT | | | | | | | 0.000857 | 9.05E-09 | TRUE | 9.05E-09 | ?+ | 0.041748 | 1 | 0.524943 | -1 | 6.99E-09 | 1 |
| 3-119480415-A-ATT | ATT | 0.000332 | 0.254422 | 0.2544 | +? | | | 0.002212 | 5.25E-15 | TRUE | 5.25E-15 | ?- | 0.243042 | 1 | 0.003214 | 1 | 2.22E-14 | 1 |
| 3-121791872-CCT-C | C | | | | | | | 0.001225 | 1.89E-12 | TRUE | 1.89E-12 | ?- | 0.014882 | 1 | 0.635616 | 1 | 5.02E-11 | 1 |
| 3-122934000-CAGGTT | C | 8.02E-05 | 0.354276 | 0.3543 | -? | | | 0.002292 | 4.4E-10 | TRUE | 4.4E-10 | ?- | 0.002034 | 1 | 0.16014 | 1 | 2.22E-08 | 1 |
| 3-124436561-G-A | A | 0.000447 | 0.320713 | 0.3207 | -? | | | 0.000733 | 3.72E-11 | TRUE | 3.72E-11 | ?+ | 0.967061 | -1 | 0.004019 | 1 | 1.38E-09 | 1 |
| 3-124436578-T-A | A | | | | | | | 0.000686 | 3.62E-08 | TRUE | 3.62E-08 | ?+ | 0.05291 | 1 | 0.914798 | -1 | 2.58E-07 | 1 |
| 3-124807408-TC-T | T | 9.75E-05 | 0.443236 | 0.4432 | -? | | | 0.000338 | 7.49E-11 | TRUE | 7.49E-11 | ?+ | 0.101664 | 1 | 0.005658 | 1 | 1.05E-07 | 1 |
| 3-124898425-C-CCT | CCT | | | | | | | 0.005099 | 1.88E-10 | TRUE | 1.88E-10 | ?+ | 0.794931 | -1 | 0.013327 | 1 | 5.08E-09 | 1 |
| 3-125521495-A-AAAA | AAAATAC | | | | | | | 0.000297 | 4.31E-08 | TRUE | 4.31E-08 | ?- | 0.017945 | 1 | 0.016625 | 1 | 7.64E-07 | 1 |
| 3-127582685-G-GAGT | GAGT | | | | | | | 0.000592 | 1.39E-10 | TRUE | 1.39E-10 | ?- | 0.002682 | 1 | 0.027678 | 1 | 1.11E-07 | 1 |
| 3-127582687-GCAA-G | G | | | | | | | 0.000405 | 3.46E-09 | TRUE | 3.46E-09 | ?+ | 0.000453 | 1 | 0.002774 | 1 | 2.02E-05 | 1 |
| 3-128645253-CCT-C | C | | | | | | | 0.000643 | 1.06E-09 | TRUE | 1.06E-09 | ?- | 0.008217 | 1 | 0.008553 | 1 | 1.56E-07 | 1 |
| 3-128696475-A-G | G | 7.81E-05 | 0.140997 | 0.141 | +? | | | 0.000471 | 2.42E-08 | TRUE | 2.42E-08 | ?- | 0.733051 | 1 | 0.006318 | 1 | 1.65E-07 | 1 |
| 3-128696485-CTT-C | C | | | | | | | 0.000676 | 5.31E-13 | TRUE | 5.31E-13 | ?- | 0.01049 | 1 | 0.450984 | 1 | 7.86E-13 | 1 |
| 3-129426499-G-T | T | | | | | 0.000319 | 0.435313 | 0.000518 | 1.09E-10 | TRUE | 2.24E-08 | ++ | 0.014782 | 1 | 0.08985 | 1 | 5.19E-08 | 1 |

| | | | | | | | | | | | | | | | | | | | | |
|---|---|---|---|---|---|---|---|---|---|---|---|---|---|---|---|---|---|---|---|---|
| 3-134701277-C-CATA1 | CATATACGTATATATGTGT | | | | | | | | | 0.319314 | 5.45E-09 | FALSE | 5.45E-09 | ?- | 0.219757 | -1 | 0.309773 | -1 | 6.41E-08 | -1 |
| 3-134701277-CAT-C | C | | | | | | | | | 0.273906 | 2.65E-12 | FALSE | 2.65E-12 | ?- | 0.114373 | 1 | 0.261706 | 1 | 2.47E-10 | 1 |
| 3-136704609-CTT-C | C | 7.8E-05 | 0.38457 | | | 0.3846 | +? | | | 0.001369 | 1.2E-08 | TRUE | 1.2E-08 | ?- | 0.592304 | 1 | 0.001078 | 1 | 3.03E-06 | 1 |
| 3-138728773-CCG-C | C | 0.000902 | 0.926752 | | | 0.9268 | +? | | | 0.001371 | 3.16E-10 | TRUE | 3.16E-10 | ?- | 0.0056 | 1 | 0.076832 | 1 | 2.34E-08 | 1 |
| 3-141640052-CT-C | C | 0.005323 | 0.019318 | | | 0.01932 | +? | 0.001074 | 0.402725 | 0.002499 | 2.79E-10 | TRUE | 1.05E-05 | +- | 0.979332 | 1 | 0.028894 | 1 | 2.32E-09 | 1 |
| 3-151417487-C-CGGG | CGGG | | | | | | | | | 0.001566 | 3.26E-08 | TRUE | 3.26E-08 | ?- | 0.237039 | -1 | 0.504998 | -1 | 1.8E-07 | -1 |
| 3-153261711-T-C | C | | | | | | | 0.000902 | 0.365319 | 0.001505 | 9.93E-11 | TRUE | 1.48E-05 | +- | 0.383731 | 1 | 0.582518 | 1 | 1.13E-09 | 1 |
| 3-155795816-T-C | C | 0.000117 | 0.078722 | | | 0.07872 | -? | 0.001757 | 0.11838 | 0.001747 | 3.39E-11 | TRUE | 1.81E-08 | -- | 0.203575 | 1 | 0.009635 | 1 | 2.03E-09 | 1 |
| 3-170509612-C-T | T | | | | | | | | | 0.000617 | 3.42E-09 | TRUE | 3.42E-09 | ?+ | 0.008642 | 1 | 0.920994 | -1 | 1.74E-09 | 1 |
| 3-170873157-C-CT | CT | | | | | | | 7.83E-05 | 0.7272 | 0.000662 | 8.52E-12 | TRUE | 1.79E-10 | -+ | 0.000105 | 1 | 0.037707 | 1 | 2.87E-08 | 1 |
| 3-174762293-A-G | G | 0.000532 | 0.024113 | | | 0.02411 | +? | 0.000194 | 0.984198 | 0.001754 | 7.17E-09 | TRUE | 7.04E-08 | +- | 0.077048 | 1 | 0.316946 | -1 | 3.55E-09 | 1 |
| 3-174762303-G-A | A | 0.001085 | 0.021198 | | | 0.0212 | -? | 0.00031 | 0.284941 | 0.002325 | 4.05E-10 | TRUE | 6.92E-10 | ++ | 0.22197 | 1 | 0.006959 | 1 | 3.13E-09 | 1 |
| 3-177225071-CAG-C | C | | | | | | | | | 0.00083 | 1.33E-10 | TRUE | 1.33E-10 | ?- | 0.090686 | 1 | 0.001174 | 1 | 5.59E-08 | 1 |
| 3-183473142-G-GGAG | GGAGT | | | | | | | | | 0.005743 | 1.05E-11 | TRUE | 1.05E-11 | ?- | 0.069402 | 1 | 0.016475 | 1 | 1.79E-09 | 1 |
| 3-184425210-C-T | T | 9.76E-05 | 0.182673 | | | 0.1827 | -? | | | 0.000973 | 1.74E-13 | TRUE | 1.74E-13 | ?+ | 0.000454 | 1 | 0.131249 | 1 | 2.67E-11 | 1 |
| 3-185258472-G-T | T | | | | | | | 0.001169 | 0.925728 | 0.001853 | 6.71E-14 | TRUE | 1.61E-08 | ++ | 0.005875 | 1 | 0.073345 | 1 | 1.69E-11 | 1 |
| 3-187807145-T-A | A | | | | | | | | | 0.001482 | 6.46E-12 | TRUE | 6.46E-12 | ?+ | 0.015021 | 1 | 0.538582 | 1 | 7.38E-12 | 1 |
| 3-192900736-T-A | A | 7.81E-05 | 0.3559 | | | 0.3559 | +? | 0.000319 | 0.427822 | 0.002311 | 2.49E-27 | TRUE | 3.5E-25 | ++ | 0.000133 | 1 | 0.017874 | 1 | 4.02E-23 | 1 |
| 3-192900746-G-T | T | | | | | | | 0.000319 | 0.427822 | 0.002164 | 8.47E-20 | TRUE | 3.2E-18 | ++ | 0.075495 | 1 | 0.012315 | 1 | 1.5E-16 | 1 |
| 3-194845601-TAATGA | T | | | | | | | | | 0.026724 | 8.37E-10 | FALSE | 8.37E-10 | ?- | 0.497416 | -1 | 0.259604 | -1 | 4.91E-09 | -1 |
| 3-194954548-G-A | A | 0.000197 | 0.445494 | | | 0.4455 | +? | 0.000316 | 0.186657 | 0.000688 | 3.62E-09 | TRUE | 2.86E-05 | -+ | 0.006194 | 1 | 0.013845 | 1 | 1.82E-07 | 1 |
| 3-195504736-C-T | T | | | | | | | | | 0.946437 | 4.16E-11 | FALSE | 4.16E-11 | ?- | 0.002112 | -1 | 0.019962 | -1 | 6.17E-08 | -1 |
| 3-195647250-C-CCGG | CCGGG | | | | | | | | | 0.01574 | 4.11E-08 | FALSE | 4.11E-08 | ?- | 0.350846 | -1 | 0.68582 | -1 | 1.51E-07 | -1 |
| 3-195969599-T-TGGC( | TGGCCAGT | 0.001973 | 0.407992 | | | 0.408 | -? | | | 0.013743 | 9.47E-15 | FALSE | 9.47E-15 | ?- | 0.572087 | 1 | 0.057479 | 1 | 9.48E-15 | 1 |
| 3-196266254-C-CCT | CCT | | | | | | | | | 0.002524 | 1.62E-11 | TRUE | 1.62E-11 | ?+ | 0.01095 | 1 | 0.212775 | 1 | 1.06E-09 | 1 |
| 3-196675754-C-CAAG | CAAGTG | | | | | | | | | 0.000678 | 1.66E-08 | TRUE | 1.66E-08 | ?+ | 0.007207 | 1 | 0.002427 | 1 | 3.31E-05 | 1 |
| 3-196852149-AC-A | A | | | | | | | | | 0.000941 | 3.75E-16 | TRUE | 3.75E-16 | ?+ | 0.000596 | 1 | 0.360603 | 1 | 1.05E-12 | 1 |
| 3-196861715-GAT-G | G | | | | | | | | | 0.000777 | 3.05E-08 | TRUE | 3.05E-08 | ?+ | 0.002216 | 1 | 0.191724 | 1 | 4.77E-06 | 1 |
| 3-196861718-G-GCA | GCA | | | | | | | | | 0.000724 | 3.03E-08 | TRUE | 3.03E-08 | ?- | 0.010202 | 1 | 0.190722 | 1 | 7.51E-07 | 1 |
| 3-193198808-C-A | A | | | | | | | 2.51E-05 | 0.60665 | | | FALSE | 2.6E-305 | -? | | | | | | |
| 3-150198383-G-A | A | | | | | | | 8.46E-05 | 0.481669 | | | FALSE | 1.5E-305 | -? | | | | | | |
| 3-24102741-G-A | A | | | | | | | 0.000147 | 0.303351 | 0.0001 | 0.994156 | FALSE | 3.4E-305 | +- | | | | | 0.9689 | 1 |
| 3-90587749-C-T | T | | | | | | | 0.00031 | 0.003102 | 0.000705 | 2.15E-06 | TRUE | 4.29E-08 | ++ | 0.000766 | 1 | 0.714271 | 1 | 0.00154 | 1 |
| 4-79608801-G-A | A | 0.000877 | 3.14E-08 | | | 3.143E-08 | +? | 0.000395 | 0.486409 | 0.001958 | 0.344918 | TRUE | 0.6344 | +- | 0.416241 | -1 | 0.56319 | 1 | 0.022743 | 1 |
| 4-109067562-G-T | T | 0.115909 | 2.73E-08 | 0.068 | 0.735506 | 2.592E-08 | -- | 0.051194 | 0.27679 | 0.09045 | 0.663405 | FALSE | 0.5643 | -+ | 0.900284 | 1 | 0.020445 | -1 | 0.705898 | 1 |
| 4-100831815-T-C | C | | | | | | | 4.07E-05 | 3.63E-08 | 0.000208 | 0.073622 | TRUE | 4.98E-07 | -- | | | | | | |
| 4-100836007-C-T | T | | | | | | | 4.07E-05 | 3.63E-08 | 0.000206 | 0.069019 | TRUE | 4.28E-07 | ++ | | | | | | |
| 4-187598649-C-A | A | | | | | | | 5.01E-05 | 1E-69 | | | FALSE | 0.6905 | -? | | | | | | |
| 4-10580-T-TG | TG | | | | | | | | | 0.939852 | 3.98E-09 | FALSE | 3.98E-09 | ?+ | 0.030063 | -1 | 0.225794 | -1 | 1.87E-09 | -1 |
| 4-634885-GCC-G | G | | | | | | | | | 0.001659 | 1.68E-09 | TRUE | 1.68E-09 | ?+ | 1.38E-05 | 1 | 0.001107 | 1 | 0.000118 | 1 |
| 4-634889-GCC-G | G | | | | | | | | | 0.001332 | 1.23E-08 | TRUE | 1.23E-08 | ?+ | 0.01006 | 1 | 0.00141 | 1 | 2.13E-05 | 1 |
| 4-920686-G-GACGT | GACGT | 7.8E-05 | 0.359887 | | | 0.3599 | +? | | | 0.001865 | 1.03E-13 | TRUE | 1.03E-13 | ?- | 0.010218 | 1 | 0.015964 | 1 | 1.34E-10 | 1 |
| 4-920689-GCCCA-G | G | | | | | | | | | 0.001343 | 2.38E-11 | TRUE | 2.38E-11 | ?+ | 0.012029 | 1 | 0.01386 | 1 | 1.6E-08 | 1 |
| 4-978148-A-G | G | | | | | | | | | 0.000151 | 2.91E-10 | TRUE | 2.91E-10 | ?- | | | | | 8.76E-09 | 1 |
| 4-1146626-G-A | A | | | | | | | | | 0.574209 | 7.79E-13 | FALSE | 7.79E-13 | ?- | 0.129311 | -1 | 6.69E-06 | -1 | 1.35E-08 | -1 |
| 4-1624880-AG-A | A | | | | | | | 0.00016 | 0.026105 | 0.00512 | 1.75E-10 | TRUE | 2.67E-09 | -+ | 0.367251 | 1 | 0.002901 | 1 | 2.88E-08 | 1 |
| 4-2440655-T-G | G | | | | | | | 0.000147 | 0.226579 | 0.000565 | 4.74E-08 | TRUE | 2.04E-05 | +- | 0.268107 | 1 | 0.785954 | 1 | 1.96E-07 | 1 |
| 4-2522168-G-A | A | 0.00041 | 0.268232 | | | 0.2682 | +? | 0.002117 | 0.982773 | 0.002707 | 2.98E-15 | TRUE | 1.79E-08 | ++ | 0.012818 | 1 | 0.00018 | 1 | 9E-17 | 1 |
| 4-2522176-T-C | C | 0.000296 | 0.554237 | | | 0.5542 | -? | 0.001923 | 0.545021 | 0.005354 | 9.46E-26 | TRUE | 2.02E-19 | -- | 0.099026 | 1 | 0.000295 | 1 | 1.9E-27 | 1 |
| 4-2522177-G-A | A | 0.000312 | 0.66755 | | | 0.6676 | +? | 0.001889 | 0.651691 | 0.004481 | 1.84E-23 | TRUE | 6.49E-17 | ++ | 0.000192 | 1 | 0.001849 | 1 | 6.65E-23 | 1 |
| 4-2522803-C-CCT | CCT | | | | | | | 0.001629 | 0.322392 | 0.003076 | 8.21E-22 | TRUE | 2.52E-16 | ++ | 0.099181 | 1 | 0.022241 | 1 | 1.44E-20 | 1 |
| 4-2525429-T-C | C | 0.000156 | 0.740893 | | | 0.7409 | -? | 0.000507 | 0.460048 | 0.00108 | 3.9E-09 | TRUE | 2.35E-07 | -- | 0.062617 | 1 | 0.582806 | 1 | 4.45E-09 | 1 |
| 4-2764348-CAG-C | C | | | | | | | | | 0.004571 | 1.9E-11 | TRUE | 1.9E-11 | ?- | 0.357961 | 1 | 0.341705 | 1 | 3.33E-11 | 1 |
| 4-3568522-T-TCA | TCA | | | | | | | | | 0.608065 | 7.28E-10 | FALSE | 7.28E-10 | ?- | 0.051367 | 1 | 0.853471 | -1 | 7.83E-08 | 1 |
| 4-8967028-A-AAC | AAC | | | | | | | | | 0.000865 | 1.66E-11 | TRUE | 1.66E-11 | ?- | 0.193238 | 1 | 0.000296 | 1 | 2.87E-09 | 1 |
| 4-9501107-A-AAC | AAC | | | | | | | | | 0.00199 | 4.69E-17 | TRUE | 4.69E-17 | ?- | 4.99E-05 | 1 | 6.32E-05 | 1 | 6.31E-10 | 1 |
| 4-9501126-T-C | C | | | | | | | | | 0.000254 | 9.75E-09 | TRUE | 9.75E-09 | ?- | 7.38E-05 | 1 | 0.000213 | 1 | 0.000297 | 1 |
| 4-9593894-T-C | C | | | | | | | | | 0.000179 | 2.38E-08 | TRUE | 2.38E-08 | ?- | 0.023986 | 1 | | | 1.59E-07 | 1 |
| 4-9650027-T-A | A | | | | | | | | | 0.000278 | 1.02E-09 | TRUE | 1.02E-09 | ?+ | 0.00095 | 1 | 0.000147 | 1 | 1.19E-05 | 1 |

| | | | | | | | | | | | | | | | | | | |
|---|---|---|---|---|---|---|---|---|---|---|---|---|---|---|---|---|---|---|
| 4-9650036-G-A | A | | | | | 3.45E-05 | 0.197965 | 0.000225 | 1.61E-09 | TRUE | 7.51E-08 | -+ | 0.000163 | 1 | | | 0.000511 | 1 |
| 4-15985889-T-TAAAT/ | TAAATATTTAATACAGATAA | | | | | | | 0.67259 | 1.52E-33 | FALSE | 1.52E-33 | ?+ | 0.395258 | -1 | 0.010317 | -1 | 8.37E-33 | -1 |
| 4-15985889-T-TGCTT/ | TGCTTATTTAATCTGTATTAAATATTTAATACAGATAA | | | | | | | 0.234347 | 4.52E-35 | FALSE | 4.52E-35 | ?- | 0.148634 | 1 | 0.010706 | 1 | 5.41E-33 | 1 |
| 4-17565394-C-T | T | | | | | | | 0.000813 | 6.25E-09 | TRUE | 6.25E-09 | ?+ | 0.013744 | 1 | 0.673011 | -1 | 9.43E-08 | 1 |
| 4-19483278-T-C | C | 0.000117 | 0.525112 | 0.5251 | +? | 0.000382 | 0.020852 | 0.000664 | 2.84E-08 | TRUE | 1.13E-08 | -- | 0.041455 | 1 | 0.000743 | 1 | 3.78E-05 | 1 |
| 4-21544845-C-CTTTTT | CTTTTTTT | | | | | | | 0.000849 | 2.56E-09 | TRUE | 2.56E-09 | ?+ | 0.596905 | -1 | 0.005236 | 1 | 1.29E-08 | 1 |
| 4-22509278-A-G | G | 0.000294 | 0.502829 | 0.5028 | +? | 9.71E-05 | 0.896124 | 0.001762 | 7.67E-15 | TRUE | 3.71E-14 | -- | 0.000749 | 1 | 0.292147 | 1 | 5.99E-14 | 1 |
| 4-26243074-A-AAC | AAC | | | | | | | 0.000268 | 2.49E-09 | TRUE | 2.49E-09 | ?- | 0.034286 | 1 | | | 1.48E-07 | 1 |
| 4-39359815-C-CCA | CCA | | | | | | | 0.000161 | 8.62E-09 | TRUE | 8.62E-09 | ?+ | | | | | 7.17E-07 | 1 |
| 4-39684712-G-GCC | GCC | | | | | | | 0.015751 | 7.48E-11 | TRUE | 7.48E-11 | ?- | 0.42356 | 1 | 0.333357 | 1 | 4.69E-10 | 1 |
| 4-39773277-GT-G | G | | | | | | | 0.006002 | 3.9E-22 | TRUE | 3.9E-22 | ?+ | 0.07573 | 1 | 0.044959 | 1 | 1.62E-19 | 1 |
| 4-39773298-T-TC | TC | | | | | 0.000175 | 0.937658 | 0.001228 | 1.63E-10 | TRUE | 4.28E-09 | +- | 0.044765 | 1 | 0.370548 | 1 | 8.38E-10 | 1 |
| 4-39831334-C-CAA | CAA | | | | | 1.57E-05 | 0.411968 | 0.000506 | 2.66E-08 | TRUE | 7.74E-08 | -+ | 0.17353 | 1 | 0.429254 | -1 | 1.57E-07 | 1 |
| 4-42081999-C-CCT | CCT | | | | | | | 0.001071 | 1.61E-13 | TRUE | 1.61E-13 | ?+ | 0.994441 | 1 | 0.188682 | 1 | 3.92E-15 | 1 |
| 4-49177311-T-A | A | | | | | | | 0.050466 | 3.54E-18 | FALSE | 3.54E-18 | ?+ | 3.91E-05 | 1 | 0.031439 | 1 | 2.81E-12 | 1 |
| 4-49177311-T-C | C | | | | | | | 0.912974 | 9.87E-14 | FALSE | 9.87E-14 | ?+ | 0.007432 | -1 | 0.479445 | -1 | 1.16E-11 | -1 |
| 4-49298389-T-G | G | | | | | | | 0.825621 | 2.46E-08 | FALSE | 2.46E-08 | ?+ | 0.005917 | -1 | 0.62554 | 1 | 2.07E-07 | -1 |
| 4-49308277-CT-C | C | | | | | | | 0.001395 | 5.67E-09 | TRUE | 5.67E-09 | ?- | 0.001862 | 1 | 0.012736 | 1 | 7.2E-06 | 1 |
| 4-49556193-C-G | G | | | | | 0.000132 | 0.258809 | 0.024874 | 1.75E-16 | FALSE | 9.66E-17 | -- | 0.000478 | 1 | 0.011095 | 1 | 2.99E-12 | 1 |
| 4-49556193-C-T | T | | | | | 0.974103 | 0.424299 | 0.972629 | 4.64E-18 | FALSE | 3.02E-06 | +- | 2.25E-05 | -1 | 0.005317 | -1 | 3.67E-12 | -1 |
| 4-49641607-AATGGA/ | A | | | | | | | 0.093879 | 4.11E-09 | TRUE | 4.11E-09 | ?- | 0.403436 | -1 | 0.055026 | -1 | 1.07E-07 | -1 |
| 4-49642557-T-A | A | | | | | | | 0.943715 | 7.55E-09 | FALSE | 7.55E-09 | ?+ | 0.074257 | -1 | 0.105827 | 1 | 1.38E-11 | 1 |
| 4-49642557-T-G | G | | | | | | | 0.050292 | 4.26E-11 | FALSE | 4.26E-11 | ?+ | 0.732285 | 1 | 0.072619 | -1 | 1.77E-11 | -1 |
| 4-49656214-T-A | A | | | | | | | 0.919883 | 2.36E-13 | FALSE | 2.37E-13 | ?- | 0.913699 | -1 | 0.043098 | -1 | 2.17E-11 | -1 |
| 4-56835539-GGTGGA- | G | | | | | | | 0.000743 | 6.48E-11 | TRUE | 6.48E-11 | ?+ | 0.006033 | 1 | 0.246381 | 1 | 4.03E-09 | 1 |
| 4-64037563-A-C | C | | | | | 0.397848 | 0.552586 | 0.451603 | 1.52E-09 | FALSE | 0.000124 | -- | 0.143116 | 1 | 0.373279 | -1 | 3.28E-09 | 1 |
| 4-69258307-G-GTTT | GTTT | | | | | | | 0.045441 | 2.55E-12 | FALSE | 2.55E-12 | ?- | 0.006916 | 1 | 0.067422 | 1 | 5.25E-10 | 1 |
| 4-70871593-CTG-C | C | 0.001014 | 0.334206 | 0.3342 | +? | | | 0.001675 | 4.48E-08 | TRUE | 4.48E-08 | ?- | 0.033074 | 1 | 0.014184 | 1 | 1.94E-05 | 1 |
| 4-70884321-C-CAT | CAT | | | | | | | 0.000457 | 1.19E-12 | TRUE | 1.19E-12 | ?+ | 0.000728 | 1 | 0.305026 | 1 | 1.65E-11 | 1 |
| 4-73674287-A-ACGTA | ACGTATATATATACACG | | | | | | | 0.264529 | 6.71E-10 | FALSE | 6.71E-10 | ?+ | 0.9256 | 1 | 0.490257 | -1 | 6.1E-11 | -1 |
| 4-75128448-T-C | C | | | | | | | 0.857591 | 4.91E-35 | FALSE | 4.91E-35 | ?- | 0.008779 | 1 | 0.023245 | 1 | 1.3E-30 | 1 |
| 4-75682763-C-CCT | CCT | | | | | | | 0.00037 | 1.26E-08 | TRUE | 1.27E-08 | ?+ | 0.007823 | 1 | 0.101694 | 1 | 3.56E-07 | 1 |
| 4-76667749-A-ACC | ACC | | | | | | | 0.001051 | 8.93E-11 | TRUE | 8.93E-11 | ?- | 0.02404 | 1 | 0.021304 | 1 | 4.67E-08 | 1 |
| 4-83130461-CA-C | C | 0.00045 | 0.35395 | 0.3539 | +? | 0.001005 | 0.784777 | 0.001956 | 9.91E-12 | TRUE | 2.61E-07 | +- | 0.000188 | 1 | 0.001763 | 1 | 5.21E-08 | 1 |
| 4-83575604-C-T | T | | | | | 0.001388 | 0.251024 | 0.003847 | 2.07E-21 | TRUE | 5.67E-13 | -+ | 0.000856 | 1 | 0.007312 | 1 | 1.63E-16 | 1 |
| 4-84770219-C-CG | CG | | | | | | | 0.00099 | 5.91E-09 | TRUE | 5.92E-09 | ?+ | 0.00615 | 1 | 0.018532 | 1 | 1.1E-05 | 1 |
| 4-86958333-T-C | C | 0.000215 | 0.651098 | 0.6511 | -? | | | 0.000841 | 1.58E-08 | TRUE | 1.58E-08 | ?- | 0.198931 | 1 | 0.01153 | 1 | 6.63E-07 | 1 |
| 4-96975693-T-C | C | | | | | 0.240187 | 0.564466 | 0.312114 | 9.14E-09 | FALSE | 0.004364 | +- | 0.023294 | 1 | 0.003787 | 1 | 1.93E-05 | 1 |
| 4-99086726-C-CCT | CCT | | | | | | | 0.002969 | 1.7E-09 | TRUE | 1.7E-09 | ?+ | 0.083257 | 1 | 0.918311 | 1 | 3.28E-09 | 1 |
| 4-102329460-T-TAA | TAA | 9.75E-05 | 0.597244 | 0.5972 | +? | | | 0.000784 | 4.09E-12 | TRUE | 4.09E-12 | ?- | 0.077831 | 1 | 0.008814 | 1 | 5.85E-11 | 1 |
| 4-102329468-C-CAA | CAA | | | | | | | 0.001794 | 2.19E-15 | TRUE | 2.19E-15 | ?+ | 0.669573 | 1 | 0.023164 | 1 | 2.94E-15 | 1 |
| 4-103431931-G-A | A | 0.000254 | 0.068658 | 0.06866 | +? | 0.001015 | 0.5353 | 0.001559 | 9.99E-10 | TRUE | 2.7E-05 | -+ | 0.117976 | 1 | 0.001165 | 1 | 9.81E-08 | 1 |
| 4-113231215-T-C | C | | | | | 0.000482 | 0.201588 | 0.000464 | 2.25E-09 | TRUE | 0.000655 | +- | 0.082516 | 1 | 0.003346 | 1 | 5.37E-08 | 1 |
| 4-113231218-ATT-A | A | | | | | | | 0.001165 | 3.75E-12 | TRUE | 3.75E-12 | ?+ | 0.605738 | 1 | 0.01752 | 1 | 7.15E-11 | 1 |
| 4-113924078-T-A | A | | | | | | | 0.855348 | 1.52E-08 | FALSE | 1.52E-08 | ?+ | 0.330217 | -1 | 0.075356 | 1 | 5.2E-09 | 1 |
| 4-113924088-T-A | A | | | | | | | 0.83168 | 3.63E-10 | FALSE | 3.63E-10 | ?+ | 0.526304 | -1 | 0.014801 | 1 | 1.07E-09 | 1 |
| 4-113924104-T-A | A | | | | | | | 0.825897 | 1.85E-10 | FALSE | 1.85E-10 | ?+ | 0.972282 | 1 | 0.094517 | 1 | 2.26E-10 | 1 |
| 4-113924113-A-G | G | | | | | | | 0.816177 | 2.67E-09 | FALSE | 2.67E-09 | ?- | 0.523103 | 1 | 0.058425 | 1 | 3.14E-08 | 1 |
| 4-114959008-T-A | A | | | | | | | 0.000586 | 4.95E-10 | TRUE | 4.95E-10 | ?+ | 0.022714 | 1 | 0.000879 | 1 | 2.01E-07 | 1 |
| 4-120281144-CA-C | C | | | | | 0.000915 | 0.141313 | 0.001649 | 6.19E-09 | TRUE | 6.46E-08 | -- | 0.005546 | 1 | 0.992751 | 1 | 4.36E-08 | 1 |
| 4-120327591-CTCAT-( | C | | | | | | | 0.002095 | 3.11E-10 | TRUE | 3.11E-10 | ?- | 1.82E-05 | 1 | 0.728759 | -1 | 4.49E-07 | 1 |
| 4-126101439-G-A | A | | | | | | | 0.839592 | 2.74E-08 | FALSE | 2.74E-08 | ?+ | 0.07691 | 1 | 0.934099 | -1 | 1.49E-07 | 1 |
| 4-126101439-G-GTAT, | GTATATATACTTA | | | | | | | 0.110291 | 2.19E-08 | FALSE | 2.19E-08 | ?+ | 0.040456 | -1 | 0.238395 | -1 | 7.05E-07 | -1 |
| 4-128104101-C-CAA | CAA | | | | | | | 0.000293 | 1.65E-09 | TRUE | 1.65E-09 | ?+ | 0.001534 | 1 | 0.042438 | 1 | 5.65E-07 | 1 |
| 4-128814728-A-AT | AT | | | | | 0.009303 | 0.382213 | 0.001128 | 1.29E-10 | TRUE | 0.2278 | +- | 0.011114 | 1 | 0.010645 | 1 | 3.26E-08 | 1 |
| 4-143887196-A-ATT | ATT | | | | | | | 0.854265 | 4.69E-19 | FALSE | 4.69E-19 | ?+ | 0.008894 | -1 | 0.008783 | -1 | 3.01E-16 | -1 |
| 4-143981449-C-G | G | | | | | 0.981931 | 0.066146 | 0.959578 | 2.68E-10 | FALSE | 0.01148 | +- | 0.333992 | -1 | 0.007596 | 1 | 1.88E-17 | 1 |
| 4-143981449-C-T | T | | | | | 0.008112 | 0.417478 | 0.009479 | 3.51E-14 | TRUE | 0.000205 | +- | 0.433513 | 1 | 0.030433 | -1 | 9.55E-14 | -1 |
| 4-144912122-C-T | T | 0.000469 | 0.181876 | 0.1819 | +? | | | 0.001191 | 2.17E-08 | TRUE | 2.17E-08 | ?+ | 0.229448 | 1 | 0.378687 | 1 | 1.21E-07 | 1 |

| | | | | | | | | | | | | | | | | | | | | |
|---|---|---|---|---|---|---|---|---|---|---|---|---|---|---|---|---|---|---|---|---|
| 4-144912136-C-G | G | 0.00049 | 0.665724 | | | 0.6657 | -? | 0.000163 | 0.671216 | 0.001421 | 1.57E-09 | TRUE | 3.66E-08 | +- | 0.023743 | 1 | 0.605468 | -1 | 3.31E-09 | 1 |
| 4-146728827-A-ATATA | ATATATACACATATATATGTGTATATATACG | | | | | | | | | 0.245081 | 1.8E-08 | FALSE | 1.8E-08 | ?+ | 0.012197 | -1 | 0.342959 | 1 | 1.42E-08 | -1 |
| 4-151065713-CAA-C | C | 0.000273 | 0.517932 | | | 0.5179 | +? | | | 0.001835 | 1.05E-09 | TRUE | 1.06E-09 | ?- | 0.088412 | 1 | 0.13314 | 1 | 1.32E-08 | 1 |
| 4-151299398-T-A | A | | | | | | | 0.000144 | 0.780394 | 0.000611 | 7.97E-10 | TRUE | 3.46E-08 | -+ | 0.002604 | 1 | 0.580262 | 1 | 4.15E-09 | 1 |
| 4-151526479-C-T | T | | | | | | | | | 0.030729 | 1.88E-09 | FALSE | 1.88E-09 | ?+ | 0.798154 | 1 | 0.021538 | 1 | 2.58E-08 | 1 |
| 4-153944779-C-CTCAT | CTCAT | | | | | | | | | 0.002381 | 4.25E-11 | TRUE | 4.25E-11 | ?+ | 0.50384 | 1 | 0.012611 | 1 | 1.23E-09 | 1 |
| 4-159437376-AG-A | A | | | | | | | | | 0.001114 | 6.07E-09 | TRUE | 6.07E-09 | ?+ | 3.25E-05 | 1 | 0.600636 | 1 | 4.08E-07 | 1 |
| 4-162815005-C-T | T | 0.00043 | 0.218284 | | | 0.2183 | +? | 7.83E-05 | 0.603129 | 0.001271 | 1.86E-09 | TRUE | 1.38E-08 | -+ | 0.114027 | 1 | 0.01484 | 1 | 1.08E-08 | 1 |
| 4-162815032-G-A | A | 0.000864 | 0.250529 | | | 0.2505 | +? | 4.7E-05 | 0.977475 | 0.00174 | 1.69E-09 | TRUE | 2.99E-09 | ++ | 0.683551 | 1 | 0.011986 | 1 | 3.04E-08 | 1 |
| 4-162815609-A-G | G | 0.000215 | 0.489259 | | | 0.4893 | -? | 0.001519 | 0.528117 | 0.000727 | 3.44E-09 | TRUE | 0.006637 | +- | 0.01386 | 1 | 0.000724 | 1 | 4.37E-05 | 1 |
| 4-168966338-C-CAGTG | CAGTG | 0.00044 | 0.899485 | | | 0.8995 | +? | | | 0.001851 | 3.95E-09 | TRUE | 3.95E-09 | ?+ | 0.292345 | 1 | 0.754144 | 1 | 5.3E-08 | 1 |
| 4-172464489-CCT-C | C | | | | | | | | | 8.82E-05 | 2.49E-08 | TRUE | 2.49E-08 | ?- | | | | | 2.56E-07 | 1 |
| 4-176800802-C-A | A | | | | | | | | | 0.009883 | 1.04E-09 | TRUE | 1.04E-09 | ?- | 0.090828 | -1 | 0.43211 | -1 | 1.66E-08 | -1 |
| 4-182621523-GA-G | G | | | | | | | | | 0.957731 | 1.69E-12 | TRUE | 1.7E-12 | ?- | 0.381952 | -1 | 0.86847 | -1 | 1.04E-18 | -1 |
| 4-183509969-CAG-C | C | | | | | | | | | 0.00502 | 1.11E-11 | TRUE | 1.11E-11 | ?- | 0.254081 | 1 | 0.026482 | 1 | 1.38E-10 | 1 |
| 4-183643226-T-A | A | 0.000475 | 0.354981 | | | 0.355 | -? | 0.000376 | 0.984512 | 0.000704 | 1.05E-08 | TRUE | 6.29E-06 | -+ | 0.370785 | 1 | 0.003701 | 1 | 2.75E-07 | 1 |
| 4-185129527-T-C | C | | | | | | | 0.371846 | 0.176629 | 0.48152 | 3.4E-08 | FALSE | 2.58E-05 | -- | 0.051721 | 1 | 0.200372 | 1 | 9.49E-06 | 1 |
| 4-186238862-CAGTTA | C | 0.000722 | 0.728981 | | | 0.729 | -? | | | 0.01493 | 7.24E-10 | FALSE | 7.24E-10 | ?- | 0.010919 | 1 | 0.102605 | 1 | 2.99E-07 | 1 |
| 4-187991573-T-A | A | | | | | | | | | 0.536019 | 4.35E-09 | FALSE | 4.35E-09 | ?+ | 0.035266 | 1 | 0.555939 | 1 | 5.01E-07 | 1 |
| 4-189761243-TCCAAG | T | | | | | | | | | 0.892213 | 2.98E-12 | FALSE | 2.98E-12 | ?- | 0.979118 | 1 | 0.340522 | -1 | 5.29E-15 | -1 |
| 4-189761883-A-AACTT | AACTTCTCCAAGGATCAGCTCCCAAATCAACCAGAACTTCTCCAAGGTTCAGCTCCCAAATCAGTCAGAACTTCTCC | | | | | | | | | 0.866107 | 7.31E-33 | FALSE | 7.31E-33 | ?+ | 0.000394 | -1 | 0.026356 | -1 | 2.15E-28 | -1 |
| 4-38320415-G-A | A | | | | | | | 2.51E-05 | 0.021242 | 0.000158 | 5.39E-07 | TRUE | 3.5E-08 | ++ | | | | | | |
| 4-38486000-C-A | A | | | | | | | 4.07E-05 | 0.154024 | 0.000472 | 7.32E-08 | TRUE | 2.61E-08 | ++ | 3.63E-05 | 1 | 0.102139 | 1 | 0.000296 | 1 |
| 4-79371965-G-C | C | | | | | | | 1.88E-05 | 0.375217 | | | FALSE | 8.8E-140 | -? | | | | | | |
| 5-59693193-A-G | G | 0.000468 | 2.07E-08 | | | 2.071E-08 | -? | 3.45E-05 | 0.574009 | 0.000274 | 0.084132 | TRUE | 0.07526 | -- | | | 0.007409 | 1 | | |
| 5-115784235-T-C | C | 0.000526 | 1.48E-08 | | | 1.477E-08 | -? | 0.000279 | 0.273652 | 0.001242 | 0.526741 | TRUE | 0.2683 | -- | | | 0.440841 | -1 | 0.470914 | -1 |
| 5-128619201-G-A | A | 0.010367 | 7.67E-09 | 0.013 | 0.525766 | 7.171E-09 | -- | 0.009574 | 0.702313 | 0.008296 | 0.771351 | TRUE | 0.8776 | -+ | 0.475158 | 1 | 0.024326 | -1 | 0.18486 | 1 |
| 5-158747-GT-G | G | | | | | | | | | 0.000689 | 1.31E-08 | TRUE | 1.31E-08 | ?+ | 0.000278 | 1 | 0.215554 | 1 | 1.7E-05 | 1 |
| 5-285364-G-A | A | | | | | | | | | 0.250208 | 2.78E-08 | TRUE | 2.78E-08 | ?+ | 0.027625 | 1 | 0.941552 | -1 | 0.000658 | 1 |
| 5-1043647-G-GGTTAG | GGTTAGTGTGTGAATGAATGGGTGA | | | | | | | | | 0.877553 | 1.81E-08 | FALSE | 1.81E-08 | ?+ | 0.251342 | 1 | 0.903611 | -1 | 7.06E-13 | -1 |
| 5-1047018-GGTGTGT( | G | | | | | | | | | 0.853741 | 2.29E-11 | FALSE | 2.3E-11 | ?- | 0.901882 | -1 | 0.719156 | -1 | 4.77E-15 | -1 |
| 5-3323646-G-A | A | | | | | | | | | 0.004033 | 1.3E-08 | TRUE | 1.3E-08 | ?+ | 0.602137 | -1 | 0.053103 | 1 | 3.6E-09 | 1 |
| 5-3323650-G-A | A | | | | | | | | | 0.002867 | 2.45E-09 | TRUE | 2.45E-09 | ?+ | 0.718548 | -1 | 0.754482 | 1 | 3.15E-11 | 1 |
| 5-5730755-TA-T | T | | | | | | | | | 0.001874 | 1.45E-08 | TRUE | 1.45E-08 | ?+ | 0.042028 | 1 | 0.409775 | 1 | 7.32E-08 | 1 |
| 5-7300656-C-CATCT | CATCT | | | | | | | | | 0.052813 | 2.01E-08 | FALSE | 2.01E-08 | ?- | 0.48246 | -1 | 0.044765 | -1 | 1.65E-08 | -1 |
| 5-20172242-A-G | G | | | | | | | | | 0.202666 | 4.74E-08 | FALSE | 4.74E-08 | ?- | 0.616375 | 1 | 0.036065 | 1 | 1.14E-05 | 1 |
| 5-21262005-C-CAAAA | CAAAAAAA | | | | | | | | | 0.037125 | 1.57E-08 | FALSE | 1.57E-08 | ?+ | 0.131992 | 1 | 0.104895 | -1 | 9.67E-11 | 1 |
| 5-23526774-C-CAA | CAA | | | | | | | | | 0.002914 | 4.42E-14 | TRUE | 4.43E-14 | ?+ | 0.004085 | 1 | 0.998271 | -1 | 1.45E-14 | 1 |
| 5-31919818-GCCA-G | G | 0.000216 | 0.461295 | | | 0.4613 | -? | | | 0.002043 | 9.92E-22 | TRUE | 9.93E-22 | ?+ | 0.000193 | 1 | 0.451221 | 1 | 9.38E-20 | 1 |
| 5-35944045-G-GGAA | GGAA | | | | | | | | | 0.964053 | 2.21E-08 | FALSE | 2.21E-08 | ?+ | 0.236964 | -1 | 0.453415 | -1 | 3.82E-08 | -1 |
| 5-35944045-G-GGAAT | GGAATC | | | | | | | | | 0.013389 | 4.15E-08 | FALSE | 4.15E-08 | ?- | 0.44287 | 1 | 0.499509 | 1 | 6.88E-08 | 1 |
| 5-35944046-T-C | C | | | | | | | | | 0.014633 | 4.17E-08 | FALSE | 4.17E-08 | ?- | 0.673896 | 1 | 0.209068 | 1 | 1.1E-07 | 1 |
| 5-37322460-C-A | A | | | | | | | | | 0.000169 | 1.24E-08 | TRUE | 1.24E-08 | ?+ | | | | | 5.67E-08 | 1 |
| 5-37396773-A-G | G | | | | | | | 0.0001 | 0.807775 | 0.000617 | 3.29E-08 | TRUE | 4.36E-07 | +- | 0.260258 | -1 | 0.054108 | 1 | 1.81E-08 | 1 |
| 5-37486551-T-G | G | | | | | | | 5.01E-05 | 0.018009 | 0.000438 | 9.75E-09 | TRUE | 5.52E-10 | -- | 0.000202 | 1 | 0.108498 | 1 | 4.42E-05 | 1 |
| 5-38394491-G-C | C | 9.78E-05 | 0.320003 | | | 0.32 | +? | | | 0.00072 | 1.72E-10 | TRUE | 1.72E-10 | ?+ | 0.011155 | 1 | 0.930967 | -1 | 4.78E-09 | 1 |
| 5-41237701-A-C | C | | | | | | | | | 0.005437 | 3.42E-10 | FALSE | 3.42E-10 | ?- | 0.000366 | 1 | 0.355112 | 1 | 9.8E-08 | 1 |
| 5-42706839-T-A | A | | | | | | | 0.000122 | 0.962605 | 0.000968 | 1.79E-08 | TRUE | 1.15E-07 | ++ | 0.112443 | 1 | 0.468273 | -1 | 2.5E-07 | 1 |
| 5-42954699-G-C | C | | | | | | | | | 0.000719 | 2.4E-11 | TRUE | 2.4E-11 | ?+ | 0.31463 | 1 | 0.00154 | 1 | 1.81E-08 | 1 |
| 5-42954706-A-G | G | | | | | | | | | 0.000874 | 1.21E-11 | TRUE | 1.21E-11 | ?- | 0.870239 | -1 | 0.794957 | -1 | 6.12E-11 | 1 |
| 5-42954730-C-G | G | | | | | | | 0.000135 | 0.326477 | 0.00085 | 2.98E-14 | TRUE | 9.69E-14 | -- | 0.006593 | 1 | 0.203183 | 1 | 4.99E-11 | 1 |
| 5-42954736-A-G | G | | | | | | | | | 0.000793 | 2.85E-09 | TRUE | 2.86E-09 | ?- | 0.000523 | 1 | 0.845718 | 1 | 1.68E-06 | 1 |
| 5-42982044-A-C | C | | | | | | | 0.002055 | 0.181836 | 0.003529 | 1.81E-10 | TRUE | 2.69E-08 | -- | 0.006591 | 1 | 0.198781 | 1 | 6.07E-08 | 1 |
| 5-43345773-G-A | A | | | | | | | | | 0.00038 | 2.19E-08 | TRUE | 2.19E-08 | ?+ | 0.002871 | 1 | 0.154307 | 1 | 1.55E-06 | 1 |
| 5-46397240-C-T | T | 0.000137 | 0.313873 | | | 0.3139 | +? | 4.07E-05 | 0.563522 | 0.000396 | 3.49E-09 | TRUE | 4.02E-08 | -+ | 0.001643 | 1 | 0.036691 | 1 | 1.95E-06 | 1 |
| 5-55688573-G-T | T | | | | | | | | | 0.001655 | 1.6E-09 | TRUE | 1.6E-09 | ?+ | 0.005541 | 1 | 0.006348 | 1 | 9.29E-05 | 1 |
| 5-55987899-G-GAGAT | GAGAT | | | | | | | | | 0.000301 | 2.42E-10 | TRUE | 2.42E-10 | ?- | 0.00879 | 1 | 0.460066 | -1 | 2.1E-10 | 1 |
| 5-57149535-C-T | T | 0.000217 | 0.794076 | | | 0.7941 | -? | | | 0.000339 | 1.71E-08 | TRUE | 1.71E-08 | ?+ | 0.148856 | 1 | 0.155078 | 1 | 1.59E-07 | 1 |
| 5-61953687-A-G | G | | | | | | | | | 0.000324 | 2.5E-11 | TRUE | 2.5E-11 | ?- | 0.327813 | 1 | 1.58E-06 | 1 | 1.18E-05 | 1 |

| | | | | | | | | | | | | | | | | | | | | |
|---|---|---|---|---|---|---|---|---|---|---|---|---|---|---|---|---|---|---|---|---|
| 5-63078868-G-GTC | GTC | | | | | | | | | 0.000703 | 3.07E-09 | TRUE | 3.07E-09 | ?- | 0.093059 | 1 | 0.269617 | 1 | 3.28E-08 | 1 |
| 5-65634189-GTTAGCC | G | | | | | | | | | 0.000569 | 1.35E-12 | TRUE | 1.35E-12 | ?+ | 0.025731 | 1 | 0.314989 | -1 | 7.24E-13 | 1 |
| 5-69337801-A-AT | AT | | | | | | | | | 0.000861 | 3.57E-09 | TRUE | 3.57E-09 | ?- | 0.019857 | 1 | 0.005367 | 1 | 3.07E-07 | 1 |
| 5-71297967-C-CA | CA | | | | | | | | | 0.90539 | 2.45E-09 | TRUE | 2.45E-09 | ?- | 0.412634 | -1 | 0.052541 | -1 | 5.61E-08 | -1 |
| 5-76509558-ATTTT-A | A | | | | | | | | | 0.000958 | 5.1E-12 | TRUE | 5.1E-12 | ?+ | 0.000773 | 1 | 0.340085 | 1 | 7.92E-10 | 1 |
| 5-78097383-A-G | G | | | | | | | 0.997663 | 0.330166 | 0.988845 | 1.72E-08 | TRUE | 1.42E-05 | -+ | 0.935521 | -1 | 0.889006 | -1 | 2.4E-10 | -1 |
| 5-80507561-A-G | G | | | | | | | 0.000479 | 0.233303 | 0.000369 | 3.02E-13 | TRUE | 1.26E-09 | -- | 0.021903 | 1 | 0.409154 | 1 | 1.62E-12 | 1 |
| 5-82726070-T-TCCTGC | TCCTGGCTAACAC | | | | | | | | | 0.000762 | 1.93E-09 | TRUE | 1.93E-09 | ?- | 0.01587 | 1 | 0.293604 | 1 | 5.76E-07 | 1 |
| 5-82972528-C-T | T | | | | | | | 1.88E-05 | 0.044276 | 0.000315 | 1.38E-09 | TRUE | 1.75E-10 | ++ | 0.431696 | 1 | 0.389434 | -1 | 4.83E-11 | 1 |
| 5-82973288-G-T | T | | | | | | | 8.77E-05 | 0.892244 | 0.000912 | 5.92E-14 | TRUE | 7.79E-13 | -+ | 0.281045 | -1 | 0.205164 | 1 | 1.93E-15 | 1 |
| 5-87257719-ATAATAT | A | | | | | | | | | 0.900229 | 1.52E-17 | FALSE | 1.52E-17 | ?+ | 0.561749 | 1 | 0.152001 | 1 | 9.17E-18 | 1 |
| 5-87257744-A-ACT | ACT | | | | | | | | | 0.8102 | 1.37E-22 | FALSE | 1.37E-22 | ?- | 0.76308 | -1 | 0.021401 | 1 | 2.54E-23 | 1 |
| 5-93000815-A-AC | AC | | | | | | | | | 0.001232 | 4.75E-10 | TRUE | 4.75E-10 | ?- | 0.000416 | 1 | 0.063928 | 1 | 3.27E-06 | 1 |
| 5-93000816-T-G | G | | | | | | | | | 0.000822 | 2.42E-10 | TRUE | 2.43E-10 | ?- | 0.004423 | 1 | 0.004047 | 1 | 1.41E-06 | 1 |
| 5-110793049-C-G | G | | | | | | | | | 0.076856 | 4.17E-11 | FALSE | 4.17E-11 | ?- | 0.027389 | 1 | 0.544351 | 1 | 1.44E-10 | 1 |
| 5-116698546-C-A | A | 0.001084 | 0.359567 | | | 0.3596 | +? | 9.4E-05 | 0.37322 | 0.001048 | 3.53E-08 | TRUE | 3.16E-08 | ++ | 0.009093 | 1 | 0.000816 | 1 | 3.8E-05 | 1 |
| 5-118177704-TTA-T | T | | | | | | | | | 0.108442 | 2.42E-20 | FALSE | 2.42E-20 | ?+ | 0.011864 | 1 | 0.055642 | 1 | 1.18E-19 | 1 |
| 5-119047353-T-TGA | TGA | | | | | | | | | 0.000806 | 6.76E-11 | TRUE | 6.76E-11 | ?- | 0.018893 | 1 | 0.016605 | 1 | 6.91E-09 | 1 |
| 5-131141917-CA-C | C | | | | | | | | | 0.734999 | 3.11E-08 | FALSE | 3.11E-08 | ?- | 0.435045 | 1 | 0.022925 | 1 | 9.84E-08 | 1 |
| 5-134615116-A-AGGC | AGGCG | 0.001033 | 0.247162 | | | 0.2472 | +? | | | 0.006471 | 9.99E-34 | TRUE | 9.99E-34 | ?- | 0.068565 | 1 | 4.83E-05 | 1 | 1.31E-28 | 1 |
| 5-134615122-GCAGA- | G | 0.000936 | 0.153604 | | | 0.1536 | -? | | | 0.005076 | 1.08E-22 | TRUE | 1.08E-22 | ?+ | 0.590182 | 1 | 0.003034 | 1 | 1.56E-19 | 1 |
| 5-134954841-C-A | A | | | | | | | | | 0.001325 | 1.58E-13 | TRUE | 1.58E-13 | ?+ | 0.002534 | 1 | 0.524058 | 1 | 1.04E-10 | 1 |
| 5-138104941-T-G | G | | | | | | | | | 0.002647 | 2.05E-11 | TRUE | 2.05E-11 | ?- | 0.622369 | 1 | 0.118262 | 1 | 3.52E-11 | 1 |
| 5-138214143-C-CT | CT | 0.000215 | 0.089625 | | | 0.08963 | -? | | | 0.001656 | 4.49E-09 | TRUE | 4.49E-09 | ?+ | 0.000365 | 1 | 0.631286 | 1 | 1.43E-06 | 1 |
| 5-138605888-T-G | G | | | | | | | 0.000226 | 0.82292 | 0.000553 | 2.99E-09 | TRUE | 9.64E-07 | +- | 0.010238 | 1 | 0.031335 | 1 | 3.73E-07 | 1 |
| 5-139242644-C-CCT | CCT | | | | | | | | | 0.001586 | 3.47E-14 | TRUE | 3.47E-14 | ?+ | 0.001627 | 1 | 0.311388 | 1 | 6.38E-13 | 1 |
| 5-139242645-A-G | G | | | | | | | | | 0.000577 | 5.66E-11 | TRUE | 5.66E-11 | ?- | 0.002145 | 1 | 0.109667 | 1 | 1.56E-09 | 1 |
| 5-139345867-TCCGC-T | T | | | | | | | | | 0.000975 | 5.2E-10 | TRUE | 5.2E-10 | ?+ | 0.002637 | 1 | 0.831884 | -1 | 9.72E-08 | 1 |
| 5-139345874-A-AACG | AACGG | | | | | | | | | 0.0011 | 4.45E-11 | TRUE | 4.45E-11 | ?- | 0.00501 | 1 | 0.383036 | 1 | 1.29E-08 | 1 |
| 5-139405218-T-TG | TG | | | | | | | | | 0.001039 | 1.39E-09 | TRUE | 1.39E-09 | ?- | 0.000153 | 1 | 0.750008 | 1 | 1.52E-06 | 1 |
| 5-139533514-GTGTC-( | G | | | | | | | | | 0.000812 | 1.04E-08 | TRUE | 1.04E-08 | ?+ | 0.018951 | 1 | 0.392789 | 1 | 2.26E-06 | 1 |
| 5-139533522-C-CACTT | CACTT | | | | | | | | | 0.001038 | 6.46E-09 | TRUE | 6.46E-09 | ?+ | 0.121339 | 1 | 0.4772 | 1 | 5.53E-07 | 1 |
| 5-139533525-C-CT | CT | | | | | | | | | 0.001042 | 4.62E-09 | TRUE | 4.63E-09 | ?+ | 0.312853 | 1 | 0.231196 | 1 | 1.45E-07 | 1 |
| 5-139533528-TA-T | T | | | | | | | | | 0.001048 | 4.46E-09 | TRUE | 4.46E-09 | ?+ | 0.086606 | 1 | 0.549337 | 1 | 9.67E-08 | 1 |
| 5-139702543-CTT-C | C | | | | | | | | | 0.000572 | 2.46E-08 | TRUE | 2.46E-08 | ?- | 0.015821 | 1 | 0.39703 | 1 | 3.25E-07 | 1 |
| 5-140229341-C-T | T | 0.192598 | 0.247081 | 0.199 | 0.438666 | 0.2692 | +- | 0.20757 | 0.861411 | 0.194638 | 4.47E-09 | FALSE | 0.000489 | ++ | 0.085536 | 1 | 0.0379 | 1 | 2.44E-08 | 1 |
| 5-141447364-CCTGAC | C | | | | | | | | | 0.002026 | 2.61E-17 | TRUE | 2.61E-17 | ?- | 0.001438 | 1 | 0.324662 | 1 | 1.18E-14 | 1 |
| 5-142336719-C-A | A | | | | | | | 0.000464 | 0.314347 | 0.002781 | 1.89E-24 | TRUE | 2.84E-19 | -+ | 0.004616 | 1 | 4.96E-06 | 1 | 3.26E-17 | 1 |
| 5-146186627-C-G | G | | | | | | | 0.003173 | 0.005647 | 0.001359 | 1.73E-11 | TRUE | 0.2226 | +- | 0.012847 | 1 | 0.381552 | -1 | 1.05E-10 | 1 |
| 5-146234861-CCA-C | C | | | | | | | | | 0.000829 | 1.2E-10 | TRUE | 1.2E-10 | ?- | 0.00233 | 1 | 0.247267 | 1 | 6.78E-10 | 1 |
| 5-146234864-T-TGC | TGC | | | | | | | | | 0.00078 | 8.21E-09 | TRUE | 8.21E-09 | ?- | 0.220553 | 1 | 0.476921 | 1 | 8.87E-10 | 1 |
| 5-146259296-ACT-A | A | 0.001192 | 0.712806 | | | 0.7128 | -? | | | 0.003801 | 1.64E-08 | TRUE | 1.64E-08 | ?+ | 0.640703 | -1 | 0.514288 | 1 | 6.53E-10 | 1 |
| 5-148209991-T-TACA( | TACACACGTACGTATGTATGTATAC | | | | | | | | | 0.379932 | 9.62E-13 | FALSE | 9.62E-13 | ?+ | 0.274769 | -1 | 0.120423 | -1 | 2.9E-12 | -1 |
| 5-148209997-C-CACG | CACGTACGTAT | | | | | | | | | 0.371341 | 3.25E-13 | FALSE | 3.25E-13 | ?- | 0.15063 | -1 | 0.294007 | -1 | 8.37E-13 | -1 |
| 5-157864857-G-A | A | | | | | | | 2.82E-05 | 0.911079 | 0.000519 | 1.45E-09 | TRUE | 3.8E-09 | ++ | 0.103557 | 1 | 0.108433 | 1 | 1.59E-07 | 1 |
| 5-159770584-G-A | A | | | | | | | | | 0.032519 | 9.31E-17 | FALSE | 9.31E-17 | ?+ | 0.230213 | -1 | 0.302588 | 1 | 1.18E-18 | 1 |
| 5-160138360-T-G | G | 0.000255 | 0.81588 | | | 0.8159 | -? | 0.000865 | 0.124206 | 0.001013 | 1.98E-11 | TRUE | 0.000158 | +- | 0.016375 | 1 | 0.026249 | 1 | 1.48E-10 | 1 |
| 5-160138763-T-C | C | 0.000843 | 0.437901 | | | 0.4379 | +? | 0.000996 | 0.385039 | 0.001294 | 4.15E-12 | TRUE | 1.53E-08 | -- | 0.001146 | 1 | 0.067799 | 1 | 9.18E-09 | 1 |
| 5-160140248-G-GGT | GGT | | | | | | | | | 0.000477 | 8.52E-10 | TRUE | 8.52E-10 | ?- | 0.000874 | 1 | 0.397281 | 1 | 6.77E-07 | 1 |
| 5-160257333-G-C | C | | | | | | | 0.001551 | 0.270922 | 0.00203 | 3.09E-15 | TRUE | 1.21E-10 | ++ | 0.09092 | 1 | 0.281118 | 1 | 2.11E-14 | 1 |
| 5-160257343-C-T | T | 0.000156 | 0.830606 | | | 0.8306 | -? | 0.001384 | 0.306034 | 0.001434 | 2.67E-16 | TRUE | 8.77E-11 | ++ | 0.057731 | 1 | 0.089 | 1 | 1.75E-15 | 1 |
| 5-163628927-CAG-C | C | | | | | | | | | 0.001815 | 1.47E-12 | TRUE | 1.47E-12 | ?- | 0.124325 | 1 | 0.005254 | 1 | 3.6E-11 | 1 |
| 5-163628931-A-ACG | ACG | | | | | | | | | 0.001591 | 8.11E-11 | TRUE | 8.11E-11 | ?- | 0.503576 | 1 | 0.012253 | 1 | 9.21E-10 | 1 |
| 5-168253265-C-CTAC( | CTACGCACTCATTATTTATA | | | | | | | | | 0.000263 | 6.97E-09 | TRUE | 6.97E-09 | ?+ | 0.018324 | 1 | | | 5.75E-08 | 1 |
| 5-168253657-C-T | T | 0.001331 | 0.934164 | | | 0.9342 | -? | 0.000219 | 0.23024 | 0.00105 | 3.65E-12 | TRUE | 8.02E-09 | -+ | 0.05917 | 1 | 0.006162 | 1 | 5.54E-09 | 1 |
| 5-168253667-C-G | G | | | | | | | 0.000216 | 0.239814 | 0.002041 | 5.5E-12 | TRUE | 1.32E-09 | +- | 0.13322 | 1 | 0.276836 | 1 | 3.51E-10 | 1 |
| 5-168253671-C-T | T | | | | | | | 2.51E-05 | 0.274923 | 0.000695 | 6.17E-10 | TRUE | 3.35E-10 | ++ | 0.004028 | 1 | 0.133243 | 1 | 5.55E-07 | 1 |
| 5-168726538-G-GAGA | GAGA | | | | | | | | | 0.802671 | 3.63E-12 | FALSE | 3.63E-12 | ?+ | 0.156605 | -1 | 0.056641 | -1 | 2.26E-11 | -1 |
| 5-169892234-T-C | C | | | | | | | 0.001256 | 0.165105 | 0.001062 | 3.87E-20 | TRUE | 8.88E-08 | +- | 0.025223 | 1 | 0.949394 | -1 | 3.85E-21 | 1 |

| | | | | | | | | | | | | | | | | | | | | |
|---|---|---|---|---|---|---|---|---|---|---|---|---|---|---|---|---|---|---|---|---|
| 5-176502362-CCT-C | C | 0.000664 | 0.272273 | | | 0.2723 | +? | | | 0.003247 | 4.42E-21 | TRUE | 4.42E-21 | ?- | 2.61E-06 | 1 | 0.38027 | 1 | 1.27E-17 | 1 |
| 5-176801068-CTG-C | C | | | | | | | | | 0.105733 | 8.6E-17 | FALSE | 8.6E-17 | ?- | 0.57049 | 1 | 0.052552 | 1 | 1.18E-16 | 1 |
| 5-176801068-CTGTGT | C | | | | | | | | | 0.688037 | 3.74E-11 | TRUE | 3.74E-11 | ?+ | 0.768297 | 1 | 0.252856 | -1 | 8.11E-12 | -1 |
| 5-176801430-C-CTG | CTG | | | | | | | | | 0.560713 | 3.81E-16 | FALSE | 3.81E-16 | ?- | 0.189254 | -1 | 0.291105 | -1 | 9.65E-16 | -1 |
| 5-177916737-A-AAT | AAT | | | | | | | | | 0.364112 | 4.21E-09 | FALSE | 4.21E-09 | ?- | 0.010424 | 1 | 0.192874 | -1 | 1.71E-09 | 1 |
| 5-178174212-ACC-A | A | | | | | | | | | 0.000214 | 2.73E-08 | TRUE | 2.73E-08 | ?+ | 3.02E-06 | 1 | | | 2.67E-06 | 1 |
| 5-179299353-T-C | C | | | | | | | | | 0.941599 | 1.16E-09 | FALSE | 1.16E-09 | ?- | 0.410465 | 1 | 0.295086 | 1 | 1.45E-10 | 1 |
| 5-179299356-T-C | C | | | | | | | | | 0.940283 | 7.61E-10 | FALSE | 7.61E-10 | ?- | 0.676718 | 1 | 0.221441 | 1 | 5.78E-11 | 1 |
| 5-179299363-G-A | A | | | | | | | | | 0.94113 | 1.52E-10 | FALSE | 1.52E-10 | ?+ | 0.684532 | 1 | 0.122298 | 1 | 1.53E-11 | 1 |
| 5-179299369-T-A | A | | | | | | | | | 0.940821 | 5.12E-10 | FALSE | 5.12E-10 | ?+ | 0.620069 | 1 | 0.107605 | 1 | 2.01E-10 | 1 |
| 5-179299381-C-T | T | | | | | | | | | 0.94002 | 3.35E-09 | FALSE | 3.35E-09 | ?+ | 0.621944 | 1 | 0.102081 | 1 | 2.29E-09 | 1 |
| 5-179544212-GC-G | G | | | | | | | | | 0.000726 | 3.69E-09 | TRUE | 3.69E-09 | ?+ | 3.12E-06 | 1 | 0.137047 | 1 | 1.09E-05 | 1 |
| 5-179591395-AG-A | A | | | | | | | | | 0.00154 | 1.35E-12 | TRUE | 1.35E-12 | ?+ | 0.010785 | 1 | 0.003112 | 1 | 3.5E-09 | 1 |
| 5-179691169-G-GCAC | GCACCC | | | | | | | | | 0.000418 | 8.07E-09 | TRUE | 8.07E-09 | ?- | 0.00013 | 1 | 0.012955 | 1 | 1.84E-06 | 1 |
| 5-179693332-GAGTTT | G | 0.008641 | 0.418192 | | | 0.4182 | +? | | | 0.019863 | 7.69E-14 | FALSE | 7.69E-14 | ?+ | 0.530539 | 1 | 0.266224 | 1 | 3.49E-12 | 1 |
| 5-179696510-AGG-A | A | | | | | | | | | 0.000751 | 5.7E-11 | TRUE | 5.7E-11 | ?+ | 0.000217 | 1 | 0.070991 | 1 | 1.17E-08 | 1 |
| 5-179696514-A-G | G | | | | | | | | | 0.000526 | 1.45E-09 | TRUE | 1.46E-09 | ?- | 0.00131 | 1 | 0.045002 | 1 | 1.44E-07 | 1 |
| 5-179789142-CT-C | C | | | | | | | | | 0.000637 | 2.44E-08 | TRUE | 2.44E-08 | ?- | 0.012476 | 1 | 0.454635 | 1 | 6.42E-08 | 1 |
| 5-181239860-A-C | C | | | | | | | 0.000316 | 0.932587 | 0.001393 | 2.06E-09 | TRUE | 8.91E-08 | -- | 0.00145 | 1 | 0.145939 | 1 | 4.31E-07 | 1 |
| 5-181240871-A-G | G | 0.158552 | 0.101036 | 0.082 | 0.255794 | 0.1122 | +- | | | 0.167282 | 1.22E-08 | TRUE | 1.22E-08 | ?- | 0.148101 | 1 | 0.76136 | -1 | 0.00796 | 1 |
| 5-94995312-G-A | A | | | | | | | 1.57E-05 | 0.415751 | | | FALSE | 2.7E-305 | +? | | | | | | |
| 5-112598407-C-A | A | | | | | | | 3.13E-05 | 0.501822 | 0.000112 | 0.970091 | FALSE | 3.9E-305 | -- | | | 0.338816 | 1 | 0.682587 | -1 |
| 5-163546704-C-A | A | | | | | | | 9.4E-06 | 0.322854 | | | FALSE | 4.8E-305 | +? | | | | | | |
| 5-36697832-G-A | A | | | | | | | 9.4E-06 | 0.239284 | | | FALSE | 4.13E-08 | +? | | | | | | |
| 5-50349753-C-T | T | | | | | | | 7.52E-05 | 0.036216 | 0.000492 | 0.016902 | TRUE | 1.9E-305 | +- | | | | | 0.081837 | 1 |
| 5-45698810-CAGAG-C | CAG | | | | | | | 2.51E-05 | 9.8E-05 | | | FALSE | 1.68E-09 | +? | | | | | | |
| 6-156801300-A-G | G | 0.016562 | 0.149698 | 0.026 | 0.132648 | 0.1385 | -- | 0.024 | 2.82E-08 | 0.018275 | 0.959189 | TRUE | 3.13E-06 | -- | 0.727173 | -1 | 0.524642 | 1 | 0.785116 | -1 |
| 6-30429738-AAAGAG( | A | 0.005417 | 0.619884 | | | 0.6199 | +? | 0.011063 | 4.04E-09 | 0.006476 | 0.594396 | TRUE | 1.94E-07 | -- | 0.156988 | -1 | 0.245225 | -1 | 0.961198 | -1 |
| 6-30495924-T-C | C | 0.005457 | 0.683536 | 0.01 | 0.374946 | 0.6708 | -- | 0.011225 | 4.32E-09 | 0.006556 | 0.527721 | TRUE | 1.51E-07 | ++ | 0.16674 | -1 | 0.128908 | -1 | 0.928124 | -1 |
| 6-30556837-A-G | G | 0.005477 | 0.742999 | 0.01 | 0.374946 | 0.7298 | -- | 0.011226 | 5.24E-09 | 0.006564 | 0.516941 | TRUE | 1.66E-07 | ++ | 0.166751 | -1 | 0.127525 | -1 | 0.916477 | -1 |
| 6-30610380-G-T | T | 0.005321 | 0.558057 | 0.01 | 0.374946 | 0.5464 | ++ | 0.011088 | 5.6E-09 | 0.006433 | 0.408124 | TRUE | 9.98E-08 | -- | 0.070765 | -1 | 0.255873 | -1 | 0.783339 | -1 |
| 6-30693413-G-A | A | | | 0.009 | 0.475466 | 0.4755 | ?+ | 0.011554 | 9.04E-09 | 0.006818 | 0.564594 | TRUE | 3E-07 | -- | 0.169252 | -1 | 0.412377 | -1 | 0.889341 | -1 |
| 6-30697582-G-T | T | 0.00511 | 0.967616 | 0.009 | 0.475466 | 0.9783 | -+ | 0.011455 | 6.13E-09 | 0.006607 | 0.577543 | TRUE | 2.4E-07 | -- | 0.167244 | -1 | 0.395952 | -1 | 0.885226 | -1 |
| 6-32381995-C-T | T | 0.194651 | 0.406854 | 0.191 | 0.565577 | 0.3865 | ++ | 0.219354 | 1.4E-08 | 0.191564 | 0.017077 | TRUE | 1.73E-09 | -- | 0.736865 | -1 | 0.436441 | -1 | 0.032687 | -1 |
| 6-32404138-C-T | T | 0.194832 | 0.394804 | 0.191 | 0.565577 | 0.3749 | ++ | 0.219438 | 1.52E-08 | 0.191755 | 0.01615 | TRUE | 1.73E-09 | -- | 0.781912 | -1 | 0.424844 | -1 | 0.029772 | -1 |
| 6-32413666-A-T | T | 0.202896 | 0.269669 | 0.202 | 0.60341 | 0.2559 | -- | 0.225787 | 2.82E-08 | 0.200748 | 0.024758 | TRUE | 5.28E-09 | ++ | 0.92418 | -1 | 0.390817 | -1 | 0.047588 | -1 |
| 6-32438044-T-C | C | 0.196348 | 0.732831 | 0.191 | 0.8645 | 0.7249 | -- | 0.22278 | 4E-08 | 0.19445 | 0.024994 | TRUE | 7.01E-09 | ++ | 0.603867 | -1 | 0.716434 | -1 | 0.040778 | -1 |
| 6-32461376-A-G | G | | | | | | | 0.208805 | 4.46E-09 | 0.193039 | 0.01651 | TRUE | 5.87E-10 | ++ | 0.648412 | -1 | 0.448449 | -1 | 0.034297 | -1 |
| 6-32485826-T-TA | TA | | | | | | | 0.167102 | 4.27E-09 | | | FALSE | 4.27E-09 | +? | | | | | | |
| 6-32600671-G-A | A | | | 0.136 | 0.905417 | 0.9054 | ?- | 0.184719 | 1.15E-08 | | | FALSE | 1.15E-08 | -? | | | | | | |
| 6-32601659-A-AAAAA | AAAAAC | | | | | | | 0.184289 | 4.22E-09 | | | FALSE | 4.22E-09 | +? | | | | | | |
| 6-32602534-T-G | G | 0.153555 | 0.986933 | 0.145 | 0.993675 | 0.9873 | -+ | 0.185596 | 4.33E-09 | 0.150322 | 0.073629 | TRUE | 4.34E-09 | ++ | 0.214207 | -1 | 0.556788 | -1 | 0.222593 | -1 |
| 6-32602640-A-C | C | 0.153615 | 0.978819 | 0.146 | 0.993675 | 0.9785 | ++ | 0.18563 | 3.72E-09 | 0.150304 | 0.072099 | TRUE | 3.71E-09 | ++ | 0.210986 | -1 | 0.557064 | -1 | 0.22025 | -1 |
| 6-32603181-G-T | T | 0.151522 | 0.908309 | 0.144 | 0.993675 | 0.9081 | -- | 0.186152 | 2E-09 | 0.1485 | 0.070655 | TRUE | 2.12E-09 | -- | 0.145289 | -1 | 0.585214 | -1 | 0.222147 | -1 |
| 6-32603333-G-A | A | 0.153287 | 0.991165 | 0.146 | 0.993675 | 0.9908 | -- | 0.18624 | 2.13E-09 | 0.150005 | 0.070943 | TRUE | 2.29E-09 | -- | 0.209285 | -1 | 0.558968 | -1 | 0.216987 | -1 |
| 6-32605402-G-A | A | 0.221645 | 0.203594 | | | 0.2036 | -? | 0.246781 | 3.2E-08 | 0.219288 | 0.085603 | TRUE | 3.42E-08 | -- | 0.179519 | -1 | 0.29313 | -1 | 0.338708 | -1 |
| 6-32605488-G-T | T | 0.168533 | 0.329938 | 0.154 | 0.54622 | 0.3496 | -+ | 0.195185 | 2.73E-08 | 0.167148 | 0.059196 | TRUE | 1.57E-08 | -- | 0.562712 | -1 | 0.261304 | -1 | 0.178145 | -1 |
| 6-32605638-A-G | G | 0.145241 | 0.998951 | 0.141 | 0.745672 | 0.9857 | +- | 0.184632 | 4.58E-09 | 0.142448 | 0.047919 | TRUE | 2.28E-09 | ++ | 0.073731 | -1 | 0.599097 | -1 | 0.201033 | -1 |
| 6-32606826-G-A | A | 0.158561 | 0.794428 | 0.148 | 0.867122 | 0.8026 | -+ | 0.186734 | 2.61E-09 | 0.154792 | 0.082916 | TRUE | 3.55E-09 | -- | 0.429542 | -1 | 0.545772 | -1 | 0.195431 | -1 |
| 6-32607091-C-T | T | 0.163599 | 0.913257 | 0.149 | 0.932733 | 0.9174 | +- | 0.186684 | 7.31E-09 | 0.161594 | 0.062939 | TRUE | 5.66E-09 | -- | 0.384896 | -1 | 0.476814 | -1 | 0.198429 | -1 |
| 6-32607119-G-A | A | 0.162655 | 0.907137 | | | 0.9071 | +? | 0.185583 | 1.1E-08 | 0.161606 | 0.062299 | TRUE | 7.83E-09 | -- | 0.385095 | -1 | 0.474488 | -1 | 0.1966 | -1 |
| 6-32609269-C-T | T | | | | | | | 0.222407 | 7.54E-09 | 0.21507 | 0.057127 | TRUE | 5.86E-09 | -- | 0.955234 | 1 | 0.24742 | -1 | 0.16833 | -1 |
| 6-32609603-A-G | G | 0.21813 | 0.973412 | 0.184 | 0.694121 | 0.9529 | -- | 0.210728 | 2.01E-08 | 0.208854 | 0.015745 | FALSE | 2.24E-09 | ++ | 0.373324 | -1 | 0.072859 | -1 | 0.147059 | -1 |
| 6-32609695-T-C | C | 0.218201 | 0.990809 | 0.184 | 0.694121 | 0.9886 | +- | 0.210693 | 2.32E-08 | 0.208848 | 0.015809 | FALSE | 2.55E-09 | ++ | 0.373307 | -1 | 0.073058 | -1 | 0.147056 | -1 |
| 6-32609784-ATG-A | A | 0.218156 | 0.992764 | | | 0.9928 | +? | 0.210819 | 2.88E-08 | 0.208841 | 0.016219 | FALSE | 3.18E-09 | -- | 0.375299 | -1 | 0.073227 | -1 | 0.149589 | -1 |
| 6-32610137-GGA-G | G | 0.218195 | 0.986541 | | | 0.9865 | -? | 0.210721 | 1.95E-08 | 0.208842 | 0.015943 | FALSE | 2.21E-09 | -- | 0.373244 | -1 | 0.073269 | -1 | 0.147846 | -1 |
| 6-32610196-T-G | G | 0.163867 | 0.893851 | 0.15 | 0.966666 | 0.892 | -- | 0.187124 | 5.39E-09 | 0.161589 | 0.062384 | TRUE | 4.33E-09 | ++ | 0.383148 | -1 | 0.477094 | -1 | 0.196369 | -1 |
| 6-32610813-A-G | G | 0.218181 | 0.987886 | 0.184 | 0.694121 | 0.9673 | -- | 0.210743 | 1.81E-08 | 0.208865 | 0.016309 | FALSE | 2.13E-09 | ++ | 0.371522 | -1 | 0.073205 | -1 | 0.150452 | -1 |
| 6-32610995-C-A | A | 0.156512 | 0.963218 | 0.146 | 0.921915 | 0.9585 | ++ | 0.185494 | 7.87E-09 | 0.154532 | 0.042427 | TRUE | 3.23E-09 | -- | 0.259689 | -1 | 0.438848 | -1 | 0.192689 | -1 |

| | | | | | | | | | | | | | | | | | | | | |
|---|---|---|---|---|---|---|---|---|---|---|---|---|---|---|---|---|---|---|---|---|
| 6-32612814-G-C | C | | | | | | | 0.26112 | 3.66E-08 | 0.260991 | 0.02221 | TRUE | 6.11E-09 | -- | 0.08672 | -1 | 0.078509 | -1 | 0.278639 | -1 |
| 6-32612840-T-A | A | 0.156565 | 0.996363 | 0.145 | 0.921915 | 0.9916 | ++ | 0.185579 | 8.7E-09 | 0.154696 | 0.045107 | TRUE | 3.84E-09 | -- | 0.239844 | -1 | 0.446187 | -1 | 0.200678 | -1 |
| 6-32612880-T-C | C | 0.320347 | 0.632688 | 0.28 | 0.59844 | 0.6616 | +- | 0.301659 | 1.42E-08 | 0.314494 | 0.019816 | TRUE | 2.46E-09 | ++ | 0.380707 | -1 | 0.034908 | -1 | 0.231892 | -1 |
| 6-32613134-T-C | C | 0.271432 | 0.742598 | | | 0.7426 | +? | 0.262318 | 2.28E-08 | | | FALSE | 2.28E-08 | +? | | | | | | |
| 6-32613231-T-C | C | 0.320215 | 0.647889 | 0.282 | 0.59844 | 0.6771 | +- | 0.301672 | 1.36E-08 | 0.314504 | 0.020371 | TRUE | 2.48E-09 | ++ | 0.378834 | -1 | 0.034671 | -1 | 0.237129 | -1 |
| 6-32613238-T-A | A | 0.320276 | 0.643284 | 0.281 | 0.59844 | 0.6724 | -+ | 0.301672 | 1.36E-08 | 0.314516 | 0.020571 | TRUE | 2.51E-09 | -- | 0.394718 | -1 | 0.034534 | -1 | 0.234418 | -1 |
| 6-32614412-A-C | C | 0.156525 | 0.999224 | 0.146 | 0.921915 | 0.9945 | -- | 0.185582 | 7.98E-09 | 0.154712 | 0.042248 | TRUE | 3.25E-09 | ++ | 0.238218 | -1 | 0.431319 | -1 | 0.192729 | -1 |
| 6-32614873-C-G | G | 0.163932 | 0.890788 | 0.15 | 0.966666 | 0.889 | -- | 0.185925 | 7.05E-09 | 0.16177 | 0.063393 | TRUE | 5.79E-09 | ++ | 0.358261 | -1 | 0.467098 | -1 | 0.200207 | -1 |
| 6-32615250-G-C | C | 0.16399 | 0.903806 | 0.15 | 0.966666 | 0.902 | ++ | 0.187124 | 4.01E-09 | 0.16177 | 0.066203 | TRUE | 3.72E-09 | -- | 0.358021 | -1 | 0.481845 | -1 | 0.205648 | -1 |
| 6-32615369-C-T | T | 0.343852 | 0.945168 | 0.301 | 0.279268 | 0.8795 | ++ | 0.309069 | 5.18E-09 | 0.33478 | 0.011798 | TRUE | 5.43E-10 | -- | 0.472141 | -1 | 0.089726 | -1 | 0.099947 | -1 |
| 6-32615429-G-A | A | 0.163997 | 0.919983 | 0.15 | 0.966666 | 0.9181 | ++ | 0.187143 | 5.09E-09 | 0.161768 | 0.063042 | TRUE | 4.2E-09 | -- | 0.357815 | -1 | 0.466556 | -1 | 0.1994 | -1 |
| 6-32616142-T-C | C | 0.295319 | 0.75615 | 0.254 | 0.133678 | 0.6769 | -- | 0.269724 | 1.48E-08 | 0.281327 | 0.01524 | TRUE | 1.81E-09 | ++ | 0.137363 | -1 | 0.180189 | -1 | 0.132395 | -1 |
| 6-32616202-A-G | G | 0.343608 | 0.901558 | 0.301 | 0.279268 | 0.8365 | -- | 0.309082 | 6.01E-09 | 0.334785 | 0.012006 | TRUE | 6.31E-10 | ++ | 0.472464 | -1 | 0.089578 | -1 | 0.10148 | -1 |
| 6-32616536-C-G | G | | | | | | | 0.307703 | 7.01E-09 | 0.340883 | 0.018483 | TRUE | 1.22E-09 | ++ | 0.585491 | -1 | 0.11928 | -1 | 0.110214 | -1 |
| 6-32617117-G-T | T | | | | | | | 0.18631 | 5.27E-09 | 0.1618 | 0.062985 | TRUE | 4.3E-09 | -- | 0.364341 | -1 | 0.464829 | -1 | 0.199405 | -1 |
| 6-32622864-T-G | G | | | | | | | 0.199452 | 2.07E-08 | 0.172353 | 0.052096 | TRUE | 1.02E-08 | ++ | 0.35822 | -1 | 0.864221 | -1 | 0.094395 | -1 |
| 6-32623007-T-TACAG | TACAG | 0.172098 | 0.435228 | | | 0.4352 | -? | 0.200367 | 1.77E-08 | 0.183624 | 0.081176 | TRUE | 1.61E-08 | ++ | 0.413133 | -1 | 0.938122 | -1 | 0.116387 | -1 |
| 6-32623436-A-G | G | 0.164315 | 0.909954 | 0.152 | 0.934526 | 0.914 | -+ | 0.187269 | 4.48E-09 | 0.166066 | 0.061802 | TRUE | 3.6E-09 | ++ | 0.370632 | -1 | 0.471268 | -1 | 0.185831 | -1 |
| 6-32624232-A-G | G | 0.146722 | 0.844312 | 0.142 | 0.914061 | 0.8496 | +- | 0.184918 | 8.1E-09 | 0.174518 | 0.063677 | FALSE | 1.34E-08 | ++ | 0.152618 | -1 | 0.431369 | -1 | 0.248412 | -1 |
| 6-32629245-G-A | A | 0.164867 | 0.938147 | 0.151 | 0.988343 | 0.9376 | -- | 0.186373 | 7.51E-09 | 0.162508 | 0.059126 | TRUE | 5.28E-09 | -- | 0.298618 | -1 | 0.436674 | -1 | 0.206903 | -1 |
| 6-32631077-G-A | A | 0.165293 | 0.864626 | 0.151 | 0.988343 | 0.8642 | -- | 0.191208 | 1.97E-08 | 0.162499 | 0.058436 | TRUE | 1.1E-08 | -- | 0.296235 | -1 | 0.445167 | -1 | 0.203166 | -1 |
| 6-32631386-C-T | T | 0.164822 | 0.94008 | 0.151 | 0.988343 | 0.9395 | -- | 0.187228 | 6.54E-09 | 0.162477 | 0.058307 | TRUE | 4.63E-09 | -- | 0.29857 | -1 | 0.444821 | -1 | 0.202515 | -1 |
| 6-32634360-C-G | G | 0.173385 | 0.555036 | 0.158 | 0.790874 | 0.5671 | -+ | 0.200364 | 1.86E-08 | 0.172996 | 0.048707 | TRUE | 8.51E-09 | ++ | 0.309326 | -1 | 0.844934 | -1 | 0.094198 | -1 |
| 6-32634784-C-T | T | | | | | | | 0.186338 | 8.56E-09 | 0.162471 | 0.058081 | TRUE | 5.74E-09 | -- | 0.298461 | -1 | 0.443833 | -1 | 0.201919 | -1 |
| 6-32636679-G-C | C | 0.174121 | 0.612204 | 0.157 | 0.822681 | 0.6229 | +- | 0.200681 | 1.85E-08 | 0.173529 | 0.049281 | TRUE | 8.56E-09 | -- | 0.249354 | -1 | 0.903178 | -1 | 0.094973 | -1 |
| 6-32636808-A-T | T | 0.174597 | 0.486762 | | | 0.4868 | -? | 0.199989 | 1.61E-08 | 0.184505 | 0.036747 | TRUE | 4.72E-09 | ++ | 0.326205 | -1 | 0.737928 | -1 | 0.065556 | -1 |
| 6-32643690-A-C | C | | | | | | | 0.181789 | 7.42E-09 | | | FALSE | 7.42E-09 | +? | | | | | | |
| 6-3003817-A-AGCCCT | AGCCCTTAT | | | | | | | | | 0.032831 | 2.24E-13 | FALSE | 2.24E-13 | ?- | 1.53E-05 | 1 | 0.336724 | 1 | 6.31E-10 | 1 |
| 6-4695703-C-T | T | | | | | | | 0.990804 | 0.41591 | 0.984134 | 1.14E-08 | TRUE | 0.001588 | +- | 0.400713 | -1 | 0.010605 | -1 | 3.97E-08 | -1 |
| 6-6716029-C-CTT | CTT | | | | | | | | | 0.001397 | 1.25E-10 | TRUE | 1.25E-10 | ?+ | 0.001498 | 1 | 0.126004 | 1 | 1.32E-08 | 1 |
| 6-7055607-G-A | A | 0.249951 | 0.388384 | 0.152 | 0.85341 | 0.3832 | ++ | | | 0.236564 | 2.58E-08 | TRUE | 2.58E-08 | ?+ | 0.005824 | 1 | 0.534644 | -1 | 2.24E-06 | 1 |
| 6-7382355-T-TG | TG | 0.000529 | 0.346572 | | | 0.3466 | +? | | | 0.001697 | 2.25E-10 | TRUE | 2.25E-10 | ?- | 0.773093 | -1 | 0.01562 | 1 | 7.44E-10 | 1 |
| 6-10786446-A-G | G | | | | | | | | | 0.00095 | 8.4E-09 | TRUE | 8.4E-09 | ?- | 0.020678 | 1 | 0.024531 | 1 | 7.65E-08 | 1 |
| 6-10810629-A-ACGCC | ACGCC | | | | | | | | | 0.000247 | 1.01E-08 | TRUE | 1.01E-08 | ?- | 0.566148 | 1 | | | 4.97E-08 | 1 |
| 6-11192289-C-CA | CA | | | | | | | 0.012318 | 0.157323 | 0.005842 | 1.84E-08 | TRUE | 0.3904 | +- | 0.157736 | -1 | 0.090609 | -1 | 2.88E-07 | -1 |
| 6-11616040-TAAAGA | T | | | | | | | | | 0.172876 | 1.67E-09 | FALSE | 1.67E-09 | ?- | 0.920902 | 1 | 0.019578 | -1 | 3.2E-08 | -1 |
| 6-12042382-C-CGG | CGG | | | | | | | | | 0.008428 | 1.46E-10 | TRUE | 1.46E-10 | ?+ | 0.056994 | 1 | 0.114354 | 1 | 6.55E-10 | 1 |
| 6-12840142-TC-T | T | | | | | | | | | 0.00121 | 2E-14 | TRUE | 2E-14 | ?+ | 0.478918 | 1 | 0.000331 | 1 | 1.47E-11 | 1 |
| 6-14817512-C-A | A | 0.000682 | 0.499422 | | | 0.4994 | -? | 5.33E-05 | 0.38453 | 0.001398 | 1.27E-08 | TRUE | 8.23E-08 | -+ | 0.014073 | 1 | 0.700908 | -1 | 8.3E-07 | 1 |
| 6-14817518-A-G | G | 0.000624 | 0.45354 | | | 0.4535 | +? | 6.58E-05 | 0.254201 | 0.001237 | 5.1E-12 | TRUE | 1.38E-10 | +- | 0.011536 | 1 | 0.245939 | -1 | 4.46E-11 | 1 |
| 6-14817529-G-GCA | GCA | 0.000409 | 0.723442 | | | 0.7234 | +? | | | 0.001146 | 9.18E-14 | TRUE | 9.18E-14 | ?- | 0.001078 | 1 | 0.340708 | 1 | 1.37E-10 | 1 |
| 6-14817530-AGG-A | A | 0.00041 | 0.721497 | | | 0.7215 | -? | | | 0.001095 | 7.49E-12 | TRUE | 7.49E-12 | ?+ | 0.000141 | 1 | 0.648097 | 1 | 9.62E-08 | 1 |
| 6-14989159-CGTG-C | C | | | | | | | | | 0.000974 | 9.18E-10 | TRUE | 9.18E-10 | ?- | 0.604319 | -1 | 0.855936 | 1 | 3.45E-10 | 1 |
| 6-15784875-G-A | A | | | | | | | 0.002631 | 0.342444 | 0.00166 | 1.16E-14 | TRUE | 6.93E-08 | ++ | 0.001004 | 1 | 0.015515 | 1 | 5.21E-12 | 1 |
| 6-16267227-T-C | C | | | | | | | 0.0013 | 0.131139 | 0.002174 | 4.15E-12 | TRUE | 5.47E-10 | -- | 0.028631 | 1 | 0.005953 | 1 | 6.73E-10 | 1 |
| 6-16267232-T-C | C | | | | | | | 0.0013 | 0.131162 | 0.002617 | 1.08E-12 | TRUE | 8.42E-11 | -- | 0.001586 | 1 | 0.050219 | 1 | 1.24E-10 | 1 |
| 6-21002980-C-CT | CT | 0.000137 | 0.545416 | | | 0.5454 | +? | | | 0.002857 | 4.83E-18 | TRUE | 4.83E-18 | ?+ | 0.274975 | 1 | 0.46787 | 1 | 1.91E-19 | 1 |
| 6-25902999-AAG-A | A | | | | | | | | | 0.000205 | 4.35E-08 | TRUE | 4.35E-08 | ?+ | | | | | 2.72E-08 | 1 |
| 6-26311267-C-CAACA | CAACAACACA | | | | | | | | | 0.000661 | 3E-08 | TRUE | 3.01E-08 | ?+ | 0.027549 | 1 | 0.96163 | -1 | 2.54E-08 | 1 |
| 6-27577093-C-CTT | CTT | | | | | | | | | 0.001925 | 2.92E-10 | TRUE | 2.92E-10 | ?+ | 0.132714 | 1 | 0.658053 | 1 | 1.81E-08 | 1 |
| 6-30621247-C-CAAA | CAAA | | | | | | | | | 0.000819 | 1.18E-12 | TRUE | 1.18E-12 | ?+ | 0.028394 | 1 | 0.005105 | 1 | 6.98E-09 | 1 |
| 6-30753762-GCA-G | G | | | | | | | | | 0.001011 | 3.22E-09 | TRUE | 3.22E-09 | ?+ | 0.047977 | 1 | 0.464741 | 1 | 5.55E-07 | 1 |
| 6-30753765-A-AGG | AGG | | | | | | | | | 0.001068 | 4.67E-09 | TRUE | 4.67E-09 | ?- | 0.012789 | 1 | 0.053284 | 1 | 3.45E-06 | 1 |
| 6-31266474-T-A | A | 0.025796 | 0.595875 | 0.017 | 0.120888 | 0.6162 | -+ | 0.006811 | 0.640408 | 0.006952 | 2.36E-10 | FALSE | 0.000664 | -+ | 0.00189 | 1 | 0.932962 | -1 | 6.28E-10 | 1 |
| 6-31808780-CCT-C | C | 0.000117 | 0.763178 | | | 0.7632 | -? | | | 0.0015 | 7.16E-13 | TRUE | 7.16E-13 | ?- | 0.137488 | 1 | 0.000351 | 1 | 5.03E-11 | 1 |
| 6-32446254-A-G | G | 0.000313 | 0.893566 | | | 0.8936 | -? | 0.000229 | 0.842475 | 0.000479 | 2.27E-08 | TRUE | 1.19E-05 | +- | 0.681729 | 1 | 0.001867 | 1 | 9.41E-06 | 1 |
| 6-34044161-T-TAC | TAC | | | | | | | | | 0.276171 | 3.09E-08 | TRUE | 3.09E-08 | ?- | 0.111927 | 1 | 0.671794 | 1 | 1.76E-05 | 1 |
| 6-34044171-T-TAGAC | TAGAC | | | | | | | 0.000116 | 0.282995 | 0.261466 | 1.27E-11 | FALSE | 1.02E-11 | -- | 0.105021 | 1 | 0.204983 | 1 | 2.07E-06 | 1 |
| 6-34212899-TTTTG-T | T | 0.000391 | 0.79046 | | | 0.7905 | +? | | | 0.003873 | 2.97E-29 | TRUE | 2.97E-29 | ?+ | 0.000561 | 1 | 0.247261 | 1 | 1.44E-27 | 1 |

| | | | | | | | | | | | | | | | | | | | | |
|---|---|---|---|---|---|---|---|---|---|---|---|---|---|---|---|---|---|---|---|---|
| 6-34702668-G-A | A | 0.000117 | 0.421958 | | | 0.422 | +? | 0.000439 | 0.167082 | 0.001589 | 6.63E-14 | TRUE | 6.58E-09 | -+ | 0.080129 | 1 | 0.001341 | 1 | 3.55E-11 | 1 |
| 6-34764875-C-G | G | | | | | | | 0.000583 | 0.657242 | 0.000553 | 1.47E-11 | TRUE | 3.45E-07 | -- | 0.009341 | 1 | 0.033638 | 1 | 6.94E-10 | 1 |
| 6-41604777-T-C | C | 0.000313 | 0.076669 | | | 0.07667 | -? | 0.000908 | 0.948278 | 0.001679 | 9.29E-13 | TRUE | 2.14E-08 | -- | 9.2E-05 | 1 | 0.057343 | 1 | 9.71E-09 | 1 |
| 6-41604785-G-A | A | 0.000195 | 0.665795 | | | 0.6658 | -? | 0.000163 | 0.695541 | 0.001346 | 1.72E-08 | TRUE | 2.87E-07 | -+ | 0.202618 | 1 | 0.012993 | 1 | 5.12E-07 | 1 |
| 6-41604799-A-G | G | 0.003141 | 0.028301 | | | 0.0283 | -? | 0.001328 | 0.632177 | 0.003967 | 8.05E-10 | TRUE | 2.16E-06 | +- | 0.000311 | 1 | 0.126695 | 1 | 8.65E-07 | 1 |
| 6-41604818-A-C | C | 0.003121 | 0.283734 | | | 0.2837 | -? | 0.007456 | 0.712866 | 0.008018 | 6.64E-13 | TRUE | 9.99E-06 | +- | 0.001143 | 1 | 0.000572 | 1 | 5.18E-08 | 1 |
| 6-41918284-ACT-A | A | 0.002715 | 0.885103 | | | 0.8851 | +? | 0.000589 | 0.755137 | 0.004379 | 7.45E-13 | TRUE | 1.41E-10 | -+ | 0.549098 | 1 | 0.148422 | 1 | 1E-11 | 1 |
| 6-42018709-T-A | A | | | | | | | 0.000454 | 0.720039 | 0.001467 | 1.82E-14 | TRUE | 1.67E-10 | -+ | 0.001271 | 1 | 0.003566 | 1 | 2.25E-10 | 1 |
| 6-43022229-GC-G | G | 0.000234 | 0.373657 | | | 0.3737 | -? | 0.001419 | 0.863369 | 0.002124 | 4.94E-11 | TRUE | 2.01E-06 | -+ | 0.685117 | 1 | 0.060519 | 1 | 5.02E-10 | 1 |
| 6-43022821-T-C | C | | | | | | | 0.013043 | 0.362499 | 0.00562 | 4.81E-10 | TRUE | 0.02931 | +- | 0.014515 | 1 | 0.000211 | 1 | 4.18E-06 | 1 |
| 6-43022827-T-C | C | 0.002776 | 0.007352 | | | 0.007352 | -? | 0.013043 | 0.362499 | 0.006063 | 3.48E-09 | TRUE | 0.0366 | +- | 0.168533 | 1 | 0.013868 | 1 | 2.89E-07 | 1 |
| 6-43165282-A-G | G | 0.23365 | 0.239278 | 0.163 | 0.042169 | 0.2933 | +- | 0.138977 | 0.051187 | 0.232378 | 1.68E-08 | FALSE | 7.49E-07 | -- | 0.05667 | 1 | 0.321673 | 1 | 0.001044 | 1 |
| 6-43329675-A-G | G | 0.015463 | 0.120292 | | | 0.1203 | -? | 0.997914 | 0.995788 | 0.977716 | 1.76E-12 | TRUE | 1.65E-10 | ++ | 0.749271 | -1 | 0.000537 | -1 | 1.01E-15 | -1 |
| 6-43534819-G-GTA | GTA | | | | | | | | | 0.000929 | 1.05E-08 | TRUE | 1.05E-08 | ?- | 0.011647 | 1 | 0.011393 | 1 | 1.19E-06 | 1 |
| 6-43738216-C-T | T | | | | | | | | | 0.022661 | 1.24E-16 | TRUE | 1.24E-16 | ?+ | 0.001212 | 1 | 0.014121 | 1 | 1.34E-13 | 1 |
| 6-61313986-G-T | T | | | | | | | 0.000373 | 0.755228 | 0.001251 | 4.48E-08 | TRUE | 1.49E-06 | ++ | 0.123125 | 1 | 0.149906 | 1 | 1.61E-07 | 1 |
| 6-66722870-T-G | G | | | | | | | | | 0.000504 | 4.45E-10 | TRUE | 4.45E-10 | ?- | 0.212456 | 1 | 0.000434 | 1 | 1.46E-06 | 1 |
| 6-73319458-CAA-C | C | | | | | | | | | 0.002887 | 2.65E-13 | TRUE | 2.65E-13 | ?- | 0.138309 | 1 | 0.014512 | 1 | 5.47E-12 | 1 |
| 6-73319481-G-A | A | | | | | | | 6.26E-05 | 0.197459 | 0.000707 | 1.07E-09 | TRUE | 4.82E-08 | -+ | 0.09553 | 1 | 0.666076 | 1 | 2.6E-09 | 1 |
| 6-87750990-CG-C | C | | | | | | | | | 0.024892 | 3.37E-11 | FALSE | 3.37E-11 | ?- | 0.867457 | 1 | 0.238604 | 1 | 9.09E-13 | 1 |
| 6-87750990-CGGGCA( | C | | | | | | | | | 0.806348 | 2.98E-08 | FALSE | 2.98E-08 | ?+ | 0.723296 | -1 | 0.098109 | -1 | 1.91E-09 | -1 |
| 6-88001571-G-GT | GT | | | | | | | | | 0.000434 | 1.89E-08 | TRUE | 1.89E-08 | ?- | 0.905734 | 1 | 0.054231 | 1 | 3.14E-08 | 1 |
| 6-101494698-A-AGGC | AGGCG | | | | | | | | | 0.00101 | 2.2E-08 | TRUE | 2.2E-08 | ?- | 0.019737 | 1 | 0.873585 | 1 | 2.24E-08 | 1 |
| 6-103320522-A-G | G | 0.000234 | 0.435109 | | | 0.4351 | -? | 0.000647 | 0.284396 | 0.001066 | 4.61E-12 | TRUE | 4.67E-07 | +- | 0.005877 | 1 | 0.983774 | 1 | 2.01E-10 | 1 |
| 6-103320550-GGAGG( | G | | | | | | | 0.000623 | 0.450862 | 0.001388 | 8.76E-13 | TRUE | 1.48E-08 | -+ | 0.006515 | 1 | 0.317809 | 1 | 1.67E-10 | 1 |
| 6-104062626-C-T | T | | | | | | | | | 0.003537 | 8.66E-11 | TRUE | 8.66E-11 | ?- | 0.112106 | -1 | 0.95249 | -1 | 2.27E-11 | -1 |
| 6-106480620-TCG-T | T | 0.005316 | 0.001142 | | | 0.001142 | -? | | | 0.001215 | 3.7E-09 | TRUE | 3.7E-09 | ?+ | 0.637131 | 1 | 0.025588 | 1 | 3.99E-08 | 1 |
| 6-106480624-A-ATG | ATG | | | | | | | | | 0.001207 | 2.94E-08 | TRUE | 2.94E-08 | ?- | 0.550212 | 1 | 0.014121 | 1 | 3.73E-07 | 1 |
| 6-118716962-ATG-A | A | | | | | | | | | 0.000952 | 3.64E-15 | TRUE | 3.64E-15 | ?+ | 0.013251 | 1 | 0.000126 | 1 | 2.03E-11 | 1 |
| 6-122138733-G-T | T | | | | | | | 0.000257 | 0.195508 | 0.002513 | 2.96E-14 | TRUE | 1.94E-11 | -+ | 4.29E-05 | 1 | 0.090517 | 1 | 2.67E-11 | 1 |
| 6-122139207-T-G | G | | | | | | | | | 0.001177 | 1.19E-09 | TRUE | 1.19E-09 | ?- | 0.215543 | 1 | 3.02E-05 | 1 | 2.67E-06 | 1 |
| 6-123729913-G-A | A | | | | | | | 0.000119 | 0.01386 | 0.000985 | 8.72E-09 | TRUE | 7.64E-07 | -+ | 0.092736 | 1 | 0.881896 | 1 | 1.62E-09 | 1 |
| 6-139484262-CAT-C | C | | | | | | | | | 0.000542 | 7.68E-13 | TRUE | 7.69E-13 | ?- | 0.02025 | 1 | 0.073244 | 1 | 3.56E-10 | 1 |
| 6-139484265-G-GGC | GGC | | | | | | | | | 0.000549 | 3.15E-13 | TRUE | 3.15E-13 | ?- | 0.002517 | 1 | 0.062083 | 1 | 6.34E-10 | 1 |
| 6-143890911-C-CTT | CTT | | | | | | | | | 0.001209 | 4.73E-09 | TRUE | 4.73E-09 | ?+ | 0.942561 | 1 | 0.223469 | 1 | 1.94E-09 | 1 |
| 6-148303578-CTT-C | C | | | | | | | | | 0.001735 | 1.08E-18 | TRUE | 1.09E-18 | ?- | 0.008788 | 1 | 0.369883 | 1 | 1.53E-16 | 1 |
| 6-151261625-C-G | G | | | | | | | 0.000188 | 0.440187 | 0.00083 | 2.64E-09 | TRUE | 1E-08 | -- | 0.732914 | 1 | 0.175739 | 1 | 2.31E-07 | 1 |
| 6-155397325-A-G | G | | | | | | | 0.002678 | 0.793851 | 0.00119 | 4.8E-15 | TRUE | 4.92E-05 | +- | 0.000139 | 1 | 0.058653 | 1 | 3.81E-11 | 1 |
| 6-156288006-G-T | T | 9.79E-05 | 0.588575 | | | 0.5886 | -? | 0.000135 | 0.516708 | 0.000188 | 1.79E-08 | TRUE | 2.73E-05 | -+ | | | | | 2.18E-05 | 1 |
| 6-157311140-CAGTGT | C | | | | | | | | | 0.120105 | 4.69E-11 | FALSE | 4.69E-11 | ?- | 0.134956 | 1 | 0.122626 | 1 | 8.38E-10 | 1 |
| 6-158349266-G-A | A | | | | | | | | | 0.834163 | 4.12E-10 | FALSE | 4.12E-10 | ?- | 0.036683 | -1 | 0.502196 | -1 | 4.97E-09 | -1 |
| 6-160841199-GTC-G | G | | | | | | | 0.566642 | 0.053381 | 0.664124 | 5.96E-10 | FALSE | 1.1E-06 | ++ | 0.988813 | -1 | 0.596847 | 1 | 9.39E-09 | 1 |
| 6-160841575-C-A | A | 0.351918 | 0.225481 | 0.418 | 0.659332 | 0.2134 | -- | 0.566595 | 0.049869 | 0.637978 | 1.82E-08 | FALSE | 2.15E-06 | ++ | 0.907279 | 1 | 0.443369 | 1 | 3.73E-07 | 1 |
| 6-160842000-A-G | G | | | | | | | 0.566855 | 0.061218 | 0.672708 | 4.24E-10 | FALSE | 1.63E-06 | -- | 0.969783 | 1 | 0.393891 | 1 | 5.98E-09 | 1 |
| 6-160847786-G-A | A | | | 0.421 | 0.659332 | 0.6593 | ?- | 0.566649 | 0.05981 | 0.673301 | 2.14E-08 | FALSE | 7.73E-06 | ++ | 0.774483 | 1 | 0.56288 | 1 | 2.42E-07 | 1 |
| 6-160852544-G-A | A | | | 0.419 | 0.659332 | 0.6593 | ?- | | | 0.652449 | 5.99E-11 | FALSE | 5.99E-11 | ?+ | 0.790584 | 1 | 0.238947 | 1 | 2.83E-09 | 1 |
| 6-160857478-G-GT | GT | | | | | | | | | 0.184413 | 2.01E-08 | FALSE | 2.01E-08 | ?+ | 0.148277 | -1 | 0.097749 | -1 | 7.18E-08 | -1 |
| 6-160858235-A-G | G | | | | | | | | | 0.422698 | 3.5E-09 | FALSE | 3.5E-09 | ?- | 0.523821 | 1 | 0.168413 | 1 | 1.95E-07 | 1 |
| 6-165408947-C-CCT | CCT | 0.003133 | 0.101866 | | | 0.1019 | -? | | | 0.002341 | 4.97E-09 | TRUE | 4.97E-09 | ?+ | 0.421651 | 1 | 0.853732 | -1 | 8.87E-10 | 1 |
| 6-168892201-GGGGC/ | G | | | | | | | | | 0.214458 | 1.98E-14 | TRUE | 1.98E-14 | ?+ | 0.016467 | 1 | 0.170178 | 1 | 3.17E-11 | 1 |
| 6-169372164-G-A | A | | | | | | | | | 0.948284 | 9.27E-22 | FALSE | 9.27E-22 | ?- | 0.174279 | -1 | 3.18E-05 | -1 | 2.8E-17 | -1 |
| 6-169781694-T-G | G | | | | | | | 0.001347 | 0.149375 | 0.000546 | 1.25E-08 | TRUE | 0.0735 | +- | 0.78788 | 1 | 0.000584 | 1 | 4.32E-07 | 1 |
| 6-169781721-G-T | T | | | | | | | 0.421994 | 0.972461 | 0.636569 | 1.36E-08 | FALSE | 0.002747 | -+ | 0.809328 | -1 | 0.951576 | -1 | 5.03E-05 | 1 |
| 6-170054847-TC-T | T | | | | | | | 2.19E-05 | 0.941482 | 0.00413 | 1.08E-14 | TRUE | 1.26E-14 | ++ | 0.459132 | 1 | 0.240634 | 1 | 1.9E-15 | 1 |
| 6-32604622-GT-G | G | | | | | | | | | | | TRUE | | | | | | | | |
| 6-77499064-C-T | T | 0.000234 | 0.285677 | | | 0.2857 | +? | 0.000175 | 0.423388 | 0.000102 | 0.29787 | TRUE | 2.9E-131 | -- | | | | | 0.349934 | -1 |
| 6-145398859-C-CAA | CAA | | | | | | | 0.000761 | 0.591018 | 0.004263 | 0.311943 | TRUE | 0.6638 | -+ | 0.364373 | 1 | 0.547268 | -1 | 0.267065 | 1 |
| 6-32369285-AT-A | A | | | | | | | | | 0.000113 | 0.496757 | TRUE | 0.4968 | ?- | 0.523805 | -1 | | | | |
| 6-77863513-C-T | T | | | | | | | 6.58E-05 | 0.190516 | | | FALSE | 2.3E-305 | +? | | | | | | |

| | | | | | | | | | | | | | | | | | | | | |
|---|---|---|---|---|---|---|---|---|---|---|---|---|---|---|---|---|---|---|---|---|
| 6-32698518-G-A | A | 0.180126 | 0.071936 | 0.13 | 0.78038 | 0.0699 | ++ | 0.129622 | 7.07E-07 | 0.159217 | 0.011738 | FALSE | 4.58E-08 | ++ | 0.044116 | -1 | 0.026691 | -1 | 0.25212 | -1 |
| 6-32714360-G-A | A | 0.126783 | 0.220628 | 0.093 | 0.361854 | 0.204 | ++ | 0.102529 | 1.82E-07 | 0.115995 | 0.032931 | TRUE | 4.74E-08 | ++ | 0.058777 | -1 | 0.36776 | -1 | 0.249851 | -1 |
| 6-32659444-G-A | A | 0.180197 | 0.064397 | | | 0.0644 | +? | 0.1284 | 2.41E-07 | 0.159344 | 0.011737 | FALSE | 1.86E-08 | ++ | 0.043416 | -1 | 0.028698 | -1 | 0.256781 | -1 |
| 6-32699122-G-A | A | 0.192988 | 0.070474 | 0.136 | 0.594368 | 0.06631 | ++ | 0.13293 | 6.34E-07 | 0.17183 | 0.012315 | FALSE | 4.71E-08 | ++ | 0.32768 | -1 | 0.0234 | -1 | 0.171592 | -1 |
| 6-38428267-G-C | C | | | | | | | 5.64E-05 | 2.93E-05 | 0.000744 | 6.37E-05 | TRUE | 3.38E-08 | ++ | 0.000107 | 1 | 0.072136 | 1 | | |
| 6-32325821-T-A | A | 0.392261 | 0.108769 | 0.401 | 0.443694 | 0.1237 | -+ | 0.377946 | 1.09E-06 | 0.377571 | 0.005101 | FALSE | 2.17E-08 | ++ | 0.587525 | -1 | 0.422639 | -1 | 0.019118 | -1 |
| 6-62897569-G-A | A | 0.001754 | 0.096926 | 0.003 | 0.182422 | 0.09527 | ++ | 0.001623 | 0.306421 | 0.001917 | 0.620998 | TRUE | 2.1E-202 | ++ | 0.271181 | -1 | 0.378366 | 1 | 0.755497 | 1 |
| 6-60731266-G-A | A | | | | | | | 1.88E-05 | 0.229798 | | | FALSE | 7.57E-36 | -? | | | | | | |
| 6-40659693-C-T | T | | | | | | | 2.82E-05 | 0.057073 | 8.78E-05 | 0.311985 | TRUE | 1.4E-305 | -- | | | | | 0.220392 | 1 |
| 6-23348508-GA-G | G | | | | | | | | | | | FALSE | | | | | | | | |
| 7-155194021-T-C | C | 0.015783 | 1.62E-08 | | | 1.618E-08 | +? | 0.000116 | 0.85764 | 0.008982 | 0.650594 | TRUE | 0.6294 | ++ | 0.469942 | -1 | 0.634362 | -1 | 0.319155 | -1 |
| 7-100276574-T-A | A | 0.136977 | 0.000938 | 0.201 | 0.203588 | 0.0007087 | -- | 0.246595 | 2.56E-08 | 0.16136 | 0.790793 | TRUE | 6.52E-06 | -+ | 0.972563 | 1 | 0.529187 | 1 | 0.897134 | 1 |
| 7-100295525-C-G | G | 0.27551 | 0.000781 | 0.229 | 0.041093 | 0.0004768 | -- | 0.733296 | 9.1E-09 | 0.74108 | 0.991055 | FALSE | 2.3E-06 | -- | 0.465168 | 1 | 0.39451 | -1 | 0.56052 | 1 |
| 7-222020-CCG-C | C | | | | | | | | | 0.012246 | 3.95E-24 | TRUE | 3.95E-24 | ?- | 0.005622 | 1 | 0.00019 | 1 | 7.69E-19 | 1 |
| 7-443821-G-GGGAT | GGGAT | | | | | | | 0.294015 | 0.996808 | 0.370019 | 4.41E-08 | TRUE | 0.002096 | +- | 0.589802 | 1 | 0.31675 | 1 | 2.35E-07 | 1 |
| 7-468291-C-T | T | | | | | | | | | 0.039504 | 1.51E-09 | FALSE | 1.52E-09 | ?+ | 0.170319 | -1 | 0.019446 | 1 | 6.05E-10 | 1 |
| 7-470179-CCCA-C | C | | | | | | | 0.008711 | 0.948577 | 0.014971 | 4.82E-11 | TRUE | 1.09E-05 | +- | 0.014033 | 1 | 0.420092 | 1 | 7.13E-09 | 1 |
| 7-606603-G-A | A | 0.000156 | 0.046783 | | | 0.04678 | +? | 0.001344 | 0.215617 | 0.001659 | 2.99E-17 | TRUE | 1.16E-07 | -+ | 0.369781 | 1 | 0.010535 | 1 | 3.19E-16 | 1 |
| 7-608598-GTCAGTGT( | G | | | | | | | | | 0.220896 | 6.09E-09 | FALSE | 6.09E-09 | ?+ | 0.058222 | -1 | 0.80165 | -1 | 5.25E-13 | 1 |
| 7-1066816-GCACCCCA | G | | | | | | | | | 0.023382 | 1.01E-08 | TRUE | 1.01E-08 | ?+ | 0.008025 | 1 | 0.781856 | 1 | 3.35E-08 | 1 |
| 7-1115481-G-GGAT | GGAT | | | | | | | 0.001002 | 0.687714 | 0.003888 | 4.58E-08 | TRUE | 9.51E-07 | -- | 0.191924 | 1 | 0.319302 | 1 | 3.34E-08 | 1 |
| 7-1216367-G-T | T | | | | | | | 0.000868 | 0.155797 | 0.003116 | 3.9E-08 | TRUE | 0.000427 | -+ | 0.225089 | 1 | 0.105964 | 1 | 6.34E-08 | 1 |
| 7-1765054-T-C | C | | | | | | | 0.000235 | 0.9368 | 0.000623 | 2.75E-12 | TRUE | 1.12E-09 | -- | 0.845138 | 1 | 0.416804 | -1 | 7.9E-14 | 1 |
| 7-2296752-T-G | G | | | | | | | 0.000323 | 0.029002 | 0.001354 | 1.1E-08 | TRUE | 1.87E-09 | -- | 0.00183 | 1 | 0.283219 | 1 | 3.53E-07 | 1 |
| 7-2455679-C-G | G | | | | | | | 4.7E-05 | 0.330179 | 0.000779 | 3.72E-10 | TRUE | 2.95E-10 | -- | 0.000465 | 1 | 0.006956 | 1 | 1.65E-06 | 1 |
| 7-2455697-C-G | G | | | | | | | 0.000404 | 0.203285 | 0.001925 | 2.68E-16 | TRUE | 1.9E-11 | +- | 3.08E-05 | 1 | 0.000553 | 1 | 6.62E-10 | 1 |
| 7-2561058-AAGGATT( | A | 0.126102 | 0.169976 | | | 0.17 | -? | | | 0.041895 | 2.28E-09 | FALSE | 2.28E-09 | ?+ | 0.018003 | 1 | 0.990402 | 1 | 1.72E-09 | 1 |
| 7-3030002-C-G | G | 9.76E-05 | 0.10395 | | | 0.1039 | -? | 0.00031 | 0.64577 | 0.001029 | 3.35E-08 | TRUE | 7.64E-07 | -- | 0.021327 | 1 | 0.220293 | 1 | 2.06E-07 | 1 |
| 7-3186156-G-T | T | | | | | | | 0.001187 | 0.020147 | 0.001986 | 1.81E-14 | TRUE | 3.54E-13 | ++ | 0.425932 | 1 | 0.000363 | 1 | 1.65E-11 | 1 |
| 7-4276876-GGA-G | G | | | | | | | | | 0.000563 | 4.14E-09 | TRUE | 4.14E-09 | ?+ | 0.020519 | 1 | 0.000182 | 1 | 7.69E-07 | 1 |
| 7-4896984-G-C | C | | | | | | | | | 0.000112 | 3.14E-08 | TRUE | 3.14E-08 | ?+ | | | | | 3.16E-06 | 1 |
| 7-4967850-A-G | G | | | | | | | | | 0.027932 | 1.79E-08 | FALSE | 1.79E-08 | ?- | 0.001009 | 1 | 0.022522 | 1 | 1.71E-06 | 1 |
| 7-5140813-G-A | A | 0.002622 | 0.784384 | | | 0.7844 | +? | 0.000736 | 0.1929 | 0.001653 | 6.55E-11 | TRUE | 8.49E-06 | -+ | 0.008919 | 1 | 0.007196 | 1 | 1.49E-08 | 1 |
| 7-5140818-G-A | A | 0.002353 | 0.574317 | | | 0.5743 | -? | 0.000316 | 0.699399 | 0.001467 | 6.03E-12 | TRUE | 2.32E-10 | ++ | 0.774795 | 1 | 0.009501 | 1 | 1.29E-10 | 1 |
| 7-5515904-C-T | T | | | | | | | 0.000326 | 0.315853 | 0.000548 | 2.92E-11 | TRUE | 1.76E-06 | -+ | 0.021658 | 1 | 0.656166 | 1 | 4.94E-11 | 1 |
| 7-5515912-G-A | A | | | | | | | 0.000323 | 0.003294 | 0.00092 | 8.76E-14 | TRUE | 5.3E-08 | -+ | 0.000275 | 1 | 0.653075 | 1 | 1.01E-11 | 1 |
| 7-5515914-C-G | G | | | | | | | 0.000301 | 0.002591 | 0.000821 | 1.22E-15 | TRUE | 2.91E-09 | +- | 0.01643 | 1 | 0.25138 | 1 | 8.65E-15 | 1 |
| 7-5515924-G-A | A | | | | | | | 0.000316 | 0.001452 | 0.000492 | 1.15E-08 | TRUE | 0.001645 | -+ | 0.017423 | 1 | 0.385468 | -1 | 3.9E-08 | 1 |
| 7-5535600-CGGGTTCA | C | | | | | | | 0.000157 | 0.580122 | 0.000493 | 5.01E-09 | TRUE | 1.42E-06 | +- | 0.000182 | 1 | 0.142762 | 1 | 3.06E-06 | 1 |
| 7-5564808-A-G | G | 7.84E-05 | 0.574031 | | | 0.574 | -? | | | 0.001508 | 5.42E-13 | TRUE | 5.42E-13 | ?- | 0.012363 | 1 | 0.007709 | 1 | 8.23E-11 | 1 |
| 7-5856871-A-AT | AT | | | | | | | | | 0.001051 | 1.63E-13 | TRUE | 1.63E-13 | ?- | 0.00938 | 1 | 0.004871 | 1 | 8.54E-10 | 1 |
| 7-6869364-C-G | G | 0.000456 | 0.112196 | | | 0.1122 | +? | 0.000405 | 0.118979 | 0.000974 | 2.55E-08 | TRUE | 6.21E-08 | -- | 0.096627 | 1 | 0.318034 | 1 | 1.67E-07 | 1 |
| 7-13820977-C-T | T | | | | | | | 1.25E-05 | 0.881387 | 0.000778 | 3.74E-08 | TRUE | 4.63E-08 | ++ | 0.361328 | 1 | 0.252729 | -1 | 2.11E-09 | 1 |
| 7-13820985-C-T | T | | | | | | | | | 0.000963 | 3.54E-14 | TRUE | 3.54E-14 | ?+ | 0.017838 | 1 | 0.139343 | 1 | 8.43E-14 | 1 |
| 7-26010871-A-T | T | | | | | | | | | 0.045517 | 2.08E-15 | FALSE | 2.08E-15 | ?- | 0.14533 | 1 | 0.469027 | 1 | 1.04E-16 | 1 |
| 7-26252136-C-CCT | CCT | | | | | | | | | 0.000275 | 8.19E-09 | TRUE | 8.19E-09 | ?+ | 0.538858 | 1 | | | 1.29E-07 | 1 |
| 7-32888710-A-ATTAC | ATTACTTG | | | | | | | | | 0.000187 | 2.77E-08 | TRUE | 2.77E-08 | ?- | 0.531836 | 1 | | | 1.43E-07 | 1 |
| 7-37456405-G-A | A | | | | | | | 0.001657 | 0.615249 | 0.001127 | 1.11E-08 | TRUE | 9.85E-05 | ++ | 0.869695 | -1 | 0.913594 | 1 | 1.19E-09 | 1 |
| 7-44676427-C-T | T | | | | | | | 0.000323 | 0.054015 | 0.000714 | 1.88E-10 | TRUE | 2.81E-05 | -+ | 0.010581 | 1 | 0.28521 | 1 | 2.49E-08 | 1 |
| 7-44816014-C-A | A | | | | | | | | | 0.031751 | 1.68E-10 | FALSE | 1.68E-10 | ?- | 0.023476 | -1 | 0.015794 | -1 | 1.03E-07 | -1 |
| 7-44916280-C-G | G | | | | | | | 0.000742 | 0.179336 | 0.000732 | 2.05E-08 | TRUE | 0.00371 | +- | 0.00059 | 1 | 0.000849 | 1 | 0.000184 | 1 |
| 7-44916283-C-A | A | | | | | | | 0.000742 | 0.179098 | 0.000926 | 7.8E-13 | TRUE | 9.98E-06 | -+ | 0.000582 | 1 | 0.010258 | 1 | 3.52E-09 | 1 |
| 7-44916302-AT-A | A | 7.8E-05 | 0.502287 | | | 0.5023 | -? | 0.000958 | 0.335755 | 0.000908 | 2.58E-10 | TRUE | 4.84E-07 | ++ | 0.00539 | 1 | 0.002233 | 1 | 1.9E-06 | 1 |
| 7-47554393-GGGT-G | G | | | | | | | | | 0.00039 | 2.88E-09 | TRUE | 2.88E-09 | ?+ | 0.002974 | 1 | 0.022273 | 1 | 9.47E-08 | 1 |
| 7-47925128-TGGAAA/ | T | | | | | | | 2.51E-05 | 0.208106 | 0.00044 | 2.58E-09 | TRUE | 3.66E-08 | -+ | 0.003408 | 1 | 5.47E-05 | 1 | 0.000247 | 1 |
| 7-48002361-T-C | C | | | | | | | | | 0.000793 | 4.68E-09 | TRUE | 4.68E-09 | ?- | 0.005924 | 1 | 0.247571 | 1 | 4.61E-07 | 1 |
| 7-49633385-C-CACAG | CACAG | | | | | | | 0.000586 | 0.008806 | 0.001001 | 1.37E-08 | TRUE | 2.71E-09 | ++ | 9.9E-05 | 1 | 0.094081 | 1 | 1.12E-05 | 1 |
| 7-51057333-TAC-T | T | | | | | | | | | 0.370149 | 2.31E-08 | TRUE | 2.31E-08 | ?+ | 0.01053 | 1 | 0.976783 | 1 | 8.29E-05 | 1 |
| 7-55835419-T-G | G | 9.77E-05 | 0.682525 | | | 0.6825 | -? | 0.000426 | 0.531286 | 0.002448 | 1.72E-19 | TRUE | 2.79E-17 | -- | 0.000401 | 1 | 0.000211 | 1 | 5.17E-13 | 1 |

| | | | | | | | | | | | | | | | | | | | | |
|---|---|---|---|---|---|---|---|---|---|---|---|---|---|---|---|---|---|---|---|---|
| 7-57783552-C-T | T | | | | | | | 0.000711 | 0.044829 | 0.00584 | 4.03E-11 | TRUE | 1.75E-11 | -- | 0.198841 | -1 | 0.30436 | -1 | 1.94E-11 | -1 |
| 7-58060060-TTTG-T | T | | | | | | | | | 0.004032 | 4.43E-14 | TRUE | 4.43E-14 | ?+ | 0.00442 | 1 | 0.246483 | 1 | 1.02E-11 | 1 |
| 7-58060068-TTC-T | T | 0.000567 | 0.562217 | | | 0.5622 | -? | | | 0.003178 | 3.24E-11 | TRUE | 3.24E-11 | ?+ | 0.00352 | 1 | 0.085395 | 1 | 1.93E-08 | 1 |
| 7-58060792-T-TTCTAT | TTCTATTC | | | | | | | | | 0.004295 | 4.32E-30 | TRUE | 4.32E-30 | ?- | 0.006849 | 1 | 0.005668 | 1 | 6.92E-30 | 1 |
| 7-62337609-T-A | A | | | | | | | | | 0.00026 | 8.39E-09 | TRUE | 8.39E-09 | ?+ | 0.012877 | 1 | 0.421254 | 1 | 1.19E-07 | 1 |
| 7-62352797-T-A | A | | | | | | | | | 0.002866 | 1.18E-12 | FALSE | 1.18E-12 | ?+ | 0.007151 | 1 | 3.64E-06 | 1 | 6.91E-08 | 1 |
| 7-62430395-T-A | A | | | | | | | 0.000686 | 0.75691 | 0.004658 | 9.11E-10 | TRUE | 4.45E-08 | -+ | 0.745861 | -1 | 0.325422 | 1 | 4.56E-11 | 1 |
| 7-64809255-G-GCGG | GCGGGGGGC | | | | | | | | | 0.158541 | 1.91E-20 | FALSE | 1.91E-20 | ?+ | 0.586515 | 1 | 0.564947 | -1 | 5.09E-21 | -1 |
| 7-65015752-C-T | T | 0.000377 | 0.837308 | | | 0.8373 | -? | 0.001651 | 0.24035 | 0.001727 | 1.7E-08 | TRUE | 4.15E-06 | ++ | 0.046531 | 1 | 0.065536 | 1 | 6.68E-07 | 1 |
| 7-65095078-AT-A | A | | | | | | | | | 0.961686 | 5.47E-12 | TRUE | 5.47E-12 | ?- | 1.87E-06 | -1 | 0.419004 | -1 | 1.1E-05 | -1 |
| 7-65095078-ATT-A | A | | | | | | | 0.012647 | 0.169934 | 0.027799 | 1.16E-11 | FALSE | 1.53E-08 | ++ | 2.83E-06 | 1 | 0.916617 | 1 | 3.28E-06 | 1 |
| 7-65520276-T-TTTTC | TTTTC | | | | | | | | | 0.008884 | 8.51E-10 | FALSE | 8.51E-10 | ?- | 0.00063 | 1 | 0.000185 | 1 | 1.86E-06 | 1 |
| 7-65711737-GAC-G | G | | | | | | | | | 0.000599 | 9.15E-14 | TRUE | 9.15E-14 | ?+ | 3.33E-05 | 1 | 0.850238 | -1 | 3.92E-12 | 1 |
| 7-65862446-AAAG-A | A | 0.000491 | 0.834091 | | | 0.8341 | +? | 0.005679 | 0.98983 | 0.001543 | 1.04E-09 | TRUE | 0.01029 | -+ | 0.347133 | 1 | 0.173716 | 1 | 2.41E-08 | 1 |
| 7-66026675-TAA-T | T | | | | | | | | | 0.001782 | 6.51E-13 | TRUE | 6.51E-13 | ?+ | 0.051582 | 1 | 0.000747 | 1 | 2.52E-10 | 1 |
| 7-66707015-G-GACCT | GACCT | 0.002701 | 0.798542 | | | 0.7985 | +? | | | 0.002603 | 6.81E-09 | TRUE | 6.81E-09 | ?- | 0.159775 | 1 | 0.013921 | 1 | 1.29E-07 | 1 |
| 7-67109039-C-T | T | 0.000156 | 0.940672 | | | 0.9407 | +? | 0.000401 | 0.893253 | 0.000559 | 2.32E-09 | TRUE | 3.79E-06 | ++ | 0.128945 | 1 | 0.749696 | -1 | 8.4E-09 | 1 |
| 7-67109054-A-ATT | ATT | | | | | | | 0.000282 | 0.83812 | 0.001497 | 1.55E-13 | TRUE | 9.38E-12 | -- | 0.000471 | 1 | 0.518369 | 1 | 1.71E-11 | 1 |
| 7-67889016-A-ATCTG | ATCTGGGAAGTGAGGAGCGCCTCTGCCCAGCCGCCAACTG | | | | | | | 0.944532 | 0.440315 | 0.832094 | 3.14E-09 | FALSE | 1.3E-06 | ++ | 0.396204 | -1 | 0.091948 | -1 | 2.24E-09 | -1 |
| 7-68108631-C-CGACA | CGACACAGT | | | | | | | | | 0.000661 | 1.2E-12 | TRUE | 1.2E-12 | ?+ | 0.131826 | 1 | 0.000414 | 1 | 2.48E-10 | 1 |
| 7-69807598-C-A | A | | | | | | | | | 0.000407 | 6.1E-11 | TRUE | 6.09E-11 | ?+ | 0.042886 | 1 | 0.39762 | -1 | 3.51E-11 | 1 |
| 7-69807601-CCT-C | C | | | | | | | | | 0.001109 | 4.66E-10 | TRUE | 4.66E-10 | ?- | 0.010328 | 1 | 0.897465 | 1 | 3.35E-09 | 1 |
| 7-70395262-G-C | C | 0.000824 | 0.626502 | | | 0.6265 | -? | 0.003843 | 0.155739 | 0.001634 | 2.03E-08 | TRUE | 8.18E-05 | ++ | 0.842778 | 1 | 0.717566 | 1 | 5.1E-10 | 1 |
| 7-71010361-TGG-T | T | | | | | | | | | 0.000393 | 1.24E-09 | TRUE | 1.24E-09 | ?+ | 0.015247 | 1 | 0.713002 | 1 | 1.86E-08 | 1 |
| 7-72247055-G-A | A | 0.367189 | 0.360778 | 0.441 | 0.52354 | 0.3901 | +- | 0.442751 | 0.015517 | 0.41352 | 1.33E-09 | FALSE | 0.1865 | -+ | 0.006654 | 1 | 0.396666 | 1 | 2.43E-09 | 1 |
| 7-72703259-C-CAA | CAA | | | | | | | | | 0.000632 | 2.99E-10 | TRUE | 2.99E-10 | ?+ | 0.041804 | 1 | 0.018133 | 1 | 2E-08 | 1 |
| 7-73239519-T-TA | TA | | | | | | | | | 0.001069 | 3.37E-15 | TRUE | 3.37E-15 | ?- | 0.870111 | -1 | 0.027809 | 1 | 3.17E-15 | 1 |
| 7-73239535-AGC-A | A | | | | | | | | | 0.000135 | 2.07E-08 | TRUE | 2.07E-08 | ?+ | | | | | 4.29E-08 | 1 |
| 7-73415961-C-CTCT | CTCT | | | | | | | | | 0.002303 | 2.36E-13 | TRUE | 2.36E-13 | ?+ | 0.008517 | 1 | 0.471976 | 1 | 3.83E-11 | 1 |
| 7-73481896-T-A | A | 0.000137 | 0.469345 | | | 0.4693 | +? | 0.001679 | 0.693204 | 0.001254 | 4.3E-12 | TRUE | 3.29E-06 | ++ | 0.006978 | 1 | 0.056093 | 1 | 2.6E-10 | 1 |
| 7-73481899-C-T | T | 0.000196 | 0.419685 | | | 0.4197 | +? | 0.00171 | 0.564469 | 0.001272 | 2.12E-14 | TRUE | 1.28E-07 | ++ | 0.178095 | 1 | 0.009868 | 1 | 4.44E-13 | 1 |
| 7-73481926-T-C | C | 0.000235 | 0.820742 | | | 0.8207 | -? | 0.000764 | 0.92193 | 0.001291 | 4.93E-14 | TRUE | 6.82E-09 | +- | 0.001343 | 1 | 0.004879 | 1 | 4.65E-10 | 1 |
| 7-73619442-CAG-C | C | | | | | | | | | 0.006506 | 6.63E-21 | TRUE | 6.63E-21 | ?- | 4.38E-06 | 1 | 0.002406 | 1 | 2.72E-16 | 1 |
| 7-73619449-C-CAA | CAA | | | | | | | | | 0.0054 | 4.62E-18 | TRUE | 4.62E-18 | ?+ | 7.99E-06 | 1 | 0.006426 | 1 | 2.75E-13 | 1 |
| 7-73763277-G-A | A | | | | | | | 7.83E-05 | 0.814753 | 0.000141 | 7.43E-09 | TRUE | 4.03E-07 | ++ | | | | | 1.25E-08 | 1 |
| 7-74071734-A-AAGTG | AAGTG | | | | | | | | | 0.000572 | 5.87E-09 | TRUE | 5.87E-09 | ?- | 0.254343 | 1 | 0.988898 | 1 | 2E-08 | 1 |
| 7-74251571-A-G | G | 0.456004 | 0.014406 | | | 0.01441 | -? | 0.40647 | 0.660966 | 0.473278 | 1.21E-08 | FALSE | 0.005246 | +- | 0.49746 | 1 | 0.416928 | 1 | 1.02E-06 | 1 |
| 7-74300691-C-CAA | CAA | 0.000273 | 0.448752 | | | 0.4488 | +? | | | 0.000703 | 1E-09 | TRUE | 1E-09 | ?+ | 0.012968 | 1 | 0.500932 | 1 | 9.64E-09 | 1 |
| 7-74303748-CT-C | C | | | | | | | | | 0.00073 | 3.61E-08 | TRUE | 3.61E-08 | ?- | 0.028722 | 1 | 0.011717 | 1 | 1.5E-06 | 1 |
| 7-74493235-G-A | A | 0.000353 | 0.306395 | | | 0.3064 | -? | 0.003283 | 0.444242 | 0.002229 | 1.6E-16 | TRUE | 1.14E-05 | -+ | 8.59E-05 | 1 | 0.096219 | 1 | 2.61E-12 | 1 |
| 7-74493250-T-C | C | 0.000313 | 0.241106 | | | 0.2411 | +? | 0.002684 | 0.727944 | 0.002032 | 1.51E-25 | TRUE | 9.66E-11 | +- | 9.32E-06 | 1 | 0.038093 | 1 | 1.37E-19 | 1 |
| 7-74784418-C-CTTG | CTTG | | | | | | | | | 0.875029 | 1.13E-09 | FALSE | 1.13E-09 | ?- | 0.003709 | -1 | 0.755461 | -1 | 4.56E-08 | -1 |
| 7-74792891-T-TA | TA | | | | | | | | | 0.00069 | 4.73E-12 | TRUE | 4.73E-12 | ?- | 0.106133 | 1 | 0.000407 | 1 | 1.85E-07 | 1 |
| 7-74999145-G-A | A | | | | | | | 0.000314 | 0.244633 | 0.000399 | 3.12E-09 | TRUE | 0.000113 | -+ | 0.420375 | 1 | 0.015705 | 1 | 3.85E-09 | 1 |
| 7-75607689-G-GT | GT | | | | | | | | | 0.002817 | 4E-32 | TRUE | 4E-32 | ?- | 1.02E-05 | 1 | 0.235079 | 1 | 1.65E-28 | 1 |
| 7-75607691-AC-A | A | | | | | | | | | 0.001359 | 5.72E-21 | TRUE | 5.72E-21 | ?+ | 7.27E-06 | 1 | 0.318778 | 1 | 2.86E-16 | 1 |
| 7-75676780-GAA-G | G | | | | | | | | | 0.623053 | 6.96E-11 | FALSE | 6.97E-11 | ?+ | 0.085204 | 1 | 0.203607 | 1 | 2.2E-08 | 1 |
| 7-75792984-ATG-A | A | 0.001514 | 0.322383 | | | 0.3224 | +? | | | 0.001538 | 4.68E-16 | TRUE | 4.68E-16 | ?+ | 0.004236 | 1 | 0.005749 | 1 | 1.81E-12 | 1 |
| 7-75792987-C-CAT | CAT | 0.001402 | 0.190182 | | | 0.1902 | +? | | | 0.001581 | 8.47E-17 | TRUE | 8.47E-17 | ?+ | 0.000949 | 1 | 0.001294 | 1 | 4.4E-12 | 1 |
| 7-75801743-G-T | T | | | | | | | | | 0.000302 | 4.28E-08 | TRUE | 4.28E-08 | ?+ | 0.020398 | 1 | | | 2.21E-06 | 1 |
| 7-75801752-ATC-A | A | | | | | | | | | 0.000171 | 6.82E-10 | TRUE | 6.82E-10 | ?+ | 0.000257 | 1 | | | 3.32E-06 | 1 |
| 7-75853377-GTTT-G | G | | | | | | | | | 0.000463 | 4.71E-08 | TRUE | 4.71E-08 | ?+ | 0.00247 | 1 | 0.830522 | 1 | 1.05E-06 | 1 |
| 7-76528574-A-AG | AG | | | | | | | | | 0.794886 | 1.43E-31 | FALSE | 1.43E-31 | ?+ | 0.001795 | -1 | 5.59E-07 | -1 | 2.61E-22 | -1 |
| 7-76528579-C-G | G | | | | | | | | | 0.800977 | 4.38E-29 | FALSE | 4.38E-29 | ?+ | 0.021309 | -1 | 1.32E-05 | -1 | 2.12E-22 | -1 |
| 7-76528593-TCACA-T | T | | | | | | | | | 0.766316 | 4.86E-30 | FALSE | 4.86E-30 | ?- | 0.020578 | -1 | 9.29E-07 | -1 | 1.02E-22 | -1 |
| 7-76528597-A-ATGTG | ATGTG | | | | | | | | | 0.805194 | 6.39E-28 | FALSE | 6.39E-28 | ?+ | 0.021149 | -1 | 0.000953 | -1 | 3.72E-23 | -1 |
| 7-89881278-AT-A | A | 9.75E-05 | 0.450102 | | | 0.4501 | +? | | | 0.00038 | 1.56E-14 | TRUE | 1.57E-14 | ?+ | 0.025092 | 1 | 1.56E-05 | 1 | 4.24E-10 | 1 |
| 7-89881280-CCTGGCT | C | | | | | | | | | 0.000263 | 3.96E-10 | TRUE | 3.96E-10 | ?- | 0.053259 | 1 | 0.001608 | 1 | 1.97E-07 | 1 |
| 7-97243876-T-TCCAC | TCCAC | | | | | | | | | 0.004568 | 1.33E-08 | TRUE | 1.33E-08 | ?- | 0.022325 | 1 | 0.190072 | 1 | 4.61E-06 | 1 |

| | | | | | | | | | | | | | | | | | | | | |
|---|---|---|---|---|---|---|---|---|---|---|---|---|---|---|---|---|---|---|---|---|
| 7-100284341-A-C | C | | | | | | | 0.000269 | 0.452824 | 0.000417 | 2.15E-08 | TRUE | 8.6E-05 | +- | 0.133599 | 1 | 0.113362 | 1 | 9.38E-08 | 1 |
| 7-100672357-CCT-C | C | 0.000274 | 0.106357 | | | 0.1064 | -? | | | 0.004255 | 6.27E-18 | TRUE | 6.27E-18 | ?- | 0.00012 | 1 | 0.542028 | 1 | 1.24E-15 | 1 |
| 7-101035140-T-A | A | 0.189345 | 0.532467 | | | 0.5325 | +? | | | 0.186878 | 3.81E-09 | TRUE | 3.81E-09 | ?+ | 0.052524 | 1 | 0.010735 | 1 | 0.000335 | 1 |
| 7-101284191-GAT-G | G | | | | | | | | | 0.000282 | 1.02E-08 | TRUE | 1.02E-08 | ?+ | 0.338474 | 1 | | | 1.3E-07 | 1 |
| 7-101284194-G-GCA | GCA | | | | | | | | | 0.000257 | 2.82E-09 | TRUE | 2.82E-09 | ?- | 0.017103 | 1 | | | 5.21E-08 | 1 |
| 7-101788981-A-T | T | | | | | | | 0.000727 | 0.868141 | 0.002162 | 1.49E-16 | TRUE | 4.87E-12 | +- | 0.01622 | 1 | 0.174622 | 1 | 4.63E-14 | 1 |
| 7-101822055-T-C | C | | | | | | | 0.00062 | 0.702338 | 0.001116 | 1.37E-09 | TRUE | 8.37E-07 | -- | 0.000572 | 1 | 0.281199 | 1 | 5.95E-08 | 1 |
| 7-102026581-AG-A | A | | | | | | | | | 0.000506 | 3.01E-09 | TRUE | 3.01E-09 | ?+ | 0.015594 | 1 | 0.000212 | 1 | 1.4E-06 | 1 |
| 7-102920935-C-CCT | CCT | | | | | | | | | 0.001292 | 2.05E-08 | TRUE | 2.05E-08 | ?+ | 0.023074 | 1 | 0.073407 | 1 | 4.63E-07 | 1 |
| 7-105596291-C-T | T | 0.000176 | 0.900101 | | | 0.9001 | -? | 0.002418 | 0.842782 | 0.002735 | 8.52E-17 | TRUE | 2.65E-08 | -+ | 1.35E-06 | 1 | 0.043919 | 1 | 1.55E-11 | 1 |
| 7-106975359-C-G | G | 0.00106 | 0.664283 | | | 0.6643 | -? | 0.005043 | 0.044755 | 0.004327 | 7.72E-33 | TRUE | 2.55E-10 | +- | 0.000136 | 1 | 0.005182 | 1 | 3.57E-26 | 1 |
| 7-111925911-C-A | A | 0.001956 | 0.947705 | | | 0.9477 | +? | 0.00089 | 0.419217 | 0.002132 | 4.93E-12 | TRUE | 1.22E-09 | ++ | 0.034665 | 1 | 0.002059 | 1 | 1.36E-07 | 1 |
| 7-111925913-C-A | A | | | 0.001 | 0.683091 | 0.6831 | ?- | 0.000887 | 0.3947 | 0.001955 | 7.64E-13 | TRUE | 3.09E-10 | ++ | 0.642458 | 1 | 0.016956 | 1 | 1.4E-09 | 1 |
| 7-116919680-A-ATG | ATG | | | | | | | | | 0.005434 | 4.06E-10 | TRUE | 4.06E-10 | ?- | 0.434853 | 1 | 0.001858 | 1 | 2.72E-08 | 1 |
| 7-117106822-G-T | T | | | | | | | 0.002174 | 0.680421 | 0.001953 | 8.95E-09 | TRUE | 0.000844 | -+ | 0.010644 | 1 | 0.206087 | 1 | 7.78E-08 | 1 |
| 7-123042619-C-A | A | | | | | | | | | 0.50618 | 1.2E-08 | FALSE | 1.2E-08 | ?+ | 0.515964 | 1 | 0.890368 | 1 | 3.03E-10 | 1 |
| 7-126391942-T-A | A | | | | | | | 0.002102 | 0.077793 | 0.002951 | 1.79E-16 | TRUE | 1.86E-06 | -+ | 0.701431 | 1 | 0.000134 | 1 | 1.34E-12 | 1 |
| 7-126391976-A-G | G | | | | | | | 0.001751 | 0.558351 | 0.002062 | 7.49E-17 | TRUE | 3.73E-08 | +- | 0.000715 | 1 | 0.014426 | 1 | 1.67E-10 | 1 |
| 7-128518959-A-G | G | 9.75E-05 | 0.173222 | | | 0.1732 | +? | 0.000417 | 0.620633 | 0.00063 | 2.76E-09 | TRUE | 1.64E-05 | +- | 0.003334 | 1 | 0.029557 | 1 | 5.12E-07 | 1 |
| 7-128552494-T-G | G | | | | | | | | | 0.000717 | 8.05E-11 | TRUE | 8.05E-11 | ?- | 0.001443 | 1 | 0.036021 | 1 | 6.27E-08 | 1 |
| 7-129358248-C-T | T | | | | | | | 0.000367 | 0.676651 | 0.002036 | 2.43E-10 | TRUE | 4.68E-09 | ++ | 0.140402 | 1 | 0.52898 | 1 | 4.19E-09 | 1 |
| 7-130087903-GGT-G | G | 9.76E-05 | 0.194326 | | | 0.1943 | -? | | | 0.001765 | 1.15E-09 | TRUE | 1.15E-09 | ?+ | 0.489523 | 1 | 0.005937 | 1 | 8.15E-08 | 1 |
| 7-130087910-G-GCA | GCA | | | | | | | | | 0.001481 | 2.52E-08 | TRUE | 2.52E-08 | ?- | 0.184194 | 1 | 0.015742 | 1 | 4.19E-06 | 1 |
| 7-132564052-C-CTGT | CTGT | | | | | | | | | 0.962751 | 2.1E-08 | FALSE | 2.1E-08 | ?- | 0.044348 | -1 | 0.538326 | -1 | 1.85E-07 | -1 |
| 7-132564131-T-G | G | | | | | | | | | 0.938219 | 5E-20 | FALSE | 5E-20 | ?+ | 0.204497 | -1 | 0.330893 | -1 | 1.42E-20 | -1 |
| 7-136789083-C-T | T | | | | | | | | | 0.096476 | 1.43E-08 | FALSE | 1.44E-08 | ?+ | 0.751208 | -1 | 0.050982 | 1 | 2.41E-09 | 1 |
| 7-138674703-A-AAT | AAT | | | | | | | | | 0.000623 | 1.63E-08 | TRUE | 1.63E-08 | ?- | 0.021971 | 1 | 0.036894 | 1 | 2.46E-06 | 1 |
| 7-140480517-GA-G | G | | | | | | | | | 0.000652 | 2.94E-10 | TRUE | 2.94E-10 | ?+ | 0.000532 | 1 | 0.67001 | 1 | 8.8E-08 | 1 |
| 7-140498699-C-A | A | | | | | | | | | 0.001779 | 1.38E-21 | TRUE | 1.38E-21 | ?+ | 0.000193 | 1 | 0.004423 | 1 | 1.22E-14 | 1 |
| 7-140619733-CAA-C | C | | | | | | | | | 0.000774 | 2.11E-11 | TRUE | 2.11E-11 | ?- | 0.003267 | 1 | 0.288578 | 1 | 5.37E-10 | 1 |
| 7-140944263-A-AT | AT | | | | | | | 0.000392 | 0.994039 | 0.003409 | 2.19E-17 | TRUE | 2.26E-15 | +- | 0.183794 | 1 | 0.082124 | 1 | 9.12E-17 | 1 |
| 7-140961641-C-CAA | CAA | | | | | | | | | 0.000759 | 5.77E-12 | TRUE | 5.77E-12 | ?+ | 0.005267 | 1 | 0.005542 | 1 | 2.23E-08 | 1 |
| 7-143802911-GACA-G | G | 0.001218 | 0.71556 | | | 0.7156 | +? | | | 0.007171 | 1.14E-10 | FALSE | 1.14E-10 | ?+ | 0.026799 | 1 | 0.000569 | 1 | 6.34E-07 | 1 |
| 7-148956743-AGG-A | A | | | | | | | | | 0.000522 | 7.74E-10 | TRUE | 7.74E-10 | ?+ | 0.015527 | 1 | 0.01494 | 1 | 1.59E-08 | 1 |
| 7-148989806-T-A | A | | | | | | | 7.52E-05 | 0.051067 | 0.001053 | 2.77E-15 | TRUE | 4.62E-16 | ++ | 0.014903 | 1 | 0.365303 | 1 | 1.79E-14 | 1 |
| 7-149095930-C-CCT | CCT | | | | | | | | | 0.001302 | 2.16E-12 | TRUE | 2.16E-12 | ?+ | 1.33E-05 | 1 | 0.026637 | 1 | 9.07E-09 | 1 |
| 7-149095962-G-C | C | | | | | | | 0.00119 | 0.232213 | 0.000705 | 5.8E-09 | TRUE | 7.35E-06 | ++ | 7.88E-06 | 1 | 0.370889 | 1 | 2.59E-06 | 1 |
| 7-149904512-C-CA | CA | | | | | | | | | 0.0022 | 3.53E-09 | TRUE | 3.53E-09 | ?+ | 0.119207 | 1 | 0.146835 | 1 | 2.11E-08 | 1 |
| 7-152402297-C-CAAT( | CAATCATA | | | | | | | | | 0.002485 | 5.98E-13 | TRUE | 5.98E-13 | ?+ | 0.001747 | 1 | 0.012765 | 1 | 3.11E-09 | 1 |
| 7-152415704-A-C | C | 0.267761 | 0.908273 | 0.488 | 0.728979 | 0.8883 | ++ | | | 0.284292 | 4.97E-09 | FALSE | 4.97E-09 | ?- | 0.201643 | 1 | 0.26939 | 1 | 8.2E-06 | 1 |
| 7-152546811-CA-C | C | | | | | | | | | 0.001449 | 3.87E-09 | TRUE | 3.87E-09 | ?- | 0.100376 | 1 | 0.118002 | 1 | 7.55E-09 | 1 |
| 7-152617758-T-C | C | 0.000333 | 0.321715 | | | 0.3217 | -? | 5.32E-05 | 0.492947 | 0.001171 | 1.49E-09 | TRUE | 1.38E-09 | -- | 0.333072 | 1 | 0.167309 | 1 | 2.71E-09 | 1 |
| 7-152617778-AT-A | A | | | | | | | 0.000984 | 0.045143 | 0.000909 | 3.09E-08 | TRUE | 3.81E-07 | ++ | 0.000515 | 1 | 0.00511 | 1 | 4.97E-05 | 1 |
| 7-154608341-A-T | T | | | | | | | | | 0.607774 | 4.87E-08 | FALSE | 4.87E-08 | ?- | 0.765302 | 1 | 0.00387 | 1 | 6.72E-07 | 1 |
| 7-155134673-C-CCT | CCT | | | | | | | | | 0.001098 | 2.44E-08 | TRUE | 2.44E-08 | ?+ | 1.98E-05 | 1 | 0.015933 | 1 | 0.000306 | 1 |
| 7-155334480-A-G | G | | | | | | | | | 0.967675 | 9.17E-09 | FALSE | 9.17E-09 | ?+ | 0.417173 | 1 | 0.753387 | -1 | 1.79E-10 | -1 |
| 7-156266423-CTGT-C | C | | | | | | | | | 0.600648 | 5.91E-36 | FALSE | 5.91E-36 | ?+ | 0.001198 | -1 | 0.648924 | -1 | 1.9E-33 | -1 |
| 7-156266427-CCTGTT | C | | | | | | | | | 0.612924 | 2.46E-38 | FALSE | 2.45E-38 | ?+ | 0.011481 | -1 | 0.785711 | -1 | 8.96E-38 | -1 |
| 7-156520347-GGGAG( | G | | | | | | | | | 0.820212 | 5.94E-11 | FALSE | 5.94E-11 | ?- | 0.401721 | -1 | 0.664509 | -1 | 6.6E-12 | -1 |
| 7-157725208-GC-G | G | | | | | | | | | 0.009749 | 3.01E-09 | TRUE | 3.01E-09 | ?+ | 0.297024 | 1 | 0.55647 | 1 | 1.12E-09 | 1 |
| 7-157933145-GGGGT( | G | | | | | | | | | 0.134843 | 2.87E-11 | FALSE | 2.87E-11 | ?- | 0.674719 | 1 | 0.274523 | -1 | 4.34E-11 | -1 |
| 7-158045747-C-A | A | | | | | | | | | 0.000266 | 1.4E-08 | TRUE | 1.4E-08 | ?+ | 0.002776 | 1 | | | 3.52E-05 | 1 |
| 7-158084639-ACGACA | A | | | | | | | | | 0.001665 | 4.38E-09 | TRUE | 4.38E-09 | ?+ | 0.006976 | 1 | 0.290402 | 1 | 1.44E-06 | 1 |
| 7-158319527-T-TCA | TCA | | | | | | | | | 0.806633 | 3.16E-08 | TRUE | 3.16E-08 | ?+ | 0.769793 | -1 | 0.033373 | -1 | 1.46E-07 | -1 |
| 7-158348495-C-G | G | | | | | | | | | 0.832645 | 4.64E-09 | FALSE | 4.64E-09 | ?+ | 0.134186 | 1 | 0.047294 | 1 | 1.21E-12 | -1 |
| 7-158365850-C-A | A | | | | | | | | | 0.038633 | 5.85E-10 | FALSE | 5.85E-10 | ?+ | 0.240496 | 1 | 0.000619 | 1 | 6.88E-08 | 1 |
| 7-158459794-CCTAGA | C | | | | | | | | | 0.11233 | 3.15E-08 | FALSE | 3.15E-08 | ?+ | 0.378589 | -1 | 0.272172 | -1 | 1.42E-07 | -1 |
| 7-158463746-CA-C | C | 0.005865 | 0.012741 | | | 0.01274 | +? | | | 0.006224 | 7.97E-15 | TRUE | 7.97E-15 | ?- | 0.006875 | 1 | 0.02243 | 1 | 1.81E-11 | 1 |
| 7-158493344-TACAC-1 | T | | | | | | | 0.004589 | 0.47237 | 0.00631 | 5.59E-10 | TRUE | 0.000239 | -+ | 0.107502 | 1 | 0.451305 | 1 | 3.33E-09 | 1 |

| | | | | | | | | | | | | | | | | | | | | |
|---|---|---|---|---|---|---|---|---|---|---|---|---|---|---|---|---|---|---|---|---|
| 7-158997976-A-G | G | | | | | | | | | 0.057524 | 2.04E-09 | FALSE | 2.04E-09 | ?- | 0.006201 | 1 | 0.012168 | 1 | 2.26E-06 | 1 |
| 7-7210415-CTTTTTTTT | CTTTTTTTTT | | | | | | | 0.000132 | 0.42805 | | | FALSE | 4.86E-26 | +? | | | | | | |
| 8-1281975-A-G | G | 0.000137 | 0.311904 | | | 0.3119 | +? | 2.51E-05 | 2.97E-08 | | | FALSE | 2.97E-08 | -? | | | | | | |
| 8-83883804-T-A | A | | | | | | | 9.4E-05 | 1E-54 | | | FALSE | 0.648 | -? | | | | | | |
| 8-91099094-G-C | C | | | | | | | 9.4E-06 | 1E-84 | | | FALSE | 0.4151 | -? | | | | | | |
| 8-695492-TCTCCTCTC | T | | | | | | | | | 0.091906 | 9.02E-09 | FALSE | 9.02E-09 | ?+ | 0.031539 | 1 | 0.558372 | 1 | 4.34E-07 | 1 |
| 8-1316426-CTCTCCAA | C | | | | | | | | | 0.024267 | 4.61E-09 | TRUE | 4.61E-09 | ?+ | 0.219228 | -1 | 0.057626 | -1 | 4.37E-07 | -1 |
| 8-1348511-G-GGT | GGT | | | | | | | | | 0.181698 | 1.15E-10 | FALSE | 1.15E-10 | ?+ | 0.457434 | -1 | 0.008309 | -1 | 2.9E-09 | -1 |
| 8-1462522-G-A | A | | | | | | | 0.000288 | 0.014192 | 0.000841 | 2.77E-08 | TRUE | 0.000284 | -+ | 0.007654 | 1 | 0.13101 | 1 | 5.77E-07 | 1 |
| 8-1603569-T-TGGGTC | TGGGTCTCAGTTCTGCAGAGGCTGGTTAGAGTGGAGGTG | | | | | | | | | 0.956239 | 1.7E-10 | TRUE | 1.7E-10 | ?+ | 0.045144 | -1 | 0.160581 | -1 | 1.98E-09 | -1 |
| 8-2452889-AT-A | A | | | | | | | | | 0.017179 | 1.49E-08 | FALSE | 1.49E-08 | ?+ | 0.051019 | 1 | 0.013426 | 1 | 8.86E-07 | 1 |
| 8-3660543-A-C | C | | | | | | | | | 0.907268 | 1.03E-14 | FALSE | 1.03E-14 | ?- | 0.032567 | 1 | 0.026495 | 1 | 1.96E-12 | 1 |
| 8-7310261-A-AAAAG | AAAAG | | | | | | | | | 0.261322 | 4.12E-10 | FALSE | 4.12E-10 | ?+ | 0.039428 | -1 | 0.860992 | -1 | 2.64E-11 | -1 |
| 8-7329835-CT-C | C | | | | | | | | | 0.770837 | 3.72E-08 | FALSE | 3.72E-08 | ?+ | 0.001708 | -1 | 0.114434 | -1 | 5.73E-06 | -1 |
| 8-7412675-CT-C | C | | | | | | | | | 0.573999 | 2.89E-22 | TRUE | 2.89E-22 | ?+ | 1E-06 | -1 | 0.161071 | -1 | 2.17E-17 | -1 |
| 8-8160877-C-CA | CA | | | | | | | 0.995295 | 0.6964 | 0.700845 | 4.65E-08 | FALSE | 1.41E-07 | +- | 0.651012 | -1 | 0.844517 | -1 | 3.94E-08 | -1 |
| 8-9493603-A-ATT | ATT | | | | | | | | | 0.000429 | 7.89E-09 | TRUE | 7.89E-09 | ?- | 0.67052 | 1 | 0.366085 | 1 | 3.79E-09 | 1 |
| 8-11026964-C-A | A | | | | | | | | | 0.419867 | 2.5E-08 | FALSE | 2.5E-08 | ?+ | 0.02731 | -1 | 0.532469 | -1 | 4.04E-09 | 1 |
| 8-12443923-C-T | T | | | | | | | 0.740606 | 0.850489 | 0.75081 | 4.37E-10 | FALSE | 0.000753 | -- | 0.632611 | -1 | 0.150646 | -1 | 2.46E-10 | -1 |
| 8-13268436-TC-T | T | | | | | | | | | 0.001866 | 7.18E-15 | TRUE | 7.18E-15 | ?+ | 0.000712 | 1 | 0.996392 | -1 | 6.8E-12 | 1 |
| 8-13324572-C-A | A | | | | | | | | | 0.04113 | 5.56E-09 | FALSE | 5.56E-09 | ?- | 0.044505 | -1 | 0.000394 | -1 | 4.88E-07 | -1 |
| 8-23267200-A-AT | AT | 0.000215 | 0.841635 | | | 0.8416 | +? | | | 0.00215 | 1.35E-12 | TRUE | 1.35E-12 | ?- | 0.922965 | -1 | 0.002455 | 1 | 5.67E-11 | 1 |
| 8-23279493-T-TGG | TGG | | | | | | | | | 0.001209 | 7.27E-09 | TRUE | 7.27E-09 | ?- | 0.011114 | 1 | 0.324069 | 1 | 1.41E-07 | 1 |
| 8-30679870-T-TC | TC | | | | | | | | | 0.001126 | 9.69E-13 | TRUE | 9.69E-13 | ?- | 0.132754 | 1 | 0.401981 | 1 | 3.18E-11 | 1 |
| 8-30912332-G-GC | GC | | | | | | | 5.64E-05 | 0.815013 | 0.896436 | 9.32E-27 | FALSE | 9.08E-27 | ++ | 0.018534 | 1 | 0.811635 | -1 | 7.35E-36 | -1 |
| 8-30912334-T-C | C | | | | | | | 0.999126 | 0.199136 | 0.042589 | 7.82E-12 | FALSE | 4.41E-11 | +- | 0.016728 | -1 | 0.656068 | -1 | 1.99E-16 | 1 |
| 8-33462169-G-GA | GA | | | | | | | | | 0.254087 | 1.99E-08 | FALSE | 1.99E-08 | ?+ | 0.110558 | -1 | 0.870941 | 1 | 2.23E-08 | -1 |
| 8-33468388-TG-T | T | 0.000273 | 0.711945 | | | 0.7119 | +? | 0.000166 | 0.679781 | 0.001551 | 3.41E-12 | TRUE | 1.54E-10 | -+ | 0.618834 | 1 | 0.007169 | 1 | 1.18E-10 | 1 |
| 8-33494173-AAG-A | A | | | | | | | | | 0.00027 | 3.09E-11 | TRUE | 3.09E-11 | ?+ | 0.018426 | 1 | 0.013058 | 1 | 5.11E-08 | 1 |
| 8-33494177-A-ACC | ACC | | | | | | | | | 0.000197 | 7.08E-11 | TRUE | 7.08E-11 | ?- | 0.006405 | 1 | 0.004339 | 1 | 1.02E-07 | 1 |
| 8-34529080-C-CAGGT | CAGGTG | | | | | | | | | 0.014209 | 1.59E-10 | FALSE | 1.59E-10 | ?+ | 0.077893 | 1 | 0.124552 | 1 | 1.06E-10 | 1 |
| 8-34529080-C-CG | CG | | | | | | | | | 0.794377 | 3.22E-34 | FALSE | 3.22E-34 | ?- | 0.249883 | -1 | 0.018191 | -1 | 2.61E-34 | -1 |
| 8-34529080-C-CGGGT | CGGGTG | | | | | | | | | 0.091268 | 1.59E-51 | FALSE | 1.59E-51 | ?+ | 0.480376 | 1 | 0.004875 | 1 | 2.05E-53 | 1 |
| 8-38194649-C-CAG | CAG | | | | | | | 0.000166 | 0.287517 | 0.001285 | 1.21E-13 | TRUE | 7.42E-11 | -+ | 0.00061 | 1 | 0.134907 | 1 | 4.89E-11 | 1 |
| 8-38390825-GGT-G | G | | | | | | | | | 0.001335 | 6.01E-10 | TRUE | 6.01E-10 | ?+ | 0.586418 | 1 | 0.010909 | 1 | 4.24E-08 | 1 |
| 8-40820765-GTGTA-G | G | | | | | | | | | 0.023478 | 1.02E-18 | FALSE | 1.02E-18 | ?- | 0.006902 | -1 | 0.096828 | -1 | 3.71E-15 | -1 |
| 8-40820780-T-C | C | | | | | | | | | 0.021698 | 1.98E-19 | TRUE | 1.98E-19 | ?+ | 0.002832 | -1 | 0.088278 | -1 | 1.31E-15 | -1 |
| 8-43008863-T-G | G | | | | | | | 0.000304 | 0.007742 | 0.001968 | 5.22E-13 | TRUE | 2.87E-09 | +- | 0.011026 | 1 | 0.265401 | 1 | 3.08E-11 | 1 |
| 8-43008878-C-T | T | | | | | | | 4.07E-05 | 0.165033 | 0.001185 | 5.01E-15 | TRUE | 1.9E-15 | ++ | 0.001404 | 1 | 0.096944 | 1 | 2.76E-11 | 1 |
| 8-43008901-C-T | T | 0.000176 | 0.186113 | | | 0.1861 | -? | 0.0001 | 0.884979 | 0.000667 | 2.54E-08 | TRUE | 1.86E-07 | ++ | 0.025443 | 1 | 0.000194 | 1 | 4.83E-05 | 1 |
| 8-43240063-AT-A | A | | | | | | | | | 0.73744 | 2.35E-11 | FALSE | 2.35E-11 | ?- | 0.959568 | 1 | 0.277236 | 1 | 1.71E-15 | -1 |
| 8-43570083-C-G | G | | | | | | | | | 0.175791 | 8.1E-10 | TRUE | 8.1E-10 | ?- | 0.78211 | -1 | 0.390888 | -1 | 0.00058 | 1 |
| 8-43927222-T-A | A | 0.000255 | 0.462561 | | | 0.4626 | +? | 0.000743 | 0.143375 | 0.001415 | 1.54E-12 | TRUE | 9.14E-11 | ++ | 0.076168 | 1 | 0.175811 | 1 | 2.62E-11 | 1 |
| 8-47081758-A-G | G | | | | | | | | | 0.000984 | 2.42E-08 | TRUE | 2.42E-08 | ?- | 0.013501 | 1 | 0.677738 | 1 | 9.75E-08 | 1 |
| 8-48012945-AAG-A | A | 0.000371 | 0.331468 | | | 0.3315 | -? | | | 0.001515 | 4.29E-08 | TRUE | 4.29E-08 | ?+ | 0.015905 | 1 | 0.009626 | 1 | 3.88E-05 | 1 |
| 8-48019002-G-T | T | | | | | | | | | 0.000328 | 1.89E-10 | TRUE | 1.89E-10 | ?+ | 0.000307 | 1 | 0.003002 | 1 | 1.34E-05 | 1 |
| 8-54278292-A-C | C | | | 0.49 | 0.771313 | 0.7713 | ?+ | 0.46711 | 0.241632 | 0.568529 | 2.84E-08 | FALSE | 5.57E-05 | -- | 0.472992 | 1 | 0.993079 | 1 | 7.27E-08 | 1 |
| 8-55827470-AACGCCA | A | | | | | | | | | 0.002339 | 3.9E-11 | TRUE | 3.9E-11 | ?+ | 0.110576 | 1 | 0.000336 | 1 | 4.26E-08 | 1 |
| 8-60523306-C-CAT | CAT | | | | | | | | | 0.000424 | 3.74E-08 | TRUE | 3.74E-08 | ?+ | 0.020302 | 1 | 0.180588 | 1 | 2.36E-06 | 1 |
| 8-66557723-G-GA | GA | | | | | | | 0.000592 | 0.436892 | 0.000758 | 1.01E-08 | TRUE | 2.63E-06 | -- | 0.005906 | 1 | 0.050073 | 1 | 8.78E-07 | 1 |
| 8-66708539-C-A | A | 0.113088 | 0.484688 | 0.062 | 0.039083 | 0.4419 | ++ | | | 0.129068 | 1.41E-08 | TRUE | 1.41E-08 | ?+ | 0.643555 | 1 | 0.543959 | -1 | 0.084192 | 1 |
| 8-67370567-C-CCA | CCA | | | | | | | 0.000132 | 0.18898 | 0.001878 | 2.07E-12 | TRUE | 1.06E-12 | ++ | 0.000699 | 1 | 0.554383 | 1 | 1.63E-11 | 1 |
| 8-80570527-GC-G | G | | | | | | | | | 0.001418 | 3.46E-16 | TRUE | 3.46E-16 | ?+ | 0.074688 | 1 | 0.460127 | 1 | 2.61E-15 | 1 |
| 8-83440735-C-CT | CT | | | | | | | | | 0.045934 | 1.35E-13 | FALSE | 1.35E-13 | ?+ | 0.104764 | 1 | 0.012082 | 1 | 6.07E-12 | 1 |
| 8-90506149-C-A | A | | | | | | | 0.000351 | 0.9022 | 0.00234 | 2.95E-22 | TRUE | 3.04E-19 | -+ | 0.005746 | 1 | 0.000951 | 1 | 8.1E-18 | 1 |
| 8-90506156-C-T | T | 0.000214 | 0.95254 | | | 0.9525 | +? | 9.71E-05 | 0.532693 | 0.00096 | 5.8E-09 | TRUE | 9.78E-09 | ++ | 0.001208 | 1 | 0.451678 | 1 | 9.71E-08 | 1 |
| 8-90875247-C-T | T | 0.000236 | 0.025776 | | | 0.02578 | +? | 0.000865 | 0.326151 | 0.001549 | 2.01E-08 | TRUE | 1.01E-06 | ++ | 0.377795 | 1 | 0.003343 | 1 | 1.49E-06 | 1 |
| 8-93926331-C-CA | CA | 0.000234 | 0.481793 | | | 0.4818 | +? | | | 0.001992 | 1.41E-09 | TRUE | 1.41E-09 | ?+ | 0.395328 | 1 | 0.369245 | 1 | 4.09E-09 | 1 |
| 8-93926333-G-GCAT | GCAT | 0.000216 | 0.343382 | | | 0.3434 | -? | | | 0.002363 | 1.46E-14 | TRUE | 1.46E-14 | ?- | 0.271483 | 1 | 0.022367 | 1 | 1.92E-12 | 1 |

| | | | | | | | | | | | | | | | | | | | | |
|---|---|---|---|---|---|---|---|---|---|---|---|---|---|---|---|---|---|---|---|---|
| 8-94591747-CT-C | C | | | | | | | | | 0.001002 | 4.39E-09 | TRUE | 4.39E-09 | ?- | 0.000614 | 1 | 0.665423 | 1 | 4.32E-08 | 1 |
| 8-97732251-ACT-A | A | | | | | | | | | 0.000507 | 4.08E-10 | TRUE | 4.08E-10 | ?+ | 4.64E-05 | 1 | 0.079534 | 1 | 3.7E-07 | 1 |
| 8-99128207-G-GGT | GGT | | | | | | | | | 0.002024 | 1.61E-17 | TRUE | 1.61E-17 | ?- | 0.003367 | 1 | 0.586291 | 1 | 1.79E-14 | 1 |
| 8-101230534-T-TG | TG | 0.000176 | 0.903431 | | | 0.9034 | -? | | | 0.001617 | 1.81E-08 | TRUE | 1.81E-08 | ?- | 0.045887 | 1 | 0.46451 | 1 | 4.56E-07 | 1 |
| 8-102240933-A-G | G | 0.000156 | 0.987232 | | | 0.9872 | -? | 0.000489 | 0.543149 | 0.001358 | 4.23E-11 | TRUE | 1.91E-07 | +- | 0.002077 | 1 | 0.215054 | 1 | 1.26E-07 | 1 |
| 8-102887720-ATTT-A | A | | | | | | | | | 0.002123 | 6.83E-09 | TRUE | 6.83E-09 | ?+ | 0.00563 | 1 | 0.066062 | 1 | 1.2E-05 | 1 |
| 8-104680838-G-GGAG | GGAGTTC | 0.000137 | 0.133182 | | | 0.1332 | -? | | | 0.001398 | 2.62E-09 | TRUE | 2.62E-09 | ?- | 0.692866 | 1 | 0.307924 | 1 | 5.36E-09 | 1 |
| 8-125402911-AAG-A | A | 0.000274 | 0.145531 | | | 0.1455 | -? | | | 0.00056 | 2.88E-08 | TRUE | 2.89E-08 | ?+ | 0.007402 | 1 | 0.185434 | 1 | 1.23E-07 | 1 |
| 8-127843609-AT-A | A | | | | | | | 0.002215 | 0.945532 | 0.001608 | 3.58E-12 | TRUE | 1.47E-05 | ++ | 0.208964 | 1 | 0.239285 | 1 | 2.34E-12 | 1 |
| 8-130031600-C-CT | CT | 0.002355 | 0.302483 | | | 0.3025 | +? | | | 0.003869 | 1.72E-13 | TRUE | 1.72E-13 | ?+ | 0.709431 | 1 | 0.021115 | 1 | 1.82E-12 | 1 |
| 8-134499573-GGATG | G | | | | | | | | | 0.00396 | 1.16E-14 | TRUE | 1.16E-14 | ?+ | 0.007211 | 1 | 0.746313 | -1 | 2.2E-15 | 1 |
| 8-138085971-G-A | A | | | | | | | 0.001754 | 0.857856 | 0.000904 | 8.54E-09 | TRUE | 0.001963 | -+ | 0.06588 | 1 | 0.007659 | 1 | 7.72E-07 | 1 |
| 8-138743351-T-TTGAT | TTGATGG | | | | | | | | | 0.048552 | 1.59E-10 | FALSE | 1.59E-10 | ?- | 0.004492 | 1 | 0.162731 | 1 | 6.6E-08 | 1 |
| 8-142110390-AAAAG | A | | | | | | | | | 0.075065 | 3.56E-08 | FALSE | 3.56E-08 | ?+ | 0.02295 | 1 | 0.019134 | 1 | 1.23E-06 | 1 |
| 8-142223129-T-C | C | | | | | | | | | 0.133896 | 7.54E-12 | FALSE | 7.54E-12 | ?- | 0.709805 | 1 | 0.000554 | 1 | 5.32E-11 | 1 |
| 8-142223313-C-CTCAT | CTCATTCACTCACTCGT | | | | | | | | | 0.292244 | 1.18E-11 | FALSE | 1.18E-11 | ?- | 0.866452 | -1 | 0.027408 | -1 | 3.76E-13 | -1 |
| 8-142223313-C-T | T | | | | | | | | | 0.107472 | 2.43E-11 | FALSE | 2.43E-11 | ?+ | 0.016705 | 1 | 0.114179 | 1 | 1.08E-09 | 1 |
| 8-142804223-A-G | G | | | | | | | 0.602888 | 0.940732 | 0.687491 | 7.93E-09 | FALSE | 0.002053 | -- | 0.160499 | 1 | 0.600483 | -1 | 9.56E-10 | 1 |
| 8-143960466-A-AT | AT | | | | | | | | | 0.002752 | 2.76E-22 | TRUE | 2.77E-22 | ?- | 0.000217 | 1 | 0.003013 | 1 | 6.75E-16 | 1 |
| 8-144124603-CA-C | C | | | | | | | | | 0.002427 | 2E-22 | TRUE | 2E-22 | ?- | 4.15E-05 | 1 | 0.233853 | 1 | 3.82E-19 | 1 |
| 8-144216367-A-G | G | | | 0.024 | 0.663912 | 0.6639 | ?+ | | | 0.070699 | 5.92E-10 | TRUE | 5.92E-10 | ?- | 0.346442 | 1 | 0.127208 | -1 | 9.26E-09 | 1 |
| 8-144226688-A-G | G | | | | | | | 0.00036 | 0.821745 | 0.000584 | 1.39E-08 | TRUE | 1.49E-05 | +- | 0.002233 | 1 | 0.015587 | 1 | 3.45E-06 | 1 |
| 8-144301660-G-T | T | | | | | | | | | 0.00024 | 4.33E-08 | TRUE | 4.33E-08 | ?+ | 0.43282 | 1 | | | 2.16E-08 | 1 |
| 8-144626587-T-G | G | | | | | | | | | 0.858009 | 1.58E-13 | FALSE | 1.58E-13 | ?+ | 0.483455 | 1 | 0.207061 | -1 | 5.6E-16 | -1 |
| 8-144693358-C-CAGC | CAGCCGCCT | | | | | | | 0.002456 | 0.695519 | 0.006452 | 7.86E-09 | TRUE | 1.59E-05 | -+ | 0.957536 | -1 | 0.370724 | 1 | 3.04E-09 | 1 |
| 8-144817546-GGCCG | G | | | | | | | | | 0.063828 | 7.8E-18 | FALSE | 7.8E-18 | ?+ | 0.026814 | 1 | 0.162415 | 1 | 3.22E-17 | 1 |
| 8-22398364-C-T | T | | | | | | | 9.4E-06 | 0.753369 | | | FALSE | 1.5E-305 | -? | | | | | | |
| 8-61598069-C-A | A | | | | | | | 2.19E-05 | 0.719668 | 0.000225 | 0.041659 | FALSE | Inf | +- | 0.086195 | 1 | | | | |
| 8-56147154-T-A | A | | | | | | | 0.000103 | 0.995411 | | | FALSE | 1.81E-54 | +? | | | | | | |
| 9-2528493-A-AC | AC | | | | | | | | | 0.000911 | 6.48E-11 | TRUE | 6.48E-11 | ?- | 0.35746 | 1 | 0.08024 | 1 | 9.21E-09 | 1 |
| 9-6626205-A-C | C | 0.002427 | 0.855775 | | | 0.8558 | -? | 0.002731 | 0.942914 | 0.001631 | 6.66E-15 | TRUE | 5.01E-06 | -- | 0.751201 | -1 | 0.675151 | 1 | 5.84E-16 | 1 |
| 9-6662926-CTG-C | C | | | | | | | 4.07E-05 | 0.818951 | 0.0017 | 5.57E-09 | TRUE | 1.29E-08 | +- | 0.718711 | 1 | 0.657932 | 1 | 5.35E-09 | 1 |
| 9-6698604-GCC-G | G | | | | | | | | | 0.000231 | 3E-09 | TRUE | 3E-09 | ?+ | | | 0.015909 | 1 | 1.38E-06 | 1 |
| 9-16934631-TTC-T | T | 0.001605 | 0.663819 | | | 0.6638 | +? | | | 0.003026 | 6.26E-09 | TRUE | 6.26E-09 | ?+ | 0.688625 | 1 | 0.047237 | 1 | 2.19E-08 | 1 |
| 9-26493399-A-G | G | | | 0.49 | 0.848995 | 0.849 | ?+ | 0.543695 | 0.334928 | 0.489736 | 4.67E-09 | FALSE | 4.88E-05 | -- | 0.152791 | 1 | 0.014421 | 1 | 1.58E-07 | 1 |
| 9-26890151-CCT-C | C | | | | | | | | | 0.001047 | 6.31E-10 | TRUE | 6.31E-10 | ?- | 0.001406 | 1 | 0.094026 | 1 | 1.14E-07 | 1 |
| 9-31320715-CGG-C | C | | | | | | | | | 0.000186 | 7.12E-09 | TRUE | 7.12E-09 | ?- | 0.371405 | 1 | | | 8.1E-07 | 1 |
| 9-31320722-T-TCAGC | TCAGC | | | | | | | | | 0.00024 | 1.6E-09 | TRUE | 1.6E-09 | ?- | | | 0.024264 | 1 | 2.37E-09 | 1 |
| 9-33744596-A-T | T | | | | | | | 0.000501 | 0.186294 | 0.001324 | 7.92E-15 | TRUE | 4.15E-13 | -- | 0.00128 | 1 | 0.092332 | 1 | 8.58E-12 | 1 |
| 9-34034655-A-ACC | ACC | | | | | | | 6.58E-05 | 0.836381 | 0.00073 | 2.09E-15 | TRUE | 1.07E-14 | -- | 0.200863 | 1 | 0.53945 | 1 | 3.57E-16 | 1 |
| 9-34042314-C-CA | CA | | | | | | | | | 0.001075 | 4.82E-08 | TRUE | 4.82E-08 | ?+ | 0.314201 | 1 | 0.015366 | 1 | 2.08E-06 | 1 |
| 9-34042322-CA-C | C | | | | | | | | | 0.001887 | 1.86E-09 | TRUE | 1.86E-09 | ?- | 0.12801 | 1 | 0.045112 | 1 | 1.8E-08 | 1 |
| 9-34094697-AT-A | A | 0.000176 | 0.304488 | | | 0.3045 | -? | 0.00094 | 0.369773 | 0.001893 | 2.95E-15 | TRUE | 8.96E-09 | -+ | 0.133154 | 1 | 0.00163 | 1 | 1.14E-11 | 1 |
| 9-36095857-G-C | C | | | | | | | 0.001385 | 0.09361 | 0.001127 | 4.21E-08 | TRUE | 1.94E-06 | ++ | 0.2395 | 1 | 0.781365 | -1 | 6.23E-09 | 1 |
| 9-36182925-C-T | T | 0.008092 | 0.107993 | 0.003 | 0.116664 | 0.1111 | +- | 0.00301 | 0.43582 | 0.011343 | 4.45E-10 | TRUE | 2.16E-05 | -+ | 0.389442 | 1 | 0.012127 | 1 | 1.18E-12 | 1 |
| 9-36604110-C-T | T | | | | | | | 0.000388 | 0.387594 | 0.002247 | 8.92E-19 | TRUE | 1.16E-14 | -+ | 0.00074 | 1 | 0.080798 | 1 | 2.05E-15 | 1 |
| 9-38688643-G-C | C | | | | | | | | | 0.000968 | 1.34E-08 | TRUE | 1.34E-08 | ?+ | 0.356523 | 1 | 0.314371 | 1 | 7.64E-07 | 1 |
| 9-39145455-G-A | A | 0.001209 | 0.377829 | | | 0.3778 | -? | | | 0.015623 | 4.19E-08 | FALSE | 4.19E-08 | ?+ | 0.533183 | 1 | 0.370379 | 1 | 3.55E-08 | 1 |
| 9-39662906-G-A | A | | | | | | | | | 0.891426 | 7.52E-36 | FALSE | 7.52E-36 | ?- | 0.094196 | -1 | 0.218929 | -1 | 2.71E-40 | -1 |
| 9-39941434-A-AT | AT | | | | | | | | | 0.870333 | 3.34E-25 | TRUE | 3.34E-25 | ?+ | 0.016391 | -1 | 0.001163 | -1 | 2.34E-21 | -1 |
| 9-40329787-CTT-C | C | | | | | | | | | 0.501963 | 3.39E-16 | FALSE | 3.39E-16 | ?+ | 0.009075 | -1 | 0.030695 | -1 | 9.98E-13 | -1 |
| 9-40329787-CTTTT-C | C | | | | | | | | | 0.032255 | 2.31E-12 | TRUE | 2.31E-12 | ?- | 0.01232 | 1 | 0.265884 | 1 | 8.61E-10 | 1 |
| 9-40557771-T-TC | TC | | | | | | | | | 0.986054 | 3.56E-08 | TRUE | 3.56E-08 | ?+ | 0.862702 | -1 | 0.182852 | -1 | 8.71E-08 | -1 |
| 9-40894485-C-G | G | | | | | | | | | 0.000845 | 5.6E-10 | TRUE | 5.6E-10 | ?- | 0.020502 | 1 | 0.131351 | 1 | 6.41E-09 | 1 |
| 9-41353766-C-A | A | | | | | | | | | 0.549031 | 3.59E-12 | FALSE | 3.59E-12 | ?+ | 0.703317 | -1 | 0.052728 | 1 | 2.6E-09 | 1 |
| 9-41727997-T-TA | TA | | | | | | | | | 0.152004 | 4.05E-09 | TRUE | 4.05E-09 | ?- | 0.000781 | 1 | 0.148043 | 1 | 2E-06 | 1 |
| 9-41727997-T-TAA | TAA | | | | | | | | | 0.80165 | 2.15E-08 | FALSE | 2.15E-08 | ?+ | 0.004092 | -1 | 0.088206 | -1 | 7.6E-06 | -1 |
| 9-41848995-ATTT-A | A | | | | | | | | | 0.430571 | 2.17E-12 | FALSE | 2.17E-12 | ?+ | 0.021998 | 1 | 0.14971 | 1 | 1.67E-10 | 1 |
| 9-42847389-AG-A | A | | | | | | | | | 0.926707 | 9.79E-09 | TRUE | 9.79E-09 | ?+ | 0.644168 | -1 | 0.136144 | 1 | 5.08E-11 | 1 |

| | | | | | | | | | | | | | | | | | | | | |
|---|---|---|---|---|---|---|---|---|---|---|---|---|---|---|---|---|---|---|---|---|
| 9-60609104-G-A | A | | | | | | | | | 0.003752 | 1.28E-11 | FALSE | 1.28E-11 | ?+ | 0.032785 | 1 | 0.78944 | -1 | 8.23E-12 | 1 |
| 9-61670607-TAG-T | T | 0.010874 | 0.79589 | | | 0.7959 | +? | | | 0.004148 | 2.38E-10 | TRUE | 2.38E-10 | ?+ | 0.851884 | 1 | 0.592257 | 1 | 5.88E-11 | 1 |
| 9-61975304-C-CTTTTT | CTTTTTTT | | | | | | | | | 0.046304 | 8.65E-12 | FALSE | 8.65E-12 | ?+ | 0.067341 | 1 | 0.262244 | 1 | 1.37E-11 | 1 |
| 9-61975304-C-CTTTTT | CTTTTTTTT | | | | | | | | | 0.037904 | 1.29E-08 | FALSE | 1.29E-08 | ?+ | 0.917909 | -1 | 0.008581 | 1 | 2.24E-07 | 1 |
| 9-61975304-CT-C | C | | | | | | | | | 0.59409 | 4.71E-22 | FALSE | 4.7E-22 | ?+ | 0.00347 | -1 | 0.044743 | -1 | 4.6E-20 | -1 |
| 9-62838432-C-CCGGC | CCGGCGCCCCCTCCGCGCCGGCGCCCCCTCCGCGCCGGCGCCCCCTCCGCGCCGGCGCCCGGCGCT | | | | | | | | | 0.0035 | 3.66E-09 | TRUE | 3.66E-09 | ?- | 0.429863 | -1 | 0.489676 | -1 | 1.56E-08 | -1 |
| 9-64383520-TCATATT | T | | | | | | | | | 0.952091 | 1.19E-15 | FALSE | 1.19E-15 | ?- | 0.92063 | -1 | 0.910333 | -1 | 6.76E-22 | -1 |
| 9-64469826-CTTTTTTT | C | | | | | | | | | 0.235392 | 2.41E-14 | FALSE | 2.42E-14 | ?+ | 0.037335 | -1 | 0.001369 | -1 | 1.22E-10 | -1 |
| 9-66029264-A-C | C | | | | | | | | | 0.02437 | 1.69E-12 | FALSE | 1.69E-12 | ?+ | 0.041561 | -1 | 0.117958 | -1 | 1.81E-11 | -1 |
| 9-66754343-CTT-C | C | | | | | | | | | 0.230864 | 3.67E-09 | FALSE | 3.67E-09 | ?- | 0.892726 | -1 | 0.347331 | 1 | 8.47E-10 | 1 |
| 9-67090646-TC-T | T | | | | | | | | | 0.943185 | 6.27E-11 | FALSE | 6.27E-11 | ?+ | 0.987656 | -1 | 0.229218 | 1 | 1.39E-16 | 1 |
| 9-67632973-C-CAA | CAA | | | | | | | | | 0.019197 | 4.17E-08 | TRUE | 4.17E-08 | ?+ | 0.324506 | 1 | 0.00776 | 1 | 2.14E-06 | 1 |
| 9-68291643-G-C | C | | | | | | | | | 0.820742 | 1.86E-12 | FALSE | 1.86E-12 | ?- | 0.383659 | -1 | 0.681208 | -1 | 1.69E-13 | -1 |
| 9-68291643-G-T | T | | | | | | | | | 0.051542 | 3.02E-32 | FALSE | 3.02E-32 | ?+ | 0.009202 | 1 | 0.000313 | 1 | 1.55E-28 | 1 |
| 9-73226646-A-G | G | | | | | | | 0.000927 | 0.19383 | 0.001223 | 2.13E-11 | TRUE | 7.22E-09 | -- | 0.001378 | 1 | 0.948804 | -1 | 1.23E-10 | 1 |
| 9-74197756-G-GAGGT | GAGGTGGGGGGGTC | | | | | | | | | 0.735767 | 3.32E-10 | FALSE | 3.32E-10 | ?- | 0.406848 | 1 | 0.489085 | 1 | 1.33E-10 | 1 |
| 9-74197756-G-GAGGT | GAGGTGGGGGGGTCAGCCCCCCGCCCGGCCAGCCGCCCCATCCGGGAGGGAGGTGGGGGGGTC | | | | | | | | | 0.173205 | 2.17E-08 | FALSE | 2.17E-08 | ?+ | 0.366386 | -1 | 0.990358 | 1 | 1.56E-09 | -1 |
| 9-74786521-C-CT | CT | | | | | | | 0.00052 | 0.437918 | 0.002598 | 6.57E-13 | TRUE | 1.32E-11 | ++ | 0.354515 | 1 | 0.221444 | 1 | 4.7E-13 | 1 |
| 9-75417474-A-G | G | 0.392523 | 0.477076 | 0.33 | 0.959098 | 0.4808 | +- | 0.316094 | 0.683794 | 0.423374 | 1.29E-10 | FALSE | 6.31E-05 | -- | 0.05844 | 1 | 0.194126 | 1 | 2.02E-07 | 1 |
| 9-82464932-T-C | C | 0.000471 | 0.179167 | | | 0.1792 | +? | 0.000442 | 0.317783 | 0.001373 | 3.48E-13 | TRUE | 1.23E-08 | +- | 0.007401 | 1 | 0.55787 | 1 | 7.99E-12 | 1 |
| 9-84449299-C-T | T | | | | | | | 0.000172 | 0.160959 | 0.000852 | 2.46E-09 | TRUE | 1.87E-09 | ++ | 0.034002 | 1 | 0.146758 | 1 | 1.39E-08 | 1 |
| 9-87415274-A-G | G | | | | | | | 0.268596 | 0.998034 | 0.422969 | 6.42E-09 | FALSE | 0.000795 | -- | 0.955192 | 1 | 0.051743 | 1 | 2.39E-06 | 1 |
| 9-92785200-GT-G | G | | | | | | | | | 0.000437 | 8.51E-10 | TRUE | 8.51E-10 | ?+ | 3.71E-05 | 1 | 0.094441 | 1 | 4.72E-06 | 1 |
| 9-93166066-A-AT | AT | 0.000176 | 0.720736 | | | 0.7207 | +? | | | 0.001888 | 3.87E-16 | TRUE | 3.87E-16 | ?- | 0.015997 | 1 | 5.7E-05 | 1 | 1.15E-12 | 1 |
| 9-93211257-T-TCCACT | TCCACTCACTTATTCACTTACACATTCACTCACTCATTTACTCACTCATCCACTTACTCATTCACTCACTCATTCATCCA | | | | | | | | | 0.668925 | 5.74E-11 | FALSE | 5.74E-11 | ?+ | 0.448419 | -1 | 0.449293 | -1 | 2.2E-10 | -1 |
| 9-94616642-CA-C | C | | | | | | | | | 0.000412 | 1.83E-15 | TRUE | 1.84E-15 | ?- | 0.015352 | 1 | 0.313277 | 1 | 3.73E-14 | 1 |
| 9-94616646-GC-G | G | | | | | | | | | 0.000545 | 5.29E-13 | TRUE | 5.29E-13 | ?+ | 0.035982 | 1 | 0.069027 | 1 | 1.42E-11 | 1 |
| 9-94616648-A-ATG | ATG | | | | | | | | | 0.00067 | 1.5E-14 | TRUE | 1.5E-14 | ?- | 0.015989 | 1 | 0.2046 | 1 | 4.99E-13 | 1 |
| 9-96946504-C-CTT | CTT | | | | | | | | | 0.001633 | 2.09E-08 | TRUE | 2.09E-08 | ?+ | 0.01852 | 1 | 0.016461 | 1 | 3.52E-06 | 1 |
| 9-104150373-GGC-G | G | | | | | | | | | 0.000885 | 1.77E-09 | TRUE | 1.77E-09 | ?+ | 0.923824 | 1 | 0.015588 | 1 | 8.93E-09 | 1 |
| 9-104150379-T-TCC | TCC | | | | | | | | | 0.000932 | 2.23E-08 | TRUE | 2.23E-08 | ?- | 0.933743 | -1 | 0.011524 | 1 | 1.12E-07 | 1 |
| 9-110014597-A-AT | AT | | | | | | | | | 0.000622 | 1.22E-11 | TRUE | 1.22E-11 | ?- | 0.015299 | 1 | 0.011769 | 1 | 8.9E-10 | 1 |
| 9-116231766-C-T | T | | | | | | | 0.000194 | 0.946044 | 0.030355 | 1.19E-15 | FALSE | 1.66E-15 | -+ | 0.398291 | 1 | 0.429784 | 1 | 4.83E-17 | 1 |
| 9-116936095-A-ACCA | ACCAT | | | | | | | | | 0.036135 | 3.78E-08 | FALSE | 3.78E-08 | ?+ | 0.017885 | -1 | 0.256621 | -1 | 3.52E-06 | -1 |
| 9-117005080-A-AGTT | AGTT | | | | | | | | | 0.028457 | 2.1E-12 | FALSE | 2.1E-12 | ?- | 0.500245 | 1 | 0.240156 | 1 | 5.95E-12 | 1 |
| 9-118668202-CAG-C | C | | | | | | | | | 0.00076 | 1.49E-08 | TRUE | 1.49E-08 | ?- | 0.000569 | 1 | 0.217491 | 1 | 2.21E-06 | 1 |
| 9-123399029-CAA-C | C | | | | | | | | | 0.004118 | 2.37E-09 | TRUE | 2.37E-09 | ?- | 0.226793 | 1 | 0.691669 | 1 | 2.9E-08 | 1 |
| 9-123706553-C-CCT | CCT | | | | | | | | | 0.001721 | 5.43E-17 | TRUE | 5.43E-17 | ?+ | 0.000113 | 1 | 0.016757 | 1 | 1.39E-12 | 1 |
| 9-123917954-CAAAA- | C | | | | | | | | | 0.005433 | 3.66E-10 | TRUE | 3.66E-10 | ?- | 0.011152 | 1 | 0.10217 | 1 | 4.21E-08 | 1 |
| 9-124588845-G-A | A | | | | | | | 0.002074 | 0.004077 | 0.000833 | 6.28E-10 | TRUE | 0.3409 | -+ | 0.104285 | 1 | 0.009241 | 1 | 8.29E-08 | 1 |
| 9-125592686-C-CCT | CCT | | | | | | | 0.000219 | 0.86742 | 0.002607 | 1.04E-10 | TRUE | 1.38E-09 | -+ | 0.838201 | -1 | 0.019254 | 1 | 2.13E-10 | 1 |
| 9-125689462-A-C | C | | | | | | | 0.000949 | 0.948883 | 0.036145 | 4.52E-11 | TRUE | 1.09E-10 | -- | 0.317114 | -1 | 0.59561 | 1 | 4.6E-12 | 1 |
| 9-126014359-C-A | A | | | | | | | 0.00068 | 0.201084 | 0.001375 | 3.98E-10 | TRUE | 8.4E-09 | ++ | 0.932413 | -1 | 0.89449 | 1 | 1.55E-10 | 1 |
| 9-128665286-C-CAG | CAG | | | | | | | | | 0.000771 | 3.66E-08 | TRUE | 3.66E-08 | ?+ | 0.000483 | 1 | 0.786542 | 1 | 5.66E-06 | 1 |
| 9-129033772-G-GAGA | GAGA | | | | | | | | | 0.000975 | 9.75E-10 | TRUE | 9.75E-10 | ?- | 0.871833 | 1 | 0.007364 | 1 | 1.51E-08 | 1 |
| 9-129539014-A-G | G | 0.489675 | 0.734466 | 0.414 | 0.593606 | 0.7664 | -+ | 0.417323 | 0.199215 | 0.493536 | 7.12E-12 | FALSE | 0.006722 | +- | 0.247648 | 1 | 0.468952 | 1 | 7.75E-09 | 1 |
| 9-129727696-C-CCT | CCT | | | | | | | | | 0.002493 | 5.56E-23 | TRUE | 5.55E-23 | ?+ | 0.006499 | 1 | 0.015586 | 1 | 8.44E-21 | 1 |
| 9-129980341-CA-C | C | | | | | | | | | 0.000348 | 2.03E-09 | TRUE | 2.03E-09 | ?- | 0.147082 | 1 | 0.020363 | 1 | 1.48E-07 | 1 |
| 9-130373707-A-AGAT | AGATGGATGGATG | | | | | | | | | 0.177188 | 9.15E-10 | FALSE | 9.15E-10 | ?+ | 0.973315 | 1 | 0.410283 | -1 | 8.75E-09 | -1 |
| 9-130955273-C-CACC | CACCGTT | | | | | | | | | 0.004685 | 7.89E-10 | TRUE | 7.89E-10 | ?+ | 0.003419 | 1 | 0.266712 | 1 | 3.25E-08 | 1 |
| 9-131326169-T-C | C | 0.000157 | 0.30655 | | | 0.3065 | +? | | | 0.000455 | 2.49E-09 | TRUE | 2.49E-09 | ?- | 0.022278 | 1 | 0.018367 | 1 | 1.21E-07 | 1 |
| 9-131326190-A-AGCC | AGCCTGGTC | | | | | | | | | 0.001707 | 6.53E-09 | TRUE | 6.53E-09 | ?- | 0.000132 | 1 | 0.002104 | 1 | 3.3E-05 | 1 |
| 9-133482173-C-T | T | | | | | | | 0.000179 | 0.141773 | 0.000762 | 1.43E-09 | TRUE | 1.13E-09 | ++ | 0.044476 | 1 | 0.737205 | 1 | 7.46E-09 | 1 |
| 9-133482205-C-G | G | 0.000117 | 0.860764 | | | 0.8608 | -? | 0.000172 | 0.315331 | 0.001886 | 2.43E-15 | TRUE | 5.55E-13 | +- | 0.000165 | 1 | 0.161821 | 1 | 9.61E-13 | 1 |
| 9-134464507-CCT-C | C | | | | | | | 0.002287 | 0.72818 | 0.004297 | 3.17E-11 | TRUE | 1.23E-06 | +- | 0.752078 | 1 | 0.263596 | 1 | 4.92E-10 | 1 |
| 9-134599423-T-TACA | TACAC | | | | | | | | | 0.524376 | 7.22E-13 | FALSE | 7.22E-13 | ?- | 0.925032 | -1 | 0.702488 | 1 | 1.04E-09 | 1 |
| 9-135673019-TTTTG-T | T | | | | | | | | | 0.000677 | 3.38E-09 | TRUE | 3.38E-09 | ?+ | 0.128554 | 1 | 0.26662 | 1 | 2.82E-09 | 1 |
| 9-136639478-C-T | T | 0.001135 | 0.920601 | | | 0.9206 | -? | 0.000263 | 0.145337 | 0.000605 | 2.09E-10 | TRUE | 7.97E-10 | ++ | 0.001375 | 1 | 0.921547 | 1 | 3.92E-09 | 1 |
| 9-137334886-G-C | C | | | | | | | 0.000166 | 0.424351 | 0.00203 | 2.17E-15 | TRUE | 1.97E-13 | -+ | 0.000621 | 1 | 0.023852 | 1 | 1.05E-12 | 1 |

| | | | | | | | | | | | | | | | | | | | | |
|---|---|---|---|---|---|---|---|---|---|---|---|---|---|---|---|---|---|---|---|---|
| 9-137334905-A-G | G | | | | | | | 2.19E-05 | 0.822202 | 0.000541 | 2.59E-08 | TRUE | 6.54E-08 | +- | 0.035631 | 1 | 0.038574 | 1 | 8.17E-07 | 1 |
| 9-105169701-G-A | A | 0.000156 | 0.135724 | | | 0.1357 | -? | 0.000348 | 0.036444 | 0.00085 | 6.39E-08 | TRUE | 2.11E-08 | ++ | 0.337179 | 1 | 0.017291 | 1 | 2.02E-07 | 1 |
| 9-81131033-TAGAGA( | T | | | | | | | | | | | FALSE | | | | | | | | |
| 9-105169723-C-T | T | | | | | | | 5.32E-05 | 0.019272 | 0.000643 | 4.41E-07 | TRUE | 2.87E-08 | -- | 0.087712 | 1 | 0.025204 | 1 | 1.19E-05 | 1 |
| 9-8630638-G-C | C | | | | | | | 2.51E-05 | 1.06E-05 | 8.78E-05 | 0.000132 | TRUE | 2.2E-08 | -- | | | | | | |
| 9-41290198-G-T | T | | | | | | | 2.83E-05 | 0.913271 | | | FALSE | 3.27E-44 | -? | | | | | | |
| 10-80678507-T-C | C | 0.081378 | 0.129684 | 0.097 | 0.230775 | 0.1162 | -- | 0.893973 | 4.04E-08 | 0.912057 | 0.49388 | TRUE | 4.78E-05 | -+ | 0.766524 | -1 | 0.125469 | -1 | 0.554378 | -1 |
| 10-347561-C-T | T | | | | | | | | | 0.89993 | 8.26E-23 | FALSE | 8.26E-23 | ?- | 0.071831 | -1 | 0.047867 | -1 | 1.88E-20 | -1 |
| 10-448762-A-G | G | | | | | | | | | 0.814657 | 9.59E-10 | FALSE | 9.59E-10 | ?+ | 0.562578 | -1 | 0.541916 | -1 | 1.34E-09 | -1 |
| 10-976272-A-G | G | | | | | | | 0.992345 | 0.950602 | 0.986762 | 5.22E-10 | TRUE | 2.12E-05 | ++ | 0.538212 | 1 | 0.669311 | -1 | 1.19E-12 | -1 |
| 10-1340863-A-G | G | | | | | | | | | 0.868816 | 1.07E-51 | FALSE | 1.07E-51 | ?+ | 0.133027 | -1 | 0.023718 | -1 | 7.16E-52 | -1 |
| 10-1593285-ATAGGT( | A | | | | | | | | | 0.011427 | 2.32E-20 | TRUE | 2.32E-20 | ?+ | 0.864498 | 1 | 0.045202 | 1 | 4E-20 | 1 |
| 10-1674997-A-ATTCT( | ATTCTGGAGGTTTGGGTTTGGGGGTACATGGATGTTCTGGAGGTTTGGGTTCGGGGATGCATGGATG | | | | | | | | | 0.78434 | 1.93E-09 | FALSE | 1.93E-09 | ?+ | 0.077969 | 1 | 0.100588 | -1 | 2.25E-10 | -1 |
| 10-1674997-A-G | G | | | | | | | | | 0.057627 | 4.84E-08 | FALSE | 4.84E-08 | ?- | 0.122383 | -1 | 0.110838 | 1 | 1.04E-08 | 1 |
| 10-2523555-G-GGGT( | GGGTGACATCCATACCCACACCTTATCCGGGTGACACCCACATCCACACCTTATCCA | | | | | | | | | 0.185733 | 7.52E-13 | FALSE | 7.52E-13 | ?+ | 0.637708 | -1 | 0.710855 | -1 | 4.31E-13 | -1 |
| 10-2983675-A-G | G | 0.000273 | 0.242806 | | | 0.2428 | +? | | | 0.001016 | 1.85E-08 | TRUE | 1.85E-08 | ?- | 0.086788 | 1 | 0.008398 | 1 | 7.33E-07 | 1 |
| 10-2983682-A-C | C | 0.000274 | 0.045904 | | | 0.0459 | +? | 3.76E-05 | 0.903154 | 0.000834 | 2.83E-10 | TRUE | 5.87E-10 | -- | 0.150622 | 1 | 0.000814 | 1 | 2.84E-08 | 1 |
| 10-2983692-T-A | A | 0.000293 | 0.124866 | | | 0.1249 | -? | 3.76E-05 | 0.902933 | 0.000839 | 2.16E-10 | TRUE | 4.47E-10 | ++ | 0.02149 | 1 | 0.018967 | 1 | 5.35E-09 | 1 |
| 10-5378108-A-ATGGT | ATGGTG | | | | | | | | | 0.000939 | 1.32E-12 | TRUE | 1.32E-12 | ?- | 0.268368 | 1 | 3.02E-05 | 1 | 2.31E-09 | 1 |
| 10-10099500-G-A | A | | | | | | | | | 0.001106 | 2.06E-11 | TRUE | 2.06E-11 | ?+ | 0.418235 | 1 | 0.200391 | 1 | 2.45E-11 | 1 |
| 10-12148087-C-CACG | CACG | | | | | | | | | 0.00129 | 1.71E-13 | TRUE | 1.71E-13 | ?+ | 0.107576 | 1 | 0.002413 | 1 | 1.6E-11 | 1 |
| 10-12151525-T-C | C | 0.005492 | 0.936409 | | | 0.9364 | -? | 0.993209 | 0.42829 | 0.992498 | 1.7E-09 | TRUE | 4.02E-05 | ++ | 0.993868 | -1 | 0.008835 | -1 | 5.92E-13 | -1 |
| 10-12151534-T-A | A | 0.00098 | 0.018428 | | | 0.01843 | -? | 0.005319 | 0.318508 | 0.001912 | 3.57E-14 | TRUE | 7.78E-06 | ++ | 0.151959 | 1 | 0.001449 | 1 | 5.23E-12 | 1 |
| 10-12151538-T-C | C | 0.001019 | 0.085789 | | | 0.08579 | +? | 0.006214 | 0.931829 | 0.002377 | 8.2E-12 | TRUE | 0.001644 | +- | 0.043298 | 1 | 0.005125 | 1 | 3.94E-09 | 1 |
| 10-12151551-C-T | T | 0.001 | 0.677692 | | | 0.6777 | +? | 0.006214 | 0.931829 | 0.001939 | 2.56E-12 | TRUE | 0.002088 | -+ | 0.057525 | 1 | 0.008427 | 1 | 1.88E-11 | 1 |
| 10-12151556-T-C | C | 0.001216 | 0.663118 | | | 0.6631 | -? | 0.006214 | 0.931829 | 0.002035 | 6.44E-17 | TRUE | 0.000125 | +- | 0.00101 | 1 | 0.000105 | 1 | 3.35E-12 | 1 |
| 10-12279639-CT-C | C | | | | | | | | | 0.000211 | 1.67E-10 | TRUE | 1.67E-10 | ?- | 0.001512 | 1 | | | 7E-09 | 1 |
| 10-12380003-AG-A | A | | | | | | | 0.000351 | 0.936821 | 0.001939 | 2.4E-10 | TRUE | 8.22E-09 | ++ | 0.002159 | 1 | 0.883406 | 1 | 1.38E-09 | 1 |
| 10-13063272-T-TAGT( | TAGTGCTGATTGGTCAGAGA | | | | | | | | | 0.46509 | 2.17E-37 | FALSE | 2.17E-37 | ?+ | 0.020132 | -1 | 0.00175 | -1 | 4.93E-33 | -1 |
| 10-13313766-CT-C | C | 0.001246 | 0.323619 | | | 0.3236 | -? | 0.002258 | 0.541077 | 0.001835 | 6.96E-09 | TRUE | 0.001843 | +- | 0.025717 | 1 | 0.857931 | 1 | 3.11E-08 | 1 |
| 10-14933460-AGCCCC | A | | | | | | | | | 0.000432 | 2.11E-10 | TRUE | 2.11E-10 | ?+ | 0.004565 | 1 | 0.016374 | 1 | 9.79E-09 | 1 |
| 10-14933466-A-ATGT( | ATGTCG | | | | | | | | | 0.000254 | 4.68E-09 | TRUE | 4.68E-09 | ?- | 0.022787 | 1 | 0.016382 | 1 | 1.13E-07 | 1 |
| 10-17983189-A-ACCT/ | ACCTAG | | | | | | | | | 0.000846 | 3.85E-08 | TRUE | 3.85E-08 | ?- | 0.661196 | -1 | 0.138055 | 1 | 2.57E-08 | 1 |
| 10-18396114-G-A | A | 0.000666 | 0.993526 | | | 0.9935 | -? | 3.76E-05 | 0.825206 | 0.000779 | 3.42E-09 | TRUE | 5.86E-09 | ++ | 0.038508 | 1 | 0.242758 | 1 | 5.98E-08 | 1 |
| 10-22516671-C-A | A | | | | | | | | | 0.025945 | 9.84E-09 | FALSE | 9.84E-09 | ?- | 0.198564 | -1 | 0.388987 | -1 | 4.97E-08 | -1 |
| 10-23108707-T-C | C | | | | | | | | | 0.000328 | 1.46E-09 | TRUE | 1.46E-09 | ?- | 0.477802 | -1 | 0.031274 | 1 | 2.55E-08 | 1 |
| 10-23149772-G-A | A | | | | | | | | | 0.906389 | 2.39E-09 | FALSE | 2.39E-09 | ?- | 0.327958 | -1 | 0.158018 | -1 | 7.69E-09 | -1 |
| 10-27215821-TC-T | T | 0.000293 | 0.89118 | | | 0.8912 | -? | | | 0.001856 | 1.44E-11 | TRUE | 1.44E-11 | ?+ | 0.967383 | 1 | 0.018615 | 1 | 1.92E-10 | 1 |
| 10-27312007-CT-C | C | 0.248975 | 0.324623 | | | 0.3246 | +? | 0.240839 | 0.934769 | 0.283356 | 1.58E-08 | TRUE | 0.001388 | +- | 0.293772 | 1 | 0.01509 | 1 | 4.57E-06 | 1 |
| 10-31411638-C-A | A | | | | | | | | | 0.044806 | 2.8E-08 | FALSE | 2.8E-08 | ?- | 0.059416 | -1 | 0.000143 | -1 | 4.42E-06 | -1 |
| 10-35114079-TGGGTT | T | | | | | | | | | 0.002393 | 2.01E-18 | TRUE | 2.01E-18 | ?+ | 0.303926 | 1 | 0.001593 | 1 | 5.14E-15 | 1 |
| 10-35370323-T-G | G | | | | | | | 0.000911 | 0.700674 | 0.001795 | 1.44E-13 | TRUE | 8.64E-10 | -- | 0.00419 | 1 | 0.018517 | 1 | 5.52E-11 | 1 |
| 10-37902303-A-G | G | | | | | | | | | 0.033018 | 6.92E-11 | FALSE | 6.92E-11 | ?- | 0.419095 | 1 | 0.299273 | 1 | 9.93E-11 | 1 |
| 10-38930856-T-A | A | | | | | | | | | 0.002555 | 2.73E-10 | TRUE | 2.73E-10 | ?+ | 0.005393 | 1 | 0.902196 | 1 | 9.21E-10 | 1 |
| 10-39454303-C-CTG | CTG | | | | | | | | | 0.822376 | 3.94E-09 | FALSE | 3.94E-09 | ?- | 0.394493 | -1 | 0.134319 | -1 | 1.22E-09 | -1 |
| 10-39565455-C-A | A | | | | | | | 0.9859 | 0.456825 | 0.948503 | 1.31E-41 | TRUE | 5.96E-31 | -- | 0.002569 | -1 | 3.4E-05 | -1 | 8.33E-37 | -1 |
| 10-41772582-A-AAG | AAG | | | | | | | | | 0.000863 | 6.46E-14 | TRUE | 6.46E-14 | ?- | 0.807104 | -1 | 0.14031 | 1 | 9.47E-13 | 1 |
| 10-41772584-GCA-G | G | | | | | | | | | 0.000763 | 1.74E-12 | TRUE | 1.74E-12 | ?+ | 0.923628 | -1 | 0.017409 | 1 | 2.33E-11 | 1 |
| 10-41847867-AT-A | A | | | | | | | | | 0.003813 | 2.26E-12 | TRUE | 2.26E-12 | ?+ | 0.473666 | 1 | 0.000211 | 1 | 9.82E-10 | 1 |
| 10-41902876-A-AATG( | AATGG | | | | | | | | | 0.14682 | 1.07E-15 | FALSE | 1.07E-15 | ?+ | 0.480802 | -1 | 0.216775 | -1 | 4.66E-15 | -1 |
| 10-41902876-A-C | C | | | | | | | 0.976634 | 0.169485 | 0.816967 | 1.2E-08 | FALSE | 1.82E-06 | +- | 0.537096 | 1 | 0.368672 | 1 | 4.35E-08 | 1 |
| 10-42066423-AATC-A | A | | | | | | | | | 0.003215 | 5.97E-13 | TRUE | 5.97E-13 | ?+ | 0.007317 | 1 | 0.330413 | 1 | 5.38E-12 | 1 |
| 10-42093392-C-CT | CT | | | | | | | | | 0.898094 | 9.36E-87 | FALSE | 9.36E-87 | ?- | 0.000287 | -1 | 1.43E-08 | -1 | 7.12E-77 | -1 |
| 10-42102736-T-A | A | | | | | | | | | 0.002143 | 2.13E-09 | TRUE | 2.13E-09 | ?+ | 0.211754 | 1 | 0.004773 | 1 | 4.15E-08 | 1 |
| 10-42102765-C-G | G | | | | | | | | | 0.001615 | 1.44E-14 | TRUE | 1.44E-14 | ?- | 0.080401 | 1 | 5.18E-05 | 1 | 1.67E-11 | 1 |
| 10-67408882-C-A | A | | | | | | | | | 0.021952 | 3.28E-08 | FALSE | 3.28E-08 | ?- | 0.060048 | -1 | 0.046674 | -1 | 3.25E-06 | -1 |
| 10-68227393-G-T | T | | | | | | | 0.000213 | 0.702014 | 0.00112 | 6.3E-10 | TRUE | 8.03E-09 | ++ | 0.068972 | 1 | 0.25419 | 1 | 2E-09 | 1 |
| 10-68593595-A-G | G | 0.000157 | 0.381814 | | | 0.3818 | +? | 0.000937 | 0.347596 | 0.000762 | 6.61E-12 | TRUE | 0.000111 | +- | 0.000151 | 1 | 0.000166 | 1 | 6.21E-07 | 1 |
| 10-70420363-C-CT | CT | | | | | | | | | 0.000954 | 1.31E-11 | TRUE | 1.32E-11 | ?+ | 0.012585 | 1 | 0.000666 | 1 | 9.59E-08 | 1 |

| | | | | | | | | | | | | | | | | | | | | |
|---|---|---|---|---|---|---|---|---|---|---|---|---|---|---|---|---|---|---|---|---|
| 10-70420366-A-T | T | | | | | | | | | 0.0004 | 9.86E-09 | TRUE | 9.87E-09 | ?- | 0.03526 | 1 | 0.003029 | 1 | 1.1E-05 | 1 |
| 10-72191881-CAA-C | C | | | | | | | | | 0.002 | 6.46E-10 | TRUE | 6.46E-10 | ?- | 0.002871 | 1 | 0.127236 | 1 | 2.53E-08 | 1 |
| 10-73947669-TG-T | T | | | | | | | | | 0.000563 | 1.01E-10 | TRUE | 1.01E-10 | ?+ | 0.441009 | 1 | 0.055038 | 1 | 2.37E-10 | 1 |
| 10-74595727-G-A | A | | | | | | | | | 0.995444 | 4.32E-11 | TRUE | 4.32E-11 | ?- | 0.011511 | -1 | 0.027876 | -1 | 1.26E-08 | -1 |
| 10-74595729-TCC-T | T | | | | | | | | | 0.992877 | 7.23E-16 | TRUE | 7.23E-16 | ?- | 0.005375 | -1 | 0.00838 | -1 | 7.34E-12 | -1 |
| 10-74648493-A-C | C | | | | | | | 0.000611 | 0.801128 | 0.00052 | 2.32E-08 | TRUE | 0.000303 | +- | 0.014164 | 1 | 0.293523 | 1 | 1.89E-07 | 1 |
| 10-79504819-A-G | G | | | | | | | | | 0.004095 | 1.19E-12 | FALSE | 1.19E-12 | ?- | 0.261969 | 1 | 0.773653 | 1 | 4.6E-13 | 1 |
| 10-79922400-A-G | G | 0.00905 | 0.035563 | | | 0.03556 | -? | | | 0.008219 | 2E-13 | TRUE | 2E-13 | ?- | 0.024689 | 1 | 0.064294 | 1 | 3.55E-12 | 1 |
| 10-84087114-CGT-C | C | | | | | | | | | 0.00095 | 1.18E-10 | TRUE | 1.18E-10 | ?- | 0.457426 | 1 | 0.017469 | 1 | 2.07E-09 | 1 |
| 10-93848224-T-A | A | 0.457363 | 0.849418 | 0.487 | 0.383456 | 0.3835 | ?- | 0.453451 | 0.796657 | 0.58231 | 1.02E-09 | FALSE | 0.000576 | ++ | 0.382933 | 1 | 0.172676 | -1 | 8.19E-10 | 1 |
| 10-97710681-A-ACT | ACT | 0.000137 | 0.42726 | | | 0.4273 | -? | | | 0.001654 | 3.07E-10 | TRUE | 3.07E-10 | ?- | 0.006666 | 1 | 0.073084 | 1 | 2.76E-08 | 1 |
| 10-101802646-GCAA-( | G | 0.000117 | 0.603263 | | | 0.6033 | -? | | | 0.001477 | 3.19E-10 | TRUE | 3.19E-10 | ?+ | 0.000774 | 1 | 0.12008 | 1 | 1.64E-07 | 1 |
| 10-102054370-C-CG | CG | 8E-05 | 0.600753 | | | 0.6008 | -? | | | 0.001171 | 4.54E-13 | TRUE | 4.54E-13 | ?+ | 0.065357 | 1 | 0.085605 | 1 | 1.57E-11 | 1 |
| 10-102129838-CG-C | C | | | | | | | | | 0.000494 | 9.75E-11 | TRUE | 9.75E-11 | ?- | 0.104534 | 1 | 0.129169 | 1 | 8.95E-10 | 1 |
| 10-102174312-C-CT | CT | 0.00049 | 0.565577 | | | 0.5656 | -? | 0.000526 | 0.361198 | 0.0015 | 5.56E-10 | TRUE | 1.48E-08 | ++ | 0.884911 | 1 | 0.089708 | 1 | 2.73E-10 | 1 |
| 10-102206649-AAG-A | A | 0.003206 | 0.516137 | | | 0.5161 | -? | | | 0.005578 | 1.25E-11 | TRUE | 1.25E-11 | ?+ | 0.078674 | 1 | 0.2228 | 1 | 1.62E-10 | 1 |
| 10-102785427-T-TCTG | TCTGC | 0.002622 | 0.556236 | | | 0.5562 | +? | | | 0.000466 | 7.23E-09 | TRUE | 7.24E-09 | ?- | 0.008154 | 1 | 0.000406 | 1 | 2.36E-05 | 1 |
| 10-111403547-C-CTG( | CTGG | | | | | | | | | 0.00036 | 6.57E-09 | TRUE | 6.57E-09 | ?+ | | | 0.125368 | 1 | 2.41E-08 | 1 |
| 10-111586970-CA-C | C | | | | | | | | | 0.954303 | 4.33E-09 | TRUE | 4.33E-09 | ?- | 0.992332 | 1 | 0.102545 | 1 | 1.31E-11 | 1 |
| 10-114884738-C-T | T | 0.000413 | 0.094732 | | | 0.09473 | +? | | | 0.000372 | 2.54E-10 | TRUE | 2.54E-10 | ?+ | 0.081566 | 1 | 0.100228 | 1 | 2.49E-09 | 1 |
| 10-116619728-ATTTT- | A | | | | | | | | | 0.001653 | 2.3E-08 | TRUE | 2.3E-08 | ?+ | 0.019335 | 1 | 0.784836 | 1 | 7.05E-08 | 1 |
| 10-117460011-T-A | A | | | | | | | 0.000291 | 0.247876 | 0.000761 | 4.26E-10 | TRUE | 3.42E-09 | ++ | 0.205517 | 1 | 0.010594 | 1 | 7.28E-09 | 1 |
| 10-119182166-A-AC | AC | | | | | | | | | 0.974485 | 2.87E-09 | TRUE | 2.88E-09 | ?+ | 0.444848 | -1 | 0.189028 | -1 | 2.64E-09 | -1 |
| 10-119200769-T-C | C | 0.000585 | 0.654252 | | | 0.6543 | -? | 0.001165 | 0.075155 | 0.001708 | 4.07E-08 | TRUE | 1.61E-07 | -- | 0.033357 | 1 | 0.016145 | 1 | 1.88E-06 | 1 |
| 10-119200773-C-T | T | 7.8E-05 | 0.243891 | | | 0.2439 | -? | 0.000307 | 0.925359 | 0.00147 | 1.26E-14 | TRUE | 5.2E-12 | -+ | 0.000659 | 1 | 0.013095 | 1 | 1.16E-10 | 1 |
| 10-119200781-C-A | A | | | | | | | | | 0.002218 | 2.28E-19 | TRUE | 2.28E-19 | ?+ | 0.00035 | 1 | 0.037339 | 1 | 7.13E-16 | 1 |
| 10-119907249-T-TTG | TTG | | | | | | | | | 0.001136 | 1.7E-14 | TRUE | 1.7E-14 | ?- | 0.001919 | 1 | 0.036746 | 1 | 2.09E-12 | 1 |
| 10-119907251-ACG-A | A | | | | | | | | | 0.000847 | 1.78E-12 | TRUE | 1.78E-12 | ?+ | 0.001821 | 1 | 0.035905 | 1 | 1.93E-10 | 1 |
| 10-120602534-T-C | C | 0.000177 | 0.271026 | | | 0.271 | +? | | | 0.000241 | 2.79E-08 | TRUE | 2.79E-08 | ?- | 0.04671 | 1 | 0.000517 | 1 | 3.37E-05 | 1 |
| 10-124809427-GGT-G | G | 0.001132 | 0.651528 | | | 0.6515 | -? | | | 0.004561 | 8.73E-17 | TRUE | 8.73E-17 | ?+ | 0.0183 | 1 | 0.391017 | 1 | 5.54E-16 | 1 |
| 10-125834928-CG-C | C | | | | | | | 0.000279 | 0.762481 | 0.003145 | 7.79E-11 | TRUE | 1.58E-09 | +- | 0.001387 | 1 | 0.063396 | 1 | 3.68E-08 | 1 |
| 10-125924732-A-C | C | | | | | | | | | 0.112199 | 1.34E-12 | FALSE | 1.34E-12 | ?- | 0.002729 | 1 | 0.340103 | 1 | 1.71E-10 | 1 |
| 10-125924732-A-G | G | | | | | | | 0.97048 | 0.611097 | 0.871269 | 1.35E-11 | FALSE | 3.79E-07 | -+ | 0.006493 | -1 | 0.40585 | -1 | 9.69E-10 | -1 |
| 10-126892051-AC-A | A | | | | | | | | | 0.008194 | 3.12E-12 | TRUE | 3.12E-12 | ?+ | 0.000864 | 1 | 0.006714 | 1 | 1.66E-08 | 1 |
| 10-128016654-G-GA | GA | | | | | | | | | 0.000779 | 4.73E-09 | TRUE | 4.73E-09 | ?- | 0.266992 | 1 | 0.003254 | 1 | 2.12E-07 | 1 |
| 10-130301442-A-G | G | | | 0.463 | 0.718786 | 0.7188 | ?+ | | | 0.644687 | 1.78E-08 | FALSE | 1.78E-08 | ?- | 0.265049 | 1 | 0.396206 | 1 | 9.26E-07 | 1 |
| 10-131177065-C-CAG | CAG | | | | | | | | | 0.708073 | 3.54E-09 | FALSE | 3.54E-09 | ?+ | 0.976296 | -1 | 0.502976 | 1 | 2.1E-09 | 1 |
| 10-131177065-C-G | G | | | | | | | 3.45E-05 | 0.364593 | 0.264467 | 1.56E-09 | FALSE | 1.82E-09 | -+ | 0.900558 | -1 | 0.598668 | -1 | 2E-09 | -1 |
| 10-132155008-A-G | G | | | | | | | | | 0.04326 | 1.37E-12 | FALSE | 1.37E-12 | ?+ | 0.001473 | -1 | 0.007409 | -1 | 2.9E-08 | -1 |
| 10-132221588-T-C | C | | | | | | | 0.800574 | 0.03285 | 0.764707 | 2.08E-08 | FALSE | 3.89E-07 | -- | 0.364612 | -1 | 0.040518 | 1 | 9.84E-08 | 1 |
| 10-132221588-T-TGG( | TGGGTGAGTGTGAGGGCTTCAGATGGCCGTCCCCCCACACACAACTGATGTCGAC | | | | | | | | | 0.054905 | 8.94E-10 | FALSE | 8.94E-10 | ?+ | 0.784399 | -1 | 0.248014 | -1 | 9.32E-10 | -1 |
| 10-132388816-C-CG | CG | | | | | | | | | 0.911971 | 8.37E-17 | TRUE | 8.37E-17 | ?- | 0.243272 | -1 | 0.139667 | -1 | 5.4E-17 | -1 |
| 10-132611004-G-GT | GT | | | | | | | | | 0.00361 | 3.67E-11 | TRUE | 3.67E-11 | ?- | 0.216818 | 1 | 0.157282 | 1 | 6.97E-10 | 1 |
| 10-133763233-C-A | A | | | | | | | | | 0.951568 | 1.3E-24 | FALSE | 1.3E-24 | ?- | 0.006731 | -1 | 0.004364 | -1 | 2.84E-21 | -1 |
| 10-85897458-G-A | A | | | | | | | 1.88E-05 | 0.566481 | | | FALSE | 1.6E-305 | -? | | | | | | |
| 10-129697225-C-T | T | | | | | | | 2.19E-05 | 0.181654 | | | FALSE | 2.3E-305 | +? | | | | | | |
| 11-64341843-GGTTGT | G | 0.008849 | 6.72E-09 | | | 6.721E-09 | +? | | | | | FALSE | | | | | | | | |
| 11-48145557-TG-T | T | | | | | | | 1.25E-05 | 1E-52 | | | FALSE | 0.4337 | -? | | | | | | |
| 11-54823098-T-G | G | | | | | | | 1.25E-05 | 1E-13 | | | FALSE | 0.05307 | -? | | | | | | |
| 11-431597-T-G | G | | | | | | | | | 0.785924 | 2.95E-08 | FALSE | 2.95E-08 | ?+ | 0.080323 | -1 | 0.909023 | -1 | 1.04E-07 | -1 |
| 11-463371-G-GT | GT | 0.010863 | 0.014248 | | | 0.01425 | +? | | | 0.007153 | 1.13E-08 | TRUE | 1.13E-08 | ?- | 0.758654 | 1 | 0.298235 | -1 | 1.74E-10 | 1 |
| 11-652592-CTT-C | C | 0.000683 | 0.215739 | | | 0.2157 | +? | | | 0.001176 | 7.31E-09 | TRUE | 7.31E-09 | ?- | 0.093523 | 1 | 0.191366 | 1 | 2.08E-08 | 1 |
| 11-881238-G-A | A | 0.000353 | 0.104151 | | | 0.1042 | -? | | | 0.00104 | 5.35E-09 | TRUE | 5.35E-09 | ?+ | 0.082524 | 1 | 0.460491 | -1 | 4.1E-09 | 1 |
| 11-1018088-TG-T | T | | | | | | | | | 0.008667 | 6.35E-09 | TRUE | 6.35E-09 | ?+ | 0.795162 | 1 | 0.239911 | 1 | 3.25E-10 | 1 |
| 11-1018456-G-A | A | | | | | | | 0.000498 | 0.373363 | 0.003314 | 1.1E-10 | TRUE | 4.13E-10 | ++ | 0.933511 | -1 | 0.132439 | 1 | 1.34E-10 | 1 |
| 11-1095580-GAGTAC/ | G | | | | | | | | | 0.002583 | 3.54E-08 | TRUE | 3.54E-08 | ?+ | 0.020547 | 1 | 0.153933 | 1 | 1.62E-06 | 1 |
| 11-1166174-CCCTATG | C | | | | | | | | | 0.00208 | 7.27E-10 | TRUE | 7.27E-10 | ?- | 0.000766 | 1 | 0.356916 | 1 | 1.84E-07 | 1 |
| 11-1341421-AC-A | A | | | | | | | 0.000486 | 0.578834 | 0.001753 | 1.13E-08 | TRUE | 2.41E-07 | ++ | 0.42246 | 1 | 0.889142 | -1 | 5.73E-10 | 1 |
| 11-1355268-C-T | T | 0.000332 | 0.859411 | | | 0.8594 | +? | 0.003806 | 0.781415 | 0.002081 | 3.42E-15 | TRUE | 3.05E-05 | -+ | 0.071904 | 1 | 0.017262 | 1 | 2.41E-12 | 1 |

| | | | | | | | | | | | | | | | | | | | | |
|---|---|---|---|---|---|---|---|---|---|---|---|---|---|---|---|---|---|---|---|---|
| 11-1355280-A-AT | AT | | | | | | | 0.003875 | 0.882325 | 0.001371 | 7.68E-13 | TRUE | 0.00069 | +- | 0.124478 | 1 | 0.507865 | 1 | 7.44E-11 | 1 |
| 11-1355825-C-T | T | 0.000509 | 0.511793 | | | 0.5118 | +? | 0.000182 | 0.004295 | 0.001199 | 2.1E-08 | TRUE | 3.18E-10 | ++ | 0.013593 | 1 | 0.006556 | 1 | 2.95E-05 | 1 |
| 11-1355828-T-G | G | 0.000353 | 0.924129 | | | 0.9241 | +? | 0.000461 | 0.182731 | 0.001357 | 2.06E-11 | TRUE | 1.83E-10 | -- | 0.011683 | 1 | 0.02611 | 1 | 1.99E-09 | 1 |
| 11-1355844-A-T | T | 0.000627 | 0.743075 | | | 0.7431 | -? | 0.000498 | 0.373811 | 0.002239 | 3.72E-16 | TRUE | 2.13E-14 | -- | 0.163449 | 1 | 0.065995 | 1 | 3.19E-14 | 1 |
| 11-1622940-C-CAAA | CAAA | 0.000234 | 0.717823 | | | 0.7178 | +? | 0.001093 | 0.693221 | 0.001506 | 7.35E-09 | TRUE | 8.24E-06 | ++ | 0.014432 | 1 | 0.032524 | 1 | 3.15E-07 | 1 |
| 11-2124905-GA-G | G | 0.002426 | 0.18976 | | | 0.1898 | -? | | | 0.002921 | 1.75E-08 | TRUE | 1.75E-08 | ?+ | 0.539421 | 1 | 0.15203 | 1 | 6.38E-08 | 1 |
| 11-2139063-CG-C | C | | | | | | | | | 0.988237 | 3.32E-10 | TRUE | 3.32E-10 | ?+ | 0.247784 | -1 | 0.460837 | -1 | 2.6E-10 | -1 |
| 11-3855608-C-CG | CG | | | | | | | | | 0.793133 | 2.85E-25 | FALSE | 2.85E-25 | ?- | 0.841407 | -1 | 9.79E-05 | -1 | 5.91E-23 | -1 |
| 11-3855608-C-CGCG | CGCG | | | | | | | | | 0.150429 | 2.15E-30 | FALSE | 2.15E-30 | ?+ | 0.794492 | 1 | 0.000169 | 1 | 3.32E-28 | 1 |
| 11-4302959-G-C | C | | | | | | | | | 0.910122 | 2.76E-08 | FALSE | 2.76E-08 | ?- | 0.059385 | -1 | 0.484244 | 1 | 4.71E-10 | -1 |
| 11-4616358-G-T | T | | | | | | | 0.998709 | 0.664147 | 0.953357 | 3.22E-08 | FALSE | 3.98E-08 | -- | 0.099846 | -1 | 0.155908 | -1 | 1.24E-06 | -1 |
| 11-8984332-G-T | T | | | | | | | | | 0.00058 | 2.87E-10 | TRUE | 2.87E-10 | ?+ | 0.002278 | 1 | 0.990656 | 1 | 1.65E-08 | 1 |
| 11-9300701-A-C | C | 0.000372 | 0.306484 | | | 0.3065 | +? | 0.002199 | 0.531004 | 0.001732 | 6.91E-21 | TRUE | 3.97E-11 | -- | 0.409383 | 1 | 0.017183 | 1 | 1.35E-20 | 1 |
| 11-9300706-G-A | A | 0.000606 | 0.687412 | | | 0.6874 | +? | 0.002146 | 0.281046 | 0.001264 | 9.43E-18 | TRUE | 1.61E-09 | ++ | 0.062653 | 1 | 0.0007 | 1 | 1.37E-13 | 1 |
| 11-9384165-T-C | C | | | | | | | | | 0.000527 | 1.81E-11 | TRUE | 1.81E-11 | ?- | 0.00037 | 1 | 0.21775 | 1 | 1.02E-09 | 1 |
| 11-9384187-C-G | G | 0.000117 | 0.385081 | | | 0.3851 | -? | 0.000119 | 0.835013 | 0.001581 | 3.15E-20 | TRUE | 1.45E-18 | +- | 8.09E-06 | 1 | 0.047926 | 1 | 3.42E-16 | 1 |
| 11-9551023-C-CTT | CTT | | | | | | | | | 0.004232 | 2.62E-24 | TRUE | 2.63E-24 | ?+ | 2.12E-07 | 1 | 0.020917 | 1 | 4.61E-18 | 1 |
| 11-12757946-A-ATTTT | ATTTTG | | | | | | | | | 0.000985 | 6.95E-09 | TRUE | 6.95E-09 | ?- | 0.073695 | 1 | 0.743949 | 1 | 1.14E-07 | 1 |
| 11-14930887-G-A | A | 0.000295 | 0.152178 | | | 0.1522 | -? | 0.00292 | 0.714559 | 0.001533 | 1.33E-10 | TRUE | 0.000159 | ++ | 0.794618 | 1 | 0.090954 | 1 | 3.11E-09 | 1 |
| 11-18328184-CAG-C | C | | | | | | | | | 0.001637 | 6.89E-09 | TRUE | 6.89E-09 | ?- | 0.267122 | 1 | 0.112648 | 1 | 6.49E-09 | 1 |
| 11-19260622-C-CCA | CCA | | | | | | | 0.999875 | 0.420952 | 0.995944 | 9.92E-11 | TRUE | 1.3E-10 | ++ | 0.82917 | 1 | 0.438143 | 1 | 9.93E-11 | 1 |
| 11-30791372-G-GTA | GTA | | | | | | | | | 0.000694 | 1.14E-09 | TRUE | 1.14E-09 | ?- | 0.002178 | 1 | 0.646248 | 1 | 3.77E-07 | 1 |
| 11-30887986-AGAGC- | A | | | | | | | | | 0.000414 | 1.46E-11 | TRUE | 1.46E-11 | ?+ | 0.000155 | 1 | 0.225535 | 1 | 1.51E-07 | 1 |
| 11-31533623-C-CAG | CAG | | | | | | | 6.26E-05 | 0.427123 | 0.000913 | 8.07E-10 | TRUE | 7.77E-10 | ++ | 0.000118 | 1 | 0.025657 | 1 | 1.82E-07 | 1 |
| 11-31971510-T-C | C | | | | | | | 0.000166 | 0.071264 | 0.002196 | 4.79E-13 | TRUE | 9.75E-14 | -- | 0.061805 | 1 | 0.016103 | 1 | 1.92E-10 | 1 |
| 11-33250072-TA-T | T | | | | | | | | | 0.000325 | 3.67E-10 | TRUE | 3.67E-10 | ?+ | 0.511687 | 1 | 0.01921 | 1 | 1.6E-10 | 1 |
| 11-37654820-G-T | T | | | | | | | 0.000999 | 0.600947 | 0.000934 | 2.48E-08 | TRUE | 3.59E-05 | ++ | 0.779844 | 1 | 0.011221 | 1 | 8.08E-08 | 1 |
| 11-46355253-C-CATG | CATG | | | | | | | | | 0.000252 | 7.84E-10 | TRUE | 7.84E-10 | ?+ | 0.000156 | 1 | 0.000762 | 1 | 0.000117 | 1 |
| 11-46830926-A-G | G | | | | | | | 9.4E-06 | 0.415225 | 0.000408 | 8.67E-12 | TRUE | 3.17E-11 | +- | 0.002381 | 1 | 0.102297 | 1 | 2.7E-10 | 1 |
| 11-47085719-G-A | A | | | | | | | 0.000291 | 0.657503 | 0.000802 | 2.39E-10 | TRUE | 2.28E-08 | ++ | 0.173236 | 1 | 0.584775 | 1 | 4.19E-11 | 1 |
| 11-47563973-C-G | G | 0.000593 | 0.677032 | | | 0.677 | +? | 0.001554 | 0.735987 | 0.000432 | 4.24E-09 | TRUE | 0.01103 | +- | 0.001535 | 1 | 0.000968 | 1 | 2.95E-05 | 1 |
| 11-47674901-A-AT | AT | | | | | | | | | 0.000315 | 5.14E-09 | TRUE | 5.14E-09 | ?- | 0.037196 | 1 | 0.000364 | 1 | 5.46E-07 | 1 |
| 11-47677751-C-CTG | CTG | | | | | | | | | 0.001091 | 1.29E-10 | TRUE | 1.29E-10 | ?+ | 0.013399 | 1 | 0.016242 | 1 | 2.2E-07 | 1 |
| 11-48727069-T-TTTGT | TTTGTC | | | | | | | | | 0.005805 | 2.79E-15 | TRUE | 2.79E-15 | ?- | 0.003509 | 1 | 0.002324 | 1 | 3.03E-11 | 1 |
| 11-48727087-A-ATG | ATG | | | | | | | | | 0.006055 | 1.82E-16 | TRUE | 1.82E-16 | ?- | 0.018782 | 1 | 0.001006 | 1 | 9.22E-13 | 1 |
| 11-48768777-A-T | T | | | | | | | | | 0.000257 | 3.27E-10 | TRUE | 3.28E-10 | ?- | 0.407702 | 1 | 0.035408 | 1 | 3.83E-10 | 1 |
| 11-48843498-T-C | C | | | | | | | | | 0.000587 | 3.16E-08 | TRUE | 3.16E-08 | ?- | 0.016489 | 1 | 0.011657 | 1 | 2.84E-05 | 1 |
| 11-48911550-C-T | T | | | | | | | | | 0.001515 | 1.31E-12 | TRUE | 1.31E-12 | ?+ | 0.001033 | 1 | 0.000247 | 1 | 1.25E-08 | 1 |
| 11-48911855-T-A | A | | | | | | | 0.989193 | 0.980661 | 0.980838 | 4.29E-14 | TRUE | 1.38E-06 | +- | 0.628122 | -1 | 0.112491 | -1 | 1.52E-19 | -1 |
| 11-50576986-A-C | C | | | | | | | 3.76E-05 | 0.227177 | 0.000189 | 5.7E-10 | TRUE | 4.68E-08 | +- | 0.157957 | 1 | | | 1.22E-11 | 1 |
| 11-50716519-C-T | T | | | | | | | | | 0.000212 | 1.64E-08 | TRUE | 1.64E-08 | ?+ | 0.003903 | 1 | | | 3.49E-05 | 1 |
| 11-54897486-T-TGCCA | TGCCA | | | | | | | | | 0.003205 | 4.65E-09 | TRUE | 4.65E-09 | ?+ | 0.327914 | -1 | 0.239327 | -1 | 8.46E-09 | -1 |
| 11-55233749-T-C | C | | | | | | | | | 0.005955 | 2.69E-68 | TRUE | 2.7E-68 | ?- | 3.66E-09 | 1 | 4.62E-11 | 1 | 4.87E-49 | 1 |
| 11-55233796-A-C | C | | | | | | | | | 0.002106 | 1.88E-23 | TRUE | 1.89E-23 | ?- | 0.000379 | 1 | 0.000224 | 1 | 6.99E-17 | 1 |
| 11-55251455-A-G | G | 0.051503 | 0.196994 | 0.076 | 0.419003 | 0.1854 | ++ | 0.061874 | 0.627801 | 0.060364 | 1.49E-09 | TRUE | 0.3606 | +- | 0.065076 | 1 | 0.884777 | 1 | 0.076653 | 1 |
| 11-55461884-C-A | A | | | | | | | 0.017618 | 0.709282 | 0.019658 | 7.51E-10 | TRUE | 0.003299 | -+ | 0.205391 | 1 | 0.976517 | -1 | 2.12E-07 | 1 |
| 11-57768000-A-AT | AT | | | | | | | 0.002255 | 0.64311 | 0.002614 | 9.68E-21 | TRUE | 3.37E-12 | -- | 0.003368 | 1 | 0.005532 | 1 | 5.81E-19 | 1 |
| 11-61824974-C-CCT | CCT | 0.002463 | 0.8907 | | | 0.8907 | -? | | | 0.004461 | 7.07E-19 | TRUE | 7.07E-19 | ?+ | 0.110714 | 1 | 0.031935 | 1 | 2.24E-16 | 1 |
| 11-61894409-T-G | G | | | | | | | 0.000783 | 0.750836 | 0.000538 | 5.78E-13 | TRUE | 5.15E-06 | +- | 0.011843 | 1 | 0.005796 | 1 | 1.41E-10 | 1 |
| 11-61894450-T-C | C | | | | | | | 0.000119 | 0.732099 | 0.000683 | 2.2E-08 | TRUE | 4.89E-07 | +- | 0.259318 | 1 | 0.001507 | 1 | 9.9E-07 | 1 |
| 11-62004372-C-CAG | CAG | | | | | | | 5.32E-05 | 0.951223 | 0.001059 | 2.18E-08 | TRUE | 4.91E-08 | -+ | 0.727299 | -1 | 0.000795 | 1 | 5.01E-07 | 1 |
| 11-62095024-C-CAAA | CAAAA | | | | | | | | | 0.029977 | 1.55E-08 | FALSE | 1.55E-08 | ?+ | 0.816744 | 1 | 0.126058 | 1 | 2.47E-08 | 1 |
| 11-62211652-G-A | A | 0.00188 | 0.688751 | | | 0.6888 | -? | 0.00067 | 0.225421 | 0.004034 | 8.07E-15 | TRUE | 3.73E-10 | -+ | 0.003959 | 1 | 0.210891 | 1 | 1.08E-12 | 1 |
| 11-62523038-A-C | C | 0.000216 | 0.07122 | | | 0.07122 | +? | 0.000235 | 0.422813 | 0.000959 | 6.65E-10 | TRUE | 5.67E-09 | -- | 0.75208 | 1 | 0.00399 | 1 | 1.25E-08 | 1 |
| 11-62537111-GCACCA | G | | | | | | | 0.000141 | 0.592255 | 0.001406 | 3.52E-13 | TRUE | 1.72E-12 | ++ | 2.69E-05 | 1 | 0.000405 | 1 | 2.73E-08 | 1 |
| 11-62750690-G-A | A | 0.000156 | 0.309595 | | | 0.3096 | +? | 0.00026 | 0.035075 | 0.000697 | 3.72E-08 | TRUE | 0.000154 | -+ | 0.010465 | 1 | 0.63871 | -1 | 1.65E-07 | 1 |
| 11-62845315-ATT-A | A | | | | | | | | | 0.00045 | 4.29E-08 | TRUE | 4.29E-08 | ?+ | 0.011895 | 1 | 0.408638 | 1 | 1.06E-07 | 1 |
| 11-62869905-CTTGA-( | C | 0.000548 | 0.050528 | | | 0.05053 | +? | | | 0.002289 | 3.37E-17 | TRUE | 3.37E-17 | ?- | 0.362569 | 1 | 0.012169 | 1 | 5.75E-15 | 1 |
| 11-63684993-CAA-C | C | | | | | | | | | 0.000637 | 1.41E-09 | TRUE | 1.41E-09 | ?- | 0.000327 | 1 | 0.580162 | 1 | 2.74E-07 | 1 |

| | | | | | | | | | | | | | | | | | | | | |
|---|---|---|---|---|---|---|---|---|---|---|---|---|---|---|---|---|---|---|---|---|
| 11-63764272-T-C | C | | | | | | | 4.07E-05 | 0.034778 | 0.000674 | 9.13E-09 | TRUE | 9.53E-10 | -- | 0.01175 | 1 | 0.360657 | 1 | 7.6E-07 | 1 |
| 11-63860686-G-A | A | 0.000156 | 0.19946 | | | 0.1995 | -? | 5.01E-05 | 0.739983 | 0.000374 | 7.78E-09 | TRUE | 2.72E-08 | ++ | 0.656701 | 1 | 0.703084 | 1 | 6.97E-09 | 1 |
| 11-64035713-G-GCCC | GCCCCCGCCACCTCCTCCCCGCCCGCTGCTCCCCCTCC | | | | | | | | | 0.952794 | 2.31E-10 | FALSE | 2.31E-10 | ?+ | 0.409649 | 1 | 0.369911 | -1 | 2.41E-15 | -1 |
| 11-64280476-C-A | A | | | | | | | | | 0.001237 | 9.45E-15 | TRUE | 9.45E-15 | ?+ | 0.05602 | 1 | 0.004367 | 1 | 4.94E-13 | 1 |
| 11-64662034-CTT-C | C | 0.000196 | 0.112831 | | | 0.1128 | +? | | | 0.001633 | 6.99E-12 | TRUE | 6.99E-12 | ?- | 0.015463 | 1 | 0.003281 | 1 | 6.45E-09 | 1 |
| 11-64664247-CAAAT-( | C | | | | | | | | | 0.000262 | 9.08E-11 | TRUE | 9.08E-11 | ?- | 0.002068 | 1 | 0.472527 | -1 | 1.17E-09 | 1 |
| 11-65463200-TAG-T | T | | | | | | | | | 0.000365 | 7.59E-09 | TRUE | 7.59E-09 | ?+ | 0.054511 | 1 | 0.68409 | 1 | 4.44E-09 | 1 |
| 11-65527549-AC-A | A | | | | | | | | | 0.000193 | 1.98E-08 | TRUE | 1.98E-08 | ?+ | | | | | 4.37E-06 | 1 |
| 11-66402115-A-T | T | | | | | | | 0.000128 | 0.233866 | 0.000731 | 3.73E-10 | TRUE | 1.24E-07 | +- | 0.106296 | 1 | 0.772912 | 1 | 8.15E-10 | 1 |
| 11-67134652-GCGT-G | G | | | | | | | | | 0.000481 | 3.02E-09 | TRUE | 3.02E-09 | ?+ | 0.667936 | 1 | 0.441868 | 1 | 7.29E-09 | 1 |
| 11-67157185-T-C | C | | | | | | | 0.001438 | 0.081903 | 0.001497 | 1.62E-13 | TRUE | 2.49E-10 | -- | 0.000519 | 1 | 0.41744 | 1 | 1.37E-09 | 1 |
| 11-67988509-A-ATG | ATG | | | | | | | | | 0.001078 | 1E-08 | TRUE | 1E-08 | ?- | 6.35E-05 | 1 | 0.245942 | 1 | 1.73E-06 | 1 |
| 11-68207031-AGG-A | A | | | | | | | | | 0.001525 | 2.14E-09 | TRUE | 2.14E-09 | ?+ | 0.010894 | 1 | 0.232971 | 1 | 1.85E-08 | 1 |
| 11-68301868-T-TCCG( | TCCGC | 9.89E-05 | 0.482833 | | | 0.4828 | -? | | | 0.217727 | 5.2E-17 | FALSE | 5.21E-17 | ?+ | 0.150964 | -1 | 0.138902 | -1 | 2.78E-15 | -1 |
| 11-68301874-T-C | C | | | | | | | | | 0.592281 | 7.32E-15 | FALSE | 7.32E-15 | ?- | 0.224297 | 1 | 0.128001 | 1 | 6.19E-13 | 1 |
| 11-68301874-T-TTGG( | TTGGCCTCCCAAAGTGCTGGGATTACAGGCGTGAGTCACCGCGCGCGGCCATTTTTTGTATTTTTAGTAGAGATGC | | | | | | | | | 0.218808 | 6.13E-18 | FALSE | 6.13E-18 | ?+ | 0.046298 | -1 | 0.08665 | -1 | 3.48E-15 | -1 |
| 11-68453603-T-C | C | 0.380124 | 0.003202 | 0.369 | 0.387402 | 0.002651 | ++ | 0.348432 | 0.738577 | 0.417718 | 1.9E-08 | FALSE | 0.003519 | +- | 0.372625 | 1 | 0.021856 | 1 | 7.81E-05 | 1 |
| 11-69082539-A-T | T | 0.000647 | 0.707608 | | | 0.7076 | -? | 0.0001 | 0.605345 | 0.00021 | 3.07E-08 | TRUE | 4.82E-06 | +- | 0.023653 | 1 | 0.503397 | -1 | 1.08E-06 | 1 |
| 11-69577301-C-G | G | | | | | | | | | 0.000653 | 3.44E-15 | TRUE | 3.44E-15 | ?- | 0.003014 | 1 | 0.000584 | 1 | 1.13E-11 | 1 |
| 11-69720156-A-ACT | ACT | | | | | | | | | 0.000807 | 5.89E-10 | TRUE | 5.89E-10 | ?- | 0.042171 | 1 | 0.02216 | 1 | 1.98E-08 | 1 |
| 11-69723576-G-C | C | | | | | | | 4.7E-05 | 0.314411 | 0.000867 | 3.99E-09 | TRUE | 2.46E-09 | ++ | 0.122471 | 1 | 0.018702 | 1 | 3.22E-07 | 1 |
| 11-70222751-T-TA | TA | | | | | | | 0.003852 | 0.154994 | 0.001977 | 1.07E-14 | TRUE | 5.32E-08 | -- | 0.221326 | 1 | 0.006819 | 1 | 2.2E-12 | 1 |
| 11-70803334-CTA-C | C | | | | | | | | | 0.007188 | 4.62E-19 | TRUE | 4.62E-19 | ?- | 0.003922 | 1 | 0.017446 | 1 | 5.53E-16 | 1 |
| 11-70803346-C-CCGTT | CCGTT | | | | | | | | | 0.007161 | 4.35E-17 | TRUE | 4.35E-17 | ?+ | 0.00386 | 1 | 0.049404 | 1 | 2.5E-14 | 1 |
| 11-70890949-GT-G | G | | | | | | | | | 0.003312 | 1.46E-30 | TRUE | 1.46E-30 | ?+ | 0.029225 | 1 | 0.002711 | 1 | 4.22E-27 | 1 |
| 11-70890952-AT-A | A | | | | | | | | | 0.001895 | 2.51E-18 | TRUE | 2.51E-18 | ?+ | 0.012147 | 1 | 0.00986 | 1 | 9.6E-15 | 1 |
| 11-71920538-CAG-C | C | 0.003564 | 0.416454 | | | 0.4165 | +? | 0.000153 | 0.311751 | 0.004968 | 1.46E-17 | TRUE | 1.15E-17 | -- | 0.00022 | 1 | 0.973983 | 1 | 1.97E-15 | 1 |
| 11-71936008-C-T | T | 0.000137 | 0.998569 | | | 0.9986 | -? | 5.32E-05 | 0.689892 | 0.000317 | 2.86E-08 | TRUE | 5.62E-07 | -+ | 0.002572 | 1 | 0.002683 | 1 | 1.51E-05 | 1 |
| 11-72063209-CCT-C | C | 0.000469 | 0.800188 | | | 0.8002 | +? | | | 0.001556 | 1.21E-14 | TRUE | 1.21E-14 | ?- | 0.013819 | 1 | 0.573881 | 1 | 2.72E-13 | 1 |
| 11-73692269-CCGAG( | C | 0.000683 | 0.695366 | | | 0.6954 | -? | | | 0.002776 | 1.01E-12 | TRUE | 1.01E-12 | ?- | 0.367765 | 1 | 0.009141 | 1 | 3.83E-11 | 1 |
| 11-73813823-C-CCT | CCT | | | | | | | | | 0.000674 | 3.25E-11 | TRUE | 3.25E-11 | ?+ | 2.96E-06 | 1 | 0.00574 | 1 | 6.58E-06 | 1 |
| 11-74066348-TACATA | T | 0.002499 | 0.325389 | | | 0.3254 | -? | 0.000796 | 0.380788 | 0.002824 | 1.19E-08 | TRUE | 1.49E-05 | -+ | 0.129907 | 1 | 0.265692 | 1 | 3E-08 | 1 |
| 11-75748015-G-A | A | | | | | | | 0.000821 | 0.944521 | 0.001434 | 1.22E-12 | TRUE | 5.87E-08 | -+ | 0.01152 | 1 | 0.012488 | 1 | 5.18E-10 | 1 |
| 11-75748022-G-A | A | | | | | | | 0.000573 | 0.370418 | 0.001056 | 1.26E-13 | TRUE | 9.03E-08 | -+ | 0.009495 | 1 | 0.014435 | 1 | 6.32E-12 | 1 |
| 11-75748028-T-G | G | | | | | | | | | 0.001098 | 1.15E-09 | TRUE | 1.16E-09 | ?- | 0.075074 | 1 | 0.014734 | 1 | 4.04E-08 | 1 |
| 11-76255841-GGGT-G | G | 0.000274 | 0.455189 | | | 0.4552 | +? | | | 0.000946 | 8.9E-11 | TRUE | 8.9E-11 | ?+ | 0.340078 | 1 | 0.115117 | 1 | 1.63E-10 | 1 |
| 11-76255847-CA-C | C | | | | | | | | | 0.000583 | 6.75E-14 | TRUE | 6.75E-14 | ?- | 0.244235 | 1 | 0.042279 | 1 | 8.5E-13 | 1 |
| 11-77546029-CCT-C | C | | | | | | | | | 0.000392 | 8.13E-09 | TRUE | 8.13E-09 | ?- | 0.52597 | -1 | 0.000403 | 1 | 8.28E-07 | 1 |
| 11-91664514-A-G | G | | | | | | | 0.000774 | 0.484709 | 0.001016 | 3.44E-17 | TRUE | 2.41E-09 | +- | 0.010623 | 1 | 0.211666 | -1 | 8.16E-18 | 1 |
| 11-103459091-C-T | T | 7.81E-05 | 0.451986 | | | 0.452 | +? | 0.00057 | 0.958299 | 0.000844 | 7.6E-10 | TRUE | 2.21E-06 | ++ | 0.291124 | 1 | 0.007459 | 1 | 3.15E-07 | 1 |
| 11-103459116-C-T | T | 0.000117 | 0.997133 | | | 0.9971 | -? | 0.000482 | 0.881363 | 0.001003 | 9.16E-09 | TRUE | 2.7E-06 | ++ | 0.285073 | 1 | 0.007648 | 1 | 6.8E-06 | 1 |
| 11-103459123-A-C | C | 0.000176 | 0.532374 | | | 0.5324 | +? | 0.000414 | 0.684033 | 0.00186 | 5.6E-09 | TRUE | 8.18E-07 | +- | 0.071422 | 1 | 0.069627 | 1 | 9.82E-07 | 1 |
| 11-105515895-G-T | T | | | | | | | 8.15E-05 | 0.479102 | 0.002863 | 2.34E-18 | TRUE | 2.64E-18 | ++ | 0.011547 | 1 | 0.036997 | 1 | 2.3E-15 | 1 |
| 11-105515902-G-A | A | 0.000587 | 0.188546 | | | 0.1885 | -? | 7.52E-05 | 0.691041 | 0.001714 | 1.38E-17 | TRUE | 3.28E-17 | ++ | 0.020359 | 1 | 0.307397 | 1 | 2.93E-16 | 1 |
| 11-108396326-CAA-C | C | | | | | | | 0.000204 | 0.872063 | 0.000883 | 3.71E-08 | TRUE | 1.31E-06 | +- | 0.204775 | 1 | 0.234457 | 1 | 2.19E-07 | 1 |
| 11-111927797-A-AC | AC | 0.000351 | 0.293491 | | | 0.2935 | +? | 0.002813 | 0.005453 | 0.002122 | 1.61E-19 | TRUE | 0.000122 | +- | 3.54E-05 | 1 | 0.483179 | 1 | 5.54E-17 | 1 |
| 11-113771108-T-TG | TG | | | | | | | | | 0.000922 | 4.83E-15 | TRUE | 4.83E-15 | ?- | 0.000126 | 1 | 0.876028 | -1 | 8.21E-13 | 1 |
| 11-117105931-CA-C | C | | | | | | | | | 0.000215 | 3.63E-08 | TRUE | 3.63E-08 | ?- | 0.012682 | 1 | | | 8.37E-08 | 1 |
| 11-118709508-G-A | A | 0.000435 | 0.178665 | | | 0.1787 | -? | 0.002033 | 0.527094 | 0.001235 | 3.93E-08 | TRUE | 0.00899 | -+ | 0.159179 | 1 | 0.102822 | 1 | 1.12E-06 | 1 |
| 11-118736461-C-CAT | CAT | | | | | | | | | 0.002503 | 2.53E-08 | TRUE | 2.53E-08 | ?+ | 0.001109 | 1 | 0.385778 | -1 | 2.4E-07 | 1 |
| 11-121859552-C-CAG, | CAGAG | | | | | | | | | 0.854937 | 2.54E-18 | FALSE | 2.55E-18 | ?+ | 0.722362 | 1 | 0.054859 | 1 | 1.01E-17 | 1 |
| 11-123615487-G-C | C | | | | | | | 7.52E-05 | 0.781646 | 0.000608 | 3.21E-09 | TRUE | 1.3E-08 | ++ | 0.042836 | 1 | 0.677773 | -1 | 5.78E-09 | 1 |
| 11-129833604-G-A | A | | | | | | | 0.000404 | 0.094626 | 0.000962 | 1.03E-10 | TRUE | 2.66E-10 | ++ | 0.180136 | 1 | 0.115888 | 1 | 1.22E-10 | 1 |
| 11-134286572-G-C | C | 0.007344 | 0.129484 | | | 0.1295 | -? | 0.010064 | 0.140239 | 0.004271 | 3.78E-18 | TRUE | 2.6E-08 | ++ | 0.553956 | 1 | 0.031632 | 1 | 4.49E-18 | 1 |
| 11-128448500-G-A | A | | | | | | | 1.88E-05 | 0.062599 | | | FALSE | 3.7E-291 | -? | | | | | | |
| 11-48165580-G-T | T | | | | | | | 1.88E-05 | 0.671075 | | | FALSE | 2.6E-305 | -? | | | | | | |
| 11-48870192-G-T | T | | | | | | | 0.000323 | 0.000117 | 0.000258 | 6.49E-06 | TRUE | 6.08E-09 | ++ | 0.618106 | -1 | 0.450388 | 1 | 8.94E-06 | 1 |
| 11-3777426-G-T | T | | | | | | | 0.000128 | 0.068039 | 0.000399 | 5.77E-08 | TRUE | 1.61E-08 | ++ | 0.005769 | 1 | 0.006689 | 1 | 9.76E-06 | 1 |
| 12-48371822-T-A | A | 0.093977 | 4.25E-08 | 0.085 | 0.448226 | 3.572E-08 | ++ | 0.098424 | 0.8941 | 0.103403 | 0.364093 | TRUE | 0.6859 | +- | 0.760915 | 1 | 0.19214 | -1 | 0.522332 | -1 |

| | | | | | | | | | | | | | | | | | | | | |
|---|---|---|---|---|---|---|---|---|---|---|---|---|---|---|---|---|---|---|---|---|
| 12-102888270-C-T | T | 0.000195 | 0.24299 | | | 0.243 | +? | 0.000204 | 1E-83 | 0.000161 | 0.07485 | TRUE | 0.1039 | -- | | | | | 0.084703 | -1 |
| 12-127699204-G-T | T | | | | | | | 2.19E-05 | 1E-97 | | | FALSE | 0.214 | -? | | | | | | |
| 12-44644608-G-GTAT, | GTATA | | | | | | | 0.000589 | 1.63E-09 | | | FALSE | 1.63E-09 | +? | | | | | | |
| 12-77809582-C-T | T | | | | | | | 9.4E-06 | 1.64E-11 | | | FALSE | 0.06887 | +? | | | | | | |
| 12-453108-C-T | T | | | | | | | | | 0.952038 | 1.61E-11 | FALSE | 1.61E-11 | ?- | 0.054061 | -1 | 0.980127 | 1 | 4.08E-13 | -1 |
| 12-762205-A-AGG | AGG | | | | | | | | | 0.000412 | 1.41E-08 | TRUE | 1.41E-08 | ?- | 0.001895 | 1 | 0.03342 | 1 | 2.06E-05 | 1 |
| 12-762206-CAT-C | C | | | | | | | | | 0.000367 | 4.42E-09 | TRUE | 4.42E-09 | ?- | 0.001515 | 1 | 0.165782 | 1 | 5.1E-06 | 1 |
| 12-2946592-C-CTT | CTT | | | | | | | | | 0.00336 | 8.43E-16 | TRUE | 8.43E-16 | ?+ | 0.053041 | 1 | 0.079934 | 1 | 6.23E-14 | 1 |
| 12-6291548-GGTGTG1 | G | | | | | | | | | 0.071797 | 2.91E-08 | FALSE | 2.91E-08 | ?+ | 0.737081 | 1 | 0.821064 | 1 | 1.7E-10 | 1 |
| 12-7660652-AAG-A | A | 0.001202 | 0.80009 | | | 0.8001 | -? | | | 0.001002 | 3.09E-11 | TRUE | 3.09E-11 | ?+ | 0.000584 | 1 | 0.175829 | 1 | 1.83E-08 | 1 |
| 12-7734982-CTT-C | C | 0.000176 | 0.182532 | | | 0.1825 | +? | | | 0.004411 | 2.15E-21 | TRUE | 2.15E-21 | ?- | 0.13607 | 1 | 0.011309 | 1 | 7.7E-20 | 1 |
| 12-8009913-A-G | G | | | | | | | | | 0.001178 | 8.29E-11 | TRUE | 8.29E-11 | ?- | 0.043187 | 1 | 0.306074 | 1 | 3.63E-09 | 1 |
| 12-8395398-C-CGTGG | CGTGG | | | | | | | | | 0.04281 | 4.36E-10 | FALSE | 4.36E-10 | ?+ | 0.341284 | 1 | 0.054716 | 1 | 4.68E-08 | 1 |
| 12-8625760-T-C | C | | | | | | | | | 0.865143 | 8.02E-14 | FALSE | 8.02E-14 | ?+ | 0.376896 | -1 | 0.655769 | -1 | 6.66E-13 | -1 |
| 12-9418997-ACC-A | A | | | | | | | | | 0.031131 | 1.29E-08 | FALSE | 1.29E-08 | ?+ | 0.848096 | -1 | 0.335448 | 1 | 3.73E-09 | 1 |
| 12-11073944-CCCT-C | C | 0.414394 | 0.970755 | | | 0.9708 | -? | | | 0.319434 | 3.47E-09 | FALSE | 3.47E-09 | ?+ | 0.681406 | -1 | 0.010018 | -1 | 9.67E-08 | -1 |
| 12-11567230-GGT-G | G | 0.000234 | 0.114995 | | | 0.115 | -? | | | 0.002568 | 7E-14 | TRUE | 7E-14 | ?+ | 0.000112 | 1 | 0.529566 | 1 | 6.13E-11 | 1 |
| 12-12363724-A-G | G | | | | | | | | | 0.000572 | 8.16E-09 | TRUE | 8.16E-09 | ?- | 0.000785 | 1 | 0.385298 | 1 | 5.36E-07 | 1 |
| 12-12553080-C-CTCT | CTCT | | | | | | | | | 0.000157 | 2.14E-08 | TRUE | 2.14E-08 | ?+ | | | | | 9.69E-06 | 1 |
| 12-18232057-A-G | G | | | | | | | | | 0.000355 | 4.46E-08 | TRUE | 4.46E-08 | ?- | 0.416269 | 1 | 0.605507 | 1 | 9.65E-09 | 1 |
| 12-20353357-C-T | T | 0.002433 | 0.25784 | | | 0.2578 | -? | 0.002998 | 0.194077 | 0.001213 | 4.22E-08 | TRUE | 0.000149 | ++ | 0.665577 | 1 | 0.033414 | 1 | 2.37E-07 | 1 |
| 12-20353414-G-GGAA | GGAACCC | 0.000273 | 0.361875 | | | 0.3619 | +? | | | 0.004498 | 4.49E-15 | TRUE | 4.49E-15 | ?- | 0.076097 | 1 | 0.074094 | 1 | 3.01E-14 | 1 |
| 12-21662734-AC-A | A | | | | | | | | | 0.010586 | 2.49E-09 | FALSE | 2.49E-09 | ?+ | 0.734286 | 1 | 0.286053 | 1 | 1.84E-09 | 1 |
| 12-26332904-T-TGG | TGG | | | | | | | | | 0.007456 | 2.08E-08 | TRUE | 2.08E-08 | ?+ | 0.115326 | -1 | 0.721883 | -1 | 4.67E-08 | -1 |
| 12-27784354-C-G | G | | | | | | | 0.000197 | 0.909335 | 0.00061 | 4.34E-11 | TRUE | 1.25E-08 | +- | 0.005342 | 1 | 0.040287 | 1 | 5.93E-09 | 1 |
| 12-31366840-A-ACC | ACC | | | | | | | | | 0.001796 | 3.8E-13 | TRUE | 3.8E-13 | ?- | 0.283225 | 1 | 0.002559 | 1 | 6.29E-12 | 1 |
| 12-31549775-A-ACC | ACC | | | | | | | | | 0.00024 | 6.27E-09 | TRUE | 6.27E-09 | ?- | 0.475754 | -1 | 0.096843 | 1 | 1.53E-09 | 1 |
| 12-32523802-G-A | A | 0.001233 | 0.171963 | | | 0.172 | +? | 0.001886 | 0.562503 | 0.002993 | 2.28E-08 | TRUE | 0.000604 | -+ | 0.234668 | 1 | 0.634027 | 1 | 1.75E-08 | 1 |
| 12-32797701-G-A | A | 0.000196 | 0.370333 | | | 0.3703 | -? | | | 0.00082 | 1.25E-08 | TRUE | 1.25E-08 | ?+ | 0.048987 | 1 | 9.95E-05 | 1 | 3.17E-05 | 1 |
| 12-32797705-G-A | A | 0.000196 | 0.163407 | | | 0.1634 | -? | 6.58E-05 | 0.875687 | 0.000763 | 1.5E-09 | TRUE | 9.43E-09 | -+ | 0.100229 | 1 | 0.000486 | 1 | 5.82E-06 | 1 |
| 12-34297017-A-G | G | | | | | | | | | 0.204898 | 3.04E-09 | FALSE | 3.04E-09 | ?- | 0.449438 | 1 | 0.200336 | -1 | 0.035242 | 1 |
| 12-34299189-T-TG | TG | | | | | | | 0.414908 | 0.842796 | 0.494658 | 3.49E-08 | FALSE | 0.001172 | -- | 0.482764 | 1 | 0.710963 | -1 | 2.82E-06 | 1 |
| 12-34332723-A-G | G | 0.184816 | 0.562972 | | | 0.563 | -? | | | 0.195747 | 7.18E-11 | FALSE | 7.18E-11 | ?- | 0.435443 | 1 | 0.372942 | -1 | 0.032793 | 1 |
| 12-34363209-T-C | C | | | | | | | | | 0.207033 | 1.83E-21 | FALSE | 1.83E-21 | ?- | 0.321002 | 1 | 0.43588 | -1 | 0.006189 | 1 |
| 12-34395679-T-C | C | 0.188531 | 0.639784 | | | 0.6398 | -? | | | 0.202185 | 9.08E-11 | FALSE | 9.08E-11 | ?- | 0.403635 | 1 | 0.338756 | -1 | 0.02701 | 1 |
| 12-37270198-G-T | T | 9.81E-05 | 0.36029 | | | 0.3603 | -? | 0.001153 | 0.915344 | 0.000854 | 5.6E-09 | TRUE | 0.00031 | -+ | 0.405706 | 1 | 0.010197 | 1 | 3.34E-08 | 1 |
| 12-37359433-TTCTG-T | T | 0.000275 | 0.323578 | | | 0.3236 | -? | 1.88E-05 | 0.392028 | 0.001757 | 4.9E-13 | TRUE | 3.38E-13 | ++ | 0.238942 | 1 | 0.002031 | 1 | 2.61E-11 | 1 |
| 12-37831033-G-GT | GT | | | | | | | | | 0.000361 | 1.37E-09 | TRUE | 1.37E-09 | ?- | 0.001452 | 1 | 0.17845 | 1 | 8.12E-08 | 1 |
| 12-40483640-C-T | T | | | | | | | 0.203718 | 0.134069 | 0.212192 | 2.07E-09 | TRUE | 0.0248 | -+ | 0.110416 | 1 | 0.114347 | 1 | 1.47E-07 | 1 |
| 12-42207674-C-CTT | CTT | | | | | | | | | 0.000995 | 2.55E-09 | TRUE | 2.55E-09 | ?+ | 0.199448 | 1 | 0.058802 | 1 | 6.57E-08 | 1 |
| 12-48802317-A-G | G | | | 0.372 | 0.197233 | 0.1972 | ?+ | 0.35256 | 0.72139 | 0.435202 | 4.53E-13 | FALSE | 1.13E-05 | -- | 0.656788 | 1 | 0.203898 | 1 | 1.22E-10 | 1 |
| 12-48988805-C-CT | CT | | | | | | | | | 0.000576 | 3.28E-11 | TRUE | 3.28E-11 | ?+ | 0.000319 | 1 | 0.307316 | 1 | 2.24E-08 | 1 |
| 12-49123265-C-T | T | 0.000587 | 0.008676 | | | 0.008676 | +? | 0.004229 | 0.390242 | 0.002067 | 3.68E-09 | TRUE | 0.02926 | -+ | 0.036345 | 1 | 0.026974 | 1 | 8.87E-07 | 1 |
| 12-49515941-G-A | A | | | | | | | 0.001084 | 0.377129 | 0.000947 | 1.64E-09 | TRUE | 3.78E-06 | ++ | 0.002419 | 1 | 0.076752 | 1 | 7.62E-07 | 1 |
| 12-49515961-A-G | G | 0.000137 | 0.757888 | | | 0.7579 | -? | 0.003502 | 0.034456 | 0.001885 | 1.02E-15 | TRUE | 8.41E-10 | -- | 0.018528 | 1 | 0.021888 | 1 | 2.29E-13 | 1 |
| 12-49611726-G-GT | GT | 0.000123 | 0.989556 | | | 0.9896 | +? | | | 0.006759 | 6.22E-10 | TRUE | 6.22E-10 | ?- | 0.316746 | 1 | 0.008803 | 1 | 1.24E-07 | 1 |
| 12-50015024-CAG-C | C | | | | | | | | | 0.000977 | 6.21E-09 | TRUE | 6.21E-09 | ?- | 0.800517 | 1 | 0.013217 | 1 | 2.8E-07 | 1 |
| 12-50015583-C-CCT | CCT | | | | | | | | | 0.001368 | 1.17E-08 | TRUE | 1.17E-08 | ?+ | 0.71383 | -1 | 0.678394 | 1 | 7.69E-10 | 1 |
| 12-50234243-C-CT | CT | | | | | | | | | 0.000152 | 5.7E-09 | TRUE | 5.7E-09 | ?+ | | | | | 7.12E-07 | 1 |
| 12-50301831-GAGGC- | G | | | | | | | | | 0.000862 | 1.48E-08 | TRUE | 1.48E-08 | ?+ | 0.028554 | 1 | 0.379968 | 1 | 1.08E-06 | 1 |
| 12-50301840-G-GGGC | GGGCA | | | | | | | | | 0.000829 | 3.1E-08 | TRUE | 3.11E-08 | ?- | 0.098953 | 1 | 0.048993 | 1 | 1.53E-06 | 1 |
| 12-50301871-T-C | C | | | | | | | | | 0.000351 | 1.73E-08 | TRUE | 1.73E-08 | ?- | 0.346574 | 1 | 0.513013 | 1 | 1.33E-07 | 1 |
| 12-50352078-T-TGA | TGA | | | | | | | | | 0.259377 | 9.16E-09 | FALSE | 9.16E-09 | ?+ | 0.855847 | 1 | 0.740442 | -1 | 2.45E-09 | -1 |
| 12-51720075-C-CA | CA | | | | | | | | | 0.738173 | 5.86E-09 | TRUE | 5.86E-09 | ?+ | 0.464586 | 1 | 0.13423 | 1 | 1.68E-08 | 1 |
| 12-53132921-C-A | A | | | | | | | 0.00124 | 0.758606 | 0.000754 | 3.72E-09 | TRUE | 0.000811 | -+ | 0.487441 | 1 | 0.007171 | 1 | 1.63E-08 | 1 |
| 12-53434594-T-TTG | TTG | | | | | | | | | 0.000259 | 8.32E-09 | TRUE | 8.32E-09 | ?- | 0.020157 | 1 | 0.435288 | 1 | 5.62E-08 | 1 |
| 12-53443104-A-ACCC( | ACCCG | | | | | | | | | 0.00031 | 2.88E-08 | TRUE | 2.88E-08 | ?- | 0.491481 | 1 | 0.024577 | 1 | 3.99E-07 | 1 |
| 12-53529371-T-A | A | 0.000351 | 0.700403 | | | 0.7004 | -? | | | 0.001407 | 2.45E-10 | TRUE | 2.45E-10 | ?+ | 0.623975 | -1 | 0.007983 | 1 | 4.81E-07 | 1 |
| 12-53537240-TG-T | T | | | | | | | | | 0.000175 | 3.17E-11 | TRUE | 3.17E-11 | ?+ | | | | | 3.8E-10 | 1 |

| | | | | | | | | | | | | | | | | | | |
|---|---|---|---|---|---|---|---|---|---|---|---|---|---|---|---|---|---|---|
| 12-53673139-T-A | A | 0.000216 | 0.231293 | 0.2313 | -? | 0.000836 | 0.513582 | 0.000755 | 1.56E-09 | TRUE | 0.000321 | -+ | 9.96E-05 | 1 | 0.000474 | 1 | 9.57E-06 | 1 |
| 12-54310262-G-A | A | | | | | 0.0011 | 0.265957 | 0.001535 | 1.94E-13 | TRUE | 2.44E-06 | -+ | 0.006356 | 1 | 0.144131 | 1 | 2.25E-10 | 1 |
| 12-54336354-CTA-C | C | | | | | | | 0.000955 | 1.12E-11 | TRUE | 1.12E-11 | ?- | 0.010752 | 1 | 0.056339 | 1 | 2.16E-09 | 1 |
| 12-55916145-G-A | A | 0.000413 | 0.059561 | 0.05956 | -? | 0.001015 | 0.285354 | 0.001098 | 3.28E-15 | TRUE | 1.34E-10 | ++ | 0.00704 | 1 | 0.00012 | 1 | 1.83E-11 | 1 |
| 12-56536657-TGGTG-T | T | | | | | | | 0.00047 | 1.83E-09 | TRUE | 1.83E-09 | ?+ | 0.017288 | 1 | 0.787517 | 1 | 1.81E-08 | 1 |
| 12-56656740-T-A | A | | | | | | | 0.000958 | 1.35E-19 | TRUE | 1.35E-19 | ?+ | 4.84E-05 | 1 | 0.06303 | 1 | 2.44E-15 | 1 |
| 12-56656759-G-C | C | | | | | | | 0.000516 | 1.63E-10 | TRUE | 1.63E-10 | ?+ | 0.005129 | 1 | 0.003218 | 1 | 4.28E-07 | 1 |
| 12-57017048-C-CTT | CTT | | | | | | | 0.001509 | 3.33E-12 | TRUE | 3.33E-12 | ?+ | 0.127175 | 1 | 0.860161 | -1 | 2.77E-11 | 1 |
| 12-64492451-CATG-C | C | | | | | | | 0.000647 | 3.13E-10 | TRUE | 3.13E-10 | ?- | 0.007336 | 1 | 0.00847 | 1 | 7.95E-07 | 1 |
| 12-64614426-C-CTGTI | CTGTTTCTTAAAATTTCAACT | | | | | | | 9.62E-05 | 3.58E-08 | TRUE | 3.58E-08 | ?+ | | | | | 7.64E-09 | 1 |
| 12-66938156-TTG-T | T | | | | | | | 0.000996 | 3.48E-10 | TRUE | 3.48E-10 | ?+ | 0.071239 | 1 | 0.029974 | 1 | 1.16E-08 | 1 |
| 12-69200553-A-ATGA( | ATGAGGAAGAGATAGGAGATGAGGAGGAG | | | | | | | 0.211356 | 1.09E-08 | FALSE | 1.09E-08 | ?+ | 0.015436 | -1 | 0.316089 | -1 | 6.39E-08 | -1 |
| 12-69200553-A-G | G | | | | | | | 0.663312 | 1.3E-08 | FALSE | 1.3E-08 | ?- | 0.162051 | 1 | 0.821436 | 1 | 9.06E-09 | 1 |
| 12-69759216-G-GT | GT | 0.005092 | 0.387943 | 0.3879 | -? | 0.00073 | 0.64759 | 0.002501 | 4.1E-08 | TRUE | 2.67E-05 | ++ | 0.131582 | -1 | 0.809534 | 1 | 3.54E-08 | -1 |
| 12-70418756-A-G | G | | | | | 9.08E-05 | 0.969392 | 0.00035 | 4.56E-09 | TRUE | 1.66E-07 | +- | 0.004765 | 1 | 0.159557 | 1 | 1.57E-07 | 1 |
| 12-72480233-A-G | G | 0.420309 | 0.035186 | 0.03519 | +? | | | 0.44299 | 6.82E-10 | FALSE | 6.82E-10 | ?+ | 0.993528 | -1 | 0.005489 | -1 | 2.08E-07 | -1 |
| 12-73182069-TA-T | T | | | | | | | 0.000877 | 1.78E-09 | TRUE | 1.78E-09 | ?+ | 0.020057 | 1 | 0.017695 | 1 | 6.54E-08 | 1 |
| 12-82133712-C-A | A | | | | | | | 0.033646 | 4.56E-08 | FALSE | 4.56E-08 | ?- | 0.075877 | -1 | 0.173157 | -1 | 2.51E-06 | -1 |
| 12-86814258-ATATAT. | A | | | | | | | 0.359549 | 6.14E-33 | FALSE | 6.15E-33 | ?- | 0.734548 | -1 | 0.000192 | -1 | 4.72E-30 | -1 |
| 12-89389651-G-T | T | | | | | | | 0.000653 | 1.51E-13 | TRUE | 1.51E-13 | ?+ | 0.836446 | 1 | 0.118994 | 1 | 3.39E-13 | 1 |
| 12-92835051-TC-T | T | 0.000253 | 0.534377 | 0.5344 | +? | | | 0.002431 | 1.63E-14 | TRUE | 1.63E-14 | ?+ | 0.000243 | 1 | 0.307448 | 1 | 8.29E-11 | 1 |
| 12-94946026-AT-A | A | | | | | | | 0.000685 | 1.52E-09 | TRUE | 1.52E-09 | ?+ | 0.19906 | 1 | 0.015611 | 1 | 7.59E-08 | 1 |
| 12-95612573-CAT-C | C | 0.002423 | 0.46635 | 0.4663 | -? | | | 0.003517 | 1.7E-12 | TRUE | 1.7E-12 | ?- | 3.23E-05 | 1 | 0.00344 | 1 | 8.95E-08 | 1 |
| 12-101445846-C-A | A | | | | | | | 0.000514 | 8.89E-15 | TRUE | 8.89E-15 | ?+ | 0.000125 | 1 | 0.020526 | 1 | 7.23E-11 | 1 |
| 12-103321503-C-T | T | | | | | | | 0.055972 | 8.36E-11 | TRUE | 8.36E-11 | ?+ | 0.355274 | 1 | 1.87E-05 | 1 | 3.33E-07 | 1 |
| 12-108905116-G-A | A | | | | | 5.64E-05 | 0.110957 | 0.000654 | 5.66E-13 | TRUE | 1.66E-13 | ++ | 0.000712 | 1 | 0.011841 | 1 | 1.04E-10 | 1 |
| 12-110043629-G-A | A | 0.000331 | 0.872646 | 0.8726 | -? | 0.00015 | 0.702288 | 0.000431 | 1.27E-08 | TRUE | 1.2E-06 | -+ | 0.000388 | 1 | 0.092557 | 1 | 5.36E-07 | 1 |
| 12-110702738-TC-T | T | | | | | 3.13E-05 | 0.264035 | 0.000209 | 1.33E-08 | TRUE | 3.2E-07 | -+ | 0.028724 | 1 | 0.022061 | 1 | 4.81E-07 | 1 |
| 12-110747045-T-C | C | | | | | 0.37403 | 0.213917 | 0.469262 | 6.25E-09 | FALSE | 2.18E-05 | -- | 0.051477 | 1 | 0.786633 | 1 | 2.42E-06 | 1 |
| 12-111611215-ATT-A | A | 0.000156 | 0.875324 | 0.8753 | -? | | | 0.000793 | 2.92E-10 | TRUE | 2.92E-10 | ?+ | 0.281466 | 1 | 0.629426 | 1 | 7.61E-10 | 1 |
| 12-111929280-G-A | A | 0.000156 | 0.276096 | 0.2761 | -? | | | 0.000575 | 6.29E-14 | TRUE | 6.29E-14 | ?+ | 0.008572 | 1 | 0.000936 | 1 | 5.24E-10 | 1 |
| 12-111929282-G-A | A | 7.81E-05 | 0.631556 | 0.6316 | -? | | | 0.000333 | 1.37E-09 | TRUE | 1.37E-09 | ?+ | 0.008743 | 1 | 0.000891 | 1 | 7.68E-07 | 1 |
| 12-112412907-C-G | G | | | | | | | 0.001038 | 3.29E-11 | TRUE | 3.3E-11 | ?- | 0.740505 | 1 | 0.037972 | 1 | 2.9E-11 | 1 |
| 12-112412911-G-GA | GA | | | | | | | 0.000871 | 8.16E-16 | TRUE | 8.16E-16 | ?- | 0.788121 | -1 | 0.013871 | 1 | 2.35E-15 | 1 |
| 12-114298592-TCCTC- | T | | | | | | | 0.001818 | 1.76E-11 | TRUE | 1.76E-11 | ?+ | 0.010954 | 1 | 0.065342 | 1 | 1.84E-07 | 1 |
| 12-117081702-G-T | T | 7.81E-05 | 0.935899 | 0.9359 | -? | 0.000601 | 0.961506 | 0.001783 | 9.82E-12 | TRUE | 1.01E-08 | ++ | 0.000205 | 1 | 0.961551 | -1 | 8.93E-09 | 1 |
| 12-117793145-T-C | C | | | | | 0.454012 | 0.985784 | 0.601578 | 2.59E-10 | FALSE | 0.000853 | -- | 0.479569 | 1 | 0.045283 | 1 | 2.95E-08 | 1 |
| 12-118065595-A-T | T | 9.75E-05 | 0.570885 | 0.5709 | -? | 0.000821 | 0.708606 | 0.001272 | 5.54E-10 | TRUE | 9.72E-07 | -- | 2.21E-07 | 1 | 0.001494 | 1 | 0.000538 | 1 |
| 12-120311520-T-C | C | | | | | | | 0.000483 | 6.35E-10 | TRUE | 6.35E-10 | ?- | 0.168575 | 1 | 0.708991 | 1 | 2.32E-10 | 1 |
| 12-120556370-TAAAA | T | | | | | | | 0.00103 | 3.11E-10 | TRUE | 3.11E-10 | ?+ | 0.002298 | 1 | 0.158768 | 1 | 9.16E-09 | 1 |
| 12-120663475-C-T | T | | | | | 4.39E-05 | 0.438428 | 0.001427 | 7.21E-14 | TRUE | 6.3E-14 | ++ | 0.005216 | 1 | 0.13682 | 1 | 1.44E-11 | 1 |
| 12-120663495-C-G | G | 0.000137 | 0.233834 | 0.2338 | +? | | | 0.001781 | 1.96E-15 | TRUE | 1.96E-15 | ?- | 0.000827 | 1 | 0.032997 | 1 | 1.79E-13 | 1 |
| 12-120893304-T-G | G | 0.000612 | 0.149763 | 0.1498 | -? | 0.001557 | 0.112503 | 0.002141 | 4.04E-10 | TRUE | 0.000649 | +- | 0.65303 | -1 | 0.369201 | 1 | 4.51E-10 | 1 |
| 12-121034133-G-C | C | | | | | | | 0.000745 | 3.4E-12 | TRUE | 3.4E-12 | ?+ | 0.00058 | 1 | 0.008689 | 1 | 1.72E-09 | 1 |
| 12-121034789-C-G | G | | | | | 0.000429 | 0.143081 | 0.002721 | 7.85E-20 | TRUE | 3.32E-19 | -- | 0.000347 | 1 | 0.063508 | 1 | 1.89E-16 | 1 |
| 12-121076600-TG-T | T | | | | | | | 0.9497 | 2.78E-11 | TRUE | 2.78E-11 | ?- | 0.238701 | -1 | 0.938785 | -1 | 2.66E-13 | -1 |
| 12-121226389-T-TGA | TGA | | | | | 0.000498 | 0.409897 | 0.001318 | 1.21E-09 | TRUE | 4.03E-06 | +- | 0.19381 | 1 | 0.045692 | 1 | 1.05E-08 | 1 |
| 12-121514771-C-CTA | CTA | | | | | | | 0.393022 | 1.99E-10 | FALSE | 1.99E-10 | ?+ | 0.134012 | 1 | 0.170108 | 1 | 8.09E-09 | 1 |
| 12-121617997-CCA-C | C | 0.002277 | 0.839888 | 0.8399 | +? | | | 0.002136 | 1.78E-09 | TRUE | 1.78E-09 | ?- | 0.888542 | 1 | 0.051633 | 1 | 4.14E-10 | 1 |
| 12-121634293-G-A | A | 0.000844 | 0.95073 | 0.9507 | +? | 9.4E-06 | 0.250878 | 0.003298 | 8.14E-25 | TRUE | 5.03E-25 | ++ | 1.66E-05 | 1 | 0.011671 | 1 | 5.66E-20 | 1 |
| 12-121634366-G-T | T | 9.77E-05 | 0.516062 | 0.5161 | -? | 8.46E-05 | 0.508955 | 0.001094 | 1.8E-08 | TRUE | 2.01E-07 | -+ | 0.079888 | 1 | 0.213459 | 1 | 9.1E-08 | 1 |
| 12-121670590-CCT-C | C | | | | | 0.000432 | 0.77022 | 0.006403 | 4.58E-43 | TRUE | 2.11E-40 | -- | 0.007641 | 1 | 0.000959 | 1 | 4.99E-36 | 1 |
| 12-121726344-CCGAT | C | | | | | | | 0.001454 | 1.53E-15 | TRUE | 1.53E-15 | ?- | 0.073389 | 1 | 0.740778 | 1 | 1.02E-14 | 1 |
| 12-121745498-CT-C | C | | | | | | | 0.000613 | 1.11E-09 | TRUE | 1.11E-09 | ?- | 0.020014 | 1 | 0.093199 | 1 | 4.95E-09 | 1 |
| 12-121747283-C-T | T | | | | | | | 0.888514 | 1.06E-11 | FALSE | 1.06E-11 | ?- | 0.223736 | -1 | 0.024298 | -1 | 2.49E-10 | -1 |
| 12-121876914-TGA-T | T | | | | | | | 0.000351 | 2.93E-08 | TRUE | 2.93E-08 | ?+ | 0.002775 | 1 | 0.010682 | 1 | 8.26E-06 | 1 |
| 12-121914347-C-CT | CT | | | | | | | 0.001265 | 3.05E-10 | TRUE | 3.05E-10 | ?+ | 0.011738 | 1 | 1.97E-06 | 1 | 5.89E-06 | 1 |
| 12-122320066-ATATT( | A | | | | | | | 0.001202 | 4.76E-08 | TRUE | 4.77E-08 | ?+ | 0.020917 | 1 | 0.332719 | -1 | 2.26E-08 | 1 |
| 12-123173072-C-T | T | | | | | 0.000457 | 0.969602 | 0.001857 | 1.87E-13 | TRUE | 8.06E-11 | ++ | 4.77E-05 | 1 | 0.052021 | 1 | 2.67E-09 | 1 |

| | | | | | | | | | | | | | | | | | | | | |
|---|---|---|---|---|---|---|---|---|---|---|---|---|---|---|---|---|---|---|---|---|
| 12-123173111-A-G | G | 7.82E-05 | 0.521628 | | | 0.5216 | -? | 0.000749 | 0.62016 | 0.001277 | 1.03E-15 | TRUE | 7.34E-11 | -- | 0.00449 | 1 | 0.028963 | 1 | 1.18E-12 | 1 |
| 12-123717186-T-G | G | | | | | | | 0.00063 | 0.287021 | 0.000985 | 1.67E-11 | TRUE | 5.02E-09 | -- | 0.627248 | 1 | 0.540677 | 1 | 6.01E-12 | 1 |
| 12-123755449-T-TGG | TGG | | | | | | | | | 0.00075 | 3.67E-09 | TRUE | 3.67E-09 | ?- | 0.913311 | 1 | 0.008622 | 1 | 2.49E-08 | 1 |
| 12-123755925-CA-C | C | | | | | | | | | 0.000865 | 1.06E-13 | TRUE | 1.06E-13 | ?- | 0.026677 | 1 | 0.556176 | 1 | 1.99E-11 | 1 |
| 12-123755940-G-T | T | | | | | | | 0.000175 | 0.49587 | 0.000263 | 2.42E-09 | TRUE | 1.36E-07 | ++ | 0.164449 | 1 | | | 1.92E-07 | 1 |
| 12-123755941-C-T | T | | | | | | | 0.000175 | 0.49587 | 0.000469 | 4.06E-10 | TRUE | 7.92E-09 | ++ | 0.109394 | 1 | 0.764869 | 1 | 1.36E-08 | 1 |
| 12-124547213-G-A | A | | | | | | | 2.51E-05 | 0.457873 | 0.000646 | 2.04E-11 | TRUE | 1.78E-11 | ++ | 0.005299 | 1 | 0.687727 | 1 | 3.02E-09 | 1 |
| 12-124547226-CAA-C | C | | | | | | | | | 0.001383 | 2.31E-14 | TRUE | 2.31E-14 | ?- | 0.013772 | 1 | 0.478627 | 1 | 1.41E-13 | 1 |
| 12-124951817-C-CA | CA | | | | | | | | | 0.000204 | 2.44E-08 | TRUE | 2.44E-08 | ?+ | 4.41E-07 | 1 | | | 3.98E-05 | 1 |
| 12-125090196-C-CCTC | CCTCCACCCAT | | | | | | | | | 0.961029 | 7.08E-10 | FALSE | 7.08E-10 | ?- | 0.068922 | -1 | 0.14926 | -1 | 1.8E-08 | -1 |
| 12-125090200-T-C | C | | | | | | | | | 0.963429 | 9E-10 | FALSE | 9E-10 | ?+ | 0.047165 | -1 | 0.13591 | -1 | 1.71E-07 | -1 |
| 12-125090208-T-TCAT | TCATCCATCCATCCTCCATTCATCATCCCTCATCCATCATTTATCCAACTATCC | | | | | | | | | 0.953495 | 1.15E-13 | TRUE | 1.15E-13 | ?+ | 0.060559 | -1 | 0.021633 | -1 | 1.41E-10 | -1 |
| 12-125090216-T-TGTC | TGTCTACACATTCTTCTCTCCACCTACCCATCCATCCATCC | | | | | | | | | 0.915741 | 2.17E-14 | FALSE | 2.17E-14 | ?+ | 0.002737 | -1 | 0.061805 | -1 | 3.27E-11 | -1 |
| 12-125803495-G-A | A | | | | | | | 7.2E-05 | 0.331749 | 0.000429 | 3.65E-09 | TRUE | 4.72E-09 | ++ | 0.458125 | -1 | 0.012199 | 1 | 1.31E-08 | 1 |
| 12-126712968-G-C | C | | | | | | | 0.005 | 0.793387 | 0.002281 | 7.88E-11 | TRUE | 0.003985 | -+ | 0.808906 | -1 | 0.034287 | 1 | 1.42E-10 | 1 |
| 12-126713469-T-C | C | 0.000587 | 0.96452 | | | 0.9645 | -? | 0.00503 | 0.75722 | 0.002964 | 6.04E-20 | TRUE | 3.14E-07 | +- | 0.200903 | 1 | 0.001244 | 1 | 1.14E-15 | 1 |
| 12-128596107-CG-C | C | | | | | | | 0.000911 | 0.999769 | 0.002207 | 3.27E-14 | TRUE | 1.04E-09 | -- | 0.014233 | 1 | 0.239506 | 1 | 9.04E-13 | 1 |
| 12-128681911-C-CTG | CTG | | | | | | | | | 0.000753 | 1.39E-08 | TRUE | 1.39E-08 | ?+ | 0.891075 | -1 | 0.462173 | 1 | 7.81E-09 | 1 |
| 12-130798250-CA-C | C | | | | | | | | | 0.923897 | 1.75E-08 | TRUE | 1.75E-08 | ?+ | 0.203898 | -1 | 0.385625 | -1 | 5.14E-08 | -1 |
| 12-131555849-A-C | C | | | | | | | | | 0.065956 | 3.75E-09 | FALSE | 3.75E-09 | ?- | 0.521478 | 1 | 0.256788 | 1 | 1.35E-08 | 1 |
| 12-132136389-CA-C | C | | | | | | | 0.007658 | 0.007194 | 0.00742 | 3.17E-12 | TRUE | 0.01611 | +- | 0.100082 | 1 | 0.066761 | 1 | 2.18E-09 | 1 |
| 12-132381811-A-AGC | AGCACCCGATACACAGAAGACAGAAGACATACTCAGCTAAAGCT | | | | | | | | | 0.001144 | 2.71E-09 | TRUE | 2.71E-09 | ?- | 0.765323 | 1 | 0.017912 | 1 | 1.68E-08 | 1 |
| 12-132683958-CAG-C | C | | | | | | | 0.001986 | 0.960693 | 0.003751 | 5.28E-11 | TRUE | 6.3E-07 | -- | 0.87134 | -1 | 0.009778 | 1 | 1.58E-08 | 1 |
| 12-123698702-G-C | C | 0.011617 | 0.22202 | 0.017 | 0.038275 | 0.2063 | ++ | 0.01858 | 0.88518 | 0.010926 | 0.103591 | FALSE | 2.9E-305 | -+ | 0.272306 | 1 | 0.303821 | 1 | 0.260163 | 1 |
| 12-66249311-G-A | A | | | | | | | 7.83E-05 | 0.187935 | | | FALSE | 1.9E-291 | -? | | | | | | |
| 12-53198430-T-A | A | | | | | | | 2.19E-05 | 0.068881 | | | FALSE | 4E-305 | +? | | | | | | |
| 13-30500545-T-A | A | 0.007735 | 1.15E-08 | | | 1.154E-08 | +? | 0.000247 | 0.313363 | | | FALSE | 0.3134 | +? | | | | | | |
| 13-19187520-T-TGG | TGG | 0.000234 | 0.43469 | | | 0.4347 | +? | | | 0.00144 | 4.28E-08 | TRUE | 4.28E-08 | ?- | 0.039738 | 1 | 0.557516 | -1 | 7.81E-09 | 1 |
| 13-19344406-T-TTC | TTC | | | | | | | | | 0.873344 | 4.12E-09 | FALSE | 4.12E-09 | ?- | 0.142926 | 1 | 0.50135 | 1 | 1.65E-08 | 1 |
| 13-19355631-T-G | G | | | | | | | | | 0.075304 | 8.16E-09 | FALSE | 8.16E-09 | ?- | 0.348061 | 1 | 0.571302 | 1 | 6.95E-10 | 1 |
| 13-19407752-C-CA | CA | | | | | | | | | 0.02459 | 8.59E-11 | FALSE | 8.59E-11 | ?- | 0.011478 | -1 | 0.023108 | -1 | 9.39E-07 | -1 |
| 13-19922626-C-CCT | CCT | 0.000351 | 0.631213 | | | 0.6312 | -? | | | 0.003003 | 2.76E-19 | TRUE | 2.76E-19 | ?+ | 0.000293 | 1 | 0.054669 | 1 | 6.12E-16 | 1 |
| 13-20266218-T-C | C | | | | | | | 0.000423 | 0.899642 | 0.000657 | 8.87E-10 | TRUE | 1.96E-06 | +- | 0.012525 | 1 | 0.325708 | 1 | 2.54E-08 | 1 |
| 13-20470690-CCT-C | C | | | | | | | | | 0.001288 | 1.62E-11 | TRUE | 1.62E-11 | ?- | 7.11E-05 | 1 | 0.018862 | 1 | 1.04E-07 | 1 |
| 13-21290448-TATGAC | T | | | | | | | | | 0.029718 | 4.87E-08 | FALSE | 4.87E-08 | ?+ | 0.532662 | 1 | 0.255161 | 1 | 7.77E-09 | 1 |
| 13-21627637-ATT-A | A | | | | | | | | | 0.001182 | 3.39E-09 | TRUE | 3.39E-09 | ?+ | 0.028149 | 1 | 0.017883 | 1 | 2.49E-07 | 1 |
| 13-27054320-G-C | C | 0.000842 | 0.813303 | | | 0.8133 | +? | 0.000291 | 0.223738 | 0.001081 | 4.54E-09 | TRUE | 1.04E-08 | ++ | 0.012331 | 1 | 0.174995 | -1 | 4.34E-09 | 1 |
| 13-28596076-T-C | C | | | | | | | 0.000366 | 0.186211 | 0.000461 | 1.71E-08 | TRUE | 3.64E-07 | -- | 0.016433 | 1 | 0.010286 | 1 | 0.000118 | 1 |
| 13-32274495-C-CAGG | CAGG | | | | | | | 0.051188 | 0.099864 | 0.074883 | 2.57E-09 | TRUE | 0.01649 | -+ | 0.12526 | 1 | 0.254348 | -1 | 9.26E-10 | 1 |
| 13-37280427-G-A | A | | | | | | | | | 0.886193 | 1.18E-09 | FALSE | 1.18E-09 | ?- | 0.833493 | -1 | 0.055345 | -1 | 6.04E-12 | -1 |
| 13-38807189-T-A | A | | | | | | | | | 0.060375 | 5.6E-11 | FALSE | 5.6E-11 | ?+ | 0.065621 | 1 | 0.731973 | 1 | 8.34E-10 | 1 |
| 13-40685666-A-C | C | | | | | | | 9.08E-05 | 0.893019 | 0.000799 | 2.91E-10 | TRUE | 2.06E-09 | -- | 0.714705 | -1 | 0.617575 | -1 | 7.38E-12 | 1 |
| 13-41443169-C-CT | CT | | | | | | | | | 0.121999 | 4.57E-09 | FALSE | 4.57E-09 | ?+ | 0.000548 | 1 | 0.237884 | 1 | 1.77E-07 | 1 |
| 13-41850639-T-C | C | | | | | | | 7.83E-05 | 0.283844 | 0.000948 | 5.41E-15 | TRUE | 5.68E-15 | -- | 0.030159 | 1 | 0.012975 | 1 | 1.03E-12 | 1 |
| 13-42917642-G-A | A | | | | | | | | | 0.003121 | 1.97E-09 | TRUE | 1.97E-09 | ?- | 0.672851 | 1 | 0.176353 | -1 | 5.17E-09 | -1 |
| 13-44569304-G-C | C | | | | | | | 0.000846 | 0.541382 | 0.005197 | 1.77E-10 | TRUE | 2.48E-08 | -- | 0.020886 | -1 | 0.281886 | -1 | 2.54E-08 | -1 |
| 13-45383701-G-C | C | | | | | | | | | 0.010248 | 9.88E-17 | TRUE | 9.88E-17 | ?+ | 0.41383 | 1 | 0.002513 | 1 | 5.92E-16 | 1 |
| 13-47381305-CA-C | C | | | | | | | | | 0.765915 | 5.37E-11 | FALSE | 5.37E-11 | ?- | 0.868321 | -1 | 0.091063 | 1 | 3.83E-12 | 1 |
| 13-47381305-CAA-C | C | | | | | | | | | 0.030944 | 2.41E-42 | FALSE | 2.41E-42 | ?+ | 0.172726 | -1 | 0.008176 | -1 | 1.52E-41 | -1 |
| 13-47381363-A-C | C | | | | | | | | | 0.730897 | 1.15E-20 | FALSE | 1.15E-20 | ?- | 0.299942 | 1 | 0.03949 | 1 | 1.5E-20 | 1 |
| 13-48428421-G-GCAC | GCACATTTCAACACGAGTCATCTTCTACCAAACCTCACCTCCAGCATTGGGGAT | | | | | | | | | 0.884698 | 1.75E-08 | FALSE | 1.75E-08 | ?+ | 0.609394 | 1 | 0.96875 | -1 | 4.25E-13 | -1 |
| 13-49525435-ATG-A | A | | | | | | | | | 0.000356 | 2.58E-10 | TRUE | 2.58E-10 | ?+ | 0.000916 | 1 | 0.154416 | 1 | 2.69E-08 | 1 |
| 13-49525439-A-AAG | AAG | | | | | | | | | 0.000395 | 2.11E-09 | TRUE | 2.12E-09 | ?- | 0.001228 | 1 | 0.157979 | 1 | 1.78E-07 | 1 |
| 13-72705499-GCACC-( | G | | | | | | | | | 0.875934 | 1.11E-08 | FALSE | 1.12E-08 | ?+ | 0.978064 | -1 | 0.00338 | 1 | 2.27E-12 | 1 |
| 13-72705503-C-CGGA | CGGAAATTT | | | | | | | | | 0.889033 | 2.04E-08 | FALSE | 2.04E-08 | ?+ | 0.849534 | 1 | 0.002263 | 1 | 1E-10 | 1 |
| 13-79091376-A-G | G | 0.00257 | 0.480703 | | | 0.4807 | -? | | | 0.002462 | 9.48E-09 | TRUE | 9.48E-09 | ?+ | 0.028969 | -1 | 0.352692 | -1 | 5.66E-07 | -1 |
| 13-82155975-G-A | A | | | | | | | | | 0.000398 | 3.41E-08 | TRUE | 3.41E-08 | ?+ | 0.248201 | 1 | 0.074979 | 1 | 5.1E-08 | 1 |
| 13-85751762-T-A | A | | | | | | | | | 0.007615 | 2.32E-08 | FALSE | 2.32E-08 | ?+ | 0.054776 | 1 | 0.425152 | 1 | 2.59E-07 | 1 |
| 13-97948951-CTG-C | C | | | | | | | | | 0.000645 | 4.99E-08 | TRUE | 4.99E-08 | ?- | 0.900364 | 1 | 0.830005 | -1 | 3.57E-08 | 1 |

| | | | | | | | | | | | | | | | | | | | | |
|---|---|---|---|---|---|---|---|---|---|---|---|---|---|---|---|---|---|---|---|---|
| 13-98116951-GGA-G | G | 0.001074 | 0.399705 | | | 0.3997 | -? | | | 0.002612 | 7.17E-14 | TRUE | 7.17E-14 | ?+ | 0.10005 | 1 | 4.81E-05 | 1 | 4.09E-10 | 1 |
| 13-98643115-A-C | C | | | | | | | 9.9E-05 | 0.732376 | 0.001149 | 7.97E-13 | TRUE | 3.07E-12 | -- | 0.006585 | 1 | 0.155003 | 1 | 1.08E-10 | 1 |
| 13-99952110-C-CTT | CTT | | | | | | | | | 0.002569 | 3.18E-20 | TRUE | 3.18E-20 | ?+ | 0.002308 | 1 | 1.63E-05 | 1 | 1.93E-13 | 1 |
| 13-99952112-CAG-C | C | | | | | | | | | 0.00252 | 5.3E-26 | TRUE | 5.3E-26 | ?- | 0.000581 | 1 | 1.14E-05 | 1 | 4.89E-18 | 1 |
| 13-99952132-AT-A | A | | | | | | | | | 0.001252 | 1.22E-10 | TRUE | 1.22E-10 | ?+ | 0.008406 | 1 | 0.006285 | 1 | 1.92E-07 | 1 |
| 13-100092421-TG-T | T | | | | | | | | | 0.001021 | 2.74E-08 | TRUE | 2.74E-08 | ?+ | 0.112101 | 1 | 0.517596 | -1 | 5.01E-09 | 1 |
| 13-101198906-G-T | T | | | | | | | 0.000561 | 0.826955 | 0.000374 | 2.03E-08 | TRUE | 0.000509 | -+ | 0.463052 | -1 | 0.011526 | 1 | 8.92E-09 | 1 |
| 13-107175091-G-A | A | | | | | | | 0.002594 | 0.308101 | 0.000793 | 1.61E-08 | TRUE | 0.000502 | ++ | 0.01007 | 1 | 0.007594 | 1 | 1.47E-05 | 1 |
| 13-110656083-ACC-A | A | | | | | | | | | 0.000553 | 4.8E-11 | TRUE | 4.8E-11 | ?+ | 0.029494 | 1 | 0.058699 | 1 | 9.65E-09 | 1 |
| 13-111960813-CGATG | C | | | | | | | | | 0.170547 | 1.5E-08 | TRUE | 1.5E-08 | ?- | 0.016961 | 1 | 0.695934 | 1 | 0.008817 | 1 |
| 13-112147484-C-T | T | | | | | | | | | 0.905953 | 1.88E-15 | FALSE | 1.88E-15 | ?- | 0.159908 | -1 | 0.069376 | -1 | 2.57E-14 | -1 |
| 13-113370479-G-C | C | | | | | | | 0.000714 | 0.865158 | 0.001106 | 1.33E-08 | TRUE | 1.21E-05 | ++ | 0.150417 | 1 | 0.200885 | 1 | 1.84E-07 | 1 |
| 13-113600427-T-C | C | | | | | | | | | 0.013848 | 1.63E-09 | TRUE | 1.63E-09 | ?- | 0.144535 | 1 | 0.817539 | 1 | 4.65E-10 | 1 |
| 13-113795470-AC-A | A | | | | | | | | | 0.003926 | 1.2E-11 | TRUE | 1.2E-11 | ?+ | 0.859054 | 1 | 0.832161 | 1 | 4.74E-13 | 1 |
| 13-114010763-G-GGA | GGAGGGGGCCGC | | | | | | | | | 0.860895 | 9.32E-13 | FALSE | 9.32E-13 | ?+ | 0.009465 | -1 | 0.111999 | -1 | 3.32E-09 | -1 |
| 13-114083511-CTGAG | C | | | | | | | | | 0.056597 | 9.89E-13 | FALSE | 9.89E-13 | ?- | 0.066341 | 1 | 0.083004 | 1 | 6.6E-11 | 1 |
| 13-114113552-CCAAT | C | | | | | | | | | 0.010262 | 4.61E-10 | TRUE | 4.61E-10 | ?- | 0.213684 | -1 | 0.730954 | 1 | 3.75E-11 | 1 |
| 13-61297289-C-T | T | | | | | | | 5.33E-05 | 0.869008 | | | FALSE | 3.2E-305 | -? | | | | | | |
| 13-100996739-GACAC | G | | | | | | | 0.000216 | 0.871009 | | | FALSE | 0.871 | +? | | | | | | |
| 13-45755202-C-A | A | | | | | | | 1.25E-05 | 0.079228 | | | FALSE | 2.4E-305 | -? | | | | | | |
| 13-100866498-G-C | C | | | | | | | 1.25E-05 | 0.37161 | | | FALSE | 2.53E-97 | -? | | | | | | |
| 13-18757253-G-A | A | | | | | | | 1.03E-05 | 0.5524 | | | FALSE | | | | | | | | |
| 14-20342296-CTCG-C | C | 0.000117 | 0.256855 | | | 0.2569 | +? | | | 0.00117 | 3.35E-15 | TRUE | 3.35E-15 | ?- | 0.017995 | 1 | 0.000935 | 1 | 8.29E-11 | 1 |
| 14-22727949-T-C | C | | | | | | | 6.89E-05 | 0.392326 | 0.000311 | 1.36E-10 | TRUE | 2.33E-08 | +- | 0.020486 | 1 | 0.542646 | -1 | 7.86E-11 | 1 |
| 14-22801564-A-ACT | ACT | | | | | | | | | 0.001035 | 1.26E-11 | TRUE | 1.26E-11 | ?- | 0.000835 | 1 | 0.015307 | 1 | 9.59E-08 | 1 |
| 14-23706574-T-C | C | 0.334583 | 0.705247 | | | 0.7052 | -? | 0.278494 | 0.6203 | 0.350575 | 1.68E-10 | FALSE | 4.74E-05 | -- | 0.594199 | -1 | 0.195864 | 1 | 6.22E-10 | 1 |
| 14-23706575-G-A | A | 0.000215 | 0.964141 | | | 0.9641 | +? | 2.51E-05 | 0.097764 | 0.003768 | 5.57E-19 | TRUE | 1.98E-19 | ++ | 0.000614 | 1 | 0.94011 | 1 | 1.11E-18 | 1 |
| 14-23706600-G-A | A | 0.000743 | 0.459433 | | | 0.4594 | -? | 0.005265 | 0.869328 | 0.005422 | 1.57E-23 | TRUE | 3.73E-11 | -+ | 0.056679 | 1 | 0.000161 | 1 | 1.71E-20 | 1 |
| 14-23706607-G-A | A | | | | | | | 0.005012 | 0.806871 | 0.011214 | 1.3E-24 | TRUE | 1.56E-14 | ++ | 0.074295 | 1 | 0.013272 | 1 | 3.5E-25 | 1 |
| 14-24170048-GT-G | G | | | | | | | | | 0.000803 | 1.27E-08 | TRUE | 1.27E-08 | ?+ | 0.021932 | 1 | 0.04869 | 1 | 4.06E-06 | 1 |
| 14-28854133-GGC-G | G | | | | | | | | | 0.001324 | 7.29E-11 | TRUE | 7.29E-11 | ?+ | 2.8E-06 | 1 | 0.000324 | 1 | 3.51E-06 | 1 |
| 14-28854137-CGT-C | C | | | | | | | | | 0.000857 | 1.83E-08 | TRUE | 1.83E-08 | ?- | 0.0014 | 1 | 0.00125 | 1 | 1.44E-05 | 1 |
| 14-28854140-G-GCCC. | GCCCA | | | | | | | | | 0.000781 | 2E-08 | TRUE | 2E-08 | ?- | 0.000949 | 1 | 0.001628 | 1 | 1.2E-05 | 1 |
| 14-31245601-G-A | A | 0.34215 | 0.477393 | 0.227 | 0.319323 | 0.5199 | -+ | | | 0.33388 | 7.97E-10 | TRUE | 7.97E-10 | ?+ | 0.010164 | 1 | 0.303985 | 1 | 0.000273 | 1 |
| 14-34487122-TA-T | T | 0.000235 | 0.100645 | | | 0.1006 | +? | | | 0.000618 | 1.89E-09 | TRUE | 1.89E-09 | ?+ | 0.812445 | 1 | 0.765389 | 1 | 3.53E-09 | 1 |
| 14-34487130-TTC-T | T | 0.000236 | 0.100466 | | | 0.1005 | +? | | | 0.001145 | 3.99E-11 | TRUE | 3.99E-11 | ?+ | 0.759041 | 1 | 0.889035 | 1 | 1.05E-10 | 1 |
| 14-34514025-CGTG-C | C | | | | | | | | | 0.000277 | 1.5E-10 | TRUE | 1.5E-10 | ?- | 0.01521 | 1 | 0.657785 | 1 | 1.92E-09 | 1 |
| 14-34565234-C-T | T | 7.81E-05 | 0.357317 | | | 0.3573 | +? | 0.003602 | 0.071581 | 0.001461 | 3.05E-16 | TRUE | 7.19E-09 | ++ | 0.156062 | 1 | 0.002305 | 1 | 1.25E-12 | 1 |
| 14-34565238-C-T | T | 9.76E-05 | 0.497903 | | | 0.4979 | -? | 0.003627 | 0.048577 | 0.00157 | 5.15E-14 | TRUE | 3.01E-08 | ++ | 0.003369 | 1 | 0.12232 | 1 | 2.13E-10 | 1 |
| 14-34565241-C-T | T | | | | | | | 0.003586 | 0.069 | 0.001782 | 1.36E-18 | TRUE | 1.53E-10 | ++ | 0.00946 | 1 | 0.103678 | 1 | 4.3E-15 | 1 |
| 14-34878995-T-G | G | | | | | | | 0.353799 | 0.573929 | 0.448076 | 2.12E-13 | FALSE | 0.000237 | +- | 0.010034 | 1 | 0.110858 | 1 | 1.06E-08 | 1 |
| 14-37236493-T-G | G | 0.12143 | 0.870825 | | | 0.8708 | -? | | | 0.137061 | 6.1E-09 | FALSE | 6.1E-09 | ?- | 0.133197 | 1 | 0.525534 | 1 | 0.254508 | 1 |
| 14-39101400-A-ACC | ACC | | | | | | | | | 0.001953 | 2.71E-12 | TRUE | 2.71E-12 | ?- | 0.001793 | 1 | 0.024242 | 1 | 4.99E-09 | 1 |
| 14-49652152-TAAGAA | T | | | | | | | | | 0.26763 | 2.46E-20 | FALSE | 2.46E-20 | ?- | 0.086167 | -1 | 0.866875 | -1 | 8.1E-21 | -1 |
| 14-54925838-C-A | A | | | | | | | 4.07E-05 | 0.957478 | 0.000456 | 5.07E-09 | TRUE | 1.65E-08 | ++ | 0.069451 | 1 | 0.314582 | 1 | 2.16E-09 | 1 |
| 14-55344909-TTGTC-1 | T | 0.000117 | 0.268904 | | | 0.2689 | +? | 0.000379 | 0.259637 | 0.00136 | 6.86E-09 | TRUE | 2.34E-08 | ++ | 0.0293 | 1 | 0.017073 | 1 | 5.48E-07 | 1 |
| 14-63519957-G-GCCA | GCCAAGAAGAAC | | | | | | | | | 0.001849 | 3.64E-08 | TRUE | 3.64E-08 | ?- | 0.023637 | 1 | 0.600334 | -1 | 1.05E-07 | 1 |
| 14-63585644-G-T | T | 0.000117 | 0.881826 | | | 0.8818 | -? | 0.000426 | 0.241319 | 0.001783 | 3.43E-17 | TRUE | 4.06E-12 | -+ | 0.010807 | 1 | 0.435815 | 1 | 4.21E-16 | 1 |
| 14-63585648-G-T | T | 9.76E-05 | 0.927106 | | | 0.9271 | +? | 0.000426 | 0.241182 | 0.001255 | 2.18E-13 | TRUE | 1.75E-08 | -+ | 0.002389 | 1 | 0.010345 | 1 | 1.25E-10 | 1 |
| 14-64547356-ACT-A | A | 0.000488 | 0.742191 | | | 0.7422 | +? | | | 0.002992 | 1.33E-09 | TRUE | 1.33E-09 | ?+ | 0.617846 | -1 | 9.14E-05 | 1 | 1.3E-07 | 1 |
| 14-65504716-ATTT-A | A | | | | | | | | | 0.000707 | 2.32E-08 | TRUE | 2.32E-08 | ?+ | 0.013067 | 1 | 0.133604 | 1 | 3.25E-06 | 1 |
| 14-66995017-TTA-T | T | | | | | | | | | 0.001156 | 4.16E-12 | TRUE | 4.16E-12 | ?+ | 0.006859 | 1 | 0.015421 | 1 | 6.13E-10 | 1 |
| 14-71326334-G-A | A | 0.000138 | 0.119264 | | | 0.1193 | -? | 0.000282 | 0.787902 | 0.000833 | 1.22E-08 | TRUE | 6.13E-07 | ++ | 0.092198 | 1 | 0.52107 | 1 | 1.96E-08 | 1 |
| 14-71326352-GCACA- | G | 0.000117 | 0.13831 | | | 0.1383 | -? | | | 0.001092 | 4.72E-10 | TRUE | 4.73E-10 | ?+ | 0.033812 | 1 | 0.598552 | -1 | 1.67E-10 | 1 |
| 14-71326357-C-CGTGT | CGTGT | | | | | | | | | 0.00088 | 1.19E-10 | TRUE | 1.19E-10 | ?+ | 0.012107 | 1 | 0.286626 | -1 | 1.97E-11 | 1 |
| 14-72837563-CCCCA-( | C | | | | | | | | | 0.001463 | 3.41E-08 | TRUE | 3.41E-08 | ?- | 0.009555 | 1 | 0.709364 | 1 | 3.39E-06 | 1 |
| 14-73480223-CTT-C | C | | | | | | | | | 0.00047 | 5.28E-09 | TRUE | 5.28E-09 | ?- | 0.005891 | 1 | 0.010333 | 1 | 6.63E-07 | 1 |
| 14-73952289-A-ATT | ATT | 0.000195 | 0.594516 | | | 0.5945 | -? | | | 0.001498 | 2.38E-14 | TRUE | 2.38E-14 | ?- | 0.001856 | 1 | 0.000723 | 1 | 1.17E-09 | 1 |
| 14-74540803-A-ATATT | ATATT | | | | | | | | | 0.072483 | 2.21E-09 | FALSE | 2.21E-09 | ?+ | 0.898858 | -1 | 0.481753 | -1 | 3.64E-10 | -1 |

| | | | | | | | | | | | | | | | | | | | | |
|---|---|---|---|---|---|---|---|---|---|---|---|---|---|---|---|---|---|---|---|---|
| 14-74540830-A-T | T | | | | | | | | | 0.08531 | 3.89E-11 | FALSE | 3.89E-11 | ?- | 0.374026 | 1 | 0.121506 | 1 | 6.99E-10 | 1 |
| 14-77308036-A-T | T | 0.000774 | 0.934735 | | | 0.9347 | +? | 0.00046 | 0.663789 | 0.001101 | 6.99E-09 | TRUE | 6.91E-07 | -- | 0.507816 | 1 | 0.002622 | 1 | 1.28E-07 | 1 |
| 14-77850817-ATTTTTT | A | | | | | | | | | 0.003443 | 7.88E-18 | TRUE | 7.88E-18 | ?+ | 0.020362 | 1 | 0.000922 | 1 | 2.37E-13 | 1 |
| 14-78832014-A-G | G | 0.000195 | 0.095015 | | | 0.09501 | +? | 0.000191 | 0.307203 | 0.001431 | 2.46E-27 | TRUE | 1.01E-22 | +- | 1.02E-05 | 1 | 0.001125 | 1 | 3.59E-20 | 1 |
| 14-81923555-AATTTT- | A | 0.000823 | 0.846851 | | | 0.8469 | -? | 0.001444 | 0.818303 | 0.005684 | 3.23E-13 | TRUE | 1.22E-09 | -+ | 0.002561 | 1 | 0.882596 | 1 | 1.43E-10 | 1 |
| 14-88557948-T-C | C | | | | | | | 0.000457 | 0.167637 | 0.001521 | 2.71E-11 | TRUE | 1.29E-10 | -- | 0.000569 | 1 | 0.030214 | 1 | 1.02E-07 | 1 |
| 14-88557971-G-A | A | | | | | | | | | 0.001407 | 5.47E-10 | TRUE | 5.47E-10 | ?+ | 0.002184 | 1 | 0.222598 | 1 | 1.52E-07 | 1 |
| 14-91460172-C-CCT | CCT | | | | | | | | | 0.00077 | 1.45E-08 | TRUE | 1.45E-08 | ?+ | 0.02721 | 1 | 0.020138 | 1 | 6.58E-07 | 1 |
| 14-91460178-ATG-A | A | | | | | | | | | 0.000453 | 3.83E-08 | TRUE | 3.83E-08 | ?+ | 0.328104 | 1 | 0.005717 | 1 | 5.84E-07 | 1 |
| 14-91507096-ATCAC-/ | A | | | | | | | | | 0.000678 | 8.29E-15 | TRUE | 8.29E-15 | ?+ | 0.738343 | -1 | 0.083282 | 1 | 8.2E-15 | 1 |
| 14-92741777-G-T | T | 9.75E-05 | 0.518888 | | | 0.5189 | -? | | | 0.000498 | 4.28E-14 | TRUE | 4.28E-14 | ?+ | 0.003966 | 1 | 4.29E-06 | 1 | 3.22E-09 | 1 |
| 14-92804358-G-GC | GC | | | | | | | | | 0.000272 | 5.19E-12 | TRUE | 5.19E-12 | ?- | 0.497501 | -1 | 0.015069 | 1 | 3.33E-11 | 1 |
| 14-92804364-C-CAAC/ | CAACA | | | | | | | | | 0.000125 | 5.16E-09 | TRUE | 5.16E-09 | ?+ | | | | | 1.5E-08 | 1 |
| 14-92804368-CTT-C | C | | | | | | | | | 0.000174 | 3.14E-08 | TRUE | 3.14E-08 | ?- | | | | | 3.33E-07 | 1 |
| 14-100301031-C-A | A | 7.81E-05 | 0.379235 | | | 0.3792 | -? | 0.000175 | 0.70354 | 0.001075 | 2.21E-11 | TRUE | 2.83E-10 | ++ | 0.056324 | 1 | 0.02244 | 1 | 7.13E-10 | 1 |
| 14-100546852-T-C | C | 0.430853 | 0.017668 | 0.434 | 0.237412 | 0.0229 | +- | | | 0.602656 | 1.55E-08 | FALSE | 1.56E-08 | ?- | 0.948919 | -1 | 0.339772 | 1 | 3.62E-05 | 1 |
| 14-102136332-G-GCT | GCT | | | | | | | | | 0.000266 | 2.2E-09 | TRUE | 2.2E-09 | ?- | 0.451081 | 1 | 0.015372 | 1 | 1.38E-08 | 1 |
| 14-102541608-A-AG | AG | 0.000176 | 0.186308 | | | 0.1863 | +? | | | 0.001182 | 5.04E-15 | TRUE | 5.04E-15 | ?- | 0.000111 | 1 | 0.017268 | 1 | 2.52E-11 | 1 |
| 14-102809721-G-T | T | 0.000235 | 0.516701 | | | 0.5167 | +? | 0.00172 | 0.674686 | 0.001789 | 4.07E-13 | TRUE | 2.03E-07 | ++ | 0.555547 | 1 | 0.084271 | 1 | 9.54E-12 | 1 |
| 14-103272252-CCT-C | C | | | | | | | | | 0.0004 | 5.36E-10 | TRUE | 5.36E-10 | ?- | 0.009493 | 1 | 0.148231 | 1 | 3.91E-08 | 1 |
| 14-103601925-G-GGT | GGT | 0.001505 | 0.281685 | | | 0.2817 | +? | | | 0.003402 | 2.12E-11 | TRUE | 2.12E-11 | ?- | 0.049247 | 1 | 0.109974 | 1 | 8.28E-11 | 1 |
| 14-103601927-CAG-C | C | | | | | | | | | 0.003848 | 1.52E-11 | TRUE | 1.52E-11 | ?- | 0.051483 | 1 | 0.599374 | 1 | 5.11E-12 | 1 |
| 14-103655446-G-GCA | GCA | 0.006838 | 0.218776 | | | 0.2188 | +? | | | 0.0026 | 1.67E-12 | TRUE | 1.67E-12 | ?- | 0.009545 | 1 | 0.101151 | 1 | 5.04E-10 | 1 |
| 14-104209450-C-G | G | | | | | | | | | 0.000462 | 1.32E-09 | TRUE | 1.32E-09 | ?- | 0.159112 | 1 | 0.050255 | 1 | 7.99E-08 | 1 |
| 14-104720429-GGA-G | G | | | | | | | | | 0.231388 | 4.63E-11 | FALSE | 4.63E-11 | ?- | 0.000398 | -1 | 0.669163 | -1 | 3.28E-09 | -1 |
| 14-104847993-G-T | T | | | | | | | 0.00015 | 0.272319 | 0.00125 | 4.92E-16 | TRUE | 1.93E-13 | -+ | 0.016831 | 1 | 0.800469 | -1 | 7.09E-15 | 1 |
| 14-104945575-C-A | A | 0.0008 | 0.643603 | | | 0.6436 | -? | 0.004398 | 0.437536 | 0.002057 | 3.27E-10 | TRUE | 0.005798 | -+ | 0.005851 | 1 | 0.264707 | 1 | 1.82E-09 | 1 |
| 14-104951987-T-G | G | | | | | | | 0.000132 | 0.333787 | 0.001347 | 4.41E-08 | TRUE | 3.79E-08 | -- | 0.940826 | -1 | 0.15971 | 1 | 1.43E-08 | 1 |
| 14-105296067-C-CAA | CAA | | | | | | | | | 0.00144 | 1.48E-16 | TRUE | 1.49E-16 | ?+ | 0.004449 | 1 | 0.019943 | 1 | 1.03E-13 | 1 |
| 14-105296075-T-C | C | | | | | | | | | 0.997488 | 6.77E-13 | TRUE | 6.77E-13 | ?+ | 0.001142 | -1 | 0.01261 | -1 | 1.61E-09 | -1 |
| 14-105623618-T-A | A | | | | | | | | | 0.770314 | 2.57E-19 | FALSE | 2.57E-19 | ?- | 0.079312 | -1 | 0.890691 | 1 | 3.45E-22 | -1 |
| 14-105650853-CATGG | C | | | | | | | | | 0.013519 | 1.21E-08 | TRUE | 1.21E-08 | ?- | 0.077085 | 1 | 0.717333 | 1 | 4.63E-08 | 1 |
| 14-105709520-C-A | A | | | | | | | | | 0.53158 | 3.4E-08 | FALSE | 3.4E-08 | ?+ | 0.068547 | 1 | 0.538338 | 1 | 0.000348 | 1 |
| 14-105744716-G-A | A | | | | | | | | | 0.847658 | 3.14E-09 | FALSE | 3.14E-09 | ?- | 0.576152 | -1 | 0.873797 | -1 | 5.14E-12 | -1 |
| 14-105859763-C-T | T | | | | | | | | | 0.064182 | 6.47E-25 | FALSE | 6.48E-25 | ?+ | 0.091021 | 1 | 0.566115 | 1 | 1.42E-27 | 1 |
| 14-106708795-C-T | T | | | | | | | | | 0.02806 | 1.33E-10 | FALSE | 1.33E-10 | ?+ | 0.300977 | 1 | 0.274525 | 1 | 2.67E-11 | 1 |
| 14-74333212-G-A | A | | | | | | | 0.000282 | 0.00637 | 0.000591 | 1.31E-06 | TRUE | 4.97E-08 | ++ | 0.005218 | 1 | 0.002526 | 1 | 0.001738 | 1 |
| 14-102272953-C-CCT | CCT | | | | | | | 9.4E-06 | 0.222708 | 0.000115 | 8.19E-08 | TRUE | 3.84E-08 | ++ | | | | | 4.87E-06 | 1 |
| 14-29947961-G-A | A | | | | | | | 1.88E-05 | 0.449906 | | | FALSE | 4.1E-242 | -? | | | | | | |
| 15-73168712-C-T | T | 0.000779 | 8.84E-09 | | | 8.839E-09 | +? | 9.4E-06 | 0.338221 | 0.000127 | 0.955528 | TRUE | 0.6662 | +- | | | 0.847933 | -1 | | |
| 15-20341944-T-A | A | | | | | | | | | 0.780738 | 2.53E-16 | FALSE | 2.53E-16 | ?+ | 0.046281 | 1 | 0.528438 | -1 | 6.05E-17 | 1 |
| 15-20341946-T-C | C | | | | | | | | | 0.821078 | 9.23E-21 | FALSE | 9.23E-21 | ?- | 0.008702 | 1 | 0.48269 | 1 | 2.41E-19 | 1 |
| 15-21323221-A-ACAA/ | ACAAAAGCACCCTGGGAAGCCATAATTAATGGGAATTCTTTGGAATACCCAACCTAAAGCAAAAGTTCACATGCCT | | | | | | | | | 0.703952 | 1.17E-11 | FALSE | 1.17E-11 | ?+ | 0.004929 | -1 | 0.006203 | -1 | 3.84E-07 | -1 |
| 15-22543717-CT-C | C | | | | | | | | | 0.97686 | 3.49E-30 | FALSE | 3.49E-30 | ?+ | 0.000437 | -1 | 0.010377 | -1 | 7.11E-24 | -1 |
| 15-22986557-C-A | A | | | | | | | 0.000473 | 0.922051 | 0.00072 | 3.72E-11 | TRUE | 1.89E-07 | ++ | 0.007831 | 1 | 0.002498 | 1 | 2.06E-08 | 1 |
| 15-23066619-CTTTTTT | C | | | | | | | | | 0.031759 | 1.4E-09 | FALSE | 1.4E-09 | ?- | 0.178331 | 1 | 0.810401 | 1 | 5.62E-08 | 1 |
| 15-23448602-CAT-C | C | | | | | | | | | 0.001338 | 2.14E-09 | TRUE | 2.14E-09 | ?- | 0.071374 | 1 | 0.028746 | 1 | 3.2E-08 | 1 |
| 15-23448605-G-GCC | GCC | | | | | | | | | 0.001294 | 5.06E-10 | TRUE | 5.07E-10 | ?- | 0.011497 | 1 | 0.011304 | 1 | 1.48E-08 | 1 |
| 15-23448633-A-AG | AG | | | | | | | | | 0.001104 | 1.4E-11 | TRUE | 1.4E-11 | ?- | 0.165744 | 1 | 0.995575 | 1 | 5.84E-12 | 1 |
| 15-23838964-CCT-C | C | | | | | | | | | 0.000811 | 2.08E-11 | TRUE | 2.08E-11 | ?- | 0.153638 | 1 | 0.001839 | 1 | 9.1E-09 | 1 |
| 15-24330166-G-C | C | | | | | | | | | 0.174424 | 4.49E-09 | FALSE | 4.49E-09 | ?- | 0.953111 | 1 | 0.935655 | 1 | 3.27E-11 | -1 |
| 15-24330166-G-T | T | | | | | | | | | 0.819782 | 1.38E-08 | FALSE | 1.39E-08 | ?+ | 0.580282 | -1 | 0.892979 | -1 | 3.23E-11 | 1 |
| 15-25006938-G-A | A | 7.8E-05 | 0.405552 | | | 0.4056 | -? | 5.32E-05 | 0.163188 | 0.000361 | 2.01E-10 | TRUE | 1.12E-10 | ++ | 0.001259 | 1 | 0.398966 | 1 | 1.13E-08 | 1 |
| 15-25131879-T-A | A | | | | | | | | | 0.073142 | 3.62E-09 | FALSE | 3.62E-09 | ?+ | 0.139239 | 1 | 0.905058 | 1 | 1.65E-09 | 1 |
| 15-25293267-T-A | A | 0.000549 | 0.141538 | | | 0.1415 | -? | 0.00259 | 0.877751 | 0.000551 | 1.5E-08 | TRUE | 0.01218 | ++ | 0.045242 | 1 | 0.901368 | -1 | 3.85E-08 | 1 |
| 15-25293273-A-C | C | 0.00053 | 0.157277 | | | 0.1573 | +? | | | 0.000735 | 4.57E-08 | TRUE | 4.57E-08 | ?- | 0.00057 | 1 | 0.78499 | -1 | 6.66E-06 | 1 |
| 15-28367284-ATTTTT- | A | | | | | | | | | 0.711893 | 4.99E-08 | TRUE | 4.99E-08 | ?- | 0.156996 | -1 | 0.339295 | -1 | 5.88E-07 | -1 |
| 15-28401117-G-A | A | | | | | | | | | 0.920044 | 9.08E-18 | FALSE | 9.08E-18 | ?- | 0.001178 | -1 | 0.042775 | -1 | 9E-15 | -1 |
| 15-28784698-T-C | C | 0.052848 | 0.600978 | 0.019 | 0.441792 | 0.5895 | ++ | | | 0.05733 | 6.12E-09 | TRUE | 6.12E-09 | ?- | 6.21E-05 | 1 | 0.864654 | 1 | 0.124655 | 1 |

| | | | | | | | | | | | | | | | | | | | | |
|---|---|---|---|---|---|---|---|---|---|---|---|---|---|---|---|---|---|---|---|---|
| 15-29739562-G-A | A | | | | | | | | | 0.00169 | 2.4E-17 | TRUE | 2.4E-17 | ?+ | 0.007404 | 1 | 0.030887 | 1 | 5.57E-15 | 1 |
| 15-30132891-C-CA | CA | | | | | | | | | 0.911833 | 2.75E-25 | FALSE | 2.75E-25 | ?- | 0.063671 | -1 | 0.564994 | -1 | 2.56E-24 | -1 |
| 15-30173080-G-GGT | GGT | | | | | | | | | 0.973474 | 4.34E-14 | FALSE | 4.34E-14 | ?+ | 0.525482 | 1 | 0.760563 | 1 | 2.38E-18 | -1 |
| 15-30689377-TAATC-T | T | | | | | | | | | 0.621682 | 1.08E-08 | FALSE | 1.08E-08 | ?- | 0.025429 | -1 | 0.001585 | -1 | 3.24E-05 | -1 |
| 15-30689423-A-ATGTA | ATGTAT | | | | | | | | | 0.656965 | 2.86E-10 | FALSE | 2.86E-10 | ?+ | 0.030851 | -1 | 0.000355 | -1 | 3.84E-06 | -1 |
| 15-32216534-TA-T | T | | | | | | | | | 0.773197 | 1.17E-09 | FALSE | 1.17E-09 | ?- | 2.75E-05 | -1 | 0.821664 | 1 | 2.54E-07 | -1 |
| 15-32275965-C-T | T | | | | | | | | | 0.99189 | 3.15E-10 | TRUE | 3.15E-10 | ?- | 0.014943 | -1 | 0.001926 | -1 | 8.19E-08 | -1 |
| 15-34175801-G-T | T | | | | | | | 0.000473 | 0.541198 | 0.000732 | 5.16E-09 | TRUE | 8.35E-07 | ++ | 0.001253 | 1 | 0.238201 | 1 | 2.37E-07 | 1 |
| 15-34175843-T-TAAGA | TAAGAA | | | | | | | | | 0.001602 | 6.23E-14 | TRUE | 6.23E-14 | ?- | 0.053797 | 1 | 0.005272 | 1 | 5.46E-12 | 1 |
| 15-40502504-TCA-T | T | 0.000156 | 0.776153 | | | 0.7762 | -? | | | 0.002036 | 2.07E-09 | TRUE | 2.07E-09 | ?+ | 0.56084 | 1 | 0.879896 | -1 | 6.93E-10 | 1 |
| 15-40646627-G-C | C | | | | | | | | | 0.000133 | 1.45E-09 | TRUE | 1.45E-09 | ?+ | | | | | 4.68E-08 | 1 |
| 15-41344055-T-TA | TA | | | | | | | | | 0.000689 | 9.82E-14 | TRUE | 9.82E-14 | ?- | 0.132021 | 1 | 0.004048 | 1 | 3.3E-11 | 1 |
| 15-41378616-C-CAA | CAA | | | | | | | | | 0.000388 | 2.94E-09 | TRUE | 2.94E-09 | ?+ | 0.0041 | 1 | 0.740979 | 1 | 4.55E-08 | 1 |
| 15-41525481-GGCAT-G | G | | | | | | | | | 0.001995 | 2.12E-11 | TRUE | 2.12E-11 | ?+ | 0.000635 | 1 | 0.125527 | 1 | 6.68E-09 | 1 |
| 15-43485576-C-A | A | | | | | | | | | 0.016206 | 3.12E-08 | FALSE | 3.12E-08 | ?- | 0.681864 | 1 | 0.001182 | -1 | 1.66E-06 | -1 |
| 15-43723919-G-T | T | 8.05E-05 | 0.180289 | | | 0.1803 | +? | | | 0.00071 | 8.28E-09 | TRUE | 8.28E-09 | ?+ | 0.02765 | 1 | 0.893911 | -1 | 8.96E-08 | 1 |
| 15-44183022-TCTC-T | T | 0.000352 | 0.244713 | | | 0.2447 | +? | | | 0.000685 | 3.65E-12 | TRUE | 3.65E-12 | ?+ | 0.357061 | 1 | 0.912359 | -1 | 9.32E-12 | 1 |
| 15-44777318-G-T | T | 0.00049 | 0.862823 | | | 0.8628 | +? | | | 0.001234 | 4.41E-08 | TRUE | 4.41E-08 | ?+ | 0.045657 | 1 | 0.785764 | 1 | 3.79E-07 | 1 |
| 15-44777319-T-TG | TG | 0.000489 | 0.855923 | | | 0.8559 | -? | | | 0.001626 | 2.62E-08 | TRUE | 2.62E-08 | ?- | 0.029876 | 1 | 0.675058 | 1 | 4.86E-08 | 1 |
| 15-49117476-G-T | T | | | | | | | 0.000592 | 0.746148 | 0.000595 | 8.74E-09 | TRUE | 1.2E-05 | ++ | 0.022943 | 1 | 0.191306 | 1 | 6.96E-08 | 1 |
| 15-52015119-G-A | A | 0.000354 | 0.774191 | | | 0.7742 | +? | 0.000861 | 0.216002 | 0.000608 | 6.87E-11 | TRUE | 0.001096 | -+ | 0.000644 | 1 | 0.030551 | 1 | 1.12E-07 | 1 |
| 15-52015126-G-A | A | | | | | | | | | 0.000397 | 1.01E-10 | TRUE | 1.01E-10 | ?+ | 0.000427 | 1 | 0.00569 | 1 | 9.6E-07 | 1 |
| 15-52427442-C-G | G | | | 0.173 | 0.633491 | 0.6335 | ?+ | 0.1722 | 0.407304 | 0.179498 | 9.94E-13 | FALSE | 1.51E-06 | -- | 0.612445 | 1 | 0.155786 | 1 | 2.93E-14 | 1 |
| 15-55413666-A-G | G | 0.482472 | 0.245296 | 0.438 | 0.524783 | 0.2684 | +- | 0.434955 | 0.229368 | 0.481706 | 4.97E-08 | FALSE | 0.05111 | +- | 0.597691 | 1 | 0.839562 | 1 | 4.65E-08 | 1 |
| 15-55441294-A-T | T | | | | | | | | | 0.000331 | 3.34E-09 | TRUE | 3.34E-09 | ?- | 0.396513 | 1 | 0.108478 | 1 | 1.12E-08 | 1 |
| 15-55641955-A-ACC | ACC | 0.00043 | 0.315665 | | | 0.3157 | -? | | | 0.000644 | 4.71E-12 | TRUE | 4.71E-12 | ?- | 0.008915 | 1 | 0.001883 | 1 | 3.33E-09 | 1 |
| 15-62898468-G-GCGC | GCGC | | | | | | | 0.006005 | 0.551485 | 0.022043 | 7.14E-11 | FALSE | 1.7E-08 | -- | 0.360085 | 1 | 0.041936 | 1 | 5.6E-12 | 1 |
| 15-63272088-C-CTT | CTT | | | | | | | | | 0.000604 | 2.66E-09 | TRUE | 2.66E-09 | ?+ | 0.014483 | 1 | 0.880268 | -1 | 7.31E-09 | 1 |
| 15-64372438-C-T | T | 0.013186 | 0.155935 | | | 0.1559 | -? | | | 0.004783 | 8.06E-09 | TRUE | 8.06E-09 | ?+ | 0.864308 | -1 | 0.008698 | 1 | 1.6E-07 | 1 |
| 15-64541359-G-A | A | | | | | | | 0.007455 | 0.674627 | 0.001781 | 6.66E-09 | TRUE | 0.0656 | -+ | 0.012014 | 1 | 0.139943 | 1 | 7.64E-08 | 1 |
| 15-64731383-T-C | C | 0.000156 | 0.451108 | | | 0.4511 | +? | 0.001485 | 0.679829 | 0.000742 | 1.13E-10 | TRUE | 0.000126 | -- | 0.1596 | 1 | 0.010844 | 1 | 1.77E-08 | 1 |
| 15-65454698-TG-T | T | | | | | | | | | 0.000748 | 1.55E-11 | TRUE | 1.55E-11 | ?+ | 0.003069 | 1 | 0.269737 | 1 | 4.87E-10 | 1 |
| 15-65454700-T-TAACT | TAACTC | | | | | | | | | 0.00048 | 2.07E-08 | TRUE | 2.07E-08 | ?- | 0.074877 | 1 | 0.974026 | 1 | 2.55E-08 | 1 |
| 15-66380772-A-G | G | | | | | | | 0.005463 | 0.825091 | 0.001287 | 1.36E-11 | TRUE | 0.002261 | -- | 0.359165 | 1 | 0.005118 | 1 | 2.82E-09 | 1 |
| 15-66853149-T-C | C | 0.463551 | 0.535238 | | | 0.5352 | +? | 0.353181 | 0.263547 | 0.477052 | 1.27E-11 | FALSE | 0.004143 | +- | 0.025226 | 1 | 0.209844 | 1 | 3.78E-07 | 1 |
| 15-69587320-G-T | T | 0.000117 | 0.43176 | | | 0.4318 | +? | 0.001958 | 0.286118 | 0.000633 | 1.73E-10 | TRUE | 0.02364 | -+ | 0.073141 | 1 | 0.375574 | 1 | 4.87E-10 | 1 |
| 15-70684100-C-T | T | 0.000157 | 0.698897 | | | 0.6989 | -? | 0.00213 | 0.643348 | 0.001455 | 4.09E-10 | TRUE | 0.000846 | -+ | 0.001392 | 1 | 0.014155 | 1 | 4.15E-07 | 1 |
| 15-74757553-AT-A | A | 0.000332 | 0.796056 | | | 0.7961 | -? | | | 0.003805 | 1.38E-14 | TRUE | 1.38E-14 | ?+ | 0.02007 | 1 | 0.00088 | 1 | 1.49E-09 | 1 |
| 15-74921556-CCG-C | C | | | | | | | | | 0.001176 | 6.93E-09 | TRUE | 6.93E-09 | ?- | 0.128324 | 1 | 0.654201 | 1 | 4.03E-08 | 1 |
| 15-74953400-G-C | C | | | | | | | | | 0.000289 | 5.27E-10 | TRUE | 5.27E-10 | ?+ | 0.032615 | 1 | 0.044715 | 1 | 2.32E-08 | 1 |
| 15-75575726-TGG-T | T | 0.019406 | 0.276992 | | | 0.277 | +? | 0.000291 | 0.521318 | 0.006327 | 2.2E-10 | TRUE | 2.91E-10 | ++ | 0.697487 | -1 | 0.044074 | 1 | 3.65E-10 | 1 |
| 15-76148723-CT-C | C | | | | | | | | | 0.98622 | 2.81E-09 | TRUE | 2.81E-09 | ?+ | 0.003769 | -1 | 0.014834 | -1 | 3.22E-07 | -1 |
| 15-76317437-G-A | A | 0.000137 | 0.255318 | | | 0.2553 | -? | 0.000163 | 0.227395 | 0.000799 | 6.05E-09 | TRUE | 8.49E-09 | ++ | 3.11E-05 | 1 | 0.023654 | 1 | 3.05E-05 | 1 |
| 15-77699496-TCC-T | T | | | | | | | | | 0.001345 | 7.29E-12 | TRUE | 7.3E-12 | ?+ | 0.086308 | 1 | 0.021524 | 1 | 1.95E-10 | 1 |
| 15-78856334-T-TTA | TTA | | | | | | | | | 0.001311 | 3.64E-09 | TRUE | 3.64E-09 | ?- | 0.104042 | 1 | 0.995008 | 1 | 5.7E-09 | 1 |
| 15-79951126-C-CAGA | CAGAGCGG | | | | | | | | | 0.002801 | 2.86E-15 | TRUE | 2.86E-15 | ?+ | 0.12313 | 1 | 0.009688 | 1 | 1.53E-13 | 1 |
| 15-82515559-A-AAAA | AAAAAAAAG | | | | | | | | | 0.936104 | 3.02E-12 | FALSE | 3.02E-12 | ?+ | 0.699117 | -1 | 0.255735 | -1 | 1.91E-15 | -1 |
| 15-82515564-G-A | A | | | | | | | | | 0.95579 | 5.23E-25 | FALSE | 5.23E-25 | ?- | 0.003595 | -1 | 0.072232 | -1 | 2.64E-23 | -1 |
| 15-89020231-C-G | G | 0.315122 | 0.864032 | | | 0.864 | +? | | | 0.317236 | 3.95E-08 | TRUE | 3.95E-08 | ?- | 0.069891 | 1 | 0.477936 | -1 | 0.103246 | 1 |
| 15-90157089-G-GCA | GCA | | | | | | | | | 0.000229 | 4.57E-09 | TRUE | 4.57E-09 | ?- | 0.012269 | 1 | 0.10597 | 1 | 3.66E-07 | 1 |
| 15-90157090-T-TG | TG | | | | | | | | | 0.00017 | 2.48E-09 | TRUE | 2.48E-09 | ?- | | | 0.00784 | 1 | 2.52E-06 | 1 |
| 15-90157092-T-TA | TA | | | | | | | | | 0.000173 | 7.95E-10 | TRUE | 7.95E-10 | ?- | | | 0.043314 | 1 | 1.21E-06 | 1 |
| 15-90505467-C-CAA | CAA | | | | | | | 0.000244 | 0.362848 | 0.002337 | 8.49E-13 | TRUE | 1.98E-12 | ++ | 0.003075 | 1 | 0.134746 | 1 | 9.12E-11 | 1 |
| 15-92850975-AT-A | A | 0.000137 | 0.382052 | | | 0.3821 | -? | | | 0.000665 | 2.51E-08 | TRUE | 2.51E-08 | ?+ | 0.11562 | 1 | 0.033456 | 1 | 1.4E-06 | 1 |
| 15-99043874-T-C | C | | | | | | | | | 0.000255 | 1.03E-09 | TRUE | 1.03E-09 | ?- | 0.010417 | 1 | | | 1.28E-08 | 1 |
| 15-26057496-G-T | T | | | | | | | 1.57E-05 | 0.343817 | | | FALSE | 2.3E-305 | +? | | | | | | |
| 15-61803501-TACACA | TACACACACAC | | | | | | | 0.000103 | 0.426704 | | | FALSE | 7.3E-69 | +? | | | | | | |
| 16-85939758-G-A | A | 0.035624 | 1.05E-08 | 0.004 | 0.352542 | 9.917E-09 | -- | 0.000677 | 0.44429 | 0.021517 | 0.44352 | FALSE | 0.6347 | -+ | 0.067199 | -1 | 0.812218 | 1 | 0.797991 | -1 |
| 16-7791458-T-A | A | | | | | | | 2.19E-05 | 1E-75 | | | FALSE | 0.4484 | -? | | | | | | |

| | | | | | | | | | | | | | | | | | | | | |
|---|---|---|---|---|---|---|---|---|---|---|---|---|---|---|---|---|---|---|---|---|
| 16-199543-T-A | A | 0.000549 | 0.41813 | | | 0.4181 | +? | 0.000185 | 0.940299 | 0.001141 | 2.86E-11 | TRUE | 9.95E-10 | -+ | 0.015427 | 1 | 0.141112 | 1 | 1.27E-08 | 1 |
| 16-311114-G-GCT | GCT | | | | | | | | | 0.99922 | 7.89E-11 | TRUE | 7.89E-11 | ?+ | 0.006421 | -1 | 0.612817 | -1 | 7.23E-09 | -1 |
| 16-311117-G-GC | GC | | | | | | | 0.999555 | 0.430101 | 0.999138 | 8.8E-10 | TRUE | 5.71E-08 | ++ | 0.006525 | -1 | 0.042095 | -1 | 2.77E-07 | -1 |
| 16-422947-C-CAG | CAG | | | | | | | | | 0.000521 | 1.64E-08 | TRUE | 1.64E-08 | ?+ | 0.013233 | 1 | 0.003689 | 1 | 1.18E-05 | 1 |
| 16-619328-ATG-A | A | | | | | | | | | 0.001361 | 6.54E-09 | TRUE | 6.54E-09 | ?+ | 0.039913 | 1 | 0.014342 | 1 | 1.3E-05 | 1 |
| 16-754494-C-CA | CA | 9.75E-05 | 0.149841 | | | 0.1498 | +? | | | 0.000981 | 1.4E-12 | TRUE | 1.4E-12 | ?+ | 0.105496 | 1 | 0.1884 | 1 | 5.22E-12 | 1 |
| 16-948420-C-T | T | | | | | | | | | 0.513849 | 7.79E-09 | FALSE | 7.79E-09 | ?+ | 0.763765 | 1 | 0.181412 | 1 | 7.97E-06 | 1 |
| 16-948553-C-G | G | | | | | | | | | 0.504358 | 1.23E-08 | FALSE | 1.23E-08 | ?- | 0.916737 | -1 | 0.098805 | 1 | 3.83E-06 | 1 |
| 16-954892-T-C | C | | | | | | | | | 0.526237 | 2.42E-09 | FALSE | 2.42E-09 | ?- | 0.624456 | 1 | 0.108651 | 1 | 2.56E-06 | 1 |
| 16-1190872-GTC-G | G | | | | | | | | | 0.01273 | 2.39E-09 | TRUE | 2.39E-09 | ?+ | 0.261023 | 1 | 0.005174 | 1 | 3.73E-08 | 1 |
| 16-1258683-G-GC | GC | | | | | | | | | 0.861369 | 3.95E-12 | FALSE | 3.95E-12 | ?- | 0.400965 | 1 | 0.29494 | 1 | 1.75E-12 | 1 |
| 16-1478801-CCA-C | C | | | | | | | | | 0.000332 | 3.98E-08 | TRUE | 3.98E-08 | ?- | 0.122987 | 1 | 0.031953 | 1 | 1.05E-06 | 1 |
| 16-1569073-C-T | T | | | | | | | 0.112941 | 0.599786 | 0.214545 | 1.86E-08 | TRUE | 0.002108 | -+ | 0.582186 | 1 | 0.877915 | 1 | 0.000296 | 1 |
| 16-2099049-C-CT | CT | | | | | | | 0.000927 | 0.317753 | 0.001436 | 1.07E-09 | TRUE | 2.42E-07 | ++ | 0.346828 | 1 | 0.009391 | 1 | 2.81E-09 | 1 |
| 16-2099051-C-G | G | | | | | | | 0.000924 | 0.308084 | 0.001186 | 4.19E-09 | FALSE | 1.19E-06 | -- | 0.046457 | 1 | 0.006096 | 1 | 2.24E-07 | 1 |
| 16-2891973-CTT-C | C | | | | | | | | | 0.00262 | 5.19E-12 | TRUE | 5.19E-12 | ?- | 0.276944 | 1 | 0.001608 | 1 | 4.22E-10 | 1 |
| 16-2891980-TTGGG-T | T | | | | | | | | | 0.001968 | 1.13E-15 | TRUE | 1.13E-15 | ?+ | 0.017385 | 1 | 0.000106 | 1 | 3.41E-12 | 1 |
| 16-2891984-G-GCAAC | GCAAC | | | | | | | | | 0.001517 | 5.94E-12 | TRUE | 5.94E-12 | ?- | 0.278733 | 1 | 0.000103 | 1 | 2.53E-09 | 1 |
| 16-2983500-C-CA | CA | | | | | | | | | 0.001305 | 2.1E-09 | TRUE | 2.1E-09 | ?+ | 0.555817 | 1 | 0.75665 | -1 | 7.98E-10 | 1 |
| 16-3923860-C-G | G | | | | | | | | | 0.0004 | 1.14E-09 | TRUE | 1.14E-09 | ?- | 0.525274 | 1 | 0.05696 | 1 | 1.14E-07 | 1 |
| 16-4022490-C-A | A | | | | | | | 0.00597 | 0.530685 | 0.04202 | 5.84E-10 | FALSE | 1.35E-08 | -- | 0.068959 | -1 | 0.742222 | -1 | 6.66E-10 | -1 |
| 16-4590542-GA-G | G | | | | | | | | | 0.000503 | 3.9E-08 | TRUE | 3.9E-08 | ?+ | 0.001209 | 1 | 0.048725 | 1 | 9.78E-05 | 1 |
| 16-4771009-C-CAA | CAA | | | | | | | | | 0.000847 | 2.94E-09 | TRUE | 2.94E-09 | ?+ | 0.267445 | 1 | 0.73199 | 1 | 3.39E-08 | 1 |
| 16-5121705-C-A | A | | | | | | | | | 0.034671 | 3.21E-08 | FALSE | 3.21E-08 | ?- | 0.873413 | -1 | 0.536015 | -1 | 2.92E-08 | -1 |
| 16-8985095-A-G | G | 0.227656 | 0.96506 | 0.202 | 0.342329 | 0.9842 | +- | 0.176994 | 0.051076 | 0.265772 | 6.17E-09 | FALSE | 4.83E-07 | -- | 0.245645 | 1 | 0.31371 | 1 | 3.16E-06 | 1 |
| 16-10645333-A-G | G | | | | | | | | | 0.000467 | 1.13E-11 | TRUE | 1.13E-11 | ?- | 0.002772 | 1 | 0.022503 | 1 | 6.38E-10 | 1 |
| 16-10645335-A-C | C | | | | | | | | | 0.00035 | 1.74E-08 | TRUE | 1.74E-08 | ?- | 0.483426 | 1 | 0.080249 | 1 | 1.57E-08 | 1 |
| 16-10734878-G-A | A | | | | | | | | | 0.00089 | 2.94E-08 | TRUE | 2.94E-08 | ?+ | 0.234176 | 1 | 0.031331 | 1 | 1.13E-06 | 1 |
| 16-11232146-T-TGC | TGC | | | | | | | | | 0.000199 | 2.3E-08 | TRUE | 2.3E-08 | ?- | 0.01787 | 1 | | | 7.32E-05 | 1 |
| 16-11808833-C-CA | CA | | | | | | | 0.000698 | 0.502887 | 0.000557 | 5.26E-10 | TRUE | 3.63E-06 | ++ | 0.335524 | -1 | 1.25E-05 | 1 | 4.85E-07 | 1 |
| 16-14532898-G-GC | GC | | | | | | | | | 0.001843 | 2.54E-08 | TRUE | 2.54E-08 | ?- | 0.003925 | 1 | 0.008728 | 1 | 2.06E-05 | 1 |
| 16-14806559-CTT-C | C | | | | | | | | | 0.957306 | 2.33E-09 | FALSE | 2.33E-09 | ?+ | 0.011272 | -1 | 0.240239 | -1 | 7.32E-08 | -1 |
| 16-15014930-CTA-C | C | 0.450487 | 0.101762 | | | 0.1018 | -? | | | 0.913797 | 1.13E-14 | FALSE | 1.13E-14 | ?+ | 0.000773 | -1 | 0.012888 | -1 | 1.77E-09 | -1 |
| 16-15359814-GCACAC | G | | | | | | | | | 0.005612 | 5.39E-10 | TRUE | 5.39E-10 | ?+ | 0.003898 | 1 | 0.175526 | 1 | 2.08E-07 | 1 |
| 16-16171817-GGGAT- | G | | | | | | | 0.000542 | 0.078946 | 0.004542 | 4.83E-08 | TRUE | 1.77E-05 | -+ | 0.19858 | 1 | 0.067015 | 1 | 3.46E-07 | 1 |
| 16-16994683-C-T | T | 0.000604 | 0.219067 | | | 0.2191 | -? | 0.001939 | 0.557708 | 0.00237 | 1.4E-08 | TRUE | 0.000566 | -+ | 0.049239 | 1 | 0.00642 | 1 | 3.33E-06 | 1 |
| 16-17470922-C-CG | CG | | | | | | | | | 0.008585 | 6.73E-10 | FALSE | 6.73E-10 | ?+ | 0.014701 | 1 | 0.387851 | 1 | 1.16E-08 | 1 |
| 16-18158393-T-A | A | | | | | | | | | 0.000345 | 6.69E-11 | TRUE | 6.69E-11 | ?+ | 0.003103 | 1 | 0.681149 | 1 | 5.68E-09 | 1 |
| 16-20408737-GCA-G | G | 0.000156 | 0.953972 | | | 0.954 | -? | | | 0.002154 | 6.29E-16 | TRUE | 6.3E-16 | ?+ | 0.028463 | 1 | 0.036815 | 1 | 1.38E-12 | 1 |
| 16-21343238-C-CAAA | CAAA | | | | | | | | | 0.064277 | 1.63E-08 | TRUE | 1.63E-08 | ?+ | 0.363619 | 1 | 0.370224 | -1 | 1.22E-10 | 1 |
| 16-21894065-TTGAG-T | T | | | | | | | | | 0.000645 | 6.36E-12 | TRUE | 6.36E-12 | ?+ | 0.0082 | 1 | 0.907113 | 1 | 6.73E-11 | 1 |
| 16-23588279-A-G | G | | | | | | | 0.000536 | 0.468786 | 0.000876 | 1.38E-12 | TRUE | 1.5E-09 | -- | 0.008878 | 1 | 0.072714 | 1 | 8.39E-11 | 1 |
| 16-23736646-A-AAGT( | AAGTGT | | | | | | | | | 0.001289 | 6.36E-14 | TRUE | 6.36E-14 | ?- | 0.972939 | -1 | 0.002758 | 1 | 2.73E-13 | 1 |
| 16-23736647-GGCAC( | G | | | | | | | | | 0.000978 | 9.34E-12 | TRUE | 9.34E-12 | ?+ | 0.772209 | 1 | 0.011056 | 1 | 5.37E-11 | 1 |
| 16-24868596-C-A | A | | | | | | | | | 0.035381 | 5.81E-09 | FALSE | 5.82E-09 | ?+ | 0.407661 | 1 | 0.279689 | -1 | 1.98E-11 | 1 |
| 16-24905612-A-AGG | AGG | | | | | | | | | 0.000311 | 2.01E-09 | TRUE | 2.01E-09 | ?- | 0.293666 | 1 | 0.064844 | 1 | 5.66E-09 | 1 |
| 16-25154176-AT-A | A | | | | | | | 0.000392 | 0.457416 | 0.00208 | 6.24E-09 | TRUE | 3.51E-08 | ++ | 0.017776 | 1 | 0.048818 | 1 | 6.2E-06 | 1 |
| 16-28441940-C-CG | CG | | | | | | | | | 0.085089 | 3.59E-09 | FALSE | 3.59E-09 | ?- | 0.046927 | -1 | 0.046667 | -1 | 3.8E-07 | -1 |
| 16-28817462-CAT-C | C | | | | | | | | | 0.001251 | 5.59E-15 | TRUE | 5.59E-15 | ?- | 0.000431 | 1 | 0.014221 | 1 | 7.58E-12 | 1 |
| 16-28817467-G-GCA | GCA | | | | | | | | | 0.001086 | 2.65E-12 | TRUE | 2.65E-12 | ?- | 2.43E-05 | 1 | 0.417447 | 1 | 1.77E-10 | 1 |
| 16-28993307-A-G | G | | | | | | | | | 0.207352 | 6.38E-18 | TRUE | 6.38E-18 | ?- | 0.040963 | 1 | 0.162456 | -1 | 0.048072 | 1 |
| 16-29573680-A-G | G | 0.000626 | 0.820423 | | | 0.8204 | +? | 0.002199 | 0.926787 | 0.001237 | 2.79E-11 | TRUE | 0.000128 | -- | 0.087133 | 1 | 0.589948 | 1 | 9.9E-12 | 1 |
| 16-29786163-CA-C | C | | | | | | | 0.000229 | 0.061174 | 0.001001 | 1.67E-13 | TRUE | 1.02E-13 | -- | 0.003684 | 1 | 0.749694 | -1 | 8.43E-13 | 1 |
| 16-29831442-C-CCG | CCG | | | | | | | | | 0.00057 | 2.66E-15 | TRUE | 2.66E-15 | ?+ | 0.047902 | 1 | 0.446986 | 1 | 8.53E-15 | 1 |
| 16-30142456-A-AGG | AGG | | | | | | | | | 0.000463 | 2.92E-08 | TRUE | 2.92E-08 | ?- | 0.70908 | 1 | 0.040904 | 1 | 3.23E-08 | 1 |
| 16-30441816-CAG-C | C | | | | | | | | | 0.001065 | 4.88E-12 | TRUE | 4.88E-12 | ?- | 0.016508 | 1 | 0.002594 | 1 | 4.91E-09 | 1 |
| 16-30625405-C-CCT | CCT | | | | | | | | | 0.002105 | 6.82E-10 | TRUE | 6.82E-10 | ?+ | 0.988504 | -1 | 0.461195 | 1 | 4.01E-11 | 1 |
| 16-31041327-A-G | G | 0.000805 | 0.96805 | | | 0.968 | +? | 0.013281 | 0.586219 | 0.00183 | 1.35E-09 | TRUE | 0.1762 | +- | 0.080493 | 1 | 0.016306 | 1 | 2.98E-08 | 1 |
| 16-32421947-C-CA | CA | | | | | | | | | 0.561722 | 2.54E-14 | TRUE | 2.54E-14 | ?- | 0.098311 | -1 | 0.029678 | -1 | 4.15E-13 | -1 |

| | | | | | | | | | | | | | | | | | | | | |
|---|---|---|---|---|---|---|---|---|---|---|---|---|---|---|---|---|---|---|---|---|
| 16-32421947-CAAAAA | C | | | | | | | | | 0.110806 | 4.45E-09 | FALSE | 4.45E-09 | ?- | 0.801162 | 1 | 0.016675 | 1 | 1.38E-08 | 1 |
| 16-34249700-CTT-C | C | | | | | | | | | 0.000316 | 3.97E-08 | TRUE | 3.97E-08 | ?- | 0.558613 | -1 | 0.073923 | 1 | 8E-08 | 1 |
| 16-34260272-A-C | C | 0.001799 | 0.791756 | | | 0.7918 | +? | | | 0.000677 | 1.23E-12 | TRUE | 1.24E-12 | ?- | 9.06E-05 | 1 | 0.003727 | 1 | 1.76E-06 | 1 |
| 16-34284927-TTC-T | T | | | | | | | | | 0.002508 | 5.69E-12 | TRUE | 5.69E-12 | ?+ | 0.008752 | 1 | 0.005706 | 1 | 2.63E-09 | 1 |
| 16-34574822-T-TTGAA | TTGAAAACAAA | | | | | | | | | 0.004466 | 1.16E-08 | TRUE | 1.16E-08 | ?- | 0.000111 | 1 | 0.705102 | 1 | 2.32E-06 | 1 |
| 16-34594634-G-A | A | | | | | | | | | 0.956747 | 2.9E-10 | FALSE | 2.9E-10 | ?+ | 0.155436 | 1 | 0.419992 | 1 | 8.81E-09 | 1 |
| 16-34630340-AGTCCA | A | | | | | | | | | 0.005181 | 2.44E-46 | TRUE | 2.44E-46 | ?+ | 2.62E-05 | 1 | 0.001495 | 1 | 3.36E-36 | 1 |
| 16-35901414-A-G | G | | | | | | | | | 0.000616 | 1.7E-10 | TRUE | 1.7E-10 | ?- | 0.437189 | 1 | 0.005023 | 1 | 9.16E-09 | 1 |
| 16-35901425-A-G | G | | | | | | | | | 0.000595 | 1.67E-09 | TRUE | 1.67E-09 | ?- | 0.304327 | -1 | 0.020241 | 1 | 3.65E-08 | 1 |
| 16-35901442-G-A | A | | | | | | | 2.19E-05 | 0.319765 | 0.000361 | 1.73E-08 | TRUE | 1.09E-08 | ++ | 0.616225 | 1 | 0.181435 | 1 | 3.87E-08 | 1 |
| 16-35971942-G-T | T | | | 0.007 | 0.317311 | 0.3173 | ?- | 0.000507 | 0.253278 | 0.00041 | 2.59E-08 | TRUE | 4.33E-06 | ++ | 0.045848 | 1 | 0.848164 | 1 | 4.18E-08 | 1 |
| 16-36256874-T-TAC | TAC | | | | | | | | | 0.0018 | 6.11E-11 | TRUE | 6.11E-11 | ?- | 0.000444 | 1 | 0.039122 | 1 | 6.19E-08 | 1 |
| 16-46395598-C-T | T | | | | | | | | | 0.999387 | 7.84E-11 | TRUE | 7.84E-11 | ?- | 4.76E-05 | -1 | 0.000931 | -1 | 3.13E-06 | -1 |
| 16-46395604-C-A | A | | | | | | | | | 0.999279 | 1.49E-12 | TRUE | 1.49E-12 | ?- | 0.002655 | -1 | 0.044182 | -1 | 1E-09 | -1 |
| 16-46399335-C-A | A | | | | | | | | | 0.689888 | 1.07E-17 | FALSE | 1.07E-17 | ?- | 0.006733 | -1 | 0.400851 | -1 | 1.78E-15 | -1 |
| 16-46399335-C-G | G | | | | | | | | | 0.310112 | 1.07E-17 | FALSE | 1.07E-17 | ?- | 0.006733 | 1 | 0.400851 | 1 | 1.78E-15 | 1 |
| 16-46402078-C-G | G | | | | | | | | | 0.365908 | 1.05E-08 | FALSE | 1.05E-08 | ?- | 0.00198 | 1 | 0.155265 | 1 | 1.41E-07 | 1 |
| 16-48925550-C-T | T | | | | | | | 0.000758 | 0.029226 | 0.001545 | 1.04E-14 | TRUE | 2.82E-07 | -+ | 0.009239 | 1 | 0.001622 | 1 | 1.5E-11 | 1 |
| 16-50150533-ATT-A | A | | | | | | | | | 0.002107 | 2.23E-18 | TRUE | 2.23E-18 | ?+ | 0.259542 | 1 | 0.000943 | 1 | 1.53E-16 | 1 |
| 16-50803378-G-T | T | 0.000313 | 0.768639 | | | 0.7686 | -? | 0.00353 | 0.748552 | 0.000856 | 2.87E-11 | TRUE | 0.01102 | -+ | 0.055205 | 1 | 0.140634 | 1 | 5.25E-10 | 1 |
| 16-50803398-A-C | C | 0.00055 | 0.266318 | | | 0.2663 | +? | 0.005713 | 0.201125 | 0.00194 | 2.45E-13 | TRUE | 0.02381 | +- | 0.001753 | 1 | 0.225866 | 1 | 2.68E-09 | 1 |
| 16-57482602-T-C | C | 0.000548 | 0.902942 | | | 0.9029 | -? | 0.000232 | 0.008383 | 0.00176 | 4.2E-14 | TRUE | 1.06E-10 | +- | 0.139809 | 1 | 0.113111 | 1 | 3.48E-12 | 1 |
| 16-58440874-C-CAT | CAT | 0.000316 | 0.351766 | | | 0.3518 | -? | | | 0.000526 | 4.63E-08 | TRUE | 4.63E-08 | ?+ | 0.000434 | 1 | 0.945037 | -1 | 7.77E-07 | 1 |
| 16-67096756-C-CCT | CCT | | | | | | | | | 0.000437 | 7.72E-09 | TRUE | 7.72E-09 | ?+ | 0.017101 | 1 | 0.740387 | 1 | 1.06E-07 | 1 |
| 16-67606533-G-A | A | 0.03839 | 0.395179 | 0.017 | 0.129146 | 0.3788 | -- | | | 0.044646 | 1.67E-17 | TRUE | 1.67E-17 | ?+ | 0.017084 | 1 | 0.524222 | -1 | 2.73E-22 | 1 |
| 16-67610589-C-CCG | CCG | | | | | | | 4.7E-05 | 0.659432 | 0.000877 | 4.77E-09 | TRUE | 2.58E-08 | -+ | 0.000301 | 1 | 0.064676 | 1 | 5.17E-06 | 1 |
| 16-67849799-C-T | T | | | | | | | 7.52E-05 | 0.243695 | 0.000451 | 8.89E-11 | TRUE | 2.46E-08 | -+ | 0.022068 | 1 | 0.01978 | 1 | 9.88E-09 | 1 |
| 16-67849804-G-A | A | | | | | | | 0.000157 | 0.430058 | 0.000622 | 6.96E-14 | TRUE | 1.33E-12 | ++ | 0.057435 | 1 | 0.002104 | 1 | 5.32E-11 | 1 |
| 16-68401966-C-G | G | | | | | | | 0.001084 | 0.362994 | 0.000897 | 1.09E-10 | TRUE | 0.000364 | +- | 0.002303 | 1 | 0.265495 | 1 | 3.75E-09 | 1 |
| 16-69576084-C-CTT | CTT | | | | | | | | | 0.000937 | 6.03E-10 | TRUE | 6.03E-10 | ?+ | 0.014155 | 1 | 0.046955 | 1 | 1.49E-08 | 1 |
| 16-70218561-CA-C | C | | | | | | | | | 0.59332 | 1.05E-08 | TRUE | 1.05E-08 | ?- | 0.940338 | -1 | 0.008907 | 1 | 4.06E-07 | 1 |
| 16-70384477-G-C | C | 0.000158 | 0.608154 | | | 0.6082 | +? | 0.000107 | 0.011317 | 0.001474 | 9.52E-15 | TRUE | 3.74E-16 | ++ | 0.007189 | 1 | 0.062229 | 1 | 1.31E-13 | 1 |
| 16-73895079-AG-A | A | | | | | | | | | 0.000336 | 4.07E-08 | TRUE | 4.07E-08 | ?+ | 0.695458 | 1 | 0.000892 | 1 | 4.57E-07 | 1 |
| 16-74615312-C-CAG | CAG | | | | | | | | | 0.000971 | 1.57E-08 | TRUE | 1.57E-08 | ?+ | 0.12724 | 1 | 0.016486 | 1 | 1.54E-07 | 1 |
| 16-81105873-G-GGTC | GGTC | | | | | | | 0.000445 | 0.34038 | 0.003901 | 1.01E-20 | TRUE | 1.39E-19 | -- | 0.000843 | 1 | 0.161758 | 1 | 2.43E-16 | 1 |
| 16-82142519-C-G | G | | | | | | | 0.053745 | 0.263284 | 0.039786 | 1.96E-21 | FALSE | 4.37E-06 | +- | 0.010946 | 1 | 0.35394 | 1 | 4.27E-18 | 1 |
| 16-84114744-C-T | T | 0.332754 | 0.111813 | 0.36 | 0.277897 | 0.1334 | +- | 0.346864 | 0.175613 | 0.365593 | 1.25E-08 | FALSE | 0.03753 | -+ | 0.261639 | 1 | 0.853645 | 1 | 6.62E-10 | 1 |
| 16-86746743-G-A | A | | | | | | | 0.382962 | 0.464467 | 0.114293 | 2.89E-08 | FALSE | 0.002477 | ++ | 0.516677 | 1 | 0.046205 | 1 | 2.62E-07 | 1 |
| 16-87884404-G-C | C | | | | | | | 7.83E-05 | 0.959602 | 0.000536 | 6.13E-09 | TRUE | 4.86E-08 | ++ | 0.001183 | 1 | 0.303621 | 1 | 5.81E-07 | 1 |
| 16-89188362-CGG-C | C | | | | | | | | | 0.002694 | 6.86E-10 | TRUE | 6.86E-10 | ?- | 0.337475 | 1 | 0.20606 | 1 | 2.87E-09 | 1 |
| 16-89479635-C-A | A | | | | | | | | | 0.000621 | 1.89E-08 | TRUE | 1.89E-08 | ?+ | 0.227202 | 1 | 0.018775 | 1 | 8.9E-07 | 1 |
| 16-89650787-C-CAG | CAG | | | | | | | | | 0.000818 | 1.25E-10 | TRUE | 1.25E-10 | ?+ | 0.007256 | 1 | 0.023915 | 1 | 1.98E-08 | 1 |
| 16-89709021-C-CCCCC | CCCCG | | | | | | | | | 0.928024 | 7.36E-11 | FALSE | 7.36E-11 | ?- | 0.052993 | -1 | 0.163839 | -1 | 8.84E-12 | -1 |
| 16-89742386-AGAGGC | A | | | | | | | 0.323876 | 0.70973 | 0.427661 | 1.41E-09 | FALSE | 0.001738 | -+ | 0.011815 | 1 | 0.881968 | -1 | 7.75E-05 | 1 |
| 16-89831052-G-T | T | | | | | | | 0.011285 | 0.724107 | 0.003503 | 7.05E-26 | TRUE | 3.27E-07 | ++ | 9.91E-05 | 1 | 0.002094 | 1 | 1.94E-20 | 1 |
| 16-89831588-T-G | G | 0.000415 | 0.579572 | | | 0.5796 | -? | 0.001597 | 0.506263 | 0.002521 | 8.21E-14 | TRUE | 3.26E-07 | +- | 0.002469 | 1 | 0.035214 | 1 | 8.33E-11 | 1 |
| 16-89831598-T-C | C | 0.001028 | 0.431538 | | | 0.4315 | +? | 0.001293 | 0.743064 | 0.002135 | 7.68E-14 | TRUE | 6.23E-08 | +- | 0.004657 | 1 | 0.077568 | 1 | 1.21E-10 | 1 |
| 16-89831610-A-G | G | | | | | | | | | 0.002049 | 4.28E-10 | TRUE | 4.28E-10 | ?- | 0.155815 | 1 | 0.238998 | -1 | 1.08E-10 | 1 |
| 16-89879111-TTGTCC | T | | | | | | | | | 0.004391 | 1.98E-09 | TRUE | 1.98E-09 | ?+ | 0.113487 | 1 | 0.300345 | 1 | 2.29E-08 | 1 |
| 16-89908259-CCAGCT | C | | | | | | | | | 0.086822 | 1.14E-08 | FALSE | 1.14E-08 | ?+ | 0.115694 | -1 | 0.025306 | -1 | 4.41E-07 | -1 |
| 16-82658281-T-A | A | | | | | | | 0.000157 | 0.004692 | 0.000565 | 6.95E-08 | TRUE | 1.3E-09 | ++ | 0.850534 | 1 | 0.03274 | 1 | 2.89E-07 | 1 |
| 16-82658267-T-A | A | | | | | | | 0.000157 | 0.004692 | 0.000719 | 5.68E-08 | TRUE | 9.82E-10 | ++ | 0.016017 | 1 | 0.058094 | 1 | 2.28E-06 | 1 |
| 16-3448877-C-T | T | 0.000877 | 0.888365 | | | 0.8884 | -? | 9.71E-05 | 0.573237 | 0.000786 | 0.874336 | TRUE | 1.94E-16 | -+ | 0.869968 | -1 | 0.445611 | -1 | | |
| 16-85583635-G-A | A | | | | | | | 4.39E-05 | 0.51605 | 0.000143 | 0.977114 | TRUE | 4.48E-10 | -- | | | 0.745703 | 1 | | |
| 17-35209114-T-G | G | 0.000604 | 2.21E-08 | | | 2.208E-08 | -? | 9.4E-06 | 0.27679 | 0.000147 | 0.48017 | TRUE | 0.2605 | -- | | | 0.604291 | 1 | | |
| 17-114123-GA-G | G | | | | | | | | | 0.947795 | 2.92E-12 | FALSE | 2.92E-12 | ?- | 0.867969 | 1 | 0.062676 | -1 | 6.39E-13 | -1 |
| 17-397831-G-T | T | | | | | | | 0.000285 | 0.828572 | 0.002112 | 2.41E-18 | TRUE | 1.16E-15 | -+ | 0.00609 | 1 | 0.008353 | 1 | 1.32E-14 | 1 |
| 17-582381-C-T | T | | | 0.471 | 0.697445 | 0.6974 | ?+ | | | 0.56345 | 2.09E-08 | FALSE | 2.09E-08 | ?+ | 0.255799 | -1 | 0.953939 | 1 | 1.92E-07 | 1 |
| 17-783556-C-G | G | | | | | | | | | 0.000549 | 3.79E-15 | TRUE | 3.79E-15 | ?- | 0.002291 | 1 | 0.060944 | 1 | 2.95E-11 | 1 |

| | | | | | | | | | | | | | | | | | | | | |
|---|---|---|---|---|---|---|---|---|---|---|---|---|---|---|---|---|---|---|---|---|
| 17-839242-G-C | C | | | | | | | 1.57E-05 | 0.382387 | 0.00035 | 2.42E-09 | TRUE | 1.15E-08 | -+ | 0.05809 | 1 | 0.088556 | 1 | 2.93E-08 | 1 |
| 17-899026-T-C | C | 0.000119 | 0.23154 | | | 0.2315 | -? | 0.000448 | 0.549774 | 0.00392 | 3.25E-12 | FALSE | 1.37E-11 | -- | 0.282567 | 1 | 0.008255 | 1 | 6.53E-09 | 1 |
| 17-971686-GGTGA-G | G | | | | | | | | | 0.001006 | 5.4E-14 | TRUE | 5.4E-14 | ?+ | 6.57E-05 | 1 | 0.0334 | 1 | 1.48E-10 | 1 |
| 17-1014620-C-CCT | CCT | | | | | | | | | 0.00089 | 1.27E-09 | TRUE | 1.27E-09 | ?+ | 0.247256 | 1 | 0.489637 | 1 | 3.08E-09 | 1 |
| 17-1195651-C-A | A | 7.85E-05 | 0.874055 | | | 0.8741 | -? | 0.000724 | 0.116815 | 0.000909 | 1.97E-09 | TRUE | 5.88E-08 | ++ | 0.050471 | 1 | 0.260475 | 1 | 5.46E-09 | 1 |
| 17-1195697-A-G | G | 7.86E-05 | 0.680564 | | | 0.6806 | -? | 0.000545 | 0.809979 | 0.000247 | 1.24E-08 | TRUE | 0.000359 | -- | 0.334101 | 1 | 0.005264 | 1 | 1.02E-07 | 1 |
| 17-1203433-G-GGGG( | GGGGC | | | | | | | | | 0.544399 | 1.06E-18 | FALSE | 1.06E-18 | ?+ | 0.115661 | -1 | 0.206979 | -1 | 9.63E-19 | -1 |
| 17-1230579-G-A | A | 0.000371 | 0.600453 | | | 0.6005 | +? | 0.001046 | 0.409325 | 0.001405 | 2.9E-09 | TRUE | 1.05E-06 | ++ | 0.956475 | 1 | 0.00195 | 1 | 5.86E-08 | 1 |
| 17-1230592-C-A | A | | | | | | | 0.001096 | 0.293081 | 0.001721 | 1.01E-08 | TRUE | 7.41E-07 | ++ | 0.717082 | -1 | 0.246219 | 1 | 3.81E-08 | 1 |
| 17-1354830-T-TTTG | TTTG | | | | | | | | | 0.001782 | 1.56E-13 | TRUE | 1.56E-13 | ?- | 6.24E-05 | 1 | 0.049906 | 1 | 2.04E-09 | 1 |
| 17-1430883-C-T | T | | | | | | | | | 0.975036 | 5.38E-17 | TRUE | 5.38E-17 | ?- | 0.003799 | -1 | 0.203212 | -1 | 9.69E-16 | -1 |
| 17-1430884-G-A | A | 0.002113 | 0.022287 | 0.005 | 0.548506 | 0.02192 | -- | 0.00379 | 0.164919 | 0.003238 | 2.51E-13 | TRUE | 1.24E-08 | ++ | 8.5E-05 | 1 | 0.013336 | 1 | 1.16E-09 | 1 |
| 17-1596998-C-CCT | CCT | 0.000156 | 0.207327 | | | 0.2073 | -? | 8.46E-05 | 0.985869 | 0.001227 | 3.07E-14 | TRUE | 2.16E-13 | -+ | 0.004524 | 1 | 0.001844 | 1 | 2.94E-11 | 1 |
| 17-1758445-A-ACT | ACT | | | | | | | 0.000564 | 0.444551 | 0.004118 | 6.12E-28 | TRUE | 5.17E-23 | +- | 0.001097 | 1 | 0.002874 | 1 | 3.55E-23 | 1 |
| 17-1788527-C-CCT | CCT | | | | | | | | | 0.000219 | 4.13E-10 | TRUE | 4.13E-10 | ?+ | | | | | 1.88E-09 | 1 |
| 17-2584417-GGC-G | G | 0.000449 | 0.406756 | | | 0.4068 | +? | | | 0.002504 | 3.25E-11 | TRUE | 3.25E-11 | ?+ | 0.014225 | 1 | 5.11E-07 | 1 | 2.66E-05 | 1 |
| 17-2823905-C-CCT | CCT | | | | | | | | | 0.001443 | 1.7E-09 | TRUE | 1.7E-09 | ?+ | 0.00949 | 1 | 0.131721 | 1 | 3.85E-07 | 1 |
| 17-2846555-CAT-C | C | | | | | | | | | 0.000366 | 7.34E-09 | TRUE | 7.34E-09 | ?- | 0.019132 | 1 | 0.161818 | 1 | 4.5E-08 | 1 |
| 17-2846555-CATGA-C | C | | | | | | | | | 0.001721 | 1.41E-19 | TRUE | 1.41E-19 | ?- | 0.005107 | 1 | 0.011648 | 1 | 3.65E-16 | 1 |
| 17-2846560-G-GC | GC | | | | | | | | | 0.00071 | 1.75E-13 | TRUE | 1.75E-13 | ?- | 0.013194 | 1 | 0.013432 | 1 | 2.23E-10 | 1 |
| 17-2846563-A-G | G | | | | | | | 2.82E-05 | 0.18275 | 0.00079 | 3.66E-11 | TRUE | 1.48E-11 | -- | 0.015292 | 1 | 0.0208 | 1 | 8.44E-08 | 1 |
| 17-2846566-A-ACGT | ACGT | | | | | | | | | 0.00078 | 2.37E-09 | TRUE | 2.37E-09 | ?- | 0.465024 | 1 | 0.518165 | 1 | 4.7E-08 | 1 |
| 17-3112620-G-GGGT | GGGT | | | | | | | | | 0.002702 | 6.92E-10 | TRUE | 6.92E-10 | ?- | 6.3E-07 | 1 | 0.002241 | 1 | 0.000415 | 1 |
| 17-3765193-CA-C | C | | | | | | | | | 0.000836 | 7.54E-11 | TRUE | 7.54E-11 | ?- | 0.008301 | 1 | 0.258522 | 1 | 3.78E-09 | 1 |
| 17-3973712-G-GCTCC | GCTCCTGC | | | | | | | | | 0.195058 | 8.29E-12 | FALSE | 8.29E-12 | ?+ | 0.059409 | -1 | 0.68822 | -1 | 2.99E-12 | -1 |
| 17-4725163-AT-A | A | 0.000176 | 0.916085 | | | 0.9161 | -? | 0.000263 | 0.010035 | 0.000724 | 2.49E-08 | TRUE | 9.51E-05 | -+ | 0.091252 | 1 | 0.19507 | 1 | 1.19E-06 | 1 |
| 17-4820089-CAT-C | C | 0.001354 | 0.034397 | | | 0.0344 | +? | | | 0.003832 | 1.33E-17 | TRUE | 1.33E-17 | ?- | 0.000268 | 1 | 0.01985 | 1 | 7.11E-14 | 1 |
| 17-4923566-A-C | C | | | | | | | | | 0.000764 | 6.86E-13 | TRUE | 6.86E-13 | ?- | 0.002767 | 1 | 0.001436 | 1 | 2.68E-09 | 1 |
| 17-4993046-A-ATT | ATT | | | | | | | | | 0.000843 | 7.15E-13 | TRUE | 7.15E-13 | ?- | 0.003516 | 1 | 0.000385 | 1 | 9.38E-09 | 1 |
| 17-4995776-CA-C | C | | | | | | | | | 0.000108 | 3.53E-13 | TRUE | 3.53E-13 | ?- | | | | | 1.35E-09 | 1 |
| 17-5013837-C-A | A | | | | | | | 0.002797 | 0.983411 | 0.002077 | 1.01E-16 | TRUE | 2.3E-07 | -+ | 0.163802 | 1 | 0.882321 | -1 | 1.86E-17 | 1 |
| 17-5181517-T-C | C | | | | | | | 0.002885 | 0.05207 | 0.000281 | 2.15E-09 | TRUE | 0.9104 | +- | 0.039892 | 1 | 0.009637 | 1 | 2.43E-06 | 1 |
| 17-5265867-C-CCT | CCT | | | | | | | | | 0.003333 | 2.96E-10 | TRUE | 2.96E-10 | ?+ | 0.010512 | 1 | 0.562141 | 1 | 4.13E-09 | 1 |
| 17-5439856-C-CCTT | CCTT | | | | | | | | | 0.000621 | 2.1E-10 | TRUE | 2.1E-10 | ?+ | 0.01828 | 1 | 0.016184 | 1 | 2.96E-08 | 1 |
| 17-7055430-C-CCT | CCT | | | | | | | | | 0.001594 | 3.81E-19 | TRUE | 3.81E-19 | ?+ | 0.270672 | 1 | 0.124743 | 1 | 1.29E-17 | 1 |
| 17-7176529-A-ACACA | ACACACACT | | | | | | | | | 0.16078 | 2.23E-08 | FALSE | 2.23E-08 | ?- | 0.051073 | 1 | 0.290816 | 1 | 0.005716 | 1 |
| 17-7268492-T-G | G | | | | | | | 0.000767 | 0.661078 | 0.001669 | 2.21E-13 | TRUE | 1.02E-09 | -- | 0.021619 | 1 | 0.171964 | 1 | 2.35E-11 | 1 |
| 17-7681502-A-AT | AT | 0.000957 | 0.91975 | | | 0.9197 | +? | 0.001124 | 0.905228 | 0.004422 | 1.43E-19 | TRUE | 2.63E-15 | -- | 0.02921 | 1 | 0.15388 | 1 | 5.41E-18 | 1 |
| 17-7681961-T-TTTTTT( | TTTTTG | 0.001765 | 0.20768 | | | 0.2077 | +? | | | 0.002858 | 5.25E-09 | TRUE | 5.25E-09 | ?- | 0.056483 | 1 | 0.017271 | 1 | 1.32E-06 | 1 |
| 17-8131304-G-T | T | 0.001193 | 0.353562 | | | 0.3536 | -? | 0.002133 | 0.527806 | 0.00265 | 1.31E-11 | TRUE | 9.99E-07 | ++ | 0.071788 | 1 | 0.093711 | 1 | 2.35E-10 | 1 |
| 17-8166150-CG-C | C | | | | | | | | | 0.064127 | 3.32E-09 | FALSE | 3.32E-09 | ?- | 0.577809 | 1 | 0.778798 | 1 | 2.47E-09 | 1 |
| 17-8853714-G-A | A | 0.072168 | 0.835342 | 0.029 | 0.742987 | 0.8296 | -- | | | 0.079081 | 1.03E-08 | TRUE | 1.03E-08 | ?+ | 0.053394 | 1 | 0.394355 | -1 | 0.317307 | 1 |
| 17-9113990-C-CT | CT | | | | | | | | | 0.000542 | 2.47E-08 | TRUE | 2.47E-08 | ?+ | 0.011485 | 1 | 0.198512 | 1 | 3.02E-07 | 1 |
| 17-9688444-A-G | G | | | 0.367 | 0.126951 | 0.127 | ?- | | | 0.508079 | 4.45E-09 | FALSE | 4.45E-09 | ?- | 0.777512 | 1 | 0.249733 | -1 | 4.71E-07 | 1 |
| 17-15799943-A-G | G | | | | | | | | | 0.507687 | 4.36E-12 | FALSE | 4.36E-12 | ?- | 0.948555 | 1 | 0.253657 | 1 | 8.63E-09 | 1 |
| 17-16371521-G-C | C | | | | | | | 0.000993 | 0.99688 | 0.001588 | 4.82E-16 | TRUE | 4.77E-10 | -+ | 0.005803 | 1 | 2.09E-05 | 1 | 1.69E-11 | 1 |
| 17-16382000-G-A | A | 0.002859 | 0.86458 | | | 0.8646 | +? | 0.004085 | 0.190867 | 0.001337 | 4.17E-09 | TRUE | 0.000127 | ++ | 0.002809 | 1 | 0.88281 | 1 | 2.23E-08 | 1 |
| 17-16382525-C-T | T | 0.001025 | 0.264439 | | | 0.2644 | -? | 0.006089 | 0.583322 | 0.002652 | 3.54E-11 | TRUE | 0.006312 | -+ | 0.052301 | 1 | 0.748194 | -1 | 3.49E-11 | 1 |
| 17-16451843-C-CAA | CAA | | | | | | | | | 0.00034 | 3.69E-08 | TRUE | 3.69E-08 | ?+ | 5.22E-05 | 1 | 0.050544 | 1 | 5.94E-05 | 1 |
| 17-17001857-G-GCA | GCA | | | | | | | | | 0.000527 | 1.87E-10 | TRUE | 1.87E-10 | ?- | 0.003484 | 1 | 0.001115 | 1 | 6.27E-07 | 1 |
| 17-17002395-G-A | A | | | | | | | 0.000141 | 0.017196 | 0.000952 | 1.09E-17 | TRUE | 1.67E-18 | ++ | 0.024413 | 1 | 0.032718 | 1 | 1.19E-14 | 1 |
| 17-18080932-A-T | T | | | | | | | 0.00041 | 0.578681 | 0.000942 | 7.15E-13 | TRUE | 3.1E-10 | -- | 0.093823 | 1 | 0.062332 | 1 | 2.17E-11 | 1 |
| 17-18084733-C-G | G | | | | | | | 0.000116 | 0.217919 | 0.001021 | 2.59E-12 | TRUE | 4.84E-10 | +- | 0.000558 | 1 | 0.835439 | 1 | 8.37E-11 | 1 |
| 17-18762161-A-T | T | | | | | | | | | 0.000494 | 4.8E-09 | TRUE | 4.8E-09 | ?- | 0.050004 | 1 | 0.147471 | 1 | 3.64E-07 | 1 |
| 17-19025179-TTTTTC- | T | | | | | | | 0.000442 | 0.952708 | 0.004792 | 4.21E-15 | TRUE | 1.48E-13 | ++ | 0.056564 | 1 | 0.025214 | 1 | 7.96E-14 | 1 |
| 17-21260860-A-AC | AC | | | | | | | | | 0.001854 | 1.14E-15 | TRUE | 1.14E-15 | ?- | 0.000319 | 1 | 0.000516 | 1 | 1.65E-10 | 1 |
| 17-21684971-C-CAGG | CAGG | 0.115286 | 0.637837 | | | 0.6378 | -? | | | 0.005678 | 6.51E-09 | TRUE | 6.51E-09 | ?+ | 0.015873 | 1 | 0.225661 | 1 | 5.45E-07 | 1 |
| 17-21887270-G-GAAT | GAATTGAATGGAATCGAATGGAA | | | | | | | | | 0.086978 | 9.8E-12 | FALSE | 9.8E-12 | ?+ | 0.029408 | -1 | 0.003464 | -1 | 8.65E-09 | -1 |
| 17-21887283-T-TCGA/ | TCGAAAGGAATGGAA | | | | | | | | | 0.06839 | 2.38E-11 | FALSE | 2.38E-11 | ?+ | 0.021767 | -1 | 0.01299 | -1 | 6.97E-09 | -1 |

| | | | | | | | | | | | | | | | | | | | | |
|---|---|---|---|---|---|---|---|---|---|---|---|---|---|---|---|---|---|---|---|---|
| 17-21895277-G-GCAT | GCATGGAC | | | | | | | | | 0.164004 | 3.84E-13 | FALSE | 3.84E-13 | ?+ | 0.076521 | -1 | 0.008252 | -1 | 7.01E-11 | -1 |
| 17-21895278-AGT-A | A | | | | | | | | | 0.157447 | 2.6E-14 | FALSE | 2.6E-14 | ?- | 0.068193 | -1 | 0.00197 | -1 | 6.5E-12 | -1 |
| 17-21920958-A-AT | AT | | | | | | | | | 0.044555 | 7E-12 | FALSE | 7E-12 | ?+ | 0.767354 | -1 | 0.002672 | -1 | 1.1E-09 | -1 |
| 17-22064831-C-CAAA | CAAAAAAAAAAAAA | | | | | | | | | 0.028043 | 1.6E-09 | TRUE | 1.6E-09 | ?+ | 0.678414 | 1 | 0.960108 | 1 | 5.43E-10 | 1 |
| 17-22155863-G-T | T | | | | | | | | | 0.45895 | 1.92E-16 | FALSE | 1.92E-16 | ?+ | 0.123466 | 1 | 0.097652 | 1 | 6.92E-11 | 1 |
| 17-22204256-G-C | C | | | | | | | | | 0.000528 | 4.05E-10 | TRUE | 4.05E-10 | ?+ | 0.136178 | 1 | 0.030527 | 1 | 3.63E-09 | 1 |
| 17-26682500-A-T | T | 0.000899 | 0.026787 | | | 0.02679 | +? | 3.76E-05 | 0.453771 | 0.003183 | 3.54E-21 | TRUE | 1.71E-20 | +- | 0.0053 | 1 | 0.001953 | 1 | 1.34E-17 | 1 |
| 17-26682529-G-C | C | | | | | | | 9.4E-06 | 0.545566 | 0.00225 | 8.79E-29 | TRUE | 7.41E-29 | ++ | 1.06E-08 | 1 | 0.020116 | 1 | 5.15E-19 | 1 |
| 17-26747843-T-C | C | 0.444938 | 0.602053 | | | 0.6021 | +? | 0.35023 | 0.60791 | 0.468459 | 2.21E-13 | FALSE | 4.58E-06 | -- | 0.057302 | 1 | 0.381854 | 1 | 2.13E-08 | 1 |
| 17-26989948-AGC-A | A | 0.000214 | 0.641394 | | | 0.6414 | -? | | | 0.004809 | 2.48E-12 | TRUE | 2.48E-12 | ?+ | 0.080392 | 1 | 0.311765 | 1 | 3.17E-11 | 1 |
| 17-28266171-G-C | C | | | | | | | 0.000454 | 0.652248 | 0.000978 | 1.84E-08 | TRUE | 2.2E-05 | -+ | 0.036381 | 1 | 0.230422 | 1 | 4.13E-08 | 1 |
| 17-28709778-CTT-C | C | 0.001114 | 0.144595 | | | 0.1446 | +? | | | 0.002439 | 1.37E-10 | TRUE | 1.37E-10 | ?- | 1.14E-05 | 1 | 0.029919 | 1 | 3.44E-07 | 1 |
| 17-28771881-T-C | C | 0.00037 | 0.096521 | | | 0.09652 | +? | 0.000113 | 0.559424 | 0.00125 | 1.61E-10 | TRUE | 3.92E-09 | +- | 0.41568 | 1 | 0.014181 | 1 | 2.03E-09 | 1 |
| 17-29356267-A-G | G | 0.000138 | 0.625627 | | | 0.6256 | -? | 0.001175 | 0.197652 | 0.000658 | 2.5E-08 | TRUE | 1.42E-05 | -- | 0.012854 | 1 | 0.006265 | 1 | 4.32E-06 | 1 |
| 17-29356278-T-C | C | | | | | | | 0.000915 | 0.724955 | 0.000628 | 2.3E-13 | TRUE | 5.51E-06 | +- | 0.198842 | 1 | 0.27605 | 1 | 1E-12 | 1 |
| 17-29356294-T-C | C | | | | | | | 0.001081 | 0.86587 | 0.000506 | 1.42E-11 | TRUE | 0.000122 | +- | 0.810955 | 1 | 0.027974 | 1 | 6.76E-11 | 1 |
| 17-29359896-A-ACT | ACT | | | | | | | 0.000304 | 0.340699 | 0.000538 | 2.54E-08 | TRUE | 5.88E-07 | -- | 0.020668 | 1 | 0.670551 | 1 | 1.11E-07 | 1 |
| 17-29813599-CTT-C | C | | | | | | | | | 0.003269 | 7.25E-17 | TRUE | 7.25E-17 | ?- | 1.33E-05 | 1 | 0.113775 | 1 | 9.75E-13 | 1 |
| 17-29852694-GGCGC- | G | | | | | | | | | 0.000867 | 6.34E-12 | TRUE | 6.34E-12 | ?+ | 0.008025 | 1 | 0.001927 | 1 | 3.74E-08 | 1 |
| 17-29852702-C-CA | CA | | | | | | | | | 0.000561 | 4.62E-09 | TRUE | 4.62E-09 | ?+ | 0.00523 | 1 | 0.007337 | 1 | 1.25E-06 | 1 |
| 17-30815346-A-T | T | 0.000398 | 0.400849 | | | 0.4008 | -? | 0.001391 | 0.226334 | 0.001094 | 9.74E-11 | TRUE | 0.000994 | +- | 0.098931 | 1 | 0.071097 | 1 | 1.97E-10 | 1 |
| 17-31929615-C-CCT | CCT | 0.002583 | 0.296482 | | | 0.2965 | -? | | | 0.007059 | 4.65E-10 | TRUE | 4.65E-10 | ?+ | 0.012995 | 1 | 0.112684 | 1 | 1.01E-06 | 1 |
| 17-35682510-TG-T | T | 0.000156 | 0.975353 | | | 0.9754 | -? | | | 0.002187 | 2.67E-10 | TRUE | 2.67E-10 | ?+ | 0.021705 | 1 | 0.287447 | 1 | 2.47E-08 | 1 |
| 17-35882107-G-A | A | 0.000119 | 0.734149 | | | 0.7341 | -? | 0.001873 | 0.734981 | 0.001471 | 4.41E-10 | TRUE | 3.18E-05 | ++ | 0.008542 | 1 | 0.00792 | 1 | 4.12E-08 | 1 |
| 17-35882115-G-A | A | | | | | | | 0.001861 | 0.833819 | 0.001082 | 3.01E-12 | TRUE | 1.87E-05 | ++ | 4.1E-05 | 1 | 0.082792 | 1 | 1.2E-08 | 1 |
| 17-35882151-C-G | G | | | | | | | 0.001491 | 0.264791 | 0.000661 | 8.35E-11 | TRUE | 6.84E-06 | -- | 0.000804 | 1 | 0.033531 | 1 | 2.39E-07 | 1 |
| 17-38175250-CT-C | C | | | | | | | | | 0.776202 | 4.45E-11 | FALSE | 4.45E-11 | ?- | 0.116968 | 1 | 0.014014 | 1 | 1.14E-09 | 1 |
| 17-38809283-T-C | C | 0.115525 | 0.11103 | | | 0.111 | +? | | | 0.130607 | 7.75E-09 | TRUE | 7.75E-09 | ?- | 0.303192 | 1 | 0.463343 | 1 | 0.196939 | 1 |
| 17-38862181-C-T | T | 0.000137 | 0.045912 | | | 0.04591 | +? | | | 0.000796 | 1.56E-14 | TRUE | 1.56E-14 | ?+ | 0.000352 | 1 | 0.260926 | -1 | 7.79E-13 | 1 |
| 17-38955163-A-G | G | 9.76E-05 | 0.291911 | | | 0.2919 | +? | | | 0.000404 | 1.6E-09 | TRUE | 1.6E-09 | ?- | 0.053962 | 1 | 0.000647 | 1 | 1.15E-06 | 1 |
| 17-39154845-C-CA | CA | | | | | | | | | 0.00095 | 7.37E-11 | TRUE | 7.37E-11 | ?+ | 0.239557 | 1 | 0.018437 | 1 | 2E-08 | 1 |
| 17-39369988-A-C | C | 0.302401 | 0.906981 | 0.269 | 0.265052 | 0.9735 | -+ | 0.251929 | 0.132707 | 0.34235 | 6.03E-09 | FALSE | 0.02878 | +- | 0.17973 | 1 | 0.51658 | 1 | 1.24E-08 | 1 |
| 17-39956807-C-CAG | CAG | | | | | | | | | 0.000521 | 5.6E-09 | TRUE | 5.6E-09 | ?+ | 0.995612 | 1 | 0.762039 | 1 | 4.45E-10 | 1 |
| 17-40013510-CCT-C | C | | | | | | | | | 0.001634 | 8.51E-17 | TRUE | 8.51E-17 | ?- | 0.000902 | 1 | 0.315161 | 1 | 1.17E-14 | 1 |
| 17-40044663-C-CTT | CTT | | | | | | | | | 0.000349 | 9.01E-09 | TRUE | 9.02E-09 | ?+ | 0.412521 | -1 | 0.904298 | 1 | 2.78E-09 | 1 |
| 17-40544234-G-A | A | | | | | | | | | 0.137912 | 9.67E-11 | TRUE | 9.67E-11 | ?+ | 0.618891 | 1 | 0.484702 | -1 | 0.007738 | 1 |
| 17-41722828-G-GTCCC | GTCCCCAA | 9.76E-05 | 0.359043 | | | 0.359 | +? | | | 0.002488 | 5.59E-10 | TRUE | 5.59E-10 | ?- | 0.37831 | 1 | 0.192483 | 1 | 1.64E-09 | 1 |
| 17-41780474-ACT-A | A | 0.00053 | 0.288318 | | | 0.2883 | -? | | | 0.001032 | 1.05E-08 | TRUE | 1.05E-08 | ?+ | 0.256763 | 1 | 0.975721 | -1 | 4.63E-09 | 1 |
| 17-41843928-AC-A | A | 0.000468 | 0.331296 | | | 0.3313 | -? | | | 0.001006 | 7.47E-10 | TRUE | 7.47E-10 | ?+ | 9.69E-05 | 1 | 0.089294 | 1 | 1.72E-06 | 1 |
| 17-41857400-C-G | G | | | | | | | 0.001666 | 0.912437 | 0.000559 | 1.44E-08 | TRUE | 0.003249 | -- | 0.345676 | -1 | 0.06981 | 1 | 7.61E-08 | 1 |
| 17-42081656-A-G | G | | | | | | | 7.2E-05 | 0.410191 | 0.000546 | 7.92E-09 | TRUE | 1.2E-08 | -- | 0.001437 | 1 | 0.272765 | 1 | 3.24E-05 | 1 |
| 17-42101152-AG-A | A | | | | | | | | | 0.000616 | 4.15E-12 | TRUE | 4.15E-12 | ?+ | 0.115228 | 1 | 0.005696 | 1 | 3.62E-10 | 1 |
| 17-42606002-A-ATTA | ATTAATT | | | | | | | | | 0.001956 | 1.63E-10 | TRUE | 1.63E-10 | ?- | 0.001782 | 1 | 0.049888 | 1 | 5.97E-08 | 1 |
| 17-42794089-C-CAG | CAG | | | | | | | | | 0.000513 | 2.16E-11 | TRUE | 2.16E-11 | ?+ | 0.212064 | 1 | 0.002977 | 1 | 6.85E-10 | 1 |
| 17-43470088-G-A | A | | | | | | | 0.000194 | 0.759479 | 0.000737 | 8.36E-09 | TRUE | 7.87E-07 | -+ | 0.001961 | 1 | 0.027832 | 1 | 7.6E-07 | 1 |
| 17-43567458-A-T | T | | | | | | | | | 0.001646 | 9.49E-11 | TRUE | 9.49E-11 | ?- | 0.002194 | 1 | 0.428909 | 1 | 2.22E-09 | 1 |
| 17-44139757-G-A | A | 0.000568 | 0.257149 | | | 0.2571 | +? | | | 0.001338 | 1.41E-08 | TRUE | 1.41E-08 | ?+ | 0.904923 | 1 | 0.034831 | 1 | 1.11E-07 | 1 |
| 17-44189985-TG-T | T | 0.010297 | 0.03941 | | | 0.03941 | -? | 0.000576 | 0.203881 | 0.002057 | 1.09E-08 | TRUE | 3.62E-08 | ++ | 0.365608 | 1 | 0.064032 | 1 | 1.16E-08 | 1 |
| 17-44372763-G-GAA | GAA | | | | | | | | | 0.001172 | 2.65E-10 | TRUE | 2.65E-10 | ?- | 0.002534 | 1 | 0.000548 | 1 | 6.06E-06 | 1 |
| 17-44372764-GTT-G | G | | | | | | | | | 0.000961 | 1.19E-08 | TRUE | 1.19E-08 | ?+ | 0.023108 | 1 | 0.013137 | 1 | 4.74E-06 | 1 |
| 17-44513203-C-CCGA | CCGA | | | | | | | | | 0.000836 | 3.95E-10 | TRUE | 3.95E-10 | ?+ | 0.011079 | 1 | 0.004384 | 1 | 2.45E-07 | 1 |
| 17-45057514-G-C | C | 0.000781 | 0.309256 | | | 0.3093 | -? | | | 0.00086 | 6.9E-12 | TRUE | 6.91E-12 | ?+ | 0.18307 | 1 | 0.016871 | 1 | 2.31E-10 | 1 |
| 17-45079271-A-ATT | ATT | | | | | | | | | 0.000801 | 3.23E-12 | TRUE | 3.23E-12 | ?- | 0.030544 | 1 | 0.01697 | 1 | 3.14E-10 | 1 |
| 17-47255122-G-GCA | GCA | 0.00078 | 0.979157 | | | 0.9792 | -? | | | 0.002397 | 4.5E-13 | TRUE | 4.5E-13 | ?- | 0.000688 | 1 | 0.048592 | 1 | 1.25E-08 | 1 |
| 17-47255125-CCT-C | C | 0.000801 | 0.934709 | | | 0.9347 | +? | | | 0.001929 | 3.05E-12 | TRUE | 3.05E-12 | ?- | 0.011719 | 1 | 0.017004 | 1 | 3.15E-08 | 1 |
| 17-47530102-ATTTTTT | A | | | | | | | | | 0.008127 | 7.66E-11 | TRUE | 7.66E-11 | ?+ | 0.054848 | 1 | 0.319356 | 1 | 1.06E-09 | 1 |
| 17-48098623-A-G | G | | | | | | | | | 0.000612 | 9.66E-10 | TRUE | 9.66E-10 | ?- | 0.014254 | 1 | 0.042641 | 1 | 9.44E-09 | 1 |
| 17-48866581-C-CCT | CCT | | | | | | | | | 0.001254 | 1.57E-10 | TRUE | 1.57E-10 | ?+ | 0.426027 | -1 | 0.119428 | 1 | 1.18E-10 | 1 |
| 17-48965289-A-G | G | 0.45215 | 0.054229 | 0.478 | 0.306914 | 0.3069 | ?+ | 0.448786 | 0.15124 | 0.567071 | 1.59E-09 | FALSE | 6.26E-06 | -- | 0.156417 | 1 | 0.52062 | -1 | 3.58E-08 | 1 |

| | | | | | | | | | | | | | | | | | | | | |
|---|---|---|---|---|---|---|---|---|---|---|---|---|---|---|---|---|---|---|---|---|
| 17-49143921-GTGAA- | G | | | | | | | | | 0.000121 | 3.6E-09 | TRUE | 3.6E-09 | ?+ | | | | | 5.71E-09 | 1 |
| 17-49345263-C-T | T | | | | | | | 0.000733 | 0.723394 | 0.001158 | 1.83E-12 | TRUE | 1.48E-08 | ++ | 0.009696 | 1 | 0.010463 | 1 | 1.8E-10 | 1 |
| 17-49641837-A-C | C | 0.000392 | 0.952699 | | | 0.9527 | -? | 1.88E-05 | 0.280772 | 0.000482 | 3.54E-08 | TRUE | 1.95E-07 | +- | 0.195963 | 1 | 0.267426 | 1 | 1.14E-06 | 1 |
| 17-49678884-C-CCAT( | CCATCCTGGCTAA | | | | | | | | | 0.000503 | 3.12E-08 | TRUE | 3.12E-08 | ?+ | 0.002097 | 1 | 0.018002 | 1 | 0.000406 | 1 |
| 17-50126919-GT-G | G | | | | | | | | | 0.001802 | 3.69E-08 | TRUE | 3.69E-08 | ?+ | 0.009963 | 1 | 0.622356 | -1 | 1.66E-07 | 1 |
| 17-50156079-A-ACG | ACG | 0.001463 | 0.109279 | | | 0.1093 | +? | 0.000298 | 0.271093 | 0.002641 | 4.02E-11 | TRUE | 6.95E-09 | +- | 0.01056 | 1 | 0.012376 | 1 | 4.91E-09 | 1 |
| 17-50759908-G-GAC | GAC | | | | | | | | | 0.006687 | 1.56E-15 | TRUE | 1.56E-15 | ?- | 0.069849 | 1 | 0.305096 | 1 | 8.53E-14 | 1 |
| 17-58478215-C-CCT | CCT | | | | | | | | | 0.002764 | 4.76E-09 | TRUE | 4.76E-09 | ?+ | 0.03496 | 1 | 0.358867 | 1 | 1.07E-08 | 1 |
| 17-58685271-T-TA | TA | | | | | | | 0.005954 | 0.841849 | 0.004782 | 1.08E-17 | TRUE | 4.21E-07 | +- | 0.005508 | 1 | 0.807792 | -1 | 2.51E-17 | 1 |
| 17-59120928-A-G | G | 0.000176 | 0.100788 | | | 0.1008 | +? | 4.7E-05 | 0.643843 | 0.000797 | 1.64E-08 | TRUE | 9.86E-08 | +- | 1.9E-06 | 1 | 0.773781 | 1 | 1.58E-05 | 1 |
| 17-59965446-C-CT | CT | | | | | | | | | 0.001177 | 2.78E-10 | TRUE | 2.78E-10 | ?+ | 0.015649 | 1 | 0.001781 | 1 | 8.26E-08 | 1 |
| 17-60340884-C-G | G | | | | | | | 0.004345 | 0.574971 | 0.003266 | 2.77E-19 | TRUE | 5.36E-07 | +- | 8.52E-05 | 1 | 0.041504 | 1 | 3.23E-14 | 1 |
| 17-60341469-A-ATTC1 | ATTCTT | 0.000626 | 0.181872 | | | 0.1819 | +? | | | 0.001918 | 2.83E-10 | TRUE | 2.83E-10 | ?- | 0.006815 | 1 | 0.262358 | 1 | 2.47E-09 | 1 |
| 17-60589164-G-GGGT | GGGT | 0.000546 | 0.433764 | | | 0.4338 | +? | | | 0.002616 | 6.02E-11 | TRUE | 6.02E-11 | ?- | 0.173795 | 1 | 0.290377 | 1 | 4.38E-11 | 1 |
| 17-60703056-CCT-C | C | | | | | | | | | 0.001316 | 1.38E-08 | TRUE | 1.38E-08 | ?- | 0.111398 | 1 | 0.061443 | 1 | 8.66E-07 | 1 |
| 17-61890402-C-G | G | 0.000197 | 0.540373 | | | 0.5404 | -? | | | 0.000794 | 8.85E-09 | TRUE | 8.85E-09 | ?- | 0.009158 | 1 | 0.552296 | 1 | 8.16E-08 | 1 |
| 17-62007671-A-AC | AC | 0.000624 | 0.590158 | | | 0.5902 | +? | 0.003007 | 0.649111 | 0.006707 | 3.69E-22 | TRUE | 3.14E-13 | +- | 0.003102 | 1 | 0.010219 | 1 | 6.47E-19 | 1 |
| 17-62157437-C-A | A | | | | | | | 0.000207 | 0.668676 | 0.001061 | 3.13E-18 | TRUE | 3.21E-16 | ++ | 0.021906 | 1 | 0.00337 | 1 | 2.48E-16 | 1 |
| 17-62157438-A-G | G | | | | | | | 0.00011 | 0.800837 | 0.000775 | 3.28E-14 | TRUE | 1.91E-12 | +- | 0.011719 | 1 | 0.059246 | 1 | 1.55E-12 | 1 |
| 17-62235247-G-A | A | 0.001678 | 0.697882 | | | 0.6979 | +? | 0.000426 | 0.42173 | 0.003678 | 5.33E-09 | TRUE | 9.46E-07 | -+ | 0.376081 | 1 | 0.866502 | -1 | 1.21E-11 | 1 |
| 17-62235962-A-C | C | 0.003009 | 0.55219 | | | 0.5522 | +? | | | 0.002018 | 3.16E-13 | TRUE | 3.16E-13 | ?- | 0.000296 | 1 | 0.242587 | 1 | 4.74E-11 | 1 |
| 17-62235981-T-C | C | 0.003203 | 0.452126 | | | 0.4521 | +? | | | 0.003602 | 5.91E-21 | TRUE | 5.91E-21 | ?- | 0.00415 | 1 | 0.000283 | 1 | 3.03E-16 | 1 |
| 17-62249528-A-ATCA" | ATCAT | | | | | | | | | 0.000795 | 2.92E-08 | TRUE | 2.92E-08 | ?- | 0.062273 | 1 | 0.033863 | 1 | 6.21E-07 | 1 |
| 17-62388645-C-G | G | | | | | | | | | 0.000295 | 1.72E-08 | TRUE | 1.72E-08 | ?- | 0.02146 | 1 | 0.505399 | 1 | 4.47E-07 | 1 |
| 17-62411588-A-G | G | | | | | | | 0.000254 | 0.070427 | 0.000374 | 9.73E-10 | TRUE | 0.000169 | +- | 0.329165 | 1 | 0.003551 | 1 | 9.16E-09 | 1 |
| 17-62466398-A-G | G | | | | | | | 0.000345 | 0.481966 | 0.000461 | 4.39E-08 | TRUE | 3.42E-06 | -- | 0.0076 | 1 | 0.034939 | 1 | 1.47E-05 | 1 |
| 17-62498597-C-G | G | | | | | | | | | 0.000635 | 4.65E-08 | TRUE | 4.65E-08 | ?- | 0.001763 | 1 | 0.26006 | 1 | 1.08E-05 | 1 |
| 17-62604736-CCGTG-( | C | 0.000862 | 0.410331 | | | 0.4103 | -? | | | 0.001794 | 5.16E-09 | TRUE | 5.16E-09 | ?- | 0.598226 | -1 | 0.056526 | 1 | 3.18E-08 | 1 |
| 17-63851546-G-C | C | 0.000294 | 0.26259 | | | 0.2626 | +? | | | 0.001397 | 3.09E-12 | TRUE | 3.09E-12 | ?+ | 0.949833 | 1 | | | 4.66E-12 | 1 |
| 17-64347887-G-T | T | | | | | | | 0.001269 | 0.074886 | 0.002124 | 2.86E-21 | TRUE | 3.35E-17 | ++ | 0.001577 | 1 | | | 7.38E-16 | 1 |
| 17-64347888-A-G | G | | | | | | | 0.000949 | 0.590512 | 0.001547 | 9.28E-14 | TRUE | 2.01E-09 | -- | 0.002827 | 1 | | | 2.58E-10 | 1 |
| 17-64347889-G-C | C | | | | | | | 0.000905 | 0.781658 | 0.001783 | 5.52E-15 | TRUE | 2.38E-10 | ++ | 0.003798 | 1 | | | 5.02E-11 | 1 |
| 17-67064485-A-G | G | | | | | | | | | 0.00033 | 6.63E-09 | TRUE | 6.63E-09 | ?- | 0.040351 | 1 | | | 3.11E-07 | 1 |
| 17-67634352-G-A | A | 0.00051 | 0.302698 | | | 0.3027 | -? | | | 0.000431 | 1.79E-08 | TRUE | 1.79E-08 | ?+ | 0.071056 | 1 | | | 8.98E-08 | 1 |
| 17-67956115-G-A | A | | | | | | | 0.000893 | 0.875483 | 0.000799 | 1.52E-09 | TRUE | 7.34E-05 | -+ | 0.416742 | 1 | | | 4.15E-09 | 1 |
| 17-67956117-G-A | A | | | | | | | 0.000883 | 0.928741 | 0.000706 | 1.52E-08 | TRUE | 0.000242 | -+ | 0.67491 | 1 | | | 9.04E-09 | 1 |
| 17-68002426-A-AT | AT | 7.96E-05 | 0.574154 | | | 0.5742 | -? | | | 0.000884 | 3.78E-15 | TRUE | 3.78E-15 | ?- | 0.658864 | 1 | | | 9.38E-13 | 1 |
| 17-68627016-A-T | T | | | | | | | | | 0.000958 | 1.35E-10 | TRUE | 1.35E-10 | ?- | 0.289286 | 1 | | | 5.12E-09 | 1 |
| 17-69099080-C-CTTT | CTTT | 0.000137 | 0.659426 | | | 0.6594 | +? | | | 0.000659 | 5.43E-09 | TRUE | 5.43E-09 | ?+ | 0.164186 | 1 | | | 5.81E-09 | 1 |
| 17-69514705-G-GCA | GCA | | | | | | | | | 0.000322 | 4.89E-10 | TRUE | 4.89E-10 | ?- | 0.007029 | 1 | | | 7.5E-06 | 1 |
| 17-73051724-G-GGT | GGT | | | | | | | | | 0.001527 | 1.1E-09 | TRUE | 1.1E-09 | ?- | 0.000672 | 1 | | | 1.53E-06 | 1 |
| 17-74976489-C-CCT | CCT | | | | | | | | | 0.000757 | 1.59E-08 | TRUE | 1.59E-08 | ?+ | 0.18517 | 1 | | | 3.95E-07 | 1 |
| 17-75206797-A-AAG | AAG | 0.000742 | 0.188219 | | | 0.1882 | +? | | | 0.004098 | 3.96E-22 | TRUE | 3.96E-22 | ?- | 0.09202 | 1 | | | 1.12E-19 | 1 |
| 17-75206799-ACC-A | A | | | | | | | | | 0.003778 | 2.44E-25 | TRUE | 2.44E-25 | ?+ | 0.013279 | 1 | | | 2.28E-21 | 1 |
| 17-75529672-C-CTAG( | CTAGCA | | | | | | | | | 0.001324 | 2.24E-09 | TRUE | 2.24E-09 | ?+ | 0.038167 | 1 | | | 7.93E-07 | 1 |
| 17-75801049-G-GCA | GCA | | | | | | | | | 0.002206 | 1.46E-19 | TRUE | 1.46E-19 | ?- | 0.442818 | -1 | | | 1.32E-16 | 1 |
| 17-75801051-CAG-C | C | | | | | | | | | 0.001802 | 2.54E-16 | TRUE | 2.54E-16 | ?- | 0.437738 | -1 | | | 3.31E-14 | 1 |
| 17-75801061-G-A | A | 0.000137 | 0.933668 | | | 0.9337 | -? | 0.001009 | 0.641255 | 0.001083 | 1.96E-10 | TRUE | 4.56E-05 | -+ | 0.08817 | 1 | | | 1.01E-08 | 1 |
| 17-75801070-ATTT-A | A | | | | | | | 0.001081 | 0.839552 | 0.004226 | 3.08E-30 | TRUE | 3.9E-23 | -+ | 0.000581 | 1 | | | 7.72E-21 | 1 |
| 17-75968218-C-CCT | CCT | | | | | | | | | 0.006614 | 4.04E-08 | TRUE | 4.04E-08 | ?+ | 0.205025 | 1 | | | 1.71E-08 | 1 |
| 17-75993291-AT-A | A | | | | | | | | | 0.001037 | 1.04E-11 | TRUE | 1.04E-11 | ?+ | 0.001944 | 1 | | | 7.68E-08 | 1 |
| 17-75993927-CGGA-C | C | | | | | | | | | 0.001005 | 8.22E-11 | TRUE | 8.22E-11 | ?- | 0.133905 | 1 | | | 5.8E-10 | 1 |
| 17-75993955-T-C | C | 0.476828 | 0.671475 | 0.433 | 0.083836 | 0.7756 | -+ | | | 0.508946 | 2.9E-10 | FALSE | 2.9E-10 | ?- | 0.941058 | -1 | | | 1.97E-07 | 1 |
| 17-76098643-C-CCT | CCT | | | | | | | | | 0.001572 | 2.15E-11 | TRUE | 2.15E-11 | ?+ | 0.335524 | 1 | | | 1.96E-11 | 1 |
| 17-78351265-G-GTAA | GTAATCCCA | | | | | | | | | 0.03571 | 1.91E-11 | FALSE | 1.91E-11 | ?+ | 0.190096 | -1 | | | 3.2E-09 | -1 |
| 17-78864868-A-AC | AC | | | | | | | | | 0.000972 | 3.27E-08 | TRUE | 3.27E-08 | ?- | 0.010914 | 1 | | | 2.1E-07 | 1 |
| 17-79512696-A-G | G | | | | | | | | | 0.88675 | 2.12E-25 | FALSE | 2.12E-25 | ?+ | 0.084446 | -1 | | | 2.88E-22 | -1 |
| 17-80875759-G-T | T | | | | | | | 0.003142 | 0.785684 | 0.001574 | 6.55E-09 | TRUE | 0.005042 | -+ | 0.05032 | 1 | | | 9.38E-09 | 1 |
| 17-81357652-C-T | T | | | | | | | 0.00026 | 0.305022 | 0.000602 | 7.89E-10 | TRUE | 3.55E-06 | -+ | 0.007554 | 1 | | | 1.59E-08 | 1 |

| | | | | | | | | | | | | | | | | | | | | |
|---|---|---|---|---|---|---|---|---|---|---|---|---|---|---|---|---|---|---|---|---|
| 17-81594086-A-G | G | 0.000747 | 0.008041 | | | 0.008041 | +? | 0.001738 | 0.006592 | 0.002078 | 2.14E-09 | TRUE | 0.01737 | +- | 0.053555 | 1 | | | 4.32E-08 | 1 |
| 17-81848677-G-A | A | 0.001986 | 0.769441 | | | 0.7694 | -? | 0.000307 | 0.605935 | 0.000883 | 2.48E-08 | TRUE | 1.16E-06 | ++ | 0.000297 | 1 | | | 0.000181 | 1 |
| 17-81849383-C-G | G | | | | | | | 0.000558 | 0.302558 | 0.000972 | 3.98E-13 | TRUE | 2.07E-10 | -- | 0.025285 | 1 | | | 1.26E-12 | 1 |
| 17-82148088-G-GGAT | GGATA | | | | | | | | | 0.963662 | 2.11E-11 | FALSE | 2.11E-11 | ?+ | 0.109449 | -1 | | | 4.31E-12 | -1 |
| 17-82725483-G-GCT | GCT | | | | | | | | | 0.001389 | 8.45E-12 | TRUE | 8.45E-12 | ?- | 0.007407 | 1 | | | 1.67E-10 | 1 |
| 17-82762297-T-C | C | | | 0.429 | 0.508973 | 0.509 | ?+ | | | 0.491852 | 8.02E-09 | FALSE | 8.02E-09 | ?- | 0.138504 | 1 | | | 1.29E-05 | 1 |
| 17-82827811-GC-G | G | | | | | | | 0.002672 | 0.904053 | 0.00495 | 9.74E-09 | TRUE | 1.59E-05 | ++ | 0.192957 | 1 | | | 1.04E-08 | 1 |
| 17-83103876-CCCGCC | C | | | | | | | | | 0.058666 | 1.66E-08 | FALSE | 1.66E-08 | ?- | 0.393306 | 1 | | | 5.37E-08 | 1 |
| 17-33399927-G-A | A | | | | | | | 1.88E-05 | 0.08908 | | | FALSE | 1.9E-305 | -? | | | | | | |
| 17-18762165-C-T | T | | | | | | | 2.19E-05 | 0.03984 | 0.000308 | 3.16E-07 | TRUE | 3.66E-08 | ++ | 0.243593 | 1 | 0.516076 | 1 | 2.47E-06 | 1 |
| 18-49233661-A-G | G | 0.026482 | 2.36E-08 | 0.002 | 0.362126 | 2.479E-08 | +- | 0.000144 | 0.319086 | 0.013954 | 0.225767 | TRUE | 0.1661 | -- | 0.969101 | -1 | 0.714638 | 1 | 0.497187 | -1 |
| 18-49235132-G-T | T | 0.026053 | 1.29E-08 | | | 1.295E-08 | -? | 0.000147 | 0.126335 | 0.013834 | 0.085897 | TRUE | 0.04734 | ++ | 0.982111 | 1 | 0.623232 | 1 | 0.762479 | -1 |
| 18-49261602-GCA-G | G | 0.025841 | 5.99E-09 | | | 5.985E-09 | -? | 0.000147 | 0.131501 | 0.013781 | 0.095669 | TRUE | 0.05368 | ++ | 0.982194 | 1 | 0.656594 | 1 | 0.762181 | -1 |
| 18-49326113-A-G | G | 0.026249 | 1.64E-08 | 0.002 | 0.362126 | 1.717E-08 | +- | 0.000147 | 0.134878 | 0.013963 | 0.089027 | TRUE | 0.04997 | -- | 0.999328 | -1 | 0.634499 | 1 | 0.762211 | -1 |
| 18-49403096-A-G | G | 0.026637 | 7.77E-09 | 0.003 | 0.362126 | 8.158E-09 | +- | 0.00021 | 0.186273 | 0.014102 | 0.111054 | TRUE | 0.06329 | -- | 0.901609 | -1 | 0.643726 | 1 | 0.43001 | -1 |
| 18-122970-T-TGAGA | TGAGA | | | | | | | | | 0.013393 | 1.81E-12 | FALSE | 1.81E-12 | ?- | 0.856448 | 1 | 0.008873 | 1 | 4.6E-10 | 1 |
| 18-218980-CA-C | C | | | | | | | 0.001698 | 0.196996 | 0.010936 | 2.55E-08 | TRUE | 4.71E-08 | -- | 0.292391 | -1 | 0.310092 | 1 | 1.17E-12 | 1 |
| 18-2996644-AT-A | A | 0.000234 | 0.442243 | | | 0.4422 | -? | 0.000761 | 0.195103 | 0.000908 | 8.4E-10 | TRUE | 0.000444 | -+ | 0.012312 | 1 | 0.120982 | 1 | 3.1E-06 | 1 |
| 18-3550114-A-G | G | | | | | | | | | 0.000497 | 4.9E-09 | TRUE | 4.9E-09 | ?- | 0.093543 | 1 | 0.294355 | 1 | 4.96E-09 | 1 |
| 18-7107368-T-TTGG | TTGG | | | | | | | | | 0.000361 | 4.49E-08 | TRUE | 4.49E-08 | ?- | 0.002765 | 1 | 0.010552 | 1 | 0.000104 | 1 |
| 18-7274664-A-AT | AT | | | | | | | | | 0.001114 | 3.18E-08 | TRUE | 3.18E-08 | ?- | 0.585719 | -1 | 0.005369 | 1 | 1.84E-07 | 1 |
| 18-9037196-GGT-G | G | 7.8E-05 | 0.858703 | | | 0.8587 | +? | | | 0.000442 | 2.68E-08 | TRUE | 2.68E-08 | ?+ | 0.062096 | 1 | 0.059776 | 1 | 2.21E-07 | 1 |
| 18-9777902-C-A | A | | | | | | | | | 0.000558 | 3.5E-10 | TRUE | 3.5E-10 | ?+ | 0.002111 | 1 | 0.019416 | 1 | 6.52E-07 | 1 |
| 18-12866210-AGGTG- | A | | | | | | | | | 0.001905 | 5.67E-21 | TRUE | 5.67E-21 | ?+ | 0.010276 | 1 | 0.405386 | 1 | 2.39E-20 | 1 |
| 18-14501440-G-C | C | | | | | | | 0.000204 | 0.036962 | 0.00126 | 3.48E-08 | TRUE | 1.05E-05 | -+ | 0.538412 | -1 | 0.049684 | 1 | 2.8E-07 | 1 |
| 18-14501562-T-C | C | | | | | | | 0.000175 | 0.513893 | 0.000564 | 7.11E-09 | TRUE | 1E-07 | -- | 0.423707 | 1 | 0.264584 | 1 | 3.47E-09 | 1 |
| 18-14502080-C-G | G | | | | | | | | | 0.000495 | 2.78E-09 | TRUE | 2.78E-09 | ?- | 0.67759 | 1 | 0.37768 | -1 | 1.02E-10 | 1 |
| 18-21616146-C-T | T | 0.000905 | 0.601936 | | | 0.6019 | -? | 0.000119 | 0.377088 | 0.000427 | 2.45E-09 | TRUE | 1.14E-08 | ++ | 0.086298 | 1 | 0.450769 | -1 | 3.79E-08 | 1 |
| 18-21653077-A-G | G | | | | | | | 0.06166 | 0.900881 | 0.228428 | 3.7E-10 | FALSE | 2.25E-06 | +- | 0.001469 | 1 | 0.032624 | 1 | 2.69E-06 | 1 |
| 18-21731482-C-A | A | | | | | | | | | 0.00339 | 8.83E-24 | TRUE | 8.83E-24 | ?+ | 0.014956 | 1 | 0.001847 | 1 | 3.86E-20 | 1 |
| 18-21974281-G-C | C | 0.000137 | 0.732885 | | | 0.7329 | -? | 0.001275 | 0.670013 | 0.001901 | 5.28E-16 | TRUE | 1.36E-10 | ++ | 0.02489 | 1 | 0.146333 | 1 | 1.99E-15 | 1 |
| 18-23093206-AT-A | A | | | | | | | | | 0.001017 | 5E-11 | TRUE | 5E-11 | ?+ | 0.558453 | -1 | 0.674895 | 1 | 2.37E-13 | 1 |
| 18-23093208-CA-C | C | | | | | | | | | 0.000928 | 1.23E-08 | TRUE | 1.23E-08 | ?- | 0.200261 | -1 | 0.453295 | -1 | 7.33E-12 | 1 |
| 18-33147978-C-CT | CT | | | | | | | 5.64E-05 | 0.343818 | 0.006503 | 1.37E-43 | TRUE | 1.12E-42 | -+ | 3.31E-05 | 1 | 1.37E-05 | 1 | 8.01E-36 | 1 |
| 18-36077637-A-G | G | | | | | | | | | 0.048518 | 4.54E-10 | FALSE | 4.54E-10 | ?- | 0.477418 | 1 | 0.447637 | 1 | 1.76E-11 | 1 |
| 18-47400508-TCCACA | T | | | | | | | | | 0.010317 | 1.23E-08 | TRUE | 1.23E-08 | ?+ | 0.101426 | 1 | 0.323142 | 1 | 1.58E-07 | 1 |
| 18-54138189-A-AT | AT | | | | | | | | | 0.001789 | 5.35E-09 | TRUE | 5.35E-09 | ?- | 0.00285 | 1 | 0.02569 | 1 | 3.11E-06 | 1 |
| 18-54207819-C-CCT | CCT | | | | | | | | | 0.002025 | 4.12E-08 | TRUE | 4.12E-08 | ?+ | 0.029393 | 1 | 0.016146 | 1 | 1.89E-06 | 1 |
| 18-54537923-C-CTACT | CTACTGAACCCACAACCACAGCAGCCAGGACTGTAACTGAACAGTCTGCCATCACAACTGTAGGCATTTACACTGA | | | | | | | | | 0.000494 | 3.39E-08 | TRUE | 3.39E-08 | ?+ | 0.81058 | 1 | 0.003368 | 1 | 1.78E-06 | 1 |
| 18-54758841-C-G | G | | | | | | | 0.000495 | 0.983058 | 0.00041 | 1.8E-08 | TRUE | 9.93E-05 | +- | 0.025124 | 1 | 0.014871 | 1 | 2.41E-07 | 1 |
| 18-56903567-G-C | C | | | | | | | 0.000207 | 0.659043 | 0.001287 | 1.04E-16 | TRUE | 3.32E-15 | ++ | 0.100524 | 1 | 0.010975 | 1 | 7.88E-14 | 1 |
| 18-58129746-CGGTGT | C | | | | | | | | | 0.003143 | 3.11E-08 | TRUE | 3.11E-08 | ?- | 0.23784 | 1 | 0.488396 | 1 | | |
| 18-61699837-C-T | T | 0.042592 | 0.763633 | | | 0.7636 | -? | | | 0.019758 | 1.07E-10 | TRUE | 1.07E-10 | ?- | 0.00398 | -1 | 0.010302 | -1 | | |
| 18-62195984-C-CTG | CTG | | | | | | | | | 0.001381 | 3.05E-08 | TRUE | 3.05E-08 | ?+ | 0.012451 | 1 | 0.949697 | 1 | | |
| 18-62195985-CAA-C | C | | | | | | | | | 0.001972 | 1.14E-13 | TRUE | 1.14E-13 | ?- | 0.025556 | 1 | 0.302142 | 1 | | |
| 18-67758970-G-A | A | | | | | | | 0.001344 | 0.960147 | 0.001121 | 8.71E-13 | TRUE | 1.68E-06 | ++ | 0.015332 | 1 | | | | |
| 18-76571552-A-ACC | ACC | | | | | | | 0.000188 | 0.33332 | 0.000439 | 1.1E-08 | TRUE | 7.85E-08 | -- | 0.019028 | 1 | | | | |
| 18-78514878-C-T | T | | | | | | | | | 0.744438 | 3.75E-11 | FALSE | 3.75E-11 | ?- | 0.098931 | 1 | | | | |
| 18-78591455-C-CCT | CCT | 7.8E-05 | 0.452807 | | | 0.4528 | -? | | | 0.001042 | 4.15E-09 | TRUE | 4.15E-09 | ?+ | 0.006057 | 1 | | | | |
| 18-78781929-GATGG/ | G | | | | | | | | | 0.033781 | 1.63E-09 | FALSE | 1.63E-09 | ?- | 0.155158 | -1 | | | | |
| 18-79473554-GAGGG/ | G | | | | | | | | | 0.031682 | 5.53E-11 | FALSE | 5.54E-11 | ?+ | 0.299475 | 1 | | | | |
| 18-79894704-C-T | T | | | | | | | | | 0.904228 | 3.06E-14 | FALSE | 3.06E-14 | ?- | 0.677748 | 1 | | | | |
| 18-80185678-A-G | G | | | | | | | | | 0.00085 | 1.17E-10 | TRUE | 1.17E-10 | ?- | 0.193603 | 1 | | | | |
| 18-9556511-G-C | C | | | | | | | 1.25E-05 | 0.176973 | | | FALSE | 2.5E-49 | -? | | | | | | |
| 18-21899459-T-A | A | | | | | | | 1.25E-05 | 0.087315 | | | FALSE | 1.2E-305 | -? | | | | | | |
| 19-42748176-A-G | G | 0.025962 | 3.52E-08 | | | 3.517E-08 | +? | 0.004736 | 0.229674 | | | FALSE | 0.2297 | -? | | | | | | |
| 19-44746404-C-T | T | 0.205885 | 4.69E-08 | 0.28 | 0.608472 | 4.125E-08 | ++ | 0.296818 | 1E-06 | 0.207674 | 0.000198 | TRUE | 8.65E-10 | ++ | 0.455577 | -1 | 0.667855 | 1 | 3.31E-05 | 1 |
| 19-44750911-C-A | A | 0.082385 | 3.31E-08 | 0.147 | 0.934838 | 3.416E-08 | ++ | 0.131424 | 6.37E-15 | 0.085136 | 0.01398 | TRUE | 2.72E-15 | ++ | 0.650374 | -1 | 0.379263 | -1 | 0.002655 | 1 |
| 19-44781370-G-A | A | 0.009237 | 2.84E-10 | 0.015 | 0.136875 | 2.343E-10 | ++ | 0.013565 | 1.94E-13 | 0.007409 | 0.004804 | TRUE | 8.09E-15 | ++ | 0.048335 | 1 | 0.662296 | 1 | 0.007805 | 1 |

| | | | | | | | | | | | | | | | | | | | | |
|---|---|---|---|---|---|---|---|---|---|---|---|---|---|---|---|---|---|---|---|---|
| 19-44792629-G-A | A | 0.024653 | 1.59E-09 | 0.026 | 0.102834 | 1.183E-09 | -- | 0.034433 | 1.76E-06 | 0.032914 | 0.005318 | TRUE | 3.51E-08 | -- | 0.246837 | -1 | 0.134444 | -1 | 0.059964 | -1 |
| 19-44793107-G-A | A | 0.00727 | 1.68E-08 | 0.01 | 0.05943 | 1.308E-08 | -- | 0.018672 | 1.16E-05 | 0.012662 | 0.002268 | TRUE | 9.29E-08 | -- | 0.235304 | -1 | 0.503209 | -1 | 0.017671 | -1 |
| 19-44793549-C-T | T | 0.024689 | 1.66E-09 | 0.027 | 0.102834 | 1.229E-09 | -- | 0.034433 | 1.78E-06 | 0.032947 | 0.004623 | TRUE | 3.06E-08 | -- | 0.238008 | -1 | 0.121316 | -1 | 0.055232 | -1 |
| 19-44795942-G-T | T | 0.025063 | 4.03E-08 | 0.028 | 0.12442 | 3.133E-08 | -- | 0.033345 | 1.36E-06 | 0.032684 | 0.003612 | TRUE | 1.85E-08 | -- | 0.996595 | 1 | 0.043707 | -1 | 0.024578 | -1 |
| 19-44799865-AAAAG- | A | 0.019372 | 1.13E-09 | | | 1.132E-09 | -? | 0.033454 | 8.88E-07 | 0.026789 | 0.018528 | TRUE | 7.6E-08 | -- | 0.53958 | -1 | 0.604436 | -1 | 0.054878 | -1 |
| 19-44813331-G-A | A | 0.015883 | 4.82E-12 | 0.022 | 0.032572 | 3.081E-12 | -- | 0.031896 | 6.12E-07 | 0.022517 | 0.015605 | TRUE | 4.23E-08 | -- | 0.745125 | 1 | 0.52407 | -1 | 0.03222 | -1 |
| 19-44820881-G-A | A | 0.232244 | 1.63E-19 | 0.339 | 0.020289 | 4.149E-20 | ++ | 0.308936 | 1.07E-33 | 0.221855 | 0.287699 | TRUE | 1.31E-26 | ++ | 0.184817 | -1 | 0.799075 | -1 | 0.084186 | 1 |
| 19-44822960-T-C | C | 0.32857 | 4.04E-09 | 0.37 | 0.315519 | 2.841E-09 | -- | | | 0.307038 | 0.033279 | TRUE | 0.03328 | ?- | 0.266253 | -1 | 0.758649 | -1 | 0.0085 | 1 |
| 19-44823407-C-T | T | 0.325431 | 1.04E-08 | 0.367 | 0.346166 | 7.553E-09 | ++ | | | 0.303366 | 0.042249 | TRUE | 0.04225 | ?+ | 0.224707 | -1 | 0.510033 | -1 | 0.008466 | 1 |
| 19-44824052-C-T | T | 0.339203 | 4.47E-08 | 0.378 | 0.285433 | 3.102E-08 | ++ | | | 0.317284 | 0.072756 | TRUE | 0.07276 | ?+ | 0.151643 | -1 | 0.454556 | -1 | 0.011936 | 1 |
| 19-44825122-T-A | A | 0.335387 | 4.47E-08 | 0.373 | 0.285433 | 3.098E-08 | ++ | 0.318945 | 8.33E-27 | 0.313805 | 0.086004 | TRUE | 1.63E-22 | ++ | 0.137384 | -1 | 0.381746 | -1 | 0.013088 | 1 |
| 19-44825123-C-A | A | 0.335374 | 4.85E-08 | 0.373 | 0.285433 | 3.365E-08 | ++ | 0.318945 | 8.33E-27 | 0.313842 | 0.086978 | TRUE | 1.68E-22 | ++ | 0.134136 | -1 | 0.381746 | -1 | 0.013105 | 1 |
| 19-44827852-G-C | C | 0.006643 | 9.3E-10 | 0.02 | 0.000266 | 5.485E-10 | ++ | 0.009163 | 4.91E-22 | 0.004906 | 0.141143 | TRUE | 3.89E-20 | ++ | 0.994943 | 1 | 0.513688 | 1 | 0.15239 | 1 |
| 19-44828468-C-T | T | 0.287281 | 1.65E-08 | | | 1.646E-08 | +? | | | 0.263704 | 0.02155 | FALSE | 0.02155 | ?+ | 0.074599 | -1 | 0.711243 | -1 | 0.005428 | 1 |
| 19-44842366-G-A | A | 0.274326 | 1.59E-08 | 0.312 | 0.50522 | 1.302E-08 | ++ | 0.281689 | 6.1E-24 | 0.253234 | 0.029654 | FALSE | 1.43E-21 | ++ | 0.092181 | -1 | 0.986704 | -1 | 0.013547 | 1 |
| 19-44843409-C-G | G | 0.009243 | 5.62E-13 | 0.015 | 0.007555 | 3.717E-13 | -- | 0.013602 | 5.77E-19 | 0.007307 | 0.030667 | TRUE | 8.13E-19 | -- | 0.239837 | 1 | 0.076849 | 1 | 0.062417 | 1 |
| 19-44845265-G-T | T | 0.274379 | 7.69E-09 | 0.313 | 0.44118 | 6.015E-09 | ++ | | | 0.253526 | 0.04256 | FALSE | 0.04256 | ?+ | 0.085288 | -1 | 0.923363 | -1 | 0.018733 | 1 |
| 19-44848259-C-G | G | 0.03103 | 4.94E-30 | 0.063 | 0.367582 | 3.555E-30 | -- | 0.04046 | 9.43E-65 | 0.023875 | 0.002147 | TRUE | 6.64E-59 | -- | 0.15338 | -1 | 0.408483 | -1 | 0.000167 | 1 |
| 19-44851039-A-G | G | 0.423155 | 1.95E-09 | 0.45 | 0.066974 | 9.115E-10 | ++ | 0.51095 | 5.39E-10 | 0.433103 | 0.331867 | TRUE | 1.7E-08 | ++ | 0.660196 | -1 | 0.965147 | -1 | 0.289742 | -1 |
| 19-44852031-C-T | T | 0.008361 | 1.69E-09 | 0.016 | 0.139607 | 1.42E-09 | ++ | 0.010083 | 1.66E-17 | 0.005794 | 0.333973 | TRUE | 1.9E-15 | ++ | 0.179943 | -1 | 0.226945 | -1 | 0.176953 | 1 |
| 19-44852464-C-G | G | 0.280648 | 1.44E-11 | 0.304 | 0.237637 | 8.952E-12 | -- | 0.259766 | 3.11E-29 | 0.257845 | 0.016487 | TRUE | 4.19E-26 | -- | 0.269836 | -1 | 0.649044 | 1 | 0.005672 | 1 |
| 19-44853207-T-A | A | 0.252982 | 1.63E-10 | | | 1.632E-10 | +? | 0.254379 | 8.31E-31 | 0.229266 | 0.010627 | FALSE | 6.14E-28 | ++ | 0.174853 | -1 | 0.826416 | 1 | 0.004778 | 1 |
| 19-44853746-C-G | G | 0.282663 | 1.24E-08 | 0.319 | 0.361936 | 9.136E-09 | -- | 0.275362 | 5.28E-25 | 0.259787 | 0.035454 | TRUE | 3.58E-22 | -- | 0.185694 | -1 | 0.861809 | 1 | 0.014888 | 1 |
| 19-44854682-G-A | A | 0.491905 | 4.16E-08 | 0.437 | 0.038323 | 1.77E-08 | -- | 0.518775 | 6.37E-32 | 0.498369 | 0.004745 | TRUE | 2.83E-29 | -- | 0.254606 | 1 | 0.04171 | 1 | 0.075825 | 1 |
| 19-44855049-C-T | T | 0.008886 | 1.15E-10 | | | 1.146E-10 | +? | 0.009253 | 3.3E-10 | 0.006205 | 0.007583 | TRUE | 1.75E-11 | ++ | 0.011634 | 1 | 0.661747 | -1 | 0.017584 | 1 |
| 19-44856329-C-T | T | 0.289699 | 4E-11 | 0.304 | 0.192425 | 2.368E-11 | ++ | 0.257226 | 2.28E-31 | 0.266409 | 0.008669 | TRUE | 2.99E-28 | ++ | 0.586246 | -1 | 0.768932 | 1 | 0.003807 | 1 |
| 19-44856449-G-A | A | 0.287366 | 8.49E-12 | 0.303 | 0.244986 | 5.326E-12 | ++ | 0.2567 | 1.31E-31 | 0.264455 | 0.006275 | TRUE | 9.65E-29 | ++ | 0.728937 | -1 | 0.875651 | 1 | 0.003193 | 1 |
| 19-44856688-G-A | A | 0.007075 | 4.4E-11 | 0.01 | 0.164491 | 3.659E-11 | ++ | 0.010935 | 3.52E-14 | 0.00552 | 0.011575 | TRUE | 3.51E-15 | ++ | 0.180448 | 1 | 0.010606 | 1 | 0.052706 | 1 |
| 19-44860563-T-G | G | 0.006686 | 5.35E-10 | 0.021 | 0.000218 | 3.11E-10 | -- | 0.009059 | 8.21E-24 | 0.004755 | 0.01687 | TRUE | 1.58E-23 | -- | 0.68018 | 1 | 0.296554 | 1 | 0.029377 | 1 |
| 19-44862190-G-A | A | 0.016944 | 5.81E-21 | | | 5.811E-21 | +? | 0.020933 | 3.97E-50 | 0.012189 | 0.000294 | TRUE | 1.03E-48 | ++ | 0.25148 | -1 | 0.539547 | -1 | 3.36E-05 | 1 |
| 19-44863241-G-A | A | 0.005594 | 7.93E-10 | 0.022 | 0.549418 | 8.641E-10 | +- | 0.006048 | 3.63E-12 | 0.003878 | 0.532183 | TRUE | 2.95E-10 | ++ | 0.794347 | -1 | 0.877536 | -1 | 0.506369 | 1 |
| 19-44864520-T-C | C | 0.106877 | 7.74E-10 | 0.119 | 0.656235 | 7.023E-10 | -- | 0.878793 | 0.060889 | 0.907205 | 0.729206 | TRUE | 0.07928 | -- | 0.908137 | -1 | 0.028074 | -1 | 0.689016 | 1 |
| 19-44864753-G-A | A | 0.164477 | 1.48E-09 | | | 1.484E-09 | +? | 0.845015 | 0.02539 | 0.852079 | 0.532228 | TRUE | 0.02804 | ++ | 0.472557 | -1 | 0.073746 | -1 | 0.720246 | 1 |
| 19-44865946-G-A | A | 0.014731 | 1.48E-18 | 0.017 | 0.028643 | 9.3E-19 | -- | 0.032497 | 2.23E-07 | 0.024576 | 0.011587 | TRUE | 1.33E-08 | -- | 0.041232 | -1 | 0.157182 | -1 | 0.13055 | -1 |
| 19-44874887-C-T | T | 0.101185 | 1.49E-12 | 0.119 | 0.28569 | 1.038E-12 | ++ | 0.118986 | 0.017446 | 0.088857 | 0.257194 | TRUE | 0.009079 | ++ | 0.806889 | 1 | 0.01095 | 1 | 0.666021 | 1 |
| 19-44876259-G-A | A | 0.138003 | 1.6E-11 | 0.139 | 0.443667 | 1.265E-11 | ++ | 0.11993 | 0.016714 | 0.12976 | 0.319418 | TRUE | 0.01134 | ++ | 0.818735 | 1 | 0.080393 | 1 | 0.806505 | 1 |
| 19-44877704-C-A | A | 0.095717 | 4.26E-12 | 0.116 | 0.329914 | 3.115E-12 | ++ | 0.117338 | 0.016931 | 0.08626 | 0.260168 | TRUE | 0.008909 | ++ | 0.726753 | 1 | 0.010516 | 1 | 0.815409 | 1 |
| 19-44877713-T-G | G | 0.482169 | 3.24E-14 | 0.444 | 0.005632 | 7.157E-15 | -- | 0.463769 | 5.12E-17 | 0.496081 | 0.009763 | TRUE | 7.49E-17 | -- | 0.130089 | 1 | 0.010157 | -1 | 0.013276 | -1 |
| 19-44878777-G-A | A | 0.397661 | 2.72E-34 | 0.476 | 0.004984 | 2.722E-35 | ++ | | | 0.601134 | 1.07E-05 | TRUE | 1.07E-05 | ?+ | 0.505225 | -1 | 0.017389 | -1 | 0.000206 | -1 |
| 19-44879709-C-T | T | 0.458075 | 5.92E-15 | 0.377 | 0.031974 | 1.983E-15 | -- | 0.422235 | 1.01E-14 | 0.436256 | 0.000256 | FALSE | 3.39E-17 | -- | 0.529382 | 1 | 0.005414 | -1 | 0.00091 | -1 |
| 19-44879804-G-T | T | 0.458298 | 5.29E-15 | 0.378 | 0.028888 | 1.728E-15 | -- | 0.422526 | 9.6E-15 | 0.436499 | 0.000293 | FALSE | 3.84E-17 | -- | 0.554067 | 1 | 0.005774 | -1 | 0.001081 | -1 |
| 19-44881674-G-A | A | 0.009237 | 1.27E-12 | 0.016 | 0.022019 | 8.818E-13 | ++ | 0.013001 | 4.5E-20 | 0.007365 | 0.042705 | TRUE | 2.46E-19 | ++ | 0.426671 | 1 | 0.08681 | 1 | 0.052022 | 1 |
| 19-44882502-G-C | C | 0.344534 | 2.02E-31 | 0.333 | 0.003429 | 2.246E-32 | -- | | | 0.358836 | 0.15418 | TRUE | 0.1542 | ?- | 0.136154 | -1 | 0.629216 | -1 | 0.56384 | -1 |
| 19-44883210-G-GTAA | GTAA | 0.139655 | 1.6E-136 | | | 1.61E-136 | -? | 0.144627 | 4.2E-210 | 0.102932 | 2.42E-15 | TRUE | 9.1E-202 | -- | 0.716689 | 1 | 0.036926 | 1 | 3E-14 | 1 |
| 19-44883377-C-T | T | 0.032452 | 4.72E-32 | 0.055 | 0.002769 | 1.348E-32 | ++ | 0.025099 | 3.7E-52 | 0.022097 | 0.00027 | TRUE | 5.35E-49 | ++ | 0.511744 | 1 | 0.783883 | 1 | 0.000364 | 1 |
| 19-44883598-A-G | G | 0.034277 | 1.29E-14 | 0.015 | 0.375725 | 1.445E-14 | -+ | | | 0.029754 | 0.001778 | TRUE | 0.001778 | ?- | 0.010323 | 1 | 0.703341 | 1 | 0.677462 | 1 |
| 19-44884202-C-G | G | 0.139129 | 3.1E-136 | 0.247 | 2.29E-09 | 4.86E-140 | -- | 0.144515 | 3.2E-210 | 0.102507 | 2.6E-15 | TRUE | 7.7E-202 | -- | 0.694831 | 1 | 0.054895 | 1 | 1.6E-14 | 1 |
| 19-44884339-G-A | A | 0.139117 | 2E-136 | 0.247 | 1.57E-09 | 2.69E-140 | ++ | 0.144486 | 1.1E-209 | 0.102911 | 1.06E-15 | TRUE | 3.3E-202 | ++ | 0.690442 | 1 | 0.04829 | 1 | 6.11E-15 | 1 |
| 19-44884873-G-A | A | 0.14254 | 2.7E-131 | 0.25 | 2.92E-09 | 4.9E-135 | ++ | 0.148201 | 1.8E-203 | 0.106709 | 3E-14 | TRUE | 7.3E-194 | ++ | 0.532027 | 1 | 0.08142 | 1 | 1.69E-13 | 1 |
| 19-44885243-A-G | G | 0.321414 | 7.2E-120 | | | 7.23E-120 | -? | 0.219943 | 8.1E-195 | 0.270535 | 6.03E-11 | TRUE | 3.1E-174 | -- | 0.071983 | 1 | 0.574852 | 1 | 1.63E-10 | 1 |
| 19-44888197-C-T | T | 0.004619 | 1.58E-10 | | | 1.576E-10 | +? | 0.000316 | 0.559206 | 0.004223 | 0.071813 | TRUE | 0.1791 | -+ | 0.039355 | 1 | 0.545783 | -1 | 0.459474 | 1 |
| 19-44888997-C-T | T | 0.172049 | 1.2E-183 | 0.305 | 4.95E-13 | 2.33E-189 | ++ | 0.166629 | 1.8E-287 | 0.125297 | 1.25E-21 | TRUE | 2.4E-277 | ++ | 0.573295 | 1 | 0.022514 | 1 | 2.36E-20 | 1 |
| 19-44890259-C-T | T | 0.010543 | 4.88E-18 | 0.016 | 0.121432 | 3.589E-18 | ++ | 0.01148 | 1.79E-29 | 0.007058 | 0.028844 | TRUE | 1.4E-27 | ++ | 0.748027 | 1 | 0.570028 | -1 | 0.022762 | 1 |
| 19-44891079-T-C | C | 0.139143 | 6.9E-139 | 0.252 | 2.84E-10 | 4.62E-143 | -- | 0.144772 | 3.3E-214 | 0.103325 | 1.97E-16 | TRUE | 3.4E-207 | -- | 0.667842 | 1 | 0.076651 | 1 | 1.94E-15 | 1 |
| 19-44891712-T-G | G | 0.325458 | 4.6E-120 | 0.38 | 1.57E-11 | 1.44E-124 | -- | 0.217634 | 5.1E-203 | 0.275395 | 1.11E-12 | TRUE | 6.7E-184 | -- | 0.03888 | 1 | 0.456522 | 1 | 9.12E-12 | 1 |
| 19-44892073-A-G | G | 0.316874 | 7.56E-25 | 0.302 | 0.002583 | 1.001E-25 | ++ | | | 0.333282 | 0.191267 | TRUE | 0.1913 | ?+ | 0.365039 | -1 | 0.689758 | -1 | 0.582011 | -1 |
| 19-44892362-A-G | G | 0.164348 | 9E-126 | 0.259 | 2.94E-10 | 7.79E-130 | -- | 0.143531 | 8E-222 | 0.127479 | 8.91E-18 | TRUE | 8E-214 | -- | 0.205959 | 1 | 0.062663 | 1 | 9.02E-17 | 1 |
| 19-44892587-G-A | A | 0.087816 | 3.52E-83 | 0.188 | 2.46E-06 | 2.391E-85 | ++ | 0.108473 | 7.5E-165 | 0.071553 | 7.83E-12 | TRUE | 7.7E-158 | ++ | 0.672227 | -1 | 0.897143 | -1 | 4.1E-13 | 1 |
| 19-44892652-C-G | G | 0.155869 | 8.6E-131 | 0.256 | 1.41E-10 | 5.35E-135 | -- | 0.143106 | 4.2E-224 | 0.11953 | 2.89E-18 | TRUE | 2.7E-217 | -- | 0.295465 | 1 | 0.034163 | 1 | 3.96E-17 | 1 |

| | | | | | | | | | | | | | | | | | | | | |
|---|---|---|---|---|---|---|---|---|---|---|---|---|---|---|---|---|---|---|---|---|
| 19-44892887-C-T | T | 0.159618 | 2.5E-129 | 0.258 | 2.94E-10 | 1.93E-133 | ++ | 0.143155 | 1.3E-222 | 0.123134 | 4.98E-18 | TRUE | 2.6E-215 | ++ | 0.257318 | 1 | 0.041985 | 1 | 6.43E-17 | 1 |
| 19-44892962-C-T | T | 0.327305 | 9.3E-131 | 0.382 | 4.15E-11 | 2.46E-135 | ++ | 0.217558 | 6.4E-208 | 0.280219 | 3.2E-13 | TRUE | 5.8E-189 | ++ | 0.0871 | 1 | 0.204187 | 1 | 4.42E-12 | 1 |
| 19-44893416-G-T | T | 0.037826 | 2.68E-24 | 0.018 | 0.876927 | 2.646E-24 | ++ | | | 0.034266 | 0.037008 | TRUE | 0.03701 | ?+ | 0.130902 | 1 | 0.682691 | 1 | 0.526863 | 1 |
| 19-44893642-C-T | T | 0.434971 | 9.24E-14 | 0.47 | 0.024005 | 4.006E-13 | +- | 0.468873 | 8E-12 | 0.575693 | 0.284955 | TRUE | 4.9E-10 | ++ | 0.717034 | 1 | 0.276208 | -1 | 0.534695 | -1 |
| 19-44893716-G-A | A | 0.038099 | 2.61E-53 | 0.054 | 0.006838 | 5.579E-54 | ++ | 0.02617 | 6.28E-53 | 0.027074 | 9.7E-05 | TRUE | 3.72E-50 | ++ | 0.369769 | 1 | 0.346772 | 1 | 0.000236 | 1 |
| 19-44893972-G-A | A | 0.033519 | 1.64E-13 | 0.024 | 0.037303 | 1.06E-13 | -- | 0.030978 | 1.43E-13 | 0.046255 | 6.81E-05 | TRUE | 1.5E-16 | -- | 0.030025 | -1 | 0.256317 | -1 | 0.004175 | -1 |
| 19-44894050-C-T | T | 0.038078 | 2.44E-54 | 0.053 | 0.006838 | 5.136E-55 | ++ | 0.02613 | 2.09E-53 | 0.026947 | 0.000145 | TRUE | 3.28E-50 | ++ | 0.389372 | 1 | 0.443915 | 1 | 0.00026 | 1 |
| 19-44894695-T-C | C | 0.03812 | 8.12E-53 | 0.053 | 0.006838 | 1.744E-53 | -- | 0.026227 | 5.42E-54 | 0.026958 | 0.000134 | TRUE | 9.89E-51 | -- | 0.458132 | 1 | 0.506459 | 1 | 0.000229 | 1 |
| 19-44894944-C-T | T | 0.004463 | 4.66E-11 | | | 4.658E-11 | +? | 0.000326 | 0.687016 | 0.004001 | 0.14948 | TRUE | 0.2771 | -+ | 0.080388 | 1 | 0.532887 | -1 | 0.67492 | 1 |
| 19-44895007-T-C | C | 0.437278 | 3.14E-17 | 0.463 | 0.00929 | 6.807E-18 | -- | 0.463581 | 3.32E-16 | 0.575243 | 0.228368 | TRUE | 1.53E-13 | -- | 0.602239 | 1 | 0.375858 | -1 | 0.393018 | -1 |
| 19-44895208-C-G | G | 0.004405 | 4.22E-11 | | | 4.225E-11 | -? | 0.000307 | 0.40995 | 0.003999 | 0.149057 | TRUE | 0.3622 | +- | 0.079289 | 1 | 0.533011 | -1 | 0.67496 | 1 |
| 19-44895376-G-C | C | 0.102496 | 4.12E-21 | 0.127 | 0.002291 | 8.595E-22 | -- | 0.173948 | 2.96E-30 | 0.125659 | 0.00578 | TRUE | 3.06E-28 | -- | 0.486564 | -1 | 0.102625 | 1 | 0.00144 | -1 |
| 19-44895459-C-A | A | 0.428828 | 1.24E-19 | 0.47 | 0.011135 | 9.478E-19 | +- | 0.47022 | 6.39E-18 | 0.582715 | 0.26959 | TRUE | 1.26E-14 | ++ | 0.719974 | 1 | 0.438938 | -1 | 0.426088 | -1 |
| 19-44895528-C-T | T | 0.037865 | 3.47E-54 | 0.053 | 0.006838 | 7.326E-55 | ++ | 0.02612 | 8.59E-54 | 0.026677 | 0.000256 | TRUE | 4.56E-50 | ++ | 0.450098 | 1 | 0.622263 | 1 | 0.000329 | 1 |
| 19-44897468-C-T | T | 0.037839 | 6.54E-54 | 0.053 | 0.006838 | 1.386E-54 | ++ | 0.026086 | 1.97E-53 | 0.026653 | 0.000226 | TRUE | 6.94E-50 | ++ | 0.441801 | 1 | 0.622299 | 1 | 0.000262 | 1 |
| 19-44897490-T-A | A | 0.03975 | 2.77E-17 | 0.024 | 0.023627 | 1.638E-17 | -- | 0.031683 | 9.9E-14 | 0.052993 | 0.000293 | TRUE | 8.89E-16 | -- | 0.114051 | -1 | 0.195313 | -1 | 0.005946 | -1 |
| 19-44897518-T-G | G | 0.032764 | 1.11E-21 | 0.017 | 0.877704 | 1.093E-21 | -- | | | 0.02897 | 0.00685 | TRUE | 0.00685 | ?- | 0.058174 | 1 | 0.386561 | 1 | 0.747589 | 1 |
| 19-44898409-A-G | G | 0.33986 | 1.14E-38 | 0.31 | 0.000222 | 4.969E-40 | ++ | 0.390276 | 4.72E-30 | 0.363813 | 0.232396 | TRUE | 1.39E-23 | ++ | 0.634273 | -1 | 0.603999 | -1 | 0.548404 | -1 |
| 19-44899005-T-G | G | 0.041773 | 1.23E-17 | | | 1.231E-17 | +? | 0.031799 | 6E-14 | 0.056696 | 0.003113 | TRUE | 2.02E-14 | ++ | 0.131402 | -1 | 0.213311 | -1 | 0.02093 | -1 |
| 19-44899213-C-T | T | 0.031267 | 6.13E-22 | 0.017 | 0.877704 | 6.062E-22 | ++ | | | 0.027867 | 0.006487 | TRUE | 0.006487 | ?+ | 0.087434 | 1 | 0.324739 | 1 | 0.725322 | 1 |
| 19-44899925-G-A | A | 0.004524 | 3.7E-10 | | | 3.697E-10 | +? | 0.000329 | 0.242715 | 0.004089 | 0.199711 | TRUE | 0.5643 | -+ | 0.115167 | 1 | 0.498921 | -1 | 0.995879 | -1 |
| 19-44899959-C-T | T | 0.039348 | 1.7E-56 | 0.056 | 0.003468 | 3.016E-57 | ++ | 0.027815 | 1.02E-55 | 0.027794 | 0.000401 | TRUE | 2.5E-51 | ++ | 0.716285 | 1 | 0.578298 | 1 | 0.000322 | 1 |
| 19-44900155-C-T | T | 0.48187 | 6.67E-83 | 0.385 | 0.000021 | 3.798E-85 | -- | 0.433887 | 1.1E-44 | 0.510603 | 0.00203 | TRUE | 7.1E-40 | -- | 0.151311 | -1 | 0.132753 | -1 | 0.043609 | -1 |
| 19-44901174-T-C | C | 0.482148 | 4.8E-82 | 0.385 | 0.000097 | 5.139E-84 | ++ | 0.433903 | 3.13E-44 | 0.510809 | 0.002199 | TRUE | 1.92E-39 | ++ | 0.169935 | -1 | 0.1253 | -1 | 0.046147 | -1 |
| 19-44901434-G-A | A | 0.23203 | 5.85E-20 | 0.198 | 0.009801 | 1.392E-20 | -- | 0.72331 | 3.6E-29 | 0.761169 | 0.002029 | TRUE | 5.09E-28 | -- | 0.615433 | 1 | 0.342213 | 1 | 0.001906 | 1 |
| 19-44901548-AACACC | A | 0.043245 | 1.15E-55 | | | 1.152E-55 | +? | 0.028106 | 1.93E-57 | 0.031151 | 4.69E-05 | TRUE | 2.87E-54 | ++ | 0.324635 | 1 | 0.051804 | 1 | 0.000307 | 1 |
| 19-44901715-T-C | C | 0.481927 | 5.95E-82 | 0.384 | 0.000011 | 2.802E-84 | ++ | 0.433997 | 1.47E-44 | 0.510775 | 0.002393 | TRUE | 1.3E-39 | ++ | 0.171119 | -1 | 0.1272 | -1 | 0.048508 | -1 |
| 19-44901805-A-G | G | 0.490034 | 7.2E-84 | 0.389 | 0.000016 | 3.503E-86 | ++ | | | 0.519942 | 0.053184 | TRUE | 0.05318 | ?+ | 0.112809 | -1 | 0.128974 | -1 | 0.347303 | -1 |
| 19-44901924-T-C | C | 0.004367 | 2.06E-10 | | | 2.055E-10 | -? | 0.000304 | 0.235459 | 0.00393 | 0.212 | TRUE | 0.5715 | +- | 0.114894 | 1 | 0.487688 | -1 | 0.775724 | 1 |
| 19-44902264-G-C | C | 0.367813 | 1.23E-45 | 0.325 | 0.000292 | 4.689E-47 | -- | 0.390856 | 5.31E-31 | 0.393637 | 0.095291 | TRUE | 2.67E-25 | -- | 0.399686 | -1 | 0.340832 | -1 | 0.415118 | -1 |
| 19-44903416-G-A | A | 0.307464 | 4.7E-157 | 0.418 | 3.61E-09 | 1.67E-161 | ++ | 0.289288 | 1.8E-143 | 0.274355 | 3.21E-09 | TRUE | 4.1E-131 | ++ | 0.090646 | 1 | 0.00664 | 1 | 1.49E-06 | 1 |
| 19-44903861-G-A | A | 0.031769 | 6.09E-22 | 0.017 | 0.877704 | 6.023E-22 | ++ | | | 0.027882 | 0.005987 | TRUE | 0.005987 | ?+ | 0.066036 | 1 | 0.359663 | 1 | 0.615466 | 1 |
| 19-44904069-G-A | A | 0.004365 | 2.04E-10 | | | 2.035E-10 | +? | 0.000307 | 0.227301 | 0.003913 | 0.230904 | TRUE | 0.6109 | -+ | 0.116123 | 1 | 0.485175 | -1 | 0.781869 | 1 |
| 19-44904152-C-T | T | 0.004366 | 2.06E-10 | | | 2.059E-10 | +? | 0.000304 | 0.235147 | 0.003917 | 0.255783 | TRUE | 0.6394 | -+ | 0.114503 | 1 | 0.4851 | -1 | 0.971539 | 1 |
| 19-44904180-G-T | T | 0.031572 | 5.38E-22 | 0.017 | 0.877704 | 5.318E-22 | ++ | | | 0.027741 | 0.008961 | TRUE | 0.008961 | ?+ | 0.086982 | 1 | 0.342358 | 1 | 0.734607 | 1 |
| 19-44904531-G-A | A | 0.434228 | 1.38E-68 | 0.359 | 7.88E-06 | 8.172E-71 | -- | | | 0.457824 | 0.004671 | TRUE | 0.00467 | ?- | 0.357825 | -1 | 0.079174 | -1 | 0.074631 | -1 |
| 19-44905218-A-G | G | 0.029395 | 2.74E-17 | 0.013 | 0.815873 | 2.696E-17 | -- | | | 0.024372 | 0.016733 | TRUE | 0.01673 | ?- | 0.074223 | 1 | 0.884953 | 1 | 0.971892 | 1 |
| 19-44905579-G-T | T | 0.476856 | 7.48E-35 | 0.461 | 0.000725 | 4.385E-36 | ++ | 0.512748 | 4.22E-30 | 0.564826 | 0.007666 | TRUE | 1.75E-27 | ++ | 0.956185 | -1 | 0.66957 | -1 | 0.023603 | -1 |
| 19-44905910-G-C | C | 0.325301 | 4.5E-32 | 0.284 | 0.001146 | 3.983E-33 | -- | 0.641561 | 1.1E-46 | 0.671857 | 4.38E-05 | TRUE | 4.3E-45 | -- | 0.344451 | 1 | 0.363955 | 1 | 2.51E-05 | 1 |
| 19-44907187-G-A | A | 0.339032 | 9.8E-48 | 0.322 | 0.000113 | 2.699E-49 | -- | 0.394539 | 2.6E-30 | 0.378218 | 0.051863 | TRUE | 1.41E-25 | -- | 0.311312 | -1 | 0.389633 | -1 | 0.324146 | -1 |
| 19-44908684-T-C | C | 0.209742 | 1.9E-293 | 0.33 | 2.22E-16 | 7.24E-305 | -- | 0.153306 | 0 | 0.149277 | 8.37E-31 | TRUE | 3.3E-305 | -- | 0.002831 | 1 | 0.004782 | 1 | 4.96E-26 | 1 |
| 19-44908822-C-T | T | 0.05208 | 5.3E-39 | | | 5.297E-39 | -? | 0.079457 | 7.89E-31 | 0.078222 | 1.53E-07 | TRUE | 1.32E-35 | -- | 0.084588 | -1 | 0.189172 | -1 | 5.15E-06 | -1 |
| 19-44909665-AC-A | A | 0.008525 | 7.91E-14 | | | 7.909E-14 | +? | 0.007646 | 2.58E-16 | 0.005532 | 0.010115 | TRUE | 1.56E-16 | ++ | 0.103157 | 1 | 0.603036 | 1 | 0.015231 | 1 |
| 19-44909698-A-C | C | 0.041342 | 1.29E-61 | 0.06 | 0.000614 | 1.351E-62 | -- | 0.027699 | 1.92E-58 | 0.02887 | 0.000112 | TRUE | 1.42E-54 | -- | 0.288525 | 1 | 0.339988 | 1 | 0.000505 | 1 |
| 19-44910319-C-T | T | 0.185068 | 1.1E-81 | 0.278 | 1.73E-08 | 1.148E-84 | ++ | 0.201638 | 5.8E-120 | 0.168812 | 7.29E-06 | TRUE | 4E-106 | ++ | 0.470334 | -1 | 0.122162 | 1 | 1.13E-06 | 1 |
| 19-44911194-C-T | T | 0.366517 | 1.21E-39 | 0.302 | 0.000033 | 3.16E-41 | -- | 0.635333 | 2.25E-51 | 0.64679 | 0.000917 | FALSE | 5.39E-46 | -- | 0.255533 | 1 | 0.895508 | 1 | 0.000415 | 1 |
| 19-44912383-G-A | A | 0.130764 | 3.72E-11 | 0.114 | 0.741412 | 3.524E-11 | ++ | 0.110033 | 0.597661 | 0.141785 | 0.198991 | TRUE | 0.237 | -- | 0.604572 | 1 | 0.917632 | 1 | 0.039921 | -1 |
| 19-44912456-G-A | A | 0.163756 | 3.7E-141 | 0.262 | 5.69E-13 | 3.56E-146 | ++ | 0.125946 | 3.1E-289 | 0.126349 | 1.51E-20 | TRUE | 7.9E-275 | ++ | 0.649074 | 1 | 0.028477 | 1 | 8.82E-22 | 1 |
| 19-44912921-G-T | T | 0.290103 | 1E-134 | 0.372 | 1.54E-10 | 3.72E-139 | ++ | 0.236478 | 9.3E-181 | 0.262982 | 1.79E-09 | TRUE | 3.7E-161 | ++ | 0.5857 | 1 | 0.100086 | 1 | 5.87E-09 | 1 |
| 19-44913034-C-T | T | 0.301758 | 2.88E-38 | 0.304 | 0.007392 | 3.511E-39 | -- | 0.394896 | 1.05E-26 | 0.341572 | 0.009163 | TRUE | 1.09E-24 | -- | 0.282912 | -1 | 0.116165 | -1 | 0.107029 | -1 |
| 19-44913221-G-A | A | 0.36443 | 1.04E-39 | 0.299 | 0.000014 | 2.222E-41 | -- | 0.638763 | 1.4E-51 | 0.649302 | 0.000381 | FALSE | 5.23E-47 | -- | 0.236529 | 1 | 0.830514 | 1 | 0.000193 | 1 |
| 19-44913484-C-T | T | 0.299563 | 4.9E-156 | 0.379 | 1.8E-10 | 9.06E-161 | ++ | 0.236534 | 6.7E-182 | 0.272328 | 1.84E-10 | TRUE | 2.6E-164 | ++ | 0.20471 | 1 | 0.096937 | 1 | 3.49E-09 | 1 |
| 19-44914381-C-CTTC | CTTCG | 0.240913 | 3.3E-124 | | | 3.27E-124 | +? | 0.233026 | 1.1E-178 | 0.218016 | 8.53E-11 | TRUE | 1.6E-163 | ++ | 0.253734 | 1 | 0.019915 | 1 | 1.66E-09 | 1 |
| 19-44915533-T-C | C | 0.240084 | 1.7E-124 | 0.347 | 1.01E-10 | 8.74E-129 | -- | 0.233158 | 9.4E-177 | 0.217646 | 3.62E-10 | TRUE | 1.3E-160 | -- | 0.265878 | 1 | 0.128807 | 1 | 1.48E-09 | 1 |
| 19-44915704-C-T | T | 0.361185 | 3.58E-35 | 0.305 | 0.000027 | 1.114E-36 | -- | 0.637279 | 2.75E-50 | 0.705884 | 4.16E-07 | FALSE | 1.3E-53 | -- | 0.263598 | 1 | 0.840329 | -1 | 7.39E-08 | 1 |
| 19-44917843-G-A | A | 0.325755 | 2.23E-41 | 0.312 | 0.00037 | 1.001E-42 | -- | 0.415525 | 1.95E-35 | 0.359217 | 0.00121 | TRUE | 2.29E-33 | -- | 0.528163 | -1 | 0.128856 | -1 | 0.009429 | -1 |
| 19-44917947-C-T | T | 0.041551 | 4.82E-59 | 0.058 | 0.000185 | 4.119E-60 | ++ | 0.028683 | 1.28E-56 | 0.0298 | 0.001165 | TRUE | 7.28E-51 | ++ | 0.297544 | 1 | 0.908085 | 1 | 0.003648 | 1 |
| 19-44917997-G-A | A | 0.136364 | 3.8E-154 | 0.263 | 1.21E-11 | 6.77E-159 | ++ | 0.153488 | 1.7E-230 | 0.10093 | 3.32E-21 | TRUE | 5E-231 | ++ | 0.885254 | -1 | 0.153523 | 1 | 7.75E-21 | 1 |
| 19-44918620-G-A | A | 0.380255 | 1.13E-42 | 0.313 | 2.46E-06 | 1.475E-44 | -- | 0.614165 | 4.39E-65 | 0.630595 | 2.99E-05 | FALSE | 4.41E-60 | -- | 0.166131 | 1 | 0.965452 | 1 | 9.14E-06 | 1 |

| | | | | | | | | | | | | | | | | | | | | |
|---|---|---|---|---|---|---|---|---|---|---|---|---|---|---|---|---|---|---|---|---|
| 19-44918715-AG-A | A | 0.272772 | 2.64E-31 | | | 2.636E-31 | -? | 0.340413 | 5.04E-20 | 0.302388 | 0.027151 | TRUE | 1.87E-18 | -- | 0.781765 | -1 | 0.46451 | -1 | 0.063132 | -1 |
| 19-44918903-C-G | G | 0.181878 | 2.7E-221 | 0.324 | 2.44E-15 | 2.14E-228 | -- | 0.185478 | 6.4E-292 | 0.135264 | 2.61E-22 | TRUE | 1.6E-290 | -- | 0.648926 | 1 | 0.140177 | 1 | 1.16E-21 | 1 |
| 19-44919589-G-A | A | 0.207919 | 9.5E-196 | 0.336 | 4E-15 | 1.89E-202 | ++ | 0.187414 | 2.1E-291 | 0.164886 | 1.15E-23 | TRUE | 7.7E-288 | ++ | 0.11223 | 1 | 0.033648 | 1 | 1.03E-21 | 1 |
| 19-44919689-A-G | G | 0.209406 | 1.3E-195 | 0.337 | 2.66E-15 | 2.22E-202 | -- | 0.187542 | 1.5E-297 | 0.166259 | 8.06E-24 | TRUE | 3.8E-288 | -- | 0.122644 | 1 | 0.019588 | 1 | 1.03E-21 | 1 |
| 19-44920677-G-A | A | 0.034037 | 1.45E-20 | 0.019 | 0.583236 | 1.37E-20 | ++ | | | 0.029822 | 0.044055 | TRUE | 0.04405 | ?+ | 0.038046 | 1 | 0.719886 | 1 | 0.764622 | -1 |
| 19-44921558-C-CA | CA | 0.406611 | 3.35E-40 | | | 3.349E-40 | -? | | | | | FALSE | 0.00058 | ?- | | | | | | |
| 19-44922203-G-A | A | 0.376301 | 1.17E-41 | 0.313 | 2.51E-06 | 1.56E-43 | -- | 0.615754 | 1.23E-63 | 0.631704 | 6.63E-05 | FALSE | 3.52E-58 | -- | 0.164521 | 1 | 0.888165 | -1 | 1.43E-05 | 1 |
| 19-44923535-G-A | A | 0.042937 | 1.72E-24 | 0.027 | 0.000276 | 5.358E-25 | -- | 0.052879 | 1.55E-20 | 0.058784 | 0.000222 | TRUE | 4.79E-22 | -- | 0.118804 | -1 | 0.037245 | -1 | 0.007718 | -1 |
| 19-44923556-C-T | T | 0.029998 | 2.15E-16 | 0.018 | 0.894462 | 2.138E-16 | ++ | | | 0.026184 | 0.035167 | TRUE | 0.03517 | ?+ | 0.037269 | 1 | 0.602304 | 1 | 0.987192 | -1 |
| 19-44923868-T-A | A | 0.138015 | 8.8E-154 | 0.264 | 2.05E-11 | 1.84E-158 | ++ | 0.154929 | 9.4E-232 | 0.101597 | 3.29E-19 | TRUE | 2.1E-228 | ++ | 0.753479 | -1 | 0.179665 | 1 | 7.22E-19 | 1 |
| 19-44924096-C-G | G | 0.29223 | 8.28E-32 | 0.287 | 0.013624 | 1.435E-32 | ++ | | | 0.328917 | 0.007863 | TRUE | 0.007863 | ?+ | 0.613772 | -1 | 0.303449 | -1 | 0.042082 | -1 |
| 19-44924977-G-A | A | 0.158908 | 1.2E-126 | 0.277 | 5.99E-10 | 1.17E-130 | ++ | 0.15595 | 3.1E-227 | 0.126568 | 1.71E-19 | TRUE | 5.8E-223 | ++ | 0.417717 | 1 | 0.074114 | 1 | 7.97E-19 | 1 |
| 19-44925202-C-T | T | 0.267952 | 1E-30 | 0.253 | 0.039148 | 2.536E-31 | -- | 0.33382 | 9.25E-20 | 0.296825 | 0.030274 | TRUE | 3.77E-18 | -- | 0.796798 | -1 | 0.321288 | -1 | 0.093592 | -1 |
| 19-44925842-G-A | A | 0.031129 | 1.96E-13 | 0.016 | 0.618201 | 1.889E-13 | ++ | | | 0.027988 | 0.521114 | TRUE | 0.5211 | ?+ | 0.364713 | 1 | 0.815401 | 1 | 0.89554 | 1 |
| 19-44926451-G-C | C | 0.135047 | 4.49E-23 | | | 4.495E-23 | +? | 0.080172 | 9.94E-24 | 0.101823 | 0.00795 | TRUE | 3.93E-22 | ++ | 0.820464 | 1 | 0.197324 | -1 | 0.00601 | 1 |
| 19-44935297-C-T | T | 0.319903 | 5.68E-14 | 0.372 | 0.179036 | 3.048E-14 | ++ | 0.311897 | 2.84E-30 | 0.30664 | 0.016581 | TRUE | 6.18E-27 | ++ | 0.694429 | 1 | 0.092897 | 1 | 0.08067 | 1 |
| 19-44942260-G-A | A | 0.013018 | 4.61E-08 | 0.023 | 0.015442 | 3.208E-08 | ++ | 0.012733 | 3.51E-06 | 0.00945 | 0.733637 | TRUE | 0.000203 | +- | 0.504016 | 1 | 0.451943 | -1 | 0.86149 | -1 |
| 19-44960129-G-A | A | 0.011518 | 6.78E-09 | 0.021 | 0.015442 | 4.626E-09 | ++ | 0.011538 | 1.29E-06 | 0.00762 | 0.633143 | TRUE | 1.18E-05 | ++ | 0.579017 | 1 | 0.776446 | -1 | 0.666498 | 1 |
| 19-45019031-G-A | A | 0.013157 | 2.5E-09 | 0.023 | 0.0361 | 1.769E-09 | ++ | 0.012858 | 6.14E-09 | 0.008586 | 0.83386 | TRUE | 4.03E-07 | ++ | 0.251017 | 1 | 0.685761 | -1 | 0.987742 | 1 |
| 19-44825110-T-A | A | 0.335503 | 6.78E-08 | 0.374 | 0.285433 | 4.717E-08 | -- | 0.318938 | 8.07E-27 | 0.313691 | 0.085196 | TRUE | 1.55E-22 | -- | 0.138499 | -1 | 0.38616 | -1 | 0.01301 | 1 |
| 19-44887076-G-A | A | | | 0.384 | 3.05E-11 | 3.05E-11 | ?- | 0.218752 | 1.2E-203 | 0.284534 | 1.55E-12 | TRUE | 5.1E-184 | -- | 0.052854 | 1 | 0.394576 | 1 | 1.43E-11 | 1 |
| 19-44906745-G-A | A | | | 0.237 | 6.74E-13 | 6.74E-13 | ?+ | 0.121868 | 2.1E-299 | 0.081516 | 4.75E-23 | TRUE | 1.2E-291 | ++ | 0.66115 | 1 | 0.072056 | 1 | 1.19E-22 | 1 |
| 19-44893408-G-T | T | | | 0.357 | 4.58E-11 | 4.58E-11 | ?+ | 0.206646 | 1E-211 | 0.230871 | 1.83E-11 | TRUE | 1.1E-188 | ++ | 0.954286 | 1 | 0.36786 | 1 | 1.05E-12 | 1 |
| 19-44842530-T-A | A | 0.044929 | 5.95E-08 | 0.044 | 0.162193 | 4.612E-08 | ++ | 0.063457 | 3.87E-09 | 0.048582 | 0.003659 | TRUE | 7.72E-11 | ++ | 0.272888 | -1 | 0.260537 | 1 | 0.001261 | -1 |
| 19-44673285-A-G | G | 0.010153 | 0.000214 | 0.015 | 0.907453 | 0.0002124 | -- | 0.015261 | 5.78E-10 | 0.008988 | 0.114836 | TRUE | 8.58E-10 | -- | 0.065782 | 1 | 0.386419 | -1 | 0.072322 | 1 |
| 19-44718265-T-C | C | 0.11684 | 0.124115 | | | 0.1241 | +? | 0.849816 | 4.05E-08 | 0.873766 | 0.101429 | TRUE | 4.63E-08 | ++ | 0.960632 | -1 | 0.69736 | 1 | 0.012923 | -1 |
| 19-44719517-G-C | C | 0.100296 | 2.99E-06 | 0.163 | 0.959472 | 0.000003162 | +- | 0.155267 | 1.66E-09 | 0.110473 | 0.023509 | TRUE | 3.36E-10 | ++ | 0.655254 | -1 | 0.940223 | 1 | 0.011771 | 1 |
| 19-44720227-G-A | A | 0.009753 | 0.00016 | 0.014 | 0.989451 | 0.0001601 | ++ | 0.016153 | 4.98E-09 | 0.00902 | 0.004182 | TRUE | 8.6E-11 | ++ | 0.936613 | -1 | 0.119132 | 1 | 0.011599 | 1 |
| 19-44726752-CAAA-C | CAA | | | | | | | 0.199519 | 4.39E-12 | | | FALSE | 4.39E-12 | +? | | | | | | |
| 19-44728895-G-A | A | 0.01612 | 8.17E-05 | 0.022 | 0.43297 | 0.00009026 | -+ | 0.044043 | 1.15E-09 | 0.024531 | 0.001297 | TRUE | 6.23E-12 | -- | 0.359972 | -1 | 0.790892 | 1 | 0.001877 | -1 |
| 19-44731001-A-AAAA | AAAAAAAAAAAAAAAAGAAAAG | | | | | | | 0.176247 | 5.13E-09 | 0.137115 | 0.019179 | TRUE | 7.11E-10 | -- | 0.701263 | -1 | 0.85331 | -1 | 0.009775 | 1 |
| 19-44733473-C-T | T | 0.150234 | 0.000134 | | | 0.000134 | +? | 0.192209 | 4.41E-14 | 0.155906 | 0.015193 | TRUE | 2.54E-14 | ++ | 0.642182 | -1 | 0.952426 | 1 | 0.007695 | 1 |
| 19-44736813-C-CT | CT | | | | | | | 0.203724 | 3.1E-15 | 0.168553 | 0.008123 | TRUE | 1.09E-15 | ++ | 0.675958 | -1 | 0.863298 | -1 | 0.002568 | 1 |
| 19-44737327-T-C | C | 0.076974 | 2.91E-06 | 0.123 | 0.508097 | 0.000003661 | -+ | 0.132232 | 5.13E-09 | 0.082028 | 0.006218 | TRUE | 1.56E-10 | -- | 0.33757 | -1 | 0.78104 | -1 | 0.000675 | 1 |
| 19-44738381-G-A | A | 0.150957 | 4.45E-06 | | | 0.000004448 | +? | 0.189159 | 6.85E-17 | 0.154968 | 0.004637 | TRUE | 1.92E-17 | ++ | 0.631069 | -1 | 0.940102 | 1 | 0.002106 | 1 |
| 19-44738850-G-A | A | 0.196963 | 0.000405 | | | 0.0004052 | +? | 0.214871 | 9.35E-17 | 0.19012 | 0.015781 | TRUE | 2.1E-16 | ++ | 0.783738 | 1 | 0.548425 | -1 | 0.007474 | 1 |
| 19-44738916-G-A | A | 0.106923 | 0.237507 | 0.07 | 0.764314 | 0.2331 | -- | 0.076933 | 4.81E-09 | 0.106757 | 0.105996 | TRUE | 1.55E-08 | -- | 0.898287 | 1 | 0.264699 | 1 | 0.026827 | -1 |
| 19-44739483-A-G | G | 0.376569 | 0.000511 | 0.34 | 0.812599 | 0.0004962 | ++ | 0.337901 | 7.26E-09 | 0.378452 | 0.000567 | TRUE | 1.95E-11 | ++ | 0.39152 | 1 | 0.706889 | 1 | 0.000158 | -1 |
| 19-44739710-C-T | T | 0.06851 | 0.008532 | 0.051 | 0.937115 | 0.008649 | -+ | 0.074057 | 2.11E-08 | 0.065411 | 0.034097 | TRUE | 5.97E-09 | -- | 0.114802 | -1 | 0.132153 | 1 | 0.028908 | -1 |
| 19-44740137-T-TATAC | TATACACACACACACAC | | | | | | | 0.055756 | 2.47E-21 | 0.050503 | 0.004364 | TRUE | 4.02E-21 | -- | 0.532529 | 1 | 0.447522 | 1 | 0.003449 | 1 |
| 19-44743791-C-T | T | 0.499863 | 1.28E-06 | 0.404 | 0.668024 | 0.000001162 | -- | 0.38887 | 3.49E-12 | 0.495606 | 0.000491 | TRUE | 1.48E-14 | -- | 0.088206 | 1 | 0.029463 | -1 | 0.00086 | -1 |
| 19-44744370-A-G | G | 0.099563 | 0.007442 | 0.058 | 0.907108 | 0.007579 | +- | 0.074154 | 5.45E-09 | 0.084591 | 0.00134 | TRUE | 3.68E-11 | ++ | 0.068453 | -1 | 0.266442 | -1 | 0.01089 | -1 |
| 19-44747899-A-C | C | 0.492564 | 2.17E-07 | 0.413 | 0.988246 | 2.379E-07 | +- | 0.394542 | 1.28E-11 | 0.501886 | 3.08E-05 | TRUE | 2.15E-15 | ++ | 0.212134 | 1 | 0.026495 | -1 | 0.000133 | -1 |
| 19-44750234-C-T | T | 0.172332 | 0.000189 | 0.129 | 0.62859 | 0.0001742 | -- | 0.130353 | 8.15E-11 | 0.164439 | 0.010885 | TRUE | 1.68E-11 | -- | 0.954743 | 1 | 0.657921 | -1 | 0.00749 | -1 |
| 19-44750354-ATTGGC | A | | | | | | | 0.129506 | 1.06E-10 | 0.158973 | 0.00564 | TRUE | 7.72E-12 | -- | 0.934928 | 1 | 0.364222 | -1 | 0.006064 | -1 |
| 19-44752009-T-C | C | 0.057152 | 0.002004 | 0.05 | 0.980926 | 0.002023 | +- | 0.073458 | 5.19E-09 | 0.057433 | 0.013723 | TRUE | 4.74E-10 | ++ | 0.170106 | -1 | 0.151143 | 1 | 0.010436 | -1 |
| 19-44752078-GCACAC | GCACACACACACACA | | | | | | | 0.133786 | 5.87E-10 | | | FALSE | 5.87E-10 | -? | | | | | | |
| 19-44752422-G-C | C | 0.059485 | 0.003766 | 0.051 | 0.980926 | 0.0038 | -+ | 0.073869 | 4.28E-09 | 0.060196 | 0.012994 | TRUE | 3.83E-10 | -- | 0.096376 | -1 | 0.143189 | 1 | 0.013136 | -1 |
| 19-44766291-C-T | T | 0.005244 | 0.000496 | 0.01 | 0.369801 | 0.0004719 | ++ | 0.006525 | 4.13E-10 | 0.004316 | 0.032331 | TRUE | 1.07E-10 | ++ | 0.011962 | 1 | 0.882637 | -1 | 0.058322 | 1 |
| 19-44766468-T-C | C | 0.013414 | 4.89E-06 | 0.029 | 0.034409 | 0.000003711 | -- | 0.023975 | 3.62E-10 | 0.013643 | 0.38753 | TRUE | 4.92E-09 | -- | 0.938865 | -1 | 0.43945 | -1 | 0.276766 | 1 |
| 19-44775001-G-A | A | 0.003749 | 1.56E-06 | 0.015 | 0.078559 | 0.000001298 | ++ | 0.005544 | 1.06E-09 | 0.002826 | 0.990993 | TRUE | 6.86E-08 | ++ | 0.636967 | 1 | 0.070249 | -1 | 0.743442 | 1 |
| 19-44784672-CTCCAT | C | 0.004814 | 0.000101 | | | 0.0001013 | -? | 0.007185 | 8.18E-10 | 0.003829 | 0.553701 | TRUE | 1.39E-08 | -- | 0.68102 | 1 | 0.726444 | 1 | 0.558594 | 1 |
| 19-44785532-GTAGAT | GTAGATAGATAGATAGA | | | | | | | 0.139857 | 1.84E-12 | | | FALSE | 1.84E-12 | +? | | | | | | |
| 19-44785532-GTAGAT | GTAGATAGATAGATAGATAGA | | | | | | | 0.512572 | 1.2E-08 | | | FALSE | 1.2E-08 | -? | | | | | | |
| 19-44799020-C-T | T | 0.006177 | 1.23E-07 | 0.016 | 0.000213 | 7.851E-08 | ++ | 0.008297 | 1.16E-20 | 0.004517 | 0.189998 | TRUE | 1.35E-18 | ++ | 0.96117 | -1 | 0.335502 | 1 | 0.233215 | 1 |
| 19-44807827-G-A | A | 0.018732 | 3.85E-07 | 0.031 | 0.025901 | 2.738E-07 | ++ | 0.026026 | 1.68E-10 | 0.01527 | 0.211157 | TRUE | 8.68E-10 | ++ | 0.465499 | 1 | 0.728851 | -1 | 0.21651 | 1 |
| 19-44812964-A-G | G | | | | | | | 0.014006 | 4.84E-24 | 0.008113 | 0.070312 | TRUE | 3.54E-22 | -- | 0.191915 | 1 | 0.325106 | -1 | 0.074276 | 1 |
| 19-44815238-A-G | G | 0.003528 | 0.000721 | 0.006 | 0.369801 | 0.0006873 | -- | 0.003702 | 1.02E-10 | 0.002455 | 0.011046 | TRUE | 6.67E-12 | -- | 0.014174 | 1 | 0.318379 | -1 | 0.012993 | 1 |
| 19-44816374-G-A | A | 0.015142 | 7.66E-07 | 0.016 | 0.101854 | 6.472E-07 | -- | 0.04533 | 4.21E-12 | 0.024865 | 0.00329 | TRUE | 8.95E-14 | -- | 0.138094 | -1 | 0.944947 | 1 | 0.005038 | -1 |

| | | | | | | | | | | | | | | | | | | | | |
|---|---|---|---|---|---|---|---|---|---|---|---|---|---|---|---|---|---|---|---|---|
| 19-44819414-C-T | T | 0.002104 | 0.004805 | 0.003 | 0.150167 | 0.005085 | +- | 0.001391 | 3.55E-08 | 0.001399 | 0.194619 | TRUE | 1.54E-07 | ++ | 0.004776 | 1 | 0.236155 | 1 | 0.380618 | 1 |
| 19-44819913-CCATCC | C | | | | | | | 0.535363 | 4.29E-22 | 0.597525 | 0.014798 | TRUE | 1.2E-20 | -- | 0.585952 | 1 | 0.373509 | 1 | 0.033592 | 1 |
| 19-44821499-C-G | G | | | 0.034 | 0.068397 | 0.0684 | ?- | 0.020864 | 2.51E-44 | 0.011563 | 0.002044 | TRUE | 2.88E-42 | -- | 0.565592 | -1 | 0.682072 | -1 | 0.000451 | 1 |
| 19-44825150-GAC-G | G | | | | | | | 0.463606 | 5.03E-16 | 0.54573 | 0.085684 | TRUE | 2.48E-14 | ++ | 0.525255 | -1 | 0.449258 | -1 | 0.035229 | 1 |
| 19-44827846-A-G | G | | | | | | | 0.557319 | 9.15E-10 | 0.61602 | 0.017058 | TRUE | 1.68E-10 | -- | 0.293279 | -1 | 0.592199 | 1 | 0.009613 | 1 |
| 19-44828285-G-C | C | | | 0.01 | 0.00099 | 0.00099 | ?+ | 0.005951 | 3.46E-12 | | | FALSE | 3.47E-12 | +? | | | | | | |
| 19-44829763-TAA-TA | TA | | | | | | | 0.30133 | 1.68E-23 | | | FALSE | 1.69E-23 | +? | | | | | | |
| 19-44829763-T-TA | TA | | | | | | | 0.228419 | 1.06E-08 | 0.24761 | 0.036301 | TRUE | 4.3E-09 | ++ | 0.858971 | 1 | 0.648736 | -1 | 0.015569 | -1 |
| 19-44831701-T-A | A | | | | | | | 0.001951 | 2.64E-08 | | | FALSE | 2.64E-08 | +? | | | | | | |
| 19-44832419-A-G | G | 0.461257 | 0.00484 | 0.486 | 0.859281 | 0.005204 | -+ | 0.441462 | 8.44E-21 | 0.458573 | 0.009124 | TRUE | 5.33E-20 | -- | 0.625559 | -1 | 0.939754 | 1 | 0.006454 | 1 |
| 19-44832778-T-C | C | 0.339683 | 1.3E-05 | 0.343 | 0.732661 | 0.00001223 | -- | 0.301369 | 2.59E-24 | 0.318772 | 0.007045 | TRUE | 6.74E-23 | -- | 0.617358 | -1 | 0.969931 | 1 | 0.003305 | 1 |
| 19-44833186-G-A | A | 0.318714 | 2.94E-05 | 0.338 | 0.747984 | 0.0000278 | ++ | 0.299412 | 4.36E-24 | 0.299853 | 0.006242 | TRUE | 7.13E-23 | ++ | 0.400845 | -1 | 0.858785 | 1 | 0.002958 | 1 |
| 19-44833336-C-T | T | 0.004816 | 0.000333 | 0.005 | 0.563703 | 0.0003382 | +- | 0.004602 | 4.09E-14 | 0.003849 | 0.002434 | TRUE | 1.09E-15 | ++ | 0.053281 | 1 | 0.876097 | 1 | 0.010657 | 1 |
| 19-44834128-G-C | C | | | | | | | 0.298275 | 4.21E-24 | 0.286094 | 0.006474 | TRUE | 6.1E-23 | ++ | 0.054268 | -1 | 0.722139 | 1 | 0.002576 | 1 |
| 19-44834247-G-C | C | 0.340047 | 4.21E-05 | 0.361 | 0.752752 | 0.00004 | ++ | 0.314491 | 1.21E-20 | 0.321933 | 0.022191 | TRUE | 4.36E-19 | ++ | 0.214147 | -1 | 0.99803 | 1 | 0.006594 | 1 |
| 19-44834661-G-A | A | | | 0.058 | 0.233314 | 0.2333 | ?+ | 0.035047 | 4.52E-63 | 0.019081 | 0.000683 | TRUE | 3.39E-59 | ++ | 0.083366 | -1 | 0.758408 | -1 | 6.81E-05 | 1 |
| 19-44834993-AT-A | A | 0.301237 | 1.83E-06 | | | 0.000001829 | +? | 0.298046 | 1.5E-23 | 0.280999 | 0.006143 | TRUE | 1.53E-22 | ++ | 0.044423 | -1 | 0.685034 | 1 | 0.002043 | 1 |
| 19-44835266-G-C | C | 0.285444 | 3.95E-07 | 0.323 | 0.747457 | 3.752E-07 | ++ | 0.297416 | 1.01E-23 | 0.264412 | 0.007056 | FALSE | 1.23E-22 | ++ | 0.042549 | -1 | 0.861713 | 1 | 0.002743 | 1 |
| 19-44836881-G-A | A | 0.308562 | 0.000102 | 0.285 | 0.222332 | 0.00007397 | ++ | 0.692664 | 4.87E-16 | 0.668278 | 0.144106 | TRUE | 9.29E-14 | ++ | 0.265904 | -1 | 0.978768 | -1 | 0.108581 | 1 |
| 19-44837479-G-A | A | 0.316506 | 1.09E-05 | 0.336 | 0.704428 | 0.0000101 | ++ | 0.299687 | 1.55E-23 | 0.29727 | 0.013334 | TRUE | 7.54E-22 | ++ | 0.033099 | -1 | 0.73246 | 1 | 0.003145 | 1 |
| 19-44838512-C-CAA | CAA | | | | | | | 0.011488 | 2.23E-16 | | | FALSE | 2.23E-16 | +? | | | | | | |
| 19-44838647-G-A | A | 0.316976 | 1.45E-05 | 0.336 | 0.704428 | 0.00001349 | ++ | 0.299703 | 8.89E-24 | 0.297664 | 0.010368 | TRUE | 2.9E-22 | ++ | 0.034233 | -1 | 0.758377 | 1 | 0.002111 | 1 |
| 19-44838691-A-G | G | 0.28068 | 0.009062 | | | 0.009062 | -? | 0.699273 | 5.65E-17 | 0.698793 | 0.101298 | TRUE | 7.28E-15 | -- | 0.938557 | 1 | 0.733841 | -1 | 0.146217 | 1 |
| 19-44839902-CTATAT, | CTATATATATATATA | | | | | | | 0.164915 | 1.34E-13 | | | FALSE | 1.34E-13 | +? | | | | | | |
| 19-44839902-CTATAT, | CTATATATATATATATA | | | | | | | 0.037586 | 1.12E-30 | | | FALSE | 1.12E-30 | +? | | | | | | |
| 19-44839902-CTATAT, | CTATATATATATATATATATATATA | | | | | | | 0.038009 | 3.82E-10 | | | FALSE | 3.82E-10 | -? | | | | | | |
| 19-44840476-GAAAA/ | GAAAAAAAA | | | | | | | 0.011193 | 2.23E-14 | | | FALSE | 2.23E-14 | +? | | | | | | |
| 19-44840782-CAA-CA | CA | | | | | | | 0.302705 | 2.21E-22 | | | FALSE | 2.21E-22 | +? | | | | | | |
| 19-44841241-T-TTAA/ | TTAAAAA | | | | | | | 0.010354 | 7.35E-16 | 0.0137 | 0.466616 | TRUE | 5.99E-13 | -- | 0.683337 | -1 | 0.466968 | -1 | 0.240408 | 1 |
| 19-44842026-A-C | C | 0.014187 | 3.22E-05 | 0.019 | 0.2733 | 0.00002925 | -- | 0.010615 | 4.9E-17 | 0.013023 | 0.180988 | TRUE | 6.36E-15 | -- | 0.407003 | 1 | 0.263739 | -1 | 0.232231 | 1 |
| 19-44844304-T-G | G | 0.15272 | 0.683014 | 0.146 | 0.813398 | 0.6935 | -+ | 0.830597 | 5.33E-09 | 0.844247 | 0.113785 | TRUE | 1.2E-08 | -- | 0.585196 | -1 | 0.956153 | -1 | 0.115765 | 1 |
| 19-44844403-G-GA | GA | | | | | | | 0.439119 | 3.74E-11 | 0.41386 | 0.432262 | TRUE | 3.09E-09 | ++ | 0.535698 | 1 | 0.198767 | 1 | 0.10888 | -1 |
| 19-44844654-C-A | A | 0.45807 | 0.020674 | 0.466 | 0.586396 | 0.02366 | +- | 0.415965 | 9.05E-11 | 0.452015 | 0.066257 | TRUE | 2.24E-10 | ++ | 0.33731 | -1 | 0.657508 | -1 | 0.019409 | 1 |
| 19-44846145-T-C | C | | | | | | | 0.415378 | 8.25E-11 | | | FALSE | 8.25E-11 | -? | | | | | | |
| 19-44849230-A-T | T | 0.435319 | 0.0023 | 0.451 | 0.754235 | 0.002585 | -+ | 0.405086 | 1.08E-12 | 0.427041 | 0.019276 | TRUE | 8.31E-13 | -- | 0.633387 | -1 | 0.952454 | 1 | 0.009392 | 1 |
| 19-44850787-T-G | G | | | 0.451 | 0.066974 | 0.06697 | ?+ | 0.510847 | 5.34E-10 | 0.432926 | 0.351864 | TRUE | 1.93E-08 | ++ | 0.675078 | -1 | 0.975218 | -1 | 0.304202 | -1 |
| 19-44850981-G-C | C | | | 0.163 | 0.955356 | 0.9554 | ?+ | 0.815806 | 4.12E-10 | 0.819759 | 0.025487 | TRUE | 1.63E-10 | ++ | 0.616897 | -1 | 0.286738 | 1 | 0.078386 | 1 |
| 19-44852010-T-A | A | 0.025397 | 0.003259 | 0.025 | 0.411093 | 0.003059 | ++ | 0.011267 | 4.67E-15 | 0.025563 | 0.313311 | TRUE | 7.51E-12 | ++ | 0.59761 | 1 | 0.555514 | -1 | 0.334814 | 1 |
| 19-44852378-A-C | C | 0.003782 | 1.52E-07 | 0.009 | 0.902554 | 1.506E-07 | -- | 0.004063 | 4.37E-09 | 0.00249 | 0.630737 | TRUE | 1.53E-06 | -+ | 0.657358 | -1 | 0.151849 | -1 | 0.946876 | -1 |
| 19-44852435-GTT-GT | GT | | | | | | | 0.683561 | 3.42E-10 | | | FALSE | 3.43E-10 | +? | | | | | | |
| 19-44852486-A-C | C | 0.18156 | 0.000182 | 0.136 | 0.305277 | 0.0001444 | ++ | 0.174368 | 1.05E-08 | 0.182928 | 0.091571 | TRUE | 1.53E-08 | ++ | 0.11482 | -1 | 0.005997 | -1 | 0.646167 | -1 |
| 19-44852526-CAA-CA | CA | | | | | | | 0.279576 | 5.61E-37 | | | FALSE | 5.62E-37 | +? | | | | | | |
| 19-44853495-C-CT | CT | | | | | | | 0.260263 | 4.74E-30 | 0.28171 | 0.005881 | FALSE | 6.23E-28 | ++ | 0.36473 | -1 | 0.27492 | 1 | 0.002791 | 1 |
| 19-44854034-G-C | C | | | | | | | 0.259019 | 1.54E-29 | 0.247385 | 0.011312 | TRUE | 9.02E-27 | ++ | 0.353251 | -1 | 0.639862 | 1 | 0.00589 | 1 |
| 19-44854120-T-C | C | | | 0.289 | 0.187292 | 0.1873 | ?- | 0.254335 | 6.39E-31 | 0.229539 | 0.007272 | FALSE | 2.57E-28 | -- | 0.178522 | -1 | 0.787517 | 1 | 0.003276 | 1 |
| 19-44854769-C-T | T | 0.016401 | 0.000978 | 0.02 | 0.178049 | 0.000884 | ++ | 0.010691 | 5.81E-16 | 0.015366 | 0.159859 | TRUE | 6.08E-14 | ++ | 0.473716 | 1 | 0.423044 | -1 | 0.257213 | 1 |
| 19-44855096-T-C | C | | | | | | | 0.518712 | 3.32E-32 | | | FALSE | 3.32E-32 | -? | | | | | | |
| 19-44856410-A-G | G | 0.014344 | 6.41E-06 | 0.02 | 0.139607 | 0.000005607 | -- | 0.01054 | 1.49E-16 | 0.012748 | 0.153395 | TRUE | 1.09E-14 | -- | 0.276259 | 1 | 0.38135 | -1 | 0.322824 | 1 |
| 19-44857231-T-G | G | 0.019646 | 2.87E-05 | | | 0.00002872 | -? | 0.011091 | 5.95E-17 | | | FALSE | 5.95E-17 | -? | | | | | | |
| 19-44858325-C-CT | CT | | | | | | | 0.178323 | 9.74E-11 | 0.156348 | 0.067805 | TRUE | 2.04E-10 | -- | 0.816425 | -1 | 0.144267 | -1 | 0.104482 | -1 |
| 19-44858389-A-G | G | 0.151303 | 4.55E-06 | 0.117 | 0.055506 | 0.000002902 | ++ | 0.118948 | 2.49E-16 | 0.153998 | 0.045724 | TRUE | 9.82E-15 | ++ | 0.29679 | 1 | 0.959407 | 1 | 0.003971 | -1 |
| 19-44858568-T-C | C | 0.314063 | 0.001655 | | | 0.001655 | -? | 0.369268 | 1.57E-18 | 0.338408 | 0.001459 | TRUE | 1.84E-19 | -- | 0.251523 | -1 | 0.172095 | 1 | 0.000504 | 1 |
| 19-44858703-T-C | C | 0.299138 | 0.000174 | 0.377 | 0.121502 | 0.0001149 | -- | 0.368407 | 1.31E-18 | 0.321207 | 0.002519 | TRUE | 3.5E-19 | -- | 0.16005 | -1 | 0.134133 | 1 | 0.000962 | 1 |
| 19-44859012-G-T | T | 0.292183 | 4.95E-05 | 0.372 | 0.097595 | 0.00003081 | ++ | 0.367145 | 9.07E-19 | 0.31265 | 0.001636 | TRUE | 1.29E-19 | ++ | 0.178226 | -1 | 0.10958 | 1 | 0.000748 | 1 |
| 19-44859410-G-A | A | 0.298055 | 0.000189 | 0.374 | 0.10701 | 0.0001232 | ++ | 0.367617 | 1.33E-18 | 0.319552 | 0.001739 | TRUE | 2.01E-19 | ++ | 0.180943 | -1 | 0.118404 | 1 | 0.000727 | 1 |
| 19-44859997-G-A | A | 0.29722 | 0.000378 | 0.373 | 0.114289 | 0.0002523 | ++ | 0.36674 | 2E-18 | 0.318346 | 0.001734 | TRUE | 2.81E-19 | ++ | 0.173703 | -1 | 0.110268 | 1 | 0.000727 | 1 |
| 19-44860443-A-G | G | 0.192889 | 0.000793 | 0.158 | 0.417284 | 0.0006718 | ++ | 0.195952 | 2.84E-12 | 0.19251 | 0.064 | TRUE | 1.27E-11 | ++ | 0.729703 | 1 | 0.157221 | -1 | 0.054685 | -1 |
| 19-44860534-T-G | G | 0.299657 | 0.001066 | 0.373 | 0.09479 | 0.0007124 | -- | 0.367073 | 1.38E-18 | 0.320645 | 0.001355 | TRUE | 1.44E-19 | -- | 0.146075 | -1 | 0.051396 | 1 | 0.000702 | 1 |
| 19-44861366-C-T | T | 0.296066 | 0.000491 | 0.372 | 0.09479 | 0.0003212 | ++ | 0.367724 | 8.37E-19 | 0.317249 | 0.001636 | TRUE | 1.23E-19 | ++ | 0.114818 | -1 | 0.061856 | 1 | 0.000771 | 1 |

| | | | | | | | | | | | | | | | | | | | | |
|---|---|---|---|---|---|---|---|---|---|---|---|---|---|---|---|---|---|---|---|---|
| 19-44861558-A-G | G | 0.295992 | 0.000471 | 0.373 | 0.09479 | 0.000308 | -- | 0.367771 | 6.54E-19 | 0.31726 | 0.001624 | TRUE | 9.84E-20 | -- | 0.114124 | -1 | 0.062461 | 1 | 0.00077 | 1 |
| 19-44861785-A-G | G | 0.003761 | 1.29E-05 | 0.01 | 0.159854 | 0.00001178 | -- | 0.00473 | 1.25E-11 | 0.00269 | 0.085424 | TRUE | 1.43E-11 | -- | 0.992527 | -1 | 0.422985 | -1 | 0.047103 | 1 |
| 19-44861815-A-G | G | 0.29602 | 0.000575 | 0.373 | 0.09479 | 0.000378 | -- | 0.367761 | 7.15E-19 | 0.317314 | 0.001389 | TRUE | 8.36E-20 | -- | 0.131135 | -1 | 0.059789 | 1 | 0.000708 | 1 |
| 19-44861991-T-C | C | 0.295424 | 0.000733 | 0.373 | 0.09479 | 0.0004852 | -- | 0.367752 | 9.15E-19 | 0.316545 | 0.001671 | TRUE | 1.36E-19 | -- | 0.112562 | -1 | 0.059384 | 1 | 0.000806 | 1 |
| 19-44862268-CACTGT | C | | | | | | | 0.367201 | 5.57E-19 | 0.000366 | 0.717028 | TRUE | 5.23E-19 | -- | 0.462977 | -1 | 0.684031 | 1 | 0.910619 | 1 |
| 19-44862293-T-G | G | | | | | | | 0.367201 | 5.57E-19 | | | FALSE | 5.57E-19 | -? | | | | | | |
| 19-44862384-A-C | C | 0.295786 | 0.000442 | | | 0.0004417 | -? | 0.367718 | 8.15E-19 | 0.317874 | 0.001187 | TRUE | 7.29E-20 | -- | 0.159133 | -1 | 0.04945 | 1 | 0.000797 | 1 |
| 19-44862475-A-AAAG | AAAG | | | | | | | 0.367994 | 9.83E-19 | 0.317348 | 0.001567 | TRUE | 1.31E-19 | -- | 0.10954 | -1 | 0.070904 | 1 | 0.000671 | 1 |
| 19-44862560-G-A | A | | | | | | | 0.368059 | 1.2E-18 | 0.317632 | 0.001464 | TRUE | 1.4E-19 | ++ | 0.120154 | -1 | 0.07458 | 1 | 0.000644 | 1 |
| 19-44863088-G-A | A | 0.295705 | 0.001178 | 0.373 | 0.109569 | 0.0008026 | ++ | 0.367833 | 1.45E-18 | 0.316956 | 0.001353 | TRUE | 1.45E-19 | ++ | 0.09824 | -1 | 0.070564 | 1 | 0.000509 | 1 |
| 19-44863346-G-A | A | 0.294291 | 0.000149 | 0.374 | 0.139449 | 0.0001002 | ++ | 0.368432 | 7.52E-19 | 0.316315 | 0.001637 | TRUE | 1.1E-19 | ++ | 0.144137 | -1 | 0.156826 | 1 | 0.000511 | 1 |
| 19-44863522-T-C | C | 0.29418 | 0.000197 | 0.374 | 0.139449 | 0.0001335 | -- | 0.368447 | 9.43E-19 | 0.316162 | 0.001713 | TRUE | 1.44E-19 | -- | 0.136396 | -1 | 0.153182 | 1 | 0.000538 | 1 |
| 19-44864245-G-C | C | 0.154836 | 2.32E-06 | 0.127 | 0.313437 | 0.000001784 | -- | 0.178503 | 2.71E-10 | 0.153057 | 0.033944 | TRUE | 1.55E-10 | -- | 0.79746 | -1 | 0.028655 | -1 | 0.089244 | -1 |
| 19-44864825-C-A | A | 0.292494 | 0.000141 | | | 0.0001408 | +? | 0.36851 | 1.34E-18 | 0.314492 | 0.002168 | TRUE | 2.76E-19 | ++ | 0.121592 | -1 | 0.131741 | 1 | 0.000654 | 1 |
| 19-44866173-TAA-TA | TA | | | | | | | 0.180209 | 3.6E-10 | | | FALSE | 3.6E-10 | -? | | | | | | |
| 19-44866307-AG-A | A | 0.005068 | 7.4E-08 | | | 7.399E-08 | +? | 0.007271 | 1.82E-11 | 0.004102 | 0.401822 | TRUE | 3.43E-10 | ++ | 0.345879 | -1 | 0.631629 | -1 | 0.27877 | 1 |
| 19-44867313-A-C | C | | | 0.374 | 0.135081 | 0.1351 | ?+ | 0.633395 | 5.65E-19 | 0.686514 | 0.001885 | TRUE | 1.08E-19 | ++ | 0.19727 | 1 | 0.117647 | -1 | 0.000732 | -1 |
| 19-44867392-A-G | G | 0.300639 | 0.000112 | 0.38 | 0.210058 | 0.0000802 | ++ | 0.633119 | 6.06E-19 | 0.678483 | 0.000862 | TRUE | 3.61E-20 | ++ | 0.650918 | 1 | 0.12761 | -1 | 0.000813 | -1 |
| 19-44867416-C-A | A | 0.301173 | 0.000152 | 0.38 | 0.210058 | 0.0001096 | -- | 0.633016 | 7E-19 | 0.677315 | 0.000889 | TRUE | 4.3E-20 | -- | 0.609642 | 1 | 0.145312 | -1 | 0.000697 | -1 |
| 19-44867581-G-A | A | 0.157189 | 1.45E-06 | 0.132 | 0.310136 | 0.000001111 | -- | 0.18259 | 2.74E-10 | 0.1573 | 0.023362 | TRUE | 8.96E-11 | -- | 0.485345 | -1 | 0.022434 | -1 | 0.083336 | -1 |
| 19-44868428-A-AT | AT | 0.187339 | 5.58E-07 | | | 5.583E-07 | +? | 0.188286 | 5.86E-11 | 0.189974 | 0.043718 | TRUE | 7.76E-11 | ++ | 0.938153 | -1 | 0.037843 | -1 | 0.104811 | -1 |
| 19-44873636-A-AAC | AAC | | | | | | | 0.07974 | 1.46E-65 | 0.100295 | 0.008022 | TRUE | 3.43E-54 | -- | 0.551783 | -1 | 0.221428 | -1 | 6.48E-05 | 1 |
| 19-44874077-G-A | A | | | | | | | 0.359914 | 2.8E-12 | 0.354529 | 0.029524 | TRUE | 3.21E-12 | -- | 0.821355 | 1 | 0.747174 | -1 | 0.010056 | -1 |
| 19-44875268-C-CT | CT | | | | | | | 0.191988 | 9.05E-10 | 0.208391 | 0.030745 | TRUE | 4.38E-10 | -- | 0.714781 | 1 | 0.064899 | -1 | 0.047339 | -1 |
| 19-44875803-A-C | C | 0.362039 | 0.169046 | 0.311 | 0.09056 | 0.1333 | ++ | 0.359304 | 3.53E-12 | 0.365364 | 0.018995 | TRUE | 1.96E-12 | ++ | 0.427161 | 1 | 0.390012 | -1 | 0.005592 | -1 |
| 19-44879418-G-A | A | | | 0.017 | 0.030732 | 0.03073 | ?- | 0.047091 | 9.03E-16 | 0.028789 | 0.012804 | TRUE | 5.53E-16 | -- | 0.665869 | -1 | 0.834752 | 1 | 0.012903 | -1 |
| 19-44879460-G-A | A | 0.349684 | 3.58E-07 | 0.408 | 0.045949 | 1.714E-07 | -- | 0.618341 | 1.28E-28 | 0.642879 | 0.000157 | TRUE | 2.29E-29 | -- | 0.719688 | 1 | 0.234461 | -1 | 5.9E-05 | -1 |
| 19-44879727-T-G | G | 0.078263 | 0.006267 | 0.089 | 0.047336 | 0.00468 | ++ | 0.093389 | 5.24E-10 | 0.086195 | 0.779466 | TRUE | 1.54E-07 | ++ | 0.457404 | 1 | 0.036815 | 1 | 0.218434 | -1 |
| 19-44879780-A-T | T | 0.078254 | 0.006152 | 0.089 | 0.047336 | 0.004591 | ++ | 0.09338 | 5.45E-10 | 0.086175 | 0.763783 | TRUE | 1.48E-07 | ++ | 0.454484 | 1 | 0.03667 | 1 | 0.21035 | -1 |
| 19-44880774-AT-A | A | | | | | | | 0.013018 | 8.08E-14 | 0.014382 | 0.052561 | TRUE | 2.85E-13 | ++ | 0.432151 | -1 | 0.504861 | -1 | 0.011964 | 1 |
| 19-44880859-G-A | A | 0.280656 | 1.1E-06 | 0.372 | 0.011958 | 4.532E-07 | -- | 0.638231 | 1.48E-24 | 0.698778 | 0.000231 | TRUE | 8.01E-26 | -- | 0.302159 | 1 | 0.304662 | -1 | 2.81E-05 | -1 |
| 19-44881148-T-C | C | | | 0.111 | 0.025027 | 0.02503 | ?+ | 0.147455 | 3.75E-25 | 0.114337 | 0.033998 | TRUE | 1.58E-22 | ++ | 0.263518 | 1 | 0.089962 | 1 | 0.002544 | -1 |
| 19-44881523-CAA-CA | CA | | | | | | | 0.212392 | 1.43E-94 | | | FALSE | 1.43E-94 | +? | | | | | | |
| 19-44882099-C-A | A | | | 0.018 | 0.168281 | 0.1683 | ?+ | 0.011849 | 3.1E-28 | 0.007395 | 0.028966 | TRUE | 1.68E-26 | ++ | 0.863647 | 1 | 0.47847 | -1 | 0.019817 | 1 |
| 19-44882783-ATTT-AT | AT | | | | | | | 0.162845 | 1.2E-181 | | | FALSE | 1.2E-181 | +? | | | | | | |
| 19-44882783-ATTT-AT | ATT | | | | | | | 0.748023 | 8.8E-130 | | | FALSE | 8.8E-130 | -? | | | | | | |
| 19-44885311-T-C | C | | | | | | | 0.216784 | 2.2E-194 | | | FALSE | 2.2E-194 | -? | | | | | | |
| 19-44885324-T-A | A | | | | | | | 0.02493 | 2.79E-52 | | | FALSE | 2.79E-52 | +? | | | | | | |
| 19-44885325-T-A | A | | | | | | | 0.02493 | 2.79E-52 | | | FALSE | 2.79E-52 | +? | | | | | | |
| 19-44885327-TTTAA-T | T | | | | | | | 0.02493 | 2.79E-52 | | | FALSE | 2.79E-52 | +? | | | | | | |
| 19-44885328-T-A | A | | | | | | | 0.196048 | 6.9E-136 | | | FALSE | 6.9E-136 | +? | | | | | | |
| 19-44885329-T-A | A | | | | | | | 0.195979 | 3.3E-137 | | | FALSE | 3.3E-137 | +? | | | | | | |
| 19-44885330-A-T | T | | | | | | | 0.195456 | 9.6E-136 | | | FALSE | 9.6E-136 | -? | | | | | | |
| 19-44885331-A-T | T | | | | | | | 0.19655 | 3.1E-137 | | | FALSE | 3.2E-137 | -? | | | | | | |
| 19-44886339-G-A | A | | | | | | | 0.033584 | 6.75E-13 | 0.060033 | 0.000436 | TRUE | 8.43E-15 | -- | 0.507062 | -1 | 0.1003 | -1 | 0.00507 | -1 |
| 19-44889415-ATTTT-A | ATTT | | | | | | | 0.030046 | 2.18E-11 | | | FALSE | 2.19E-11 | -? | | | | | | |
| 19-44890947-G-A | A | 0.002692 | 3.9E-06 | 0.005 | 0.005052 | 0.000003295 | ++ | 0.003029 | 1.49E-11 | 0.001756 | 0.2552 | TRUE | 1.17E-10 | ++ | 0.291977 | -1 | 0.194786 | -1 | 0.121472 | 1 |
| 19-44892009-G-A | A | | | | | | | 0.606171 | 8.95E-64 | 0.677097 | 8.51E-06 | TRUE | 1.93E-60 | ++ | 0.5532 | 1 | 0.973994 | 1 | 6.96E-07 | 1 |
| 19-44892457-T-C | C | | | | | | | 0.219044 | 1.4E-207 | 0.282366 | 7.23E-13 | TRUE | 8.8E-188 | -- | 0.050086 | 1 | 0.309697 | 1 | 7.83E-12 | 1 |
| 19-44894255-A-C | C | | | | | | | 0.463485 | 6.49E-16 | 0.579236 | 0.197835 | TRUE | 1.6E-13 | ++ | 0.686033 | 1 | 0.353722 | -1 | 0.393497 | -1 |
| 19-44894261-C-CTT | CTT | | | | | | | 0.382053 | 3.09E-10 | 0.48293 | 0.007278 | TRUE | 2.25E-11 | -- | 0.191813 | -1 | 0.100179 | -1 | 0.10034 | -1 |
| 19-44896087-G-T | T | | | 0.053 | 0.006838 | 0.006838 | ?+ | 0.026117 | 2.46E-53 | 0.026842 | 0.000188 | TRUE | 6.06E-50 | ++ | 0.368119 | 1 | 0.620988 | 1 | 0.000278 | 1 |
| 19-44896639-G-A | A | | | 0.053 | 0.006838 | 0.006838 | ?+ | 0.026143 | 2.08E-53 | 0.026854 | 0.000213 | TRUE | 6.77E-50 | ++ | 0.429077 | 1 | 0.628688 | 1 | 0.000264 | 1 |
| 19-44897227-T-TG | TG | | | | | | | 0.026324 | 8.46E-55 | 0.028648 | 1.95E-05 | TRUE | 7.97E-53 | -- | 0.071148 | 1 | 0.24703 | 1 | 0.000236 | 1 |
| 19-44897776-C-CA | CA | | | | | | | 0.398268 | 3.55E-31 | 0.44433 | 0.408197 | TRUE | 8.94E-24 | -- | 0.313295 | -1 | 0.398126 | -1 | 0.88097 | 1 |
| 19-44897790-A-AG | AG | | | | | | | 0.136204 | 7.7E-266 | 0.093105 | 5.32E-19 | TRUE | 3.2E-257 | -- | 0.301623 | 1 | 0.065828 | 1 | 7.14E-18 | 1 |
| 19-44898611-T-C | C | | | 0.073 | 0.035575 | 0.03557 | ?- | 0.041628 | 2.12E-36 | | | FALSE | 2.12E-36 | -? | | | | | | |
| 19-44899220-C-T | T | | | 0.331 | 0.000301 | 0.000301 | ?- | 0.391831 | 1.42E-30 | 0.411492 | 0.169324 | TRUE | 3.29E-24 | -- | 0.627915 | -1 | 0.444186 | -1 | 0.47329 | -1 |
| 19-44899618-ATTTTTT | ATTTT | | | | | | | 0.438981 | 7.52E-15 | | | FALSE | 7.52E-15 | -? | | | | | | |

| | | | | | | | | | | | | | | | | | | | | |
|---|---|---|---|---|---|---|---|---|---|---|---|---|---|---|---|---|---|---|---|---|
| 19-44899791-CTTTTTT | CTTTTTTTTTTT | | | | | | | 0.021584 | 3.94E-44 | | | FALSE | 3.94E-44 | +? | | | | | | |
| 19-44901600-T-C | C | | | 0.059 | 0.00277 | 0.00277 | ?- | 0.028093 | 2.11E-57 | 0.031151 | 4.65E-05 | TRUE | 3.06E-54 | -- | 0.325288 | 1 | 0.051914 | 1 | 0.000304 | 1 |
| 19-44905307-A-T | T | 0.205332 | 0.000404 | 0.166 | 0.001915 | 0.0002009 | ++ | 0.166597 | 8.16E-27 | 0.212664 | 0.064019 | TRUE | 4.52E-22 | ++ | 0.27506 | 1 | 0.799142 | 1 | 0.00279 | -1 |
| 19-44909967-TGGG-T | TGG | | | | | | | 0.385343 | 3.6E-26 | | | FALSE | 3.6E-26 | -? | | | | | | |
| 19-44909976-G-T | T | | | | | | | 0.079793 | 3.06E-30 | 0.080958 | 1.65E-07 | TRUE | 5.15E-35 | -- | 0.04829 | -1 | 0.35058 | -1 | 5.73E-06 | -1 |
| 19-44910674-CTTTTTT | CTTTTTTTTTTTTTTTTTT | | | | | | | 0.24958 | 1.26E-17 | | | FALSE | 1.26E-17 | -? | | | | | | |
| 19-44912678-G-T | T | | | | | | | 0.125822 | 2.2E-288 | 0.126365 | 1.61E-20 | TRUE | 5.4E-274 | ++ | 0.663755 | 1 | 0.028395 | 1 | 8.48E-22 | 1 |
| 19-44915229-G-A | A | | | 0.036 | 0.065459 | 0.06546 | ?+ | 0.022054 | 1.63E-53 | 0.012035 | 0.001741 | TRUE | 2.59E-50 | ++ | 0.333863 | -1 | 0.338499 | -1 | 0.000212 | 1 |
| 19-44916137-C-CAAA | CAAAAAAAAA | | | | | | | 0.290851 | 2.44E-98 | | | FALSE | 2.44E-98 | +? | | | | | | |
| 19-44916968-T-TA | TA | | | | | | | 0.028636 | 1.65E-11 | 0.023644 | 0.040917 | TRUE | 1.64E-11 | ++ | 0.208216 | 1 | 0.126605 | 1 | 0.004815 | -1 |
| 19-44917961-G-GA | GA | | | | | | | 0.012615 | 4.29E-31 | 0.007497 | 0.042697 | TRUE | 1.1E-28 | -- | 0.889146 | 1 | 0.627282 | -1 | 0.045549 | 1 |
| 19-44918337-CTTTTTT | CTTTTTTTTTTTTT | | | | | | | 0.688539 | 6.9E-137 | | | FALSE | 6.9E-137 | -? | | | | | | |
| 19-44918393-G-A | A | | | | | | | 0.028407 | 2.27E-54 | | | FALSE | 2.27E-54 | +? | | | | | | |
| 19-44919304-T-G | G | 0.031396 | 0.002731 | 0.04 | 0.171202 | 0.00241 | ++ | 0.042298 | 4.04E-08 | 0.035543 | 0.256311 | TRUE | 3.5E-07 | ++ | 0.868698 | -1 | 0.799666 | -1 | 0.252862 | -1 |
| 19-44920379-G-A | A | | | 0.262 | 0.014949 | 0.01495 | ?- | 0.338565 | 3.9E-20 | 0.324828 | 0.094881 | FALSE | 1.83E-17 | -- | 0.639376 | -1 | 0.291372 | -1 | 0.325559 | -1 |
| 19-44920730-C-CA | CA | | | | | | | 0.163869 | 3.3E-226 | 0.181196 | 3.92E-17 | TRUE | 1.6E-215 | ++ | 0.469857 | 1 | 0.28659 | 1 | 2.84E-18 | 1 |
| 19-44921082-G-GTT | GTT | | | | | | | 0.180021 | 4.8E-291 | | | FALSE | 4.8E-291 | -? | | | | | | |
| 19-44921094-A-T | T | | | | | | | 0.188624 | 9.6E-284 | | | FALSE | 9.6E-284 | -? | | | | | | |
| 19-44921095-A-T | T | | | | | | | 0.157284 | 6.8E-228 | 0.000139 | 0.683605 | TRUE | 6.6E-228 | -- | | | | | 0.850501 | 1 |
| 19-44921257-A-C | C | | | | | | | 0.193458 | 7.7E-292 | | | FALSE | 7.7E-292 | -? | | | | | | |
| 19-44921809-G-A | A | | | | | | | 0.028241 | 3.89E-55 | 0.029262 | 0.000198 | TRUE | 1.87E-51 | ++ | 0.305058 | 1 | 0.831442 | 1 | 0.000589 | 1 |
| 19-44921918-T-G | G | | | | | | | 0.615468 | 5.71E-64 | 0.633356 | 4.11E-05 | FALSE | 5.39E-59 | -- | 0.062295 | 1 | 0.725616 | -1 | 8.81E-06 | 1 |
| 19-44921921-G-A | A | | | | | | | 0.052832 | 9.77E-21 | 0.0588 | 0.000105 | TRUE | 1.06E-22 | -- | 0.08843 | -1 | 0.020759 | -1 | 0.006338 | -1 |
| 19-44925417-T-C | C | 0.001501 | 0.003395 | 0.003 | 0.694887 | 0.003351 | -- | 0.001713 | 8.15E-11 | 0.000954 | 0.05508 | TRUE | 2.95E-11 | -- | 0.519491 | -1 | 0.733976 | -1 | 0.024175 | 1 |
| 19-44928426-G-GAA | GAA | | | | | | | 0.011929 | 5.06E-12 | 0.012636 | 0.013102 | TRUE | 1.05E-12 | -- | 0.105248 | -1 | 0.548061 | -1 | 0.008675 | 1 |
| 19-44933496-C-T | T | 0.027656 | 0.001109 | 0.023 | 0.045582 | 0.0009272 | ++ | 0.012547 | 8.08E-11 | 0.027109 | 0.006468 | TRUE | 1.44E-11 | ++ | 0.281782 | 1 | 0.405821 | 1 | 0.011802 | 1 |
| 19-44934080-G-GA | GA | | | | | | | 0.007439 | 7.22E-09 | | | FALSE | 7.22E-09 | -? | | | | | | |
| 19-44935318-C-A | A | 0.013916 | 7.49E-08 | 0.019 | 0.045757 | 5.826E-08 | ++ | 0.011909 | 6.88E-11 | 0.012221 | 0.015409 | TRUE | 1.27E-11 | ++ | 0.169496 | 1 | 0.501559 | -1 | 0.022397 | 1 |
| 19-44935386-A-G | G | | | 0.301 | 0.194165 | 0.1942 | ?+ | 0.296339 | 1.73E-09 | 0.357051 | 0.055414 | FALSE | 1.91E-09 | ++ | 0.256774 | -1 | 0.058988 | -1 | 0.236776 | -1 |
| 19-44973974-C-G | G | | | 0.276 | 0.001831 | 0.001831 | ?- | 0.209923 | 5.2E-10 | 0.293666 | 0.143058 | TRUE | 4.54E-09 | -- | 0.000151 | 1 | 0.101175 | -1 | 0.562248 | 1 |
| 19-45050800-T-G | G | | | 0.014 | 0.046139 | 0.04614 | ?- | 0.008088 | 2.91E-13 | | | FALSE | 2.91E-13 | -? | | | | | | |
| 19-45094524-T-TA | TA | 0.101731 | 0.002281 | | | 0.002281 | -? | 0.092741 | 1.99E-09 | 0.09103 | 0.008637 | TRUE | 1.4E-10 | -- | 0.278885 | -1 | 0.540136 | 1 | 0.001374 | 1 |
| 19-45102867-C-T | T | 0.099431 | 0.001029 | 0.117 | 0.064693 | 0.0007487 | ++ | 0.091635 | 8.81E-10 | 0.08857 | 0.007024 | TRUE | 5.2E-11 | ++ | 0.109608 | -1 | 0.50067 | 1 | 0.000709 | 1 |
| 19-45108968-C-T | T | 0.097383 | 0.001076 | 0.115 | 0.101107 | 0.0008119 | ++ | 0.091767 | 1.76E-09 | 0.086598 | 0.006878 | TRUE | 9.13E-11 | ++ | 0.13534 | -1 | 0.486645 | 1 | 0.000902 | 1 |
| 19-45111343-C-T | T | 0.100733 | 0.00313 | 0.116 | 0.101107 | 0.002414 | ++ | 0.092042 | 7.19E-10 | 0.090255 | 0.004438 | TRUE | 2.46E-11 | ++ | 0.177719 | -1 | 0.452802 | 1 | 0.000497 | 1 |
| 19-45115701-G-A | A | | | | | | | 0.092086 | 6.12E-10 | 0.090797 | 0.006538 | TRUE | 3.5E-11 | ++ | 0.154319 | -1 | 0.464463 | 1 | 0.000715 | 1 |
| 19-45118499-TG-T | T | 0.095951 | 0.004282 | | | 0.004282 | +? | 0.091181 | 4.98E-10 | 0.085147 | 0.003829 | TRUE | 1.43E-11 | ++ | 0.150868 | -1 | 0.317173 | 1 | 0.00063 | 1 |
| 19-45123977-C-T | T | 0.11075 | 0.003431 | 0.115 | 0.136697 | 0.002723 | ++ | 0.091601 | 5.18E-10 | 0.093117 | 0.004553 | TRUE | 1.94E-11 | ++ | 0.08884 | -1 | 0.42335 | 1 | 0.000828 | 1 |
| 19-45128558-G-A | A | 0.113755 | 0.012258 | 0.117 | 0.103194 | 0.009767 | ++ | 0.091507 | 1.9E-10 | 0.095929 | 0.003778 | TRUE | 6.51E-12 | ++ | 0.099791 | -1 | 0.419825 | 1 | 0.000508 | 1 |
| 19-45130428-G-A | A | 0.114987 | 0.01326 | 0.119 | 0.101802 | 0.01058 | ++ | 0.091958 | 1.83E-10 | 0.098233 | 0.004432 | TRUE | 7.93E-12 | ++ | 0.111712 | -1 | 0.439071 | 1 | 0.000374 | 1 |
| 19-45132943-C-T | T | 0.095046 | 0.000786 | 0.117 | 0.077235 | 0.0005812 | ++ | 0.094564 | 2.88E-09 | 0.08535 | 0.005988 | TRUE | 1.16E-10 | ++ | 0.052807 | -1 | 0.328856 | 1 | 0.00116 | 1 |
| 19-45133061-C-A | A | | | 0.117 | 0.115599 | 0.1156 | ?+ | 0.091312 | 2.41E-10 | 0.086042 | 0.004305 | TRUE | 8.71E-12 | ++ | 0.121818 | -1 | 0.380538 | 1 | 0.000483 | 1 |
| 19-45134987-C-T | T | 0.09275 | 0.000574 | 0.114 | 0.115599 | 0.0004345 | ++ | 0.091143 | 5.16E-10 | 0.082075 | 0.004525 | TRUE | 1.76E-11 | ++ | 0.058232 | -1 | 0.314123 | 1 | 0.000613 | 1 |
| 19-45137848-C-A | A | | | | | | | 0.093922 | 4.81E-09 | 0.089459 | 0.005497 | TRUE | 1.68E-10 | ++ | 0.10565 | -1 | 0.479069 | 1 | 0.000556 | 1 |
| 19-45138786-G-A | A | 0.093923 | 0.001369 | 0.114 | 0.115599 | 0.001052 | ++ | 0.091225 | 3.14E-10 | 0.083371 | 0.002211 | TRUE | 4.75E-12 | ++ | 0.160457 | -1 | 0.24807 | 1 | 0.000481 | 1 |
| 19-45139287-C-G | G | | | 0.117 | 0.12323 | 0.1232 | ?- | 0.091701 | 4.74E-10 | 0.08562 | 0.006445 | TRUE | 2.63E-11 | -- | 0.13855 | -1 | 0.390728 | 1 | 0.000969 | 1 |
| 19-45141096-A-T | T | 0.092599 | 0.000476 | 0.114 | 0.115599 | 0.0003586 | -- | 0.091613 | 7.37E-10 | 0.082089 | 0.005501 | TRUE | 3.07E-11 | -- | 0.164091 | -1 | 0.321393 | 1 | 0.001231 | 1 |
| 19-45146841-G-T | T | 0.094087 | 0.001321 | 0.114 | 0.114063 | 0.001013 | ++ | 0.092265 | 2.53E-10 | 0.083648 | 0.005053 | TRUE | 1.08E-11 | ++ | 0.103126 | -1 | 0.277575 | 1 | 0.00094 | 1 |
| 19-45147128-C-T | T | 0.093207 | 0.000545 | 0.114 | 0.157745 | 0.0004258 | ++ | 0.092393 | 4.02E-10 | 0.082714 | 0.00577 | TRUE | 1.9E-11 | ++ | 0.115573 | -1 | 0.289124 | 1 | 0.001163 | 1 |
| 19-45147979-G-A | A | 0.102687 | 0.001668 | 0.118 | 0.159377 | 0.00132 | ++ | 0.092575 | 8.41E-11 | 0.092102 | 0.009227 | TRUE | 9.85E-12 | ++ | 0.246377 | -1 | 0.554884 | 1 | 0.001096 | 1 |
| 19-45148728-G-GGTA | GGTAT | 0.093265 | 0.000729 | | | 0.0007294 | -? | 0.092547 | 5.55E-10 | 0.083018 | 0.007919 | TRUE | 3.79E-11 | -- | 0.116087 | -1 | 0.356404 | 1 | 0.001451 | 1 |
| 19-45152075-T-C | C | 0.094133 | 0.000747 | 0.115 | 0.115599 | 0.000568 | -- | 0.092578 | 2.96E-10 | 0.083221 | 0.005847 | TRUE | 1.48E-11 | -- | 0.113668 | -1 | 0.369813 | 1 | 0.000844 | 1 |
| 19-45157381-C-T | T | | | | | | | 0.076322 | 5.74E-10 | 0.080322 | 0.001368 | TRUE | 5.21E-12 | ++ | 0.062114 | -1 | 0.329956 | -1 | 6.68E-05 | 1 |
| 19-45165069-C-T | T | | | | | | | 0.076542 | 5.64E-10 | 0.08833 | 0.001738 | TRUE | 7.27E-12 | ++ | 0.34687 | -1 | 0.365858 | -1 | 0.000122 | 1 |
| 19-45169864-G-A | A | 0.03083 | 0.004917 | 0.039 | 0.697115 | 0.004742 | -- | 0.05017 | 8.77E-09 | 0.035483 | 0.404494 | TRUE | 1.66E-07 | -- | 0.22754 | -1 | 0.563792 | 1 | 0.502761 | -1 |
| 19-45170443-G-A | A | 0.104003 | 0.40006 | 0.101 | 0.132675 | 0.3631 | ++ | 0.076816 | 1.22E-09 | 0.088603 | 0.001385 | TRUE | 1.11E-11 | ++ | 0.335304 | -1 | 0.426825 | -1 | 0.000106 | 1 |
| 19-45171299-CACAAA | CACAAAACAAAACAAA | | | | | | | 0.079784 | 8.45E-09 | | | FALSE | 8.45E-09 | +? | | | | | | |
| 19-45171305-A-T | T | 0.132126 | 0.229306 | | | 0.2293 | -? | 0.078215 | 1.27E-08 | 0.118778 | 0.000715 | TRUE | 4.99E-11 | -- | 0.544015 | 1 | 0.310042 | -1 | 0.000151 | 1 |
| 19-45177191-GCACAC | GCACACACACACA | | | | | | | 0.082228 | 4.11E-12 | | | FALSE | 4.11E-12 | +? | | | | | | |

| | | | | | | | | | | | | | | | | | | | | |
|---|---|---|---|---|---|---|---|---|---|---|---|---|---|---|---|---|---|---|---|---|
| 19-45181810-T-C | C | | | | | | | 0.075022 | 4.14E-09 | 0.071459 | 0.0019 | TRUE | 4.22E-11 | -- | 0.337735 | -1 | 0.638459 | -1 | 0.000279 | 1 |
| 19-45186634-C-T | T | 0.179309 | 0.003352 | 0.166 | 0.040377 | 0.002292 | ++ | 0.11544 | 3.71E-09 | 0.16548 | 0.078206 | TRUE | 7.41E-09 | ++ | 0.508618 | -1 | 0.175446 | -1 | 0.002562 | 1 |
| 19-45186718-A-T | T | | | 0.163 | 0.031194 | 0.03119 | ?- | 0.115331 | 9.25E-09 | 0.161829 | 0.056335 | TRUE | 8.94E-09 | -- | 0.715137 | -1 | 0.216871 | -1 | 0.003073 | 1 |
| 19-45186719-A-C | C | 0.177836 | 0.003852 | 0.163 | 0.0342 | 0.002611 | -- | 0.115938 | 1.61E-08 | 0.162955 | 0.053255 | TRUE | 1.29E-08 | -- | 0.696359 | -1 | 0.236669 | -1 | 0.003161 | 1 |
| 19-45187287-C-T | T | 0.09339 | 0.048114 | 0.093 | 0.066399 | 0.03969 | ++ | 0.077782 | 3.35E-09 | 0.079436 | 0.010733 | TRUE | 2.76E-10 | ++ | 0.208532 | -1 | 0.539492 | -1 | 0.000635 | 1 |
| 19-45187517-A-G | G | 0.125721 | 0.042106 | 0.104 | 0.130924 | 0.03556 | -- | 0.080547 | 2.09E-08 | 0.113839 | 0.011909 | TRUE | 1.97E-09 | -- | 0.988663 | -1 | 0.512011 | -1 | 0.000789 | 1 |
| 19-45188312-C-T | T | 0.064928 | 0.837751 | 0.046 | 0.142751 | 0.8751 | -+ | 0.033907 | 1.78E-08 | 0.05343 | 0.536492 | TRUE | 1.42E-06 | ++ | 0.593938 | -1 | 0.313008 | -1 | 0.217394 | 1 |
| 19-45199680-CTTCTC | C | | | | | | | 0.014724 | 1.27E-09 | 0.021865 | 0.094875 | TRUE | 2.95E-09 | -- | 0.151117 | 1 | 0.203257 | -1 | 0.124811 | 1 |
| 19-45207281-C-T | T | 0.269444 | 0.119835 | 0.207 | 0.350587 | 0.1067 | ++ | 0.168208 | 6.14E-10 | 0.238825 | 0.04076 | FALSE | 5.68E-10 | ++ | 0.117768 | 1 | 0.509381 | -1 | 0.085751 | 1 |
| 19-54288504-A-G | G | 0.459422 | 0.935947 | 0.403 | 0.342844 | 0.8788 | -- | 0.600881 | 1.82E-08 | 0.579981 | 0.714266 | FALSE | 1.46E-06 | ++ | 0.297967 | -1 | 0.715387 | 1 | 0.745168 | 1 |
| 19-54288511-TG-T | T | | | | | | | 0.599795 | 1.89E-08 | 0.578906 | 0.753128 | FALSE | 1.75E-06 | -- | 0.260406 | -1 | 0.670164 | 1 | 0.693847 | 1 |
| 19-449659-C-G | G | 0.407881 | 0.665712 | 0.492 | 0.188323 | 0.5933 | -- | | | 0.590286 | 2.24E-08 | FALSE | 2.24E-08 | ?- | 0.108077 | 1 | 0.807114 | 1 | 2.46E-05 | 1 |
| 19-560603-T-C | C | | | | | | | | | 0.497507 | 7.9E-13 | FALSE | 7.9E-13 | ?- | 0.700122 | 1 | 0.600833 | 1 | 5.73E-09 | 1 |
| 19-613718-G-GCACC | GCACC | | | | | | | | | 0.165956 | 7.35E-09 | FALSE | 7.35E-09 | ?+ | 0.071471 | -1 | 0.493466 | 1 | 5.98E-10 | -1 |
| 19-780466-ATT-A | A | 0.001172 | 0.875754 | | | 0.8758 | -? | 0.001244 | 0.293829 | 0.000845 | 2.8E-09 | TRUE | 0.003651 | -+ | 0.478356 | 1 | 0.031006 | 1 | 1.11E-08 | 1 |
| 19-1430887-ATTT-A | A | | | | | | | | | 0.000675 | 2.5E-10 | TRUE | 2.5E-10 | ?+ | 2.81E-05 | 1 | 0.927709 | 1 | 3.74E-07 | 1 |
| 19-1536950-A-T | T | | | | | | | 0.00011 | 0.649421 | 0.01976 | 2.54E-10 | FALSE | 4.88E-10 | -+ | 0.267773 | -1 | 0.032641 | -1 | 2.27E-08 | -1 |
| 19-1666683-G-GC | GC | 0.001739 | 0.377257 | | | 0.3773 | -? | | | 0.005077 | 1.08E-13 | TRUE | 1.08E-13 | ?- | 0.0119 | 1 | 0.002234 | 1 | 6.73E-11 | 1 |
| 19-1893632-T-C | C | 0.001017 | 0.640825 | | | 0.6408 | +? | 0.001281 | 0.337528 | 0.0028 | 5.2E-12 | TRUE | 3.94E-09 | -- | 0.022694 | 1 | 0.265223 | 1 | 8.22E-11 | 1 |
| 19-1899530-T-TC | TC | | | | | | | | | 0.001469 | 3.79E-11 | TRUE | 3.79E-11 | ?- | 0.009695 | 1 | 0.207763 | 1 | 2.14E-09 | 1 |
| 19-1927018-C-G | G | | | 0.23 | 0.644153 | 0.6442 | ?- | 0.214696 | 0.442573 | 0.310601 | 4.45E-08 | TRUE | 0.009245 | +- | 0.051205 | 1 | 0.478311 | -1 | 3.45E-06 | 1 |
| 19-2654412-A-G | G | | | | | | | 0.000113 | 0.745937 | 0.001279 | 2.13E-10 | TRUE | 7.76E-10 | -- | 0.45217 | 1 | 0.05087 | 1 | 4.27E-08 | 1 |
| 19-2673038-C-CCT | CCT | | | | | | | | | 0.002334 | 1.07E-12 | TRUE | 1.07E-12 | ?+ | 0.208864 | 1 | 0.009631 | 1 | 8.32E-11 | 1 |
| 19-2870227-AGTTG-A | A | | | | | | | | | 0.000749 | 4.27E-11 | TRUE | 4.27E-11 | ?+ | 0.617366 | 1 | 0.08024 | 1 | 3.75E-11 | 1 |
| 19-2870231-G-GCA | GCA | | | | | | | | | 0.000586 | 1.62E-09 | TRUE | 1.62E-09 | ?- | 0.361194 | 1 | 0.0158 | 1 | 1.27E-08 | 1 |
| 19-3259781-G-GA | GA | 0.001589 | 0.120973 | | | 0.121 | +? | | | 0.001862 | 5.06E-12 | TRUE | 5.06E-12 | ?- | 0.017381 | 1 | 0.052133 | 1 | 1.59E-10 | 1 |
| 19-3479270-A-AAAG | AAAG | 0.001736 | 0.196953 | | | 0.197 | +? | | | 0.002039 | 1.89E-08 | TRUE | 1.89E-08 | ?- | 0.024082 | 1 | 0.044159 | 1 | 6.61E-07 | 1 |
| 19-3587428-C-CCCG | CCCG | 0.000175 | 0.063817 | | | 0.06382 | +? | | | 0.000485 | 4.65E-08 | TRUE | 4.65E-08 | ?+ | 0.479265 | 1 | 0.868548 | 1 | 3.32E-09 | 1 |
| 19-3719608-T-C | C | 0.000118 | 0.188009 | | | 0.188 | +? | 0.000407 | 0.407335 | 0.000414 | 1.9E-09 | TRUE | 9.3E-07 | -- | 0.663841 | 1 | 0.012323 | 1 | 8.24E-08 | 1 |
| 19-3843252-CTT-C | C | 0.004032 | 0.479929 | | | 0.4799 | +? | | | 0.007233 | 1.86E-20 | TRUE | 1.86E-20 | ?- | 0.013878 | 1 | 0.104574 | 1 | 1.02E-19 | 1 |
| 19-4228508-G-GTCTC | GTCTC | | | | | | | 0.359091 | 0.979821 | 0.447928 | 1.17E-08 | FALSE | 0.001565 | -- | 0.070702 | 1 | 0.085018 | -1 | 1.66E-08 | 1 |
| 19-4289268-G-A | A | 0.000217 | 0.593166 | | | 0.5932 | -? | 0.004614 | 0.745157 | 0.000764 | 2.49E-11 | TRUE | 0.003803 | ++ | 0.522835 | -1 | 0.236433 | 1 | 4.52E-11 | 1 |
| 19-4289272-G-A | A | 0.000555 | 0.671455 | | | 0.6715 | -? | 0.005616 | 0.985852 | 0.001573 | 8.41E-10 | TRUE | 0.01016 | -+ | 0.793582 | -1 | 0.213426 | 1 | 8.47E-10 | 1 |
| 19-4302852-CA-C | C | | | | | | | | | 0.000164 | 7.98E-09 | TRUE | 7.98E-09 | ?- | 0.000385 | 1 | | | 8.39E-06 | 1 |
| 19-4331320-CCT-C | C | | | | | | | | | 0.001257 | 5.99E-14 | TRUE | 5.99E-14 | ?- | 2.48E-07 | 1 | 0.004852 | 1 | 1.26E-08 | 1 |
| 19-4331323-T-TCC | TCC | | | | | | | | | 0.001119 | 1.53E-13 | TRUE | 1.53E-13 | ?- | 6.25E-06 | 1 | 0.074545 | 1 | 4.93E-10 | 1 |
| 19-4417457-C-CGG | CGG | | | | | | | | | 0.062366 | 1.48E-14 | FALSE | 1.48E-14 | ?+ | 0.795923 | -1 | 0.058696 | 1 | 1.78E-14 | 1 |
| 19-4450003-T-G | G | 0.000118 | 0.272716 | | | 0.2727 | +? | 0.001785 | 0.660341 | 0.005188 | 5.38E-27 | TRUE | 5.92E-20 | -- | 0.001207 | 1 | 0.001934 | 1 | 8.86E-22 | 1 |
| 19-4520591-A-G | G | 0.085617 | 0.255368 | | | 0.2554 | -? | | | 0.097889 | 1.55E-08 | TRUE | 1.55E-08 | ?- | 0.038248 | 1 | 0.61346 | -1 | 0.04749 | 1 |
| 19-4528057-C-A | A | | | | | | | 1.25E-05 | 0.43248 | 0.001072 | 3.57E-16 | TRUE | 9.69E-16 | -+ | 0.126624 | 1 | 0.172444 | 1 | 6.34E-16 | 1 |
| 19-4562943-GA-G | G | 7.87E-05 | 0.459313 | | | 0.4593 | +? | | | 0.000699 | 4.51E-08 | TRUE | 4.51E-08 | ?+ | 0.216109 | 1 | 0.026193 | 1 | 1.61E-07 | 1 |
| 19-4666569-C-T | T | 0.226213 | 0.104659 | 0.167 | 0.508114 | 0.09779 | ++ | | | 0.251031 | 7.31E-10 | TRUE | 7.31E-10 | ?+ | 0.205598 | 1 | 0.232616 | -1 | 7.33E-05 | 1 |
| 19-4755334-CCT-C | C | | | | | | | | | 0.00102 | 1.25E-14 | TRUE | 1.25E-14 | ?- | 0.003859 | 1 | 0.15792 | 1 | 3E-11 | 1 |
| 19-4788589-ACT-A | A | | | | | | | | | 0.002892 | 1.86E-12 | TRUE | 1.86E-12 | ?+ | 0.082011 | 1 | 0.014303 | 1 | 5.04E-10 | 1 |
| 19-4862940-C-CCT | CCT | | | | | | | | | 0.001535 | 4.99E-08 | TRUE | 4.99E-08 | ?+ | 0.063885 | 1 | 0.023092 | 1 | 1.69E-06 | 1 |
| 19-4899269-G-A | A | 0.000391 | 0.39471 | | | 0.3947 | -? | 0.001156 | 0.418919 | 0.001244 | 1.74E-08 | TRUE | 0.000741 | -+ | 0.224985 | 1 | 0.008616 | 1 | 8.79E-07 | 1 |
| 19-5175397-G-C | C | | | | | | | 1.88E-05 | 0.2232 | 0.000295 | 3.42E-09 | TRUE | 5.55E-08 | -+ | 0.01636 | 1 | 0.141265 | 1 | 2.01E-08 | 1 |
| 19-5175404-G-A | A | | | | | | | | | 0.000478 | 5.31E-09 | TRUE | 5.31E-09 | ?+ | 0.895301 | 1 | 0.040684 | 1 | 4.04E-09 | 1 |
| 19-5192343-G-A | A | | | | | | | | | 0.001394 | 3.66E-14 | TRUE | 3.66E-14 | ?+ | 0.203649 | 1 | 0.112764 | 1 | 1.02E-13 | 1 |
| 19-5192353-C-A | A | | | | | | | | | 0.00093 | 5.87E-13 | TRUE | 5.87E-13 | ?+ | 0.159028 | 1 | 0.050108 | 1 | 1.86E-12 | 1 |
| 19-5192936-G-A | A | | | | | | | 1.57E-05 | 0.40179 | 0.000265 | 2.67E-08 | TRUE | 1.2E-07 | -+ | | | | | 1.78E-07 | 1 |
| 19-5810053-CAGG-C | C | 0.000156 | 0.339321 | | | 0.3393 | +? | | | 0.001881 | 4.25E-11 | TRUE | 4.25E-11 | ?- | 0.11119 | 1 | 0.172127 | 1 | 3.92E-10 | 1 |
| 19-6104375-C-T | T | | | | | | | 6.26E-05 | 0.550938 | 0.0004 | 3.26E-09 | TRUE | 8.6E-08 | -+ | 0.081907 | 1 | 0.044249 | 1 | 1.52E-08 | 1 |
| 19-6104380-T-C | C | | | | | | | | | 0.000833 | 9.02E-14 | TRUE | 9.02E-14 | ?- | 0.094755 | 1 | 0.464955 | 1 | 5.01E-13 | 1 |
| 19-6104382-A-G | G | | | | | | | | | 0.000919 | 3.59E-10 | TRUE | 3.59E-10 | ?- | 0.04395 | 1 | 0.184298 | 1 | 3.92E-09 | 1 |
| 19-6308254-G-A | A | | | | | | | 9.71E-05 | 0.368815 | 0.000946 | 5.11E-11 | TRUE | 2.46E-09 | -+ | 4.04E-05 | 1 | 0.009874 | 1 | 3.98E-07 | 1 |
| 19-6348245-ACC-A | A | 0.00039 | 0.231188 | | | 0.2312 | -? | | | 0.004888 | 5.33E-31 | TRUE | 5.33E-31 | ?+ | 0.000125 | 1 | 0.008478 | 1 | 2.85E-25 | 1 |
| 19-8042051-G-A | A | 0.000273 | 0.775544 | | | 0.7755 | -? | 0.000564 | 0.524986 | 0.001168 | 4.85E-08 | TRUE | 8.23E-05 | -+ | 0.978782 | -1 | 0.019597 | 1 | 1.75E-07 | 1 |
| 19-8042058-A-G | G | | | | | | | | | 0.00148 | 4.06E-08 | TRUE | 4.06E-08 | ?- | 0.402483 | 1 | 0.00088 | 1 | 2.19E-06 | 1 |

| | | | | | | | | | | | | | | | | | | | | |
|---|---|---|---|---|---|---|---|---|---|---|---|---|---|---|---|---|---|---|---|---|
| 19-8165431-T-G | G | | | | | | | | | 0.000598 | 4.6E-08 | TRUE | 4.6E-08 | ?- | 0.000214 | 1 | 0.466256 | 1 | 5.7E-06 | 1 |
| 19-8165439-CTT-C | C | | | | | | | | | 0.000548 | 8.18E-10 | TRUE | 8.18E-10 | ?- | 0.000844 | 1 | 0.17757 | 1 | 1.94E-07 | 1 |
| 19-8600010-AT-A | A | | | | | | | | | 0.001017 | 1.21E-11 | TRUE | 1.21E-11 | ?+ | 0.03341 | 1 | 0.247915 | 1 | 4.29E-10 | 1 |
| 19-9766170-G-A | A | 0.000216 | 0.553227 | | | 0.5532 | +? | 0.00016 | 0.746928 | 0.000445 | 2.54E-09 | TRUE | 6.18E-07 | -+ | 0.03775 | 1 | 0.308903 | -1 | 7.48E-11 | 1 |
| 19-10263242-GCGCC- | G | | | | | | | | | 0.003509 | 4.88E-08 | TRUE | 4.88E-08 | ?+ | 0.439253 | 1 | 0.915816 | 1 | 2.8E-09 | 1 |
| 19-11071676-G-GC | GC | 0.001223 | 0.213201 | | | 0.2132 | -? | | | 0.006829 | 1.75E-35 | TRUE | 1.75E-35 | ?- | 0.001021 | 1 | 0.005671 | 1 | 1.07E-30 | 1 |
| 19-11473295-A-G | G | | | | | | | | | 0.306218 | 3.87E-10 | TRUE | 3.87E-10 | ?- | 0.29381 | 1 | 0.976865 | 1 | 0.008846 | 1 |
| 19-11582992-T-G | G | | | | | | | | | 0.000635 | 1.61E-09 | TRUE | 1.61E-09 | ?- | 0.000139 | 1 | 0.164727 | 1 | 8.97E-05 | 1 |
| 19-11648684-AAC-A | A | | | | | | | | | 0.000539 | 9.04E-10 | TRUE | 9.04E-10 | ?+ | 0.703539 | 1 | 0.058386 | 1 | 1.64E-09 | 1 |
| 19-11668061-G-A | A | 0.002411 | 0.652143 | | | 0.6521 | -? | 0.003881 | 0.424244 | 0.003554 | 2.32E-13 | TRUE | 8.53E-08 | ++ | 0.007098 | 1 | 0.26003 | 1 | 2.69E-12 | 1 |
| 19-12129144-TA-T | T | | | | | | | | | 0.00051 | 5.85E-09 | TRUE | 5.85E-09 | ?+ | 0.745293 | 1 | 0.360103 | 1 | 1.63E-09 | 1 |
| 19-12286479-C-CAA | CAA | | | | | | | | | 0.000903 | 8.62E-09 | TRUE | 8.62E-09 | ?+ | 0.427075 | 1 | 0.058529 | 1 | 4.51E-08 | 1 |
| 19-13180657-A-C | C | 0.00243 | 0.340815 | | | 0.3408 | -? | 0.000445 | 0.839454 | 0.000986 | 1.78E-10 | TRUE | 5.31E-07 | +- | 0.931249 | 1 | 0.000609 | 1 | 8.85E-09 | 1 |
| 19-13661533-CAT-C | C | 0.000175 | 0.457073 | | | 0.4571 | +? | 0.000207 | 0.363768 | 0.001519 | 1.02E-09 | TRUE | 1.08E-07 | +- | 0.002814 | 1 | 0.020674 | 1 | 1.71E-07 | 1 |
| 19-13736813-CCT-C | C | | | | | | | | | 0.001222 | 9.21E-11 | TRUE | 9.21E-11 | ?- | 0.008318 | 1 | 0.600778 | 1 | 1.35E-09 | 1 |
| 19-13745457-A-G | G | | | | | | | | | 0.000593 | 7.49E-09 | TRUE | 7.49E-09 | ?- | 0.917978 | -1 | 0.000538 | 1 | 1.27E-07 | 1 |
| 19-13786952-A-G | G | | | | | | | | | 0.411533 | 2.41E-12 | FALSE | 2.42E-12 | ?- | 0.101746 | 1 | 0.653081 | 1 | 3.06E-06 | 1 |
| 19-13893919-A-AT | AT | | | | | | | | | 0.001735 | 6.48E-19 | TRUE | 6.48E-19 | ?- | 0.077857 | 1 | 0.00115 | 1 | 7.09E-16 | 1 |
| 19-13893922-A-AT | AT | | | | | | | | | 0.001099 | 3.67E-11 | TRUE | 3.67E-11 | ?- | 0.262753 | 1 | 0.291031 | 1 | 3.59E-11 | 1 |
| 19-13946219-A-C | C | | | | | | | 0.001153 | 0.29139 | 0.002176 | 3.5E-14 | TRUE | 1.84E-07 | +- | 0.000121 | 1 | 0.04242 | 1 | 1.54E-10 | 1 |
| 19-14258228-C-CATA | CATA | 0.000139 | 0.649407 | | | 0.6494 | +? | | | 0.001785 | 1.2E-10 | TRUE | 1.2E-10 | ?+ | 0.236039 | 1 | 0.414644 | 1 | 4.17E-11 | 1 |
| 19-14648966-TCCC-T | T | | | | | | | | | 0.000984 | 1.32E-11 | TRUE | 1.32E-11 | ?+ | 0.0117 | 1 | 0.009202 | 1 | 8.58E-10 | 1 |
| 19-14707697-GCCCA-( | G | | | | | | | | | 0.00061 | 1.16E-08 | TRUE | 1.16E-08 | ?+ | 1.94E-05 | 1 | 0.812976 | -1 | 3.34E-06 | 1 |
| 19-15173421-C-A | A | | | | | | | 0.004454 | 0.408266 | 0.014821 | 2.6E-08 | FALSE | 0.000145 | +- | 0.290424 | -1 | 0.020809 | -1 | 7.38E-07 | -1 |
| 19-15822925-G-GGGC | GGGCT | | | | | | | | | 0.001832 | 2.58E-26 | TRUE | 2.58E-26 | ?- | 0.004766 | 1 | 0.804793 | 1 | 1.76E-25 | 1 |
| 19-15822927-CTTAT-C | C | | | | | | | | | 0.00094 | 3E-12 | TRUE | 3E-12 | ?- | 0.015593 | 1 | 0.17477 | 1 | 4.25E-11 | 1 |
| 19-15873260-CGCACC | C | 0.008184 | 0.675661 | | | 0.6757 | -? | | | 0.023542 | 1.42E-11 | FALSE | 1.42E-11 | ?- | 0.192899 | 1 | 0.020147 | 1 | 8.43E-10 | 1 |
| 19-16311726-A-G | G | 0.303477 | 0.200953 | 0.25 | 0.467639 | 0.1847 | ++ | | | 0.325083 | 3.57E-12 | FALSE | 3.57E-12 | ?- | 0.695626 | 1 | 0.739464 | 1 | 1.17E-09 | 1 |
| 19-16557360-AT-A | A | | | | | | | 0.000197 | 0.354923 | 0.001081 | 1.27E-10 | TRUE | 4E-10 | ++ | 0.000124 | 1 | 0.077204 | 1 | 7.7E-07 | 1 |
| 19-17354058-GGT-G | G | | | | | | | | | 0.000336 | 2.97E-08 | TRUE | 2.97E-08 | ?+ | 0.32886 | 1 | 0.357769 | 1 | 7.08E-08 | 1 |
| 19-18528048-C-CT | CT | | | | | | | | | 0.000663 | 2.81E-10 | TRUE | 2.81E-10 | ?+ | 0.035833 | 1 | 0.078244 | 1 | 5.87E-09 | 1 |
| 19-18619797-CCA-C | C | 0.001179 | 0.302148 | | | 0.3021 | +? | | | 0.001376 | 3.01E-09 | TRUE | 3.01E-09 | ?- | 0.098817 | 1 | 0.369059 | -1 | 8.06E-10 | 1 |
| 19-18689093-G-GCC | GCC | 0.000137 | 0.374492 | | | 0.3745 | -? | | | 0.002099 | 1.83E-08 | TRUE | 1.83E-08 | ?- | 0.457991 | 1 | 0.041241 | 1 | 1.79E-07 | 1 |
| 19-19363796-G-GTT | GTT | | | | | | | | | 0.00171 | 4.86E-13 | TRUE | 4.86E-13 | ?- | 0.560311 | 1 | 0.019881 | 1 | 1.67E-12 | 1 |
| 19-19382660-C-T | T | | | | | | | 0.000125 | 0.80841 | 0.000452 | 2.61E-10 | TRUE | 8.87E-09 | ++ | 0.002425 | 1 | 0.251517 | 1 | 3.34E-07 | 1 |
| 19-19572678-AACGTG | A | 0.000587 | 0.616021 | | | 0.616 | +? | | | 0.002363 | 1.36E-09 | TRUE | 1.36E-09 | ?+ | 0.014362 | 1 | 0.913163 | 1 | 6.34E-09 | 1 |
| 19-20023807-CCT-C | C | | | | | | | | | 0.000174 | 1.08E-08 | TRUE | 1.08E-08 | ?- | 8.57E-05 | 1 | | | 5.2E-07 | 1 |
| 19-20076324-C-A | A | | | | | | | 0.000107 | 0.965211 | 0.000455 | 8.82E-15 | TRUE | 9.1E-13 | -+ | 0.00017 | 1 | 0.368903 | -1 | 1.95E-14 | 1 |
| 19-20143926-A-G | G | 0.000157 | 0.604082 | | | 0.6041 | -? | 0.00098 | 0.757766 | 0.002427 | 6.88E-19 | TRUE | 1.24E-13 | -- | 0.000394 | 1 | 0.000502 | 1 | 4.54E-12 | 1 |
| 19-20143939-G-A | A | | | | | | | 0.001833 | 0.28918 | 0.00397 | 7.88E-27 | TRUE | 2.94E-15 | -+ | 5.51E-07 | 1 | 2.68E-05 | 1 | 1.67E-15 | 1 |
| 19-20144534-CA-C | C | | | | | | | | | 0.001499 | 1.19E-08 | TRUE | 1.19E-08 | ?- | 0.186625 | 1 | 0.11317 | 1 | 2.54E-08 | 1 |
| 19-22712186-CAT-C | C | | | | | | | | | 0.000813 | 5.31E-10 | TRUE | 5.31E-10 | ?- | 0.035555 | 1 | 1.54E-05 | 1 | 2.95E-06 | 1 |
| 19-22712189-G-GCC | GCC | | | | | | | | | 0.000759 | 1.06E-09 | TRUE | 1.06E-09 | ?- | 0.093265 | 1 | 0.00024 | 1 | 8.48E-07 | 1 |
| 19-22831340-TGAG-T | T | 0.009294 | 0.918018 | | | 0.918 | -? | 0.000166 | 0.741934 | 0.005076 | 3.31E-10 | TRUE | 1.33E-09 | -+ | 0.809775 | -1 | 0.012864 | 1 | 6.72E-10 | 1 |
| 19-23676840-C-CAA | CAA | | | | | | | | | 0.000436 | 1.41E-08 | TRUE | 1.41E-08 | ?+ | 0.155436 | 1 | 0.000309 | 1 | 1.15E-06 | 1 |
| 19-23676856-C-CCT | CCT | | | | | | | | | 0.00034 | 3.76E-09 | TRUE | 3.76E-09 | ?+ | 0.492292 | -1 | 0.000627 | 1 | 9.39E-07 | 1 |
| 19-29308706-T-C | C | 0.486988 | 0.679237 | 0.448 | 0.739306 | 0.7393 | ?- | 0.433082 | 0.673598 | 0.541673 | 4.09E-13 | FALSE | 1.77E-05 | -- | 0.334252 | 1 | 0.140686 | 1 | 1.4E-09 | 1 |
| 19-29900215-A-G | G | | | | | | | | | 0.880261 | 4.5E-13 | FALSE | 4.5E-13 | ?+ | 0.021295 | -1 | 0.687885 | 1 | 8.94E-14 | -1 |
| 19-32728929-A-G | G | | | | | | | 0.003321 | 0.005701 | 0.00463 | 1.16E-17 | TRUE | 2.09E-05 | +- | 0.304858 | 1 | 0.046598 | 1 | 2.13E-17 | 1 |
| 19-33033650-CCT-C | C | | | | | | | | | 0.000533 | 1.94E-08 | TRUE | 1.94E-08 | ?- | 0.016088 | 1 | 0.325746 | 1 | 4.62E-07 | 1 |
| 19-33141241-C-T | T | | | | | | | | | 0.579042 | 4.48E-08 | TRUE | 4.48E-08 | ?+ | 0.086557 | 1 | 0.509826 | 1 | 1.84E-05 | 1 |
| 19-33283110-T-C | C | 0.000273 | 0.001541 | | | 0.001541 | -? | 0.000166 | 0.398578 | 0.000898 | 6.11E-10 | TRUE | 2.18E-09 | -- | 0.012415 | 1 | 0.979287 | -1 | 2.23E-08 | 1 |
| 19-33837044-AC-A | A | | | | | | | | | 0.000474 | 1.45E-13 | TRUE | 1.45E-13 | ?+ | 0.753959 | 1 | 0.016428 | 1 | 6.03E-14 | 1 |
| 19-34031357-G-C | C | 0.000805 | 0.701241 | | | 0.7012 | +? | 0.002979 | 0.001682 | 0.001815 | 2.67E-09 | TRUE | 4.55E-09 | ++ | 0.012287 | 1 | 0.037617 | 1 | 1.19E-07 | 1 |
| 19-34031358-C-A | A | | | | | | | 0.003017 | 0.000694 | 0.001734 | 4.13E-08 | TRUE | 8.57E-09 | ++ | 0.057392 | 1 | 0.168084 | 1 | 3.23E-07 | 1 |
| 19-34371991-C-CCT | CCT | | | | | | | | | 0.000315 | 3.43E-08 | TRUE | 3.43E-08 | ?+ | 0.11531 | 1 | 3.91E-05 | 1 | 1.19E-05 | 1 |
| 19-34465420-CG-C | C | | | | | | | | | 0.000596 | 7.81E-11 | TRUE | 7.81E-11 | ?- | 0.854492 | -1 | 0.008074 | 1 | 8.26E-11 | 1 |
| 19-35386346-C-T | T | 0.000512 | 0.057842 | | | 0.05784 | -? | 0.000973 | 0.938541 | 0.001236 | 1.59E-10 | TRUE | 3.8E-06 | -+ | 0.545837 | 1 | 0.054892 | 1 | 2.09E-09 | 1 |
| 19-35386354-C-T | T | 0.000551 | 0.117261 | | | 0.1173 | -? | 0.000936 | 0.871465 | 0.000874 | 1.71E-10 | TRUE | 1.84E-05 | -+ | 0.670056 | -1 | 0.000209 | 1 | 8.73E-09 | 1 |

| | | | | | | | | | | | | | | | | | | | | |
|---|---|---|---|---|---|---|---|---|---|---|---|---|---|---|---|---|---|---|---|---|
| 19-35630490-A-C | C | 0.207025 | 0.916069 | 0.166 | 0.957243 | 0.9188 | +- | 0.146824 | 0.400601 | 0.218054 | 1.69E-08 | TRUE | 5.18E-05 | -- | 0.066362 | 1 | 0.781358 | -1 | 4.47E-06 | 1 |
| 19-35911172-CAT-C | C | | | | | | | | | 0.000402 | 3.32E-08 | TRUE | 3.32E-08 | ?- | 0.000171 | 1 | 0.304815 | 1 | 1.28E-06 | 1 |
| 19-36204814-ACC-A | A | 0.000273 | 0.385018 | | | 0.385 | +? | | | 0.002922 | 4.35E-26 | TRUE | 4.35E-26 | ?+ | 0.025773 | 1 | 0.001905 | 1 | 1.4E-22 | 1 |
| 19-36204819-G-GCA | GCA | 0.000489 | 0.486643 | | | 0.4866 | -? | | | 0.002711 | 4.97E-25 | TRUE | 4.97E-25 | ?- | 0.016068 | 1 | 0.002217 | 1 | 1.89E-20 | 1 |
| 19-36516403-CTTGG-( | C | | | | | | | | | 0.000804 | 1E-10 | TRUE | 1E-10 | ?- | 0.00164 | 1 | 0.000657 | 1 | 9.68E-07 | 1 |
| 19-36868169-AT-A | A | | | | | | | | | 0.001316 | 4.08E-13 | TRUE | 4.08E-13 | ?+ | 0.005404 | 1 | 0.316096 | 1 | 8.5E-11 | 1 |
| 19-36868792-A-AT | AT | | | | | | | 5.01E-05 | 0.155975 | 0.000972 | 4.68E-11 | TRUE | 1.72E-09 | +- | 0.016935 | 1 | 0.113215 | 1 | 2.46E-09 | 1 |
| 19-36950324-A-G | G | 9.75E-05 | 0.905648 | | | 0.9056 | +? | | | 0.000561 | 8.16E-10 | TRUE | 8.16E-10 | ?- | 0.001572 | 1 | 0.005874 | 1 | 6.53E-07 | 1 |
| 19-37687722-C-CAG | CAG | | | | | | | | | 0.000467 | 7.47E-09 | TRUE | 7.47E-09 | ?+ | 0.053208 | 1 | 0.169759 | 1 | 3.75E-07 | 1 |
| 19-38243249-GCC-G | G | 0.000136 | 0.568543 | | | 0.5685 | -? | | | 0.000896 | 6.09E-09 | TRUE | 6.09E-09 | ?+ | 0.974464 | 1 | 0.00014 | 1 | 3.01E-06 | 1 |
| 19-39105162-T-TGC | TGC | 0.000469 | 0.43203 | | | 0.432 | +? | | | 0.002739 | 1.24E-11 | TRUE | 1.24E-11 | ?- | 0.000961 | 1 | 0.000607 | 1 | 3.48E-08 | 1 |
| 19-39105164-CGT-C | C | | | | | | | | | 0.002371 | 3.54E-11 | TRUE | 3.54E-11 | ?- | 0.000925 | 1 | 0.000568 | 1 | 6.02E-08 | 1 |
| 19-39108282-GGC-G | G | | | | | | | | | 0.001961 | 2.85E-19 | TRUE | 2.85E-19 | ?+ | 4.36E-05 | 1 | 0.091056 | 1 | 1.23E-15 | 1 |
| 19-39108289-A-ATG | ATG | | | | | | | | | 0.001611 | 1.08E-12 | TRUE | 1.08E-12 | ?- | 0.00231 | 1 | 0.06414 | 1 | 6.3E-10 | 1 |
| 19-39148690-C-CAG | CAG | | | | | | | | | 0.00245 | 5.18E-11 | TRUE | 5.18E-11 | ?+ | 0.980391 | 1 | 0.005296 | 1 | 2.15E-09 | 1 |
| 19-39301024-T-TCC | TCC | | | | | | | 0.00011 | 0.252972 | 0.001581 | 2.06E-18 | TRUE | 2.68E-18 | -- | 0.001208 | 1 | 0.013301 | 1 | 6.48E-14 | 1 |
| 19-40402196-CAA-C | C | | | | | | | | | 0.002395 | 4.7E-10 | TRUE | 4.7E-10 | ?- | 0.370907 | 1 | 0.312124 | 1 | 5.69E-09 | 1 |
| 19-40791098-A-T | T | 0.000553 | 0.575195 | | | 0.5752 | -? | 0.002142 | 0.264481 | 0.002324 | 1.46E-10 | TRUE | 2.89E-07 | -- | 0.197319 | 1 | 0.08141 | 1 | 6.76E-10 | 1 |
| 19-41157626-AGT-A | A | | | | | | | | | 0.000307 | 2.07E-10 | TRUE | 2.07E-10 | ?+ | 0.002288 | 1 | 0.035467 | 1 | 2.47E-07 | 1 |
| 19-41509971-C-CT | CT | | | | | | | | | 0.005887 | 1.17E-15 | TRUE | 1.17E-15 | ?+ | 0.440937 | 1 | 0.228065 | 1 | 7.29E-16 | 1 |
| 19-41806330-T-TTG | TTG | | | | | | | | | 0.001284 | 3.95E-11 | TRUE | 3.95E-11 | ?- | 0.514738 | 1 | 0.915682 | 1 | 4.31E-12 | 1 |
| 19-42482163-A-AT | AT | 0.000156 | 0.720627 | | | 0.7206 | +? | 0.001084 | 0.60929 | 0.001195 | 8.4E-09 | TRUE | 1.33E-05 | -- | 0.018043 | 1 | 0.10193 | 1 | 7.72E-06 | 1 |
| 19-43061627-AT-A | A | 0.000137 | 0.792596 | | | 0.7926 | -? | | | 0.0008 | 2.74E-08 | TRUE | 2.74E-08 | ?+ | 0.569864 | 1 | 7.48E-05 | 1 | 3.6E-06 | 1 |
| 19-44457678-G-A | A | | | | | | | 0.000179 | 0.830066 | 0.001656 | 7.08E-14 | TRUE | 1.14E-12 | ++ | 0.129681 | 1 | 0.265304 | 1 | 2.16E-12 | 1 |
| 19-44862219-T-C | C | 0.296104 | 0.000578 | 0.374 | 0.09479 | 0.0003802 | -- | | | 0.325429 | 2.51E-08 | TRUE | 2.51E-08 | ?- | 0.591218 | -1 | 0.022993 | 1 | 8.34E-09 | 1 |
| 19-44882783-AT-A | A | | | | | | | | | 0.752852 | 1.59E-09 | FALSE | 1.59E-09 | ?- | 0.33823 | 1 | 0.174222 | -1 | 1.25E-10 | -1 |
| 19-44882783-ATT-A | A | | | | | | | | | 0.19067 | 1.66E-08 | TRUE | 1.66E-08 | ?+ | 0.352655 | -1 | 0.612858 | 1 | 7.32E-11 | 1 |
| 19-44885313-T-C | C | | | | | | | | | 0.254201 | 9.84E-13 | FALSE | 9.84E-13 | ?- | 0.108641 | 1 | 0.633009 | 1 | 2.31E-12 | 1 |
| 19-44909521-CT-C | C | | | | | | | | | 0.043531 | 1.47E-14 | TRUE | 1.47E-14 | ?- | 0.982686 | -1 | 0.206445 | -1 | 4.6E-17 | 1 |
| 19-44909967-TGG-T | T | | | | | | | | | 0.110931 | 9.29E-28 | TRUE | 9.29E-28 | ?+ | 0.205379 | 1 | 0.019301 | 1 | 6.61E-26 | 1 |
| 19-44916825-A-C | C | | | | | | | | | 0.089321 | 7.55E-21 | TRUE | 7.55E-21 | ?- | 0.837118 | 1 | 0.124506 | 1 | 1.24E-20 | 1 |
| 19-44921093-TAA-T | T | | | | | | | | | 0.095653 | 3.33E-19 | TRUE | 3.34E-19 | ?+ | 0.845196 | -1 | 0.152392 | 1 | 7.92E-19 | 1 |
| 19-44921095-A-ATTTT | ATTTT | | | | | | | 5.95E-05 | 0.946693 | 0.110797 | 2.64E-19 | TRUE | 2.68E-19 | -- | 0.649637 | 1 | 0.228405 | 1 | 1.79E-18 | 1 |
| 19-45086157-A-G | G | 0.439369 | 0.289745 | 0.466 | 0.844857 | 0.8449 | ?+ | 0.5243 | 0.011399 | 0.607371 | 5.61E-09 | FALSE | 1.92E-07 | -- | 0.127965 | 1 | 0.176479 | 1 | 5.89E-08 | 1 |
| 19-45108380-TC-T | T | 0.000918 | 0.769252 | | | 0.7693 | -? | | | 0.00114 | 1.17E-09 | TRUE | 1.17E-09 | ?+ | 0.011509 | 1 | 0.002638 | 1 | 6.88E-07 | 1 |
| 19-45260078-CT-C | C | | | | | | | | | 0.000822 | 3.1E-13 | TRUE | 3.1E-13 | ?- | 0.001243 | 1 | 6.28E-05 | 1 | 2.04E-09 | 1 |
| 19-45260080-CT-C | C | | | | | | | | | 0.000807 | 1.57E-10 | TRUE | 1.57E-10 | ?- | 0.013309 | 1 | 7.69E-05 | 1 | 5.88E-08 | 1 |
| 19-45436408-C-CCT | CCT | | | | | | | | | 0.000871 | 7.8E-09 | TRUE | 7.81E-09 | ?+ | 0.696936 | 1 | 0.177593 | 1 | 1.43E-07 | 1 |
| 19-45594721-GCCTGT | G | 0.037749 | 0.96823 | | | 0.9682 | -? | | | 0.053933 | 3.37E-09 | FALSE | 3.37E-09 | ?+ | 0.123764 | 1 | 0.576824 | 1 | 1.18E-08 | 1 |
| 19-46832405-GGC-G | G | | | | | | | | | 0.006123 | 2.68E-11 | TRUE | 2.68E-11 | ?+ | 0.438007 | 1 | 0.180719 | 1 | 4.56E-11 | 1 |
| 19-46832408-C-CAG | CAG | | | | | | | | | 0.003457 | 1.06E-12 | TRUE | 1.06E-12 | ?+ | 0.124969 | 1 | 0.159706 | 1 | 1.81E-11 | 1 |
| 19-46844565-A-AGG | AGG | | | | | | | | | 0.001115 | 7.85E-09 | TRUE | 7.85E-09 | ?- | 1.48E-06 | 1 | 0.427463 | 1 | 4.21E-05 | 1 |
| 19-46844567-GCA-G | G | | | | | | | | | 0.000477 | 8.21E-10 | TRUE | 8.21E-10 | ?+ | 0.00013 | 1 | 0.958107 | -1 | 2.29E-07 | 1 |
| 19-47213512-T-TC | TC | 0.000234 | 0.464375 | | | 0.4644 | -? | | | 0.001485 | 7.6E-10 | TRUE | 7.6E-10 | ?- | 0.666478 | 1 | 0.192678 | 1 | 2.05E-09 | 1 |
| 19-47458598-CTCT-C | C | | | | | | | | | 0.003011 | 4.3E-08 | TRUE | 4.3E-08 | ?- | 0.28819 | 1 | 0.013067 | 1 | 4.91E-07 | 1 |
| 19-47604922-G-A | A | 0.00272 | 0.23491 | | | 0.2349 | -? | 0.001723 | 0.187892 | 0.000846 | 3.13E-10 | TRUE | 0.0166 | -+ | 0.049632 | 1 | 0.69054 | -1 | 1.11E-09 | 1 |
| 19-48188862-T-A | A | 0.000235 | 0.079083 | | | 0.07908 | -? | 0.001215 | 0.390055 | 0.001307 | 4.94E-14 | TRUE | 2.77E-06 | -+ | 0.037734 | 1 | 0.013227 | 1 | 5.37E-12 | 1 |
| 19-48189428-C-T | T | 0.000195 | 0.771062 | | | 0.7711 | -? | 0.001704 | 0.850937 | 0.000694 | 3.88E-09 | TRUE | 0.003146 | -+ | 0.061311 | 1 | 0.574963 | 1 | 1.39E-08 | 1 |
| 19-48564935-TGC-T | T | | | | | | | | | 0.003156 | 3.79E-28 | TRUE | 3.79E-28 | ?+ | 1.58E-08 | 1 | 0.000974 | 1 | 5.31E-20 | 1 |
| 19-48690221-C-T | T | | | | | | | | | 0.120672 | 4.33E-13 | TRUE | 4.33E-13 | ?+ | 0.027212 | 1 | 0.468805 | 1 | 0.781047 | 1 |
| 19-48691269-T-C | C | 0.109161 | 0.991622 | | | 0.9916 | +? | | | 0.122513 | 7.4E-11 | FALSE | 7.4E-11 | ?- | 0.101771 | 1 | 0.516858 | 1 | 0.763417 | 1 |
| 19-48777085-G-C | C | 0.000176 | 0.065263 | | | 0.06526 | -? | | | 0.00063 | 1.92E-10 | TRUE | 1.92E-10 | ?+ | 0.190182 | 1 | 0.016876 | 1 | 1.5E-09 | 1 |
| 19-48916454-C-CG | CG | | | | | | | | | 0.000671 | 1.75E-10 | TRUE | 1.75E-10 | ?+ | 0.918221 | -1 | 0.106254 | 1 | 1.24E-10 | 1 |
| 19-49206127-ATTT-A | A | 0.000569 | 0.610342 | | | 0.6103 | +? | | | 0.002808 | 1.56E-14 | TRUE | 1.56E-14 | ?+ | 0.000123 | 1 | 0.098408 | 1 | 5.2E-12 | 1 |
| 19-49291839-C-T | T | | | | | | | 0.007226 | 0.559414 | 0.001404 | 3.18E-11 | TRUE | 0.00224 | ++ | 0.081148 | 1 | 5.18E-05 | 1 | 1.03E-06 | 1 |
| 19-49714558-GCA-G | G | | | | | | | | | 0.000671 | 3.54E-11 | TRUE | 3.54E-11 | ?+ | 0.000775 | 1 | 0.010746 | 1 | 5.46E-08 | 1 |
| 19-49905891-C-CCT | CCT | | | | | | | | | 0.001162 | 2.19E-10 | TRUE | 2.19E-10 | ?+ | 0.004806 | 1 | 0.001664 | 1 | 4.26E-07 | 1 |
| 19-52384389-GT-G | G | | | | | | | | | 0.004467 | 1.82E-09 | TRUE | 1.82E-09 | ?+ | 0.312566 | 1 | 0.010978 | 1 | 3.76E-08 | 1 |
| 19-53629086-GGC-G | G | | | | | | | | | 0.000273 | 8.03E-10 | TRUE | 8.03E-10 | ?+ | | | 0.003432 | 1 | 5.3E-08 | 1 |

| | | | | | | | | | | | | | | | | | | | | |
|---|---|---|---|---|---|---|---|---|---|---|---|---|---|---|---|---|---|---|---|---|
| 19-53632731-T-A | A | 0.426458 | 0.104623 | 0.4 | 0.143297 | 0.08384 | ++ | 0.377919 | 0.33713 | 0.463464 | 5.41E-10 | FALSE | 0.008821 | -+ | 0.173476 | 1 | 0.556182 | -1 | 1.94E-09 | 1 |
| 19-53947162-G-T | T | | | | | | | | | 0.000527 | 3.43E-10 | TRUE | 3.43E-10 | ?+ | 0.012162 | 1 | 0.00252 | 1 | 1.77E-07 | 1 |
| 19-53950357-CTG-C | C | | | | | | | | | 0.001194 | 2.15E-10 | TRUE | 2.15E-10 | ?- | 0.403665 | 1 | 0.104348 | 1 | 7.91E-10 | 1 |
| 19-54059216-A-AAGC | AAGCC | | | | | | | | | 0.000829 | 2.61E-08 | TRUE | 2.61E-08 | ?- | 0.255934 | 1 | 0.514657 | 1 | 1.9E-08 | 1 |
| 19-54953825-A-ATC | ATC | 0.000371 | 0.88896 | | | 0.889 | -? | | | 0.001814 | 6.4E-10 | TRUE | 6.4E-10 | ?- | 0.089218 | 1 | 0.084801 | 1 | 1.33E-08 | 1 |
| 19-55397554-GT-G | G | | | | | | | | | 0.000641 | 1.99E-09 | TRUE | 1.99E-09 | ?+ | 0.030802 | 1 | 0.200201 | 1 | 4.76E-08 | 1 |
| 19-55421214-TTAAGA | T | 0.000353 | 0.480108 | | | 0.4801 | -? | | | 0.000636 | 2.41E-08 | TRUE | 2.41E-08 | ?+ | 0.87198 | -1 | 0.48899 | 1 | 9.14E-10 | 1 |
| 19-55422561-G-GGT | GGT | | | | | | | | | 0.001367 | 1.06E-19 | TRUE | 1.06E-19 | ?- | 0.529535 | 1 | 0.005906 | 1 | 6.4E-18 | 1 |
| 19-55422571-A-ATGC | ATGC | | | | | | | | | 0.000807 | 6.12E-11 | TRUE | 6.12E-11 | ?- | 0.83399 | -1 | 0.314122 | 1 | 5.68E-10 | 1 |
| 19-55422574-ACGCCT | A | | | | | | | | | 0.000471 | 4.04E-08 | TRUE | 4.04E-08 | ?+ | 0.385473 | -1 | 0.997076 | -1 | 1.73E-09 | 1 |
| 19-56879866-GT-G | G | | | | | | | | | 0.001029 | 4.96E-14 | TRUE | 4.96E-14 | ?+ | 0.006189 | 1 | 0.869249 | 1 | 3.91E-13 | 1 |
| 19-57064425-A-C | C | | | | | | | | | 8.71E-05 | 1.08E-09 | TRUE | 1.08E-09 | ?- | | | | | 9.08E-06 | 1 |
| 19-57066412-CA-C | C | | | | | | | | | 0.002177 | 8.12E-14 | TRUE | 8.12E-14 | ?- | 0.026942 | 1 | 0.214952 | 1 | 1.61E-12 | 1 |
| 19-58524353-A-G | G | 0.000394 | 0.800816 | | | 0.8008 | +? | | | 0.000399 | 7.95E-10 | TRUE | 7.95E-10 | ?- | 0.117257 | 1 | 0.040021 | 1 | 1.28E-09 | 1 |
| 19-58542262-GA-G | G | | | | | | | 0.99656 | 0.54052 | 0.988054 | 2.01E-09 | TRUE | 1.59E-07 | -- | 0.017448 | -1 | 0.138409 | -1 | 1.82E-08 | -1 |
| 19-44730062-AAATTT. | A | 0.08739 | 0.002298 | | | 0.002298 | -? | 0.090385 | 1.06E-07 | 0.08507 | 0.024845 | TRUE | 1.7E-08 | -- | 0.163766 | -1 | 0.395045 | 1 | 0.030029 | -1 |
| 19-45008063-G-A | A | 0.003692 | 0.000192 | 0.007 | 0.005858 | 0.0001676 | ++ | 0.005315 | 2.1E-06 | 0.002993 | 0.001696 | TRUE | 1.31E-08 | ++ | 0.581197 | 1 | 0.040476 | 1 | 0.003563 | 1 |
| 19-56055154-ACT-A | A | | | | | | | 4.07E-05 | 0.060619 | 0.00092 | 5.34E-08 | TRUE | 1.13E-08 | ++ | 0.00256 | 1 | 0.004235 | 1 | 2.83E-05 | 1 |
| 19-44732593-AAG-A | A | 0.085763 | 0.001493 | | | 0.001493 | -? | 0.090272 | 6.35E-08 | 0.083504 | 0.016805 | TRUE | 6.58E-09 | -- | 0.099681 | -1 | 0.33001 | 1 | 0.02074 | -1 |
| 19-44960141-G-A | A | | | 0.01 | 0.002381 | 0.002381 | ?- | 0.006218 | 9.51E-07 | 0.003891 | 0.003205 | TRUE | 1.09E-08 | -- | 0.082589 | 1 | 0.159619 | 1 | 0.00903 | 1 |
| 19-44909967-TGGG-T | TG | | | | | | | 0.149744 | 0 | | | FALSE | 5.9E-305 | +? | | | | | | |
| 19-44736280-C-A | A | 0.101576 | 0.000491 | 0.081 | 0.910854 | 0.0005059 | -+ | 0.091234 | 8.25E-08 | 0.102645 | 0.048557 | TRUE | 3.84E-08 | -- | 0.781927 | -1 | 0.437983 | 1 | 0.024556 | -1 |
| 19-44799247-G-A | A | 0.007405 | 2.13E-06 | 0.007 | 0.380053 | 0.000002 | -- | 0.022947 | 1.47E-07 | 0.012799 | 0.010288 | TRUE | 5.97E-09 | -- | 0.891187 | -1 | 0.931662 | -1 | 0.01094 | -1 |
| 19-45176297-C-CA | CA | | | | | | | 0.094119 | 2.59E-06 | 0.113348 | 2.01E-06 | TRUE | 4.24E-11 | ++ | 0.498414 | -1 | 0.741541 | -1 | 1.61E-07 | 1 |
| 19-44900801-C-T | T | 0.00113 | 0.004724 | | | 0.004724 | +? | 0.001735 | 9.49E-08 | 0.000668 | 0.010016 | TRUE | 3.22E-09 | ++ | 0.036277 | 1 | 0.18623 | -1 | 0.016062 | 1 |
| 19-45226980-C-T | T | | | 0.065 | 0.233 | 0.233 | ?- | 0.044525 | 6.98E-07 | 0.073408 | 0.004155 | TRUE | 1.44E-08 | ++ | 0.010954 | 1 | 0.216498 | 1 | 0.221895 | 1 |
| 19-7236842-C-T | T | 9.75E-05 | 0.829837 | | | 0.8298 | +? | 1.88E-05 | 0.097089 | 0.000508 | 1.19E-07 | TRUE | 3.77E-08 | ++ | 0.013701 | 1 | 0.355414 | -1 | 9.94E-07 | 1 |
| 19-45410565-TTGTGT | T | | | | | | | | | | | FALSE | | | | | | | | |
| 19-44732598-TGAA-T | T | 0.085757 | 0.001524 | | | 0.001524 | -? | 0.090272 | 6.35E-08 | 0.083504 | 0.016787 | TRUE | 6.57E-09 | -- | 0.099547 | -1 | 0.330143 | 1 | 0.020733 | -1 |
| 19-44869757-GT-G | G | 0.001111 | 0.003089 | | | 0.003089 | +? | 0.001688 | 3.86E-07 | 0.000668 | 0.0202 | TRUE | 2.39E-08 | ++ | | | 0.175162 | -1 | 0.016308 | 1 |
| 19-44728555-G-A | A | 0.085582 | 0.004941 | 0.071 | 0.91233 | 0.005049 | -+ | 0.090075 | 8.71E-08 | 0.083811 | 0.014381 | TRUE | 7.16E-09 | -- | 0.10193 | -1 | 0.438651 | 1 | 0.027954 | -1 |
| 19-45208252-G-A | A | 0.328468 | 0.110053 | 0.376 | 0.996685 | 0.111 | -- | 0.620882 | 1.82E-06 | 0.694076 | 2.01E-06 | FALSE | 5.41E-11 | -- | 0.058757 | 1 | 0.721639 | 1 | 4.35E-05 | 1 |
| 19-38498805-G-T | T | | | | | | | 1.25E-05 | 0.633286 | | | FALSE | 2.9E-305 | +? | | | | | | |
| 19-44882164-C-T | T | 0.001111 | 0.003097 | | | 0.003097 | -? | 0.001751 | 4.62E-07 | 0.000701 | 0.027219 | TRUE | 3.72E-08 | -- | | | 0.168273 | -1 | 0.019689 | 1 |
| 19-44693694-G-A | A | 0.010814 | 0.000615 | 0.01 | 0.057771 | 0.0007302 | -+ | 0.019868 | 2.44E-06 | 0.011821 | 0.0042 | TRUE | 3.5E-08 | -- | 0.867567 | -1 | 0.251774 | 1 | 0.001115 | -1 |
| 19-44731001-AAAAA | A | | | | | | | | | | | TRUE | | | | | | | | |
| 19-44730118-G-A | A | 0.076429 | 0.001064 | 0.07 | 0.781813 | 0.001118 | +- | 0.089633 | 9.11E-08 | 0.078352 | 0.011131 | TRUE | 5.22E-09 | ++ | 0.122137 | -1 | 0.623488 | 1 | 0.028847 | -1 |
| 19-44748549-G-T | T | 0.008315 | 3.6E-05 | | | 0.00003602 | -? | 0.022834 | 6.22E-08 | 0.012853 | 0.001954 | TRUE | 4.53E-10 | -- | 0.535683 | -1 | 0.682028 | 1 | 0.001882 | -1 |
| 19-45223710-G-A | A | 0.073219 | 0.004096 | 0.065 | 0.184471 | 0.00475 | +- | 0.044898 | 8.02E-07 | 0.077629 | 0.00104 | TRUE | 3.61E-09 | ++ | 0.00676 | 1 | 0.172311 | 1 | 0.155527 | 1 |
| 19-44898730-G-A | A | | | 0.004 | 0.311118 | 0.3111 | ?+ | 0.003527 | 6.27E-08 | 0.001959 | 0.072424 | TRUE | 2.01E-08 | ++ | 0.014703 | 1 | 0.812375 | 1 | 0.159452 | 1 |
| 19-45209858-C-A | A | 0.01024 | 0.03452 | 0.019 | 0.187152 | 0.03249 | -- | 0.014221 | 2.48E-07 | 0.008627 | 0.020505 | TRUE | 1.97E-08 | -- | 0.573142 | 1 | 0.807059 | -1 | 0.009148 | 1 |
| 19-44714520-G-C | C | 0.1011 | 0.057795 | | | 0.05779 | +? | 0.849914 | 7.13E-08 | 0.890458 | 0.082528 | TRUE | 4.97E-08 | ++ | 0.684797 | -1 | 0.673418 | 1 | 0.017631 | -1 |
| 19-44735929-G-A | A | 0.374506 | 0.00152 | 0.334 | 0.741141 | 0.7411 | ?+ | 0.329287 | 4.02E-07 | 0.373931 | 0.004164 | TRUE | 7.09E-09 | ++ | 0.343833 | 1 | 0.515535 | 1 | 0.000739 | -1 |
| 19-45171922-G-A | A | 0.011147 | 0.024056 | | | 0.02406 | -? | 0.015279 | 3.76E-07 | 0.008949 | 0.030631 | TRUE | 4.47E-08 | -- | 0.648902 | 1 | 0.98316 | 1 | 0.02504 | 1 |
| 20-51028527-T-C | C | 0.298425 | 3.11E-08 | 0.202 | 0.29021 | 2.269E-08 | ++ | | | 0.279474 | 0.708753 | TRUE | 0.7088 | ?+ | 0.854512 | -1 | 0.307041 | -1 | 0.72175 | -1 |
| 20-51033011-T-C | C | 0.301697 | 6E-09 | 0.212 | 0.315701 | 4.384E-09 | ++ | | | 0.283286 | 0.647046 | TRUE | 0.647 | ?+ | 0.712415 | -1 | 0.236435 | -1 | 0.741099 | -1 |
| 20-51036345-T-A | A | 0.284364 | 1.27E-08 | 0.205 | 0.483236 | 1.044E-08 | -- | | | 0.264266 | 0.473965 | TRUE | 0.474 | ?- | 0.359545 | -1 | 0.315153 | -1 | 0.669971 | -1 |
| 20-51038035-A-G | G | 0.291175 | 2.75E-09 | 0.203 | 0.459513 | 2.23E-09 | ++ | | | 0.271064 | 0.454512 | TRUE | 0.4545 | ?+ | 0.471189 | -1 | 0.303459 | -1 | 0.594492 | -1 |
| 20-51041177-A-G | G | 0.293673 | 9.02E-09 | 0.202 | 0.424146 | 7.163E-09 | ++ | | | 0.274521 | 0.390661 | TRUE | 0.3907 | ?+ | 0.616103 | -1 | 0.241715 | -1 | 0.47917 | -1 |
| 20-506117-C-T | T | | | | | | | 0.000874 | 0.835076 | 0.001326 | 9.93E-12 | TRUE | 4.46E-07 | -+ | 0.020877 | 1 | 0.023258 | 1 | 5.38E-10 | 1 |
| 20-506120-C-T | T | 0.001357 | 0.794458 | | | 0.7945 | -? | 0.000871 | 0.873483 | 0.00141 | 3.06E-10 | TRUE | 2.92E-06 | -+ | 0.284323 | 1 | 0.115815 | 1 | 2.97E-09 | 1 |
| 20-3850050-G-A | A | 0.02368 | 0.018174 | | | 0.01817 | -? | | | 0.030363 | 6.72E-10 | TRUE | 6.72E-10 | ?+ | 0.018252 | 1 | 0.20121 | -1 | 1.57E-11 | 1 |
| 20-3850101-A-AC | AC | 0.000177 | 0.043757 | | | 0.04376 | +? | 0.002096 | 0.307591 | 0.00338 | 1.8E-28 | TRUE | 4.97E-15 | +- | 0.002324 | 1 | 0.004646 | 1 | 9.94E-25 | 1 |
| 20-3978903-CA-C | C | | | | | | | 0.000279 | 0.915307 | 0.001368 | 3.07E-20 | TRUE | 6.56E-17 | +- | 2.69E-05 | 1 | 0.120948 | 1 | 1.54E-16 | 1 |
| 20-4081212-AAT-A | A | 0.002059 | 0.914616 | | | 0.9146 | -? | | | 0.001828 | 4.02E-08 | TRUE | 4.02E-08 | ?+ | 0.246458 | 1 | 0.082651 | 1 | 5.07E-07 | 1 |
| 20-16241716-C-A | A | | | | | | | 0.002725 | 0.905911 | 0.001243 | 6.8E-14 | TRUE | 2E-05 | ++ | 0.477755 | 1 | 0.017302 | 1 | 1.47E-11 | 1 |
| 20-16258104-G-C | C | | | | | | | 0.998608 | 0.750428 | 0.916713 | 4.64E-08 | FALSE | 1.22E-07 | -+ | 0.191147 | -1 | 0.725092 | 1 | 1.28E-09 | 1 |
| 20-16636840-A-T | T | | | | | | | 0.001526 | 0.667896 | 0.001376 | 2.8E-08 | TRUE | 0.001064 | +- | 0.13513 | 1 | 0.094619 | 1 | 8.79E-08 | 1 |
| 20-20356085-A-G | G | | | | | | | | | 0.92907 | 9.49E-17 | FALSE | 9.49E-17 | ?+ | 0.661483 | -1 | 0.719313 | 1 | 1.1E-26 | -1 |

| | | | | | | | | | | | | | | | | | |
|---|---|---|---|---|---|---|---|---|---|---|---|---|---|---|---|---|---|
| 20-20356115-C-CTG | CTG | | | | | | 0.894517 | 5.2E-15 | FALSE | 5.2E-15 | ?- | 0.740285 | 1 | 0.492182 | -1 | 2.91E-19 | -1 |
| 20-24625974-C-T | T | | | | | 0.254564 | 0.792594 | 0.244087 | 1.17E-09 | FALSE | 0.000297 | ++ | 0.077861 | 1 | 0.143131 | 1 | 1.37E-07 | 1 |
| 20-25885058-G-GT | GT | | | | | | 0.027226 | 1.39E-10 | FALSE | 1.39E-10 | ?- | 0.059411 | 1 | 0.394557 | 1 | 1.32E-09 | 1 |
| 20-26363358-T-A | A | | | | | 9.41E-06 | 0.464988 | 0.000163 | 2.06E-08 | TRUE | 8.38E-08 | -+ | | | | | 2.42E-07 | 1 |
| 20-30361946-A-C | C | | | | | 0.491045 | 0.522025 | 0.586424 | 1.7E-08 | FALSE | 0.01278 | +- | 0.783183 | 1 | 0.168366 | 1 | 1.17E-07 | 1 |
| 20-30397502-T-TA | TA | | | | | | 0.504417 | 5.14E-12 | FALSE | 5.14E-12 | ?- | 0.196931 | 1 | 0.017353 | 1 | 4.69E-10 | 1 |
| 20-30397502-T-TAA | TAA | | | | | | 0.483441 | 1.08E-14 | FALSE | 1.08E-14 | ?+ | 0.006498 | -1 | 0.013437 | -1 | 1.68E-10 | -1 |
| 20-30424031-T-TTGTG | TTGTGTG | | | | | | 0.008143 | 1.19E-20 | TRUE | 1.19E-20 | ?- | 0.076805 | 1 | 0.286494 | 1 | 3.32E-18 | 1 |
| 20-31053559-A-G | G | | | | | | 0.878989 | 1.17E-08 | FALSE | 1.17E-08 | ?+ | 0.011015 | -1 | 0.324508 | -1 | 2.15E-07 | -1 |
| 20-31073278-A-T | T | | | | | | 0.928109 | 4.52E-16 | FALSE | 4.52E-16 | ?+ | 0.006239 | -1 | 0.012875 | -1 | 7.23E-13 | -1 |
| 20-31073302-T-A | A | | | | | | 0.908347 | 4.1E-22 | FALSE | 4.1E-22 | ?- | 0.063766 | -1 | 0.020078 | -1 | 1.11E-19 | -1 |
| 20-31073335-G-C | C | | | | | | 0.878506 | 7.59E-12 | FALSE | 7.59E-12 | ?- | 0.134008 | -1 | 0.048259 | -1 | 2.15E-10 | -1 |
| 20-31074716-A-T | T | | | | | | 0.005813 | 2.51E-08 | TRUE | 2.51E-08 | ?- | 0.269863 | 1 | 0.004659 | 1 | 4.59E-06 | 1 |
| 20-31185833-GAATGC | G | | | | | | 0.060191 | 4.98E-10 | FALSE | 4.98E-10 | ?- | 0.384378 | -1 | 0.515111 | 1 | 1.57E-10 | -1 |
| 20-31582540-A-G | G | | | | | 0.000238 | 0.517815 | 0.000509 | 2.71E-08 | TRUE | 7.86E-07 | -- | 0.013309 | 1 | 0.388516 | 1 | 7.32E-06 | 1 |
| 20-31656375-GGC-G | G | 0.005409 | 0.045832 | 0.04583 | -? | | | 0.004074 | 1E-08 | TRUE | 1E-08 | ?+ | 0.762929 | -1 | 0.372351 | -1 | 6.8E-12 | 1 |
| 20-31703178-G-GA | GA | 0.000411 | 0.66486 | 0.6649 | +? | | | 0.002869 | 7.04E-11 | TRUE | 7.04E-11 | ?- | 0.010184 | 1 | 0.698855 | -1 | 1.74E-11 | 1 |
| 20-31750080-GGC-G | G | 9.75E-05 | 0.36705 | 0.367 | -? | | | 0.002986 | 1.38E-16 | TRUE | 1.38E-16 | ?+ | 0.000375 | 1 | 0.195027 | 1 | 4.04E-15 | 1 |
| 20-32019470-CACCTT | C | | | | | | | 0.005667 | 2.65E-08 | TRUE | 2.65E-08 | ?- | 0.629043 | 1 | 0.488671 | 1 | 2.08E-08 | 1 |
| 20-32296095-C-G | G | | | | | 2.51E-05 | 0.213378 | 0.000655 | 7.61E-10 | TRUE | 7.29E-09 | +- | 0.038413 | 1 | 5.27E-06 | 1 | 3.66E-05 | 1 |
| 20-32735144-AG-A | A | | | | | | | 0.000336 | 3.59E-10 | TRUE | 3.59E-10 | ?+ | 0.094956 | 1 | 0.105653 | 1 | 1.37E-09 | 1 |
| 20-34646551-G-GCC | GCC | | | | | | | 0.000784 | 6.01E-14 | TRUE | 6.01E-14 | ?- | 0.901872 | -1 | 0.231973 | 1 | 3.51E-13 | 1 |
| 20-34646555-CCT-C | C | | | | | | | 0.000561 | 1.74E-11 | TRUE | 1.74E-11 | ?- | 0.358264 | -1 | 0.064829 | 1 | 1.23E-11 | 1 |
| 20-35097276-CAT-C | C | | | | | | | 0.026314 | 1.34E-09 | TRUE | 1.34E-09 | ?- | 0.845359 | 1 | 0.021037 | 1 | 1.99E-09 | 1 |
| 20-36058190-T-C | C | | | | | 0.001544 | 0.865026 | 0.000454 | 2.57E-08 | TRUE | 0.01047 | +- | 0.713896 | 1 | 0.871586 | 1 | 2.46E-08 | 1 |
| 20-36058191-G-A | A | | | | | 0.001657 | 0.965526 | 0.000689 | 2.66E-10 | TRUE | 0.000441 | ++ | 0.323776 | 1 | 0.501031 | 1 | 3.2E-10 | 1 |
| 20-36058218-AT-A | A | | | | | 0.00063 | 0.244962 | 0.000985 | 5.53E-09 | TRUE | 2.34E-07 | ++ | 0.311008 | 1 | 0.326956 | 1 | 7.98E-09 | 1 |
| 20-36190592-C-A | A | 0.000195 | 0.827256 | 0.8273 | +? | 0.000705 | 0.69562 | 0.002657 | 6.45E-22 | TRUE | 1.84E-16 | -+ | 0.000342 | 1 | 3.05E-05 | 1 | 3.41E-13 | 1 |
| 20-36190632-C-CTTAC | CTTACA | | | | | | | 0.001496 | 8.14E-17 | TRUE | 8.14E-17 | ?+ | 0.014239 | 1 | 0.019792 | 1 | 1.89E-15 | 1 |
| 20-36191164-A-C | C | 0.000313 | 0.350784 | 0.3508 | +? | 0.000345 | 0.040447 | 0.001458 | 5.7E-13 | TRUE | 7.36E-09 | +- | 0.015016 | 1 | 0.367127 | 1 | 2.86E-12 | 1 |
| 20-36191165-C-T | T | 0.000293 | 0.542816 | 0.5428 | -? | 0.000341 | 0.041266 | 0.001038 | 2.73E-08 | TRUE | 7.63E-05 | -+ | 0.067169 | 1 | 0.041215 | 1 | 5.46E-07 | 1 |
| 20-36728254-C-CCG | CCG | | | | | | | 0.000628 | 1.81E-08 | TRUE | 1.81E-08 | ?+ | 0.447159 | 1 | 0.003985 | 1 | 3.36E-07 | 1 |
| 20-36767785-T-C | C | | | | | 1.88E-05 | 0.920236 | 0.000293 | 1.74E-08 | TRUE | 5.19E-08 | +- | 0.025294 | 1 | 0.504389 | -1 | 1.52E-08 | 1 |
| 20-36787692-CCTGGC | C | | | | | | | 0.001555 | 2.3E-16 | TRUE | 2.3E-16 | ?- | 0.000343 | 1 | 0.010077 | 1 | 2.77E-13 | 1 |
| 20-36949029-C-CTG | CTG | 0.000371 | 0.958116 | 0.9581 | +? | | | 0.005247 | 2.16E-36 | TRUE | 2.16E-36 | ?+ | 0.919581 | 1 | 3.82E-08 | 1 | 1.4E-31 | 1 |
| 20-36949031-G-GGC | GGC | 0.000371 | 0.95811 | 0.9581 | -? | | | 0.004609 | 2.44E-29 | TRUE | 2.44E-29 | ?- | 0.841297 | 1 | 3.31E-06 | 1 | 6.84E-26 | 1 |
| 20-36949037-GATGC- | G | | | | | | | 0.001679 | 3.74E-15 | TRUE | 3.74E-15 | ?+ | 0.841247 | -1 | 0.00017 | 1 | 2.88E-12 | 1 |
| 20-36949046-CCT-C | C | 0.000137 | 0.797772 | 0.7978 | -? | | | 0.001617 | 3.96E-16 | TRUE | 3.96E-16 | ?- | 0.899083 | -1 | 0.003119 | 1 | 7.92E-15 | 1 |
| 20-37109452-CCT-C | C | 0.000215 | 0.223075 | 0.2231 | +? | | | 0.001834 | 1.66E-08 | TRUE | 1.66E-08 | ?- | 0.051065 | 1 | 0.646025 | 1 | 2.13E-08 | 1 |
| 20-38850804-TA-T | T | | | | | | | 0.000975 | 1.64E-10 | TRUE | 1.64E-10 | ?+ | 0.215637 | -1 | 0.014889 | 1 | 1.47E-09 | 1 |
| 20-39708990-TAAAAA | T | 0.427558 | 0.641374 | 0.6414 | +? | | | 0.605596 | 4.29E-14 | FALSE | 4.29E-14 | ?- | 0.334045 | -1 | 0.141709 | -1 | 5.08E-13 | -1 |
| 20-43469358-A-ATTT | ATTT | | | | | | | 0.002189 | 1.48E-16 | TRUE | 1.48E-16 | ?- | 0.023715 | 1 | 0.000323 | 1 | 1.93E-13 | 1 |
| 20-43652786-A-ACC | ACC | | | | | | | 0.000846 | 5.18E-11 | TRUE | 5.18E-11 | ?- | 0.005688 | 1 | 0.303511 | 1 | 2.43E-09 | 1 |
| 20-43652787-TGA-T | T | | | | | | | 0.000483 | 2.66E-09 | TRUE | 2.66E-09 | ?+ | 0.020826 | 1 | 0.262484 | 1 | 4.63E-08 | 1 |
| 20-45931856-ACC-A | A | | | | | | | 0.000757 | 4.09E-10 | TRUE | 4.09E-10 | ?+ | 0.192294 | 1 | 0.399772 | 1 | 6.23E-10 | 1 |
| 20-47194165-GCA-G | G | | | | | | | 0.001073 | 1.15E-08 | TRUE | 1.16E-08 | ?+ | 0.649431 | -1 | 0.164133 | 1 | 4.06E-09 | 1 |
| 20-47381012-A-ACT | ACT | | | | | | | 0.001063 | 2.67E-13 | TRUE | 2.67E-13 | ?- | 0.003187 | 1 | 0.01283 | 1 | 3.46E-10 | 1 |
| 20-48501959-AG-A | A | 0.000488 | 0.372408 | 0.3724 | -? | | | 0.002587 | 2.43E-09 | TRUE | 2.43E-09 | ?+ | 0.011915 | 1 | 0.048672 | 1 | 6.89E-07 | 1 |
| 20-48501962-A-AG | AG | 0.000473 | 0.438574 | 0.4386 | +? | | | 0.002083 | 3.17E-11 | TRUE | 3.17E-11 | ?- | 0.002731 | 1 | 0.013388 | 1 | 1.25E-07 | 1 |
| 20-49061328-ATG-A | A | | | | | | | 0.004325 | 4.05E-11 | TRUE | 4.05E-11 | ?+ | 0.151192 | 1 | 0.004234 | 1 | 2.31E-08 | 1 |
| 20-49061331-CCT-C | C | 0.002212 | 0.289827 | 0.2898 | +? | | | 0.003382 | 1.37E-10 | TRUE | 1.37E-10 | ?- | 0.199105 | 1 | 0.003674 | 1 | 1.45E-07 | 1 |
| 20-49198329-C-CA | CA | 0.000391 | 0.342341 | 0.3423 | -? | | | 0.001324 | 4.39E-08 | TRUE | 4.39E-08 | ?+ | 0.012124 | 1 | 0.038257 | 1 | 1.05E-05 | 1 |
| 20-49202549-GCA-G | G | 9.75E-05 | 0.693791 | 0.6938 | +? | | | 0.000453 | 3.42E-08 | TRUE | 3.42E-08 | ?+ | 0.020728 | 1 | 0.298669 | 1 | 7.82E-07 | 1 |
| 20-51520826-A-AT | AT | | | | | | | 0.000265 | 4.97E-08 | TRUE | 4.97E-08 | ?- | 0.012274 | 1 | 0.000449 | 1 | 6.08E-05 | 1 |
| 20-51520827-A-T | T | | | | | | | 0.000502 | 7.78E-10 | TRUE | 7.78E-10 | ?- | 0.00018 | 1 | 0.021371 | 1 | 3.4E-07 | 1 |
| 20-53482754-A-G | G | | | | | 0.000865 | 0.222995 | 0.003473 | 4.27E-10 | TRUE | 2.4E-09 | -- | 0.146764 | 1 | 0.01518 | 1 | 1.98E-07 | 1 |
| 20-53482760-G-A | A | 0.000176 | 0.738393 | 0.7384 | +? | 0.000273 | 0.837682 | 0.001489 | 1.03E-08 | TRUE | 1.69E-07 | ++ | 0.157606 | 1 | 0.635732 | 1 | 3.07E-07 | 1 |
| 20-53698443-G-C | C | | | | | | | 0.001015 | 7.67E-12 | TRUE | 7.67E-12 | ?+ | 0.055942 | 1 | 0.000251 | 1 | 2.5E-08 | 1 |
| 20-59401313-GCA-G | G | | | | | | | 0.983482 | 8.69E-11 | TRUE | 8.69E-11 | ?- | 0.012625 | -1 | 0.905537 | -1 | 2.08E-11 | -1 |

| | | | | | | | | | | | | | | | | | | | | |
|---|---|---|---|---|---|---|---|---|---|---|---|---|---|---|---|---|---|---|---|---|
| 20-61290436-C-CCGT | CCGTATCCAAT | | | | | | | | | 0.024467 | 1.18E-09 | FALSE | 1.18E-09 | ?- | 0.1071 | -1 | 0.172951 | -1 | 4.21E-08 | -1 |
| 20-62278275-T-TTG | TTG | | | | | | | 0.996363 | 0.270412 | 0.968847 | 7.67E-10 | TRUE | 6.84E-07 | -+ | 0.03231 | -1 | 0.313755 | -1 | 1.74E-08 | -1 |
| 20-62399827-C-T | T | | | | | | | 0.000241 | 0.673279 | 0.001363 | 2.28E-10 | TRUE | 2.66E-08 | -+ | 0.016947 | 1 | 0.358538 | 1 | 9.15E-10 | 1 |
| 20-62865769-C-CTT | CTT | | | | | | | | | 0.000925 | 1.1E-12 | TRUE | 1.1E-12 | ?+ | 2.22E-05 | 1 | 0.014485 | 1 | 1.28E-08 | 1 |
| 20-62865772-G-T | T | | | | | | | | | 0.000398 | 1.93E-09 | TRUE | 1.93E-09 | ?+ | 0.001137 | 1 | 0.035408 | 1 | 7.78E-07 | 1 |
| 20-63461661-GGGA-G | G | | | | | | | 9.4E-06 | 0.461209 | 0.00432 | 3.04E-09 | TRUE | 4.14E-09 | -+ | 0.174683 | 1 | 0.008705 | 1 | 1.53E-07 | 1 |
| 20-63821488-G-A | A | | | | | | | 0.000742 | 0.144102 | 0.000701 | 1.85E-08 | TRUE | 1.2E-06 | ++ | 0.804423 | -1 | 0.879517 | -1 | 3.09E-10 | 1 |
| 20-63902200-G-A | A | 7.81E-05 | 0.492236 | | | 0.4922 | -? | 0.000507 | 0.985258 | 0.000856 | 1.95E-11 | TRUE | 1.56E-07 | ++ | 0.695447 | 1 | 0.017145 | 1 | 3.19E-11 | 1 |
| 20-63902207-A-G | G | | | | | | | 0.000495 | 0.847461 | 0.000558 | 1.01E-08 | TRUE | 2.1E-05 | -- | 0.330573 | -1 | 0.970729 | 1 | 5.23E-10 | 1 |
| 20-64125830-TACTGT | T | | | | | | | | | 0.876662 | 4.11E-16 | FALSE | 4.11E-16 | ?- | 0.640273 | 1 | 0.246916 | -1 | 6.85E-18 | -1 |
| 20-64173439-GTAGTT | G | | | | | | | | | 0.027457 | 2.37E-17 | FALSE | 2.37E-17 | ?+ | 0.060051 | 1 | 0.301165 | 1 | 1.67E-15 | 1 |
| 20-13349551-G-A | A | 7.8E-05 | 0.316939 | | | 0.3169 | +? | 0.000257 | 6.58E-07 | 0.000114 | 0.00329 | TRUE | 7.67E-09 | ++ | | | | | 0.007696 | 1 |
| 21-31398874-A-G | G | 0.031551 | 4.4E-08 | | | 0.000000044 | +? | | | 0.020417 | 0.268082 | TRUE | 0.2681 | ?- | 0.586578 | -1 | 0.506385 | -1 | 0.995671 | 1 |
| 21-31402009-T-G | G | 0.032647 | 3.69E-08 | | | 3.687E-08 | +? | | | 0.021546 | 0.257265 | TRUE | 0.2573 | ?- | 0.363288 | -1 | 0.378186 | -1 | 0.959651 | 1 |
| 21-31403761-T-C | C | 0.032679 | 2.79E-08 | | | 2.793E-08 | +? | | | 0.021502 | 0.281877 | TRUE | 0.2819 | ?- | 0.368375 | -1 | 0.376869 | -1 | 0.979957 | 1 |
| 21-31403765-G-C | C | 0.032621 | 2.16E-08 | | | 2.159E-08 | -? | | | 0.021499 | 0.279346 | TRUE | 0.2793 | ?+ | 0.367171 | -1 | 0.377824 | -1 | 0.978665 | 1 |
| 21-5328547-CACAAAC | C | | | | | | | | | 0.752617 | 8.26E-09 | FALSE | 8.26E-09 | ?+ | 0.000456 | -1 | 0.502468 | -1 | 3.93E-05 | -1 |
| 21-5329312-AGTGG-A | A | | | | | | | | | 0.02779 | 4.67E-11 | FALSE | 4.67E-11 | ?- | 0.019616 | -1 | 0.205056 | -1 | 2.48E-09 | -1 |
| 21-5329317-AG-A | A | | | | | | | | | 0.025821 | 1.21E-09 | FALSE | 1.21E-09 | ?- | 0.004772 | -1 | 0.245869 | -1 | 7.17E-08 | -1 |
| 21-8449995-TTGCC-T | T | | | | | | | | | 0.091912 | 6.65E-11 | FALSE | 6.65E-11 | ?+ | 0.002572 | 1 | 0.959118 | -1 | 1.52E-10 | 1 |
| 21-8449995-TTGCCTG | T | | | | | | | | | 0.070512 | 2.57E-08 | FALSE | 2.57E-08 | ?+ | 0.096396 | 1 | 0.142743 | 1 | 3.65E-07 | 1 |
| 21-8449995-TTGCCTG | T | | | | | | | | | 0.613034 | 7.52E-38 | FALSE | 7.52E-38 | ?- | 0.000456 | -1 | 0.030805 | -1 | 7.88E-34 | -1 |
| 21-9124671-AACCCAA | A | | | | | | | | | 0.000727 | 4.72E-08 | TRUE | 4.72E-08 | ?+ | 0.015022 | 1 | 0.88687 | 1 | 9.35E-07 | 1 |
| 21-9817035-GAA-G | G | | | | | | | | | 0.004794 | 1.95E-10 | TRUE | 1.95E-10 | ?+ | 0.420503 | 1 | 0.033094 | 1 | 4.2E-09 | 1 |
| 21-9833806-T-A | A | | | | | | | | | 0.88764 | 2.73E-16 | FALSE | 2.73E-16 | ?- | 0.010823 | -1 | 0.866255 | -1 | 2.9E-17 | -1 |
| 21-10415599-T-C | C | | | | | | | | | 0.003538 | 6.41E-12 | TRUE | 6.41E-12 | ?+ | 0.409827 | -1 | 0.377166 | -1 | 3.08E-12 | -1 |
| 21-10415600-G-A | A | | | | | | | | | 0.003851 | 6.14E-12 | TRUE | 6.14E-12 | ?- | 0.695216 | -1 | 0.309485 | -1 | 3.04E-12 | -1 |
| 21-10717092-C-A | A | | | | | | | | | 0.995601 | 2.07E-09 | TRUE | 2.07E-09 | ?- | 0.012362 | -1 | 0.018804 | -1 | 4.11E-06 | -1 |
| 21-10756490-AG-A | A | | | | | | | | | 0.003095 | 3.52E-14 | TRUE | 3.52E-14 | ?+ | 0.02267 | 1 | 0.005401 | 1 | 1.39E-11 | 1 |
| 21-13083839-T-C | C | | | | | | | | | 0.030811 | 2.4E-09 | FALSE | 2.4E-09 | ?- | 0.000148 | 1 | 0.020234 | 1 | 5.46E-06 | 1 |
| 21-23757294-CCT-C | C | | | | | | | | | 0.000465 | 6.73E-10 | TRUE | 6.73E-10 | ?- | 0.017311 | 1 | 0.002306 | 1 | 4.35E-07 | 1 |
| 21-28685091-AAAC-A | A | | | | | | | | | 0.000731 | 1.6E-09 | TRUE | 1.6E-09 | ?+ | 0.012164 | 1 | 0.171946 | 1 | 8.01E-08 | 1 |
| 21-28995088-GTGA-G | G | 0.000117 | 0.498187 | | | 0.4982 | -? | | | 0.000581 | 4.42E-09 | TRUE | 4.42E-09 | ?+ | 0.694949 | -1 | 0.001042 | 1 | 5.02E-07 | 1 |
| 21-31690330-C-CCAA | CCAA | | | | | | | | | 0.000715 | 3.33E-09 | TRUE | 3.34E-09 | ?+ | 0.00606 | 1 | 0.014078 | 1 | 6.77E-07 | 1 |
| 21-39324090-AG-A | A | 0.001174 | 0.896031 | | | 0.896 | +? | | | 0.003607 | 1.65E-15 | TRUE | 1.66E-15 | ?+ | 0.008325 | 1 | 0.789223 | 1 | 1.09E-15 | 1 |
| 21-39383050-A-AAG | AAG | | | | | | | | | 0.000585 | 2.17E-10 | TRUE | 2.17E-10 | ?- | 0.088844 | 1 | 0.010678 | 1 | 2.57E-08 | 1 |
| 21-39383052-ACC-A | A | | | | | | | | | 0.000485 | 2.17E-11 | TRUE | 2.17E-11 | ?+ | 0.253135 | 1 | 0.001439 | 1 | 1.4E-09 | 1 |
| 21-42718089-C-CG | CG | 0.000647 | 0.957797 | | | 0.9578 | -? | | | 0.001939 | 1.56E-16 | TRUE | 1.56E-16 | ?+ | 0.002526 | 1 | 0.016389 | 1 | 2.8E-13 | 1 |
| 21-42718092-GA-G | G | 0.000628 | 0.599125 | | | 0.5991 | +? | | | 0.002628 | 2.27E-16 | TRUE | 2.27E-16 | ?+ | 0.002121 | 1 | 0.148142 | 1 | 2.93E-13 | 1 |
| 21-43788382-G-C | C | 0.000255 | 0.19004 | | | 0.19 | +? | 0.000204 | 0.963143 | 0.000772 | 1.16E-08 | TRUE | 6.39E-07 | -+ | 0.027384 | 1 | 0.002063 | 1 | 1.17E-06 | 1 |
| 21-44409661-G-A | A | | | | | | | | | 0.032231 | 3.29E-10 | FALSE | 3.29E-10 | ?+ | 0.006985 | 1 | 0.559838 | 1 | 1.08E-08 | 1 |
| 21-45155722-TCCACC | T | | | | | | | | | 0.003075 | 1.21E-11 | TRUE | 1.21E-11 | ?+ | 0.557702 | 1 | 0.164109 | 1 | 1.79E-10 | 1 |
| 21-45500131-GGGTGG | G | | | | | | | | | 0.000852 | 1.03E-08 | TRUE | 1.03E-08 | ?+ | 0.248194 | 1 | 0.004104 | 1 | 1.31E-06 | 1 |
| 21-46085044-CT-C | C | | | | | | | | | 0.001375 | 2.72E-08 | TRUE | 2.72E-08 | ?- | 0.120652 | 1 | 0.011643 | 1 | 9.69E-06 | 1 |
| 21-46390054-A-G | G | | | | | | | 9.4E-06 | 0.304421 | 0.000711 | 5.26E-11 | TRUE | 3.22E-11 | -- | 0.016244 | 1 | 0.014091 | 1 | 6.29E-09 | 1 |
| 21-9878744-CTTTTTTT | CTTTTTTCTTTTTTT | | | | | | | 3.45E-05 | 0.791786 | | | FALSE | 4.92E-10 | -? | | | | | | |
| 22-30295559-C-G | G | | | | | | | 1.25E-05 | 1E-08 | | | FALSE | 0.1438 | -? | | | | | | |
| 22-47711777-T-C | C | 0.032983 | 0.447531 | 0.03 | 0.007442 | 0.4052 | -- | 0.026812 | 1.5E-08 | 0.031607 | 0.217798 | TRUE | 1.45E-07 | -- | 0.063586 | 1 | 0.509365 | 1 | 0.607686 | 1 |
| 22-11368837-T-G | G | | | | | | | 0.000241 | 0.43741 | 0.00732 | 7.03E-10 | TRUE | 7.49E-09 | +- | 0.763052 | -1 | 0.002916 | 1 | 2.03E-12 | 1 |
| 22-12556740-CACACA | C | | | | | | | | | 0.001515 | 1.46E-08 | TRUE | 1.46E-08 | ?- | 0.03307 | 1 | 0.035167 | 1 | 2.32E-06 | 1 |
| 22-12595798-T-TGTA | TGTA | | | | | | | | | 0.143218 | 2.43E-10 | FALSE | 2.43E-10 | ?- | 0.212659 | 1 | 0.086103 | 1 | 2.77E-09 | 1 |
| 22-15213063-CATTT-C | C | | | | | | | | | 0.856528 | 2.55E-26 | FALSE | 2.55E-26 | ?+ | 4.59E-06 | -1 | 0.091264 | -1 | 2.34E-20 | -1 |
| 22-16279325-G-C | C | | | | | | | | | 0.0002 | 4.66E-08 | TRUE | 4.66E-08 | ?+ | | | | | 6.48E-08 | 1 |
| 22-17618008-C-A | A | | | | | | | 0.001353 | 0.160312 | 0.001554 | 1.61E-13 | TRUE | 2.56E-05 | -+ | 0.092276 | 1 | 0.005767 | 1 | 3.38E-11 | 1 |
| 22-18160517-A-T | T | | | | | | | 0.000839 | 0.974412 | 0.001086 | 3.03E-10 | TRUE | 3.04E-06 | -- | 0.297798 | 1 | 0.244121 | 1 | 2.5E-10 | 1 |
| 22-19300549-CCT-C | C | 0.000586 | 0.364591 | | | 0.3646 | -? | | | 0.001996 | 1.55E-14 | TRUE | 1.56E-14 | ?- | 0.09386 | 1 | 0.001955 | 1 | 5.84E-12 | 1 |
| 22-19300566-G-A | A | | | | | | | 4.7E-05 | 0.392246 | 0.00032 | 5.45E-12 | TRUE | 1.17E-11 | ++ | 0.021777 | 1 | 0.402622 | 1 | 3.07E-11 | 1 |
| 22-19311896-C-CAAA | CAAAA | | | | | | | | | 0.022395 | 8.99E-09 | TRUE | 8.99E-09 | ?+ | 0.436453 | 1 | 0.65559 | -1 | 1.03E-10 | 1 |
| 22-20050079-TGG-T | T | | | | | | | | | 0.00052 | 4.26E-11 | TRUE | 4.26E-11 | ?+ | 0.016528 | 1 | 0.919452 | -1 | 1.28E-10 | 1 |

| | | | | | | | | | | | | | | | | | | | | |
|---|---|---|---|---|---|---|---|---|---|---|---|---|---|---|---|---|---|---|---|---|
| 22-20359454-C-G | G | | | | | | | | | 0.961465 | 1.45E-21 | FALSE | 1.45E-21 | ?+ | 0.031373 | -1 | 0.361134 | -1 | 3.99E-21 | -1 |
| 22-20503118-ACC-A | A | | | | | | | | | 0.000918 | 1.12E-09 | TRUE | 1.12E-09 | ?+ | 0.439586 | 1 | 0.1199 | 1 | 1.61E-08 | 1 |
| 22-21924568-G-GCGA | GCGAAC | | | | | | | | | 0.000466 | 7.45E-15 | TRUE | 7.45E-15 | ?- | 0.135165 | 1 | 0.109976 | 1 | 1.16E-13 | 1 |
| 22-22251998-T-C | C | | | | | | | | | 0.000451 | 1.02E-08 | TRUE | 1.02E-08 | ?- | 0.008869 | 1 | 0.906733 | 1 | 1.2E-07 | 1 |
| 22-22518612-AC-A | A | | | | | | | | | 0.001163 | 8.5E-16 | TRUE | 8.5E-16 | ?+ | 0.000468 | 1 | 0.056185 | 1 | 7.35E-13 | 1 |
| 22-22732613-A-T | T | 0.002436 | 0.801962 | | | 0.802 | +? | 0.010163 | 0.718562 | 0.001738 | 4.71E-13 | TRUE | 0.003911 | -- | 0.939098 | -1 | 0.636237 | 1 | 2.56E-13 | 1 |
| 22-23515705-A-G | G | 0.093673 | 0.851176 | | | 0.8512 | +? | | | 0.098783 | 3.54E-10 | TRUE | 3.54E-10 | ?- | 0.001634 | 1 | 0.460224 | 1 | 0.683973 | -1 |
| 22-23646338-A-ATAA( | ATAAGAAAGATTTT | | | | | | | | | 0.000522 | 1.7E-08 | TRUE | 1.7E-08 | ?- | 0.001411 | 1 | 0.795454 | -1 | 2E-07 | 1 |
| 22-23719208-T-C | C | | | | | | | 0.000958 | 0.796841 | 0.001296 | 5.72E-14 | TRUE | 9.07E-09 | -- | 0.117451 | 1 | 0.004206 | 1 | 3.16E-12 | 1 |
| 22-23797852-CAG-C | C | | | | | | | | | 0.000562 | 2.84E-13 | TRUE | 2.84E-13 | ?- | 0.000562 | 1 | 0.319501 | 1 | 2.81E-10 | 1 |
| 22-24490949-CAA-C | C | | | | | | | | | 0.000729 | 6.89E-12 | TRUE | 6.89E-12 | ?- | 0.00023 | 1 | 0.025366 | 1 | 3.32E-08 | 1 |
| 22-24777872-A-ATT | ATT | | | | | | | | | 0.001715 | 1.08E-13 | TRUE | 1.08E-13 | ?- | 0.05822 | 1 | 0.00025 | 1 | 1.92E-10 | 1 |
| 22-25252791-C-CCA | CCA | 0.000176 | 0.676586 | | | 0.6766 | -? | | | 0.004184 | 1.49E-31 | TRUE | 1.49E-31 | ?+ | 0.000702 | 1 | 0.008296 | 1 | 1.54E-27 | 1 |
| 22-25252792-AGAGG | A | 0.000156 | 0.792489 | | | 0.7925 | +? | | | 0.003461 | 3.09E-28 | TRUE | 3.09E-28 | ?+ | 0.000511 | 1 | 0.005963 | 1 | 6.17E-24 | 1 |
| 22-25252802-A-AAGC | AAGCG | | | | | | | | | 0.003307 | 7.61E-23 | TRUE | 7.61E-23 | ?- | 0.002258 | 1 | 0.005862 | 1 | 4.92E-20 | 1 |
| 22-25324817-G-T | T | | | | | | | | | 0.000818 | 1.65E-09 | TRUE | 1.66E-09 | ?+ | 0.002741 | 1 | 0.036368 | 1 | 3.01E-06 | 1 |
| 22-25345441-AGGGG | A | | | | | | | | | 0.007681 | 5.07E-14 | TRUE | 5.07E-14 | ?+ | 0.03201 | 1 | 0.016114 | 1 | 8.14E-12 | 1 |
| 22-28089534-G-GAA | GAA | 0.000529 | 0.900337 | | | 0.9003 | -? | | | 0.001008 | 2.23E-08 | TRUE | 2.23E-08 | ?- | 0.468892 | -1 | 0.020705 | 1 | 1.02E-07 | 1 |
| 22-28536011-C-CCAT( | CCATCCTGGCTAA | | | | | | | | | 0.003717 | 3.58E-10 | TRUE | 3.58E-10 | ?+ | 0.002456 | 1 | 0.121913 | 1 | 5.74E-08 | 1 |
| 22-28536396-G-A | A | 0.000195 | 0.884184 | | | 0.8842 | +? | 0.001146 | 0.032122 | 0.001235 | 1.58E-08 | TRUE | 0.01092 | -+ | 6.36E-05 | 1 | 0.000806 | 1 | 0.000518 | 1 |
| 22-28536542-T-G | G | 0.000476 | 0.895991 | | | 0.896 | +? | 0.001294 | 0.895935 | 0.001572 | 9.09E-13 | TRUE | 6.14E-07 | +- | 0.322305 | 1 | 0.152309 | 1 | 5.62E-12 | 1 |
| 22-28645472-C-CA | CA | 0.000664 | 0.133321 | | | 0.1333 | +? | | | 0.002373 | 1.71E-11 | TRUE | 1.71E-11 | ?+ | 0.793338 | -1 | 0.004379 | 1 | 8.64E-12 | 1 |
| 22-28770546-A-AT | AT | 0.000508 | 0.054765 | | | 0.05477 | +? | | | 0.001771 | 5.78E-11 | TRUE | 5.78E-11 | ?- | 0.000712 | 1 | 0.109296 | 1 | 4.86E-09 | 1 |
| 22-29148578-CG-C | C | | | | | | | | | 0.000623 | 4.85E-12 | TRUE | 4.85E-12 | ?- | 0.08292 | 1 | 0.125935 | 1 | 4.07E-10 | 1 |
| 22-30913268-CG-C | C | | | | | | | | | 0.003759 | 1.26E-18 | TRUE | 1.26E-18 | ?- | 0.952617 | 1 | 0.000171 | 1 | 6.36E-17 | 1 |
| 22-31293758-G-GT | GT | | | | | | | | | 0.000569 | 3.08E-08 | TRUE | 3.08E-08 | ?- | 0.010491 | 1 | 0.000263 | 1 | 0.000373 | 1 |
| 22-35050426-C-A | A | | | | | | | | | 0.000249 | 2.49E-09 | TRUE | 2.49E-09 | ?+ | 0.003433 | 1 | 0.039411 | 1 | 2.1E-07 | 1 |
| 22-35699725-GCCCC-( | G | | | | | | | | | 0.004465 | 2.3E-10 | TRUE | 2.3E-10 | ?+ | 0.060407 | 1 | 0.611329 | 1 | 6.54E-09 | 1 |
| 22-36068076-G-A | A | 7.8E-05 | 0.150952 | | | 0.151 | -? | 0.000426 | 0.160306 | 0.000766 | 3.04E-10 | TRUE | 5.49E-09 | ++ | 0.006421 | 1 | 0.46319 | 1 | 6.18E-10 | 1 |
| 22-36068090-G-A | A | 9.76E-05 | 0.169895 | | | 0.1699 | -? | 0.000229 | 0.737581 | 0.001092 | 1.47E-16 | TRUE | 2.83E-14 | ++ | 5.3E-05 | 1 | 0.125815 | 1 | 1.29E-12 | 1 |
| 22-36389311-CCT-C | C | | | | | | | | | 0.000572 | 1.5E-08 | TRUE | 1.5E-08 | ?- | 0.001805 | 1 | 0.020763 | 1 | 6.78E-06 | 1 |
| 22-37939702-T-A | A | | | | | | | | | 0.000458 | 9.1E-10 | TRUE | 9.1E-10 | ?+ | 0.015626 | 1 | 0.201315 | 1 | 1.25E-08 | 1 |
| 22-38242246-TTG-T | T | | | | | | | | | 0.000281 | 1.39E-08 | TRUE | 1.39E-08 | ?+ | 0.022496 | 1 | | | 1.19E-07 | 1 |
| 22-38342004-C-CCA | CCA | | | | | | | | | 0.0012 | 5.16E-12 | TRUE | 5.16E-12 | ?+ | 0.001527 | 1 | 0.717124 | -1 | 2.19E-09 | 1 |
| 22-38405984-A-AT | AT | 0.000176 | 0.154445 | | | 0.1544 | +? | | | 0.001163 | 1.33E-09 | TRUE | 1.33E-09 | ?- | 0.039321 | 1 | 0.005224 | 1 | 2.72E-07 | 1 |
| 22-38654128-G-C | C | | | | | | | 0.002688 | 0.946907 | 0.002537 | 4.83E-28 | TRUE | 3.78E-14 | ++ | 0.000319 | 1 | 0.061284 | 1 | 3.1E-26 | 1 |
| 22-39166463-C-CCT | CCT | | | | | | | | | 0.000751 | 1.69E-09 | TRUE | 1.69E-09 | ?+ | 0.535396 | 1 | 0.086433 | 1 | 1.1E-08 | 1 |
| 22-39166662-A-AG | AG | | | | | | | | | 0.000577 | 4.37E-08 | TRUE | 4.38E-08 | ?- | 0.006313 | 1 | 0.03177 | 1 | 2.94E-06 | 1 |
| 22-40796022-CAG-C | C | | | | | | | 8.77E-05 | 0.493337 | 0.001425 | 1.83E-13 | TRUE | 3.59E-12 | +- | 0.322347 | 1 | 0.005714 | 1 | 3.12E-12 | 1 |
| 22-40935856-GT-G | G | | | | | | | | | 0.000972 | 1.89E-09 | TRUE | 1.89E-09 | ?+ | 0.000145 | 1 | 0.024166 | 1 | 5.88E-06 | 1 |
| 22-41212458-T-C | C | 0.385507 | 0.566884 | 0.384 | 0.25166 | 0.6315 | +- | 0.36804 | 0.87755 | 0.44051 | 7.96E-09 | FALSE | 0.000799 | -- | 0.600825 | 1 | 0.016283 | 1 | 6.66E-08 | 1 |
| 22-41293017-CG-C | C | 0.000195 | 0.279656 | | | 0.2797 | +? | | | 0.001343 | 4.5E-09 | TRUE | 4.5E-09 | ?- | 0.011917 | 1 | 0.418244 | 1 | 1.06E-07 | 1 |
| 22-41379102-A-ATGC( | ATGCC | 0.000156 | 0.542749 | | | 0.5427 | +? | | | 0.002719 | 1.98E-10 | TRUE | 1.98E-10 | ?- | 0.636677 | 1 | 0.407653 | 1 | 2.85E-10 | 1 |
| 22-41609575-CTTG-C | C | | | | | | | | | 0.000473 | 3.18E-08 | TRUE | 3.18E-08 | ?- | 0.098762 | 1 | 0.011503 | 1 | 6.3E-05 | 1 |
| 22-41712999-C-T | T | 0.000137 | 0.866873 | | | 0.8669 | -? | 0.000179 | 0.479323 | 0.001121 | 3.05E-08 | TRUE | 7.43E-08 | ++ | 0.002523 | 1 | 0.47319 | 1 | 1.97E-06 | 1 |
| 22-41786143-C-T | T | | | | | | | | | 0.00031 | 8.5E-10 | TRUE | 8.5E-10 | ?+ | 0.61668 | -1 | 0.362057 | -1 | 1.19E-11 | 1 |
| 22-41786146-G-A | A | | | | | | | 1.57E-05 | 0.263165 | 0.000258 | 6.75E-09 | TRUE | 7.18E-08 | -+ | 0.610021 | -1 | | | 1.3E-09 | 1 |
| 22-41786153-G-T | T | | | | | | | 4.07E-05 | 0.937115 | 0.000601 | 1.62E-10 | TRUE | 6.88E-10 | -+ | 0.432336 | -1 | 0.796955 | -1 | 1.38E-12 | 1 |
| 22-41795898-T-TG | TG | 9.77E-05 | 0.29623 | | | 0.2962 | +? | 0.006352 | 0.54594 | 0.001433 | 2.17E-11 | TRUE | 0.03323 | +- | 0.00916 | 1 | 0.784357 | 1 | 6.02E-10 | 1 |
| 22-41795903-C-T | T | 0.000118 | 0.224602 | | | 0.2246 | -? | 0.006352 | 0.54594 | 0.00134 | 6.41E-15 | TRUE | 0.009858 | -+ | 0.009543 | 1 | 0.46369 | 1 | 3.49E-12 | 1 |
| 22-42547037-ATGCC-/ | A | 0.007858 | 0.030594 | | | 0.03059 | -? | | | 0.03583 | 6.13E-09 | FALSE | 6.13E-09 | ?+ | 0.714722 | 1 | 0.790668 | -1 | 1.73E-10 | 1 |
| 22-42547043-AGCTAA | A | 0.007603 | 0.024147 | | | 0.02415 | -? | | | 0.021322 | 1.3E-08 | FALSE | 1.3E-08 | ?+ | 0.995256 | -1 | 0.64008 | -1 | 1.11E-10 | 1 |
| 22-43016634-C-T | T | | | 0.036 | 0.028985 | 0.02898 | ?- | | | 0.098691 | 3.58E-08 | TRUE | 3.58E-08 | ?+ | 0.120179 | 1 | 0.063887 | 1 | 0.716805 | 1 |
| 22-43499248-GTGGTG | G | | | | | | | | | 0.035731 | 5.58E-11 | FALSE | 5.58E-11 | ?+ | 0.398879 | 1 | 0.334239 | 1 | 5.83E-12 | 1 |
| 22-44675142-C-A | A | 0.000613 | 0.588327 | | | 0.5883 | -? | 0.00286 | 0.695644 | 0.001356 | 1.2E-11 | TRUE | 0.000881 | -+ | 0.000355 | 1 | 0.00167 | 1 | 5.16E-08 | 1 |
| 22-44749165-A-AC | AC | 0.000353 | 0.040265 | | | 0.04027 | +? | | | 0.001462 | 5.55E-09 | TRUE | 5.55E-09 | ?- | 0.001963 | 1 | 0.988408 | 1 | 2.71E-07 | 1 |
| 22-44886986-C-CCA | CCA | | | | | | | | | 0.000744 | 2.32E-09 | TRUE | 2.32E-09 | ?+ | 0.076731 | 1 | 0.013429 | 1 | 1.44E-06 | 1 |
| 22-45229338-G-C | C | | | | | | | | | 0.801234 | 3.09E-17 | FALSE | 3.09E-17 | ?+ | 0.104056 | 1 | 0.795111 | 1 | 2.74E-17 | 1 |
| 22-45229338-G-T | T | | | | | | | 0.002741 | 0.136939 | 0.171825 | 1.15E-13 | FALSE | 3.77E-14 | -- | 0.119802 | -1 | 0.769104 | -1 | 5.18E-14 | -1 |

| | | | | | | | | | | | | | | | | | | | | |
|---|---|---|---|---|---|---|---|---|---|---|---|---|---|---|---|---|---|---|---|---|
| 22-45519419-G-C | C | | | | | | | 0.001629 | 0.849274 | 0.000952 | 2.62E-08 | TRUE | 0.002129 | -+ | 0.011192 | 1 | 0.755959 | -1 | 8.92E-08 | 1 |
| 22-45519431-G-A | A | 9.77E-05 | 0.405675 | | | 0.4057 | +? | 0.001629 | 0.849449 | 0.000781 | 2E-10 | TRUE | 0.000547 | -+ | 0.000282 | 1 | 0.441285 | 1 | 3.08E-08 | 1 |
| 22-46263081-T-C | C | 0.217086 | 0.500054 | | | 0.5001 | +? | | | 0.250238 | 3.97E-09 | FALSE | 3.97E-09 | ?- | 0.436184 | 1 | 0.067086 | 1 | 3.17E-05 | 1 |
| 22-46713286-C-CTT | CTT | | | | | | | | | 0.000435 | 1.15E-11 | TRUE | 1.15E-11 | ?+ | 0.026747 | 1 | 0.306803 | 1 | 1.09E-10 | 1 |
| 22-47212498-G-A | A | | | | | | | | | 0.497543 | 1.48E-10 | FALSE | 1.48E-10 | ?+ | 0.741891 | 1 | 0.037335 | 1 | 3.28E-08 | 1 |
| 22-48399528-T-C | C | 0.075472 | 0.559836 | 0.03 | 0.682776 | 0.5675 | -+ | | | 0.083165 | 2.07E-13 | TRUE | 2.07E-13 | ?- | 0.018287 | 1 | 0.617837 | 1 | 0.283941 | 1 |
| 22-48910553-CCCCTT | C | | | | | | | | | 0.000694 | 3.77E-09 | TRUE | 3.77E-09 | ?- | 0.033477 | 1 | 0.000188 | 1 | 1.53E-05 | 1 |
| 22-49399428-CCA-C | C | | | | | | | | | 0.000165 | 3.98E-10 | TRUE | 3.98E-10 | ?- | | | | | 5.45E-06 | 1 |
| 22-49879291-AT-A | A | | | | | | | | | 0.000506 | 4.9E-10 | TRUE | 4.91E-10 | ?+ | | | 0.000456 | 1 | 5.03E-08 | 1 |
| 22-50335161-G-C | C | 0.472302 | 0.196479 | 0.393 | 0.824983 | 0.204 | -+ | | | 0.481417 | 2.9E-16 | FALSE | 2.9E-16 | ?+ | 0.028256 | 1 | 0.054557 | 1 | 7.38E-11 | 1 |
| 22-50354109-ACC-A | A | | | | | | | | | 0.001892 | 2.87E-11 | TRUE | 2.87E-11 | ?+ | 0.256055 | 1 | 0.647984 | 1 | 1.38E-11 | 1 |
| 22-50354112-G-GGC | GGC | 0.00051 | 0.023179 | | | 0.02318 | +? | | | 0.002179 | 6.48E-11 | TRUE | 6.48E-11 | ?- | 0.295717 | 1 | 0.746897 | 1 | 4.76E-11 | 1 |
| 22-50372984-GT-G | G | | | | | | | | | 0.00057 | 2.7E-10 | TRUE | 2.7E-10 | ?+ | 0.001722 | 1 | 0.415399 | 1 | 2.21E-08 | 1 |
| 22-50412310-A-G | G | 0.394935 | 0.29736 | 0.363 | 0.018805 | 0.389 | +- | 0.334215 | 0.284742 | 0.404936 | 1.24E-08 | FALSE | 0.01789 | +- | 0.176359 | 1 | 0.6978 | 1 | 1.06E-06 | 1 |
| 22-50543535-T-TGCA | TGCA | 0.006301 | 0.019725 | | | 0.01972 | +? | | | 0.003647 | 3.16E-08 | TRUE | 3.16E-08 | ?- | 0.617229 | 1 | 0.546198 | 1 | 4.47E-08 | 1 |
| 22-50543540-CGGT-C | C | | | | | | | | | 0.004846 | 1.28E-14 | TRUE | 1.28E-14 | ?- | 0.502606 | 1 | 0.246 | 1 | 4E-15 | 1 |
| 22-25895426-T-A | A | | | | | | | 2.51E-05 | 0.99096 | | | FALSE | 1E-305 | +? | | | | | | |
| 22-25789433-C-T | T | 0.000216 | 0.346681 | | | 0.3467 | -? | 3.45E-05 | 0.064919 | 0.0005 | 9.91E-08 | TRUE | 1.81E-08 | ++ | 0.038672 | 1 | 0.61271 | 1 | 2.99E-07 | 1 |
| 22-35049660-G-T | T | | | | | | | 9.4E-06 | 0.305431 | | | FALSE | 1.2E-305 | +? | | | | | | |
| 22-15619654-G-T | T | | | | | | | 2.51E-05 | 0.008266 | | | FALSE | | | | | | | | |
