## Supplementary Table 3 for "Identification of 16 novel Alzheimer’s disease susceptibility loci using multi-ancestry meta-analyses of clinical Alzheimer’s disease and AD-by-proxy cases from four whole genome sequencing datasets"

**Supplementary table 3:** Independent loci that passed QC in our Clinical AD meta-analysis.
MAF - minor allele frequency; P - p-value; META_P - p-value for the meta analysis.

| Locus | Chromosome | Variant ID | Rsid | Gene | MAF | Effect allele | NIA_P | NIMH_P | Direction (NIA, NIMH) | NIA_NIMH_META_P | UKB_AOU_META_P | New or Old |
|---|---|---|---|---|---|---|---|---|---|---|---|---|
| 1 | 1 | 1-165164548-G-A | rs573181360 | RNU6-755P, LMX1A | 0.0005 | G | 2.09E-08 | | +? | 2.093E-08 | 0.9857 | New |
| 2 | 1 | 1-20328237-TG-T | rs141744862 | VWA5B1 | 0.0031 | T | 4.6E-08 | | +? | 4.604E-08 | 0.1779 | New |
| 3 | 1 | 1-207510847-G-T | | CR1 | 0.1274 | T | 1.6E-09 | 0.059625 | ++ | 8.17E-10 | 0.00398 | Old |
| 3 | 1 | 1-207512441-C-T | | CR1 | 0.1453 | T | 3E-10 | 0.163644 | ++ | 1.819E-10 | 0.02247 | Old |
| 3 | 1 | 1-207524699-C-T | rs6661489 | CR1 | 0.1425 | T | 1.47E-10 | | +? | 1.468E-10 | 0.02257 | Old |
| 3 | 1 | 1-207573951-G-A | | AL137789.1 | 0.1344 | A | 3.63E-10 | 0.046681 | ++ | 1.726E-10 | 0.009715 | Old |
| 3 | 1 | 1-207577223-C-T | | AL137789.1 | 0.1248 | T | 2.23E-09 | 0.036457 | ++ | 1.052E-09 | 0.03141 | Old |
| 3 | 1 | 1-207611623-G-A | | CR1 | 0.2226 | A | 4.72E-08 | 0.139348 | ++ | 2.885E-08 | 0.008675 | Old |
| 3 | 1 | 1-207612944-G-A | | CR1 | 0.2227 | A | 7.86E-08 | 0.12557 | ++ | 4.733E-08 | 0.009994 | Old |
| 3 | 1 | 1-207613197-G-A | | CR1 | 0.1635 | A | 7.68E-09 | 0.07148 | ++ | 4.063E-09 | 0.01409 | Old |
| 3 | 1 | 1-207613483-G-A | | CR1 | 0.2227 | A | 6.61E-08 | 0.139348 | ++ | 4.052E-08 | 0.01044 | Old |
| 3 | 1 | 1-207621975-G-A | | CR1 | 0.1649 | A | 2.19E-09 | 0.081119 | ++ | 1.164E-09 | 0.01154 | Old |
| 3 | 1 | 1-207623552-T-A | | CR1 | 0.1327 | A | 3.2E-10 | 0.081085 | ++ | 1.683E-10 | 0.01818 | Old |
| 3 | 1 | 1-207625349-C-T | | CR1 | 0.1647 | C | 2.63E-09 | | +? | 2.627E-09 | 0.01112 | Old |
| 3 | 1 | 1-207626529-C-A | | CR1 | 0.1648 | A | 2.34E-09 | 0.084246 | ++ | 1.253E-09 | 0.009343 | Old |
| 3 | 1 | 1-207627210-C-T | | CR1 | 0.1425 | T | 2.42E-09 | 0.072722 | ++ | 1.268E-09 | 0.01954 | Old |
| 3 | 1 | 1-207629207-C-A | | CR1 | 0.1236 | A | 3.62E-09 | 0.066785 | ++ | 1.907E-09 | 0.004591 | Old |
| 3 | 1 | 1-207630796-C-A | | CR1 | 0.1422 | A | 2.71E-09 | 0.079471 | ++ | 1.447E-09 | 0.01894 | Old |
| 3 | 1 | 1-207633385-G-A | | CR1 | 0.1419 | G | 2.86E-09 | 0.079471 | ++ | 1.528E-09 | 0.01796 | Old |
| 4 | 2 | 2-127090354-G-A | rs35114168 | BIN1 | 0.2455 | A | 1.47E-09 | 0.251863 | ++ | 9.432E-10 | 0.0001673 | Old |
| 4 | 2 | 2-127124439-T-A | rs6720234 | BIN1, AC110926.2 | 0.1174 | A | 1.57E-08 | 0.687451 | ++ | 1.444E-08 | 0.002833 | Old |
| 4 | 2 | 2-127130409-C-A | rs12989701 | BIN1, AC110926.2 | 0.1211 | A | 4.21E-08 | 0.811491 | ++ | 4.111E-08 | 0.0002707 | Old |
| 4 | 2 | 2-127131181-C-A | rs11680911 | BIN1, AC110926.2 | 0.2667 | C | 4.34E-10 | 0.2664 | ++ | 2.797E-10 | 0.000007868 | Old |
| 4 | 2 | 2-127131542-C-T | rs13009551 | BIN1, AC110926.2 | 0.107 | T | 3.07E-08 | 0.897678 | ++ | 3.124E-08 | 0.04924 | Old |
| 4 | 2 | 2-127133851-C-A | rs4663105 | BIN1, AC110926.2 | 0.4789 | C | 1.77E-10 | 0.077272 | ++ | 8.114E-11 | 6.348E-11 | Old |
| 4 | 2 | 2-127135234-C-T | rs6733839 | BIN1, AC110926.2 | 0.4168 | T | 1.19E-10 | 0.039481 | ++ | 4.673E-11 | 6.137E-12 | Old |
| 5 | 2 | 2-74159003-G-A | rs150214656 | MOB1A | 0.0003 | G | 2.1E-08 | | +? | 2.102E-08 | 0.5875 | New |
| 6 | 3 | 3-109106242-C-T | rs557495347 | MORC1-AS1 | 0.0008 | T | 4.71E-08 | | +? | 4.707E-08 | 0.9534 | New |
| 7 | 4 | 4-79608801-G-A | rs117010230 | LINC00989 | 0.0009 | A | 3.14E-08 | | +? | 3.143E-08 | 0.6344 | New |
| 8 | 5 | 5-115784235-C-T | rs56918975 | RNU2-49P, CDO1 | 0.0005 | C | 1.48E-08 | | +? | 1.477E-08 | 0.2683 | New |
| 9 | 5 | 5-128619201-G-A | rs147450666 | FBN2, SLC27A6 | 0.0104 | A | 7.67E-09 | 0.525766 | -- | 7.171E-09 | 0.8776 | New |
| 10 | 5 | 5-59693193-G-A | rs182525847 | PDE4D | 0.0005 | G | 2.07E-08 | | +? | 2.071E-08 | 0.07526 | New |
| 11 | 7 | 7-155194021-C-T | rs186724723 | RN7SKP280, AC099552.1 | 0.0158 | C | 1.62E-08 | | -? | 1.618E-08 | 0.6294 | Old |
| 12 | 12 | 12-48371822-T-A | rs56098445 | AC090115.1 | 0.094 | A | 4.25E-08 | 0.448226 | ++ | 3.572E-08 | 0.6859 | New |

| | | | | | | | | | | | | |
|---|---|---|---|---|---|---|---|---|---|---|---|---|
| 13 | 15 | 15-73168712-C-T | rs541189631 | NEO1 | 0.0008 | T | 8.84E-09 | | +? | 8.839E-09 | 0.6662 | New |
| 14 | 17 | 17-35209114-G-T | rs553129131 | SLC35G3, AC022916.1 | 0.0006 | G | 2.21E-08 | | +? | 2.208E-08 | 0.2605 | New |
| 15 | 18 | 18-49233661-G-A | rs142978359 | DYM | 0.0265 | G | 2.36E-08 | 0.362126 | -+ | 2.479E-08 | 0.1661 | New |
| 15 | 18 | 18-49235132-G-T | rs147440553 | DYM | 0.0261 | T | 1.29E-08 | | -? | 1.295E-08 | 0.04734 | New |
| 15 | 18 | 18-49261602-GCA-G | rs200388554 | DYM | 0.0258 | G | 5.99E-09 | | -? | 5.985E-09 | 0.05368 | New |
| 15 | 18 | 18-49326113-G-A | rs138229840 | DYM | 0.0262 | G | 1.64E-08 | 0.362126 | -+ | 1.717E-08 | 0.04997 | New |
| 15 | 18 | 18-49403096-G-A | rs146444978 | DYM | 0.0266 | G | 7.77E-09 | 0.362126 | -+ | 8.158E-09 | 0.06329 | New |
| 16 | 19 | 19-44746404-C-T | rs12459810 | RF00285, BCL3 | 0.2063 | T | 4.69E-08 | 0.608472 | ++ | 4.125E-08 | 8.646E-10 | Old |
| 16 | 19 | 19-44750911-C-A | rs8103315 | BCL3 | 0.0826 | A | 3.31E-08 | 0.934838 | ++ | 3.416E-08 | 2.719E-15 | Old |
| 16 | 19 | 19-44895007-C-T | rs157588 | TOMM40 | 0.4379 | C | 3.14E-17 | 0.00929 | ++ | 6.807E-18 | 1.529E-13 | Old |
| 17 | 19 | 19-44781370-G-A | rs80168591 | CBLC | 0.0092 | A | 2.84E-10 | 0.136875 | ++ | 2.343E-10 | 8.087E-15 | Old |
| 17 | 19 | 19-44820881-G-A | rs28399637 | BCAM | 0.2328 | A | 1.63E-19 | 0.020289 | ++ | 4.149E-20 | 1.305E-26 | Old |
| 17 | 19 | 19-44827852-G-C | rs905342119 | BCAM, NECTIN2 | 0.0067 | C | 9.3E-10 | 0.000266 | ++ | 5.485E-10 | 3.893E-20 | Old |
| 17 | 19 | 19-44843409-G-C | rs148601586 | BCAM, NECTIN2 | 0.0092 | G | 5.62E-13 | 0.007555 | ++ | 3.717E-13 | 8.133E-19 | Old |
| 17 | 19 | 19-44848259-G-C | rs41289512 | NECTIN2 | 0.0311 | G | 4.94E-30 | 0.367582 | ++ | 3.555E-30 | 6.644E-59 | Old |
| 17 | 19 | 19-44851039-G-A | rs11666329 | NECTIN2 | 0.4233 | G | 1.95E-09 | 0.066974 | -- | 9.115E-10 | 1.699E-08 | Old |
| 17 | 19 | 19-44852031-C-T | rs149661872 | NECTIN2 | 0.0084 | T | 1.69E-09 | 0.139607 | ++ | 1.42E-09 | 1.897E-15 | Old |
| 17 | 19 | 19-44855049-C-T | rs551048812 | NECTIN2 | 0.0089 | T | 1.15E-10 | | +? | 1.146E-10 | 1.752E-11 | Old |
| 17 | 19 | 19-44856329-C-T | rs56317818 | NECTIN2 | 0.2898 | T | 4E-11 | 0.192425 | ++ | 2.368E-11 | 2.994E-28 | Old |
| 17 | 19 | 19-44856688-G-A | rs183610051 | NECTIN2 | 0.0071 | A | 4.4E-11 | 0.164491 | ++ | 3.659E-11 | 3.512E-15 | Old |
| 17 | 19 | 19-44860563-G-T | rs138607350 | NECTIN2 | 0.0067 | G | 5.35E-10 | 0.000218 | ++ | 3.11E-10 | 1.579E-23 | Old |
| 17 | 19 | 19-44862190-G-A | rs146275714 | NECTIN2 | 0.0169 | A | 5.81E-21 | | +? | 5.811E-21 | 1.034E-48 | Old |
| 17 | 19 | 19-44863241-G-A | rs183427010 | NECTIN2 | 0.0056 | A | 7.93E-10 | 0.549418 | +- | 8.641E-10 | 2.946E-10 | Old |
| 17 | 19 | 19-44874887-C-T | rs34278513 | NECTIN2 | 0.1012 | T | 1.49E-12 | 0.28569 | ++ | 1.038E-12 | 0.009079 | Old |
| 17 | 19 | 19-44876259-G-A | rs412776 | NECTIN2 | 0.138 | A | 1.6E-11 | 0.443667 | ++ | 1.265E-11 | 0.01134 | Old |
| 17 | 19 | 19-44877704-C-A | rs3865427 | NECTIN2 | 0.0958 | A | 4.26E-12 | 0.329914 | ++ | 3.115E-12 | 0.008909 | Old |
| 17 | 19 | 19-44877713-G-T | rs11668861 | NECTIN2 | 0.4826 | G | 3.24E-14 | 0.005632 | ++ | 7.157E-15 | 7.49E-17 | Old |
| 17 | 19 | 19-44878777-G-A | | NECTIN2 | 0.3982 | A | 2.72E-34 | 0.004984 | ++ | 2.722E-35 | 0.0000107 | Old |
| 17 | 19 | 19-44881674-G-A | rs79701229 | NECTIN2 | 0.0092 | A | 1.27E-12 | 0.022019 | ++ | 8.818E-13 | 2.464E-19 | Old |
| 17 | 19 | 19-44882502-G-C | | AC011481.2 | 0.3445 | C | 2.02E-31 | 0.003429 | -- | 2.246E-32 | 0.1542 | Old |
| 17 | 19 | 19-44883210-GTAA-G | rs142042446 | AC011481.2 | 0.1397 | GTAA | 1.6E-136 | | +? | 1.61E-136 | 9.09E-202 | Old |
| 17 | 19 | 19-44883377-C-T | rs147636938 | AC011481.2 | 0.0325 | T | 4.72E-32 | 0.002769 | ++ | 1.348E-32 | 5.353E-49 | Old |
| 17 | 19 | 19-44883598-G-A | rs166907 | AC011481.2 | 0.0343 | G | 1.29E-14 | 0.375725 | +- | 1.445E-14 | 0.001778 | Old |
| 17 | 19 | 19-44884202-G-C | rs12972156 | AC011481.2 | 0.1396 | G | 3.1E-136 | 2.29E-09 | ++ | 4.86E-140 | 7.7E-202 | Old |
| 17 | 19 | 19-44884339-G-A | rs12972970 | AC011481.2 | 0.1396 | A | 2E-136 | 1.57E-09 | ++ | 2.69E-140 | 3.3E-202 | Old |
| 17 | 19 | 19-44884873-G-A | rs34342646 | AC011481.2 | 0.143 | A | 2.7E-131 | 2.92E-09 | ++ | 4.9E-135 | 7.26E-194 | Old |
| 17 | 19 | 19-44885243-G-A | rs283811 | AC011481.2 | 0.3214 | G | 7.2E-120 | | +? | 7.23E-120 | 3.12E-174 | Old |
| 17 | 19 | 19-44887076-A-G | rs283815 | AC011481.2 | 0.384 | A | | 3.05E-11 | ?+ | 3.05E-11 | 5.14E-184 | Old |
| 17 | 19 | 19-44888197-C-T | rs145654351 | NECTIN2 | 0.0046 | T | 1.58E-10 | | +? | 1.576E-10 | 0.1791 | Old |
| 17 | 19 | 19-44888997-C-T | rs6857 | AC011481.2 | 0.1728 | T | 1.2E-183 | 4.95E-13 | ++ | 2.33E-189 | 2.42E-277 | Old |
| 17 | 19 | 19-44890259-C-T | rs117310449 | AC011481.2 | 0.0105 | T | 4.88E-18 | 0.121432 | ++ | 3.589E-18 | 1.399E-27 | Old |
| 17 | 19 | 19-44891079-C-T | rs71352238 | TOMM40 | 0.1397 | C | 6.9E-139 | 2.84E-10 | ++ | 4.62E-143 | 3.36E-207 | Old |
| 17 | 19 | 19-44891712-G-T | rs184017 | TOMM40 | 0.3258 | G | 4.6E-120 | 1.57E-11 | ++ | 1.44E-124 | 6.68E-184 | Old |
| 17 | 19 | 19-44892073-G-A | rs2075649 | TOMM40 | 0.3168 | G | 7.56E-25 | 0.002583 | -- | 1.001E-25 | 0.1913 | Old |
| 17 | 19 | 19-44892362-G-A | rs2075650 | TOMM40 | 0.1648 | G | 9E-126 | 2.94E-10 | ++ | 7.79E-130 | 8.01E-214 | Old |
| 17 | 19 | 19-44892587-G-A | rs34095326 | TOMM40 | 0.0882 | A | 3.52E-83 | 2.46E-06 | ++ | 2.391E-85 | 7.73E-158 | Old |

| | | | | | | | | | | | | |
|---|---|---|---|---|---|---|---|---|---|---|---|---|
| 17 | 19 | 19-44892652-G-C | rs34404554 | TOMM40 | 0.1564 | G | 8.6E-131 | 1.41E-10 | ++ | 5.35E-135 | 2.72E-217 | Old |
| 17 | 19 | 19-44892887-C-T | rs11556505 | TOMM40 | 0.1601 | T | 2.5E-129 | 2.94E-10 | ++ | 1.93E-133 | 2.63E-215 | Old |
| 17 | 19 | 19-44892962-C-T | rs157582 | TOMM40 | 0.3276 | T | 9.3E-131 | 4.15E-11 | ++ | 2.46E-135 | 5.76E-189 | Old |
| 17 | 19 | 19-44893408-G-T | rs59007384 | TOMM40 | 0.357 | T | | 4.58E-11 | ?+ | 4.58E-11 | 1.14E-188 | Old |
| 17 | 19 | 19-44893416-G-T | rs157583 | TOMM40 | 0.0378 | T | 2.68E-24 | 0.876927 | ++ | 2.646E-24 | 0.03701 | Old |
| 17 | 19 | 19-44893716-G-A | rs77301115 | TOMM40 | 0.0381 | A | 2.61E-53 | 0.006838 | ++ | 5.579E-54 | 3.72E-50 | Old |
| 17 | 19 | 19-44894050-C-T | rs112849259 | TOMM40 | 0.0381 | T | 2.44E-54 | 0.006838 | ++ | 5.136E-55 | 3.279E-50 | Old |
| 17 | 19 | 19-44894695-C-T | rs116881820 | TOMM40 | 0.0381 | C | 8.12E-53 | 0.006838 | ++ | 1.744E-53 | 9.889E-51 | Old |
| 17 | 19 | 19-44894944-C-T | rs115908094 | TOMM40 | 0.0045 | T | 4.66E-11 | | +? | 4.658E-11 | 0.2771 | Old |
| 17 | 19 | 19-44895208-G-C | | TOMM40 | 0.0044 | G | 4.22E-11 | | +? | 4.225E-11 | 0.3622 | Old |
| 17 | 19 | 19-44895376-G-C | rs11668327 | TOMM40 | 0.1026 | C | 4.12E-21 | 0.002291 | -- | 8.595E-22 | 3.057E-28 | Old |
| 17 | 19 | 19-44895528-C-T | rs79398853 | TOMM40 | 0.0379 | T | 3.47E-54 | 0.006838 | ++ | 7.326E-55 | 4.564E-50 | Old |
| 17 | 19 | 19-44897468-C-T | rs114536010 | TOMM40 | 0.0379 | T | 6.54E-54 | 0.006838 | ++ | 1.386E-54 | 6.941E-50 | Old |
| 17 | 19 | 19-44897518-G-T | rs205909 | TOMM40 | 0.0328 | G | 1.11E-21 | 0.877704 | ++ | 1.093E-21 | 0.00685 | Old |
| 17 | 19 | 19-44898409-G-A | rs8106922 | TOMM40 | 0.3397 | G | 1.14E-38 | 0.000222 | -- | 4.969E-40 | 1.39E-23 | Old |
| 17 | 19 | 19-44899213-C-T | rs490243 | TOMM40 | 0.0313 | T | 6.13E-22 | 0.877704 | ++ | 6.062E-22 | 0.006487 | Old |
| 17 | 19 | 19-44899925-G-A | rs148998607 | TOMM40 | 0.0045 | A | 3.7E-10 | | +? | 3.697E-10 | 0.5643 | Old |
| 17 | 19 | 19-44899959-C-T | rs115881343 | TOMM40 | 0.0394 | T | 1.7E-56 | 0.003468 | ++ | 3.016E-57 | 2.504E-51 | Old |
| 17 | 19 | 19-44900155-C-T | rs1160985 | TOMM40 | 0.4813 | T | 6.67E-83 | 0.000021 | -- | 3.798E-85 | 7.095E-40 | Old |
| 17 | 19 | 19-44901174-C-T | rs741780 | TOMM40 | 0.4816 | C | 4.8E-82 | 0.000097 | -- | 5.139E-84 | 1.917E-39 | Old |
| 17 | 19 | 19-44901434-G-A | | TOMM40 | 0.2319 | A | 5.85E-20 | 0.009801 | -- | 1.392E-20 | 5.088E-28 | Old |
| 17 | 19 | 19-44901548-AACACGGTGAAACTCCGTCTCTACT-A | rs113492558 | TOMM40 | 0.0432 | A | 1.15E-55 | | +? | 1.152E-55 | 2.867E-54 | Old |
| 17 | 19 | 19-44901715-C-T | rs1038025 | TOMM40 | 0.4814 | C | 5.95E-82 | 0.000011 | -- | 2.802E-84 | 1.298E-39 | Old |
| 17 | 19 | 19-44901805-G-A | rs1038026 | TOMM40 | 0.4895 | G | 7.2E-84 | 0.000016 | -- | 3.503E-86 | 0.05318 | Old |
| 17 | 19 | 19-44901924-C-T | rs144618582 | TOMM40 | 0.0044 | C | 2.06E-10 | | +? | 2.055E-10 | 0.5715 | Old |
| 17 | 19 | 19-44902264-G-C | rs1305062 | TOMM40 | 0.3676 | C | 1.23E-45 | 0.000292 | -- | 4.689E-47 | 2.671E-25 | Old |
| 17 | 19 | 19-44903416-G-A | rs10119 | TOMM40 | 0.3081 | A | 4.7E-157 | 3.61E-09 | ++ | 1.67E-161 | 4.07E-131 | Old |
| 17 | 19 | 19-44903861-G-A | rs435380 | TOMM40 | 0.0318 | A | 6.09E-22 | 0.877704 | ++ | 6.023E-22 | 0.005987 | Old |
| 17 | 19 | 19-44904069-G-A | rs72654461 | TOMM40 | 0.0044 | A | 2.04E-10 | | +? | 2.035E-10 | 0.6109 | Old |
| 17 | 19 | 19-44904152-C-T | rs72654463 | TOMM40 | 0.0044 | T | 2.06E-10 | | +? | 2.059E-10 | 0.6394 | Old |
| 17 | 19 | 19-44904180-G-T | rs446037 | TOMM40 | 0.0316 | T | 5.38E-22 | 0.877704 | ++ | 5.318E-22 | 0.008961 | Old |
| 17 | 19 | 19-44904531-G-A | rs7259620 | TOMM40 | 0.4338 | A | 1.38E-68 | 7.88E-06 | -- | 8.172E-71 | 0.00467 | Old |
| 17 | 19 | 19-44905218-G-A | rs439382 | APOE | 0.0294 | G | 2.74E-17 | 0.815873 | ++ | 2.696E-17 | 0.01673 | Old |
| 17 | 19 | 19-44905579-G-T | | APOE | 0.4772 | T | 7.48E-35 | 0.000725 | ++ | 4.385E-36 | 1.754E-27 | Old |
| 17 | 19 | 19-44905910-G-C | | APOE | 0.3251 | C | 4.5E-32 | 0.001146 | -- | 3.983E-33 | 4.297E-45 | Old |
| 17 | 19 | 19-44906745-G-A | rs769449 | APOE | 0.237 | A | | 6.74E-13 | ?+ | 6.74E-13 | 1.23E-291 | Old |
| 17 | 19 | 19-44907187-G-A | rs769450 | APOE | 0.3389 | A | 9.8E-48 | 0.000113 | -- | 2.699E-49 | 1.41E-25 | Old |
| 17 | 19 | 19-44908684-C-T | rs429358 | APOE | 0.2104 | C | 1.9E-293 | 2.22E-16 | ++ | 7.24E-305 | 3.26E-305 | Old |
| 17 | 19 | 19-44908822-C-T | rs7412 | APOE | 0.0521 | T | 5.3E-39 | | -? | 5.297E-39 | 1.321E-35 | Old |
| 17 | 19 | 19-44909665-AC-A | rs537741299 | AC011481.3 | 0.0085 | A | 7.91E-14 | | +? | 7.909E-14 | 1.56E-16 | Old |
| 17 | 19 | 19-44909698-C-A | rs1081105 | AC011481.3 | 0.0414 | C | 1.29E-61 | 0.000614 | ++ | 1.351E-62 | 1.418E-54 | Old |
| 17 | 19 | 19-44910319-C-T | rs75627662 | AC011481.3 | 0.1855 | T | 1.1E-81 | 1.73E-08 | ++ | 1.148E-84 | 3.96E-106 | Old |
| 17 | 19 | 19-44912383-G-A | rs445925 | AC011481.3 | 0.1307 | A | 3.72E-11 | 0.741412 | ++ | 3.524E-11 | 0.237 | Old |
| 17 | 19 | 19-44912456-G-A | rs10414043 | AC011481.3 | 0.1643 | A | 3.7E-141 | 5.69E-13 | ++ | 3.56E-146 | 7.93E-275 | Old |
| 17 | 19 | 19-44912921-G-T | rs483082 | AC011481.3 | 0.2906 | T | 1E-134 | 1.54E-10 | ++ | 3.72E-139 | 3.74E-161 | Old |

| | | | | | | | | | | | | |
|---|---|---|---|---|---|---|---|---|---|---|---|---|
| 17 | 19 | 19-44913034-C-T | rs59325138 | AC011481.3 | 0.3018 | T | 2.88E-38 | 0.007392 | -- | 3.511E-39 | 1.092E-24 | Old |
| 17 | 19 | 19-44913484-C-T | rs438811 | AC011481.3 | 0.3 | T | 4.9E-156 | 1.8E-10 | ++ | 9.06E-161 | 2.59E-164 | Old |
| 17 | 19 | 19-44914381-C-CTTCG | rs11568822 | AC011481.3 | 0.2409 | CTTCG | 3.3E-124 | | +? | 3.27E-124 | 1.61E-163 | Old |
| 17 | 19 | 19-44915533-C-T | rs5117 | APOC1 | 0.2407 | C | 1.7E-124 | 1.01E-10 | ++ | 8.74E-129 | 1.25E-160 | Old |
| 17 | 19 | 19-44917843-G-A | rs3925681 | APOC1 | 0.3257 | A | 2.23E-41 | 0.00037 | -- | 1.001E-42 | 2.294E-33 | Old |
| 17 | 19 | 19-44917947-C-T | rs150966173 | APOC1 | 0.0416 | T | 4.82E-59 | 0.000185 | ++ | 4.119E-60 | 7.279E-51 | Old |
| 17 | 19 | 19-44917997-G-A | rs12721046 | APOC1 | 0.137 | A | 3.8E-154 | 1.21E-11 | ++ | 6.77E-159 | 4.97E-231 | Old |
| 17 | 19 | 19-44918715-AG-A | rs12721052 | APOC1 | 0.2728 | A | 2.64E-31 | | -? | 2.636E-31 | 1.867E-18 | Old |
| 17 | 19 | 19-44918903-G-C | rs12721051 | APOC1 | 0.1827 | G | 2.7E-221 | 2.44E-15 | ++ | 2.14E-228 | 1.57E-290 | Old |
| 17 | 19 | 19-44919589-G-A | rs56131196 | APOC1 | 0.2087 | A | 9.5E-196 | 4E-15 | ++ | 1.89E-202 | 7.74E-288 | Old |
| 17 | 19 | 19-44919689-G-A | rs4420638 | APOC1 | 0.2102 | G | 1.3E-195 | 2.66E-15 | ++ | 2.22E-202 | 3.77E-288 | Old |
| 17 | 19 | 19-44920677-G-A | rs157591 | APOC1, APOC4 | 0.034 | A | 1.45E-20 | 0.583236 | ++ | 1.37E-20 | 0.04405 | Old |
| 17 | 19 | 19-44923556-C-T | rs157598 | APOC1, APOC4 | 0.03 | T | 2.15E-16 | 0.894462 | ++ | 2.138E-16 | 0.03517 | Old |
| 17 | 19 | 19-44923868-T-A | rs111789331 | APOC1, APOC4 | 0.1386 | A | 8.8E-154 | 2.05E-11 | ++ | 1.84E-158 | 2.14E-228 | Old |
| 17 | 19 | 19-44924096-G-C | rs4803770 | APOC1, APOC4 | 0.2922 | G | 8.28E-32 | 0.013624 | -- | 1.435E-32 | 0.007863 | Old |
| 17 | 19 | 19-44924977-G-A | rs66626994 | APOC1, APOC4 | 0.1595 | A | 1.2E-126 | 5.99E-10 | ++ | 1.17E-130 | 5.84E-223 | Old |
| 17 | 19 | 19-44925202-C-T | rs4803772 | APOC1, APOC4 | 0.2679 | T | 1E-30 | 0.039148 | -- | 2.536E-31 | 3.768E-18 | Old |
| 17 | 19 | 19-44925842-G-A | rs10424663 | APOC1, APOC4 | 0.0311 | A | 1.96E-13 | 0.618201 | ++ | 1.889E-13 | 0.5211 | Old |
| 17 | 19 | 19-44926451-G-C | rs60049679 | APOC1, APOC4 | 0.135 | C | 4.49E-23 | | +? | 4.495E-23 | 3.926E-22 | Old |
| 17 | 19 | 19-44935297-C-T | rs7254133 | APOC1, APOC4 | 0.3202 | T | 5.68E-14 | 0.179036 | ++ | 3.048E-14 | 6.177E-27 | Old |
| 17 | 19 | 19-45019031-G-A | rs74359223 | RELB | 0.0132 | A | 2.5E-09 | 0.0361 | ++ | 1.769E-09 | 4.032E-07 | Old |
| 18 | 19 | 19-44792629-G-A | rs113330691 | CBLC | 0.0247 | A | 1.59E-09 | 0.102834 | -- | 1.183E-09 | 3.506E-08 | Old |
| 18 | 19 | 19-44793107-G-A | rs112450640 | CBLC | 0.0073 | A | 1.68E-08 | 0.05943 | -- | 1.308E-08 | 9.294E-08 | Old |
| 18 | 19 | 19-44793549-C-T | rs3208856 | CBLC | 0.0247 | T | 1.66E-09 | 0.102834 | -- | 1.229E-09 | 3.06E-08 | Old |
| 18 | 19 | 19-44795942-G-T | rs76560105 | CBLC | 0.0251 | T | 4.03E-08 | 0.12442 | -- | 3.133E-08 | 1.852E-08 | Old |
| 18 | 19 | 19-44799865-AAAAG-A | | CBLC | 0.0194 | A | 1.13E-09 | | -? | 1.132E-09 | 7.595E-08 | Old |
| 18 | 19 | 19-44813331-G-A | rs28399654 | BCAM | 0.0159 | A | 4.82E-12 | 0.032572 | -- | 3.081E-12 | 4.233E-08 | Old |
| 18 | 19 | 19-44822960-C-T | rs10406338 | BCAM, NECTIN2 | 0.3288 | C | 4.04E-09 | 0.315519 | ++ | 2.841E-09 | 0.03328 | Old |
| 18 | 19 | 19-44823407-C-T | rs10405693 | BCAM, NECTIN2 | 0.3257 | T | 1.04E-08 | 0.346166 | ++ | 7.553E-09 | 0.04225 | Old |
| 18 | 19 | 19-44824052-C-T | rs10412413 | BCAM, NECTIN2 | 0.3394 | T | 4.47E-08 | 0.285433 | ++ | 3.102E-08 | 0.07276 | Old |
| 18 | 19 | 19-44825110-A-T | rs58132661 | BCAM, NECTIN2 | 0.3357 | A | 6.78E-08 | 0.285433 | ++ | 4.717E-08 | 1.551E-22 | Old |
| 18 | 19 | 19-44825122-T-A | rs58826447 | BCAM, NECTIN2 | 0.3356 | A | 4.47E-08 | 0.285433 | ++ | 3.098E-08 | 1.627E-22 | Old |
| 18 | 19 | 19-44825123-C-A | rs58446550 | BCAM, NECTIN2 | 0.3356 | A | 4.85E-08 | 0.285433 | ++ | 3.365E-08 | 1.676E-22 | Old |
| 18 | 19 | 19-44842530-A-T | rs111371860 | BCAM, NECTIN2 | 0.0449 | A | 5.95E-08 | 0.162193 | -- | 4.612E-08 | 7.723E-11 | Old |
| 18 | 19 | 19-44852464-G-C | rs2972559 | NECTIN2 | 0.2808 | G | 1.44E-11 | 0.237637 | ++ | 8.952E-12 | 4.187E-26 | Old |
| 18 | 19 | 19-44853746-G-C | rs35396326 | NECTIN2 | 0.2829 | G | 1.24E-08 | 0.361936 | ++ | 9.136E-09 | 3.578E-22 | Old |
| 18 | 19 | 19-44856449-G-A | rs12462573 | NECTIN2 | 0.2874 | A | 8.49E-12 | 0.244986 | ++ | 5.326E-12 | 9.646E-29 | Old |
| 18 | 19 | 19-44865946-G-A | rs112422902 | NECTIN2 | 0.0147 | A | 1.48E-18 | 0.028643 | -- | 9.3E-19 | 1.327E-08 | Old |
| 18 | 19 | 19-44893972-G-A | rs1160983 | TOMM40 | 0.0335 | A | 1.64E-13 | 0.037303 | -- | 1.06E-13 | 1.499E-16 | Old |
| 18 | 19 | 19-44897490-T-A | rs61679753 | TOMM40 | 0.0397 | A | 2.77E-17 | 0.023627 | -- | 1.638E-17 | 8.891E-16 | Old |
| 18 | 19 | 19-44899005-G-T | rs111784051 | TOMM40 | 0.0418 | G | 1.23E-17 | | -? | 1.231E-17 | 2.023E-14 | Old |
| 18 | 19 | 19-44923535-G-A | rs141622900 | APOC1, APOC4 | 0.0429 | A | 1.72E-24 | 0.000276 | -- | 5.358E-25 | 4.792E-22 | Old |
| 19 | 19 | 19-44854682-G-A | rs2927468 | NECTIN2 | 0.4915 | A | 4.16E-08 | 0.038323 | -- | 1.77E-08 | 2.827E-29 | Old |
| 19 | 19 | 19-44864520-C-T | rs8105340 | NECTIN2 | 0.1069 | C | 7.74E-10 | 0.656235 | ++ | 7.023E-10 | 0.07928 | Old |
| 19 | 19 | 19-44864753-G-A | | NECTIN2 | 0.1645 | A | 1.48E-09 | | +? | 1.484E-09 | 0.02804 | Old |
| 20 | 19 | 19-44942260-G-A | rs79429216 | APOC4, APOC4-APOC2 | 0.013 | A | 4.61E-08 | 0.015442 | ++ | 3.208E-08 | 0.0002029 | Old |

| | | | | | | | | | | | | |
|---|---|---|---|---|---|---|---|---|---|---|---|---|
| 20 | 19 | 19-44960129-G-A | rs116949436 | CLPTM1 | 0.0115 | A | 6.78E-09 | 0.015442 | ++ | 4.626E-09 | 0.00001176 | Old |
| 21 | 20 | 20-51028527-C-T | rs1570009 | KCNG1, AL121785.1 | 0.2981 | C | 3.11E-08 | 0.29021 | -- | 2.269E-08 | 0.7088 | New |
| 21 | 20 | 20-51033011-C-T | rs6020921 | KCNG1, AL121785.1 | 0.3013 | C | 6E-09 | 0.315701 | -- | 4.384E-09 | 0.647 | New |
| 21 | 20 | 20-51036345-T-A | rs12480959 | KCNG1, AL121785.1 | 0.284 | A | 1.27E-08 | 0.483236 | -- | 1.044E-08 | 0.474 | New |
| 21 | 20 | 20-51038035-G-A | rs4809823 | KCNG1, AL121785.1 | 0.2908 | G | 2.75E-09 | 0.459513 | -- | 2.23E-09 | 0.4545 | New |
| 21 | 20 | 20-51041177-G-A | rs4811115 | KCNG1, AL121785.1 | 0.2933 | G | 9.02E-09 | 0.424146 | -- | 7.163E-09 | 0.3907 | New |
| 22 | 21 | 21-31398874-G-A | rs77728772 | TIAM1 | 0.0316 | G | 4.4E-08 | | -? | 0.000000044 | 0.2681 | New |
| 22 | 21 | 21-31402009-G-T | rs117323901 | TIAM1 | 0.0326 | G | 3.69E-08 | | -? | 3.687E-08 | 0.2573 | New |
| 22 | 21 | 21-31403761-C-T | rs117806270 | TIAM1 | 0.0327 | C | 2.79E-08 | | -? | 2.793E-08 | 0.2819 | New |
| 22 | 21 | 21-31403765-G-C | rs77589046 | TIAM1 | 0.0326 | C | 2.16E-08 | | -? | 2.159E-08 | 0.2793 | New |
