## Supplementary Table 4 for "Identification of 16 novel Alzheimer’s disease susceptibility loci using multi-ancestry meta-analyses of clinical Alzheimer’s disease and AD-by-proxy cases from four whole genome sequencing datasets"

**Supplementary table 4:** Independent loci that passed QC in our AD-by-proxy meta-analysis, including variants that were not nominally significant in our clinical AD meta-analysis. MAF - minor allele frequency; P - p-value; META_P - p-value for the meta analysis.

| Locus | Chromosome | Variant ID | Rsid | Gene | MAF | Effect allele | UKB_P | AOU_P | Direction (UKB, AOU) | UKB_AOU_META_P | NIA_NIMH_META_P | New or Old |
|---|---|---|---|---|---|---|---|---|---|---|---|---|
| 1 | 1 | 1-10152033-C-T | rs975197152 | UBE4B | 0.001 | T | 0.421838 | 1.6E-09 | -+ | 9.642E-09 | | New |
| 1 | 1 | 1-10152040-C-CCT | rs1367265225 | UBE4B | 0.0033 | CCT | | 7.74E-24 | ?+ | 7.735E-24 | | New |
| 1 | 1 | 1-10152050-AATTT-A | | NA | 0.0012 | AATTT | | 3.35E-10 | ?+ | 3.351E-10 | | New |
| 2 | 1 | 1-10237828-G-A | rs561741258 | KIF1B | 0.0007 | A | 0.550187 | 2E-10 | ++ | 1.05E-09 | | New |
| 3 | 1 | 1-10508359-G-A | rs1298189613 | PEX14 | 0.0002 | G | | 1.19E-10 | ?+ | 1.186E-10 | | New |
| 3 | 1 | 1-10508363-C-T | rs879878849 | PEX14 | 0.0002 | C | | 1.9E-10 | ?+ | 1.9E-10 | | New |
| 4 | 1 | 1-112574476-G-T | rs370350367 | ST7L | 0.0021 | G | 0.021861 | 1.85E-10 | ++ | 1.805E-10 | 1 | New |
| 5 | 1 | 1-11709990-G-T | rs190224580 | DRAXIN | 0.0012 | T | 0.464033 | 3.38E-10 | ++ | 5.115E-10 | 0.806 | New |
| 6 | 1 | 1-121400518-GAA-G | rs1553361219 | AC244021.1 | 0.0028 | G | | 1.18E-08 | ?+ | 1.185E-08 | | New |
| 7 | 1 | 1-125068456-G-T | rs1445112456 | NONE, NONE | 0.0005 | G | | 1.18E-09 | ?+ | 1.177E-09 | | New |
| 8 | 1 | 1-125176581-G-T | rs1452815376 | NONE, NONE | 0.0007 | T | | 1.79E-08 | ?+ | 1.791E-08 | | New |
| 9 | 1 | 1-1277184-ATG-A | | NA | 0.0006 | ATG | | 3.11E-08 | ?+ | 3.11E-08 | | Old |
| 10 | 1 | 1-13530355-G-A | rs1010617612 | AL354712.1, PDPN | 0.001 | A | 0.96944 | 6.63E-10 | -+ | 6.943E-09 | | New |
| 11 | 1 | 1-1370032-C-CAG | | NA | 0.0008 | CAG | | 3.22E-10 | ?+ | 3.223E-10 | | Old |
| 12 | 1 | 1-143195687-TGAATGGAATCATCATC-T | | NA | 0.0012 | TGAATGGAATCATCAT | | 1.41E-11 | ?+ | 1.413E-11 | | New |
| 13 | 1 | 1-143202364-A-G | rs1163770121 | NONE, RNA5SP533 | 0.006 | A | | 6.47E-12 | ?+ | 6.47E-12 | | New |
| 14 | 1 | 1-143210077-A-T | rs1225279127 | NONE, RNA5SP533 | 0.012 | T | 0.728836 | 9.7E-118 | -+ | 2.49E-116 | | New |
| 15 | 1 | 1-143253866-C-T | rs1648038996 | NONE, RNA5SP533 | 0.0031 | T | | 2.19E-11 | ?+ | 2.19E-11 | | New |
| 16 | 1 | 1-143272094-G-A | rs1237780657 | NONE, RNA5SP533 | 0.0008 | G | 0.149567 | 1.14E-11 | -+ | 1.789E-10 | | New |

| | | | | | | | | | | | | |
|---|---|---|---|---|---|---|---|---|---|---|---|---|
| 17 | 1 | 1-143272167-C-CG | | NA | 0.0008 | C | | 3.94E-08 | ?+ | 3.939E-08 | | New |
| 18 | 1 | 1-144565462-C-T | rs1658150431 | RF00003, PPIAL4F | 0.001 | C | | 1.88E-11 | ?+ | 1.877E-11 | 0.3152 | New |
| 19 | 1 | 1-144636407-C-CA | | NA | 0.0101 | CA | | 8.17E-12 | ?+ | 8.167E-12 | | New |
| 19 | 1 | 1-144636407-CTTCA-C | | NA | 0.013 | CTTCA | | 1.34E-10 | ?+ | 1.337E-10 | | New |
| 20 | 1 | 1-148017581-AC-A | | NA | 0.001 | A | | 4.53E-09 | ?+ | 4.533E-09 | | New |
| 20 | 1 | 1-148017587-AAT-A | | NA | 0.0009 | A | | 3.67E-08 | ?+ | 3.672E-08 | | New |
| 21 | 1 | 1-148066853-GCAT-G | | RNVU1-7, NBPF11 | 0.0004 | GCAT | | 4.13E-10 | ?+ | 4.13E-10 | | New |
| 21 | 1 | 1-148066859-GCA-G | | RNVU1-7, NBPF11 | 0.0003 | GCA | | 1.35E-08 | ?+ | 1.354E-08 | | New |
| 22 | 1 | 1-149920006-GCTGGGATTACAGGCGTGAGCCACTGCGCCTGGCTAACTTTTGTATTGTTAATGGAGACAGGGTTTCACCGTGTTGGCCAGGCTGCTCTCGAACTCCGGACCTCAAGTGATCCACTCACCTCT-G | | NA | 0.0062 | G | | 7.48E-28 | ?+ | 7.481E-28 | | New |
| 23 | 1 | 1-1500804-AAAAT-A | rs1264912848 | ATAD3B, ATAD3A | 0.0032 | A | 0.87353 | 1.75E-15 | ++ | 1.39E-10 | 0.1118 | Old |
| 24 | 1 | 1-150304119-TG-T | | NA | 0.0009 | T | 0.497618 | 4.53E-10 | -+ | 2.234E-09 | | New |
| 25 | 1 | 1-150526800-ATT-A | | NA | 0.0008 | A | | 3.68E-12 | ?+ | 3.684E-12 | | New |
| 26 | 1 | 1-150671030-AG-A | rs1300768323 | GOLPH3L | 0.0021 | A | | 1.31E-14 | ?+ | 1.31E-14 | | New |
| 27 | 1 | 1-150676899-C-T | rs990322706 | GOLPH3L | 0.0037 | C | 0.303001 | 9.78E-18 | ++ | 3.709E-08 | | New |
| 28 | 1 | 1-150960054-C-CA | rs1398444737 | SETDB1 | 0.0042 | C | | 1.28E-16 | ?+ | 1.275E-16 | | New |
| 29 | 1 | 1-151083907-ATTT-A | | NA | 0.002 | ATTT | | 6.95E-09 | ?+ | 6.953E-09 | | New |

| | | | | | | | | | | | | |
|---|---|---|---|---|---|---|---|---|---|---|---|---|
| 30 | 1 | 1-152109643-GT-G | | TCHH | 0.0016 | GT | | 8.29E-09 | ?+ | 8.293E-09 | | New |
| 30 | 1 | 1-152109644-C-CTT | | TCHH | 0.0019 | CTT | | 1.88E-10 | ?+ | 1.878E-10 | | New |
| 31 | 1 | 1-155274178-ATTAG-A | rs1224276847 | CLK2 | 0.0014 | A | | 1.81E-13 | ?+ | 1.805E-13 | | New |
| 32 | 1 | 1-155819544-C-CTA | | NA | 0.0009 | CTA | | 1.25E-10 | ?+ | 1.251E-10 | | New |
| 32 | 1 | 1-155819546-AAG-A | | NA | 0.0009 | A | | 1.59E-10 | ?+ | 1.592E-10 | | New |
| 33 | 1 | 1-155877614-C-CT | rs1213636743 | SYT11 | 0.0014 | CT | 0.342595 | 4.13E-19 | ++ | 3.477E-15 | | New |
| 34 | 1 | 1-155878634-GAC-G | | NA | 0.0008 | G | | 5.36E-10 | ?+ | 5.363E-10 | | New |
| 35 | 1 | 1-155948467-TA-T | rs1247800597 | ARHGEF2 | 0.0013 | TA | | 4.41E-10 | ?+ | 4.413E-10 | | New |
| 36 | 1 | 1-155955307-TAG-T | | NA | 0.0007 | TAG | | 3.18E-08 | ?+ | 3.179E-08 | | New |
| 37 | 1 | 1-155963917-GGT-G | | NA | 0.0014 | G | | 1.09E-09 | ?+ | 1.088E-09 | | New |
| 37 | 1 | 1-155963921-C-CAG | | NA | 0.0013 | CAG | | 1.78E-09 | ?+ | 1.777E-09 | | New |
| 38 | 1 | 1-15860133-G-T | rs1015482461 | SPEN | 0.0051 | T | 0.865415 | 2.18E-45 | ++ | 2.403E-30 | 0.116 | New |
| 38 | 1 | 1-15860146-C-T | rs995145323 | SPEN | 0.0033 | T | 0.901398 | 1.87E-26 | ++ | 2.196E-15 | | New |
| 39 | 1 | 1-15869308-GC-G | | NA | 0.0005 | G | | 1.84E-08 | ?+ | 1.843E-08 | | New |
| 40 | 1 | 1-15977177-C-CAA | rs1471550107 | ZBTB17, SRARP | 0.0013 | C | | 5.38E-10 | ?+ | 5.377E-10 | | New |
| 41 | 1 | 1-161348819-C-CAG | | NA | 0.0007 | C | | 9.94E-14 | ?+ | 9.94E-14 | | Old |
| 42 | 1 | 1-161372350-ATC-A | | NA | 0.0015 | A | | 1.53E-14 | ?+ | 1.531E-14 | | Old |
| 43 | 1 | 1-16382341-C-CCA | | NA | 0.0005 | CCA | | 7.08E-11 | ?+ | 7.081E-11 | | New |
| 44 | 1 | 1-16429296-C-CCCGG | rs1231827674 | SPATA21 | 0.0014 | CCCGG | | 1.31E-10 | ?+ | 1.309E-10 | 0.4158 | New |
| 45 | 1 | 1-16732342-G-A | rs4394648 | AL021920.1, AL021920.3 | 0.0068 | A | | 4.05E-09 | ?+ | 4.051E-09 | 0.6696 | New |
| 46 | 1 | 1-17006395-C-CATGCACCACCAT | | NA | 0.0025 | C | | 4.9E-25 | ?+ | 4.901E-25 | | New |

| | | | | | | | | | | | | |
|---|---|---|---|---|---|---|---|---|---|---|---|---|
| 47 | 1 | 1-17054484-C-CT | rs1281628355 | SDHB | 0.0023 | C | | 1.66E-08 | ?+ | 1.662E-08 | 0.5208 | New |
| 48 | 1 | 1-171621577-G-T | rs552223287 | MYOCOS | 0.0011 | T | 0.790158 | 1.85E-13 | -+ | 8.537E-13 | | New |
| 48 | 1 | 1-171621582-G-A | rs1652576846 | MYOCOS | 0.001 | G | | 7.35E-13 | ?+ | 7.348E-13 | | New |
| 49 | 1 | 1-173835689-C-CCT | rs1553202098 | DARS2 | 0.0006 | C | | 3.1E-08 | ?+ | 3.096E-08 | | New |
| 50 | 1 | 1-17543895-GCCCA-G | | NA | 0.001 | GCCCA | | 2.25E-08 | ?+ | 2.246E-08 | | New |
| 51 | 1 | 1-17544491-C-T | rs1223058726 | ARHGEF10L | 0.0015 | T | 0.112974 | 5.15E-13 | ++ | 3.833E-11 | | New |
| 51 | 1 | 1-17544499-G-T | rs2996647 | ARHGEF10L | 0.0837 | T | 1.52E-05 | 2.37E-16 | ++ | 7.192E-17 | 0.7419 | New |
| 52 | 1 | 1-17544542-ACCTG-A | | NA | 0.0006 | ACCTG | | 1.47E-10 | ?+ | 1.473E-10 | | New |
| 53 | 1 | 1-179070132-G-A | rs889994155 | FAM20B | 0.0013 | A | 0.688914 | 3.31E-11 | ++ | 3.653E-10 | 0.9853 | New |
| 54 | 1 | 1-179070191-ACC-A | rs1351651013 | FAM20B | 0.001 | ACC | | 2.49E-08 | ?+ | 2.485E-08 | | New |
| 55 | 1 | 1-179174275-AG-A | | NA | 0.0007 | AG | | 3.76E-09 | ?+ | 3.764E-09 | | New |
| 56 | 1 | 1-197576732-C-T | | DENND1B | 0.0007 | C | | 9.26E-11 | ?+ | 9.256E-11 | | Old |
| 57 | 1 | 1-200258342-T-A | | LINC00862 | 0.0006 | A | | 3.07E-08 | ?+ | 3.074E-08 | | New |
| 58 | 1 | 1-200284721-C-CTG | | NA | 0.0009 | C | | 1.45E-08 | ?+ | 1.454E-08 | | New |
| 59 | 1 | 1-200345290-C-CGG | | NA | 0.0009 | CGG | | 2.28E-11 | ?+ | 2.282E-11 | | New |
| 59 | 1 | 1-200345291-ACC-A | | NA | 0.0008 | A | | 1.51E-10 | ?+ | 1.513E-10 | | New |
| 60 | 1 | 1-202740096-GCA-G | rs879685982 | KDM5B | 0.0037 | G | | 1.51E-09 | ?+ | 1.506E-09 | | New |
| 61 | 1 | 1-203503088-T-A | rs1661417525 | OPTC | 0.0007 | T | | 1.11E-09 | ?+ | 1.112E-09 | | New |
| 62 | 1 | 1-203921617-AGG-A | | NA | 0.0003 | AGG | | 1.36E-08 | ?+ | 1.36E-08 | | New |
| 63 | 1 | 1-20463538-ACC-A | rs2051320721 | LINC01141, AL139254.2 | 0.0118 | ACC | | 5.43E-15 | ?+ | 5.424E-15 | | New |

| | | | | | | | | | | | | |
|---|---|---|---|---|---|---|---|---|---|---|---|---|
| 64 | 1 | 1-20647193-AC-A | | NA | 0.0003 | A | | 1.37E-08 | ?+ | 1.374E-08 | | New |
| 65 | 1 | 1-20796590-C-CCT | rs1423973664 | HP1BP3, EIF4G3 | 0.0062 | CCT | | 1.85E-12 | ?+ | 1.845E-12 | | New |
| 66 | 1 | 1-210976986-G-C | rs572893139 | AL590132.1, KCNH1 | 0.0006 | G | | 4.53E-08 | ?+ | 4.53E-08 | 0.581 | New |
| 67 | 1 | 1-214897967-G-A | | CENPF, AC099563.2 | 0.0011 | A | | 3.77E-15 | ?+ | 3.767E-15 | | New |
| 68 | 1 | 1-218455927-G-A | rs190180107 | TGFB2, C1orf143 | 0.0019 | A | 0.163756 | 2.23E-10 | ++ | 8.967E-11 | 0.8605 | New |
| 69 | 1 | 1-218561848-G-C | rs1324865687 | RF00012, LINC01710 | 0.0039 | C | 0.372117 | 2.65E-32 | -+ | 2.009E-22 | 0.6764 | New |
| 70 | 1 | 1-220048846-C-CAA | | NA | 0.0006 | CAA | | 1.5E-10 | ?+ | 1.501E-10 | | New |
| 70 | 1 | 1-220048851-T-A | | EPRS, BPNT1 | 0.0004 | A | | 7.16E-09 | ?+ | 7.161E-09 | | New |
| 70 | 1 | 1-220048860-C-T | rs1264199617 | EPRS, BPNT1 | 0.0004 | T | 0.6469 | 6.14E-09 | ++ | 1.391E-08 | | New |
| 71 | 1 | 1-220140384-TGA-T | rs1290134825 | IARS2 | 0.0035 | T | | 2.69E-08 | ?+ | 2.693E-08 | 0.1619 | New |
| 72 | 1 | 1-22058633-C-CCACCA | | NA | 0.001 | CCACCA | | 1.07E-10 | ?+ | 1.075E-10 | | New |
| 73 | 1 | 1-223119009-C-A | rs1264236841 | TLR5 | 0.0015 | A | 0.970535 | 2.89E-14 | ++ | 7.218E-09 | 0.637 | New |
| 74 | 1 | 1-224206818-C-CCT | | NA | 0.0007 | CCT | | 1.35E-08 | ?+ | 1.352E-08 | | New |
| 75 | 1 | 1-22429164-C-CCT | | NA | 0.0005 | CCT | | 3.24E-08 | ?+ | 3.24E-08 | | New |
| 76 | 1 | 1-226142102-C-CCT | | NA | 0.0013 | CCT | | 4.08E-16 | ?+ | 4.08E-16 | | New |
| 77 | 1 | 1-228892872-C-T | rs1657042371 | RHOU, AL137793.1 | 0.0011 | C | | 3.29E-08 | ?+ | 3.29E-08 | | New |
| 78 | 1 | 1-241063951-AGGTG-A | | NA | 0.0005 | A | | 1.18E-11 | ?+ | 1.184E-11 | | New |
| 79 | 1 | 1-246376445-G-T | rs1368723920 | SMYD3 | 0.0006 | T | | 5.79E-09 | ?+ | 5.787E-09 | | New |
| 80 | 1 | 1-24812865-C-CAT | rs1358228775 | CLIC4 | 0.0018 | C | | 1.69E-16 | ?+ | 1.695E-16 | | New |
| 80 | 1 | 1-24812868-GCA-G | rs1261343213 | CLIC4 | 0.0018 | GCA | | 6.34E-14 | ?+ | 6.342E-14 | | New |

| | | | | | | | | | | | | |
|---|---|---|---|---|---|---|---|---|---|---|---|---|
| 81 | 1 | 1-25925787-AGTGC-A | | NA | 0.0005 | A | | 3.09E-10 | ?+ | 3.091E-10 | | New |
| 82 | 1 | 1-26108552-TG-T | rs1213491372 | TRIM63, PDIK1L | 0.0016 | T | 0.447481 | 1.91E-10 | ++ | 3.979E-10 | 0.1473 | New |
| 83 | 1 | 1-26334705-ACC-A | | NA | 0.0013 | ACC | | 4.5E-11 | ?+ | 4.495E-11 | | New |
| 83 | 1 | 1-26334710-GTCAA-G | | NA | 0.0005 | G | | 9.13E-10 | ?+ | 9.132E-10 | | New |
| 84 | 1 | 1-26648341-C-CCT | rs2079977919 | RF01210, AL512408.1 | 0.001 | C | | 1.09E-08 | ?+ | 1.085E-08 | 0.1683 | New |
| 85 | 1 | 1-26651469-AGC-A | | NA | 0.0008 | A | | 4.22E-10 | ?+ | 4.215E-10 | | New |
| 85 | 1 | 1-26651472-C-CTT | | NA | 0.0006 | CTT | | 1.55E-08 | ?+ | 1.552E-08 | | New |
| 86 | 1 | 1-27232615-C-CA | rs1210025951 | AL590640.1 | 0.0029 | CA | | 9.46E-09 | ?+ | 9.463E-09 | | New |
| 87 | 1 | 1-28350749-GTA-G | | NA | 0.0007 | G | | 4.1E-10 | ?+ | 4.103E-10 | | New |
| 88 | 1 | 1-28391247-AAAT-A | rs547260551 | PHACTR4 | 0.006 | A | | 3.3E-09 | ?+ | 3.295E-09 | 0.07281 | New |
| 89 | 1 | 1-28649458-C-CAG | rs1408113834 | AL360012.1, LINC01715, RNU11 | 0.0037 | CAG | | 3.4E-15 | ?+ | 3.398E-15 | | New |
| 89 | 1 | 1-28649461-GAT-G | rs1439737489 | AL360012.1, LINC01715, RNU11 | 0.0031 | G | | 3.36E-14 | ?+ | 3.363E-14 | 0.5987 | New |
| 90 | 1 | 1-28649645-GGCA-G | | NA | 0.0008 | G | | 1.64E-14 | ?+ | 1.638E-14 | | New |
| 91 | 1 | 1-28715380-C-CCT | rs1421604029 | GMEB1 | 0.0008 | CCT | | 3.47E-08 | ?+ | 3.474E-08 | | New |
| 92 | 1 | 1-28733121-AT-A | | NA | 0.0004 | AT | | 3.39E-12 | ?+ | 3.389E-12 | | New |
| 93 | 1 | 1-28994406-C-CAA | | NA | 0.0012 | CAA | | 1.23E-08 | ?+ | 1.23E-08 | | New |
| 94 | 1 | 1-31578988-T-C | rs12736080 | LINC01226 | 0.008 | C | | 1.58E-16 | ?+ | 1.58E-16 | | New |
| 95 | 1 | 1-35117555-C-CAG | | NA | 0.001 | C | | 3.86E-11 | ?+ | 3.856E-11 | | New |
| 96 | 1 | 1-35579988-C-CCT | | NA | 0.0008 | C | | 4.69E-09 | ?+ | 4.687E-09 | 0.8455 | New |

| | | | | | | | | | | | | |
|---|---|---|---|---|---|---|---|---|---|---|---|---|
| 97 | 1 | 1-36102439-C-A | rs1643690279 | COL8A2 | 0.0016 | A | 0.750459 | 1.18E-14 | ++ | 4.662E-10 | | New |
| 98 | 1 | 1-36571154-C-CT | rs201208642 | CSF3R, AL596257.1 | 0.0013 | C | 0.486388 | 2.47E-09 | ++ | 1.986E-09 | | New |
| 99 | 1 | 1-37598789-C-CTCAA | | NA | 0 | C | | 2.04E-08 | ?+ | 2.037E-08 | | New |
| 100 | 1 | 1-37884763-AAT-A | rs1178610444 | INPP5B | 0.0008 | AAT | | 4.6E-09 | ?+ | 4.601E-09 | 0.2432 | New |
| 100 | 1 | 1-37884766-ATG-A | 1-37884766-ATG-/ | NA | 0.0007 | A | | 4.22E-09 | ?+ | 4.216E-09 | | New |
| 101 | 1 | 1-37935848-C-CCA | rs1488544082 | INPP5B | 0.0054 | CCA | 0.518856 | 1.27E-27 | -+ | 1.63E-23 | | New |
| 102 | 1 | 1-3798817-C-CAAA | | NA | 0.0008 | CAAA | | 3.43E-15 | ?+ | 3.434E-15 | | New |
| 103 | 1 | 1-38869984-T-A | rs1642424605 | AL139260.1, AL139260.2 | 0.0008 | A | 0.745338 | 6.08E-15 | -+ | 2.112E-12 | | New |
| 104 | 1 | 1-39067815-G-T | | NDUFS5, MACF1 | 1E-04 | G | 0.844572 | 3.03E-09 | ++ | 1.045E-08 | | New |
| 105 | 1 | 1-39179844-G-A | rs903391718 | MACF1 | 0.0017 | A | 0.82374 | 5.74E-23 | ++ | 7.818E-18 | | New |
| 106 | 1 | 1-39504208-GAT-G | rs548532112 | BMP8A | 0.0034 | G | 0.781916 | 1.42E-19 | ++ | 3.523E-11 | 0.1694 | New |
| 107 | 1 | 1-39579706-C-CGTG | | NA | 0.0006 | C | | 7.97E-09 | ?+ | 7.974E-09 | | New |
| 108 | 1 | 1-39606527-C-A | rs991345515 | PABPC4, HEYL | 0.0014 | C | 0.929636 | 1.85E-11 | -+ | 1.65E-10 | 0.06786 | New |
| 108 | 1 | 1-39606529-C-T | rs568528046 | PABPC4, HEYL | 0.0009 | T | | 1.9E-11 | ?+ | 1.898E-11 | 0.01975 | New |
| 109 | 1 | 1-39993465-TTC-T | | NA | 0.0011 | T | | 3.34E-09 | ?+ | 3.338E-09 | | New |
| 110 | 1 | 1-40590743-C-CAA | | NA | 0.0005 | CAA | | 8.47E-09 | ?+ | 8.471E-09 | | New |
| 110 | 1 | 1-40590746-TATCG-T | | NA | 0.0003 | T | | 3.29E-08 | ?+ | 3.287E-08 | | New |
| 111 | 1 | 1-42614311-ACCT-A | | NA | 0.0021 | A | | 1.45E-14 | ?+ | 1.448E-14 | | New |
| 112 | 1 | 1-43248985-C-A | | CFAP57 | 0.0018 | A | 0.789124 | 2.94E-15 | ++ | 9.166E-12 | | New |
| 113 | 1 | 1-44148272-C-CTACT | rs1553173615 | KLF18, RF00019 | 0.0007 | CTACT | | 4.14E-08 | ?+ | 4.145E-08 | | New |
| 114 | 1 | 1-44952753-C-T | rs987270886 | EIF2B3 | 0.0041 | C | 0.125232 | 3.51E-26 | ++ | 2.898E-16 | 0.4301 | New |
| 114 | 1 | 1-44952782-G-T | rs946244174 | EIF2B3 | 0.0064 | G | 0.974855 | 2.69E-35 | ++ | 5.092E-18 | | New |

| | | | | | | | | | | | | |
|---|---|---|---|---|---|---|---|---|---|---|---|---|
| 115 | 1 | 1-46311457-GC-G | | NA | 0.0033 | G | | 1.41E-10 | ?+ | 1.405E-10 | | New |
| 116 | 1 | 1-46323003-ACC-A | | NA | 0.0012 | ACC | | 3.65E-10 | ?+ | 3.645E-10 | | New |
| 117 | 1 | 1-46337062-C-A | rs903253466 | UQCRH, NSUN4 | 0.0024 | A | 0.861843 | 8.51E-20 | ++ | 6.43E-13 | | New |
| 117 | 1 | 1-46337077-G-A | rs1661880714 | UQCRH, NSUN4 | 0.003 | A | 0.650541 | 8.98E-31 | ++ | 3.967E-21 | | New |
| 117 | 1 | 1-46337088-G-A | rs1035837513 | UQCRH, NSUN4 | 0.0033 | A | 0.984205 | 7.11E-29 | -+ | 5.953E-19 | | New |
| 118 | 1 | 1-52623572-C-T | | GPX7, SHISAL2A | 0.0001 | T | | 1.16E-08 | ?+ | 1.165E-08 | | New |
| 119 | 1 | 1-52910506-C-T | rs1376370722 | ECHDC2 | 0.0005 | C | | 1.31E-09 | ?+ | 1.306E-09 | | New |
| 119 | 1 | 1-52910512-GCAC-G | | NA | 0.0006 | G | | 3.37E-09 | ?+ | 3.368E-09 | | New |
| 120 | 1 | 1-54466273-ACC-A | | NA | 0.0003 | ACC | | 4.74E-10 | ?+ | 4.74E-10 | | New |
| 120 | 1 | 1-54466274-TTG-T | | NA | 0.0005 | T | | 3.69E-14 | ?+ | 3.693E-14 | | New |
| 121 | 1 | 1-54504924-C-CA | rs889487643 | AL035415.1, AC099796.3 | 0.0008 | C | | 1.51E-08 | ?+ | 1.51E-08 | | New |
| 122 | 1 | 1-5674860-GAAT-G | | NA | 0.0026 | G | | 2.83E-08 | ?+ | 2.828E-08 | 0.5474 | New |
| 123 | 1 | 1-5884481-AGC-A | rs1386709764 | NPHP4 | 0.0025 | A | | 1.36E-08 | ?+ | 1.363E-08 | | New |
| 124 | 1 | 1-61155454-ACC-A | rs1182012625 | NFIA | 0.0034 | ACC | | 4.31E-12 | ?+ | 4.306E-12 | 0.3071 | New |
| 125 | 1 | 1-63237607-C-CA | | NA | 0.0007 | CA | | 1.78E-09 | ?+ | 1.781E-09 | | New |
| 126 | 1 | 1-85367115-G-A | rs1431252608 | AL078459.1 | 0.0003 | A | | 4.81E-09 | ?+ | 4.807E-09 | | New |
| 127 | 1 | 1-86848155-T-A | rs1658217273 | AL355981.1, SELENOF | 0.0005 | T | | 9.51E-10 | ?+ | 9.512E-10 | | New |
| 128 | 1 | 1-8774498-C-CAT | | NA | 0.0012 | C | | 4.74E-08 | ?+ | 4.738E-08 | | New |
| 128 | 1 | 1-8774520-AT-A | rs2124551100 | NA | 0.0005 | AT | 0.027594 | 2.49E-12 | ++ | 2.957E-11 | | New |
| 129 | 1 | 1-88806458-C-CCG | | NA | 0.001 | CCG | | 1.52E-08 | ?+ | 1.522E-08 | | New |
| 130 | 1 | 1-90050171-AGG-A | | NA | 0.0002 | AGG | | 6.99E-10 | ?+ | 6.987E-10 | | New |

| | | | | | | | | | | | | |
|---|---|---|---|---|---|---|---|---|---|---|---|---|
| 131 | 1 | 1-9158215-G-A | rs1409228577 | MIR34AHG | 0.0008 | G | 0.299098 | 4.41E-10 | ++ | 4.433E-10 | 0.9312 | New |
| 131 | 1 | 1-9158256-AG-A | rs1359100579 | MIR34AHG | 0.0005 | A | | 2.92E-09 | ?+ | 2.916E-09 | | New |
| 132 | 1 | 1-9623342-C-CCT | rs1200379345 | TMEM201, PIK3CD | 0.003 | CCT | | 1.73E-25 | ?+ | 1.733E-25 | | New |
| 132 | 1 | 1-9623353-G-A | rs1644420296 | TMEM201, PIK3CD | 0.0005 | A | | 4.55E-08 | ?+ | 4.546E-08 | | New |
| 133 | 1 | 1-9702141-T-A | rs1646657652 | PIK3CD | 0.0017 | T | | 6.5E-18 | ?+ | 6.504E-18 | | New |
| 133 | 1 | 1-9702143-T-A | rs1646657755 | PIK3CD | 0.0017 | T | | 3.16E-15 | ?+ | 3.154E-15 | | New |
| 134 | 1 | 1-9830445-C-T | rs11121495 | AL357140.3, CTNNBIP1 | 0.3177 | C | | 2.58E-13 | ?+ | 2.576E-13 | 0.9323 | New |
| 135 | 2 | 2-101207215-ATT-A | | NA | 0.0011 | A | | 6.71E-12 | ?+ | 6.711E-12 | | New |
| 136 | 2 | 2-101578677-T-A | rs1052317533 | AC093894.2, MAP4K4 | 0.0011 | T | | 3.71E-08 | ?+ | 3.706E-08 | | New |
| 137 | 2 | 2-106888375-G-T | rs1208010228 | ST6GAL2, AC005040.2 | 0.0005 | G | | 2.52E-08 | ?+ | 2.523E-08 | | New |
| 138 | 2 | 2-11052875-TCG-T | rs1455283239 | KCNF1, AC062028.1 | 0.0009 | TCG | | 1.68E-08 | ?+ | 1.676E-08 | 0.99 | Old |
| 138 | 2 | 2-11052877-GCA-G | rs1173935228 | KCNF1, AC062028.1 | 0.0008 | G | | 2E-08 | ?+ | 1.996E-08 | | Old |
| 139 | 2 | 2-111295877-ATTTTTT-A | | MIR4435-2HG | 0.0791 | A | | 1.17E-12 | ?- | 1.167E-12 | | New |
| 140 | 2 | 2-111880255-C-T | rs1465759459 | ANAPC1 | 0.0008 | T | 0.006346 | 3.03E-09 | ++ | 6.677E-11 | | New |
| 141 | 2 | 2-113864927-TA-T | | NA | 0.0023 | T | | 4.84E-13 | ?+ | 4.843E-13 | | New |
| 142 | 2 | 2-118093497-AGATGATGATGAT-A | | INSIG2 | 0.3283 | A | | 2.98E-14 | ?- | 2.979E-14 | | New |
| 143 | 2 | 2-120197492-ACT-A | rs1408747551 | AC012363.1 | 0.0035 | ACT | | 1.02E-18 | ?+ | 1.015E-18 | 0.151 | New |
| 144 | 2 | 2-126871297-G-A | rs1679763294 | RNU6-675P, TEX51 | 0.0009 | G | 0.539144 | 2.48E-15 | -+ | 5.943E-12 | | Old |

| | | | | | | | | | | | | |
|---|---|---|---|---|---|---|---|---|---|---|---|---|
| 145 | 2 | 2-127128582-C-T | rs13025717 | BIN1, AC110926.2 | 0.2947 | T | 4.4E-08 | 0.024032 | ++ | 7.457E-09 | 0.00001185 | Old |
| 145 | 2 | 2-127128657-C-T | rs13025765 | BIN1, AC110926.2 | 0.2942 | T | 4.17E-08 | 0.019492 | ++ | 5.418E-09 | 0.00001229 | Old |
| 145 | 2 | 2-127128840-A-C | rs12617835 | BIN1, AC110926.2 | 0.2944 | A | 5.39E-08 | 0.023627 | ++ | 8.693E-09 | 0.1052 | Old |
| 145 | 2 | 2-127133851-C-A | rs4663105 | BIN1, AC110926.2 | 0.4334 | C | 2.51E-10 | 0.01738 | ++ | 6.348E-11 | 8.114E-11 | Old |
| 145 | 2 | 2-127135234-C-T | rs6733839 | BIN1, AC110926.2 | 0.3923 | T | 4.4E-12 | 0.033562 | ++ | 6.137E-12 | 4.673E-11 | Old |
| 146 | 2 | 2-127858638-C-CTG | rs1274020366 | POLR2D | 0.0026 | CTG | | 2.36E-15 | ?+ | 2.357E-15 | 0.2504 | New |
| 146 | 2 | 2-127858655-G-C | rs1690388098 | POLR2D | 0.0009 | C | | 8.83E-11 | ?+ | 8.827E-11 | 0.1055 | New |
| 147 | 2 | 2-128059121-C-CTT | | SAP130, RF00019 | 0.0018 | CTT | | 1.65E-12 | ?+ | 1.647E-12 | | New |
| 148 | 2 | 2-128082350-GCACA-G | | RF00019, UGGT1 | 0.0021 | G | | 7.5E-09 | ?+ | 7.501E-09 | | New |
| 149 | 2 | 2-130469139-C-CA | | POTEI | 0.03 | CA | | 2.28E-11 | ?+ | 2.275E-11 | | New |
| 150 | 2 | 2-130863966-C-CTA | | NA | 0.0021 | CTA | | 7.07E-17 | ?+ | 7.076E-17 | | New |
| 151 | 2 | 2-131634722-G-C | rs1001830266 | RNU6-617P, LINC01087 | 0.0009 | G | 0.167768 | 1.5E-10 | ++ | 1.596E-10 | | New |
| 152 | 2 | 2-131756355-G-A | rs1305401557 | C2orf27A | 0.0041 | G | | 9.49E-09 | ?+ | 9.487E-09 | | New |
| 153 | 2 | 2-133308914-G-T | rs1681023547 | NCKAP5 | 0.0012 | T | 0.527445 | 6.25E-12 | ++ | 1.423E-09 | | New |
| 153 | 2 | 2-133308921-G-A | rs1681024656 | NCKAP5 | 0.0009 | G | 0.609481 | 1.04E-11 | ++ | 8.118E-09 | | New |
| 153 | 2 | 2-133308945-C-T | rs1338208268 | NCKAP5 | 0.0011 | T | 0.960418 | 5.64E-15 | -+ | 4.207E-08 | | New |
| 153 | 2 | 2-133308946-G-A | rs1475548985 | NCKAP5 | 0.0014 | G | 0.931604 | 3.95E-20 | -+ | 8.106E-12 | | New |
| 153 | 2 | 2-133308953-G-A | rs1041743937 | NCKAP5 | 0.0013 | G | 0.946634 | 1.55E-15 | -+ | 8.234E-09 | | New |

| | | | | | | | | | | | | |
|---|---|---|---|---|---|---|---|---|---|---|---|---|
| 154 | 2 | 2-134710815-C-T | rs10164986 | TMEM163 | 0.3221 | C | | 4.4E-08 | ?+ | 4.401E-08 | 0.4644 | Old |
| 155 | 2 | 2-135470542-C-CT | | NA | 0.0011 | C | | 6.69E-14 | ?+ | 6.694E-14 | | New |
| 156 | 2 | 2-151415128-ACC-A | rs1296731650 | RIF1 | 0.0023 | ACC | | 8.08E-14 | ?+ | 8.079E-14 | 0.1995 | New |
| 157 | 2 | 2-157549149-T-A | | ACVR1C | 0.0002 | A | | 3.5E-09 | ?+ | 3.504E-09 | | New |
| 157 | 2 | 2-157549175-GA-G | | ACVR1C | 0.0001 | G | | 3.02E-09 | ?+ | 3.024E-09 | | New |
| 158 | 2 | 2-169608315-G-C | rs905835670 | PPIG | 0.0029 | C | 0.32132 | 1.41E-20 | ++ | 5.098E-20 | | New |
| 159 | 2 | 2-174296329-TA-T | rs1242963535 | RF00019, LINC01305 | 0.0021 | TA | 0.506122 | 6.78E-23 | ++ | 1.771E-15 | | New |
| 160 | 2 | 2-179792562-G-A | rs367921783 | ZNF385B | 0.0006 | G | | 3.76E-08 | ?+ | 3.765E-08 | | New |
| 160 | 2 | 2-179792566-G-A | rs1296008867 | ZNF385B | 0.0006 | G | | 4.46E-10 | ?+ | 4.459E-10 | | New |
| 161 | 2 | 2-183479004-C-CTG | | AC021851.2 | 0.0019 | C | | 5.77E-11 | ?+ | 5.768E-11 | | New |
| 162 | 2 | 2-198215952-C-CCT | rs1404822667 | AC020719.1, LINC01923 | 0.002 | CCT | | 1.39E-11 | ?+ | 1.385E-11 | | New |
| 163 | 2 | 2-202003854-C-CCT | | NA | 0.0003 | C | | 3.38E-09 | ?+ | 3.382E-09 | | New |
| 164 | 2 | 2-202294096-C-CCT | | NA | 0.0012 | C | | 2.61E-09 | ?+ | 2.614E-09 | | Old |
| 165 | 2 | 2-202567965-C-CGT | | NA | 0.0005 | CGT | | 2.45E-08 | ?+ | 2.446E-08 | | Old |
| 166 | 2 | 2-202813151-C-T | | ICA1L | 0.0004 | C | | 2.83E-13 | ?+ | 2.83E-13 | | Old |
| 167 | 2 | 2-202861728-AAT-A | | NA | 0.0004 | AAT | | 2.83E-08 | ?+ | 2.827E-08 | | Old |
| 168 | 2 | 2-205992418-ATGG-A | | NA | 0.0004 | ATGG | | 5.67E-10 | ?+ | 5.667E-10 | | New |
| 168 | 2 | 2-205992420-AGGG-A | | NA | 0.0002 | A | | 4.47E-09 | ?+ | 4.47E-09 | | New |
| 169 | 2 | 2-206050883-G-C | rs1575851053 | INO80D | 0.0007 | C | 0.802356 | 4.28E-13 | -+ | 1.175E-10 | | New |
| 170 | 2 | 2-213584240-TA-T | rs1385628861 | SPAG16 | 0.0024 | TA | 0.7376 | 5.92E-20 | -+ | 9.462E-19 | | New |

| | | | | | | | | | | | | |
|---|---|---|---|---|---|---|---|---|---|---|---|---|
| 171 | 2 | 2-219581029-AGGTG-A | | NA | 0.0006 | A | | 1.22E-08 | ?+ | 1.216E-08 | | New |
| 171 | 2 | 2-219581037-AGGTG-A | | NA | 0.0007 | AGGTG | | 1.91E-12 | ?+ | 1.909E-12 | | New |
| 172 | 2 | 2-224529423-C-A | rs1219833709 | CUL3 | 0.0009 | A | | 4.38E-10 | ?+ | 4.379E-10 | | New |
| 173 | 2 | 2-224755811-C-CCT | rs1553553653 | CCDC195, DOCK10 | 0.0026 | CCT | | 1.43E-08 | ?+ | 1.432E-08 | | New |
| 174 | 2 | 2-231440470-ATG-A | rs1264652607 | B3GNT7, AC017104.3 | 0.0015 | ATG | | 1.26E-16 | ?+ | 1.255E-16 | 0.5342 | New |
| 174 | 2 | 2-231440472-ACG-A | rs1473940278 | B3GNT7, AC017104.3 | 0.001 | A | | 1.31E-09 | ?+ | 1.308E-09 | 0.4514 | New |
| 175 | 2 | 2-231501097-G-C | | RNU2-22P | 0.0004 | C | | 1.32E-09 | ?+ | 1.321E-09 | | New |
| 176 | 2 | 2-231565270-G-A | rs1360681730 | NMUR1, AC104634.2 | 0.0006 | A | | 3.38E-09 | ?+ | 3.381E-09 | 0.825 | New |
| 176 | 2 | 2-231565277-G-A | rs1308706830 | NMUR1, AC104634.2 | 0.0006 | A | 0.618733 | 9.46E-11 | ++ | 1.237E-08 | 0.5981 | New |
| 176 | 2 | 2-231565279-G-C | rs1375871492 | NMUR1, AC104634.2 | 0.0008 | C | 0.508329 | 7.88E-16 | ++ | 9.499E-14 | 0.411 | New |
| 177 | 2 | 2-234316318-G-A | rs569481169 | AC122134.1, AC097713.3 | 0.0009 | A | 0.197389 | 1.09E-11 | -+ | 1.472E-08 | 0.1614 | New |
| 178 | 2 | 2-238119146-G-GAA | rs142229517 | ESPNL | 0.0143 | G | | 1.6E-09 | ?+ | 1.595E-09 | | New |
| 179 | 2 | 2-238794552-C-T | rs1469424262 | AC145625.1, TWIST2 | 0.0009 | T | | 1.09E-11 | ?+ | 1.09E-11 | | New |
| 180 | 2 | 2-239815908-GA-G | rs1231605654 | AC093802.1 | 0.0069 | GA | | 1.22E-10 | ?+ | 1.218E-10 | | New |
| 181 | 2 | 2-241488609-G-C | rs938331976 | FARP2 | 0.0031 | G | 0.717588 | 1.67E-44 | ++ | 2.178E-42 | | New |
| 182 | 2 | 2-24152050-G-T | rs762345986 | AC008073.3, FAM228B | 0.0001 | T | | 4.32E-08 | ?+ | 4.316E-08 | | New |
| 183 | 2 | 2-25534570-C-CT | rs1295517516 | DTNB | 0.0035 | C | | 7.17E-11 | ?+ | 7.166E-11 | | Old |

| | | | | | | | | | | | | |
|---|---|---|---|---|---|---|---|---|---|---|---|---|
| 184 | 2 | 2-27126918-A-G | rs12614792 | ABHD1 | 0.0165 | G | 0.251618 | 4.73E-19 | ++ | 2.225E-14 | 0.7722 | New |
| 185 | 2 | 2-27141043-T-A | | PRR30, TCF23 | 0.0003 | A | | 2.48E-08 | ?+ | 2.476E-08 | | New |
| 186 | 2 | 2-27273508-GAT-G | rs1259975228 | SLC30A3 | 0.0058 | G | | 2.09E-11 | ?+ | 2.09E-11 | 0.0362 | New |
| 187 | 2 | 2-27511478-C-CCT | | NA | 0.001 | C | | 1.47E-10 | ?+ | 1.466E-10 | | New |
| 188 | 2 | 2-33306351-C-CTT | rs1195724496 | LTBP1 | 0.0009 | C | | 3.71E-09 | ?+ | 3.714E-09 | 0.7061 | New |
| 189 | 2 | 2-34321033-C-CA | rs1465685846 | LINC01320 | 0.0036 | C | | 1.08E-10 | ?+ | 1.082E-10 | 0.7088 | New |
| 190 | 2 | 2-38696978-G-T | rs1340901888 | GALM | 0.0003 | T | 0.867679 | 2.54E-09 | ++ | 1.056E-08 | | New |
| 191 | 2 | 2-39089972-C-CAAAAAAAAAAAA | | NA | 0.0007 | C | | 3.23E-11 | ?+ | 3.226E-11 | | New |
| 192 | 2 | 2-39126426-AT-A | | NA | 0.0017 | A | | 7.9E-11 | ?+ | 7.905E-11 | | New |
| 193 | 2 | 2-46273971-C-CAAAGAAAGA | | NA | 0.0007 | CAAAGAAAGA | | 1.98E-09 | ?+ | 1.977E-09 | | New |
| 194 | 2 | 2-47353179-AT-A | rs1344764494 | EPCAM | 0.001 | A | | 1.06E-10 | ?+ | 1.058E-10 | | New |
| 195 | 2 | 2-47768888-TGG-T | rs1321375918 | MSH6 | 0.0014 | TGG | | 2.16E-09 | ?+ | 2.158E-09 | | New |
| 196 | 2 | 2-53970819-C-CGAA | rs1390937290 | PSME4 | 0.0064 | C | | 2.31E-12 | ?+ | 2.312E-12 | | New |
| 197 | 2 | 2-53978264-ACTT-A | | NA | 0.0006 | ACTT | | 1.16E-08 | ?+ | 1.162E-08 | | New |
| 198 | 2 | 2-54004201-G-T | | ACYP2 | 0.0003 | T | | 4.07E-08 | ?+ | 4.073E-08 | | New |
| 199 | 2 | 2-61148337-C-CAA | | NA | 0.0003 | CAA | | 2.49E-08 | ?+ | 2.488E-08 | | New |
| 200 | 2 | 2-62167572-ATT-A | rs1394091179 | COMMD1, B3GNT2 | 0.0018 | A | | 8.19E-12 | ?+ | 8.188E-12 | 0.1757 | New |
| 201 | 2 | 2-6420447-C-CATGAGGTCAGGAGATTGAG | rs1241385483 | LINC01247, LINC01824 | 0.0052 | C | | 1.36E-08 | ?+ | 1.364E-08 | | New |
| 202 | 2 | 2-65157396-TA-T | rs1233069191 | RAB1A, RF00090 | 0.0008 | TA | | 6.16E-10 | ?+ | 6.16E-10 | 0.299 | Old |
| 203 | 2 | 2-65168546-C-CCT | rs1395183799 | RF00090, ACTR2 | 0.0042 | CCT | | 4.06E-10 | ?+ | 4.063E-10 | 0.07489 | Old |
| 204 | 2 | 2-65280182-ACC-A | | ACTR2, SPRED2 | 0.0014 | ACC | | 9E-10 | ?+ | 9.004E-10 | | Old |

| | | | | | | | | | | | | |
|---|---|---|---|---|---|---|---|---|---|---|---|---|
| 205 | 2 | 2-65446107-C-A | rs944659757 | AC007389.1, AC012370.2 | 0.0014 | A | | 2.54E-21 | ?+ | 2.537E-21 | | Old |
| 205 | 2 | 2-65446110-G-C | rs1669197221 | AC007389.1, AC012370.2 | 0.0012 | G | | 5.69E-16 | ?+ | 5.687E-16 | | Old |
| 206 | 2 | 2-70188373-T-A | rs1265024412 | C2orf42 | 0.0013 | A | 0.846904 | 1.11E-14 | ++ | 9.119E-11 | 0.9363 | New |
| 207 | 2 | 2-79011165-C-CT | rs1439643019 | AC092604.1, REG3G | 0.0015 | C | | 2.39E-12 | ?+ | 2.385E-12 | | Old |
| 208 | 2 | 2-79818457-C-CAT | | NA | 0.0008 | CAT | | 1.88E-10 | ?+ | 1.884E-10 | | New |
| 209 | 2 | 2-85532753-C-CACCG | | NA | 0.0019 | CACCG | | 4.04E-13 | ?+ | 4.035E-13 | | New |
| 210 | 2 | 2-86191352-ATT-A | | NA | 0.0009 | ATT | | 1.12E-09 | ?+ | 1.122E-09 | | New |
| 211 | 2 | 2-86191352-ATTAAT-A | | IMMT | 0.0006 | ATTAAT | | 8.72E-09 | ?+ | 8.725E-09 | | New |
| 212 | 2 | 2-88005024-G-A | rs1205506154 | RGPD2, RNU2-63P | 0.001 | G | 0.714893 | 2.18E-12 | ++ | 3.082E-12 | 0.5392 | New |
| 212 | 2 | 2-88005034-T-A | rs1232324269 | RGPD2, RNU2-63P | 0.0009 | T | | 3.7E-12 | ?+ | 3.699E-12 | | New |
| 212 | 2 | 2-88005040-C-T | rs1479717350 | RGPD2, RNU2-63P | 0.0007 | T | | 3.1E-16 | ?+ | 3.103E-16 | 0.3969 | New |
| 213 | 2 | 2-88005607-C-CA | | RGPD2, RNU2-63P | 0.0005 | C | | 7.6E-09 | ?+ | 7.601E-09 | 0.4046 | New |
| 214 | 2 | 2-89636593-GA-G | | RF00001, IGKV2D-40 | 0.4534 | GA | | 3.98E-09 | ?- | 3.98E-09 | | New |
| 214 | 2 | 2-89636593-GAAAA-G | | RF00001, IGKV2D-40 | 0.3313 | G | | 2.66E-27 | ?+ | 2.659E-27 | | New |
| 215 | 2 | 2-90331433-G-C | rs1221908556 | IGKV3D-7, AC233263.6 | 0.002 | G | | 2.82E-09 | ?+ | 2.823E-09 | | New |

| | | | | | | | | | | | | |
|---|---|---|---|---|---|---|---|---|---|---|---|---|
| 216 | 2 | 2-90396303-T-C | rs62141985 | AC233263.6, AC233266.2 | 0.0054 | C | | 4.07E-11 | ?- | 4.073E-11 | | New |
| 217 | 2 | 2-9503812-TG-T | rs1318606417 | AC080162.1 | 0.0004 | T | | 4.69E-10 | ?+ | 4.689E-10 | | Old |
| 218 | 2 | 2-99650376-G-T | rs566358991 | AFF3 | 0.0007 | G | 0.59274 | 1.9E-14 | ++ | 4.03E-14 | | New |
| 219 | 3 | 3-10030681-C-CCT | rs1484030501 | FANCD2 | 0.0035 | C | | 2.14E-27 | ?+ | 2.136E-27 | | New |
| 220 | 3 | 3-10102824-C-CA | rs1227758043 | FANCD2OS | 0.0006 | CA | | 7.12E-11 | ?+ | 7.122E-11 | | New |
| 221 | 3 | 3-10239992-C-CAA | rs1349978361 | IRAK2 | 0.0057 | CAA | | 1.07E-17 | ?+ | 1.068E-17 | | New |
| 222 | 3 | 3-103419699-GT-G | rs1707023343 | RNU1-43P, MIR548AB | 0.0043 | G | 0.094139 | 1.66E-31 | ++ | 7.12E-20 | | New |
| 223 | 3 | 3-109285911-C-CGT | | NA | 0.0009 | CGT | | 9.05E-09 | ?+ | 9.05E-09 | | New |
| 224 | 3 | 3-119480415-ATT-A | | NA | 0.0022 | ATT | | 5.25E-15 | ?+ | 5.253E-15 | 0.2544 | New |
| 225 | 3 | 3-121791872-C-CCT | | IQCB1 | 0.0012 | C | | 1.89E-12 | ?+ | 1.894E-12 | | New |
| 226 | 3 | 3-122934000-C-CAGGTTCA | rs61012123 | SEMA5B | 0.0023 | C | | 4.4E-10 | ?+ | 4.402E-10 | 0.3543 | New |
| 227 | 3 | 3-124436561-G-A | rs1375566941 | KALRN | 0.0007 | A | | 3.72E-11 | ?+ | 3.719E-11 | 0.3207 | New |
| 227 | 3 | 3-124436578-T-A | rs2093468087 | KALRN | 0.0007 | A | | 3.62E-08 | ?+ | 3.624E-08 | | New |
| 228 | 3 | 3-124807408-TC-T | | ITGB5 | 0.0003 | T | | 7.49E-11 | ?+ | 7.492E-11 | 0.4432 | New |
| 229 | 3 | 3-124898425-C-CCT | rs1254559012 | ITGB5 | 0.0051 | CCT | | 1.88E-10 | ?+ | 1.876E-10 | | New |
| 230 | 3 | 3-12502323-ATC-A | rs1267492347 | TSEN2 | 0.0016 | A | | 1.15E-11 | ?+ | 1.151E-11 | 0.9146 | New |
| 231 | 3 | 3-125521495-AAAATAC-A | | NA | 0.0003 | AAAATAC | | 4.31E-08 | ?+ | 4.306E-08 | | New |
| 232 | 3 | 3-127582685-GAGT-G | | NA | 0.0006 | GAGT | | 1.39E-10 | ?+ | 1.389E-10 | | New |
| 232 | 3 | 3-127582687-GCAA-G | | NA | 0.0004 | G | | 3.46E-09 | ?+ | 3.464E-09 | | New |
| 233 | 3 | 3-128645253-C-CCT | | NA | 0.0006 | C | | 1.06E-09 | ?+ | 1.056E-09 | | New |

| | | | | | | | | | | | |
|---|---|---|---|---|---|---|---|---|---|---|---|
| 234 | 3 | 3-128696475-G-A | rs1325254595 | RPN1, RF00399 | 0.0005 | G | | 2.42E-08 | ?+ | 2.418E-08 | 0.141 | New |
| 234 | 3 | 3-128696485-C-CTT | | NA | 0.0007 | C | | 5.31E-13 | ?+ | 5.31E-13 | | New |
| 235 | 3 | 3-129426499-G-T | rs1396035227 | EFCAB12 | 0.0004 | T | 0.435313 | 1.09E-10 | ++ | 2.243E-08 | | New |
| 236 | 3 | 3-136704609-C-CTT | rs1292259832 | STAG1 | 0.0014 | C | | 1.2E-08 | ?+ | 1.201E-08 | 0.3846 | New |
| 237 | 3 | 3-138728773-C-CCG | | NA | 0.0014 | C | | 3.16E-10 | ?+ | 3.156E-10 | 0.9268 | New |
| 238 | 3 | 3-151417487-C-CGGG | | NA | 0.0016 | CGGG | | 3.26E-08 | ?- | 3.258E-08 | | New |
| 239 | 3 | 3-15491870-C-T | rs1198422174 | COLQ | 0.0007 | T | | 6.47E-09 | ?+ | 6.466E-09 | 0.4823 | New |
| 239 | 3 | 3-15491876-G-A | rs1378323638 | COLQ | 0.0008 | A | 0.979003 | 1.35E-13 | ++ | 7.723E-13 | | New |
| 240 | 3 | 3-15569473-C-T | rs1424900334 | HACL1 | 0.001 | C | | 8.02E-10 | ?+ | 8.02E-10 | | New |
| 240 | 3 | 3-15569482-G-A | rs1306953234 | HACL1 | 0.0006 | A | 0.593871 | 9.45E-09 | -+ | 1.473E-08 | | New |
| 240 | 3 | 3-15569486-C-T | rs1336810492 | HACL1 | 0.0005 | C | 0.408286 | 5.66E-09 | -+ | 1.548E-08 | | New |
| 241 | 3 | 3-155795816-C-T | rs902725266 | AC104472.3, C3orf33 | 0.0018 | C | 0.11838 | 3.39E-11 | ++ | 1.813E-08 | 0.07872 | New |
| 242 | 3 | 3-15592997-C-CATGTGTGCGTGTATACACATGTACGCAT | rs1487030640 | HACL1 | 0.0095 | C | | 1.52E-08 | ?+ | 1.52E-08 | | New |
| 243 | 3 | 3-15593380-AT-A | | NA | 0.0013 | A | | 4.92E-14 | ?+ | 4.925E-14 | | New |
| 243 | 3 | 3-15593389-ATTTT-A | | NA | 0.0012 | A | | 9.19E-12 | ?+ | 9.191E-12 | | New |
| 244 | 3 | 3-170509612-C-T | rs1201387997 | SLC7A14-AS1 | 0.0006 | T | | 3.42E-09 | ?+ | 3.417E-09 | | New |
| 245 | 3 | 3-170873157-C-CT | | NA | 0.0006 | CT | 0.7272 | 8.52E-12 | -+ | 1.789E-10 | | New |
| 246 | 3 | 3-174762303-G-A | rs1423662667 | NAALADL2 | 0.002 | A | 0.284941 | 4.05E-10 | ++ | 6.916E-10 | 0.0212 | New |
| 247 | 3 | 3-177225071-C-CAG | rs1371401738 | TBL1XR1 | 0.0008 | C | | 1.33E-10 | ?+ | 1.327E-10 | | New |
| 248 | 3 | 3-183473142-GGAGT-G | rs1287529016 | RF00394, KLHL6 | 0.0057 | GGAGT | | 1.05E-11 | ?+ | 1.046E-11 | | New |
| 249 | 3 | 3-184425210-C-T | rs1298828254 | LINC02054 | 0.001 | T | | 1.74E-13 | ?+ | 1.736E-13 | 0.1827 | New |
| 250 | 3 | 3-185258472-G-T | rs62288696 | EHHADH, MAP3K13 | 0.0015 | T | 0.925728 | 6.71E-14 | ++ | 1.612E-08 | | New |

| | | | | | | | | | | | | |
|---|---|---|---|---|---|---|---|---|---|---|---|---|
| 251 | 3 | 3-187807145-T-A | rs1239483384 | AC108681.1, AC068295.1 | 0.0015 | A | | 6.46E-12 | ?+ | 6.456E-12 | | New |
| 252 | 3 | 3-192900736-T-A | rs1242199581 | MB21D2 | 0.002 | A | 0.427822 | 2.49E-27 | ++ | 3.502E-25 | 0.3559 | New |
| 252 | 3 | 3-192900746-G-T | rs1257773292 | MB21D2 | 0.0019 | T | 0.427822 | 8.47E-20 | ++ | 3.198E-18 | | New |
| 253 | 3 | 3-196266254-C-CCT | | AC069257.3, PCYT1A | 0.0025 | CCT | | 1.62E-11 | ?+ | 1.619E-11 | | New |
| 254 | 3 | 3-196675754-C-CAAGTG | | PIGX | 0.0007 | CAAGTG | | 1.66E-08 | ?+ | 1.658E-08 | | New |
| 255 | 3 | 3-196852149-AC-A | | NA | 0.0009 | A | | 3.75E-16 | ?+ | 3.752E-16 | | New |
| 256 | 3 | 3-196861715-GAT-G | | NA | 0.0008 | G | | 3.05E-08 | ?+ | 3.048E-08 | | New |
| 256 | 3 | 3-196861718-GCA-G | rs1297156851 | PAK2, SENP5 | 0.0007 | GCA | | 3.03E-08 | ?+ | 3.032E-08 | | New |
| 257 | 3 | 3-3431293-G-T | rs1304462752 | AC034195.1 | 0.0017 | T | 0.122209 | 4.34E-21 | -+ | 1.752E-08 | 0.9713 | New |
| 258 | 3 | 3-36963928-C-T | rs1170846268 | TRANK1, RNU6ATAC4P | 0.0014 | T | | 5.88E-09 | ?+ | 5.88E-09 | 0.4509 | Old |
| 258 | 3 | 3-36963941-G-T | rs536145993 | TRANK1, RNU6ATAC4P | 0.0007 | T | 0.528123 | 3.73E-08 | ++ | 4.126E-08 | | Old |
| 259 | 3 | 3-40344008-AGATG-A | | NA | 0.0005 | AGATG | | 5.17E-12 | ?+ | 5.171E-12 | | New |
| 260 | 3 | 3-41061991-AT-A | rs1277391500 | AC099541.1, AC009743.1 | 0.0011 | AT | | 2.6E-10 | ?+ | 2.599E-10 | 0.2223 | New |
| 260 | 3 | 3-41061993-AG-A | rs1349409080 | AC099541.1, AC009743.1 | 0.0013 | A | | 1.06E-10 | ?+ | 1.061E-10 | | New |
| 261 | 3 | 3-41971464-C-CA | rs1208311413 | ULK4, TRAK1 | 0.0019 | C | | 6.17E-11 | ?+ | 6.174E-11 | 0.8239 | New |
| 262 | 3 | 3-47755019-GAT-G | rs1485429079 | SMARCC1 | 0.002 | G | | 6.48E-11 | ?+ | 6.483E-11 | 0.8586 | New |
| 262 | 3 | 3-47755022-GCA-G | rs1191304654 | SMARCC1 | 0.0019 | GCA | | 1.48E-10 | ?+ | 1.482E-10 | | New |

| | | | | | | | | | | | |
|---|---|---|---|---|---|---|---|---|---|---|---|
| 263 | 3 | 3-47789474-G-T | | SMARCC1, DHX30 | 0.0001 | T | | 3.78E-08 | ?+ | 3.781E-08 | | New |
| 264 | 3 | 3-48876877-C-CCT | | NA | 0.0007 | C | | 1.65E-11 | ?+ | 1.649E-11 | | New |
| 265 | 3 | 3-48925935-G-C | rs2085530621 | ARIH2 | 0.0008 | G | | 1.54E-09 | ?+ | 1.543E-09 | | New |
| 266 | 3 | 3-48939528-C-CGGGTGGATCATG | | NA | 0.0029 | C | | 7.08E-09 | ?+ | 7.082E-09 | | New |
| 267 | 3 | 3-49210704-TCTAA-T | | NA | 0.0013 | TCTAA | | 7.16E-11 | ?+ | 7.159E-11 | | New |
| 268 | 3 | 3-49947243-C-CATGAGAA | | NA | 0.0019 | CATGAGAA | | 2.7E-16 | ?+ | 2.699E-16 | | New |
| 269 | 3 | 3-51353784-C-CCTCT | rs1262184171 | DOCK3 | 0.0021 | C | 0.18184 | 8.99E-18 | -+ | 5.872E-17 | 0.9227 | New |
| 270 | 3 | 3-73151147-G-A | rs761486125 | RNU2-64P, RNU6-1270P | 0.0001 | A | 0.46582 | 1.56E-09 | -+ | 1.425E-08 | 0.4679 | New |
| 271 | 3 | 3-75612176-C-CG | | LINC02018, MIR1324 | 0.1081 | CG | | 1.23E-10 | ?+ | 1.234E-10 | | New |
| 272 | 3 | 3-75616418-ATG-A | | NA | 0.0014 | ATG | 0.413409 | 1.33E-20 | ++ | 8.457E-17 | | New |
| 273 | 3 | 3-75674095-TCAAATATGGGTCAAATATGGCTTAG-T | | NA | 0.0034 | T | | 4.02E-13 | ?+ | 4.022E-13 | | New |
| 274 | 3 | 3-75734134-T-A | rs79804154 | LINC00960 | 0.0001 | A | | 1.47E-08 | ?+ | 1.468E-08 | | New |
| 275 | 3 | 3-77292683-GGGTAAGCTGAGGCTAGATCACCCCAGACATAAAGTAAAATTGATGGTTAAACGGGAAGTTGAGGCTAGAGCACTAAAGACATAAAGTAAATTTGACGGTTAAACA-G | rs1560455525 | ROBO2 | 0.2612 | G | | 1.24E-10 | ?- | 1.243E-10 | | New |
| 276 | 3 | 3-90283097-C-CT | rs373822597 | RNU6-712P, RF01699 | 0.0018 | CT | | 6.09E-09 | ?+ | 6.091E-09 | | New |

| | | | | | | | | | | | | |
|---|---|---|---|---|---|---|---|---|---|---|---|---|
| 277 | 3 | 3-90498243-G-C | rs1196020873 | RNU6-712P, RF01699 | 0.001 | G | | 2.01E-08 | ?+ | 2.007E-08 | | New |
| 278 | 3 | 3-90498313-G-T | rs1249242045 | RNU6-712P, RF01699 | 0.0008 | T | 0.385347 | 1.62E-09 | ++ | 1.52E-09 | | New |
| 279 | 3 | 3-90587749-C-T | rs1206123205 | RNU6-712P, RF01699 | 0.0005 | T | 0.003102 | 2.15E-06 | ++ | 4.287E-08 | | New |
| 280 | 3 | 3-9803125-C-T | rs3872719 | ARPC4, ARPC4-TTLL3 | 0.001 | T | 0.708871 | 1.9E-09 | ++ | 3.016E-09 | 0.2456 | New |
| 281 | 4 | 4-102329460-TAA-T | | NA | 0.0008 | TAA | | 4.09E-12 | ?+ | 4.093E-12 | 0.5972 | New |
| 281 | 4 | 4-102329468-C-CAA | | NA | 0.0018 | CAA | | 2.19E-15 | ?+ | 2.186E-15 | | New |
| 282 | 4 | 4-113231218-ATT-A | | ANK2 | 0.0012 | A | | 3.75E-12 | ?+ | 3.749E-12 | | New |
| 283 | 4 | 4-114959008-T-A | rs1303093395 | NDST4 | 0.0006 | A | | 4.95E-10 | ?+ | 4.947E-10 | | New |
| 284 | 4 | 4-120327591-C-CTCAT | | AC073475.1 | 0.0021 | C | | 3.11E-10 | ?+ | 3.113E-10 | | New |
| 285 | 4 | 4-128104101-C-CAA | | NA | 0.0003 | CAA | | 1.65E-09 | ?+ | 1.647E-09 | | New |
| 286 | 4 | 4-144912122-C-T | rs1441804178 | AC098588.1, ANAPC10 | 0.0012 | T | | 2.17E-08 | ?+ | 2.173E-08 | 0.1819 | New |
| 286 | 4 | 4-144912136-G-C | rs574655882 | AC098588.1, ANAPC10 | 0.0013 | G | 0.671216 | 1.57E-09 | -+ | 3.657E-08 | 0.6657 | New |
| 287 | 4 | 4-151065713-C-CAA | | LRBA, RPS3A | 0.0018 | C | | 1.05E-09 | ?+ | 1.055E-09 | 0.5179 | New |
| 288 | 4 | 4-151299398-T-A | rs903887193 | SH3D19 | 0.0005 | A | 0.780394 | 7.97E-10 | -+ | 3.457E-08 | | New |
| 289 | 4 | 4-153944779-C-CTCAT | | SFRP2, AC079298.3 | 0.0024 | CTCAT | | 4.25E-11 | ?+ | 4.247E-11 | | New |
| 290 | 4 | 4-159437376-AG-A | | NA | 0.0011 | A | | 6.07E-09 | ?+ | 6.069E-09 | | New |
| 291 | 4 | 4-162815005-C-T | rs570743232 | AC021134.1, AC021134.2 | 0.0012 | T | 0.603129 | 1.86E-09 | -+ | 1.377E-08 | 0.2183 | New |

| | | | | | | | | | | | | |
|---|---|---|---|---|---|---|---|---|---|---|---|---|
| 291 | 4 | 4-162815032-G-A | rs556209078 | AC021134.1, AC021134.2 | 0.0017 | A | 0.977475 | 1.69E-09 | ++ | 2.991E-09 | 0.2505 | New |
| 292 | 4 | 4-168966338-C-CAGTG | rs1261220185 | CBR4 | 0.0019 | CAGTG | | 3.95E-09 | ?+ | 3.946E-09 | 0.8995 | New |
| 293 | 4 | 4-172464489-C-CCT | | NA | 1E-04 | C | | 2.49E-08 | ?+ | 2.485E-08 | | New |
| 294 | 4 | 4-17565394-C-T | rs1368642558 | CLRN2, LAP3 | 0.0008 | T | | 6.25E-09 | ?+ | 6.252E-09 | | New |
| 295 | 4 | 4-176800802-C-A | rs866049921 | VEGFC, AC097518.2 | 0.0099 | A | | 1.04E-09 | ?- | 1.038E-09 | | New |
| 296 | 4 | 4-182621523-G-GA | rs11291088 | TENM3 | 0.0423 | G | | 1.69E-12 | ?+ | 1.695E-12 | | New |
| 297 | 4 | 4-183509969-C-CAG | rs1483773823 | ING2 | 0.005 | C | | 1.11E-11 | ?+ | 1.108E-11 | | New |
| 298 | 4 | 4-19483278-C-T | rs1163397286 | AC024230.1 | 0.0005 | C | 0.020852 | 2.84E-08 | ++ | 1.132E-08 | 0.5251 | New |
| 299 | 4 | 4-21544845-C-CTTTTTTT | | NA | 0.0008 | CTTTTTTT | | 2.56E-09 | ?+ | 2.559E-09 | | New |
| 300 | 4 | 4-22509278-G-A | rs1254742144 | ADGRA3 | 0.0017 | G | 0.896124 | 7.67E-15 | ++ | 3.712E-14 | 0.5028 | New |
| 301 | 4 | 4-2522168-G-A | rs546019839 | RNF4 | 0.0024 | A | 0.982773 | 2.98E-15 | ++ | 1.785E-08 | 0.2682 | New |
| 301 | 4 | 4-2522176-C-T | rs576468917 | RNF4 | 0.0043 | C | 0.545021 | 9.46E-26 | ++ | 2.015E-19 | 0.5542 | New |
| 301 | 4 | 4-2522177-G-A | rs541977342 | RNF4 | 0.0036 | A | 0.651691 | 1.84E-23 | ++ | 6.485E-17 | 0.6676 | New |
| 302 | 4 | 4-2522803-C-CCT | rs1340008592 | RNF4 | 0.0025 | CCT | 0.322392 | 8.21E-22 | ++ | 2.521E-16 | | New |
| 303 | 4 | 4-26243074-AAC-A | | NA | 0.0003 | AAC | | 2.49E-09 | ?+ | 2.489E-09 | | New |
| 304 | 4 | 4-2764348-C-CAG | | NA | 0.0046 | C | | 1.9E-11 | ?+ | 1.899E-11 | | New |
| 305 | 4 | 4-38320415-G-A | rs79769504 | AC098680.1, LINC02513 | 0.0001 | A | 0.021242 | 5.39E-07 | ++ | 0.000000035 | | New |
| 306 | 4 | 4-38486000-C-A | | LINC01258 | 0.0004 | A | 0.154024 | 7.32E-08 | ++ | 2.612E-08 | | New |
| 307 | 4 | 4-39359815-C-CCA | | NA | 0.0002 | CCA | | 8.62E-09 | ?+ | 8.616E-09 | | New |
| 308 | 4 | 4-39684712-GCC-G | rs1717431194 | AC108471.2, UBE2K | 0.0158 | GCC | | 7.48E-11 | ?+ | 7.477E-11 | | New |
| 309 | 4 | 4-39773277-GT-G | rs1182056131 | UBE2K | 0.006 | G | | 3.9E-22 | ?+ | 3.896E-22 | | Old |

| | | | | | | | | | | | | |
|---|---|---|---|---|---|---|---|---|---|---|---|---|
| 309 | 4 | 4-39773298-TC-T | rs879792800 | UBE2K | 0.0011 | TC | 0.937658 | 1.63E-10 | -+ | 4.277E-09 | | Old |
| 310 | 4 | 4-42081999-C-CCT | | NA | 0.0011 | CCT | | 1.61E-13 | ?+ | 1.608E-13 | | New |
| 311 | 4 | 4-49308277-C-CT | | NA | 0.0014 | C | | 5.67E-09 | ?+ | 5.67E-09 | | New |
| 312 | 4 | 4-49641607-AATGGAATAAAAGGTCATGGAATGGAAT-A | | NA | 0.0939 | A | | 4.11E-09 | ?- | 4.114E-09 | | New |
| 313 | 4 | 4-56835539-GGTGGA-G | | NA | 0.0007 | G | | 6.48E-11 | ?+ | 6.481E-11 | | New |
| 314 | 4 | 4-634885-GCC-G | rs1224926429 | PDE6B | 0.0017 | G | | 1.68E-09 | ?+ | 1.676E-09 | | Old |
| 314 | 4 | 4-634889-GCC-G | rs1478230788 | PDE6B | 0.0013 | G | | 1.23E-08 | ?+ | 1.228E-08 | | Old |
| 315 | 4 | 4-70871593-C-CTG | rs1457264965 | RNU6-891P, MOB1B | 0.0017 | C | | 4.48E-08 | ?+ | 4.481E-08 | 0.3342 | New |
| 316 | 4 | 4-70884321-C-CAT | | NA | 0.0005 | CAT | | 1.19E-12 | ?+ | 1.189E-12 | | New |
| 317 | 4 | 4-75682763-C-CCT | rs1240119130 | G3BP2 | 0.0004 | CCT | | 1.26E-08 | ?+ | 1.265E-08 | | Old |
| 318 | 4 | 4-76667749-ACC-A | rs1381828529 | SHROOM3 | 0.0011 | ACC | | 8.93E-11 | ?+ | 8.929E-11 | | Old |
| 319 | 4 | 4-83575604-C-T | rs1335344166 | GPAT3 | 0.0031 | T | 0.251024 | 2.07E-21 | -+ | 5.665E-13 | | New |
| 320 | 4 | 4-84770219-C-CG | | NA | 0.001 | CG | | 5.91E-09 | ?+ | 5.915E-09 | | New |
| 321 | 4 | 4-86958333-C-T | rs903117011 | AFF1 | 0.0008 | C | | 1.58E-08 | ?+ | 1.584E-08 | 0.6511 | New |
| 322 | 4 | 4-8967028-AAC-A | | NA | 0.0009 | AAC | | 1.66E-11 | ?+ | 1.658E-11 | | New |
| 323 | 4 | 4-920686-GACGT-G | rs1470992668 | GAK | 0.0019 | GACGT | | 1.03E-13 | ?+ | 1.027E-13 | 0.3599 | Old |
| 323 | 4 | 4-920689-GCCCA-G | rs1723794096 | GAK | 0.0013 | G | | 2.38E-11 | ?+ | 2.382E-11 | | Old |
| 324 | 4 | 4-9501107-AAC-A | | DEFB131A, MIR548I2 | 0.002 | AAC | | 4.69E-17 | ?+ | 4.686E-17 | | New |
| 324 | 4 | 4-9501126-C-T | | DEFB131A, MIR548I2 | 0.0003 | C | | 9.75E-09 | ?+ | 9.747E-09 | | New |
| 325 | 4 | 4-9593894-C-T | rs1213918414 | AC105916.1 | 0.0002 | C | | 2.38E-08 | ?+ | 2.381E-08 | | New |
| 326 | 4 | 4-9650027-T-A | | AC105916.1 | 0.0003 | A | | 1.02E-09 | ?+ | 1.019E-09 | | New |
| 327 | 4 | 4-978148-G-A | rs1713432349 | DGKQ | 0.0002 | G | | 2.91E-10 | ?+ | 2.91E-10 | | Old |

| | | | | | | | | | | | | |
|---|---|---|---|---|---|---|---|---|---|---|---|---|
| 328 | 4 | 4-99086726-C-CCT | rs1351881089 | ADH5 | 0.003 | CCT | | 1.7E-09 | ?+ | 1.697E-09 | | New |
| 329 | 5 | 5-116698546-C-A | rs1218851442 | SEMA6A-AS2, LINC02214 | 0.001 | A | 0.37322 | 3.53E-08 | ++ | 3.158E-08 | 0.3596 | New |
| 330 | 5 | 5-119047353-TGA-T | | NA | 0.0008 | TGA | | 6.76E-11 | ?+ | 6.761E-11 | | New |
| 331 | 5 | 5-134615116-AGGCG-A | rs1298939524 | SAR1B | 0.0065 | AGGCG | | 9.99E-34 | ?+ | 9.988E-34 | 0.2472 | New |
| 331 | 5 | 5-134615122-GCAGA-G | rs1233962092 | SAR1B | 0.0051 | G | | 1.08E-22 | ?+ | 1.08E-22 | 0.1536 | New |
| 332 | 5 | 5-134954841-C-A | rs920377342 | PCBD2 | 0.0013 | A | | 1.58E-13 | ?+ | 1.576E-13 | | New |
| 333 | 5 | 5-138104941-G-T | rs113532033 | WNT8A, NME5 | 0.0026 | G | | 2.05E-11 | ?+ | 2.048E-11 | | New |
| 334 | 5 | 5-138214143-C-CT | rs1284394562 | CDC23 | 0.0017 | CT | | 4.49E-09 | ?+ | 4.487E-09 | 0.08963 | New |
| 335 | 5 | 5-139242644-C-CCT | rs1311463499 | RNU6-572P, MATR3 | 0.0016 | CCT | | 3.47E-14 | ?+ | 3.473E-14 | | New |
| 335 | 5 | 5-139242645-G-A | rs1355274306 | RNU6-572P, MATR3 | 0.0006 | G | | 5.66E-11 | ?+ | 5.66E-11 | | New |
| 336 | 5 | 5-139345867-TCCGC-T | | PAIP2 | 0.001 | T | | 5.2E-10 | ?+ | 5.196E-10 | | New |
| 336 | 5 | 5-139345874-AACGG-A | | PAIP2 | 0.0011 | AACGG | | 4.45E-11 | ?+ | 4.447E-11 | | New |
| 337 | 5 | 5-139405218-TG-T | | NA | 0.001 | TG | | 1.39E-09 | ?+ | 1.394E-09 | | New |
| 338 | 5 | 5-139533514-GTGTC-G | | NA | 0.0008 | G | | 1.04E-08 | ?+ | 1.042E-08 | | New |
| 338 | 5 | 5-139533522-C-CACTT | | NA | 0.001 | CACTT | | 6.46E-09 | ?+ | 6.464E-09 | | New |
| 338 | 5 | 5-139533525-C-CT | | NA | 0.001 | CT | | 4.62E-09 | ?+ | 4.625E-09 | | New |
| 338 | 5 | 5-139533528-TA-T | | NA | 0.001 | T | | 4.46E-09 | ?+ | 4.463E-09 | | New |
| 339 | 5 | 5-139702543-C-CTT | | NA | 0.0006 | C | | 2.46E-08 | ?+ | 2.464E-08 | | New |
| 340 | 5 | 5-141447364-C-CCTGACCCTGG | | NA | 0.002 | C | | 2.61E-17 | ?+ | 2.611E-17 | | New |

| | | | | | | | | | | | | |
|---|---|---|---|---|---|---|---|---|---|---|---|---|
| 341 | 5 | 5-142336719-C-A | rs1229916131 | SPRY4-AS1 | 0.0024 | A | 0.314347 | 1.89E-24 | -+ | 2.843E-19 | | New |
| 342 | 5 | 5-146234861-C-CCA | | AC091959.3, RBM27 | 0.0008 | C | | 1.2E-10 | ?+ | 1.198E-10 | | New |
| 342 | 5 | 5-146234864-TGC-T | | AC091959.3, RBM27 | 0.0008 | TGC | | 8.21E-09 | ?+ | 8.212E-09 | | New |
| 343 | 5 | 5-146259296-ACT-A | rs1170313679 | AC091959.3, RBM27 | 0.0038 | A | | 1.64E-08 | ?+ | 1.644E-08 | 0.7128 | New |
| 344 | 5 | 5-157864857-G-A | rs1273153391 | CLINT1, RNU2-48P | 0.0005 | A | 0.911079 | 1.45E-09 | ++ | 3.802E-09 | | New |
| 345 | 5 | 5-158747-GT-G | rs1245037762 | PLEKHG4B | 0.0007 | G | | 1.31E-08 | ?+ | 1.312E-08 | | New |
| 346 | 5 | 5-160138763-C-T | rs893629637 | PWWP2A, FABP6 | 0.0012 | C | 0.385039 | 4.15E-12 | ++ | 1.533E-08 | 0.4379 | New |
| 347 | 5 | 5-160140248-GGT-G | | NA | 0.0005 | GGT | | 8.52E-10 | ?+ | 8.524E-10 | | New |
| 348 | 5 | 5-160257333-G-C | rs559986131 | CCNJL | 0.0018 | C | 0.270922 | 3.09E-15 | ++ | 1.21E-10 | | New |
| 348 | 5 | 5-160257343-C-T | rs1307496930 | CCNJL | 0.0014 | T | 0.306034 | 2.67E-16 | ++ | 8.768E-11 | 0.8306 | New |
| 349 | 5 | 5-163628927-C-CAG | | NA | 0.0018 | C | | 1.47E-12 | ?+ | 1.47E-12 | | New |
| 349 | 5 | 5-163628931-ACG-A | | NA | 0.0016 | ACG | | 8.11E-11 | ?+ | 8.109E-11 | | New |
| 350 | 5 | 5-168253265-C-CTACGCACTCATTATTTATA | | NA | 0.0003 | CTACGCACTCATTATTT | | 6.97E-09 | ?+ | 6.968E-09 | | New |
| 351 | 5 | 5-168253657-C-T | rs1281813435 | AC026689.1 | 0.0009 | T | 0.23024 | 3.65E-12 | -+ | 8.022E-09 | 0.9342 | New |
| 351 | 5 | 5-168253667-G-C | rs1274427570 | AC026689.1 | 0.0018 | G | 0.239814 | 5.5E-12 | -+ | 1.322E-09 | | New |
| 351 | 5 | 5-168253671-C-T | rs1305260347 | AC026689.1 | 0.0007 | T | 0.274923 | 6.17E-10 | ++ | 3.35E-10 | | New |
| 352 | 5 | 5-176502362-C-CCT | | FAF2 | 0.0032 | C | | 4.42E-21 | ?+ | 4.418E-21 | 0.2723 | New |
| 353 | 5 | 5-176801068-CTGTGTG-C | | LINC01574, UNC5A | 0.312 | C | | 3.74E-11 | ?+ | 3.736E-11 | | New |
| 354 | 5 | 5-178174212-ACC-A | | NA | 0.0002 | A | | 2.73E-08 | ?+ | 2.733E-08 | | New |

| | | | | | | | | | | | | |
|---|---|---|---|---|---|---|---|---|---|---|---|---|
| 355 | 5 | 5-179544212-GC-G | | NA | 0.0007 | G | | 3.69E-09 | ?+ | 3.693E-09 | | New |
| 356 | 5 | 5-179591395-AG-A | | NA | 0.0015 | A | | 1.35E-12 | ?+ | 1.346E-12 | | New |
| 357 | 5 | 5-179691169-GCACCC-G | | NA | 0.0004 | GCACCC | | 8.07E-09 | ?+ | 8.074E-09 | | Old |
| 358 | 5 | 5-179696510-AGG-A | | NA | 0.0008 | A | | 5.7E-11 | ?+ | 5.696E-11 | | Old |
| 358 | 5 | 5-179696514-G-A | rs1459112573 | CANX | 0.0005 | G | | 1.45E-09 | ?+ | 1.455E-09 | | Old |
| 359 | 5 | 5-179789142-C-CT | | NA | 0.0006 | C | | 2.44E-08 | ?+ | 2.44E-08 | | Old |
| 360 | 5 | 5-181240871-G-A | rs2546403 | RACK1 | 0.1673 | G | | 1.22E-08 | ?+ | 1.218E-08 | 0.1122 | New |
| 361 | 5 | 5-23526774-C-CAA | rs1491369607 | PRDM9 | 0.0029 | CAA | | 4.42E-14 | ?+ | 4.425E-14 | | New |
| 362 | 5 | 5-285364-G-A | rs28463544 | AC021087.5, AHRR, PDCD6 | 0.2502 | A | | 2.78E-08 | ?+ | 2.784E-08 | | New |
| 363 | 5 | 5-31919818-GCCA-G | rs1343350294 | PDZD2 | 0.002 | G | | 9.92E-22 | ?+ | 9.926E-22 | 0.4613 | New |
| 364 | 5 | 5-3323646-G-A | rs374903395 | AC091832.1, LINC01019 | 0.004 | A | | 1.3E-08 | ?+ | 1.302E-08 | | New |
| 364 | 5 | 5-3323650-G-A | rs369041631 | AC091832.1, LINC01019 | 0.0029 | A | | 2.45E-09 | ?+ | 2.449E-09 | | New |
| 365 | 5 | 5-37322460-C-A | rs1744306563 | NUP155 | 0.0002 | A | | 1.24E-08 | ?+ | 1.236E-08 | | New |
| 366 | 5 | 5-37486551-G-T | rs1739878299 | WDR70 | 0.0004 | G | 0.018009 | 9.75E-09 | ++ | 5.517E-10 | | New |
| 367 | 5 | 5-38394491-G-C | rs1443689893 | EGFLAM | 0.0007 | C | | 1.72E-10 | ?+ | 1.722E-10 | 0.32 | New |
| 368 | 5 | 5-42954699-G-C | rs950675668 | AC008945.2, AC008875.3 | 0.0007 | C | | 2.4E-11 | ?+ | 2.402E-11 | | New |
| 368 | 5 | 5-42954706-G-A | rs1740607393 | AC008945.2, AC008875.3 | 0.0009 | G | | 1.21E-11 | ?+ | 1.21E-11 | | New |

| | | | | | | | | | | | | |
|---|---|---|---|---|---|---|---|---|---|---|---|---|
| 368 | 5 | 5-42954730-G-C | rs949204282 | AC008945.2, AC008875.3 | 0.0008 | G | 0.326477 | 2.98E-14 | ++ | 9.691E-14 | | New |
| 368 | 5 | 5-42954736-G-A | rs1046138470 | AC008945.2, AC008875.3 | 0.0008 | G | | 2.85E-09 | ?+ | 2.855E-09 | | New |
| 369 | 5 | 5-42982044-C-A | rs13159363 | AC008945.2, AC008875.3 | 0.0028 | C | 0.181836 | 1.81E-10 | ++ | 2.689E-08 | | New |
| 370 | 5 | 5-43345773-G-A | rs1334993567 | HMGCS1, CCL28 | 0.0004 | A | | 2.19E-08 | ?+ | 2.189E-08 | | New |
| 371 | 5 | 5-46397240-C-T | rs1164552231 | AC122694.1, NONE | 0.0004 | T | 0.563522 | 3.49E-09 | -+ | 4.022E-08 | 0.3139 | New |
| 372 | 5 | 5-55688573-G-T | rs1179471099 | SLC38A9 | 0.0017 | T | | 1.6E-09 | ?+ | 1.6E-09 | | New |
| 373 | 5 | 5-55987899-GAGAT-G | | NA | 0.0003 | GAGAT | | 2.42E-10 | ?+ | 2.42E-10 | | New |
| 374 | 5 | 5-57149535-C-T | rs959586683 | MIER3, RF00019 | 0.0003 | T | | 1.71E-08 | ?+ | 1.709E-08 | 0.7941 | New |
| 375 | 5 | 5-5730755-TA-T | rs1246009187 | AC026736.1, AC010266.2 | 0.0019 | T | | 1.45E-08 | ?+ | 1.451E-08 | | New |
| 376 | 5 | 5-61953687-G-A | rs1400393603 | AC008877.1, RN7SKP157 | 0.0003 | G | | 2.5E-11 | ?+ | 2.502E-11 | | Old |
| 377 | 5 | 5-63078868-GTC-G | | NA | 0.0007 | GTC | | 3.07E-09 | ?+ | 3.066E-09 | | New |
| 378 | 5 | 5-65634189-GTTAGCCAGGATA-G | | NA | 0.0006 | G | | 1.35E-12 | ?+ | 1.353E-12 | | New |
| 379 | 5 | 5-69337801-AT-A | | NA | 0.0009 | AT | | 3.57E-09 | ?+ | 3.565E-09 | | New |
| 380 | 5 | 5-71297967-CA-C | | NA | 0.0946 | CA | | 2.45E-09 | ?+ | 2.452E-09 | | New |
| 381 | 5 | 5-76509558-ATTTT-A | | NA | 0.001 | A | | 5.1E-12 | ?+ | 5.099E-12 | | New |
| 382 | 5 | 5-80507561-G-A | rs761833241 | FAM151B | 0.0004 | G | 0.233303 | 3.02E-13 | ++ | 1.258E-09 | | New |

| | | | | | | | | | | | | |
|---|---|---|---|---|---|---|---|---|---|---|---|---|
| 383 | 5 | 5-82726070-TCCTGGCTAACAC-T | | AC026782.2, AC008885.2 | 0.0008 | TCCTGGCTAACAC | | 1.93E-09 | ?+ | 1.932E-09 | | New |
| 384 | 5 | 5-82972528-C-T | rs1743330765 | AC027338.2, AC027338.1 | 0.0003 | T | 0.044276 | 1.38E-09 | ++ | 1.751E-10 | | New |
| 385 | 5 | 5-82973288-G-T | rs974742724 | AC027338.2, AC027338.1 | 0.0008 | T | 0.892244 | 5.92E-14 | -+ | 7.789E-13 | | New |
| 386 | 5 | 5-93000815-AC-A | | NA | 0.0012 | AC | | 4.75E-10 | ?+ | 4.752E-10 | | New |
| 386 | 5 | 5-93000816-G-T | | LINC02058, AC012625.1 | 0.0008 | G | | 2.42E-10 | ?+ | 2.425E-10 | | New |
| 387 | 6 | 6-101494698-AGGCG-A | | GRIK2 | 0.001 | AGGCG | | 2.2E-08 | ?+ | 0.000000022 | | New |
| 388 | 6 | 6-103320550-GGAGGCT-G | rs1445466240 | AL357139.2, RF00438 | 0.0012 | G | 0.450862 | 8.76E-13 | -+ | 1.478E-08 | | New |
| 389 | 6 | 6-104062626-C-T | rs1168094839 | AL590608.1, AL357522.1 | 0.0035 | T | | 8.66E-11 | ?- | 8.663E-11 | | New |
| 390 | 6 | 6-106480620-TCG-T | rs1352922508 | CRYBG1 | 0.0012 | T | | 3.7E-09 | ?+ | 3.7E-09 | 0.001142 | New |
| 390 | 6 | 6-106480624-ATG-A | rs1288116823 | CRYBG1 | 0.0012 | ATG | | 2.94E-08 | ?+ | 2.941E-08 | | New |
| 391 | 6 | 6-10786446-G-A | rs868538362 | AL024498.2, MAK, TMEM14B | 0.0009 | G | | 8.4E-09 | ?+ | 8.398E-09 | | New |
| 392 | 6 | 6-10810629-ACGCC-A | | NA | 0.0002 | ACGCC | | 1.01E-08 | ?+ | 1.012E-08 | | New |
| 393 | 6 | 6-118716962-ATG-A | | CEP85L, MCM9 | 0.001 | A | | 3.64E-15 | ?+ | 3.639E-15 | | New |
| 394 | 6 | 6-12042382-C-CGG | | NA | 0.0084 | CGG | | 1.46E-10 | ?+ | 1.456E-10 | | New |

| | | | | | | | | | | | | |
|---|---|---|---|---|---|---|---|---|---|---|---|---|
| 395 | 6 | 6-122138733-G-T | rs1369563818 | RNU2-8P, RNU1-18P | 0.0023 | T | 0.195508 | 2.96E-14 | -+ | 1.937E-11 | | New |
| 396 | 6 | 6-122139207-G-T | rs1776228526 | RNU2-8P, RNU1-18P | 0.0012 | G | | 1.19E-09 | ?+ | 1.191E-09 | | New |
| 397 | 6 | 6-12840142-TC-T | | NA | 0.0012 | T | | 2E-14 | ?+ | 1.997E-14 | | New |
| 398 | 6 | 6-139484262-C-CAT | | NA | 0.0005 | C | | 7.68E-13 | ?+ | 7.686E-13 | | New |
| 398 | 6 | 6-139484265-GGC-G | | NA | 0.0005 | GGC | | 3.15E-13 | ?+ | 3.149E-13 | | New |
| 399 | 6 | 6-143890911-C-CTT | | NA | 0.0012 | CTT | | 4.73E-09 | ?+ | 4.732E-09 | | New |
| 400 | 6 | 6-14817518-G-A | rs532415821 | AL138720.1 | 0.0012 | G | 0.254201 | 5.1E-12 | -+ | 1.383E-10 | 0.4535 | New |
| 400 | 6 | 6-14817529-GCA-G | rs759633872 | AL138720.1 | 0.0011 | GCA | | 9.18E-14 | ?+ | 9.181E-14 | 0.7234 | New |
| 400 | 6 | 6-14817530-AGG-A | rs1249641444 | AL138720.1 | 0.0011 | A | | 7.49E-12 | ?+ | 7.494E-12 | 0.7215 | New |
| 401 | 6 | 6-148303578-C-CTT | | NA | 0.0017 | C | | 1.08E-18 | ?+ | 1.085E-18 | | New |
| 402 | 6 | 6-14989159-C-CGTG | | NA | 0.001 | C | | 9.18E-10 | ?+ | 9.176E-10 | | New |
| 403 | 6 | 6-151261625-G-C | | AKAP12 | 0.0007 | G | 0.440187 | 2.64E-09 | ++ | 0.00000001 | | New |
| 404 | 6 | 6-158692007-C-A | | SYTL3 | 0.0001 | A | 0.839753 | 0.336519 | ++ | 1.24E-305 | | Old |
| 405 | 6 | 6-16267227-C-T | rs1293514950 | GMPR | 0.0018 | C | 0.131139 | 4.15E-12 | ++ | 5.466E-10 | | New |
| 405 | 6 | 6-16267232-C-T | rs1218372872 | GMPR | 0.0021 | C | 0.131162 | 1.08E-12 | ++ | 8.424E-11 | | New |
| 406 | 6 | 6-165408947-C-CCT | rs1428838377 | PDE10A | 0.0023 | CCT | | 4.97E-09 | ?+ | 4.97E-09 | 0.1019 | New |
| 407 | 6 | 6-168892201-GGGGCAGCCCGAGACTCAGGCTCAGGTTTTCCACCTGGACCTTGGAAAGAGACATCACACAGCTCAGCGTCCAGGAGGC-G | rs1562494598 | AL513210.2, AL513210.1 | 0.2145 | G | | 1.98E-14 | ?+ | 1.981E-14 | | New |
| 408 | 6 | 6-170054847-TC-T | rs202108135 | AL049612.1, AL603783.1 | 0.0041 | T | 0.941482 | 1.08E-14 | ++ | 1.256E-14 | | New |

| | | | | | | | | | | | | |
|---|---|---|---|---|---|---|---|---|---|---|---|---|
| 409 | 6 | 6-21002980-C-CT | rs1295446575 | CDKAL1 | 0.0029 | CT | | 4.83E-18 | ?+ | 4.832E-18 | 0.5454 | New |
| 410 | 6 | 6-25902999-AAG-A | | SLC17A3, SLC17A2 | 0.0002 | A | | 4.35E-08 | ?+ | 4.348E-08 | | New |
| 411 | 6 | 6-26311267-C-CAACAACACA | rs1201608045 | HIST1H4H, AL021917.1 | 0.0007 | CAACAACACA | | 3E-08 | ?+ | 3.005E-08 | | New |
| 412 | 6 | 6-27577093-C-CTT | rs1348229080 | AL021918.3, RNU6-471P | 0.0019 | CTT | | 2.92E-10 | ?+ | 2.92E-10 | | Old |
| 413 | 6 | 6-30621247-C-CAAA | | MRPS18B | 0.0008 | CAAA | | 1.18E-12 | ?+ | 1.178E-12 | | New |
| 414 | 6 | 6-30753762-GCA-G | | NA | 0.001 | G | | 3.22E-09 | ?+ | 3.221E-09 | | New |
| 414 | 6 | 6-30753765-AGG-A | | HCG20 | 0.0011 | AGG | | 4.67E-09 | ?+ | 4.671E-09 | | New |
| 415 | 6 | 6-31808780-C-CCT | rs1315875949 | HSPA1L | 0.0015 | C | | 7.16E-13 | ?+ | 7.159E-13 | 0.7632 | Old |
| 416 | 6 | 6-32381995-C-T | rs9268455 | TSBP1-AS1 | 0.2103 | T | 1.4E-08 | 0.017077 | -- | 1.731E-09 | 0.3865 | Old |
| 416 | 6 | 6-32404138-C-T | rs3793127 | TSBP1-AS1 | 0.2104 | T | 1.52E-08 | 0.01615 | -- | 1.731E-09 | 0.3749 | Old |
| 416 | 6 | 6-32413666-T-A | rs9268522 | TSBP1-AS1, HLA-DRA | 0.2176 | T | 2.82E-08 | 0.024758 | -- | 5.275E-09 | 0.2559 | Old |
| 416 | 6 | 6-32438044-C-T | rs9268627 | TSBP1-AS1, HLA-DRA | 0.2136 | C | 4E-08 | 0.024994 | -- | 7.009E-09 | 0.7249 | Old |
| 416 | 6 | 6-32461376-G-A | rs9268844 | HLA-DRA, HLA-DRB5 | 0.2038 | G | 4.46E-09 | 0.01651 | -- | 5.871E-10 | | Old |
| 416 | 6 | 6-32602534-G-T | rs34039593 | HLA-DRB1, HLA-DQA1 | 0.1746 | G | 4.33E-09 | 0.073629 | -- | 4.341E-09 | 0.9873 | Old |
| 416 | 6 | 6-32602640-C-A | rs2647062 | HLA-DRB1, HLA-DQA1 | 0.1746 | C | 3.72E-09 | 0.072099 | -- | 3.706E-09 | 0.9785 | Old |
| 416 | 6 | 6-32603181-G-T | rs679242 | HLA-DRB1, HLA-DQA1 | 0.1745 | T | 2E-09 | 0.070655 | -- | 2.118E-09 | 0.9081 | Old |

| | | | | | | | | | | | | |
|---|---|---|---|---|---|---|---|---|---|---|---|---|
| 416 | 6 | 6-32603333-G-A | rs2760990 | HLA-DRB1, HLA-DQA1 | 0.175 | A | 2.13E-09 | 0.070943 | -- | 2.285E-09 | 0.9908 | Old |
| 416 | 6 | 6-32605402-G-A | rs601020 | HLA-DRB1, HLA-DQA1 | 0.2378 | A | 3.2E-08 | 0.085603 | -- | 3.416E-08 | 0.2036 | Old |
| 416 | 6 | 6-32605488-G-T | rs601148 | HLA-DRB1, HLA-DQA1 | 0.1863 | T | 2.73E-08 | 0.059196 | -- | 1.574E-08 | 0.3496 | Old |
| 416 | 6 | 6-32605638-G-A | rs601945 | HLA-DRB1, HLA-DQA1 | 0.1717 | G | 4.58E-09 | 0.047919 | -- | 2.277E-09 | 0.9857 | Old |
| 416 | 6 | 6-32606826-G-A | rs617578 | HLA-DRB1, HLA-DQA1 | 0.1767 | A | 2.61E-09 | 0.082916 | -- | 3.545E-09 | 0.8026 | Old |
| 416 | 6 | 6-32607091-C-T | rs7760841 | HLA-DRB1, HLA-DQA1 | 0.1787 | T | 7.31E-09 | 0.062939 | -- | 5.661E-09 | 0.9174 | Old |
| 416 | 6 | 6-32607119-G-A | rs7770010 | HLA-DRB1, HLA-DQA1 | 0.178 | A | 1.1E-08 | 0.062299 | -- | 7.834E-09 | 0.9071 | Old |
| 416 | 6 | 6-32609269-C-T | rs562289 | HLA-DRB1, HLA-DQA1 | 0.22 | T | 7.54E-09 | 0.057127 | -- | 5.861E-09 | | Old |
| 416 | 6 | 6-32610196-G-T | rs532965 | HLA-DRB1, HLA-DQA1 | 0.179 | G | 5.39E-09 | 0.062384 | -- | 4.327E-09 | 0.892 | Old |
| 416 | 6 | 6-32610995-C-A | rs504594 | HLA-DRB1, HLA-DQA1 | 0.1758 | A | 7.87E-09 | 0.042427 | -- | 3.231E-09 | 0.9585 | Old |

| | | | | | | | | | | | | |
|---|---|---|---|---|---|---|---|---|---|---|---|---|
| 416 | 6 | 6-32612814-G-C | rs7449585 | HLA-DRB1, HLA-DQA1 | 0.2611 | C | 3.66E-08 | 0.02221 | -- | 6.112E-09 | | Old |
| 416 | 6 | 6-32612840-T-A | rs3997872 | HLA-DRB1, HLA-DQA1 | 0.1759 | A | 8.7E-09 | 0.045107 | -- | 3.838E-09 | 0.9916 | Old |
| 416 | 6 | 6-32612880-C-T | rs2395516 | HLA-DRB1, HLA-DQA1 | 0.306 | C | 1.42E-08 | 0.019816 | -- | 2.463E-09 | 0.6616 | Old |
| 416 | 6 | 6-32613231-C-T | rs3129747 | HLA-DRB1, HLA-DQA1 | 0.3061 | C | 1.36E-08 | 0.020371 | -- | 2.475E-09 | 0.6771 | Old |
| 416 | 6 | 6-32613238-T-A | rs3129748 | HLA-DRB1, HLA-DQA1 | 0.3061 | A | 1.36E-08 | 0.020571 | -- | 2.507E-09 | 0.6724 | Old |
| 416 | 6 | 6-32614412-C-A | rs3129751 | HLA-DRB1, HLA-DQA1 | 0.1759 | C | 7.98E-09 | 0.042248 | -- | 3.252E-09 | 0.9945 | Old |
| 416 | 6 | 6-32614873-G-C | rs3104413 | HLA-DRB1, HLA-DQA1 | 0.1782 | G | 7.05E-09 | 0.063393 | -- | 5.794E-09 | 0.889 | Old |
| 416 | 6 | 6-32615250-G-C | rs3129753 | HLA-DRB1, HLA-DQA1 | 0.1791 | C | 4.01E-09 | 0.066203 | -- | 3.715E-09 | 0.902 | Old |
| 416 | 6 | 6-32615369-C-T | rs4959105 | HLA-DRB1, HLA-DQA1 | 0.3179 | T | 5.18E-09 | 0.011798 | -- | 5.425E-10 | 0.8795 | Old |
| 416 | 6 | 6-32615429-G-A | rs6941395 | HLA-DRB1, HLA-DQA1 | 0.1791 | A | 5.09E-09 | 0.063042 | -- | 4.201E-09 | 0.9181 | Old |

| | | | | | | | | | | | | |
|---|---|---|---|---|---|---|---|---|---|---|---|---|
| 416 | 6 | 6-32616142-C-T | rs33964890 | HLA-DRB1, HLA-DQA1 | 0.2737 | C | 1.48E-08 | 0.01524 | -- | 1.805E-09 | 0.6769 | Old |
| 416 | 6 | 6-32616202-G-A | rs33915496 | HLA-DRB1, HLA-DQA1 | 0.318 | G | 6.01E-09 | 0.012006 | -- | 6.305E-10 | 0.8365 | Old |
| 416 | 6 | 6-32616536-G-C | rs33932178 | HLA-DRB1, HLA-DQA1 | 0.319 | G | 7.01E-09 | 0.018483 | -- | 1.22E-09 | | Old |
| 416 | 6 | 6-32617117-G-T | rs508318 | HLA-DRB1, HLA-DQA1 | 0.1785 | T | 5.27E-09 | 0.062985 | -- | 4.304E-09 | | Old |
| 416 | 6 | 6-32622864-G-T | rs9271580 | HLA-DRB1, HLA-DQA1 | 0.1909 | G | 2.07E-08 | 0.052096 | -- | 1.02E-08 | | Old |
| 416 | 6 | 6-32623007-TACAG-T | rs5875381 | HLA-DRB1, HLA-DQA1 | 0.1952 | TACAG | 1.77E-08 | 0.081176 | -- | 1.613E-08 | 0.4352 | Old |
| 416 | 6 | 6-32623436-G-A | rs9271594 | HLA-DRB1, HLA-DQA1 | 0.1806 | G | 4.48E-09 | 0.061802 | -- | 3.597E-09 | 0.914 | Old |
| 416 | 6 | 6-32629245-G-A | rs3129769 | HLA-DQA1 | 0.1788 | A | 7.51E-09 | 0.059126 | -- | 5.281E-09 | 0.9376 | Old |
| 416 | 6 | 6-32631077-G-A | rs3104381 | HLA-DQA1 | 0.1822 | A | 1.97E-08 | 0.058436 | -- | 1.104E-08 | 0.8642 | Old |
| 416 | 6 | 6-32631386-C-T | rs3104378 | HLA-DQA1 | 0.1794 | T | 6.54E-09 | 0.058307 | -- | 4.625E-09 | 0.9395 | Old |
| 416 | 6 | 6-32634360-G-C | rs3104371 | HLA-DQA1 | 0.1917 | G | 1.86E-08 | 0.048707 | -- | 8.514E-09 | 0.5671 | Old |
| 416 | 6 | 6-32634784-C-T | rs3104368 | HLA-DQA1 | 0.1788 | T | 8.56E-09 | 0.058081 | -- | 5.735E-09 | | Old |
| 416 | 6 | 6-32636679-G-C | rs9272353 | HLA-DQA1 | 0.1921 | C | 1.85E-08 | 0.049281 | -- | 8.559E-09 | 0.6229 | Old |
| 416 | 6 | 6-32636808-T-A | rs9272363 | HLA-DQA1 | 0.1952 | T | 1.61E-08 | 0.036747 | -- | 4.715E-09 | 0.4868 | Old |

| | | | | | | | | | | | | |
|---|---|---|---|---|---|---|---|---|---|---|---|---|
| 416 | 6 | 6-32714360-A-G | rs3957148 | HLA-DQB1, AL662789.1 | 0.1073 | A | 1.82E-07 | 0.032931 | -- | 4.744E-08 | 0.204 | Old |
| 417 | 6 | 6-34044161-TAC-T | | GRM4 | 0.2762 | TAC | | 3.09E-08 | ?+ | 3.094E-08 | | New |
| 418 | 6 | 6-34212899-TTTTG-T | rs1177909976 | GRM4, HMGA1 | 0.0039 | T | | 2.97E-29 | ?+ | 2.966E-29 | 0.7905 | New |
| 419 | 6 | 6-34702668-G-A | rs931906248 | AL451165.2, SNRPC | 0.0013 | A | 0.167082 | 6.63E-14 | -+ | 6.579E-09 | 0.422 | New |
| 420 | 6 | 6-38428267-G-C | rs78128943 | BTBD9 | 0.0006 | C | 2.93E-05 | 6.37E-05 | ++ | 3.376E-08 | | New |
| 421 | 6 | 6-40659693-T-C | | LRFN2, AL583854.1 | 0 | T | 0.057073 | 0.311985 | ++ | 1.41E-305 | | Old |
| 422 | 6 | 6-41604777-C-T | rs1397842965 | FOXP4, MDFI | 0.0014 | C | 0.948278 | 9.29E-13 | ++ | 2.135E-08 | 0.07667 | Old |
| 423 | 6 | 6-41918284-ACT-A | rs1491105501 | MED20 | 0.0037 | A | 0.755137 | 7.45E-13 | -+ | 1.406E-10 | 0.8851 | New |
| 424 | 6 | 6-42018709-T-A | rs1483765759 | CCND3 | 0.0012 | A | 0.720039 | 1.82E-14 | -+ | 1.668E-10 | | New |
| 425 | 6 | 6-43329675-A-G | rs7766978 | ZNF318 | 0.0187 | G | 0.995788 | 1.76E-12 | ++ | 1.646E-10 | 0.1203 | New |
| 426 | 6 | 6-43534819-GTA-G | | NA | 0.0009 | GTA | | 1.05E-08 | ?+ | 1.047E-08 | | New |
| 427 | 6 | 6-43738216-C-T | rs112260099 | AL136131.2 | 0.0227 | T | | 1.24E-16 | ?+ | 1.24E-16 | | New |
| 428 | 6 | 6-62897569-G-A | | KHDRBS2, FKBP1C | 0.0016 | A | 0.306421 | 0.620998 | ++ | 2.05E-202 | 0.09527 | New |
| 429 | 6 | 6-66722870-G-T | | EYS, RNU7-66P | 0.0005 | G | | 4.45E-10 | ?+ | 4.452E-10 | | New |
| 430 | 6 | 6-6716029-C-CTT | | NA | 0.0014 | CTT | | 1.25E-10 | ?+ | 1.252E-10 | | New |
| 431 | 6 | 6-7055607-G-A | rs62393574 | AL139390.1, RREB1 | 0.2366 | A | | 2.58E-08 | ?+ | 2.578E-08 | 0.3832 | New |
| 432 | 6 | 6-73319458-C-CAA | | NA | 0.0029 | C | | 2.65E-13 | ?+ | 2.647E-13 | | New |
| 432 | 6 | 6-73319481-G-A | rs1283541332 | KHDC1, DPPA5 | 0.0007 | A | 0.197459 | 1.07E-09 | -+ | 4.822E-08 | | New |
| 433 | 6 | 6-7382355-TG-T | rs1332897454 | CAGE1 | 0.0017 | TG | | 2.25E-10 | ?+ | 2.253E-10 | 0.3466 | New |
| 434 | 6 | 6-77499064-C-T | | HTR1B, MEI4 | 0.0002 | T | 0.423388 | 0.29787 | -- | 2.92E-131 | 0.2857 | New |
| 435 | 6 | 6-88001571-GT-G | | RN7SL183P, AL136096.1 | 0.0004 | GT | | 1.89E-08 | ?+ | 1.887E-08 | | New |

| | | | | | | | | | | | | |
|---|---|---|---|---|---|---|---|---|---|---|---|---|
| 436 | 7 | 7-100672357-C-CCT | rs1270383603 | ACTL6B, GNB2 | 0.0043 | C | | 6.27E-18 | ?+ | 6.27E-18 | 0.1064 | Old |
| 437 | 7 | 7-101035140-T-A | rs10265276 | MUC17 | 0.1869 | A | | 3.81E-09 | ?+ | 3.814E-09 | 0.5325 | Old |
| 438 | 7 | 7-101284191-GAT-G | | NA | 0.0003 | G | | 1.02E-08 | ?+ | 1.017E-08 | | New |
| 438 | 7 | 7-101284194-GCA-G | | NA | 0.0003 | GCA | | 2.82E-09 | ?+ | 2.818E-09 | | New |
| 439 | 7 | 7-101788981-T-A | rs891280128 | MYL10, CUX1 | 0.0018 | T | 0.868141 | 1.49E-16 | -+ | 4.866E-12 | | New |
| 440 | 7 | 7-102026581-AG-A | | NA | 0.0005 | A | | 3.01E-09 | ?+ | 3.007E-09 | | New |
| 441 | 7 | 7-102920935-C-CCT | | FBXL13, LRRC17 | 0.0013 | CCT | | 2.05E-08 | ?+ | 2.054E-08 | | New |
| 442 | 7 | 7-105596291-C-T | rs947078247 | EFCAB10 | 0.0026 | T | 0.842782 | 8.52E-17 | -+ | 2.646E-08 | 0.9001 | New |
| 443 | 7 | 7-1066816-GCACCCCAGAGGTGAGGGTTTGGGGCACAGTCTGTTGGCGGAGGCAGGAGTA-G | rs1563072994 | C7orf50 | 0.0234 | G | | 1.01E-08 | ?+ | 1.006E-08 | | New |
| 444 | 7 | 7-111925911-C-A | rs574995791 | DOCK4 | 0.0017 | A | 0.419217 | 4.93E-12 | ++ | 1.222E-09 | 0.9477 | New |
| 444 | 7 | 7-111925913-C-A | rs535943537 | DOCK4 | 0.0016 | A | 0.3947 | 7.64E-13 | ++ | 3.086E-10 | 0.6831 | New |
| 445 | 7 | 7-116919680-ATG-A | rs1414168601 | CAPZA2 | 0.0054 | ATG | | 4.06E-10 | ?+ | 4.061E-10 | | New |
| 446 | 7 | 7-126391976-G-A | rs1045685494 | AC000372.1 | 0.0019 | G | 0.558351 | 7.49E-17 | -+ | 3.73E-08 | | New |
| 447 | 7 | 7-128552494-G-T | | AC090114.2, AC108010.1 | 0.0007 | G | | 8.05E-11 | ?+ | 8.054E-11 | | New |
| 448 | 7 | 7-129358248-C-T | rs891954249 | AHCYL2 | 0.0017 | T | 0.676651 | 2.43E-10 | ++ | 4.675E-09 | | New |
| 449 | 7 | 7-130087903-GGT-G | rs1235961115 | KLHDC10 | 0.0018 | G | | 1.15E-09 | ?+ | 1.149E-09 | 0.1943 | New |
| 449 | 7 | 7-130087910-GCA-G | rs1347602558 | KLHDC10 | 0.0015 | GCA | | 2.52E-08 | ?+ | 2.523E-08 | | New |

| | | | | | | | | | | | | |
|---|---|---|---|---|---|---|---|---|---|---|---|---|
| 450 | 7 | 7-13820977-C-T | rs1421858724 | AC011287.1, AC005019.2 | 0.0008 | T | 0.881387 | 3.74E-08 | ++ | 4.631E-08 | | New |
| 450 | 7 | 7-13820985-C-T | rs1412811394 | AC011287.1, AC005019.2 | 0.001 | T | | 3.54E-14 | ?+ | 3.541E-14 | | New |
| 451 | 7 | 7-138674703-AAT-A | | NA | 0.0006 | AAT | | 1.63E-08 | ?+ | 1.634E-08 | | New |
| 452 | 7 | 7-140480517-GA-G | rs1455936159 | MKRN1, DENND2A | 0.0007 | G | | 2.94E-10 | ?+ | 2.939E-10 | | New |
| 453 | 7 | 7-140498699-C-A | rs918393304 | MKRN1, DENND2A | 0.0018 | A | | 1.38E-21 | ?+ | 1.377E-21 | | New |
| 454 | 7 | 7-140619733-C-CAA | | NA | 0.0008 | C | | 2.11E-11 | ?+ | 2.114E-11 | | New |
| 455 | 7 | 7-140944263-AT-A | rs1292982927 | BRAF, MRPS33 | 0.003 | AT | 0.994039 | 2.19E-17 | -+ | 2.264E-15 | | New |
| 456 | 7 | 7-140961641-C-CAA | | NA | 0.0008 | CAA | | 5.77E-12 | ?+ | 5.765E-12 | | New |
| 457 | 7 | 7-148956743-AGG-A | | NA | 0.0005 | A | | 7.74E-10 | ?+ | 7.74E-10 | | New |
| 458 | 7 | 7-148989806-T-A | | GHET1 | 0.001 | A | 0.051067 | 2.77E-15 | ++ | 4.624E-16 | | New |
| 459 | 7 | 7-149095930-C-CCT | | NA | 0.0013 | CCT | | 2.16E-12 | ?+ | 2.155E-12 | | New |
| 460 | 7 | 7-149904512-C-CA | rs1159657792 | AC092681.3, AC092681.2 | 0.0022 | CA | | 3.53E-09 | ?+ | 3.53E-09 | | New |
| 461 | 7 | 7-152402297-C-CAATCATA | rs71260329 | KMT2C | 0.0025 | CAATCATA | | 5.98E-13 | ?+ | 5.976E-13 | | New |
| 462 | 7 | 7-152546811-C-CA | rs1165639521 | LINC01003, RNA5SP250 | 0.0014 | C | | 3.87E-09 | ?+ | 3.865E-09 | | New |
| 463 | 7 | 7-152617758-C-T | rs1385078687 | RF00568, XRCC2 | 0.0011 | C | 0.492947 | 1.49E-09 | ++ | 1.378E-09 | 0.3217 | New |
| 464 | 7 | 7-155134673-C-CCT | | NA | 0.0011 | CCT | | 2.44E-08 | ?+ | 2.441E-08 | | Old |
| 465 | 7 | 7-157725208-GC-G | rs746693788 | PTPRN2 | 0.0097 | G | | 3.01E-09 | ?+ | 3.012E-09 | | New |

| | | | | | | | | | | | | |
|---|---|---|---|---|---|---|---|---|---|---|---|---|
| 466 | 7 | 7-158045747-C-A | rs1585262241 | PTPRN2 | 0.0003 | A | | 1.4E-08 | ?+ | 1.403E-08 | | New |
| 467 | 7 | 7-158084639-ACGACACTCATCCACATCCT-A | rs1387462349 | PTPRN2 | 0.0017 | A | | 4.38E-09 | ?+ | 4.376E-09 | | New |
| 468 | 7 | 7-158319527-T-TCA | | PTPRN2 | 0.1934 | TCA | | 3.16E-08 | ?+ | 3.164E-08 | | New |
| 469 | 7 | 7-158463746-C-CA | rs1349126896 | PTPRN2 | 0.0062 | C | | 7.97E-15 | ?+ | 7.973E-15 | 0.01274 | New |
| 470 | 7 | 7-1765054-C-T | | ELFN1, MAD1L1 | 0.0005 | C | 0.9368 | 2.75E-12 | ++ | 1.12E-09 | | New |
| 471 | 7 | 7-222020-C-CCG | rs1350645584 | FAM20C | 0.0122 | C | | 3.95E-24 | ?+ | 3.948E-24 | | New |
| 472 | 7 | 7-2296752-G-T | rs1208805700 | SNX8 | 0.0011 | G | 0.029002 | 1.1E-08 | ++ | 1.865E-09 | | New |
| 473 | 7 | 7-2455679-G-C | rs1778672500 | CHST12, GRIFIN | 0.0007 | G | 0.330179 | 3.72E-10 | ++ | 2.95E-10 | | New |
| 473 | 7 | 7-2455697-G-C | rs939252020 | CHST12, GRIFIN | 0.0016 | G | 0.203285 | 2.68E-16 | -+ | 1.904E-11 | | New |
| 474 | 7 | 7-26252136-C-CCT | | NA | 0.0003 | CCT | | 8.19E-09 | ?+ | 8.192E-09 | | New |
| 475 | 7 | 7-3186156-G-T | rs112236497 | AC073316.1, AC073316.2 | 0.0016 | T | 0.020147 | 1.81E-14 | ++ | 3.539E-13 | | New |
| 476 | 7 | 7-32888710-ATTACTTG-A | | NA | 0.0002 | ATTACTTG | | 2.77E-08 | ?+ | 2.772E-08 | | New |
| 477 | 7 | 7-4276876-GGA-G | | NA | 0.0006 | G | | 4.14E-09 | ?+ | 4.142E-09 | | New |
| 478 | 7 | 7-47554393-GGGT-G | | NA | 0.0004 | G | | 2.88E-09 | ?+ | 2.882E-09 | | New |
| 479 | 7 | 7-47925128-TGGAAAAAAAAA-T | | NA | 0.0004 | T | 0.208106 | 2.58E-09 | -+ | 3.662E-08 | | New |
| 480 | 7 | 7-48002361-C-T | rs1789406952 | SUN3 | 0.0008 | C | | 4.68E-09 | ?+ | 4.68E-09 | | New |
| 481 | 7 | 7-4896984-G-C | rs1784729937 | RADIL, MMD2 | 0.0001 | C | | 3.14E-08 | ?+ | 3.14E-08 | | New |
| 482 | 7 | 7-49633385-C-CACAG | rs1232901130 | AC092448.1, AC093775.1 | 0.0008 | CACAG | 0.008806 | 1.37E-08 | ++ | 2.713E-09 | | New |
| 483 | 7 | 7-51057333-TAC-T | | COBL | 0.3701 | T | | 2.31E-08 | ?+ | 2.306E-08 | | New |

| | | | | | | | | | | | | |
|---|---|---|---|---|---|---|---|---|---|---|---|---|
| 484 | 7 | 7-5140818-G-A | rs1388898746 | RBAKDN, RNU6-215P | 0.0012 | A | 0.699399 | 6.03E-12 | ++ | 2.322E-10 | 0.5743 | New |
| 485 | 7 | 7-5564808-G-A | rs1206340214 | ACTB, FSCN1 | 0.0015 | G | | 5.42E-13 | ?+ | 5.419E-13 | 0.574 | New |
| 486 | 7 | 7-55835419-G-T | rs774698563 | SEPT14 | 0.0021 | G | 0.531286 | 1.72E-19 | ++ | 2.789E-17 | 0.6825 | New |
| 487 | 7 | 7-57783552-C-T | rs143180426 | ZNF716, NONE | 0.0046 | T | 0.044829 | 4.03E-11 | -- | 1.745E-11 | | New |
| 488 | 7 | 7-58060060-TTTG-T | | NA | 0.004 | T | | 4.43E-14 | ?+ | 4.429E-14 | | New |
| 488 | 7 | 7-58060068-TTC-T | | NA | 0.0032 | T | | 3.24E-11 | ?+ | 3.237E-11 | 0.5622 | New |
| 489 | 7 | 7-58060792-TTCTATTC-T | | NA | 0.0043 | TTCTATTC | | 4.32E-30 | ?+ | 4.324E-30 | | New |
| 490 | 7 | 7-5856871-AT-A | | NA | 0.0011 | AT | | 1.63E-13 | ?+ | 1.629E-13 | | New |
| 491 | 7 | 7-62337609-T-A | rs200628965 | NONE, RNU6-417P | 0.0003 | A | | 8.39E-09 | ?+ | 8.391E-09 | | New |
| 492 | 7 | 7-62430395-T-A | rs879025335 | NONE, RNU6-417P | 0.004 | A | 0.75691 | 9.11E-10 | -+ | 4.446E-08 | | New |
| 493 | 7 | 7-65095078-A-AT | | SNORA15B-1, AC104073.4 | 0.0383 | A | | 5.47E-12 | ?+ | 5.466E-12 | | New |
| 494 | 7 | 7-65711737-GAC-G | | NA | 0.0006 | G | | 9.15E-14 | ?+ | 9.146E-14 | | New |
| 495 | 7 | 7-66026675-TAA-T | rs1402152841 | GUSB, ASL | 0.0018 | T | | 6.51E-13 | ?+ | 6.512E-13 | | New |
| 496 | 7 | 7-66707015-GACCT-G | rs1185379301 | AC027644.4, RABGEF1 | 0.0026 | GACCT | | 6.81E-09 | ?+ | 6.81E-09 | 0.7985 | New |
| 497 | 7 | 7-67109054-ATT-A | | NA | 0.0013 | ATT | 0.83812 | 1.55E-13 | ++ | 9.376E-12 | | New |
| 498 | 7 | 7-68108631-C-CGACACAGT | | NA | 0.0007 | CGACACAGT | | 1.2E-12 | ?+ | 1.201E-12 | | New |
| 499 | 7 | 7-69807598-C-A | | AUTS2 | 0.0004 | A | | 6.1E-11 | ?+ | 6.094E-11 | | New |
| 499 | 7 | 7-69807601-C-CCT | | NA | 0.0011 | C | | 4.66E-10 | ?+ | 4.66E-10 | | New |
| 500 | 7 | 7-71010361-TGG-T | | NA | 0.0004 | T | | 1.24E-09 | ?+ | 1.243E-09 | | New |
| 501 | 7 | 7-72703259-C-CAA | | NA | 0.0006 | CAA | | 2.99E-10 | ?+ | 2.988E-10 | | New |

| | | | | | | | | | | | | |
|---|---|---|---|---|---|---|---|---|---|---|---|---|
| 502 | 7 | 7-73239519-TA-T | | NA | 0.0011 | TA | | 3.37E-15 | ?+ | 3.371E-15 | | New |
| 502 | 7 | 7-73239535-AGC-A | | NA | 0.0001 | A | | 2.07E-08 | ?+ | 2.071E-08 | | New |
| 503 | 7 | 7-73415961-C-CTCT | | NA | 0.0023 | CTCT | | 2.36E-13 | ?+ | 2.358E-13 | | New |
| 504 | 7 | 7-73481926-C-T | rs1554573754 | BAZ1B | 0.0011 | C | 0.92193 | 4.93E-14 | -+ | 6.822E-09 | 0.8207 | New |
| 505 | 7 | 7-73619442-C-CAG | rs1554601495 | MLXIPL | 0.0065 | C | | 6.63E-21 | ?+ | 6.626E-21 | | New |
| 505 | 7 | 7-73619449-C-CAA | rs1554601497 | MLXIPL | 0.0054 | CAA | | 4.62E-18 | ?+ | 4.618E-18 | | New |
| 506 | 7 | 7-74071734-AAGTG-A | | NA | 0.0006 | AAGTG | | 5.87E-09 | ?+ | 5.872E-09 | | New |
| 507 | 7 | 7-74300691-C-CAA | rs1554726695 | CLIP2 | 0.0007 | CAA | | 1E-09 | ?+ | 1.001E-09 | 0.4488 | New |
| 508 | 7 | 7-74303748-C-CT | | NA | 0.0007 | C | | 3.61E-08 | ?+ | 3.608E-08 | | New |
| 509 | 7 | 7-74493250-C-T | rs1554337191 | GTF2IRD1 | 0.0024 | C | 0.727944 | 1.51E-25 | -+ | 9.657E-11 | 0.2411 | New |
| 510 | 7 | 7-74792891-TA-T | | NA | 0.0007 | TA | | 4.73E-12 | ?+ | 4.732E-12 | | New |
| 511 | 7 | 7-75607689-GT-G | | NA | 0.0028 | GT | | 4E-32 | ?+ | 4.002E-32 | | New |
| 511 | 7 | 7-75607691-AC-A | | NA | 0.0014 | A | | 5.72E-21 | ?+ | 5.718E-21 | | New |
| 512 | 7 | 7-75792984-ATG-A | rs1554531069 | CCL26, CCL24 | 0.0015 | A | | 4.68E-16 | ?+ | 4.677E-16 | 0.3224 | New |
| 512 | 7 | 7-75792987-C-CAT | rs1554531072 | CCL26, CCL24 | 0.0016 | CAT | | 8.47E-17 | ?+ | 8.47E-17 | 0.1902 | New |
| 513 | 7 | 7-75801743-G-T | rs1554532051 | CCL26, CCL24 | 0.0003 | T | | 4.28E-08 | ?+ | 4.284E-08 | | New |
| 513 | 7 | 7-75801752-ATC-A | | NA | 0.0002 | A | | 6.82E-10 | ?+ | 6.816E-10 | | New |
| 514 | 7 | 7-75853377-GTTT-G | | NA | 0.0005 | G | | 4.71E-08 | ?+ | 4.714E-08 | | New |
| 515 | 7 | 7-89881278-AT-A | | NA | 0.0004 | A | | 1.56E-14 | ?+ | 1.565E-14 | 0.4501 | New |
| 515 | 7 | 7-89881280-C-CCTGGCTAACAT | | NA | 0.0003 | C | | 3.96E-10 | ?+ | 3.955E-10 | | New |
| 516 | 7 | 7-97243876-TCCAC-T | rs1240859920 | SDHAF3, RN7SKP104 | 0.0046 | TCCAC | | 1.33E-08 | ?+ | 1.332E-08 | | New |
| 517 | 8 | 8-101230534-TG-T | | NA | 0.0016 | TG | | 1.81E-08 | ?+ | 1.812E-08 | 0.9034 | New |

| | | | | | | | | | | |
|---|---|---|---|---|---|---|---|---|---|---|
| 518 | 8 | 8-102887720-ATTT-A | | NA | 0.0021 | A | 6.83E-09 | ?+ | 6.828E-09 | | New |
| 519 | 8 | 8-104680838-GGAGTTC-G | | NA | 0.0014 | GGAGTTC | 2.62E-09 | ?+ | 2.622E-09 | 0.1332 | New |
| 520 | 8 | 8-125402911-AAG-A | rs1246130952 | NSMCE2, TRIB1 | 0.0006 | A | 2.88E-08 | ?+ | 2.885E-08 | 0.1455 | New |
| 521 | 8 | 8-130031600-C-CT | rs1370965392 | FAM49B, ASAP1 | 0.0039 | CT | 1.72E-13 | ?+ | 1.724E-13 | 0.3025 | New |
| 522 | 8 | 8-1316426-C-CTCTCCAACAGTGGTCTACACTCGAGAAACTCGGCAGCTTTTAAAAATAGAGCGTGTGCGAGTGCAGCGTCTCTCCAACAGTGGTCTACACTCGAGACACTCGGCAGCGTTTAAAAATAGAGGCTGTGCGAGTGCAGCG | rs1563078617 | DLGAP2 | 0.0243 | C | 4.61E-09 | ?- | 4.605E-09 | | New |
| 523 | 8 | 8-13268436-TC-T | | NA | 0.0019 | T | 7.18E-15 | ?+ | 7.18E-15 | | New |
| 524 | 8 | 8-134499573-GGATGCCCCCGCTGCTGGTTACACACAGAGCCTGATTTGGGAGGGTCGGGGTGGAGCCGT-G | rs1818800972 | ZFAT | 0.004 | G | 1.16E-14 | ?+ | 1.155E-14 | | New |
| 525 | 8 | 8-143960466-AT-A | | NA | 0.0028 | AT | 2.76E-22 | ?+ | 2.765E-22 | | Old |
| 526 | 8 | 8-144124603-C-CA | rs1374854652 | WDR97, HGH1 | 0.0024 | C | 2E-22 | ?+ | 1.996E-22 | | Old |
| 527 | 8 | 8-144216367-G-A | rs146192223 | MROH1 | 0.0707 | G | 5.92E-10 | ?+ | 5.924E-10 | 0.6639 | Old |
| 528 | 8 | 8-144301660-G-T | | HSF1 | 0.0002 | T | 4.33E-08 | ?+ | 4.325E-08 | | Old |
| 529 | 8 | 8-1603569-T-TGGGTCTCAGTTCTGCAGAGGCTGGTTAGAGTGGAGGTG | rs71190749 | DLGAP2-AS1 | 0.0438 | TGGGTCTCAGTTCTGC. | 1.7E-10 | ?+ | 1.698E-10 | | New |

| | | | | | | | | | | | | |
|---|---|---|---|---|---|---|---|---|---|---|---|---|
| 530 | 8 | 8-23267200-AT-A | rs1200144773 | CHMP7, R3HCC1 | 0.0022 | AT | | 1.35E-12 | ?+ | 1.353E-12 | 0.8416 | New |
| 531 | 8 | 8-23279493-TGG-T | rs1802665945 | R3HCC1 | 0.0012 | TGG | | 7.27E-09 | ?+ | 7.269E-09 | | New |
| 532 | 8 | 8-30679870-TC-T | | NA | 0.0011 | TC | | 9.69E-13 | ?+ | 9.69E-13 | | New |
| 533 | 8 | 8-33468388-TG-T | rs1456752008 | FUT10 | 0.0014 | T | 0.679781 | 3.41E-12 | -+ | 1.544E-10 | 0.7119 | New |
| 534 | 8 | 8-33494173-AAG-A | | NA | 0.0003 | A | | 3.09E-11 | ?+ | 3.085E-11 | | New |
| 534 | 8 | 8-33494177-ACC-A | | NA | 0.0002 | ACC | | 7.08E-11 | ?+ | 7.076E-11 | | New |
| 535 | 8 | 8-38194649-C-CAG | | NA | 0.0011 | CAG | 0.287517 | 1.21E-13 | -+ | 7.418E-11 | | New |
| 536 | 8 | 8-38390825-GGT-G | | LETM2 | 0.0013 | G | | 6.01E-10 | ?+ | 6.014E-10 | | New |
| 537 | 8 | 8-40820780-C-T | rs114236584 | ZMAT4 | 0.0217 | C | | 1.98E-19 | ?- | 1.983E-19 | | New |
| 538 | 8 | 8-43008878-C-T | rs1179689749 | HOOK3 | 0.0011 | T | 0.165033 | 5.01E-15 | ++ | 1.899E-15 | | New |
| 539 | 8 | 8-43570083-G-C | rs10097868 | RF00012, NONE | 0.1758 | G | | 8.1E-10 | ?+ | 8.103E-10 | | New |
| 540 | 8 | 8-43927222-T-A | rs1470310616 | RF00012, NONE | 0.0012 | A | 0.143375 | 1.54E-12 | ++ | 9.141E-11 | 0.4626 | New |
| 541 | 8 | 8-47081758-G-A | | RN7SKP32, AC120036.5 | 0.001 | G | | 2.42E-08 | ?+ | 2.42E-08 | | New |
| 542 | 8 | 8-48012945-AAG-A | rs1452849601 | UBE2V2 | 0.0015 | A | | 4.29E-08 | ?+ | 4.286E-08 | 0.3315 | New |
| 543 | 8 | 8-48019002-G-T | rs771656340 | UBE2V2 | 0.0003 | T | | 1.89E-10 | ?+ | 1.886E-10 | | New |
| 544 | 8 | 8-55827470-AACGCCATTCTCCTGCCTCAGCCTCCCGAGTAGCTG-A | | NA | 0.0023 | A | | 3.9E-11 | ?+ | 3.898E-11 | | New |
| 545 | 8 | 8-60523306-C-CAT | | RAB2A | 0.0004 | CAT | | 3.74E-08 | ?+ | 3.743E-08 | | New |
| 546 | 8 | 8-66708539-C-A | rs113771521 | SGK3 | 0.1291 | A | | 1.41E-08 | ?+ | 1.405E-08 | 0.4419 | New |
| 547 | 8 | 8-67370567-C-CCA | rs1388282779 | AC021321.1, CPA6 | 0.0017 | CCA | 0.18898 | 2.07E-12 | ++ | 1.058E-12 | | New |
| 548 | 8 | 8-7412675-CT-C | | ZNF705G, DEFB4B | 0.426 | C | | 2.89E-22 | ?+ | 2.893E-22 | | New |
| 549 | 8 | 8-80570527-GC-G | | RNU7-174P, RNU2-71P | 0.0014 | G | | 3.46E-16 | ?+ | 3.464E-16 | | Old |

| | | | | | | | | | | | | |
|---|---|---|---|---|---|---|---|---|---|---|---|---|
| 550 | 8 | 8-90506149-C-A | rs1329444417 | AC004083.1, LINC00534 | 0.0021 | A | 0.9022 | 2.95E-22 | -+ | 3.043E-19 | | New |
| 550 | 8 | 8-90506156-C-T | rs1008588893 | AC004083.1, LINC00534 | 0.0009 | T | 0.532693 | 5.8E-09 | ++ | 9.776E-09 | 0.9525 | New |
| 551 | 8 | 8-93926331-C-CA | rs1470817156 | PDP1 | 0.002 | CA | | 1.41E-09 | ?+ | 1.407E-09 | 0.4818 | New |
| 551 | 8 | 8-93926333-GCAT-G | rs1168533702 | PDP1 | 0.0024 | GCAT | | 1.46E-14 | ?+ | 1.46E-14 | 0.3434 | New |
| 552 | 8 | 8-94591747-C-CT | | NA | 0.001 | C | | 4.39E-09 | ?+ | 4.387E-09 | | Old |
| 553 | 8 | 8-9493603-ATT-A | | NA | 0.0004 | ATT | | 7.89E-09 | ?+ | 7.888E-09 | | New |
| 554 | 8 | 8-97732251-ACT-A | rs1172668780 | MTDH, RF00019 | 0.0005 | A | | 4.08E-10 | ?+ | 4.076E-10 | | New |
| 555 | 8 | 8-99128207-GGT-G | | NA | 0.002 | GGT | | 1.61E-17 | ?+ | 1.61E-17 | | New |
| 556 | 9 | 9-104150373-GGC-G | | NA | 0.0009 | G | | 1.77E-09 | ?+ | 1.768E-09 | | New |
| 556 | 9 | 9-104150379-TCC-T | | SMC2, OR13F1 | 0.0009 | TCC | | 2.23E-08 | ?+ | 2.233E-08 | | New |
| 557 | 9 | 9-105169701-G-A | rs906834295 | AL591506.1 | 0.0007 | A | 0.036444 | 6.39E-08 | ++ | 2.114E-08 | 0.1357 | Old |
| 557 | 9 | 9-105169723-T-C | rs957545383 | AL591506.1 | 0.0006 | T | 0.019272 | 4.41E-07 | ++ | 2.873E-08 | | Old |
| 558 | 9 | 9-110014597-AT-A | | NA | 0.0006 | AT | | 1.22E-11 | ?+ | 1.219E-11 | | New |
| 559 | 9 | 9-118668202-C-CAG | | NA | 0.0008 | C | | 1.49E-08 | ?+ | 1.489E-08 | | New |
| 560 | 9 | 9-123399029-C-CAA | rs1314416129 | DENND1A | 0.0041 | C | | 2.37E-09 | ?+ | 2.371E-09 | | New |
| 561 | 9 | 9-123706553-C-CCT | | NA | 0.0017 | CCT | | 5.43E-17 | ?+ | 5.427E-17 | | New |
| 562 | 9 | 9-123917954-C-CAAAA | rs1225330562 | DENND1A | 0.0054 | C | | 3.66E-10 | ?+ | 3.657E-10 | | New |
| 563 | 9 | 9-125592686-C-CCT | rs1470069776 | MAPKAP1 | 0.0024 | CCT | 0.86742 | 1.04E-10 | -+ | 1.382E-09 | | New |
| 564 | 9 | 9-125689462-C-A | rs77961302 | MAPKAP1 | 0.0346 | C | 0.948883 | 4.52E-11 | ++ | 1.085E-10 | | New |

| | | | | | | | | | | | | |
|---|---|---|---|---|---|---|---|---|---|---|---|---|
| 565 | 9 | 9-126014359-C-A | rs553736939 | PBX3, AL589923.1 | 0.0011 | A | 0.201084 | 3.98E-10 | ++ | 8.404E-09 | | New |
| 566 | 9 | 9-128665286-C-CAG | | NA | 0.0008 | CAG | | 3.66E-08 | ?+ | 3.659E-08 | | New |
| 567 | 9 | 9-129033772-GAGA-G | | NA | 0.001 | GAGA | | 9.75E-10 | ?+ | 9.751E-10 | | New |
| 568 | 9 | 9-129727696-C-CCT | | NA | 0.0025 | CCT | | 5.56E-23 | ?+ | 5.552E-23 | | New |
| 569 | 9 | 9-129980341-C-CA | | NA | 0.0003 | C | | 2.03E-09 | ?+ | 2.026E-09 | | New |
| 570 | 9 | 9-130955273-C-CACCGTT | rs1156259878 | AL161733.1, LAMC3 | 0.0047 | CACCGTT | | 7.89E-10 | ?+ | 7.886E-10 | | New |
| 571 | 9 | 9-131326169-C-T | rs1019641964 | PLPP7 | 0.0005 | C | | 2.49E-09 | ?+ | 2.493E-09 | 0.3065 | New |
| 571 | 9 | 9-131326190-AGCCTGGTC-A | rs1836070657 | PLPP7 | 0.0017 | AGCCTGGTC | | 6.53E-09 | ?+ | 6.533E-09 | | New |
| 572 | 9 | 9-133482173-C-T | rs1010545857 | SLC2A6, MYMK | 0.0007 | T | 0.141773 | 1.43E-09 | ++ | 1.128E-09 | | New |
| 572 | 9 | 9-133482205-G-C | rs959227807 | SLC2A6, MYMK | 0.0017 | G | 0.315331 | 2.43E-15 | -+ | 5.55E-13 | 0.8608 | New |
| 573 | 9 | 9-135673019-TTTTG-T | | LCN9, SOHLH1 | 0.0007 | T | | 3.38E-09 | ?+ | 3.377E-09 | | New |
| 574 | 9 | 9-136639478-C-T | rs1417363683 | AL590226.2, AL590226.1 | 0.0005 | T | 0.145337 | 2.09E-10 | ++ | 7.972E-10 | 0.9206 | New |
| 575 | 9 | 9-137334886-G-C | rs949943356 | EXD3 | 0.0019 | C | 0.424351 | 2.17E-15 | -+ | 1.966E-13 | | New |
| 576 | 9 | 9-16934631-TTC-T | rs1364541871 | AL162725.2 | 0.003 | T | | 6.26E-09 | ?+ | 6.262E-09 | 0.6638 | New |
| 577 | 9 | 9-2528493-AC-A | | NA | 0.0009 | AC | | 6.48E-11 | ?+ | 6.476E-11 | | New |
| 578 | 9 | 9-26890151-C-CCT | | NA | 0.001 | C | | 6.31E-10 | ?+ | 6.308E-10 | | New |
| 579 | 9 | 9-31320715-C-CGG | | NA | 0.0002 | C | | 7.12E-09 | ?+ | 7.123E-09 | | New |
| 579 | 9 | 9-31320722-TCAGC-T | | NA | 0.0002 | TCAGC | | 1.6E-09 | ?+ | 1.596E-09 | | New |
| 580 | 9 | 9-33744596-T-A | rs1822346958 | LINC01251, PRSS3 | 0.0011 | T | 0.186294 | 7.92E-15 | ++ | 4.146E-13 | | New |
| 581 | 9 | 9-34034655-ACC-A | | NA | 0.0007 | ACC | 0.836381 | 2.09E-15 | ++ | 1.065E-14 | | New |

| | | | | | | | | | | | | |
|---|---|---|---|---|---|---|---|---|---|---|---|---|
| 582 | 9 | 9-34042314-C-CA | rs1407033483 | UBAP2 | 0.0011 | CA | | 4.82E-08 | ?+ | 4.824E-08 | | New |
| 582 | 9 | 9-34042322-C-CA | rs765534905 | UBAP2 | 0.0019 | C | | 1.86E-09 | ?+ | 1.863E-09 | | New |
| 583 | 9 | 9-34094697-AT-A | rs1354347537 | AL354989.1 | 0.0015 | A | 0.369773 | 2.95E-15 | -+ | 8.962E-09 | 0.3045 | New |
| 584 | 9 | 9-36604110-C-T | rs1587397386 | MELK | 0.0019 | T | 0.387594 | 8.92E-19 | -+ | 1.162E-14 | | New |
| 585 | 9 | 9-38688643-G-C | rs1203948313 | RNU6-765P, FAM240B | 0.001 | C | | 1.34E-08 | ?+ | 1.338E-08 | | New |
| 586 | 9 | 9-39941434-A-AT | | RF00156, AL772307.1 | 0.1297 | AT | | 3.34E-25 | ?+ | 3.337E-25 | | New |
| 587 | 9 | 9-40329787-C-CTTTT | | BX664727.3 | 0.0323 | C | | 2.31E-12 | ?+ | 2.306E-12 | | New |
| 588 | 9 | 9-40557771-T-TC | rs1232620381 | BMS1P14 | 0.0139 | TC | | 3.56E-08 | ?+ | 3.559E-08 | | New |
| 589 | 9 | 9-40894485-G-C | rs143962334 | AL353626.1, MIR1299 | 0.0008 | G | | 5.6E-10 | ?+ | 5.603E-10 | | New |
| 590 | 9 | 9-41727997-TA-T | | FAM242F, AL162731.1 | 0.152 | TA | | 4.05E-09 | ?+ | 4.054E-09 | | New |
| 591 | 9 | 9-42847389-A-AG | | BX664718.1, RNU6-599P | 0.0733 | A | | 9.79E-09 | ?- | 9.793E-09 | | New |
| 592 | 9 | 9-61670607-TAG-T | rs1822106862 | AL935212.1, RN7SL722P | 0.0041 | T | | 2.38E-10 | ?+ | 2.384E-10 | 0.7959 | New |
| 593 | 9 | 9-62838432-C-CCGGCGCCCCCTCCGCGCCGGCGCCCCCTCCGCGCCGGCGCCCCCTCCGCGCCGGCGCCCGGCGCT | | NA | 0.0035 | CCGGCGCCCCCTCCGC | | 3.66E-09 | ?- | 3.658E-09 | | New |
| 594 | 9 | 9-6662926-C-CTG | rs1316966968 | AL162411.1 | 0.0016 | C | 0.818951 | 5.57E-09 | -+ | 1.293E-08 | | New |
| 595 | 9 | 9-6698604-GCC-G | | NA | 0.0002 | G | | 3E-09 | ?+ | 3.004E-09 | | New |
| 596 | 9 | 9-67632973-C-CAA | | BX664730.1, RF00019 | 0.0192 | CAA | | 4.17E-08 | ?+ | 4.168E-08 | | New |

| | | | | | | | | | | | | |
|---|---|---|---|---|---|---|---|---|---|---|---|---|
| 597 | 9 | 9-73226646-G-A | rs1822289798 | ANXA1, AL451127.2 | 0.0011 | G | 0.19383 | 2.13E-11 | ++ | 7.22E-09 | | New |
| 598 | 9 | 9-74786521-C-CT | rs1290866150 | TRPM6 | 0.0022 | CT | 0.437918 | 6.57E-13 | ++ | 1.315E-11 | | New |
| 599 | 9 | 9-82464932-C-T | rs950049780 | AL162726.4, AL356490.1 | 0.0011 | C | 0.317783 | 3.48E-13 | -+ | 1.232E-08 | 0.1792 | New |
| 600 | 9 | 9-84449299-C-T | rs548469999 | AL157886.1 | 0.0007 | T | 0.160959 | 2.46E-09 | ++ | 1.869E-09 | | New |
| 601 | 9 | 9-8630638-C-G | rs142283076 | PTPRD | 1E-04 | C | 1.06E-05 | 0.000132 | ++ | 2.197E-08 | | New |
| 602 | 9 | 9-92785200-GT-G | | NA | 0.0004 | G | | 8.51E-10 | ?+ | 8.509E-10 | | New |
| 603 | 9 | 9-93166066-AT-A | rs1183474232 | AL390760.1, WNK2 | 0.0019 | AT | | 3.87E-16 | ?+ | 3.872E-16 | 0.7207 | New |
| 604 | 9 | 9-94616642-C-CA | rs1272598863 | FBP1 | 0.0004 | C | | 1.83E-15 | ?+ | 1.835E-15 | | New |
| 604 | 9 | 9-94616646-GC-G | rs1206929678 | FBP1 | 0.0005 | G | | 5.29E-13 | ?+ | 5.29E-13 | | New |
| 604 | 9 | 9-94616648-ATG-A | rs1231327791 | FBP1 | 0.0007 | ATG | | 1.5E-14 | ?+ | 1.495E-14 | | New |
| 605 | 9 | 9-96946504-C-CTT | rs1332932956 | MFSD14C | 0.0016 | CTT | | 2.09E-08 | ?+ | 2.091E-08 | | New |
| 606 | 10 | 10-10099500-G-A | rs994051438 | LINC02670, RF00019 | 0.0011 | A | | 2.06E-11 | ?+ | 2.059E-11 | | New |
| 607 | 10 | 10-101802646-GCAA-G | rs1243800619 | OGA | 0.0015 | G | | 3.19E-10 | ?+ | 3.189E-10 | 0.6033 | New |
| 608 | 10 | 10-102054370-C-CG | rs1432629908 | ARMH3 | 0.0012 | CG | | 4.54E-13 | ?+ | 4.535E-13 | 0.6008 | New |
| 609 | 10 | 10-102129838-C-CG | | NA | 0.0005 | C | | 9.75E-11 | ?+ | 9.753E-11 | | New |
| 610 | 10 | 10-102174312-C-CT | rs1469263388 | NOLC1, ELOVL3 | 0.0012 | CT | 0.361198 | 5.56E-10 | ++ | 1.478E-08 | 0.5656 | New |
| 611 | 10 | 10-102206649-AAG-A | rs1472108032 | NOLC1, ELOVL3 | 0.0056 | A | | 1.25E-11 | ?+ | 1.252E-11 | 0.5161 | New |
| 612 | 10 | 10-102785427-TCTGC-T | rs1172108720 | WBP1L | 0.0005 | TCTGC | | 7.23E-09 | ?+ | 7.235E-09 | 0.5562 | New |
| 613 | 10 | 10-111403547-C-CTGG | | NA | 0.0004 | CTGG | | 6.57E-09 | ?+ | 6.57E-09 | | New |

| | | | | | | | | | | | | |
|---|---|---|---|---|---|---|---|---|---|---|---|---|
| 614 | 10 | 10-111586970-CA-C | | AC021035.1, AL136119.1 | 0.0457 | C | | 4.33E-09 | ?- | 4.326E-09 | | New |
| 615 | 10 | 10-114884738-C-T | rs1321925127 | FAM160B1 | 0.0004 | T | | 2.54E-10 | ?+ | 2.543E-10 | 0.09473 | New |
| 616 | 10 | 10-116619728-ATTTT-A | rs1185918181 | PNLIPRP1, PNLIPRP2 | 0.0017 | A | | 2.3E-08 | ?+ | 2.298E-08 | | New |
| 617 | 10 | 10-117460011-T-A | rs1278738107 | AC005871.2 | 0.0006 | A | 0.247876 | 4.26E-10 | ++ | 3.421E-09 | | New |
| 618 | 10 | 10-119182166-A-AC | | PRDX3, GRK5 | 0.0255 | AC | | 2.87E-09 | ?+ | 2.875E-09 | | New |
| 619 | 10 | 10-119200773-C-T | rs771041661 | PRDX3, GRK5 | 0.0012 | T | 0.925359 | 1.26E-14 | -+ | 5.2E-12 | 0.2439 | New |
| 619 | 10 | 10-119200781-C-A | rs1368039384 | PRDX3, GRK5 | 0.0022 | A | | 2.28E-19 | ?+ | 2.275E-19 | | New |
| 620 | 10 | 10-119907249-TTG-T | | SEC23IP | 0.0011 | TTG | | 1.7E-14 | ?+ | 1.703E-14 | | New |
| 620 | 10 | 10-119907251-ACG-A | | SEC23IP | 0.0008 | A | | 1.78E-12 | ?+ | 1.784E-12 | | New |
| 621 | 10 | 10-120602534-C-T | rs1184974643 | LINC01561, AC023282.1 | 0.0002 | C | | 2.79E-08 | ?+ | 2.792E-08 | 0.271 | New |
| 622 | 10 | 10-12148087-C-CACG | | SEC61A2 | 0.0013 | CACG | | 1.71E-13 | ?+ | 1.711E-13 | | Old |
| 623 | 10 | 10-12279639-C-CT | | NA | 0.0002 | C | | 1.67E-10 | ?+ | 1.673E-10 | | New |
| 624 | 10 | 10-12380003-AG-A | rs1281842209 | CAMK1D | 0.0017 | A | 0.936821 | 2.4E-10 | ++ | 8.224E-09 | | New |
| 625 | 10 | 10-124809427-GGT-G | rs1458056517 | ABRAXAS2 | 0.0046 | G | | 8.73E-17 | ?+ | 8.727E-17 | 0.6515 | New |
| 626 | 10 | 10-125834928-C-CG | rs1382795820 | BCCIP | 0.0028 | C | 0.762481 | 7.79E-11 | -+ | 1.583E-09 | | New |
| 627 | 10 | 10-126892051-AC-A | rs2030065142 | RF00271, DOCK1 | 0.0082 | A | | 3.12E-12 | ?+ | 3.116E-12 | | New |
| 628 | 10 | 10-128016654-GA-G | | PTPRE | 0.0008 | GA | | 4.73E-09 | ?+ | 4.727E-09 | | New |
| 629 | 10 | 10-132388816-CG-C | | LRRC27, PWWP2B | 0.088 | CG | | 8.37E-17 | ?+ | 8.366E-17 | | New |
| 630 | 10 | 10-132611004-GT-G | rs113740336 | INPP5A | 0.0036 | GT | | 3.67E-11 | ?+ | 3.669E-11 | | New |

| | | | | | | | | | | | | |
|---|---|---|---|---|---|---|---|---|---|---|---|---|
| 631 | 10 | 10-14933460-AGCCCC-A | | NA | 0.0004 | A | | 2.11E-10 | ?+ | 2.108E-10 | | New |
| 631 | 10 | 10-14933466-ATGTCG-A | | NA | 0.0003 | ATGTCG | | 4.68E-09 | ?+ | 4.683E-09 | | New |
| 632 | 10 | 10-1593285-ATAGGTCTCCTCTCTGGCATGGTCCCCTCTGTCACCCAGCTCCCCTGGCCCAAACCACACTCTG-A | rs1833286573 | ADARB2 | 0.0114 | A | | 2.32E-20 | ?+ | 2.317E-20 | | New |
| 633 | 10 | 10-17983189-ACCTAG-A | | NA | 0.0008 | ACCTAG | | 3.85E-08 | ?+ | 3.854E-08 | | New |
| 634 | 10 | 10-18396114-G-A | rs997658060 | CACNB2 | 0.0007 | A | 0.825206 | 3.42E-09 | ++ | 5.859E-09 | 0.9935 | New |
| 635 | 10 | 10-23108707-C-T | rs1840010844 | AL139815.1 | 0.0003 | C | | 1.46E-09 | ?+ | 1.459E-09 | | New |
| 636 | 10 | 10-27215821-TC-T | rs1247157380 | ACBD5 | 0.0019 | T | | 1.44E-11 | ?+ | 1.437E-11 | 0.8912 | New |
| 637 | 10 | 10-2983675-G-A | rs1413323051 | AL355361.1, AL731533.1 | 0.001 | G | | 1.85E-08 | ?+ | 1.85E-08 | 0.2428 | New |
| 637 | 10 | 10-2983682-C-A | rs1351359673 | AL355361.1, AL731533.1 | 0.0008 | C | 0.903154 | 2.83E-10 | ++ | 5.873E-10 | 0.0459 | New |
| 637 | 10 | 10-2983692-T-A | rs1349859144 | AL355361.1, AL731533.1 | 0.0008 | A | 0.902933 | 2.16E-10 | ++ | 4.468E-10 | 0.1249 | New |
| 638 | 10 | 10-35114079-TGGGTTTTTTTTTTTTG-T | rs1207599796 | AL392046.1 | 0.0024 | T | | 2.01E-18 | ?+ | 2.013E-18 | | New |
| 639 | 10 | 10-35370323-G-T | rs1836903150 | CCNY | 0.0015 | G | 0.700674 | 1.44E-13 | ++ | 8.642E-10 | | New |
| 640 | 10 | 10-38930856-T-A | rs879031925 | AL133216.1, NONE | 0.0026 | A | | 2.73E-10 | ?+ | 2.73E-10 | | New |
| 641 | 10 | 10-39565455-A-C | rs1355114191 | AL133216.1, NONE | 0.0396 | A | 0.456825 | 1.31E-41 | ++ | 5.956E-31 | | New |
| 642 | 10 | 10-41772582-AAG-A | | NA | 0.0009 | AAG | | 6.46E-14 | ?+ | 6.457E-14 | | New |

| | | | | | | | | | | | |
|---|---|---|---|---|---|---|---|---|---|---|---|
| 642 | 10 | 10-41772584-GCA-G | | NA | 0.0008 | G | | 1.74E-12 | ?+ | 1.735E-12 | | New |
| 643 | 10 | 10-41847867-AT-A | rs1157245202 | NONE, LINC00839 | 0.0038 | A | | 2.26E-12 | ?+ | 2.264E-12 | | New |
| 643 | 10 | 10-42066423-AATC-A | rs1252500162 | NONE, LINC00839 | 0.0032 | A | | 5.97E-13 | ?+ | 5.972E-13 | | New |
| 644 | 10 | 10-42102736-T-A | rs143943467 | NONE, LINC00839 | 0.0021 | A | | 2.13E-09 | ?+ | 2.134E-09 | | New |
| 644 | 10 | 10-42102765-G-C | rs57742507 | NONE, LINC00839 | 0.0016 | G | | 1.44E-14 | ?+ | 1.439E-14 | | New |
| 645 | 10 | 10-5378108-ATGGTG-A | | NA | 0.0009 | ATGGTG | | 1.32E-12 | ?+ | 1.323E-12 | | New |
| 646 | 10 | 10-68227393-G-T | rs935637877 | MYPN, ATOH7 | 0.001 | T | 0.702014 | 6.3E-10 | ++ | 8.034E-09 | | New |
| 647 | 10 | 10-70420363-C-CT | | NA | 0.001 | CT | | 1.31E-11 | ?+ | 1.315E-11 | | New |
| 647 | 10 | 10-70420366-T-A | rs1007847836 | EIF4EBP2 | 0.0004 | T | | 9.86E-09 | ?+ | 9.866E-09 | | New |
| 648 | 10 | 10-72191881-C-CAA | rs1275756507 | ASCC1 | 0.002 | C | | 6.46E-10 | ?+ | 6.463E-10 | | New |
| 649 | 10 | 10-73947669-TG-T | | NA | 0.0006 | T | | 1.01E-10 | ?+ | 1.011E-10 | | New |
| 650 | 10 | 10-74595727-A-G | rs368042471 | ADK | 0.0046 | A | | 4.32E-11 | ?+ | 4.316E-11 | | New |
| 650 | 10 | 10-74595729-T-TCC | rs377744233 | ADK | 0.0071 | T | | 7.23E-16 | ?+ | 7.234E-16 | | New |
| 651 | 10 | 10-79922400-G-A | rs2758559 | AL512662.2 | 0.0082 | G | | 2E-13 | ?+ | 1.996E-13 | 0.03556 | Old |
| 652 | 10 | 10-84087114-C-CGT | | NA | 0.001 | C | | 1.18E-10 | ?+ | 1.18E-10 | | New |
| 653 | 10 | 10-97710681-ACT-A | rs1374926972 | AVPI1, MARVELD1 | 0.0017 | ACT | | 3.07E-10 | ?+ | 3.072E-10 | 0.4273 | New |
| 654 | 11 | 11-1018088-TG-T | rs368342230 | MUC6 | 0.0087 | T | | 6.35E-09 | ?+ | 6.353E-09 | | New |
| 655 | 11 | 11-1018456-G-A | rs79920422 | MUC6 | 0.0028 | A | 0.373363 | 1.1E-10 | ++ | 4.132E-10 | | New |
| 656 | 11 | 11-105515895-G-T | rs1053968571 | CARD18 | 0.0028 | T | 0.479102 | 2.34E-18 | ++ | 2.641E-18 | | New |

| | | | | | | | | | | | | |
|---|---|---|---|---|---|---|---|---|---|---|---|---|
| 656 | 11 | 11-105515902-G-A | rs914982993 | CARD18 | 0.0016 | A | 0.691041 | 1.38E-17 | ++ | 3.275E-17 | 0.1885 | New |
| 657 | 11 | 11-1095580-GAGTACA-G | | NA | 0.0026 | G | | 3.54E-08 | ?+ | 3.537E-08 | | New |
| 658 | 11 | 11-113771108-TG-T | | NA | 0.0009 | TG | | 4.83E-15 | ?+ | 4.825E-15 | | New |
| 659 | 11 | 11-1166174-C-CCCTATGGTGAGACCCTGCACCCAACACACAGTCTA | | NA | 0.0021 | C | | 7.27E-10 | ?+ | 7.273E-10 | | New |
| 660 | 11 | 11-117105931-C-CA | | NA | 0.0002 | C | | 3.63E-08 | ?+ | 3.625E-08 | | New |
| 661 | 11 | 11-118736461-C-CAT | rs1555154730 | AP002954.1 | 0.0025 | CAT | | 2.53E-08 | ?+ | 2.529E-08 | | New |
| 662 | 11 | 11-123615487-G-C | rs1349200192 | GRAMD1B | 0.0006 | C | 0.781646 | 3.21E-09 | ++ | 1.296E-08 | | New |
| 663 | 11 | 11-12757946-ATTTTG-A | | NA | 0.001 | ATTTTG | | 6.95E-09 | ?+ | 6.946E-09 | | New |
| 664 | 11 | 11-129833604-G-A | rs1316952114 | TMEM45B | 0.0008 | A | 0.094626 | 1.03E-10 | ++ | 2.658E-10 | | New |
| 665 | 11 | 11-134286572-G-C | rs1243638640 | GLB1L3 | 0.0087 | C | 0.140239 | 3.78E-18 | ++ | 2.599E-08 | 0.1295 | New |
| 666 | 11 | 11-1355825-C-T | rs890196119 | LINC02689 | 0.001 | T | 0.004295 | 2.1E-08 | ++ | 3.183E-10 | 0.5118 | New |
| 666 | 11 | 11-1355828-G-T | rs1379529633 | LINC02689 | 0.0011 | G | 0.182731 | 2.06E-11 | ++ | 1.831E-10 | 0.9241 | New |
| 666 | 11 | 11-1355844-T-A | rs1023546743 | LINC02689 | 0.0019 | T | 0.373811 | 3.72E-16 | ++ | 2.126E-14 | 0.7431 | New |
| 667 | 11 | 11-18328184-C-CAG | rs1484419955 | GTF2H1 | 0.0016 | C | | 6.89E-09 | ?+ | 6.89E-09 | | New |
| 668 | 11 | 11-19260622-CCA-C | rs113522288 | CSRP3-AS1 | 0.0037 | CCA | 0.420952 | 9.92E-11 | -- | 1.303E-10 | | New |
| 669 | 11 | 11-2124905-GA-G | rs1276903792 | H19, IGF2 | 0.0029 | G | | 1.75E-08 | ?+ | 1.754E-08 | 0.1898 | New |
| 670 | 11 | 11-2139063-CG-C | | IGF2 | 0.0118 | C | | 3.32E-10 | ?+ | 3.319E-10 | | New |
| 671 | 11 | 11-30791372-GTA-G | | NA | 0.0007 | GTA | | 1.14E-09 | ?+ | 1.14E-09 | | New |
| 672 | 11 | 11-30887986-AGAGC-A | | NA | 0.0004 | A | | 1.46E-11 | ?+ | 1.457E-11 | | New |
| 673 | 11 | 11-31533623-C-CAG | | NA | 0.0009 | CAG | 0.427123 | 8.07E-10 | ++ | 7.772E-10 | | New |

| | | | | | | | | | | | | |
|---|---|---|---|---|---|---|---|---|---|---|---|---|
| 674 | 11 | 11-31971510-C-T | rs905950847 | PAUPAR | 0.002 | C | 0.071264 | 4.79E-13 | ++ | 9.747E-14 | | New |
| 675 | 11 | 11-33250072-TA-T | | NA | 0.0003 | T | | 3.67E-10 | ?+ | 3.67E-10 | | New |
| 676 | 11 | 11-3777426-G-T | | NUP98 | 0.0003 | T | 0.068039 | 5.77E-08 | ++ | 1.611E-08 | | New |
| 677 | 11 | 11-463371-GT-G | rs1311117768 | AC138230.1 | 0.0072 | GT | | 1.13E-08 | ?+ | 1.127E-08 | 0.01425 | New |
| 678 | 11 | 11-46355253-C-CATG | | NA | 0.0003 | CATG | | 7.84E-10 | ?+ | 7.843E-10 | | New |
| 679 | 11 | 11-46830926-G-A | rs1421155854 | CKAP5 | 0.0004 | G | 0.415225 | 8.67E-12 | -+ | 3.172E-11 | | Old |
| 680 | 11 | 11-47085719-G-A | rs564101029 | C11orf49 | 0.0007 | A | 0.657503 | 2.39E-10 | ++ | 2.284E-08 | | Old |
| 681 | 11 | 11-47674901-AT-A | | AGBL2 | 0.0003 | AT | | 5.14E-09 | ?+ | 5.138E-09 | | Old |
| 682 | 11 | 11-47677751-C-CTG | | NA | 0.0011 | CTG | | 1.29E-10 | ?+ | 1.289E-10 | | Old |
| 683 | 11 | 11-48727069-TTTGTC-T | rs1359118268 | OR4A47, TRIM51GP | 0.0058 | TTTGTC | | 2.79E-15 | ?+ | 2.786E-15 | | New |
| 683 | 11 | 11-48727087-ATG-A | rs1198553262 | OR4A47, TRIM51GP | 0.0061 | ATG | | 1.82E-16 | ?+ | 1.817E-16 | | New |
| 684 | 11 | 11-48768777-T-A | rs958560696 | OR4A47, TRIM51GP | 0.0003 | T | | 3.27E-10 | ?+ | 3.275E-10 | | New |
| 685 | 11 | 11-48843498-C-T | rs796329474 | OR4A47, TRIM51GP | 0.0006 | C | | 3.16E-08 | ?+ | 3.158E-08 | | New |
| 686 | 11 | 11-48870192-G-T | rs199945687 | OR4A47, TRIM51GP | 0.0003 | T | 0.000117 | 6.49E-06 | ++ | 6.084E-09 | | New |
| 687 | 11 | 11-48911550-C-T | rs1442475248 | OR4A47, TRIM51GP | 0.0015 | T | | 1.31E-12 | ?+ | 1.313E-12 | | New |
| 688 | 11 | 11-50576986-C-A | rs558450407 | AC109635.2, NONE | 0.0002 | C | 0.227177 | 5.7E-10 | -+ | 4.678E-08 | | New |
| 689 | 11 | 11-50716519-C-T | rs867108694 | AC109635.2, NONE | 0.0002 | T | | 1.64E-08 | ?+ | 1.643E-08 | | New |
| 690 | 11 | 11-54897486-TGCCA-T | | NA | 0.0032 | TGCCA | | 4.65E-09 | ?- | 4.65E-09 | | New |
| 691 | 11 | 11-55233749-C-T | rs2120001636 | OR4A5, TRIM48 | 0.006 | C | | 2.69E-68 | ?+ | 2.696E-68 | | New |

| | | | | | | | | | | | | |
|---|---|---|---|---|---|---|---|---|---|---|---|---|
| 691 | 11 | 11-55233796-C-A | | OR4A5, TRIM48 | 0.0021 | C | | 1.88E-23 | ?+ | 1.885E-23 | | New |
| 692 | 11 | 11-57768000-AT-A | rs1475471251 | AP001931.1, CTNND1, TMX2-CTNND1 | 0.0024 | AT | 0.64311 | 9.68E-21 | ++ | 3.371E-12 | | New |
| 693 | 11 | 11-61824974-C-CCT | rs1555073412 | FADS1, FADS2 | 0.0045 | CCT | | 7.07E-19 | ?+ | 7.074E-19 | 0.8907 | New |
| 694 | 11 | 11-62004372-C-CAG | | NA | 0.001 | CAG | 0.951223 | 2.18E-08 | -+ | 4.912E-08 | | New |
| 695 | 11 | 11-62211652-G-A | rs545361786 | SCGB2A1 | 0.0033 | A | 0.225421 | 8.07E-15 | -+ | 3.729E-10 | 0.6888 | New |
| 696 | 11 | 11-62523038-C-A | rs780466682 | AHNAK | 0.0008 | C | 0.422813 | 6.65E-10 | ++ | 5.665E-09 | 0.07122 | New |
| 697 | 11 | 11-62537111-GCACCACA-G | | NA | 0.0013 | G | 0.592255 | 3.52E-13 | ++ | 1.719E-12 | | New |
| 698 | 11 | 11-62845315-ATT-A | | NA | 0.0005 | A | | 4.29E-08 | ?+ | 4.289E-08 | | New |
| 699 | 11 | 11-62869905-C-CTTGA | rs1395280731 | SLC3A2 | 0.0023 | C | | 3.37E-17 | ?+ | 3.37E-17 | 0.05053 | New |
| 700 | 11 | 11-63684993-C-CAA | | NA | 0.0006 | C | | 1.41E-09 | ?+ | 1.411E-09 | | New |
| 701 | 11 | 11-63764272-C-T | rs1590909847 | C11orf95 | 0.0006 | C | 0.034778 | 9.13E-09 | ++ | 9.531E-10 | | New |
| 702 | 11 | 11-63860686-G-A | rs535744882 | MARK2 | 0.0003 | A | 0.739983 | 7.78E-09 | ++ | 2.715E-08 | 0.1995 | New |
| 703 | 11 | 11-64280476-C-A | rs1325649112 | BAD, GPR137 | 0.0012 | A | | 9.45E-15 | ?+ | 9.454E-15 | | New |
| 704 | 11 | 11-64662034-C-CTT | rs1413522490 | NRXN2 | 0.0016 | C | | 6.99E-12 | ?+ | 6.986E-12 | 0.1128 | New |
| 705 | 11 | 11-64664247-C-CAAAT | | NA | 0.0003 | C | | 9.08E-11 | ?+ | 9.082E-11 | | New |
| 706 | 11 | 11-652592-C-CTT | rs1210224922 | DEAF1 | 0.0012 | C | | 7.31E-09 | ?+ | 7.309E-09 | 0.2157 | New |
| 707 | 11 | 11-65463200-TAG-T | | NA | 0.0004 | T | | 7.59E-09 | ?+ | 7.588E-09 | | New |
| 708 | 11 | 11-65527549-AC-A | | NA | 0.0002 | A | | 1.98E-08 | ?+ | 1.976E-08 | | New |
| 709 | 11 | 11-67134652-GCGT-G | | NA | 0.0005 | G | | 3.02E-09 | ?+ | 3.019E-09 | | New |
| 710 | 11 | 11-67157185-C-T | rs1273071898 | KDM2A | 0.0015 | C | 0.081903 | 1.62E-13 | ++ | 2.486E-10 | | New |

| | | | | | | | | | | | | |
|---|---|---|---|---|---|---|---|---|---|---|---|---|
| 711 | 11 | 11-67988509-ATG-A | | NA | 0.0011 | ATG | | 1E-08 | ?+ | 1.004E-08 | | New |
| 712 | 11 | 11-68207031-AGG-A | rs1341660992 | KMT5B | 0.0015 | A | | 2.14E-09 | ?+ | 2.14E-09 | | New |
| 713 | 11 | 11-69577301-G-C | rs1430663358 | LINC01488, CCND1 | 0.0007 | G | | 3.44E-15 | ?+ | 3.44E-15 | | New |
| 714 | 11 | 11-69720156-ACT-A | rs1855026528 | FGF19, FGF4 | 0.0008 | ACT | | 5.89E-10 | ?+ | 5.889E-10 | | New |
| 715 | 11 | 11-69723576-G-C | rs1333742182 | FGF19, FGF4 | 0.0008 | C | 0.314411 | 3.99E-09 | ++ | 2.456E-09 | | New |
| 716 | 11 | 11-70803334-C-CTA | rs1555050988 | SHANK2 | 0.0072 | C | | 4.62E-19 | ?+ | 4.621E-19 | | New |
| 716 | 11 | 11-70803346-C-CCGTT | rs1555051013 | SHANK2 | 0.0072 | CCGTT | | 4.35E-17 | ?+ | 4.354E-17 | | New |
| 717 | 11 | 11-70890949-GT-G | rs1555074574 | AP003783.1 | 0.0033 | G | | 1.46E-30 | ?+ | 1.46E-30 | | New |
| 717 | 11 | 11-70890952-AT-A | rs1555074577 | AP003783.1 | 0.0019 | A | | 2.51E-18 | ?+ | 2.512E-18 | | New |
| 718 | 11 | 11-71920538-C-CAG | rs1168992261 | AP002495.1 | 0.0048 | C | 0.311751 | 1.46E-17 | ++ | 1.145E-17 | 0.4165 | New |
| 719 | 11 | 11-72063209-C-CCT | rs1243122814 | NUMA1 | 0.0016 | C | | 1.21E-14 | ?+ | 1.208E-14 | 0.8002 | New |
| 720 | 11 | 11-73692269-C-CCGAGGT | rs1342643975 | RAB6A | 0.0028 | C | | 1.01E-12 | ?+ | 1.012E-12 | 0.6954 | Old |
| 721 | 11 | 11-73813823-C-CCT | | NA | 0.0007 | CCT | | 3.25E-11 | ?+ | 3.246E-11 | | Old |
| 722 | 11 | 11-75748028-G-T | rs1269046383 | RN7SL786P, AP001922.1 | 0.0011 | G | | 1.15E-09 | ?+ | 1.155E-09 | | New |
| 723 | 11 | 11-76255841-GGGT-G | rs1331553624 | AP000785.2, THAP12 | 0.0009 | G | | 8.9E-11 | ?+ | 8.897E-11 | 0.4552 | New |
| 723 | 11 | 11-76255847-C-CA | rs1401992092 | AP000785.2, THAP12 | 0.0006 | C | | 6.75E-14 | ?+ | 6.751E-14 | | New |
| 724 | 11 | 11-77546029-C-CCT | | NA | 0.0004 | C | | 8.13E-09 | ?+ | 8.131E-09 | | New |
| 725 | 11 | 11-881238-G-A | rs1485554844 | CHID1 | 0.001 | A | | 5.35E-09 | ?+ | 5.347E-09 | 0.1042 | New |
| 726 | 11 | 11-8984332-G-T | rs534052504 | NRIP3 | 0.0006 | T | | 2.87E-10 | ?+ | 2.873E-10 | | New |

| | | | | | | | | | | | | |
|---|---|---|---|---|---|---|---|---|---|---|---|---|
| 727 | 11 | 11-91664514-G-A | rs372237806 | AP002791.1, AP003055.1 | 0.0009 | G | 0.484709 | 3.44E-17 | -+ | 2.413E-09 | | New |
| 728 | 11 | 11-9300701-C-A | rs1443840515 | TMEM41B | 0.002 | C | 0.531004 | 6.91E-21 | ++ | 3.965E-11 | 0.3065 | New |
| 728 | 11 | 11-9300706-G-A | rs368848940 | TMEM41B | 0.0018 | A | 0.281046 | 9.43E-18 | ++ | 1.606E-09 | 0.6874 | New |
| 729 | 11 | 11-9384165-C-T | rs1590420364 | IPO7 | 0.0005 | C | | 1.81E-11 | ?+ | 1.813E-11 | | New |
| 729 | 11 | 11-9384187-G-C | rs1303095550 | IPO7 | 0.0015 | G | 0.835013 | 3.15E-20 | -+ | 1.449E-18 | 0.3851 | New |
| 730 | 11 | 11-9551023-C-CTT | rs1318500235 | ZNF143, WEE1 | 0.0042 | CTT | | 2.62E-24 | ?+ | 2.625E-24 | | New |
| 731 | 12 | 12-101445846-C-A | | RF00019 | 0.0005 | A | | 8.89E-15 | ?+ | 8.887E-15 | | New |
| 732 | 12 | 12-103321503-C-T | rs202037943 | C12orf42 | 0.056 | T | | 8.36E-11 | ?+ | 8.362E-11 | | New |
| 733 | 12 | 12-108905116-G-A | rs528671660 | DAO, SVOP | 0.0006 | A | 0.110957 | 5.66E-13 | ++ | 1.664E-13 | | New |
| 734 | 12 | 12-111611215-ATT-A | rs1382775283 | ATXN2-AS, BRAP | 0.0008 | A | | 2.92E-10 | ?+ | 2.917E-10 | 0.8753 | New |
| 735 | 12 | 12-111929280-G-A | rs1297364356 | MAPKAPK5, TMEM116 | 0.0006 | A | | 6.29E-14 | ?+ | 6.29E-14 | 0.2761 | New |
| 735 | 12 | 12-111929282-G-A | rs1340910297 | MAPKAPK5, TMEM116 | 0.0003 | A | | 1.37E-09 | ?+ | 1.372E-09 | 0.6316 | New |
| 736 | 12 | 12-112412907-G-C | rs2037357773 | AC004086.1 | 0.001 | G | | 3.29E-11 | ?+ | 3.295E-11 | | New |
| 736 | 12 | 12-112412911-GA-G | | NA | 0.0009 | GA | | 8.16E-16 | ?+ | 8.16E-16 | | New |
| 737 | 12 | 12-114298592-TCCTC-T | rs1371643882 | LINC02459, TBX5 | 0.0018 | T | | 1.76E-11 | ?+ | 1.756E-11 | | New |
| 738 | 12 | 12-11567230-GGT-G | rs1491387339 | LINC01252 | 0.0026 | G | | 7E-14 | ?+ | 7E-14 | 0.115 | New |
| 739 | 12 | 12-117081702-G-T | rs1282630843 | TESC | 0.0014 | T | 0.961506 | 9.82E-12 | ++ | 1.008E-08 | 0.9359 | New |
| 740 | 12 | 12-120311520-C-T | rs1482211676 | SIRT4 | 0.0005 | C | | 6.35E-10 | ?+ | 6.353E-10 | | New |
| 741 | 12 | 12-120556370-TAAAAA-T | | NA | 0.001 | T | | 3.11E-10 | ?+ | 3.111E-10 | | New |

| | | | | | | | | | | |
|---|---|---|---|---|---|---|---|---|---|---|
| 742 | 12 | 12-120663475-C-T | rs1880782601 | CABP1 | 0.0014 | T | 0.438428 | 7.21E-14 | ++ | 6.297E-14 | | New |
| 742 | 12 | 12-120663495-G-C | rs1039962800 | CABP1 | 0.0018 | G | | 1.96E-15 | ?+ | 1.962E-15 | 0.2338 | New |
| 743 | 12 | 12-121034133-G-C | rs1592939473 | OASL | 0.0007 | C | | 3.4E-12 | ?+ | 3.398E-12 | | New |
| 744 | 12 | 12-121034789-G-C | rs1366382657 | OASL | 0.0024 | G | 0.143081 | 7.85E-20 | ++ | 3.316E-19 | | New |
| 745 | 12 | 12-121076600-T-TG | | OASL, AC079602.3 | 0.0503 | T | | 2.78E-11 | ?+ | 2.776E-11 | | New |
| 746 | 12 | 12-121617997-C-CCA | rs1555321571 | RNU6-1004P, ORAI1 | 0.0021 | C | | 1.78E-09 | ?+ | 1.776E-09 | 0.8399 | New |
| 747 | 12 | 12-121634293-G-A | rs1230863855 | ORAI1 | 0.0033 | A | 0.250878 | 8.14E-25 | ++ | 5.033E-25 | 0.9507 | New |
| 748 | 12 | 12-121670590-C-CCT | rs1408180684 | MORN3 | 0.006 | C | 0.77022 | 4.58E-43 | ++ | 2.111E-40 | | New |
| 749 | 12 | 12-121726344-C-CCGATGCAGGTGGATCA | | NA | 0.0015 | C | | 1.53E-15 | ?+ | 1.528E-15 | | New |
| 750 | 12 | 12-121745498-C-CT | rs1555331243 | TMEM120B | 0.0006 | C | | 1.11E-09 | ?+ | 1.111E-09 | | New |
| 751 | 12 | 12-121876914-TGA-T | | NA | 0.0004 | T | | 2.93E-08 | ?+ | 2.927E-08 | | New |
| 752 | 12 | 12-121914347-C-CT | | NA | 0.0013 | CT | | 3.05E-10 | ?+ | 3.047E-10 | | New |
| 753 | 12 | 12-122320066-ATATTGAGACCATCCTG-A | rs1353668650 | CLIP1 | 0.0012 | A | | 4.76E-08 | ?+ | 4.765E-08 | | New |
| 754 | 12 | 12-123173072-C-T | rs967043661 | MPHOSPH9 | 0.0015 | T | 0.969602 | 1.87E-13 | ++ | 8.057E-11 | | New |
| 754 | 12 | 12-123173111-G-A | rs557051451 | MPHOSPH9 | 0.0011 | G | 0.62016 | 1.03E-15 | ++ | 7.335E-11 | 0.5216 | New |
| 755 | 12 | 12-12363724-G-A | rs1168684570 | BORCS5 | 0.0006 | G | | 8.16E-09 | ?+ | 8.158E-09 | | New |
| 756 | 12 | 12-123717186-G-T | rs1310494836 | AC117503.5 | 0.0008 | G | 0.287021 | 1.67E-11 | ++ | 5.019E-09 | | New |
| 757 | 12 | 12-123755449-TGG-T | | NA | 0.0007 | TGG | | 3.67E-09 | ?+ | 3.669E-09 | | New |
| 758 | 12 | 12-123755925-C-CA | | NA | 0.0009 | C | | 1.06E-13 | ?+ | 1.064E-13 | | New |

| | | | | | | | | | | | | |
|---|---|---|---|---|---|---|---|---|---|---|---|---|
| 758 | 12 | 12-123755941-C-T | rs562305514 | ATP6V0A2 | 0.0004 | T | 0.49587 | 4.06E-10 | ++ | 7.919E-09 | | New |
| 759 | 12 | 12-124547213-G-A | rs565046070 | NCOR2 | 0.0006 | A | 0.457873 | 2.04E-11 | ++ | 1.783E-11 | | New |
| 759 | 12 | 12-124547226-C-CAA | | NCOR2 | 0.0014 | C | | 2.31E-14 | ?+ | 2.313E-14 | | New |
| 760 | 12 | 12-124951817-C-CA | | NA | 0.0002 | CA | | 2.44E-08 | ?+ | 2.438E-08 | | New |
| 761 | 12 | 12-125090208-T-TCATCCATCCATCCTCCATTCATCATCCCTCATCCATCATTTATCCAACTATCC | | AACS | 0.0465 | TCATCCATCCATCCTCC | | 1.15E-13 | ?+ | 1.154E-13 | | New |
| 762 | 12 | 12-12553080-C-CTCT | | NA | 0.0002 | CTCT | | 2.14E-08 | ?+ | 2.143E-08 | | New |
| 763 | 12 | 12-125803495-G-A | rs1042663288 | TMEM132B, AC005252.2 | 0.0004 | A | 0.331749 | 3.65E-09 | ++ | 4.722E-09 | | New |
| 764 | 12 | 12-128596107-C-CG | rs1875936746 | TMEM132C | 0.0017 | C | 0.999769 | 3.27E-14 | ++ | 1.039E-09 | | New |
| 765 | 12 | 12-128681911-C-CTG | rs1467612191 | TMEM132C | 0.0008 | CTG | | 1.39E-08 | ?+ | 1.388E-08 | | New |
| 766 | 12 | 12-130798250-CA-C | | STX2 | 0.0761 | C | | 1.75E-08 | ?+ | 1.748E-08 | | New |
| 767 | 12 | 12-132381811-AGCACCCGATACACAGAAGACAGAAGACATACTCAGCTAAAGCT-A | | NA | 0.0011 | AGCACCCGATACACAG | | 2.71E-09 | ?+ | 2.707E-09 | | New |
| 768 | 12 | 12-18232057-G-A | | NONE, PIK3C2G | 0.0004 | G | | 4.46E-08 | ?+ | 4.457E-08 | | New |
| 769 | 12 | 12-20353414-GGAACCC-G | rs1425955889 | LINC02468, AC129102.1 | 0.0045 | GGAACCC | | 4.49E-15 | ?+ | 4.492E-15 | 0.3619 | New |
| 770 | 12 | 12-26332904-TGG-T | | NA | 0.0075 | TGG | | 2.08E-08 | ?- | 2.079E-08 | | New |
| 771 | 12 | 12-27784354-G-C | rs1271765809 | KLHL42 | 0.0005 | G | 0.909335 | 4.34E-11 | -+ | 1.245E-08 | | New |

| | | | | | | | | | | | |
|---|---|---|---|---|---|---|---|---|---|---|---|
| 772 | 12 | 12-2946592-C-CTT | rs1221147047 | TULP3, RNU6-1315P | 0.0034 | CTT | | 8.43E-16 | ?+ | 8.429E-16 | | New |
| 773 | 12 | 12-31366840-ACC-A | | NA | 0.0018 | ACC | | 3.8E-13 | ?+ | 3.798E-13 | | New |
| 774 | 12 | 12-31549775-ACC-A | | NA | 0.0002 | ACC | | 6.27E-09 | ?+ | 6.268E-09 | | New |
| 775 | 12 | 12-32797701-G-A | rs1456374074 | PKP2 | 0.0008 | A | | 1.25E-08 | ?+ | 1.249E-08 | 0.3703 | New |
| 775 | 12 | 12-32797705-G-A | rs527605160 | PKP2 | 0.0007 | A | 0.875687 | 1.5E-09 | -+ | 9.433E-09 | 0.1634 | New |
| 776 | 12 | 12-37359433-TTCTG-T | rs1385472106 | NONE, RF01518 | 0.0017 | T | 0.392028 | 4.9E-13 | ++ | 3.383E-13 | 0.3236 | New |
| 777 | 12 | 12-37831033-GT-G | | NA | 0.0004 | GT | | 1.37E-09 | ?+ | 1.372E-09 | | New |
| 778 | 12 | 12-41796537-C-T | | LINC02400, AC090630.1 | 0.0001 | T | 0.935215 | 0.144834 | +- | 2.34E-223 | 0.5567 | New |
| 779 | 12 | 12-42207674-C-CTT | | NA | 0.001 | CTT | | 2.55E-09 | ?+ | 2.548E-09 | | New |
| 780 | 12 | 12-48988805-C-CT | | NA | 0.0006 | CT | | 3.28E-11 | ?+ | 3.279E-11 | | New |
| 781 | 12 | 12-49515961-G-A | rs1179715974 | SPATS2 | 0.003 | G | 0.034456 | 1.02E-15 | ++ | 8.412E-10 | 0.7579 | New |
| 782 | 12 | 12-49611726-GT-G | rs1403284517 | PRPF40B | 0.0068 | GT | | 6.22E-10 | ?+ | 6.218E-10 | 0.9896 | New |
| 783 | 12 | 12-50015024-C-CAG | | NA | 0.001 | C | | 6.21E-09 | ?+ | 6.214E-09 | | New |
| 784 | 12 | 12-50015583-C-CCT | | NA | 0.0014 | CCT | | 1.17E-08 | ?+ | 1.172E-08 | | New |
| 785 | 12 | 12-50234243-C-CT | | NA | 0.0002 | CT | | 5.7E-09 | ?+ | 5.704E-09 | | New |
| 786 | 12 | 12-50301831-GAGGC-G | | NA | 0.0009 | G | | 1.48E-08 | ?+ | 1.475E-08 | | New |
| 786 | 12 | 12-50301840-GGGCA-G | | NA | 0.0008 | GGGCA | | 3.1E-08 | ?+ | 3.105E-08 | | New |
| 786 | 12 | 12-50301871-C-T | rs1179018119 | LIMA1, FAM186A | 0.0004 | C | | 1.73E-08 | ?+ | 1.726E-08 | | New |
| 787 | 12 | 12-51720075-CA-C | | SCN8A | 0.2618 | CA | | 5.86E-09 | ?- | 5.862E-09 | | New |
| 788 | 12 | 12-53434594-TTG-T | | NA | 0.0003 | TTG | | 8.32E-09 | ?+ | 8.319E-09 | | New |

| | | | | | | | | | | | | |
|---|---|---|---|---|---|---|---|---|---|---|---|---|
| 789 | 12 | 12-53443104-ACCCG-A | | NA | 0.0003 | ACCCG | | 2.88E-08 | ?+ | 2.878E-08 | | New |
| 790 | 12 | 12-53529371-T-A | rs1458520743 | ATF7, ATF7-NPFF | 0.0014 | A | | 2.45E-10 | ?+ | 2.449E-10 | 0.7004 | New |
| 791 | 12 | 12-53537240-TG-T | | NA | 0.0002 | T | | 3.17E-11 | ?+ | 3.166E-11 | | New |
| 792 | 12 | 12-54336354-C-CTA | | NA | 0.001 | C | | 1.12E-11 | ?+ | 1.123E-11 | | New |
| 793 | 12 | 12-55916145-G-A | rs993749175 | PYM1 | 0.0011 | A | 0.285354 | 3.28E-15 | ++ | 1.335E-10 | 0.05956 | New |
| 794 | 12 | 12-56536657-TGGTG-T | | NA | 0.0005 | T | | 1.83E-09 | ?+ | 1.83E-09 | | New |
| 795 | 12 | 12-56656740-T-A | rs1951611943 | ATP5F1B, PTGES3 | 0.001 | A | | 1.35E-19 | ?+ | 1.348E-19 | | New |
| 795 | 12 | 12-56656759-G-C | | ATP5F1B, PTGES3 | 0.0005 | C | | 1.63E-10 | ?+ | 1.634E-10 | | New |
| 796 | 12 | 12-57017048-C-CTT | | NA | 0.0015 | CTT | | 3.33E-12 | ?+ | 3.327E-12 | | New |
| 797 | 12 | 12-64492451-C-CATG | | NA | 0.0006 | C | | 3.13E-10 | ?+ | 3.133E-10 | | New |
| 798 | 12 | 12-64614426-C-CTGTTTCTTAAAATTTCAACT | | NA | 0.0001 | CTGTTTCTTAAAATTTC | | 3.58E-08 | ?+ | 3.58E-08 | | New |
| 799 | 12 | 12-66938156-TTG-T | rs2041518089 | GRIP1 | 0.001 | T | | 3.48E-10 | ?+ | 3.48E-10 | | New |
| 800 | 12 | 12-73182069-TA-T | | NA | 0.0009 | T | | 1.78E-09 | ?+ | 1.783E-09 | | New |
| 801 | 12 | 12-762205-AGG-A | | NA | 0.0004 | AGG | | 1.41E-08 | ?+ | 1.408E-08 | | New |
| 801 | 12 | 12-762206-C-CAT | | NA | 0.0004 | C | | 4.42E-09 | ?+ | 4.416E-09 | | New |
| 802 | 12 | 12-7660652-AAG-A | rs1376194951 | APOBEC1 | 0.001 | A | | 3.09E-11 | ?+ | 3.094E-11 | 0.8001 | New |
| 803 | 12 | 12-7734982-C-CTT | | NA | 0.0044 | C | | 2.15E-21 | ?+ | 2.153E-21 | 0.1825 | New |
| 804 | 12 | 12-8009913-G-A | rs976572617 | RF00019, AC006511.4 | 0.0012 | G | | 8.29E-11 | ?+ | 8.293E-11 | | New |
| 805 | 12 | 12-89389651-G-T | rs1302697184 | AC010201.3 | 0.0007 | T | | 1.51E-13 | ?+ | 1.513E-13 | | New |
| 806 | 12 | 12-92835051-TC-T | rs1397586721 | EEA1 | 0.0024 | T | | 1.63E-14 | ?+ | 1.627E-14 | 0.5344 | New |

| | | | | | | | | | | | | |
|---|---|---|---|---|---|---|---|---|---|---|---|---|
| 807 | 12 | 12-94946026-AT-A | | NA | 0.0007 | A | | 1.52E-09 | ?+ | 1.516E-09 | | New |
| 808 | 12 | 12-95612573-C-CAT | rs1461072994 | USP44, RF00019 | 0.0035 | C | | 1.7E-12 | ?+ | 1.696E-12 | 0.4663 | New |
| 809 | 13 | 13-100092421-TG-T | rs1196563547 | PCCA | 0.001 | T | | 2.74E-08 | ?+ | 2.741E-08 | | New |
| 810 | 13 | 13-110656083-ACC-A | | CARS2 | 0.0006 | A | | 4.8E-11 | ?+ | 4.802E-11 | | New |
| 811 | 13 | 13-111960813-C-CGATG | | LINC00354, RF00287 | 0.1705 | C | | 1.5E-08 | ?+ | 1.501E-08 | | New |
| 812 | 13 | 13-113600427-C-T | rs9604171 | TFDP1 | 0.0138 | C | | 1.63E-09 | ?+ | 1.631E-09 | | New |
| 813 | 13 | 13-113795470-AC-A | rs1401920475 | TMEM255B | 0.0039 | A | | 1.2E-11 | ?+ | 1.203E-11 | | New |
| 814 | 13 | 13-114113552-C-CCAATCCACCCACCACGGCGAGTGATGTCCGTACAGCTCACACCGCATCCAT | rs1566582849 | RASA3 | 0.0103 | C | | 4.61E-10 | ?+ | 4.614E-10 | | New |
| 815 | 13 | 13-19187520-TGG-T | rs1306405136 | TUBA3C, AL139327.2 | 0.0014 | TGG | | 4.28E-08 | ?+ | 4.281E-08 | 0.4347 | New |
| 816 | 13 | 13-19922626-C-CCT | rs1485351806 | AL355001.2, ZMYM2 | 0.003 | CCT | | 2.76E-19 | ?+ | 2.76E-19 | 0.6312 | New |
| 817 | 13 | 13-20470690-C-CCT | | NA | 0.0013 | C | | 1.62E-11 | ?+ | 1.621E-11 | | New |
| 818 | 13 | 13-21627637-ATT-A | | NA | 0.0012 | A | | 3.39E-09 | ?+ | 3.392E-09 | | New |
| 819 | 13 | 13-27054320-G-C | rs1392734500 | AL160035.1, USP12 | 0.0009 | C | 0.223738 | 4.54E-09 | ++ | 1.039E-08 | 0.8133 | New |
| 820 | 13 | 13-40685666-C-A | rs1033826869 | FOXO1, MIR320D1 | 0.0007 | C | 0.893019 | 2.91E-10 | ++ | 2.055E-09 | | New |
| 821 | 13 | 13-41850639-C-T | rs1327088179 | VWA8 | 0.0009 | C | 0.283844 | 5.41E-15 | ++ | 5.675E-15 | | New |
| 822 | 13 | 13-42917642-G-A | rs199595584 | EPSTI1 | 0.0031 | A | | 1.97E-09 | ?- | 1.971E-09 | | New |
| 823 | 13 | 13-44569304-G-C | rs73465766 | TSC22D1 | 0.0038 | C | 0.541382 | 1.77E-10 | -- | 2.476E-08 | | New |

| | | | | | | | | | | | | |
|---|---|---|---|---|---|---|---|---|---|---|---|---|
| 824 | 13 | 13-45383701-G-C | rs111407307 | TPT1-AS1 | 0.0102 | C | | 9.88E-17 | ?+ | 9.878E-17 | | New |
| 825 | 13 | 13-49525435-ATG-A | | NA | 0.0004 | A | | 2.58E-10 | ?+ | 2.576E-10 | | New |
| 825 | 13 | 13-49525439-AAG-A | | NA | 0.0004 | AAG | | 2.11E-09 | ?+ | 2.115E-09 | | New |
| 826 | 13 | 13-79091376-G-A | rs1435170216 | LINC00331, RBM26 | 0.0025 | G | | 9.48E-09 | ?- | 9.475E-09 | 0.4807 | New |
| 827 | 13 | 13-82155975-G-A | rs1017075640 | AL353633.1, AL445255.1 | 0.0004 | A | | 3.41E-08 | ?+ | 3.413E-08 | | New |
| 828 | 13 | 13-97948951-C-CTG | rs1202324392 | AL356580.1 | 0.0006 | C | | 4.99E-08 | ?+ | 4.989E-08 | | New |
| 829 | 13 | 13-98116951-GGA-G | rs1304078336 | IPO5, FARP1 | 0.0026 | G | | 7.17E-14 | ?+ | 7.169E-14 | 0.3997 | New |
| 830 | 13 | 13-98643115-C-A | rs577038368 | STK24-AS1, RN7SL60P | 0.0011 | C | 0.732376 | 7.97E-13 | ++ | 3.071E-12 | | New |
| 831 | 13 | 13-99952110-C-CTT | rs2052996149 | AL137139.1 | 0.0026 | CTT | | 3.18E-20 | ?+ | 3.18E-20 | | New |
| 831 | 13 | 13-99952112-C-CAG | rs1286972096 | AL137139.1 | 0.0025 | C | | 5.3E-26 | ?+ | 5.302E-26 | | New |
| 831 | 13 | 13-99952132-AT-A | | NA | 0.0013 | A | | 1.22E-10 | ?+ | 1.223E-10 | | New |
| 832 | 14 | 14-100301031-C-A | rs1167605935 | SLC25A29 | 0.0009 | A | 0.70354 | 2.21E-11 | ++ | 2.826E-10 | 0.3792 | New |
| 833 | 14 | 14-102136332-GCT-G | | NA | 0.0003 | GCT | | 2.2E-09 | ?+ | 2.2E-09 | | New |
| 834 | 14 | 14-102272953-C-CCT | | NA | 0.0001 | CCT | 0.222708 | 8.19E-08 | ++ | 3.836E-08 | | New |
| 835 | 14 | 14-102541608-AG-A | rs1389132849 | MIR4309, LINC02323 | 0.0012 | AG | | 5.04E-15 | ?+ | 5.043E-15 | 0.1863 | New |
| 836 | 14 | 14-103272252-C-CCT | | NA | 0.0004 | C | | 5.36E-10 | ?+ | 5.36E-10 | | New |
| 837 | 14 | 14-103601925-GGT-G | rs1290948092 | AL139300.1, KLC1 | 0.0034 | GGT | | 2.12E-11 | ?+ | 2.115E-11 | 0.2817 | New |
| 837 | 14 | 14-103601927-C-CAG | rs1385983626 | AL139300.1, KLC1 | 0.0038 | C | | 1.52E-11 | ?+ | 1.523E-11 | | New |
| 838 | 14 | 14-103655446-GCA-G | rs1444368239 | AL139300.1, KLC1 | 0.0026 | GCA | | 1.67E-12 | ?+ | 1.674E-12 | 0.2188 | New |

| | | | | | | | | | | | | |
|---|---|---|---|---|---|---|---|---|---|---|---|---|
| 839 | 14 | 14-104209450-G-C | rs1223739912 | KIF26A, LINC02691 | 0.0005 | G | | 1.32E-09 | ?+ | 1.321E-09 | | New |
| 840 | 14 | 14-104847993-G-T | rs186218196 | AL583810.2, AL583810.1 | 0.0011 | T | 0.272319 | 4.92E-16 | -+ | 1.934E-13 | | New |
| 841 | 14 | 14-104951987-G-T | rs770911369 | AHNAK2 | 0.0012 | G | 0.333787 | 4.41E-08 | ++ | 3.788E-08 | | New |
| 842 | 14 | 14-105296067-C-CAA | | NA | 0.0014 | CAA | | 1.48E-16 | ?+ | 1.485E-16 | | Old |
| 842 | 14 | 14-105296075-T-C | rs4347564 | BRF1 | 0.0025 | C | | 6.77E-13 | ?+ | 6.768E-13 | | Old |
| 843 | 14 | 14-105650853-C-CATGGAACAGGAAGCATCCAGGATGGAACAGGAAGCATCCAGT | rs1265782083 | AL928742.1, IGHA1 | 0.0135 | C | | 1.21E-08 | ?+ | 1.212E-08 | | Old |
| 844 | 14 | 14-20342296-C-CTCG | | AL355075.4, RPPH1 | 0.0012 | C | | 3.35E-15 | ?+ | 3.351E-15 | 0.2569 | New |
| 845 | 14 | 14-22727949-C-T | rs2038153903 | AL160314.2 | 0.0003 | C | 0.392326 | 1.36E-10 | -+ | 2.329E-08 | | New |
| 846 | 14 | 14-22801564-ACT-A | | NA | 0.001 | ACT | | 1.26E-11 | ?+ | 1.259E-11 | | New |
| 847 | 14 | 14-23706575-G-A | rs1482168704 | AL160237.1 | 0.0037 | A | 0.097764 | 5.57E-19 | ++ | 1.978E-19 | 0.9641 | New |
| 847 | 14 | 14-23706600-G-A | rs903337086 | AL160237.1 | 0.0053 | A | 0.869328 | 1.57E-23 | -+ | 3.728E-11 | 0.4594 | New |
| 847 | 14 | 14-23706607-G-A | rs146896044 | AL160237.1 | 0.0084 | A | 0.806871 | 1.3E-24 | ++ | 1.56E-14 | | New |
| 848 | 14 | 14-24170048-GT-G | rs1281146788 | IRF9, REC8 | 0.0008 | G | | 1.27E-08 | ?+ | 1.266E-08 | | New |
| 849 | 14 | 14-28854133-GGC-G | | NA | 0.0013 | G | | 7.29E-11 | ?+ | 7.293E-11 | | New |
| 849 | 14 | 14-28854137-C-CGT | | NA | 0.0009 | C | | 1.83E-08 | ?+ | 1.83E-08 | | New |
| 849 | 14 | 14-28854140-GCCCA-G | | NA | 0.0008 | GCCCA | | 2E-08 | ?+ | 2.001E-08 | | New |
| 850 | 14 | 14-31245601-G-A | rs142188527 | RF00019 | 0.3339 | A | | 7.97E-10 | ?+ | 7.967E-10 | 0.5199 | New |
| 851 | 14 | 14-34487122-TA-T | rs1264221554 | SPTSSA, EAPP | 0.0006 | T | | 1.89E-09 | ?+ | 1.894E-09 | 0.1006 | New |

| | | | | | | | | | | | | |
|---|---|---|---|---|---|---|---|---|---|---|---|---|
| 851 | 14 | 14-34487130-TTC-T | rs1175681088 | SPTSSA, EAPP | 0.0011 | T | | 3.99E-11 | ?+ | 3.993E-11 | 0.1005 | New |
| 852 | 14 | 14-34514025-C-CGTG | | NA | 0.0003 | C | | 1.5E-10 | ?+ | 1.497E-10 | | New |
| 853 | 14 | 14-34565234-C-T | rs1012535274 | SNX6 | 0.003 | T | 0.071581 | 3.05E-16 | ++ | 7.185E-09 | 0.3573 | New |
| 853 | 14 | 14-34565238-C-T | rs376690477 | SNX6 | 0.0031 | T | 0.048577 | 5.15E-14 | ++ | 3.009E-08 | 0.4979 | New |
| 853 | 14 | 14-34565241-C-T | rs1370017161 | SNX6 | 0.003 | T | 0.069 | 1.36E-18 | ++ | 1.534E-10 | | New |
| 854 | 14 | 14-39101400-ACC-A | | NA | 0.002 | ACC | | 2.71E-12 | ?+ | 2.706E-12 | | Old |
| 855 | 14 | 14-54925838-C-A | rs147252057 | GCH1, WDHD1 | 0.0004 | A | 0.957478 | 5.07E-09 | ++ | 1.654E-08 | | New |
| 856 | 14 | 14-55344909-TTGTC-T | rs1393967520 | FBXO34 | 0.0011 | T | 0.259637 | 6.86E-09 | ++ | 2.336E-08 | 0.2689 | New |
| 857 | 14 | 14-63519957-GCCAAGAAGAAC-G | | NA | 0.0018 | GCCAAGAAGAAC | | 3.64E-08 | ?+ | 3.636E-08 | | New |
| 858 | 14 | 14-63585644-G-T | rs374049962 | AL136038.3, WDR89 | 0.0015 | T | 0.241319 | 3.43E-17 | -+ | 4.064E-12 | 0.8818 | New |
| 858 | 14 | 14-63585648-G-T | rs773334844 | AL136038.3, WDR89 | 0.001 | T | 0.241182 | 2.18E-13 | -+ | 1.745E-08 | 0.9271 | New |
| 859 | 14 | 14-64547356-ACT-A | | HSPA2, PPP1R36 | 0.003 | A | | 1.33E-09 | ?+ | 1.328E-09 | 0.7422 | New |
| 860 | 14 | 14-65504716-ATTT-A | | NA | 0.0007 | A | | 2.32E-08 | ?+ | 2.322E-08 | | New |
| 861 | 14 | 14-66995017-TTA-T | | NA | 0.0012 | T | | 4.16E-12 | ?+ | 4.161E-12 | | New |
| 862 | 14 | 14-71326352-GCACA-G | | SIPA1L1 | 0.0011 | G | | 4.72E-10 | ?+ | 4.725E-10 | 0.1383 | New |
| 862 | 14 | 14-71326357-C-CGTGT | | SIPA1L1 | 0.0009 | CGTGT | | 1.19E-10 | ?+ | 1.189E-10 | | New |
| 863 | 14 | 14-72837563-C-CCCCA | | NA | 0.0015 | C | | 3.41E-08 | ?+ | 3.405E-08 | | Old |
| 864 | 14 | 14-73480223-C-CTT | | NA | 0.0005 | C | | 5.28E-09 | ?+ | 5.28E-09 | | Old |
| 865 | 14 | 14-73952289-ATT-A | rs1339702970 | COQ6 | 0.0015 | ATT | | 2.38E-14 | ?+ | 2.383E-14 | 0.5945 | New |
| 866 | 14 | 14-74333212-G-A | rs577330509 | VRTN | 0.0005 | A | 0.00637 | 1.31E-06 | ++ | 4.971E-08 | | New |

| | | | | | | | | | | | |
|---|---|---|---|---|---|---|---|---|---|---|---|
| 867 | 14 | 14-77850817-ATTTTTT-A | rs879739678 | ADCK1 | 0.0034 | A | | 7.88E-18 | ?+ | 7.884E-18 | | New |
| 868 | 14 | 14-78832014-G-A | rs1311911699 | NRXN3 | 0.0013 | G | 0.307203 | 2.46E-27 | -+ | 1.01E-22 | 0.09501 | New |
| 869 | 14 | 14-81923555-AATTTT-A | rs1378802782 | AL355838.1 | 0.0045 | A | 0.818303 | 3.23E-13 | -+ | 1.223E-09 | 0.8469 | Old |
| 870 | 14 | 14-88557948-C-T | rs1217460643 | AL162171.1, ZC3H14 | 0.0012 | C | 0.167637 | 2.71E-11 | ++ | 1.291E-10 | | New |
| 870 | 14 | 14-88557971-G-A | rs1299853547 | AL162171.1, ZC3H14 | 0.0014 | A | | 5.47E-10 | ?+ | 5.467E-10 | | New |
| 871 | 14 | 14-91460172-C-CCT | | NA | 0.0008 | CCT | | 1.45E-08 | ?+ | 1.454E-08 | | New |
| 871 | 14 | 14-91460178-ATG-A | | NA | 0.0005 | A | | 3.83E-08 | ?+ | 3.829E-08 | | New |
| 872 | 14 | 14-91507096-ATCAC-A | | NA | 0.0007 | A | | 8.29E-15 | ?+ | 8.287E-15 | | New |
| 873 | 14 | 14-92741777-G-T | rs1044208812 | LGMN | 0.0005 | T | | 4.28E-14 | ?+ | 4.284E-14 | 0.5189 | Old |
| 874 | 14 | 14-92804358-GC-G | | NA | 0.0003 | GC | | 5.19E-12 | ?+ | 5.188E-12 | | Old |
| 874 | 14 | 14-92804364-C-CAACA | | NA | 0.0001 | CAACA | | 5.16E-09 | ?+ | 5.162E-09 | | Old |
| 874 | 14 | 14-92804368-C-CTT | | NA | 0.0002 | C | | 3.14E-08 | ?+ | 3.141E-08 | | Old |
| 875 | 15 | 15-23448602-C-CAT | | NA | 0.0013 | C | | 2.14E-09 | ?+ | 2.138E-09 | | New |
| 875 | 15 | 15-23448605-GCC-G | | NA | 0.0013 | GCC | | 5.06E-10 | ?+ | 5.065E-10 | | New |
| 875 | 15 | 15-23448633-AG-A | | NA | 0.0011 | AG | | 1.4E-11 | ?+ | 1.398E-11 | | New |
| 876 | 15 | 15-23838964-C-CCT | | NA | 0.0008 | C | | 2.08E-11 | ?+ | 2.081E-11 | | New |
| 877 | 15 | 15-25006938-G-A | rs1267131333 | SNHG14 | 0.0003 | A | 0.163188 | 2.01E-10 | ++ | 1.124E-10 | 0.4056 | New |
| 878 | 15 | 15-25293273-C-A | rs1191974147 | SNHG14 | 0.0007 | C | | 4.57E-08 | ?+ | 4.571E-08 | 0.1573 | New |
| 879 | 15 | 15-28367284-A-ATTTTT | | HERC2, GOLGA8F | 0.2881 | A | | 4.99E-08 | ?+ | 4.991E-08 | | New |

| | | | | | | | | | | |
|---|---|---|---|---|---|---|---|---|---|---|
| 880 | 15 | 15-28784698-C-T | rs56994395 | GOLGA8M, GOLGA6L7 | 0.0573 | C | 6.12E-09 | ?+ | 6.116E-09 | 0.5895 | New |
| 881 | 15 | 15-29739562-G-A | rs1464017073 | TJP1 | 0.0017 | A | 2.4E-17 | ?+ | 2.403E-17 | | New |
| 882 | 15 | 15-32275965-T-C | rs76918399 | AC068448.1, RNU6-18P | 0.0081 | T | 3.15E-10 | ?+ | 3.147E-10 | | New |
| 883 | 15 | 15-34175843-TAAGAA-T | | NA | 0.0016 | TAAGAA | 6.23E-14 | ?+ | 6.229E-14 | | New |
| 884 | 15 | 15-40502504-TCA-T | rs1172600966 | AC091045.1 | 0.002 | T | 2.07E-09 | ?+ | 2.073E-09 | 0.7762 | New |
| 885 | 15 | 15-40646627-G-C | | AC022405.1 | 0.0001 | C | 1.45E-09 | ?+ | 1.453E-09 | | New |
| 886 | 15 | 15-41344055-TA-T | | NA | 0.0007 | TA | 9.82E-14 | ?+ | 9.823E-14 | | New |
| 887 | 15 | 15-41378616-C-CAA | | NA | 0.0004 | CAA | 2.94E-09 | ?+ | 2.939E-09 | | New |
| 888 | 15 | 15-41525481-GGCAT-G | | NA | 0.002 | G | 2.12E-11 | ?+ | 2.121E-11 | | New |
| 889 | 15 | 15-43723919-G-T | rs1385582538 | RNU6-353P, RNU6-354P | 0.0007 | T | 8.28E-09 | ?+ | 8.28E-09 | 0.1803 | New |
| 890 | 15 | 15-44183022-TCTC-T | rs1291362090 | FRMD5 | 0.0007 | T | 3.65E-12 | ?+ | 3.647E-12 | 0.2447 | New |
| 891 | 15 | 15-44777318-G-T | rs1199092550 | AC122108.2 | 0.0012 | T | 4.41E-08 | ?+ | 4.414E-08 | 0.8628 | New |
| 891 | 15 | 15-44777319-TG-T | rs1255811814 | AC122108.2 | 0.0016 | TG | 2.62E-08 | ?+ | 2.62E-08 | 0.8559 | New |
| 892 | 15 | 15-52015126-G-A | rs1031529869 | MAPK6-DT | 0.0004 | A | 1.01E-10 | ?+ | 1.012E-10 | | New |
| 893 | 15 | 15-55441294-T-A | | DNAAF4-CCPG1 | 0.0003 | T | 3.34E-09 | ?+ | 3.337E-09 | | New |
| 894 | 15 | 15-55641955-ACC-A | | NA | 0.0006 | ACC | 4.71E-12 | ?+ | 4.713E-12 | 0.3157 | New |
| 895 | 15 | 15-63272088-C-CTT | | NA | 0.0006 | CTT | 2.66E-09 | ?+ | 2.659E-09 | | Old |
| 896 | 15 | 15-64372438-C-T | rs1371331260 | AC087632.1, PCLAF | 0.0048 | T | 8.06E-09 | ?+ | 8.063E-09 | 0.1559 | Old |
| 897 | 15 | 15-65454698-TG-T | | NA | 0.0007 | T | 1.55E-11 | ?+ | 1.546E-11 | | New |

| | | | | | | | | | | | | |
|---|---|---|---|---|---|---|---|---|---|---|---|---|
| 897 | 15 | 15-65454700-TAACTC-T | | NA | 0.0005 | TAACTC | | 2.07E-08 | ?+ | 2.065E-08 | | New |
| 898 | 15 | 15-74757553-AT-A | rs1330271270 | CYP1A2 | 0.0038 | A | | 1.38E-14 | ?+ | 1.376E-14 | 0.7961 | New |
| 899 | 15 | 15-74921556-C-CCG | rs1283490962 | COX5A | 0.0012 | C | | 6.93E-09 | ?+ | 6.931E-09 | | New |
| 900 | 15 | 15-74953400-G-C | | COX5A, RPP25 | 0.0003 | C | | 5.27E-10 | ?+ | 5.272E-10 | | New |
| 901 | 15 | 15-75575726-TGG-T | rs1295714207 | AC105036.3 | 0.0059 | T | 0.521318 | 2.2E-10 | ++ | 2.913E-10 | 0.277 | New |
| 902 | 15 | 15-76148723-CT-C | rs5813819 | TMEM266 | 0.0138 | C | | 2.81E-09 | ?+ | 2.806E-09 | | New |
| 903 | 15 | 15-76317437-G-A | rs1450411286 | ETFA, ISL2 | 0.0007 | A | 0.227395 | 6.05E-09 | ++ | 8.493E-09 | 0.2553 | New |
| 904 | 15 | 15-77699496-TCC-T | | NA | 0.0013 | T | | 7.29E-12 | ?+ | 7.295E-12 | | New |
| 905 | 15 | 15-78856334-TTA-T | | MORF4L1 | 0.0013 | TTA | | 3.64E-09 | ?+ | 3.636E-09 | | Old |
| 906 | 15 | 15-79951126-C-CAGAGCGG | rs1252579567 | ST20-AS1, BCL2A1 | 0.0028 | CAGAGCGG | | 2.86E-15 | ?+ | 2.858E-15 | | New |
| 907 | 15 | 15-89020231-G-C | rs1985691 | MFGE8, AC013565.1 | 0.3172 | G | | 3.95E-08 | ?+ | 3.945E-08 | 0.864 | New |
| 908 | 15 | 15-90157089-GCA-G | | NA | 0.0002 | GCA | | 4.57E-09 | ?+ | 4.573E-09 | | New |
| 908 | 15 | 15-90157090-TG-T | | NA | 0.0002 | TG | | 2.48E-09 | ?+ | 2.479E-09 | | New |
| 908 | 15 | 15-90157092-TA-T | | NA | 0.0002 | TA | | 7.95E-10 | ?+ | 7.952E-10 | | New |
| 909 | 15 | 15-90505467-C-CAA | rs1476023478 | IQGAP1, CRTC3 | 0.0021 | CAA | 0.362848 | 8.49E-13 | ++ | 1.977E-12 | | New |
| 910 | 15 | 15-92850975-AT-A | | NA | 0.0007 | A | | 2.51E-08 | ?+ | 2.513E-08 | 0.3821 | New |
| 911 | 15 | 15-99043874-C-T | rs1555476996 | AC036108.1 | 0.0003 | C | | 1.03E-09 | ?+ | 1.031E-09 | | Old |
| 912 | 16 | 16-10645333-G-A | rs1555465179 | TEKT5 | 0.0005 | G | | 1.13E-11 | ?+ | 1.127E-11 | | New |
| 912 | 16 | 16-10645335-C-A | rs568319760 | TEKT5 | 0.0003 | C | | 1.74E-08 | ?+ | 1.735E-08 | | New |
| 913 | 16 | 16-10734878-G-A | rs955981614 | TEKT5, NUBP1 | 0.0009 | A | | 2.94E-08 | ?+ | 2.936E-08 | | New |
| 914 | 16 | 16-11232146-TGC-T | | NA | 0.0002 | TGC | | 2.3E-08 | ?+ | 0.000000023 | | New |

| | | | | | | | | | | | | |
|---|---|---|---|---|---|---|---|---|---|---|---|---|
| 915 | 16 | 16-1190872-GTC-G | rs759417989 | CACNA1H | 0.0127 | G | | 2.39E-09 | ?+ | 2.389E-09 | | New |
| 916 | 16 | 16-14532898-GC-G | rs1438440014 | PARN | 0.0018 | GC | | 2.54E-08 | ?+ | 2.543E-08 | | New |
| 917 | 16 | 16-1478801-C-CCA | | NA | 0.0003 | C | | 3.98E-08 | ?+ | 3.98E-08 | | New |
| 918 | 16 | 16-15359814-GCACACACACACA-G | | NA | 0.0056 | G | | 5.39E-10 | ?+ | 5.388E-10 | | New |
| 919 | 16 | 16-18158393-T-A | | AC091489.1 | 0.0003 | A | | 6.69E-11 | ?+ | 6.686E-11 | | New |
| 920 | 16 | 16-199543-T-A | rs1350229578 | LUC7L | 0.001 | A | 0.940299 | 2.86E-11 | -+ | 9.954E-10 | 0.4181 | New |
| 921 | 16 | 16-20408737-GCA-G | | NA | 0.0022 | G | | 6.29E-16 | ?+ | 6.295E-16 | 0.954 | New |
| 922 | 16 | 16-21343238-C-CAAA | | CRYM, NPIPB3 | 0.0643 | CAAA | | 1.63E-08 | ?+ | 1.631E-08 | | New |
| 923 | 16 | 16-21894065-TTGAG-T | | NA | 0.0006 | T | | 6.36E-12 | ?+ | 6.358E-12 | | New |
| 924 | 16 | 16-23588279-G-A | rs945379630 | NDUFAB1 | 0.0007 | G | 0.468786 | 1.38E-12 | ++ | 1.5E-09 | | New |
| 925 | 16 | 16-23736646-AAGTGT-A | | NA | 0.0013 | AAGTGT | | 6.36E-14 | ?+ | 6.361E-14 | | New |
| 925 | 16 | 16-23736647-GGCACC-G | | NA | 0.001 | G | | 9.34E-12 | ?+ | 9.343E-12 | | New |
| 926 | 16 | 16-24905612-AGG-A | | NA | 0.0003 | AGG | | 2.01E-09 | ?+ | 2.01E-09 | | New |
| 927 | 16 | 16-25154176-AT-A | rs1248734336 | LCMT1 | 0.0017 | A | 0.457416 | 6.24E-09 | ++ | 3.505E-08 | | New |
| 928 | 16 | 16-28817462-C-CAT | | NA | 0.0013 | C | | 5.59E-15 | ?+ | 5.592E-15 | | New |
| 928 | 16 | 16-28817467-GCA-G | | NA | 0.0011 | GCA | | 2.65E-12 | ?+ | 2.647E-12 | | New |
| 929 | 16 | 16-2891973-C-CTT | | NA | 0.0026 | C | | 5.19E-12 | ?+ | 5.191E-12 | | New |
| 929 | 16 | 16-2891980-TTGGG-T | | NA | 0.002 | T | | 1.13E-15 | ?+ | 1.128E-15 | | New |
| 929 | 16 | 16-2891984-GCAAC-G | | NA | 0.0015 | GCAAC | | 5.94E-12 | ?+ | 5.942E-12 | | New |
| 930 | 16 | 16-28993307-G-A | rs111708809 | LAT, RRN3P2 | 0.2074 | G | | 6.38E-18 | ?+ | 6.379E-18 | | New |
| 931 | 16 | 16-29786163-C-CA | rs1567352683 | ZG16, KIF22 | 0.0008 | C | 0.061174 | 1.67E-13 | ++ | 1.015E-13 | | Old |

| | | | | | | | | | | | | |
|---|---|---|---|---|---|---|---|---|---|---|---|---|
| 932 | 16 | 16-29831442-C-CCG | | NA | 0.0006 | CCG | | 2.66E-15 | ?+ | 2.659E-15 | | Old |
| 933 | 16 | 16-2983500-C-CA | | AC004233.3, LINC00514 | 0.0013 | CA | | 2.1E-09 | ?+ | 2.099E-09 | | New |
| 934 | 16 | 16-30142456-AGG-A | | NA | 0.0005 | AGG | | 2.92E-08 | ?+ | 2.919E-08 | | Old |
| 935 | 16 | 16-30441816-C-CAG | | NA | 0.0011 | C | | 4.88E-12 | ?+ | 4.883E-12 | | Old |
| 936 | 16 | 16-30625405-C-CCT | rs2052164865 | ZNF689, PRR14 | 0.0021 | CCT | | 6.82E-10 | ?+ | 6.816E-10 | | Old |
| 937 | 16 | 16-311114-G-GCT | rs58810411 | AXIN1 | 0.0008 | GCT | | 7.89E-11 | ?+ | 7.889E-11 | | New |
| 938 | 16 | 16-32421947-CA-C | | AC133485.2, AC138915.3 | 0.4383 | CA | | 2.54E-14 | ?+ | 2.539E-14 | | New |
| 939 | 16 | 16-34249700-C-CTT | | NA | 0.0003 | C | | 3.97E-08 | ?+ | 3.974E-08 | | New |
| 940 | 16 | 16-34260272-C-A | rs1394896178 | LINC00273, AC135776.4 | 0.0007 | C | | 1.23E-12 | ?+ | 1.235E-12 | 0.7918 | New |
| 941 | 16 | 16-34284927-TTC-T | | NA | 0.0025 | T | | 5.69E-12 | ?+ | 5.691E-12 | | New |
| 942 | 16 | 16-3448877-C-T | | AC025283.3, NAA60 | 0.0001 | T | 0.573237 | 0.874336 | -+ | 1.935E-16 | 0.8884 | New |
| 943 | 16 | 16-34574822-TTGAAAACAAA-T | | NA | 0.0045 | TTGAAAACAAA | | 1.16E-08 | ?+ | 1.159E-08 | | New |
| 944 | 16 | 16-34630340-AGTCCATT-A | | NA | 0.0052 | A | | 2.44E-46 | ?+ | 2.439E-46 | | New |
| 945 | 16 | 16-35901414-G-A | rs1596645121 | AC106785.2, AC116553.2 | 0.0006 | G | | 1.7E-10 | ?+ | 1.704E-10 | | New |
| 945 | 16 | 16-35901425-G-A | rs1462977024 | AC106785.2, AC116553.2 | 0.0006 | G | | 1.67E-09 | ?+ | 1.674E-09 | | New |

| | | | | | | | | | | | | |
|---|---|---|---|---|---|---|---|---|---|---|---|---|
| 945 | 16 | 16-35901442-G-A | rs1391812258 | AC106785.2, AC116553.2 | 0.0003 | A | 0.319765 | 1.73E-08 | ++ | 1.089E-08 | | New |
| 946 | 16 | 16-36256874-TAC-T | | NA | 0.0018 | TAC | | 6.11E-11 | ?+ | 6.112E-11 | | New |
| 947 | 16 | 16-3923860-G-C | | CREBBP, AC005736.1 | 0.0004 | G | | 1.14E-09 | ?+ | 1.138E-09 | | New |
| 948 | 16 | 16-422947-C-CAG | | NA | 0.0005 | CAG | | 1.64E-08 | ?+ | 1.637E-08 | | New |
| 949 | 16 | 16-4590542-GA-G | | NA | 0.0005 | G | | 3.9E-08 | ?+ | 3.899E-08 | | New |
| 950 | 16 | 16-46395598-T-C | rs10221139 | NONE, RNU6-845P | 0.0006 | T | | 7.84E-11 | ?+ | 7.835E-11 | | New |
| 950 | 16 | 16-46395604-A-C | rs13337595 | NONE, RNU6-845P | 0.0007 | A | | 1.49E-12 | ?+ | 1.489E-12 | | New |
| 951 | 16 | 16-4771009-C-CAA | | NA | 0.0008 | CAA | | 2.94E-09 | ?+ | 2.94E-09 | | New |
| 952 | 16 | 16-50150533-ATT-A | rs2037732492 | AC007610.5, RF00156 | 0.0021 | A | | 2.23E-18 | ?+ | 2.232E-18 | | New |
| 953 | 16 | 16-58440874-C-CAT | rs1407236131 | LINC02137 | 0.0005 | CAT | | 4.63E-08 | ?+ | 4.633E-08 | 0.3518 | New |
| 954 | 16 | 16-619328-ATG-A | rs1244433189 | RAB40C | 0.0014 | A | | 6.54E-09 | ?+ | 6.542E-09 | | New |
| 955 | 16 | 16-67096756-C-CCT | | NA | 0.0004 | CCT | | 7.72E-09 | ?+ | 7.722E-09 | | New |
| 956 | 16 | 16-67606533-G-A | rs57337864 | CTCF | 0.0446 | A | | 1.67E-17 | ?+ | 1.668E-17 | 0.3788 | New |
| 957 | 16 | 16-67610589-C-CCG | | NA | 0.0008 | CCG | 0.659432 | 4.77E-09 | -+ | 2.584E-08 | | New |
| 958 | 16 | 16-67849799-C-T | rs1461907371 | NUTF2 | 0.0004 | T | 0.243695 | 8.89E-11 | -+ | 2.461E-08 | | New |
| 958 | 16 | 16-67849804-G-A | rs763087992 | NUTF2 | 0.0005 | A | 0.430058 | 6.96E-14 | ++ | 1.328E-12 | | New |
| 959 | 16 | 16-69576084-C-CTT | | NA | 0.0009 | CTT | | 6.03E-10 | ?+ | 6.033E-10 | | New |
| 960 | 16 | 16-70218561-CA-C | | CLEC18C, EXOSC6 | 0.4067 | C | | 1.05E-08 | ?- | 1.047E-08 | | Old |

| | | | | | | | | | | | | |
|---|---|---|---|---|---|---|---|---|---|---|---|---|
| 961 | 16 | 16-70384477-G-C | rs1267581371 | AC012184.1 | 0.0013 | C | 0.011317 | 9.52E-15 | ++ | 3.742E-16 | 0.6082 | Old |
| 962 | 16 | 16-73895079-AG-A | | NA | 0.0003 | A | | 4.07E-08 | ?+ | 4.07E-08 | | New |
| 963 | 16 | 16-74615312-C-CAG | | NA | 0.001 | CAG | | 1.57E-08 | ?+ | 1.57E-08 | | New |
| 964 | 16 | 16-754494-C-CA | | NA | 0.001 | CA | | 1.4E-12 | ?+ | 1.403E-12 | 0.1498 | New |
| 965 | 16 | 16-81105873-GGTC-G | | NA | 0.0035 | GGTC | 0.34038 | 1.01E-20 | ++ | 1.385E-19 | | New |
| 966 | 16 | 16-82658267-T-A | rs2150920593 | CDH13 | 0.0006 | A | 0.004692 | 5.68E-08 | ++ | 9.821E-10 | | New |
| 966 | 16 | 16-82658281-T-A | rs1174607704 | CDH13 | 0.0004 | A | 0.004692 | 6.95E-08 | ++ | 1.304E-09 | | New |
| 967 | 16 | 16-85583635-G-A | | GSE1 | 0.0001 | A | 0.51605 | 0.977114 | -- | 4.481E-10 | | New |
| 968 | 16 | 16-87884404-G-C | rs1013953075 | CA5A | 0.0005 | C | 0.959602 | 6.13E-09 | ++ | 4.858E-08 | | New |
| 969 | 16 | 16-89188362-C-CGG | rs199638393 | CDH15 | 0.0027 | C | | 6.86E-10 | ?+ | 6.861E-10 | | New |
| 970 | 16 | 16-89479635-C-A | rs368776529 | ANKRD11 | 0.0006 | A | | 1.89E-08 | ?+ | 1.885E-08 | | New |
| 971 | 16 | 16-89650787-C-CAG | | NA | 0.0008 | CAG | | 1.25E-10 | ?+ | 1.252E-10 | | Old |
| 972 | 16 | 16-89831610-G-A | rs544301146 | SPIRE2 | 0.002 | G | | 4.28E-10 | ?+ | 4.275E-10 | | Old |
| 973 | 16 | 16-89879111-TTGTCCGTGTACACAGATGGGCTTCGGGGCCTGTCATACGTGCTGTTCGTGTACACAGACGGGCTCCTGGGCCTATCACCCGTGC- | rs2042404240 | TCF25 | 0.0044 | T | | 1.98E-09 | ?+ | 1.979E-09 | | Old |
| 974 | 17 | 17-1014620-C-CCT | | ABR | 0.0009 | CCT | | 1.27E-09 | ?+ | 1.267E-09 | | New |
| 975 | 17 | 17-1354830-TTTG-T | | NA | 0.0018 | TTTG | | 1.56E-13 | ?+ | 1.557E-13 | | Old |
| 976 | 17 | 17-1430883-T-C | rs4547394 | CRK | 0.025 | T | | 5.38E-17 | ?+ | 5.38E-17 | | Old |
| 976 | 17 | 17-1430884-G-A | rs533822042 | CRK | 0.0036 | A | 0.164919 | 2.51E-13 | ++ | 1.244E-08 | 0.02192 | Old |
| 977 | 17 | 17-1596998-C-CCT | rs1251919664 | SLC43A2 | 0.0012 | CCT | 0.985869 | 3.07E-14 | -+ | 2.156E-13 | 0.2073 | Old |
| 978 | 17 | 17-16371521-G-C | | CENPV, UBB | 0.0013 | C | 0.99688 | 4.82E-16 | -+ | 4.773E-10 | | New |

| | | | | | | | | | | | | |
|---|---|---|---|---|---|---|---|---|---|---|---|---|
| 979 | 17 | 17-16451843-C-CAA | | NA | 0.0003 | CAA | | 3.69E-08 | ?+ | 3.694E-08 | | New |
| 980 | 17 | 17-17001857-GCA-G | | NA | 0.0005 | GCA | | 1.87E-10 | ?+ | 1.865E-10 | | New |
| 981 | 17 | 17-17002395-G-A | rs1476682091 | LINC02090, AC104024.2 | 0.0008 | A | 0.017196 | 1.09E-17 | ++ | 1.674E-18 | | New |
| 982 | 17 | 17-1758445-ACT-A | rs1306536849 | SERPINF2, SERPINF1 | 0.0036 | ACT | 0.444551 | 6.12E-28 | -+ | 5.17E-23 | | Old |
| 983 | 17 | 17-1788527-C-CCT | | NA | 0.0002 | CCT | | 4.13E-10 | ?+ | 4.131E-10 | | Old |
| 984 | 17 | 17-18080932-T-A | rs1597705854 | GID4, DRG2 | 0.0008 | T | 0.578681 | 7.15E-13 | ++ | 3.097E-10 | | Old |
| 985 | 17 | 17-18084733-G-C | rs941572665 | GID4, DRG2 | 0.0009 | G | 0.217919 | 2.59E-12 | -+ | 4.837E-10 | | Old |
| 986 | 17 | 17-18762161-T-A | | FBXW10 | 0.0005 | T | | 4.8E-09 | ?+ | 4.802E-09 | | New |
| 986 | 17 | 17-18762165-C-T | rs1268187921 | FBXW10 | 0.0003 | T | 0.03984 | 3.16E-07 | ++ | 3.659E-08 | | New |
| 987 | 17 | 17-19025179-TTTTTC-T | rs1293661154 | GRAP | 0.0043 | T | 0.952708 | 4.21E-15 | ++ | 1.477E-13 | | New |
| 988 | 17 | 17-21260860-AC-A | rs1372704444 | NATD1, MAP2K3 | 0.0019 | AC | | 1.14E-15 | ?+ | 1.143E-15 | | New |
| 989 | 17 | 17-21684971-C-CAGG | | NA | 0.0057 | CAGG | | 6.51E-09 | ?+ | 6.508E-09 | 0.6378 | New |
| 990 | 17 | 17-22064831-C-CAAAAAAAAAAAAA | | NA | 0.028 | CAAAAAAAAAAAAA | | 1.6E-09 | ?+ | 1.603E-09 | | New |
| 991 | 17 | 17-22204256-G-C | rs1363962018 | KCNJ18, LINC02002 | 0.0005 | C | | 4.05E-10 | ?+ | 4.047E-10 | | New |
| 992 | 17 | 17-2584417-GGC-G | rs1241129008 | RN7SL33P, PAFAH1B1 | 0.0025 | G | | 3.25E-11 | ?+ | 3.25E-11 | 0.4068 | New |
| 993 | 17 | 17-26682500-T-A | rs1223720719 | NONE, AC069061.2 | 0.0031 | T | 0.453771 | 3.54E-21 | -+ | 1.71E-20 | 0.02679 | New |
| 994 | 17 | 17-26682529-G-C | rs1906375469 | NONE, AC069061.2 | 0.0022 | C | 0.545566 | 8.79E-29 | ++ | 7.414E-29 | | New |

| | | | | | | | | | | | | |
|---|---|---|---|---|---|---|---|---|---|---|---|---|
| 995 | 17 | 17-26989948-AGC-A | | NA | 0.0048 | A | | 2.48E-12 | ?+ | 2.482E-12 | 0.6414 | New |
| 996 | 17 | 17-2823905-C-CCT | | RAP1GAP2 | 0.0014 | CCT | | 1.7E-09 | ?+ | 1.697E-09 | | New |
| 997 | 17 | 17-2846555-C-CAT | | NA | 0.0004 | C | | 7.34E-09 | ?+ | 7.336E-09 | | New |
| 998 | 17 | 17-2846555-C-CATGA | | NA | 0.0017 | C | | 1.41E-19 | ?+ | 1.408E-19 | | New |
| 998 | 17 | 17-2846560-GC-G | | NA | 0.0007 | GC | | 1.75E-13 | ?+ | 1.747E-13 | | New |
| 998 | 17 | 17-2846563-G-A | rs1031320110 | RAP1GAP2 | 0.0008 | G | 0.18275 | 3.66E-11 | ++ | 1.481E-11 | | New |
| 998 | 17 | 17-2846566-ACGT-A | | NA | 0.0008 | ACGT | | 2.37E-09 | ?+ | 2.368E-09 | | New |
| 999 | 17 | 17-28709778-C-CTT | rs1441508801 | PROCA1 | 0.0024 | C | | 1.37E-10 | ?+ | 1.372E-10 | 0.1446 | New |
| 1000 | 17 | 17-28771881-C-T | rs902214963 | FAM222B | 0.0011 | C | 0.559424 | 1.61E-10 | -+ | 3.919E-09 | 0.09652 | New |
| 1001 | 17 | 17-29813599-C-CTT | | NA | 0.0033 | C | | 7.25E-17 | ?+ | 7.247E-17 | | New |
| 1002 | 17 | 17-29852694-GGCGC-G | | NA | 0.0009 | G | | 6.34E-12 | ?+ | 6.342E-12 | | New |
| 1002 | 17 | 17-29852702-C-CA | | NA | 0.0006 | CA | | 4.62E-09 | ?+ | 4.62E-09 | | New |
| 1003 | 17 | 17-3112620-GGGT-G | rs1334630696 | OR1D2, OR1G1 | 0.0027 | GGGT | | 6.92E-10 | ?+ | 6.915E-10 | | New |
| 1004 | 17 | 17-31929615-C-CCT | rs1282750627 | UTP6, SUZ12 | 0.0071 | CCT | | 4.65E-10 | ?+ | 4.646E-10 | 0.2965 | New |
| 1005 | 17 | 17-35682510-TG-T | | NA | 0.0022 | T | | 2.67E-10 | ?+ | 2.672E-10 | 0.9754 | New |
| 1006 | 17 | 17-3765193-C-CA | | NA | 0.0008 | C | | 7.54E-11 | ?+ | 7.542E-11 | | New |
| 1007 | 17 | 17-38809283-C-T | rs61422252 | CWC25 | 0.1306 | C | | 7.75E-09 | ?+ | 7.751E-09 | 0.111 | New |
| 1008 | 17 | 17-38862181-C-T | rs1269322417 | RPL23, LASP1 | 0.0008 | T | | 1.56E-14 | ?+ | 1.562E-14 | 0.04591 | New |
| 1009 | 17 | 17-38955163-G-A | rs1905486762 | FBXO47 | 0.0004 | G | | 1.6E-09 | ?+ | 1.599E-09 | 0.2919 | New |
| 1010 | 17 | 17-39154845-C-CA | rs1331156689 | PLXDC1 | 0.001 | CA | | 7.37E-11 | ?+ | 7.368E-11 | | New |
| 1011 | 17 | 17-397831-G-T | rs1257925295 | RPH3AL, AC141424.1 | 0.0019 | T | 0.828572 | 2.41E-18 | -+ | 1.155E-15 | | New |

| | | | | | | | | | | | | |
|---|---|---|---|---|---|---|---|---|---|---|---|---|
| 1012 | 17 | 17-39956807-C-CAG | | NA | 0.0005 | CAG | | 5.6E-09 | ?+ | 5.601E-09 | | New |
| 1013 | 17 | 17-40013510-C-CCT | | NA | 0.0016 | C | | 8.51E-17 | ?+ | 8.507E-17 | | New |
| 1014 | 17 | 17-40044663-C-CTT | | NA | 0.0003 | CTT | | 9.01E-09 | ?+ | 9.015E-09 | | New |
| 1015 | 17 | 17-40544234-G-A | rs8066507 | AC004585.1, CCR7 | 0.1379 | A | | 9.67E-11 | ?+ | 9.672E-11 | | New |
| 1016 | 17 | 17-41722828-GTCCCCAA-G | | NA | 0.0025 | GTCCCCAA | | 5.59E-10 | ?+ | 5.594E-10 | 0.359 | New |
| 1017 | 17 | 17-41780474-ACT-A | rs1555609987 | JUP | 0.001 | A | | 1.05E-08 | ?+ | 1.048E-08 | 0.2883 | New |
| 1018 | 17 | 17-41843928-AC-A | rs1555620989 | KLHL10 | 0.001 | A | | 7.47E-10 | ?+ | 7.473E-10 | 0.3313 | New |
| 1019 | 17 | 17-42081656-G-A | rs2053750497 | ZNF385C | 0.0005 | G | 0.410191 | 7.92E-09 | ++ | 1.196E-08 | | New |
| 1020 | 17 | 17-42101152-AG-A | rs1391504551 | DHX58 | 0.0006 | A | | 4.15E-12 | ?+ | 4.153E-12 | | New |
| 1021 | 17 | 17-42606002-ATTAATT-A | | NA | 0.002 | ATTAATT | | 1.63E-10 | ?+ | 1.626E-10 | | New |
| 1022 | 17 | 17-42794089-C-CAG | | NA | 0.0005 | CAG | | 2.16E-11 | ?+ | 2.155E-11 | | New |
| 1023 | 17 | 17-43567458-T-A | rs530555010 | DHX8, ETV4 | 0.0016 | T | | 9.49E-11 | ?+ | 9.488E-11 | | New |
| 1024 | 17 | 17-44139757-G-A | rs1442217664 | RNU6-131P, C17orf53 | 0.0013 | A | | 1.41E-08 | ?+ | 1.408E-08 | 0.2571 | Old |
| 1025 | 17 | 17-44189985-TG-T | rs1216572284 | TMUB2 | 0.0016 | T | 0.203881 | 1.09E-08 | ++ | 3.618E-08 | 0.03941 | Old |
| 1026 | 17 | 17-44372763-GAA-G | | NA | 0.0012 | GAA | | 2.65E-10 | ?+ | 2.65E-10 | | Old |
| 1026 | 17 | 17-44372764-GTT-G | | NA | 0.001 | G | | 1.19E-08 | ?+ | 1.185E-08 | | Old |
| 1027 | 17 | 17-44513203-C-CCGA | | NA | 0.0008 | CCGA | | 3.95E-10 | ?+ | 3.951E-10 | | Old |
| 1028 | 17 | 17-45057514-G-C | rs1244239771 | DCAKD, NMT1 | 0.0009 | C | | 6.9E-12 | ?+ | 6.905E-12 | 0.3093 | New |
| 1029 | 17 | 17-45079271-ATT-A | | NA | 0.0008 | ATT | | 3.23E-12 | ?+ | 3.233E-12 | | New |
| 1030 | 17 | 17-47255122-GCA-G | rs1239388321 | AC068234.1, ITGB3 | 0.0024 | GCA | | 4.5E-13 | ?+ | 4.495E-13 | 0.9792 | Old |
| 1030 | 17 | 17-47255125-C-CCT | rs1355670372 | AC068234.1, ITGB3 | 0.0019 | C | | 3.05E-12 | ?+ | 3.047E-12 | 0.9347 | Old |

| | | | | | | | | | | | | |
|---|---|---|---|---|---|---|---|---|---|---|---|---|
| 1031 | 17 | 17-47530102-ATTTTTTT-A | | NPEPPS | 0.0081 | A | | 7.66E-11 | ?+ | 7.662E-11 | | New |
| 1032 | 17 | 17-48098623-G-A | | CBX1 | 0.0006 | G | | 9.66E-10 | ?+ | 9.663E-10 | | New |
| 1033 | 17 | 17-4820089-C-CAT | rs1331439083 | PLD2 | 0.0038 | C | | 1.33E-17 | ?+ | 1.333E-17 | 0.0344 | Old |
| 1034 | 17 | 17-48866581-C-CCT | | NA | 0.0013 | CCT | | 1.57E-10 | ?+ | 1.573E-10 | | Old |
| 1035 | 17 | 17-49143921-GTGAA-G | | NA | 0.0001 | G | | 3.6E-09 | ?+ | 3.602E-09 | | Old |
| 1036 | 17 | 17-4923566-C-A | rs1970262824 | CHRNE | 0.0008 | C | | 6.86E-13 | ?+ | 6.861E-13 | | Old |
| 1037 | 17 | 17-49345263-C-T | rs906741775 | ZNF652 | 0.001 | T | 0.723394 | 1.83E-12 | ++ | 1.482E-08 | | Old |
| 1038 | 17 | 17-49678884-C-CCATCCTGGCTAA | | NA | 0.0005 | CCATCCTGGCTAA | | 3.12E-08 | ?+ | 3.122E-08 | | Old |
| 1039 | 17 | 17-4993046-ATT-A | | NA | 0.0008 | ATT | | 7.15E-13 | ?+ | 7.151E-13 | | Old |
| 1040 | 17 | 17-4995776-C-CA | | NA | 1E-04 | C | | 3.53E-13 | ?+ | 3.526E-13 | | Old |
| 1041 | 17 | 17-50126919-GT-G | | NA | 0.0018 | G | | 3.69E-08 | ?+ | 3.69E-08 | | New |
| 1042 | 17 | 17-50156079-ACG-A | rs1295331050 | PPP1R9B, AC015909.2 | 0.0023 | ACG | 0.271093 | 4.02E-11 | -+ | 6.95E-09 | 0.1093 | New |
| 1043 | 17 | 17-50759908-GAC-G | rs1474019140 | LUC7L3, ANKRD40CL | 0.0067 | GAC | | 1.56E-15 | ?+ | 1.564E-15 | | New |
| 1044 | 17 | 17-5265867-C-CCT | rs1484578739 | AC087500.1, RABEP1 | 0.0033 | CCT | | 2.96E-10 | ?+ | 2.955E-10 | | Old |
| 1045 | 17 | 17-5439856-C-CCTT | | NA | 0.0006 | CCTT | | 2.1E-10 | ?+ | 2.098E-10 | | Old |
| 1046 | 17 | 17-58478215-C-CCT | | HSF5 | 0.0028 | CCT | | 4.76E-09 | ?+ | 4.757E-09 | | Old |
| 1047 | 17 | 17-59965446-C-CT | | NA | 0.0012 | CT | | 2.78E-10 | ?+ | 2.783E-10 | | New |
| 1048 | 17 | 17-60341469-ATTCTT-A | rs1214054796 | USP32 | 0.0019 | ATTCTT | | 2.83E-10 | ?+ | 2.83E-10 | 0.1819 | New |
| 1049 | 17 | 17-60589164-GGGT-G | rs1404224919 | LINC01999, PPM1D | 0.0026 | GGGT | | 6.02E-11 | ?+ | 6.022E-11 | 0.4338 | New |
| 1050 | 17 | 17-60703056-C-CCT | | NA | 0.0013 | C | | 1.38E-08 | ?+ | 1.378E-08 | | New |

| | | | | | | | | | | | | |
|---|---|---|---|---|---|---|---|---|---|---|---|---|
| 1051 | 17 | 17-61890402-G-C | rs1429724769 | INTS2 | 0.0008 | G | | 8.85E-09 | ?+ | 8.85E-09 | 0.5404 | New |
| 1052 | 17 | 17-62007671-AC-A | rs1345200633 | MED13 | 0.0053 | AC | 0.649111 | 3.69E-22 | -+ | 3.143E-13 | 0.5902 | New |
| 1053 | 17 | 17-62157437-C-A | rs1427302619 | RF00019, EFCAB3 | 0.0009 | A | 0.668676 | 3.13E-18 | ++ | 3.212E-16 | | New |
| 1053 | 17 | 17-62157438-G-A | rs1414985603 | RF00019, EFCAB3 | 0.0007 | G | 0.800837 | 3.28E-14 | -+ | 1.907E-12 | | New |
| 1054 | 17 | 17-62235962-C-A | rs536039270 | RF00019, EFCAB3 | 0.002 | C | | 3.16E-13 | ?+ | 3.161E-13 | 0.5522 | New |
| 1054 | 17 | 17-62235981-C-T | rs1347265103 | RF00019, EFCAB3 | 0.0036 | C | | 5.91E-21 | ?+ | 5.91E-21 | 0.4521 | New |
| 1055 | 17 | 17-62249528-ATCAT-A | | NA | 0.0008 | ATCAT | | 2.92E-08 | ?+ | 2.922E-08 | | New |
| 1056 | 17 | 17-62388645-G-C | | EFCAB3 | 0.0003 | G | | 1.72E-08 | ?+ | 1.718E-08 | | New |
| 1057 | 17 | 17-62498597-G-C | rs920463313 | TLK2 | 0.0006 | G | | 4.65E-08 | ?+ | 4.651E-08 | | Old |
| 1058 | 17 | 17-62604736-C-CCGTG | | TLK2 | 0.0018 | C | | 5.16E-09 | ?+ | 5.162E-09 | 0.4103 | Old |
| 1059 | 17 | 17-63851546-G-C | rs1904751435 | SMARCD2, CSH2 | 0.0014 | C | | 3.09E-12 | ?+ | 3.091E-12 | 0.2626 | Old |
| 1060 | 17 | 17-64347887-G-T | rs2035619712 | PECAM1 | 0.0018 | T | 0.074886 | 2.86E-21 | ++ | 3.35E-17 | | New |
| 1060 | 17 | 17-64347888-G-A | rs2035619840 | PECAM1 | 0.0013 | G | 0.590512 | 9.28E-14 | ++ | 2.008E-09 | | New |
| 1060 | 17 | 17-64347889-G-C | rs2035619952 | PECAM1 | 0.0015 | C | 0.781658 | 5.52E-15 | ++ | 2.375E-10 | | New |
| 1061 | 17 | 17-67064485-G-A | | CACNG1, HELZ | 0.0003 | G | | 6.63E-09 | ?+ | 6.628E-09 | | New |
| 1062 | 17 | 17-67634352-G-A | rs1389719881 | PITPNC1 | 0.0004 | A | | 1.79E-08 | ?+ | 1.792E-08 | 0.3027 | New |
| 1063 | 17 | 17-68002426-AT-A | rs1480643073 | C17orf58, KPNA2 | 0.0009 | AT | | 3.78E-15 | ?+ | 3.782E-15 | 0.5742 | New |
| 1064 | 17 | 17-68627016-T-A | rs1179509817 | LINC01482 | 0.001 | T | | 1.35E-10 | ?+ | 1.351E-10 | | New |
| 1065 | 17 | 17-69099080-C-CTTT | rs1200022958 | ABCA6 | 0.0007 | CTTT | | 5.43E-09 | ?+ | 5.429E-09 | 0.6594 | New |
| 1066 | 17 | 17-69514705-GCA-G | | NA | 0.0003 | GCA | | 4.89E-10 | ?+ | 4.886E-10 | | New |
| 1067 | 17 | 17-7055430-C-CCT | | NA | 0.0016 | CCT | | 3.81E-19 | ?+ | 3.813E-19 | | New |

| | | | | | | | | | | | | |
|---|---|---|---|---|---|---|---|---|---|---|---|---|
| 1068 | 17 | 17-7268492-G-T | rs1404018243 | RF00019, SLC2A4 | 0.0013 | G | 0.661078 | 2.21E-13 | ++ | 1.016E-09 | | New |
| 1069 | 17 | 17-73051724-GGT-G | rs150326641 | SLC39A11 | 0.0015 | GGT | | 1.1E-09 | ?+ | 1.096E-09 | | New |
| 1070 | 17 | 17-74976489-C-CCT | | NA | 0.0008 | CCT | | 1.59E-08 | ?+ | 1.589E-08 | | New |
| 1071 | 17 | 17-75206797-AAG-A | rs1489928357 | NUP85 | 0.0041 | AAG | | 3.96E-22 | ?+ | 3.959E-22 | 0.1882 | New |
| 1071 | 17 | 17-75206799-ACC-A | rs1201639171 | NUP85 | 0.0038 | A | | 2.44E-25 | ?+ | 2.435E-25 | | New |
| 1072 | 17 | 17-75529672-C-CTAGCA | | NA | 0.0013 | CTAGCA | | 2.24E-09 | ?+ | 2.244E-09 | | New |
| 1073 | 17 | 17-75711868-C-CTG | | NA | 0.0001 | CTG | | 5.83E-09 | ?+ | 5.829E-09 | | New |
| 1074 | 17 | 17-75801049-GCA-G | | NA | 0.0022 | GCA | | 1.46E-19 | ?+ | 1.463E-19 | | New |
| 1074 | 17 | 17-75801051-C-CAG | | NA | 0.0018 | C | | 2.54E-16 | ?+ | 2.544E-16 | | New |
| 1074 | 17 | 17-75801070-ATTT-A | rs1416175390 | NA | 0.0035 | A | 0.839552 | 3.08E-30 | -+ | 3.902E-23 | | New |
| 1075 | 17 | 17-75968218-C-CCT | rs2065957087 | ACOX1 | 0.0066 | CCT | | 4.04E-08 | ?+ | 4.04E-08 | | New |
| 1076 | 17 | 17-75993291-AT-A | | NA | 0.001 | A | | 1.04E-11 | ?+ | 1.042E-11 | | New |
| 1077 | 17 | 17-75993927-C-CGGA | | TEN1, TEN1-CDK3 | 0.001 | C | | 8.22E-11 | ?+ | 8.223E-11 | | New |
| 1078 | 17 | 17-76098643-C-CCT | | NA | 0.0016 | CCT | | 2.15E-11 | ?+ | 2.146E-11 | | New |
| 1079 | 17 | 17-7681502-AT-A | rs1475747079 | TP53 | 0.0036 | AT | 0.905228 | 1.43E-19 | ++ | 2.63E-15 | 0.9197 | New |
| 1080 | 17 | 17-7681961-TTTTTTG-T | rs1180886598 | TP53 | 0.0029 | TTTTTTG | | 5.25E-09 | ?+ | 5.246E-09 | 0.2077 | New |
| 1081 | 17 | 17-783556-G-C | | MRM3 | 0.0005 | G | | 3.79E-15 | ?+ | 3.79E-15 | | New |
| 1082 | 17 | 17-78864868-AC-A | rs1265889286 | TIMP2 | 0.001 | AC | | 3.27E-08 | ?+ | 3.27E-08 | | New |
| 1083 | 17 | 17-81849383-G-C | | P4HB | 0.0008 | G | 0.302558 | 3.98E-13 | ++ | 2.074E-10 | | New |
| 1084 | 17 | 17-82725483-GCT-G | | NA | 0.0014 | GCT | | 8.45E-12 | ?+ | 8.447E-12 | | New |
| 1085 | 17 | 17-839242-G-C | | NXN | 0.0003 | C | 0.382387 | 2.42E-09 | -+ | 1.154E-08 | | New |
| 1086 | 17 | 17-8853714-G-A | rs60256690 | PIK3R6 | 0.0791 | A | | 1.03E-08 | ?+ | 1.033E-08 | 0.8296 | New |

| | | | | | | | | | | | | |
|---|---|---|---|---|---|---|---|---|---|---|---|---|
| 1087 | 17 | 17-9113990-C-CT | | NA | 0.0005 | CT | | 2.47E-08 | ?+ | 2.467E-08 | | New |
| 1088 | 17 | 17-971686-GGTGA-G | | NA | 0.001 | G | | 5.4E-14 | ?+ | 5.397E-14 | | New |
| 1089 | 18 | 18-12866210-AGGTG-A | | PTPN2 | 0.0019 | A | | 5.67E-21 | ?+ | 5.669E-21 | | New |
| 1090 | 18 | 18-14502080-G-C | rs1909747014 | AP006261.1, POTEC | 0.0005 | G | | 2.78E-09 | ?+ | 2.778E-09 | | New |
| 1091 | 18 | 18-21616146-C-T | rs758518862 | SNRPD1 | 0.0004 | T | 0.377088 | 2.45E-09 | ++ | 1.141E-08 | 0.6019 | New |
| 1092 | 18 | 18-21731482-C-A | rs890005603 | RF00019, MIB1 | 0.0034 | A | | 8.83E-24 | ?+ | 8.832E-24 | | New |
| 1093 | 18 | 18-218980-C-CA | rs11333392 | THOC1 | 0.0086 | C | 0.196996 | 2.55E-08 | ++ | 4.711E-08 | | New |
| 1094 | 18 | 18-21974281-G-C | rs1568254901 | AC103987.2, LINC01900 | 0.0016 | C | 0.670013 | 5.28E-16 | ++ | 1.362E-10 | 0.7329 | New |
| 1095 | 18 | 18-23093206-AT-A | | NA | 0.001 | A | | 5E-11 | ?+ | 4.996E-11 | | New |
| 1095 | 18 | 18-23093208-C-CA | | NA | 0.0009 | C | | 1.23E-08 | ?+ | 1.234E-08 | | New |
| 1096 | 18 | 18-33147978-C-CT | rs1220304375 | CCDC178 | 0.0064 | CT | 0.343818 | 1.37E-43 | -+ | 1.123E-42 | | New |
| 1097 | 18 | 18-3550114-G-A | | DLGAP1 | 0.0005 | G | | 4.9E-09 | ?+ | 4.902E-09 | | New |
| 1098 | 18 | 18-47400508-TCCACAGTCATCTTCCCACCCGAGGCCACCACACTGTGCCTTC-T | rs1416466766 | MIR4527HG | 0.0103 | T | | 1.23E-08 | ?+ | 1.231E-08 | | New |
| 1099 | 18 | 18-54138189-AT-A | | NA | 0.0018 | AT | | 5.35E-09 | ?+ | 5.354E-09 | | New |
| 1100 | 18 | 18-54207819-C-CCT | | NA | 0.002 | CCT | | 4.12E-08 | ?+ | 4.116E-08 | | New |
| 1101 | 18 | 18-54537923-C-CTACTGAACCCACAACCACAGCAGCCAGGACTGTAACTGAACAGTCTGCCATCACAACTGTAGGCATTTACACTGAGACTACAACCCCATCAGCCACCACTACCACT | | NA | 0.0005 | CTACTGAACCCACAAC | | 3.39E-08 | ?+ | 3.389E-08 | | New |

| | | | | | | | | | | | | |
|---|---|---|---|---|---|---|---|---|---|---|---|---|
| 1102 | 18 | 18-56903567-G-C | rs1296656080 | WDR7 | 0.0011 | C | 0.659043 | 1.04E-16 | ++ | 3.316E-15 | | New |
| 1103 | 18 | 18-58129746-C-CGGTGTTGGGCTCTGTTGGGGTTTGGTTGTGAGCGGAACTGT | | NA | 0.0031 | C | | 3.11E-08 | ?+ | 3.11E-08 | | Old |
| 1104 | 18 | 18-61699837-C-T | rs140886023 | AC105094.2 | 0.0198 | T | | 1.07E-10 | ?- | 1.07E-10 | 0.7636 | New |
| 1105 | 18 | 18-62195984-C-CTG | | RELCH | 0.0014 | CTG | | 3.05E-08 | ?+ | 3.052E-08 | | New |
| 1105 | 18 | 18-62195985-C-CAA | | RELCH | 0.002 | C | | 1.14E-13 | ?+ | 1.14E-13 | | New |
| 1106 | 18 | 18-7107368-TTGG-T | | NA | 0.0004 | TTGG | | 4.49E-08 | ?+ | 4.486E-08 | | New |
| 1107 | 18 | 18-7274664-AT-A | | NA | 0.0011 | AT | | 3.18E-08 | ?+ | 3.181E-08 | | New |
| 1108 | 18 | 18-76817728-GCCT-G | | NA | 0 | GCCT | | 1.1E-08 | ?+ | 1.095E-08 | | New |
| 1109 | 18 | 18-78591455-C-CCT | | NA | 0.001 | CCT | | 4.15E-09 | ?+ | 4.146E-09 | 0.4528 | New |
| 1110 | 18 | 18-80185678-G-A | rs1340254507 | AC139100.1 | 0.0008 | G | | 1.17E-10 | ?+ | 1.168E-10 | | New |
| 1111 | 18 | 18-9037196-GGT-G | | NA | 0.0004 | G | | 2.68E-08 | ?+ | 2.676E-08 | 0.8587 | New |
| 1112 | 18 | 18-9777902-C-A | | RAB31 | 0.0006 | A | | 3.5E-10 | ?+ | 3.498E-10 | | New |
| 1113 | 19 | 19-10263242-GCGCC-G | rs1377788735 | AC011511.2 | 0.0035 | G | | 4.88E-08 | ?+ | 4.876E-08 | | New |
| 1114 | 19 | 19-11071676-GC-G | rs1404913201 | SMARCA4 | 0.0068 | GC | | 1.75E-35 | ?+ | 1.753E-35 | 0.2132 | New |
| 1115 | 19 | 19-11473295-G-A | rs664477 | AC008481.3, ELAVL3 | 0.3062 | G | | 3.87E-10 | ?+ | 3.871E-10 | | New |
| 1116 | 19 | 19-11582992-G-T | rs6511738 | ZNF627 | 0.0006 | G | | 1.61E-09 | ?+ | 1.605E-09 | | New |
| 1117 | 19 | 19-11648684-AAC-A | | NA | 0.0005 | A | | 9.04E-10 | ?+ | 9.036E-10 | | New |
| 1118 | 19 | 19-12129144-TA-T | | NA | 0.0005 | T | | 5.85E-09 | ?+ | 5.849E-09 | | New |
| 1119 | 19 | 19-12286479-C-CAA | | NA | 0.0009 | CAA | | 8.62E-09 | ?+ | 8.616E-09 | | New |
| 1120 | 19 | 19-13736813-C-CCT | rs1481218172 | CCDC130 | 0.0012 | C | | 9.21E-11 | ?+ | 9.211E-11 | | New |

| | | | | | | | | | | | | |
|---|---|---|---|---|---|---|---|---|---|---|---|---|
| 1121 | 19 | 19-13745457-G-A | | CCDC130 | 0.0006 | G | | 7.49E-09 | ?+ | 7.485E-09 | | New |
| 1122 | 19 | 19-13893919-AT-A | | NA | 0.0017 | AT | | 6.48E-19 | ?+ | 6.478E-19 | | New |
| 1122 | 19 | 19-13893922-AT-A | | NA | 0.0011 | AT | | 3.67E-11 | ?+ | 3.672E-11 | | New |
| 1123 | 19 | 19-14258228-C-CATA | | AC011509.2 | 0.0018 | CATA | | 1.2E-10 | ?+ | 1.198E-10 | 0.6494 | New |
| 1124 | 19 | 19-1430887-ATTT-A | rs1358250852 | DAZAP1 | 0.0007 | A | | 2.5E-10 | ?+ | 2.495E-10 | | Old |
| 1125 | 19 | 19-14648966-TCCC-T | rs1352071596 | ADGRE3 | 0.001 | T | | 1.32E-11 | ?+ | 1.318E-11 | | New |
| 1126 | 19 | 19-14707697-GCCCA-G | | NA | 0.0006 | G | | 1.16E-08 | ?+ | 1.163E-08 | | New |
| 1127 | 19 | 19-15822925-GGGCT-G | | NA | 0.0018 | GGGCT | | 2.58E-26 | ?+ | 2.578E-26 | | New |
| 1127 | 19 | 19-15822927-C-CTTAT | | NA | 0.0009 | C | | 3E-12 | ?+ | 2.999E-12 | | New |
| 1128 | 19 | 19-16557360-AT-A | rs1255709951 | AC008764.1, SLC35E1 | 0.0009 | A | 0.354923 | 1.27E-10 | ++ | 4E-10 | | New |
| 1129 | 19 | 19-1666683-GC-G | rs1374443291 | TCF3, RF00017 | 0.0051 | GC | | 1.08E-13 | ?+ | 1.081E-13 | 0.3773 | Old |
| 1130 | 19 | 19-17354058-GGT-G | | NA | 0.0003 | G | | 2.97E-08 | ?+ | 2.969E-08 | | New |
| 1131 | 19 | 19-18528048-C-CT | | NA | 0.0007 | CT | | 2.81E-10 | ?+ | 2.805E-10 | | New |
| 1132 | 19 | 19-18619797-C-CCA | rs1253147039 | AC003112.1 | 0.0014 | C | | 3.01E-09 | ?+ | 3.008E-09 | 0.3021 | New |
| 1133 | 19 | 19-18689093-GCC-G | rs1209568272 | CRTC1 | 0.0021 | GCC | | 1.83E-08 | ?+ | 1.828E-08 | 0.3745 | New |
| 1134 | 19 | 19-1893632-C-T | rs560268575 | ABHD17A, SCAMP4 | 0.0022 | C | 0.337528 | 5.2E-12 | ++ | 3.94E-09 | 0.6408 | Old |
| 1135 | 19 | 19-1899530-TC-T | | NA | 0.0015 | TC | | 3.79E-11 | ?+ | 3.787E-11 | | Old |
| 1136 | 19 | 19-19363796-GTT-G | | MAU2, GATAD2A | 0.0017 | GTT | | 4.86E-13 | ?+ | 4.861E-13 | | New |
| 1137 | 19 | 19-19382660-C-T | rs140390502 | MAU2, GATAD2A | 0.0004 | T | 0.80841 | 2.61E-10 | ++ | 8.87E-09 | | New |
| 1138 | 19 | 19-19572678-AACGTG-A | rs1483340178 | PBX4 | 0.0024 | A | | 1.36E-09 | ?+ | 1.355E-09 | 0.616 | New |
| 1139 | 19 | 19-20023807-C-CCT | | NA | 0.0002 | C | | 1.08E-08 | ?+ | 1.077E-08 | | New |

| | | | | | | | | | | | | |
|---|---|---|---|---|---|---|---|---|---|---|---|---|
| 1140 | 19 | 19-20076324-C-A | | ZNF682, ZNF90 | 0.0004 | A | 0.965211 | 8.82E-15 | -+ | 9.098E-13 | | New |
| 1141 | 19 | 19-20143926-G-A | rs62107009 | AC011447.3 | 0.0019 | G | 0.757766 | 6.88E-19 | ++ | 1.237E-13 | 0.6041 | New |
| 1141 | 19 | 19-20143939-G-A | rs575933971 | AC011447.3 | 0.0032 | A | 0.28918 | 7.88E-27 | -+ | 2.942E-15 | | New |
| 1142 | 19 | 19-20144534-C-CA | rs1555709908 | AC011447.3 | 0.0015 | C | | 1.19E-08 | ?+ | 1.191E-08 | | New |
| 1143 | 19 | 19-22712186-C-CAT | | NA | 0.0008 | C | | 5.31E-10 | ?+ | 5.309E-10 | | New |
| 1143 | 19 | 19-22712189-GCC-G | | NA | 0.0008 | GCC | | 1.06E-09 | ?+ | 1.062E-09 | | New |
| 1144 | 19 | 19-22831340-TGAG-T | rs1160098113 | ZNF723 | 0.0048 | T | 0.741934 | 3.31E-10 | -+ | 1.332E-09 | 0.918 | New |
| 1145 | 19 | 19-23676840-C-CAA | | NA | 0.0004 | CAA | | 1.41E-08 | ?+ | 1.406E-08 | | New |
| 1145 | 19 | 19-23676856-C-CCT | | NA | 0.0003 | CCT | | 3.76E-09 | ?+ | 3.755E-09 | | New |
| 1146 | 19 | 19-2654412-G-A | rs1250990056 | GNG7 | 0.0012 | G | 0.745937 | 2.13E-10 | ++ | 7.761E-10 | | New |
| 1147 | 19 | 19-2673038-C-CCT | rs1189518681 | GNG7 | 0.0023 | CCT | | 1.07E-12 | ?+ | 1.067E-12 | | New |
| 1148 | 19 | 19-27461080-G-A | rs1474832594 | NONE, ERVK-28 | 0.0001 | A | | 2.55E-08 | ?+ | 2.549E-08 | | New |
| 1149 | 19 | 19-2870227-AGTTG-A | | NA | 0.0007 | A | | 4.27E-11 | ?+ | 4.269E-11 | | New |
| 1149 | 19 | 19-2870231-GCA-G | | NA | 0.0006 | GCA | | 1.62E-09 | ?+ | 1.623E-09 | | New |
| 1150 | 19 | 19-3259781-GA-G | rs1210150502 | CELF5 | 0.0019 | GA | | 5.06E-12 | ?+ | 5.06E-12 | 0.121 | New |
| 1151 | 19 | 19-33033650-C-CCT | | NA | 0.0005 | C | | 1.94E-08 | ?+ | 1.936E-08 | | New |
| 1152 | 19 | 19-33141241-T-C | rs7249607 | WDR88 | 0.421 | T | | 4.48E-08 | ?- | 4.482E-08 | | New |
| 1153 | 19 | 19-33283110-C-T | rs548960608 | AC008738.6 | 0.0008 | C | 0.398578 | 6.11E-10 | ++ | 2.175E-09 | 0.001541 | New |
| 1154 | 19 | 19-33837044-AC-A | | NA | 0.0005 | A | | 1.45E-13 | ?+ | 1.45E-13 | | New |
| 1155 | 19 | 19-34031357-G-C | rs1237948850 | AC016587.1 | 0.0026 | C | 0.001682 | 2.67E-09 | ++ | 4.554E-09 | 0.7012 | New |
| 1155 | 19 | 19-34031358-C-A | rs1283930218 | AC016587.1 | 0.0027 | A | 0.000694 | 4.13E-08 | ++ | 8.569E-09 | | New |
| 1156 | 19 | 19-34371991-C-CCT | | NA | 0.0003 | CCT | | 3.43E-08 | ?+ | 3.429E-08 | | New |

| | | | | | | | | | | | | |
|---|---|---|---|---|---|---|---|---|---|---|---|---|
| 1157 | 19 | 19-34465420-C-CG | | NA | 0.0006 | C | | 7.81E-11 | ?+ | 7.812E-11 | | New |
| 1158 | 19 | 19-3479270-AAAG-A | rs1339749404 | SMIM24 | 0.002 | AAAG | | 1.89E-08 | ?+ | 1.89E-08 | 0.197 | New |
| 1159 | 19 | 19-3587428-C-CCCG | rs1221662457 | GIPC3 | 0.0005 | CCCG | | 4.65E-08 | ?+ | 4.647E-08 | 0.06382 | New |
| 1160 | 19 | 19-35911172-C-CAT | | NA | 0.0004 | C | | 3.32E-08 | ?+ | 3.316E-08 | | New |
| 1161 | 19 | 19-36204814-ACC-A | rs1403003350 | ZNF565 | 0.0029 | A | | 4.35E-26 | ?+ | 4.347E-26 | 0.385 | New |
| 1161 | 19 | 19-36204819-GCA-G | rs1362397835 | ZNF565 | 0.0027 | GCA | | 4.97E-25 | ?+ | 4.972E-25 | 0.4866 | New |
| 1162 | 19 | 19-36516403-C-CTTGG | | NA | 0.0008 | C | | 1E-10 | ?+ | 1.003E-10 | | New |
| 1163 | 19 | 19-36868169-AT-A | | NA | 0.0013 | A | | 4.08E-13 | ?+ | 4.078E-13 | | New |
| 1164 | 19 | 19-36868792-AT-A | | NA | 0.0009 | AT | 0.155975 | 4.68E-11 | -+ | 1.72E-09 | | New |
| 1165 | 19 | 19-36950324-G-A | rs1314452460 | ZNF568 | 0.0006 | G | | 8.16E-10 | ?+ | 8.158E-10 | 0.9056 | New |
| 1166 | 19 | 19-37687722-C-CAG | | NA | 0.0005 | CAG | | 7.47E-09 | ?+ | 7.469E-09 | | New |
| 1167 | 19 | 19-38243249-GCC-G | | NA | 0.0009 | G | | 6.09E-09 | ?+ | 6.09E-09 | 0.5685 | New |
| 1168 | 19 | 19-3843252-C-CTT | rs1332151591 | ZFR2 | 0.0072 | C | | 1.86E-20 | ?+ | 1.86E-20 | 0.4799 | New |
| 1169 | 19 | 19-39105162-TGC-T | rs1357694804 | ACP7 | 0.0027 | TGC | | 1.24E-11 | ?+ | 1.237E-11 | 0.432 | New |
| 1169 | 19 | 19-39105164-C-CGT | rs2073397987 | ACP7 | 0.0024 | C | | 3.54E-11 | ?+ | 3.537E-11 | | New |
| 1170 | 19 | 19-39108282-GGC-G | rs1294325936 | ACP7 | 0.002 | G | | 2.85E-19 | ?+ | 2.854E-19 | | New |
| 1170 | 19 | 19-39108289-ATG-A | rs1282434985 | ACP7 | 0.0016 | ATG | | 1.08E-12 | ?+ | 1.079E-12 | | New |
| 1171 | 19 | 19-39148690-C-CAG | rs1184157092 | PAK4 | 0.0025 | CAG | | 5.18E-11 | ?+ | 5.18E-11 | | New |
| 1172 | 19 | 19-39301024-TCC-T | rs1465355970 | IFNL1, LRFN1 | 0.0015 | TCC | 0.252972 | 2.06E-18 | ++ | 2.675E-18 | | New |
| 1173 | 19 | 19-40402196-C-CAA | | NA | 0.0024 | C | | 4.7E-10 | ?+ | 4.701E-10 | | New |
| 1174 | 19 | 19-41157626-AGT-A | | NA | 0.0003 | A | | 2.07E-10 | ?+ | 2.067E-10 | | New |

| | | | | | | | | | | | | |
|---|---|---|---|---|---|---|---|---|---|---|---|---|
| 1175 | 19 | 19-41509971-C-CT | | NA | 0.0059 | CT | | 1.17E-15 | ?+ | 1.169E-15 | | Old |
| 1176 | 19 | 19-41806330-TTG-T | | NA | 0.0013 | TTG | | 3.95E-11 | ?+ | 3.954E-11 | | Old |
| 1177 | 19 | 19-4302852-C-CA | | NA | 0.0002 | C | | 7.98E-09 | ?+ | 7.982E-09 | | New |
| 1178 | 19 | 19-43061627-AT-A | rs1396957395 | AC004784.1 | 0.0008 | A | | 2.74E-08 | ?+ | 2.736E-08 | 0.7926 | Old |
| 1179 | 19 | 19-4331320-C-CCT | | NA | 0.0013 | C | | 5.99E-14 | ?+ | 5.986E-14 | | New |
| 1179 | 19 | 19-4331323-TCC-T | | NA | 0.0011 | TCC | | 1.53E-13 | ?+ | 1.53E-13 | | New |
| 1180 | 19 | 19-44457678-G-A | rs1205865847 | ZNF229, ZNF180 | 0.0015 | A | 0.830066 | 7.08E-14 | ++ | 1.137E-12 | | Old |
| 1181 | 19 | 19-4450003-G-T | rs969036491 | AC011498.7 | 0.0041 | G | 0.660341 | 5.38E-27 | ++ | 5.918E-20 | 0.2727 | New |
| 1182 | 19 | 19-44673285-G-A | rs142034848 | AC243964.2 | 0.0136 | G | 5.78E-10 | 0.114836 | ++ | 8.579E-10 | 0.0002124 | Old |
| 1182 | 19 | 19-44733473-C-T | rs2927434 | RF00285, BCL3 | 0.1809 | T | 4.41E-14 | 0.015193 | ++ | 2.538E-14 | 0.000134 | Old |
| 1182 | 19 | 19-44736813-C-CT | rs58455045 | RF00285, BCL3 | 0.1927 | CT | 3.1E-15 | 0.008123 | ++ | 1.086E-15 | | Old |
| 1182 | 19 | 19-44738381-G-A | rs2927437 | RF00285, BCL3 | 0.1784 | A | 6.85E-17 | 0.004637 | ++ | 1.915E-17 | 0.000004448 | Old |
| 1182 | 19 | 19-44740137-TATACACACACACAC-T | rs34562807 | RF00285, BCL3 | 0.0541 | TATACACA | 2.47E-21 | 0.004364 | ++ | 4.017E-21 | | Old |
| 1182 | 19 | 19-44781370-G-A | rs80168591 | CBLC | 0.012 | A | 1.94E-13 | 0.004804 | ++ | 8.087E-15 | 2.343E-10 | Old |
| 1182 | 19 | 19-44799020-C-T | rs139997344 | CBLC | 0.0074 | T | 1.16E-20 | 0.189998 | ++ | 1.352E-18 | 7.851E-08 | Old |
| 1182 | 19 | 19-44812964-G-A | rs528070791 | BCAM | 0.0126 | G | 4.84E-24 | 0.070312 | ++ | 3.541E-22 | | Old |
| 1182 | 19 | 19-44819913-CCATCCCCAACTCATCCT-C | rs3842409 | BCAM | 0.4442 | C | 4.29E-22 | 0.014798 | -- | 1.2E-20 | | Old |
| 1182 | 19 | 19-44820881-G-A | rs28399637 | BCAM | 0.2824 | A | 1.07E-33 | 0.287699 | ++ | 1.305E-26 | 4.149E-20 | Old |
| 1182 | 19 | 19-44821499-G-C | rs28399664 | BCAM | 0.0185 | G | 2.51E-44 | 0.002044 | ++ | 2.879E-42 | 0.0684 | Old |
| 1182 | 19 | 19-44827852-G-C | rs905342119 | BCAM, NECTIN2 | 0.0082 | C | 4.91E-22 | 0.141143 | ++ | 3.893E-20 | 5.485E-10 | Old |

| | | | | | | | | | | | | |
|---|---|---|---|---|---|---|---|---|---|---|---|---|
| 1182 | 19 | 19-44834661-G-A | rs147711004 | BCAM, NECTIN2 | 0.0311 | A | 4.52E-63 | 0.000683 | ++ | 3.393E-59 | 0.2333 | Old |
| 1182 | 19 | 19-44843409-G-C | rs148601586 | BCAM, NECTIN2 | 0.0122 | G | 5.77E-19 | 0.030667 | ++ | 8.133E-19 | 3.717E-13 | Old |
| 1182 | 19 | 19-44848259-G-C | rs41289512 | NECTIN2 | 0.0362 | G | 9.43E-65 | 0.002147 | ++ | 6.644E-59 | 3.555E-30 | Old |
| 1182 | 19 | 19-44850787-G-T | rs57537848 | NECTIN2 | 0.4852 | G | 5.34E-10 | 0.351864 | -- | 1.929E-08 | 0.06697 | Old |
| 1182 | 19 | 19-44851039-G-A | rs11666329 | NECTIN2 | 0.4853 | G | 5.39E-10 | 0.331867 | -- | 1.699E-08 | 9.115E-10 | Old |
| 1182 | 19 | 19-44852031-C-T | rs149661872 | NECTIN2 | 0.0091 | T | 1.66E-17 | 0.333973 | ++ | 1.897E-15 | 1.42E-09 | Old |
| 1182 | 19 | 19-44852486-C-A | rs4802241 | NECTIN2 | 0.1772 | C | 1.05E-08 | 0.091571 | -- | 1.531E-08 | 0.0001444 | Old |
| 1182 | 19 | 19-44855049-C-T | rs551048812 | NECTIN2 | 0.0084 | T | 3.3E-10 | 0.007583 | ++ | 1.752E-11 | 1.146E-10 | Old |
| 1182 | 19 | 19-44856329-C-T | rs56317818 | NECTIN2 | 0.2603 | T | 2.28E-31 | 0.008669 | ++ | 2.994E-28 | 2.368E-11 | Old |
| 1182 | 19 | 19-44856688-G-A | rs183610051 | NECTIN2 | 0.0097 | A | 3.52E-14 | 0.011575 | ++ | 3.512E-15 | 3.659E-11 | Old |
| 1182 | 19 | 19-44858325-C-CT | | NECTIN2 | 0.1712 | CT | 9.74E-11 | 0.067805 | -- | 2.042E-10 | | Old |
| 1182 | 19 | 19-44860563-G-T | rs138607350 | NECTIN2 | 0.0081 | G | 8.21E-24 | 0.01687 | ++ | 1.579E-23 | 3.11E-10 | Old |
| 1182 | 19 | 19-44861785-G-A | rs538568659 | NECTIN2 | 0.0042 | G | 1.25E-11 | 0.085424 | ++ | 1.433E-11 | 0.00001178 | Old |
| 1182 | 19 | 19-44862190-G-A | rs146275714 | NECTIN2 | 0.0187 | A | 3.97E-50 | 0.000294 | ++ | 1.034E-48 | 5.811E-21 | Old |
| 1182 | 19 | 19-44863241-G-A | rs183427010 | NECTIN2 | 0.0055 | A | 3.63E-12 | 0.532183 | ++ | 2.946E-10 | 8.641E-10 | Old |
| 1182 | 19 | 19-44864245-G-C | rs73050216 | NECTIN2 | 0.1703 | C | 2.71E-10 | 0.033944 | -- | 1.546E-10 | 0.000001784 | Old |
| 1182 | 19 | 19-44866307-AG-A | rs34111552 | NECTIN2 | 0.0065 | A | 1.82E-11 | 0.401822 | ++ | 3.432E-10 | 7.399E-08 | Old |
| 1182 | 19 | 19-44867581-G-A | rs12610605 | NECTIN2 | 0.1744 | A | 2.74E-10 | 0.023362 | -- | 8.964E-11 | 0.000001111 | Old |
| 1182 | 19 | 19-44868428-AT-A | rs34165484 | NECTIN2 | 0.1889 | AT | 5.86E-11 | 0.043718 | -- | 7.762E-11 | 5.583E-07 | Old |
| 1182 | 19 | 19-44873636-AAC-A | rs34606745 | NECTIN2 | 0.0866 | AAC | 1.46E-65 | 0.008022 | ++ | 3.427E-54 | | Old |
| 1182 | 19 | 19-44875268-C-CT | rs57907894 | NECTIN2 | 0.1975 | CT | 9.05E-10 | 0.030745 | -- | 4.378E-10 | | Old |

| | | | | | | | | | | | | |
|---|---|---|---|---|---|---|---|---|---|---|---|---|
| 1182 | 19 | 19-44877713-T-G | | NECTIN2 | 0.4745 | G | 5.12E-17 | 0.009763 | -- | 7.49E-17 | 7.157E-15 | Old |
| 1182 | 19 | 19-44879460-A-G | rs406456 | NECTIN2 | 0.3735 | A | 1.28E-28 | 0.000157 | ++ | 2.292E-29 | 1.714E-07 | Old |
| 1182 | 19 | 19-44880774-AT-A | | NECTIN2 | 0.0134 | A | 8.08E-14 | 0.052561 | ++ | 2.853E-13 | | Old |
| 1182 | 19 | 19-44881674-G-A | rs79701229 | NECTIN2 | 0.0116 | A | 4.5E-20 | 0.042705 | ++ | 2.464E-19 | 8.818E-13 | Old |
| 1182 | 19 | 19-44882099-C-A | rs144261139 | AC011481.2 | 0.0108 | A | 3.1E-28 | 0.028966 | ++ | 1.681E-26 | 0.1683 | Old |
| 1182 | 19 | 19-44882783-ATT-A | | AC011481.2 | 0.1907 | A | | 1.66E-08 | ?+ | 1.656E-08 | | Old |
| 1182 | 19 | 19-44883210-GTAA-G | rs142042446 | AC011481.2 | 0.1325 | GTAA | 4.2E-210 | 2.42E-15 | ++ | 9.09E-202 | 1.61E-136 | Old |
| 1182 | 19 | 19-44883377-C-T | rs147636938 | AC011481.2 | 0.0242 | T | 3.7E-52 | 0.00027 | ++ | 5.353E-49 | 1.348E-32 | Old |
| 1182 | 19 | 19-44884202-G-C | rs12972156 | AC011481.2 | 0.1323 | G | 3.2E-210 | 2.6E-15 | ++ | 7.7E-202 | 4.86E-140 | Old |
| 1182 | 19 | 19-44884339-G-A | rs12972970 | AC011481.2 | 0.1324 | A | 1.1E-209 | 1.06E-15 | ++ | 3.3E-202 | 2.69E-140 | Old |
| 1182 | 19 | 19-44884873-G-A | rs34342646 | AC011481.2 | 0.136 | A | 1.8E-203 | 3E-14 | ++ | 7.26E-194 | 4.9E-135 | Old |
| 1182 | 19 | 19-44885243-G-A | rs283811 | AC011481.2 | 0.2366 | G | 8.1E-195 | 6.03E-11 | ++ | 3.12E-174 | 7.23E-120 | Old |
| 1182 | 19 | 19-44887076-A-G | rs283815 | AC011481.2 | 0.2408 | A | 1.2E-203 | 1.55E-12 | ++ | 5.14E-184 | 3.05E-11 | Old |
| 1182 | 19 | 19-44888997-C-T | rs6857 | AC011481.2 | 0.1543 | T | 1.8E-287 | 1.25E-21 | ++ | 2.42E-277 | 2.33E-189 | Old |
| 1182 | 19 | 19-44890259-C-T | rs117310449 | AC011481.2 | 0.0104 | T | 1.79E-29 | 0.028844 | ++ | 1.399E-27 | 3.589E-18 | Old |
| 1182 | 19 | 19-44891079-C-T | rs71352238 | TOMM40 | 0.1326 | C | 3.3E-214 | 1.97E-16 | ++ | 3.36E-207 | 4.62E-143 | Old |
| 1182 | 19 | 19-44891712-G-T | rs184017 | TOMM40 | 0.2369 | G | 5.1E-203 | 1.11E-12 | ++ | 6.68E-184 | 1.44E-124 | Old |
| 1182 | 19 | 19-44892009-A-G | rs157580 | TOMM40 | 0.3709 | A | 8.95E-64 | 8.51E-06 | -- | 1.931E-60 | | Old |
| 1182 | 19 | 19-44892362-G-A | rs2075650 | TOMM40 | 0.1386 | G | 8E-222 | 8.91E-18 | ++ | 8.01E-214 | 7.79E-130 | Old |
| 1182 | 19 | 19-44892457-C-T | rs157581 | TOMM40 | 0.2402 | C | 1.4E-207 | 7.23E-13 | ++ | 8.82E-188 | | Old |
| 1182 | 19 | 19-44892587-G-A | rs34095326 | TOMM40 | 0.0979 | A | 7.5E-165 | 7.83E-12 | ++ | 7.73E-158 | 2.391E-85 | Old |

| | | | | | | | | | | | | |
|---|---|---|---|---|---|---|---|---|---|---|---|---|
| 1182 | 19 | 19-44892652-G-C | rs34404554 | TOMM40 | 0.1359 | G | 4.2E-224 | 2.89E-18 | ++ | 2.72E-217 | 5.35E-135 | Old |
| 1182 | 19 | 19-44892887-C-T | rs11556505 | TOMM40 | 0.137 | T | 1.3E-222 | 4.98E-18 | ++ | 2.63E-215 | 1.93E-133 | Old |
| 1182 | 19 | 19-44892962-C-T | rs157582 | TOMM40 | 0.2385 | T | 6.4E-208 | 3.2E-13 | ++ | 5.76E-189 | 2.46E-135 | Old |
| 1182 | 19 | 19-44893408-G-T | rs59007384 | TOMM40 | 0.2146 | T | 1E-211 | 1.83E-11 | ++ | 1.14E-188 | 4.58E-11 | Old |
| 1182 | 19 | 19-44893716-G-A | rs77301115 | TOMM40 | 0.0265 | A | 6.28E-53 | 9.7E-05 | ++ | 3.72E-50 | 5.579E-54 | Old |
| 1182 | 19 | 19-44894050-C-T | rs112849259 | TOMM40 | 0.0264 | T | 2.09E-53 | 0.000145 | ++ | 3.279E-50 | 5.136E-55 | Old |
| 1182 | 19 | 19-44894261-C-CTT | rs35647923 | TOMM40 | 0.4157 | CTT | 3.09E-10 | 0.007278 | -- | 2.254E-11 | | Old |
| 1182 | 19 | 19-44894695-C-T | rs116881820 | TOMM40 | 0.0265 | C | 5.42E-54 | 0.000134 | ++ | 9.889E-51 | 1.744E-53 | Old |
| 1182 | 19 | 19-44895376-G-C | rs11668327 | TOMM40 | 0.1589 | C | 2.96E-30 | 0.00578 | -- | 3.057E-28 | 8.595E-22 | Old |
| 1182 | 19 | 19-44895528-C-T | rs79398853 | TOMM40 | 0.0263 | T | 8.59E-54 | 0.000256 | ++ | 4.564E-50 | 7.326E-55 | Old |
| 1182 | 19 | 19-44896087-G-T | rs75687619 | TOMM40 | 0.0263 | T | 2.46E-53 | 0.000188 | ++ | 6.064E-50 | 0.006838 | Old |
| 1182 | 19 | 19-44896639-G-A | rs76366838 | TOMM40 | 0.0264 | A | 2.08E-53 | 0.000213 | ++ | 6.766E-50 | 0.006838 | Old |
| 1182 | 19 | 19-44897227-TG-T | | TOMM40 | 0.0271 | TG | 8.46E-55 | 1.95E-05 | ++ | 7.966E-53 | | Old |
| 1182 | 19 | 19-44897468-C-T | rs114536010 | TOMM40 | 0.0263 | T | 1.97E-53 | 0.000226 | ++ | 6.941E-50 | 1.386E-54 | Old |
| 1182 | 19 | 19-44897776-C-CA | | TOMM40 | 0.4129 | CA | 3.55E-31 | 0.408197 | -- | 8.94E-24 | | Old |
| 1182 | 19 | 19-44897790-AG-A | rs1555789087 | TOMM40 | 0.1241 | AG | 7.7E-266 | 5.32E-19 | ++ | 3.22E-257 | | Old |
| 1182 | 19 | 19-44898409-G-A | rs8106922 | TOMM40 | 0.3815 | G | 4.72E-30 | 0.232396 | -- | 1.39E-23 | 4.969E-40 | Old |
| 1182 | 19 | 19-44898730-G-A | | TOMM40 | 0.0032 | A | 6.27E-08 | 0.072424 | ++ | 2.012E-08 | 0.3111 | Old |
| 1182 | 19 | 19-44899220-C-T | rs34878901 | TOMM40 | 0.3985 | T | 1.42E-30 | 0.169324 | -- | 3.294E-24 | 0.000301 | Old |
| 1182 | 19 | 19-44899959-C-T | rs115881343 | TOMM40 | 0.0278 | T | 1.02E-55 | 0.000401 | ++ | 2.504E-51 | 3.016E-57 | Old |
| 1182 | 19 | 19-44900155-C-T | rs1160985 | TOMM40 | 0.4597 | T | 1.1E-44 | 0.00203 | -- | 7.095E-40 | 3.798E-85 | Old |

| | | | | | | | | | | | | |
|---|---|---|---|---|---|---|---|---|---|---|---|---|
| 1182 | 19 | 19-44901174-C-T | rs741780 | TOMM40 | 0.4598 | C | 3.13E-44 | 0.002199 | -- | 1.917E-39 | 5.139E-84 | Old |
| 1182 | 19 | 19-44901434-G-A | | TOMM40 | 0.2644 | A | 3.6E-29 | 0.002029 | -- | 5.088E-28 | 1.392E-20 | Old |
| 1182 | 19 | 19-44901548-AACACGGTGAAACTCCGTCTCTACT-A | rs113492558 | TOMM40 | 0.0291 | A | 1.93E-57 | 4.69E-05 | ++ | 2.867E-54 | 1.152E-55 | Old |
| 1182 | 19 | 19-44901600-C-T | rs112019714 | TOMM40 | 0.0291 | C | 2.11E-57 | 4.65E-05 | ++ | 3.059E-54 | 0.00277 | Old |
| 1182 | 19 | 19-44901715-C-T | rs1038025 | TOMM40 | 0.4598 | C | 1.47E-44 | 0.002393 | -- | 1.298E-39 | 2.802E-84 | Old |
| 1182 | 19 | 19-44902264-G-C | rs1305062 | TOMM40 | 0.3918 | C | 5.31E-31 | 0.095291 | -- | 2.671E-25 | 4.689E-47 | Old |
| 1182 | 19 | 19-44903416-G-A | rs10119 | TOMM40 | 0.2845 | A | 1.8E-143 | 3.21E-09 | ++ | 4.07E-131 | 1.67E-161 | Old |
| 1182 | 19 | 19-44905579-G-T | | APOE | 0.4701 | T | 4.22E-30 | 0.007666 | ++ | 1.754E-27 | 4.385E-36 | Old |
| 1182 | 19 | 19-44905910-G-C | | APOE | 0.3484 | C | 1.1E-46 | 4.38E-05 | -- | 4.297E-45 | 3.983E-33 | Old |
| 1182 | 19 | 19-44906745-G-A | rs769449 | APOE | 0.1106 | A | 2.1E-299 | 4.75E-23 | ++ | 1.23E-291 | 6.74E-13 | Old |
| 1182 | 19 | 19-44907187-G-A | rs769450 | APOE | 0.3891 | A | 2.6E-30 | 0.051863 | -- | 1.41E-25 | 2.699E-49 | Old |
| 1182 | 19 | 19-44908684-C-T | rs429358 | APOE | 0.1521 | C | 0 | 8.37E-31 | ++ | 3.26E-305 | 7.24E-305 | Old |
| 1182 | 19 | 19-44908822-C-T | rs7412 | APOE | 0.079 | T | 7.89E-31 | 1.53E-07 | -- | 1.321E-35 | 5.297E-39 | Old |
| 1182 | 19 | 19-44909521-C-CT | | AC011481.3 | 0.0435 | C | | 1.47E-14 | ?+ | 1.468E-14 | | Old |
| 1182 | 19 | 19-44909665-AC-A | rs537741299 | AC011481.3 | 0.007 | A | 2.58E-16 | 0.010115 | ++ | 1.56E-16 | 7.909E-14 | Old |
| 1182 | 19 | 19-44909698-C-A | rs1081105 | AC011481.3 | 0.0281 | C | 1.92E-58 | 0.000112 | ++ | 1.418E-54 | 1.351E-62 | Old |
| 1182 | 19 | 19-44909967-TGG-T | | AC011481.3 | 0.1109 | T | | 9.29E-28 | ?+ | 9.288E-28 | | Old |
| 1182 | 19 | 19-44909976-G-T | rs1065853 | AC011481.3 | 0.0802 | T | 3.06E-30 | 1.65E-07 | -- | 5.145E-35 | | Old |
| 1182 | 19 | 19-44910319-C-T | rs75627662 | AC011481.3 | 0.1916 | T | 5.8E-120 | 7.29E-06 | ++ | 3.96E-106 | 1.148E-84 | Old |
| 1182 | 19 | 19-44912456-G-A | rs10414043 | AC011481.3 | 0.1261 | A | 3.1E-289 | 1.51E-20 | ++ | 7.93E-275 | 3.56E-146 | Old |

| | | | | | | | | | | | | |
|---|---|---|---|---|---|---|---|---|---|---|---|---|
| 1182 | 19 | 19-44912678-G-T | rs7256200 | AC011481.3 | 0.126 | T | 2.2E-288 | 1.61E-20 | ++ | 5.4E-274 | | Old |
| 1182 | 19 | 19-44912921-G-T | rs483082 | AC011481.3 | 0.2449 | T | 9.3E-181 | 1.79E-09 | ++ | 3.74E-161 | 3.72E-139 | Old |
| 1182 | 19 | 19-44913034-C-T | rs59325138 | AC011481.3 | 0.3774 | T | 1.05E-26 | 0.009163 | -- | 1.092E-24 | 3.511E-39 | Old |
| 1182 | 19 | 19-44913484-C-T | rs438811 | AC011481.3 | 0.248 | T | 6.7E-182 | 1.84E-10 | ++ | 2.59E-164 | 9.06E-161 | Old |
| 1182 | 19 | 19-44914381-C-CTTCG | rs11568822 | AC011481.3 | 0.2283 | CTTCG | 1.1E-178 | 8.53E-11 | ++ | 1.61E-163 | 3.27E-124 | Old |
| 1182 | 19 | 19-44915229-G-A | rs12691088 | APOC1 | 0.0196 | A | 1.63E-53 | 0.001741 | ++ | 2.587E-50 | 0.06546 | Old |
| 1182 | 19 | 19-44915533-C-T | rs5117 | APOC1 | 0.2283 | C | 9.4E-177 | 3.62E-10 | ++ | 1.25E-160 | 8.74E-129 | Old |
| 1182 | 19 | 19-44916825-C-A | rs73052335 | APOC1 | 0.0893 | C | | 7.55E-21 | ?+ | 7.552E-21 | | Old |
| 1182 | 19 | 19-44917843-G-A | rs3925681 | APOC1 | 0.397 | A | 1.95E-35 | 0.00121 | -- | 2.294E-33 | 1.001E-42 | Old |
| 1182 | 19 | 19-44917947-C-T | rs150966173 | APOC1 | 0.029 | T | 1.28E-56 | 0.001165 | ++ | 7.279E-51 | 4.119E-60 | Old |
| 1182 | 19 | 19-44917961-GA-G | rs374095935 | APOC1 | 0.0114 | GA | 4.29E-31 | 0.042697 | ++ | 1.099E-28 | | Old |
| 1182 | 19 | 19-44917997-G-A | rs12721046 | APOC1 | 0.1385 | A | 1.7E-230 | 3.32E-21 | ++ | 4.97E-231 | 6.77E-159 | Old |
| 1182 | 19 | 19-44918715-AG-A | rs12721052 | APOC1 | 0.3278 | A | 5.04E-20 | 0.027151 | -- | 1.867E-18 | 2.636E-31 | Old |
| 1182 | 19 | 19-44918903-G-C | rs12721051 | APOC1 | 0.1707 | G | 6.4E-292 | 2.61E-22 | ++ | 1.57E-290 | 2.14E-228 | Old |
| 1182 | 19 | 19-44919589-G-A | rs56131196 | APOC1 | 0.1805 | A | 2.1E-291 | 1.15E-23 | ++ | 7.74E-288 | 1.89E-202 | Old |
| 1182 | 19 | 19-44919689-G-A | rs4420638 | APOC1 | 0.181 | G | 1.5E-297 | 8.06E-24 | ++ | 3.77E-288 | 2.22E-202 | Old |
| 1182 | 19 | 19-44920730-C-CA | rs35733971 | APOC1, APOC4 | 0.1693 | CA | 3.3E-226 | 3.92E-17 | ++ | 1.59E-215 | | Old |
| 1182 | 19 | 19-44921093-TAA-T | rs368340812 | APOC1, APOC4 | 0.0957 | T | | 3.33E-19 | ?+ | 3.335E-19 | | Old |
| 1182 | 19 | 19-44921095-ATTTT-A | rs759515809 | APOC1, APOC4 | 0.1107 | ATTTT | 0.946693 | 2.64E-19 | ++ | 2.675E-19 | | Old |
| 1182 | 19 | 19-44921809-G-A | rs188535946 | APOC1, APOC4 | 0.0286 | A | 3.89E-55 | 0.000198 | ++ | 1.871E-51 | | Old |
| 1182 | 19 | 19-44923868-T-A | rs111789331 | APOC1, APOC4 | 0.1397 | A | 9.4E-232 | 3.29E-19 | ++ | 2.14E-228 | 1.84E-158 | Old |

| | | | | | | | | | | | | |
|---|---|---|---|---|---|---|---|---|---|---|---|---|
| 1182 | 19 | 19-44924977-G-A | rs66626994 | APOC1, APOC4 | 0.1471 | A | 3.1E-227 | 1.71E-19 | ++ | 5.84E-223 | 1.17E-130 | Old |
| 1182 | 19 | 19-44925202-C-T | rs4803772 | APOC1, APOC4 | 0.3216 | T | 9.25E-20 | 0.030274 | -- | 3.768E-18 | 2.536E-31 | Old |
| 1182 | 19 | 19-44926451-G-C | rs60049679 | APOC1, APOC4 | 0.0877 | C | 9.94E-24 | 0.00795 | ++ | 3.926E-22 | 4.495E-23 | Old |
| 1182 | 19 | 19-44928426-GAA-G | rs569925552 | APOC1, APOC4 | 0.0121 | GAA | 5.06E-12 | 0.013102 | ++ | 1.046E-12 | | Old |
| 1182 | 19 | 19-44935297-C-T | rs7254133 | APOC1, APOC4 | 0.3102 | T | 2.84E-30 | 0.016581 | ++ | 6.177E-27 | 3.048E-14 | Old |
| 1182 | 19 | 19-44935318-C-A | rs141441332 | APOC1, APOC4 | 0.012 | A | 6.88E-11 | 0.015409 | ++ | 1.273E-11 | 5.826E-08 | Old |
| 1182 | 19 | 19-44973974-G-C | rs57465754 | CLPTM1 | 0.2396 | G | 5.2E-10 | 0.143058 | ++ | 4.541E-09 | 0.001831 | Old |
| 1183 | 19 | 19-44693694-G-A | rs151330717 | AC243964.2 | 0.0178 | A | 2.44E-06 | 0.0042 | -- | 3.499E-08 | 0.0007302 | Old |
| 1183 | 19 | 19-44728555-G-A | rs1551891 | RF00285, BCL3 | 0.088 | A | 8.71E-08 | 0.014381 | -- | 7.159E-09 | 0.005049 | Old |
| 1183 | 19 | 19-44730062-AAATTTATAG-A | | RF00285, BCL3 | 0.0886 | A | 1.06E-07 | 0.024845 | -- | 1.703E-08 | 0.002298 | Old |
| 1183 | 19 | 19-44732593-AAG-A | | RF00285, BCL3 | 0.088 | A | 6.35E-08 | 0.016805 | -- | 6.575E-09 | 0.001493 | Old |
| 1183 | 19 | 19-44732598-TGAA-T | | RF00285, BCL3 | 0.088 | T | 6.35E-08 | 0.016787 | -- | 6.566E-09 | 0.001524 | Old |
| 1183 | 19 | 19-44736280-C-A | rs62117162 | RF00285, BCL3 | 0.0952 | A | 8.25E-08 | 0.048557 | -- | 3.844E-08 | 0.0005059 | Old |
| 1183 | 19 | 19-44739710-C-T | rs62117204 | RF00285, BCL3 | 0.0713 | T | 2.11E-08 | 0.034097 | -- | 5.973E-09 | 0.008649 | Old |
| 1183 | 19 | 19-44744370-G-A | rs4803750 | RF00285, BCL3 | 0.0776 | G | 5.45E-09 | 0.00134 | -- | 3.684E-11 | 0.007579 | Old |
| 1183 | 19 | 19-44748549-G-T | rs531660643 | BCL3 | 0.0202 | T | 6.22E-08 | 0.001954 | -- | 4.527E-10 | 0.00003602 | Old |
| 1183 | 19 | 19-44750234-C-T | rs10401176 | BCL3 | 0.1425 | T | 8.15E-11 | 0.010885 | -- | 1.682E-11 | 0.0001742 | Old |
| 1183 | 19 | 19-44750354-ATTGGC-A | rs66586168 | BCL3 | 0.1398 | A | 1.06E-10 | 0.00564 | -- | 7.723E-12 | | Old |
| 1183 | 19 | 19-44752009-C-T | rs62117205 | BCL3 | 0.0685 | C | 5.19E-09 | 0.013723 | -- | 4.738E-10 | 0.002023 | Old |
| 1183 | 19 | 19-44752422-G-C | rs62117206 | BCL3 | 0.0696 | C | 4.28E-09 | 0.012994 | -- | 3.834E-10 | 0.0038 | Old |
| 1183 | 19 | 19-44799247-G-A | rs148933445 | CBLC | 0.0201 | A | 1.47E-07 | 0.010288 | -- | 5.973E-09 | 0.000002 | Old |

| | | | | | | | | | | | | |
|---|---|---|---|---|---|---|---|---|---|---|---|---|
| 1183 | 19 | 19-44844304-T-G | rs11668738 | BCAM, NECTIN2 | 0.165 | G | 5.33E-09 | 0.113785 | -- | 1.195E-08 | 0.6935 | Old |
| 1183 | 19 | 19-44850981-C-G | rs2972566 | NECTIN2 | 0.1829 | C | 4.12E-10 | 0.025487 | -- | 1.626E-10 | 0.9554 | Old |
| 1183 | 19 | 19-44881148-C-T | rs73052307 | NECTIN2 | 0.137 | C | 3.75E-25 | 0.033998 | -- | 1.579E-22 | 0.02503 | Old |
| 1183 | 19 | 19-44916968-TA-T | rs753649749 | APOC1 | 0.0271 | TA | 1.65E-11 | 0.040917 | -- | 1.639E-11 | | Old |
| 1184 | 19 | 19-44714520-G-C | | AC243964.2 | 0.1378 | C | 7.13E-08 | 0.082528 | ++ | 4.967E-08 | 0.05779 | Old |
| 1184 | 19 | 19-44718265-T-C | rs56198711 | AC243964.2 | 0.1427 | C | 4.05E-08 | 0.101429 | ++ | 4.63E-08 | 0.1241 | Old |
| 1184 | 19 | 19-44719517-G-C | rs73037426 | AC243964.2 | 0.1418 | C | 1.66E-09 | 0.023509 | ++ | 3.362E-10 | 0.000003162 | Old |
| 1184 | 19 | 19-44731001-AAAAAAAAAAAAAAAGAAAAG-A | | RF00285, BCL3 | 0.1643 | AAAAAAA | 5.13E-09 | 0.019179 | ++ | 7.108E-10 | | Old |
| 1184 | 19 | 19-44735929-A-G | rs2927436 | RF00285, BCL3 | 0.3444 | A | 4.02E-07 | 0.004164 | -- | 7.088E-09 | 0.7411 | Old |
| 1184 | 19 | 19-44737327-C-T | rs55923289 | RF00285, BCL3 | 0.1178 | C | 5.13E-09 | 0.006218 | ++ | 1.562E-10 | 0.000003661 | Old |
| 1184 | 19 | 19-44738850-G-A | rs2927438 | RF00285, BCL3 | 0.2069 | A | 9.35E-17 | 0.015781 | ++ | 2.102E-16 | 0.0004052 | Old |
| 1184 | 19 | 19-44746404-C-T | rs12459810 | RF00285, BCL3 | 0.2696 | T | 1E-06 | 0.000198 | ++ | 8.646E-10 | 4.125E-08 | Old |
| 1184 | 19 | 19-44750911-C-A | rs8103315 | BCL3 | 0.1182 | A | 6.37E-15 | 0.01398 | ++ | 2.719E-15 | 3.416E-08 | Old |
| 1185 | 19 | 19-44720227-G-A | rs111740474 | AC243964.2 | 0.0143 | A | 4.98E-09 | 0.004182 | ++ | 8.6E-11 | 0.0001601 | Old |
| 1185 | 19 | 19-44833336-C-T | rs112616980 | BCAM, NECTIN2 | 0.0044 | T | 4.09E-14 | 0.002434 | ++ | 1.087E-15 | 0.0003382 | Old |
| 1186 | 19 | 19-44728895-G-A | rs62117160 | RF00285, BCL3 | 0.0389 | A | 1.15E-09 | 0.001297 | -- | 6.229E-12 | 0.00009026 | Old |
| 1186 | 19 | 19-44730118-A-G | rs62117161 | RF00285, BCL3 | 0.086 | A | 9.11E-08 | 0.011131 | -- | 5.22E-09 | 0.001118 | Old |
| 1186 | 19 | 19-44739483-G-A | rs2927439 | RF00285, BCL3 | 0.3517 | G | 7.26E-09 | 0.000567 | -- | 1.954E-11 | 0.0004962 | Old |
| 1186 | 19 | 19-44743791-C-T | rs4803748 | RF00285, BCL3 | 0.4247 | T | 3.49E-12 | 0.000491 | -- | 1.478E-14 | 0.000001162 | Old |
| 1186 | 19 | 19-44747899-C-A | rs2965169 | BCL3 | 0.4307 | C | 1.28E-11 | 3.08E-05 | -- | 2.151E-15 | 2.379E-07 | Old |
| 1186 | 19 | 19-44792629-G-A | rs113330691 | CBLC | 0.0339 | A | 1.76E-06 | 0.005318 | -- | 3.506E-08 | 1.183E-09 | Old |

| | | | | | | | | | | | | |
|---|---|---|---|---|---|---|---|---|---|---|---|---|
| 1186 | 19 | 19-44793549-C-T | rs3208856 | CBLC | 0.0339 | T | 1.78E-06 | 0.004623 | -- | 3.06E-08 | 1.229E-09 | Old |
| 1186 | 19 | 19-44795942-G-T | rs76560105 | CBLC | 0.0331 | T | 1.36E-06 | 0.003612 | -- | 1.852E-08 | 3.133E-08 | Old |
| 1186 | 19 | 19-44813331-G-A | rs28399654 | BCAM | 0.0289 | A | 6.12E-07 | 0.015605 | -- | 4.233E-08 | 3.081E-12 | Old |
| 1186 | 19 | 19-44816374-G-A | rs118147862 | BCAM | 0.0399 | A | 4.21E-12 | 0.00329 | -- | 8.954E-14 | 6.472E-07 | Old |
| 1186 | 19 | 19-44865946-G-A | rs112422902 | NECTIN2 | 0.0298 | A | 2.23E-07 | 0.011587 | -- | 1.327E-08 | 9.3E-19 | Old |
| 1186 | 19 | 19-44879418-G-A | rs41290120 | NECTIN2 | 0.0417 | A | 9.03E-16 | 0.012804 | -- | 5.526E-16 | 0.03073 | Old |
| 1186 | 19 | 19-44886339-G-A | rs7254892 | AC011481.2 | 0.0446 | A | 6.75E-13 | 0.000436 | -- | 8.434E-15 | | Old |
| 1186 | 19 | 19-44893972-G-A | rs1160983 | TOMM40 | 0.037 | A | 1.43E-13 | 6.81E-05 | -- | 1.499E-16 | 1.06E-13 | Old |
| 1186 | 19 | 19-44897490-T-A | rs61679753 | TOMM40 | 0.0404 | A | 9.9E-14 | 0.000293 | -- | 8.891E-16 | 1.638E-17 | Old |
| 1186 | 19 | 19-44899005-G-T | rs111784051 | TOMM40 | 0.0421 | G | 6E-14 | 0.003113 | -- | 2.023E-14 | 1.231E-17 | Old |
| 1186 | 19 | 19-44921921-G-A | rs190712692 | APOC1, APOC4 | 0.055 | A | 9.77E-21 | 0.000105 | -- | 1.064E-22 | | Old |
| 1186 | 19 | 19-44923535-G-A | rs141622900 | APOC1, APOC4 | 0.055 | A | 1.55E-20 | 0.000222 | -- | 4.792E-22 | 5.358E-25 | Old |
| 1187 | 19 | 19-44738916-G-A | rs1531517 | RF00285, BCL3 | 0.0876 | A | 4.81E-09 | 0.105996 | -- | 1.551E-08 | 0.2331 | Old |
| 1188 | 19 | 19-44766291-C-T | rs193249943 | RF00156, CBLC | 0.0059 | T | 4.13E-10 | 0.032331 | ++ | 1.067E-10 | 0.0004719 | Old |
| 1188 | 19 | 19-44815238-G-A | rs180887453 | BCAM | 0.0034 | G | 1.02E-10 | 0.011046 | ++ | 6.671E-12 | 0.0006873 | Old |
| 1188 | 19 | 19-44869757-GT-G | | NECTIN2 | 0.0015 | G | 3.86E-07 | 0.0202 | ++ | 2.387E-08 | 0.003089 | Old |
| 1188 | 19 | 19-44882164-T-C | rs369077933 | AC011481.2 | 0.0016 | T | 4.62E-07 | 0.027219 | ++ | 3.716E-08 | 0.003097 | Old |
| 1188 | 19 | 19-44900801-C-T | rs142412517 | TOMM40 | 0.0016 | T | 9.49E-08 | 0.010016 | ++ | 3.223E-09 | 0.004724 | Old |
| 1189 | 19 | 19-44766468-C-T | rs139326841 | RF00156, CBLC | 0.0213 | C | 3.62E-10 | 0.38753 | ++ | 4.921E-09 | 0.000003711 | Old |
| 1189 | 19 | 19-44807827-G-A | rs140824606 | AC092306.1, BCAM | 0.0232 | A | 1.68E-10 | 0.211157 | ++ | 8.684E-10 | 2.738E-07 | Old |
| 1189 | 19 | 19-44841241-TTAAAAA-T | rs71171294 | BCAM, NECTIN2 | 0.0113 | TTAAAAA | 7.35E-16 | 0.466616 | ++ | 5.992E-13 | | Old |

| | | | | | | | | | | | | |
|---|---|---|---|---|---|---|---|---|---|---|---|---|
| 1189 | 19 | 19-44842026-C-A | rs61642202 | BCAM, NECTIN2 | 0.0113 | C | 4.9E-17 | 0.180988 | ++ | 6.361E-15 | 0.00002925 | Old |
| 1189 | 19 | 19-44852010-T-A | rs76205446 | NECTIN2 | 0.0165 | A | 4.67E-15 | 0.313311 | ++ | 7.512E-12 | 0.003059 | Old |
| 1189 | 19 | 19-44854769-C-T | rs143459034 | NECTIN2 | 0.0121 | T | 5.81E-16 | 0.159859 | ++ | 6.083E-14 | 0.000884 | Old |
| 1189 | 19 | 19-44856410-G-A | rs41289514 | NECTIN2 | 0.0112 | G | 1.49E-16 | 0.153395 | ++ | 1.093E-14 | 0.000005607 | Old |
| 1190 | 19 | 19-44784672-C-CTCCAT | rs569705402 | CBLC | 0.0064 | C | 8.18E-10 | 0.553701 | ++ | 1.387E-08 | 0.0001013 | Old |
| 1191 | 19 | 19-44825110-A-T | rs58132661 | BCAM, NECTIN2 | 0.3172 | A | 8.07E-27 | 0.085196 | ++ | 1.551E-22 | 4.717E-08 | Old |
| 1191 | 19 | 19-44825122-T-A | rs58826447 | BCAM, NECTIN2 | 0.3173 | A | 8.33E-27 | 0.086004 | ++ | 1.627E-22 | 3.098E-08 | Old |
| 1191 | 19 | 19-44825123-C-A | rs58446550 | BCAM, NECTIN2 | 0.3173 | A | 8.33E-27 | 0.086978 | ++ | 1.676E-22 | 3.365E-08 | Old |
| 1191 | 19 | 19-44825150-GAC-G | rs34798982 | BCAM, NECTIN2 | 0.4908 | G | 5.03E-16 | 0.085684 | ++ | 2.48E-14 | | Old |
| 1191 | 19 | 19-44827846-A-G | rs7343130 | BCAM, NECTIN2 | 0.4235 | G | 9.15E-10 | 0.017058 | -- | 1.679E-10 | | Old |
| 1191 | 19 | 19-44829763-TA-T | | BCAM, NECTIN2 | 0.2349 | TA | 1.06E-08 | 0.036301 | -- | 4.298E-09 | | Old |
| 1191 | 19 | 19-44832419-G-A | rs56394238 | BCAM, NECTIN2 | 0.4471 | G | 8.44E-21 | 0.009124 | ++ | 5.332E-20 | 0.005204 | Old |
| 1191 | 19 | 19-44832778-C-T | rs7359852 | BCAM, NECTIN2 | 0.3072 | C | 2.59E-24 | 0.007045 | ++ | 6.742E-23 | 0.00001223 | Old |
| 1191 | 19 | 19-44833186-G-A | rs3021439 | BCAM, NECTIN2 | 0.2996 | A | 4.36E-24 | 0.006242 | ++ | 7.125E-23 | 0.0000278 | Old |
| 1191 | 19 | 19-44834128-G-C | rs2927480 | BCAM, NECTIN2 | 0.2943 | C | 4.21E-24 | 0.006474 | ++ | 6.096E-23 | | Old |
| 1191 | 19 | 19-44834247-G-C | rs11667241 | BCAM, NECTIN2 | 0.317 | C | 1.21E-20 | 0.022191 | ++ | 4.364E-19 | 0.00004 | Old |
| 1191 | 19 | 19-44834993-AT-A | rs200722375 | BCAM, NECTIN2 | 0.2925 | A | 1.5E-23 | 0.006143 | ++ | 1.526E-22 | 0.000001829 | Old |
| 1191 | 19 | 19-44837479-G-A | rs73048293 | BCAM, NECTIN2 | 0.2989 | A | 1.55E-23 | 0.013334 | ++ | 7.543E-22 | 0.0000101 | Old |
| 1191 | 19 | 19-44838647-G-A | rs12459575 | BCAM, NECTIN2 | 0.299 | A | 8.89E-24 | 0.010368 | ++ | 2.896E-22 | 0.00001349 | Old |
| 1191 | 19 | 19-44842530-A-T | rs111371860 | BCAM, NECTIN2 | 0.0588 | A | 3.87E-09 | 0.003659 | -- | 7.723E-11 | 4.612E-08 | Old |
| 1191 | 19 | 19-44844403-GA-G | | BCAM, NECTIN2 | 0.4313 | GA | 3.74E-11 | 0.432262 | -- | 3.086E-09 | | Old |

| | | | | | | | | | | | | |
|---|---|---|---|---|---|---|---|---|---|---|---|---|
| 1191 | 19 | 19-44844654-C-A | rs4452060 | BCAM, NECTIN2 | 0.4281 | A | 9.05E-11 | 0.066257 | ++ | 2.242E-10 | 0.02366 | Old |
| 1191 | 19 | 19-44849230-T-A | rs12974942 | NECTIN2 | 0.4125 | T | 1.08E-12 | 0.019276 | ++ | 8.305E-13 | 0.002585 | Old |
| 1191 | 19 | 19-44852464-G-C | rs2972559 | NECTIN2 | 0.2591 | G | 3.11E-29 | 0.016487 | ++ | 4.187E-26 | 8.952E-12 | Old |
| 1191 | 19 | 19-44853746-G-C | rs35396326 | NECTIN2 | 0.2703 | G | 5.28E-25 | 0.035454 | ++ | 3.578E-22 | 9.136E-09 | Old |
| 1191 | 19 | 19-44854034-G-C | rs4803763 | NECTIN2 | 0.2552 | C | 1.54E-29 | 0.011312 | ++ | 9.02E-27 | | Old |
| 1191 | 19 | 19-44854682-G-A | rs2927468 | NECTIN2 | 0.488 | A | 6.37E-32 | 0.004745 | -- | 2.827E-29 | 1.77E-08 | Old |
| 1191 | 19 | 19-44856449-G-A | rs12462573 | NECTIN2 | 0.2593 | A | 1.31E-31 | 0.006275 | ++ | 9.646E-29 | 5.326E-12 | Old |
| 1191 | 19 | 19-44858389-G-A | rs365653 | NECTIN2 | 0.1317 | G | 2.49E-16 | 0.045724 | -- | 9.822E-15 | 0.000002902 | Old |
| 1191 | 19 | 19-44858568-C-T | rs418227 | NECTIN2 | 0.3591 | C | 1.57E-18 | 0.001459 | ++ | 1.84E-19 | 0.001655 | Old |
| 1191 | 19 | 19-44858703-C-T | rs417193 | NECTIN2 | 0.353 | C | 1.31E-18 | 0.002519 | ++ | 3.503E-19 | 0.0001149 | Old |
| 1191 | 19 | 19-44859012-G-T | rs2436474 | NECTIN2 | 0.3494 | T | 9.07E-19 | 0.001636 | ++ | 1.285E-19 | 0.00003081 | Old |
| 1191 | 19 | 19-44859410-G-A | rs377702 | NECTIN2 | 0.3519 | A | 1.33E-18 | 0.001739 | ++ | 2.005E-19 | 0.0001232 | Old |
| 1191 | 19 | 19-44859997-G-A | rs387369 | NECTIN2 | 0.3509 | A | 2E-18 | 0.001734 | ++ | 2.813E-19 | 0.0002523 | Old |
| 1191 | 19 | 19-44860443-G-A | rs12978931 | NECTIN2 | 0.1948 | G | 2.84E-12 | 0.064 | -- | 1.27E-11 | 0.0006718 | Old |
| 1191 | 19 | 19-44860534-G-T | rs384973 | NECTIN2 | 0.3519 | G | 1.38E-18 | 0.001355 | ++ | 1.435E-19 | 0.0007124 | Old |
| 1191 | 19 | 19-44861366-C-T | rs411856 | NECTIN2 | 0.3512 | T | 8.37E-19 | 0.001636 | ++ | 1.225E-19 | 0.0003212 | Old |
| 1191 | 19 | 19-44861558-G-A | rs395710 | NECTIN2 | 0.3513 | G | 6.54E-19 | 0.001624 | ++ | 9.844E-20 | 0.000308 | Old |
| 1191 | 19 | 19-44861815-G-A | rs555608 | NECTIN2 | 0.3513 | G | 7.15E-19 | 0.001389 | ++ | 8.364E-20 | 0.000378 | Old |
| 1191 | 19 | 19-44861991-C-T | rs2436475 | NECTIN2 | 0.351 | C | 9.15E-19 | 0.001671 | ++ | 1.362E-19 | 0.0004852 | Old |
| 1191 | 19 | 19-44862219-C-T | rs11669109 | NECTIN2 | 0.3254 | C | | 2.51E-08 | ?+ | 2.512E-08 | 0.0003802 | Old |
| 1191 | 19 | 19-44862384-C-A | rs12980631 | NECTIN2 | 0.3515 | C | 8.15E-19 | 0.001187 | ++ | 7.292E-20 | 0.0004417 | Old |

| | | | | | | | | | | | | |
|---|---|---|---|---|---|---|---|---|---|---|---|---|
| 1191 | 19 | 19-44862475-AAAG-A | rs56283909 | NECTIN2 | 0.3515 | AAAG | 9.83E-19 | 0.001567 | ++ | 1.311E-19 | | Old |
| 1191 | 19 | 19-44862560-G-A | rs11665829 | NECTIN2 | 0.3516 | A | 1.2E-18 | 0.001464 | ++ | 1.397E-19 | | Old |
| 1191 | 19 | 19-44863088-G-A | rs416116 | NECTIN2 | 0.3512 | A | 1.45E-18 | 0.001353 | ++ | 1.452E-19 | 0.0008026 | Old |
| 1191 | 19 | 19-44863346-G-A | rs521629 | NECTIN2 | 0.3515 | A | 7.52E-19 | 0.001637 | ++ | 1.104E-19 | 0.0001002 | Old |
| 1191 | 19 | 19-44863522-C-T | rs519825 | NECTIN2 | 0.3514 | C | 9.43E-19 | 0.001713 | ++ | 1.438E-19 | 0.0001335 | Old |
| 1191 | 19 | 19-44864825-C-A | rs520283 | NECTIN2 | 0.3509 | A | 1.34E-18 | 0.002168 | ++ | 2.76E-19 | 0.0001408 | Old |
| 1191 | 19 | 19-44867313-A-C | rs565566 | NECTIN2 | 0.3493 | C | 5.65E-19 | 0.001885 | ++ | 1.077E-19 | 0.1351 | Old |
| 1191 | 19 | 19-44867392-A-G | rs564724 | NECTIN2 | 0.352 | G | 6.06E-19 | 0.000862 | ++ | 3.607E-20 | 0.0000802 | Old |
| 1191 | 19 | 19-44867416-A-C | rs510297 | NECTIN2 | 0.3525 | A | 7E-19 | 0.000889 | ++ | 4.299E-20 | 0.0001096 | Old |
| 1191 | 19 | 19-44874077-G-A | rs393584 | NECTIN2 | 0.3581 | A | 2.8E-12 | 0.029524 | -- | 3.209E-12 | | Old |
| 1191 | 19 | 19-44875803-C-A | rs387976 | NECTIN2 | 0.3613 | C | 3.53E-12 | 0.018995 | -- | 1.956E-12 | 0.1333 | Old |
| 1191 | 19 | 19-44880859-A-G | rs406315 | NECTIN2 | 0.3422 | A | 1.48E-24 | 0.000231 | ++ | 8.012E-26 | 4.532E-07 | Old |
| 1191 | 19 | 19-44893642-C-T | | TOMM40 | 0.4962 | T | 8E-12 | 0.284955 | ++ | 4.903E-10 | 4.006E-13 | Old |
| 1191 | 19 | 19-44894255-A-C | rs157585 | TOMM40 | 0.4991 | C | 6.49E-16 | 0.197835 | ++ | 1.599E-13 | | Old |
| 1191 | 19 | 19-44895007-C-T | rs157588 | TOMM40 | 0.4999 | C | 3.32E-16 | 0.228368 | ++ | 1.529E-13 | 6.807E-18 | Old |
| 1191 | 19 | 19-44895459-C-A | | TOMM40 | 0.4931 | A | 6.39E-18 | 0.26959 | ++ | 1.26E-14 | 9.478E-19 | Old |
| 1191 | 19 | 19-44905307-T-A | rs449647 | APOE | 0.1834 | T | 8.16E-27 | 0.064019 | -- | 4.518E-22 | 0.0002009 | Old |
| 1192 | 19 | 19-44836881-A-G | | BCAM, NECTIN2 | 0.3156 | A | 4.87E-16 | 0.144106 | -- | 9.291E-14 | 0.00007397 | Old |
| 1192 | 19 | 19-44838691-A-G | rs10407439 | BCAM, NECTIN2 | 0.3009 | G | 5.65E-17 | 0.101298 | -- | 7.281E-15 | 0.009062 | Old |
| 1193 | 19 | 19-44862268-C-CACTGTGTGTGGTGGCGGGCACCTG | rs144767421 | NECTIN2 | 0.3669 | C | 5.57E-19 | 0.717028 | ++ | 5.228E-19 | | Old |

| | | | | | | | | | | | | |
|---|---|---|---|---|---|---|---|---|---|---|---|---|
| 1194 | 19 | 19-44890947-G-A | rs561654715 | TOMM40 | 0.0027 | A | 1.49E-11 | 0.2552 | ++ | 1.171E-10 | 0.000003295 | Old |
| 1195 | 19 | 19-44921095-T-A | rs56369833 | APOC1, APOC4 | 0.1572 | T | 6.8E-228 | 0.683605 | ++ | 6.58E-228 | | Old |
| 1196 | 19 | 19-44925417-C-T | rs565334527 | APOC1, APOC4 | 0.0016 | C | 8.15E-11 | 0.05508 | ++ | 2.945E-11 | 0.003351 | Old |
| 1197 | 19 | 19-44933496-C-T | rs114533385 | APOC1, APOC4 | 0.0184 | T | 8.08E-11 | 0.006468 | ++ | 1.442E-11 | 0.0009272 | Old |
| 1197 | 19 | 19-44960141-A-G | rs576651896 | CLPTM1 | 0.0055 | A | 9.51E-07 | 0.003205 | ++ | 1.09E-08 | 0.002381 | Old |
| 1197 | 19 | 19-45008063-G-A | rs531438766 | RELB | 0.0047 | A | 2.1E-06 | 0.001696 | ++ | 1.306E-08 | 0.0001676 | Old |
| 1198 | 19 | 19-45094524-TA-T | rs149151450 | MARK4, PPP1R37 | 0.0922 | TA | 1.99E-09 | 0.008637 | ++ | 1.397E-10 | 0.002281 | Old |
| 1198 | 19 | 19-45102867-C-T | rs7248421 | MARK4, PPP1R37 | 0.0906 | T | 8.81E-10 | 0.007024 | ++ | 5.204E-11 | 0.0007487 | Old |
| 1198 | 19 | 19-45108968-C-T | rs12462040 | MARK4, PPP1R37 | 0.0901 | T | 1.76E-09 | 0.006878 | ++ | 9.131E-11 | 0.0008119 | Old |
| 1198 | 19 | 19-45111343-C-T | rs34545713 | MARK4, PPP1R37 | 0.0914 | T | 7.19E-10 | 0.004438 | ++ | 2.456E-11 | 0.002414 | Old |
| 1198 | 19 | 19-45115701-G-A | rs2004357 | MARK4, PPP1R37 | 0.0917 | A | 6.12E-10 | 0.006538 | ++ | 3.497E-11 | | Old |
| 1198 | 19 | 19-45118499-TG-T | rs144328302 | MARK4, PPP1R37 | 0.0892 | T | 4.98E-10 | 0.003829 | ++ | 1.426E-11 | 0.004282 | Old |
| 1198 | 19 | 19-45123977-C-T | rs10405086 | MARK4, PPP1R37 | 0.0921 | T | 5.18E-10 | 0.004553 | ++ | 1.939E-11 | 0.002723 | Old |
| 1198 | 19 | 19-45128558-G-A | rs17643262 | MARK4, PPP1R37 | 0.093 | A | 1.9E-10 | 0.003778 | ++ | 6.506E-12 | 0.009767 | Old |
| 1198 | 19 | 19-45130428-G-A | rs754366 | MARK4, PPP1R37 | 0.0941 | A | 1.83E-10 | 0.004432 | ++ | 7.927E-12 | 0.01058 | Old |
| 1198 | 19 | 19-45132943-C-T | rs1114832 | MARK4, PPP1R37 | 0.0916 | T | 2.88E-09 | 0.005988 | ++ | 1.155E-10 | 0.0005812 | Old |
| 1198 | 19 | 19-45133061-C-A | rs1114831 | MARK4, PPP1R37 | 0.0896 | A | 2.41E-10 | 0.004305 | ++ | 8.714E-12 | 0.1156 | Old |
| 1198 | 19 | 19-45134987-C-T | rs28620490 | MARK4, PPP1R37 | 0.0882 | T | 5.16E-10 | 0.004525 | ++ | 1.761E-11 | 0.0004345 | Old |
| 1198 | 19 | 19-45137848-C-A | rs77401305 | MARK4, PPP1R37 | 0.0925 | A | 4.81E-09 | 0.005497 | ++ | 1.675E-10 | | Old |
| 1198 | 19 | 19-45138786-G-A | rs10401157 | MARK4, PPP1R37 | 0.0887 | A | 3.14E-10 | 0.002211 | ++ | 4.753E-12 | 0.001052 | Old |
| 1198 | 19 | 19-45139287-G-C | rs10401823 | MARK4, PPP1R37 | 0.0897 | G | 4.74E-10 | 0.006445 | ++ | 2.625E-11 | 0.1232 | Old |

| | | | | | | | | | | | |
|---|---|---|---|---|---|---|---|---|---|---|---|
| 1198 | 19 | 19-45141096-T-A | rs78273125 | MARK4, PPP1R37 | 0.0885 | T | 7.37E-10 | 0.005501 | ++ | 3.074E-11 | 0.0003586 | Old |
| 1198 | 19 | 19-45146841-G-T | rs74846209 | PPP1R37 | 0.0895 | T | 2.53E-10 | 0.005053 | ++ | 1.077E-11 | 0.001013 | Old |
| 1198 | 19 | 19-45147128-C-T | rs1048699 | PPP1R37 | 0.0893 | T | 4.02E-10 | 0.00577 | ++ | 1.895E-11 | 0.0004258 | Old |
| 1198 | 19 | 19-45147979-G-A | rs113321260 | MARK4 | 0.0924 | A | 8.41E-11 | 0.009227 | ++ | 9.853E-12 | 0.00132 | Old |
| 1198 | 19 | 19-45148728-GGTAT-G | rs143019611 | MARK4 | 0.0895 | GGTAT | 5.55E-10 | 0.007919 | ++ | 3.789E-11 | 0.0007294 | Old |
| 1198 | 19 | 19-45152075-C-T | rs28469095 | NKPD1 | 0.0895 | C | 2.96E-10 | 0.005847 | ++ | 1.476E-11 | 0.000568 | Old |
| 1198 | 19 | 19-45157381-C-T | rs10417602 | MARK4, NKPD1 | 0.0777 | T | 5.74E-10 | 0.001368 | ++ | 5.211E-12 | | Old |
| 1198 | 19 | 19-45165069-C-T | rs10410833 | MARK4, TRAPPC6A | 0.0807 | T | 5.64E-10 | 0.001738 | ++ | 7.271E-12 | | Old |
| 1198 | 19 | 19-45170443-G-A | rs28367893 | MARK4, TRAPPC6A | 0.0809 | A | 1.22E-09 | 0.001385 | ++ | 1.107E-11 | 0.3631 | Old |
| 1198 | 19 | 19-45171305-T-A | rs201189859 | MARK4, TRAPPC6A | 0.0934 | T | 1.27E-08 | 0.000715 | ++ | 4.986E-11 | 0.2293 | Old |
| 1198 | 19 | 19-45171922-A-G | rs150685845 | MARK4, TRAPPC6A | 0.0136 | A | 3.76E-07 | 0.030631 | ++ | 4.47E-08 | 0.02406 | Old |
| 1198 | 19 | 19-45176297-C-CA | | MARK4, TRAPPC6A | 0.1008 | CA | 2.59E-06 | 2.01E-06 | ++ | 4.235E-11 | | Old |
| 1198 | 19 | 19-45181810-C-T | rs78979751 | MARK4 | 0.0738 | C | 4.14E-09 | 0.0019 | ++ | 4.216E-11 | | Old |
| 1198 | 19 | 19-45186634-C-T | rs10415850 | MARK4 | 0.1333 | T | 3.71E-09 | 0.078206 | ++ | 7.41E-09 | 0.002292 | Old |
| 1198 | 19 | 19-45186718-T-A | rs59678362 | MARK4 | 0.1318 | T | 9.25E-09 | 0.056335 | ++ | 8.938E-09 | 0.03119 | Old |
| 1198 | 19 | 19-45186719-C-A | rs59839536 | MARK4 | 0.1327 | C | 1.61E-08 | 0.053255 | ++ | 1.292E-08 | 0.002611 | Old |
| 1198 | 19 | 19-45187287-C-T | rs75858218 | MARK4 | 0.0783 | T | 3.35E-09 | 0.010733 | ++ | 2.764E-10 | 0.03969 | Old |
| 1198 | 19 | 19-45187517-G-A | rs12462536 | MARK4 | 0.0923 | G | 2.09E-08 | 0.011909 | ++ | 1.969E-09 | 0.03556 | Old |
| 1198 | 19 | 19-45209858-A-C | rs183110257 | MARK4 | 0.0127 | A | 2.48E-07 | 0.020505 | ++ | 1.965E-08 | 0.03249 | Old |

| | | | | | | | | | | | | |
|---|---|---|---|---|---|---|---|---|---|---|---|---|
| 1199 | 19 | 19-45108380-TC-T | rs1428666075 | MARK4, PPP1R37 | 0.0011 | T | | 1.17E-09 | ?+ | 1.167E-09 | 0.7693 | Old |
| 1200 | 19 | 19-45199680-C-CTTCTCTTCTG | rs200726585 | MARK4 | 0.0171 | C | 1.27E-09 | 0.094875 | ++ | 2.952E-09 | | Old |
| 1201 | 19 | 19-4520591-G-A | rs183251182 | PLIN4, PLIN5 | 0.0979 | G | | 1.55E-08 | ?+ | 1.551E-08 | 0.2554 | New |
| 1201 | 19 | 19-4666569-C-T | rs78455862 | MYDGF | 0.251 | T | | 7.31E-10 | ?+ | 7.307E-10 | 0.09779 | New |
| 1202 | 19 | 19-45223710-G-A | rs112909419 | EXOC3L2, MARK4 | 0.0577 | A | 8.02E-07 | 0.00104 | ++ | 3.608E-09 | 0.00475 | Old |
| 1202 | 19 | 19-45226980-C-T | rs112405270 | EXOC3L2, MARK4 | 0.0557 | T | 6.98E-07 | 0.004155 | ++ | 1.439E-08 | 0.233 | Old |
| 1203 | 19 | 19-45260078-C-CT | | NA | 0.0008 | C | | 3.1E-13 | ?+ | 3.1E-13 | | Old |
| 1203 | 19 | 19-45260080-C-CT | | NA | 0.0008 | C | | 1.57E-10 | ?+ | 1.569E-10 | | Old |
| 1204 | 19 | 19-4528057-C-A | | PLIN5 | 0.0011 | A | 0.43248 | 3.57E-16 | -+ | 9.687E-16 | | New |
| 1205 | 19 | 19-45436408-C-CCT | | NA | 0.0009 | CCT | | 7.8E-09 | ?+ | 7.806E-09 | | Old |
| 1206 | 19 | 19-4562943-GA-G | rs1328984647 | SEMA6B, TNFAIP8L1 | 0.0007 | G | | 4.51E-08 | ?+ | 4.509E-08 | 0.4593 | New |
| 1207 | 19 | 19-46832405-GGC-G | rs1304344790 | AC008622.2, AP2S1 | 0.0061 | G | | 2.68E-11 | ?+ | 2.684E-11 | | Old |
| 1207 | 19 | 19-46832408-C-CAG | rs1395049388 | AC008622.2, AP2S1 | 0.0035 | CAG | | 1.06E-12 | ?+ | 1.06E-12 | | Old |
| 1208 | 19 | 19-46844565-AGG-A | | NA | 0.0011 | AGG | | 7.85E-09 | ?+ | 7.848E-09 | | Old |
| 1208 | 19 | 19-46844567-GCA-G | | NA | 0.0005 | G | | 8.21E-10 | ?+ | 8.207E-10 | | Old |
| 1209 | 19 | 19-47213512-TC-T | rs1454695433 | SAE1, BBC3 | 0.0015 | TC | | 7.6E-10 | ?+ | 7.601E-10 | 0.4644 | New |
| 1210 | 19 | 19-47458598-C-CTCT | | SLC8A2 | 0.003 | C | | 4.3E-08 | ?+ | 4.299E-08 | | New |
| 1211 | 19 | 19-4755334-C-CCT | | NA | 0.001 | C | | 1.25E-14 | ?+ | 1.251E-14 | | New |
| 1212 | 19 | 19-4788589-ACT-A | rs1197488771 | AC005523.1 | 0.0029 | A | | 1.86E-12 | ?+ | 1.86E-12 | | New |
| 1213 | 19 | 19-48564935-TGC-T | rs1291738594 | AC008403.4 | 0.0032 | T | | 3.79E-28 | ?+ | 3.789E-28 | | Old |
| 1214 | 19 | 19-4862940-C-CCT | rs1215624998 | PLIN3 | 0.0015 | CCT | | 4.99E-08 | ?+ | 4.988E-08 | | New |
| 1215 | 19 | 19-48690221-C-T | rs74589403 | NTN5, FUT2 | 0.1207 | T | | 4.33E-13 | ?+ | 4.332E-13 | | Old |

| | | | | | | | | | | | | |
|---|---|---|---|---|---|---|---|---|---|---|---|---|
| 1216 | 19 | 19-48777085-G-C | rs1445575576 | FGF21, RNU6-317P | 0.0006 | C | | 1.92E-10 | ?+ | 1.918E-10 | 0.06526 | Old |
| 1217 | 19 | 19-48916454-C-CG | | NA | 0.0007 | CG | | 1.75E-10 | ?+ | 1.748E-10 | | Old |
| 1218 | 19 | 19-49206127-ATTT-A | rs1252354207 | TRPM4 | 0.0028 | A | | 1.56E-14 | ?+ | 1.557E-14 | 0.6103 | Old |
| 1219 | 19 | 19-49714558-GCA-G | | NA | 0.0007 | G | | 3.54E-11 | ?+ | 3.537E-11 | | Old |
| 1220 | 19 | 19-49905891-C-CCT | | NA | 0.0012 | CCT | | 2.19E-10 | ?+ | 2.192E-10 | | Old |
| 1221 | 19 | 19-5175404-G-A | rs2040076095 | KDM4B, AC022517.1 | 0.0005 | A | | 5.31E-09 | ?+ | 5.306E-09 | | New |
| 1222 | 19 | 19-5192343-G-A | rs2040147005 | AC022517.1, PTPRS | 0.0014 | A | | 3.66E-14 | ?+ | 3.659E-14 | | New |
| 1222 | 19 | 19-5192353-C-A | rs537418221 | AC022517.1, PTPRS | 0.0009 | A | | 5.87E-13 | ?+ | 5.87E-13 | | New |
| 1223 | 19 | 19-52384389-GT-G | rs759082743 | ZNF880 | 0.0045 | G | | 1.82E-09 | ?+ | 1.817E-09 | | New |
| 1224 | 19 | 19-53629086-GGC-G | | NA | 0.0003 | G | | 8.03E-10 | ?+ | 8.031E-10 | | New |
| 1225 | 19 | 19-53947162-G-T | rs1415730747 | CACNG7, CACNG8 | 0.0005 | T | | 3.43E-10 | ?+ | 3.425E-10 | | Old |
| 1226 | 19 | 19-53950357-C-CTG | rs1323565438 | CACNG7, CACNG8 | 0.0012 | C | | 2.15E-10 | ?+ | 2.146E-10 | | Old |
| 1227 | 19 | 19-54059216-AAGCC-A | | NA | 0.0008 | AAGCC | | 2.61E-08 | ?+ | 2.611E-08 | | Old |
| 1228 | 19 | 19-54953825-ATC-A | rs1213132600 | NLRP2, NLRP7 | 0.0018 | ATC | | 6.4E-10 | ?+ | 6.395E-10 | 0.889 | Old |
| 1229 | 19 | 19-55397554-GT-G | rs1190483560 | RPL28 | 0.0006 | G | | 1.99E-09 | ?+ | 1.991E-09 | | New |
| 1230 | 19 | 19-55421214-TTAAGA-T | rs1212560546 | UBE2S, SHISA7 | 0.0006 | T | | 2.41E-08 | ?+ | 2.411E-08 | 0.4801 | New |
| 1231 | 19 | 19-55422561-GGT-G | | NA | 0.0014 | GGT | | 1.06E-19 | ?+ | 1.062E-19 | | New |
| 1231 | 19 | 19-55422571-ATGC-A | | NA | 0.0008 | ATGC | | 6.12E-11 | ?+ | 6.12E-11 | | New |
| 1231 | 19 | 19-55422574-ACGCCTG-A | | NA | 0.0005 | A | | 4.04E-08 | ?+ | 4.036E-08 | | New |
| 1232 | 19 | 19-56055154-ACT-A | | NA | 0.0009 | A | 0.060619 | 5.34E-08 | ++ | 1.132E-08 | | New |

| | | | | | | | | | | | | |
|---|---|---|---|---|---|---|---|---|---|---|---|---|
| 1233 | 19 | 19-56879866-GT-G | rs1323625018 | MIMT1, USP29 | 0.001 | G | | 4.96E-14 | ?+ | 4.956E-14 | | New |
| 1234 | 19 | 19-57064425-C-A | rs1983727065 | MIMT1, USP29 | 1E-04 | C | | 1.08E-09 | ?+ | 1.083E-09 | | New |
| 1235 | 19 | 19-57066412-C-CA | rs1306363346 | MIMT1, USP29 | 0.0022 | C | | 8.12E-14 | ?+ | 8.119E-14 | | New |
| 1236 | 19 | 19-5810053-C-CAGG | rs1157699025 | DUS3L, NRTN | 0.0019 | C | | 4.25E-11 | ?+ | 4.254E-11 | 0.3393 | New |
| 1237 | 19 | 19-58524353-G-A | rs1477026225 | ZBTB45 | 0.0004 | G | | 7.95E-10 | ?+ | 7.952E-10 | 0.8008 | New |
| 1238 | 19 | 19-6104380-C-T | rs1259238984 | RFX2 | 0.0008 | C | | 9.02E-14 | ?+ | 9.015E-14 | | New |
| 1238 | 19 | 19-6104382-G-A | rs1318048165 | RFX2 | 0.0009 | G | | 3.59E-10 | ?+ | 3.591E-10 | | New |
| 1239 | 19 | 19-6308254-G-A | rs1411623251 | ACER1 | 0.0009 | A | 0.368815 | 5.11E-11 | -+ | 2.457E-09 | | New |
| 1240 | 19 | 19-6348245-ACC-A | rs1264751901 | AC011491.3 | 0.0049 | A | | 5.33E-31 | ?+ | 5.334E-31 | 0.2312 | New |
| 1241 | 19 | 19-7236842-C-T | | INSR | 0.0005 | T | 0.097089 | 1.19E-07 | ++ | 3.772E-08 | 0.8298 | New |
| 1242 | 19 | 19-8042058-G-A | rs573469061 | AC008946.1, CCL25 | 0.0015 | G | | 4.06E-08 | ?+ | 4.06E-08 | | New |
| 1243 | 19 | 19-8165431-G-T | rs2083750537 | FBN3, CERS4 | 0.0006 | G | | 4.6E-08 | ?+ | 0.000000046 | | New |
| 1243 | 19 | 19-8165439-C-CTT | | NA | 0.0005 | C | | 8.18E-10 | ?+ | 8.184E-10 | | New |
| 1244 | 19 | 19-8600010-AT-A | | NA | 0.001 | A | | 1.21E-11 | ?+ | 1.21E-11 | | New |
| 1245 | 20 | 20-13349551-G-A | rs537298290 | ISM1, AL121782.1 | 0.0002 | A | 6.58E-07 | 0.00329 | ++ | 7.671E-09 | 0.3169 | New |
| 1246 | 20 | 20-30424031-TTGTGTG-T | rs1209602347 | NA | 0.0081 | TTGTGTG | | 1.19E-20 | ?+ | 1.186E-20 | | New |
| 1247 | 20 | 20-31074716-T-A | rs1194281258 | RNA5SP528, DEFB115 | 0.0058 | T | | 2.51E-08 | ?+ | 2.513E-08 | | New |
| 1248 | 20 | 20-31656375-GGC-G | rs1347052251 | COX4I2, BCL2L1 | 0.0041 | G | | 1E-08 | ?+ | 1.001E-08 | 0.04583 | New |
| 1249 | 20 | 20-31703178-GA-G | | NA | 0.0029 | GA | | 7.04E-11 | ?+ | 7.043E-11 | 0.6649 | New |
| 1250 | 20 | 20-31750080-GGC-G | | TPX2 | 0.003 | G | | 1.38E-16 | ?+ | 1.377E-16 | 0.367 | New |
| 1251 | 20 | 20-32019470-C-CACCTTG | rs1473834293 | AL031658.1 | 0.0057 | C | | 2.65E-08 | ?+ | 2.654E-08 | | New |
| 1252 | 20 | 20-32296095-G-C | rs1472081286 | KIF3B | 0.0006 | G | 0.213378 | 7.61E-10 | -+ | 7.288E-09 | | New |

| | | | | | | | | | | | | |
|---|---|---|---|---|---|---|---|---|---|---|---|---|
| 1253 | 20 | 20-32735144-AG-A | | NA | 0.0003 | A | | 3.59E-10 | ?+ | 3.59E-10 | | New |
| 1254 | 20 | 20-34646551-GCC-G | | NA | 0.0008 | GCC | | 6.01E-14 | ?+ | 6.009E-14 | | New |
| 1254 | 20 | 20-34646555-C-CCT | | NA | 0.0006 | C | | 1.74E-11 | ?+ | 1.74E-11 | | New |
| 1255 | 20 | 20-35097276-C-CAT | | TRPC4AP, EDEM2 | 0.0263 | C | | 1.34E-09 | ?+ | 1.337E-09 | | New |
| 1256 | 20 | 20-36190592-C-A | rs1034010324 | EPB41L1 | 0.0022 | A | 0.69562 | 6.45E-22 | -+ | 1.842E-16 | 0.8273 | New |
| 1256 | 20 | 20-36190632-C-CTTACA | rs1407756628 | EPB41L1 | 0.0015 | CTTACA | | 8.14E-17 | ?+ | 8.143E-17 | | New |
| 1257 | 20 | 20-36728254-C-CCG | | NA | 0.0006 | CCG | | 1.81E-08 | ?+ | 1.813E-08 | | New |
| 1258 | 20 | 20-36787692-C-CCTGGCCAACATGG | | NA | 0.0016 | C | | 2.3E-16 | ?+ | 2.302E-16 | | New |
| 1259 | 20 | 20-36949029-C-CTG | rs1376535353 | SAMHD1 | 0.0052 | CTG | | 2.16E-36 | ?+ | 2.164E-36 | 0.9581 | New |
| 1259 | 20 | 20-36949031-GGC-G | rs1311427302 | SAMHD1 | 0.0046 | GGC | | 2.44E-29 | ?+ | 2.438E-29 | 0.9581 | New |
| 1259 | 20 | 20-36949037-GATGC-G | | SAMHD1 | 0.0017 | G | | 3.74E-15 | ?+ | 3.74E-15 | | New |
| 1259 | 20 | 20-36949046-C-CCT | | SAMHD1 | 0.0016 | C | | 3.96E-16 | ?+ | 3.956E-16 | 0.7978 | New |
| 1260 | 20 | 20-37109452-C-CCT | | MROH8 | 0.0018 | C | | 1.66E-08 | ?+ | 1.659E-08 | 0.2231 | New |
| 1261 | 20 | 20-3850050-G-A | rs138238485 | MAVS | 0.0304 | A | | 6.72E-10 | ?+ | 6.72E-10 | 0.01817 | New |
| 1262 | 20 | 20-3850101-AC-A | rs1257128018 | MAVS | 0.0029 | AC | 0.307591 | 1.8E-28 | -+ | 4.971E-15 | 0.04376 | New |
| 1263 | 20 | 20-38850804-TA-T | | NA | 0.001 | T | | 1.64E-10 | ?+ | 1.637E-10 | | New |
| 1264 | 20 | 20-3978903-C-CA | rs1472697447 | RNF24 | 0.0012 | C | 0.915307 | 3.07E-20 | -+ | 6.556E-17 | | New |
| 1265 | 20 | 20-4081212-AAT-A | rs1411904646 | AL356414.1, SMOX | 0.0018 | A | | 4.02E-08 | ?+ | 4.015E-08 | 0.9146 | New |
| 1266 | 20 | 20-43469358-ATTT-A | | NA | 0.0022 | ATTT | | 1.48E-16 | ?+ | 1.482E-16 | | New |
| 1267 | 20 | 20-43652786-ACC-A | | NA | 0.0008 | ACC | | 5.18E-11 | ?+ | 5.179E-11 | | New |
| 1267 | 20 | 20-43652787-TGA-T | | NA | 0.0005 | T | | 2.66E-09 | ?+ | 2.658E-09 | | New |

| | | | | | | | | | | | | |
|---|---|---|---|---|---|---|---|---|---|---|---|---|
| 1268 | 20 | 20-45931856-ACC-A | | NA | 0.0008 | A | | 4.09E-10 | ?+ | 4.093E-10 | | New |
| 1269 | 20 | 20-47194165-GCA-G | | NA | 0.0011 | G | | 1.15E-08 | ?+ | 1.155E-08 | | New |
| 1270 | 20 | 20-47381012-ACT-A | | ZMYND8, LINC01754 | 0.0011 | ACT | | 2.67E-13 | ?+ | 2.67E-13 | | New |
| 1271 | 20 | 20-48501959-AG-A | rs1445715340 | RNU7-144P, PREX1 | 0.0026 | A | | 2.43E-09 | ?+ | 2.432E-09 | 0.3724 | New |
| 1271 | 20 | 20-48501962-AG-A | rs1306818289 | RNU7-144P, PREX1 | 0.0021 | AG | | 3.17E-11 | ?+ | 3.168E-11 | 0.4386 | New |
| 1272 | 20 | 20-49061328-ATG-A | rs1555821715 | CSE1L | 0.0043 | A | | 4.05E-11 | ?+ | 4.048E-11 | | New |
| 1272 | 20 | 20-49061331-C-CCT | rs1214091512 | CSE1L | 0.0034 | C | | 1.37E-10 | ?+ | 1.367E-10 | 0.2898 | New |
| 1273 | 20 | 20-49198329-C-CA | rs1219080044 | STAU1, DDX27 | 0.0013 | CA | | 4.39E-08 | ?+ | 4.393E-08 | 0.3423 | New |
| 1274 | 20 | 20-49202549-GCA-G | rs1197599157 | STAU1, DDX27 | 0.0005 | G | | 3.42E-08 | ?+ | 3.421E-08 | 0.6938 | New |
| 1275 | 20 | 20-51520826-AT-A | | NA | 0.0003 | AT | | 4.97E-08 | ?+ | 4.974E-08 | | New |
| 1275 | 20 | 20-51520827-T-A | rs2076432782 | NFATC2 | 0.0005 | T | | 7.78E-10 | ?+ | 7.775E-10 | | New |
| 1276 | 20 | 20-53482754-G-A | rs1057509217 | AL109930.1, AL354993.2 | 0.0028 | G | 0.222995 | 4.27E-10 | ++ | 2.4E-09 | | New |
| 1277 | 20 | 20-53698443-G-C | rs1237656557 | RNU7-14P, AC005914.1 | 0.001 | C | | 7.67E-12 | ?+ | 7.672E-12 | | New |
| 1278 | 20 | 20-59401313-G-GCA | | AL160410.1, AL389889.2 | 0.0165 | G | | 8.69E-11 | ?+ | 8.691E-11 | | New |
| 1279 | 20 | 20-62399827-C-T | rs1162057875 | CABLES2 | 0.0012 | T | 0.673279 | 2.28E-10 | -+ | 2.663E-08 | | New |
| 1280 | 20 | 20-62865769-C-CTT | rs2064056060 | TCFL5, DIDO1 | 0.0009 | CTT | | 1.1E-12 | ?+ | 1.104E-12 | | New |
| 1280 | 20 | 20-62865772-G-T | rs2064056109 | TCFL5, DIDO1 | 0.0004 | T | | 1.93E-09 | ?+ | 1.93E-09 | | New |

| | | | | | | | | | | | | |
|---|---|---|---|---|---|---|---|---|---|---|---|---|
| 1281 | 20 | 20-63461661-GGGA-G | rs1409928359 | KCNQ2 | 0.0043 | G | 0.461209 | 3.04E-09 | -+ | 4.141E-09 | | Old |
| 1282 | 21 | 21-10415599-C-T | | AP003900.1, AF254983.1 | 0.0035 | C | | 6.41E-12 | ?- | 6.412E-12 | | New |
| 1282 | 21 | 21-10415600-G-A | | AP003900.1, AF254983.1 | 0.0039 | A | | 6.14E-12 | ?- | 6.143E-12 | | New |
| 1283 | 21 | 21-10717092-A-C | rs28368484 | IGHV1OR21-1, NONE | 0.0044 | A | | 2.07E-09 | ?+ | 2.067E-09 | | New |
| 1284 | 21 | 21-10756490-AG-A | | NA | 0.0031 | A | | 3.52E-14 | ?+ | 3.518E-14 | | New |
| 1285 | 21 | 21-23757294-C-CCT | | NA | 0.0005 | C | | 6.73E-10 | ?+ | 6.734E-10 | | New |
| 1286 | 21 | 21-28685091-AAAC-A | | AF165147.1, RF00026 | 0.0007 | A | | 1.6E-09 | ?+ | 1.603E-09 | | New |
| 1287 | 21 | 21-28995088-GTGA-G | rs1461827110 | LTN1, RWDD2B | 0.0006 | G | | 4.42E-09 | ?+ | 4.417E-09 | 0.4982 | New |
| 1288 | 21 | 21-31690330-C-CCAA | | SCAF4 | 0.0007 | CCAA | | 3.33E-09 | ?+ | 3.335E-09 | | New |
| 1289 | 21 | 21-39324090-AG-A | rs1372913625 | BRWD1-AS1 | 0.0036 | A | | 1.65E-15 | ?+ | 1.655E-15 | 0.896 | New |
| 1290 | 21 | 21-39383050-AAG-A | | NA | 0.0006 | AAG | | 2.17E-10 | ?+ | 2.169E-10 | | New |
| 1290 | 21 | 21-39383052-ACC-A | | NA | 0.0005 | A | | 2.17E-11 | ?+ | 2.167E-11 | | New |
| 1291 | 21 | 21-42718089-C-CG | rs1477539403 | PDE9A | 0.0019 | CG | | 1.56E-16 | ?+ | 1.563E-16 | 0.9578 | New |
| 1291 | 21 | 21-42718092-GA-G | rs1172964571 | PDE9A | 0.0026 | G | | 2.27E-16 | ?+ | 2.266E-16 | 0.5991 | New |
| 1292 | 21 | 21-45155722-TCCACCCAC-T | rs199551441 | ADARB1 | 0.0031 | T | | 1.21E-11 | ?+ | 1.207E-11 | | New |
| 1293 | 21 | 21-45500131-GGGTGGA-G | | NA | 0.0009 | G | | 1.03E-08 | ?+ | 1.032E-08 | | New |
| 1294 | 21 | 21-46085044-C-CT | rs1168284838 | AP001476.2, AP001471.1 | 0.0014 | C | | 2.72E-08 | ?+ | 2.724E-08 | | New |

| | | | | | | | | | | | | |
|---|---|---|---|---|---|---|---|---|---|---|---|---|
| 1295 | 21 | 21-46390054-G-A | rs1039886548 | PCNT | 0.0007 | G | 0.304421 | 5.26E-11 | ++ | 3.221E-11 | | New |
| 1296 | 21 | 21-9124671-AACCCAAAACAATGGGAGTGACGTGCTAAAACCATT-A | | NA | 0.0007 | A | | 4.72E-08 | ?+ | 4.723E-08 | | New |
| 1297 | 21 | 21-9817035-GAA-G | | NA | 0.0048 | G | | 1.95E-10 | ?+ | 1.948E-10 | | New |
| 1298 | 22 | 22-11368837-G-T | rs1174371704 | RF00002, NONE | 0.0069 | G | 0.43741 | 7.03E-10 | -+ | 7.49E-09 | | New |
| 1299 | 22 | 22-12556740-C-CACACACACACACAGAG | | NA | 0.0015 | C | | 1.46E-08 | ?+ | 1.463E-08 | | New |
| 1300 | 22 | 22-16279325-G-C | rs1265426063 | RF00026, AP000547.2 | 0.0002 | C | | 4.66E-08 | ?+ | 4.66E-08 | | New |
| 1301 | 22 | 22-19300549-C-CCT | rs1555998308 | AC000085.1 | 0.002 | C | | 1.55E-14 | ?+ | 1.555E-14 | 0.3646 | New |
| 1301 | 22 | 22-19300566-G-A | rs921890878 | AC000085.1 | 0.0003 | A | 0.392246 | 5.45E-12 | ++ | 1.174E-11 | | New |
| 1302 | 22 | 22-19311896-C-CAAAA | | AC000085.1 | 0.0224 | CAAAA | | 8.99E-09 | ?+ | 8.989E-09 | | New |
| 1303 | 22 | 22-20050079-TGG-T | | NA | 0.0005 | T | | 4.26E-11 | ?+ | 4.259E-11 | | New |
| 1304 | 22 | 22-20503118-ACC-A | | NA | 0.0009 | A | | 1.12E-09 | ?+ | 1.118E-09 | | New |
| 1305 | 22 | 22-21924568-GCGAAC-G | | NA | 0.0005 | GCGAAC | | 7.45E-15 | ?+ | 7.453E-15 | | New |
| 1306 | 22 | 22-22251998-C-T | rs1404067778 | VPREB1, AC245060.6 | 0.0005 | C | | 1.02E-08 | ?+ | 1.02E-08 | | New |
| 1307 | 22 | 22-22518612-AC-A | | NA | 0.0012 | A | | 8.5E-16 | ?+ | 8.5E-16 | | New |
| 1308 | 22 | 22-23515705-G-A | rs150291783 | LINC02557, PCAT14 | 0.0988 | G | | 3.54E-10 | ?+ | 3.541E-10 | 0.8512 | New |
| 1309 | 22 | 22-23646338-ATAAGAAAGATTTT-A | | NA | 0.0005 | ATAAGAAAGATTTT | | 1.7E-08 | ?+ | 1.698E-08 | | New |
| 1310 | 22 | 22-23719208-C-T | rs1269074462 | GUSBP11, ZNF70 | 0.0011 | C | 0.796841 | 5.72E-14 | ++ | 9.068E-09 | | New |
| 1311 | 22 | 22-23797852-C-CAG | | NA | 0.0006 | C | | 2.84E-13 | ?+ | 2.844E-13 | | New |

| | | | | | | | | | | | | |
|---|---|---|---|---|---|---|---|---|---|---|---|---|
| 1312 | 22 | 22-24490949-C-CAA | | NA | 0.0007 | C | | 6.89E-12 | ?+ | 6.889E-12 | | New |
| 1313 | 22 | 22-24777872-ATT-A | | NA | 0.0017 | ATT | | 1.08E-13 | ?+ | 1.083E-13 | | New |
| 1314 | 22 | 22-25252791-C-CCA | rs1177181245 | Z99916.3 | 0.0042 | CCA | | 1.49E-31 | ?+ | 1.488E-31 | 0.6766 | New |
| 1314 | 22 | 22-25252792-AGAGGTG-A | rs1372844864 | Z99916.3 | 0.0035 | A | | 3.09E-28 | ?+ | 3.092E-28 | 0.7925 | New |
| 1314 | 22 | 22-25252802-AAGCG-A | rs1299512285 | Z99916.3 | 0.0033 | AAGCG | | 7.61E-23 | ?+ | 7.61E-23 | | New |
| 1315 | 22 | 22-25324817-G-T | rs1203341863 | AL022324.4, LRP5L | 0.0008 | T | | 1.65E-09 | ?+ | 1.655E-09 | | New |
| 1316 | 22 | 22-25345441-AGGGGTGAGCCCCTGCTCTCAGCCTCCCACAGTGCTGGGATTACT-A | | AL022324.4, LRP5L | 0.0077 | A | | 5.07E-14 | ?+ | 5.067E-14 | | New |
| 1317 | 22 | 22-25789433-C-T | | MYO18B | 0.0005 | T | 0.064919 | 9.91E-08 | ++ | 1.81E-08 | 0.3467 | New |
| 1318 | 22 | 22-28089534-GAA-G | | NA | 0.001 | GAA | | 2.23E-08 | ?+ | 2.228E-08 | 0.9003 | New |
| 1319 | 22 | 22-28536011-C-CCATCCTGGCTAA | rs1339933971 | TTC28 | 0.0037 | CCATCCTGGCTAA | | 3.58E-10 | ?+ | 3.579E-10 | | New |
| 1320 | 22 | 22-28645472-C-CA | rs1312893620 | TTC28 | 0.0024 | CA | | 1.71E-11 | ?+ | 1.71E-11 | 0.1333 | New |
| 1321 | 22 | 22-28770546-AT-A | rs1469740920 | HSCB, CCDC117 | 0.0018 | AT | | 5.78E-11 | ?+ | 5.776E-11 | 0.05477 | New |
| 1322 | 22 | 22-29148578-C-CG | | NA | 0.0006 | C | | 4.85E-12 | ?+ | 4.852E-12 | | New |
| 1323 | 22 | 22-30913268-C-CG | rs1312458002 | OSBP2, MORC2-AS1 | 0.0038 | C | | 1.26E-18 | ?+ | 1.258E-18 | | New |
| 1324 | 22 | 22-31293758-GT-G | rs1308349799 | PIK3IP1-AS1 | 0.0006 | GT | | 3.08E-08 | ?+ | 3.075E-08 | | New |
| 1325 | 22 | 22-35050426-C-A | rs1162255140 | AL024495.1, ISX | 0.0002 | A | | 2.49E-09 | ?+ | 2.493E-09 | | New |
| 1326 | 22 | 22-35699725-GCCCC-G | | NA | 0.0045 | G | | 2.3E-10 | ?+ | 2.304E-10 | | New |
| 1327 | 22 | 22-36068076-G-A | rs1317768370 | Z95114.4 | 0.0006 | A | 0.160306 | 3.04E-10 | ++ | 5.489E-09 | 0.151 | New |
| 1327 | 22 | 22-36068090-G-A | rs557990794 | Z95114.4 | 0.0009 | A | 0.737581 | 1.47E-16 | ++ | 2.825E-14 | 0.1699 | New |

| | | | | | | | | | | | | |
|---|---|---|---|---|---|---|---|---|---|---|---|---|
| 1328 | 22 | 22-36389311-C-CCT | | NA | 0.0006 | C | | 1.5E-08 | ?+ | 1.503E-08 | | New |
| 1329 | 22 | 22-37939702-T-A | | MICALL1 | 0.0005 | A | | 9.1E-10 | ?+ | 9.096E-10 | | New |
| 1330 | 22 | 22-38242246-TTG-T | | NA | 0.0003 | T | | 1.39E-08 | ?+ | 1.394E-08 | | New |
| 1331 | 22 | 22-38342004-C-CCA | | NA | 0.0012 | CCA | | 5.16E-12 | ?+ | 5.159E-12 | | New |
| 1332 | 22 | 22-38405984-AT-A | | NA | 0.0012 | AT | | 1.33E-09 | ?+ | 1.328E-09 | 0.1544 | New |
| 1333 | 22 | 22-38654128-G-C | rs942934493 | FAM227A | 0.0026 | C | 0.946907 | 4.83E-28 | ++ | 3.783E-14 | | New |
| 1334 | 22 | 22-39166463-C-CCT | | NA | 0.0008 | CCT | | 1.69E-09 | ?+ | 1.688E-09 | | New |
| 1335 | 22 | 22-39166662-AG-A | | NA | 0.0006 | AG | | 4.37E-08 | ?+ | 4.375E-08 | | New |
| 1336 | 22 | 22-40796022-C-CAG | | NA | 0.0013 | C | 0.493337 | 1.83E-13 | -+ | 3.587E-12 | | New |
| 1337 | 22 | 22-40935856-GT-G | | NA | 0.001 | G | | 1.89E-09 | ?+ | 1.89E-09 | | New |
| 1338 | 22 | 22-41293017-C-CG | rs1186350373 | RANGAP1, ZC3H7B | 0.0013 | C | | 4.5E-09 | ?+ | 4.497E-09 | 0.2797 | New |
| 1339 | 22 | 22-41379102-ATGCC-A | | TEF | 0.0027 | ATGCC | | 1.98E-10 | ?+ | 1.98E-10 | 0.5427 | New |
| 1340 | 22 | 22-41609575-C-CTTG | | NA | 0.0005 | C | | 3.18E-08 | ?+ | 3.176E-08 | | New |
| 1341 | 22 | 22-41786143-C-T | rs2075977616 | MEI1 | 0.0003 | T | | 8.5E-10 | ?+ | 8.5E-10 | | New |
| 1341 | 22 | 22-41786153-G-T | rs1364149896 | MEI1 | 0.0006 | T | 0.937115 | 1.62E-10 | -+ | 6.88E-10 | | New |
| 1342 | 22 | 22-43016634-C-T | rs7290470 | PACSIN2, AL022476.1 | 0.0987 | T | | 3.58E-08 | ?+ | 3.579E-08 | 0.02898 | New |
| 1343 | 22 | 22-44749165-AC-A | rs1388705722 | PRR5-ARHGAP8 | 0.0015 | AC | | 5.55E-09 | ?+ | 5.552E-09 | 0.04027 | New |
| 1344 | 22 | 22-44886986-C-CCA | | PHF21B | 0.0007 | CCA | | 2.32E-09 | ?+ | 2.319E-09 | | New |
| 1345 | 22 | 22-46713286-C-CTT | | NA | 0.0004 | CTT | | 1.15E-11 | ?+ | 1.151E-11 | | New |
| 1346 | 22 | 22-48399528-C-T | rs113717364 | MIR3201, Z72006.1 | 0.0832 | C | | 2.07E-13 | ?+ | 2.069E-13 | 0.5675 | New |

| | | | | | | | | | | | |
|---|---|---|---|---|---|---|---|---|---|---|---|
| 1347 | 22 | 22-48910553-C-CCCCTT | rs1169595633 | LINC01310, AL078622.1 | 0.0007 | C | 3.77E-09 | ?+ | 3.774E-09 | | New |
| 1348 | 22 | 22-49399428-C-CCA | | NA | 0.0002 | C | 3.98E-10 | ?+ | 3.978E-10 | | New |
| 1349 | 22 | 22-49879291-AT-A | | NA | 0.0005 | A | 4.9E-10 | ?+ | 4.905E-10 | | New |
| 1350 | 22 | 22-50354109-ACC-A | rs1555964346 | PPP6R2 | 0.0019 | A | 2.87E-11 | ?+ | 2.872E-11 | | New |
| 1350 | 22 | 22-50354112-GGC-G | rs1240189301 | PPP6R2 | 0.0022 | GGC | 6.48E-11 | ?+ | 6.482E-11 | 0.02318 | New |
| 1351 | 22 | 22-50372984-GT-G | | NA | 0.0006 | G | 2.7E-10 | ?+ | 2.697E-10 | | New |
| 1352 | 22 | 22-50543535-TGCA-T | rs1356120401 | U62317.1; U62317.4 | 0.0036 | TGCA | 3.16E-08 | ?+ | 3.162E-08 | 0.01972 | New |
| 1352 | 22 | 22-50543540-C-CGGT | rs1483504988 | U62317.1; U62317.4 | 0.0048 | C | 1.28E-14 | ?+ | 1.284E-14 | | New |
