## Supplementary Table 5 for "Identification of 16 novel Alzheimer’s disease susceptibility loci using multi-ancestry meta-analyses of clinical Alzheimer’s disease and AD-by-proxy cases from four whole genome sequencing datasets"

**Supplementary table 5:** Independent loci that passed QC in our UKB GWAS, including variants that were not nominally significant in our clinical AD meta-analysis.
MAF - minor allele frequency; P - p-value; META_P - p-value for the meta analysis.

| Locus | Chromosome | Variant ID | Rsid | Gene | MAF | Effect allele | UKB_P | AOU_P | Direction (UKB, AOU) | UKB_AOU_META_P | NIA_NIMH_META_P | New or Old |
|---|---|---|---|---|---|---|---|---|---|---|---|---|
| 1 | 2 | 2-127128582-C-T | rs13025717 | BIN1, AC110926.2 | 0.2947 | T | 4.4E-08 | 0.024032 | ++ | 7.457E-09 | 0.00001185 | Old |
| 1 | 2 | 2-127128657-C-T | rs13025765 | BIN1, AC110926.2 | 0.2942 | T | 4.17E-08 | 0.019492 | ++ | 5.418E-09 | 0.00001229 | Old |
| 1 | 2 | 2-127133851-C-A | rs4663105 | BIN1, AC110926.2 | 0.4334 | C | 2.51E-10 | 0.01738 | ++ | 6.348E-11 | 8.114E-11 | Old |
| 1 | 2 | 2-127135234-C-T | rs6733839 | BIN1, AC110926.2 | 0.3923 | T | 4.4E-12 | 0.033562 | ++ | 6.137E-12 | 4.673E-11 | Old |
| 2 | 2 | 2-30994688-C-T | rs763350901 | GALNT14 | 0.0001 | T | 3.08E-08 | 0.429695 | ++ | 7.316E-08 | | New |
| 3 | 4 | 4-100831815-C-T | rs148360344 | LINC01218, PPP3CA | 1E-04 | C | 3.63E-08 | 0.073622 | ++ | 4.984E-07 | | New |
| 3 | 4 | 4-100836007-C-T | rs77324998 | LINC01218, PPP3CA | 0.0001 | T | 3.63E-08 | 0.069019 | ++ | 4.284E-07 | | New |
| 4 | 6 | 6-156801300-G-A | rs138681527 | ARID1B | 0.0223 | G | 2.82E-08 | 0.959189 | ++ | 0.000003129 | 0.1385 | New |
| 5 | 6 | 6-30429738-AAAGAGCCACATGAAGAGAT-A | rs368223591 | RPP21, AL662873.1 | 0.0097 | A | 4.04E-09 | 0.594396 | -- | 1.943E-07 | 0.6199 | New |
| 5 | 6 | 6-30495924-C-T | rs138246546 | HLA-E, LINC02569 | 0.0098 | C | 4.32E-09 | 0.527721 | -- | 1.505E-07 | 0.6708 | New |
| 5 | 6 | 6-30556837-G-A | rs41272611 | GNL1 | 0.0098 | G | 5.24E-09 | 0.516941 | -- | 1.656E-07 | 0.7298 | New |
| 5 | 6 | 6-30610380-G-T | rs187886061 | PPP1R10 | 0.0097 | T | 5.6E-09 | 0.408124 | -- | 9.977E-08 | 0.5464 | New |
| 6 | 6 | 6-30693413-G-A | rs192046402 | NRM, MDC1 | 0.0101 | A | 9.04E-09 | 0.564594 | -- | 2.999E-07 | 0.4755 | New |
| 6 | 6 | 6-30697582-G-T | rs115906270 | NRM, MDC1 | 0.01 | T | 6.13E-09 | 0.577543 | -- | 2.399E-07 | 0.9783 | New |
| 7 | 6 | 6-32381995-C-T | rs9268455 | TSBP1-AS1 | 0.2103 | T | 1.4E-08 | 0.017077 | -- | 1.731E-09 | 0.3865 | Old |
| 7 | 6 | 6-32404138-C-T | rs3793127 | TSBP1-AS1 | 0.2104 | T | 1.52E-08 | 0.01615 | -- | 1.731E-09 | 0.3749 | Old |
| 7 | 6 | 6-32413666-T-A | rs9268522 | TSBP1-AS1, HLA-DRA | 0.2176 | T | 2.82E-08 | 0.024758 | -- | 5.275E-09 | 0.2559 | Old |
| 7 | 6 | 6-32438044-C-T | rs9268627 | TSBP1-AS1, HLA-DRA | 0.2136 | C | 4E-08 | 0.024994 | -- | 7.009E-09 | 0.7249 | Old |
| 7 | 6 | 6-32461376-G-A | rs9268844 | HLA-DRA, HLA-DRB5 | 0.2038 | G | 4.46E-09 | 0.01651 | -- | 5.871E-10 | | Old |
| 7 | 6 | 6-32602534-G-T | rs34039593 | HLA-DRB1, HLA-DQA1 | 0.1746 | G | 4.33E-09 | 0.073629 | -- | 4.341E-09 | 0.9873 | Old |
| 7 | 6 | 6-32602640-C-A | rs2647062 | HLA-DRB1, HLA-DQA1 | 0.1746 | C | 3.72E-09 | 0.072099 | -- | 3.706E-09 | 0.9785 | Old |
| 7 | 6 | 6-32603181-G-T | rs679242 | HLA-DRB1, HLA-DQA1 | 0.1745 | T | 2E-09 | 0.070655 | -- | 2.118E-09 | 0.9081 | Old |
| 7 | 6 | 6-32603333-G-A | rs2760990 | HLA-DRB1, HLA-DQA1 | 0.175 | A | 2.13E-09 | 0.070943 | -- | 2.285E-09 | 0.9908 | Old |
| 7 | 6 | 6-32605402-G-A | rs601020 | HLA-DRB1, HLA-DQA1 | 0.2378 | A | 3.2E-08 | 0.085603 | -- | 3.416E-08 | 0.2036 | Old |
| 7 | 6 | 6-32605488-G-T | rs601148 | HLA-DRB1, HLA-DQA1 | 0.1863 | T | 2.73E-08 | 0.059196 | -- | 1.574E-08 | 0.3496 | Old |
| 7 | 6 | 6-32605638-G-A | rs601945 | HLA-DRB1, HLA-DQA1 | 0.1717 | G | 4.58E-09 | 0.047919 | -- | 2.277E-09 | 0.9857 | Old |
| 7 | 6 | 6-32606826-G-A | rs617578 | HLA-DRB1, HLA-DQA1 | 0.1767 | A | 2.61E-09 | 0.082916 | -- | 3.545E-09 | 0.8026 | Old |
| 7 | 6 | 6-32607091-C-T | rs7760841 | HLA-DRB1, HLA-DQA1 | 0.1787 | T | 7.31E-09 | 0.062939 | -- | 5.661E-09 | 0.9174 | Old |
| 7 | 6 | 6-32607119-G-A | rs7770010 | HLA-DRB1, HLA-DQA1 | 0.178 | A | 1.1E-08 | 0.062299 | -- | 7.834E-09 | 0.9071 | Old |
| 7 | 6 | 6-32609269-C-T | rs562289 | HLA-DRB1, HLA-DQA1 | 0.22 | T | 7.54E-09 | 0.057127 | -- | 5.861E-09 | | Old |
| 7 | 6 | 6-32610196-G-T | rs532965 | HLA-DRB1, HLA-DQA1 | 0.179 | G | 5.39E-09 | 0.062384 | -- | 4.327E-09 | 0.892 | Old |
| 7 | 6 | 6-32610995-C-A | rs504594 | HLA-DRB1, HLA-DQA1 | 0.1758 | A | 7.87E-09 | 0.042427 | -- | 3.231E-09 | 0.9585 | Old |
| 7 | 6 | 6-32612814-G-C | rs7449585 | HLA-DRB1, HLA-DQA1 | 0.2611 | C | 3.66E-08 | 0.02221 | -- | 6.112E-09 | | Old |
| 7 | 6 | 6-32612840-T-A | rs3997872 | HLA-DRB1, HLA-DQA1 | 0.1759 | A | 8.7E-09 | 0.045107 | -- | 3.838E-09 | 0.9916 | Old |
| 7 | 6 | 6-32612880-C-T | rs2395516 | HLA-DRB1, HLA-DQA1 | 0.306 | C | 1.42E-08 | 0.019816 | -- | 2.463E-09 | 0.6616 | Old |
| 7 | 6 | 6-32613231-C-T | rs3129747 | HLA-DRB1, HLA-DQA1 | 0.3061 | C | 1.36E-08 | 0.020371 | -- | 2.475E-09 | 0.6771 | Old |
| 7 | 6 | 6-32613238-T-A | rs3129748 | HLA-DRB1, HLA-DQA1 | 0.3061 | A | 1.36E-08 | 0.020571 | -- | 2.507E-09 | 0.6724 | Old |
| 7 | 6 | 6-32614412-C-A | rs3129751 | HLA-DRB1, HLA-DQA1 | 0.1759 | C | 7.98E-09 | 0.042248 | -- | 3.252E-09 | 0.9945 | Old |

| | | | | | | | | | | | | |
|---|---|---|---|---|---|---|---|---|---|---|---|---|
| 7 | 6 | 6-32614873-G-C | rs3104413 | HLA-DRB1, HLA-DQA1 | 0.1782 | G | 7.05E-09 | 0.063393 | -- | 5.794E-09 | 0.889 | Old |
| 7 | 6 | 6-32615250-G-C | rs3129753 | HLA-DRB1, HLA-DQA1 | 0.1791 | C | 4.01E-09 | 0.066203 | -- | 3.715E-09 | 0.902 | Old |
| 7 | 6 | 6-32615369-C-T | rs4959105 | HLA-DRB1, HLA-DQA1 | 0.3179 | T | 5.18E-09 | 0.011798 | -- | 5.425E-10 | 0.8795 | Old |
| 7 | 6 | 6-32615429-G-A | rs6941395 | HLA-DRB1, HLA-DQA1 | 0.1791 | A | 5.09E-09 | 0.063042 | -- | 4.201E-09 | 0.9181 | Old |
| 7 | 6 | 6-32616142-C-T | rs33964890 | HLA-DRB1, HLA-DQA1 | 0.2737 | C | 1.48E-08 | 0.01524 | -- | 1.805E-09 | 0.6769 | Old |
| 7 | 6 | 6-32616202-G-A | rs33915496 | HLA-DRB1, HLA-DQA1 | 0.318 | G | 6.01E-09 | 0.012006 | -- | 6.305E-10 | 0.8365 | Old |
| 7 | 6 | 6-32616536-G-C | rs33932178 | HLA-DRB1, HLA-DQA1 | 0.319 | G | 7.01E-09 | 0.018483 | -- | 1.22E-09 | | Old |
| 7 | 6 | 6-32617117-G-T | rs508318 | HLA-DRB1, HLA-DQA1 | 0.1785 | T | 5.27E-09 | 0.062985 | -- | 4.304E-09 | | Old |
| 7 | 6 | 6-32622864-G-T | rs9271580 | HLA-DRB1, HLA-DQA1 | 0.1909 | G | 2.07E-08 | 0.052096 | -- | 1.02E-08 | | Old |
| 7 | 6 | 6-32623007-TACAG-T | rs5875381 | HLA-DRB1, HLA-DQA1 | 0.1952 | TACAG | 1.77E-08 | 0.081176 | -- | 1.613E-08 | 0.4352 | Old |
| 7 | 6 | 6-32623436-G-A | rs9271594 | HLA-DRB1, HLA-DQA1 | 0.1806 | G | 4.48E-09 | 0.061802 | -- | 3.597E-09 | 0.914 | Old |
| 7 | 6 | 6-32629245-G-A | rs3129769 | HLA-DQA1 | 0.1788 | A | 7.51E-09 | 0.059126 | -- | 5.281E-09 | 0.9376 | Old |
| 7 | 6 | 6-32631077-G-A | rs3104381 | HLA-DQA1 | 0.1822 | A | 1.97E-08 | 0.058436 | -- | 1.104E-08 | 0.8642 | Old |
| 7 | 6 | 6-32631386-C-T | rs3104378 | HLA-DQA1 | 0.1794 | T | 6.54E-09 | 0.058307 | -- | 4.625E-09 | 0.9395 | Old |
| 7 | 6 | 6-32634360-G-C | rs3104371 | HLA-DQA1 | 0.1917 | G | 1.86E-08 | 0.048707 | -- | 8.514E-09 | 0.5671 | Old |
| 7 | 6 | 6-32634784-C-T | rs3104368 | HLA-DQA1 | 0.1788 | T | 8.56E-09 | 0.058081 | -- | 5.735E-09 | | Old |
| 7 | 6 | 6-32636679-G-C | rs9272353 | HLA-DQA1 | 0.1921 | C | 1.85E-08 | 0.049281 | -- | 8.559E-09 | 0.6229 | Old |
| 7 | 6 | 6-32636808-T-A | rs9272363 | HLA-DQA1 | 0.1952 | T | 1.61E-08 | 0.036747 | -- | 4.715E-09 | 0.4868 | Old |
| 8 | 7 | 7-100276574-T-A | rs34857299 | CASTOR3, SPDYE3 | 0.2209 | A | 2.56E-08 | 0.790793 | -+ | 0.000006518 | 0.0007087 | Old |
| 9 | 10 | 10-80678507-T-C | rs4406769 | LINC02655, AL096706.1 | 0.0999 | C | 4.04E-08 | 0.49388 | -+ | 0.00004779 | 0.1162 | Old |
| 10 | 12 | 12-102888270-C-T | rs968665225 | PAH | 0.0002 | T | 1E-83 | 0.07485 | -- | 0.1039 | 0.243 | New |
| 11 | 19 | 19-44673285-G-A | rs142034848 | AC243964.2 | 0.0136 | G | 5.78E-10 | 0.114836 | ++ | 8.579E-10 | 0.0002124 | Old |
| 11 | 19 | 19-44733473-C-T | rs2927434 | RF00285, BCL3 | 0.1809 | T | 4.41E-14 | 0.015193 | ++ | 2.538E-14 | 0.000134 | Old |
| 11 | 19 | 19-44736813-C-CT | rs58455045 | RF00285, BCL3 | 0.1927 | CT | 3.1E-15 | 0.008123 | ++ | 1.086E-15 | | Old |
| 11 | 19 | 19-44738381-G-A | rs2927437 | RF00285, BCL3 | 0.1784 | A | 6.85E-17 | 0.004637 | ++ | 1.915E-17 | 0.000004448 | Old |
| 11 | 19 | 19-44740137-TATACACACACACAC AC-T | rs34562807 | RF00285, BCL3 | 0.0541 | TATACACA | 2.47E-21 | 0.004364 | ++ | 4.017E-21 | | Old |
| 11 | 19 | 19-44781370-G-A | rs80168591 | CBLC | 0.012 | A | 1.94E-13 | 0.004804 | ++ | 8.087E-15 | 2.343E-10 | Old |
| 11 | 19 | 19-44799020-C-T | rs139997344 | CBLC | 0.0074 | T | 1.16E-20 | 0.189998 | ++ | 1.352E-18 | 7.851E-08 | Old |
| 11 | 19 | 19-44812964-G-A | rs528070791 | BCAM | 0.0126 | G | 4.84E-24 | 0.070312 | ++ | 3.541E-22 | | Old |
| 11 | 19 | 19-44819913-CCATCCCCAACTCAT CCT-C | rs3842409 | BCAM | 0.4442 | C | 4.29E-22 | 0.014798 | -- | 1.2E-20 | | Old |
| 11 | 19 | 19-44820881-G-A | rs28399637 | BCAM | 0.2824 | A | 1.07E-33 | 0.287699 | ++ | 1.305E-26 | 4.149E-20 | Old |
| 11 | 19 | 19-44821499-G-C | rs28399664 | BCAM | 0.0185 | G | 2.51E-44 | 0.002044 | ++ | 2.879E-42 | 0.0684 | Old |
| 11 | 19 | 19-44827852-G-C | rs905342119 | BCAM, NECTIN2 | 0.0082 | C | 4.91E-22 | 0.141143 | ++ | 3.893E-20 | 5.485E-10 | Old |
| 11 | 19 | 19-44834661-G-A | rs147711004 | BCAM, NECTIN2 | 0.0311 | A | 4.52E-63 | 0.000683 | ++ | 3.393E-59 | 0.2333 | Old |
| 11 | 19 | 19-44843409-G-C | rs148601586 | BCAM, NECTIN2 | 0.0122 | G | 5.77E-19 | 0.030667 | ++ | 8.133E-19 | 3.717E-13 | Old |
| 11 | 19 | 19-44848259-G-C | rs41289512 | NECTIN2 | 0.0362 | G | 9.43E-65 | 0.002147 | ++ | 6.644E-59 | 3.555E-30 | Old |
| 11 | 19 | 19-44850787-G-T | rs57537848 | NECTIN2 | 0.4852 | G | 5.34E-10 | 0.351864 | -- | 1.929E-08 | 0.06697 | Old |
| 11 | 19 | 19-44851039-G-A | rs11666329 | NECTIN2 | 0.4853 | G | 5.39E-10 | 0.331867 | -- | 1.699E-08 | 9.115E-10 | Old |
| 11 | 19 | 19-44852031-C-T | rs149661872 | NECTIN2 | 0.0091 | T | 1.66E-17 | 0.333973 | ++ | 1.897E-15 | 1.42E-09 | Old |
| 11 | 19 | 19-44852378-C-A | rs557293140 | NECTIN2 | 0.0037 | C | 4.37E-09 | 0.630737 | +- | 0.00000153 | 1.506E-07 | Old |

| | | | | | | | | | | | | |
|---|---|---|---|---|---|---|---|---|---|---|---|---|
| 11 | 19 | 19-44852486-C-A | rs4802241 | NECTIN2 | 0.1772 | C | 1.05E-08 | 0.091571 | -- | 1.531E-08 | 0.0001444 | Old |
| 11 | 19 | 19-44855049-C-T | rs551048812 | NECTIN2 | 0.0084 | T | 3.3E-10 | 0.007583 | ++ | 1.752E-11 | 1.146E-10 | Old |
| 11 | 19 | 19-44856329-C-T | rs56317818 | NECTIN2 | 0.2603 | T | 2.28E-31 | 0.008669 | ++ | 2.994E-28 | 2.368E-11 | Old |
| 11 | 19 | 19-44856688-G-A | rs183610051 | NECTIN2 | 0.0097 | A | 3.52E-14 | 0.011575 | ++ | 3.512E-15 | 3.659E-11 | Old |
| 11 | 19 | 19-44858325-C-CT | | NECTIN2 | 0.1712 | CT | 9.74E-11 | 0.067805 | -- | 2.042E-10 | | Old |
| 11 | 19 | 19-44860563-G-T | rs138607350 | NECTIN2 | 0.0081 | G | 8.21E-24 | 0.01687 | ++ | 1.579E-23 | 3.11E-10 | Old |
| 11 | 19 | 19-44861785-G-A | rs538568659 | NECTIN2 | 0.0042 | G | 1.25E-11 | 0.085424 | ++ | 1.433E-11 | 0.00001178 | Old |
| 11 | 19 | 19-44862190-G-A | rs146275714 | NECTIN2 | 0.0187 | A | 3.97E-50 | 0.000294 | ++ | 1.034E-48 | 5.811E-21 | Old |
| 11 | 19 | 19-44863241-G-A | rs183427010 | NECTIN2 | 0.0055 | A | 3.63E-12 | 0.532183 | ++ | 2.946E-10 | 8.641E-10 | Old |
| 11 | 19 | 19-44864245-G-C | rs73050216 | NECTIN2 | 0.1703 | C | 2.71E-10 | 0.033944 | -- | 1.546E-10 | 0.000001784 | Old |
| 11 | 19 | 19-44866307-AG-A | rs34111552 | NECTIN2 | 0.0065 | A | 1.82E-11 | 0.401822 | ++ | 3.432E-10 | 7.399E-08 | Old |
| 11 | 19 | 19-44867581-G-A | rs12610605 | NECTIN2 | 0.1744 | A | 2.74E-10 | 0.023362 | -- | 8.964E-11 | 0.000001111 | Old |
| 11 | 19 | 19-44868428-AT-A | rs34165484 | NECTIN2 | 0.1889 | AT | 5.86E-11 | 0.043718 | -- | 7.762E-11 | 5.583E-07 | Old |
| 11 | 19 | 19-44873636-AAC-A | rs34606745 | NECTIN2 | 0.0866 | AAC | 1.46E-65 | 0.008022 | ++ | 3.427E-54 | | Old |
| 11 | 19 | 19-44875268-C-CT | rs57907894 | NECTIN2 | 0.1975 | CT | 9.05E-10 | 0.030745 | -- | 4.378E-10 | | Old |
| 11 | 19 | 19-44877713-T-G | | NECTIN2 | 0.4745 | G | 5.12E-17 | 0.009763 | -- | 7.49E-17 | 7.157E-15 | Old |
| 11 | 19 | 19-44879460-A-G | rs406456 | NECTIN2 | 0.3735 | A | 1.28E-28 | 0.000157 | ++ | 2.292E-29 | 1.714E-07 | Old |
| 11 | 19 | 19-44880774-AT-A | | NECTIN2 | 0.0134 | A | 8.08E-14 | 0.052561 | ++ | 2.853E-13 | | Old |
| 11 | 19 | 19-44881674-G-A | rs79701229 | NECTIN2 | 0.0116 | A | 4.5E-20 | 0.042705 | ++ | 2.464E-19 | 8.818E-13 | Old |
| 11 | 19 | 19-44882099-C-A | rs144261139 | AC011481.2 | 0.0108 | A | 3.1E-28 | 0.028966 | ++ | 1.681E-26 | 0.1683 | Old |
| 11 | 19 | 19-44883210-GTAA-G | rs142042446 | AC011481.2 | 0.1325 | GTAA | 4.2E-210 | 2.42E-15 | ++ | 9.09E-202 | 1.61E-136 | Old |
| 11 | 19 | 19-44883377-C-T | rs147636938 | AC011481.2 | 0.0242 | T | 3.7E-52 | 0.00027 | ++ | 5.353E-49 | 1.348E-32 | Old |
| 11 | 19 | 19-44884202-G-C | rs12972156 | AC011481.2 | 0.1323 | G | 3.2E-210 | 2.6E-15 | ++ | 7.7E-202 | 4.86E-140 | Old |
| 11 | 19 | 19-44884339-G-A | rs12972970 | AC011481.2 | 0.1324 | A | 1.1E-209 | 1.06E-15 | ++ | 3.3E-202 | 2.69E-140 | Old |
| 11 | 19 | 19-44884873-G-A | rs34342646 | AC011481.2 | 0.136 | A | 1.8E-203 | 3E-14 | ++ | 7.26E-194 | 4.9E-135 | Old |
| 11 | 19 | 19-44885243-G-A | rs283811 | AC011481.2 | 0.2366 | G | 8.1E-195 | 6.03E-11 | ++ | 3.12E-174 | 7.23E-120 | Old |
| 11 | 19 | 19-44887076-A-G | rs283815 | AC011481.2 | 0.2408 | A | 1.2E-203 | 1.55E-12 | ++ | 5.14E-184 | 3.05E-11 | Old |
| 11 | 19 | 19-44888997-C-T | rs6857 | AC011481.2 | 0.1543 | T | 1.8E-287 | 1.25E-21 | ++ | 2.42E-277 | 2.33E-189 | Old |
| 11 | 19 | 19-44890259-C-T | rs117310449 | AC011481.2 | 0.0104 | T | 1.79E-29 | 0.028844 | ++ | 1.399E-27 | 3.589E-18 | Old |
| 11 | 19 | 19-44891079-C-T | rs71352238 | TOMM40 | 0.1326 | C | 3.3E-214 | 1.97E-16 | ++ | 3.36E-207 | 4.62E-143 | Old |
| 11 | 19 | 19-44891712-G-T | rs184017 | TOMM40 | 0.2369 | G | 5.1E-203 | 1.11E-12 | ++ | 6.68E-184 | 1.44E-124 | Old |
| 11 | 19 | 19-44892009-A-G | rs157580 | TOMM40 | 0.3709 | A | 8.95E-64 | 8.51E-06 | -- | 1.931E-60 | | Old |
| 11 | 19 | 19-44892362-G-A | rs2075650 | TOMM40 | 0.1386 | G | 8E-222 | 8.91E-18 | ++ | 8.01E-214 | 7.79E-130 | Old |
| 11 | 19 | 19-44892457-C-T | rs157581 | TOMM40 | 0.2402 | C | 1.4E-207 | 7.23E-13 | ++ | 8.82E-188 | | Old |
| 11 | 19 | 19-44892587-G-A | rs34095326 | TOMM40 | 0.0979 | A | 7.5E-165 | 7.83E-12 | ++ | 7.73E-158 | 2.391E-85 | Old |
| 11 | 19 | 19-44892652-G-C | rs34404554 | TOMM40 | 0.1359 | G | 4.2E-224 | 2.89E-18 | ++ | 2.72E-217 | 5.35E-135 | Old |
| 11 | 19 | 19-44892887-C-T | rs11556505 | TOMM40 | 0.137 | T | 1.3E-222 | 4.98E-18 | ++ | 2.63E-215 | 1.93E-133 | Old |
| 11 | 19 | 19-44892962-C-T | rs157582 | TOMM40 | 0.2385 | T | 6.4E-208 | 3.2E-13 | ++ | 5.76E-189 | 2.46E-135 | Old |
| 11 | 19 | 19-44893408-G-T | rs59007384 | TOMM40 | 0.2146 | T | 1E-211 | 1.83E-11 | ++ | 1.14E-188 | 4.58E-11 | Old |
| 11 | 19 | 19-44893716-G-A | rs77301115 | TOMM40 | 0.0265 | A | 6.28E-53 | 9.7E-05 | ++ | 3.72E-50 | 5.579E-54 | Old |
| 11 | 19 | 19-44894050-C-T | rs112849259 | TOMM40 | 0.0264 | T | 2.09E-53 | 0.000145 | ++ | 3.279E-50 | 5.136E-55 | Old |
| 11 | 19 | 19-44894261-C-CTT | rs35647923 | TOMM40 | 0.4157 | CTT | 3.09E-10 | 0.007278 | -- | 2.254E-11 | | Old |
| 11 | 19 | 19-44894695-C-T | rs116881820 | TOMM40 | 0.0265 | C | 5.42E-54 | 0.000134 | ++ | 9.889E-51 | 1.744E-53 | Old |
| 11 | 19 | 19-44895376-G-C | rs11668327 | TOMM40 | 0.1589 | C | 2.96E-30 | 0.00578 | -- | 3.057E-28 | 8.595E-22 | Old |

| | | | | | | | | | | | | |
|---|---|---|---|---|---|---|---|---|---|---|---|---|
| 11 | 19 | 19-44895528-C-T | rs79398853 | TOMM40 | 0.0263 | T | 8.59E-54 | 0.000256 | ++ | 4.564E-50 | 7.326E-55 | Old |
| 11 | 19 | 19-44896087-G-T | rs75687619 | TOMM40 | 0.0263 | T | 2.46E-53 | 0.000188 | ++ | 6.064E-50 | 0.006838 | Old |
| 11 | 19 | 19-44896639-G-A | rs76366838 | TOMM40 | 0.0264 | A | 2.08E-53 | 0.000213 | ++ | 6.766E-50 | 0.006838 | Old |
| 11 | 19 | 19-44897227-TG-T | | TOMM40 | 0.0271 | TG | 8.46E-55 | 1.95E-05 | ++ | 7.966E-53 | | Old |
| 11 | 19 | 19-44897468-C-T | rs114536010 | TOMM40 | 0.0263 | T | 1.97E-53 | 0.000226 | ++ | 6.941E-50 | 1.386E-54 | Old |
| 11 | 19 | 19-44897776-C-CA | | TOMM40 | 0.4129 | CA | 3.55E-31 | 0.408197 | -- | 8.94E-24 | | Old |
| 11 | 19 | 19-44897790-AG-A | rs1555789087 | TOMM40 | 0.1241 | AG | 7.7E-266 | 5.32E-19 | ++ | 3.22E-257 | | Old |
| 11 | 19 | 19-44898409-G-A | rs8106922 | TOMM40 | 0.3815 | G | 4.72E-30 | 0.232396 | -- | 1.39E-23 | 4.969E-40 | Old |
| 11 | 19 | 19-44899220-C-T | rs34878901 | TOMM40 | 0.3985 | T | 1.42E-30 | 0.169324 | -- | 3.294E-24 | 0.000301 | Old |
| 11 | 19 | 19-44899959-C-T | rs115881343 | TOMM40 | 0.0278 | T | 1.02E-55 | 0.000401 | ++ | 2.504E-51 | 3.016E-57 | Old |
| 11 | 19 | 19-44900155-C-T | rs1160985 | TOMM40 | 0.4597 | T | 1.1E-44 | 0.00203 | -- | 7.095E-40 | 3.798E-85 | Old |
| 11 | 19 | 19-44901174-C-T | rs741780 | TOMM40 | 0.4598 | C | 3.13E-44 | 0.002199 | -- | 1.917E-39 | 5.139E-84 | Old |
| 11 | 19 | 19-44901434-G-A | | TOMM40 | 0.2644 | A | 3.6E-29 | 0.002029 | -- | 5.088E-28 | 1.392E-20 | Old |
| 11 | 19 | 19-44901548-AACACGGTGAAACTCCGTCTCTACT-A | rs113492558 | TOMM40 | 0.0291 | A | 1.93E-57 | 4.69E-05 | ++ | 2.867E-54 | 1.152E-55 | Old |
| 11 | 19 | 19-44901600-C-T | rs112019714 | TOMM40 | 0.0291 | C | 2.11E-57 | 4.65E-05 | ++ | 3.059E-54 | 0.00277 | Old |
| 11 | 19 | 19-44901715-C-T | rs1038025 | TOMM40 | 0.4598 | C | 1.47E-44 | 0.002393 | -- | 1.298E-39 | 2.802E-84 | Old |
| 11 | 19 | 19-44902264-G-C | rs1305062 | TOMM40 | 0.3918 | C | 5.31E-31 | 0.095291 | -- | 2.671E-25 | 4.689E-47 | Old |
| 11 | 19 | 19-44903416-G-A | rs10119 | TOMM40 | 0.2845 | A | 1.8E-143 | 3.21E-09 | ++ | 4.07E-131 | 1.67E-161 | Old |
| 11 | 19 | 19-44905579-G-T | | APOE | 0.4701 | T | 4.22E-30 | 0.007666 | ++ | 1.754E-27 | 4.385E-36 | Old |
| 11 | 19 | 19-44905910-G-C | | APOE | 0.3484 | C | 1.1E-46 | 4.38E-05 | -- | 4.297E-45 | 3.983E-33 | Old |
| 11 | 19 | 19-44906745-G-A | rs769449 | APOE | 0.1106 | A | 2.1E-299 | 4.75E-23 | ++ | 1.23E-291 | 6.74E-13 | Old |
| 11 | 19 | 19-44907187-G-A | rs769450 | APOE | 0.3891 | A | 2.6E-30 | 0.051863 | -- | 1.41E-25 | 2.699E-49 | Old |
| 11 | 19 | 19-44908822-C-T | rs7412 | APOE | 0.079 | T | 7.89E-31 | 1.53E-07 | -- | 1.321E-35 | 5.297E-39 | Old |
| 11 | 19 | 19-44909665-AC-A | rs537741299 | AC011481.3 | 0.007 | A | 2.58E-16 | 0.010115 | ++ | 1.56E-16 | 7.909E-14 | Old |
| 11 | 19 | 19-44909698-C-A | rs1081105 | AC011481.3 | 0.0281 | C | 1.92E-58 | 0.000112 | ++ | 1.418E-54 | 1.351E-62 | Old |
| 11 | 19 | 19-44909976-G-T | rs1065853 | AC011481.3 | 0.0802 | T | 3.06E-30 | 1.65E-07 | -- | 5.145E-35 | | Old |
| 11 | 19 | 19-44910319-C-T | rs75627662 | AC011481.3 | 0.1916 | T | 5.8E-120 | 7.29E-06 | ++ | 3.96E-106 | 1.148E-84 | Old |
| 11 | 19 | 19-44912456-G-A | rs10414043 | AC011481.3 | 0.1261 | A | 3.1E-289 | 1.51E-20 | ++ | 7.93E-275 | 3.56E-146 | Old |
| 11 | 19 | 19-44912678-G-T | rs7256200 | AC011481.3 | 0.126 | T | 2.2E-288 | 1.61E-20 | ++ | 5.4E-274 | | Old |
| 11 | 19 | 19-44912921-G-T | rs483082 | AC011481.3 | 0.2449 | T | 9.3E-181 | 1.79E-09 | ++ | 3.74E-161 | 3.72E-139 | Old |
| 11 | 19 | 19-44913034-C-T | rs59325138 | AC011481.3 | 0.3774 | T | 1.05E-26 | 0.009163 | -- | 1.092E-24 | 3.511E-39 | Old |
| 11 | 19 | 19-44913484-C-T | rs438811 | AC011481.3 | 0.248 | T | 6.7E-182 | 1.84E-10 | ++ | 2.59E-164 | 9.06E-161 | Old |
| 11 | 19 | 19-44914381-C-CTTCG | rs11568822 | AC011481.3 | 0.2283 | CTTCG | 1.1E-178 | 8.53E-11 | ++ | 1.61E-163 | 3.27E-124 | Old |
| 11 | 19 | 19-44915229-G-A | rs12691088 | APOC1 | 0.0196 | A | 1.63E-53 | 0.001741 | ++ | 2.587E-50 | 0.06546 | Old |
| 11 | 19 | 19-44915533-C-T | rs5117 | APOC1 | 0.2283 | C | 9.4E-177 | 3.62E-10 | ++ | 1.25E-160 | 8.74E-129 | Old |
| 11 | 19 | 19-44917843-G-A | rs3925681 | APOC1 | 0.397 | A | 1.95E-35 | 0.00121 | -- | 2.294E-33 | 1.001E-42 | Old |
| 11 | 19 | 19-44917947-C-T | rs150966173 | APOC1 | 0.029 | T | 1.28E-56 | 0.001165 | ++ | 7.279E-51 | 4.119E-60 | Old |
| 11 | 19 | 19-44917961-GA-G | rs374095935 | APOC1 | 0.0114 | GA | 4.29E-31 | 0.042697 | ++ | 1.099E-28 | | Old |
| 11 | 19 | 19-44917997-G-A | rs12721046 | APOC1 | 0.1385 | A | 1.7E-230 | 3.32E-21 | ++ | 4.97E-231 | 6.77E-159 | Old |
| 11 | 19 | 19-44918715-AG-A | rs12721052 | APOC1 | 0.3278 | A | 5.04E-20 | 0.027151 | -- | 1.867E-18 | 2.636E-31 | Old |
| 11 | 19 | 19-44918903-G-C | rs12721051 | APOC1 | 0.1707 | G | 6.4E-292 | 2.61E-22 | ++ | 1.57E-290 | 2.14E-228 | Old |
| 11 | 19 | 19-44919589-G-A | rs56131196 | APOC1 | 0.1805 | A | 2.1E-291 | 1.15E-23 | ++ | 7.74E-288 | 1.89E-202 | Old |

| | | | | | | | | | | | | |
|---|---|---|---|---|---|---|---|---|---|---|---|---|
| 11 | 19 | 19-44919689-G-A | rs4420638 | APOC1 | 0.181 | G | 1.5E-297 | 8.06E-24 | ++ | 3.77E-288 | 2.22E-202 | Old |
| 11 | 19 | 19-44920730-C-CA | rs35733971 | APOC1, APOC4 | 0.1693 | CA | 3.3E-226 | 3.92E-17 | ++ | 1.59E-215 | | Old |
| 11 | 19 | 19-44921809-G-A | rs188535946 | APOC1, APOC4 | 0.0286 | A | 3.89E-55 | 0.000198 | ++ | 1.871E-51 | | Old |
| 11 | 19 | 19-44923868-T-A | rs111789331 | APOC1, APOC4 | 0.1397 | A | 9.4E-232 | 3.29E-19 | ++ | 2.14E-228 | 1.84E-158 | Old |
| 11 | 19 | 19-44924977-G-A | rs66626994 | APOC1, APOC4 | 0.1471 | A | 3.1E-227 | 1.71E-19 | ++ | 5.84E-223 | 1.17E-130 | Old |
| 11 | 19 | 19-44925202-C-T | rs4803772 | APOC1, APOC4 | 0.3216 | T | 9.25E-20 | 0.030274 | -- | 3.768E-18 | 2.536E-31 | Old |
| 11 | 19 | 19-44926451-G-C | rs60049679 | APOC1, APOC4 | 0.0877 | C | 9.94E-24 | 0.00795 | ++ | 3.926E-22 | 4.495E-23 | Old |
| 11 | 19 | 19-44928426-GAA-G | rs569925552 | APOC1, APOC4 | 0.0121 | GAA | 5.06E-12 | 0.013102 | ++ | 1.046E-12 | | Old |
| 11 | 19 | 19-44935297-C-T | rs7254133 | APOC1, APOC4 | 0.3102 | T | 2.84E-30 | 0.016581 | ++ | 6.177E-27 | 3.048E-14 | Old |
| 11 | 19 | 19-44935318-C-A | rs141441332 | APOC1, APOC4 | 0.012 | A | 6.88E-11 | 0.015409 | ++ | 1.273E-11 | 5.826E-08 | Old |
| 11 | 19 | 19-44973974-G-C | rs57465754 | CLPTM1 | 0.2396 | G | 5.2E-10 | 0.143058 | ++ | 4.541E-09 | 0.001831 | Old |
| 11 | 19 | 19-45019031-G-A | rs74359223 | RELB | 0.0117 | A | 6.14E-09 | 0.83386 | ++ | 4.032E-07 | 1.769E-09 | Old |
| 12 | 19 | 19-44718265-T-C | rs56198711 | AC243964.2 | 0.1427 | C | 4.05E-08 | 0.101429 | ++ | 4.63E-08 | 0.1241 | Old |
| 12 | 19 | 19-44719517-G-C | rs73037426 | AC243964.2 | 0.1418 | C | 1.66E-09 | 0.023509 | ++ | 3.362E-10 | 0.000003162 | Old |
| 12 | 19 | 19-44731001-AAAAAAAAAAAAAAAGAAAAG-A | | RF00285, BCL3 | 0.1643 | AAAAAAA | 5.13E-09 | 0.019179 | ++ | 7.108E-10 | | Old |
| 12 | 19 | 19-44737327-C-T | rs55923289 | RF00285, BCL3 | 0.1178 | C | 5.13E-09 | 0.006218 | ++ | 1.562E-10 | 0.000003661 | Old |
| 12 | 19 | 19-44738850-G-A | rs2927438 | RF00285, BCL3 | 0.2069 | A | 9.35E-17 | 0.015781 | ++ | 2.102E-16 | 0.0004052 | Old |
| 12 | 19 | 19-44750911-C-A | rs8103315 | BCL3 | 0.1182 | A | 6.37E-15 | 0.01398 | ++ | 2.719E-15 | 3.416E-08 | Old |
| 13 | 19 | 19-44720227-G-A | rs111740474 | AC243964.2 | 0.0143 | A | 4.98E-09 | 0.004182 | ++ | 8.6E-11 | 0.0001601 | Old |
| 13 | 19 | 19-44833336-C-T | rs112616980 | BCAM, NECTIN2 | 0.0044 | T | 4.09E-14 | 0.002434 | ++ | 1.087E-15 | 0.0003382 | Old |
| 14 | 19 | 19-44728895-G-A | rs62117160 | RF00285, BCL3 | 0.0389 | A | 1.15E-09 | 0.001297 | -- | 6.229E-12 | 0.00009026 | Old |
| 14 | 19 | 19-44738916-G-A | rs1531517 | RF00285, BCL3 | 0.0876 | A | 4.81E-09 | 0.105996 | -- | 1.551E-08 | 0.2331 | Old |
| 14 | 19 | 19-44739710-C-T | rs62117204 | RF00285, BCL3 | 0.0713 | T | 2.11E-08 | 0.034097 | -- | 5.973E-09 | 0.008649 | Old |
| 14 | 19 | 19-44744370-G-A | rs4803750 | RF00285, BCL3 | 0.0776 | G | 5.45E-09 | 0.00134 | -- | 3.684E-11 | 0.007579 | Old |
| 14 | 19 | 19-44750234-C-T | rs10401176 | BCL3 | 0.1425 | T | 8.15E-11 | 0.010885 | -- | 1.682E-11 | 0.0001742 | Old |
| 14 | 19 | 19-44750354-ATTGGC-A | rs66586168 | BCL3 | 0.1398 | A | 1.06E-10 | 0.00564 | -- | 7.723E-12 | | Old |
| 14 | 19 | 19-44752009-C-T | rs62117205 | BCL3 | 0.0685 | C | 5.19E-09 | 0.013723 | -- | 4.738E-10 | 0.002023 | Old |
| 14 | 19 | 19-44752422-G-C | rs62117206 | BCL3 | 0.0696 | C | 4.28E-09 | 0.012994 | -- | 3.834E-10 | 0.0038 | Old |
| 14 | 19 | 19-44816374-G-A | rs118147862 | BCAM | 0.0399 | A | 4.21E-12 | 0.00329 | -- | 8.954E-14 | 6.472E-07 | Old |
| 14 | 19 | 19-44844304-T-G | rs11668738 | BCAM, NECTIN2 | 0.165 | G | 5.33E-09 | 0.113785 | -- | 1.195E-08 | 0.6935 | Old |
| 14 | 19 | 19-44850981-C-G | rs2972566 | NECTIN2 | 0.1829 | C | 4.12E-10 | 0.025487 | -- | 1.626E-10 | 0.9554 | Old |
| 14 | 19 | 19-44879418-G-A | rs41290120 | NECTIN2 | 0.0417 | A | 9.03E-16 | 0.012804 | -- | 5.526E-16 | 0.03073 | Old |
| 14 | 19 | 19-44881148-C-T | rs73052307 | NECTIN2 | 0.137 | C | 3.75E-25 | 0.033998 | -- | 1.579E-22 | 0.02503 | Old |
| 14 | 19 | 19-44916968-TA-T | rs753649749 | APOC1 | 0.0271 | TA | 1.65E-11 | 0.040917 | -- | 1.639E-11 | | Old |
| 14 | 19 | 19-44919304-G-T | rs1064725 | APOC1 | 0.0399 | G | 4.04E-08 | 0.256311 | -- | 3.504E-07 | 0.00241 | Old |
| 15 | 19 | 19-44739483-G-A | rs2927439 | RF00285, BCL3 | 0.3517 | G | 7.26E-09 | 0.000567 | -- | 1.954E-11 | 0.0004962 | Old |
| 15 | 19 | 19-44743791-C-T | rs4803748 | RF00285, BCL3 | 0.4247 | T | 3.49E-12 | 0.000491 | -- | 1.478E-14 | 0.000001162 | Old |
| 15 | 19 | 19-44747899-C-A | rs2965169 | BCL3 | 0.4307 | C | 1.28E-11 | 3.08E-05 | -- | 2.151E-15 | 2.379E-07 | Old |
| 15 | 19 | 19-44886339-G-A | rs7254892 | AC011481.2 | 0.0446 | A | 6.75E-13 | 0.000436 | -- | 8.434E-15 | | Old |
| 15 | 19 | 19-44893972-G-A | rs1160983 | TOMM40 | 0.037 | A | 1.43E-13 | 6.81E-05 | -- | 1.499E-16 | 1.06E-13 | Old |
| 15 | 19 | 19-44897490-T-A | rs61679753 | TOMM40 | 0.0404 | A | 9.9E-14 | 0.000293 | -- | 8.891E-16 | 1.638E-17 | Old |

| | | | | | | | | | | | | |
|---|---|---|---|---|---|---|---|---|---|---|---|---|
| 15 | 19 | 19-44899005-G-T | rs111784051 | TOMM40 | 0.0421 | G | 6E-14 | 0.003113 | -- | 2.023E-14 | 1.231E-17 | Old |
| 15 | 19 | 19-44921921-G-A | rs190712692 | APOC1, APOC4 | 0.055 | A | 9.77E-21 | 0.000105 | -- | 1.064E-22 | | Old |
| 15 | 19 | 19-44923535-G-A | rs141622900 | APOC1, APOC4 | 0.055 | A | 1.55E-20 | 0.000222 | -- | 4.792E-22 | 5.358E-25 | Old |
| 16 | 19 | 19-44766291-C-T | rs193249943 | RF00156, CBLC | 0.0059 | T | 4.13E-10 | 0.032331 | ++ | 1.067E-10 | 0.0004719 | Old |
| 16 | 19 | 19-44815238-G-A | rs180887453 | BCAM | 0.0034 | G | 1.02E-10 | 0.011046 | ++ | 6.671E-12 | 0.0006873 | Old |
| 16 | 19 | 19-44819414-C-T | rs117012738 | BCAM | 0.0014 | T | 3.55E-08 | 0.194619 | ++ | 1.541E-07 | 0.005085 | Old |
| 17 | 19 | 19-44766468-C-T | rs139326841 | RF00156, CBLC | 0.0213 | C | 3.62E-10 | 0.38753 | ++ | 4.921E-09 | 0.000003711 | Old |
| 17 | 19 | 19-44807827-G-A | rs140824606 | AC092306.1, BCAM | 0.0232 | A | 1.68E-10 | 0.211157 | ++ | 8.684E-10 | 2.738E-07 | Old |
| 17 | 19 | 19-44841241-TTAAAAA-T | rs71171294 | BCAM, NECTIN2 | 0.0113 | TTAAAAA | 7.35E-16 | 0.466616 | ++ | 5.992E-13 | | Old |
| 17 | 19 | 19-44842026-C-A | rs61642202 | BCAM, NECTIN2 | 0.0113 | C | 4.9E-17 | 0.180988 | ++ | 6.361E-15 | 0.00002925 | Old |
| 17 | 19 | 19-44852010-T-A | rs76205446 | NECTIN2 | 0.0165 | A | 4.67E-15 | 0.313311 | ++ | 7.512E-12 | 0.003059 | Old |
| 17 | 19 | 19-44854769-C-T | rs143459034 | NECTIN2 | 0.0121 | T | 5.81E-16 | 0.159859 | ++ | 6.083E-14 | 0.000884 | Old |
| 17 | 19 | 19-44856410-G-A | rs41289514 | NECTIN2 | 0.0112 | G | 1.49E-16 | 0.153395 | ++ | 1.093E-14 | 0.000005607 | Old |
| 18 | 19 | 19-44775001-G-A | rs187010311 | RF00156, CBLC | 0.0049 | A | 1.06E-09 | 0.990993 | ++ | 6.858E-08 | 0.000001298 | Old |
| 19 | 19 | 19-44784672-C-CTCCAT | rs569705402 | CBLC | 0.0064 | C | 8.18E-10 | 0.553701 | ++ | 1.387E-08 | 0.0001013 | Old |
| 20 | 19 | 19-44825110-A-T | rs58132661 | BCAM, NECTIN2 | 0.3172 | A | 8.07E-27 | 0.085196 | ++ | 1.551E-22 | 4.717E-08 | Old |
| 20 | 19 | 19-44825122-T-A | rs58826447 | BCAM, NECTIN2 | 0.3173 | A | 8.33E-27 | 0.086004 | ++ | 1.627E-22 | 3.098E-08 | Old |
| 20 | 19 | 19-44825123-C-A | rs58446550 | BCAM, NECTIN2 | 0.3173 | A | 8.33E-27 | 0.086978 | ++ | 1.676E-22 | 3.365E-08 | Old |
| 20 | 19 | 19-44825150-GAC-G | rs34798982 | BCAM, NECTIN2 | 0.4908 | G | 5.03E-16 | 0.085684 | ++ | 2.48E-14 | | Old |
| 20 | 19 | 19-44827846-A-G | rs7343130 | BCAM, NECTIN2 | 0.4235 | G | 9.15E-10 | 0.017058 | -- | 1.679E-10 | | Old |
| 20 | 19 | 19-44829763-TA-T | | BCAM, NECTIN2 | 0.2349 | TA | 1.06E-08 | 0.036301 | -- | 4.298E-09 | | Old |
| 20 | 19 | 19-44832419-G-A | rs56394238 | BCAM, NECTIN2 | 0.4471 | G | 8.44E-21 | 0.009124 | ++ | 5.332E-20 | 0.005204 | Old |
| 20 | 19 | 19-44832778-C-T | rs7359852 | BCAM, NECTIN2 | 0.3072 | C | 2.59E-24 | 0.007045 | ++ | 6.742E-23 | 0.00001223 | Old |
| 20 | 19 | 19-44833186-G-A | rs3021439 | BCAM, NECTIN2 | 0.2996 | A | 4.36E-24 | 0.006242 | ++ | 7.125E-23 | 0.0000278 | Old |
| 20 | 19 | 19-44834128-G-C | rs2927480 | BCAM, NECTIN2 | 0.2943 | C | 4.21E-24 | 0.006474 | ++ | 6.096E-23 | | Old |
| 20 | 19 | 19-44834247-G-C | rs11667241 | BCAM, NECTIN2 | 0.317 | C | 1.21E-20 | 0.022191 | ++ | 4.364E-19 | 0.00004 | Old |
| 20 | 19 | 19-44834993-AT-A | rs200722375 | BCAM, NECTIN2 | 0.2925 | A | 1.5E-23 | 0.006143 | ++ | 1.526E-22 | 0.000001829 | Old |
| 20 | 19 | 19-44837479-G-A | rs73048293 | BCAM, NECTIN2 | 0.2989 | A | 1.55E-23 | 0.013334 | ++ | 7.543E-22 | 0.0000101 | Old |
| 20 | 19 | 19-44838647-G-A | rs12459575 | BCAM, NECTIN2 | 0.299 | A | 8.89E-24 | 0.010368 | ++ | 2.896E-22 | 0.00001349 | Old |
| 20 | 19 | 19-44842530-A-T | rs111371860 | BCAM, NECTIN2 | 0.0588 | A | 3.87E-09 | 0.003659 | -- | 7.723E-11 | 4.612E-08 | Old |
| 20 | 19 | 19-44844403-GA-G | | BCAM, NECTIN2 | 0.4313 | GA | 3.74E-11 | 0.432262 | -- | 3.086E-09 | | Old |
| 20 | 19 | 19-44844654-C-A | rs4452060 | BCAM, NECTIN2 | 0.4281 | A | 9.05E-11 | 0.066257 | ++ | 2.242E-10 | 0.02366 | Old |
| 20 | 19 | 19-44849230-T-A | rs12974942 | NECTIN2 | 0.4125 | T | 1.08E-12 | 0.019276 | ++ | 8.305E-13 | 0.002585 | Old |
| 20 | 19 | 19-44852464-G-C | rs2972559 | NECTIN2 | 0.2591 | G | 3.11E-29 | 0.016487 | ++ | 4.187E-26 | 8.952E-12 | Old |
| 20 | 19 | 19-44853746-G-C | rs35396326 | NECTIN2 | 0.2703 | G | 5.28E-25 | 0.035454 | ++ | 3.578E-22 | 9.136E-09 | Old |
| 20 | 19 | 19-44854034-G-C | rs4803763 | NECTIN2 | 0.2552 | C | 1.54E-29 | 0.011312 | ++ | 9.02E-27 | | Old |
| 20 | 19 | 19-44854682-G-A | rs2927468 | NECTIN2 | 0.488 | A | 6.37E-32 | 0.004745 | -- | 2.827E-29 | 1.77E-08 | Old |
| 20 | 19 | 19-44856449-G-A | rs12462573 | NECTIN2 | 0.2593 | A | 1.31E-31 | 0.006275 | ++ | 9.646E-29 | 5.326E-12 | Old |
| 20 | 19 | 19-44858389-G-A | rs365653 | NECTIN2 | 0.1317 | G | 2.49E-16 | 0.045724 | -- | 9.822E-15 | 0.000002902 | Old |
| 20 | 19 | 19-44858568-C-T | rs418227 | NECTIN2 | 0.3591 | C | 1.57E-18 | 0.001459 | ++ | 1.84E-19 | 0.001655 | Old |
| 20 | 19 | 19-44858703-C-T | rs417193 | NECTIN2 | 0.353 | C | 1.31E-18 | 0.002519 | ++ | 3.503E-19 | 0.0001149 | Old |
| 20 | 19 | 19-44859012-G-T | rs2436474 | NECTIN2 | 0.3494 | T | 9.07E-19 | 0.001636 | ++ | 1.285E-19 | 0.00003081 | Old |
| 20 | 19 | 19-44859410-G-A | rs377702 | NECTIN2 | 0.3519 | A | 1.33E-18 | 0.001739 | ++ | 2.005E-19 | 0.0001232 | Old |

| | | | | | | | | | | | | |
|---|---|---|---|---|---|---|---|---|---|---|---|---|
| 20 | 19 | 19-44859997-G-A | rs387369 | NECTIN2 | 0.3509 | A | 2E-18 | 0.001734 | ++ | 2.813E-19 | 0.0002523 | Old |
| 20 | 19 | 19-44860443-G-A | rs12978931 | NECTIN2 | 0.1948 | G | 2.84E-12 | 0.064 | -- | 1.27E-11 | 0.0006718 | Old |
| 20 | 19 | 19-44860534-G-T | rs384973 | NECTIN2 | 0.3519 | G | 1.38E-18 | 0.001355 | ++ | 1.435E-19 | 0.0007124 | Old |
| 20 | 19 | 19-44861366-C-T | rs411856 | NECTIN2 | 0.3512 | T | 8.37E-19 | 0.001636 | ++ | 1.225E-19 | 0.0003212 | Old |
| 20 | 19 | 19-44861558-G-A | rs395710 | NECTIN2 | 0.3513 | G | 6.54E-19 | 0.001624 | ++ | 9.844E-20 | 0.000308 | Old |
| 20 | 19 | 19-44861815-G-A | rs555608 | NECTIN2 | 0.3513 | G | 7.15E-19 | 0.001389 | ++ | 8.364E-20 | 0.000378 | Old |
| 20 | 19 | 19-44861991-C-T | rs2436475 | NECTIN2 | 0.351 | C | 9.15E-19 | 0.001671 | ++ | 1.362E-19 | 0.0004852 | Old |
| 20 | 19 | 19-44862384-C-A | rs12980631 | NECTIN2 | 0.3515 | C | 8.15E-19 | 0.001187 | ++ | 7.292E-20 | 0.0004417 | Old |
| 20 | 19 | 19-44862475-AAAG-A | rs56283909 | NECTIN2 | 0.3515 | AAAG | 9.83E-19 | 0.001567 | ++ | 1.311E-19 | | Old |
| 20 | 19 | 19-44862560-G-A | rs11665829 | NECTIN2 | 0.3516 | A | 1.2E-18 | 0.001464 | ++ | 1.397E-19 | | Old |
| 20 | 19 | 19-44863088-G-A | rs416116 | NECTIN2 | 0.3512 | A | 1.45E-18 | 0.001353 | ++ | 1.452E-19 | 0.0008026 | Old |
| 20 | 19 | 19-44863346-G-A | rs521629 | NECTIN2 | 0.3515 | A | 7.52E-19 | 0.001637 | ++ | 1.104E-19 | 0.0001002 | Old |
| 20 | 19 | 19-44863522-C-T | rs519825 | NECTIN2 | 0.3514 | C | 9.43E-19 | 0.001713 | ++ | 1.438E-19 | 0.0001335 | Old |
| 20 | 19 | 19-44864825-C-A | rs520283 | NECTIN2 | 0.3509 | A | 1.34E-18 | 0.002168 | ++ | 2.76E-19 | 0.0001408 | Old |
| 20 | 19 | 19-44867313-A-C | rs565566 | NECTIN2 | 0.3493 | C | 5.65E-19 | 0.001885 | ++ | 1.077E-19 | 0.1351 | Old |
| 20 | 19 | 19-44867392-A-G | rs564724 | NECTIN2 | 0.352 | G | 6.06E-19 | 0.000862 | ++ | 3.607E-20 | 0.0000802 | Old |
| 20 | 19 | 19-44867416-A-C | rs510297 | NECTIN2 | 0.3525 | A | 7E-19 | 0.000889 | ++ | 4.299E-20 | 0.0001096 | Old |
| 20 | 19 | 19-44874077-G-A | rs393584 | NECTIN2 | 0.3581 | A | 2.8E-12 | 0.029524 | -- | 3.209E-12 | | Old |
| 20 | 19 | 19-44875803-C-A | rs387976 | NECTIN2 | 0.3613 | C | 3.53E-12 | 0.018995 | -- | 1.956E-12 | 0.1333 | Old |
| 20 | 19 | 19-44879727-G-T | rs11669338 | NECTIN2 | 0.091 | G | 5.24E-10 | 0.779466 | -- | 0.000000154 | 0.00468 | Old |
| 20 | 19 | 19-44879780-T-A | rs11673139 | NECTIN2 | 0.091 | T | 5.45E-10 | 0.763783 | -- | 1.483E-07 | 0.004591 | Old |
| 20 | 19 | 19-44880859-A-G | rs406315 | NECTIN2 | 0.3422 | A | 1.48E-24 | 0.000231 | ++ | 8.012E-26 | 4.532E-07 | Old |
| 20 | 19 | 19-44893642-C-T | | TOMM40 | 0.4962 | T | 8E-12 | 0.284955 | ++ | 4.903E-10 | 4.006E-13 | Old |
| 20 | 19 | 19-44894255-A-C | rs157585 | TOMM40 | 0.4991 | C | 6.49E-16 | 0.197835 | ++ | 1.599E-13 | | Old |
| 20 | 19 | 19-44895007-C-T | rs157588 | TOMM40 | 0.4999 | C | 3.32E-16 | 0.228368 | ++ | 1.529E-13 | 6.807E-18 | Old |
| 20 | 19 | 19-44895459-C-A | | TOMM40 | 0.4931 | A | 6.39E-18 | 0.26959 | ++ | 1.26E-14 | 9.478E-19 | Old |
| 20 | 19 | 19-44905307-T-A | rs449647 | APOE | 0.1834 | T | 8.16E-27 | 0.064019 | -- | 4.518E-22 | 0.0002009 | Old |
| 21 | 19 | 19-44836881-A-G | | BCAM, NECTIN2 | 0.3156 | A | 4.87E-16 | 0.144106 | -- | 9.291E-14 | 0.00007397 | Old |
| 21 | 19 | 19-44838691-A-G | rs10407439 | BCAM, NECTIN2 | 0.3009 | G | 5.65E-17 | 0.101298 | -- | 7.281E-15 | 0.009062 | Old |
| 22 | 19 | 19-44862268-C-CACTGTGTGTGGTGGCGGGCACCTG | rs144767421 | NECTIN2 | 0.3669 | C | 5.57E-19 | 0.717028 | ++ | 5.228E-19 | | Old |
| 23 | 19 | 19-44890947-G-A | rs561654715 | TOMM40 | 0.0027 | A | 1.49E-11 | 0.2552 | ++ | 1.171E-10 | 0.000003295 | Old |
| 24 | 19 | 19-44921095-T-A | rs56369833 | APOC1, APOC4 | 0.1572 | T | 6.8E-228 | 0.683605 | ++ | 6.58E-228 | | Old |
| 25 | 19 | 19-44925417-C-T | rs565334527 | APOC1, APOC4 | 0.0016 | C | 8.15E-11 | 0.05508 | ++ | 2.945E-11 | 0.003351 | Old |
| 26 | 19 | 19-44933496-C-T | rs114533385 | APOC1, APOC4 | 0.0184 | T | 8.08E-11 | 0.006468 | ++ | 1.442E-11 | 0.0009272 | Old |
| 27 | 19 | 19-45094524-TA-T | rs149151450 | MARK4, PPP1R37 | 0.0922 | TA | 1.99E-09 | 0.008637 | ++ | 1.397E-10 | 0.002281 | Old |
| 27 | 19 | 19-45102867-C-T | rs7248421 | MARK4, PPP1R37 | 0.0906 | T | 8.81E-10 | 0.007024 | ++ | 5.204E-11 | 0.0007487 | Old |
| 27 | 19 | 19-45108968-C-T | rs12462040 | MARK4, PPP1R37 | 0.0901 | T | 1.76E-09 | 0.006878 | ++ | 9.131E-11 | 0.0008119 | Old |
| 27 | 19 | 19-45111343-C-T | rs34545713 | MARK4, PPP1R37 | 0.0914 | T | 7.19E-10 | 0.004438 | ++ | 2.456E-11 | 0.002414 | Old |
| 27 | 19 | 19-45115701-G-A | rs2004357 | MARK4, PPP1R37 | 0.0917 | A | 6.12E-10 | 0.006538 | ++ | 3.497E-11 | | Old |
| 27 | 19 | 19-45118499-TG-T | rs144328302 | MARK4, PPP1R37 | 0.0892 | T | 4.98E-10 | 0.003829 | ++ | 1.426E-11 | 0.004282 | Old |
| 27 | 19 | 19-45123977-C-T | rs10405086 | MARK4, PPP1R37 | 0.0921 | T | 5.18E-10 | 0.004553 | ++ | 1.939E-11 | 0.002723 | Old |
| 27 | 19 | 19-45128558-G-A | rs17643262 | MARK4, PPP1R37 | 0.093 | A | 1.9E-10 | 0.003778 | ++ | 6.506E-12 | 0.009767 | Old |

| | | | | | | | | | | | | |
|---|---|---|---|---|---|---|---|---|---|---|---|---|
| 27 | 19 | 19-45130428-G-A | rs754366 | MARK4, PPP1R37 | 0.0941 | A | 1.83E-10 | 0.004432 | ++ | 7.927E-12 | 0.01058 | Old |
| 27 | 19 | 19-45132943-C-T | rs1114832 | MARK4, PPP1R37 | 0.0916 | T | 2.88E-09 | 0.005988 | ++ | 1.155E-10 | 0.0005812 | Old |
| 27 | 19 | 19-45133061-C-A | rs1114831 | MARK4, PPP1R37 | 0.0896 | A | 2.41E-10 | 0.004305 | ++ | 8.714E-12 | 0.1156 | Old |
| 27 | 19 | 19-45134987-C-T | rs28620490 | MARK4, PPP1R37 | 0.0882 | T | 5.16E-10 | 0.004525 | ++ | 1.761E-11 | 0.0004345 | Old |
| 27 | 19 | 19-45137848-C-A | rs77401305 | MARK4, PPP1R37 | 0.0925 | A | 4.81E-09 | 0.005497 | ++ | 1.675E-10 | | Old |
| 27 | 19 | 19-45138786-G-A | rs10401157 | MARK4, PPP1R37 | 0.0887 | A | 3.14E-10 | 0.002211 | ++ | 4.753E-12 | 0.001052 | Old |
| 27 | 19 | 19-45139287-G-C | rs10401823 | MARK4, PPP1R37 | 0.0897 | G | 4.74E-10 | 0.006445 | ++ | 2.625E-11 | 0.1232 | Old |
| 27 | 19 | 19-45141096-T-A | rs78273125 | MARK4, PPP1R37 | 0.0885 | T | 7.37E-10 | 0.005501 | ++ | 3.074E-11 | 0.0003586 | Old |
| 27 | 19 | 19-45146841-G-T | rs74846209 | PPP1R37 | 0.0895 | T | 2.53E-10 | 0.005053 | ++ | 1.077E-11 | 0.001013 | Old |
| 27 | 19 | 19-45147128-C-T | rs1048699 | PPP1R37 | 0.0893 | T | 4.02E-10 | 0.00577 | ++ | 1.895E-11 | 0.0004258 | Old |
| 27 | 19 | 19-45147979-G-A | rs113321260 | MARK4 | 0.0924 | A | 8.41E-11 | 0.009227 | ++ | 9.853E-12 | 0.00132 | Old |
| 27 | 19 | 19-45148728-GGTATG | rs143019611 | MARK4 | 0.0895 | GGTAT | 5.55E-10 | 0.007919 | ++ | 3.789E-11 | 0.0007294 | Old |
| 27 | 19 | 19-45152075-C-T | rs28469095 | NKPD1 | 0.0895 | C | 2.96E-10 | 0.005847 | ++ | 1.476E-11 | 0.000568 | Old |
| 27 | 19 | 19-45157381-C-T | rs10417602 | MARK4, NKPD1 | 0.0777 | T | 5.74E-10 | 0.001368 | ++ | 5.211E-12 | | Old |
| 27 | 19 | 19-45165069-C-T | rs10410833 | MARK4, TRAPPC6A | 0.0807 | T | 5.64E-10 | 0.001738 | ++ | 7.271E-12 | | Old |
| 27 | 19 | 19-45170443-G-A | rs28367893 | MARK4, TRAPPC6A | 0.0809 | A | 1.22E-09 | 0.001385 | ++ | 1.107E-11 | 0.3631 | Old |
| 27 | 19 | 19-45171305-T-A | rs201189859 | MARK4, TRAPPC6A | 0.0934 | T | 1.27E-08 | 0.000715 | ++ | 4.986E-11 | 0.2293 | Old |
| 27 | 19 | 19-45181810-C-T | rs78979751 | MARK4 | 0.0738 | C | 4.14E-09 | 0.0019 | ++ | 4.216E-11 | | Old |
| 27 | 19 | 19-45186634-C-T | rs10415850 | MARK4 | 0.1333 | T | 3.71E-09 | 0.078206 | ++ | 7.41E-09 | 0.002292 | Old |
| 27 | 19 | 19-45186718-T-A | rs59678362 | MARK4 | 0.1318 | T | 9.25E-09 | 0.056335 | ++ | 8.938E-09 | 0.03119 | Old |
| 27 | 19 | 19-45186719-C-A | rs59839536 | MARK4 | 0.1327 | C | 1.61E-08 | 0.053255 | ++ | 1.292E-08 | 0.002611 | Old |
| 27 | 19 | 19-45187287-C-T | rs75858218 | MARK4 | 0.0783 | T | 3.35E-09 | 0.010733 | ++ | 2.764E-10 | 0.03969 | Old |
| 27 | 19 | 19-45187517-G-A | rs12462536 | MARK4 | 0.0923 | G | 2.09E-08 | 0.011909 | ++ | 1.969E-09 | 0.03556 | Old |
| 27 | 19 | 19-45188312-C-T | rs111243475 | MARK4 | 0.0413 | T | 1.78E-08 | 0.536492 | ++ | 0.00000142 | 0.8751 | Old |
| 28 | 19 | 19-45169864-G-A | rs76971643 | MARK4, TRAPPC6A | 0.0456 | A | 8.77E-09 | 0.404494 | -- | 1.656E-07 | 0.004742 | Old |
| 29 | 19 | 19-45199680-C-CTTCTCTTCTG | rs200726585 | MARK4 | 0.0171 | C | 1.27E-09 | 0.094875 | ++ | 2.952E-09 | | Old |
| 30 | 22 | 22-47711777-C-T | rs115880709 | AL117329.1 | 0.0286 | C | 1.5E-08 | 0.217798 | ++ | 1.449E-07 | 0.4052 | New |
