## Supplementary Table 6 for "Identification of 16 novel Alzheimer’s disease susceptibility loci using multi-ancestry meta-analyses of clinical Alzheimer’s disease and AD-by-proxy cases from four whole genome sequencing datasets"

**Supplementary table 6:** Independent loci that passed QC in our AoU GWAS, including variants that were not nominally significant in our clinical AD meta-analysis.
MAF - minor allele frequency; P - p-value; META_P - p-value for the meta analysis.

| Locus | Chromosome | Variant ID | Rsid | Gene | MAF | Effect allele | UKB_P | AOU_P | Direction (UKB, AOU) | UKB_AOU_META_P | NIA_NIMH_META_P | New or Old |
|---|---|---|---|---|---|---|---|---|---|---|---|---|
| 1 | 1 | 1-100385985-G-A | rs1203190 | CDC14A | 0.0012 | A | 0.210958 | 3.53E-11 | -+ | 0.0002627 | | New |
| 2 | 1 | 1-10152033-C-T | rs9751971 | UBE4B | 0.001 | T | 0.421838 | 1.6E-09 | -+ | 9.642E-09 | | New |
| 2 | 1 | 1-10152040-C-CCT | rs1367265 | UBE4B | 0.0033 | CCT | | 7.74E-24 | ?+ | 7.735E-24 | | New |
| 2 | 1 | 1-10152050-AATTT-A | | NA | 0.0012 | AATTT | | 3.35E-10 | ?+ | 3.351E-10 | | New |
| 3 | 1 | 1-10237828-G-A | rs5617412 | KIF1B | 0.0007 | A | 0.550187 | 2E-10 | ++ | 1.05E-09 | | New |
| 4 | 1 | 1-10508359-G-A | rs1298189 | PEX14 | 0.0002 | G | | 1.19E-10 | ?+ | 1.186E-10 | | New |
| 4 | 1 | 1-10508363-C-T | rs8798788 | PEX14 | 0.0002 | C | | 1.9E-10 | ?+ | 1.9E-10 | | New |
| 5 | 1 | 1-112574467-G-A | rs5744944 | ST7L | 0.0015 | A | 0.436548 | 4.55E-09 | -+ | 0.00006484 | 0.3173 | New |
| 5 | 1 | 1-112574476-G-T | rs3703503 | ST7L | 0.0021 | G | 0.021861 | 1.85E-10 | ++ | 1.805E-10 | 1 | New |
| 6 | 1 | 1-11709990-G-T | rs1902245 | DRAXIN | 0.0012 | T | 0.464033 | 3.38E-10 | ++ | 5.115E-10 | 0.806 | New |
| 7 | 1 | 1-121400518-GAA-G | rs1553361 | AC244021.1 | 0.0028 | G | | 1.18E-08 | ?+ | 1.185E-08 | | New |
| 8 | 1 | 1-125068456-G-T | rs1445112 | NONE, NONE | 0.0005 | G | | 1.18E-09 | ?+ | 1.177E-09 | | New |
| 9 | 1 | 1-125176581-G-T | rs1452815 | NONE, NONE | 0.0007 | T | | 1.79E-08 | ?+ | 1.791E-08 | | New |
| 10 | 1 | 1-1277184-ATG-A | | NA | 0.0006 | ATG | | 3.11E-08 | ?+ | 3.11E-08 | | Old |
| 11 | 1 | 1-13530355-G-A | rs1010617 | AL354712.1, PDPN | 0.001 | A | 0.96944 | 6.63E-10 | -+ | 6.943E-09 | | New |
| 12 | 1 | 1-1370032-C-CAG | | NA | 0.0008 | CAG | | 3.22E-10 | ?+ | 3.223E-10 | | Old |
| 13 | 1 | 1-143195687-TGAATGGAATCATCATC-T | | NA | 0.0012 | TGAATGGAATCATCAT | | 1.41E-11 | ?+ | 1.413E-11 | | New |
| 14 | 1 | 1-143202364-A-G | rs1163770 | NONE, RNA5SP533 | 0.006 | A | | 6.47E-12 | ?+ | 6.47E-12 | | New |
| 15 | 1 | 1-143210077-A-T | rs1225279 | NONE, RNA5SP533 | 0.012 | T | 0.728836 | 9.7E-118 | -+ | 2.49E-116 | | New |
| 16 | 1 | 1-143253866-C-T | rs1648038 | NONE, RNA5SP533 | 0.0031 | T | | 2.19E-11 | ?+ | 2.19E-11 | | New |
| 17 | 1 | 1-143272094-G-A | rs1237780 | NONE, RNA5SP533 | 0.0008 | G | 0.149567 | 1.14E-11 | -+ | 1.789E-10 | | New |

| | | | | | | | | | | | | |
|---|---|---|---|---|---|---|---|---|---|---|---|---|
| 18 | 1 | 1-143272167-C-CG | | NA | 0.0008 | C | | 3.94E-08 | ?+ | 3.939E-08 | | New |
| 19 | 1 | 1-143273397-G-A | rs1360957 | NONE, RNA5SP533 | 0.001 | G | 0.997336 | 4.13E-08 | -+ | 0.002372 | | New |
| 20 | 1 | 1-144565462-C-T | rs1658150 | RF00003, PPIAL4F | 0.001 | C | | 1.88E-11 | ?+ | 1.877E-11 | 0.3152 | New |
| 21 | 1 | 1-144636407-C-CA | | NA | 0.0101 | CA | | 8.17E-12 | ?+ | 8.167E-12 | | New |
| 21 | 1 | 1-144636407-CTTCA-C | | NA | 0.013 | CTTCA | | 1.34E-10 | ?+ | 1.337E-10 | | New |
| 22 | 1 | 1-148017581-AC-A | | NA | 0.001 | A | | 4.53E-09 | ?+ | 4.533E-09 | | New |
| 22 | 1 | 1-148017587-AAT-A | | NA | 0.0009 | A | | 3.67E-08 | ?+ | 3.672E-08 | | New |
| 23 | 1 | 1-148066853-GCAT-G | | RNVU1-7, NBPF11 | 0.0004 | GCAT | | 4.13E-10 | ?+ | 4.13E-10 | | New |
| 23 | 1 | 1-148066859-GCA-G | | RNVU1-7, NBPF11 | 0.0003 | GCA | | 1.35E-08 | ?+ | 1.354E-08 | | New |
| 24 | 1 | GCTGGGATTACAGGCGTGAGCCACTGCGCCTGGCTAACTTTTGTATTGTTAATGGAGACAGGGTTTCACCGTGTTGGCCAGGCTGCTCTCGAACTCCGGACCTCAAGTGATCCACTCACCTCT-G | | NA | 0.0062 | G | | 7.48E-28 | ?+ | 7.481E-28 | | New |
| 25 | 1 | 1-149945859-G-C | rs1559823 | OTUD7B | 0.0003 | G | 0.626237 | 3.27E-08 | -+ | 7.789E-07 | | New |
| 26 | 1 | 1-1500804-AAAAT-A | rs1264912 | ATAD3B, ATAD3A | 0.0032 | A | 0.87353 | 1.75E-15 | ++ | 1.39E-10 | 0.1118 | Old |
| 27 | 1 | 1-150304119-TG-T | | NA | 0.0009 | T | 0.497618 | 4.53E-10 | -+ | 2.234E-09 | | New |
| 28 | 1 | 1-150526800-ATT-A | | NA | 0.0008 | A | | 3.68E-12 | ?+ | 3.684E-12 | | New |
| 29 | 1 | 1-150671030-AG-A | rs1300768 | GOLPH3L | 0.0021 | A | | 1.31E-14 | ?+ | 1.31E-14 | | New |
| 30 | 1 | 1-150676899-C-T | rs9903227 | GOLPH3L | 0.0037 | C | 0.303001 | 9.78E-18 | ++ | 3.709E-08 | | New |
| 31 | 1 | 1-150960054-C-CA | rs1398444 | SETDB1 | 0.0042 | C | | 1.28E-16 | ?+ | 1.275E-16 | | New |
| 32 | 1 | 1-151083907-ATTT-A | | NA | 0.002 | ATTT | | 6.95E-09 | ?+ | 6.953E-09 | | New |
| 33 | 1 | 1-152109643-GT-G | | TCHH | 0.0016 | GT | | 8.29E-09 | ?+ | 8.293E-09 | | New |
| 33 | 1 | 1-152109644-C-CTT | | TCHH | 0.0019 | CTT | | 1.88E-10 | ?+ | 1.878E-10 | | New |

| | | | | | | | | | | | | |
|---|---|---|---|---|---|---|---|---|---|---|---|---|
| 34 | 1 | 1-152218911-G-A | | HRNR | 0.0097 | G | 0.386072 | 4.93E-09 | -+ | 8.804E-07 | 0.04487 | New |
| 35 | 1 | 1-155274178-ATTAG-A | rs1224276 | CLK2 | 0.0014 | A | | 1.81E-13 | ?+ | 1.805E-13 | | New |
| 36 | 1 | 1-155819544-C-CTA | | NA | 0.0009 | CTA | | 1.25E-10 | ?+ | 1.251E-10 | | New |
| 36 | 1 | 1-155819546-AAG-A | | NA | 0.0009 | A | | 1.59E-10 | ?+ | 1.592E-10 | | New |
| 37 | 1 | 1-155825397-AG-A | rs1348893 | GON4L | 0.0014 | A | 0.990467 | 1.71E-10 | ++ | 1.322E-07 | 0.7258 | New |
| 38 | 1 | 1-155877614-C-CT | rs1213636 | SYT11 | 0.0014 | CT | 0.342595 | 4.13E-19 | ++ | 3.477E-15 | | New |
| 38 | 1 | 1-155877639-C-T | rs9536581 | SYT11 | 0.0006 | T | 0.105511 | 2.44E-09 | -+ | 8.866E-08 | | New |
| 39 | 1 | 1-155878634-GAC-G | | NA | 0.0008 | G | | 5.36E-10 | ?+ | 5.363E-10 | | New |
| 40 | 1 | 1-155948467-TA-T | rs1247800 | ARHGEF2 | 0.0013 | TA | | 4.41E-10 | ?+ | 4.413E-10 | | New |
| 41 | 1 | 1-155955307-TAG-T | | NA | 0.0007 | TAG | | 3.18E-08 | ?+ | 3.179E-08 | | New |
| 42 | 1 | 1-155963917-GGT-G | | NA | 0.0014 | G | | 1.09E-09 | ?+ | 1.088E-09 | | New |
| 42 | 1 | 1-155963921-C-CAG | | NA | 0.0013 | CAG | | 1.78E-09 | ?+ | 1.777E-09 | | New |
| 43 | 1 | 1-15860102-G-A | rs8910654 | SPEN | 0.0016 | A | 0.992715 | 2.2E-11 | -+ | 0.000002692 | 0.1307 | New |
| 43 | 1 | 1-15860133-G-T | rs1015482 | SPEN | 0.0051 | T | 0.865415 | 2.18E-45 | ++ | 2.403E-30 | 0.116 | New |
| 43 | 1 | 1-15860146-C-T | rs9951453 | SPEN | 0.0033 | T | 0.901398 | 1.87E-26 | ++ | 2.196E-15 | | New |
| 44 | 1 | 1-15869308-GC-G | | NA | 0.0005 | G | | 1.84E-08 | ?+ | 1.843E-08 | | New |
| 45 | 1 | 1-15977137-T-A | rs1482849 | ZBTB17, SRARP | 0.0005 | A | 0.322022 | 7.93E-09 | -+ | 3.519E-07 | | New |
| 45 | 1 | 1-15977177-C-CAA | rs1471550 | ZBTB17, SRARP | 0.0013 | C | | 5.38E-10 | ?+ | 5.377E-10 | | New |
| 46 | 1 | 1-161348819-C-CAG | | NA | 0.0007 | C | | 9.94E-14 | ?+ | 9.94E-14 | | Old |
| 47 | 1 | 1-161372350-ATC-A | | NA | 0.0015 | A | | 1.53E-14 | ?+ | 1.531E-14 | | Old |
| 48 | 1 | 1-16382341-C-CCA | | NA | 0.0005 | CCA | | 7.08E-11 | ?+ | 7.081E-11 | | New |
| 49 | 1 | 1-16429296-C-CCCGG | rs1231827 | SPATA21 | 0.0014 | CCCGG | | 1.31E-10 | ?+ | 1.309E-10 | 0.4158 | New |
| 50 | 1 | 1-16516729-GC-G | rs7612763 | AL137802.3 | 0.0033 | G | 0.114071 | 7.42E-10 | +- | 0.07768 | 0.8192 | New |
| 51 | 1 | 1-16732342-G-A | rs4394648 | AL021920.1, AL021920.3 | 0.0068 | A | | 4.05E-09 | ?+ | 4.051E-09 | 0.6696 | New |

| | | | | | | | | | | | | |
|---|---|---|---|---|---|---|---|---|---|---|---|---|
| 52 | 1 | 1-17006395-C-CATGCACCACCAT | | NA | 0.0025 | C | | 4.9E-25 | ?+ | 4.901E-25 | | New |
| 53 | 1 | 1-17054484-C-CT | rs1281628 | SDHB | 0.0023 | C | | 1.66E-08 | ?+ | 1.662E-08 | 0.5208 | New |
| 54 | 1 | 1-171621577-G-T | rs5522232 | MYOCOS | 0.0011 | T | 0.790158 | 1.85E-13 | -+ | 8.537E-13 | | New |
| 54 | 1 | 1-171621582-G-A | rs1652576 | MYOCOS | 0.001 | G | | 7.35E-13 | ?+ | 7.348E-13 | | New |
| 55 | 1 | 1-173835689-C-CCT | rs1553202 | DARS2 | 0.0006 | C | | 3.1E-08 | ?+ | 3.096E-08 | | New |
| 56 | 1 | 1-17543895-GCCCA-G | | NA | 0.001 | GCCCA | | 2.25E-08 | ?+ | 2.246E-08 | | New |
| 57 | 1 | 1-17544491-C-T | rs1223058 | ARHGEF10L | 0.0015 | T | 0.112974 | 5.15E-13 | ++ | 3.833E-11 | | New |
| 57 | 1 | 1-17544499-G-T | rs2996647 | ARHGEF10L | 0.0837 | T | 1.52E-05 | 2.37E-16 | ++ | 7.192E-17 | 0.7419 | New |
| 58 | 1 | 1-17544542-ACCTG-A | | NA | 0.0006 | ACCTG | | 1.47E-10 | ?+ | 1.473E-10 | | New |
| 59 | 1 | 1-178078516-G-T | rs1658204 | CRYZL2P-SEC16B, RASAL2-AS1 | 0.001 | T | 0.809812 | 3.01E-13 | ++ | 0.000001617 | | New |
| 60 | 1 | 1-179070132-G-A | rs8899941 | FAM20B | 0.0013 | A | 0.688914 | 3.31E-11 | ++ | 3.653E-10 | 0.9853 | New |
| 61 | 1 | 1-179070191-ACC-A | rs1351651 | FAM20B | 0.001 | ACC | | 2.49E-08 | ?+ | 2.485E-08 | | New |
| 62 | 1 | 1-179174275-AG-A | | NA | 0.0007 | AG | | 3.76E-09 | ?+ | 3.764E-09 | | New |
| 63 | 1 | 1-197576732-C-T | | DENND1B | 0.0007 | C | | 9.26E-11 | ?+ | 9.256E-11 | | Old |
| 64 | 1 | 1-200258342-T-A | | LINC00862 | 0.0006 | A | | 3.07E-08 | ?+ | 3.074E-08 | | New |
| 65 | 1 | 1-200284721-C-CTG | | NA | 0.0009 | C | | 1.45E-08 | ?+ | 1.454E-08 | | New |
| 66 | 1 | 1-200345290-C-CGG | | NA | 0.0009 | CGG | | 2.28E-11 | ?+ | 2.282E-11 | | New |
| 66 | 1 | 1-200345291-ACC-A | | NA | 0.0008 | A | | 1.51E-10 | ?+ | 1.513E-10 | | New |
| 67 | 1 | 1-200386443-C-T | rs1929769 | AC104461.1, LINC00862 | 0.0014 | T | 0.343766 | 4.77E-08 | -+ | 0.05078 | | New |
| 68 | 1 | 1-202740096-GCA-G | rs8796859 | KDM5B | 0.0037 | G | | 1.51E-09 | ?+ | 1.506E-09 | | New |
| 69 | 1 | 1-203503088-T-A | rs1661417 | OPTC | 0.0007 | T | | 1.11E-09 | ?+ | 1.112E-09 | | New |

| | | | | | | | | | | | | |
|---|---|---|---|---|---|---|---|---|---|---|---|---|
| 70 | 1 | 1-203921617-AGG-A | | NA | 0.0003 | AGG | | 1.36E-08 | ?+ | 1.36E-08 | | New |
| 71 | 1 | 1-20463538-ACC-A | rs2051320 | LINC01141, AL139254.2 | 0.0118 | ACC | | 5.43E-15 | ?+ | 5.424E-15 | | New |
| 72 | 1 | 1-20647193-AC-A | | NA | 0.0003 | A | | 1.37E-08 | ?+ | 1.374E-08 | | New |
| 73 | 1 | 1-20796590-C-CCT | rs1423973 | HP1BP3, EIF4G3 | 0.0062 | CCT | | 1.85E-12 | ?+ | 1.845E-12 | | New |
| 74 | 1 | 1-210976986-G-C | rs5728931 | AL590132.1, KCNH1 | 0.0006 | G | | 4.53E-08 | ?+ | 4.53E-08 | 0.581 | New |
| 75 | 1 | 1-214897967-G-A | | CENPF, AC099563.2 | 0.0011 | A | | 3.77E-15 | ?+ | 3.767E-15 | | New |
| 76 | 1 | 1-218455409-G-C | rs1269307 | TGFB2, C1orf143 | 0.0006 | C | 0.209681 | 2.23E-09 | -+ | 0.000001693 | 0.9816 | New |
| 77 | 1 | 1-218455927-G-A | rs1901801 | TGFB2, C1orf143 | 0.0019 | A | 0.163756 | 2.23E-10 | ++ | 8.967E-11 | 0.8605 | New |
| 78 | 1 | 1-218561848-G-C | rs1324865 | RF00012, LINC01710 | 0.0039 | C | 0.372117 | 2.65E-32 | -+ | 2.009E-22 | 0.6764 | New |
| 79 | 1 | 1-220048846-C-CAA | | NA | 0.0006 | CAA | | 1.5E-10 | ?+ | 1.501E-10 | | New |
| 79 | 1 | 1-220048851-T-A | | EPRS, BPNT1 | 0.0004 | A | | 7.16E-09 | ?+ | 7.161E-09 | | New |
| 79 | 1 | 1-220048860-C-T | rs1264199 | EPRS, BPNT1 | 0.0004 | T | 0.6469 | 6.14E-09 | ++ | 1.391E-08 | | New |
| 80 | 1 | 1-220140384-TGA-T | rs1290134 | IARS2 | 0.0035 | T | | 2.69E-08 | ?+ | 2.693E-08 | 0.1619 | New |
| 81 | 1 | 1-22058633-C-CCACCA | | NA | 0.001 | CCACCA | | 1.07E-10 | ?+ | 1.075E-10 | | New |
| 82 | 1 | 1-223118344-G-T | rs1225356 | TLR5 | 0.0008 | T | 0.372023 | 9.8E-11 | -+ | 3.986E-07 | | New |
| 82 | 1 | 1-223118351-G-A | rs1214165 | TLR5 | 0.0005 | A | 0.256513 | 1.03E-08 | -+ | 0.0001138 | 0.2428 | New |
| 83 | 1 | 1-223119009-C-A | rs1264236 | TLR5 | 0.0015 | A | 0.970535 | 2.89E-14 | ++ | 7.218E-09 | 0.637 | New |
| 84 | 1 | 1-224206818-C-CCT | | NA | 0.0007 | CCT | | 1.35E-08 | ?+ | 1.352E-08 | | New |
| 85 | 1 | 1-22429164-C-CCT | | NA | 0.0005 | CCT | | 3.24E-08 | ?+ | 3.24E-08 | | New |
| 86 | 1 | 1-226142102-C-CCT | | NA | 0.0013 | CCT | | 4.08E-16 | ?+ | 4.08E-16 | | New |

| | | | | | | | | | | | | |
|---|---|---|---|---|---|---|---|---|---|---|---|---|
| 87 | 1 | 1-228892872-C-T | rs1657042 | RHOU, AL137793.1 | 0.0011 | C | | 3.29E-08 | ?+ | 3.29E-08 | | New |
| 88 | 1 | 1-241063951-AGGTG-A | | NA | 0.0005 | A | | 1.18E-11 | ?+ | 1.184E-11 | | New |
| 89 | 1 | 1-246376445-G-T | rs1368723 | SMYD3 | 0.0006 | T | | 5.79E-09 | ?+ | 5.787E-09 | | New |
| 90 | 1 | 1-24812865-C-CAT | rs1358228 | CLIC4 | 0.0018 | C | | 1.69E-16 | ?+ | 1.695E-16 | | New |
| 90 | 1 | 1-24812868-GCA-G | rs1261343 | CLIC4 | 0.0018 | GCA | | 6.34E-14 | ?+ | 6.342E-14 | | New |
| 91 | 1 | 1-25925787-AGTGC-A | | NA | 0.0005 | A | | 3.09E-10 | ?+ | 3.091E-10 | | New |
| 92 | 1 | 1-26108552-TG-T | rs1213491 | TRIM63, PDIK1L | 0.0016 | T | 0.447481 | 1.91E-10 | ++ | 3.979E-10 | 0.1473 | New |
| 93 | 1 | 1-26334705-ACC-A | | NA | 0.0013 | ACC | | 4.5E-11 | ?+ | 4.495E-11 | | New |
| 93 | 1 | 1-26334710-GTCAA-G | | NA | 0.0005 | G | | 9.13E-10 | ?+ | 9.132E-10 | | New |
| 94 | 1 | 1-26648341-C-CCT | rs2079977 | RF01210, AL512408.1 | 0.001 | C | | 1.09E-08 | ?+ | 1.085E-08 | 0.1683 | New |
| 95 | 1 | 1-26651469-AGC-A | | NA | 0.0008 | A | | 4.22E-10 | ?+ | 4.215E-10 | | New |
| 95 | 1 | 1-26651472-C-CTT | | NA | 0.0006 | CTT | | 1.55E-08 | ?+ | 1.552E-08 | | New |
| 96 | 1 | 1-27232615-C-CA | rs1210025 | AL590640.1 | 0.0029 | CA | | 9.46E-09 | ?+ | 9.463E-09 | | New |
| 97 | 1 | 1-27865762-ATT-A | rs1264039 | PPP1R8, THEMIS2 | 0.0014 | A | 0.184637 | 3.18E-08 | -+ | 0.003287 | 0.4593 | New |
| 98 | 1 | 1-28350749-GTA-G | | NA | 0.0007 | G | | 4.1E-10 | ?+ | 4.103E-10 | | New |
| 99 | 1 | 1-28391247-AAAT-A | rs5472605 | PHACTR4 | 0.006 | A | | 3.3E-09 | ?+ | 3.295E-09 | 0.07281 | New |
| 100 | 1 | 1-28649458-C-CAG | rs1408113 | AL360012.1, LINC01715, RNU11 | 0.0037 | CAG | | 3.4E-15 | ?+ | 3.398E-15 | | New |
| 100 | 1 | 1-28649461-GAT-G | rs1439737 | AL360012.1, LINC01715, RNU11 | 0.0031 | G | | 3.36E-14 | ?+ | 3.363E-14 | 0.5987 | New |
| 101 | 1 | 1-28649645-GGCA-G | | NA | 0.0008 | G | | 1.64E-14 | ?+ | 1.638E-14 | | New |
| 102 | 1 | 1-28715380-C-CCT | rs1421604 | GMEB1 | 0.0008 | CCT | | 3.47E-08 | ?+ | 3.474E-08 | | New |

| | | | | | | | | | | | | |
|---|---|---|---|---|---|---|---|---|---|---|---|---|
| 103 | 1 | 1-28733121-AT-A | | NA | 0.0004 | AT | | 3.39E-12 | ?+ | 3.389E-12 | | New |
| 104 | 1 | 1-28994406-C-CAA | | NA | 0.0012 | CAA | | 1.23E-08 | ?+ | 1.23E-08 | | New |
| 105 | 1 | 1-29175504-G-C | rs1252460 | SRSF4 | 0.0034 | C | 0.682523 | 5.22E-14 | -+ | 0.0002275 | 0.1117 | New |
| 105 | 1 | 1-29175505-G-C | rs9511675 | SRSF4 | 0.0034 | C | 0.682523 | 5.25E-12 | -+ | 0.001454 | | New |
| 106 | 1 | 1-31578988-T-C | rs1273608 | LINC01226 | 0.008 | C | | 1.58E-16 | ?+ | 1.58E-16 | | New |
| 107 | 1 | 1-35117555-C-CAG | | NA | 0.001 | C | | 3.86E-11 | ?+ | 3.856E-11 | | New |
| 108 | 1 | 1-35579988-C-CCT | | NA | 0.0008 | C | | 4.69E-09 | ?+ | 4.687E-09 | 0.8455 | New |
| 109 | 1 | 1-36102439-C-A | rs1643690 | COL8A2 | 0.0016 | A | 0.750459 | 1.18E-14 | ++ | 4.662E-10 | | New |
| 109 | 1 | 1-36102460-C-CTGGCTAATTTTTTCTATTT | | NA | 0.0009 | C | 0.252577 | 5.5E-14 | -+ | 0.00000104 | | New |
| 110 | 1 | 1-36571154-C-CT | rs2012086 | CSF3R, AL596257.1 | 0.0013 | C | 0.486388 | 2.47E-09 | ++ | 1.986E-09 | | New |
| 111 | 1 | 1-37598789-C-CTCAA | | NA | 0 | C | | 2.04E-08 | ?+ | 2.037E-08 | | New |
| 112 | 1 | 1-37884763-AAT-A | rs1178610 | INPP5B | 0.0008 | AAT | | 4.6E-09 | ?+ | 4.601E-09 | 0.2432 | New |
| 112 | 1 | 1-37884766-ATG-A | 1-3788476 | NA | 0.0007 | A | | 4.22E-09 | ?+ | 4.216E-09 | | New |
| 113 | 1 | 1-37935848-C-CCA | rs1488544 | INPP5B | 0.0054 | CCA | 0.518856 | 1.27E-27 | -+ | 1.63E-23 | | New |
| 114 | 1 | 1-37941708-GT-G | rs1302759 | INPP5B | 0.0022 | GT | 0.767317 | 1.18E-08 | ++ | 0.00000308 | 0.7676 | New |
| 115 | 1 | 1-3798817-C-CAAA | | NA | 0.0008 | CAAA | | 3.43E-15 | ?+ | 3.434E-15 | | New |
| 116 | 1 | 1-38869984-T-A | rs1642424 | AL139260.1, AL139260.2 | 0.0008 | A | 0.745338 | 6.08E-15 | -+ | 2.112E-12 | | New |
| 117 | 1 | 1-39067815-G-T | | NDUFS5, MACF1 | 1E-04 | G | 0.844572 | 3.03E-09 | ++ | 1.045E-08 | | New |
| 118 | 1 | 1-39179844-G-A | rs9033917 | MACF1 | 0.0017 | A | 0.82374 | 5.74E-23 | ++ | 7.818E-18 | | New |
| 119 | 1 | 1-39504208-GAT-G | rs5485321 | BMP8A | 0.0034 | G | 0.781916 | 1.42E-19 | ++ | 3.523E-11 | 0.1694 | New |
| 120 | 1 | 1-39533298-C-T | rs1300889 | BMP8A, PABPC4 | 0.0021 | C | 0.943951 | 1.28E-10 | ++ | 0.00003494 | | New |
| 120 | 1 | 1-39533299-C-A | rs5740013 | BMP8A, PABPC4 | 0.0021 | A | 0.884992 | 7.91E-10 | ++ | 0.00005984 | | New |
| 121 | 1 | 1-39579706-C-CGTG | | NA | 0.0006 | C | | 7.97E-09 | ?+ | 7.974E-09 | | New |
| 122 | 1 | 1-39606520-C-T | rs1016657 | PABPC4, HEYL | 0.0011 | T | 0.775827 | 1.16E-08 | -+ | 1.054E-07 | 0.4177 | New |
| 122 | 1 | 1-39606527-C-A | rs9913455 | PABPC4, HEYL | 0.0014 | C | 0.929636 | 1.85E-11 | -+ | 1.65E-10 | 0.06786 | New |

| | | | | | | | | | | | | |
|---|---|---|---|---|---|---|---|---|---|---|---|---|
| 122 | 1 | 1-39606529-C-T | rs5685280 | PABPC4, HEYL | 0.0009 | T | | 1.9E-11 | ?+ | 1.898E-11 | 0.01975 | New |
| 123 | 1 | 1-39993465-TTC-T | | NA | 0.0011 | T | | 3.34E-09 | ?+ | 3.338E-09 | | New |
| 124 | 1 | 1-40590743-C-CAA | | NA | 0.0005 | CAA | | 8.47E-09 | ?+ | 8.471E-09 | | New |
| 124 | 1 | 1-40590746-TATCG-T | | NA | 0.0003 | T | | 3.29E-08 | ?+ | 3.287E-08 | | New |
| 125 | 1 | 1-42614311-ACCT-A | | NA | 0.0021 | A | | 1.45E-14 | ?+ | 1.448E-14 | | New |
| 126 | 1 | 1-43248985-C-A | | CFAP57 | 0.0018 | A | 0.789124 | 2.94E-15 | ++ | 9.166E-12 | | New |
| 127 | 1 | 1-44148272-C-CTACT | rs1553173 | KLF18, RF00019 | 0.0007 | CTACT | | 4.14E-08 | ?+ | 4.145E-08 | | New |
| 128 | 1 | 1-44952753-C-T | rs9872708 | EIF2B3 | 0.0041 | C | 0.125232 | 3.51E-26 | ++ | 2.898E-16 | 0.4301 | New |
| 128 | 1 | 1-44952782-G-T | rs9462441 | EIF2B3 | 0.0064 | G | 0.974855 | 2.69E-35 | ++ | 5.092E-18 | | New |
| 129 | 1 | 1-46311457-GC-G | | NA | 0.0033 | G | | 1.41E-10 | ?+ | 1.405E-10 | | New |
| 130 | 1 | 1-46323003-ACC-A | | NA | 0.0012 | ACC | | 3.65E-10 | ?+ | 3.645E-10 | | New |
| 131 | 1 | 1-46337062-C-A | rs9032534 | UQCRH, NSUN4 | 0.0024 | A | 0.861843 | 8.51E-20 | ++ | 6.43E-13 | | New |
| 131 | 1 | 1-46337077-G-A | rs1661880 | UQCRH, NSUN4 | 0.003 | A | 0.650541 | 8.98E-31 | ++ | 3.967E-21 | | New |
| 131 | 1 | 1-46337088-G-A | rs1035837 | UQCRH, NSUN4 | 0.0033 | A | 0.984205 | 7.11E-29 | -+ | 5.953E-19 | | New |
| 132 | 1 | 1-49135071-G-T | rs1377249 | AC099788.1 | 0.0029 | G | 0.588821 | 6.87E-16 | ++ | 6.018E-07 | | New |
| 133 | 1 | 1-52623572-C-T | | GPX7, SHISAL2A | 0.0001 | T | | 1.16E-08 | ?+ | 1.165E-08 | | New |
| 134 | 1 | 1-52910506-C-T | rs1376370 | ECHDC2 | 0.0005 | C | | 1.31E-09 | ?+ | 1.306E-09 | | New |
| 134 | 1 | 1-52910512-GCAC-G | | NA | 0.0006 | G | | 3.37E-09 | ?+ | 3.368E-09 | | New |
| 135 | 1 | 1-54466273-ACC-A | | NA | 0.0003 | ACC | | 4.74E-10 | ?+ | 4.74E-10 | | New |
| 135 | 1 | 1-54466274-TTG-T | | NA | 0.0005 | T | | 3.69E-14 | ?+ | 3.693E-14 | | New |
| 136 | 1 | 1-54504924-C-CA | rs8894876 | AL035415.1, AC099796.3 | 0.0008 | C | | 1.51E-08 | ?+ | 1.51E-08 | | New |
| 137 | 1 | 1-54505400-C-CA | rs1463170 | AL035415.1, AC099796.3 | 0.0016 | CA | 0.11527 | 2.2E-08 | ++ | 8.534E-07 | 0.3789 | New |
| 138 | 1 | 1-5674860-GAAT-G | | NA | 0.0026 | G | | 2.83E-08 | ?+ | 2.828E-08 | 0.5474 | New |
| 139 | 1 | 1-5884481-AGC-A | rs1386709 | NPHP4 | 0.0025 | A | | 1.36E-08 | ?+ | 1.363E-08 | | New |

| | | | | | | | | | | | | |
|---|---|---|---|---|---|---|---|---|---|---|---|---|
| 140 | 1 | 1-61155454-ACC-A | rs1182012 | NFIA | 0.0034 | ACC | | 4.31E-12 | ?+ | 4.306E-12 | 0.3071 | New |
| 141 | 1 | 1-63237607-C-CA | | NA | 0.0007 | CA | | 1.78E-09 | ?+ | 1.781E-09 | | New |
| 142 | 1 | 1-7197400-G-A | rs8861184 | CAMTA1 | 0.0041 | G | 0.552694 | 2.24E-18 | ++ | 0.000000081 | | New |
| 143 | 1 | 1-85367115-G-A | rs1431252 | AL078459.1 | 0.0003 | A | | 4.81E-09 | ?+ | 4.807E-09 | | New |
| 144 | 1 | 1-86848155-T-A | rs1658217 | AL355981.1, SELENOF | 0.0005 | T | | 9.51E-10 | ?+ | 9.512E-10 | | New |
| 145 | 1 | 1-8774498-C-CAT | | NA | 0.0012 | C | | 4.74E-08 | ?+ | 4.738E-08 | | New |
| 145 | 1 | 1-8774520-AT-A | rs2124551 | NA | 0.0005 | AT | 0.027594 | 2.49E-12 | ++ | 2.957E-11 | | New |
| 146 | 1 | 1-88806458-C-CCG | | NA | 0.001 | CCG | | 1.52E-08 | ?+ | 1.522E-08 | | New |
| 147 | 1 | 1-90050171-AGG-A | | NA | 0.0002 | AGG | | 6.99E-10 | ?+ | 6.987E-10 | | New |
| 148 | 1 | 1-9158215-G-A | rs1409228 | MIR34AHG | 0.0008 | G | 0.299098 | 4.41E-10 | ++ | 4.433E-10 | 0.9312 | New |
| 148 | 1 | 1-9158256-AG-A | rs1359100 | MIR34AHG | 0.0005 | A | | 2.92E-09 | ?+ | 2.916E-09 | | New |
| 149 | 1 | 1-9623342-C-CCT | rs1200379 | TMEM201, PIK3CD | 0.003 | CCT | | 1.73E-25 | ?+ | 1.733E-25 | | New |
| 149 | 1 | 1-9623353-G-A | rs1644420 | TMEM201, PIK3CD | 0.0005 | A | | 4.55E-08 | ?+ | 4.546E-08 | | New |
| 150 | 1 | 1-9702141-T-A | rs1646657 | PIK3CD | 0.0017 | T | | 6.5E-18 | ?+ | 6.504E-18 | | New |
| 150 | 1 | 1-9702143-T-A | rs1646657 | PIK3CD | 0.0017 | T | | 3.16E-15 | ?+ | 3.154E-15 | | New |
| 151 | 1 | 1-9830445-C-T | rs1112149 | AL357140.3, CTNNBIP1 | 0.3177 | C | | 2.58E-13 | ?+ | 2.576E-13 | 0.9323 | New |
| 152 | 2 | 2-101207215-ATT-A | | NA | 0.0011 | A | | 6.71E-12 | ?+ | 6.711E-12 | | New |
| 153 | 2 | 2-101578677-T-A | rs1052317 | AC093894.2, MAP4K4 | 0.0011 | T | | 3.71E-08 | ?+ | 3.706E-08 | | New |
| 154 | 2 | 2-106888365-T-A | rs1355697 | ST6GAL2, AC005040.2 | 0.0008 | T | 0.910539 | 7.3E-11 | ++ | 2.153E-07 | | New |
| 154 | 2 | 2-106888375-G-T | rs1208010 | ST6GAL2, AC005040.2 | 0.0005 | G | | 2.52E-08 | ?+ | 2.523E-08 | | New |
| 155 | 2 | 2-109658261-G-C | rs1700662 | SOWAHC, AC074387.1 | 0.0021 | C | 0.330235 | 1.84E-12 | ++ | 6.434E-07 | | New |

| | | | | | | | | | | | | |
|---|---|---|---|---|---|---|---|---|---|---|---|---|
| 156 | 2 | 2-11052875-TCG-T | rs1455283 | KCNF1, AC062028.1 | 0.0009 | TCG | | 1.68E-08 | ?+ | 1.676E-08 | 0.99 | Old |
| 156 | 2 | 2-11052877-GCA-G | rs1173935 | KCNF1, AC062028.1 | 0.0008 | G | | 2E-08 | ?+ | 1.996E-08 | | Old |
| 157 | 2 | 2-111295877-ATTTTTT-A | | MIR4435-2HG | 0.0791 | A | | 1.17E-12 | ?- | 1.167E-12 | | New |
| 158 | 2 | 2-111880255-C-T | rs1465759 | ANAPC1 | 0.0008 | T | 0.006346 | 3.03E-09 | ++ | 6.677E-11 | | New |
| 159 | 2 | 2-113864927-TA-T | | NA | 0.0023 | T | | 4.84E-13 | ?+ | 4.843E-13 | | New |
| 160 | 2 | 2-118093497-AGATGATGATGAT-A | | INSIG2 | 0.3283 | A | | 2.98E-14 | ?- | 2.979E-14 | | New |
| 161 | 2 | 2-120197492-ACT-A | rs1408747 | AC012363.1 | 0.0035 | ACT | | 1.02E-18 | ?+ | 1.015E-18 | 0.151 | New |
| 162 | 2 | 2-126871297-G-A | rs1679763 | RNU6-675P, TEX51 | 0.0009 | G | 0.539144 | 2.48E-15 | -+ | 5.943E-12 | | Old |
| 162 | 2 | 2-126871303-G-T | | RNU6-675P, TEX51 | 0.0009 | T | 0.770985 | 9.19E-10 | -+ | 9.835E-08 | | Old |
| 163 | 2 | 2-127858638-C-CTG | rs1274020 | POLR2D | 0.0026 | CTG | | 2.36E-15 | ?+ | 2.357E-15 | 0.2504 | New |
| 163 | 2 | 2-127858655-G-C | rs1690388 | POLR2D | 0.0009 | C | | 8.83E-11 | ?+ | 8.827E-11 | 0.1055 | New |
| 164 | 2 | 2-128059121-C-CTT | | SAP130, RF00019 | 0.0018 | CTT | | 1.65E-12 | ?+ | 1.647E-12 | | New |
| 165 | 2 | 2-128082350-GCACA-G | | RF00019, UGGT1 | 0.0021 | G | | 7.5E-09 | ?+ | 7.501E-09 | | New |
| 166 | 2 | 2-130469139-C-CA | | POTEI | 0.03 | CA | | 2.28E-11 | ?+ | 2.275E-11 | | New |
| 167 | 2 | 2-130863966-C-CTA | | NA | 0.0021 | CTA | | 7.07E-17 | ?+ | 7.076E-17 | | New |
| 168 | 2 | 2-131634219-G-C | rs1235862 | RNU6-617P, LINC01087 | 0.0046 | G | 0.690714 | 5.03E-09 | -+ | 0.03352 | 0.5165 | New |
| 169 | 2 | 2-131634249-C-T | rs1683744 | RNU6-617P, LINC01087 | 0.0033 | C | 0.446437 | 2.73E-08 | ++ | 0.006057 | | New |

| | | | | | | | | | | | | |
|---|---|---|---|---|---|---|---|---|---|---|---|---|
| 170 | 2 | 2-131634722-G-C | rs1001830 | RNU6-617P, LINC01087 | 0.0009 | G | 0.167768 | 1.5E-10 | ++ | 1.596E-10 | | New |
| 171 | 2 | 2-131636079-C-A | rs1301482 | LINC01087 | 0.0011 | A | 0.431759 | 4.03E-08 | ++ | 1.108E-07 | | New |
| 172 | 2 | 2-131756355-G-A | rs1305401 | C2orf27A | 0.0041 | G | | 9.49E-09 | ?+ | 9.487E-09 | | New |
| 173 | 2 | 2-132075422-T-A | rs1997153 | AC093787.2, ANKRD30BL | 0.0007 | T | 0.477468 | 3.15E-11 | ++ | 1.694E-07 | | New |
| 174 | 2 | 2-133308914-G-T | rs1681023 | NCKAP5 | 0.0012 | T | 0.527445 | 6.25E-12 | ++ | 1.423E-09 | | New |
| 174 | 2 | 2-133308921-G-A | rs1681024 | NCKAP5 | 0.0009 | G | 0.609481 | 1.04E-11 | ++ | 8.118E-09 | | New |
| 174 | 2 | 2-133308945-C-T | rs1338208 | NCKAP5 | 0.0011 | T | 0.960418 | 5.64E-15 | -+ | 4.207E-08 | | New |
| 174 | 2 | 2-133308946-G-A | rs1475548 | NCKAP5 | 0.0014 | G | 0.931604 | 3.95E-20 | -+ | 8.106E-12 | | New |
| 174 | 2 | 2-133308953-G-A | rs1041743 | NCKAP5 | 0.0013 | G | 0.946634 | 1.55E-15 | -+ | 8.234E-09 | | New |
| 175 | 2 | 2-134152483-G-C | rs1025240 | MGAT5 | 0.0006 | G | 0.563896 | 4.07E-08 | -+ | 0.00000251 | | Old |
| 176 | 2 | 2-134710815-C-T | rs1016498 | TMEM163 | 0.3221 | C | | 4.4E-08 | ?+ | 4.401E-08 | 0.4644 | Old |
| 177 | 2 | 2-135470542-C-CT | | NA | 0.0011 | C | | 6.69E-14 | ?+ | 6.694E-14 | | New |
| 178 | 2 | 2-151415128-ACC-A | rs1296731 | RIF1 | 0.0023 | ACC | | 8.08E-14 | ?+ | 8.079E-14 | 0.1995 | New |
| 179 | 2 | 2-156856973-T-A | rs1249378 | AC096589.1, AC096589.2 | 0.0003 | A | 0.448627 | 3.41E-10 | -+ | 0.000000481 | | New |
| 180 | 2 | 2-157549149-T-A | | ACVR1C | 0.0002 | A | | 3.5E-09 | ?+ | 3.504E-09 | | New |
| 180 | 2 | 2-157549175-GA-G | | ACVR1C | 0.0001 | G | | 3.02E-09 | ?+ | 3.024E-09 | | New |
| 181 | 2 | 2-169608315-G-C | rs9058356 | PPIG | 0.0029 | C | 0.32132 | 1.41E-20 | ++ | 5.098E-20 | | New |
| 182 | 2 | 2-174296329-TA-T | rs1242963 | RF00019, LINC01305 | 0.0021 | TA | 0.506122 | 6.78E-23 | ++ | 1.771E-15 | | New |
| 183 | 2 | 2-179792562-G-A | rs3679217 | ZNF385B | 0.0006 | G | | 3.76E-08 | ?+ | 3.765E-08 | | New |
| 183 | 2 | 2-179792566-G-A | rs1296008 | ZNF385B | 0.0006 | G | | 4.46E-10 | ?+ | 4.459E-10 | | New |
| 184 | 2 | 2-183479004-C-CTG | | AC021851.2 | 0.0019 | C | | 5.77E-11 | ?+ | 5.768E-11 | | New |
| 185 | 2 | 2-198215952-C-CCT | rs1404822 | AC020719.1, LINC01923 | 0.002 | CCT | | 1.39E-11 | ?+ | 1.385E-11 | | New |

| | | | | | | | | | | | | |
|---|---|---|---|---|---|---|---|---|---|---|---|---|
| 186 | 2 | 2-202003854-C-CCT | | NA | 0.0003 | C | | 3.38E-09 | ?+ | 3.382E-09 | | New |
| 187 | 2 | 2-202294096-C-CCT | | NA | 0.0012 | C | | 2.61E-09 | ?+ | 2.614E-09 | | Old |
| 188 | 2 | 2-202567965-C-CGT | | NA | 0.0005 | CGT | | 2.45E-08 | ?+ | 2.446E-08 | | Old |
| 189 | 2 | 2-202813151-C-T | | ICA1L | 0.0004 | C | | 2.83E-13 | ?+ | 2.83E-13 | | Old |
| 190 | 2 | 2-202861728-AAT-A | | NA | 0.0004 | AAT | | 2.83E-08 | ?+ | 2.827E-08 | | Old |
| 191 | 2 | 2-205992418-ATGG-A | | NA | 0.0004 | ATGG | | 5.67E-10 | ?+ | 5.667E-10 | | New |
| 191 | 2 | 2-205992420-AGGG-A | | NA | 0.0002 | A | | 4.47E-09 | ?+ | 4.47E-09 | | New |
| 192 | 2 | 2-206050883-G-C | rs1575851 | INO80D | 0.0007 | C | 0.802356 | 4.28E-13 | -+ | 1.175E-10 | | New |
| 193 | 2 | 2-213584240-TA-T | rs1385628 | SPAG16 | 0.0024 | TA | 0.7376 | 5.92E-20 | -+ | 9.462E-19 | | New |
| 194 | 2 | 2-219315661-G-A | rs1016783 | PTPRN, RESP18 | 0.0084 | A | 0.545234 | 3.58E-10 | ++ | 0.006004 | | New |
| 194 | 2 | 2-219315696-T-A | rs9369824 | PTPRN, RESP18 | 0.0017 | A | 0.69334 | 8.09E-11 | ++ | 0.00001432 | | New |
| 195 | 2 | 2-219581029-AGGTG-A | | NA | 0.0006 | A | | 1.22E-08 | ?+ | 1.216E-08 | | New |
| 195 | 2 | 2-219581037-AGGTG-A | | NA | 0.0007 | AGGTG | | 1.91E-12 | ?+ | 1.909E-12 | | New |
| 196 | 2 | 2-221892456-A-AG | rs7105036 | AC068489.1, PAX3 | 0.0079 | AG | 0.050605 | 4.63E-09 | -+ | 1.272E-07 | | New |
| 197 | 2 | 2-223825837-G-T | rs1388440 | AC093884.1, AP1S3 | 0.0013 | T | 0.238963 | 3.5E-19 | -+ | 1.161E-07 | | New |
| 198 | 2 | 2-224529423-C-A | rs1219833 | CUL3 | 0.0009 | A | | 4.38E-10 | ?+ | 4.379E-10 | | New |
| 199 | 2 | 2-224755811-C-CCT | rs1553553 | CCDC195, DOCK10 | 0.0026 | CCT | | 1.43E-08 | ?+ | 1.432E-08 | | New |
| 200 | 2 | 2-231440470-ATG-A | rs1264652 | B3GNT7, AC017104.3 | 0.0015 | ATG | | 1.26E-16 | ?+ | 1.255E-16 | 0.5342 | New |
| 200 | 2 | 2-231440472-ACG-A | rs1473940 | B3GNT7, AC017104.3 | 0.001 | A | | 1.31E-09 | ?+ | 1.308E-09 | 0.4514 | New |
| 201 | 2 | 2-231501097-G-C | | RNU2-22P | 0.0004 | C | | 1.32E-09 | ?+ | 1.321E-09 | | New |
| 202 | 2 | 2-231565270-G-A | rs1360681 | NMUR1, AC104634.2 | 0.0006 | A | | 3.38E-09 | ?+ | 3.381E-09 | 0.825 | New |

| | | | | | | | | | | | | |
|---|---|---|---|---|---|---|---|---|---|---|---|---|
| 202 | 2 | 2-231565277-G-A | rs1308706 | NMUR1, AC104634.2 | 0.0006 | A | 0.618733 | 9.46E-11 | ++ | 1.237E-08 | 0.5981 | New |
| 202 | 2 | 2-231565279-G-C | rs1375871 | NMUR1, AC104634.2 | 0.0008 | C | 0.508329 | 7.88E-16 | ++ | 9.499E-14 | 0.411 | New |
| 203 | 2 | 2-234316318-G-A | rs5694811 | AC122134.1, AC097713.3 | 0.0009 | A | 0.197389 | 1.09E-11 | -+ | 1.472E-08 | 0.1614 | New |
| 204 | 2 | 2-236272485-G-A | rs1574680 | AC019068.1 | 0.0012 | A | 0.894413 | 3.55E-08 | ++ | 0.00001058 | | New |
| 205 | 2 | 2-238119146-G-GAA | rs1422295 | ESPNL | 0.0143 | G | | 1.6E-09 | ?+ | 1.595E-09 | | New |
| 206 | 2 | 2-238794552-C-T | rs1469424 | AC145625.1, TWIST2 | 0.0009 | T | | 1.09E-11 | ?+ | 1.09E-11 | | New |
| 207 | 2 | 2-239815908-GA-G | rs1231605 | AC093802.1 | 0.0069 | GA | | 1.22E-10 | ?+ | 1.218E-10 | | New |
| 208 | 2 | 2-241488609-G-C | rs9383319 | FARP2 | 0.0031 | G | 0.717588 | 1.67E-44 | ++ | 2.178E-42 | | New |
| 209 | 2 | 2-24152050-G-T | rs7623459 | AC008073.3, FAM228B | 0.0001 | T | | 4.32E-08 | ?+ | 4.316E-08 | | New |
| 210 | 2 | 2-25534570-C-CT | rs1295517 | DTNB | 0.0035 | C | | 7.17E-11 | ?+ | 7.166E-11 | | Old |
| 211 | 2 | 2-26010180-T-A | rs1665200 | KIF3C, RAB10 | 0.0007 | A | 0.752837 | 4.56E-08 | -+ | 5.085E-07 | | New |
| 212 | 2 | 2-27126918-A-G | rs1261479 | ABHD1 | 0.0165 | G | 0.251618 | 4.73E-19 | ++ | 2.225E-14 | 0.7722 | New |
| 213 | 2 | 2-27141043-T-A | | PRR30, TCF23 | 0.0003 | A | | 2.48E-08 | ?+ | 2.476E-08 | | New |
| 214 | 2 | 2-27273508-GAT-G | rs1259975 | SLC30A3 | 0.0058 | G | | 2.09E-11 | ?+ | 2.09E-11 | 0.0362 | New |
| 215 | 2 | 2-27511478-C-CCT | | NA | 0.001 | C | | 1.47E-10 | ?+ | 1.466E-10 | | New |
| 216 | 2 | 2-33306351-C-CTT | rs1195724 | LTBP1 | 0.0009 | C | | 3.71E-09 | ?+ | 3.714E-09 | 0.7061 | New |
| 217 | 2 | 2-34321033-C-CA | rs1465685 | LINC01320 | 0.0036 | C | | 1.08E-10 | ?+ | 1.082E-10 | 0.7088 | New |
| 218 | 2 | 2-38696978-G-T | rs1340901 | GALM | 0.0003 | T | 0.867679 | 2.54E-09 | ++ | 1.056E-08 | | New |
| 219 | 2 | 2-39089972-C-CAAAAAAAAAAAA | | NA | 0.0007 | C | | 3.23E-11 | ?+ | 3.226E-11 | | New |
| 220 | 2 | 2-39126426-AT-A | | NA | 0.0017 | A | | 7.9E-11 | ?+ | 7.905E-11 | | New |
| 221 | 2 | 2-46273971-C-CAAAGAAAGA | | NA | 0.0007 | CAAAGAAAGA | | 1.98E-09 | ?+ | 1.977E-09 | | New |

| | | | | | | | | | | | | |
|---|---|---|---|---|---|---|---|---|---|---|---|---|
| 222 | 2 | 2-47353179-AT-A | rs1344764 | EPCAM | 0.001 | A | | 1.06E-10 | ?+ | 1.058E-10 | | New |
| 223 | 2 | 2-47768888-TGG-T | rs1321375 | MSH6 | 0.0014 | TGG | | 2.16E-09 | ?+ | 2.158E-09 | | New |
| 224 | 2 | 2-52905737-G-A | rs9702552 | AC010967.1 | 0.0007 | A | 0.742768 | 2.9E-11 | -+ | 3.806E-07 | 0.9323 | New |
| 225 | 2 | 2-53970819-C-CGAA | rs1390937 | PSME4 | 0.0064 | C | | 2.31E-12 | ?+ | 2.312E-12 | | New |
| 226 | 2 | 2-53978264-ACTT-A | | NA | 0.0006 | ACTT | | 1.16E-08 | ?+ | 1.162E-08 | | New |
| 227 | 2 | 2-54004201-G-T | | ACYP2 | 0.0003 | T | | 4.07E-08 | ?+ | 4.073E-08 | | New |
| 228 | 2 | 2-61148337-C-CAA | | NA | 0.0003 | CAA | | 2.49E-08 | ?+ | 2.488E-08 | | New |
| 229 | 2 | 2-62167572-ATT-A | rs1394091 | COMMD1, B3GNT2 | 0.0018 | A | | 8.19E-12 | ?+ | 8.188E-12 | 0.1757 | New |
| 230 | 2 | 2-6420447-C-CATGAGGTCAGGAGATTGAG | rs1241385 | LINC01247, LINC01824 | 0.0052 | C | | 1.36E-08 | ?+ | 1.364E-08 | | New |
| 231 | 2 | 2-65157396-TA-T | rs1233069 | RAB1A, RF00090 | 0.0008 | TA | | 6.16E-10 | ?+ | 6.16E-10 | 0.299 | Old |
| 232 | 2 | 2-65157939-T-A | rs1035885 | RF00090 | 0.0046 | T | 0.268906 | 1.97E-13 | -+ | 0.001583 | 0.9227 | Old |
| 233 | 2 | 2-65168546-C-CCT | rs1395183 | RF00090, ACTR2 | 0.0042 | CCT | | 4.06E-10 | ?+ | 4.063E-10 | 0.07489 | Old |
| 234 | 2 | 2-65280182-ACC-A | | ACTR2, SPRED2 | 0.0014 | ACC | | 9E-10 | ?+ | 9.004E-10 | | Old |
| 235 | 2 | 2-65446107-C-A | rs9446597 | AC007389.1, AC012370.2 | 0.0014 | A | | 2.54E-21 | ?+ | 2.537E-21 | | Old |
| 235 | 2 | 2-65446110-G-C | rs1669197 | AC007389.1, AC012370.2 | 0.0012 | G | | 5.69E-16 | ?+ | 5.687E-16 | | Old |
| 236 | 2 | 2-70188373-T-A | rs1265024 | C2orf42 | 0.0013 | A | 0.846904 | 1.11E-14 | ++ | 9.119E-11 | 0.9363 | New |
| 237 | 2 | 2-79011165-C-CT | rs1439643 | AC092604.1, REG3G | 0.0015 | C | | 2.39E-12 | ?+ | 2.385E-12 | | Old |
| 238 | 2 | 2-79818457-C-CAT | | NA | 0.0008 | CAT | | 1.88E-10 | ?+ | 1.884E-10 | | New |
| 239 | 2 | 2-85532753-C-CACCG | | NA | 0.0019 | CACCG | | 4.04E-13 | ?+ | 4.035E-13 | | New |
| 240 | 2 | 2-86191352-ATT-A | | NA | 0.0009 | ATT | | 1.12E-09 | ?+ | 1.122E-09 | | New |
| 241 | 2 | 2-86191352-ATTAAT-A | | IMMT | 0.0006 | ATTAAT | | 8.72E-09 | ?+ | 8.725E-09 | | New |
| 242 | 2 | 2-87798773-G-C | rs1477485 | RGPD2 | 0.0028 | C | 0.165882 | 1.23E-09 | -+ | 0.08589 | | New |

| | | | | | | | | | | | | |
|---|---|---|---|---|---|---|---|---|---|---|---|---|
| 243 | 2 | 2-88005024-G-A | rs1205506 | RGPD2, RNU2-63P | 0.001 | G | 0.714893 | 2.18E-12 | ++ | 3.082E-12 | 0.5392 | New |
| 243 | 2 | 2-88005034-T-A | rs1232324 | RGPD2, RNU2-63P | 0.0009 | T | | 3.7E-12 | ?+ | 3.699E-12 | | New |
| 243 | 2 | 2-88005040-C-T | rs1479717 | RGPD2, RNU2-63P | 0.0007 | T | | 3.1E-16 | ?+ | 3.103E-16 | 0.3969 | New |
| 244 | 2 | 2-88005607-C-CA | | RGPD2, RNU2-63P | 0.0005 | C | | 7.6E-09 | ?+ | 7.601E-09 | 0.4046 | New |
| 245 | 2 | 2-89636593-GA-G | | RF00001, IGKV2D-40 | 0.4534 | GA | | 3.98E-09 | ?- | 3.98E-09 | | New |
| 245 | 2 | 2-89636593-GAAAA-G | | RF00001, IGKV2D-40 | 0.3313 | G | | 2.66E-27 | ?+ | 2.659E-27 | | New |
| 246 | 2 | 2-90331433-G-C | rs1221908 | IGKV3D-7, AC233263.6 | 0.002 | G | | 2.82E-09 | ?+ | 2.823E-09 | | New |
| 247 | 2 | 2-90396303-T-C | rs6214198 | AC233263.6, AC233266.2 | 0.0054 | C | | 4.07E-11 | ?- | 4.073E-11 | | New |
| 248 | 2 | 2-94499647-C-A | rs1674905 | NONE, AL845331.2 | 0.0007 | A | 0.40263 | 1.67E-09 | ++ | 7.753E-08 | 0.4255 | New |
| 249 | 2 | 2-9503812-TG-T | rs1318606 | AC080162.1 | 0.0004 | T | | 4.69E-10 | ?+ | 4.689E-10 | | Old |
| 250 | 2 | 2-99650376-G-T | rs5663589 | AFF3 | 0.0007 | G | 0.59274 | 1.9E-14 | ++ | 4.03E-14 | | New |
| 251 | 3 | 3-10030681-C-CCT | rs1484030 | FANCD2 | 0.0035 | C | | 2.14E-27 | ?+ | 2.136E-27 | | New |
| 252 | 3 | 3-10102824-C-CA | rs1227758 | FANCD2OS | 0.0006 | CA | | 7.12E-11 | ?+ | 7.122E-11 | | New |
| 253 | 3 | 3-10239992-C-CAA | rs1349978 | IRAK2 | 0.0057 | CAA | | 1.07E-17 | ?+ | 1.068E-17 | | New |
| 254 | 3 | 3-103419699-GT-G | rs1707023 | RNU1-43P, MIR548AB | 0.0043 | G | 0.094139 | 1.66E-31 | ++ | 7.12E-20 | | New |
| 255 | 3 | 3-109285911-C-CGT | | NA | 0.0009 | CGT | | 9.05E-09 | ?+ | 9.05E-09 | | New |

| | | | | | | | | | | | | |
|---|---|---|---|---|---|---|---|---|---|---|---|---|
| 256 | 3 | 3-11166550-C-CG | rs2020708 | HRH1 | 0.0009 | CG | 0.677911 | 3.01E-08 | -+ | 0.00001256 | | New |
| 257 | 3 | 3-11823413-G-A | | TAMM41 | 0.0063 | A | 0.285145 | 2.94E-14 | ++ | 0.00004888 | 0.04224 | New |
| 258 | 3 | 3-119480415-ATT-A | | NA | 0.0022 | ATT | | 5.25E-15 | ?+ | 5.253E-15 | 0.2544 | New |
| 259 | 3 | 3-121791872-C-CCT | | IQCB1 | 0.0012 | C | | 1.89E-12 | ?+ | 1.894E-12 | | New |
| 260 | 3 | 3-122934000-C-CAGGTTCA | rs6101212 | SEMA5B | 0.0023 | C | | 4.4E-10 | ?+ | 4.402E-10 | 0.3543 | New |
| 261 | 3 | 3-124436561-G-A | rs1375566 | KALRN | 0.0007 | A | | 3.72E-11 | ?+ | 3.719E-11 | 0.3207 | New |
| 261 | 3 | 3-124436578-T-A | rs2093468 | KALRN | 0.0007 | A | | 3.62E-08 | ?+ | 3.624E-08 | | New |
| 262 | 3 | 3-124807408-TC-T | | ITGB5 | 0.0003 | T | | 7.49E-11 | ?+ | 7.492E-11 | 0.4432 | New |
| 263 | 3 | 3-124898425-C-CCT | rs1254559 | ITGB5 | 0.0051 | CCT | | 1.88E-10 | ?+ | 1.876E-10 | | New |
| 264 | 3 | 3-12502323-ATC-A | rs1267492 | TSEN2 | 0.0016 | A | | 1.15E-11 | ?+ | 1.151E-11 | 0.9146 | New |
| 265 | 3 | 3-125521495-AAAATAC-A | | NA | 0.0003 | AAAATAC | | 4.31E-08 | ?+ | 4.306E-08 | | New |
| 266 | 3 | 3-127582685-GAGT-G | | NA | 0.0006 | GAGT | | 1.39E-10 | ?+ | 1.389E-10 | | New |
| 266 | 3 | 3-127582687-GCAA-G | | NA | 0.0004 | G | | 3.46E-09 | ?+ | 3.464E-09 | | New |
| 267 | 3 | 3-128645253-C-CCT | | NA | 0.0006 | C | | 1.06E-09 | ?+ | 1.056E-09 | | New |
| 268 | 3 | 3-128696475-G-A | rs1325254 | RPN1, RF00399 | 0.0005 | G | | 2.42E-08 | ?+ | 2.418E-08 | 0.141 | New |
| 268 | 3 | 3-128696485-C-CTT | | NA | 0.0007 | C | | 5.31E-13 | ?+ | 5.31E-13 | | New |
| 269 | 3 | 3-129426499-G-T | rs1396035 | EFCAB12 | 0.0004 | T | 0.435313 | 1.09E-10 | ++ | 2.243E-08 | | New |
| 270 | 3 | 3-136704609-C-CTT | rs1292259 | STAG1 | 0.0014 | C | | 1.2E-08 | ?+ | 1.201E-08 | 0.3846 | New |
| 271 | 3 | 3-138728773-C-CCG | | NA | 0.0014 | C | | 3.16E-10 | ?+ | 3.156E-10 | 0.9268 | New |
| 272 | 3 | 3-141640052-C-CT | rs1198946 | RASA2, RNF7 | 0.0019 | C | 0.402725 | 2.79E-10 | -+ | 0.00001049 | 0.01932 | New |
| 273 | 3 | 3-151417487-C-CGGG | | NA | 0.0016 | CGGG | | 3.26E-08 | ?- | 3.258E-08 | | New |
| 274 | 3 | 3-153261711-C-T | rs1250164 | RN7SL300P, AC078788.1 | 0.0013 | C | 0.365319 | 9.93E-11 | -+ | 0.00001478 | | New |
| 275 | 3 | 3-15491870-C-T | rs1198422 | COLQ | 0.0007 | T | | 6.47E-09 | ?+ | 6.466E-09 | 0.4823 | New |
| 275 | 3 | 3-15491876-G-A | rs1378323 | COLQ | 0.0008 | A | 0.979003 | 1.35E-13 | ++ | 7.723E-13 | | New |

| | | | | | | | | | | | | |
|---|---|---|---|---|---|---|---|---|---|---|---|---|
| 276 | 3 | 3-15569473-C-T | rs1424900 | HACL1 | 0.001 | C | | 8.02E-10 | ?+ | 8.02E-10 | | New |
| 276 | 3 | 3-15569482-G-A | rs1306953 | HACL1 | 0.0006 | A | 0.593871 | 9.45E-09 | -+ | 1.473E-08 | | New |
| 276 | 3 | 3-15569486-C-T | rs1336810 | HACL1 | 0.0005 | C | 0.408286 | 5.66E-09 | -+ | 1.548E-08 | | New |
| 277 | 3 | 3-155795816-C-T | rs9027252 | AC104472.3, C3orf33 | 0.0018 | C | 0.11838 | 3.39E-11 | ++ | 1.813E-08 | 0.07872 | New |
| 278 | 3 | 3-15592997-C-CATGTGTGCGTGTATACACATGTACGCAT | rs1487030 | HACL1 | 0.0095 | C | | 1.52E-08 | ?+ | 1.52E-08 | | New |
| 279 | 3 | 3-15593380-AT-A | | NA | 0.0013 | A | | 4.92E-14 | ?+ | 4.925E-14 | | New |
| 279 | 3 | 3-15593389-ATTTT-A | | NA | 0.0012 | A | | 9.19E-12 | ?+ | 9.191E-12 | | New |
| 280 | 3 | 3-16422426-G-C | rs2075203 | RFTN1 | 0.001 | C | 0.248755 | 2.28E-10 | -+ | 0.000001247 | 0.1421 | New |
| 281 | 3 | 3-170509612-C-T | rs1201387 | SLC7A14-AS1 | 0.0006 | T | | 3.42E-09 | ?+ | 3.417E-09 | | New |
| 282 | 3 | 3-170873157-C-CT | | NA | 0.0006 | CT | 0.7272 | 8.52E-12 | -+ | 1.789E-10 | | New |
| 283 | 3 | 3-174762293-G-A | | NAALADL2 | 0.0016 | G | 0.984198 | 7.17E-09 | -+ | 7.043E-08 | 0.02411 | New |
| 283 | 3 | 3-174762303-G-A | rs1423662 | NAALADL2 | 0.002 | A | 0.284941 | 4.05E-10 | ++ | 6.916E-10 | 0.0212 | New |
| 284 | 3 | 3-177225071-C-CAG | rs1371401 | TBL1XR1 | 0.0008 | C | | 1.33E-10 | ?+ | 1.327E-10 | | New |
| 285 | 3 | 3-183473142-GGAGT-G | rs1287529 | RF00394, KLHL6 | 0.0057 | GGAGT | | 1.05E-11 | ?+ | 1.046E-11 | | New |
| 286 | 3 | 3-184425210-C-T | rs1298828 | LINC02054 | 0.001 | T | | 1.74E-13 | ?+ | 1.736E-13 | 0.1827 | New |
| 287 | 3 | 3-185258472-G-T | rs6228869 | EHHADH, MAP3K13 | 0.0015 | T | 0.925728 | 6.71E-14 | ++ | 1.612E-08 | | New |
| 288 | 3 | 3-187807145-T-A | rs1239483 | AC108681.1, AC068295.1 | 0.0015 | A | | 6.46E-12 | ?+ | 6.456E-12 | | New |
| 289 | 3 | 3-192900736-T-A | rs1242199 | MB21D2 | 0.002 | A | 0.427822 | 2.49E-27 | ++ | 3.502E-25 | 0.3559 | New |
| 289 | 3 | 3-192900746-G-T | rs1257773 | MB21D2 | 0.0019 | T | 0.427822 | 8.47E-20 | ++ | 3.198E-18 | | New |
| 290 | 3 | 3-194954548-G-A | rs1022617 | AC090505.2, XXYLT1 | 0.0006 | A | 0.186657 | 3.62E-09 | -+ | 0.00002855 | 0.4455 | New |
| 291 | 3 | 3-196266254-C-CCT | | AC069257.3, PCYT1A | 0.0025 | CCT | | 1.62E-11 | ?+ | 1.619E-11 | | New |

| | | | | | | | | | | | |
|---|---|---|---|---|---|---|---|---|---|---|---|
| 292 | 3 | 3-196675754-C-CAAGTG | | PIGX | 0.0007 | CAAGTG | | 1.66E-08 | ?+ | 1.658E-08 | | New |
| 293 | 3 | 3-196852149-AC-A | | NA | 0.0009 | A | | 3.75E-16 | ?+ | 3.752E-16 | | New |
| 294 | 3 | 3-196861715-GAT-G | | NA | 0.0008 | G | | 3.05E-08 | ?+ | 3.048E-08 | | New |
| 294 | 3 | 3-196861718-GCA-G | rs1297156 | PAK2, SENP5 | 0.0007 | GCA | | 3.03E-08 | ?+ | 3.032E-08 | | New |
| 295 | 3 | 3-29143871-C-A | rs1290441 | RBMS3-AS3 | 0.0003 | A | 0.893975 | 3.12E-08 | -+ | 1.789E-07 | | New |
| 296 | 3 | 3-3430731-C-T | rs5572056 | AC034195.1 | 0.0005 | T | 0.484819 | 4.32E-08 | ++ | 0.0000551 | | New |
| 297 | 3 | 3-3431293-G-T | rs1304462 | AC034195.1 | 0.0017 | T | 0.122209 | 4.34E-21 | -+ | 1.752E-08 | 0.9713 | New |
| 297 | 3 | 3-3431321-C-T | rs1848461 | AC034195.1 | 0.0002 | C | 0.386242 | 1.14E-08 | -+ | 5.791E-08 | | New |
| 298 | 3 | 3-36963928-C-T | rs1170846 | TRANK1, RNU6ATAC4P | 0.0014 | T | | 5.88E-09 | ?+ | 5.88E-09 | 0.4509 | Old |
| 298 | 3 | 3-36963941-G-T | rs5361459 | TRANK1, RNU6ATAC4P | 0.0007 | T | 0.528123 | 3.73E-08 | ++ | 4.126E-08 | | Old |
| 299 | 3 | 3-40344008-AGATG-A | | NA | 0.0005 | AGATG | | 5.17E-12 | ?+ | 5.171E-12 | | New |
| 300 | 3 | 3-41061991-AT-A | rs1277391 | AC099541.1, AC009743.1 | 0.0011 | AT | | 2.6E-10 | ?+ | 2.599E-10 | 0.2223 | New |
| 300 | 3 | 3-41061993-AG-A | rs1349409 | AC099541.1, AC009743.1 | 0.0013 | A | | 1.06E-10 | ?+ | 1.061E-10 | | New |
| 301 | 3 | 3-41971464-C-CA | rs1208311 | ULK4, TRAK1 | 0.0019 | C | | 6.17E-11 | ?+ | 6.174E-11 | 0.8239 | New |
| 302 | 3 | 3-47755019-GAT-G | rs1485429 | SMARCC1 | 0.002 | G | | 6.48E-11 | ?+ | 6.483E-11 | 0.8586 | New |
| 302 | 3 | 3-47755022-GCA-G | rs1191304 | SMARCC1 | 0.0019 | GCA | | 1.48E-10 | ?+ | 1.482E-10 | | New |
| 303 | 3 | 3-47789462-G-C | | SMARCC1, DHX30 | 0.0004 | G | 0.897148 | 3.8E-08 | ++ | 0.0009337 | | New |
| 303 | 3 | 3-47789474-G-T | | SMARCC1, DHX30 | 0.0001 | T | | 3.78E-08 | ?+ | 3.781E-08 | | New |

| | | | | | | | | | | | | |
|---|---|---|---|---|---|---|---|---|---|---|---|---|
| 304 | 3 | 3-48201876-G-A | rs1309645 | MIR4443, CAMP | 0.0056 | G | 0.983892 | 7.41E-13 | -+ | 0.001856 | 0.7998 | New |
| 304 | 3 | 3-48201926-G-C | rs1179799 | MIR4443, CAMP | 0.0025 | G | 0.009898 | 1.3E-08 | ++ | 3.529E-07 | 0.2613 | New |
| 305 | 3 | 3-48876877-C-CCT | | NA | 0.0007 | C | | 1.65E-11 | ?+ | 1.649E-11 | | New |
| 306 | 3 | 3-48925935-G-C | rs2085530 | ARIH2 | 0.0008 | G | | 1.54E-09 | ?+ | 1.543E-09 | | New |
| 307 | 3 | 3-48939528-C-CGGGTGGATCATG | | NA | 0.0029 | C | | 7.08E-09 | ?+ | 7.082E-09 | | New |
| 308 | 3 | 3-49210704-TCTAA-T | | NA | 0.0013 | TCTAA | | 7.16E-11 | ?+ | 7.159E-11 | | New |
| 309 | 3 | 3-49399235-C-T | rs1352013 | RHOA | 0.0004 | T | 0.318581 | 6.73E-09 | -+ | 0.00003978 | | New |
| 310 | 3 | 3-49947243-C-CATGAGAA | | NA | 0.0019 | CATGAGAA | | 2.7E-16 | ?+ | 2.699E-16 | | New |
| 311 | 3 | 3-51353784-C-CCTCT | rs1262184 | DOCK3 | 0.0021 | C | 0.18184 | 8.99E-18 | -+ | 5.872E-17 | 0.9227 | New |
| 312 | 3 | 3-71313726-C-T | rs1021098 | AC097634.4, FOXP1 | 0.0025 | T | 0.937193 | 7.92E-10 | -+ | 0.0004507 | 0.8745 | New |
| 313 | 3 | 3-73151147-G-A | rs7614861 | RNU2-64P, RNU6-1270P | 0.0001 | A | 0.46582 | 1.56E-09 | -+ | 1.425E-08 | 0.4679 | New |
| 314 | 3 | 3-75612176-C-CG | | LINC02018, MIR1324 | 0.1081 | CG | | 1.23E-10 | ?+ | 1.234E-10 | | New |
| 315 | 3 | 3-75616418-ATG-A | | NA | 0.0014 | ATG | 0.413409 | 1.33E-20 | ++ | 8.457E-17 | | New |
| 316 | 3 | 3-75674095-TCAAATATGGGTCAAATATGGCTTAG-T | | NA | 0.0034 | T | | 4.02E-13 | ?+ | 4.022E-13 | | New |
| 317 | 3 | 3-75734134-T-A | rs7980415 | LINC00960 | 0.0001 | A | | 1.47E-08 | ?+ | 1.468E-08 | | New |
| 318 | 3 | 3-77292683-GGGTAAGCTGAGGCTAGATCACCCCAGACATAAAGTAAAATTGATGGTTAAACGGGAAGTTGAGGCTAGAGCACTAAAGACATAAAGTAAATTTGACGGTTAAACA-G | rs1560455 | ROBO2 | 0.2612 | G | | 1.24E-10 | ?- | 1.243E-10 | | New |

| | | | | | | | | | | | | |
|---|---|---|---|---|---|---|---|---|---|---|---|---|
| 319 | 3 | 3-80420455-G-T | rs1575784 | ROBO1, AC108690 .1 | 0.0006 | T | 0.34103 | 8.47E-09 | -+ | 1.058E-07 | | New |
| 320 | 3 | 3-90283097-C-CT | rs3738225 | RNU6-712P, RF01699 | 0.0018 | CT | | 6.09E-09 | ?+ | 6.091E-09 | | New |
| 321 | 3 | 3-90291826-C-T | rs1705599 | RNU6-712P, RF01699 | 0.001 | C | 0.116525 | 1.24E-08 | -+ | 0.0554 | | New |
| 322 | 3 | 3-90498243-G-C | rs1196020 | RNU6-712P, RF01699 | 0.001 | G | | 2.01E-08 | ?+ | 2.007E-08 | | New |
| 323 | 3 | 3-90498313-G-T | rs1249242 | RNU6-712P, RF01699 | 0.0008 | T | 0.385347 | 1.62E-09 | ++ | 1.52E-09 | | New |
| 324 | 3 | 3-9803104-C-T | rs5617934 | ARPC4, ARPC4-TTLL3 | 0.0009 | C | 0.900714 | 3.77E-09 | -+ | 5.222E-07 | 0.7829 | New |
| 324 | 3 | 3-9803125-C-T | rs3872719 | ARPC4, ARPC4-TTLL3 | 0.001 | T | 0.708871 | 1.9E-09 | ++ | 3.016E-09 | 0.2456 | New |
| 325 | 4 | 4-102329460-TAA-T | | NA | 0.0008 | TAA | | 4.09E-12 | ?+ | 4.093E-12 | 0.5972 | New |
| 325 | 4 | 4-102329468-C-CAA | | NA | 0.0018 | CAA | | 2.19E-15 | ?+ | 2.186E-15 | | New |
| 326 | 4 | 4-103431931-G-A | rs5509112 | LINC0242 8 | 0.0013 | A | 0.5353 | 9.99E-10 | -+ | 0.00002699 | 0.06866 | New |
| 327 | 4 | 4-113231215-C-T | | ANK2 | 0.0005 | C | 0.201588 | 2.25E-09 | -+ | 0.0006554 | | New |
| 327 | 4 | 4-113231218-ATT-A | | ANK2 | 0.0012 | A | | 3.75E-12 | ?+ | 3.749E-12 | | New |
| 328 | 4 | 4-114959008-T-A | rs1303093 | NDST4 | 0.0006 | A | | 4.95E-10 | ?+ | 4.947E-10 | | New |
| 329 | 4 | 4-120281144-C-CA | rs1403703 | AC073475 .1 | 0.0013 | C | 0.141313 | 6.19E-09 | ++ | 6.46E-08 | | New |
| 330 | 4 | 4-120327591-C-CTCAT | | AC073475 .1 | 0.0021 | C | | 3.11E-10 | ?+ | 3.113E-10 | | New |
| 331 | 4 | 4-128104101-C-CAA | | NA | 0.0003 | CAA | | 1.65E-09 | ?+ | 1.647E-09 | | New |
| 332 | 4 | 4-128814728-AT-A | | NA | 0.0085 | AT | 0.382213 | 1.29E-10 | -+ | 0.2278 | | New |
| 333 | 4 | 4-143981449-C-T | rs4125557 | AC107223 .1 | 0.0086 | T | 0.417478 | 3.51E-14 | +- | 0.0002046 | | New |

| | | | | | | | | | | | | |
|---|---|---|---|---|---|---|---|---|---|---|---|---|
| 334 | 4 | 4-144912122-C-T | rs1441804 | AC098588 .1, ANAPC10 | 0.0012 | T | | 2.17E-08 | ?+ | 2.173E-08 | 0.1819 | New |
| 334 | 4 | 4-144912136-G-C | rs5746558 | AC098588 .1, ANAPC10 | 0.0013 | G | 0.671216 | 1.57E-09 | -+ | 3.657E-08 | 0.6657 | New |
| 335 | 4 | 4-151065713-C-CAA | | LRBA, RPS3A | 0.0018 | C | | 1.05E-09 | ?+ | 1.055E-09 | 0.5179 | New |
| 336 | 4 | 4-151299398-T-A | rs9038871 | SH3D19 | 0.0005 | A | 0.780394 | 7.97E-10 | -+ | 3.457E-08 | | New |
| 337 | 4 | 4-153944779-C-CTCAT | | SFRP2, AC079298 .3 | 0.0024 | CTCAT | | 4.25E-11 | ?+ | 4.247E-11 | | New |
| 338 | 4 | 4-159437376-AG-A | | NA | 0.0011 | A | | 6.07E-09 | ?+ | 6.069E-09 | | New |
| 339 | 4 | 4-162815005-C-T | rs5707432 | AC021134 .1, AC021134 .2 | 0.0012 | T | 0.603129 | 1.86E-09 | -+ | 1.377E-08 | 0.2183 | New |
| 339 | 4 | 4-162815032-G-A | rs5562090 | AC021134 .1, AC021134 .2 | 0.0017 | A | 0.977475 | 1.69E-09 | ++ | 2.991E-09 | 0.2505 | New |
| 340 | 4 | 4-162815609-G-A | rs9400185 | AC021134 .1, AC021134 .2 | 0.0013 | G | 0.528117 | 3.44E-09 | -+ | 0.006637 | 0.4893 | New |
| 341 | 4 | 4-168966338-C-CAGTG | rs1261220 | CBR4 | 0.0019 | CAGTG | | 3.95E-09 | ?+ | 3.946E-09 | 0.8995 | New |
| 342 | 4 | 4-172464489-C-CCT | | NA | 1E-04 | C | | 2.49E-08 | ?+ | 2.485E-08 | | New |
| 343 | 4 | 4-17565394-C-T | rs1368642 | CLRN2, LAP3 | 0.0008 | T | | 6.25E-09 | ?+ | 6.252E-09 | | New |
| 344 | 4 | 4-176800802-C-A | rs8660499 | VEGFC, AC097518 .2 | 0.0099 | A | | 1.04E-09 | ?- | 1.038E-09 | | New |
| 345 | 4 | 4-182621523-G-GA | rs1129108 | TENM3 | 0.0423 | G | | 1.69E-12 | ?+ | 1.695E-12 | | New |
| 346 | 4 | 4-183509969-C-CAG | rs1483773 | ING2 | 0.005 | C | | 1.11E-11 | ?+ | 1.108E-11 | | New |
| 347 | 4 | 4-183643226-T-A | rs1300537 | RWDD4 | 0.0006 | A | 0.984512 | 1.05E-08 | -+ | 0.000006291 | 0.355 | New |

| | | | | | | | | | | | |
|---|---|---|---|---|---|---|---|---|---|---|---|
| 348 | 4 | 4-19483278-C-T | rs1163397 | AC024230.1 | 0.0005 | C | 0.020852 | 2.84E-08 | ++ | 1.132E-08 | 0.5251 New |
| 349 | 4 | 4-21544845-C-CTTTTTTT | | NA | 0.0008 | CTTTTTTT | | 2.56E-09 | ?+ | 2.559E-09 | New |
| 350 | 4 | 4-22509278-G-A | rs1254742 | ADGRA3 | 0.0017 | G | 0.896124 | 7.67E-15 | ++ | 3.712E-14 | 0.5028 New |
| 351 | 4 | 4-2440655-G-T | rs1193262 | CFAP99 | 0.0005 | G | 0.226579 | 4.74E-08 | -+ | 0.00002036 | New |
| 352 | 4 | 4-2522168-G-A | rs5460198 | RNF4 | 0.0024 | A | 0.982773 | 2.98E-15 | ++ | 1.785E-08 | 0.2682 New |
| 352 | 4 | 4-2522176-C-T | rs5764689 | RNF4 | 0.0043 | C | 0.545021 | 9.46E-26 | ++ | 2.015E-19 | 0.5542 New |
| 352 | 4 | 4-2522177-G-A | rs5419773 | RNF4 | 0.0036 | A | 0.651691 | 1.84E-23 | ++ | 6.485E-17 | 0.6676 New |
| 353 | 4 | 4-2522803-C-CCT | rs1340008 | RNF4 | 0.0025 | CCT | 0.322392 | 8.21E-22 | ++ | 2.521E-16 | New |
| 354 | 4 | 4-2525429-C-T | rs9243937 | RNF4 | 0.0009 | C | 0.460048 | 3.9E-09 | ++ | 2.353E-07 | 0.7409 New |
| 355 | 4 | 4-26243074-AAC-A | | NA | 0.0003 | AAC | | 2.49E-09 | ?+ | 2.489E-09 | New |
| 356 | 4 | 4-2764348-C-CAG | | NA | 0.0046 | C | | 1.9E-11 | ?+ | 1.899E-11 | New |
| 357 | 4 | 4-39359815-C-CCA | | NA | 0.0002 | CCA | | 8.62E-09 | ?+ | 8.616E-09 | New |
| 358 | 4 | 4-39684712-GCC-G | rs1717431 | AC108471.2, UBE2K | 0.0158 | GCC | | 7.48E-11 | ?+ | 7.477E-11 | New |
| 359 | 4 | 4-39773277-GT-G | rs1182056 | UBE2K | 0.006 | G | | 3.9E-22 | ?+ | 3.896E-22 | Old |
| 359 | 4 | 4-39773298-TC-T | rs8797928 | UBE2K | 0.0011 | TC | 0.937658 | 1.63E-10 | -+ | 4.277E-09 | Old |
| 360 | 4 | 4-39831334-C-CAA | | NA | 0.0005 | CAA | 0.411968 | 2.66E-08 | -+ | 7.737E-08 | Old |
| 361 | 4 | 4-42081999-C-CCT | | NA | 0.0011 | CCT | | 1.61E-13 | ?+ | 1.608E-13 | New |
| 362 | 4 | 4-49308277-C-CT | | NA | 0.0014 | C | | 5.67E-09 | ?+ | 5.67E-09 | New |
| 363 | 4 | 4-49641607-AATGGAATAAAAGGTCATGGAATGGAAT-A | | NA | 0.0939 | A | | 4.11E-09 | ?- | 4.114E-09 | New |
| 364 | 4 | 4-56835539-GGTGGA-G | | NA | 0.0007 | G | | 6.48E-11 | ?+ | 6.481E-11 | New |
| 365 | 4 | 4-634885-GCC-G | rs1224926 | PDE6B | 0.0017 | G | | 1.68E-09 | ?+ | 1.676E-09 | Old |
| 365 | 4 | 4-634889-GCC-G | rs1478230 | PDE6B | 0.0013 | G | | 1.23E-08 | ?+ | 1.228E-08 | Old |
| 366 | 4 | 4-70871593-C-CTG | rs1457264 | RNU6-891P, MOB1B | 0.0017 | C | | 4.48E-08 | ?+ | 4.481E-08 | 0.3342 New |
| 367 | 4 | 4-70884321-C-CAT | | NA | 0.0005 | CAT | | 1.19E-12 | ?+ | 1.189E-12 | New |
| 368 | 4 | 4-75682763-C-CCT | rs1240119 | G3BP2 | 0.0004 | CCT | | 1.26E-08 | ?+ | 1.265E-08 | Old |
| 369 | 4 | 4-76667749-ACC-A | rs1381828 | SHROOM3 | 0.0011 | ACC | | 8.93E-11 | ?+ | 8.929E-11 | Old |
| 370 | 4 | 4-83130461-C-CA | rs1029109 | PLAC8 | 0.0016 | C | 0.784777 | 9.91E-12 | -+ | 2.614E-07 | 0.3539 New |
| 371 | 4 | 4-83575604-C-T | rs1335344 | GPAT3 | 0.0031 | T | 0.251024 | 2.07E-21 | -+ | 5.665E-13 | New |
| 372 | 4 | 4-84770219-C-CG | | NA | 0.001 | CG | | 5.91E-09 | ?+ | 5.915E-09 | New |
| 373 | 4 | 4-86958333-C-T | rs9031170 | AFF1 | 0.0008 | C | | 1.58E-08 | ?+ | 1.584E-08 | 0.6511 New |
| 374 | 4 | 4-8967028-AAC-A | | NA | 0.0009 | AAC | | 1.66E-11 | ?+ | 1.658E-11 | New |

| | | | | | | | | | | | |
|---|---|---|---|---|---|---|---|---|---|---|---|
| 375 | 4 | 4-920686-GACGT-G | rs1470992 | GAK | 0.0019 | GACGT | | 1.03E-13 | ?+ | 1.027E-13 | 0.3599 | Old |
| 375 | 4 | 4-920689-GCCCA-G | rs1723794 | GAK | 0.0013 | G | | 2.38E-11 | ?+ | 2.382E-11 | | Old |
| 376 | 4 | 4-9501107-AAC-A | | DEFB131A, MIR548I2 | 0.002 | AAC | | 4.69E-17 | ?+ | 4.686E-17 | | New |
| 376 | 4 | 4-9501126-C-T | | DEFB131A, MIR548I2 | 0.0003 | C | | 9.75E-09 | ?+ | 9.747E-09 | | New |
| 377 | 4 | 4-9593894-C-T | rs1213918 | AC105916.1 | 0.0002 | C | | 2.38E-08 | ?+ | 2.381E-08 | | New |
| 378 | 4 | 4-9650027-T-A | | AC105916.1 | 0.0003 | A | | 1.02E-09 | ?+ | 1.019E-09 | | New |
| 378 | 4 | 4-9650036-G-A | rs2108924 | AC105916.1 | 0.0002 | A | 0.197965 | 1.61E-09 | -+ | 7.505E-08 | | New |
| 379 | 4 | 4-978148-G-A | rs1713432 | DGKQ | 0.0002 | G | | 2.91E-10 | ?+ | 2.91E-10 | | Old |
| 380 | 4 | 4-99086726-C-CCT | rs1351881 | ADH5 | 0.003 | CCT | | 1.7E-09 | ?+ | 1.697E-09 | | New |
| 381 | 5 | 5-116698546-C-A | rs1218851 | SEMA6A-AS2, LINC02214 | 0.001 | A | 0.37322 | 3.53E-08 | ++ | 3.158E-08 | 0.3596 | New |
| 382 | 5 | 5-119047353-TGA-T | | NA | 0.0008 | TGA | | 6.76E-11 | ?+ | 6.761E-11 | | New |
| 383 | 5 | 5-134615116-AGGCG-A | rs1298939 | SAR1B | 0.0065 | AGGCG | | 9.99E-34 | ?+ | 9.988E-34 | 0.2472 | New |
| 383 | 5 | 5-134615122-GCAGA-G | rs1233962 | SAR1B | 0.0051 | G | | 1.08E-22 | ?+ | 1.08E-22 | 0.1536 | New |
| 384 | 5 | 5-134954841-C-A | rs9203773 | PCBD2 | 0.0013 | A | | 1.58E-13 | ?+ | 1.576E-13 | | New |
| 385 | 5 | 5-138104941-G-T | rs1135320 | WNT8A, NME5 | 0.0026 | G | | 2.05E-11 | ?+ | 2.048E-11 | | New |
| 386 | 5 | 5-138214143-C-CT | rs1284394 | CDC23 | 0.0017 | CT | | 4.49E-09 | ?+ | 4.487E-09 | 0.08963 | New |
| 387 | 5 | 5-138605888-G-T | rs1241247 | HSPA9, CTNNA1 | 0.0005 | G | 0.82292 | 2.99E-09 | -+ | 0.000000964 | | New |
| 388 | 5 | 5-139242644-C-CCT | rs1311463 | RNU6-572P, MATR3 | 0.0016 | CCT | | 3.47E-14 | ?+ | 3.473E-14 | | New |
| 388 | 5 | 5-139242645-G-A | rs1355274 | RNU6-572P, MATR3 | 0.0006 | G | | 5.66E-11 | ?+ | 5.66E-11 | | New |

| | | | | | | | | | | | | |
|---|---|---|---|---|---|---|---|---|---|---|---|---|
| 389 | 5 | 5-139345867-TCCGC-T | | PAIP2 | 0.001 | T | | 5.2E-10 | ?+ | 5.196E-10 | | New |
| 389 | 5 | 5-139345874-AACGG-A | | PAIP2 | 0.0011 | AACGG | | 4.45E-11 | ?+ | 4.447E-11 | | New |
| 390 | 5 | 5-139405218-TG-T | | NA | 0.001 | TG | | 1.39E-09 | ?+ | 1.394E-09 | | New |
| 391 | 5 | 5-139533514-GTGTC-G | | NA | 0.0008 | G | | 1.04E-08 | ?+ | 1.042E-08 | | New |
| 391 | 5 | 5-139533522-C-CACTT | | NA | 0.001 | CACTT | | 6.46E-09 | ?+ | 6.464E-09 | | New |
| 391 | 5 | 5-139533525-C-CT | | NA | 0.001 | CT | | 4.62E-09 | ?+ | 4.625E-09 | | New |
| 391 | 5 | 5-139533528-TA-T | | NA | 0.001 | T | | 4.46E-09 | ?+ | 4.463E-09 | | New |
| 392 | 5 | 5-139702543-C-CTT | | NA | 0.0006 | C | | 2.46E-08 | ?+ | 2.464E-08 | | New |
| 393 | 5 | 5-141447364-C-CCTGACCCTGG | | NA | 0.002 | C | | 2.61E-17 | ?+ | 2.611E-17 | | New |
| 394 | 5 | 5-142336719-C-A | rs1229916 | SPRY4-AS1 | 0.0024 | A | 0.314347 | 1.89E-24 | -+ | 2.843E-19 | | New |
| 395 | 5 | 5-146234861-C-CCA | | AC091959.3, RBM27 | 0.0008 | C | | 1.2E-10 | ?+ | 1.198E-10 | | New |
| 395 | 5 | 5-146234864-TGC-T | | AC091959.3, RBM27 | 0.0008 | TGC | | 8.21E-09 | ?+ | 8.212E-09 | | New |
| 396 | 5 | 5-146259296-ACT-A | rs1170313 | AC091959.3, RBM27 | 0.0038 | A | | 1.64E-08 | ?+ | 1.644E-08 | 0.7128 | New |
| 397 | 5 | 5-157864857-G-A | rs1273153 | CLINT1, RNU2-48P | 0.0005 | A | 0.911079 | 1.45E-09 | ++ | 3.802E-09 | | New |
| 398 | 5 | 5-158747-GT-G | rs1245037 | PLEKHG4B | 0.0007 | G | | 1.31E-08 | ?+ | 1.312E-08 | | New |
| 399 | 5 | 5-160138360-G-T | rs5285575 | PWWP2A, FABP6 | 0.0009 | G | 0.124206 | 1.98E-11 | -+ | 0.0001582 | 0.8159 | New |
| 400 | 5 | 5-160138763-C-T | rs8936296 | PWWP2A, FABP6 | 0.0012 | C | 0.385039 | 4.15E-12 | ++ | 1.533E-08 | 0.4379 | New |
| 401 | 5 | 5-160140248-GGT-G | | NA | 0.0005 | GGT | | 8.52E-10 | ?+ | 8.524E-10 | | New |
| 402 | 5 | 5-160257333-G-C | rs5599861 | CCNJL | 0.0018 | C | 0.270922 | 3.09E-15 | ++ | 1.21E-10 | | New |
| 402 | 5 | 5-160257343-C-T | rs1307496 | CCNJL | 0.0014 | T | 0.306034 | 2.67E-16 | ++ | 8.768E-11 | 0.8306 | New |
| 403 | 5 | 5-163628927-C-CAG | | NA | 0.0018 | C | | 1.47E-12 | ?+ | 1.47E-12 | | New |

| | | | | | | | | | | | | |
|---|---|---|---|---|---|---|---|---|---|---|---|---|
| 403 | 5 | 5-163628931-ACG-A | | NA | 0.0016 | ACG | | 8.11E-11 | ?+ | 8.109E-11 | | New |
| 404 | 5 | 5-168253265-C-CTACGCACTCATTATTTATA | | NA | 0.0003 | CTACGCACTCATTATTT | | 6.97E-09 | ?+ | 6.968E-09 | | New |
| 405 | 5 | 5-168253657-C-T | rs1281813 | AC026689.1 | 0.0009 | T | 0.23024 | 3.65E-12 | -+ | 8.022E-09 | 0.9342 | New |
| 405 | 5 | 5-168253667-G-C | rs1274427 | AC026689.1 | 0.0018 | G | 0.239814 | 5.5E-12 | -+ | 1.322E-09 | | New |
| 405 | 5 | 5-168253671-C-T | rs1305260 | AC026689.1 | 0.0007 | T | 0.274923 | 6.17E-10 | ++ | 3.35E-10 | | New |
| 406 | 5 | 5-169892234-C-T | rs1398423 | DOCK2, INSYN2B | 0.0012 | C | 0.165105 | 3.87E-20 | -+ | 8.877E-08 | | New |
| 407 | 5 | 5-176502362-C-CCT | | FAF2 | 0.0032 | C | | 4.42E-21 | ?+ | 4.418E-21 | 0.2723 | New |
| 408 | 5 | 5-176801068-CTGTGTG-C | | LINC01574, UNC5A | 0.312 | C | | 3.74E-11 | ?+ | 3.736E-11 | | New |
| 409 | 5 | 5-178174212-ACC-A | | NA | 0.0002 | A | | 2.73E-08 | ?+ | 2.733E-08 | | New |
| 410 | 5 | 5-179544212-GC-G | | NA | 0.0007 | G | | 3.69E-09 | ?+ | 3.693E-09 | | New |
| 411 | 5 | 5-179591395-AG-A | | NA | 0.0015 | A | | 1.35E-12 | ?+ | 1.346E-12 | | New |
| 412 | 5 | 5-179691169-GCACCC-G | | NA | 0.0004 | GCACCC | | 8.07E-09 | ?+ | 8.074E-09 | | Old |
| 413 | 5 | 5-179696510-AGG-A | | NA | 0.0008 | A | | 5.7E-11 | ?+ | 5.696E-11 | | Old |
| 413 | 5 | 5-179696514-G-A | rs1459112 | CANX | 0.0005 | G | | 1.45E-09 | ?+ | 1.455E-09 | | Old |
| 414 | 5 | 5-179789142-C-CT | | NA | 0.0006 | C | | 2.44E-08 | ?+ | 2.44E-08 | | Old |
| 415 | 5 | 5-181239860-C-A | rs1277187 | RACK1 | 0.0012 | C | 0.932587 | 2.06E-09 | ++ | 8.905E-08 | | New |
| 416 | 5 | 5-181240871-G-A | rs2546403 | RACK1 | 0.1673 | G | | 1.22E-08 | ?+ | 1.218E-08 | 0.1122 | New |
| 417 | 5 | 5-23526774-C-CAA | rs1491369 | PRDM9 | 0.0029 | CAA | | 4.42E-14 | ?+ | 4.425E-14 | | New |
| 418 | 5 | 5-285364-G-A | rs2846354 | AC021087.5, AHRR, PDCD6 | 0.2502 | A | | 2.78E-08 | ?+ | 2.784E-08 | | New |
| 419 | 5 | 5-31919818-GCCA-G | rs1343350 | PDZD2 | 0.002 | G | | 9.92E-22 | ?+ | 9.926E-22 | 0.4613 | New |
| 420 | 5 | 5-3323646-G-A | rs3749033 | AC091832.1, LINC01019 | 0.004 | A | | 1.3E-08 | ?+ | 1.302E-08 | | New |

| | | | | | | | | | | | |
|---|---|---|---|---|---|---|---|---|---|---|---|
| 420 | 5 | 5-3323650-G-A | rs3690416 | AC091832.1, LINC01019 | 0.0029 | A | | 2.45E-09 | ?+ | 2.449E-09 | | New |
| 421 | 5 | 5-37322460-C-A | rs1744306 | NUP155 | 0.0002 | A | | 1.24E-08 | ?+ | 1.236E-08 | | New |
| 422 | 5 | 5-37396773-G-A | rs1388224 | WDR70 | 0.0005 | G | 0.807775 | 3.29E-08 | -+ | 4.363E-07 | | New |
| 423 | 5 | 5-37486551-G-T | rs1739878 | WDR70 | 0.0004 | G | 0.018009 | 9.75E-09 | ++ | 5.517E-10 | | New |
| 424 | 5 | 5-38394491-G-C | rs1443689 | EGFLAM | 0.0007 | C | | 1.72E-10 | ?+ | 1.722E-10 | 0.32 | New |
| 425 | 5 | 5-42706839-T-A | rs1249385 | GHR | 0.0009 | A | 0.962605 | 1.79E-08 | ++ | 1.148E-07 | | New |
| 426 | 5 | 5-42954699-G-C | rs9506756 | AC008945.2, AC008875.3 | 0.0007 | C | | 2.4E-11 | ?+ | 2.402E-11 | | New |
| 426 | 5 | 5-42954706-G-A | rs1740607 | AC008945.2, AC008875.3 | 0.0009 | G | | 1.21E-11 | ?+ | 1.21E-11 | | New |
| 426 | 5 | 5-42954730-G-C | rs9492042 | AC008945.2, AC008875.3 | 0.0008 | G | 0.326477 | 2.98E-14 | ++ | 9.691E-14 | | New |
| 426 | 5 | 5-42954736-G-A | rs1046138 | AC008945.2, AC008875.3 | 0.0008 | G | | 2.85E-09 | ?+ | 2.855E-09 | | New |
| 427 | 5 | 5-42982044-C-A | rs1315936 | AC008945.2, AC008875.3 | 0.0028 | C | 0.181836 | 1.81E-10 | ++ | 2.689E-08 | | New |
| 428 | 5 | 5-43345773-G-A | rs1334993 | HMGCS1, CCL28 | 0.0004 | A | | 2.19E-08 | ?+ | 2.189E-08 | | New |
| 429 | 5 | 5-46397240-C-T | rs1164552 | AC122694.1, NONE | 0.0004 | T | 0.563522 | 3.49E-09 | -+ | 4.022E-08 | 0.3139 | New |
| 430 | 5 | 5-55688573-G-T | rs1179471 | SLC38A9 | 0.0017 | T | | 1.6E-09 | ?+ | 1.6E-09 | | New |
| 431 | 5 | 5-55987899-GAGAT-G | | NA | 0.0003 | GAGAT | | 2.42E-10 | ?+ | 2.42E-10 | | New |
| 432 | 5 | 5-57149535-C-T | rs9595866 | MIER3, RF00019 | 0.0003 | T | | 1.71E-08 | ?+ | 1.709E-08 | 0.7941 | New |

| | | | | | | | | | | | | |
|---|---|---|---|---|---|---|---|---|---|---|---|---|
| 433 | 5 | 5-5730755-TA-T | rs1246009 | AC026736.1, AC010266.2 | 0.0019 | T | | 1.45E-08 | ?+ | 1.451E-08 | | New |
| 434 | 5 | 5-61953687-G-A | rs1400393 | AC008877.1, RN7SKP157 | 0.0003 | G | | 2.5E-11 | ?+ | 2.502E-11 | | Old |
| 435 | 5 | 5-63078868-GTC-G | | NA | 0.0007 | GTC | | 3.07E-09 | ?+ | 3.066E-09 | | New |
| 436 | 5 | 5-65634189-GTTAGCCAGGATA-G | | NA | 0.0006 | G | | 1.35E-12 | ?+ | 1.353E-12 | | New |
| 437 | 5 | 5-69337801-AT-A | | NA | 0.0009 | AT | | 3.57E-09 | ?+ | 3.565E-09 | | New |
| 438 | 5 | 5-71297967-CA-C | | NA | 0.0946 | CA | | 2.45E-09 | ?+ | 2.452E-09 | | New |
| 439 | 5 | 5-76509558-ATTTT-A | | NA | 0.001 | A | | 5.1E-12 | ?+ | 5.099E-12 | | New |
| 440 | 5 | 5-78097383-A-G | rs4703750 | AP3B1 | 0.0088 | G | 0.330166 | 1.72E-08 | -+ | 0.00001423 | | New |
| 441 | 5 | 5-80507561-G-A | rs7618332 | FAM151B | 0.0004 | G | 0.233303 | 3.02E-13 | ++ | 1.258E-09 | | New |
| 442 | 5 | 5-82726070-TCCTGGCTAACAC-T | | AC026782.2, AC008885.2 | 0.0008 | TCCTGGCTAACAC | | 1.93E-09 | ?+ | 1.932E-09 | | New |
| 443 | 5 | 5-82972528-C-T | rs1743330 | AC027338.2, AC027338.1 | 0.0003 | T | 0.044276 | 1.38E-09 | ++ | 1.751E-10 | | New |
| 444 | 5 | 5-82973288-G-T | rs9747427 | AC027338.2, AC027338.1 | 0.0008 | T | 0.892244 | 5.92E-14 | -+ | 7.789E-13 | | New |
| 445 | 5 | 5-93000815-AC-A | | NA | 0.0012 | AC | | 4.75E-10 | ?+ | 4.752E-10 | | New |
| 445 | 5 | 5-93000816-G-T | | LINC02058, AC012625.1 | 0.0008 | G | | 2.42E-10 | ?+ | 2.425E-10 | | New |
| 446 | 6 | 6-101494698-AGGCG-A | | GRIK2 | 0.001 | AGGCG | | 2.2E-08 | ?+ | 0.000000022 | | New |
| 447 | 6 | 6-103320522-G-A | rs1441885 | AL357139.2, RF00438 | 0.0009 | G | 0.284396 | 4.61E-12 | -+ | 4.673E-07 | 0.4351 | New |

| | | | | | | | | | | | | |
|---|---|---|---|---|---|---|---|---|---|---|---|---|
| 447 | 6 | 6-103320550-GGAGGCT-G | rs1445466 | AL357139.2, RF00438 | 0.0012 | G | 0.450862 | 8.76E-13 | -+ | 1.478E-08 | | New |
| 448 | 6 | 6-104062626-C-T | rs1168094 | AL590608.1, AL357522.1 | 0.0035 | T | | 8.66E-11 | ?- | 8.663E-11 | | New |
| 449 | 6 | 6-106480620-TCG-T | rs1352922 | CRYBG1 | 0.0012 | T | | 3.7E-09 | ?+ | 3.7E-09 | 0.001142 | New |
| 449 | 6 | 6-106480624-ATG-A | rs1288116 | CRYBG1 | 0.0012 | ATG | | 2.94E-08 | ?+ | 2.941E-08 | | New |
| 450 | 6 | 6-10786446-G-A | rs8685383 | AL024498.2, MAK, TMEM14B | 0.0009 | G | | 8.4E-09 | ?+ | 8.398E-09 | | New |
| 451 | 6 | 6-10810629-ACGCC-A | | NA | 0.0002 | ACGCC | | 1.01E-08 | ?+ | 1.012E-08 | | New |
| 452 | 6 | 6-11192289-C-CA | | NA | 0.0114 | CA | 0.157323 | 1.84E-08 | +- | 0.3904 | | New |
| 453 | 6 | 6-118716962-ATG-A | | CEP85L, MCM9 | 0.001 | A | | 3.64E-15 | ?+ | 3.639E-15 | | New |
| 454 | 6 | 6-12042382-C-CGG | | NA | 0.0084 | CGG | | 1.46E-10 | ?+ | 1.456E-10 | | New |
| 455 | 6 | 6-122138733-G-T | rs1369563 | RNU2-8P, RNU1-18P | 0.0023 | T | 0.195508 | 2.96E-14 | -+ | 1.937E-11 | | New |
| 456 | 6 | 6-122139207-G-T | rs1776228 | RNU2-8P, RNU1-18P | 0.0012 | G | | 1.19E-09 | ?+ | 1.191E-09 | | New |
| 457 | 6 | 6-12840142-TC-T | | NA | 0.0012 | T | | 2E-14 | ?+ | 1.997E-14 | | New |
| 458 | 6 | 6-139484262-C-CAT | | NA | 0.0005 | C | | 7.68E-13 | ?+ | 7.686E-13 | | New |
| 458 | 6 | 6-139484265-GGC-G | | NA | 0.0005 | GGC | | 3.15E-13 | ?+ | 3.149E-13 | | New |
| 459 | 6 | 6-143890911-C-CTT | | NA | 0.0012 | CTT | | 4.73E-09 | ?+ | 4.732E-09 | | New |
| 460 | 6 | 6-14817512-C-A | rs5698770 | AL138720.1 | 0.0013 | A | 0.38453 | 1.27E-08 | -+ | 8.225E-08 | 0.4994 | New |
| 460 | 6 | 6-14817518-G-A | rs5324158 | AL138720.1 | 0.0012 | G | 0.254201 | 5.1E-12 | -+ | 1.383E-10 | 0.4535 | New |
| 460 | 6 | 6-14817529-GCA-G | rs7596338 | AL138720.1 | 0.0011 | GCA | | 9.18E-14 | ?+ | 9.181E-14 | 0.7234 | New |

| | | | | | | | | | | | | |
|---|---|---|---|---|---|---|---|---|---|---|---|---|
| 460 | 6 | 6-14817530-AGG-A | rs1249641 | AL138720.1 | 0.0011 | A | | 7.49E-12 | ?+ | 7.494E-12 | 0.7215 | New |
| 461 | 6 | 6-148303578-C-CTT | | NA | 0.0017 | C | | 1.08E-18 | ?+ | 1.085E-18 | | New |
| 462 | 6 | 6-14989159-C-CGTG | | NA | 0.001 | C | | 9.18E-10 | ?+ | 9.176E-10 | | New |
| 463 | 6 | 6-151261625-G-C | | AKAP12 | 0.0007 | G | 0.440187 | 2.64E-09 | ++ | 0.00000001 | | New |
| 464 | 6 | 6-155397325-G-A | rs1129006 | NOX3 | 0.0022 | G | 0.793851 | 4.8E-15 | -+ | 0.00004919 | | New |
| 465 | 6 | 6-156288006-G-T | rs1302203 | AL589693.1 | 0.0002 | T | 0.516708 | 1.79E-08 | -+ | 0.0000273 | 0.5886 | New |
| 466 | 6 | 6-15784875-G-A | rs1444258 | DTNBP1, LINC02543 | 0.0023 | A | 0.342444 | 1.16E-14 | ++ | 6.933E-08 | | New |
| 467 | 6 | 6-16267227-C-T | rs1293514 | GMPR | 0.0018 | C | 0.131139 | 4.15E-12 | ++ | 5.466E-10 | | New |
| 467 | 6 | 6-16267232-C-T | rs1218372 | GMPR | 0.0021 | C | 0.131162 | 1.08E-12 | ++ | 8.424E-11 | | New |
| 468 | 6 | 6-165408947-C-CCT | rs1428838 | PDE10A | 0.0023 | CCT | | 4.97E-09 | ?+ | 4.97E-09 | 0.1019 | New |
| 469 | 6 | 6-168892201-GGGGCAGCCCGAGACTCAGGCTCAGGTTTTCCACCTGGACCTTGGAAAGAGACATCACACAGCTCAGCGTCCAGGAGGC-G | rs1562494 | AL513210.2, AL513210.1 | 0.2145 | G | | 1.98E-14 | ?+ | 1.981E-14 | | New |
| 470 | 6 | 6-169781694-G-T | rs1410200 | ERMARD | 0.0011 | G | 0.149375 | 1.25E-08 | -+ | 0.0735 | | New |
| 471 | 6 | 6-170054847-TC-T | rs2021081 | AL049612.1, AL603783.1 | 0.0041 | T | 0.941482 | 1.08E-14 | ++ | 1.256E-14 | | New |
| 472 | 6 | 6-21002980-C-CT | rs1295446 | CDKAL1 | 0.0029 | CT | | 4.83E-18 | ?+ | 4.832E-18 | 0.5454 | New |
| 473 | 6 | 6-25902999-AAG-A | | SLC17A3, SLC17A2 | 0.0002 | A | | 4.35E-08 | ?+ | 4.348E-08 | | New |
| 474 | 6 | 6-26311267-C-CAACAACACA | rs1201608 | HIST1H4H, AL021917.1 | 0.0007 | CAACAACACA | | 3E-08 | ?+ | 3.005E-08 | | New |
| 475 | 6 | 6-27577093-C-CTT | rs1348229 | AL021918.3, RNU6-471P | 0.0019 | CTT | | 2.92E-10 | ?+ | 2.92E-10 | | Old |
| 476 | 6 | 6-30621247-C-CAAA | | MRPS18B | 0.0008 | CAAA | | 1.18E-12 | ?+ | 1.178E-12 | | New |

| | | | | | | | | | | | | |
|---|---|---|---|---|---|---|---|---|---|---|---|---|
| 477 | 6 | 6-30753762-GCA-G | | NA | 0.001 | G | | 3.22E-09 | ?+ | 3.221E-09 | | New |
| 477 | 6 | 6-30753765-AGG-A | | HCG20 | 0.0011 | AGG | | 4.67E-09 | ?+ | 4.671E-09 | | New |
| 478 | 6 | 6-31808780-C-CCT | rs1315875 | HSPA1L | 0.0015 | C | | 7.16E-13 | ?+ | 7.159E-13 | 0.7632 | Old |
| 479 | 6 | 6-32446254-G-A | rs9485011 | HLA-DRA, HLA-DRB5 | 0.0004 | G | 0.842475 | 2.27E-08 | -+ | 0.00001192 | 0.8936 | Old |
| 480 | 6 | 6-34044161-TAC-T | | GRM4 | 0.2762 | TAC | | 3.09E-08 | ?+ | 3.094E-08 | | New |
| 481 | 6 | 6-34212899-TTTTG-T | rs1177909 | GRM4, HMGA1 | 0.0039 | T | | 2.97E-29 | ?+ | 2.966E-29 | 0.7905 | New |
| 482 | 6 | 6-34702668-G-A | rs9319062 | AL451165.2, SNRPC | 0.0013 | A | 0.167082 | 6.63E-14 | -+ | 6.579E-09 | 0.422 | New |
| 483 | 6 | 6-34764875-G-C | | SNRPC | 0.0006 | G | 0.657242 | 1.47E-11 | ++ | 3.448E-07 | | New |
| 484 | 6 | 6-41604777-C-T | rs1397842 | FOXP4, MDFI | 0.0014 | C | 0.948278 | 9.29E-13 | ++ | 2.135E-08 | 0.07667 | Old |
| 484 | 6 | 6-41604785-G-A | rs9302597 | FOXP4, MDFI | 0.0012 | A | 0.695541 | 1.72E-08 | -+ | 2.866E-07 | 0.6658 | Old |
| 484 | 6 | 6-41604799-G-A | rs1878780 | FOXP4, MDFI | 0.0031 | G | 0.632177 | 8.05E-10 | -+ | 0.000002156 | 0.0283 | Old |
| 484 | 6 | 6-41604818-C-A | rs5445366 | FOXP4, MDFI | 0.0077 | C | 0.712866 | 6.64E-13 | -+ | 0.000009985 | 0.2837 | Old |
| 485 | 6 | 6-41918284-ACT-A | rs1491105 | MED20 | 0.0037 | A | 0.755137 | 7.45E-13 | -+ | 1.406E-10 | 0.8851 | New |
| 486 | 6 | 6-42018709-T-A | rs1483765 | CCND3 | 0.0012 | A | 0.720039 | 1.82E-14 | -+ | 1.668E-10 | | New |
| 487 | 6 | 6-43022229-GC-G | rs1259726 | RRP36 | 0.0018 | G | 0.863369 | 4.94E-11 | -+ | 0.000002008 | 0.3737 | New |
| 488 | 6 | 6-43022821-C-T | rs5283276 | RRP36 | 0.0113 | C | 0.362499 | 4.81E-10 | -+ | 0.02931 | | New |
| 488 | 6 | 6-43022827-C-T | rs5483812 | RRP36 | 0.0114 | C | 0.362499 | 3.48E-09 | -+ | 0.0366 | 0.007352 | New |
| 489 | 6 | 6-43329675-A-G | rs7766978 | ZNF318 | 0.0187 | G | 0.995788 | 1.76E-12 | ++ | 1.646E-10 | 0.1203 | New |
| 490 | 6 | 6-43534819-GTA-G | | NA | 0.0009 | GTA | | 1.05E-08 | ?+ | 1.047E-08 | | New |
| 491 | 6 | 6-43738216-C-T | rs1122600 | AL136131.2 | 0.0227 | T | | 1.24E-16 | ?+ | 1.24E-16 | | New |
| 492 | 6 | 6-4695703-T-C | rs6915530 | AL034376.1, CDYL | 0.0121 | T | 0.41591 | 1.14E-08 | -+ | 0.001588 | | New |
| 493 | 6 | 6-61313986-G-T | rs1381355 | AL512427.2, KHDRBS2-OT | 0.001 | T | 0.755228 | 4.48E-08 | ++ | 0.000001493 | | New |
| 494 | 6 | 6-66722870-G-T | | EYS, RNU7-66P | 0.0005 | G | | 4.45E-10 | ?+ | 4.452E-10 | | New |
| 495 | 6 | 6-6716029-C-CTT | | NA | 0.0014 | CTT | | 1.25E-10 | ?+ | 1.252E-10 | | New |

| | | | | | | | | | | | | |
|---|---|---|---|---|---|---|---|---|---|---|---|---|
| 496 | 6 | 6-7055607-G-A | rs6239357 | AL139390.1, RREB1 | 0.2366 | A | | 2.58E-08 | ?+ | 2.578E-08 | 0.3832 | New |
| 497 | 6 | 6-73319458-C-CAA | | NA | 0.0029 | C | | 2.65E-13 | ?+ | 2.647E-13 | | New |
| 497 | 6 | 6-73319481-G-A | rs1283541 | KHDC1, DPPA5 | 0.0007 | A | 0.197459 | 1.07E-09 | -+ | 4.822E-08 | | New |
| 498 | 6 | 6-7382355-TG-T | rs1332897 | CAGE1 | 0.0017 | TG | | 2.25E-10 | ?+ | 2.253E-10 | 0.3466 | New |
| 499 | 6 | 6-88001571-GT-G | | RN7SL183P, AL136096.1 | 0.0004 | GT | | 1.89E-08 | ?+ | 1.887E-08 | | New |
| 500 | 7 | 7-100284341-C-A | rs1181626 | CASTOR3, SPDYE3 | 0.0004 | C | 0.452824 | 2.15E-08 | -+ | 0.00008595 | | Old |
| 501 | 7 | 7-100672357-C-CCT | rs1270383 | ACTL6B, GNB2 | 0.0043 | C | | 6.27E-18 | ?+ | 6.27E-18 | 0.1064 | Old |
| 502 | 7 | 7-101035140-T-A | rs1026527 | MUC17 | 0.1869 | A | | 3.81E-09 | ?+ | 3.814E-09 | 0.5325 | Old |
| 503 | 7 | 7-101284191-GAT-G | | NA | 0.0003 | G | | 1.02E-08 | ?+ | 1.017E-08 | | New |
| 503 | 7 | 7-101284194-GCA-G | | NA | 0.0003 | GCA | | 2.82E-09 | ?+ | 2.818E-09 | | New |
| 504 | 7 | 7-101788981-T-A | rs8912801 | MYL10, CUX1 | 0.0018 | T | 0.868141 | 1.49E-16 | -+ | 4.866E-12 | | New |
| 505 | 7 | 7-101822055-C-T | rs1032695 | CUX1 | 0.0009 | C | 0.702338 | 1.37E-09 | ++ | 8.369E-07 | | New |
| 506 | 7 | 7-102026581-AG-A | | NA | 0.0005 | A | | 3.01E-09 | ?+ | 3.007E-09 | | New |
| 507 | 7 | 7-102920935-C-CCT | | FBXL13, LRRC17 | 0.0013 | CCT | | 2.05E-08 | ?+ | 2.054E-08 | | New |
| 508 | 7 | 7-105596291-C-T | rs9470782 | EFCAB10 | 0.0026 | T | 0.842782 | 8.52E-17 | -+ | 2.646E-08 | 0.9001 | New |
| 509 | 7 | 7-1066816-GCACCCCAGAGGTGAGGGTTTGGGGCACAGTCTGTTGGCGGAGGCAGGAGTA-G | rs1563072 | C7orf50 | 0.0234 | G | | 1.01E-08 | ?+ | 1.006E-08 | | New |
| 510 | 7 | 7-1115481-GGAT-G | rs1433330 | C7orf50 | 0.0031 | GGAT | 0.687714 | 4.58E-08 | ++ | 9.507E-07 | | New |
| 511 | 7 | 7-111925911-C-A | rs5749957 | DOCK4 | 0.0017 | A | 0.419217 | 4.93E-12 | ++ | 1.222E-09 | 0.9477 | New |
| 511 | 7 | 7-111925913-C-A | rs5359435 | DOCK4 | 0.0016 | A | 0.3947 | 7.64E-13 | ++ | 3.086E-10 | 0.6831 | New |
| 512 | 7 | 7-116919680-ATG-A | rs1414168 | CAPZA2 | 0.0054 | ATG | | 4.06E-10 | ?+ | 4.061E-10 | | New |
| 513 | 7 | 7-117106822-G-T | rs1292647 | ST7-AS2 | 0.0021 | T | 0.680421 | 8.95E-09 | -+ | 0.0008444 | | New |

| | | | | | | | | | | | | |
|---|---|---|---|---|---|---|---|---|---|---|---|---|
| 514 | 7 | 7-1216367-G-T | rs9976708 | AC091729.3, UNCX | 0.0023 | T | 0.155797 | 3.9E-08 | -+ | 0.0004272 | | New |
| 515 | 7 | 7-126391942-T-A | rs5627886 | AC000372.1 | 0.0025 | A | 0.077793 | 1.79E-16 | -+ | 0.000001857 | | New |
| 515 | 7 | 7-126391976-G-A | rs1045685 | AC000372.1 | 0.0019 | G | 0.558351 | 7.49E-17 | -+ | 3.73E-08 | | New |
| 516 | 7 | 7-128518959-G-A | | METTL2B, AC090114.2 | 0.0005 | G | 0.620633 | 2.76E-09 | -+ | 0.00001635 | 0.1732 | New |
| 517 | 7 | 7-128552494-G-T | | AC090114.2, AC108010.1 | 0.0007 | G | | 8.05E-11 | ?+ | 8.054E-11 | | New |
| 518 | 7 | 7-129358248-C-T | rs8919542 | AHCYL2 | 0.0017 | T | 0.676651 | 2.43E-10 | ++ | 4.675E-09 | | New |
| 519 | 7 | 7-130087903-GGT-G | rs1235961 | KLHDC10 | 0.0018 | G | | 1.15E-09 | ?+ | 1.149E-09 | 0.1943 | New |
| 519 | 7 | 7-130087910-GCA-G | rs1347602 | KLHDC10 | 0.0015 | GCA | | 2.52E-08 | ?+ | 2.523E-08 | | New |
| 520 | 7 | 7-13820977-C-T | rs1421858 | AC011287.1, AC005019.2 | 0.0008 | T | 0.881387 | 3.74E-08 | ++ | 4.631E-08 | | New |
| 520 | 7 | 7-13820985-C-T | rs1412811 | AC011287.1, AC005019.2 | 0.001 | T | | 3.54E-14 | ?+ | 3.541E-14 | | New |
| 521 | 7 | 7-138674703-AAT-A | | NA | 0.0006 | AAT | | 1.63E-08 | ?+ | 1.634E-08 | | New |
| 522 | 7 | 7-140480517-GA-G | rs1455936 | MKRN1, DENND2A | 0.0007 | G | | 2.94E-10 | ?+ | 2.939E-10 | | New |
| 523 | 7 | 7-140498699-C-A | rs9183933 | MKRN1, DENND2A | 0.0018 | A | | 1.38E-21 | ?+ | 1.377E-21 | | New |
| 524 | 7 | 7-140619733-C-CAA | | NA | 0.0008 | C | | 2.11E-11 | ?+ | 2.114E-11 | | New |
| 525 | 7 | 7-140944263-AT-A | rs1292982 | BRAF, MRPS33 | 0.003 | AT | 0.994039 | 2.19E-17 | -+ | 2.264E-15 | | New |
| 526 | 7 | 7-140961641-C-CAA | | NA | 0.0008 | CAA | | 5.77E-12 | ?+ | 5.765E-12 | | New |
| 527 | 7 | 7-148956743-AGG-A | | NA | 0.0005 | A | | 7.74E-10 | ?+ | 7.74E-10 | | New |
| 528 | 7 | 7-148989806-T-A | | GHET1 | 0.001 | A | 0.051067 | 2.77E-15 | ++ | 4.624E-16 | | New |

| | | | | | | | | | | | | |
|---|---|---|---|---|---|---|---|---|---|---|---|---|
| 529 | 7 | 7-149095930-C-CCT | | NA | 0.0013 | CCT | | 2.16E-12 | ?+ | 2.155E-12 | | New |
| 529 | 7 | 7-149095962-G-C | rs9659130 | ZNF786, ZNF425 | 0.001 | C | 0.232213 | 5.8E-09 | ++ | 0.00000735 | | New |
| 530 | 7 | 7-149904512-C-CA | rs1159657 | AC092681.3, AC092681.2 | 0.0022 | CA | | 3.53E-09 | ?+ | 3.53E-09 | | New |
| 531 | 7 | 7-152402297-C-CAATCATA | rs7126032 | KMT2C | 0.0025 | CAATCATA | | 5.98E-13 | ?+ | 5.976E-13 | | New |
| 532 | 7 | 7-152546811-C-CA | rs1165639 | LINC01003, RNA5SP250 | 0.0014 | C | | 3.87E-09 | ?+ | 3.865E-09 | | New |
| 533 | 7 | 7-152617758-C-T | rs1385078 | RF00568, XRCC2 | 0.0011 | C | 0.492947 | 1.49E-09 | ++ | 1.378E-09 | 0.3217 | New |
| 533 | 7 | 7-152617778-AT-A | rs2116963 | NA | 0.001 | A | 0.045143 | 3.09E-08 | ++ | 0.000000381 | | New |
| 534 | 7 | 7-155134673-C-CCT | | NA | 0.0011 | CCT | | 2.44E-08 | ?+ | 2.441E-08 | | Old |
| 535 | 7 | 7-157725208-GC-G | rs7466937 | PTPRN2 | 0.0097 | G | | 3.01E-09 | ?+ | 3.012E-09 | | New |
| 536 | 7 | 7-158045747-C-A | rs1585262 | PTPRN2 | 0.0003 | A | | 1.4E-08 | ?+ | 1.403E-08 | | New |
| 537 | 7 | 7-158084639-ACGACACTCATCCACATCCT-A | rs1387462 | PTPRN2 | 0.0017 | A | | 4.38E-09 | ?+ | 4.376E-09 | | New |
| 538 | 7 | 7-158319527-T-TCA | | PTPRN2 | 0.1934 | TCA | | 3.16E-08 | ?+ | 3.164E-08 | | New |
| 539 | 7 | 7-158463746-C-CA | rs1349126 | PTPRN2 | 0.0062 | C | | 7.97E-15 | ?+ | 7.973E-15 | 0.01274 | New |
| 540 | 7 | 7-158493344-TACAC-T | rs1348179 | PTPRN2 | 0.0054 | T | 0.47237 | 5.59E-10 | -+ | 0.0002393 | | New |
| 541 | 7 | 7-1765054-C-T | | ELFN1, MAD1L1 | 0.0005 | C | 0.9368 | 2.75E-12 | ++ | 1.12E-09 | | New |
| 542 | 7 | 7-222020-C-CCG | rs1350645 | FAM20C | 0.0122 | C | | 3.95E-24 | ?+ | 3.948E-24 | | New |
| 543 | 7 | 7-2296752-G-T | rs1208805 | SNX8 | 0.0011 | G | 0.029002 | 1.1E-08 | ++ | 1.865E-09 | | New |
| 544 | 7 | 7-2455679-G-C | rs1778672 | CHST12, GRIFIN | 0.0007 | G | 0.330179 | 3.72E-10 | ++ | 2.95E-10 | | New |
| 544 | 7 | 7-2455697-G-C | rs9392520 | CHST12, GRIFIN | 0.0016 | G | 0.203285 | 2.68E-16 | -+ | 1.904E-11 | | New |
| 545 | 7 | 7-26252136-C-CCT | | NA | 0.0003 | CCT | | 8.19E-09 | ?+ | 8.192E-09 | | New |
| 546 | 7 | 7-3030002-G-C | rs9466886 | CARD11 | 0.0008 | G | 0.64577 | 3.35E-08 | ++ | 7.643E-07 | 0.1039 | New |

| | | | | | | | | | | | | |
|---|---|---|---|---|---|---|---|---|---|---|---|---|
| 547 | 7 | 7-3186156-G-T | rs1122364 | AC073316.1, AC073316.2 | 0.0016 | T | 0.020147 | 1.81E-14 | ++ | 3.539E-13 | | New |
| 548 | 7 | 7-32888710-ATTACTTG-A | | NA | 0.0002 | ATTACTTG | | 2.77E-08 | ?+ | 2.772E-08 | | New |
| 549 | 7 | 7-36704566-G-T | rs7724406 | AOAH | 0.0001 | T | 0.075183 | 3.42E-08 | ++ | 2.625E-07 | | New |
| 550 | 7 | 7-37456405-G-A | rs1028428 | ELMO1, GPR141 | 0.0015 | A | 0.615249 | 1.11E-08 | ++ | 0.00009848 | | Old |
| 551 | 7 | 7-4276876-GGA-G | | NA | 0.0006 | G | | 4.14E-09 | ?+ | 4.142E-09 | | New |
| 552 | 7 | 7-443821-GGGAT-G | | AC188617.1, AC188617.2 | 0.3181 | GGGAT | 0.996808 | 4.41E-08 | -+ | 0.002096 | | New |
| 553 | 7 | 7-44676427-C-T | rs9466922 | OGDH | 0.0006 | T | 0.054015 | 1.88E-10 | -+ | 0.00002811 | | New |
| 554 | 7 | 7-44916280-G-C | | AC004854.2, AC004847.1 | 0.0007 | G | 0.179336 | 2.05E-08 | -+ | 0.00371 | | New |
| 554 | 7 | 7-44916283-C-A | | AC004854.2, AC004847.1 | 0.0008 | A | 0.179098 | 7.8E-13 | -+ | 0.000009981 | | New |
| 554 | 7 | 7-44916302-AT-A | | AC004854.2, AC004847.1 | 0.0009 | A | 0.335755 | 2.58E-10 | ++ | 4.835E-07 | 0.5023 | New |
| 555 | 7 | 7-470179-C-CCCA | rs5487141 | AC188617.2, AC147651.3 | 0.0116 | C | 0.948577 | 4.82E-11 | -+ | 0.00001085 | | New |
| 556 | 7 | 7-47554393-GGGT-G | | NA | 0.0004 | G | | 2.88E-09 | ?+ | 2.882E-09 | | New |
| 557 | 7 | 7-47925128-TGGAAAAAAAAA-T | | NA | 0.0004 | T | 0.208106 | 2.58E-09 | -+ | 3.662E-08 | | New |
| 558 | 7 | 7-48002361-C-T | rs1789406 | SUN3 | 0.0008 | C | | 4.68E-09 | ?+ | 4.68E-09 | | New |
| 559 | 7 | 7-4896984-G-C | rs1784729 | RADIL, MMD2 | 0.0001 | C | | 3.14E-08 | ?+ | 3.14E-08 | | New |

| | | | | | | | | | | | | |
|---|---|---|---|---|---|---|---|---|---|---|---|---|
| 560 | 7 | 7-49633385-C-CACAG | rs1232901 | AC092448.1, AC093775.1 | 0.0008 | CACAG | 0.008806 | 1.37E-08 | ++ | 2.713E-09 | | New |
| 561 | 7 | 7-51057333-TAC-T | | COBL | 0.3701 | T | | 2.31E-08 | ?+ | 2.306E-08 | | New |
| 562 | 7 | 7-5140813-G-A | rs5753956 | RBAKDN, RNU6-215P | 0.0013 | A | 0.1929 | 6.55E-11 | -+ | 0.000008492 | 0.7844 | New |
| 562 | 7 | 7-5140818-G-A | rs1388898 | RBAKDN, RNU6-215P | 0.0012 | A | 0.699399 | 6.03E-12 | ++ | 2.322E-10 | 0.5743 | New |
| 563 | 7 | 7-5515904-C-T | rs1784644 | FBXL18, ACTB | 0.0005 | T | 0.315853 | 2.92E-11 | -+ | 0.000001764 | | New |
| 564 | 7 | 7-5535600-C-CGGGTTCA | rs1390981 | ACTB | 0.0004 | C | 0.580122 | 5.01E-09 | -+ | 0.000001421 | | New |
| 565 | 7 | 7-5564808-G-A | rs1206340 | ACTB, FSCN1 | 0.0015 | G | | 5.42E-13 | ?+ | 5.419E-13 | 0.574 | New |
| 566 | 7 | 7-55835419-G-T | rs7746985 | SEPT14 | 0.0021 | G | 0.531286 | 1.72E-19 | ++ | 2.789E-17 | 0.6825 | New |
| 567 | 7 | 7-57783552-C-T | rs1431804 | ZNF716, NONE | 0.0046 | T | 0.044829 | 4.03E-11 | -- | 1.745E-11 | | New |
| 568 | 7 | 7-58060060-TTTG-T | | NA | 0.004 | T | | 4.43E-14 | ?+ | 4.429E-14 | | New |
| 568 | 7 | 7-58060068-TTC-T | | NA | 0.0032 | T | | 3.24E-11 | ?+ | 3.237E-11 | 0.5622 | New |
| 569 | 7 | 7-58060792-TTCTATTC-T | | NA | 0.0043 | TTCTATTC | | 4.32E-30 | ?+ | 4.324E-30 | | New |
| 570 | 7 | 7-5856871-AT-A | | NA | 0.0011 | AT | | 1.63E-13 | ?+ | 1.629E-13 | | New |
| 571 | 7 | 7-606603-G-A | rs1398858 | AC147651.4 | 0.0015 | A | 0.215617 | 2.99E-17 | -+ | 1.163E-07 | 0.04678 | New |
| 572 | 7 | 7-62337609-T-A | rs2006289 | NONE, RNU6-417P | 0.0003 | A | | 8.39E-09 | ?+ | 8.391E-09 | | New |
| 573 | 7 | 7-62430395-T-A | rs8790253 | NONE, RNU6-417P | 0.004 | A | 0.75691 | 9.11E-10 | -+ | 4.446E-08 | | New |
| 574 | 7 | 7-65015752-C-T | rs1908903 | ERV3-1, RNU6-1229P | 0.0017 | T | 0.24035 | 1.7E-08 | ++ | 0.000004148 | 0.8373 | New |
| 575 | 7 | 7-65095078-A-AT | | SNORA15B-1, AC104073.4 | 0.0383 | A | | 5.47E-12 | ?+ | 5.466E-12 | | New |

| | | | | | | | | | | | |
|---|---|---|---|---|---|---|---|---|---|---|---|
| 576 | 7 | 7-65711737-GAC-G | | NA | 0.0006 | G | | 9.15E-14 | ?+ | 9.146E-14 | | New |
| 577 | 7 | 7-65862446-AAAG-A | rs1168592 | RNU6-973P, VKORC1L1 | 0.0049 | A | 0.98983 | 1.04E-09 | -+ | 0.01029 | 0.8341 | New |
| 578 | 7 | 7-66026675-TAA-T | rs1402152 | GUSB, ASL | 0.0018 | T | | 6.51E-13 | ?+ | 6.512E-13 | | New |
| 579 | 7 | 7-66707015-GACCT-G | rs1185379 | AC027644.4, RABGEF1 | 0.0026 | GACCT | | 6.81E-09 | ?+ | 6.81E-09 | 0.7985 | New |
| 580 | 7 | 7-67109039-C-T | rs1286512 | TYW1 | 0.0005 | T | 0.893253 | 2.32E-09 | ++ | 0.000003789 | 0.9407 | New |
| 580 | 7 | 7-67109054-ATT-A | | NA | 0.0013 | ATT | 0.83812 | 1.55E-13 | ++ | 9.376E-12 | | New |
| 581 | 7 | 7-68108631-C-CGACACAGT | | NA | 0.0007 | CGACACAGT | | 1.2E-12 | ?+ | 1.201E-12 | | New |
| 582 | 7 | 7-6869364-G-C | rs1249014 | CCZ1B, AC079804.3 | 0.0008 | G | 0.118979 | 2.55E-08 | ++ | 6.205E-08 | 0.1122 | New |
| 583 | 7 | 7-69807598-C-A | | AUTS2 | 0.0004 | A | | 6.1E-11 | ?+ | 6.094E-11 | | New |
| 583 | 7 | 7-69807601-C-CCT | | NA | 0.0011 | C | | 4.66E-10 | ?+ | 4.66E-10 | | New |
| 584 | 7 | 7-70395262-G-C | rs5494273 | AUTS2 | 0.0033 | C | 0.155739 | 2.03E-08 | ++ | 0.00008184 | 0.6265 | New |
| 585 | 7 | 7-71010361-TGG-T | | NA | 0.0004 | T | | 1.24E-09 | ?+ | 1.243E-09 | | New |
| 586 | 7 | 7-72703259-C-CAA | | NA | 0.0006 | CAA | | 2.99E-10 | ?+ | 2.988E-10 | | New |
| 587 | 7 | 7-73239519-TA-T | | NA | 0.0011 | TA | | 3.37E-15 | ?+ | 3.371E-15 | | New |
| 587 | 7 | 7-73239535-AGC-A | | NA | 0.0001 | A | | 2.07E-08 | ?+ | 2.071E-08 | | New |
| 588 | 7 | 7-73415961-C-CTCT | | NA | 0.0023 | CTCT | | 2.36E-13 | ?+ | 2.358E-13 | | New |
| 589 | 7 | 7-73481896-T-A | rs1789216 | BAZ1B | 0.0015 | A | 0.693204 | 4.3E-12 | ++ | 0.000003293 | 0.4693 | New |
| 589 | 7 | 7-73481899-C-T | rs5685877 | BAZ1B | 0.0015 | T | 0.564469 | 2.12E-14 | ++ | 1.281E-07 | 0.4197 | New |
| 589 | 7 | 7-73481926-C-T | rs1554573 | BAZ1B | 0.0011 | C | 0.92193 | 4.93E-14 | -+ | 6.822E-09 | 0.8207 | New |
| 590 | 7 | 7-73619442-C-CAG | rs1554601 | MLXIPL | 0.0065 | C | | 6.63E-21 | ?+ | 6.626E-21 | | New |
| 590 | 7 | 7-73619449-C-CAA | rs1554601 | MLXIPL | 0.0054 | CAA | | 4.62E-18 | ?+ | 4.618E-18 | | New |
| 591 | 7 | 7-73763277-G-A | rs1477643 | ABHD11, CLDN3 | 0.0001 | A | 0.814753 | 7.43E-09 | ++ | 4.025E-07 | | New |
| 592 | 7 | 7-74071734-AAGTG-A | | NA | 0.0006 | AAGTG | | 5.87E-09 | ?+ | 5.872E-09 | | New |
| 593 | 7 | 7-74300691-C-CAA | rs1554726 | CLIP2 | 0.0007 | CAA | | 1E-09 | ?+ | 1.001E-09 | 0.4488 | New |
| 594 | 7 | 7-74303748-C-CT | | NA | 0.0007 | C | | 3.61E-08 | ?+ | 3.608E-08 | | New |
| 595 | 7 | 7-74493235-G-A | rs9287852 | GTF2IRD1 | 0.0029 | A | 0.444242 | 1.6E-16 | -+ | 0.00001141 | 0.3064 | New |
| 595 | 7 | 7-74493250-C-T | rs1554337 | GTF2IRD1 | 0.0024 | C | 0.727944 | 1.51E-25 | -+ | 9.657E-11 | 0.2411 | New |
| 596 | 7 | 7-74792891-TA-T | | NA | 0.0007 | TA | | 4.73E-12 | ?+ | 4.732E-12 | | New |

| | | | | | | | | | | | | |
|---|---|---|---|---|---|---|---|---|---|---|---|---|
| 597 | 7 | 7-74999145-G-A | rs1379444 | CASTOR2 | 0.0004 | A | 0.244633 | 3.12E-09 | -+ | 0.0001126 | | New |
| 598 | 7 | 7-75607689-GT-G | | NA | 0.0028 | GT | | 4E-32 | ?+ | 4.002E-32 | | New |
| 598 | 7 | 7-75607691-AC-A | | NA | 0.0014 | A | | 5.72E-21 | ?+ | 5.718E-21 | | New |
| 599 | 7 | 7-75792984-ATG-A | rs1554531 | CCL26, CCL24 | 0.0015 | A | | 4.68E-16 | ?+ | 4.677E-16 | 0.3224 | New |
| 599 | 7 | 7-75792987-C-CAT | rs1554531 | CCL26, CCL24 | 0.0016 | CAT | | 8.47E-17 | ?+ | 8.47E-17 | 0.1902 | New |
| 600 | 7 | 7-75801743-G-T | rs1554532 | CCL26, CCL24 | 0.0003 | T | | 4.28E-08 | ?+ | 4.284E-08 | | New |
| 600 | 7 | 7-75801752-ATC-A | | NA | 0.0002 | A | | 6.82E-10 | ?+ | 6.816E-10 | | New |
| 601 | 7 | 7-75853377-GTTT-G | | NA | 0.0005 | G | | 4.71E-08 | ?+ | 4.714E-08 | | New |
| 602 | 7 | 7-89881278-AT-A | | NA | 0.0004 | A | | 1.56E-14 | ?+ | 1.565E-14 | 0.4501 | New |
| 602 | 7 | 7-89881280-C-CCTGGCTAACAT | | NA | 0.0003 | C | | 3.96E-10 | ?+ | 3.955E-10 | | New |
| 603 | 7 | 7-97243876-TCCAC-T | rs1240859 | SDHAF3, RN7SKP104 | 0.0046 | TCCAC | | 1.33E-08 | ?+ | 1.332E-08 | | New |
| 604 | 8 | 8-101230534-TG-T | | NA | 0.0016 | TG | | 1.81E-08 | ?+ | 1.812E-08 | 0.9034 | New |
| 605 | 8 | 8-102240933-G-A | rs1313099 | UBR5-AS1 | 0.0011 | G | 0.543149 | 4.23E-11 | -+ | 1.911E-07 | 0.9872 | New |
| 606 | 8 | 8-102887720-ATTT-A | | NA | 0.0021 | A | | 6.83E-09 | ?+ | 6.828E-09 | | New |
| 607 | 8 | 8-104680838-GGAGTTC-G | | NA | 0.0014 | GGAGTTC | | 2.62E-09 | ?+ | 2.622E-09 | 0.1332 | New |
| 608 | 8 | 8-125402911-AAG-A | rs1246130 | NSMCE2, TRIB1 | 0.0006 | A | | 2.88E-08 | ?+ | 2.885E-08 | 0.1455 | New |
| 609 | 8 | 8-127843609-AT-A | rs5326479 | PVT1 | 0.002 | A | 0.945532 | 3.58E-12 | ++ | 0.00001471 | | New |
| 610 | 8 | 8-130031600-C-CT | rs1370965 | FAM49B, ASAP1 | 0.0039 | CT | | 1.72E-13 | ?+ | 1.724E-13 | 0.3025 | New |
| 611 | 8 | 8-1316426-C-CTCTCCAACAGTGGTCTACACTCGAGAAACTCGGCAGCTTTTAAAAATAGAGCGTGTGCGAGTGCAGCGTCTCTCCAACAGTGGTCTACACTCGAGACACTCGGCAGCGTTTAAAAATAGAGGCTGTGCGAGTGCAGCG | rs1563078 | DLGAP2 | 0.0243 | C | | 4.61E-09 | ?- | 4.605E-09 | | New |

| | | | | | | | | | | | | |
|---|---|---|---|---|---|---|---|---|---|---|---|---|
| 612 | 8 | 8-13268436-TC-T | | NA | 0.0019 | T | | 7.18E-15 | ?+ | 7.18E-15 | | New |
| 613 | 8 | 8-134499573-GGATGCCCCCGCTGCTGGTTACACACAGAGCCTGATTTGGGAGGGTCGGGGTGGAGCCGT-G | rs1818800 | ZFAT | 0.004 | G | | 1.16E-14 | ?+ | 1.155E-14 | | New |
| 614 | 8 | 8-138085971-G-A | rs9799868 | AC046195.1, FAM135B | 0.0015 | A | 0.857856 | 8.54E-09 | -+ | 0.001963 | | New |
| 615 | 8 | 8-143960466-AT-A | | NA | 0.0028 | AT | | 2.76E-22 | ?+ | 2.765E-22 | | Old |
| 616 | 8 | 8-144124603-C-CA | rs1374854 | WDR97, HGH1 | 0.0024 | C | | 2E-22 | ?+ | 1.996E-22 | | Old |
| 617 | 8 | 8-144216367-G-A | rs1461922 | MROH1 | 0.0707 | G | | 5.92E-10 | ?+ | 5.924E-10 | 0.6639 | Old |
| 618 | 8 | 8-144226688-G-A | rs9098193 | MROH1 | 0.0005 | G | 0.821745 | 1.39E-08 | -+ | 0.00001485 | | Old |
| 619 | 8 | 8-144301660-G-T | | HSF1 | 0.0002 | T | | 4.33E-08 | ?+ | 4.325E-08 | | Old |
| 620 | 8 | 8-144693358-C-CAGCCGCCT | rs1430358 | RF00017, AF186192.1 | 0.0049 | CAGCCGC( | 0.695519 | 7.86E-09 | -+ | 0.00001591 | | New |
| 621 | 8 | 8-1603569-T-TGGGTCTCAGTTCTGCAGAGGCTGGTTAGAGTGGAGGTG | rs7119074 | DLGAP2-AS1 | 0.0438 | TGGGTCTCAGTTCTGC. | | 1.7E-10 | ?+ | 1.698E-10 | | New |
| 622 | 8 | 8-23267200-AT-A | rs1200144 | CHMP7, R3HCC1 | 0.0022 | AT | | 1.35E-12 | ?+ | 1.353E-12 | 0.8416 | New |
| 623 | 8 | 8-23279493-TGG-T | rs1802665 | R3HCC1 | 0.0012 | TGG | | 7.27E-09 | ?+ | 7.269E-09 | | New |
| 624 | 8 | 8-30679870-TC-T | | NA | 0.0011 | TC | | 9.69E-13 | ?+ | 9.69E-13 | | New |
| 625 | 8 | 8-33468388-TG-T | rs1456752 | FUT10 | 0.0014 | T | 0.679781 | 3.41E-12 | -+ | 1.544E-10 | 0.7119 | New |
| 626 | 8 | 8-33494173-AAG-A | | NA | 0.0003 | A | | 3.09E-11 | ?+ | 3.085E-11 | | New |
| 626 | 8 | 8-33494177-ACC-A | | NA | 0.0002 | ACC | | 7.08E-11 | ?+ | 7.076E-11 | | New |
| 627 | 8 | 8-38194649-C-CAG | | NA | 0.0011 | CAG | 0.287517 | 1.21E-13 | -+ | 7.418E-11 | | New |
| 628 | 8 | 8-38390825-GGT-G | | LETM2 | 0.0013 | G | | 6.01E-10 | ?+ | 6.014E-10 | | New |
| 629 | 8 | 8-40820780-C-T | rs1142365 | ZMAT4 | 0.0217 | C | | 1.98E-19 | ?- | 1.983E-19 | | New |
| 630 | 8 | 8-43008878-C-T | rs1179689 | HOOK3 | 0.0011 | T | 0.165033 | 5.01E-15 | ++ | 1.899E-15 | | New |
| 630 | 8 | 8-43008901-C-T | rs8675903 | HOOK3 | 0.0006 | T | 0.884979 | 2.54E-08 | ++ | 1.864E-07 | 0.1861 | New |
| 631 | 8 | 8-43570083-G-C | rs1009786 | RF00012, NONE | 0.1758 | G | | 8.1E-10 | ?+ | 8.103E-10 | | New |
| 632 | 8 | 8-43927222-T-A | rs1470310 | RF00012, NONE | 0.0012 | A | 0.143375 | 1.54E-12 | ++ | 9.141E-11 | 0.4626 | New |

| | | | | | | | | | | | | |
|---|---|---|---|---|---|---|---|---|---|---|---|---|
| 633 | 8 | 8-47081758-G-A | | RN7SKP32, AC120036.5 | 0.001 | G | | 2.42E-08 | ?+ | 2.42E-08 | | New |
| 634 | 8 | 8-48012945-AAG-A | rs1452849 | UBE2V2 | 0.0015 | A | | 4.29E-08 | ?+ | 4.286E-08 | 0.3315 | New |
| 635 | 8 | 8-48019002-G-T | rs7716563 | UBE2V2 | 0.0003 | T | | 1.89E-10 | ?+ | 1.886E-10 | | New |
| 636 | 8 | 8-55827470-AACGCCATTCTCCTGCCTCAGCCTCCCGAGTAGCTG-A | | NA | 0.0023 | A | | 3.9E-11 | ?+ | 3.898E-11 | | New |
| 637 | 8 | 8-60523306-C-CAT | | RAB2A | 0.0004 | CAT | | 3.74E-08 | ?+ | 3.743E-08 | | New |
| 638 | 8 | 8-66557723-GA-G | | AC009879.3, MYBL1 | 0.0007 | GA | 0.436892 | 1.01E-08 | ++ | 0.000002625 | | New |
| 639 | 8 | 8-66708539-C-A | rs1137715 | SGK3 | 0.1291 | A | | 1.41E-08 | ?+ | 1.405E-08 | 0.4419 | New |
| 640 | 8 | 8-67370567-C-CCA | rs1388282 | AC021321.1, CPA6 | 0.0017 | CCA | 0.18898 | 2.07E-12 | ++ | 1.058E-12 | | New |
| 641 | 8 | 8-7412675-CT-C | | ZNF705G, DEFB4B | 0.426 | C | | 2.89E-22 | ?+ | 2.893E-22 | | New |
| 642 | 8 | 8-80570527-GC-G | | RNU7-174P, RNU2-71P | 0.0014 | G | | 3.46E-16 | ?+ | 3.464E-16 | | Old |
| 643 | 8 | 8-90506149-C-A | rs1329444 | AC004083.1, LINC00534 | 0.0021 | A | 0.9022 | 2.95E-22 | -+ | 3.043E-19 | | New |
| 643 | 8 | 8-90506156-C-T | rs1008588 | AC004083.1, LINC00534 | 0.0009 | T | 0.532693 | 5.8E-09 | ++ | 9.776E-09 | 0.9525 | New |
| 644 | 8 | 8-90875247-C-T | rs1218355 | NECAB1 | 0.0013 | T | 0.326151 | 2.01E-08 | ++ | 0.000001009 | 0.02578 | New |
| 645 | 8 | 8-93926331-C-CA | rs1470817 | PDP1 | 0.002 | CA | | 1.41E-09 | ?+ | 1.407E-09 | 0.4818 | New |
| 645 | 8 | 8-93926333-GCAT-G | rs1168533 | PDP1 | 0.0024 | GCAT | | 1.46E-14 | ?+ | 1.46E-14 | 0.3434 | New |
| 646 | 8 | 8-94591747-C-CT | | NA | 0.001 | C | | 4.39E-09 | ?+ | 4.387E-09 | | Old |
| 647 | 8 | 8-9493603-ATT-A | | NA | 0.0004 | ATT | | 7.89E-09 | ?+ | 7.888E-09 | | New |
| 648 | 8 | 8-97732251-ACT-A | rs1172668 | MTDH, RF00019 | 0.0005 | A | | 4.08E-10 | ?+ | 4.076E-10 | | New |
| 649 | 8 | 8-99128207-GGT-G | | NA | 0.002 | GGT | | 1.61E-17 | ?+ | 1.61E-17 | | New |

| | | | | | | | | | | | | |
|---|---|---|---|---|---|---|---|---|---|---|---|---|
| 650 | 9 | 9-104150373-GGC-G | | NA | 0.0009 | G | | 1.77E-09 | ?+ | 1.768E-09 | | New |
| 650 | 9 | 9-104150379-TCC-T | | SMC2, OR13F1 | 0.0009 | TCC | | 2.23E-08 | ?+ | 2.233E-08 | | New |
| 651 | 9 | 9-110014597-AT-A | | NA | 0.0006 | AT | | 1.22E-11 | ?+ | 1.219E-11 | | New |
| 652 | 9 | 9-118668202-C-CAG | | NA | 0.0008 | C | | 1.49E-08 | ?+ | 1.489E-08 | | New |
| 653 | 9 | 9-123399029-C-CAA | rs1314416 | DENND1A | 0.0041 | C | | 2.37E-09 | ?+ | 2.371E-09 | | New |
| 654 | 9 | 9-123706553-C-CCT | | NA | 0.0017 | CCT | | 5.43E-17 | ?+ | 5.427E-17 | | New |
| 655 | 9 | 9-123917954-C-CAAAA | rs1225330 | DENND1A | 0.0054 | C | | 3.66E-10 | ?+ | 3.657E-10 | | New |
| 656 | 9 | 9-125592686-C-CCT | rs1470069 | MAPKAP1 | 0.0024 | CCT | 0.86742 | 1.04E-10 | -+ | 1.382E-09 | | New |
| 657 | 9 | 9-125689462-C-A | rs7796130 | MAPKAP1 | 0.0346 | C | 0.948883 | 4.52E-11 | ++ | 1.085E-10 | | New |
| 658 | 9 | 9-126014359-C-A | rs5537369 | PBX3, AL589923.1 | 0.0011 | A | 0.201084 | 3.98E-10 | ++ | 8.404E-09 | | New |
| 659 | 9 | 9-128665286-C-CAG | | NA | 0.0008 | CAG | | 3.66E-08 | ?+ | 3.659E-08 | | New |
| 660 | 9 | 9-129033772-GAGA-G | | NA | 0.001 | GAGA | | 9.75E-10 | ?+ | 9.751E-10 | | New |
| 661 | 9 | 9-129727696-C-CCT | | NA | 0.0025 | CCT | | 5.56E-23 | ?+ | 5.552E-23 | | New |
| 662 | 9 | 9-129980341-C-CA | | NA | 0.0003 | C | | 2.03E-09 | ?+ | 2.026E-09 | | New |
| 663 | 9 | 9-130955273-C-CACCGTT | rs1156259 | AL161733.1, LAMC3 | 0.0047 | CACCGTT | | 7.89E-10 | ?+ | 7.886E-10 | | New |
| 664 | 9 | 9-131326169-C-T | rs1019641 | PLPP7 | 0.0005 | C | | 2.49E-09 | ?+ | 2.493E-09 | 0.3065 | New |
| 664 | 9 | 9-131326190-AGCCTGGTC-A | rs1836070 | PLPP7 | 0.0017 | AGCCTGGTC | | 6.53E-09 | ?+ | 6.533E-09 | | New |
| 665 | 9 | 9-133482173-C-T | rs1010545 | SLC2A6, MYMK | 0.0007 | T | 0.141773 | 1.43E-09 | ++ | 1.128E-09 | | New |
| 665 | 9 | 9-133482205-G-C | rs9592278 | SLC2A6, MYMK | 0.0017 | G | 0.315331 | 2.43E-15 | -+ | 5.55E-13 | 0.8608 | New |
| 666 | 9 | 9-134464507-C-CCT | rs1442239 | RXRA, AL669970.1 | 0.0035 | C | 0.72818 | 3.17E-11 | -+ | 0.000001228 | | New |
| 667 | 9 | 9-135673019-TTTTG-T | | LCN9, SOHLH1 | 0.0007 | T | | 3.38E-09 | ?+ | 3.377E-09 | | New |

| | | | | | | | | | | | | |
|---|---|---|---|---|---|---|---|---|---|---|---|---|
| 668 | 9 | 9-136639478-C-T | rs1417363 | AL590226.2, AL590226.1 | 0.0005 | T | 0.145337 | 2.09E-10 | ++ | 7.972E-10 | 0.9206 | New |
| 669 | 9 | 9-137334886-G-C | rs9499433 | EXD3 | 0.0019 | C | 0.424351 | 2.17E-15 | -+ | 1.966E-13 | | New |
| 669 | 9 | 9-137334905-G-A | rs1463475 | EXD3 | 0.0005 | G | 0.822202 | 2.59E-08 | -+ | 6.542E-08 | | New |
| 670 | 9 | 9-16934631-TTC-T | rs1364541 | AL162725.2 | 0.003 | T | | 6.26E-09 | ?+ | 6.262E-09 | 0.6638 | New |
| 671 | 9 | 9-2528493-AC-A | | NA | 0.0009 | AC | | 6.48E-11 | ?+ | 6.476E-11 | | New |
| 672 | 9 | 9-26890151-C-CCT | | NA | 0.001 | C | | 6.31E-10 | ?+ | 6.308E-10 | | New |
| 673 | 9 | 9-31320715-C-CGG | | NA | 0.0002 | C | | 7.12E-09 | ?+ | 7.123E-09 | | New |
| 673 | 9 | 9-31320722-TCAGC-T | | NA | 0.0002 | TCAGC | | 1.6E-09 | ?+ | 1.596E-09 | | New |
| 674 | 9 | 9-33744596-T-A | rs1822346 | LINC01251, PRSS3 | 0.0011 | T | 0.186294 | 7.92E-15 | ++ | 4.146E-13 | | New |
| 675 | 9 | 9-34034655-ACC-A | | NA | 0.0007 | ACC | 0.836381 | 2.09E-15 | ++ | 1.065E-14 | | New |
| 676 | 9 | 9-34042314-C-CA | rs1407033 | UBAP2 | 0.0011 | CA | | 4.82E-08 | ?+ | 4.824E-08 | | New |
| 676 | 9 | 9-34042322-C-CA | rs7655349 | UBAP2 | 0.0019 | C | | 1.86E-09 | ?+ | 1.863E-09 | | New |
| 677 | 9 | 9-34094697-AT-A | rs1354347 | AL354989.1 | 0.0015 | A | 0.369773 | 2.95E-15 | -+ | 8.962E-09 | 0.3045 | New |
| 678 | 9 | 9-36095857-G-C | rs1905555 | RECK | 0.0013 | C | 0.09361 | 4.21E-08 | ++ | 0.000001942 | | New |
| 679 | 9 | 9-36182925-C-T | rs1827224 | CCIN, CLTA | 0.0078 | T | 0.43582 | 4.45E-10 | -+ | 0.00002159 | 0.1111 | New |
| 680 | 9 | 9-36604110-C-T | rs1587397 | MELK | 0.0019 | T | 0.387594 | 8.92E-19 | -+ | 1.162E-14 | | New |
| 681 | 9 | 9-38688643-G-C | rs1203948 | RNU6-765P, FAM240B | 0.001 | C | | 1.34E-08 | ?+ | 1.338E-08 | | New |
| 682 | 9 | 9-39941434-A-AT | | RF00156, AL772307.1 | 0.1297 | AT | | 3.34E-25 | ?+ | 3.337E-25 | | New |
| 683 | 9 | 9-40329787-C-CTTTT | | BX664727.3 | 0.0323 | C | | 2.31E-12 | ?+ | 2.306E-12 | | New |
| 684 | 9 | 9-40557771-T-TC | rs1232620 | BMS1P14 | 0.0139 | TC | | 3.56E-08 | ?+ | 3.559E-08 | | New |
| 685 | 9 | 9-40894485-G-C | rs1439623 | AL353626.1, MIR1299 | 0.0008 | G | | 5.6E-10 | ?+ | 5.603E-10 | | New |
| 686 | 9 | 9-41727997-TA-T | | FAM242F, AL162731.1 | 0.152 | TA | | 4.05E-09 | ?+ | 4.054E-09 | | New |

| | | | | | | | | | | | | |
|---|---|---|---|---|---|---|---|---|---|---|---|---|
| 687 | 9 | 9-42847389-A-AG | | BX664718.1, RNU6-599P | 0.0733 | A | | 9.79E-09 | ?- | 9.793E-09 | | New |
| 688 | 9 | 9-61670607-TAG-T | rs1822106 | AL935212.1, RN7SL722P | 0.0041 | T | | 2.38E-10 | ?+ | 2.384E-10 | 0.7959 | New |
| 689 | 9 | 9-62838432-C-CCGGCGCCCCCTCCGCGCCGGCGCCCCCCTCCGCGCCGGCGCCCCCTCCGCGCCGGCGCCCGGCGCT | | NA | 0.0035 | CCGGCGCCCCCTCCGC | | 3.66E-09 | ?- | 3.658E-09 | | New |
| 690 | 9 | 9-6626205-C-A | rs9010894 | GLDC | 0.0024 | C | 0.942914 | 6.66E-15 | ++ | 0.000005007 | 0.8558 | New |
| 691 | 9 | 9-6662926-C-CTG | rs1316966 | AL162411.1 | 0.0016 | C | 0.818951 | 5.57E-09 | -+ | 1.293E-08 | | New |
| 692 | 9 | 9-6698604-GCC-G | | NA | 0.0002 | G | | 3E-09 | ?+ | 3.004E-09 | | New |
| 693 | 9 | 9-67632973-C-CAA | | BX664730.1, RF00019 | 0.0192 | CAA | | 4.17E-08 | ?+ | 4.168E-08 | | New |
| 694 | 9 | 9-73226646-G-A | rs1822289 | ANXA1, AL451127.2 | 0.0011 | G | 0.19383 | 2.13E-11 | ++ | 7.22E-09 | | New |
| 695 | 9 | 9-74786521-C-CT | rs1290866 | TRPM6 | 0.0022 | CT | 0.437918 | 6.57E-13 | ++ | 1.315E-11 | | New |
| 696 | 9 | 9-82464932-C-T | rs9500497 | AL162726.4, AL356490.1 | 0.0011 | C | 0.317783 | 3.48E-13 | -+ | 1.232E-08 | 0.1792 | New |
| 697 | 9 | 9-84449299-C-T | rs5484699 | AL157886.1 | 0.0007 | T | 0.160959 | 2.46E-09 | ++ | 1.869E-09 | | New |
| 698 | 9 | 9-92785200-GT-G | | NA | 0.0004 | G | | 8.51E-10 | ?+ | 8.509E-10 | | New |
| 699 | 9 | 9-93166066-AT-A | rs1183474 | AL390760.1, WNK2 | 0.0019 | AT | | 3.87E-16 | ?+ | 3.872E-16 | 0.7207 | New |
| 700 | 9 | 9-94616642-C-CA | rs1272598 | FBP1 | 0.0004 | C | | 1.83E-15 | ?+ | 1.835E-15 | | New |
| 700 | 9 | 9-94616646-GC-G | rs1206929 | FBP1 | 0.0005 | G | | 5.29E-13 | ?+ | 5.29E-13 | | New |
| 700 | 9 | 9-94616648-ATG-A | rs1231327 | FBP1 | 0.0007 | ATG | | 1.5E-14 | ?+ | 1.495E-14 | | New |
| 701 | 9 | 9-96946504-C-CTT | rs1332932 | MFSD14C | 0.0016 | CTT | | 2.09E-08 | ?+ | 2.091E-08 | | New |
| 702 | 10 | 10-10099500-G-A | rs9940514 | LINC02670, RF00019 | 0.0011 | A | | 2.06E-11 | ?+ | 2.059E-11 | | New |

| | | | | | | | | | | | | |
|---|---|---|---|---|---|---|---|---|---|---|---|---|
| 703 | 10 | 10-101802646-GCAA-G | rs1243800 | OGA | 0.0015 | G | | 3.19E-10 | ?+ | 3.189E-10 | 0.6033 | New |
| 704 | 10 | 10-102054370-C-CG | rs1432629 | ARMH3 | 0.0012 | CG | | 4.54E-13 | ?+ | 4.535E-13 | 0.6008 | New |
| 705 | 10 | 10-102129838-C-CG | | NA | 0.0005 | C | | 9.75E-11 | ?+ | 9.753E-11 | | New |
| 706 | 10 | 10-102174312-C-CT | rs1469263 | NOLC1, ELOVL3 | 0.0012 | CT | 0.361198 | 5.56E-10 | ++ | 1.478E-08 | 0.5656 | New |
| 707 | 10 | 10-102206649-AAG-A | rs1472108 | NOLC1, ELOVL3 | 0.0056 | A | | 1.25E-11 | ?+ | 1.252E-11 | 0.5161 | New |
| 708 | 10 | 10-102785427-TCTGC-T | rs1172108 | WBP1L | 0.0005 | TCTGC | | 7.23E-09 | ?+ | 7.235E-09 | 0.5562 | New |
| 709 | 10 | 10-111403547-C-CTGG | | NA | 0.0004 | CTGG | | 6.57E-09 | ?+ | 6.57E-09 | | New |
| 710 | 10 | 10-111586970-CA-C | | AC021035.1, AL136119.1 | 0.0457 | C | | 4.33E-09 | ?- | 4.326E-09 | | New |
| 711 | 10 | 10-114884738-C-T | rs1321925 | FAM160B1 | 0.0004 | T | | 2.54E-10 | ?+ | 2.543E-10 | 0.09473 | New |
| 712 | 10 | 10-116619728-ATTTT-A | rs1185918 | PNLIPRP1, PNLIPRP2 | 0.0017 | A | | 2.3E-08 | ?+ | 2.298E-08 | | New |
| 713 | 10 | 10-117460011-T-A | rs1278738 | AC005871.2 | 0.0006 | A | 0.247876 | 4.26E-10 | ++ | 3.421E-09 | | New |
| 714 | 10 | 10-119182166-A-AC | | PRDX3, GRK5 | 0.0255 | AC | | 2.87E-09 | ?+ | 2.875E-09 | | New |
| 715 | 10 | 10-119200769-C-T | rs5512605 | PRDX3, GRK5 | 0.0015 | C | 0.075155 | 4.07E-08 | ++ | 1.607E-07 | 0.6543 | New |
| 715 | 10 | 10-119200773-C-T | rs7710416 | PRDX3, GRK5 | 0.0012 | T | 0.925359 | 1.26E-14 | -+ | 5.2E-12 | 0.2439 | New |
| 715 | 10 | 10-119200781-C-A | rs1368039 | PRDX3, GRK5 | 0.0022 | A | | 2.28E-19 | ?+ | 2.275E-19 | | New |
| 716 | 10 | 10-119907249-TTG-T | | SEC23IP | 0.0011 | TTG | | 1.7E-14 | ?+ | 1.703E-14 | | New |
| 716 | 10 | 10-119907251-ACG-A | | SEC23IP | 0.0008 | A | | 1.78E-12 | ?+ | 1.784E-12 | | New |
| 717 | 10 | 10-120602534-C-T | rs1184974 | LINC01561, AC023282.1 | 0.0002 | C | | 2.79E-08 | ?+ | 2.792E-08 | 0.271 | New |

| | | | | | | | | | | | | |
|---|---|---|---|---|---|---|---|---|---|---|---|---|
| 718 | 10 | 10-12148087-C-CACG | | SEC61A2 | 0.0013 | CACG | | 1.71E-13 | ?+ | 1.711E-13 | | Old |
| 719 | 10 | 10-12151525-T-C | rs4237421 | SEC61A2 | 0.007 | C | 0.42829 | 1.7E-09 | ++ | 0.00004015 | 0.9364 | Old |
| 719 | 10 | 10-12151534-T-A | rs1432331 | SEC61A2 | 0.0046 | A | 0.318508 | 3.57E-14 | ++ | 0.000007775 | 0.01843 | Old |
| 719 | 10 | 10-12151538-C-T | rs1299170 | SEC61A2 | 0.0054 | C | 0.931829 | 8.2E-12 | -+ | 0.001644 | 0.08579 | Old |
| 719 | 10 | 10-12151551-C-T | rs1232456 | SEC61A2 | 0.0053 | T | 0.931829 | 2.56E-12 | -+ | 0.002088 | 0.6777 | Old |
| 719 | 10 | 10-12151556-C-T | rs1346896 | SEC61A2 | 0.0053 | C | 0.931829 | 6.44E-17 | -+ | 0.0001252 | 0.6631 | Old |
| 720 | 10 | 10-12279639-C-CT | | NA | 0.0002 | C | | 1.67E-10 | ?+ | 1.673E-10 | | New |
| 721 | 10 | 10-12380003-AG-A | rs1281842 | CAMK1D | 0.0017 | A | 0.936821 | 2.4E-10 | ++ | 8.224E-09 | | New |
| 722 | 10 | 10-124809427-GGT-G | rs1458056 | ABRAXAS2 | 0.0046 | G | | 8.73E-17 | ?+ | 8.727E-17 | 0.6515 | New |
| 723 | 10 | 10-125834928-C-CG | rs1382795 | BCCIP | 0.0028 | C | 0.762481 | 7.79E-11 | -+ | 1.583E-09 | | New |
| 724 | 10 | 10-126892051-AC-A | rs2030065 | RF00271, DOCK1 | 0.0082 | A | | 3.12E-12 | ?+ | 3.116E-12 | | New |
| 725 | 10 | 10-128016654-GA-G | | PTPRE | 0.0008 | GA | | 4.73E-09 | ?+ | 4.727E-09 | | New |
| 726 | 10 | 10-132388816-CG-C | | LRRC27, PWWP2B | 0.088 | CG | | 8.37E-17 | ?+ | 8.366E-17 | | New |
| 727 | 10 | 10-132611004-GT-G | rs1137403 | INPP5A | 0.0036 | GT | | 3.67E-11 | ?+ | 3.669E-11 | | New |
| 728 | 10 | 10-13313766-C-CT | rs1254477 | PHYH, SEPHS1 | 0.0021 | C | 0.541077 | 6.96E-09 | -+ | 0.001843 | 0.3236 | New |
| 729 | 10 | 10-14933460-AGCCCC-A | | NA | 0.0004 | A | | 2.11E-10 | ?+ | 2.108E-10 | | New |
| 729 | 10 | 10-14933466-ATGTCG-A | | NA | 0.0003 | ATGTCG | | 4.68E-09 | ?+ | 4.683E-09 | | New |
| 730 | 10 | 10-1593285-ATAGGTCTCCTCTCTGGCATGGTCCCCTCTGTCACCCAGCTCCCCTGGCCCAAACCACACTCTG-A | rs1833286 | ADARB2 | 0.0114 | A | | 2.32E-20 | ?+ | 2.317E-20 | | New |
| 731 | 10 | 10-17983189-ACCTAG-A | | NA | 0.0008 | ACCTAG | | 3.85E-08 | ?+ | 3.854E-08 | | New |
| 732 | 10 | 10-18396114-G-A | rs9976580 | CACNB2 | 0.0007 | A | 0.825206 | 3.42E-09 | ++ | 5.859E-09 | 0.9935 | New |
| 733 | 10 | 10-23108707-C-T | rs1840010 | AL139815.1 | 0.0003 | C | | 1.46E-09 | ?+ | 1.459E-09 | | New |
| 734 | 10 | 10-27215821-TC-T | rs1247157 | ACBD5 | 0.0019 | T | | 1.44E-11 | ?+ | 1.437E-11 | 0.8912 | New |

| | | | | | | | | | | | | |
|---|---|---|---|---|---|---|---|---|---|---|---|---|
| 735 | 10 | 10-27312007-C-CT | rs7767287 | AL160291.1, RNU6-666P | 0.255 | C | 0.934769 | 1.58E-08 | -+ | 0.001388 | 0.3246 | New |
| 736 | 10 | 10-2983675-G-A | rs1413323 | AL355361.1, AL731533.1 | 0.001 | G | | 1.85E-08 | ?+ | 1.85E-08 | 0.2428 | New |
| 736 | 10 | 10-2983682-C-A | rs1351359 | AL355361.1, AL731533.1 | 0.0008 | C | 0.903154 | 2.83E-10 | ++ | 5.873E-10 | 0.0459 | New |
| 736 | 10 | 10-2983692-T-A | rs1349859 | AL355361.1, AL731533.1 | 0.0008 | A | 0.902933 | 2.16E-10 | ++ | 4.468E-10 | 0.1249 | New |
| 737 | 10 | 10-35114079-TGGGTTTTTTTTTTTTG-T | rs1207599 | AL392046.1 | 0.0024 | T | | 2.01E-18 | ?+ | 2.013E-18 | | New |
| 738 | 10 | 10-35370323-G-T | rs1836903 | CCNY | 0.0015 | G | 0.700674 | 1.44E-13 | ++ | 8.642E-10 | | New |
| 739 | 10 | 10-38930856-T-A | rs8790319 | AL133216.1, NONE | 0.0026 | A | | 2.73E-10 | ?+ | 2.73E-10 | | New |
| 740 | 10 | 10-39565455-A-C | rs1355114 | AL133216.1, NONE | 0.0396 | A | 0.456825 | 1.31E-41 | ++ | 5.956E-31 | | New |
| 741 | 10 | 10-41772582-AAG-A | | NA | 0.0009 | AAG | | 6.46E-14 | ?+ | 6.457E-14 | | New |
| 741 | 10 | 10-41772584-GCA-G | | NA | 0.0008 | G | | 1.74E-12 | ?+ | 1.735E-12 | | New |
| 742 | 10 | 10-41847867-AT-A | rs1157245 | NONE, LINC00839 | 0.0038 | A | | 2.26E-12 | ?+ | 2.264E-12 | | New |
| 742 | 10 | 10-42066423-AATC-A | rs1252500 | NONE, LINC00839 | 0.0032 | A | | 5.97E-13 | ?+ | 5.972E-13 | | New |
| 743 | 10 | 10-42102736-T-A | rs1439434 | NONE, LINC00839 | 0.0021 | A | | 2.13E-09 | ?+ | 2.134E-09 | | New |
| 743 | 10 | 10-42102765-G-C | rs5774250 | NONE, LINC00839 | 0.0016 | G | | 1.44E-14 | ?+ | 1.439E-14 | | New |
| 744 | 10 | 10-5378108-ATGGTG-A | | NA | 0.0009 | ATGGTG | | 1.32E-12 | ?+ | 1.323E-12 | | New |

| | | | | | | | | | | | | |
|---|---|---|---|---|---|---|---|---|---|---|---|---|
| 745 | 10 | 10-68227393-G-T | rs9356378 | MYPN, ATOH7 | 0.001 | T | 0.702014 | 6.3E-10 | ++ | 8.034E-09 | | New |
| 746 | 10 | 10-68593595-G-A | rs1008341 | TET1 | 0.0009 | G | 0.347596 | 6.61E-12 | -+ | 0.0001112 | 0.3818 | New |
| 747 | 10 | 10-70420363-C-CT | | NA | 0.001 | CT | | 1.31E-11 | ?+ | 1.315E-11 | | New |
| 747 | 10 | 10-70420366-T-A | rs1007847 | EIF4EBP2 | 0.0004 | T | | 9.86E-09 | ?+ | 9.866E-09 | | New |
| 748 | 10 | 10-72191881-C-CAA | rs1275756 | ASCC1 | 0.002 | C | | 6.46E-10 | ?+ | 6.463E-10 | | New |
| 749 | 10 | 10-73947669-TG-T | | NA | 0.0006 | T | | 1.01E-10 | ?+ | 1.011E-10 | | New |
| 750 | 10 | 10-74595727-A-G | rs3680424 | ADK | 0.0046 | A | | 4.32E-11 | ?+ | 4.316E-11 | | New |
| 750 | 10 | 10-74595729-T-TCC | rs3777442 | ADK | 0.0071 | T | | 7.23E-16 | ?+ | 7.234E-16 | | New |
| 751 | 10 | 10-74648493-C-A | rs1202013 | ADK | 0.0006 | C | 0.801128 | 2.32E-08 | -+ | 0.0003033 | | New |
| 752 | 10 | 10-79922400-G-A | rs2758559 | AL512662.2 | 0.0082 | G | | 2E-13 | ?+ | 1.996E-13 | 0.03556 | Old |
| 753 | 10 | 10-84087114-C-CGT | | NA | 0.001 | C | | 1.18E-10 | ?+ | 1.18E-10 | | New |
| 754 | 10 | 10-976272-A-G | rs1250895 | AL359878.1 | 0.0102 | G | 0.950602 | 5.22E-10 | ++ | 0.00002117 | | New |
| 755 | 10 | 10-97710681-ACT-A | rs1374926 | AVPI1, MARVELD1 | 0.0017 | ACT | | 3.07E-10 | ?+ | 3.072E-10 | 0.4273 | New |
| 756 | 11 | 11-1018088-TG-T | rs3683422 | MUC6 | 0.0087 | T | | 6.35E-09 | ?+ | 6.353E-09 | | New |
| 757 | 11 | 11-1018456-G-A | rs7992042 | MUC6 | 0.0028 | A | 0.373363 | 1.1E-10 | ++ | 4.132E-10 | | New |
| 758 | 11 | 11-103459091-C-T | rs1030089 | DYNC2H1 | 0.0007 | T | 0.958299 | 7.6E-10 | ++ | 0.000002208 | 0.452 | New |
| 758 | 11 | 11-103459116-C-T | rs9725471 | DYNC2H1 | 0.0008 | T | 0.881363 | 9.16E-09 | ++ | 0.000002698 | 0.9971 | New |
| 758 | 11 | 11-103459123-C-A | rs9499912 | DYNC2H1 | 0.0015 | C | 0.684033 | 5.6E-09 | -+ | 8.184E-07 | 0.5324 | New |
| 759 | 11 | 11-105515895-G-T | rs1053968 | CARD18 | 0.0028 | T | 0.479102 | 2.34E-18 | ++ | 2.641E-18 | | New |
| 759 | 11 | 11-105515902-G-A | rs9149829 | CARD18 | 0.0016 | A | 0.691041 | 1.38E-17 | ++ | 3.275E-17 | 0.1885 | New |
| 760 | 11 | 11-108396326-C-CAA | | NA | 0.0007 | C | 0.872063 | 3.71E-08 | -+ | 0.000001314 | | New |
| 761 | 11 | 11-1095580-GAGTACA-G | | NA | 0.0026 | G | | 3.54E-08 | ?+ | 3.537E-08 | | New |
| 762 | 11 | 11-113771108-TG-T | | NA | 0.0009 | TG | | 4.83E-15 | ?+ | 4.825E-15 | | New |
| 763 | 11 | 11-1166174-C-CCCTATGGTGAGACCCTGCACCCAACACACAGTCTA | | NA | 0.0021 | C | | 7.27E-10 | ?+ | 7.273E-10 | | New |
| 764 | 11 | 11-117105931-C-CA | | NA | 0.0002 | C | | 3.63E-08 | ?+ | 3.625E-08 | | New |

| | | | | | | | | | | | | |
|---|---|---|---|---|---|---|---|---|---|---|---|---|
| 765 | 11 | 11-118709508-G-A | rs1240319 | AP002954.1 | 0.0018 | A | 0.527094 | 3.93E-08 | -+ | 0.00899 | 0.1787 | New |
| 766 | 11 | 11-118736461-C-CAT | rs1555154 | AP002954.1 | 0.0025 | CAT | | 2.53E-08 | ?+ | 2.529E-08 | | New |
| 767 | 11 | 11-123615487-G-C | rs1349200 | GRAMD1B | 0.0006 | C | 0.781646 | 3.21E-09 | ++ | 1.296E-08 | | New |
| 768 | 11 | 11-12757946-ATTTTG-A | | NA | 0.001 | ATTTTG | | 6.95E-09 | ?+ | 6.946E-09 | | New |
| 769 | 11 | 11-129833604-G-A | rs1316952 | TMEM45B | 0.0008 | A | 0.094626 | 1.03E-10 | ++ | 2.658E-10 | | New |
| 770 | 11 | 11-1341421-AC-A | rs1394780 | AC136297.1, LINC02689 | 0.0014 | A | 0.578834 | 1.13E-08 | ++ | 0.000000241 | | New |
| 771 | 11 | 11-134286572-G-C | rs1243638 | GLB1L3 | 0.0087 | C | 0.140239 | 3.78E-18 | ++ | 2.599E-08 | 0.1295 | New |
| 772 | 11 | 11-1355268-C-T | rs1272815 | LINC02689 | 0.0033 | T | 0.781415 | 3.42E-15 | -+ | 0.00003054 | 0.8594 | New |
| 772 | 11 | 11-1355280-AT-A | | LINC02689 | 0.0033 | AT | 0.882325 | 7.68E-13 | -+ | 0.00069 | | New |
| 773 | 11 | 11-1355825-C-T | rs8901961 | LINC02689 | 0.001 | T | 0.004295 | 2.1E-08 | ++ | 3.183E-10 | 0.5118 | New |
| 773 | 11 | 11-1355828-G-T | rs1379529 | LINC02689 | 0.0011 | G | 0.182731 | 2.06E-11 | ++ | 1.831E-10 | 0.9241 | New |
| 773 | 11 | 11-1355844-T-A | rs1023546 | LINC02689 | 0.0019 | T | 0.373811 | 3.72E-16 | ++ | 2.126E-14 | 0.7431 | New |
| 774 | 11 | 11-14930887-G-A | rs9295909 | CALCB | 0.0025 | A | 0.714559 | 1.33E-10 | ++ | 0.0001585 | 0.1522 | New |
| 775 | 11 | 11-1622940-C-CAAA | rs1435812 | KRTAP5-4 | 0.0013 | CAAA | 0.693221 | 7.35E-09 | ++ | 0.00000824 | 0.7178 | New |
| 776 | 11 | 11-18328184-C-CAG | rs1484419 | GTF2H1 | 0.0016 | C | | 6.89E-09 | ?+ | 6.89E-09 | | New |
| 777 | 11 | 11-19260622-CCA-C | rs1135222 | CSRP3-AS1 | 0.0037 | CCA | 0.420952 | 9.92E-11 | -- | 1.303E-10 | | New |
| 778 | 11 | 11-2124905-GA-G | rs1276903 | H19, IGF2 | 0.0029 | G | | 1.75E-08 | ?+ | 1.754E-08 | 0.1898 | New |
| 779 | 11 | 11-2139063-CG-C | | IGF2 | 0.0118 | C | | 3.32E-10 | ?+ | 3.319E-10 | | New |
| 780 | 11 | 11-30791372-GTA-G | | NA | 0.0007 | GTA | | 1.14E-09 | ?+ | 1.14E-09 | | New |
| 781 | 11 | 11-30887986-AGAGC-A | | NA | 0.0004 | A | | 1.46E-11 | ?+ | 1.457E-11 | | New |
| 782 | 11 | 11-31533623-C-CAG | | NA | 0.0009 | CAG | 0.427123 | 8.07E-10 | ++ | 7.772E-10 | | New |
| 783 | 11 | 11-31971510-C-T | rs9059508 | PAUPAR | 0.002 | C | 0.071264 | 4.79E-13 | ++ | 9.747E-14 | | New |

| | | | | | | | | | | | | |
|---|---|---|---|---|---|---|---|---|---|---|---|---|
| 784 | 11 | 11-33250072-TA-T | | NA | 0.0003 | T | | 3.67E-10 | ?+ | 3.67E-10 | | New |
| 785 | 11 | 11-37654820-G-T | rs1234469 | C11orf74, RF00322 | 0.001 | T | 0.600947 | 2.48E-08 | ++ | 0.00003587 | | New |
| 786 | 11 | 11-463371-GT-G | rs1311117 | AC138230 .1 | 0.0072 | GT | | 1.13E-08 | ?+ | 1.127E-08 | 0.01425 | New |
| 787 | 11 | 11-46355253-C-CATG | | NA | 0.0003 | CATG | | 7.84E-10 | ?+ | 7.843E-10 | | New |
| 788 | 11 | 11-46830926-G-A | rs1421155 | CKAP5 | 0.0004 | G | 0.415225 | 8.67E-12 | -+ | 3.172E-11 | | Old |
| 789 | 11 | 11-47085719-G-A | rs5641010 | C11orf49 | 0.0007 | A | 0.657503 | 2.39E-10 | ++ | 2.284E-08 | | Old |
| 790 | 11 | 11-47563973-G-C | rs1043274 | CELF1 | 0.0013 | G | 0.735987 | 4.24E-09 | -+ | 0.01103 | 0.677 | Old |
| 791 | 11 | 11-47674901-AT-A | | AGBL2 | 0.0003 | AT | | 5.14E-09 | ?+ | 5.138E-09 | | Old |
| 792 | 11 | 11-47677751-C-CTG | | NA | 0.0011 | CTG | | 1.29E-10 | ?+ | 1.289E-10 | | Old |
| 793 | 11 | 11-48727069-TTTGTC-T | rs1359118 | OR4A47, TRIM51GP | 0.0058 | TTTGTC | | 2.79E-15 | ?+ | 2.786E-15 | | New |
| 793 | 11 | 11-48727087-ATG-A | rs1198553 | OR4A47, TRIM51GP | 0.0061 | ATG | | 1.82E-16 | ?+ | 1.817E-16 | | New |
| 794 | 11 | 11-48768777-T-A | rs9585606 | OR4A47, TRIM51GP | 0.0003 | T | | 3.27E-10 | ?+ | 3.275E-10 | | New |
| 795 | 11 | 11-48843498-C-T | rs7963294 | OR4A47, TRIM51GP | 0.0006 | C | | 3.16E-08 | ?+ | 3.158E-08 | | New |
| 796 | 11 | 11-48911550-C-T | rs1442475 | OR4A47, TRIM51GP | 0.0015 | T | | 1.31E-12 | ?+ | 1.313E-12 | | New |
| 797 | 11 | 11-48911855-A-T | rs6485924 | OR4A47, TRIM51GP | 0.0142 | A | 0.980661 | 4.29E-14 | -+ | 0.000001383 | | New |
| 798 | 11 | 11-50576986-C-A | rs5584504 | AC109635 .2, NONE | 0.0002 | C | 0.227177 | 5.7E-10 | -+ | 4.678E-08 | | New |
| 799 | 11 | 11-50716519-C-T | rs8671086 | AC109635 .2, NONE | 0.0002 | T | | 1.64E-08 | ?+ | 1.643E-08 | | New |
| 800 | 11 | 11-54897486-TGCCA-T | | NA | 0.0032 | TGCCA | | 4.65E-09 | ?- | 4.65E-09 | | New |
| 801 | 11 | 11-55233749-C-T | rs2120001 | OR4A5, TRIM48 | 0.006 | C | | 2.69E-68 | ?+ | 2.696E-68 | | New |
| 801 | 11 | 11-55233796-C-A | | OR4A5, TRIM48 | 0.0021 | C | | 1.88E-23 | ?+ | 1.885E-23 | | New |

| | | | | | | | | | | | |
|---|---|---|---|---|---|---|---|---|---|---|---|
| 802 | 11 | 11-55251455-G-A | rs6189837 | OR4A5, TRIM48 | 0.0618 | G | 0.627801 | 1.49E-09 | -+ | 0.3606 | 0.1854 New |
| 802 | 11 | 11-55461884-C-A | rs5354997 | OR4A15, OR4C15 | 0.0182 | A | 0.709282 | 7.51E-10 | -+ | 0.003299 | New |
| 803 | 11 | 11-57768000-AT-A | rs1475471 | AP001931.1, CTNND1, TMX2-CTNND1 | 0.0024 | AT | 0.64311 | 9.68E-21 | ++ | 3.371E-12 | New |
| 804 | 11 | 11-61824974-C-CCT | rs1555073 | FADS1, FADS2 | 0.0045 | CCT | | 7.07E-19 | ?+ | 7.074E-19 | 0.8907 New |
| 805 | 11 | 11-61894409-G-T | rs1938621 | FADS3, RAB3IL1 | 0.0007 | G | 0.750836 | 5.78E-13 | -+ | 0.000005148 | New |
| 805 | 11 | 11-61894450-C-T | rs1555084 | FADS3, RAB3IL1 | 0.0006 | C | 0.732099 | 2.2E-08 | -+ | 4.891E-07 | New |
| 806 | 11 | 11-62004372-C-CAG | | NA | 0.001 | CAG | 0.951223 | 2.18E-08 | -+ | 4.912E-08 | New |
| 807 | 11 | 11-62211652-G-A | rs5453617 | SCGB2A1 | 0.0033 | A | 0.225421 | 8.07E-15 | -+ | 3.729E-10 | 0.6888 New |
| 808 | 11 | 11-62523038-C-A | rs7804666 | AHNAK | 0.0008 | C | 0.422813 | 6.65E-10 | ++ | 5.665E-09 | 0.07122 New |
| 809 | 11 | 11-62537111-GCACCACA-G | | NA | 0.0013 | G | 0.592255 | 3.52E-13 | ++ | 1.719E-12 | New |
| 810 | 11 | 11-62845315-ATT-A | | NA | 0.0005 | A | | 4.29E-08 | ?+ | 4.289E-08 | New |
| 811 | 11 | 11-62869905-C-CTTGA | rs1395280 | SLC3A2 | 0.0023 | C | | 3.37E-17 | ?+ | 3.37E-17 | 0.05053 New |
| 812 | 11 | 11-63684993-C-CAA | | NA | 0.0006 | C | | 1.41E-09 | ?+ | 1.411E-09 | New |
| 813 | 11 | 11-63764272-C-T | rs1590909 | C11orf95 | 0.0006 | C | 0.034778 | 9.13E-09 | ++ | 9.531E-10 | New |
| 814 | 11 | 11-63860686-G-A | rs5357448 | MARK2 | 0.0003 | A | 0.739983 | 7.78E-09 | ++ | 2.715E-08 | 0.1995 New |
| 815 | 11 | 11-64280476-C-A | rs1325649 | BAD, GPR137 | 0.0012 | A | | 9.45E-15 | ?+ | 9.454E-15 | New |
| 816 | 11 | 11-64662034-C-CTT | rs1413522 | NRXN2 | 0.0016 | C | | 6.99E-12 | ?+ | 6.986E-12 | 0.1128 New |
| 817 | 11 | 11-64664247-C-CAAAT | | NA | 0.0003 | C | | 9.08E-11 | ?+ | 9.082E-11 | New |
| 818 | 11 | 11-652592-C-CTT | rs1210224 | DEAF1 | 0.0012 | C | | 7.31E-09 | ?+ | 7.309E-09 | 0.2157 New |
| 819 | 11 | 11-65463200-TAG-T | | NA | 0.0004 | T | | 7.59E-09 | ?+ | 7.588E-09 | New |
| 820 | 11 | 11-65527549-AC-A | | NA | 0.0002 | A | | 1.98E-08 | ?+ | 1.976E-08 | New |

| | | | | | | | | | | | |
|---|---|---|---|---|---|---|---|---|---|---|---|
| 821 | 11 | 11-66402115-T-A | rs1335619 | RNU1-84P, AP001107.5 | 0.0006 | T | 0.233866 | 3.73E-10 | -+ | 1.239E-07 | | New |
| 822 | 11 | 11-67134652-GCGT-G | | NA | 0.0005 | G | | 3.02E-09 | ?+ | 3.019E-09 | | New |
| 823 | 11 | 11-67157185-C-T | rs1273071 | KDM2A | 0.0015 | C | 0.081903 | 1.62E-13 | ++ | 2.486E-10 | | New |
| 824 | 11 | 11-67988509-ATG-A | | NA | 0.0011 | ATG | | 1E-08 | ?+ | 1.004E-08 | | New |
| 825 | 11 | 11-68207031-AGG-A | rs1341660 | KMT5B | 0.0015 | A | | 2.14E-09 | ?+ | 2.14E-09 | | New |
| 826 | 11 | 11-69082539-T-A | rs9991779 | AP003071.5, TPCN2 | 0.0002 | T | 0.605345 | 3.07E-08 | -+ | 0.000004815 | 0.7076 | New |
| 827 | 11 | 11-69577301-G-C | rs1430663 | LINC01488, CCND1 | 0.0007 | G | | 3.44E-15 | ?+ | 3.44E-15 | | New |
| 828 | 11 | 11-69720156-ACT-A | rs1855026 | FGF19, FGF4 | 0.0008 | ACT | | 5.89E-10 | ?+ | 5.889E-10 | | New |
| 829 | 11 | 11-69723576-G-C | rs1333742 | FGF19, FGF4 | 0.0008 | C | 0.314411 | 3.99E-09 | ++ | 2.456E-09 | | New |
| 830 | 11 | 11-70222751-TA-T | rs1287705 | FADD, PPFIA1 | 0.0033 | TA | 0.154994 | 1.07E-14 | ++ | 5.324E-08 | | New |
| 831 | 11 | 11-70803334-C-CTA | rs1555050 | SHANK2 | 0.0072 | C | | 4.62E-19 | ?+ | 4.621E-19 | | New |
| 831 | 11 | 11-70803346-C-CCGTT | rs1555051 | SHANK2 | 0.0072 | CCGTT | | 4.35E-17 | ?+ | 4.354E-17 | | New |
| 832 | 11 | 11-70890949-GT-G | rs1555074 | AP003783.1 | 0.0033 | G | | 1.46E-30 | ?+ | 1.46E-30 | | New |
| 832 | 11 | 11-70890952-AT-A | rs1555074 | AP003783.1 | 0.0019 | A | | 2.51E-18 | ?+ | 2.512E-18 | | New |
| 833 | 11 | 11-71920538-C-CAG | rs1168992 | AP002495.1 | 0.0048 | C | 0.311751 | 1.46E-17 | ++ | 1.145E-17 | 0.4165 | New |
| 834 | 11 | 11-71936008-C-T | rs9789076 | RNF121 | 0.0003 | T | 0.689892 | 2.86E-08 | -+ | 5.618E-07 | 0.9986 | New |
| 835 | 11 | 11-72063209-C-CCT | rs1243122 | NUMA1 | 0.0016 | C | | 1.21E-14 | ?+ | 1.208E-14 | 0.8002 | New |
| 836 | 11 | 11-73692269-C-CCGAGGT | rs1342643 | RAB6A | 0.0028 | C | | 1.01E-12 | ?+ | 1.012E-12 | 0.6954 | Old |
| 837 | 11 | 11-73813823-C-CCT | | NA | 0.0007 | CCT | | 3.25E-11 | ?+ | 3.246E-11 | | Old |
| 838 | 11 | 11-74066348-TACATATGTAACAAATCTGCAC-T | rs1249620 | C2CD3 | 0.0022 | T | 0.380788 | 1.19E-08 | -+ | 0.00001488 | 0.3254 | Old |

| | | | | | | | | | | | | |
|---|---|---|---|---|---|---|---|---|---|---|---|---|
| 839 | 11 | 11-75748015-G-A | rs9197375 | RN7SL786P, AP001922.1 | 0.0012 | A | 0.944521 | 1.22E-12 | -+ | 5.868E-08 | | New |
| 839 | 11 | 11-75748022-G-A | rs1944580 | RN7SL786P, AP001922.1 | 0.0009 | A | 0.370418 | 1.26E-13 | -+ | 9.029E-08 | | New |
| 839 | 11 | 11-75748028-G-T | rs1269046 | RN7SL786P, AP001922.1 | 0.0011 | G | | 1.15E-09 | ?+ | 1.155E-09 | | New |
| 840 | 11 | 11-76255841-GGGT-G | rs1331553 | AP000785.2, THAP12 | 0.0009 | G | | 8.9E-11 | ?+ | 8.897E-11 | 0.4552 | New |
| 840 | 11 | 11-76255847-C-CA | rs1401992 | AP000785.2, THAP12 | 0.0006 | C | | 6.75E-14 | ?+ | 6.751E-14 | | New |
| 841 | 11 | 11-77546029-C-CCT | | NA | 0.0004 | C | | 8.13E-09 | ?+ | 8.131E-09 | | New |
| 842 | 11 | 11-881238-G-A | rs1485554 | CHID1 | 0.001 | A | | 5.35E-09 | ?+ | 5.347E-09 | 0.1042 | New |
| 843 | 11 | 11-8984332-G-T | rs5340525 | NRIP3 | 0.0006 | T | | 2.87E-10 | ?+ | 2.873E-10 | | New |
| 844 | 11 | 11-91664514-G-A | rs3722378 | AP002791.1, AP003055.1 | 0.0009 | G | 0.484709 | 3.44E-17 | -+ | 2.413E-09 | | New |
| 845 | 11 | 11-9300701-C-A | rs1443840 | TMEM41B | 0.002 | C | 0.531004 | 6.91E-21 | ++ | 3.965E-11 | 0.3065 | New |
| 845 | 11 | 11-9300706-G-A | rs3688489 | TMEM41B | 0.0018 | A | 0.281046 | 9.43E-18 | ++ | 1.606E-09 | 0.6874 | New |
| 846 | 11 | 11-9384165-C-T | rs1590420 | IPO7 | 0.0005 | C | | 1.81E-11 | ?+ | 1.813E-11 | | New |
| 846 | 11 | 11-9384187-G-C | rs1303095 | IPO7 | 0.0015 | G | 0.835013 | 3.15E-20 | -+ | 1.449E-18 | 0.3851 | New |
| 847 | 11 | 11-9551023-C-CTT | rs1318500 | ZNF143, WEE1 | 0.0042 | CTT | | 2.62E-24 | ?+ | 2.625E-24 | | New |
| 848 | 12 | 12-101445846-C-A | | RF00019 | 0.0005 | A | | 8.89E-15 | ?+ | 8.887E-15 | | New |
| 849 | 12 | 12-103321503-C-T | rs2020379 | C12orf42 | 0.056 | T | | 8.36E-11 | ?+ | 8.362E-11 | | New |
| 850 | 12 | 12-108905116-G-A | rs5286716 | DAO, SVOP | 0.0006 | A | 0.110957 | 5.66E-13 | ++ | 1.664E-13 | | New |
| 851 | 12 | 12-110043629-G-A | rs9215243 | C12orf76 | 0.0004 | A | 0.702288 | 1.27E-08 | -+ | 0.000001204 | 0.8726 | New |
| 852 | 12 | 12-110702738-TC-T | | NA | 0.0002 | T | 0.264035 | 1.33E-08 | -+ | 3.202E-07 | | New |

| | | | | | | | | | | | |
|---|---|---|---|---|---|---|---|---|---|---|---|
| 853 | 12 | 12-111611215-ATT-A | rs1382775 | ATXN2-AS, BRAP | 0.0008 | A | | 2.92E-10 | ?+ | 2.917E-10 | 0.8753 | New |
| 854 | 12 | 12-111929280-G-A | rs1297364 | MAPKAPK5, TMEM116 | 0.0006 | A | | 6.29E-14 | ?+ | 6.29E-14 | 0.2761 | New |
| 854 | 12 | 12-111929282-G-A | rs1340910 | MAPKAPK5, TMEM116 | 0.0003 | A | | 1.37E-09 | ?+ | 1.372E-09 | 0.6316 | New |
| 855 | 12 | 12-112412907-G-C | rs2037357 | AC004086.1 | 0.001 | G | | 3.29E-11 | ?+ | 3.295E-11 | | New |
| 855 | 12 | 12-112412911-GA-G | | NA | 0.0009 | GA | | 8.16E-16 | ?+ | 8.16E-16 | | New |
| 856 | 12 | 12-114298592-TCCTC-T | rs1371643 | LINC02459, TBX5 | 0.0018 | T | | 1.76E-11 | ?+ | 1.756E-11 | | New |
| 857 | 12 | 12-11567230-GGT-G | rs1491387 | LINC01252 | 0.0026 | G | | 7E-14 | ?+ | 7E-14 | 0.115 | New |
| 858 | 12 | 12-117081702-G-T | rs1282630 | TESC | 0.0014 | T | 0.961506 | 9.82E-12 | ++ | 1.008E-08 | 0.9359 | New |
| 859 | 12 | 12-118065595-T-A | rs1334803 | VSIG10 | 0.0011 | T | 0.708606 | 5.54E-10 | ++ | 9.719E-07 | 0.5709 | New |
| 860 | 12 | 12-120311520-C-T | rs1482211 | SIRT4 | 0.0005 | C | | 6.35E-10 | ?+ | 6.353E-10 | | New |
| 861 | 12 | 12-120556370-TAAAAA-T | | NA | 0.001 | T | | 3.11E-10 | ?+ | 3.111E-10 | | New |
| 862 | 12 | 12-120663475-C-T | rs1880782 | CABP1 | 0.0014 | T | 0.438428 | 7.21E-14 | ++ | 6.297E-14 | | New |
| 862 | 12 | 12-120663495-G-C | rs1039962 | CABP1 | 0.0018 | G | | 1.96E-15 | ?+ | 1.962E-15 | 0.2338 | New |
| 863 | 12 | 12-120893304-G-T | rs1406406 | SPPL3 | 0.0019 | G | 0.112503 | 4.04E-10 | -+ | 0.000649 | 0.1498 | New |
| 864 | 12 | 12-121034133-G-C | rs1592939 | OASL | 0.0007 | C | | 3.4E-12 | ?+ | 3.398E-12 | | New |
| 865 | 12 | 12-121034789-G-C | rs1366382 | OASL | 0.0024 | G | 0.143081 | 7.85E-20 | ++ | 3.316E-19 | | New |
| 866 | 12 | 12-121076600-T-TG | | OASL, AC079602.3 | 0.0503 | T | | 2.78E-11 | ?+ | 2.776E-11 | | New |
| 867 | 12 | 12-121226389-TGA-T | | P2RX4 | 0.0011 | TGA | 0.409897 | 1.21E-09 | -+ | 0.000004027 | | New |
| 868 | 12 | 12-121617997-C-CCA | rs1555321 | RNU6-1004P, ORAI1 | 0.0021 | C | | 1.78E-09 | ?+ | 1.776E-09 | 0.8399 | New |
| 869 | 12 | 12-121634293-G-A | rs1230863 | ORAI1 | 0.0033 | A | 0.250878 | 8.14E-25 | ++ | 5.033E-25 | 0.9507 | New |
| 869 | 12 | 12-121634366-G-T | rs1361982 | ORAI1 | 0.001 | T | 0.508955 | 1.8E-08 | -+ | 2.013E-07 | 0.5161 | New |
| 870 | 12 | 12-121670590-C-CCT | rs1408180 | MORN3 | 0.006 | C | 0.77022 | 4.58E-43 | ++ | 2.111E-40 | | New |

| | | | | | | | | | | | | |
|---|---|---|---|---|---|---|---|---|---|---|---|---|
| 871 | 12 | 12-121726344-C-CCGATGCAGGTGGATCA | | NA | 0.0015 | C | | 1.53E-15 | ?+ | 1.528E-15 | | New |
| 872 | 12 | 12-121745498-C-CT | rs1555331 | TMEM120B | 0.0006 | C | | 1.11E-09 | ?+ | 1.111E-09 | | New |
| 873 | 12 | 12-121876914-TGA-T | | NA | 0.0004 | T | | 2.93E-08 | ?+ | 2.927E-08 | | New |
| 874 | 12 | 12-121914347-C-CT | | NA | 0.0013 | CT | | 3.05E-10 | ?+ | 3.047E-10 | | New |
| 875 | 12 | 12-122320066-ATATTGAGACCATCCTG-A | rs1353668 | CLIP1 | 0.0012 | A | | 4.76E-08 | ?+ | 4.765E-08 | | New |
| 876 | 12 | 12-123173072-C-T | rs9670436 | MPHOSPH9 | 0.0015 | T | 0.969602 | 1.87E-13 | ++ | 8.057E-11 | | New |
| 876 | 12 | 12-123173111-G-A | rs5570514 | MPHOSPH9 | 0.0011 | G | 0.62016 | 1.03E-15 | ++ | 7.335E-11 | 0.5216 | New |
| 877 | 12 | 12-12363724-G-A | rs1168684 | BORCS5 | 0.0006 | G | | 8.16E-09 | ?+ | 8.158E-09 | | New |
| 878 | 12 | 12-123717186-G-T | rs1310494 | AC117503.5 | 0.0008 | G | 0.287021 | 1.67E-11 | ++ | 5.019E-09 | | New |
| 879 | 12 | 12-123755449-TGG-T | | NA | 0.0007 | TGG | | 3.67E-09 | ?+ | 3.669E-09 | | New |
| 880 | 12 | 12-123755925-C-CA | | NA | 0.0009 | C | | 1.06E-13 | ?+ | 1.064E-13 | | New |
| 880 | 12 | 12-123755940-G-T | rs1425062 | ATP6V0A2 | 0.0002 | T | 0.49587 | 2.42E-09 | ++ | 1.361E-07 | | New |
| 880 | 12 | 12-123755941-C-T | rs5623055 | ATP6V0A2 | 0.0004 | T | 0.49587 | 4.06E-10 | ++ | 7.919E-09 | | New |
| 881 | 12 | 12-124547213-G-A | rs5650460 | NCOR2 | 0.0006 | A | 0.457873 | 2.04E-11 | ++ | 1.783E-11 | | New |
| 881 | 12 | 12-124547226-C-CAA | | NCOR2 | 0.0014 | C | | 2.31E-14 | ?+ | 2.313E-14 | | New |
| 882 | 12 | 12-124951817-C-CA | | NA | 0.0002 | CA | | 2.44E-08 | ?+ | 2.438E-08 | | New |
| 883 | 12 | 12-125090208-T-TCATCCATCCATCCTCCATTCATCATCCCTCATCCATCATTTATCCAACTATCC | | AACS | 0.0465 | TCATCCATCCATCCTCC | | 1.15E-13 | ?+ | 1.154E-13 | | New |
| 884 | 12 | 12-12553080-C-CTCT | | NA | 0.0002 | CTCT | | 2.14E-08 | ?+ | 2.143E-08 | | New |

| | | | | | | | | | | | | |
|---|---|---|---|---|---|---|---|---|---|---|---|---|
| 885 | 12 | 12-125803495-G-A | rs1042663 | TMEM132B, AC005252.2 | 0.0004 | A | 0.331749 | 3.65E-09 | ++ | 4.722E-09 | | New |
| 886 | 12 | 12-126712968-G-C | rs5390224 | AC069235.1 | 0.0044 | C | 0.793387 | 7.88E-11 | -+ | 0.003985 | | New |
| 887 | 12 | 12-126713469-C-T | rs1484997 | AC069235.1 | 0.0043 | C | 0.75722 | 6.04E-20 | -+ | 3.141E-07 | 0.9645 | New |
| 888 | 12 | 12-128596107-C-CG | rs1875936 | TMEM132C | 0.0017 | C | 0.999769 | 3.27E-14 | ++ | 1.039E-09 | | New |
| 889 | 12 | 12-128681911-C-CTG | rs1467612 | TMEM132C | 0.0008 | CTG | | 1.39E-08 | ?+ | 1.388E-08 | | New |
| 890 | 12 | 12-130798250-CA-C | | STX2 | 0.0761 | C | | 1.75E-08 | ?+ | 1.748E-08 | | New |
| 891 | 12 | 12-132381811-AGCACCCGATACACAGAAGACAGAAGACATACTCAGCTAAAGCT-A | | NA | 0.0011 | AGCACCCGATACACAG | | 2.71E-09 | ?+ | 2.707E-09 | | New |
| 892 | 12 | 12-132683958-C-CAG | rs2005417 | POLE | 0.003 | C | 0.960693 | 5.28E-11 | ++ | 6.304E-07 | | New |
| 893 | 12 | 12-18232057-G-A | | NONE, PIK3C2G | 0.0004 | G | | 4.46E-08 | ?+ | 4.457E-08 | | New |
| 894 | 12 | 12-20353357-C-T | rs1027696 | LINC02468, AC129102.1 | 0.0026 | T | 0.194077 | 4.22E-08 | ++ | 0.0001485 | 0.2578 | New |
| 895 | 12 | 12-20353414-GGAACCC-G | rs1425955 | LINC02468, AC129102.1 | 0.0045 | GGAACCC | | 4.49E-15 | ?+ | 4.492E-15 | 0.3619 | New |
| 896 | 12 | 12-26332904-TGG-T | | NA | 0.0075 | TGG | | 2.08E-08 | ?- | 2.079E-08 | | New |
| 897 | 12 | 12-27784354-G-C | rs1271765 | KLHL42 | 0.0005 | G | 0.909335 | 4.34E-11 | -+ | 1.245E-08 | | New |
| 898 | 12 | 12-2946592-C-CTT | rs1221147 | TULP3, RNU6-1315P | 0.0034 | CTT | | 8.43E-16 | ?+ | 8.429E-16 | | New |
| 899 | 12 | 12-31366840-ACC-A | | NA | 0.0018 | ACC | | 3.8E-13 | ?+ | 3.798E-13 | | New |
| 900 | 12 | 12-31549775-ACC-A | | NA | 0.0002 | ACC | | 6.27E-09 | ?+ | 6.268E-09 | | New |

| | | | | | | | | | | | | |
|---|---|---|---|---|---|---|---|---|---|---|---|---|
| 901 | 12 | 12-32523802-G-A | rs5543691 | FGD4 | 0.0024 | A | 0.562503 | 2.28E-08 | -+ | 0.0006041 | 0.172 | New |
| 902 | 12 | 12-32797701-G-A | rs1456374 | PKP2 | 0.0008 | A | | 1.25E-08 | ?+ | 1.249E-08 | 0.3703 | New |
| 902 | 12 | 12-32797705-G-A | rs5276051 | PKP2 | 0.0007 | A | 0.875687 | 1.5E-09 | -+ | 9.433E-09 | 0.1634 | New |
| 903 | 12 | 12-37270198-G-T | rs1378297 | NONE, RF01518 | 0.001 | T | 0.915344 | 5.6E-09 | -+ | 0.0003097 | 0.3603 | New |
| 904 | 12 | 12-37359433-TTCTG-T | rs1385472 | NONE, RF01518 | 0.0017 | T | 0.392028 | 4.9E-13 | ++ | 3.383E-13 | 0.3236 | New |
| 905 | 12 | 12-37831033-GT-G | | NA | 0.0004 | GT | | 1.37E-09 | ?+ | 1.372E-09 | | New |
| 906 | 12 | 12-40483640-C-T | rs7407659 | AC107023.1, MUC19 | 0.2066 | T | 0.134069 | 2.07E-09 | -+ | 0.0248 | | New |
| 907 | 12 | 12-42207674-C-CTT | | NA | 0.001 | CTT | | 2.55E-09 | ?+ | 2.548E-09 | | New |
| 908 | 12 | 12-48988805-C-CT | | NA | 0.0006 | CT | | 3.28E-11 | ?+ | 3.279E-11 | | New |
| 909 | 12 | 12-49123265-C-T | rs5617750 | LMBR1L, AC011603.2 | 0.0037 | T | 0.390242 | 3.68E-09 | -+ | 0.02926 | 0.008676 | New |
| 910 | 12 | 12-49515941-G-A | rs1195385 | SPATS2 | 0.001 | A | 0.377129 | 1.64E-09 | ++ | 0.000003781 | | New |
| 910 | 12 | 12-49515961-G-A | rs1179715 | SPATS2 | 0.003 | G | 0.034456 | 1.02E-15 | ++ | 8.412E-10 | 0.7579 | New |
| 911 | 12 | 12-49611726-GT-G | rs1403284 | PRPF40B | 0.0068 | GT | | 6.22E-10 | ?+ | 6.218E-10 | 0.9896 | New |
| 912 | 12 | 12-50015024-C-CAG | | NA | 0.001 | C | | 6.21E-09 | ?+ | 6.214E-09 | | New |
| 913 | 12 | 12-50015583-C-CCT | | NA | 0.0014 | CCT | | 1.17E-08 | ?+ | 1.172E-08 | | New |
| 914 | 12 | 12-50234243-C-CT | | NA | 0.0002 | CT | | 5.7E-09 | ?+ | 5.704E-09 | | New |
| 915 | 12 | 12-50301831-GAGGC-G | | NA | 0.0009 | G | | 1.48E-08 | ?+ | 1.475E-08 | | New |
| 915 | 12 | 12-50301840-GGGCA-G | | NA | 0.0008 | GGGCA | | 3.1E-08 | ?+ | 3.105E-08 | | New |
| 915 | 12 | 12-50301871-C-T | rs1179018 | LIMA1, FAM186A | 0.0004 | C | | 1.73E-08 | ?+ | 1.726E-08 | | New |
| 916 | 12 | 12-51720075-CA-C | | SCN8A | 0.2618 | CA | | 5.86E-09 | ?- | 5.862E-09 | | New |
| 917 | 12 | 12-53132921-C-A | rs5367197 | SOAT2, RNU6-333P | 0.0011 | A | 0.758606 | 3.72E-09 | -+ | 0.0008108 | | New |
| 918 | 12 | 12-53434594-TTG-T | | NA | 0.0003 | TTG | | 8.32E-09 | ?+ | 8.319E-09 | | New |
| 919 | 12 | 12-53443104-ACCCG-A | | NA | 0.0003 | ACCCG | | 2.88E-08 | ?+ | 2.878E-08 | | New |

| | | | | | | | | | | | | |
|---|---|---|---|---|---|---|---|---|---|---|---|---|
| 920 | 12 | 12-53529371-T-A | rs1458520 | ATF7, ATF7-NPFF | 0.0014 | A | | 2.45E-10 | ?+ | 2.449E-10 | 0.7004 | New |
| 921 | 12 | 12-53537240-TG-T | | NA | 0.0002 | T | | 3.17E-11 | ?+ | 3.166E-11 | | New |
| 922 | 12 | 12-53673139-T-A | rs1373827 | ATP5MC2 | 0.0008 | A | 0.513582 | 1.56E-09 | -+ | 0.0003211 | 0.2313 | New |
| 923 | 12 | 12-54310262-G-A | rs5675002 | AC078778.1 | 0.0013 | A | 0.265957 | 1.94E-13 | -+ | 0.000002442 | | New |
| 924 | 12 | 12-54336354-C-CTA | | NA | 0.001 | C | | 1.12E-11 | ?+ | 1.123E-11 | | New |
| 925 | 12 | 12-55916145-G-A | rs9937491 | PYM1 | 0.0011 | A | 0.285354 | 3.28E-15 | ++ | 1.335E-10 | 0.05956 | New |
| 926 | 12 | 12-56536657-TGGTG-T | | NA | 0.0005 | T | | 1.83E-09 | ?+ | 1.83E-09 | | New |
| 927 | 12 | 12-56656740-T-A | rs1951611 | ATP5F1B, PTGES3 | 0.001 | A | | 1.35E-19 | ?+ | 1.348E-19 | | New |
| 927 | 12 | 12-56656759-G-C | | ATP5F1B, PTGES3 | 0.0005 | C | | 1.63E-10 | ?+ | 1.634E-10 | | New |
| 928 | 12 | 12-57017048-C-CTT | | NA | 0.0015 | CTT | | 3.33E-12 | ?+ | 3.327E-12 | | New |
| 929 | 12 | 12-64492451-C-CATG | | NA | 0.0006 | C | | 3.13E-10 | ?+ | 3.133E-10 | | New |
| 930 | 12 | 12-64614426-C-CTGTTTCTTAAAATTTCAACT | | NA | 0.0001 | CTGTTTCTTAAAATTTC | | 3.58E-08 | ?+ | 3.58E-08 | | New |
| 931 | 12 | 12-66938156-TTG-T | rs2041518 | GRIP1 | 0.001 | T | | 3.48E-10 | ?+ | 3.48E-10 | | New |
| 932 | 12 | 12-69759216-GT-G | rs1420997 | RAB3IP | 0.0016 | GT | 0.64759 | 4.1E-08 | -- | 0.00002672 | 0.3879 | New |
| 933 | 12 | 12-70418756-G-A | rs1868975 | KCNMB4 | 0.0003 | G | 0.969392 | 4.56E-09 | -+ | 0.000000166 | | New |
| 934 | 12 | 12-73182069-TA-T | | NA | 0.0009 | T | | 1.78E-09 | ?+ | 1.783E-09 | | New |
| 935 | 12 | 12-762205-AGG-A | | NA | 0.0004 | AGG | | 1.41E-08 | ?+ | 1.408E-08 | | New |
| 935 | 12 | 12-762206-C-CAT | | NA | 0.0004 | C | | 4.42E-09 | ?+ | 4.416E-09 | | New |
| 936 | 12 | 12-7660652-AAG-A | rs1376194 | APOBEC1 | 0.001 | A | | 3.09E-11 | ?+ | 3.094E-11 | 0.8001 | New |
| 937 | 12 | 12-7734982-C-CTT | | NA | 0.0044 | C | | 2.15E-21 | ?+ | 2.153E-21 | 0.1825 | New |
| 938 | 12 | 12-8009913-G-A | rs9765726 | RF00019, AC006511.4 | 0.0012 | G | | 8.29E-11 | ?+ | 8.293E-11 | | New |
| 939 | 12 | 12-89389651-G-T | rs1302697 | AC010201.3 | 0.0007 | T | | 1.51E-13 | ?+ | 1.513E-13 | | New |
| 940 | 12 | 12-92835051-TC-T | rs1397586 | EEA1 | 0.0024 | T | | 1.63E-14 | ?+ | 1.627E-14 | 0.5344 | New |
| 941 | 12 | 12-94946026-AT-A | | NA | 0.0007 | A | | 1.52E-09 | ?+ | 1.516E-09 | | New |

| | | | | | | | | | | | |
|---|---|---|---|---|---|---|---|---|---|---|---|
| 942 | 12 | 12-95612573-C-CAT | rs1461072 | USP44, RF00019 | 0.0035 | C | | 1.7E-12 | ?+ | 1.696E-12 | 0.4663 New |
| 943 | 13 | 13-100092421-TG-T | rs1196563 | PCCA | 0.001 | T | | 2.74E-08 | ?+ | 2.741E-08 | New |
| 944 | 13 | 13-101198906-G-T | rs1400411 | NALCN | 0.0005 | T | 0.826955 | 2.03E-08 | -+ | 0.0005086 | Old |
| 945 | 13 | 13-107175091-G-A | rs1311419 | FAM155A | 0.0022 | A | 0.308101 | 1.61E-08 | ++ | 0.0005019 | New |
| 946 | 13 | 13-110656083-ACC-A | | CARS2 | 0.0006 | A | | 4.8E-11 | ?+ | 4.802E-11 | New |
| 947 | 13 | 13-111960813-C-CGATG | | LINC00354, RF00287 | 0.1705 | C | | 1.5E-08 | ?+ | 1.501E-08 | New |
| 948 | 13 | 13-113370479-G-C | rs1118537 | GRTP1, ADPRHL1 | 0.0009 | C | 0.865158 | 1.33E-08 | ++ | 0.00001214 | New |
| 949 | 13 | 13-113600427-C-T | rs9604171 | TFDP1 | 0.0138 | C | | 1.63E-09 | ?+ | 1.631E-09 | New |
| 950 | 13 | 13-113795470-AC-A | rs1401920 | TMEM255B | 0.0039 | A | | 1.2E-11 | ?+ | 1.203E-11 | New |
| 951 | 13 | 13-114113552-C-CCAATCCACCCACCACGGCGAGTGATGTCCGTACAGCTCACACCGCATCCAT | rs1566582 | RASA3 | 0.0103 | C | | 4.61E-10 | ?+ | 4.614E-10 | New |
| 952 | 13 | 13-19187520-TGG-T | rs1306405 | TUBA3C, AL139327.2 | 0.0014 | TGG | | 4.28E-08 | ?+ | 4.281E-08 | 0.4347 New |
| 953 | 13 | 13-19922626-C-CCT | rs1485351 | AL355001.2, ZMYM2 | 0.003 | CCT | | 2.76E-19 | ?+ | 2.76E-19 | 0.6312 New |
| 954 | 13 | 13-20266218-C-T | rs1870974 | GJB6, CRYL1 | 0.0006 | C | 0.899642 | 8.87E-10 | -+ | 0.000001959 | New |
| 955 | 13 | 13-20470690-C-CCT | | NA | 0.0013 | C | | 1.62E-11 | ?+ | 1.621E-11 | New |
| 956 | 13 | 13-21627637-ATT-A | | NA | 0.0012 | A | | 3.39E-09 | ?+ | 3.392E-09 | New |
| 957 | 13 | 13-27054320-G-C | rs1392734 | AL160035.1, USP12 | 0.0009 | C | 0.223738 | 4.54E-09 | ++ | 1.039E-08 | 0.8133 New |
| 958 | 13 | 13-28596076-C-T | rs1166327 | AL139005.1, RF00019 | 0.0004 | C | 0.186211 | 1.71E-08 | ++ | 3.643E-07 | New |
| 959 | 13 | 13-32274495-C-CAGG | rs3608015 | FRY | 0.0603 | CAGG | 0.099864 | 2.57E-09 | -+ | 0.01649 | New |

| | | | | | | | | | | | | |
|---|---|---|---|---|---|---|---|---|---|---|---|---|
| 960 | 13 | 13-40685666-C-A | rs1033826 | FOXO1, MIR320D1 | 0.0007 | C | 0.893019 | 2.91E-10 | ++ | 2.055E-09 | | New |
| 961 | 13 | 13-41850639-C-T | rs1327088 | VWA8 | 0.0009 | C | 0.283844 | 5.41E-15 | ++ | 5.675E-15 | | New |
| 962 | 13 | 13-42917642-G-A | rs1995955 | EPSTI1 | 0.0031 | A | | 1.97E-09 | ?- | 1.971E-09 | | New |
| 963 | 13 | 13-44569304-G-C | rs7346576 | TSC22D1 | 0.0038 | C | 0.541382 | 1.77E-10 | -- | 2.476E-08 | | New |
| 964 | 13 | 13-45383701-G-C | rs1114073 | TPT1-AS1 | 0.0102 | C | | 9.88E-17 | ?+ | 9.878E-17 | | New |
| 965 | 13 | 13-49525435-ATG-A | | NA | 0.0004 | A | | 2.58E-10 | ?+ | 2.576E-10 | | New |
| 965 | 13 | 13-49525439-AAG-A | | NA | 0.0004 | AAG | | 2.11E-09 | ?+ | 2.115E-09 | | New |
| 966 | 13 | 13-79091376-G-A | rs1435170 | LINC00331, RBM26 | 0.0025 | G | | 9.48E-09 | ?- | 9.475E-09 | 0.4807 | New |
| 967 | 13 | 13-82155975-G-A | rs1017075 | AL353633.1, AL445255.1 | 0.0004 | A | | 3.41E-08 | ?+ | 3.413E-08 | | New |
| 968 | 13 | 13-97948951-C-CTG | rs1202324 | AL356580.1 | 0.0006 | C | | 4.99E-08 | ?+ | 4.989E-08 | | New |
| 969 | 13 | 13-98116951-GGA-G | rs1304078 | IPO5, FARP1 | 0.0026 | G | | 7.17E-14 | ?+ | 7.169E-14 | 0.3997 | New |
| 970 | 13 | 13-98643115-C-A | rs5770383 | STK24-AS1, RN7SL60P | 0.0011 | C | 0.732376 | 7.97E-13 | ++ | 3.071E-12 | | New |
| 971 | 13 | 13-99952110-C-CTT | rs2052996 | AL137139.1 | 0.0026 | CTT | | 3.18E-20 | ?+ | 3.18E-20 | | New |
| 971 | 13 | 13-99952112-C-CAG | rs1286972 | AL137139.1 | 0.0025 | C | | 5.3E-26 | ?+ | 5.302E-26 | | New |
| 971 | 13 | 13-99952132-AT-A | | NA | 0.0013 | A | | 1.22E-10 | ?+ | 1.223E-10 | | New |
| 972 | 14 | 14-100301031-C-A | rs1167605 | SLC25A29 | 0.0009 | A | 0.70354 | 2.21E-11 | ++ | 2.826E-10 | 0.3792 | New |
| 973 | 14 | 14-102136332-GCT-G | | NA | 0.0003 | GCT | | 2.2E-09 | ?+ | 2.2E-09 | | New |
| 974 | 14 | 14-102541608-AG-A | rs1389132 | MIR4309, LINC02323 | 0.0012 | AG | | 5.04E-15 | ?+ | 5.043E-15 | 0.1863 | New |
| 975 | 14 | 14-102809721-G-T | rs1040914 | TRAF3 | 0.0018 | T | 0.674686 | 4.07E-13 | ++ | 2.034E-07 | 0.5167 | New |
| 976 | 14 | 14-103272252-C-CCT | | NA | 0.0004 | C | | 5.36E-10 | ?+ | 5.36E-10 | | New |
| 977 | 14 | 14-103601925-GGT-G | rs1290948 | AL139300.1, KLC1 | 0.0034 | GGT | | 2.12E-11 | ?+ | 2.115E-11 | 0.2817 | New |

| | | | | | | | | | | | | |
|---|---|---|---|---|---|---|---|---|---|---|---|---|
| 977 | 14 | 14-103601927-C-CAG | rs1385983 | AL139300.1, KLC1 | 0.0038 | C | | 1.52E-11 | ?+ | 1.523E-11 | | New |
| 978 | 14 | 14-103655446-GCA-G | rs1444368 | AL139300.1, KLC1 | 0.0026 | GCA | | 1.67E-12 | ?+ | 1.674E-12 | 0.2188 | New |
| 979 | 14 | 14-104209450-G-C | rs1223739 | KIF26A, LINC02691 | 0.0005 | G | | 1.32E-09 | ?+ | 1.321E-09 | | New |
| 980 | 14 | 14-104847993-G-T | rs1862181 | AL583810.2, AL583810.1 | 0.0011 | T | 0.272319 | 4.92E-16 | -+ | 1.934E-13 | | New |
| 981 | 14 | 14-104945575-C-A | rs3732076 | AHNAK2 | 0.0037 | A | 0.437536 | 3.27E-10 | -+ | 0.005798 | 0.6436 | New |
| 982 | 14 | 14-104951987-G-T | rs7709113 | AHNAK2 | 0.0012 | G | 0.333787 | 4.41E-08 | ++ | 3.788E-08 | | New |
| 983 | 14 | 14-105296067-C-CAA | | NA | 0.0014 | CAA | | 1.48E-16 | ?+ | 1.485E-16 | | Old |
| 983 | 14 | 14-105296075-T-C | rs4347564 | BRF1 | 0.0025 | C | | 6.77E-13 | ?+ | 6.768E-13 | | Old |
| 984 | 14 | 14-105650853-C-CATGGAACAGGAAGCATCCAGGATGGAACAGGAAGCATCCAGT | rs1265782 | AL928742.1, IGHA1 | 0.0135 | C | | 1.21E-08 | ?+ | 1.212E-08 | | Old |
| 985 | 14 | 14-20342296-C-CTCG | | AL355075.4, RPPH1 | 0.0012 | C | | 3.35E-15 | ?+ | 3.351E-15 | 0.2569 | New |
| 986 | 14 | 14-22727949-C-T | rs2038153 | AL160314.2 | 0.0003 | C | 0.392326 | 1.36E-10 | -+ | 2.329E-08 | | New |
| 987 | 14 | 14-22801564-ACT-A | | NA | 0.001 | ACT | | 1.26E-11 | ?+ | 1.259E-11 | | New |
| 988 | 14 | 14-23706575-G-A | rs1482168 | AL160237.1 | 0.0037 | A | 0.097764 | 5.57E-19 | ++ | 1.978E-19 | 0.9641 | New |
| 988 | 14 | 14-23706600-G-A | rs9033370 | AL160237.1 | 0.0053 | A | 0.869328 | 1.57E-23 | -+ | 3.728E-11 | 0.4594 | New |
| 988 | 14 | 14-23706607-G-A | rs1468960 | AL160237.1 | 0.0084 | A | 0.806871 | 1.3E-24 | ++ | 1.56E-14 | | New |
| 989 | 14 | 14-24170048-GT-G | rs1281146 | IRF9, REC8 | 0.0008 | G | | 1.27E-08 | ?+ | 1.266E-08 | | New |
| 990 | 14 | 14-28854133-GGC-G | | NA | 0.0013 | G | | 7.29E-11 | ?+ | 7.293E-11 | | New |
| 990 | 14 | 14-28854137-C-CGT | | NA | 0.0009 | C | | 1.83E-08 | ?+ | 1.83E-08 | | New |
| 990 | 14 | 14-28854140-GCCCA-G | | NA | 0.0008 | GCCCA | | 2E-08 | ?+ | 2.001E-08 | | New |

| | | | | | | | | | | | | |
|---|---|---|---|---|---|---|---|---|---|---|---|---|
| 991 | 14 | 14-31245601-G-A | rs1421885 | RF00019 | 0.3339 | A | | 7.97E-10 | ?+ | 7.967E-10 | 0.5199 | New |
| 992 | 14 | 14-34487122-TA-T | rs1264221 | SPTSSA, EAPP | 0.0006 | T | | 1.89E-09 | ?+ | 1.894E-09 | 0.1006 | New |
| 992 | 14 | 14-34487130-TTC-T | rs1175681 | SPTSSA, EAPP | 0.0011 | T | | 3.99E-11 | ?+ | 3.993E-11 | 0.1005 | New |
| 993 | 14 | 14-34514025-C-CGTG | | NA | 0.0003 | C | | 1.5E-10 | ?+ | 1.497E-10 | | New |
| 994 | 14 | 14-34565234-C-T | rs1012535 | SNX6 | 0.003 | T | 0.071581 | 3.05E-16 | ++ | 7.185E-09 | 0.3573 | New |
| 994 | 14 | 14-34565238-C-T | rs3766904 | SNX6 | 0.0031 | T | 0.048577 | 5.15E-14 | ++ | 3.009E-08 | 0.4979 | New |
| 994 | 14 | 14-34565241-C-T | rs1370017 | SNX6 | 0.003 | T | 0.069 | 1.36E-18 | ++ | 1.534E-10 | | New |
| 995 | 14 | 14-39101400-ACC-A | | NA | 0.002 | ACC | | 2.71E-12 | ?+ | 2.706E-12 | | Old |
| 996 | 14 | 14-54925838-C-A | rs1472520 | GCH1, WDHD1 | 0.0004 | A | 0.957478 | 5.07E-09 | ++ | 1.654E-08 | | New |
| 997 | 14 | 14-55344909-TTGTC-T | rs1393967 | FBXO34 | 0.0011 | T | 0.259637 | 6.86E-09 | ++ | 2.336E-08 | 0.2689 | New |
| 998 | 14 | GCCAAGAAGAAC-G | | NA | 0.0018 | GCCAAGAAGAAC | | 3.64E-08 | ?+ | 3.636E-08 | | New |
| 999 | 14 | 14-63585644-G-T | rs3740499 | AL136038.3, WDR89 | 0.0015 | T | 0.241319 | 3.43E-17 | -+ | 4.064E-12 | 0.8818 | New |
| 999 | 14 | 14-63585648-G-T | rs7733348 | AL136038.3, WDR89 | 0.001 | T | 0.241182 | 2.18E-13 | -+ | 1.745E-08 | 0.9271 | New |
| 1000 | 14 | 14-64547356-ACT-A | | HSPA2, PPP1R36 | 0.003 | A | | 1.33E-09 | ?+ | 1.328E-09 | 0.7422 | New |
| 1001 | 14 | 14-65504716-ATTT-A | | NA | 0.0007 | A | | 2.32E-08 | ?+ | 2.322E-08 | | New |
| 1002 | 14 | 14-66995017-TTA-T | | NA | 0.0012 | T | | 4.16E-12 | ?+ | 4.161E-12 | | New |
| 1003 | 14 | 14-71326334-G-A | rs1327449 | SIPA1L1 | 0.0007 | A | 0.787902 | 1.22E-08 | ++ | 6.131E-07 | 0.1193 | New |
| 1003 | 14 | 14-71326352-GCACA-G | | SIPA1L1 | 0.0011 | G | | 4.72E-10 | ?+ | 4.725E-10 | 0.1383 | New |
| 1003 | 14 | 14-71326357-C-CGTGT | | SIPA1L1 | 0.0009 | CGTGT | | 1.19E-10 | ?+ | 1.189E-10 | | New |
| 1004 | 14 | 14-72837563-C-CCCCA | | NA | 0.0015 | C | | 3.41E-08 | ?+ | 3.405E-08 | | Old |
| 1005 | 14 | 14-73480223-C-CTT | | NA | 0.0005 | C | | 5.28E-09 | ?+ | 5.28E-09 | | Old |
| 1006 | 14 | 14-73952289-ATT-A | rs1339702 | COQ6 | 0.0015 | ATT | | 2.38E-14 | ?+ | 2.383E-14 | 0.5945 | New |

| | | | | | | | | | | | | |
|---|---|---|---|---|---|---|---|---|---|---|---|---|
| 1007 | 14 | 14-77308036-T-A | rs1346324 | POMT2 | 0.0009 | T | 0.663789 | 6.99E-09 | ++ | 6.912E-07 | 0.9347 | New |
| 1008 | 14 | 14-77850817-ATTTTTT-A | rs8797396 | ADCK1 | 0.0034 | A | | 7.88E-18 | ?+ | 7.884E-18 | | New |
| 1009 | 14 | 14-78832014-G-A | rs1311911 | NRXN3 | 0.0013 | G | 0.307203 | 2.46E-27 | -+ | 1.01E-22 | 0.09501 | New |
| 1010 | 14 | 14-81923555-AATTTT-A | rs1378802 | AL355838.1 | 0.0045 | A | 0.818303 | 3.23E-13 | -+ | 1.223E-09 | 0.8469 | Old |
| 1011 | 14 | 14-88557948-C-T | rs1217460 | AL162171.1, ZC3H14 | 0.0012 | C | 0.167637 | 2.71E-11 | ++ | 1.291E-10 | | New |
| 1011 | 14 | 14-88557971-G-A | rs1299853 | AL162171.1, ZC3H14 | 0.0014 | A | | 5.47E-10 | ?+ | 5.467E-10 | | New |
| 1012 | 14 | 14-91460172-C-CCT | | NA | 0.0008 | CCT | | 1.45E-08 | ?+ | 1.454E-08 | | New |
| 1012 | 14 | 14-91460178-ATG-A | | NA | 0.0005 | A | | 3.83E-08 | ?+ | 3.829E-08 | | New |
| 1013 | 14 | 14-91507096-ATCAC-A | | NA | 0.0007 | A | | 8.29E-15 | ?+ | 8.287E-15 | | New |
| 1014 | 14 | 14-92741777-G-T | rs1044208 | LGMN | 0.0005 | T | | 4.28E-14 | ?+ | 4.284E-14 | 0.5189 | Old |
| 1015 | 14 | 14-92804358-GC-G | | NA | 0.0003 | GC | | 5.19E-12 | ?+ | 5.188E-12 | | Old |
| 1015 | 14 | 14-92804364-C-CAACA | | NA | 0.0001 | CAACA | | 5.16E-09 | ?+ | 5.162E-09 | | Old |
| 1015 | 14 | 14-92804368-C-CTT | | NA | 0.0002 | C | | 3.14E-08 | ?+ | 3.141E-08 | | Old |
| 1016 | 15 | 15-22986557-C-A | rs1163607 | TUBGCP5 | 0.0006 | A | 0.922051 | 3.72E-11 | ++ | 1.886E-07 | | New |
| 1017 | 15 | 15-23448602-C-CAT | | NA | 0.0013 | C | | 2.14E-09 | ?+ | 2.138E-09 | | New |
| 1017 | 15 | 15-23448605-GCC-G | | NA | 0.0013 | GCC | | 5.06E-10 | ?+ | 5.065E-10 | | New |
| 1017 | 15 | 15-23448633-AG-A | | NA | 0.0011 | AG | | 1.4E-11 | ?+ | 1.398E-11 | | New |
| 1018 | 15 | 15-23838964-C-CCT | | NA | 0.0008 | C | | 2.08E-11 | ?+ | 2.081E-11 | | New |
| 1019 | 15 | 15-25006938-G-A | rs1267131 | SNHG14 | 0.0003 | A | 0.163188 | 2.01E-10 | ++ | 1.124E-10 | 0.4056 | New |
| 1020 | 15 | 15-25293267-T-A | rs5373742 | SNHG14 | 0.0022 | A | 0.877751 | 1.5E-08 | ++ | 0.01218 | 0.1415 | New |
| 1020 | 15 | 15-25293273-C-A | rs1191974 | SNHG14 | 0.0007 | C | | 4.57E-08 | ?+ | 4.571E-08 | 0.1573 | New |
| 1021 | 15 | 15-28367284-A-ATTTTT | | HERC2, GOLGA8F | 0.2881 | A | | 4.99E-08 | ?+ | 4.991E-08 | | New |

| | | | | | | | | | | | | |
|---|---|---|---|---|---|---|---|---|---|---|---|---|
| 1022 | 15 | 15-28784698-C-T | rs5699439 | GOLGA8M, GOLGA6L7 | 0.0573 | C | | 6.12E-09 | ?+ | 6.116E-09 | 0.5895 | New |
| 1023 | 15 | 15-29739562-G-A | rs1464017 | TJP1 | 0.0017 | A | | 2.4E-17 | ?+ | 2.403E-17 | | New |
| 1024 | 15 | 15-32275965-T-C | rs7691839 | AC068448.1, RNU6-18P | 0.0081 | T | | 3.15E-10 | ?+ | 3.147E-10 | | New |
| 1025 | 15 | 15-34175801-G-T | rs1889312 | KATNBL1 | 0.0006 | T | 0.541198 | 5.16E-09 | ++ | 8.354E-07 | | New |
| 1026 | 15 | 15-34175843-TAAGAA-T | | NA | 0.0016 | TAAGAA | | 6.23E-14 | ?+ | 6.229E-14 | | New |
| 1027 | 15 | 15-40502504-TCA-T | rs1172600 | AC091045.1 | 0.002 | T | | 2.07E-09 | ?+ | 2.073E-09 | 0.7762 | New |
| 1028 | 15 | 15-40646627-G-C | | AC022405.1 | 0.0001 | C | | 1.45E-09 | ?+ | 1.453E-09 | | New |
| 1029 | 15 | 15-41344055-TA-T | | NA | 0.0007 | TA | | 9.82E-14 | ?+ | 9.823E-14 | | New |
| 1030 | 15 | 15-41378616-C-CAA | | NA | 0.0004 | CAA | | 2.94E-09 | ?+ | 2.939E-09 | | New |
| 1031 | 15 | 15-41525481-GGCAT-G | | NA | 0.002 | G | | 2.12E-11 | ?+ | 2.121E-11 | | New |
| 1032 | 15 | 15-43723919-G-T | rs1385582 | RNU6-353P, RNU6-354P | 0.0007 | T | | 8.28E-09 | ?+ | 8.28E-09 | 0.1803 | New |
| 1033 | 15 | 15-44183022-TCTC-T | rs1291362 | FRMD5 | 0.0007 | T | | 3.65E-12 | ?+ | 3.647E-12 | 0.2447 | New |
| 1034 | 15 | 15-44777318-G-T | rs1199092 | AC122108.2 | 0.0012 | T | | 4.41E-08 | ?+ | 4.414E-08 | 0.8628 | New |
| 1034 | 15 | 15-44777319-TG-T | rs1255811 | AC122108.2 | 0.0016 | TG | | 2.62E-08 | ?+ | 2.62E-08 | 0.8559 | New |
| 1035 | 15 | 15-49117476-G-T | rs1029888 | COPS2 | 0.0006 | T | 0.746148 | 8.74E-09 | ++ | 0.00001197 | | New |
| 1036 | 15 | 15-52015119-G-A | rs1003542 | MAPK6-DT | 0.0008 | A | 0.216002 | 6.87E-11 | -+ | 0.001096 | 0.7742 | New |
| 1036 | 15 | 15-52015126-G-A | rs1031529 | MAPK6-DT | 0.0004 | A | | 1.01E-10 | ?+ | 1.012E-10 | | New |
| 1037 | 15 | 15-55441294-T-A | | DNAAF4-CCPG1 | 0.0003 | T | | 3.34E-09 | ?+ | 3.337E-09 | | New |
| 1038 | 15 | 15-55641955-ACC-A | | NA | 0.0006 | ACC | | 4.71E-12 | ?+ | 4.713E-12 | 0.3157 | New |
| 1039 | 15 | 15-63272088-C-CTT | | NA | 0.0006 | CTT | | 2.66E-09 | ?+ | 2.659E-09 | | Old |

| | | | | | | | | | | | | |
|---|---|---|---|---|---|---|---|---|---|---|---|---|
| 1040 | 15 | 15-64372438-C-T | rs1371331 | AC087632.1, PCLAF | 0.0048 | T | | 8.06E-09 | ?+ | 8.063E-09 | 0.1559 | Old |
| 1041 | 15 | 15-64541359-G-A | rs1408797 | ZNF609 | 0.0066 | A | 0.674627 | 6.66E-09 | -+ | 0.0656 | | Old |
| 1042 | 15 | 15-64731383-C-T | rs5646245 | AC100830.1, RBPMS2 | 0.0013 | C | 0.679829 | 1.13E-10 | ++ | 0.0001259 | 0.4511 | Old |
| 1043 | 15 | 15-65454698-TG-T | | NA | 0.0007 | T | | 1.55E-11 | ?+ | 1.546E-11 | | New |
| 1043 | 15 | 15-65454700-TAACTC-T | | NA | 0.0005 | TAACTC | | 2.07E-08 | ?+ | 2.065E-08 | | New |
| 1044 | 15 | 15-66380772-G-A | rs1440223 | TIPIN | 0.0047 | G | 0.825091 | 1.36E-11 | ++ | 0.002261 | | New |
| 1045 | 15 | 15-69587320-G-T | rs1421805 | DRAIC | 0.0016 | T | 0.286118 | 1.73E-10 | -+ | 0.02364 | 0.4318 | New |
| 1046 | 15 | 15-70684100-C-T | rs1015955 | UACA | 0.0019 | T | 0.643348 | 4.09E-10 | -+ | 0.0008459 | 0.6989 | New |
| 1047 | 15 | 15-74757553-AT-A | rs1330271 | CYP1A2 | 0.0038 | A | | 1.38E-14 | ?+ | 1.376E-14 | 0.7961 | New |
| 1048 | 15 | 15-74921556-C-CCG | rs1283490 | COX5A | 0.0012 | C | | 6.93E-09 | ?+ | 6.931E-09 | | New |
| 1049 | 15 | 15-74953400-G-C | | COX5A, RPP25 | 0.0003 | C | | 5.27E-10 | ?+ | 5.272E-10 | | New |
| 1050 | 15 | 15-75575726-TGG-T | rs1295714 | AC105036.3 | 0.0059 | T | 0.521318 | 2.2E-10 | ++ | 2.913E-10 | 0.277 | New |
| 1051 | 15 | 15-76148723-CT-C | rs5813819 | TMEM266 | 0.0138 | C | | 2.81E-09 | ?+ | 2.806E-09 | | New |
| 1052 | 15 | 15-76317437-G-A | rs1450411 | ETFA, ISL2 | 0.0007 | A | 0.227395 | 6.05E-09 | ++ | 8.493E-09 | 0.2553 | New |
| 1053 | 15 | 15-77699496-TCC-T | | NA | 0.0013 | T | | 7.29E-12 | ?+ | 7.295E-12 | | New |
| 1054 | 15 | 15-78856334-TTA-T | | MORF4L1 | 0.0013 | TTA | | 3.64E-09 | ?+ | 3.636E-09 | | Old |
| 1055 | 15 | 15-79951126-C-CAGAGCGG | rs1252579 | ST20-AS1, BCL2A1 | 0.0028 | CAGAGCGG | | 2.86E-15 | ?+ | 2.858E-15 | | New |
| 1056 | 15 | 15-89020231-G-C | rs1985691 | MFGE8, AC013565.1 | 0.3172 | G | | 3.95E-08 | ?+ | 3.945E-08 | 0.864 | New |
| 1057 | 15 | 15-90157089-GCA-G | | NA | 0.0002 | GCA | | 4.57E-09 | ?+ | 4.573E-09 | | New |
| 1057 | 15 | 15-90157090-TG-T | | NA | 0.0002 | TG | | 2.48E-09 | ?+ | 2.479E-09 | | New |
| 1057 | 15 | 15-90157092-TA-T | | NA | 0.0002 | TA | | 7.95E-10 | ?+ | 7.952E-10 | | New |
| 1058 | 15 | 15-90505467-C-CAA | rs1476023 | IQGAP1, CRTC3 | 0.0021 | CAA | 0.362848 | 8.49E-13 | ++ | 1.977E-12 | | New |
| 1059 | 15 | 15-92850975-AT-A | | NA | 0.0007 | A | | 2.51E-08 | ?+ | 2.513E-08 | 0.3821 | New |
| 1060 | 15 | 15-99043874-C-T | rs1555476 | AC036108.1 | 0.0003 | C | | 1.03E-09 | ?+ | 1.031E-09 | | Old |
| 1061 | 16 | 16-10645333-G-A | rs1555465 | TEKT5 | 0.0005 | G | | 1.13E-11 | ?+ | 1.127E-11 | | New |

| | | | | | | | | | | | | |
|---|---|---|---|---|---|---|---|---|---|---|---|---|
| 1061 | 16 | 16-10645335-C-A | rs5683197 | TEKT5 | 0.0003 | C | | 1.74E-08 | ?+ | 1.735E-08 | | New |
| 1062 | 16 | 16-10734878-G-A | rs9559816 | TEKT5, NUBP1 | 0.0009 | A | | 2.94E-08 | ?+ | 2.936E-08 | | New |
| 1063 | 16 | 16-11232146-TGC-T | | NA | 0.0002 | TGC | | 2.3E-08 | ?+ | 0.000000023 | | New |
| 1064 | 16 | 16-11808833-C-CA | | NA | 0.0006 | CA | 0.502887 | 5.26E-10 | ++ | 0.000003627 | | New |
| 1065 | 16 | 16-1190872-GTC-G | rs7594179 | CACNA1H | 0.0127 | G | | 2.39E-09 | ?+ | 2.389E-09 | | New |
| 1066 | 16 | 16-14532898-GC-G | rs1438440 | PARN | 0.0018 | GC | | 2.54E-08 | ?+ | 2.543E-08 | | New |
| 1067 | 16 | 16-1478801-C-CCA | | NA | 0.0003 | C | | 3.98E-08 | ?+ | 3.98E-08 | | New |
| 1068 | 16 | 16-15359814-GCACACACACACA-G | | NA | 0.0056 | G | | 5.39E-10 | ?+ | 5.388E-10 | | New |
| 1069 | 16 | 16-1569073-C-T | rs9927187 | IFT140 | 0.152 | T | 0.599786 | 1.86E-08 | -+ | 0.002108 | | New |
| 1070 | 16 | 16-16171817-GGGAT-G | rs1260579 | ABCC6 | 0.0039 | G | 0.078946 | 4.83E-08 | -+ | 0.00001767 | | New |
| 1071 | 16 | 16-16994683-C-T | rs5409807 | AC092326.1, AC109446.4 | 0.0021 | T | 0.557708 | 1.4E-08 | -+ | 0.0005663 | 0.2191 | New |
| 1072 | 16 | 16-18158393-T-A | | AC091489.1 | 0.0003 | A | | 6.69E-11 | ?+ | 6.686E-11 | | New |
| 1073 | 16 | 16-199543-T-A | rs1350229 | LUC7L | 0.001 | A | 0.940299 | 2.86E-11 | -+ | 9.954E-10 | 0.4181 | New |
| 1074 | 16 | 16-20408737-GCA-G | | NA | 0.0022 | G | | 6.29E-16 | ?+ | 6.295E-16 | 0.954 | New |
| 1075 | 16 | 16-2099049-C-CT | rs1375283 | PKD1 | 0.0012 | CT | 0.317753 | 1.07E-09 | ++ | 2.421E-07 | | New |
| 1076 | 16 | 16-21343238-C-CAAA | | CRYM, NPIPB3 | 0.0643 | CAAA | | 1.63E-08 | ?+ | 1.631E-08 | | New |
| 1077 | 16 | 16-21894065-TTGAG-T | | NA | 0.0006 | T | | 6.36E-12 | ?+ | 6.358E-12 | | New |
| 1078 | 16 | 16-23588279-G-A | rs9453796 | NDUFAB1 | 0.0007 | G | 0.468786 | 1.38E-12 | ++ | 1.5E-09 | | New |
| 1079 | 16 | 16-23736646-AAGTGT-A | | NA | 0.0013 | AAGTGT | | 6.36E-14 | ?+ | 6.361E-14 | | New |
| 1079 | 16 | 16-23736647-GGCACC-G | | NA | 0.001 | G | | 9.34E-12 | ?+ | 9.343E-12 | | New |
| 1080 | 16 | 16-24905612-AGG-A | | NA | 0.0003 | AGG | | 2.01E-09 | ?+ | 2.01E-09 | | New |
| 1081 | 16 | 16-25154176-AT-A | rs1248734 | LCMT1 | 0.0017 | A | 0.457416 | 6.24E-09 | ++ | 3.505E-08 | | New |
| 1082 | 16 | 16-28817462-C-CAT | | NA | 0.0013 | C | | 5.59E-15 | ?+ | 5.592E-15 | | New |
| 1082 | 16 | 16-28817467-GCA-G | | NA | 0.0011 | GCA | | 2.65E-12 | ?+ | 2.647E-12 | | New |

| | | | | | | | | | | | | |
|---|---|---|---|---|---|---|---|---|---|---|---|---|
| 1083 | 16 | 16-2891973-C-CTT | | NA | 0.0026 | C | | 5.19E-12 | ?+ | 5.191E-12 | | New |
| 1083 | 16 | 16-2891980-TTGGG-T | | NA | 0.002 | T | | 1.13E-15 | ?+ | 1.128E-15 | | New |
| 1083 | 16 | 16-2891984-GCAAC-G | | NA | 0.0015 | GCAAC | | 5.94E-12 | ?+ | 5.942E-12 | | New |
| 1084 | 16 | 16-28993307-G-A | rs1117088 | LAT, RRN3P2 | 0.2074 | G | | 6.38E-18 | ?+ | 6.379E-18 | | New |
| 1085 | 16 | 16-29573680-G-A | rs5577089 | NPIPB12, AC009086.2 | 0.0019 | G | 0.926787 | 2.79E-11 | ++ | 0.0001275 | 0.8204 | Old |
| 1086 | 16 | 16-29786163-C-CA | rs1567352 | ZG16, KIF22 | 0.0008 | C | 0.061174 | 1.67E-13 | ++ | 1.015E-13 | | Old |
| 1087 | 16 | 16-29831442-C-CCG | | NA | 0.0006 | CCG | | 2.66E-15 | ?+ | 2.659E-15 | | Old |
| 1088 | 16 | 16-2983500-C-CA | | AC004233.3, LINC00514 | 0.0013 | CA | | 2.1E-09 | ?+ | 2.099E-09 | | New |
| 1089 | 16 | 16-30142456-AGG-A | | NA | 0.0005 | AGG | | 2.92E-08 | ?+ | 2.919E-08 | | Old |
| 1090 | 16 | 16-30441816-C-CAG | | NA | 0.0011 | C | | 4.88E-12 | ?+ | 4.883E-12 | | Old |
| 1091 | 16 | 16-30625405-C-CCT | rs2052164 | ZNF689, PRR14 | 0.0021 | CCT | | 6.82E-10 | ?+ | 6.816E-10 | | Old |
| 1092 | 16 | 16-31041327-G-A | rs1241213 | STX4, AC135050.3 | 0.0122 | G | 0.586219 | 1.35E-09 | -+ | 0.1762 | 0.968 | Old |
| 1093 | 16 | 16-311114-G-GCT | rs5881041 | AXIN1 | 0.0008 | GCT | | 7.89E-11 | ?+ | 7.889E-11 | | New |
| 1093 | 16 | 16-311117-G-GC | rs6781615 | AXIN1 | 0.0007 | GC | 0.430101 | 8.8E-10 | ++ | 5.706E-08 | | New |
| 1094 | 16 | 16-32421947-CA-C | | AC133485.2, AC138915.3 | 0.4383 | CA | | 2.54E-14 | ?+ | 2.539E-14 | | New |
| 1095 | 16 | 16-34249700-C-CTT | | NA | 0.0003 | C | | 3.97E-08 | ?+ | 3.974E-08 | | New |
| 1096 | 16 | 16-34260272-C-A | rs1394896 | LINC00273, AC135776.4 | 0.0007 | C | | 1.23E-12 | ?+ | 1.235E-12 | 0.7918 | New |
| 1097 | 16 | 16-34284927-TTC-T | | NA | 0.0025 | T | | 5.69E-12 | ?+ | 5.691E-12 | | New |

| | | | | | | | | | | | | |
|---|---|---|---|---|---|---|---|---|---|---|---|---|
| 1098 | 16 | 16-34574822-TTGAAAACAAA-T | | NA | 0.0045 | TTGAAAACAAA | | 1.16E-08 | ?+ | 1.159E-08 | | New |
| 1099 | 16 | 16-34630340-AGTCCATT-A | | NA | 0.0052 | A | | 2.44E-46 | ?+ | 2.439E-46 | | New |
| 1100 | 16 | 16-35901414-G-A | rs1596645 | AC106785.2, AC116553.2 | 0.0006 | G | | 1.7E-10 | ?+ | 1.704E-10 | | New |
| 1100 | 16 | 16-35901425-G-A | rs1462977 | AC106785.2, AC116553.2 | 0.0006 | G | | 1.67E-09 | ?+ | 1.674E-09 | | New |
| 1100 | 16 | 16-35901442-G-A | rs1391812 | AC106785.2, AC116553.2 | 0.0003 | A | 0.319765 | 1.73E-08 | ++ | 1.089E-08 | | New |
| 1101 | 16 | 16-35971942-G-T | rs6205915 | AC116553.2, NONE | 0.0005 | T | 0.253278 | 2.59E-08 | ++ | 0.000004332 | 0.3173 | New |
| 1102 | 16 | 16-36256874-TAC-T | | NA | 0.0018 | TAC | | 6.11E-11 | ?+ | 6.112E-11 | | New |
| 1103 | 16 | 16-3923860-G-C | | CREBBP, AC005736.1 | 0.0004 | G | | 1.14E-09 | ?+ | 1.138E-09 | | New |
| 1104 | 16 | 16-422947-C-CAG | | NA | 0.0005 | CAG | | 1.64E-08 | ?+ | 1.637E-08 | | New |
| 1105 | 16 | 16-4590542-GA-G | | NA | 0.0005 | G | | 3.9E-08 | ?+ | 3.899E-08 | | New |
| 1106 | 16 | 16-46395598-T-C | rs1022113 | NONE, RNU6-845P | 0.0006 | T | | 7.84E-11 | ?+ | 7.835E-11 | | New |
| 1106 | 16 | 16-46395604-A-C | rs1333759 | NONE, RNU6-845P | 0.0007 | A | | 1.49E-12 | ?+ | 1.489E-12 | | New |
| 1107 | 16 | 16-4771009-C-CAA | | NA | 0.0008 | CAA | | 2.94E-09 | ?+ | 2.94E-09 | | New |
| 1108 | 16 | 16-50150533-ATT-A | rs2037732 | AC007610.5, RF00156 | 0.0021 | A | | 2.23E-18 | ?+ | 2.232E-18 | | New |
| 1109 | 16 | 16-50803378-G-T | rs1240963 | AC007728.1 | 0.003 | T | 0.748552 | 2.87E-11 | -+ | 0.01102 | 0.7686 | New |
| 1109 | 16 | 16-50803398-C-A | rs9414517 | AC007728.1 | 0.0049 | C | 0.201125 | 2.45E-13 | -+ | 0.02381 | 0.2663 | New |
| 1110 | 16 | 16-58440874-C-CAT | rs1407236 | LINC02137 | 0.0005 | CAT | | 4.63E-08 | ?+ | 4.633E-08 | 0.3518 | New |

| | | | | | | | | | | | | |
|---|---|---|---|---|---|---|---|---|---|---|---|---|
| 1111 | 16 | 16-619328-ATG-A | rs1244433 | RAB40C | 0.0014 | A | | 6.54E-09 | ?+ | 6.542E-09 | | New |
| 1112 | 16 | 16-67096756-C-CCT | | NA | 0.0004 | CCT | | 7.72E-09 | ?+ | 7.722E-09 | | New |
| 1113 | 16 | 16-67606533-G-A | rs5733786 | CTCF | 0.0446 | A | | 1.67E-17 | ?+ | 1.668E-17 | 0.3788 | New |
| 1114 | 16 | 16-67610589-C-CCG | | NA | 0.0008 | CCG | 0.659432 | 4.77E-09 | -+ | 2.584E-08 | | New |
| 1115 | 16 | 16-67849799-C-T | rs1461907 | NUTF2 | 0.0004 | T | 0.243695 | 8.89E-11 | -+ | 2.461E-08 | | New |
| 1115 | 16 | 16-67849804-G-A | rs7630879 | NUTF2 | 0.0005 | A | 0.430058 | 6.96E-14 | ++ | 1.328E-12 | | New |
| 1116 | 16 | 16-68401966-G-C | rs1220097 | SMPD3 | 0.001 | G | 0.362994 | 1.09E-10 | -+ | 0.000364 | | New |
| 1117 | 16 | 16-69576084-C-CTT | | NA | 0.0009 | CTT | | 6.03E-10 | ?+ | 6.033E-10 | | New |
| 1118 | 16 | 16-70218561-CA-C | | CLEC18C, EXOSC6 | 0.4067 | C | | 1.05E-08 | ?- | 1.047E-08 | | Old |
| 1119 | 16 | 16-70384477-G-C | rs1267581 | AC012184.1 | 0.0013 | C | 0.011317 | 9.52E-15 | ++ | 3.742E-16 | 0.6082 | Old |
| 1120 | 16 | 16-73895079-AG-A | | NA | 0.0003 | A | | 4.07E-08 | ?+ | 4.07E-08 | | New |
| 1121 | 16 | 16-74615312-C-CAG | | NA | 0.001 | CAG | | 1.57E-08 | ?+ | 1.57E-08 | | New |
| 1122 | 16 | 16-754494-C-CA | | NA | 0.001 | CA | | 1.4E-12 | ?+ | 1.403E-12 | 0.1498 | New |
| 1123 | 16 | 16-81105873-GGTC-G | | NA | 0.0035 | GGTC | 0.34038 | 1.01E-20 | ++ | 1.385E-19 | | New |
| 1124 | 16 | 16-87884404-G-C | rs1013953 | CA5A | 0.0005 | C | 0.959602 | 6.13E-09 | ++ | 4.858E-08 | | New |
| 1125 | 16 | 16-89188362-C-CGG | rs1996383 | CDH15 | 0.0027 | C | | 6.86E-10 | ?+ | 6.861E-10 | | New |
| 1126 | 16 | 16-89479635-C-A | rs3687765 | ANKRD11 | 0.0006 | A | | 1.89E-08 | ?+ | 1.885E-08 | | New |
| 1127 | 16 | 16-89650787-C-CAG | | NA | 0.0008 | CAG | | 1.25E-10 | ?+ | 1.252E-10 | | Old |
| 1128 | 16 | 16-89831052-G-T | rs1220150 | SPIRE2 | 0.0097 | T | 0.724107 | 7.05E-26 | ++ | 3.267E-07 | | Old |
| 1129 | 16 | 16-89831588-G-T | rs9607019 | SPIRE2 | 0.0021 | G | 0.506263 | 8.21E-14 | -+ | 3.259E-07 | 0.5796 | Old |
| 1129 | 16 | 16-89831598-C-T | rs9931732 | SPIRE2 | 0.0018 | C | 0.743064 | 7.68E-14 | -+ | 6.226E-08 | 0.4315 | Old |
| 1129 | 16 | 16-89831610-G-A | rs5443011 | SPIRE2 | 0.002 | G | | 4.28E-10 | ?+ | 4.275E-10 | | Old |
| 1130 | 16 | 16-89879111-TTGTCCGTGTACACAGATGGGCTTCGGGGCCTGTCATACGTGCTGTTCGTGTACACAGACGGGCTCCTGGGCCTATCACCCGTGC-T | rs2042404 | TCF25 | 0.0044 | T | | 1.98E-09 | ?+ | 1.979E-09 | | Old |
| 1131 | 17 | 17-1014620-C-CCT | | ABR | 0.0009 | CCT | | 1.27E-09 | ?+ | 1.267E-09 | | New |

| | | | | | | | | | | | | |
|---|---|---|---|---|---|---|---|---|---|---|---|---|
| 1132 | 17 | 17-1195651-C-A | rs8913131 | ABR | 0.0008 | A | 0.116815 | 1.97E-09 | ++ | 5.876E-08 | 0.8741 | New |
| 1132 | 17 | 17-1195697-G-A | rs1409937 | ABR | 0.0004 | G | 0.809979 | 1.24E-08 | ++ | 0.0003591 | 0.6806 | New |
| 1133 | 17 | 17-1230579-G-A | rs1313799 | ABR | 0.0012 | A | 0.409325 | 2.9E-09 | ++ | 0.000001051 | 0.6005 | Old |
| 1133 | 17 | 17-1230592-C-A | rs5421390 | ABR | 0.0014 | A | 0.293081 | 1.01E-08 | ++ | 7.406E-07 | | Old |
| 1134 | 17 | 17-1354830-TTTG-T | | NA | 0.0018 | TTTG | | 1.56E-13 | ?+ | 1.557E-13 | | Old |
| 1135 | 17 | 17-1430883-T-C | rs4547394 | CRK | 0.025 | T | | 5.38E-17 | ?+ | 5.38E-17 | | Old |
| 1135 | 17 | 17-1430884-G-A | rs5338220 | CRK | 0.0036 | A | 0.164919 | 2.51E-13 | ++ | 1.244E-08 | 0.02192 | Old |
| 1136 | 17 | 17-1596998-C-CCT | rs1251919 | SLC43A2 | 0.0012 | CCT | 0.985869 | 3.07E-14 | -+ | 2.156E-13 | 0.2073 | Old |
| 1137 | 17 | 17-16371521-G-C | | CENPV, UBB | 0.0013 | C | 0.99688 | 4.82E-16 | -+ | 4.773E-10 | | New |
| 1138 | 17 | 17-16382000-G-A | rs1265896 | UBB | 0.0035 | A | 0.190867 | 4.17E-09 | ++ | 0.0001269 | 0.8646 | New |
| 1139 | 17 | 17-16382525-C-T | rs7518839 | UBB | 0.0053 | T | 0.583322 | 3.54E-11 | -+ | 0.006312 | 0.2644 | New |
| 1140 | 17 | 17-16451843-C-CAA | | NA | 0.0003 | CAA | | 3.69E-08 | ?+ | 3.694E-08 | | New |
| 1141 | 17 | 17-17001857-GCA-G | | NA | 0.0005 | GCA | | 1.87E-10 | ?+ | 1.865E-10 | | New |
| 1142 | 17 | 17-17002395-G-A | rs1476682 | LINC02090, AC104024.2 | 0.0008 | A | 0.017196 | 1.09E-17 | ++ | 1.674E-18 | | New |
| 1143 | 17 | 17-1758445-ACT-A | rs1306536 | SERPINF2, SERPINF1 | 0.0036 | ACT | 0.444551 | 6.12E-28 | -+ | 5.17E-23 | | Old |
| 1144 | 17 | 17-1788527-C-CCT | | NA | 0.0002 | CCT | | 4.13E-10 | ?+ | 4.131E-10 | | Old |
| 1145 | 17 | 17-18080932-T-A | rs1597705 | GID4, DRG2 | 0.0008 | T | 0.578681 | 7.15E-13 | ++ | 3.097E-10 | | Old |
| 1146 | 17 | 17-18084733-G-C | rs9415726 | GID4, DRG2 | 0.0009 | G | 0.217919 | 2.59E-12 | -+ | 4.837E-10 | | Old |
| 1147 | 17 | 17-18762161-T-A | | FBXW10 | 0.0005 | T | | 4.8E-09 | ?+ | 4.802E-09 | | New |
| 1148 | 17 | 17-19025179-TTTTTC-T | rs1293661 | GRAP | 0.0043 | T | 0.952708 | 4.21E-15 | ++ | 1.477E-13 | | New |
| 1149 | 17 | 17-21260860-AC-A | rs1372704 | NATD1, MAP2K3 | 0.0019 | AC | | 1.14E-15 | ?+ | 1.143E-15 | | New |
| 1150 | 17 | 17-21684971-C-CAGG | | NA | 0.0057 | CAGG | | 6.51E-09 | ?+ | 6.508E-09 | 0.6378 | New |
| 1151 | 17 | 17-22064831-C-CAAAAAAAAAAAAA | | NA | 0.028 | CAAAAAAAAAAAAA | | 1.6E-09 | ?+ | 1.603E-09 | | New |
| 1152 | 17 | 17-22204256-G-C | rs1363962 | KCNJ18, LINC02002 | 0.0005 | C | | 4.05E-10 | ?+ | 4.047E-10 | | New |

| | | | | | | | | | | | | |
|---|---|---|---|---|---|---|---|---|---|---|---|---|
| 1153 | 17 | 17-2584417-GGC-G | rs1241129 | RN7SL33P, PAFAH1B1 | 0.0025 | G | | 3.25E-11 | ?+ | 3.25E-11 | 0.4068 | New |
| 1154 | 17 | 17-26682500-T-A | rs1223720 | NONE, AC069061.2 | 0.0031 | T | 0.453771 | 3.54E-21 | -+ | 1.71E-20 | 0.02679 | New |
| 1155 | 17 | 17-26682529-G-C | rs1906375 | NONE, AC069061.2 | 0.0022 | C | 0.545566 | 8.79E-29 | ++ | 7.414E-29 | | New |
| 1156 | 17 | 17-26989948-AGC-A | | NA | 0.0048 | A | | 2.48E-12 | ?+ | 2.482E-12 | 0.6414 | New |
| 1157 | 17 | 17-2823905-C-CCT | | RAP1GAP2 | 0.0014 | CCT | | 1.7E-09 | ?+ | 1.697E-09 | | New |
| 1158 | 17 | 17-28266171-G-C | rs9940657 | AC061975.1 | 0.0008 | C | 0.652248 | 1.84E-08 | -+ | 0.00002202 | | New |
| 1159 | 17 | 17-2846555-C-CAT | | NA | 0.0004 | C | | 7.34E-09 | ?+ | 7.336E-09 | | New |
| 1160 | 17 | 17-2846555-C-CATGA | | NA | 0.0017 | C | | 1.41E-19 | ?+ | 1.408E-19 | | New |
| 1160 | 17 | 17-2846560-GC-G | | NA | 0.0007 | GC | | 1.75E-13 | ?+ | 1.747E-13 | | New |
| 1160 | 17 | 17-2846563-G-A | rs1031320 | RAP1GAP2 | 0.0008 | G | 0.18275 | 3.66E-11 | ++ | 1.481E-11 | | New |
| 1160 | 17 | 17-2846566-ACGT-A | | NA | 0.0008 | ACGT | | 2.37E-09 | ?+ | 2.368E-09 | | New |
| 1161 | 17 | 17-28709778-C-CTT | rs1441508 | PROCA1 | 0.0024 | C | | 1.37E-10 | ?+ | 1.372E-10 | 0.1446 | New |
| 1162 | 17 | 17-28771881-C-T | rs9022149 | FAM222B | 0.0011 | C | 0.559424 | 1.61E-10 | -+ | 3.919E-09 | 0.09652 | New |
| 1163 | 17 | 17-29356267-G-A | rs5748471 | AC068025.2 | 0.001 | G | 0.197652 | 2.5E-08 | ++ | 0.00001422 | 0.6256 | New |
| 1163 | 17 | 17-29356278-C-T | rs2069525 | AC068025.2 | 0.0008 | C | 0.724955 | 2.3E-13 | -+ | 0.000005511 | | New |
| 1163 | 17 | 17-29356294-C-T | rs9836846 | AC068025.2 | 0.0009 | C | 0.86587 | 1.42E-11 | -+ | 0.0001219 | | New |
| 1164 | 17 | 17-29359896-ACT-A | | NA | 0.0005 | ACT | 0.340699 | 2.54E-08 | ++ | 5.875E-07 | | New |
| 1165 | 17 | 17-29813599-C-CTT | | NA | 0.0033 | C | | 7.25E-17 | ?+ | 7.247E-17 | | New |
| 1166 | 17 | 17-29852694-GGCGC-G | | NA | 0.0009 | G | | 6.34E-12 | ?+ | 6.342E-12 | | New |
| 1166 | 17 | 17-29852702-C-CA | | NA | 0.0006 | CA | | 4.62E-09 | ?+ | 4.62E-09 | | New |
| 1167 | 17 | 17-30815346-T-A | rs1445598 | CRLF3 | 0.0013 | T | 0.226334 | 9.74E-11 | -+ | 0.0009938 | 0.4008 | New |

| | | | | | | | | | | | |
|---|---|---|---|---|---|---|---|---|---|---|---|
| 1168 | 17 | 17-3112620-GGGT-G | rs1334630 | OR1D2, OR1G1 | 0.0027 | GGGT | | 6.92E-10 | ?+ | 6.915E-10 | | New |
| 1169 | 17 | 17-31929615-C-CCT | rs1282750 | UTP6, SUZ12 | 0.0071 | CCT | | 4.65E-10 | ?+ | 4.646E-10 | 0.2965 | New |
| 1170 | 17 | 17-35682510-TG-T | | NA | 0.0022 | T | | 2.67E-10 | ?+ | 2.672E-10 | 0.9754 | New |
| 1171 | 17 | 17-35882107-G-A | rs1207364 | AC015849.1 | 0.0017 | A | 0.734981 | 4.41E-10 | ++ | 0.0000318 | 0.7341 | New |
| 1171 | 17 | 17-35882115-G-A | rs1197081 | AC015849.1 | 0.0016 | A | 0.833819 | 3.01E-12 | ++ | 0.00001871 | | New |
| 1171 | 17 | 17-35882151-G-C | rs1268418 | AC015849.1 | 0.0012 | G | 0.264791 | 8.35E-11 | ++ | 0.000006836 | | New |
| 1172 | 17 | 17-3765193-C-CA | | NA | 0.0008 | C | | 7.54E-11 | ?+ | 7.542E-11 | | New |
| 1173 | 17 | 17-38809283-C-T | rs6142225 | CWC25 | 0.1306 | C | | 7.75E-09 | ?+ | 7.751E-09 | 0.111 | New |
| 1174 | 17 | 17-38862181-C-T | rs1269322 | RPL23, LASP1 | 0.0008 | T | | 1.56E-14 | ?+ | 1.562E-14 | 0.04591 | New |
| 1175 | 17 | 17-38955163-G-A | rs1905486 | FBXO47 | 0.0004 | G | | 1.6E-09 | ?+ | 1.599E-09 | 0.2919 | New |
| 1176 | 17 | 17-39154845-C-CA | rs1331156 | PLXDC1 | 0.001 | CA | | 7.37E-11 | ?+ | 7.368E-11 | | New |
| 1177 | 17 | 17-397831-G-T | rs1257925 | RPH3AL, AC141424.1 | 0.0019 | T | 0.828572 | 2.41E-18 | -+ | 1.155E-15 | | New |
| 1178 | 17 | 17-39956807-C-CAG | | NA | 0.0005 | CAG | | 5.6E-09 | ?+ | 5.601E-09 | | New |
| 1179 | 17 | 17-40013510-C-CCT | | NA | 0.0016 | C | | 8.51E-17 | ?+ | 8.507E-17 | | New |
| 1180 | 17 | 17-40044663-C-CTT | | NA | 0.0003 | CTT | | 9.01E-09 | ?+ | 9.015E-09 | | New |
| 1181 | 17 | 17-40544234-G-A | rs8066507 | AC004585.1, CCR7 | 0.1379 | A | | 9.67E-11 | ?+ | 9.672E-11 | | New |
| 1182 | 17 | 17-41722828-GTCCCCAA-G | | NA | 0.0025 | GTCCCCAA | | 5.59E-10 | ?+ | 5.594E-10 | 0.359 | New |
| 1183 | 17 | 17-41780474-ACT-A | rs1555609 | JUP | 0.001 | A | | 1.05E-08 | ?+ | 1.048E-08 | 0.2883 | New |
| 1184 | 17 | 17-41843928-AC-A | rs1555620 | KLHL10 | 0.001 | A | | 7.47E-10 | ?+ | 7.473E-10 | 0.3313 | New |
| 1185 | 17 | 17-41857400-G-C | rs1597948 | KLHL11 | 0.0014 | G | 0.912437 | 1.44E-08 | ++ | 0.003249 | | New |
| 1186 | 17 | 17-42081656-G-A | rs2053750 | ZNF385C | 0.0005 | G | 0.410191 | 7.92E-09 | ++ | 1.196E-08 | | New |
| 1187 | 17 | 17-42101152-AG-A | rs1391504 | DHX58 | 0.0006 | A | | 4.15E-12 | ?+ | 4.153E-12 | | New |
| 1188 | 17 | 17-42606002-ATTAATT-A | | NA | 0.002 | ATTAATT | | 1.63E-10 | ?+ | 1.626E-10 | | New |
| 1189 | 17 | 17-42794089-C-CAG | | NA | 0.0005 | CAG | | 2.16E-11 | ?+ | 2.155E-11 | | New |

| | | | | | | | | | | | | |
|---|---|---|---|---|---|---|---|---|---|---|---|---|
| 1190 | 17 | 17-43470088-G-A | rs5397789 | MIR2117HG, RNU6-971P | 0.0006 | A | 0.759479 | 8.36E-09 | -+ | 7.865E-07 | | New |
| 1191 | 17 | 17-43567458-T-A | rs5305550 | DHX8, ETV4 | 0.0016 | T | | 9.49E-11 | ?+ | 9.488E-11 | | New |
| 1192 | 17 | 17-44139757-G-A | rs1442217 | RNU6-131P, C17orf53 | 0.0013 | A | | 1.41E-08 | ?+ | 1.408E-08 | 0.2571 | Old |
| 1193 | 17 | 17-44189985-TG-T | rs1216572 | TMUB2 | 0.0016 | T | 0.203881 | 1.09E-08 | ++ | 3.618E-08 | 0.03941 | Old |
| 1194 | 17 | 17-44372763-GAA-G | | NA | 0.0012 | GAA | | 2.65E-10 | ?+ | 2.65E-10 | | Old |
| 1194 | 17 | 17-44372764-GTT-G | | NA | 0.001 | G | | 1.19E-08 | ?+ | 1.185E-08 | | Old |
| 1195 | 17 | 17-44513203-C-CCGA | | NA | 0.0008 | CCGA | | 3.95E-10 | ?+ | 3.951E-10 | | Old |
| 1196 | 17 | 17-45057514-G-C | rs1244239 | DCAKD, NMT1 | 0.0009 | C | | 6.9E-12 | ?+ | 6.905E-12 | 0.3093 | New |
| 1197 | 17 | 17-45079271-ATT-A | | NA | 0.0008 | ATT | | 3.23E-12 | ?+ | 3.233E-12 | | New |
| 1198 | 17 | 17-47255122-GCA-G | rs1239388 | AC068234.1, ITGB3 | 0.0024 | GCA | | 4.5E-13 | ?+ | 4.495E-13 | 0.9792 | Old |
| 1198 | 17 | 17-47255125-C-CCT | rs1355670 | AC068234.1, ITGB3 | 0.0019 | C | | 3.05E-12 | ?+ | 3.047E-12 | 0.9347 | Old |
| 1199 | 17 | 17-47530102-ATTTTTTT-A | | NPEPPS | 0.0081 | A | | 7.66E-11 | ?+ | 7.662E-11 | | New |
| 1200 | 17 | 17-48098623-G-A | | CBX1 | 0.0006 | G | | 9.66E-10 | ?+ | 9.663E-10 | | New |
| 1201 | 17 | 17-4820089-C-CAT | rs1331439 | PLD2 | 0.0038 | C | | 1.33E-17 | ?+ | 1.333E-17 | 0.0344 | Old |
| 1202 | 17 | 17-48866581-C-CCT | | NA | 0.0013 | CCT | | 1.57E-10 | ?+ | 1.573E-10 | | Old |
| 1203 | 17 | 17-49143921-GTGAA-G | | NA | 0.0001 | G | | 3.6E-09 | ?+ | 3.602E-09 | | Old |
| 1204 | 17 | 17-4923566-C-A | rs1970262 | CHRNE | 0.0008 | C | | 6.86E-13 | ?+ | 6.861E-13 | | Old |
| 1205 | 17 | 17-49345263-C-T | rs9067417 | ZNF652 | 0.001 | T | 0.723394 | 1.83E-12 | ++ | 1.482E-08 | | Old |
| 1206 | 17 | 17-49641837-C-A | rs1460518 | SPOP | 0.0005 | C | 0.280772 | 3.54E-08 | -+ | 1.951E-07 | 0.9527 | Old |
| 1207 | 17 | 17-49678884-C-CCATCCTGGCTAA | | NA | 0.0005 | CCATCCTGGCTAA | | 3.12E-08 | ?+ | 3.122E-08 | | Old |
| 1208 | 17 | 17-4993046-ATT-A | | NA | 0.0008 | ATT | | 7.15E-13 | ?+ | 7.151E-13 | | Old |
| 1209 | 17 | 17-4995776-C-CA | | NA | 1E-04 | C | | 3.53E-13 | ?+ | 3.526E-13 | | Old |
| 1210 | 17 | 17-50126919-GT-G | | NA | 0.0018 | G | | 3.69E-08 | ?+ | 3.69E-08 | | New |
| 1211 | 17 | 17-5013837-C-A | rs1378994 | KIF1C | 0.0025 | A | 0.983411 | 1.01E-16 | -+ | 0.00000023 | | Old |

| | | | | | | | | | | | | |
|---|---|---|---|---|---|---|---|---|---|---|---|---|
| 1212 | 17 | 17-50156079-ACG-A | rs1295331 | PPP1R9B, AC015909.2 | 0.0023 | ACG | 0.271093 | 4.02E-11 | -+ | 6.95E-09 | 0.1093 | New |
| 1213 | 17 | 17-50759908-GAC-G | rs1474019 | LUC7L3, ANKRD40CL | 0.0067 | GAC | | 1.56E-15 | ?+ | 1.564E-15 | | New |
| 1214 | 17 | 17-5181517-C-T | rs7702105 | ZNF594 | 0.0026 | C | 0.05207 | 2.15E-09 | -+ | 0.9104 | | Old |
| 1215 | 17 | 17-5265867-C-CCT | rs1484578 | AC087500.1, RABEP1 | 0.0033 | CCT | | 2.96E-10 | ?+ | 2.955E-10 | | Old |
| 1216 | 17 | 17-5439856-C-CCTT | | NA | 0.0006 | CCTT | | 2.1E-10 | ?+ | 2.098E-10 | | Old |
| 1217 | 17 | 17-58478215-C-CCT | | HSF5 | 0.0028 | CCT | | 4.76E-09 | ?+ | 4.757E-09 | | Old |
| 1218 | 17 | 17-58685271-TA-T | | AC011195.2, TEX14 | 0.0055 | TA | 0.841849 | 1.08E-17 | -+ | 4.214E-07 | | Old |
| 1219 | 17 | 17-59120928-G-A | rs1393003 | SKA2 | 0.0007 | G | 0.643843 | 1.64E-08 | -+ | 9.858E-08 | 0.1008 | New |
| 1220 | 17 | 17-59965446-C-CT | | NA | 0.0012 | CT | | 2.78E-10 | ?+ | 2.783E-10 | | New |
| 1221 | 17 | 17-60340884-G-C | rs2009305 | USP32 | 0.0039 | G | 0.574971 | 2.77E-19 | -+ | 5.357E-07 | | New |
| 1222 | 17 | 17-60341469-ATTCTT-A | rs1214054 | USP32 | 0.0019 | ATTCTT | | 2.83E-10 | ?+ | 2.83E-10 | 0.1819 | New |
| 1223 | 17 | 17-60589164-GGGT-G | rs1404224 | LINC01999, PPM1D | 0.0026 | GGGT | | 6.02E-11 | ?+ | 6.022E-11 | 0.4338 | New |
| 1224 | 17 | 17-60703056-C-CCT | | NA | 0.0013 | C | | 1.38E-08 | ?+ | 1.378E-08 | | New |
| 1225 | 17 | 17-61890402-G-C | rs1429724 | INTS2 | 0.0008 | G | | 8.85E-09 | ?+ | 8.85E-09 | 0.5404 | New |
| 1226 | 17 | 17-62007671-AC-A | rs1345200 | MED13 | 0.0053 | AC | 0.649111 | 3.69E-22 | -+ | 3.143E-13 | 0.5902 | New |
| 1227 | 17 | 17-62157437-C-A | rs1427302 | RF00019, EFCAB3 | 0.0009 | A | 0.668676 | 3.13E-18 | ++ | 3.212E-16 | | New |
| 1227 | 17 | 17-62157438-G-A | rs1414985 | RF00019, EFCAB3 | 0.0007 | G | 0.800837 | 3.28E-14 | -+ | 1.907E-12 | | New |
| 1228 | 17 | 17-62235247-G-A | rs5851362 | RF00019, EFCAB3 | 0.0031 | A | 0.42173 | 5.33E-09 | -+ | 9.457E-07 | 0.6979 | New |
| 1229 | 17 | 17-62235962-C-A | rs5360392 | RF00019, EFCAB3 | 0.002 | C | | 3.16E-13 | ?+ | 3.161E-13 | 0.5522 | New |
| 1229 | 17 | 17-62235981-C-T | rs1347265 | RF00019, EFCAB3 | 0.0036 | C | | 5.91E-21 | ?+ | 5.91E-21 | 0.4521 | New |
| 1230 | 17 | 17-62249528-ATCAT-A | | NA | 0.0008 | ATCAT | | 2.92E-08 | ?+ | 2.922E-08 | | New |
| 1231 | 17 | 17-62388645-G-C | | EFCAB3 | 0.0003 | G | | 1.72E-08 | ?+ | 1.718E-08 | | New |
| 1232 | 17 | 17-62411588-G-A | rs167491 | EFCAB3 | 0.0003 | G | 0.070427 | 9.73E-10 | -+ | 0.0001685 | | New |

| | | | | | | | | | | | | |
|---|---|---|---|---|---|---|---|---|---|---|---|---|
| 1233 | 17 | 17-62466398-G-A | rs2070881 | TLK2 | 0.0004 | G | 0.481966 | 4.39E-08 | ++ | 0.000003417 | | Old |
| 1234 | 17 | 17-62498597-G-C | rs9204633 | TLK2 | 0.0006 | G | | 4.65E-08 | ?+ | 4.651E-08 | | Old |
| 1235 | 17 | 17-62604736-C-CCGTG | | TLK2 | 0.0018 | C | | 5.16E-09 | ?+ | 5.162E-09 | 0.4103 | Old |
| 1236 | 17 | 17-63851546-G-C | rs1904751 | SMARCD2, CSH2 | 0.0014 | C | | 3.09E-12 | ?+ | 3.091E-12 | 0.2626 | Old |
| 1237 | 17 | 17-64347887-G-T | rs2035619 | PECAM1 | 0.0018 | T | 0.074886 | 2.86E-21 | ++ | 3.35E-17 | | New |
| 1237 | 17 | 17-64347888-G-A | rs2035619 | PECAM1 | 0.0013 | G | 0.590512 | 9.28E-14 | ++ | 2.008E-09 | | New |
| 1237 | 17 | 17-64347889-G-C | rs2035619 | PECAM1 | 0.0015 | C | 0.781658 | 5.52E-15 | ++ | 2.375E-10 | | New |
| 1238 | 17 | 17-67064485-G-A | | CACNG1, HELZ | 0.0003 | G | | 6.63E-09 | ?+ | 6.628E-09 | | New |
| 1239 | 17 | 17-67634352-G-A | rs1389719 | PITPNC1 | 0.0004 | A | | 1.79E-08 | ?+ | 1.792E-08 | 0.3027 | New |
| 1240 | 17 | 17-67956115-G-A | rs1280964 | AC134407.2 | 0.0008 | A | 0.875483 | 1.52E-09 | -+ | 0.00007338 | | New |
| 1240 | 17 | 17-67956117-G-A | rs1598902 | AC134407.2 | 0.0008 | A | 0.928741 | 1.52E-08 | -+ | 0.0002419 | | New |
| 1241 | 17 | 17-68002426-AT-A | rs1480643 | C17orf58, KPNA2 | 0.0009 | AT | | 3.78E-15 | ?+ | 3.782E-15 | 0.5742 | New |
| 1242 | 17 | 17-68627016-T-A | rs1179509 | LINC01482 | 0.001 | T | | 1.35E-10 | ?+ | 1.351E-10 | | New |
| 1243 | 17 | 17-69099080-C-CTTT | rs1200022 | ABCA6 | 0.0007 | CTTT | | 5.43E-09 | ?+ | 5.429E-09 | 0.6594 | New |
| 1244 | 17 | 17-69514705-GCA-G | | NA | 0.0003 | GCA | | 4.89E-10 | ?+ | 4.886E-10 | | New |
| 1245 | 17 | 17-7055430-C-CCT | | NA | 0.0016 | CCT | | 3.81E-19 | ?+ | 3.813E-19 | | New |
| 1246 | 17 | 17-7268492-G-T | rs1404018 | RF00019, SLC2A4 | 0.0013 | G | 0.661078 | 2.21E-13 | ++ | 1.016E-09 | | New |
| 1247 | 17 | 17-73051724-GGT-G | rs1503266 | SLC39A11 | 0.0015 | GGT | | 1.1E-09 | ?+ | 1.096E-09 | | New |
| 1248 | 17 | 17-74976489-C-CCT | | NA | 0.0008 | CCT | | 1.59E-08 | ?+ | 1.589E-08 | | New |
| 1249 | 17 | 17-75206797-AAG-A | rs1489928 | NUP85 | 0.0041 | AAG | | 3.96E-22 | ?+ | 3.959E-22 | 0.1882 | New |
| 1249 | 17 | 17-75206799-ACC-A | rs1201639 | NUP85 | 0.0038 | A | | 2.44E-25 | ?+ | 2.435E-25 | | New |
| 1250 | 17 | 17-75529672-C-CTAGCA | | NA | 0.0013 | CTAGCA | | 2.24E-09 | ?+ | 2.244E-09 | | New |
| 1251 | 17 | 17-75711868-C-CTG | | NA | 0.0001 | CTG | | 5.83E-09 | ?+ | 5.829E-09 | | New |
| 1252 | 17 | 17-75801049-GCA-G | | NA | 0.0022 | GCA | | 1.46E-19 | ?+ | 1.463E-19 | | New |

| | | | | | | | | | | | | |
|---|---|---|---|---|---|---|---|---|---|---|---|---|
| 1252 | 17 | 17-75801051-C-CAG | | NA | 0.0018 | C | | 2.54E-16 | ?+ | 2.544E-16 | | New |
| 1252 | 17 | 17-75801061-G-A | rs1157209 | UNK | 0.001 | A | 0.641255 | 1.96E-10 | -+ | 0.00004563 | 0.9337 | New |
| 1252 | 17 | 17-75801070-ATTT-A | rs1416175 | NA | 0.0035 | A | 0.839552 | 3.08E-30 | -+ | 3.902E-23 | | New |
| 1253 | 17 | 17-75968218-C-CCT | rs2065957 | ACOX1 | 0.0066 | CCT | | 4.04E-08 | ?+ | 4.04E-08 | | New |
| 1254 | 17 | 17-75993291-AT-A | | NA | 0.001 | A | | 1.04E-11 | ?+ | 1.042E-11 | | New |
| 1255 | 17 | 17-75993927-C-CGGA | | TEN1, TEN1-CDK3 | 0.001 | C | | 8.22E-11 | ?+ | 8.223E-11 | | New |
| 1256 | 17 | 17-76098643-C-CCT | | NA | 0.0016 | CCT | | 2.15E-11 | ?+ | 2.146E-11 | | New |
| 1257 | 17 | 17-7681502-AT-A | rs1475747 | TP53 | 0.0036 | AT | 0.905228 | 1.43E-19 | ++ | 2.63E-15 | 0.9197 | New |
| 1258 | 17 | 17-7681961-TTTTTTG-T | rs1180886 | TP53 | 0.0029 | TTTTTTG | | 5.25E-09 | ?+ | 5.246E-09 | 0.2077 | New |
| 1259 | 17 | 17-783556-G-C | | MRM3 | 0.0005 | G | | 3.79E-15 | ?+ | 3.79E-15 | | New |
| 1260 | 17 | 17-78864868-AC-A | rs1265889 | TIMP2 | 0.001 | AC | | 3.27E-08 | ?+ | 3.27E-08 | | New |
| 1261 | 17 | 17-80875759-G-T | rs1175028 | RPTOR | 0.0027 | T | 0.785684 | 6.55E-09 | -+ | 0.005042 | | New |
| 1262 | 17 | 17-8131304-G-T | rs3732616 | HES7, AC129492.1 | 0.0024 | T | 0.527806 | 1.31E-11 | ++ | 9.994E-07 | 0.3536 | New |
| 1263 | 17 | 17-81357652-C-T | rs1052674 | AC027601.6, AC110285.3 | 0.0005 | T | 0.305022 | 7.89E-10 | -+ | 0.000003554 | | New |
| 1264 | 17 | 17-81848677-G-A | rs1821448 | P4HB | 0.0007 | A | 0.605935 | 2.48E-08 | ++ | 0.000001156 | 0.7694 | New |
| 1265 | 17 | 17-81849383-G-C | | P4HB | 0.0008 | G | 0.302558 | 3.98E-13 | ++ | 2.074E-10 | | New |
| 1266 | 17 | 17-82725483-GCT-G | | NA | 0.0014 | GCT | | 8.45E-12 | ?+ | 8.447E-12 | | New |
| 1267 | 17 | 17-82827811-GC-G | rs1404767 | TBCD | 0.0039 | G | 0.904053 | 9.74E-09 | ++ | 0.00001588 | | New |
| 1268 | 17 | 17-839242-G-C | | NXN | 0.0003 | C | 0.382387 | 2.42E-09 | -+ | 1.154E-08 | | New |
| 1269 | 17 | 17-8853714-G-A | rs6025669 | PIK3R6 | 0.0791 | A | | 1.03E-08 | ?+ | 1.033E-08 | 0.8296 | New |
| 1270 | 17 | 17-9113990-C-CT | | NA | 0.0005 | CT | | 2.47E-08 | ?+ | 2.467E-08 | | New |
| 1271 | 17 | 17-971686-GGTGA-G | | NA | 0.001 | G | | 5.4E-14 | ?+ | 5.397E-14 | | New |
| 1272 | 18 | 18-12866210-AGGTG-A | | PTPN2 | 0.0019 | A | | 5.67E-21 | ?+ | 5.669E-21 | | New |
| 1273 | 18 | 18-14501562-C-T | rs1373487 | AP006261.1, POTEC | 0.0005 | C | 0.513893 | 7.11E-09 | ++ | 1.002E-07 | | New |

| | | | | | | | | | | | | |
|---|---|---|---|---|---|---|---|---|---|---|---|---|
| 1274 | 18 | 18-14502080-G-C | rs1909747 | AP006261.1, POTEC | 0.0005 | G | | 2.78E-09 | ?+ | 2.778E-09 | | New |
| 1275 | 18 | 18-21616146-C-T | rs7585188 | SNRPD1 | 0.0004 | T | 0.377088 | 2.45E-09 | ++ | 1.141E-08 | 0.6019 | New |
| 1276 | 18 | 18-21731482-C-A | rs8900056 | RF00019, MIB1 | 0.0034 | A | | 8.83E-24 | ?+ | 8.832E-24 | | New |
| 1277 | 18 | 18-218980-C-CA | rs1133339 | THOC1 | 0.0086 | C | 0.196996 | 2.55E-08 | ++ | 4.711E-08 | | New |
| 1278 | 18 | 18-21974281-G-C | rs1568254 | AC103987.2, LINC01900 | 0.0016 | C | 0.670013 | 5.28E-16 | ++ | 1.362E-10 | 0.7329 | New |
| 1279 | 18 | 18-23093206-AT-A | | NA | 0.001 | A | | 5E-11 | ?+ | 4.996E-11 | | New |
| 1279 | 18 | 18-23093208-C-CA | | NA | 0.0009 | C | | 1.23E-08 | ?+ | 1.234E-08 | | New |
| 1280 | 18 | 18-2996644-AT-A | | NA | 0.0008 | A | 0.195103 | 8.4E-10 | -+ | 0.0004439 | 0.4422 | New |
| 1281 | 18 | 18-33147978-C-CT | rs1220304 | CCDC178 | 0.0064 | CT | 0.343818 | 1.37E-43 | -+ | 1.123E-42 | | New |
| 1282 | 18 | 18-3550114-G-A | | DLGAP1 | 0.0005 | G | | 4.9E-09 | ?+ | 4.902E-09 | | New |
| 1283 | 18 | 18-47400508-TCCACAGTCATCTTCCCACCCGAGGCCACCACACTGTGCCTTC-T | rs1416466 | MIR4527HG | 0.0103 | T | | 1.23E-08 | ?+ | 1.231E-08 | | New |
| 1284 | 18 | 18-54138189-AT-A | | NA | 0.0018 | AT | | 5.35E-09 | ?+ | 5.354E-09 | | New |
| 1285 | 18 | 18-54207819-C-CCT | | NA | 0.002 | CCT | | 4.12E-08 | ?+ | 4.116E-08 | | New |
| 1286 | 18 | 18-54537923-C-CTACTGAACCCACAACCACAGCAGCCAGGACTGTAACTGAACAGTCTGCCATCACAACTGTAGGCATTTACACTGAGACTACAACCCCATCAGCCACCACTACCACT | | NA | 0.0005 | CTACTGAACCCACAAC | | 3.39E-08 | ?+ | 3.389E-08 | | New |
| 1287 | 18 | 18-54758841-G-C | rs1297030 | RAB27B | 0.0005 | G | 0.983058 | 1.8E-08 | -+ | 0.00009929 | | New |
| 1288 | 18 | 18-56903567-G-C | rs1296656 | WDR7 | 0.0011 | C | 0.659043 | 1.04E-16 | ++ | 3.316E-15 | | New |
| 1289 | 18 | 18-58129746-C-CGGTGTTGGGCTCTGTTGGGGTTTGGTTGTGAGCGGAACTGT | | NA | 0.0031 | C | | 3.11E-08 | ?+ | 3.11E-08 | | Old |
| 1290 | 18 | 18-61699837-C-T | rs1408860 | AC105094.2 | 0.0198 | T | | 1.07E-10 | ?- | 1.07E-10 | 0.7636 | New |

| | | | | | | | | | | | | |
|---|---|---|---|---|---|---|---|---|---|---|---|---|
| 1291 | 18 | 18-62195984-C-CTG | | RELCH | 0.0014 | CTG | | 3.05E-08 | ?+ | 3.052E-08 | | New |
| 1291 | 18 | 18-62195985-C-CAA | | RELCH | 0.002 | C | | 1.14E-13 | ?+ | 1.14E-13 | | New |
| 1292 | 18 | 18-67758970-G-A | rs1283088 | AC022655.1, AC114689.3 | 0.0012 | A | 0.960147 | 8.71E-13 | ++ | 0.000001678 | | New |
| 1293 | 18 | 18-7107368-TTGG-T | | NA | 0.0004 | TTGG | | 4.49E-08 | ?+ | 4.486E-08 | | New |
| 1294 | 18 | 18-7274664-AT-A | | NA | 0.0011 | AT | | 3.18E-08 | ?+ | 3.181E-08 | | New |
| 1295 | 18 | 18-76571552-ACC-A | rs1205462 | LINC00683 | 0.0004 | ACC | 0.33332 | 1.1E-08 | ++ | 7.846E-08 | | New |
| 1296 | 18 | 18-76817728-GCCT-G | | NA | 0 | GCCT | | 1.1E-08 | ?+ | 1.095E-08 | | New |
| 1297 | 18 | 18-78591455-C-CCT | | NA | 0.001 | CCT | | 4.15E-09 | ?+ | 4.146E-09 | 0.4528 | New |
| 1298 | 18 | 18-80185678-G-A | rs1340254 | AC139100.1 | 0.0008 | G | | 1.17E-10 | ?+ | 1.168E-10 | | New |
| 1299 | 18 | 18-9037196-GGT-G | | NA | 0.0004 | G | | 2.68E-08 | ?+ | 2.676E-08 | 0.8587 | New |
| 1300 | 18 | 18-9777902-C-A | | RAB31 | 0.0006 | A | | 3.5E-10 | ?+ | 3.498E-10 | | New |
| 1301 | 19 | 19-10263242-GCGCC-G | rs1377788 | AC011511.2 | 0.0035 | G | | 4.88E-08 | ?+ | 4.876E-08 | | New |
| 1302 | 19 | 19-11071676-GC-G | rs1404913 | SMARCA4 | 0.0068 | GC | | 1.75E-35 | ?+ | 1.753E-35 | 0.2132 | New |
| 1303 | 19 | 19-11473295-G-A | rs664477 | AC008481.3, ELAVL3 | 0.3062 | G | | 3.87E-10 | ?+ | 3.871E-10 | | New |
| 1304 | 19 | 19-11582992-G-T | rs6511738 | ZNF627 | 0.0006 | G | | 1.61E-09 | ?+ | 1.605E-09 | | New |
| 1305 | 19 | 19-11648684-AAC-A | | NA | 0.0005 | A | | 9.04E-10 | ?+ | 9.036E-10 | | New |
| 1306 | 19 | 19-11668061-G-A | rs5710266 | AC008543.1 | 0.0037 | A | 0.424244 | 2.32E-13 | ++ | 8.532E-08 | 0.6521 | New |
| 1307 | 19 | 19-12129144-TA-T | | NA | 0.0005 | T | | 5.85E-09 | ?+ | 5.849E-09 | | New |
| 1308 | 19 | 19-12286479-C-CAA | | NA | 0.0009 | CAA | | 8.62E-09 | ?+ | 8.616E-09 | | New |
| 1309 | 19 | 19-13180657-C-A | rs9980128 | IER2, CACNA1A | 0.0008 | C | 0.839454 | 1.78E-10 | -+ | 5.314E-07 | 0.3408 | New |
| 1310 | 19 | 19-13661533-C-CAT | rs1214400 | CACNA1A, CCDC130 | 0.0013 | C | 0.363768 | 1.02E-09 | -+ | 1.079E-07 | 0.4571 | New |
| 1311 | 19 | 19-13736813-C-CCT | rs1481218 | CCDC130 | 0.0012 | C | | 9.21E-11 | ?+ | 9.211E-11 | | New |

| | | | | | | | | | | | | |
|---|---|---|---|---|---|---|---|---|---|---|---|---|
| 1312 | 19 | 19-13745457-G-A | | CCDC130 | 0.0006 | G | | 7.49E-09 | ?+ | 7.485E-09 | | New |
| 1313 | 19 | 19-13893919-AT-A | | NA | 0.0017 | AT | | 6.48E-19 | ?+ | 6.478E-19 | | New |
| 1313 | 19 | 19-13893922-AT-A | | NA | 0.0011 | AT | | 3.67E-11 | ?+ | 3.672E-11 | | New |
| 1314 | 19 | 19-13946219-C-A | rs1450781 | PODNL1 | 0.0018 | C | 0.29139 | 3.5E-14 | -+ | 1.836E-07 | | New |
| 1315 | 19 | 19-14258228-C-CATA | | AC011509.2 | 0.0018 | CATA | | 1.2E-10 | ?+ | 1.198E-10 | 0.6494 | New |
| 1316 | 19 | 19-1430887-ATTT-A | rs1358250 | DAZAP1 | 0.0007 | A | | 2.5E-10 | ?+ | 2.495E-10 | | Old |
| 1317 | 19 | 19-14648966-TCCC-T | rs1352071 | ADGRE3 | 0.001 | T | | 1.32E-11 | ?+ | 1.318E-11 | | New |
| 1318 | 19 | 19-14707697-GCCCA-G | | NA | 0.0006 | G | | 1.16E-08 | ?+ | 1.163E-08 | | New |
| 1319 | 19 | 19-15822925-GGGCT-G | | NA | 0.0018 | GGGCT | | 2.58E-26 | ?+ | 2.578E-26 | | New |
| 1319 | 19 | 19-15822927-C-CTTAT | | NA | 0.0009 | C | | 3E-12 | ?+ | 2.999E-12 | | New |
| 1320 | 19 | 19-16557360-AT-A | rs1255709 | AC008764.1, SLC35E1 | 0.0009 | A | 0.354923 | 1.27E-10 | ++ | 4E-10 | | New |
| 1321 | 19 | 19-1666683-GC-G | rs1374443 | TCF3, RF00017 | 0.0051 | GC | | 1.08E-13 | ?+ | 1.081E-13 | 0.3773 | Old |
| 1322 | 19 | 19-17354058-GGT-G | | NA | 0.0003 | G | | 2.97E-08 | ?+ | 2.969E-08 | | New |
| 1323 | 19 | 19-18528048-C-CT | | NA | 0.0007 | CT | | 2.81E-10 | ?+ | 2.805E-10 | | New |
| 1324 | 19 | 19-18619797-C-CCA | rs1253147 | AC003112.1 | 0.0014 | C | | 3.01E-09 | ?+ | 3.008E-09 | 0.3021 | New |
| 1325 | 19 | 19-18689093-GCC-G | rs1209568 | CRTC1 | 0.0021 | GCC | | 1.83E-08 | ?+ | 1.828E-08 | 0.3745 | New |
| 1326 | 19 | 19-1893632-C-T | rs5602685 | ABHD17A, SCAMP4 | 0.0022 | C | 0.337528 | 5.2E-12 | ++ | 3.94E-09 | 0.6408 | Old |
| 1327 | 19 | 19-1899530-TC-T | | NA | 0.0015 | TC | | 3.79E-11 | ?+ | 3.787E-11 | | Old |
| 1328 | 19 | 19-1927018-G-C | rs6728003 | SCAMP4, CSNK1G2 | 0.248 | G | 0.442573 | 4.45E-08 | -+ | 0.009245 | 0.6442 | Old |
| 1329 | 19 | 19-19363796-GTT-G | | MAU2, GATAD2A | 0.0017 | GTT | | 4.86E-13 | ?+ | 4.861E-13 | | New |
| 1330 | 19 | 19-19382660-C-T | rs1403905 | MAU2, GATAD2A | 0.0004 | T | 0.80841 | 2.61E-10 | ++ | 8.87E-09 | | New |
| 1331 | 19 | 19-19572678-AACGTG-A | rs1483340 | PBX4 | 0.0024 | A | | 1.36E-09 | ?+ | 1.355E-09 | 0.616 | New |
| 1332 | 19 | 19-20023807-C-CCT | | NA | 0.0002 | C | | 1.08E-08 | ?+ | 1.077E-08 | | New |

| | | | | | | | | | | | | |
|---|---|---|---|---|---|---|---|---|---|---|---|---|
| 1333 | 19 | 19-20076324-C-A | | ZNF682, ZNF90 | 0.0004 | A | 0.965211 | 8.82E-15 | -+ | 9.098E-13 | | New |
| 1334 | 19 | 19-20143926-G-A | rs6210700 | AC011447.3 | 0.0019 | G | 0.757766 | 6.88E-19 | ++ | 1.237E-13 | 0.6041 | New |
| 1334 | 19 | 19-20143939-G-A | rs5759339 | AC011447.3 | 0.0032 | A | 0.28918 | 7.88E-27 | -+ | 2.942E-15 | | New |
| 1335 | 19 | 19-20144534-C-CA | rs1555709 | AC011447.3 | 0.0015 | C | | 1.19E-08 | ?+ | 1.191E-08 | | New |
| 1336 | 19 | 19-22712186-C-CAT | | NA | 0.0008 | C | | 5.31E-10 | ?+ | 5.309E-10 | | New |
| 1336 | 19 | 19-22712189-GCC-G | | NA | 0.0008 | GCC | | 1.06E-09 | ?+ | 1.062E-09 | | New |
| 1337 | 19 | 19-22831340-TGAG-T | rs1160098 | ZNF723 | 0.0048 | T | 0.741934 | 3.31E-10 | -+ | 1.332E-09 | 0.918 | New |
| 1338 | 19 | 19-23676840-C-CAA | | NA | 0.0004 | CAA | | 1.41E-08 | ?+ | 1.406E-08 | | New |
| 1338 | 19 | 19-23676856-C-CCT | | NA | 0.0003 | CCT | | 3.76E-09 | ?+ | 3.755E-09 | | New |
| 1339 | 19 | 19-2654412-G-A | rs1250990 | GNG7 | 0.0012 | G | 0.745937 | 2.13E-10 | ++ | 7.761E-10 | | New |
| 1340 | 19 | 19-2673038-C-CCT | rs1189518 | GNG7 | 0.0023 | CCT | | 1.07E-12 | ?+ | 1.067E-12 | | New |
| 1341 | 19 | 19-27461080-G-A | rs1474832 | NONE, ERVK-28 | 0.0001 | A | | 2.55E-08 | ?+ | 2.549E-08 | | New |
| 1342 | 19 | 19-2870227-AGTTG-A | | NA | 0.0007 | A | | 4.27E-11 | ?+ | 4.269E-11 | | New |
| 1342 | 19 | 19-2870231-GCA-G | | NA | 0.0006 | GCA | | 1.62E-09 | ?+ | 1.623E-09 | | New |
| 1343 | 19 | 19-3259781-GA-G | rs1210150 | CELF5 | 0.0019 | GA | | 5.06E-12 | ?+ | 5.06E-12 | 0.121 | New |
| 1344 | 19 | 19-33033650-C-CCT | | NA | 0.0005 | C | | 1.94E-08 | ?+ | 1.936E-08 | | New |
| 1345 | 19 | 19-33141241-T-C | rs7249607 | WDR88 | 0.421 | T | | 4.48E-08 | ?- | 4.482E-08 | | New |
| 1346 | 19 | 19-33283110-C-T | rs5489606 | AC008738.6 | 0.0008 | C | 0.398578 | 6.11E-10 | ++ | 2.175E-09 | 0.001541 | New |
| 1347 | 19 | 19-33837044-AC-A | | NA | 0.0005 | A | | 1.45E-13 | ?+ | 1.45E-13 | | New |
| 1348 | 19 | 19-34031357-G-C | rs1237948 | AC016587.1 | 0.0026 | C | 0.001682 | 2.67E-09 | ++ | 4.554E-09 | 0.7012 | New |
| 1348 | 19 | 19-34031358-C-A | rs1283930 | AC016587.1 | 0.0027 | A | 0.000694 | 4.13E-08 | ++ | 8.569E-09 | | New |
| 1349 | 19 | 19-34371991-C-CCT | | NA | 0.0003 | CCT | | 3.43E-08 | ?+ | 3.429E-08 | | New |
| 1350 | 19 | 19-34465420-C-CG | | NA | 0.0006 | C | | 7.81E-11 | ?+ | 7.812E-11 | | New |

| | | | | | | | | | | | | |
|---|---|---|---|---|---|---|---|---|---|---|---|---|
| 1351 | 19 | 19-3479270-AAAG-A | rs1339749 | SMIM24 | 0.002 | AAAG | | 1.89E-08 | ?+ | 1.89E-08 | 0.197 | New |
| 1352 | 19 | 19-35386346-C-T | rs1444638 | GPR42, LINC01531 | 0.0011 | T | 0.938541 | 1.59E-10 | -+ | 0.000003799 | 0.05784 | New |
| 1352 | 19 | 19-35386354-C-T | rs1218821 | GPR42, LINC01531 | 0.0009 | T | 0.871465 | 1.71E-10 | -+ | 0.0000184 | 0.1173 | New |
| 1353 | 19 | 19-35630490-C-A | rs1297569 | RBM42 | 0.1723 | C | 0.400601 | 1.69E-08 | ++ | 0.00005176 | 0.9188 | New |
| 1354 | 19 | 19-3587428-C-CCCG | rs1221662 | GIPC3 | 0.0005 | CCCG | | 4.65E-08 | ?+ | 4.647E-08 | 0.06382 | New |
| 1355 | 19 | 19-35911172-C-CAT | | NA | 0.0004 | C | | 3.32E-08 | ?+ | 3.316E-08 | | New |
| 1356 | 19 | 19-36204814-ACC-A | rs1403003 | ZNF565 | 0.0029 | A | | 4.35E-26 | ?+ | 4.347E-26 | 0.385 | New |
| 1356 | 19 | 19-36204819-GCA-G | rs1362397 | ZNF565 | 0.0027 | GCA | | 4.97E-25 | ?+ | 4.972E-25 | 0.4866 | New |
| 1357 | 19 | 19-36516403-C-CTTGG | | NA | 0.0008 | C | | 1E-10 | ?+ | 1.003E-10 | | New |
| 1358 | 19 | 19-36868169-AT-A | | NA | 0.0013 | A | | 4.08E-13 | ?+ | 4.078E-13 | | New |
| 1359 | 19 | 19-36868792-AT-A | | NA | 0.0009 | AT | 0.155975 | 4.68E-11 | -+ | 1.72E-09 | | New |
| 1360 | 19 | 19-36950324-G-A | rs1314452 | ZNF568 | 0.0006 | G | | 8.16E-10 | ?+ | 8.158E-10 | 0.9056 | New |
| 1361 | 19 | 19-3719608-C-T | rs1182924 | TJP3 | 0.0004 | C | 0.407335 | 1.9E-09 | ++ | 9.298E-07 | 0.188 | New |
| 1362 | 19 | 19-37687722-C-CAG | | NA | 0.0005 | CAG | | 7.47E-09 | ?+ | 7.469E-09 | | New |
| 1363 | 19 | 19-38243249-GCC-G | | NA | 0.0009 | G | | 6.09E-09 | ?+ | 6.09E-09 | 0.5685 | New |
| 1364 | 19 | 19-3843252-C-CTT | rs1332151 | ZFR2 | 0.0072 | C | | 1.86E-20 | ?+ | 1.86E-20 | 0.4799 | New |
| 1365 | 19 | 19-39105162-TGC-T | rs1357694 | ACP7 | 0.0027 | TGC | | 1.24E-11 | ?+ | 1.237E-11 | 0.432 | New |
| 1365 | 19 | 19-39105164-C-CGT | rs2073397 | ACP7 | 0.0024 | C | | 3.54E-11 | ?+ | 3.537E-11 | | New |
| 1366 | 19 | 19-39108282-GGC-G | rs1294325 | ACP7 | 0.002 | G | | 2.85E-19 | ?+ | 2.854E-19 | | New |
| 1366 | 19 | 19-39108289-ATG-A | rs1282434 | ACP7 | 0.0016 | ATG | | 1.08E-12 | ?+ | 1.079E-12 | | New |
| 1367 | 19 | 19-39148690-C-CAG | rs1184157 | PAK4 | 0.0025 | CAG | | 5.18E-11 | ?+ | 5.18E-11 | | New |
| 1368 | 19 | 19-39301024-TCC-T | rs1465355 | IFNL1, LRFN1 | 0.0015 | TCC | 0.252972 | 2.06E-18 | ++ | 2.675E-18 | | New |

| | | | | | | | | | | | | |
|---|---|---|---|---|---|---|---|---|---|---|---|---|
| 1369 | 19 | 19-40402196-C-CAA | | NA | 0.0024 | C | | 4.7E-10 | ?+ | 4.701E-10 | | New |
| 1370 | 19 | 19-40791098-T-A | rs1013402 | AC008537.4 | 0.0022 | T | 0.264481 | 1.46E-10 | ++ | 2.894E-07 | 0.5752 | New |
| 1371 | 19 | 19-41157626-AGT-A | | NA | 0.0003 | A | | 2.07E-10 | ?+ | 2.067E-10 | | New |
| 1372 | 19 | 19-41509971-C-CT | | NA | 0.0059 | CT | | 1.17E-15 | ?+ | 1.169E-15 | | Old |
| 1373 | 19 | 19-41806330-TTG-T | | NA | 0.0013 | TTG | | 3.95E-11 | ?+ | 3.954E-11 | | Old |
| 1374 | 19 | 19-42482163-AT-A | rs1435969 | LIPE-AS1 | 0.0011 | AT | 0.60929 | 8.4E-09 | ++ | 0.00001325 | 0.7206 | New |
| 1375 | 19 | 19-4289268-G-A | rs1464924 | SHD | 0.004 | A | 0.745157 | 2.49E-11 | ++ | 0.003803 | 0.5932 | New |
| 1375 | 19 | 19-4289272-G-A | rs9910684 | SHD | 0.0049 | A | 0.985852 | 8.41E-10 | -+ | 0.01016 | 0.6715 | New |
| 1376 | 19 | 19-4302852-C-CA | | NA | 0.0002 | C | | 7.98E-09 | ?+ | 7.982E-09 | | New |
| 1377 | 19 | 19-43061627-AT-A | rs1396957 | AC004784.1 | 0.0008 | A | | 2.74E-08 | ?+ | 2.736E-08 | 0.7926 | Old |
| 1378 | 19 | 19-4331320-C-CCT | | NA | 0.0013 | C | | 5.99E-14 | ?+ | 5.986E-14 | | New |
| 1378 | 19 | 19-4331323-TCC-T | | NA | 0.0011 | TCC | | 1.53E-13 | ?+ | 1.53E-13 | | New |
| 1379 | 19 | 19-44457678-G-A | rs1205865 | ZNF229, ZNF180 | 0.0015 | A | 0.830066 | 7.08E-14 | ++ | 1.137E-12 | | Old |
| 1380 | 19 | 19-4450003-G-T | rs9690364 | AC011498.7 | 0.0041 | G | 0.660341 | 5.38E-27 | ++ | 5.918E-20 | 0.2727 | New |
| 1381 | 19 | 19-44862219-C-T | rs1166910 | NECTIN2 | 0.3254 | C | | 2.51E-08 | ?+ | 2.512E-08 | 0.0003802 | Old |
| 1382 | 19 | 19-44882783-ATT-A | | AC011481.2 | 0.1907 | A | | 1.66E-08 | ?+ | 1.656E-08 | | Old |
| 1382 | 19 | 19-44883210-GTAA-G | rs1420424 | AC011481.2 | 0.1325 | GTAA | 4.2E-210 | 2.42E-15 | ++ | 9.09E-202 | 1.61E-136 | Old |
| 1382 | 19 | 19-44884202-G-C | rs1297215 | AC011481.2 | 0.1323 | G | 3.2E-210 | 2.6E-15 | ++ | 7.7E-202 | 4.86E-140 | Old |
| 1382 | 19 | 19-44884339-G-A | rs1297297 | AC011481.2 | 0.1324 | A | 1.1E-209 | 1.06E-15 | ++ | 3.3E-202 | 2.69E-140 | Old |
| 1382 | 19 | 19-44884873-G-A | rs3434264 | AC011481.2 | 0.136 | A | 1.8E-203 | 3E-14 | ++ | 7.26E-194 | 4.9E-135 | Old |
| 1382 | 19 | 19-44885243-G-A | rs283811 | AC011481.2 | 0.2366 | G | 8.1E-195 | 6.03E-11 | ++ | 3.12E-174 | 7.23E-120 | Old |
| 1382 | 19 | 19-44887076-A-G | rs283815 | AC011481.2 | 0.2408 | A | 1.2E-203 | 1.55E-12 | ++ | 5.14E-184 | 3.05E-11 | Old |
| 1382 | 19 | 19-44888997-C-T | rs6857 | AC011481.2 | 0.1543 | T | 1.8E-287 | 1.25E-21 | ++ | 2.42E-277 | 2.33E-189 | Old |
| 1382 | 19 | 19-44891079-C-T | rs7135223 | TOMM40 | 0.1326 | C | 3.3E-214 | 1.97E-16 | ++ | 3.36E-207 | 4.62E-143 | Old |
| 1382 | 19 | 19-44891712-G-T | rs184017 | TOMM40 | 0.2369 | G | 5.1E-203 | 1.11E-12 | ++ | 6.68E-184 | 1.44E-124 | Old |
| 1382 | 19 | 19-44892362-G-A | rs2075650 | TOMM40 | 0.1386 | G | 8E-222 | 8.91E-18 | ++ | 8.01E-214 | 7.79E-130 | Old |

| | | | | | | | | | | | | |
|---|---|---|---|---|---|---|---|---|---|---|---|---|
| 1382 | 19 | 19-44892457-C-T | rs157581 | TOMM40 | 0.2402 | C | 1.4E-207 | 7.23E-13 | ++ | 8.82E-188 | | Old |
| 1382 | 19 | 19-44892587-G-A | rs3409532 | TOMM40 | 0.0979 | A | 7.5E-165 | 7.83E-12 | ++ | 7.73E-158 | 2.391E-85 | Old |
| 1382 | 19 | 19-44892652-G-C | rs3440455 | TOMM40 | 0.1359 | G | 4.2E-224 | 2.89E-18 | ++ | 2.72E-217 | 5.35E-135 | Old |
| 1382 | 19 | 19-44892887-C-T | rs1155650 | TOMM40 | 0.137 | T | 1.3E-222 | 4.98E-18 | ++ | 2.63E-215 | 1.93E-133 | Old |
| 1382 | 19 | 19-44892962-C-T | rs157582 | TOMM40 | 0.2385 | T | 6.4E-208 | 3.2E-13 | ++ | 5.76E-189 | 2.46E-135 | Old |
| 1382 | 19 | 19-44893408-G-T | rs5900738 | TOMM40 | 0.2146 | T | 1E-211 | 1.83E-11 | ++ | 1.14E-188 | 4.58E-11 | Old |
| 1382 | 19 | 19-44897790-AG-A | rs1555789 | TOMM40 | 0.1241 | AG | 7.7E-266 | 5.32E-19 | ++ | 3.22E-257 | | Old |
| 1382 | 19 | 19-44903416-G-A | rs10119 | TOMM40 | 0.2845 | A | 1.8E-143 | 3.21E-09 | ++ | 4.07E-131 | 1.67E-161 | Old |
| 1382 | 19 | 19-44906745-G-A | rs769449 | APOE | 0.1106 | A | 2.1E-299 | 4.75E-23 | ++ | 1.23E-291 | 6.74E-13 | Old |
| 1382 | 19 | 19-44908684-C-T | rs429358 | APOE | 0.1521 | C | 0 | 8.37E-31 | ++ | 3.26E-305 | 7.24E-305 | Old |
| 1382 | 19 | 19-44909521-C-CT | | AC011481.3 | 0.0435 | C | | 1.47E-14 | ?+ | 1.468E-14 | | Old |
| 1382 | 19 | 19-44909967-TGG-T | | AC011481.3 | 0.1109 | T | | 9.29E-28 | ?+ | 9.288E-28 | | Old |
| 1382 | 19 | 19-44912456-G-A | rs1041404 | AC011481.3 | 0.1261 | A | 3.1E-289 | 1.51E-20 | ++ | 7.93E-275 | 3.56E-146 | Old |
| 1382 | 19 | 19-44912678-G-T | rs7256200 | AC011481.3 | 0.126 | T | 2.2E-288 | 1.61E-20 | ++ | 5.4E-274 | | Old |
| 1382 | 19 | 19-44912921-G-T | rs483082 | AC011481.3 | 0.2449 | T | 9.3E-181 | 1.79E-09 | ++ | 3.74E-161 | 3.72E-139 | Old |
| 1382 | 19 | 19-44913484-C-T | rs438811 | AC011481.3 | 0.248 | T | 6.7E-182 | 1.84E-10 | ++ | 2.59E-164 | 9.06E-161 | Old |
| 1382 | 19 | 19-44914381-C-CTTCG | rs1156882 | AC011481.3 | 0.2283 | CTTCG | 1.1E-178 | 8.53E-11 | ++ | 1.61E-163 | 3.27E-124 | Old |
| 1382 | 19 | 19-44915533-C-T | rs5117 | APOC1 | 0.2283 | C | 9.4E-177 | 3.62E-10 | ++ | 1.25E-160 | 8.74E-129 | Old |
| 1382 | 19 | 19-44916825-C-A | rs7305233 | APOC1 | 0.0893 | C | | 7.55E-21 | ?+ | 7.552E-21 | | Old |
| 1382 | 19 | 19-44917997-G-A | rs1272104 | APOC1 | 0.1385 | A | 1.7E-230 | 3.32E-21 | ++ | 4.97E-231 | 6.77E-159 | Old |
| 1382 | 19 | 19-44918903-G-C | rs1272105 | APOC1 | 0.1707 | G | 6.4E-292 | 2.61E-22 | ++ | 1.57E-290 | 2.14E-228 | Old |
| 1382 | 19 | 19-44919589-G-A | rs5613119 | APOC1 | 0.1805 | A | 2.1E-291 | 1.15E-23 | ++ | 7.74E-288 | 1.89E-202 | Old |
| 1382 | 19 | 19-44919689-G-A | rs4420638 | APOC1 | 0.181 | G | 1.5E-297 | 8.06E-24 | ++ | 3.77E-288 | 2.22E-202 | Old |
| 1382 | 19 | 19-44920730-C-CA | rs3573397 | APOC1, APOC4 | 0.1693 | CA | 3.3E-226 | 3.92E-17 | ++ | 1.59E-215 | | Old |
| 1382 | 19 | 19-44921093-TAA-T | rs3683408 | APOC1, APOC4 | 0.0957 | T | | 3.33E-19 | ?+ | 3.335E-19 | | Old |
| 1382 | 19 | 19-44921095-ATTTT-A | rs7595158 | APOC1, APOC4 | 0.1107 | ATTTT | 0.946693 | 2.64E-19 | ++ | 2.675E-19 | | Old |
| 1382 | 19 | 19-44923868-T-A | rs1117893 | APOC1, APOC4 | 0.1397 | A | 9.4E-232 | 3.29E-19 | ++ | 2.14E-228 | 1.84E-158 | Old |
| 1382 | 19 | 19-44924977-G-A | rs6662699 | APOC1, APOC4 | 0.1471 | A | 3.1E-227 | 1.71E-19 | ++ | 5.84E-223 | 1.17E-130 | Old |

| | | | | | | | | | | | | |
|---|---|---|---|---|---|---|---|---|---|---|---|---|
| 1383 | 19 | 19-45108380-TC-T | rs1428666 | MARK4, PPP1R37 | 0.0011 | T | | 1.17E-09 | ?+ | 1.167E-09 | 0.7693 | Old |
| 1384 | 19 | 19-4520591-G-A | rs1832511 | PLIN4, PLIN5 | 0.0979 | G | | 1.55E-08 | ?+ | 1.551E-08 | 0.2554 | New |
| 1384 | 19 | 19-4666569-C-T | rs7845586 | MYDGF | 0.251 | T | | 7.31E-10 | ?+ | 7.307E-10 | 0.09779 | New |
| 1385 | 19 | 19-45260078-C-CT | | NA | 0.0008 | C | | 3.1E-13 | ?+ | 3.1E-13 | | Old |
| 1385 | 19 | 19-45260080-C-CT | | NA | 0.0008 | C | | 1.57E-10 | ?+ | 1.569E-10 | | Old |
| 1386 | 19 | 19-4528057-C-A | | PLIN5 | 0.0011 | A | 0.43248 | 3.57E-16 | -+ | 9.687E-16 | | New |
| 1387 | 19 | 19-45436408-C-CCT | | NA | 0.0009 | CCT | | 7.8E-09 | ?+ | 7.806E-09 | | Old |
| 1388 | 19 | 19-4562943-GA-G | rs1328984 | SEMA6B, TNFAIP8L1 | 0.0007 | G | | 4.51E-08 | ?+ | 4.509E-08 | 0.4593 | New |
| 1389 | 19 | 19-46832405-GGC-G | rs1304344 | AC008622.2, AP2S1 | 0.0061 | G | | 2.68E-11 | ?+ | 2.684E-11 | | Old |
| 1389 | 19 | 19-46832408-C-CAG | rs1395049 | AC008622.2, AP2S1 | 0.0035 | CAG | | 1.06E-12 | ?+ | 1.06E-12 | | Old |
| 1390 | 19 | 19-46844565-AGG-A | | NA | 0.0011 | AGG | | 7.85E-09 | ?+ | 7.848E-09 | | Old |
| 1390 | 19 | 19-46844567-GCA-G | | NA | 0.0005 | G | | 8.21E-10 | ?+ | 8.207E-10 | | Old |
| 1391 | 19 | 19-47213512-TC-T | rs1454695 | SAE1, BBC3 | 0.0015 | TC | | 7.6E-10 | ?+ | 7.601E-10 | 0.4644 | New |
| 1392 | 19 | 19-47458598-C-CTCT | | SLC8A2 | 0.003 | C | | 4.3E-08 | ?+ | 4.299E-08 | | New |
| 1393 | 19 | 19-4755334-C-CCT | | NA | 0.001 | C | | 1.25E-14 | ?+ | 1.251E-14 | | New |
| 1394 | 19 | 19-47604922-G-A | rs1473564 | RN7SL322P, AC010519.1 | 0.0015 | A | 0.187892 | 3.13E-10 | -+ | 0.0166 | 0.2349 | New |
| 1395 | 19 | 19-4788589-ACT-A | rs1197488 | AC005523.1 | 0.0029 | A | | 1.86E-12 | ?+ | 1.86E-12 | | New |
| 1396 | 19 | 19-48188862-T-A | rs1165043 | ZSWIM9 | 0.0013 | A | 0.390055 | 4.94E-14 | -+ | 0.00000277 | 0.07908 | Old |
| 1397 | 19 | 19-48189428-C-T | rs1435089 | ZSWIM9 | 0.0014 | T | 0.850937 | 3.88E-09 | -+ | 0.003146 | 0.7711 | Old |
| 1398 | 19 | 19-48564935-TGC-T | rs1291738 | AC008403.4 | 0.0032 | T | | 3.79E-28 | ?+ | 3.789E-28 | | Old |
| 1399 | 19 | 19-4862940-C-CCT | rs1215624 | PLIN3 | 0.0015 | CCT | | 4.99E-08 | ?+ | 4.988E-08 | | New |
| 1400 | 19 | 19-48690221-C-T | rs7458940 | NTN5, FUT2 | 0.1207 | T | | 4.33E-13 | ?+ | 4.332E-13 | | Old |

| | | | | | | | | | | | |
|---|---|---|---|---|---|---|---|---|---|---|---|
| 1401 | 19 | 19-48777085-G-C | rs1445575 | FGF21, RNU6-317P | 0.0006 | C | | 1.92E-10 | ?+ | 1.918E-10 | 0.06526 Old |
| 1402 | 19 | 19-48916454-C-CG | | NA | 0.0007 | CG | | 1.75E-10 | ?+ | 1.748E-10 | Old |
| 1403 | 19 | 19-4899269-G-A | rs5729819 | ARRDC5 | 0.0012 | A | 0.418919 | 1.74E-08 | -+ | 0.0007406 | 0.3947 New |
| 1404 | 19 | 19-49206127-ATTT-A | rs1252354 | TRPM4 | 0.0028 | A | | 1.56E-14 | ?+ | 1.557E-14 | 0.6103 Old |
| 1405 | 19 | 19-49291839-C-T | rs1970084 | SLC6A16 | 0.0064 | T | 0.559414 | 3.18E-11 | ++ | 0.00224 | Old |
| 1406 | 19 | 19-49714558-GCA-G | | NA | 0.0007 | G | | 3.54E-11 | ?+ | 3.537E-11 | Old |
| 1407 | 19 | 19-49905891-C-CCT | | NA | 0.0012 | CCT | | 2.19E-10 | ?+ | 2.192E-10 | Old |
| 1408 | 19 | 19-5175397-G-C | rs1472698 | KDM4B, AC022517.1 | 0.0003 | C | 0.2232 | 3.42E-09 | -+ | 5.549E-08 | New |
| 1408 | 19 | 19-5175404-G-A | rs2040076 | KDM4B, AC022517.1 | 0.0005 | A | | 5.31E-09 | ?+ | 5.306E-09 | New |
| 1409 | 19 | 19-5192343-G-A | rs2040147 | AC022517.1, PTPRS | 0.0014 | A | | 3.66E-14 | ?+ | 3.659E-14 | New |
| 1409 | 19 | 19-5192353-C-A | rs5374182 | AC022517.1, PTPRS | 0.0009 | A | | 5.87E-13 | ?+ | 5.87E-13 | New |
| 1410 | 19 | 19-5192936-G-A | rs9558701 | AC022517.1, PTPRS | 0.0003 | A | 0.40179 | 2.67E-08 | -+ | 1.203E-07 | New |
| 1411 | 19 | 19-52384389-GT-G | rs7590827 | ZNF880 | 0.0045 | G | | 1.82E-09 | ?+ | 1.817E-09 | New |
| 1412 | 19 | 19-53629086-GGC-G | | NA | 0.0003 | G | | 8.03E-10 | ?+ | 8.031E-10 | New |
| 1413 | 19 | 19-53947162-G-T | rs1415730 | CACNG7, CACNG8 | 0.0005 | T | | 3.43E-10 | ?+ | 3.425E-10 | Old |
| 1414 | 19 | 19-53950357-C-CTG | rs1323565 | CACNG7, CACNG8 | 0.0012 | C | | 2.15E-10 | ?+ | 2.146E-10 | Old |
| 1415 | 19 | 19-54059216-AAGCC-A | | NA | 0.0008 | AAGCC | | 2.61E-08 | ?+ | 2.611E-08 | Old |
| 1416 | 19 | 19-54953825-ATC-A | rs1213132 | NLRP2, NLRP7 | 0.0018 | ATC | | 6.4E-10 | ?+ | 6.395E-10 | 0.889 Old |
| 1417 | 19 | 19-55397554-GT-G | rs1190483 | RPL28 | 0.0006 | G | | 1.99E-09 | ?+ | 1.991E-09 | New |
| 1418 | 19 | 19-55421214-TTAAGA-T | rs1212560 | UBE2S, SHISA7 | 0.0006 | T | | 2.41E-08 | ?+ | 2.411E-08 | 0.4801 New |
| 1419 | 19 | 19-55422561-GGT-G | | NA | 0.0014 | GGT | | 1.06E-19 | ?+ | 1.062E-19 | New |

| | | | | | | | | | | | | |
|---|---|---|---|---|---|---|---|---|---|---|---|---|
| 1419 | 19 | 19-55422571-ATGC-A | | NA | 0.0008 | ATGC | | 6.12E-11 | ?+ | 6.12E-11 | | New |
| 1419 | 19 | 19-55422574-ACGCCTG-A | | NA | 0.0005 | A | | 4.04E-08 | ?+ | 4.036E-08 | | New |
| 1420 | 19 | 19-56879866-GT-G | rs1323625 | MIMT1, USP29 | 0.001 | G | | 4.96E-14 | ?+ | 4.956E-14 | | New |
| 1421 | 19 | 19-57064425-C-A | rs1983727 | MIMT1, USP29 | 1E-04 | C | | 1.08E-09 | ?+ | 1.083E-09 | | New |
| 1422 | 19 | 19-57066412-C-CA | rs1306363 | MIMT1, USP29 | 0.0022 | C | | 8.12E-14 | ?+ | 8.119E-14 | | New |
| 1423 | 19 | 19-5810053-C-CAGG | rs1157699 | DUS3L, NRTN | 0.0019 | C | | 4.25E-11 | ?+ | 4.254E-11 | 0.3393 | New |
| 1424 | 19 | 19-58524353-G-A | rs1477026 | ZBTB45 | 0.0004 | G | | 7.95E-10 | ?+ | 7.952E-10 | 0.8008 | New |
| 1425 | 19 | 19-58542262-G-GA | rs6747336 | ZBTB45, RN7SL525P | 0.0091 | G | 0.54052 | 2.01E-09 | ++ | 1.591E-07 | | New |
| 1426 | 19 | 19-6104375-C-T | rs1218624 | RFX2 | 0.0004 | T | 0.550938 | 3.26E-09 | -+ | 8.596E-08 | | New |
| 1426 | 19 | 19-6104380-C-T | rs1259238 | RFX2 | 0.0008 | C | | 9.02E-14 | ?+ | 9.015E-14 | | New |
| 1426 | 19 | 19-6104382-G-A | rs1318048 | RFX2 | 0.0009 | G | | 3.59E-10 | ?+ | 3.591E-10 | | New |
| 1427 | 19 | 19-6308254-G-A | rs1411623 | ACER1 | 0.0009 | A | 0.368815 | 5.11E-11 | -+ | 2.457E-09 | | New |
| 1428 | 19 | 19-6348245-ACC-A | rs1264751 | AC011491.3 | 0.0049 | A | | 5.33E-31 | ?+ | 5.334E-31 | 0.2312 | New |
| 1429 | 19 | 19-780466-ATT-A | rs1172865 | AC006273.1 | 0.0011 | A | 0.293829 | 2.8E-09 | -+ | 0.003651 | 0.8758 | Old |
| 1430 | 19 | 19-8042051-G-A | rs1045656 | AC008946.1, CCL25 | 0.0009 | A | 0.524986 | 4.85E-08 | -+ | 0.00008225 | 0.7755 | New |
| 1430 | 19 | 19-8042058-G-A | rs5734690 | AC008946.1, CCL25 | 0.0015 | G | | 4.06E-08 | ?+ | 4.06E-08 | | New |
| 1431 | 19 | 19-8165431-G-T | rs2083750 | FBN3, CERS4 | 0.0006 | G | | 4.6E-08 | ?+ | 0.000000046 | | New |
| 1431 | 19 | 19-8165439-C-CTT | | NA | 0.0005 | C | | 8.18E-10 | ?+ | 8.184E-10 | | New |
| 1432 | 19 | 19-8600010-AT-A | | NA | 0.001 | A | | 1.21E-11 | ?+ | 1.21E-11 | | New |
| 1433 | 19 | 19-9766170-G-A | rs5599806 | ZNF846 | 0.0004 | A | 0.746928 | 2.54E-09 | -+ | 6.177E-07 | 0.5532 | New |
| 1434 | 20 | 20-16241716-C-A | rs1175776 | AL161941.1 | 0.0023 | A | 0.905911 | 6.8E-14 | ++ | 0.00001997 | | New |
| 1435 | 20 | 20-16636840-T-A | rs3735584 | AL049794.2, RF00019 | 0.0015 | T | 0.667896 | 2.8E-08 | -+ | 0.001064 | | New |
| 1436 | 20 | 20-26363358-T-A | rs1198620 | MIR663AHG, NONE | 0.0002 | A | 0.464988 | 2.06E-08 | -+ | 8.382E-08 | | New |

| | | | | | | | | | | | | |
|---|---|---|---|---|---|---|---|---|---|---|---|---|
| 1437 | 20 | 20-30424031-TTGTGTG-T | rs1209602 | NA | 0.0081 | TTGTGTG | | 1.19E-20 | ?+ | 1.186E-20 | | New |
| 1438 | 20 | 20-31074716-T-A | rs1194281 | RNA5SP528, DEFB115 | 0.0058 | T | | 2.51E-08 | ?+ | 2.513E-08 | | New |
| 1439 | 20 | 20-31582540-G-A | | AL110115.2, ID1 | 0.0004 | G | 0.517815 | 2.71E-08 | ++ | 7.858E-07 | | New |
| 1440 | 20 | 20-31656375-GGC-G | rs1347052 | COX4I2, BCL2L1 | 0.0041 | G | | 1E-08 | ?+ | 1.001E-08 | 0.04583 | New |
| 1441 | 20 | 20-31703178-GA-G | | NA | 0.0029 | GA | | 7.04E-11 | ?+ | 7.043E-11 | 0.6649 | New |
| 1442 | 20 | 20-31750080-GGC-G | | TPX2 | 0.003 | G | | 1.38E-16 | ?+ | 1.377E-16 | 0.367 | New |
| 1443 | 20 | 20-32019470-C-CACCTTG | rs1473834 | AL031658.1 | 0.0057 | C | | 2.65E-08 | ?+ | 2.654E-08 | | New |
| 1444 | 20 | 20-32296095-G-C | rs1472081 | KIF3B | 0.0006 | G | 0.213378 | 7.61E-10 | -+ | 7.288E-09 | | New |
| 1445 | 20 | 20-32735144-AG-A | | NA | 0.0003 | A | | 3.59E-10 | ?+ | 3.59E-10 | | New |
| 1446 | 20 | 20-34646551-GCC-G | | NA | 0.0008 | GCC | | 6.01E-14 | ?+ | 6.009E-14 | | New |
| 1446 | 20 | 20-34646555-C-CCT | | NA | 0.0006 | C | | 1.74E-11 | ?+ | 1.74E-11 | | New |
| 1447 | 20 | 20-35097276-C-CAT | | TRPC4AP, EDEM2 | 0.0263 | C | | 1.34E-09 | ?+ | 1.337E-09 | | New |
| 1448 | 20 | 20-36058190-C-T | rs2057418 | AL035420.1 | 0.0013 | C | 0.865026 | 2.57E-08 | -+ | 0.01047 | | New |
| 1448 | 20 | 20-36058191-G-A | rs2057418 | AL035420.1 | 0.0014 | A | 0.965526 | 2.66E-10 | ++ | 0.000441 | | New |
| 1448 | 20 | 20-36058218-AT-A | | NA | 0.0008 | A | 0.244962 | 5.53E-09 | ++ | 2.341E-07 | | New |
| 1449 | 20 | 20-36190592-C-A | rs1034010 | EPB41L1 | 0.0022 | A | 0.69562 | 6.45E-22 | -+ | 1.842E-16 | 0.8273 | New |
| 1449 | 20 | 20-36190632-C-CTTACA | rs1407756 | EPB41L1 | 0.0015 | CTTACA | | 8.14E-17 | ?+ | 8.143E-17 | | New |
| 1450 | 20 | 20-36728254-C-CCG | | NA | 0.0006 | CCG | | 1.81E-08 | ?+ | 1.813E-08 | | New |
| 1451 | 20 | 20-36767785-C-T | rs1207337 | DSN1 | 0.0003 | C | 0.920236 | 1.74E-08 | -+ | 5.19E-08 | | New |
| 1452 | 20 | CCTGGCCAACATGG | | NA | 0.0016 | C | | 2.3E-16 | ?+ | 2.302E-16 | | New |
| 1453 | 20 | 20-36949029-C-CTG | rs1376535 | SAMHD1 | 0.0052 | CTG | | 2.16E-36 | ?+ | 2.164E-36 | 0.9581 | New |
| 1453 | 20 | 20-36949031-GGC-G | rs1311427 | SAMHD1 | 0.0046 | GGC | | 2.44E-29 | ?+ | 2.438E-29 | 0.9581 | New |

| | | | | | | | | | | | | |
|---|---|---|---|---|---|---|---|---|---|---|---|---|
| 1453 | 20 | 20-36949037-GATGC-G | | SAMHD1 | 0.0017 | G | | 3.74E-15 | ?+ | 3.74E-15 | | New |
| 1453 | 20 | 20-36949046-C-CCT | | SAMHD1 | 0.0016 | C | | 3.96E-16 | ?+ | 3.956E-16 | 0.7978 | New |
| 1454 | 20 | 20-37109452-C-CCT | | MROH8 | 0.0018 | C | | 1.66E-08 | ?+ | 1.659E-08 | 0.2231 | New |
| 1455 | 20 | 20-3850050-G-A | rs1382384 | MAVS | 0.0304 | A | | 6.72E-10 | ?+ | 6.72E-10 | 0.01817 | New |
| 1456 | 20 | 20-3850101-AC-A | rs1257128 | MAVS | 0.0029 | AC | 0.307591 | 1.8E-28 | -+ | 4.971E-15 | 0.04376 | New |
| 1457 | 20 | 20-38850804-TA-T | | NA | 0.001 | T | | 1.64E-10 | ?+ | 1.637E-10 | | New |
| 1458 | 20 | 20-3978903-C-CA | rs1472697 | RNF24 | 0.0012 | C | 0.915307 | 3.07E-20 | -+ | 6.556E-17 | | New |
| 1459 | 20 | 20-4081212-AAT-A | rs1411904 | AL356414.1, SMOX | 0.0018 | A | | 4.02E-08 | ?+ | 4.015E-08 | 0.9146 | New |
| 1460 | 20 | 20-43469358-ATTT-A | | NA | 0.0022 | ATTT | | 1.48E-16 | ?+ | 1.482E-16 | | New |
| 1461 | 20 | 20-43652786-ACC-A | | NA | 0.0008 | ACC | | 5.18E-11 | ?+ | 5.179E-11 | | New |
| 1461 | 20 | 20-43652787-TGA-T | | NA | 0.0005 | T | | 2.66E-09 | ?+ | 2.658E-09 | | New |
| 1462 | 20 | 20-45931856-ACC-A | | NA | 0.0008 | A | | 4.09E-10 | ?+ | 4.093E-10 | | New |
| 1463 | 20 | 20-47194165-GCA-G | | NA | 0.0011 | G | | 1.15E-08 | ?+ | 1.155E-08 | | New |
| 1464 | 20 | 20-47381012-ACT-A | | ZMYND8, LINC01754 | 0.0011 | ACT | | 2.67E-13 | ?+ | 2.67E-13 | | New |
| 1465 | 20 | 20-48501959-AG-A | rs1445715 | RNU7-144P, PREX1 | 0.0026 | A | | 2.43E-09 | ?+ | 2.432E-09 | 0.3724 | New |
| 1465 | 20 | 20-48501962-AG-A | rs1306818 | RNU7-144P, PREX1 | 0.0021 | AG | | 3.17E-11 | ?+ | 3.168E-11 | 0.4386 | New |
| 1466 | 20 | 20-49061328-ATG-A | rs1555821 | CSE1L | 0.0043 | A | | 4.05E-11 | ?+ | 4.048E-11 | | New |
| 1466 | 20 | 20-49061331-C-CCT | rs1214091 | CSE1L | 0.0034 | C | | 1.37E-10 | ?+ | 1.367E-10 | 0.2898 | New |
| 1467 | 20 | 20-49198329-C-CA | rs1219080 | STAU1, DDX27 | 0.0013 | CA | | 4.39E-08 | ?+ | 4.393E-08 | 0.3423 | New |
| 1468 | 20 | 20-49202549-GCA-G | rs1197599 | STAU1, DDX27 | 0.0005 | G | | 3.42E-08 | ?+ | 3.421E-08 | 0.6938 | New |
| 1469 | 20 | 20-506117-C-T | rs5632444 | CSNK2A1 | 0.0011 | T | 0.835076 | 9.93E-12 | -+ | 4.457E-07 | | Old |
| 1469 | 20 | 20-506120-C-T | rs1006238 | CSNK2A1 | 0.0012 | T | 0.873483 | 3.06E-10 | -+ | 0.000002923 | 0.7945 | Old |

| | | | | | | | | | | | | |
|---|---|---|---|---|---|---|---|---|---|---|---|---|
| 1470 | 20 | 20-51520826-AT-A | | NA | 0.0003 | AT | | 4.97E-08 | ?+ | 4.974E-08 | | New |
| 1470 | 20 | 20-51520827-T-A | rs2076432 | NFATC2 | 0.0005 | T | | 7.78E-10 | ?+ | 7.775E-10 | | New |
| 1471 | 20 | 20-53482754-G-A | rs1057509 | AL109930.1, AL354993.2 | 0.0028 | G | 0.222995 | 4.27E-10 | ++ | 2.4E-09 | | New |
| 1471 | 20 | 20-53482760-G-A | rs1020277 | AL109930.1, AL354993.2 | 0.0013 | A | 0.837682 | 1.03E-08 | ++ | 0.000000169 | 0.7384 | New |
| 1472 | 20 | 20-53698443-G-C | rs1237656 | RNU7-14P, AC005914.1 | 0.001 | C | | 7.67E-12 | ?+ | 7.672E-12 | | New |
| 1473 | 20 | 20-59401313-G-GCA | | AL160410.1, AL389889.2 | 0.0165 | G | | 8.69E-11 | ?+ | 8.691E-11 | | New |
| 1474 | 20 | 20-62278275-T-TTG | rs1147200 | OSBPL2 | 0.0254 | TTG | 0.270412 | 7.67E-10 | -+ | 6.835E-07 | | New |
| 1475 | 20 | 20-62399827-C-T | rs1162057 | CABLES2 | 0.0012 | T | 0.673279 | 2.28E-10 | -+ | 2.663E-08 | | New |
| 1476 | 20 | 20-62865769-C-CTT | rs2064056 | TCFL5, DIDO1 | 0.0009 | CTT | | 1.1E-12 | ?+ | 1.104E-12 | | New |
| 1476 | 20 | 20-62865772-G-T | rs2064056 | TCFL5, DIDO1 | 0.0004 | T | | 1.93E-09 | ?+ | 1.93E-09 | | New |
| 1477 | 20 | 20-63461661-GGGA-G | rs1409928 | KCNQ2 | 0.0043 | G | 0.461209 | 3.04E-09 | -+ | 4.141E-09 | | Old |
| 1478 | 20 | 20-63821488-G-A | rs1600724 | ZBTB46 | 0.0007 | A | 0.144102 | 1.85E-08 | ++ | 0.000001204 | | Old |
| 1479 | 20 | 20-63902200-G-A | rs1238646 | DNAJC5 | 0.0007 | A | 0.985258 | 1.95E-11 | ++ | 1.559E-07 | 0.4922 | Old |
| 1479 | 20 | 20-63902207-G-A | rs1379210 | DNAJC5 | 0.0005 | G | 0.847461 | 1.01E-08 | ++ | 0.00002096 | | Old |
| 1480 | 21 | 21-10415599-C-T | | AP003900.1, AF254983.1 | 0.0035 | C | | 6.41E-12 | ?- | 6.412E-12 | | New |
| 1480 | 21 | 21-10415600-G-A | | AP003900.1, AF254983.1 | 0.0039 | A | | 6.14E-12 | ?- | 6.143E-12 | | New |
| 1481 | 21 | 21-10717092-A-C | rs2836848 | IGHV1OR21-1, NONE | 0.0044 | A | | 2.07E-09 | ?+ | 2.067E-09 | | New |

| | | | | | | | | | | | | |
|---|---|---|---|---|---|---|---|---|---|---|---|---|
| 1482 | 21 | 21-10756490-AG-A | | NA | 0.0031 | A | | 3.52E-14 | ?+ | 3.518E-14 | | New |
| 1483 | 21 | 21-23757294-C-CCT | | NA | 0.0005 | C | | 6.73E-10 | ?+ | 6.734E-10 | | New |
| 1484 | 21 | 21-28685091-AAAC-A | | AF165147.1, RF00026 | 0.0007 | A | | 1.6E-09 | ?+ | 1.603E-09 | | New |
| 1485 | 21 | 21-28995088-GTGA-G | rs1461827 | LTN1, RWDD2B | 0.0006 | G | | 4.42E-09 | ?+ | 4.417E-09 | 0.4982 | New |
| 1486 | 21 | 21-31690330-C-CCAA | | SCAF4 | 0.0007 | CCAA | | 3.33E-09 | ?+ | 3.335E-09 | | New |
| 1487 | 21 | 21-39324090-AG-A | rs1372913 | BRWD1-AS1 | 0.0036 | A | | 1.65E-15 | ?+ | 1.655E-15 | 0.896 | New |
| 1488 | 21 | 21-39383050-AAG-A | | NA | 0.0006 | AAG | | 2.17E-10 | ?+ | 2.169E-10 | | New |
| 1488 | 21 | 21-39383052-ACC-A | | NA | 0.0005 | A | | 2.17E-11 | ?+ | 2.167E-11 | | New |
| 1489 | 21 | 21-42718089-C-CG | rs1477539 | PDE9A | 0.0019 | CG | | 1.56E-16 | ?+ | 1.563E-16 | 0.9578 | New |
| 1489 | 21 | 21-42718092-GA-G | rs1172964 | PDE9A | 0.0026 | G | | 2.27E-16 | ?+ | 2.266E-16 | 0.5991 | New |
| 1490 | 21 | 21-43788382-G-C | rs1445911 | CSTB, RRP1 | 0.0006 | C | 0.963143 | 1.16E-08 | -+ | 6.394E-07 | 0.19 | New |
| 1491 | 21 | 21-45155722-TCCACCCAC-T | rs1995514 | ADARB1 | 0.0031 | T | | 1.21E-11 | ?+ | 1.207E-11 | | New |
| 1492 | 21 | 21-45500131-GGGTGGA-G | | NA | 0.0009 | G | | 1.03E-08 | ?+ | 1.032E-08 | | New |
| 1493 | 21 | 21-46085044-C-CT | rs1168284 | AP001476.2, AP001471.1 | 0.0014 | C | | 2.72E-08 | ?+ | 2.724E-08 | | New |
| 1494 | 21 | 21-46390054-G-A | rs1039886 | PCNT | 0.0007 | G | 0.304421 | 5.26E-11 | ++ | 3.221E-11 | | New |
| 1495 | 21 | 21-9124671-AACCCAAAACAATGGGAGTGACGTGCTAAAACCATT-A | | NA | 0.0007 | A | | 4.72E-08 | ?+ | 4.723E-08 | | New |
| 1496 | 21 | 21-9817035-GAA-G | | NA | 0.0048 | G | | 1.95E-10 | ?+ | 1.948E-10 | | New |
| 1497 | 22 | 22-11368837-G-T | rs1174371 | RF00002, NONE | 0.0069 | G | 0.43741 | 7.03E-10 | -+ | 7.49E-09 | | New |

| | | | | | | | | | | | | |
|---|---|---|---|---|---|---|---|---|---|---|---|---|
| 1498 | 22 | 22-12556740-C-CACACACACACACAGAG | | NA | 0.0015 | C | | 1.46E-08 | ?+ | 1.463E-08 | | New |
| 1499 | 22 | 22-16279325-G-C | rs1265426 | RF00026, AP000547.2 | 0.0002 | C | | 4.66E-08 | ?+ | 4.66E-08 | | New |
| 1500 | 22 | 22-17618008-C-A | rs1393446 | ATP6V1E1 | 0.0015 | A | 0.160312 | 1.61E-13 | -+ | 0.00002559 | | New |
| 1501 | 22 | 22-18160517-T-A | rs1304142 | USP18 | 0.001 | T | 0.974412 | 3.03E-10 | ++ | 0.000003044 | | New |
| 1502 | 22 | 22-19300549-C-CCT | rs1555998 | AC000085.1 | 0.002 | C | | 1.55E-14 | ?+ | 1.555E-14 | 0.3646 | New |
| 1502 | 22 | 22-19300566-G-A | rs9218908 | AC000085.1 | 0.0003 | A | 0.392246 | 5.45E-12 | ++ | 1.174E-11 | | New |
| 1503 | 22 | 22-19311896-C-CAAAA | | AC000085.1 | 0.0224 | CAAAA | | 8.99E-09 | ?+ | 8.989E-09 | | New |
| 1504 | 22 | 22-20050079-TGG-T | | NA | 0.0005 | T | | 4.26E-11 | ?+ | 4.259E-11 | | New |
| 1505 | 22 | 22-20503118-ACC-A | | NA | 0.0009 | A | | 1.12E-09 | ?+ | 1.118E-09 | | New |
| 1506 | 22 | 22-21924568-GCGAAC-G | | NA | 0.0005 | GCGAAC | | 7.45E-15 | ?+ | 7.453E-15 | | New |
| 1507 | 22 | 22-22251998-C-T | rs1404067 | VPREB1, AC245060.6 | 0.0005 | C | | 1.02E-08 | ?+ | 1.02E-08 | | New |
| 1508 | 22 | 22-22518612-AC-A | | NA | 0.0012 | A | | 8.5E-16 | ?+ | 8.5E-16 | | New |
| 1509 | 22 | 22-22732613-T-A | rs1322705 | IGLV3-19, IGLV2-18 | 0.0091 | T | 0.718562 | 4.71E-13 | ++ | 0.003911 | 0.802 | New |
| 1510 | 22 | 22-23515705-G-A | rs1502917 | LINC02557, PCAT14 | 0.0988 | G | | 3.54E-10 | ?+ | 3.541E-10 | 0.8512 | New |
| 1511 | 22 | 22-23646338-ATAAGAAAGATTTT-A | | NA | 0.0005 | ATAAGAAAGATTTT | | 1.7E-08 | ?+ | 1.698E-08 | | New |
| 1512 | 22 | 22-23719208-C-T | rs1269074 | GUSBP11, ZNF70 | 0.0011 | C | 0.796841 | 5.72E-14 | ++ | 9.068E-09 | | New |
| 1513 | 22 | 22-23797852-C-CAG | | NA | 0.0006 | C | | 2.84E-13 | ?+ | 2.844E-13 | | New |
| 1514 | 22 | 22-24490949-C-CAA | | NA | 0.0007 | C | | 6.89E-12 | ?+ | 6.889E-12 | | New |
| 1515 | 22 | 22-24777872-ATT-A | | NA | 0.0017 | ATT | | 1.08E-13 | ?+ | 1.083E-13 | | New |
| 1516 | 22 | 22-25252791-C-CCA | rs1177181 | Z99916.3 | 0.0042 | CCA | | 1.49E-31 | ?+ | 1.488E-31 | 0.6766 | New |

| | | | | | | | | | | | | |
|---|---|---|---|---|---|---|---|---|---|---|---|---|
| 1516 | 22 | 22-25252792-AGAGGTG-A | rs1372844 | Z99916.3 | 0.0035 | A | | 3.09E-28 | ?+ | 3.092E-28 | 0.7925 | New |
| 1516 | 22 | 22-25252802-AAGCG-A | rs1299512 | Z99916.3 | 0.0033 | AAGCG | | 7.61E-23 | ?+ | 7.61E-23 | | New |
| 1517 | 22 | 22-25324817-G-T | rs1203341 | AL022324.4, LRP5L | 0.0008 | T | | 1.65E-09 | ?+ | 1.655E-09 | | New |
| 1518 | 22 | 22-25345441-AGGGGTGAGCCCCTGCTCTCAGCCTCCCACAGTGCTGGGATTACT-A | | AL022324.4, LRP5L | 0.0077 | A | | 5.07E-14 | ?+ | 5.067E-14 | | New |
| 1519 | 22 | 22-28089534-GAA-G | | NA | 0.001 | GAA | | 2.23E-08 | ?+ | 2.228E-08 | 0.9003 | New |
| 1520 | 22 | 22-28536011-C-CCATCCTGGCTAA | rs1339933 | TTC28 | 0.0037 | CCATCCTGGCTAA | | 3.58E-10 | ?+ | 3.579E-10 | | New |
| 1521 | 22 | 22-28536542-G-T | rs9171907 | TTC28 | 0.0014 | G | 0.895935 | 9.09E-13 | -+ | 6.138E-07 | 0.896 | New |
| 1522 | 22 | 22-28645472-C-CA | rs1312893 | TTC28 | 0.0024 | CA | | 1.71E-11 | ?+ | 1.71E-11 | 0.1333 | New |
| 1523 | 22 | 22-28770546-AT-A | rs1469740 | HSCB, CCDC117 | 0.0018 | AT | | 5.78E-11 | ?+ | 5.776E-11 | 0.05477 | New |
| 1524 | 22 | 22-29148578-C-CG | | NA | 0.0006 | C | | 4.85E-12 | ?+ | 4.852E-12 | | New |
| 1525 | 22 | 22-30913268-C-CG | rs1312458 | OSBP2, MORC2-AS1 | 0.0038 | C | | 1.26E-18 | ?+ | 1.258E-18 | | New |
| 1526 | 22 | 22-31293758-GT-G | rs1308349 | PIK3IP1-AS1 | 0.0006 | GT | | 3.08E-08 | ?+ | 3.075E-08 | | New |
| 1527 | 22 | 22-35050426-C-A | rs1162255 | AL024495.1, ISX | 0.0002 | A | | 2.49E-09 | ?+ | 2.493E-09 | | New |
| 1528 | 22 | 22-35699725-GCCCC-G | | NA | 0.0045 | G | | 2.3E-10 | ?+ | 2.304E-10 | | New |
| 1529 | 22 | 22-36068076-G-A | rs1317768 | Z95114.4 | 0.0006 | A | 0.160306 | 3.04E-10 | ++ | 5.489E-09 | 0.151 | New |
| 1529 | 22 | 22-36068090-G-A | rs5579907 | Z95114.4 | 0.0009 | A | 0.737581 | 1.47E-16 | ++ | 2.825E-14 | 0.1699 | New |
| 1530 | 22 | 22-36389311-C-CCT | | NA | 0.0006 | C | | 1.5E-08 | ?+ | 1.503E-08 | | New |
| 1531 | 22 | 22-37939702-T-A | | MICALL1 | 0.0005 | A | | 9.1E-10 | ?+ | 9.096E-10 | | New |
| 1532 | 22 | 22-38242246-TTG-T | | NA | 0.0003 | T | | 1.39E-08 | ?+ | 1.394E-08 | | New |
| 1533 | 22 | 22-38342004-C-CCA | | NA | 0.0012 | CCA | | 5.16E-12 | ?+ | 5.159E-12 | | New |
| 1534 | 22 | 22-38405984-AT-A | | NA | 0.0012 | AT | | 1.33E-09 | ?+ | 1.328E-09 | 0.1544 | New |
| 1535 | 22 | 22-38654128-G-C | rs9429344 | FAM227A | 0.0026 | C | 0.946907 | 4.83E-28 | ++ | 3.783E-14 | | New |

| | | | | | | | | | | | |
|---|---|---|---|---|---|---|---|---|---|---|---|
| 1536 | 22 | 22-39166463-C-CCT | | NA | 0.0008 | CCT | | 1.69E-09 | ?+ | 1.688E-09 | | New |
| 1537 | 22 | 22-39166662-AG-A | | NA | 0.0006 | AG | | 4.37E-08 | ?+ | 4.375E-08 | | New |
| 1538 | 22 | 22-40796022-C-CAG | | NA | 0.0013 | C | 0.493337 | 1.83E-13 | -+ | 3.587E-12 | | New |
| 1539 | 22 | 22-40935856-GT-G | | NA | 0.001 | G | | 1.89E-09 | ?+ | 1.89E-09 | | New |
| 1540 | 22 | 22-41293017-C-CG | rs1186350 | RANGAP1, ZC3H7B | 0.0013 | C | | 4.5E-09 | ?+ | 4.497E-09 | 0.2797 | New |
| 1541 | 22 | 22-41379102-ATGCC-A | | TEF | 0.0027 | ATGCC | | 1.98E-10 | ?+ | 1.98E-10 | 0.5427 | New |
| 1542 | 22 | 22-41609575-C-CTTG | | NA | 0.0005 | C | | 3.18E-08 | ?+ | 3.176E-08 | | New |
| 1543 | 22 | 22-41712999-C-T | rs1027884 | MEI1 | 0.001 | T | 0.479323 | 3.05E-08 | ++ | 7.426E-08 | 0.8669 | New |
| 1544 | 22 | 22-41786143-C-T | rs2075977 | MEI1 | 0.0003 | T | | 8.5E-10 | ?+ | 8.5E-10 | | New |
| 1544 | 22 | 22-41786146-G-A | rs1277118 | MEI1 | 0.0002 | A | 0.263165 | 6.75E-09 | -+ | 7.177E-08 | | New |
| 1544 | 22 | 22-41786153-G-T | rs1364149 | MEI1 | 0.0006 | T | 0.937115 | 1.62E-10 | -+ | 6.88E-10 | | New |
| 1545 | 22 | 22-41795898-TG-T | rs1395390 | MEI1 | 0.0056 | TG | 0.54594 | 2.17E-11 | -+ | 0.03323 | 0.2962 | New |
| 1545 | 22 | 22-41795903-C-T | rs9869179 | MEI1 | 0.0055 | T | 0.54594 | 6.41E-15 | -+ | 0.009858 | 0.2246 | New |
| 1546 | 22 | 22-43016634-C-T | rs7290470 | PACSIN2, AL022476 .1 | 0.0987 | T | | 3.58E-08 | ?+ | 3.579E-08 | 0.02898 | New |
| 1547 | 22 | 22-44675142-C-A | rs9829090 | PRR5 | 0.0024 | A | 0.695644 | 1.2E-11 | -+ | 0.0008807 | 0.5883 | New |
| 1548 | 22 | 22-44749165-AC-A | rs1388705 | PRR5-ARHGAP8 | 0.0015 | AC | | 5.55E-09 | ?+ | 5.552E-09 | 0.04027 | New |
| 1549 | 22 | 22-44886986-C-CCA | | PHF21B | 0.0007 | CCA | | 2.32E-09 | ?+ | 2.319E-09 | | New |
| 1550 | 22 | 22-45519419-G-C | rs1375675 | FBLN1 | 0.0014 | C | 0.849274 | 2.62E-08 | -+ | 0.002129 | | New |
| 1550 | 22 | 22-45519431-G-A | rs1198184 | FBLN1 | 0.0014 | A | 0.849449 | 2E-10 | -+ | 0.0005469 | 0.4057 | New |
| 1551 | 22 | 22-46713286-C-CTT | | NA | 0.0004 | CTT | | 1.15E-11 | ?+ | 1.151E-11 | | New |
| 1552 | 22 | 22-48399528-C-T | rs1137173 | MIR3201, Z72006.1 | 0.0832 | C | | 2.07E-13 | ?+ | 2.069E-13 | 0.5675 | New |
| 1553 | 22 | 22-48910553-C-CCCCTT | rs1169595 | LINC0131 0, AL078622 .1 | 0.0007 | C | | 3.77E-09 | ?+ | 3.774E-09 | | New |
| 1554 | 22 | 22-49399428-C-CCA | | NA | 0.0002 | C | | 3.98E-10 | ?+ | 3.978E-10 | | New |
| 1555 | 22 | 22-49879291-AT-A | | NA | 0.0005 | A | | 4.9E-10 | ?+ | 4.905E-10 | | New |

| | | | | | | | | | | | |
|---|---|---|---|---|---|---|---|---|---|---|---|
| 1556 | 22 | 22-50354109-ACC-A | rs1555964 | PPP6R2 | 0.0019 | A | 2.87E-11 | ?+ | 2.872E-11 | | New |
| 1556 | 22 | 22-50354112-GGC-G | rs1240189 | PPP6R2 | 0.0022 | GGC | 6.48E-11 | ?+ | 6.482E-11 | 0.02318 | New |
| 1557 | 22 | 22-50372984-GT-G | | NA | 0.0006 | G | 2.7E-10 | ?+ | 2.697E-10 | | New |
| 1558 | 22 | 22-50543535-TGCA-T | rs1356120 | U62317.1; U62317.4 | 0.0036 | TGCA | 3.16E-08 | ?+ | 3.162E-08 | 0.01972 | New |
| 1558 | 22 | 22-50543540-C-CGGT | rs1483504 | U62317.1; U62317.4 | 0.0048 | C | 1.28E-14 | ?+ | 1.284E-14 | | New |
