## Supplementary Table 7 for "Identification of 16 novel Alzheimer’s disease susceptibility loci using multi-ancestry meta-analyses of clinical Alzheimer’s disease and AD-by-proxy cases from four whole genome sequencing datasets"

**Supplementary table 7:** Hardy-Weinberg equilibrium statistics and power of Bellenguez et al. lead variants in the AoU cohort.

P - p-value; ALL - full AoU dataset, all ancestries; AFR - African ancestry subset; AMR - admixed American subset; EAS - east Asian subset; EUR - European subset; MID - Middle East subset; SAS - South Asian subset .

| | | | | | | | | Population Stratified P-values for AOU | | | P-value for Hardy-Weinberg equilibrium | | | | | | |
|---|---|---|---|---|---|---|---|---|---|---|---|---|---|---|---|---|---|
| Variant ID | Chromosome | Rsid | Effect alele | Bellenguez et al. P | AOU_P | UKB_P | Power AoU | AFR | AMR | EUR | ALL | AFR | AMR | EAS | EUR | MID | SAS |
| 1-109345810-C-T | 1 | rs141749679 | C | 7.54E-09 | 0.46249176 | 0.009967816 | 0.005495621 | 0.929143943 | 0.137442485 | 0.75282321 | 0.744203 | 0.507491 | 0.548672 | 0.500132 | 0.746497 | 0.5 | 0.502822 |
| 1-207577223-T-C | 1 | rs679515 | T | 7.16E-46 | 0.03141377 | | 0.93972961 | 0.413322455 | 0.036253505 | 0.046555393 | 4.9238E-99 | 0.439301 | 0.078868 | 0.720436 | 0.078812 | 0.751299 | 0.713088 |
| 10-11676714-G-A | 10 | rs7912495 | G | 9.74E-19 | 0.16346207 | 0.004067242 | 0.117428372 | 0.608567843 | 0.011810275 | 0.613955649 | 8.80075E-43 | 0.146483 | 1.5349E-05 | 0.063757 | 0.0169 | 0.30601 | 0.303644 |
| 10-122413396-G-A | 10 | rs7908662 | G | 2.59E-09 | 0.84147903 | 0.000591453 | 0.006905419 | 0.46460192 | 0.426339921 | 0.535207265 | 1.15443E-34 | 0.904011 | 6.581E-11 | 0.588189 | 0.234719 | 0.619337 | 0.325071 |
| 10-60025170-T-G | 10 | rs7068231 | T | 3.32E-13 | 0.13932402 | 0.005792153 | 0.039247268 | 0.455665345 | 0.568779585 | 0.090446132 | 7.418E-195 | 0.348634 | 1.0285E-28 | 0.328856 | 0.549479 | 0.603583 | 0.026657 |
| 10-80494228-C-T | 10 | rs6586028 | C | 1.97E-19 | 0.39540031 | 0.001123777 | 0.11120976 | 0.157397562 | 0.521584888 | 0.669112279 | 1.24461E-76 | 0.638227 | 0.440202 | 0.416164 | 0.814315 | 0.908677 | 0.347091 |
| 10-96266650-G-A | 10 | rs6584063 | G | 6.73E-11 | 0.63980997 | 0.149173267 | 0.024839234 | 0.105051196 | 0.160526597 | 0.783072549 | 0.204395 | 0.73457 | 0.214545 | 0.814755 | 0.839492 | 0.074162 | 0.300667 |
| 11-121482368-G-T | 11 | rs74685827 | G | 2.81E-11 | 0.14497596 | 0.070134225 | 0.022638326 | 0.219513382 | 0.438413582 | 0.174344399 | 1.18298E-07 | 0.777325 | 0.281269 | 0.775871 | 0.723557 | 0.501615 | 0.55371 |
| 11-121564878-C-T | 11 | rs11218343 | C | 1.40E-21 | 0.14842532 | 0.124202689 | 0.309400703 | 0.04048741 | 0.577642315 | 0.03508488 | 2.05862E-41 | 0.184323 | 0.497881 | 0.058932 | 0.503865 | 0.216344 | 0.635825 |
| 11-47370397-A-G | 11 | rs10437655 | A | 5.28E-14 | 0.44272087 | 0.007655437 | 0.103082269 | 0.105131292 | 0.238779479 | 0.21493394 | 0.000108603 | 0.00186411 | 0.0297209 | 0.002158 | 0.114147 | 0.079841 | 0.816954 |
| 11-60254475-A-G | 11 | rs1582763 | A | 3.74E-42 | 0.20955161 | 0.010911137 | 0.891576864 | 0.32639162 | 0.325674684 | 0.409302126 | 7.897E-267 | 0.03796 | 0.399147 | 0.858479 | 0.138029 | 0.713537 | 0.088086 |
| 11-86157598-T-C | 11 | rs3851179 | T | 2.95E-48 | 0.55268286 | 0.000601866 | 0.975210552 | 0.617438918 | 0.513246504 | 0.541126746 | 1.4624E-106 | 0.121769 | 0.262755 | 0.922333 | 0.03713 | 0.696688 | 0.05078 |
| 12-113281983-C-T | 12 | rs6489896 | C | 1.80E-09 | 0.05967329 | 0.343158689 | 0.007123547 | 0.242225578 | 0.795540511 | 0.173507399 | 6.62838E-16 | 0.0117766 | 0.965488 | 0.594859 | 0.612651 | 0.482886 | 0.956394 |
| 14-105761758-A-G | 14 | rs7157106 | A | 1.99E-08 | 0.42495603 | | 0.022045131 | 0.333762085 | 0.977559544 | 0.756916548 | 5.8971E-302 | 0.0216255 | 3.4796E-19 | 3.05E-06 | 7.58E-05 | 0.069873 | 0.46402 |
| 14-106665591-A-G | 14 | rs10131280 | A | 4.26E-10 | 0.52918192 | 0.072408575 | 0.008832117 | 0.413064622 | 0.558916589 | 0.618071901 | 5.20071E-05 | 0.27059 | 0.678683 | 0.004134 | 0.816628 | 0.668507 | 0.621199 |
| 14-52924962-G-A | 14 | rs17125924 | G | 8.32E-16 | 0.46726043 | 0.008223941 | 0.071551596 | 0.633839062 | 0.332933886 | 0.542090754 | 6.1332E-08 | 0.753503 | 0.0135357 | 0.715488 | 0.716784 | 0.858136 | 0.459854 |
| 14-92464917-G-A | 14 | rs7401792 | G | 4.83E-08 | 0.8634689 | | 0.003782804 | 0.566929427 | 0.83479108 | 0.547177209 | 1.9337E-302 | 0.130083 | 0.00035847 | 0.015604 | 0.207846 | 0.575825 | 0.92937 |
| 14-92472511-A-G | 14 | rs12590654 | A | 4.25E-21 | 0.70234743 | 3.96515E-06 | 0.325839582 | 0.625272027 | 0.587293237 | 0.709270668 | 0.944603 | 0.314297 | 0.137153 | 0.311721 | 0.598344 | 0.130141 | 0.928873 |
| 15-50701814-G-A | 15 | rs8025980 | G | 1.32E-08 | 0.78570226 | | 0.004610718 | 0.899553298 | 0.81487745 | 0.703075558 | 1.7416E-08 | 0.0922586 | 0.0678628 | 0.475811 | 0.877354 | 0.018332 | 0.302261 |
| 15-58764824-A-T | 15 | rs602602 | A | 2.07E-15 | 0.99210711 | 0.019780625 | 0.084375788 | 0.132665501 | 0.099632264 | 0.612518203 | 5.7981E-146 | 0.657614 | 5.8108E-22 | 0.530807 | 0.275697 | 0.674586 | 0.511815 |
| 15-63277703-T-C | 15 | rs117618017 | T | 2.15E-25 | 0.44546645 | | 0.469876279 | 0.956002218 | 0.031854442 | 0.916435069 | 3.15141E-44 | 0.35752 | 0.049697 | 0.063114 | 0.67928 | 0.894279 | |
| 15-64131307-G-A | 15 | rs3848143 | G | 8.41E-11 | 0.45947195 | | 0.006466901 | 0.040105137 | 0.750104896 | 0.627991839 | 6.97607E-82 | 0.0997139 | 0.0283712 | 0.462664 | 0.157106 | 0.536111 | 5.18E-12 |
| 15-78936857-A-G | 15 | rs12592898 | A | 4.18E-09 | 0.58015628 | 0.463562184 | 0.008879267 | 0.306282371 | 0.944127396 | 0.313993024 | 1.28664E-27 | 0.292628 | 0.358797 | 0.205995 | 0.779431 | 0.296816 | 0.511186 |
| 16-30010081-T-C | 16 | rs1140239 | T | 2.59E-13 | 0.43294228 | 0.084522046 | 0.147769343 | 0.852887207 | 0.479318289 | 0.415518151 | 4.97634E-93 | 0.731627 | 0.196752 | 0.857137 | 0.161046 | 0.248984 | 0.531415 |
| 16-31111250-T-C | 16 | rs889555 | T | 1.96E-11 | 0.16245534 | | 0.019337228 | 0.19675646 | 0.966869588 | 0.146542636 | 8.67188E-35 | 0.411134 | 0.713513 | 0.482775 | 0.099368 | 0.355693 | 6.72E-17 |
| 16-70660097-A-C | 16 | rs4985556 | A | 5.98E-10 | 0.00022081 | 0.916693179 | 0.01112911 | 0.242274106 | 0.001005565 | 0.001978882 | 4.49302E-25 | 0.242808 | 0.167126 | 0.124557 | 0.731298 | 0.104979 | 0.954898 |
| 16-79574511-C-T | 16 | rs450674 | C | 3.16E-08 | 0.13765342 | 0.023742148 | 0.005319646 | 0.719992546 | 0.709600642 | 0.246127987 | 2.32549E-37 | 0.277306 | 0.00387945 | 0.086756 | 0.476188 | 0.704596 | 0.392027 |
| 16-81739398-G-A | 16 | rs12446759 | G | 1.22E-13 | 0.16443982 | 0.017408045 | 0.03929793 | 0.156526185 | 0.676436447 | 0.441336656 | 1.1965E-302 | 0.995578 | 0.694209 | 0.321572 | 0.830527 | 0.432625 | 0.552006 |
| 16-81908423-G-C | 16 | rs72824905 | G | 8.48E-12 | 0.96203816 | 0.003192934 | 0.058936561 | 0.245634859 | 0.71237062 | 0.906041937 | 0.0277612 | 0.5452 | 0.110994 | 0.500265 | 0.300381 | 0.514666 | 0.502822 |
| 16-86420604-A-T | 16 | rs16941239 | A | 1.29E-08 | 0.36778075 | | 0.007113283 | 0.503645549 | 0.367086996 | 0.129739434 | 4.0327E-189 | 0.749869 | 0.501556 | 0.587796 | 0.594715 | 0.611259 | 0.759579 |
| 16-90103687-A-G | 16 | rs56407236 | A | 6.47E-15 | 0.00038968 | 0.029150768 | 0.059190037 | 0.469755637 | 0.344010739 | 0.001000322 | 0.926647 | 0.703459 | 0.358911 | 0.27585 | 0.903824 | 0.811545 | 0.499419 |
| 17-1728046-T-TGAG | 17 | rs35048651 | T | 7.67E-11 | 0.31813467 | 0.511802025 | 0.025835819 | 0.6898267 | 0.218866856 | 0.731880203 | 2.20419E-22 | 0.14073 | 0.0481606 | 0.039169 | 0.200075 | 0.435579 | 4.32E-05 |
| 17-18156140-A-G | 17 | rs2242595 | A | 1.11E-09 | 0.49588047 | 0.052313045 | 0.004786883 | 0.45413921 | 0.116043548 | 0.373418646 | 1.2673E-200 | 0.774033 | 2.4385E-31 | 0.402631 | 0.49756 | 0.91858 | 0.325089 |
| 17-44352876-T-C | 17 | rs5848 | T | 2.38E-20 | 0.80071064 | 0.016427822 | 0.170530329 | 0.329104636 | 0.136248585 | 0.731561767 | 2.2234E-302 | 0.074929 | 7.4026E-05 | 0.307796 | 0.014808 | 0.204475 | 0.79374 |
| 17-46779275-G-C | 17 | rs199515 | G | 9.34E-13 | 0.28411489 | 0.728076477 | 0.045201891 | 0.846658748 | 0.276719013 | 0.276462353 | 7.61659E-26 | 0.0119098 | 7.2164E-08 | 0.622384 | 0.781572 | 0.244193 | 0.617287 |
| 17-49219935-T-C | 17 | rs616338 | T | 2.82E-14 | 0.38544641 | 0.023378142 | 0.163948364 | 0.233777656 | 0.22507072 | 0.432962218 | 0.152151 | 0.554386 | 0.754342 | 0.5 | 0.495308 | 0.501615 | 0.502822 |
| 17-5233752-A-G | 17 | rs7225151 | A | 4.13E-13 | 0.08685004 | 0.02012241 | 0.042044458 | 0.899375805 | 0.130088229 | 0.074765286 | 6.45866E-34 | 0.862896 | 0.00168893 | 0.279116 | 0.773042 | 0.053972 | 0.64453 |
| 17-58332680-G-A | 17 | rs2526377 | G | 1.58E-12 | 0.51387205 | 0.001262205 | 0.043604508 | 0.280315427 | 0.346570825 | 0.662051845 | 2.97745E-09 | 0.178538 | 0.418005 | 0.660835 | 0.443027 | 0.523064 | 0.60718 |
| 17-63471557-C-T | 17 | rs4277405 | C | 8.80E-20 | 0.08510596 | 1.13865E-05 | 0.150242036 | 0.345506323 | | 0.097476516 | 0.0380635 | 0.85906 | 0.0113129 | 0.252789 | 0.80173 | 0.485548 | 0.536818 |
| 19-1050875-A-G | 19 | rs12151021 | A | 1.59E-37 | 0.02519881 | 9.2098E-05 | 0.875795073 | 0.221128643 | 0.812969214 | 0.024505863 | 6.92887E-29 | 0.471939 | 1.4496E-07 | 0.549835 | 0.526498 | 0.114907 | 0.580236 |
| 19-1854254-G-GC | 19 | rs149080927 | G | 5.09E-10 | 0.70265801 | | 0.030474012 | 0.12610404 | 0.240509923 | 0.51530695 | 3.8532E-291 | 0.701803 | 0.008453 | 0.656897 | 0.456757 | 0.714904 | 0.292875 |
| 19-49950060-T-C | 19 | rs9304690 | T | 4.74E-09 | 0.84415112 | 0.046014001 | 0.008331578 | 0.53589413 | 0.085786708 | 0.755607048 | 1.93914E-46 | 0.00061917 | 0.00109587 | 0.594217 | 0.97901 | 0.282715 | 0.914741 |
| 19-54267597-C-T | 19 | rs587709 | C | 3.63E-11 | 0.74258642 | 0.000219094 | 0.017985803 | 0.622222913 | 0.887688513 | 0.598917496 | 2.58817E-86 | 0.263856 | 0.0222994 | 2.81E-08 | 0.041419 | 0.61018 | 0.445888 |
| 2-105749599-C-T | 2 | rs143080277 | C | 2.07E-13 | 0.55608861 | 0.000740594 | 0.077259424 | 0.435120956 | 0.128912788 | 0.63523585 | 0.888765 | 0.526699 | 0.0572503 | 0.5 | 0.398338 | 0.5 | 0.501648 |
| 2-127135234-T-C | 2 | rs6733839 | T | 6.06E-118 | 0.03356217 | 4.39744E-12 | 0.999999996 | 0.323925397 | 0.356581658 | 0.17663469 | 0.0338456 | 0.442363 | 0.0244571 | 0.265068 | 0.492426 | 0.64388 | 0.898341 |
| 2-202878716-T-TC | 2 | rs139643391 | T | 1.08E-08 | 0.32273718 | | 0.008506649 | 0.16151468 | 0.261618831 | 0.472631867 | 1.71088E-20 | 0.186951 | 0.846981 | 0.563818 | 0.97493 | 0.175192 | 0.561647 |
| 2-233117202-G-C | 2 | rs10933431 | G | 3.62E-18 | 0.69846235 | | 0.173567909 | 0.187292333 | 0.418398385 | 0.304995899 | 5.2185E-292 | 0.419056 | 0.160341 | 0.222549 | 0.845931 | 0.85094 | 0.892498 |
| 2-37304796-C-T | 2 | rs17020490 | C | 3.29E-09 | 0.99475894 | | 0.007268015 | 0.083508369 | 0.913687956 | 0.798265022 | 2.0553E-291 | 0.0677137 | 1.0015E-50 | 0.861317 | 0.915652 | 0.830351 | 0.333025 |
| 2-9558882-G-A | 2 | rs72777026 | G | 2.72E-08 | 0.19821568 | 0.140604752 | 0.007095282 | 0.024810756 | 0.548759435 | 0.874082449 | 1.0047E-135 | 0.206855 | 0.0009299 | 0.044566 | 0.781113 | 0.323897 | 0.77076 |
| 20-413334-A-G | 20 | rs1358782 | A | 1.55E-08 | 0.97497819 | 0.200265436 | 0.013579497 | 0.327360292 | 0.195393 | 0.612666311 | 3.83631E-12 | 0.535677 | 0.00710236 | 0.90664 | 0.126868 | 0.301055 | 0.556372 |
| 20-56423488-G-A | 20 | rs6014724 | G | 4.13E-21 | 0.55056804 | 0.022688198 | 0.287140756 | 0.334038096 | 0.565348182 | 0.442442665 | 3.69209E-52 | 0.136037 | 0.151296 | 0.738273 | 2.94E-06 | 0.043386 | 0.834199 |
| 20-63743088-T-C | 20 | rs6742 | T | 2.58E-09 | 0.90050598 | 0.011631347 | 0.009960051 | 0.169466555 | 0.988289861 | 0.799505766 | 3.6616E-302 | 1.349E-302 | 5.577E-145 | 0.075222 | 6.3E-292 | 0.00677 | 2.14E-17 |
| 21-26101558-C-T | 21 | rs2154481 | C | 1.00E-12 | 0.27714114 | 0.073787025 | 0.045363218 | 0.156623888 | 0.79086253 | 0.466560535 | 1.0637E-291 | 0.00537067 | 1.4061E-07 | 0.545788 | 0.249407 | 0.973609 | 0.34554 |
| 21-26775872-T-C | 21 | rs2830489 | T | 1.69E-10 | 0.97886414 | 0.049917175 | 0.019319972 | 0.956212463 | 0.923403891 | 0.58809434 | 1.7032E-230 | 0.0084897 | 1.0761E-07 | 0.672121 | 0.928181 | 0.198118 | 0.65359 |
| 3-155069722-A-G | 3 | rs16824536 | A | 3.63E-08 | 0.39274259 | | 0.003673749 | 0.311941388 | 0.179219805 | 0.83408786 | 1.0525E-302 | 0.951376 | 0.00070169 | 0.885349 | 0.170081 | 0.526413 | 0.862386 |
| 3-155084189-G-A | 3 | rs61762319 | G | 2.16E-11 | 0.54281895 | 0.017935356 | 0.024673451 | 0.520724486 | 0.599220965 | 0.444319118 | 0.00020715 | 0.48054 | 0.629767 | 0.500926 | 0.069565 | 0.554 | 0.207595 |
| 4-11023507-C-T | 4 | rs6846529 | C | 2.20E-17 | 0.76229488 | 0.004725198 | 0.163456411 | 0.715069081 | 0.404461325 | 0.596108068 | 5.02158E-16 | 0.933426 | 0.861344 | 0.564031 | 0.020901 | 0.504322 | 0.428839 |
| 4-40197226-G-C | 4 | rs2245466 | G | 1.22E-09 | 0.0256059 | 0.349259317 | 0.020089424 | 0.829773002 | 0.312570509 | 0.053156943 | 4.52779E-20 | 0.285173 | 0.987752 | 0.003321 | 0.000219 | 0.444679 | 0.884497 |
| 4-993555-G-T | 4 | rs3822030 | G | 8.29E-12 | 0.06342785 | 0.059220218 | 0.042197049 | 0.088478958 | 0.132142038 | 0.289765049 | 1.231E-302 | 0.00095129 | 0.0137681 | 0.015194 | 0.32968 | 0.038239 | 3.78E-05 |
| 5-14724304-A-T | 5 | rs112403360 | A | 2.27E-09 | 0.34081445 | 0.632238589 | 0.015099748 | 0.892583594 | 0.918821396 | 0.344814881 | 0.0708148 | 0.990867 | 0.452773 | 0.899465 | 0.103138 | 0.855959 | 0.316539 |

| | | | | | | | | | | | | | | | | | |
|---|---|---|---|---|---|---|---|---|---|---|---|---|---|---|---|---|---|
| 5-151052827-T-C | 5 | rs871269 | T | 8.67E-09 | 0.08028788 | 0.063686884 | 0.004119426 | 0.485863193 | 0.194835011 | 0.098980581 | 6.75938E-70 | 0.113911 | 3.0642E-29 | 0.001782 | 0.18516 | 0.295459 | 0.95647 |
| 5-180201150-A-G | 5 | rs113706587 | A | 2.22E-16 | 0.04034967 | 0.024376424 | 0.068203533 | 0.534159553 | 0.021104254 | 0.131997285 | 1.75133E-23 | 0.0189679 | 0.0121103 | 0.044376 | 0.460206 | 0.505847 | 0.032508 |
| 5-86927378-C-T | 5 | rs62374257 | C | 1.38E-15 | 0.06441693 | 0.20735762 | 0.101016589 | 0.937408096 | 0.786735958 | 0.075363316 | 5.8465E-116 | 0.280104 | 0.00108215 | 0.791698 | 0.334411 | 0.945846 | 0.078452 |
| 6-114291731-T-C | 6 | rs785129 | T | 2.40E-09 | 0.17231497 | 0.000451305 | 0.003410309 | 0.321805898 | 0.369929529 | 0.371047919 | 1.40154E-55 | 0.530275 | 6.3281E-05 | 0.6479 | 0.066253 | 0.609037 | 0.030562 |
| 6-32615322-G-A | 6 | rs6605556 | G | 7.07E-20 | 0.05889657 | | 0.359258919 | 0.341002058 | 0.452276476 | 0.195939953 | 6.53607E-29 | 0.231042 | 0.00068576 | 0.061834 | 0.031291 | 0.952794 | 0.484137 |
| 6-41036354-A-G | 6 | rs10947943 | A | 1.13E-09 | 0.34470851 | 0.169044872 | 0.011255816 | 0.279958076 | 0.481007668 | 0.395795635 | 1.66626E-66 | 0.00262709 | 2.0272E-06 | 0.651138 | 0.836278 | 0.705616 | 0.332746 |
| 6-41161469-T-C | 6 | rs143332484 | T | 2.78E-25 | 0.8050499 | 1.95492E-06 | 0.568742109 | 0.175976226 | 0.900854184 | 0.903875053 | 0.452658 | 0.579125 | 0.200144 | 0.500441 | 0.629665 | 0.5 | 0.516271 |
| 6-41161514-T-C | 6 | rs75932628 | T | 2.53E-37 | 0.27136261 | 2.81138E-06 | 0.951265571 | 0.353676334 | 0.519249396 | 0.16424078 | 1.71498E-05 | 0.506747 | 0.00206021 | 0.5 | 0.003165 | 0.500269 | 0.509356 |
| 6-41181270-G-A | 6 | rs60755019 | G | 2.07E-08 | 0.06174048 | | 0.098223049 | 0.20632602 | 0.543000213 | 0.319846056 | 2.6841E-302 | 0.26474 | 0.0109655 | 0.65544 | 0.186373 | 0.387074 | 0.667173 |
| 6-47517390-T-C | 6 | rs7767350 | T | 7.94E-22 | 0.01694806 | 0.026047137 | 0.333020086 | 0.100242526 | 0.403289579 | 0.054006993 | 2.44384E-08 | 0.859851 | 0.0618233 | 0.549413 | 0.160611 | 0.283622 | 0.175392 |
| 7-100334426-C-T | 7 | rs7384878 | C | 1.06E-26 | 0.78482892 | 5.70493E-07 | 0.582501403 | 0.449620392 | 0.607266047 | 0.505175174 | 6.9725E-292 | 0.430723 | 9.8462E-38 | 0.000226 | 0.205278 | 0.498024 | 0.645269 |
| 7-12229967-A-C | 7 | rs13237518 | A | 4.88E-11 | 0.35602705 | 0.054555683 | 0.006156072 | 0.644127736 | 0.806710854 | 0.654211238 | 1.8196E-229 | 0.226954 | 0.012593 | 0.209134 | 0.094945 | 0.200993 | 0.097699 |
| 7-143413669-A-G | 7 | rs11771145 | A | 3.30E-14 | 0.46867847 | 0.716232461 | 0.030986813 | 0.374052016 | 0.780892047 | 0.212686047 | 1.37275E-74 | 0.278584 | 0.496877 | 0.091657 | 0.155439 | 0.297801 | 0.001812 |
| 7-28129126-G-GTCT[illegible] | 7 | rs1160871 | G | 9.83E-09 | 0.9786891 | | 0.010135567 | 0.862751256 | 0.293061656 | 0.65862013 | 1.8918E-302 | 0.0252518 | 1.5897E-13 | 0.11357 | 0.067448 | 0.016954 | 2.89E-07 |
| 7-37844191-T-C | 7 | rs6966331 | T | 4.64E-10 | 0.45052511 | 0.048891122 | 0.004731169 | 0.609605673 | 0.70592246 | 0.598674831 | 4.1578E-291 | 0.687977 | 0.00001285 | 0.216786 | 0.346613 | 0.972606 | 0.015536 |
| 7-54873635-T-C | 7 | rs76928645 | T | 1.62E-10 | 0.15188049 | | 0.013559063 | 0.034262575 | 0.205171484 | 0.095984255 | 2.64318E-25 | 0.56826 | 0.0509788 | 0.524216 | 0.646169 | 0.164483 | 0.87361 |
| 7-7817263-T-C | 7 | rs6943429 | T | 1.03E-10 | 0.01558655 | 0.478861586 | 0.027643758 | 0.523625756 | 0.031270153 | 0.075292202 | 1.54241E-08 | 0.1667 | 0.00455398 | 0.546731 | 0.086554 | 0.717564 | 0.521924 |
| 7-8204382-T-C | 7 | rs10952097 | T | 6.81E-09 | 0.82555137 | | 0.010713181 | 0.954214474 | 0.133111017 | 0.765809942 | 2.1522E-302 | 0.00026671 | 3.3155E-22 | 0.550029 | 0.71838 | 0.12437 | 0.796966 |
| 8-11844613-C-G | 8 | rs1065712 | C | 1.94E-09 | 0.24292712 | 0.086866035 | 0.004424393 | 0.870737603 | 0.049214151 | 0.288892322 | 1.10952E-17 | 0.891016 | 0.859437 | 0.502906 | 0.067528 | 0.831447 | 0.370378 |
| 8-144103704-A-G | 8 | rs34173062 | A | 1.72E-16 | 0.30455519 | 0.01153108 | 0.292352205 | 0.727114826 | 0.242645365 | 0.320483002 | 5.17891E-23 | 0.612888 | 0.291752 | 0.522819 | 0.244374 | 0.218864 | 0.713563 |
| 8-27362470-T-C | 8 | rs73223431 | T | 4.03E-22 | 0.18234044 | 0.000227536 | 0.254999039 | 0.773720315 | 0.002313503 | 0.65792744 | 4.82209E-24 | 0.408334 | 3.247E-08 | 0.420799 | 0.492072 | 0.551494 | 0.031589 |
| 8-27607795-T-C | 8 | rs11787077 | T | 1.70E-44 | 0.05830019 | 1.10632E-05 | 0.904203769 | 0.412493396 | 0.234186038 | 0.207869609 | 9.92921E-35 | 0.0896665 | 0.037124 | 0.716498 | 0.516999 | 0.471706 | 0.020834 |
| 9-104903697-G-C | 9 | rs1800978 | G | 1.59E-09 | 0.64045998 | 0.463131158 | 0.005006543 | 0.693149637 | 0.969031735 | 0.619599416 | 1.78995E-56 | 0.286465 | 0.0383721 | 0.094815 | 0.768487 | 0.001709 | 0.057243 |
